## Supplemental table S13. Associations between metabolome intensities and clinical outcomes. for "A YOUNG CHILD FORMULA SUPPLEMENTED WITH L. REUTERI AND GALACTO-OLIGOSACCHARIDES MODULATES THE COMPOSITION AND FUNCTION OF THE GUT MICROBIOME SUPPORTING BONE AND MUSCLE DEVELOPMENT IN TODDLERS"

| Category | Test type | Item | Name | Feature | Effect size | Standard e | Bias mode |
|---|---|---|---|---|---|---|---|
| SCFA_panel | abundance | 2-Methylpr | 2-Methylpr | Tibia_length | 0,05085 | 0,054087 | NA |
| SCFA_panel | abundance | 2-Methylpr | 2-Methylpr | Radius_length | -0,03881 | 0,072536 | NA |
| SCFA_panel | abundance | 2-Methylpr | 2-Methylpr | Radius_SOS | 0,024803 | 0,05179 | NA |
| SCFA_panel | abundance | 2-Methylpr | 2-Methylpr | Tibia_SOS | -0,03831 | 0,054479 | NA |
| SCFA_panel | abundance | 2-Methylpr | 2-Methylpr | Handgrip | -0,12843 | 0,062131 | NA |
| SCFA_panel | abundance | 3-Methylbu | 3-Methylbu | Tibia_length | 0,032589 | 0,054461 | NA |
| SCFA_panel | abundance | 3-Methylbu | 3-Methylbu | Radius_length | -0,03333 | 0,071189 | NA |
| SCFA_panel | abundance | 3-Methylbu | 3-Methylbu | Radius_SOS | 0,041428 | 0,052083 | NA |
| SCFA_panel | abundance | 3-Methylbu | 3-Methylbu | Tibia_SOS | -0,03173 | 0,055242 | NA |
| SCFA_panel | abundance | 3-Methylbu | 3-Methylbu | Handgrip | -0,16736 | 0,062271 | NA |
| SCFA_panel | abundance | Acetic acid | Acetic acid | Tibia_length | -0,01186 | 0,055419 | NA |
| SCFA_panel | abundance | Acetic acid | Acetic acid | Radius_length | 4,79E-04 | 0,073843 | NA |
| SCFA_panel | abundance | Acetic acid | Acetic acid | Radius_SOS | 0,030678 | 0,052606 | NA |
| SCFA_panel | abundance | Acetic acid | Acetic acid | Tibia_SOS | 0,020414 | 0,056205 | NA |
| SCFA_panel | abundance | Acetic acid | Acetic acid | Handgrip | 0,030145 | 0,062326 | NA |
| SCFA_panel | abundance | Butanoic a | Butanoic a | Tibia_length | -0,06657 | 0,052982 | NA |
| SCFA_panel | abundance | Butanoic a | Butanoic a | Radius_length | -0,0443 | 0,071763 | NA |
| SCFA_panel | abundance | Butanoic a | Butanoic a | Radius_SOS | 0,025865 | 0,050837 | NA |
| SCFA_panel | abundance | Butanoic a | Butanoic a | Tibia_SOS | 0,012665 | 0,054702 | NA |
| SCFA_panel | abundance | Butanoic a | Butanoic a | Handgrip | -0,00944 | 0,061712 | NA |
| SCFA_panel | abundance | Hexanoic a | Hexanoic a | Tibia_length | -0,01783 | 0,067127 | NA |
| SCFA_panel | abundance | Hexanoic a | Hexanoic a | Radius_length | -0,00725 | 0,083039 | NA |
| SCFA_panel | abundance | Hexanoic a | Hexanoic a | Radius_SOS | 0,085926 | 0,06696 | NA |
| SCFA_panel | abundance | Hexanoic a | Hexanoic a | Tibia_SOS | -0,07639 | 0,069285 | NA |
| SCFA_panel | abundance | Hexanoic a | Hexanoic a | Handgrip | 0,058547 | 0,081307 | NA |
| WGCNA_module | abundance | M1 | M1 | Tibia_length | -0,06523 | 0,053249 | NA |
| WGCNA_module | abundance | M1 | M1 | Radius_length | -0,02623 | 0,073013 | NA |
| WGCNA_module | abundance | M1 | M1 | Radius_SOS | -0,05494 | 0,051057 | NA |
| WGCNA_module | abundance | M1 | M1 | Tibia_SOS | 3,38E-04 | 0,054601 | NA |
| WGCNA_module | abundance | M1 | M1 | Handgrip | 0,077771 | 0,062105 | NA |
| WGCNA_module | abundance | M10 | M10 | Tibia_length | -0,01209 | 0,051803 | NA |
| WGCNA_module | abundance | M10 | M10 | Radius_length | -0,0324 | 0,071423 | NA |
| WGCNA_module | abundance | M10 | M10 | Radius_SOS | -0,03865 | 0,049929 | NA |
| WGCNA_module | abundance | M10 | M10 | Tibia_SOS | 0,116624 | 0,052847 | NA |
| WGCNA_module | abundance | M10 | M10 | Handgrip | 0,114057 | 0,061076 | NA |
| WGCNA_module | abundance | M11 | M11 | Tibia_length | -0,09965 | 0,050683 | NA |
| WGCNA_module | abundance | M11 | M11 | Radius_length | -0,11481 | 0,069207 | NA |
| WGCNA_module | abundance | M11 | M11 | Radius_SOS | 0,039247 | 0,049322 | NA |
| WGCNA_module | abundance | M11 | M11 | Tibia_SOS | -0,14646 | 0,052131 | NA |
| WGCNA_module | abundance | M11 | M11 | Handgrip | -0,18272 | 0,059742 | NA |
| WGCNA_module | abundance | M12 | M12 | Tibia_length | 0,117163 | 0,049513 | NA |
| WGCNA_module | abundance | M12 | M12 | Radius_length | 0,187866 | 0,066897 | NA |
| WGCNA_module | abundance | M12 | M12 | Radius_SOS | 0,068866 | 0,048928 | NA |
| WGCNA_module | abundance | M12 | M12 | Tibia_SOS | 0,089343 | 0,052315 | NA |
| WGCNA_module | abundance | M12 | M12 | Handgrip | 0,086003 | 0,060054 | NA |
| WGCNA_module | abundance | M13 | M13 | Tibia_length | 0,177682 | 0,051891 | NA |
| WGCNA_module | abundance | M13 | M13 | Radius_length | 0,075706 | 0,07194 | NA |

| | | | | | | | |
|---|---|---|---|---|---|---|---|
| WGCNA_module | abundance | M13 | M13 | Radius_SOS | 0,007855 | 0,050487 | NA |
| WGCNA_module | abundance | M13 | M13 | Tibia_SOS | 0,138438 | 0,054394 | NA |
| WGCNA_module | abundance | M13 | M13 | Handgrip | 0,06133 | 0,060857 | NA |
| WGCNA_module | abundance | M14 | M14 | Tibia_length | 0,025074 | 0,051874 | NA |
| WGCNA_module | abundance | M14 | M14 | Radius_length | 0,017131 | 0,07124 | NA |
| WGCNA_module | abundance | M14 | M14 | Radius_SOS | 0,046625 | 0,049915 | NA |
| WGCNA_module | abundance | M14 | M14 | Tibia_SOS | 0,010138 | 0,053591 | NA |
| WGCNA_module | abundance | M14 | M14 | Handgrip | 0,001402 | 0,060907 | NA |
| WGCNA_module | abundance | M15 | M15 | Tibia_length | -0,09048 | 0,053021 | NA |
| WGCNA_module | abundance | M15 | M15 | Radius_length | 0,018358 | 0,072291 | NA |
| WGCNA_module | abundance | M15 | M15 | Radius_SOS | 0,01215 | 0,050744 | NA |
| WGCNA_module | abundance | M15 | M15 | Tibia_SOS | -0,02125 | 0,054363 | NA |
| WGCNA_module | abundance | M15 | M15 | Handgrip | 0,035758 | 0,060642 | NA |
| WGCNA_module | abundance | M16 | M16 | Tibia_length | 0,108921 | 0,053158 | NA |
| WGCNA_module | abundance | M16 | M16 | Radius_length | 0,168578 | 0,07193 | NA |
| WGCNA_module | abundance | M16 | M16 | Radius_SOS | 0,091187 | 0,050477 | NA |
| WGCNA_module | abundance | M16 | M16 | Tibia_SOS | 0,078484 | 0,054704 | NA |
| WGCNA_module | abundance | M16 | M16 | Handgrip | 0,181445 | 0,059805 | NA |
| WGCNA_module | abundance | M17 | M17 | Tibia_length | -0,05981 | 0,056156 | NA |
| WGCNA_module | abundance | M17 | M17 | Radius_length | 0,012789 | 0,074785 | NA |
| WGCNA_module | abundance | M17 | M17 | Radius_SOS | -0,01785 | 0,053496 | NA |
| WGCNA_module | abundance | M17 | M17 | Tibia_SOS | 0,003708 | 0,058081 | NA |
| WGCNA_module | abundance | M17 | M17 | Handgrip | -2,03E-04 | 0,062841 | NA |
| WGCNA_module | abundance | M18 | M18 | Tibia_length | -0,11323 | 0,052208 | NA |
| WGCNA_module | abundance | M18 | M18 | Radius_length | -0,19213 | 0,070118 | NA |
| WGCNA_module | abundance | M18 | M18 | Radius_SOS | -0,07564 | 0,050161 | NA |
| WGCNA_module | abundance | M18 | M18 | Tibia_SOS | -0,09374 | 0,053715 | NA |
| WGCNA_module | abundance | M18 | M18 | Handgrip | -0,13414 | 0,060931 | NA |
| WGCNA_module | abundance | M19 | M19 | Tibia_length | 0,013027 | 0,049351 | NA |
| WGCNA_module | abundance | M19 | M19 | Radius_length | 0,057482 | 0,066642 | NA |
| WGCNA_module | abundance | M19 | M19 | Radius_SOS | 0,039858 | 0,049005 | NA |
| WGCNA_module | abundance | M19 | M19 | Tibia_SOS | 0,065489 | 0,052374 | NA |
| WGCNA_module | abundance | M19 | M19 | Handgrip | -0,00769 | 0,060356 | NA |
| WGCNA_module | abundance | M2 | M2 | Tibia_length | 0,123654 | 0,049128 | NA |
| WGCNA_module | abundance | M2 | M2 | Radius_length | 0,192817 | 0,065281 | NA |
| WGCNA_module | abundance | M2 | M2 | Radius_SOS | 0,022016 | 0,049033 | NA |
| WGCNA_module | abundance | M2 | M2 | Tibia_SOS | 0,152998 | 0,051884 | NA |
| WGCNA_module | abundance | M2 | M2 | Handgrip | 0,097167 | 0,059909 | NA |
| WGCNA_module | abundance | M20 | M20 | Tibia_length | -0,02035 | 0,049051 | NA |
| WGCNA_module | abundance | M20 | M20 | Radius_length | 0,108929 | 0,06452 | NA |
| WGCNA_module | abundance | M20 | M20 | Radius_SOS | -0,01291 | 0,049242 | NA |
| WGCNA_module | abundance | M20 | M20 | Tibia_SOS | 0,047559 | 0,052707 | NA |
| WGCNA_module | abundance | M20 | M20 | Handgrip | 0,019809 | 0,060182 | NA |
| WGCNA_module | abundance | M21 | M21 | Tibia_length | -0,0552 | 0,055567 | NA |
| WGCNA_module | abundance | M21 | M21 | Radius_length | -0,12045 | 0,073357 | NA |
| WGCNA_module | abundance | M21 | M21 | Radius_SOS | -0,13912 | 0,052741 | NA |
| WGCNA_module | abundance | M21 | M21 | Tibia_SOS | -0,0746 | 0,057661 | NA |
| WGCNA_module | abundance | M21 | M21 | Handgrip | -0,09737 | 0,062208 | NA |

| | | | | | | | |
|---|---|---|---|---|---|---|---|
| WGCNA_module | abundance | M22 | M22 | Tibia_length | -0,03379 | 0,056103 | NA |
| WGCNA_module | abundance | M22 | M22 | Radius_length | 0,096061 | 0,074195 | NA |
| WGCNA_module | abundance | M22 | M22 | Radius_SOS | -0,02776 | 0,053351 | NA |
| WGCNA_module | abundance | M22 | M22 | Tibia_SOS | 0,089982 | 0,056122 | NA |
| WGCNA_module | abundance | M22 | M22 | Handgrip | 0,095737 | 0,062659 | NA |
| WGCNA_module | abundance | M23 | M23 | Tibia_length | 0,08184 | 0,049445 | NA |
| WGCNA_module | abundance | M23 | M23 | Radius_length | 0,096648 | 0,066935 | NA |
| WGCNA_module | abundance | M23 | M23 | Radius_SOS | 0,007385 | 0,049028 | NA |
| WGCNA_module | abundance | M23 | M23 | Tibia_SOS | 0,066105 | 0,052372 | NA |
| WGCNA_module | abundance | M23 | M23 | Handgrip | -0,01463 | 0,060268 | NA |
| WGCNA_module | abundance | M24 | M24 | Tibia_length | -0,08644 | 0,050717 | NA |
| WGCNA_module | abundance | M24 | M24 | Radius_length | -0,10364 | 0,069253 | NA |
| WGCNA_module | abundance | M24 | M24 | Radius_SOS | -0,08589 | 0,049287 | NA |
| WGCNA_module | abundance | M24 | M24 | Tibia_SOS | -0,04169 | 0,052693 | NA |
| WGCNA_module | abundance | M24 | M24 | Handgrip | 0,004183 | 0,060837 | NA |
| WGCNA_module | abundance | M25 | M25 | Tibia_length | 0,057666 | 0,054385 | NA |
| WGCNA_module | abundance | M25 | M25 | Radius_length | 0,187379 | 0,071681 | NA |
| WGCNA_module | abundance | M25 | M25 | Radius_SOS | -0,01438 | 0,051503 | NA |
| WGCNA_module | abundance | M25 | M25 | Tibia_SOS | 0,057663 | 0,05478 | NA |
| WGCNA_module | abundance | M25 | M25 | Handgrip | 0,025347 | 0,062768 | NA |
| WGCNA_module | abundance | M26 | M26 | Tibia_length | -0,08126 | 0,050013 | NA |
| WGCNA_module | abundance | M26 | M26 | Radius_length | -0,13346 | 0,067707 | NA |
| WGCNA_module | abundance | M26 | M26 | Radius_SOS | -0,04655 | 0,04904 | NA |
| WGCNA_module | abundance | M26 | M26 | Tibia_SOS | -0,09959 | 0,052283 | NA |
| WGCNA_module | abundance | M26 | M26 | Handgrip | -0,08936 | 0,060036 | NA |
| WGCNA_module | abundance | M27 | M27 | Tibia_length | -0,04902 | 0,050562 | NA |
| WGCNA_module | abundance | M27 | M27 | Radius_length | -0,09568 | 0,069012 | NA |
| WGCNA_module | abundance | M27 | M27 | Radius_SOS | 0,074299 | 0,049113 | NA |
| WGCNA_module | abundance | M27 | M27 | Tibia_SOS | -0,04491 | 0,052807 | NA |
| WGCNA_module | abundance | M27 | M27 | Handgrip | -0,01129 | 0,060891 | NA |
| WGCNA_module | abundance | M28 | M28 | Tibia_length | -0,13017 | 0,048562 | NA |
| WGCNA_module | abundance | M28 | M28 | Radius_length | -0,17692 | 0,062616 | NA |
| WGCNA_module | abundance | M28 | M28 | Radius_SOS | -0,08595 | 0,049331 | NA |
| WGCNA_module | abundance | M28 | M28 | Tibia_SOS | -0,10031 | 0,052851 | NA |
| WGCNA_module | abundance | M28 | M28 | Handgrip | 0,001227 | 0,060355 | NA |
| WGCNA_module | abundance | M29 | M29 | Tibia_length | -0,01999 | 0,057476 | NA |
| WGCNA_module | abundance | M29 | M29 | Radius_length | -0,03508 | 0,074713 | NA |
| WGCNA_module | abundance | M29 | M29 | Radius_SOS | -0,02491 | 0,054644 | NA |
| WGCNA_module | abundance | M29 | M29 | Tibia_SOS | 0,02119 | 0,058341 | NA |
| WGCNA_module | abundance | M29 | M29 | Handgrip | -0,04709 | 0,060126 | NA |
| WGCNA_module | abundance | M3 | M3 | Tibia_length | -0,1258 | 0,051409 | NA |
| WGCNA_module | abundance | M3 | M3 | Radius_length | -0,14508 | 0,070048 | NA |
| WGCNA_module | abundance | M3 | M3 | Radius_SOS | -0,02579 | 0,049799 | NA |
| WGCNA_module | abundance | M3 | M3 | Tibia_SOS | -0,13206 | 0,052719 | NA |
| WGCNA_module | abundance | M3 | M3 | Handgrip | -0,12915 | 0,060535 | NA |
| WGCNA_module | abundance | M30 | M30 | Tibia_length | -0,07154 | 0,049495 | NA |
| WGCNA_module | abundance | M30 | M30 | Radius_length | -0,11064 | 0,066487 | NA |
| WGCNA_module | abundance | M30 | M30 | Radius_SOS | 0,001665 | 0,04903 | NA |

| | | | | | | | |
|---|---|---|---|---|---|---|---|
| WGCNA_module | abundance | M30 | M30 | Tibia_SOS | -0,04819 | 0,052438 | NA |
| WGCNA_module | abundance | M30 | M30 | Handgrip | -0,0433 | 0,060206 | NA |
| WGCNA_module | abundance | M31 | M31 | Tibia_length | -0,05555 | 0,052981 | NA |
| WGCNA_module | abundance | M31 | M31 | Radius_length | 0,020837 | 0,07219 | NA |
| WGCNA_module | abundance | M31 | M31 | Radius_SOS | 5,57E-04 | 0,050847 | NA |
| WGCNA_module | abundance | M31 | M31 | Tibia_SOS | -0,0035 | 0,053213 | NA |
| WGCNA_module | abundance | M31 | M31 | Handgrip | -0,07795 | 0,061121 | NA |
| WGCNA_module | abundance | M32 | M32 | Tibia_length | -0,11755 | 0,052994 | NA |
| WGCNA_module | abundance | M32 | M32 | Radius_length | -0,16893 | 0,071841 | NA |
| WGCNA_module | abundance | M32 | M32 | Radius_SOS | 0,007256 | 0,050777 | NA |
| WGCNA_module | abundance | M32 | M32 | Tibia_SOS | -0,12512 | 0,053447 | NA |
| WGCNA_module | abundance | M32 | M32 | Handgrip | -0,11435 | 0,061628 | NA |
| WGCNA_module | abundance | M33 | M33 | Tibia_length | -0,02887 | 0,049099 | NA |
| WGCNA_module | abundance | M33 | M33 | Radius_length | 8,58E-04 | 0,064972 | NA |
| WGCNA_module | abundance | M33 | M33 | Radius_SOS | 0,024525 | 0,049176 | NA |
| WGCNA_module | abundance | M33 | M33 | Tibia_SOS | -0,06878 | 0,052573 | NA |
| WGCNA_module | abundance | M33 | M33 | Handgrip | -0,00617 | 0,060238 | NA |
| WGCNA_module | abundance | M34 | M34 | Tibia_length | 8,93E-04 | 0,053089 | NA |
| WGCNA_module | abundance | M34 | M34 | Radius_length | -0,00203 | 0,072086 | NA |
| WGCNA_module | abundance | M34 | M34 | Radius_SOS | 0,057603 | 0,050758 | NA |
| WGCNA_module | abundance | M34 | M34 | Tibia_SOS | -0,00637 | 0,054489 | NA |
| WGCNA_module | abundance | M34 | M34 | Handgrip | 0,035054 | 0,061955 | NA |
| WGCNA_module | abundance | M4 | M4 | Tibia_length | 0,013163 | 0,048986 | NA |
| WGCNA_module | abundance | M4 | M4 | Radius_length | 0,054728 | 0,062205 | NA |
| WGCNA_module | abundance | M4 | M4 | Radius_SOS | 0,015467 | 0,049758 | NA |
| WGCNA_module | abundance | M4 | M4 | Tibia_SOS | -0,0355 | 0,052953 | NA |
| WGCNA_module | abundance | M4 | M4 | Handgrip | 0,012708 | 0,06025 | NA |
| WGCNA_module | abundance | M5 | M5 | Tibia_length | 0,026873 | 0,050822 | NA |
| WGCNA_module | abundance | M5 | M5 | Radius_length | 0,006341 | 0,069695 | NA |
| WGCNA_module | abundance | M5 | M5 | Radius_SOS | -0,02389 | 0,049369 | NA |
| WGCNA_module | abundance | M5 | M5 | Tibia_SOS | -0,022 | 0,053149 | NA |
| WGCNA_module | abundance | M5 | M5 | Handgrip | 0,041387 | 0,061182 | NA |
| WGCNA_module | abundance | M6 | M6 | Tibia_length | -0,13703 | 0,051797 | NA |
| WGCNA_module | abundance | M6 | M6 | Radius_length | -0,19212 | 0,069692 | NA |
| WGCNA_module | abundance | M6 | M6 | Radius_SOS | -0,05458 | 0,049818 | NA |
| WGCNA_module | abundance | M6 | M6 | Tibia_SOS | -0,11506 | 0,053189 | NA |
| WGCNA_module | abundance | M6 | M6 | Handgrip | -0,08852 | 0,06044 | NA |
| WGCNA_module | abundance | M7 | M7 | Tibia_length | 0,110263 | 0,052358 | NA |
| WGCNA_module | abundance | M7 | M7 | Radius_length | 0,094861 | 0,071715 | NA |
| WGCNA_module | abundance | M7 | M7 | Radius_SOS | 0,071773 | 0,050505 | NA |
| WGCNA_module | abundance | M7 | M7 | Tibia_SOS | 0,045363 | 0,054039 | NA |
| WGCNA_module | abundance | M7 | M7 | Handgrip | -0,05917 | 0,061901 | NA |
| WGCNA_module | abundance | M8 | M8 | Tibia_length | -0,16236 | 0,050775 | NA |
| WGCNA_module | abundance | M8 | M8 | Radius_length | -0,10796 | 0,070233 | NA |
| WGCNA_module | abundance | M8 | M8 | Radius_SOS | -0,02405 | 0,049574 | NA |
| WGCNA_module | abundance | M8 | M8 | Tibia_SOS | -0,07251 | 0,053315 | NA |
| WGCNA_module | abundance | M8 | M8 | Handgrip | 0,007986 | 0,060849 | NA |
| WGCNA_module | abundance | M9 | M9 | Tibia_length | -0,02759 | 0,05053 | NA |

| | | | | | | | |
|---|---|---|---|---|---|---|---|
| WGCNA_module | abundance | M9 | M9 | Radius_length | 0,055507 | 0,069388 | NA |
| WGCNA_module | abundance | M9 | M9 | Radius_SOS | -0,01354 | 0,049277 | NA |
| WGCNA_module | abundance | M9 | M9 | Tibia_SOS | 0,058617 | 0,052969 | NA |
| WGCNA_module | abundance | M9 | M9 | Handgrip | 0,044679 | 0,060614 | NA |
| SCFA_panel | abundance | Pentanoic | Pentanoic | Tibia_length | -0,01685 | 0,053447 | NA |
| SCFA_panel | abundance | Pentanoic | Pentanoic | Radius_length | -0,00914 | 0,070958 | NA |
| SCFA_panel | abundance | Pentanoic | Pentanoic | Radius_SOS | 0,026734 | 0,051158 | NA |
| SCFA_panel | abundance | Pentanoic | Pentanoic | Tibia_SOS | -0,07161 | 0,05445 | NA |
| SCFA_panel | abundance | Pentanoic | Pentanoic | Handgrip | -0,06883 | 0,062944 | NA |
| SCFA_panel | abundance | Propanoic | Propanoic | Tibia_length | 0,005412 | 0,051431 | NA |
| SCFA_panel | abundance | Propanoic | Propanoic | Radius_length | -0,05697 | 0,070241 | NA |
| SCFA_panel | abundance | Propanoic | Propanoic | Radius_SOS | -0,02462 | 0,049668 | NA |
| SCFA_panel | abundance | Propanoic | Propanoic | Tibia_SOS | 0,026991 | 0,053245 | NA |
| SCFA_panel | abundance | Propanoic | Propanoic | Handgrip | -0,02213 | 0,061287 | NA |
| 2a | abundance | SL00001 | Deoxycytid | Tibia_length | -0,11416 | 0,054475 | NA |
| 2a | abundance | SL00001 | Deoxycytid | Radius_length | -0,07379 | 0,073462 | NA |
| 2a | abundance | SL00001 | Deoxycytid | Radius_SOS | -0,05855 | 0,051853 | NA |
| 2a | abundance | SL00001 | Deoxycytid | Tibia_SOS | -0,04107 | 0,054463 | NA |
| 2a | abundance | SL00001 | Deoxycytid | Handgrip | -0,12974 | 0,061708 | NA |
| 2a | abundance | SL00009 | N-Acetylhis | Tibia_length | 0,011843 | 0,053986 | NA |
| 2a | abundance | SL00009 | N-Acetylhis | Radius_length | 0,045231 | 0,073018 | NA |
| 2a | abundance | SL00009 | N-Acetylhis | Radius_SOS | 0,003155 | 0,051457 | NA |
| 2a | abundance | SL00009 | N-Acetylhis | Tibia_SOS | 0,056383 | 0,055734 | NA |
| 2a | abundance | SL00009 | N-Acetylhis | Handgrip | 0,077633 | 0,061809 | NA |
| 2a | abundance | SL00017 | Aspartic ac | Tibia_length | 0,108429 | 0,049737 | NA |
| 2a | abundance | SL00017 | Aspartic ac | Radius_length | 0,213994 | 0,066411 | NA |
| 2a | abundance | SL00017 | Aspartic ac | Radius_SOS | 0,014617 | 0,049063 | NA |
| 2a | abundance | SL00017 | Aspartic ac | Tibia_SOS | 0,117673 | 0,052312 | NA |
| 2a | abundance | SL00017 | Aspartic ac | Handgrip | 0,076218 | 0,060239 | NA |
| 2a | abundance | SL00020 | Carnitine | Tibia_length | 0,026401 | 0,052256 | NA |
| 2a | abundance | SL00020 | Carnitine | Radius_length | -0,01274 | 0,071275 | NA |
| 2a | abundance | SL00020 | Carnitine | Radius_SOS | -0,0617 | 0,05018 | NA |
| 2a | abundance | SL00020 | Carnitine | Tibia_SOS | -0,06811 | 0,053621 | NA |
| 2a | abundance | SL00020 | Carnitine | Handgrip | 0,005252 | 0,061618 | NA |
| 2a | abundance | SL00024 | Creatine | Tibia_length | -0,06847 | 0,054177 | NA |
| 2a | abundance | SL00024 | Creatine | Radius_length | -0,08238 | 0,073133 | NA |
| 2a | abundance | SL00024 | Creatine | Radius_SOS | -0,07205 | 0,051641 | NA |
| 2a | abundance | SL00024 | Creatine | Tibia_SOS | -0,00522 | 0,055856 | NA |
| 2a | abundance | SL00024 | Creatine | Handgrip | 0,026729 | 0,061903 | NA |
| 2a | abundance | SL00035 | Glycylglyci | Tibia_length | 0,028419 | 0,054553 | NA |
| 2a | abundance | SL00035 | Glycylglyci | Radius_length | 0,045915 | 0,073529 | NA |
| 2a | abundance | SL00035 | Glycylglyci | Radius_SOS | -0,0103 | 0,051964 | NA |
| 2a | abundance | SL00035 | Glycylglyci | Tibia_SOS | 0,055319 | 0,055438 | NA |
| 2a | abundance | SL00035 | Glycylglyci | Handgrip | 0,031751 | 0,062386 | NA |
| 2a | abundance | SL00040 | Kynurenine | Tibia_length | -0,03947 | 0,050683 | NA |
| 2a | abundance | SL00040 | Kynurenine | Radius_length | -0,09895 | 0,069193 | NA |
| 2a | abundance | SL00040 | Kynurenine | Radius_SOS | -0,07539 | 0,049124 | NA |
| 2a | abundance | SL00040 | Kynurenine | Tibia_SOS | -0,07452 | 0,052682 | NA |

| | | | | | | | |
|---|---|---|---|---|---|---|---|
| 2a | abundance | SL00040 | Kynurenine | Handgrip | -0,09771 | 0,060212 | NA |
| 2a | abundance | SL00049 | N-Acetylar | Tibia_length | 0,068086 | 0,052637 | NA |
| 2a | abundance | SL00049 | N-Acetylar | Radius_length | 0,080969 | 0,0713 | NA |
| 2a | abundance | SL00049 | N-Acetylar | Radius_SOS | 0,001891 | 0,050101 | NA |
| 2a | abundance | SL00049 | N-Acetylar | Tibia_SOS | 0,064986 | 0,053687 | NA |
| 2a | abundance | SL00049 | N-Acetylar | Handgrip | 0,068815 | 0,060634 | NA |
| 2a | abundance | SL00052 | N-Isovalery | Tibia_length | 0,100728 | 0,055233 | NA |
| 2a | abundance | SL00052 | N-Isovalery | Radius_length | 0,20141 | 0,07248 | NA |
| 2a | abundance | SL00052 | N-Isovalery | Radius_SOS | 0,062503 | 0,052685 | NA |
| 2a | abundance | SL00052 | N-Isovalery | Tibia_SOS | 0,050761 | 0,056134 | NA |
| 2a | abundance | SL00052 | N-Isovalery | Handgrip | -0,01302 | 0,063589 | NA |
| 1 | abundance | SL00054 | Ornithine | Tibia_length | 0,10572 | 0,052238 | NA |
| 1 | abundance | SL00054 | Ornithine | Radius_length | 0,102708 | 0,071745 | NA |
| 1 | abundance | SL00054 | Ornithine | Radius_SOS | 0,104438 | 0,049804 | NA |
| 1 | abundance | SL00054 | Ornithine | Tibia_SOS | 0,097644 | 0,053702 | NA |
| 1 | abundance | SL00054 | Ornithine | Handgrip | -0,01654 | 0,06137 | NA |
| 1 | abundance | SL00061 | Proline | Tibia_length | 0,112599 | 0,051792 | NA |
| 1 | abundance | SL00061 | Proline | Radius_length | 0,070543 | 0,071545 | NA |
| 1 | abundance | SL00061 | Proline | Radius_SOS | -0,00561 | 0,050016 | NA |
| 1 | abundance | SL00061 | Proline | Tibia_SOS | 0,116942 | 0,053437 | NA |
| 1 | abundance | SL00061 | Proline | Handgrip | 0,089723 | 0,060268 | NA |
| 1 | abundance | SL00063 | Pyridoxal | Tibia_length | 0,005833 | 0,052027 | NA |
| 1 | abundance | SL00063 | Pyridoxal | Radius_length | 0,086878 | 0,07096 | NA |
| 1 | abundance | SL00063 | Pyridoxal | Radius_SOS | -0,03333 | 0,049977 | NA |
| 1 | abundance | SL00063 | Pyridoxal | Tibia_SOS | 0,087121 | 0,053268 | NA |
| 1 | abundance | SL00063 | Pyridoxal | Handgrip | -0,02557 | 0,061575 | NA |
| 2a | abundance | SL00074 | Valine/ 5-A | Tibia_length | 0,018432 | 0,053314 | NA |
| 2a | abundance | SL00074 | Valine/ 5-A | Radius_length | 0,049237 | 0,0726 | NA |
| 2a | abundance | SL00074 | Valine/ 5-A | Radius_SOS | -0,07147 | 0,050962 | NA |
| 2a | abundance | SL00074 | Valine/ 5-A | Tibia_SOS | 0,11463 | 0,053896 | NA |
| 2a | abundance | SL00074 | Valine/ 5-A | Handgrip | 0,098988 | 0,060852 | NA |
| 2a | abundance | SL00081 | 5-Methylcy | Tibia_length | -0,08437 | 0,051669 | NA |
| 2a | abundance | SL00081 | 5-Methylcy | Radius_length | 0,036004 | 0,071172 | NA |
| 2a | abundance | SL00081 | 5-Methylcy | Radius_SOS | -0,01615 | 0,04999 | NA |
| 2a | abundance | SL00081 | 5-Methylcy | Tibia_SOS | -0,00144 | 0,053266 | NA |
| 2a | abundance | SL00081 | 5-Methylcy | Handgrip | 0,032346 | 0,060885 | NA |
| 1 | abundance | SL00082 | Allopurinol | Tibia_length | 0,115899 | 0,056805 | NA |
| 1 | abundance | SL00082 | Allopurinol | Radius_length | 0,172989 | 0,075307 | NA |
| 1 | abundance | SL00082 | Allopurinol | Radius_SOS | 0,161732 | 0,053116 | NA |
| 1 | abundance | SL00082 | Allopurinol | Tibia_SOS | 0,085539 | 0,057739 | NA |
| 1 | abundance | SL00082 | Allopurinol | Handgrip | 0,140607 | 0,061968 | NA |
| 2a | abundance | SL00089 | Dissaccari | Tibia_length | 0,067076 | 0,051786 | NA |
| 2a | abundance | SL00089 | Dissaccari | Radius_length | 0,135754 | 0,070886 | NA |
| 2a | abundance | SL00089 | Dissaccari | Radius_SOS | -0,03375 | 0,04986 | NA |
| 2a | abundance | SL00089 | Dissaccari | Tibia_SOS | -0,01403 | 0,053306 | NA |
| 2a | abundance | SL00089 | Dissaccari | Handgrip | 0,004525 | 0,060976 | NA |
| 1 | abundance | SL00096 | Hexoses III | Tibia_length | -0,15627 | 0,054526 | NA |
| 1 | abundance | SL00096 | Hexoses III | Radius_length | -0,07404 | 0,074468 | NA |

| | | | | | | | |
|---|---|---|---|---|---|---|---|
| 1 | abundance | SL00096 | Hexoses III | Radius_SOS | -0,1168 | 0,052559 | NA |
| 1 | abundance | SL00096 | Hexoses III | Tibia_SOS | -0,07879 | 0,055852 | NA |
| 1 | abundance | SL00096 | Hexoses III | Handgrip | -0,16668 | 0,06151 | NA |
| 1 | abundance | SL00097 | Hexoses II | Tibia_length | -0,01102 | 0,055654 | NA |
| 1 | abundance | SL00097 | Hexoses II | Radius_length | 0,071268 | 0,074328 | NA |
| 1 | abundance | SL00097 | Hexoses II | Radius_SOS | -0,02189 | 0,052864 | NA |
| 1 | abundance | SL00097 | Hexoses II | Tibia_SOS | 0,020414 | 0,056451 | NA |
| 1 | abundance | SL00097 | Hexoses II | Handgrip | 0,094793 | 0,06235 | NA |
| 2a | abundance | SL00103 | Gulonic ac | Tibia_length | 0,04138 | 0,053562 | NA |
| 2a | abundance | SL00103 | Gulonic ac | Radius_length | 0,041352 | 0,072925 | NA |
| 2a | abundance | SL00103 | Gulonic ac | Radius_SOS | 0,015481 | 0,051175 | NA |
| 2a | abundance | SL00103 | Gulonic ac | Tibia_SOS | 0,045157 | 0,054338 | NA |
| 2a | abundance | SL00103 | Gulonic ac | Handgrip | 0,104073 | 0,061224 | NA |
| 1 | abundance | SL00104 | Hexoses I | Tibia_length | -0,02603 | 0,056099 | NA |
| 1 | abundance | SL00104 | Hexoses I | Radius_length | 0,015612 | 0,074824 | NA |
| 1 | abundance | SL00104 | Hexoses I | Radius_SOS | -0,04413 | 0,053339 | NA |
| 1 | abundance | SL00104 | Hexoses I | Tibia_SOS | -0,00222 | 0,057339 | NA |
| 1 | abundance | SL00104 | Hexoses I | Handgrip | 0,091103 | 0,063361 | NA |
| 2a | abundance | SL00108 | Histidinol | Tibia_length | -0,13454 | 0,053054 | NA |
| 2a | abundance | SL00108 | Histidinol | Radius_length | -0,03618 | 0,072461 | NA |
| 2a | abundance | SL00108 | Histidinol | Radius_SOS | -0,08422 | 0,051232 | NA |
| 2a | abundance | SL00108 | Histidinol | Tibia_SOS | -0,01244 | 0,053756 | NA |
| 2a | abundance | SL00108 | Histidinol | Handgrip | 0,085887 | 0,061373 | NA |
| 2a | abundance | SL00124 | Dissaccari | Tibia_length | -0,12803 | 0,054661 | NA |
| 2a | abundance | SL00124 | Dissaccari | Radius_length | -0,02392 | 0,074042 | NA |
| 2a | abundance | SL00124 | Dissaccari | Radius_SOS | -0,03104 | 0,052244 | NA |
| 2a | abundance | SL00124 | Dissaccari | Tibia_SOS | 0,013602 | 0,055792 | NA |
| 2a | abundance | SL00124 | Dissaccari | Handgrip | 0,038449 | 0,062922 | NA |
| 2a | abundance | SL00128 | N-Acetylal | Tibia_length | 0,001491 | 0,052561 | NA |
| 2a | abundance | SL00128 | N-Acetylal | Radius_length | 0,048435 | 0,071462 | NA |
| 2a | abundance | SL00128 | N-Acetylal | Radius_SOS | 0,012134 | 0,050412 | NA |
| 2a | abundance | SL00128 | N-Acetylal | Tibia_SOS | 0,103101 | 0,054187 | NA |
| 2a | abundance | SL00128 | N-Acetylal | Handgrip | -0,01294 | 0,061673 | NA |
| 2a | abundance | SL00133 | N-Acetylgl | Tibia_length | 0,082277 | 0,052888 | NA |
| 2a | abundance | SL00133 | N-Acetylgl | Radius_length | 0,02883 | 0,071701 | NA |
| 2a | abundance | SL00133 | N-Acetylgl | Radius_SOS | -0,04737 | 0,050496 | NA |
| 2a | abundance | SL00133 | N-Acetylgl | Tibia_SOS | -0,042 | 0,053907 | NA |
| 2a | abundance | SL00133 | N-Acetylgl | Handgrip | 0,015926 | 0,061462 | NA |
| 2a | abundance | SL00137 | Phenylace | Tibia_length | -0,00816 | 0,056013 | NA |
| 2a | abundance | SL00137 | Phenylace | Radius_length | -0,07722 | 0,074054 | NA |
| 2a | abundance | SL00137 | Phenylace | Radius_SOS | 0,051751 | 0,053075 | NA |
| 2a | abundance | SL00137 | Phenylace | Tibia_SOS | -0,08032 | 0,055937 | NA |
| 2a | abundance | SL00137 | Phenylace | Handgrip | -0,00328 | 0,062914 | NA |
| 1 | abundance | SL00140 | Pyridoxam | Tibia_length | 0,098317 | 0,052002 | NA |
| 1 | abundance | SL00140 | Pyridoxam | Radius_length | 0,192125 | 0,070471 | NA |
| 1 | abundance | SL00140 | Pyridoxam | Radius_SOS | 0,055673 | 0,050041 | NA |
| 1 | abundance | SL00140 | Pyridoxam | Tibia_SOS | 0,141937 | 0,052933 | NA |
| 1 | abundance | SL00140 | Pyridoxam | Handgrip | 0,065186 | 0,060184 | NA |

| | | | | | | | |
|---|---|---|---|---|---|---|---|
| 2a | abundance | SL00149 | Dissaccari | Tibia_length | 0,012652 | 0,053152 | NA |
| 2a | abundance | SL00149 | Dissaccari | Radius_length | 0,0814 | 0,072344 | NA |
| 2a | abundance | SL00149 | Dissaccari | Radius_SOS | -0,02475 | 0,05076 | NA |
| 2a | abundance | SL00149 | Dissaccari | Tibia_SOS | -0,01914 | 0,054495 | NA |
| 2a | abundance | SL00149 | Dissaccari | Handgrip | 0,007023 | 0,061698 | NA |
| 1 | abundance | SL00154 | Trigonellin | Tibia_length | -0,03817 | 0,0558 | NA |
| 1 | abundance | SL00154 | Trigonellin | Radius_length | 0,04552 | 0,074413 | NA |
| 1 | abundance | SL00154 | Trigonellin | Radius_SOS | 0,016825 | 0,053091 | NA |
| 1 | abundance | SL00154 | Trigonellin | Tibia_SOS | 0,012266 | 0,056603 | NA |
| 1 | abundance | SL00154 | Trigonellin | Handgrip | 0,094999 | 0,062669 | NA |
| 1 | abundance | SL00159 | Uracil | Tibia_length | 0,025082 | 0,052136 | NA |
| 1 | abundance | SL00159 | Uracil | Radius_length | 0,073715 | 0,072193 | NA |
| 1 | abundance | SL00159 | Uracil | Radius_SOS | 0,008478 | 0,050098 | NA |
| 1 | abundance | SL00159 | Uracil | Tibia_SOS | 0,133603 | 0,052638 | NA |
| 1 | abundance | SL00159 | Uracil | Handgrip | 0,150821 | 0,060206 | NA |
| 2a | abundance | SL00161 | Uridine | Tibia_length | 0,008068 | 0,052375 | NA |
| 2a | abundance | SL00161 | Uridine | Radius_length | 0,079816 | 0,071438 | NA |
| 2a | abundance | SL00161 | Uridine | Radius_SOS | -0,07158 | 0,050165 | NA |
| 2a | abundance | SL00161 | Uridine | Tibia_SOS | -0,00335 | 0,053757 | NA |
| 2a | abundance | SL00161 | Uridine | Handgrip | -0,00114 | 0,061294 | NA |
| 1 | abundance | SL00185 | 2-Oxo-3-pl | Tibia_length | 0,014134 | 0,057348 | NA |
| 1 | abundance | SL00185 | 2-Oxo-3-pl | Radius_length | -0,15339 | 0,074252 | NA |
| 1 | abundance | SL00185 | 2-Oxo-3-pl | Radius_SOS | -0,08692 | 0,054634 | NA |
| 1 | abundance | SL00185 | 2-Oxo-3-pl | Tibia_SOS | 0,025771 | 0,058149 | NA |
| 1 | abundance | SL00185 | 2-Oxo-3-pl | Handgrip | -0,00393 | 0,063529 | NA |
| 2a | abundance | SL00189 | 3-Hydroxyl | Tibia_length | -0,0239 | 0,055254 | NA |
| 2a | abundance | SL00189 | 3-Hydroxyl | Radius_length | -0,06249 | 0,073562 | NA |
| 2a | abundance | SL00189 | 3-Hydroxyl | Radius_SOS | -0,02686 | 0,0523 | NA |
| 2a | abundance | SL00189 | 3-Hydroxyl | Tibia_SOS | -0,05288 | 0,055552 | NA |
| 2a | abundance | SL00189 | 3-Hydroxyl | Handgrip | 0,021065 | 0,063246 | NA |
| 2a | abundance | SL00197 | 4-Methyl-2 | Tibia_length | 0,149031 | 0,055656 | NA |
| 2a | abundance | SL00197 | 4-Methyl-2 | Radius_length | 0,020848 | 0,074836 | NA |
| 2a | abundance | SL00197 | 4-Methyl-2 | Radius_SOS | -0,04537 | 0,05341 | NA |
| 2a | abundance | SL00197 | 4-Methyl-2 | Tibia_SOS | 0,117611 | 0,05733 | NA |
| 2a | abundance | SL00197 | 4-Methyl-2 | Handgrip | 0,069748 | 0,062393 | NA |
| 2a | abundance | SL00199 | 5-Hydroxyi | Tibia_length | -0,07121 | 0,052082 | NA |
| 2a | abundance | SL00199 | 5-Hydroxyi | Radius_length | -0,03116 | 0,071444 | NA |
| 2a | abundance | SL00199 | 5-Hydroxyi | Radius_SOS | -0,09835 | 0,049961 | NA |
| 2a | abundance | SL00199 | 5-Hydroxyi | Tibia_SOS | -0,01069 | 0,053853 | NA |
| 2a | abundance | SL00199 | 5-Hydroxyi | Handgrip | 0,084362 | 0,061425 | NA |
| 2a | abundance | SL00200 | 8-Aminooc | Tibia_length | 0,035841 | 0,052572 | NA |
| 2a | abundance | SL00200 | 8-Aminooc | Radius_length | 0,054387 | 0,072008 | NA |
| 2a | abundance | SL00200 | 8-Aminooc | Radius_SOS | 0,015453 | 0,050484 | NA |
| 2a | abundance | SL00200 | 8-Aminooc | Tibia_SOS | 0,03552 | 0,054745 | NA |
| 2a | abundance | SL00200 | 8-Aminooc | Handgrip | 0,047135 | 0,062308 | NA |
| 2a | abundance | SL00201 | Pentose II | Tibia_length | 0,008171 | 0,05211 | NA |
| 2a | abundance | SL00201 | Pentose II | Radius_length | 0,137927 | 0,071133 | NA |
| 2a | abundance | SL00201 | Pentose II | Radius_SOS | 0,017493 | 0,050016 | NA |

| | | | | | | | |
|---|---|---|---|---|---|---|---|
| 2a | abundance | SL00201 | Pentose II | Tibia_SOS | 0,084491 | 0,052916 | NA |
| 2a | abundance | SL00201 | Pentose II | Handgrip | 0,16256 | 0,060112 | NA |
| 2a | abundance | SL00205 | Citraconic | Tibia_length | -0,04275 | 0,056946 | NA |
| 2a | abundance | SL00205 | Citraconic | Radius_length | -0,02007 | 0,07516 | NA |
| 2a | abundance | SL00205 | Citraconic | Radius_SOS | -0,07389 | 0,054274 | NA |
| 2a | abundance | SL00205 | Citraconic | Tibia_SOS | -0,07278 | 0,056941 | NA |
| 2a | abundance | SL00205 | Citraconic | Handgrip | 0,015264 | 0,06284 | NA |
| 2a | abundance | SL00210 | Galactonic | Tibia_length | -0,05137 | 0,056398 | NA |
| 2a | abundance | SL00210 | Galactonic | Radius_length | -0,05215 | 0,074843 | NA |
| 2a | abundance | SL00210 | Galactonic | Radius_SOS | -0,14495 | 0,054081 | NA |
| 2a | abundance | SL00210 | Galactonic | Tibia_SOS | -0,02708 | 0,056797 | NA |
| 2a | abundance | SL00210 | Galactonic | Handgrip | 0,106994 | 0,063564 | NA |
| 2a | abundance | SL00212 | Galactosar | Tibia_length | -0,03692 | 0,05135 | NA |
| 2a | abundance | SL00212 | Galactosar | Radius_length | -0,18915 | 0,068997 | NA |
| 2a | abundance | SL00212 | Galactosar | Radius_SOS | -0,02064 | 0,049596 | NA |
| 2a | abundance | SL00212 | Galactosar | Tibia_SOS | -0,15177 | 0,052568 | NA |
| 2a | abundance | SL00212 | Galactosar | Handgrip | -0,15415 | 0,060012 | NA |
| 2a | abundance | SL00215 | Glycolic ac | Tibia_length | 0,076565 | 0,055323 | NA |
| 2a | abundance | SL00215 | Glycolic ac | Radius_length | 0,185742 | 0,072743 | NA |
| 2a | abundance | SL00215 | Glycolic ac | Radius_SOS | 0,04022 | 0,052399 | NA |
| 2a | abundance | SL00215 | Glycolic ac | Tibia_SOS | 0,143253 | 0,056301 | NA |
| 2a | abundance | SL00215 | Glycolic ac | Handgrip | 0,063361 | 0,062167 | NA |
| 2a | abundance | SL00216 | Glyoxylic a | Tibia_length | -0,0468 | 0,053364 | NA |
| 2a | abundance | SL00216 | Glyoxylic a | Radius_length | -0,05534 | 0,07285 | NA |
| 2a | abundance | SL00216 | Glyoxylic a | Radius_SOS | -0,03745 | 0,051107 | NA |
| 2a | abundance | SL00216 | Glyoxylic a | Tibia_SOS | 0,060596 | 0,054854 | NA |
| 2a | abundance | SL00216 | Glyoxylic a | Handgrip | 0,125939 | 0,061807 | NA |
| 1 | abundance | SL00222 | Lactic acid | Tibia_length | -0,03093 | 0,055746 | NA |
| 1 | abundance | SL00222 | Lactic acid | Radius_length | 0,003612 | 0,074205 | NA |
| 1 | abundance | SL00222 | Lactic acid | Radius_SOS | -0,02295 | 0,053301 | NA |
| 1 | abundance | SL00222 | Lactic acid | Tibia_SOS | -0,01346 | 0,056555 | NA |
| 1 | abundance | SL00222 | Lactic acid | Handgrip | 0,154658 | 0,061085 | NA |
| 2a | abundance | SL00226 | N-Acetylgly | Tibia_length | 0,096856 | 0,053905 | NA |
| 2a | abundance | SL00226 | N-Acetylgly | Radius_length | 0,156046 | 0,071969 | NA |
| 2a | abundance | SL00226 | N-Acetylgly | Radius_SOS | -0,00595 | 0,051516 | NA |
| 2a | abundance | SL00226 | N-Acetylgly | Tibia_SOS | 0,098041 | 0,055889 | NA |
| 2a | abundance | SL00226 | N-Acetylgly | Handgrip | 0,058154 | 0,06196 | NA |
| 2a | abundance | SL00239 | Pinitol | Tibia_length | -0,09634 | 0,055063 | NA |
| 2a | abundance | SL00239 | Pinitol | Radius_length | -0,03699 | 0,073517 | NA |
| 2a | abundance | SL00239 | Pinitol | Radius_SOS | 0,02458 | 0,052375 | NA |
| 2a | abundance | SL00239 | Pinitol | Tibia_SOS | -0,06518 | 0,056326 | NA |
| 2a | abundance | SL00239 | Pinitol | Handgrip | -0,06071 | 0,063407 | NA |
| 1 | abundance | SL00240 | Pyruvic aci | Tibia_length | 0,078354 | 0,055419 | NA |
| 1 | abundance | SL00240 | Pyruvic aci | Radius_length | 0,029946 | 0,074345 | NA |
| 1 | abundance | SL00240 | Pyruvic aci | Radius_SOS | -0,06008 | 0,053179 | NA |
| 1 | abundance | SL00240 | Pyruvic aci | Tibia_SOS | 0,068526 | 0,056074 | NA |
| 1 | abundance | SL00240 | Pyruvic aci | Handgrip | 0,211058 | 0,060657 | NA |
| 2a | abundance | SL00243 | Deoxysuga | Tibia_length | 0,005599 | 0,054567 | NA |

| | | | | | | | |
|---|---|---|---|---|---|---|---|
| 2a | abundance | SL00243 | Deoxysuga | Radius_length | 0,08573 | 0,073889 | NA |
| 2a | abundance | SL00243 | Deoxysuga | Radius_SOS | 0,053501 | 0,051745 | NA |
| 2a | abundance | SL00243 | Deoxysuga | Tibia_SOS | 0,00575 | 0,054985 | NA |
| 2a | abundance | SL00243 | Deoxysuga | Handgrip | 0,139053 | 0,061127 | NA |
| 2a | abundance | SL00245 | sugar alcol | Tibia_length | 0,010534 | 0,057713 | NA |
| 2a | abundance | SL00245 | sugar alcol | Radius_length | 0,081711 | 0,07603 | NA |
| 2a | abundance | SL00245 | sugar alcol | Radius_SOS | 0,078648 | 0,054424 | NA |
| 2a | abundance | SL00245 | sugar alcol | Tibia_SOS | 0,009817 | 0,057993 | NA |
| 2a | abundance | SL00245 | sugar alcol | Handgrip | 0,169216 | 0,061121 | NA |
| 2a | abundance | SL00246 | Pentose III | Tibia_length | -0,01068 | 0,052437 | NA |
| 2a | abundance | SL00246 | Pentose III | Radius_length | 0,023625 | 0,072241 | NA |
| 2a | abundance | SL00246 | Pentose III | Radius_SOS | 0,006333 | 0,050271 | NA |
| 2a | abundance | SL00246 | Pentose III | Tibia_SOS | 0,036428 | 0,053785 | NA |
| 2a | abundance | SL00246 | Pentose III | Handgrip | 0,107443 | 0,061282 | NA |
| 2a | abundance | SL00247 | Shikimic a | Tibia_length | 0,012398 | 0,058755 | NA |
| 2a | abundance | SL00247 | Shikimic a | Radius_length | -0,06695 | 0,075403 | NA |
| 2a | abundance | SL00247 | Shikimic a | Radius_SOS | -0,0571 | 0,056242 | NA |
| 2a | abundance | SL00247 | Shikimic a | Tibia_SOS | 0,065584 | 0,060221 | NA |
| 2a | abundance | SL00247 | Shikimic a | Handgrip | 0,010105 | 0,062661 | NA |
| 2a | abundance | SL00248 | Gluconic a | Tibia_length | 0,009536 | 0,05365 | NA |
| 2a | abundance | SL00248 | Gluconic a | Radius_length | 0,139038 | 0,072643 | NA |
| 2a | abundance | SL00248 | Gluconic a | Radius_SOS | -3,32E-04 | 0,051118 | NA |
| 2a | abundance | SL00248 | Gluconic a | Tibia_SOS | 0,069417 | 0,05378 | NA |
| 2a | abundance | SL00248 | Gluconic a | Handgrip | 0,113344 | 0,061552 | NA |
| 2a | abundance | SL00260 | Pentose I | Tibia_length | -0,02099 | 0,055023 | NA |
| 2a | abundance | SL00260 | Pentose I | Radius_length | 0,051073 | 0,073928 | NA |
| 2a | abundance | SL00260 | Pentose I | Radius_SOS | -0,0408 | 0,052356 | NA |
| 2a | abundance | SL00260 | Pentose I | Tibia_SOS | 0,002856 | 0,055683 | NA |
| 2a | abundance | SL00260 | Pentose I | Handgrip | 0,115027 | 0,062616 | NA |
| 2a | abundance | SL00262 | cis-Aconiti | Tibia_length | -0,11144 | 0,054552 | NA |
| 2a | abundance | SL00262 | cis-Aconiti | Radius_length | -0,02852 | 0,073558 | NA |
| 2a | abundance | SL00262 | cis-Aconiti | Radius_SOS | -0,05481 | 0,052164 | NA |
| 2a | abundance | SL00262 | cis-Aconiti | Tibia_SOS | -0,12556 | 0,056242 | NA |
| 2a | abundance | SL00262 | cis-Aconiti | Handgrip | -0,03078 | 0,062632 | NA |
| 2a | abundance | SL00264 | Methyl ace | Tibia_length | 0,108681 | 0,052703 | NA |
| 2a | abundance | SL00264 | Methyl ace | Radius_length | 0,129369 | 0,071284 | NA |
| 2a | abundance | SL00264 | Methyl ace | Radius_SOS | 0,027977 | 0,050442 | NA |
| 2a | abundance | SL00264 | Methyl ace | Tibia_SOS | 0,097809 | 0,0541 | NA |
| 2a | abundance | SL00264 | Methyl ace | Handgrip | -0,03004 | 0,061554 | NA |
| 2a | abundance | SL00265 | Purine | Tibia_length | 0,01791 | 0,054787 | NA |
| 2a | abundance | SL00265 | Purine | Radius_length | 0,107272 | 0,073304 | NA |
| 2a | abundance | SL00265 | Purine | Radius_SOS | 0,033035 | 0,051959 | NA |
| 2a | abundance | SL00265 | Purine | Tibia_SOS | 0,037921 | 0,055948 | NA |
| 2a | abundance | SL00265 | Purine | Handgrip | 0,11876 | 0,061432 | NA |
| 1 | abundance | SL00268 | Thymine | Tibia_length | 0,016409 | 0,054066 | NA |
| 1 | abundance | SL00268 | Thymine | Radius_length | -0,01377 | 0,07334 | NA |
| 1 | abundance | SL00268 | Thymine | Radius_SOS | -0,02366 | 0,051537 | NA |
| 1 | abundance | SL00268 | Thymine | Tibia_SOS | 0,156614 | 0,056199 | NA |

| | | | | | | | | |
|---|---|---|---|---|---|---|---|---|
| | 1 | abundance | SL00268 | Thymine | Handgrip | 0,021533 | 0,06367 | NA |
| 2a | | abundance | SL00270 | 3-hydroxy-: | Tibia_length | -0,07854 | 0,052996 | NA |
| 2a | | abundance | SL00270 | 3-hydroxy-: | Radius_length | 0,025357 | 0,071931 | NA |
| 2a | | abundance | SL00270 | 3-hydroxy-: | Radius_SOS | 0,037703 | 0,050825 | NA |
| 2a | | abundance | SL00270 | 3-hydroxy-: | Tibia_SOS | -0,01957 | 0,053407 | NA |
| 2a | | abundance | SL00270 | 3-hydroxy-: | Handgrip | 0,018091 | 0,060948 | NA |
| 2a | | abundance | SL00275 | Dopamine | Tibia_length | -0,15652 | 0,053522 | NA |
| 2a | | abundance | SL00275 | Dopamine | Radius_length | -0,13046 | 0,071774 | NA |
| 2a | | abundance | SL00275 | Dopamine | Radius_SOS | -0,09189 | 0,051085 | NA |
| 2a | | abundance | SL00275 | Dopamine | Tibia_SOS | -0,0907 | 0,054268 | NA |
| 2a | | abundance | SL00275 | Dopamine | Handgrip | -0,0336 | 0,061516 | NA |
| 2a | | abundance | SL00282 | 3-Hydroxyl | Tibia_length | -0,15235 | 0,058341 | NA |
| 2a | | abundance | SL00282 | 3-Hydroxyl | Radius_length | -0,16759 | 0,07484 | NA |
| 2a | | abundance | SL00282 | 3-Hydroxyl | Radius_SOS | -0,04951 | 0,055751 | NA |
| 2a | | abundance | SL00282 | 3-Hydroxyl | Tibia_SOS | -0,1374 | 0,051989 | NA |
| 2a | | abundance | SL00282 | 3-Hydroxyl | Handgrip | -0,18441 | 0,061177 | NA |
| 2a | | abundance | SL00285 | Dihydrofer | Tibia_length | -0,03433 | 0,052824 | NA |
| 2a | | abundance | SL00285 | Dihydrofer | Radius_length | 0,034876 | 0,071722 | NA |
| 2a | | abundance | SL00285 | Dihydrofer | Radius_SOS | -0,03832 | 0,050632 | NA |
| 2a | | abundance | SL00285 | Dihydrofer | Tibia_SOS | -0,12032 | 0,054519 | NA |
| 2a | | abundance | SL00285 | Dihydrofer | Handgrip | 0,124842 | 0,062391 | NA |
| 2a | | abundance | SL00287 | DOPA | Tibia_length | -0,04216 | 0,051046 | NA |
| 2a | | abundance | SL00287 | DOPA | Radius_length | -0,07406 | 0,06955 | NA |
| 2a | | abundance | SL00287 | DOPA | Radius_SOS | -0,00211 | 0,049394 | NA |
| 2a | | abundance | SL00287 | DOPA | Tibia_SOS | -0,09933 | 0,052336 | NA |
| 2a | | abundance | SL00287 | DOPA | Handgrip | -0,10213 | 0,06032 | NA |
| 2a | | abundance | SL00288 | Propionylc | Tibia_length | -0,01451 | 0,058782 | NA |
| 2a | | abundance | SL00288 | Propionylc | Radius_length | 0,018426 | 0,07681 | NA |
| 2a | | abundance | SL00288 | Propionylc | Radius_SOS | 0,084762 | 0,055609 | NA |
| 2a | | abundance | SL00288 | Propionylc | Tibia_SOS | 0,008469 | 0,057913 | NA |
| 2a | | abundance | SL00288 | Propionylc | Handgrip | 0,001225 | 0,060193 | NA |
| 2a | | abundance | SL00290 | N-Acetylm | Tibia_length | 0,017181 | 0,054518 | NA |
| 2a | | abundance | SL00290 | N-Acetylm | Radius_length | 0,061096 | 0,073785 | NA |
| 2a | | abundance | SL00290 | N-Acetylm | Radius_SOS | -0,03268 | 0,051996 | NA |
| 2a | | abundance | SL00290 | N-Acetylm | Tibia_SOS | 0,022794 | 0,055404 | NA |
| 2a | | abundance | SL00290 | N-Acetylm | Handgrip | 0,091184 | 0,062619 | NA |
| 2a | | abundance | SL00293 | Butyrylcarr | Tibia_length | -0,03089 | 0,057787 | NA |
| 2a | | abundance | SL00293 | Butyrylcarr | Radius_length | 0,096599 | 0,074835 | NA |
| 2a | | abundance | SL00293 | Butyrylcarr | Radius_SOS | 0,023373 | 0,055046 | NA |
| 2a | | abundance | SL00293 | Butyrylcarr | Tibia_SOS | 0,086278 | 0,057419 | NA |
| 2a | | abundance | SL00293 | Butyrylcarr | Handgrip | -0,0112 | 0,062653 | NA |
| 2a | | abundance | SL00295 | 5-Methylur | Tibia_length | 0,04092 | 0,054029 | NA |
| 2a | | abundance | SL00295 | 5-Methylur | Radius_length | -0,00711 | 0,073246 | NA |
| 2a | | abundance | SL00295 | 5-Methylur | Radius_SOS | -0,09428 | 0,051184 | NA |
| 2a | | abundance | SL00295 | 5-Methylur | Tibia_SOS | 0,075648 | 0,054223 | NA |
| 2a | | abundance | SL00295 | 5-Methylur | Handgrip | 0,004469 | 0,061795 | NA |
| 2a | | abundance | SL00297 | Acetylmura | Tibia_length | -0,00029 | 0,049475 | NA |
| 2a | | abundance | SL00297 | Acetylmura | Radius_length | -0,07829 | 0,066803 | NA |

| | | | | | | | |
|---|---|---|---|---|---|---|---|
| 2a | abundance | SL00297 | Acetylmura | Radius_SOS | -0,00764 | 0,049037 | NA |
| 2a | abundance | SL00297 | Acetylmura | Tibia_SOS | 0,05506 | 0,052517 | NA |
| 2a | abundance | SL00297 | Acetylmura | Handgrip | 0,033434 | 0,060306 | NA |
| 2a | abundance | SL00299 | Î²-Murichol | Tibia_length | 0,037161 | 0,050668 | NA |
| 2a | abundance | SL00299 | Î²-Murichol | Radius_length | 0,112809 | 0,068916 | NA |
| 2a | abundance | SL00299 | Î²-Murichol | Radius_SOS | 0,072249 | 0,049124 | NA |
| 2a | abundance | SL00299 | Î²-Murichol | Tibia_SOS | -0,03557 | 0,053218 | NA |
| 2a | abundance | SL00299 | Î²-Murichol | Handgrip | 0,031215 | 0,061505 | NA |
| 2a | abundance | SL00309 | 1-Aminocy | Tibia_length | 0,088526 | 0,050865 | NA |
| 2a | abundance | SL00309 | 1-Aminocy | Radius_length | 0,172024 | 0,068701 | NA |
| 2a | abundance | SL00309 | 1-Aminocy | Radius_SOS | 0,027035 | 0,049382 | NA |
| 2a | abundance | SL00309 | 1-Aminocy | Tibia_SOS | 0,161945 | 0,052253 | NA |
| 2a | abundance | SL00309 | 1-Aminocy | Handgrip | 0,102164 | 0,060353 | NA |
| 2a | abundance | SL00311 | N,N-Dimet | Tibia_length | -0,09507 | 0,053519 | NA |
| 2a | abundance | SL00311 | N,N-Dimet | Radius_length | -0,0583 | 0,072645 | NA |
| 2a | abundance | SL00311 | N,N-Dimet | Radius_SOS | 0,005618 | 0,051204 | NA |
| 2a | abundance | SL00311 | N,N-Dimet | Tibia_SOS | 0,012809 | 0,054628 | NA |
| 2a | abundance | SL00311 | N,N-Dimet | Handgrip | -0,00691 | 0,062289 | NA |
| 2a | abundance | SL00313 | 2-Aminois( | Tibia_length | -0,10501 | 0,054959 | NA |
| 2a | abundance | SL00313 | 2-Aminois( | Radius_length | -0,05205 | 0,074069 | NA |
| 2a | abundance | SL00313 | 2-Aminois( | Radius_SOS | -0,06592 | 0,052199 | NA |
| 2a | abundance | SL00313 | 2-Aminois( | Tibia_SOS | -0,09447 | 0,056459 | NA |
| 2a | abundance | SL00313 | 2-Aminois( | Handgrip | -0,06932 | 0,060739 | NA |
| 2a | abundance | SL00315 | Malonic ac | Tibia_length | 0,074932 | 0,049702 | NA |
| 2a | abundance | SL00315 | Malonic ac | Radius_length | 0,127454 | 0,067079 | NA |
| 2a | abundance | SL00315 | Malonic ac | Radius_SOS | 0,013086 | 0,049046 | NA |
| 2a | abundance | SL00315 | Malonic ac | Tibia_SOS | 0,032286 | 0,05246 | NA |
| 2a | abundance | SL00315 | Malonic ac | Handgrip | 0,128273 | 0,059701 | NA |
| 1 | abundance | SL00318 | 4-Aminoph | Tibia_length | -0,15098 | 0,053085 | NA |
| 1 | abundance | SL00318 | 4-Aminoph | Radius_length | -0,22651 | 0,070812 | NA |
| 1 | abundance | SL00318 | 4-Aminoph | Radius_SOS | -0,01327 | 0,050809 | NA |
| 1 | abundance | SL00318 | 4-Aminoph | Tibia_SOS | -0,05745 | 0,054488 | NA |
| 1 | abundance | SL00318 | 4-Aminoph | Handgrip | -0,06931 | 0,061647 | NA |
| 2a | abundance | SL00320 | Î³-Caprola( | Tibia_length | -0,12601 | 0,053226 | NA |
| 2a | abundance | SL00320 | Î³-Caprola( | Radius_length | -0,08914 | 0,072137 | NA |
| 2a | abundance | SL00320 | Î³-Caprola( | Radius_SOS | 0,014039 | 0,051008 | NA |
| 2a | abundance | SL00320 | Î³-Caprola( | Tibia_SOS | -0,10028 | 0,053929 | NA |
| 2a | abundance | SL00320 | Î³-Caprola( | Handgrip | -0,0743 | 0,06118 | NA |
| 2a | abundance | SL00325 | 2-Methylm | Tibia_length | -0,01346 | 0,056494 | NA |
| 2a | abundance | SL00325 | 2-Methylm | Radius_length | 0,046891 | 0,074239 | NA |
| 2a | abundance | SL00325 | 2-Methylm | Radius_SOS | 0,02923 | 0,053651 | NA |
| 2a | abundance | SL00325 | 2-Methylm | Tibia_SOS | 0,052754 | 0,056954 | NA |
| 2a | abundance | SL00325 | 2-Methylm | Handgrip | -0,0536 | 0,06284 | NA |
| 2a | abundance | SL00328 | 2-(hydroxy | Tibia_length | 0,07241 | 0,051169 | NA |
| 2a | abundance | SL00328 | 2-(hydroxy | Radius_length | 0,001366 | 0,070509 | NA |
| 2a | abundance | SL00328 | 2-(hydroxy | Radius_SOS | 0,018836 | 0,049602 | NA |
| 2a | abundance | SL00328 | 2-(hydroxy | Tibia_SOS | -0,04521 | 0,053191 | NA |
| 2a | abundance | SL00328 | 2-(hydroxy | Handgrip | -0,01753 | 0,061193 | NA |

| | | | | | | | |
|---|---|---|---|---|---|---|---|
| 2a | abundance | SL00329 | 2-Hydroxy- | Tibia_length | 0,094474 | 0,053647 | NA |
| 2a | abundance | SL00329 | 2-Hydroxy- | Radius_length | 0,138373 | 0,072522 | NA |
| 2a | abundance | SL00329 | 2-Hydroxy- | Radius_SOS | 0,06541 | 0,050982 | NA |
| 2a | abundance | SL00329 | 2-Hydroxy- | Tibia_SOS | 0,057765 | 0,054836 | NA |
| 2a | abundance | SL00329 | 2-Hydroxy- | Handgrip | 0,180232 | 0,0601 | NA |
| 2a | abundance | SL00334 | Imidazolea | Tibia_length | 0,026929 | 0,056191 | NA |
| 2a | abundance | SL00334 | Imidazolea | Radius_length | 0,078178 | 0,073942 | NA |
| 2a | abundance | SL00334 | Imidazolea | Radius_SOS | 0,086138 | 0,053123 | NA |
| 2a | abundance | SL00334 | Imidazolea | Tibia_SOS | 0,026731 | 0,056915 | NA |
| 2a | abundance | SL00334 | Imidazolea | Handgrip | 0,075469 | 0,063575 | NA |
| 2a | abundance | SL00346 | N-Methylni | Tibia_length | -0,07487 | 0,054864 | NA |
| 2a | abundance | SL00346 | N-Methylni | Radius_length | -0,16307 | 0,072802 | NA |
| 2a | abundance | SL00346 | N-Methylni | Radius_SOS | -0,07165 | 0,051805 | NA |
| 2a | abundance | SL00346 | N-Methylni | Tibia_SOS | -0,05598 | 0,05452 | NA |
| 2a | abundance | SL00346 | N-Methylni | Handgrip | -0,02755 | 0,060921 | NA |
| 2a | abundance | SL00349 | 1-Aminocy | Tibia_length | 0,027664 | 0,05148 | NA |
| 2a | abundance | SL00349 | 1-Aminocy | Radius_length | -0,05581 | 0,070351 | NA |
| 2a | abundance | SL00349 | 1-Aminocy | Radius_SOS | 0,00817 | 0,049768 | NA |
| 2a | abundance | SL00349 | 1-Aminocy | Tibia_SOS | 0,009514 | 0,053005 | NA |
| 2a | abundance | SL00349 | 1-Aminocy | Handgrip | -0,01972 | 0,061861 | NA |
| 1 | abundance | SL00350 | Stachydrin | Tibia_length | -0,0552 | 0,051634 | NA |
| 1 | abundance | SL00350 | Stachydrin | Radius_length | -0,03224 | 0,070586 | NA |
| 1 | abundance | SL00350 | Stachydrin | Radius_SOS | -0,06207 | 0,049756 | NA |
| 1 | abundance | SL00350 | Stachydrin | Tibia_SOS | -0,05243 | 0,053436 | NA |
| 1 | abundance | SL00350 | Stachydrin | Handgrip | 0,053936 | 0,060519 | NA |
| 1 | abundance | SL00353 | Adipic acid | Tibia_length | 0,006779 | 0,051098 | NA |
| 1 | abundance | SL00353 | Adipic acid | Radius_length | 0,095772 | 0,069781 | NA |
| 1 | abundance | SL00353 | Adipic acid | Radius_SOS | -0,00857 | 0,049511 | NA |
| 1 | abundance | SL00353 | Adipic acid | Tibia_SOS | -0,03704 | 0,053005 | NA |
| 1 | abundance | SL00353 | Adipic acid | Handgrip | -0,06198 | 0,061117 | NA |
| 2a | abundance | SL00356 | N-(5-Amin | Tibia_length | -0,00498 | 0,050128 | NA |
| 2a | abundance | SL00356 | N-(5-Amin | Radius_length | -0,03764 | 0,068219 | NA |
| 2a | abundance | SL00356 | N-(5-Amin | Radius_SOS | -0,04199 | 0,049068 | NA |
| 2a | abundance | SL00356 | N-(5-Amin | Tibia_SOS | -0,06363 | 0,052707 | NA |
| 2a | abundance | SL00356 | N-(5-Amin | Handgrip | 0,009885 | 0,060852 | NA |
| 1 | abundance | SL00368 | N-Methylty | Tibia_length | -0,03872 | 0,05789 | NA |
| 1 | abundance | SL00368 | N-Methylty | Radius_length | -0,04282 | 0,074852 | NA |
| 1 | abundance | SL00368 | N-Methylty | Radius_SOS | -0,01442 | 0,055139 | NA |
| 1 | abundance | SL00368 | N-Methylty | Tibia_SOS | 0,073177 | 0,058224 | NA |
| 1 | abundance | SL00368 | N-Methylty | Handgrip | 0,016485 | 0,060185 | NA |
| 2a | abundance | SL00373 | N1-Methyl- | Tibia_length | 0,037698 | 0,048957 | NA |
| 2a | abundance | SL00373 | N1-Methyl- | Radius_length | 0,153391 | 0,063223 | NA |
| 2a | abundance | SL00373 | N1-Methyl- | Radius_SOS | 0,052749 | 0,049351 | NA |
| 2a | abundance | SL00373 | N1-Methyl- | Tibia_SOS | -4,64E-04 | 0,052719 | NA |
| 2a | abundance | SL00373 | N1-Methyl- | Handgrip | 0,131975 | 0,059711 | NA |
| 2a | abundance | SL00376 | N-AcetylPr | Tibia_length | -0,1477 | 0,056404 | NA |
| 2a | abundance | SL00376 | N-AcetylPr | Radius_length | -0,06527 | 0,074212 | NA |
| 2a | abundance | SL00376 | N-AcetylPr | Radius_SOS | -0,0933 | 0,053314 | NA |

| | | | | | | | |
|---|---|---|---|---|---|---|---|
| 2a | abundance | SL00376 | N-AcetylPr | Tibia_SOS | -0,10165 | 0,055891 | NA |
| 2a | abundance | SL00376 | N-AcetylPr | Handgrip | -0,10658 | 0,062121 | NA |
| 2a | abundance | SL00380 | 2-Aminoad | Tibia_length | 0,033022 | 0,051481 | NA |
| 2a | abundance | SL00380 | 2-Aminoad | Radius_length | 0,144636 | 0,070157 | NA |
| 2a | abundance | SL00380 | 2-Aminoad | Radius_SOS | 0,015641 | 0,049742 | NA |
| 2a | abundance | SL00380 | 2-Aminoad | Tibia_SOS | 0,019528 | 0,053363 | NA |
| 2a | abundance | SL00380 | 2-Aminoad | Handgrip | -0,01457 | 0,0617 | NA |
| 2a | abundance | SL00383 | 3-(2-Hydro | Tibia_length | 0,040579 | 0,053681 | NA |
| 2a | abundance | SL00383 | 3-(2-Hydro | Radius_length | 0,026797 | 0,072616 | NA |
| 2a | abundance | SL00383 | 3-(2-Hydro | Radius_SOS | 0,069628 | 0,051055 | NA |
| 2a | abundance | SL00383 | 3-(2-Hydro | Tibia_SOS | 0,107294 | 0,055476 | NA |
| 2a | abundance | SL00383 | 3-(2-Hydro | Handgrip | 0,001674 | 0,062196 | NA |
| 1 | abundance | SL00384 | p-Coumari | Tibia_length | 0,057215 | 0,052527 | NA |
| 1 | abundance | SL00384 | p-Coumari | Radius_length | -0,05666 | 0,071483 | NA |
| 1 | abundance | SL00384 | p-Coumari | Radius_SOS | -0,0686 | 0,050246 | NA |
| 1 | abundance | SL00384 | p-Coumari | Tibia_SOS | -0,0362 | 0,054262 | NA |
| 1 | abundance | SL00384 | p-Coumari | Handgrip | -0,02449 | 0,062831 | NA |
| 2a | abundance | SL00390 | Gallic acid | Tibia_length | -0,14184 | 0,052928 | NA |
| 2a | abundance | SL00390 | Gallic acid | Radius_length | -0,08934 | 0,072237 | NA |
| 2a | abundance | SL00390 | Gallic acid | Radius_SOS | -0,03733 | 0,050858 | NA |
| 2a | abundance | SL00390 | Gallic acid | Tibia_SOS | -0,10199 | 0,053897 | NA |
| 2a | abundance | SL00390 | Gallic acid | Handgrip | -0,056 | 0,061532 | NA |
| 1 | abundance | SL00391 | N-Acetyllei | Tibia_length | 0,117539 | 0,052018 | NA |
| 1 | abundance | SL00391 | N-Acetyllei | Radius_length | 0,105039 | 0,071058 | NA |
| 1 | abundance | SL00391 | N-Acetyllei | Radius_SOS | 0,052039 | 0,050118 | NA |
| 1 | abundance | SL00391 | N-Acetyllei | Tibia_SOS | 0,067062 | 0,054035 | NA |
| 1 | abundance | SL00391 | N-Acetyllei | Handgrip | -0,00693 | 0,061799 | NA |
| 1 | abundance | SL00401 | Theophyllii | Tibia_length | -0,0628 | 0,054755 | NA |
| 1 | abundance | SL00401 | Theophyllii | Radius_length | -0,10832 | 0,073023 | NA |
| 1 | abundance | SL00401 | Theophyllii | Radius_SOS | -0,00796 | 0,052294 | NA |
| 1 | abundance | SL00401 | Theophyllii | Tibia_SOS | -0,0203 | 0,054883 | NA |
| 1 | abundance | SL00401 | Theophyllii | Handgrip | -0,05552 | 0,061614 | NA |
| 2a | abundance | SL00407 | Homovanil | Tibia_length | -0,05292 | 0,051488 | NA |
| 2a | abundance | SL00407 | Homovanil | Radius_length | -0,12304 | 0,07015 | NA |
| 2a | abundance | SL00407 | Homovanil | Radius_SOS | -0,04355 | 0,049687 | NA |
| 2a | abundance | SL00407 | Homovanil | Tibia_SOS | -0,07294 | 0,053425 | NA |
| 2a | abundance | SL00407 | Homovanil | Handgrip | -0,02086 | 0,060514 | NA |
| 2a | abundance | SL00420 | 4-Hydroxy- | Tibia_length | -0,05251 | 0,054628 | NA |
| 2a | abundance | SL00420 | 4-Hydroxy- | Radius_length | -0,17443 | 0,072572 | NA |
| 2a | abundance | SL00420 | 4-Hydroxy- | Radius_SOS | -0,07267 | 0,051859 | NA |
| 2a | abundance | SL00420 | 4-Hydroxy- | Tibia_SOS | -0,11234 | 0,054992 | NA |
| 2a | abundance | SL00420 | 4-Hydroxy- | Handgrip | -0,13907 | 0,061633 | NA |
| 2a | abundance | SL00421 | Asymmetri | Tibia_length | -0,13302 | 0,048973 | NA |
| 2a | abundance | SL00421 | Asymmetri | Radius_length | -0,1599 | 0,065888 | NA |
| 2a | abundance | SL00421 | Asymmetri | Radius_SOS | -0,07012 | 0,048911 | NA |
| 2a | abundance | SL00421 | Asymmetri | Tibia_SOS | -0,11276 | 0,052177 | NA |
| 2a | abundance | SL00421 | Asymmetri | Handgrip | -0,0207 | 0,060374 | NA |
| 2a | abundance | SL00430 | Homocarn | Tibia_length | -0,0569 | 0,05343 | NA |

| | | | | | | | |
|---|---|---|---|---|---|---|---|
| 2a | abundance | SL00430 | Homocarn | Radius_length | -0,01705 | 0,072397 | NA |
| 2a | abundance | SL00430 | Homocarn | Radius_SOS | 0,024722 | 0,051051 | NA |
| 2a | abundance | SL00430 | Homocarn | Tibia_SOS | 0,013088 | 0,054475 | NA |
| 2a | abundance | SL00430 | Homocarn | Handgrip | -0,01366 | 0,062179 | NA |
| 2a | abundance | SL00431 | Tiglylcarnit | Tibia_length | -0,06788 | 0,053356 | NA |
| 2a | abundance | SL00431 | Tiglylcarnit | Radius_length | -0,1715 | 0,071585 | NA |
| 2a | abundance | SL00431 | Tiglylcarnit | Radius_SOS | -0,00761 | 0,05101 | NA |
| 2a | abundance | SL00431 | Tiglylcarnit | Tibia_SOS | -0,15399 | 0,053619 | NA |
| 2a | abundance | SL00431 | Tiglylcarnit | Handgrip | -0,1166 | 0,061812 | NA |
| 2a | abundance | SL00433 | Isovalerylc | Tibia_length | -0,08338 | 0,053534 | NA |
| 2a | abundance | SL00433 | Isovalerylc | Radius_length | 0,122261 | 0,072623 | NA |
| 2a | abundance | SL00433 | Isovalerylc | Radius_SOS | -0,02281 | 0,051298 | NA |
| 2a | abundance | SL00433 | Isovalerylc | Tibia_SOS | -0,02342 | 0,054548 | NA |
| 2a | abundance | SL00433 | Isovalerylc | Handgrip | 0,01485 | 0,061988 | NA |
| 2a | abundance | SL00436 | 1-Carboxy€ | Tibia_length | 0,016997 | 0,053739 | NA |
| 2a | abundance | SL00436 | 1-Carboxy€ | Radius_length | 0,089667 | 0,072183 | NA |
| 2a | abundance | SL00436 | 1-Carboxy€ | Radius_SOS | 0,083109 | 0,051038 | NA |
| 2a | abundance | SL00436 | 1-Carboxy€ | Tibia_SOS | 0,065646 | 0,053994 | NA |
| 2a | abundance | SL00436 | 1-Carboxy€ | Handgrip | 0,031512 | 0,061049 | NA |
| 2a | abundance | SL00438 | Daidzein | Tibia_length | -0,00822 | 0,054102 | NA |
| 2a | abundance | SL00438 | Daidzein | Radius_length | -0,02087 | 0,073589 | NA |
| 2a | abundance | SL00438 | Daidzein | Radius_SOS | 0,011044 | 0,051641 | NA |
| 2a | abundance | SL00438 | Daidzein | Tibia_SOS | 0,044082 | 0,055274 | NA |
| 2a | abundance | SL00438 | Daidzein | Handgrip | 0,016786 | 0,063367 | NA |
| 2a | abundance | SL00440 | 5-Methylcy | Tibia_length | 0,017369 | 0,052785 | NA |
| 2a | abundance | SL00440 | 5-Methylcy | Radius_length | 0,195762 | 0,070411 | NA |
| 2a | abundance | SL00440 | 5-Methylcy | Radius_SOS | 0,011286 | 0,05052 | NA |
| 2a | abundance | SL00440 | 5-Methylcy | Tibia_SOS | 0,086294 | 0,05424 | NA |
| 2a | abundance | SL00440 | 5-Methylcy | Handgrip | 0,121476 | 0,060584 | NA |
| 2a | abundance | SL00445 | Glucosami | Tibia_length | -0,08718 | 0,055924 | NA |
| 2a | abundance | SL00445 | Glucosami | Radius_length | -0,06493 | 0,074222 | NA |
| 2a | abundance | SL00445 | Glucosami | Radius_SOS | -0,09879 | 0,053543 | NA |
| 2a | abundance | SL00445 | Glucosami | Tibia_SOS | -0,02486 | 0,056434 | NA |
| 2a | abundance | SL00445 | Glucosami | Handgrip | 0,001549 | 0,062895 | NA |
| 2a | abundance | SL00447 | Apigenin | Tibia_length | 0,021945 | 0,05396 | NA |
| 2a | abundance | SL00447 | Apigenin | Radius_length | -0,0033 | 0,073333 | NA |
| 2a | abundance | SL00447 | Apigenin | Radius_SOS | -0,05346 | 0,051544 | NA |
| 2a | abundance | SL00447 | Apigenin | Tibia_SOS | -0,02202 | 0,056457 | NA |
| 2a | abundance | SL00447 | Apigenin | Handgrip | 0,090721 | 0,062732 | NA |
| 2a | abundance | SL00455 | 12,13-DH( | Tibia_length | 0,106408 | 0,05165 | NA |
| 2a | abundance | SL00455 | 12,13-DH( | Radius_length | 0,175376 | 0,069624 | NA |
| 2a | abundance | SL00455 | 12,13-DH( | Radius_SOS | 0,04034 | 0,04974 | NA |
| 2a | abundance | SL00455 | 12,13-DH( | Tibia_SOS | 0,014666 | 0,053131 | NA |
| 2a | abundance | SL00455 | 12,13-DH( | Handgrip | 0,021071 | 0,060772 | NA |
| 2a | abundance | SL00467 | Sucralose | Tibia_length | -0,01487 | 0,05373 | NA |
| 2a | abundance | SL00467 | Sucralose | Radius_length | -0,02797 | 0,072539 | NA |
| 2a | abundance | SL00467 | Sucralose | Radius_SOS | 0,048315 | 0,051291 | NA |
| 2a | abundance | SL00467 | Sucralose | Tibia_SOS | -0,00145 | 0,054815 | NA |

| | | | | | | | | |
|---|---|---|---|---|---|---|---|---|
| 2a | | abundance | SL00467 | Sucralose | Handgrip | 0,040725 | 0,062307 | NA |
| | 1 | abundance | SL00502 | 2-Ketobuty | Tibia_length | 0,03935 | 0,051689 | NA |
| | 1 | abundance | SL00502 | 2-Ketobuty | Radius_length | -0,05217 | 0,070893 | NA |
| | 1 | abundance | SL00502 | 2-Ketobuty | Radius_SOS | 0,020005 | 0,049864 | NA |
| | 1 | abundance | SL00502 | 2-Ketobuty | Tibia_SOS | 0,064163 | 0,054088 | NA |
| | 1 | abundance | SL00502 | 2-Ketobuty | Handgrip | 0,03979 | 0,061062 | NA |
| 2a | | abundance | X00011 | Glutamine | Tibia_length | 0,083273 | 0,052176 | NA |
| 2a | | abundance | X00011 | Glutamine | Radius_length | 0,110833 | 0,070745 | NA |
| 2a | | abundance | X00011 | Glutamine | Radius_SOS | -0,00756 | 0,050049 | NA |
| 2a | | abundance | X00011 | Glutamine | Tibia_SOS | 0,135753 | 0,052907 | NA |
| 2a | | abundance | X00011 | Glutamine | Handgrip | 0,031328 | 0,060691 | NA |
| | 1 | abundance | X00018 | Succinic ac | Tibia_length | 0,038165 | 0,055819 | NA |
| | 1 | abundance | X00018 | Succinic ac | Radius_length | 0,033086 | 0,074017 | NA |
| | 1 | abundance | X00018 | Succinic ac | Radius_SOS | 0,035122 | 0,052991 | NA |
| | 1 | abundance | X00018 | Succinic ac | Tibia_SOS | -0,03772 | 0,055985 | NA |
| | 1 | abundance | X00018 | Succinic ac | Handgrip | 0,128902 | 0,061474 | NA |
| | 1 | abundance | X00020 | sugar alcol | Tibia_length | -0,06732 | 0,057716 | NA |
| | 1 | abundance | X00020 | sugar alcol | Radius_length | -0,06583 | 0,075423 | NA |
| | 1 | abundance | X00020 | sugar alcol | Radius_SOS | 0,032338 | 0,054422 | NA |
| | 1 | abundance | X00020 | sugar alcol | Tibia_SOS | -0,02001 | 0,057789 | NA |
| | 1 | abundance | X00020 | sugar alcol | Handgrip | 0,057451 | 0,062899 | NA |
| | 1 | abundance | X00022 | N-Methylas | Tibia_length | 0,11706 | 0,051278 | NA |
| | 1 | abundance | X00022 | N-Methylas | Radius_length | 0,222258 | 0,068789 | NA |
| | 1 | abundance | X00022 | N-Methylas | Radius_SOS | 0,048927 | 0,049519 | NA |
| | 1 | abundance | X00022 | N-Methylas | Tibia_SOS | 0,175979 | 0,052228 | NA |
| | 1 | abundance | X00022 | N-Methylas | Handgrip | 0,07412 | 0,060424 | NA |
| | 1 | abundance | X00023 | N-Formylm | Tibia_length | -0,06421 | 0,050756 | NA |
| | 1 | abundance | X00023 | N-Formylm | Radius_length | -0,09939 | 0,069918 | NA |
| | 1 | abundance | X00023 | N-Formylm | Radius_SOS | -0,03467 | 0,049376 | NA |
| | 1 | abundance | X00023 | N-Formylm | Tibia_SOS | -0,04874 | 0,052805 | NA |
| | 1 | abundance | X00023 | N-Formylm | Handgrip | -0,14638 | 0,059744 | NA |
| | 1 | abundance | X00025 | Acesulfam | Tibia_length | -0,05025 | 0,052713 | NA |
| | 1 | abundance | X00025 | Acesulfam | Radius_length | -0,14053 | 0,071539 | NA |
| | 1 | abundance | X00025 | Acesulfam | Radius_SOS | 0,034353 | 0,050389 | NA |
| | 1 | abundance | X00025 | Acesulfam | Tibia_SOS | -0,01141 | 0,054851 | NA |
| | 1 | abundance | X00025 | Acesulfam | Handgrip | -0,04856 | 0,061501 | NA |
| | 1 | abundance | X00026 | Inosine | Tibia_length | -0,0319 | 0,055859 | NA |
| | 1 | abundance | X00026 | Inosine | Radius_length | -0,06449 | 0,074004 | NA |
| | 1 | abundance | X00026 | Inosine | Radius_SOS | -0,11461 | 0,052857 | NA |
| | 1 | abundance | X00026 | Inosine | Tibia_SOS | 0,055462 | 0,056122 | NA |
| | 1 | abundance | X00026 | Inosine | Handgrip | 0,028778 | 0,062392 | NA |
| | 1 | abundance | X00027 | N-Acetylglu | Tibia_length | 0,147758 | 0,052138 | NA |
| | 1 | abundance | X00027 | N-Acetylglu | Radius_length | 0,194949 | 0,07007 | NA |
| | 1 | abundance | X00027 | N-Acetylglu | Radius_SOS | 0,029917 | 0,05017 | NA |
| | 1 | abundance | X00027 | N-Acetylglu | Tibia_SOS | 0,071167 | 0,052869 | NA |
| | 1 | abundance | X00027 | N-Acetylglu | Handgrip | 0,008762 | 0,060524 | NA |
| | 1 | abundance | X00029 | Urocanic a | Tibia_length | 0,006555 | 0,053465 | NA |
| | 1 | abundance | X00029 | Urocanic a | Radius_length | -0,10891 | 0,072467 | NA |

| | | | | | | | |
|---|---|---|---|---|---|---|---|
| 1 | abundance | X00029 | Urocanic a | Radius_SOS | -4,36E-04 | 0,051064 | NA |
| 1 | abundance | X00029 | Urocanic a | Tibia_SOS | 0,11451 | 0,055037 | NA |
| 1 | abundance | X00029 | Urocanic a | Handgrip | 0,058963 | 0,063068 | NA |
| 1 | abundance | X00030 | 3,4-Dihydr | Tibia_length | -0,1607 | 0,053077 | NA |
| 1 | abundance | X00030 | 3,4-Dihydr | Radius_length | -0,0642 | 0,072364 | NA |
| 1 | abundance | X00030 | 3,4-Dihydr | Radius_SOS | 0,006696 | 0,051007 | NA |
| 1 | abundance | X00030 | 3,4-Dihydr | Tibia_SOS | -0,05225 | 0,054194 | NA |
| 1 | abundance | X00030 | 3,4-Dihydr | Handgrip | 0,042196 | 0,062511 | NA |
| 1 | abundance | X00032 | Indole-3-pı | Tibia_length | 0,022544 | 0,054603 | NA |
| 1 | abundance | X00032 | Indole-3-pı | Radius_length | 0,071272 | 0,073134 | NA |
| 1 | abundance | X00032 | Indole-3-pı | Radius_SOS | 0,040054 | 0,051888 | NA |
| 1 | abundance | X00032 | Indole-3-pı | Tibia_SOS | 0,061372 | 0,055305 | NA |
| 1 | abundance | X00032 | Indole-3-pı | Handgrip | 0,121808 | 0,062633 | NA |
| 1 | abundance | X00033 | Deoxychol | Tibia_length | 0,026075 | 0,052178 | NA |
| 1 | abundance | X00033 | Deoxychol | Radius_length | 0,018066 | 0,071483 | NA |
| 1 | abundance | X00033 | Deoxychol | Radius_SOS | 0,097919 | 0,050044 | NA |
| 1 | abundance | X00033 | Deoxychol | Tibia_SOS | -0,00499 | 0,053605 | NA |
| 1 | abundance | X00033 | Deoxychol | Handgrip | 0,02805 | 0,060772 | NA |
| 1 | abundance | X00034 | Pyroglutam | Tibia_length | 0,11027 | 0,053098 | NA |
| 1 | abundance | X00034 | Pyroglutam | Radius_length | 0,117044 | 0,072026 | NA |
| 1 | abundance | X00034 | Pyroglutam | Radius_SOS | 0,042281 | 0,050878 | NA |
| 1 | abundance | X00034 | Pyroglutam | Tibia_SOS | 0,076614 | 0,054777 | NA |
| 1 | abundance | X00034 | Pyroglutam | Handgrip | 0,083689 | 0,060364 | NA |
| 1 | abundance | X00035 | Choline | Tibia_length | -0,06916 | 0,054051 | NA |
| 1 | abundance | X00035 | Choline | Radius_length | -0,10685 | 0,072629 | NA |
| 1 | abundance | X00035 | Choline | Radius_SOS | -0,09246 | 0,052011 | NA |
| 1 | abundance | X00035 | Choline | Tibia_SOS | -0,03166 | 0,055196 | NA |
| 1 | abundance | X00035 | Choline | Handgrip | -0,00449 | 0,062191 | NA |
| 1 | abundance | X00036 | Serine | Tibia_length | 0,025488 | 0,051632 | NA |
| 1 | abundance | X00036 | Serine | Radius_length | 0,066225 | 0,070411 | NA |
| 1 | abundance | X00036 | Serine | Radius_SOS | -0,03455 | 0,049798 | NA |
| 1 | abundance | X00036 | Serine | Tibia_SOS | 0,072152 | 0,053184 | NA |
| 1 | abundance | X00036 | Serine | Handgrip | 0,113664 | 0,061365 | NA |
| 1 | abundance | X00038 | Histamine | Tibia_length | -0,1072 | 0,048821 | NA |
| 1 | abundance | X00038 | Histamine | Radius_length | -0,15892 | 0,064049 | NA |
| 1 | abundance | X00038 | Histamine | Radius_SOS | -0,0234 | 0,049277 | NA |
| 1 | abundance | X00038 | Histamine | Tibia_SOS | -0,1336 | 0,052202 | NA |
| 1 | abundance | X00038 | Histamine | Handgrip | 0,026288 | 0,060319 | NA |
| 1 | abundance | X00040 | N-Acetyltyı | Tibia_length | 0,091833 | 0,052209 | NA |
| 1 | abundance | X00040 | N-Acetyltyı | Radius_length | 0,097689 | 0,071129 | NA |
| 1 | abundance | X00040 | N-Acetyltyı | Radius_SOS | 0,08767 | 0,050018 | NA |
| 1 | abundance | X00040 | N-Acetyltyı | Tibia_SOS | 0,083996 | 0,053246 | NA |
| 1 | abundance | X00040 | N-Acetyltyı | Handgrip | -0,03688 | 0,060931 | NA |
| 1 | abundance | X00042 | Glucuronic | Tibia_length | -0,0122 | 0,056257 | NA |
| 1 | abundance | X00042 | Glucuronic | Radius_length | -0,0165 | 0,075064 | NA |
| 1 | abundance | X00042 | Glucuronic | Radius_SOS | -0,04844 | 0,053305 | NA |
| 1 | abundance | X00042 | Glucuronic | Tibia_SOS | 0,033226 | 0,056363 | NA |
| 1 | abundance | X00042 | Glucuronic | Handgrip | 0,130063 | 0,06313 | NA |

| | | | | | | | |
|---|---|---|---|---|---|---|---|
| 1 | abundance | X00045 | Threonine | Tibia_length | 0,073615 | 0,051296 | NA |
| 1 | abundance | X00045 | Threonine | Radius_length | 0,115764 | 0,069903 | NA |
| 1 | abundance | X00045 | Threonine | Radius_SOS | -0,02265 | 0,049618 | NA |
| 1 | abundance | X00045 | Threonine | Tibia_SOS | 0,078008 | 0,052968 | NA |
| 1 | abundance | X00045 | Threonine | Handgrip | 0,106462 | 0,060346 | NA |
| 1 | abundance | X00050 | Î²-Hydroxyi | Tibia_length | -0,06128 | 0,053066 | NA |
| 1 | abundance | X00050 | Î²-Hydroxyi | Radius_length | 4,85E-04 | 0,072149 | NA |
| 1 | abundance | X00050 | Î²-Hydroxyi | Radius_SOS | -0,05394 | 0,050806 | NA |
| 1 | abundance | X00050 | Î²-Hydroxyi | Tibia_SOS | 0,026945 | 0,054199 | NA |
| 1 | abundance | X00050 | Î²-Hydroxyi | Handgrip | 0,044248 | 0,061738 | NA |
| 2b | abundance | X00051 | Î²-D-Gluco | Tibia_length | 0,015004 | 0,057117 | NA |
| 2b | abundance | X00051 | Î²-D-Gluco | Radius_length | 0,014331 | 0,07545 | NA |
| 2b | abundance | X00051 | Î²-D-Gluco | Radius_SOS | -0,00603 | 0,054218 | NA |
| 2b | abundance | X00051 | Î²-D-Gluco | Tibia_SOS | 0,058796 | 0,05838 | NA |
| 2b | abundance | X00051 | Î²-D-Gluco | Handgrip | 0,131312 | 0,059672 | NA |
| 1 | abundance | X00056 | 3-(4-hydro: | Tibia_length | 0,032282 | 0,053616 | NA |
| 1 | abundance | X00056 | 3-(4-hydro: | Radius_length | -0,04725 | 0,072741 | NA |
| 1 | abundance | X00056 | 3-(4-hydro: | Radius_SOS | 0,062286 | 0,050953 | NA |
| 1 | abundance | X00056 | 3-(4-hydro: | Tibia_SOS | 0,007527 | 0,05602 | NA |
| 1 | abundance | X00056 | 3-(4-hydro: | Handgrip | 0,052843 | 0,062083 | NA |
| 1 | abundance | X00057 | N-Acetylor | Tibia_length | 6,97E-04 | 0,054207 | NA |
| 1 | abundance | X00057 | N-Acetylor | Radius_length | 0,050254 | 0,073263 | NA |
| 1 | abundance | X00057 | N-Acetylor | Radius_SOS | 5,99E-04 | 0,051644 | NA |
| 1 | abundance | X00057 | N-Acetylor | Tibia_SOS | 0,040211 | 0,055159 | NA |
| 1 | abundance | X00057 | N-Acetylor | Handgrip | -0,04115 | 0,061502 | NA |
| 1 | abundance | X00059 | 3,4-Dihydr | Tibia_length | -0,14414 | 0,054442 | NA |
| 1 | abundance | X00059 | 3,4-Dihydr | Radius_length | -0,04117 | 0,073707 | NA |
| 1 | abundance | X00059 | 3,4-Dihydr | Radius_SOS | -0,0563 | 0,052099 | NA |
| 1 | abundance | X00059 | 3,4-Dihydr | Tibia_SOS | -0,10612 | 0,055135 | NA |
| 1 | abundance | X00059 | 3,4-Dihydr | Handgrip | 0,108751 | 0,060776 | NA |
| 1 | abundance | X00060 | Ethylmalor | Tibia_length | -0,00341 | 0,05471 | NA |
| 1 | abundance | X00060 | Ethylmalor | Radius_length | 0,035117 | 0,073244 | NA |
| 1 | abundance | X00060 | Ethylmalor | Radius_SOS | 0,068698 | 0,051877 | NA |
| 1 | abundance | X00060 | Ethylmalor | Tibia_SOS | -0,06361 | 0,054857 | NA |
| 1 | abundance | X00060 | Ethylmalor | Handgrip | -0,05711 | 0,062465 | NA |
| 1 | abundance | X00061 | Thiamine | Tibia_length | -0,00149 | 0,050104 | NA |
| 1 | abundance | X00061 | Thiamine | Radius_length | -0,04162 | 0,068256 | NA |
| 1 | abundance | X00061 | Thiamine | Radius_SOS | -0,0669 | 0,049016 | NA |
| 1 | abundance | X00061 | Thiamine | Tibia_SOS | 0,058668 | 0,052775 | NA |
| 1 | abundance | X00061 | Thiamine | Handgrip | -0,05388 | 0,060808 | NA |
| 1 | abundance | X00063 | 2-Hydroxy( | Tibia_length | 0,10413 | 0,053997 | NA |
| 1 | abundance | X00063 | 2-Hydroxy( | Radius_length | 0,138514 | 0,072888 | NA |
| 1 | abundance | X00063 | 2-Hydroxy( | Radius_SOS | 0,068549 | 0,05127 | NA |
| 1 | abundance | X00063 | 2-Hydroxy( | Tibia_SOS | 0,071645 | 0,055668 | NA |
| 1 | abundance | X00063 | 2-Hydroxy( | Handgrip | 0,188187 | 0,060442 | NA |
| 1 | abundance | X00064 | Traumatic | Tibia_length | 0,066096 | 0,053842 | NA |
| 1 | abundance | X00064 | Traumatic | Radius_length | 0,074945 | 0,072847 | NA |
| 1 | abundance | X00064 | Traumatic | Radius_SOS | 0,068049 | 0,051362 | NA |

| | | | | | | | |
|---|---|---|---|---|---|---|---|
| 1 | abundance | X00064 | Traumatic | Tibia_SOS | 0,078011 | 0,054044 | NA |
| 1 | abundance | X00064 | Traumatic | Handgrip | -0,00349 | 0,061674 | NA |
| 1 | abundance | X00066 | Tryptamine | Tibia_length | 0,037466 | 0,049441 | NA |
| 1 | abundance | X00066 | Tryptamine | Radius_length | -0,00447 | 0,06675 | NA |
| 1 | abundance | X00066 | Tryptamine | Radius_SOS | -0,04949 | 0,048979 | NA |
| 1 | abundance | X00066 | Tryptamine | Tibia_SOS | -0,02475 | 0,052496 | NA |
| 1 | abundance | X00066 | Tryptamine | Handgrip | -0,0917 | 0,059963 | NA |
| 1 | abundance | X00067 | Hypoxanth | Tibia_length | -0,09717 | 0,055088 | NA |
| 1 | abundance | X00067 | Hypoxanth | Radius_length | -0,00965 | 0,074436 | NA |
| 1 | abundance | X00067 | Hypoxanth | Radius_SOS | -0,07046 | 0,052641 | NA |
| 1 | abundance | X00067 | Hypoxanth | Tibia_SOS | 0,083143 | 0,055658 | NA |
| 1 | abundance | X00067 | Hypoxanth | Handgrip | 0,092361 | 0,062811 | NA |
| 1 | abundance | X00068 | N-Acetylm | Tibia_length | 0,1419 | 0,050285 | NA |
| 1 | abundance | X00068 | N-Acetylm | Radius_length | 0,160506 | 0,068082 | NA |
| 1 | abundance | X00068 | N-Acetylm | Radius_SOS | 0,032645 | 0,049194 | NA |
| 1 | abundance | X00068 | N-Acetylm | Tibia_SOS | 0,10788 | 0,052492 | NA |
| 1 | abundance | X00068 | N-Acetylm | Handgrip | 0,068761 | 0,060121 | NA |
| 1 | abundance | X00070 | Î±-aminobu | Tibia_length | -0,15121 | 0,051843 | NA |
| 1 | abundance | X00070 | Î±-aminobu | Radius_length | -0,12023 | 0,071261 | NA |
| 1 | abundance | X00070 | Î±-aminobu | Radius_SOS | -0,03729 | 0,050003 | NA |
| 1 | abundance | X00070 | Î±-aminobu | Tibia_SOS | -0,06254 | 0,053376 | NA |
| 1 | abundance | X00070 | Î±-aminobu | Handgrip | 0,005603 | 0,060405 | NA |
| 1 | abundance | X00071 | Tyrosine | Tibia_length | 0,172024 | 0,048699 | NA |
| 1 | abundance | X00071 | Tyrosine | Radius_length | 0,14535 | 0,065646 | NA |
| 1 | abundance | X00071 | Tyrosine | Radius_SOS | 0,059246 | 0,048959 | NA |
| 1 | abundance | X00071 | Tyrosine | Tibia_SOS | 0,103179 | 0,052254 | NA |
| 1 | abundance | X00071 | Tyrosine | Handgrip | 0,035829 | 0,060163 | NA |
| 1 | abundance | X00072 | 3,5-Dihydr | Tibia_length | -0,10341 | 0,054833 | NA |
| 1 | abundance | X00072 | 3,5-Dihydr | Radius_length | -0,05023 | 0,07357 | NA |
| 1 | abundance | X00072 | 3,5-Dihydr | Radius_SOS | -0,09287 | 0,052411 | NA |
| 1 | abundance | X00072 | 3,5-Dihydr | Tibia_SOS | -0,03743 | 0,055137 | NA |
| 1 | abundance | X00072 | 3,5-Dihydr | Handgrip | -0,01015 | 0,062739 | NA |
| 1 | abundance | X00073 | Indole-3-la | Tibia_length | 0,098404 | 0,05272 | NA |
| 1 | abundance | X00073 | Indole-3-la | Radius_length | 0,173687 | 0,071209 | NA |
| 1 | abundance | X00073 | Indole-3-la | Radius_SOS | 0,064745 | 0,050193 | NA |
| 1 | abundance | X00073 | Indole-3-la | Tibia_SOS | 0,095293 | 0,054171 | NA |
| 1 | abundance | X00073 | Indole-3-la | Handgrip | 0,235417 | 0,059314 | NA |
| 1 | abundance | X00074 | Methylsuc | Tibia_length | 0,043367 | 0,051023 | NA |
| 1 | abundance | X00074 | Methylsuc | Radius_length | 0,006046 | 0,069757 | NA |
| 1 | abundance | X00074 | Methylsuc | Radius_SOS | 0,052284 | 0,049369 | NA |
| 1 | abundance | X00074 | Methylsuc | Tibia_SOS | 0,059673 | 0,052746 | NA |
| 1 | abundance | X00074 | Methylsuc | Handgrip | 0,035955 | 0,060554 | NA |
| 1 | abundance | X00076 | Isoleucine | Tibia_length | 0,119449 | 0,050168 | NA |
| 1 | abundance | X00076 | Isoleucine | Radius_length | 0,147209 | 0,068194 | NA |
| 1 | abundance | X00076 | Isoleucine | Radius_SOS | 0,011468 | 0,049183 | NA |
| 1 | abundance | X00076 | Isoleucine | Tibia_SOS | 0,144782 | 0,052136 | NA |
| 1 | abundance | X00076 | Isoleucine | Handgrip | 0,078352 | 0,060086 | NA |
| 1 | abundance | X00078 | Acetylagm | Tibia_length | 8,37E-04 | 0,050501 | NA |

| | | | | | | | |
|---|---|---|---|---|---|---|---|
| 1 | abundance | X00078 | Acetylagm | Radius_length | -0,06439 | 0,069172 | NA |
| 1 | abundance | X00078 | Acetylagm | Radius_SOS | -0,08031 | 0,049096 | NA |
| 1 | abundance | X00078 | Acetylagm | Tibia_SOS | -0,05181 | 0,052875 | NA |
| 1 | abundance | X00078 | Acetylagm | Handgrip | 0,011341 | 0,060706 | NA |
| 1 | abundance | X00082 | Cholic acid | Tibia_length | -4,40E-05 | 0,050032 | NA |
| 1 | abundance | X00082 | Cholic acid | Radius_length | 0,060456 | 0,068122 | NA |
| 1 | abundance | X00082 | Cholic acid | Radius_SOS | -0,0068 | 0,049099 | NA |
| 1 | abundance | X00082 | Cholic acid | Tibia_SOS | -0,06909 | 0,052469 | NA |
| 1 | abundance | X00082 | Cholic acid | Handgrip | 0,026596 | 0,060755 | NA |
| 1 | abundance | X00083 | 1,7-Dimeth | Tibia_length | -0,06132 | 0,050524 | NA |
| 1 | abundance | X00083 | 1,7-Dimeth | Radius_length | -0,07951 | 0,069018 | NA |
| 1 | abundance | X00083 | 1,7-Dimeth | Radius_SOS | 0,027533 | 0,049243 | NA |
| 1 | abundance | X00083 | 1,7-Dimeth | Tibia_SOS | -0,05201 | 0,05243 | NA |
| 1 | abundance | X00083 | 1,7-Dimeth | Handgrip | 0,005264 | 0,060352 | NA |
| 1 | abundance | X00084 | Tricarballyl | Tibia_length | -0,03418 | 0,053194 | NA |
| 1 | abundance | X00084 | Tricarballyl | Radius_length | -0,08495 | 0,071801 | NA |
| 1 | abundance | X00084 | Tricarballyl | Radius_SOS | -0,02668 | 0,050817 | NA |
| 1 | abundance | X00084 | Tricarballyl | Tibia_SOS | -0,08528 | 0,05355 | NA |
| 1 | abundance | X00084 | Tricarballyl | Handgrip | -0,02332 | 0,061718 | NA |
| 1 | abundance | X00088 | Pantotheni | Tibia_length | -0,05206 | 0,055753 | NA |
| 1 | abundance | X00088 | Pantotheni | Radius_length | -0,11026 | 0,07357 | NA |
| 1 | abundance | X00088 | Pantotheni | Radius_SOS | -0,07851 | 0,053467 | NA |
| 1 | abundance | X00088 | Pantotheni | Tibia_SOS | -0,00797 | 0,056245 | NA |
| 1 | abundance | X00088 | Pantotheni | Handgrip | -0,02402 | 0,062171 | NA |
| 1 | abundance | X00089 | 4-Hydroxyb | Tibia_length | -0,0299 | 0,052431 | NA |
| 1 | abundance | X00089 | 4-Hydroxyb | Radius_length | -0,02723 | 0,071688 | NA |
| 1 | abundance | X00089 | 4-Hydroxyb | Radius_SOS | -0,06606 | 0,050249 | NA |
| 1 | abundance | X00089 | 4-Hydroxyb | Tibia_SOS | 0,029211 | 0,053043 | NA |
| 1 | abundance | X00089 | 4-Hydroxyb | Handgrip | 0,206846 | 0,059743 | NA |
| 1 | abundance | X00090 | Malic acid | Tibia_length | -0,02968 | 0,053279 | NA |
| 1 | abundance | X00090 | Malic acid | Radius_length | -0,11538 | 0,07229 | NA |
| 1 | abundance | X00090 | Malic acid | Radius_SOS | -0,05631 | 0,050932 | NA |
| 1 | abundance | X00090 | Malic acid | Tibia_SOS | 0,031112 | 0,054603 | NA |
| 1 | abundance | X00090 | Malic acid | Handgrip | 0,095452 | 0,062647 | NA |
| 1 | abundance | X00092 | 3-Methylhi | Tibia_length | -0,1028 | 0,051248 | NA |
| 1 | abundance | X00092 | 3-Methylhi | Radius_length | -0,01893 | 0,070533 | NA |
| 1 | abundance | X00092 | 3-Methylhi | Radius_SOS | 4,80E-04 | 0,049755 | NA |
| 1 | abundance | X00092 | 3-Methylhi | Tibia_SOS | 0,035759 | 0,053293 | NA |
| 1 | abundance | X00092 | 3-Methylhi | Handgrip | -0,00218 | 0,061297 | NA |
| 1 | abundance | X00093 | Deoxyinosi | Tibia_length | 0,016247 | 0,05183 | NA |
| 1 | abundance | X00093 | Deoxyinosi | Radius_length | 0,05206 | 0,070715 | NA |
| 1 | abundance | X00093 | Deoxyinosi | Radius_SOS | -0,06955 | 0,049762 | NA |
| 1 | abundance | X00093 | Deoxyinosi | Tibia_SOS | 0,04137 | 0,053875 | NA |
| 1 | abundance | X00093 | Deoxyinosi | Handgrip | 0,100349 | 0,061312 | NA |
| 1 | abundance | X00094 | Methionine | Tibia_length | 0,101042 | 0,050688 | NA |
| 1 | abundance | X00094 | Methionine | Radius_length | 0,179148 | 0,068276 | NA |
| 1 | abundance | X00094 | Methionine | Radius_SOS | 0,023745 | 0,049308 | NA |
| 1 | abundance | X00094 | Methionine | Tibia_SOS | 0,135725 | 0,052383 | NA |

| | | | | | | | |
|---|---|---|---|---|---|---|---|
| 1 | abundance | X00094 | Methionine | Handgrip | 0,111918 | 0,060024 | NA |
| 1 | abundance | X00097 | 2,6-Dihydr | Tibia_length | -0,11345 | 0,050963 | NA |
| 1 | abundance | X00097 | 2,6-Dihydr | Radius_length | -0,09682 | 0,070277 | NA |
| 1 | abundance | X00097 | 2,6-Dihydr | Radius_SOS | -0,0329 | 0,049594 | NA |
| 1 | abundance | X00097 | 2,6-Dihydr | Tibia_SOS | -0,07214 | 0,053107 | NA |
| 1 | abundance | X00097 | 2,6-Dihydr | Handgrip | 0,003524 | 0,060687 | NA |
| 1 | abundance | X00099 | 7-Methylgu | Tibia_length | -0,10301 | 0,051628 | NA |
| 1 | abundance | X00099 | 7-Methylgu | Radius_length | -0,11516 | 0,070706 | NA |
| 1 | abundance | X00099 | 7-Methylgu | Radius_SOS | -0,04702 | 0,050057 | NA |
| 1 | abundance | X00099 | 7-Methylgu | Tibia_SOS | -0,00342 | 0,053679 | NA |
| 1 | abundance | X00099 | 7-Methylgu | Handgrip | -0,02793 | 0,062416 | NA |
| 1 | abundance | X00101 | Threonic ac | Tibia_length | 0,019597 | 0,052245 | NA |
| 1 | abundance | X00101 | Threonic ac | Radius_length | 0,072193 | 0,0716 | NA |
| 1 | abundance | X00101 | Threonic ac | Radius_SOS | 0,051129 | 0,050066 | NA |
| 1 | abundance | X00101 | Threonic ac | Tibia_SOS | 0,044327 | 0,053714 | NA |
| 1 | abundance | X00101 | Threonic ac | Handgrip | 0,096946 | 0,060492 | NA |
| 1 | abundance | X00102 | Tryptophar | Tibia_length | 0,048205 | 0,052881 | NA |
| 1 | abundance | X00102 | Tryptophar | Radius_length | 0,111003 | 0,071996 | NA |
| 1 | abundance | X00102 | Tryptophar | Radius_SOS | -0,03289 | 0,050646 | NA |
| 1 | abundance | X00102 | Tryptophar | Tibia_SOS | 0,097685 | 0,053712 | NA |
| 1 | abundance | X00102 | Tryptophar | Handgrip | 0,029409 | 0,060659 | NA |
| 1 | abundance | X00106 | N-Acetyltry | Tibia_length | -0,02033 | 0,0571 | NA |
| 1 | abundance | X00106 | N-Acetyltry | Radius_length | -0,0523 | 0,075209 | NA |
| 1 | abundance | X00106 | N-Acetyltry | Radius_SOS | -0,0282 | 0,05429 | NA |
| 1 | abundance | X00106 | N-Acetyltry | Tibia_SOS | 0,015417 | 0,05706 | NA |
| 1 | abundance | X00106 | N-Acetyltry | Handgrip | -0,01297 | 0,063354 | NA |
| 1 | abundance | X00107 | N6-Acetylly | Tibia_length | 0,111332 | 0,051816 | NA |
| 1 | abundance | X00107 | N6-Acetylly | Radius_length | 0,184654 | 0,069829 | NA |
| 1 | abundance | X00107 | N6-Acetylly | Radius_SOS | 0,096608 | 0,049793 | NA |
| 1 | abundance | X00107 | N6-Acetylly | Tibia_SOS | 0,11166 | 0,053278 | NA |
| 1 | abundance | X00107 | N6-Acetylly | Handgrip | 0,059336 | 0,060861 | NA |
| 1 | abundance | X00112 | Xanthosine | Tibia_length | 0,00759 | 0,053547 | NA |
| 1 | abundance | X00112 | Xanthosine | Radius_length | -0,07258 | 0,072597 | NA |
| 1 | abundance | X00112 | Xanthosine | Radius_SOS | -0,06023 | 0,051009 | NA |
| 1 | abundance | X00112 | Xanthosine | Tibia_SOS | 0,093021 | 0,054142 | NA |
| 1 | abundance | X00112 | Xanthosine | Handgrip | -0,00524 | 0,062529 | NA |
| 1 | abundance | X00113 | N-Acetylph | Tibia_length | 0,007381 | 0,055505 | NA |
| 1 | abundance | X00113 | N-Acetylph | Radius_length | -0,05036 | 0,07382 | NA |
| 1 | abundance | X00113 | N-Acetylph | Radius_SOS | 0,03872 | 0,052743 | NA |
| 1 | abundance | X00113 | N-Acetylph | Tibia_SOS | 0,066098 | 0,05644 | NA |
| 1 | abundance | X00113 | N-Acetylph | Handgrip | -0,03472 | 0,063526 | NA |
| 1 | abundance | X00114 | Indole-3-m | Tibia_length | 0,102198 | 0,056578 | NA |
| 1 | abundance | X00114 | Indole-3-m | Radius_length | 0,08891 | 0,074333 | NA |
| 1 | abundance | X00114 | Indole-3-m | Radius_SOS | -0,07217 | 0,054148 | NA |
| 1 | abundance | X00114 | Indole-3-m | Tibia_SOS | 0,13764 | 0,056497 | NA |
| 1 | abundance | X00114 | Indole-3-m | Handgrip | 0,149222 | 0,062358 | NA |
| 1 | abundance | X00115 | Histidine | Tibia_length | 0,050284 | 0,049464 | NA |
| 1 | abundance | X00115 | Histidine | Radius_length | 0,239689 | 0,064954 | NA |

| | | | | | | | |
|---|---|---|---|---|---|---|---|
| 1 | abundance | X00115 | Histidine | Radius_SOS | 0,022313 | 0,049022 | NA |
| 1 | abundance | X00115 | Histidine | Tibia_SOS | 0,107976 | 0,05218 | NA |
| 1 | abundance | X00115 | Histidine | Handgrip | 0,103889 | 0,059867 | NA |
| 1 | abundance | X00116 | Cytidine | Tibia_length | -0,04349 | 0,054901 | NA |
| 1 | abundance | X00116 | Cytidine | Radius_length | 0,043435 | 0,073282 | NA |
| 1 | abundance | X00116 | Cytidine | Radius_SOS | -0,08906 | 0,052302 | NA |
| 1 | abundance | X00116 | Cytidine | Tibia_SOS | 0,010959 | 0,056053 | NA |
| 1 | abundance | X00116 | Cytidine | Handgrip | -0,05042 | 0,062611 | NA |
| 1 | abundance | X00120 | Leucylalan | Tibia_length | -0,03293 | 0,051473 | NA |
| 1 | abundance | X00120 | Leucylalan | Radius_length | 0,026938 | 0,070239 | NA |
| 1 | abundance | X00120 | Leucylalan | Radius_SOS | 0,029321 | 0,04962 | NA |
| 1 | abundance | X00120 | Leucylalan | Tibia_SOS | -0,09873 | 0,052795 | NA |
| 1 | abundance | X00120 | Leucylalan | Handgrip | 0,014154 | 0,060649 | NA |
| 1 | abundance | X00123 | Phenylalar | Tibia_length | 0,115299 | 0,050203 | NA |
| 1 | abundance | X00123 | Phenylalar | Radius_length | 0,112802 | 0,068561 | NA |
| 1 | abundance | X00123 | Phenylalar | Radius_SOS | -0,00723 | 0,049226 | NA |
| 1 | abundance | X00123 | Phenylalar | Tibia_SOS | 0,107552 | 0,052366 | NA |
| 1 | abundance | X00123 | Phenylalar | Handgrip | 0,051184 | 0,060146 | NA |
| 1 | abundance | X00124 | Leucine | Tibia_length | 0,1011 | 0,050415 | NA |
| 1 | abundance | X00124 | Leucine | Radius_length | 0,145916 | 0,068538 | NA |
| 1 | abundance | X00124 | Leucine | Radius_SOS | 0,002545 | 0,049262 | NA |
| 1 | abundance | X00124 | Leucine | Tibia_SOS | 0,126055 | 0,052369 | NA |
| 1 | abundance | X00124 | Leucine | Handgrip | 0,087534 | 0,060072 | NA |
| 1 | abundance | X00125 | Taurine | Tibia_length | -0,0376 | 0,053881 | NA |
| 1 | abundance | X00125 | Taurine | Radius_length | -0,10023 | 0,07311 | NA |
| 1 | abundance | X00125 | Taurine | Radius_SOS | -0,04897 | 0,051743 | NA |
| 1 | abundance | X00125 | Taurine | Tibia_SOS | -0,01511 | 0,055505 | NA |
| 1 | abundance | X00125 | Taurine | Handgrip | 0,036284 | 0,062625 | NA |
| 1 | abundance | X00127 | Nicotinic a | Tibia_length | -0,01567 | 0,056093 | NA |
| 1 | abundance | X00127 | Nicotinic a | Radius_length | 0,044877 | 0,074507 | NA |
| 1 | abundance | X00127 | Nicotinic a | Radius_SOS | -0,02251 | 0,053611 | NA |
| 1 | abundance | X00127 | Nicotinic a | Tibia_SOS | 0,079163 | 0,056208 | NA |
| 1 | abundance | X00127 | Nicotinic a | Handgrip | 0,08161 | 0,062995 | NA |
| 1 | abundance | X00129 | Quinic acic | Tibia_length | -0,03666 | 0,052648 | NA |
| 1 | abundance | X00129 | Quinic acic | Radius_length | -0,08101 | 0,071541 | NA |
| 1 | abundance | X00129 | Quinic acic | Radius_SOS | -0,10599 | 0,050257 | NA |
| 1 | abundance | X00129 | Quinic acic | Tibia_SOS | -0,17045 | 0,054759 | NA |
| 1 | abundance | X00129 | Quinic acic | Handgrip | -0,05344 | 0,060936 | NA |
| 1 | abundance | X00130 | Deoxyuridi | Tibia_length | 0,027841 | 0,050955 | NA |
| 1 | abundance | X00130 | Deoxyuridi | Radius_length | 0,082951 | 0,069456 | NA |
| 1 | abundance | X00130 | Deoxyuridi | Radius_SOS | -0,07317 | 0,049282 | NA |
| 1 | abundance | X00130 | Deoxyuridi | Tibia_SOS | 0,061594 | 0,05265 | NA |
| 1 | abundance | X00130 | Deoxyuridi | Handgrip | 0,041636 | 0,060709 | NA |
| 1 | abundance | X00132 | Glyceric ac | Tibia_length | 0,020323 | 0,052569 | NA |
| 1 | abundance | X00132 | Glyceric ac | Radius_length | 0,068423 | 0,071798 | NA |
| 1 | abundance | X00132 | Glyceric ac | Radius_SOS | 8,13E-04 | 0,050349 | NA |
| 1 | abundance | X00132 | Glyceric ac | Tibia_SOS | 0,033332 | 0,053681 | NA |
| 1 | abundance | X00132 | Glyceric ac | Handgrip | 0,037799 | 0,061178 | NA |

| | | | | | | | |
|---|---|---|---|---|---|---|---|
| 1 | abundance | X00135 | N-Acetylgl | Tibia_length | 0,011064 | 0,052674 | NA |
| 1 | abundance | X00135 | N-Acetylgl | Radius_length | 0,049756 | 0,072064 | NA |
| 1 | abundance | X00135 | N-Acetylgl | Radius_SOS | -0,02453 | 0,050523 | NA |
| 1 | abundance | X00135 | N-Acetylgl | Tibia_SOS | 0,032406 | 0,054054 | NA |
| 1 | abundance | X00135 | N-Acetylgl | Handgrip | 0,073611 | 0,06241 | NA |
| 3 | abundance | X00136 | leu-gln | Tibia_length | 0,223846 | 0,053172 | NA |
| 3 | abundance | X00136 | leu-gln | Radius_length | 0,131445 | 0,072867 | NA |
| 3 | abundance | X00136 | leu-gln | Radius_SOS | 0,086427 | 0,051787 | NA |
| 3 | abundance | X00136 | leu-gln | Tibia_SOS | 0,089745 | 0,055596 | NA |
| 3 | abundance | X00136 | leu-gln | Handgrip | 0,084751 | 0,062051 | NA |
| 3 | abundance | X00163 | primidone | Tibia_length | 0,032576 | 0,053945 | NA |
| 3 | abundance | X00163 | primidone | Radius_length | -0,00464 | 0,07357 | NA |
| 3 | abundance | X00163 | primidone | Radius_SOS | -0,09348 | 0,05118 | NA |
| 3 | abundance | X00163 | primidone | Tibia_SOS | 0,064338 | 0,05533 | NA |
| 3 | abundance | X00163 | primidone | Handgrip | 0,017282 | 0,062033 | NA |
| 3 | abundance | X00180 | 1-(2-Carbo | Tibia_length | -0,12078 | 0,055632 | NA |
| 3 | abundance | X00180 | 1-(2-Carbo | Radius_length | -0,11496 | 0,074171 | NA |
| 3 | abundance | X00180 | 1-(2-Carbo | Radius_SOS | -0,06278 | 0,052977 | NA |
| 3 | abundance | X00180 | 1-(2-Carbo | Tibia_SOS | -0,0845 | 0,055915 | NA |
| 3 | abundance | X00180 | 1-(2-Carbo | Handgrip | -0,13688 | 0,061436 | NA |
| 3 | abundance | X00182 | 3-(Butylsul | Tibia_length | 0,087168 | 0,051884 | NA |
| 3 | abundance | X00182 | 3-(Butylsul | Radius_length | 0,051176 | 0,071615 | NA |
| 3 | abundance | X00182 | 3-(Butylsul | Radius_SOS | 0,034234 | 0,0501 | NA |
| 3 | abundance | X00182 | 3-(Butylsul | Tibia_SOS | -0,01955 | 0,053327 | NA |
| 3 | abundance | X00182 | 3-(Butylsul | Handgrip | -0,18402 | 0,060219 | NA |
| 3 | abundance | X00199 | tert-Butyl 3 | Tibia_length | -0,08159 | 0,054762 | NA |
| 3 | abundance | X00199 | tert-Butyl 3 | Radius_length | -0,12476 | 0,072923 | NA |
| 3 | abundance | X00199 | tert-Butyl 3 | Radius_SOS | -0,06787 | 0,052188 | NA |
| 3 | abundance | X00199 | tert-Butyl 3 | Tibia_SOS | -0,07761 | 0,057096 | NA |
| 3 | abundance | X00199 | tert-Butyl 3 | Handgrip | -0,09088 | 0,062368 | NA |
| 3 | abundance | X00204 | 4-O-{3-O-[ | Tibia_length | -0,01725 | 0,050659 | NA |
| 3 | abundance | X00204 | 4-O-{3-O-[ | Radius_length | 0,119211 | 0,06891 | NA |
| 3 | abundance | X00204 | 4-O-{3-O-[ | Radius_SOS | 0,045785 | 0,049253 | NA |
| 3 | abundance | X00204 | 4-O-{3-O-[ | Tibia_SOS | 0,087712 | 0,052791 | NA |
| 3 | abundance | X00204 | 4-O-{3-O-[ | Handgrip | 0,079784 | 0,060789 | NA |
| 3 | abundance | X00221 | Astemizole | Tibia_length | 0,057138 | 0,049579 | NA |
| 3 | abundance | X00221 | Astemizole | Radius_length | 0,148687 | 0,066342 | NA |
| 3 | abundance | X00221 | Astemizole | Radius_SOS | -0,03799 | 0,048996 | NA |
| 3 | abundance | X00221 | Astemizole | Tibia_SOS | 0,058884 | 0,052407 | NA |
| 3 | abundance | X00221 | Astemizole | Handgrip | 0,033976 | 0,060356 | NA |
| 3 | abundance | X00224 | Nifedipine | Tibia_length | 0,001529 | 0,054104 | NA |
| 3 | abundance | X00224 | Nifedipine | Radius_length | -0,01471 | 0,07323 | NA |
| 3 | abundance | X00224 | Nifedipine | Radius_SOS | -0,02622 | 0,051651 | NA |
| 3 | abundance | X00224 | Nifedipine | Tibia_SOS | 0,003693 | 0,054297 | NA |
| 3 | abundance | X00224 | Nifedipine | Handgrip | -0,08457 | 0,0621 | NA |
| 3 | abundance | X00233 | g-Aminobu | Tibia_length | 0,042452 | 0,053881 | NA |
| 3 | abundance | X00233 | g-Aminobu | Radius_length | 0,05258 | 0,07282 | NA |
| 3 | abundance | X00233 | g-Aminobu | Radius_SOS | -0,00137 | 0,051533 | NA |

| | | | | | | | |
|---|---|---|---|---|---|---|---|
| 3 | abundance | X00233 | g-Aminobu | Tibia_SOS | -0,05241 | 0,054975 | NA |
| 3 | abundance | X00233 | g-Aminobu | Handgrip | 0,077881 | 0,06271 | NA |
| 3 | abundance | X00237 | OI170000( | Tibia_length | 0,004769 | 0,05137 | NA |
| 3 | abundance | X00237 | OI170000( | Radius_length | 0,015747 | 0,070239 | NA |
| 3 | abundance | X00237 | OI170000( | Radius_SOS | 0,047756 | 0,049563 | NA |
| 3 | abundance | X00237 | OI170000( | Tibia_SOS | 0,035211 | 0,053084 | NA |
| 3 | abundance | X00237 | OI170000( | Handgrip | -0,0545 | 0,061014 | NA |
| 3 | abundance | X00242 | IN00258 | Tibia_length | -0,11148 | 0,05608 | NA |
| 3 | abundance | X00242 | IN00258 | Radius_length | 0,028023 | 0,074773 | NA |
| 3 | abundance | X00242 | IN00258 | Radius_SOS | -0,06999 | 0,053748 | NA |
| 3 | abundance | X00242 | IN00258 | Tibia_SOS | 0,063277 | 0,057407 | NA |
| 3 | abundance | X00242 | IN00258 | Handgrip | 0,044024 | 0,063008 | NA |
| 3 | abundance | X00253 | TDP-2 | Tibia_length | -0,05233 | 0,051384 | NA |
| 3 | abundance | X00253 | TDP-2 | Radius_length | 5,15E-04 | 0,070651 | NA |
| 3 | abundance | X00253 | TDP-2 | Radius_SOS | 0,041323 | 0,049833 | NA |
| 3 | abundance | X00253 | TDP-2 | Tibia_SOS | 0,017228 | 0,053028 | NA |
| 3 | abundance | X00253 | TDP-2 | Handgrip | -0,12235 | 0,060337 | NA |
| 3 | abundance | X00261 | Mexiletine | Tibia_length | -0,06713 | 0,055755 | NA |
| 3 | abundance | X00261 | Mexiletine | Radius_length | -0,05035 | 0,073981 | NA |
| 3 | abundance | X00261 | Mexiletine | Radius_SOS | -0,01324 | 0,052874 | NA |
| 3 | abundance | X00261 | Mexiletine | Tibia_SOS | -0,00345 | 0,056799 | NA |
| 3 | abundance | X00261 | Mexiletine | Handgrip | 0,084418 | 0,062105 | NA |
| 2b | abundance | X00264 | PEG n12 | Tibia_length | 0,044173 | 0,05616 | NA |
| 2b | abundance | X00264 | PEG n12 | Radius_length | 0,139112 | 0,073448 | NA |
| 2b | abundance | X00264 | PEG n12 | Radius_SOS | -0,06757 | 0,053452 | NA |
| 2b | abundance | X00264 | PEG n12 | Tibia_SOS | 0,014748 | 0,056222 | NA |
| 2b | abundance | X00264 | PEG n12 | Handgrip | -0,00906 | 0,063332 | NA |
| 3 | abundance | X00266 | 2-Oxo-3-(p | Tibia_length | 0,08667 | 0,054647 | NA |
| 3 | abundance | X00266 | 2-Oxo-3-(p | Radius_length | 0,146981 | 0,07251 | NA |
| 3 | abundance | X00266 | 2-Oxo-3-(p | Radius_SOS | 0,032303 | 0,052125 | NA |
| 3 | abundance | X00266 | 2-Oxo-3-(p | Tibia_SOS | 0,007299 | 0,056268 | NA |
| 3 | abundance | X00266 | 2-Oxo-3-(p | Handgrip | 0,021004 | 0,063388 | NA |
| 3 | abundance | X00305 | Formylkyn | Tibia_length | 0,093341 | 0,053145 | NA |
| 3 | abundance | X00305 | Formylkyn | Radius_length | 0,057459 | 0,072535 | NA |
| 3 | abundance | X00305 | Formylkyn | Radius_SOS | -0,05581 | 0,051013 | NA |
| 3 | abundance | X00305 | Formylkyn | Tibia_SOS | -0,02556 | 0,054593 | NA |
| 3 | abundance | X00305 | Formylkyn | Handgrip | -0,0321 | 0,062632 | NA |
| 3 | abundance | X00320 | Panthenol | Tibia_length | -0,04866 | 0,051291 | NA |
| 3 | abundance | X00320 | Panthenol | Radius_length | -0,07635 | 0,070093 | NA |
| 3 | abundance | X00320 | Panthenol | Radius_SOS | 0,061502 | 0,049496 | NA |
| 3 | abundance | X00320 | Panthenol | Tibia_SOS | -2,53E-04 | 0,053018 | NA |
| 3 | abundance | X00320 | Panthenol | Handgrip | -0,05737 | 0,060985 | NA |
| 3 | abundance | X00327 | 3-Hydroxy- | Tibia_length | -0,09 | 0,051085 | NA |
| 3 | abundance | X00327 | 3-Hydroxy- | Radius_length | -0,16902 | 0,069422 | NA |
| 3 | abundance | X00327 | 3-Hydroxy- | Radius_SOS | -0,03207 | 0,049638 | NA |
| 3 | abundance | X00327 | 3-Hydroxy- | Tibia_SOS | -0,03714 | 0,052972 | NA |
| 3 | abundance | X00327 | 3-Hydroxy- | Handgrip | -0,0341 | 0,061004 | NA |
| 3 | abundance | X00331 | 4-(9H-beta | Tibia_length | -0,08144 | 0,055548 | NA |

| | | | | | | | |
|---|---|---|---|---|---|---|---|
| 3 | abundance | X00331 | 4-(9H-beta | Radius_length | -0,03787 | 0,073982 | NA |
| 3 | abundance | X00331 | 4-(9H-beta | Radius_SOS | 0,054653 | 0,052897 | NA |
| 3 | abundance | X00331 | 4-(9H-beta | Tibia_SOS | -0,06232 | 0,05583 | NA |
| 3 | abundance | X00331 | 4-(9H-beta | Handgrip | -0,1199 | 0,062593 | NA |
| 3 | abundance | X00344 | SECONAL | Tibia_length | 0,051992 | 0,054214 | NA |
| 3 | abundance | X00344 | SECONAL | Radius_length | 0,154403 | 0,07229 | NA |
| 3 | abundance | X00344 | SECONAL | Radius_SOS | 0,022303 | 0,051268 | NA |
| 3 | abundance | X00344 | SECONAL | Tibia_SOS | 0,075401 | 0,054659 | NA |
| 3 | abundance | X00344 | SECONAL | Handgrip | 0,104345 | 0,062688 | NA |
| 3 | abundance | X00361 | nitecapone | Tibia_length | -0,0489 | 0,051313 | NA |
| 3 | abundance | X00361 | nitecapone | Radius_length | -0,0033 | 0,07036 | NA |
| 3 | abundance | X00361 | nitecapone | Radius_SOS | 0,0349 | 0,049674 | NA |
| 3 | abundance | X00361 | nitecapone | Tibia_SOS | -0,03044 | 0,053971 | NA |
| 3 | abundance | X00361 | nitecapone | Handgrip | -0,11201 | 0,06099 | NA |
| 3 | abundance | X00367 | hexobarbit | Tibia_length | -0,0948 | 0,055559 | NA |
| 3 | abundance | X00367 | hexobarbit | Radius_length | -0,11972 | 0,072966 | NA |
| 3 | abundance | X00367 | hexobarbit | Radius_SOS | -0,01476 | 0,052711 | NA |
| 3 | abundance | X00367 | hexobarbit | Tibia_SOS | -0,09154 | 0,056364 | NA |
| 3 | abundance | X00367 | hexobarbit | Handgrip | -0,07366 | 0,063311 | NA |
| 3 | abundance | X00375 | N-[(2S)-2-I | Tibia_length | 0,032165 | 0,055582 | NA |
| 3 | abundance | X00375 | N-[(2S)-2-I | Radius_length | 0,152909 | 0,073181 | NA |
| 3 | abundance | X00375 | N-[(2S)-2-I | Radius_SOS | -0,01767 | 0,052807 | NA |
| 3 | abundance | X00375 | N-[(2S)-2-I | Tibia_SOS | 0,084299 | 0,056717 | NA |
| 3 | abundance | X00375 | N-[(2S)-2-I | Handgrip | 0,11047 | 0,061769 | NA |
| 3 | abundance | X00383 | pretazettin | Tibia_length | -0,04474 | 0,052652 | NA |
| 3 | abundance | X00383 | pretazettin | Radius_length | -0,08874 | 0,071238 | NA |
| 3 | abundance | X00383 | pretazettin | Radius_SOS | 0,017431 | 0,050338 | NA |
| 3 | abundance | X00383 | pretazettin | Tibia_SOS | -0,05677 | 0,053258 | NA |
| 3 | abundance | X00383 | pretazettin | Handgrip | -0,04261 | 0,062166 | NA |
| 3 | abundance | X00384 | 3-Formyl-2 | Tibia_length | -0,09333 | 0,055797 | NA |
| 3 | abundance | X00384 | 3-Formyl-2 | Radius_length | -0,13905 | 0,073387 | NA |
| 3 | abundance | X00384 | 3-Formyl-2 | Radius_SOS | -0,10139 | 0,053287 | NA |
| 3 | abundance | X00384 | 3-Formyl-2 | Tibia_SOS | -0,0407 | 0,055806 | NA |
| 3 | abundance | X00384 | 3-Formyl-2 | Handgrip | -0,08733 | 0,061829 | NA |
| 3 | abundance | X00403 | 7alpha-Hy | Tibia_length | 0,055278 | 0,054074 | NA |
| 3 | abundance | X00403 | 7alpha-Hy | Radius_length | 0,071394 | 0,07259 | NA |
| 3 | abundance | X00403 | 7alpha-Hy | Radius_SOS | -0,01193 | 0,051494 | NA |
| 3 | abundance | X00403 | 7alpha-Hy | Tibia_SOS | -0,02223 | 0,055584 | NA |
| 3 | abundance | X00403 | 7alpha-Hy | Handgrip | 0,026234 | 0,061224 | NA |
| 3 | abundance | X00404 | piscidic ac | Tibia_length | -0,13788 | 0,055266 | NA |
| 3 | abundance | X00404 | piscidic ac | Radius_length | -0,17678 | 0,072665 | NA |
| 3 | abundance | X00404 | piscidic ac | Radius_SOS | -0,04274 | 0,052837 | NA |
| 3 | abundance | X00404 | piscidic ac | Tibia_SOS | -0,104 | 0,055084 | NA |
| 3 | abundance | X00404 | piscidic ac | Handgrip | -0,11983 | 0,060857 | NA |
| 3 | abundance | X00432 | 8-(Methyls | Tibia_length | -0,09103 | 0,055768 | NA |
| 3 | abundance | X00432 | 8-(Methyls | Radius_length | -0,11654 | 0,073399 | NA |
| 3 | abundance | X00432 | 8-(Methyls | Radius_SOS | -0,00205 | 0,053091 | NA |
| 3 | abundance | X00432 | 8-(Methyls | Tibia_SOS | -0,05358 | 0,056416 | NA |

| | | | | | | | |
|---|---|---|---|---|---|---|---|
| 3 | abundance | X00432 | 8-(Methyls | Handgrip | -0,14401 | 0,06212 | NA |
| 3 | abundance | X00450 | Gly-Ser | Tibia_length | -0,09265 | 0,054129 | NA |
| 3 | abundance | X00450 | Gly-Ser | Radius_length | -0,01147 | 0,073203 | NA |
| 3 | abundance | X00450 | Gly-Ser | Radius_SOS | -0,02642 | 0,05174 | NA |
| 3 | abundance | X00450 | Gly-Ser | Tibia_SOS | -0,01809 | 0,055134 | NA |
| 3 | abundance | X00450 | Gly-Ser | Handgrip | 0,066419 | 0,062705 | NA |
| 2b | abundance | X00477 | Sparfloxac | Tibia_length | 0,017386 | 0,055479 | NA |
| 2b | abundance | X00477 | Sparfloxac | Radius_length | 0,017131 | 0,074173 | NA |
| 2b | abundance | X00477 | Sparfloxac | Radius_SOS | -0,0548 | 0,052983 | NA |
| 2b | abundance | X00477 | Sparfloxac | Tibia_SOS | -0,05675 | 0,056383 | NA |
| 2b | abundance | X00477 | Sparfloxac | Handgrip | -0,05952 | 0,061967 | NA |
| 3 | abundance | X00518 | Nisinic aci | Tibia_length | 0,124295 | 0,052328 | NA |
| 3 | abundance | X00518 | Nisinic aci | Radius_length | 0,117315 | 0,071227 | NA |
| 3 | abundance | X00518 | Nisinic aci | Radius_SOS | 0,044141 | 0,050421 | NA |
| 3 | abundance | X00518 | Nisinic aci | Tibia_SOS | -0,06242 | 0,053845 | NA |
| 3 | abundance | X00518 | Nisinic aci | Handgrip | 0,128953 | 0,061376 | NA |
| 3 | abundance | X00528 | R-(+)-Etira | Tibia_length | 0,031212 | 0,049821 | NA |
| 3 | abundance | X00528 | R-(+)-Etira | Radius_length | -0,02202 | 0,067568 | NA |
| 3 | abundance | X00528 | R-(+)-Etira | Radius_SOS | -0,03447 | 0,049018 | NA |
| 3 | abundance | X00528 | R-(+)-Etira | Tibia_SOS | -0,02285 | 0,052485 | NA |
| 3 | abundance | X00528 | R-(+)-Etira | Handgrip | -0,02518 | 0,060604 | NA |
| 3 | abundance | X00545 | 3-hydroxyc | Tibia_length | -0,05435 | 0,054777 | NA |
| 3 | abundance | X00545 | 3-hydroxyc | Radius_length | -0,10671 | 0,072886 | NA |
| 3 | abundance | X00545 | 3-hydroxyc | Radius_SOS | 0,036547 | 0,052176 | NA |
| 3 | abundance | X00545 | 3-hydroxyc | Tibia_SOS | -0,05312 | 0,054887 | NA |
| 3 | abundance | X00545 | 3-hydroxyc | Handgrip | -0,02575 | 0,061877 | NA |
| 3 | abundance | X00549 | 3-[(2Z)-1-C | Tibia_length | -0,06816 | 0,051648 | NA |
| 3 | abundance | X00549 | 3-[(2Z)-1-C | Radius_length | -0,16753 | 0,069921 | NA |
| 3 | abundance | X00549 | 3-[(2Z)-1-C | Radius_SOS | -0,05345 | 0,049796 | NA |
| 3 | abundance | X00549 | 3-[(2Z)-1-C | Tibia_SOS | -0,14544 | 0,053014 | NA |
| 3 | abundance | X00549 | 3-[(2Z)-1-C | Handgrip | -0,11276 | 0,060612 | NA |
| 3 | abundance | X00574 | miglustat | Tibia_length | -0,04392 | 0,052156 | NA |
| 3 | abundance | X00574 | miglustat | Radius_length | -0,0954 | 0,071065 | NA |
| 3 | abundance | X00574 | miglustat | Radius_SOS | 0,020308 | 0,050028 | NA |
| 3 | abundance | X00574 | miglustat | Tibia_SOS | -0,08934 | 0,05402 | NA |
| 3 | abundance | X00574 | miglustat | Handgrip | -0,07351 | 0,061991 | NA |
| 3 | abundance | X00594 | Nicotinami | Tibia_length | -0,02234 | 0,055547 | NA |
| 3 | abundance | X00594 | Nicotinami | Radius_length | 0,030035 | 0,073691 | NA |
| 3 | abundance | X00594 | Nicotinami | Radius_SOS | -7,18E-04 | 0,052939 | NA |
| 3 | abundance | X00594 | Nicotinami | Tibia_SOS | -0,09739 | 0,056931 | NA |
| 3 | abundance | X00594 | Nicotinami | Handgrip | 0,092581 | 0,062504 | NA |
| 3 | abundance | X00635 | 4-(5,6-Dihy | Tibia_length | 0,01121 | 0,053008 | NA |
| 3 | abundance | X00635 | 4-(5,6-Dihy | Radius_length | 0,087553 | 0,071841 | NA |
| 3 | abundance | X00635 | 4-(5,6-Dihy | Radius_SOS | 0,023466 | 0,050704 | NA |
| 3 | abundance | X00635 | 4-(5,6-Dihy | Tibia_SOS | 0,12414 | 0,053898 | NA |
| 3 | abundance | X00635 | 4-(5,6-Dihy | Handgrip | -0,02368 | 0,061428 | NA |
| 3 | abundance | X00637 | N~6~,N~6 | Tibia_length | -0,02298 | 0,049409 | NA |
| 3 | abundance | X00637 | N~6~,N~6 | Radius_length | -0,01958 | 0,066389 | NA |

| | | | | | | | |
|---|---|---|---|---|---|---|---|
| 3 | abundance | X00637 | N~6~,N~6 | Radius_SOS | 0,013973 | 0,049043 | NA |
| 3 | abundance | X00637 | N~6~,N~6 | Tibia_SOS | -0,04685 | 0,052468 | NA |
| 3 | abundance | X00637 | N~6~,N~6 | Handgrip | -0,03782 | 0,06015 | NA |
| 3 | abundance | X00646 | Deacetyldi | Tibia_length | -0,01039 | 0,052894 | NA |
| 3 | abundance | X00646 | Deacetyldi | Radius_length | -0,05388 | 0,071646 | NA |
| 3 | abundance | X00646 | Deacetyldi | Radius_SOS | 0,036508 | 0,050592 | NA |
| 3 | abundance | X00646 | Deacetyldi | Tibia_SOS | -0,14126 | 0,053012 | NA |
| 3 | abundance | X00646 | Deacetyldi | Handgrip | -0,13173 | 0,060342 | NA |
| 3 | abundance | X00670 | 7-Chloro-5 | Tibia_length | -0,02063 | 0,05646 | NA |
| 3 | abundance | X00670 | 7-Chloro-5 | Radius_length | 0,033057 | 0,074276 | NA |
| 3 | abundance | X00670 | 7-Chloro-5 | Radius_SOS | -0,02285 | 0,053536 | NA |
| 3 | abundance | X00670 | 7-Chloro-5 | Tibia_SOS | -0,02657 | 0,057164 | NA |
| 3 | abundance | X00670 | 7-Chloro-5 | Handgrip | 0,126357 | 0,062861 | NA |
| 3 | abundance | X00683 | Zinecard | Tibia_length | -0,13331 | 0,054513 | NA |
| 3 | abundance | X00683 | Zinecard | Radius_length | -0,1037 | 0,073132 | NA |
| 3 | abundance | X00683 | Zinecard | Radius_SOS | -0,02173 | 0,052087 | NA |
| 3 | abundance | X00683 | Zinecard | Tibia_SOS | 0,01744 | 0,055704 | NA |
| 3 | abundance | X00683 | Zinecard | Handgrip | -0,05797 | 0,063524 | NA |
| 3 | abundance | X00693 | n-Ribosylh | Tibia_length | 0,040895 | 0,056748 | NA |
| 3 | abundance | X00693 | n-Ribosylh | Radius_length | 0,080856 | 0,07451 | NA |
| 3 | abundance | X00693 | n-Ribosylh | Radius_SOS | -0,0177 | 0,053983 | NA |
| 3 | abundance | X00693 | n-Ribosylh | Tibia_SOS | -0,12438 | 0,056451 | NA |
| 3 | abundance | X00693 | n-Ribosylh | Handgrip | 0,094676 | 0,063115 | NA |
| 3 | abundance | X00698 | valgancicl | Tibia_length | -0,03777 | 0,054915 | NA |
| 3 | abundance | X00698 | valgancicl | Radius_length | -0,12389 | 0,072894 | NA |
| 3 | abundance | X00698 | valgancicl | Radius_SOS | 0,043701 | 0,052363 | NA |
| 3 | abundance | X00698 | valgancicl | Tibia_SOS | -0,06158 | 0,054605 | NA |
| 3 | abundance | X00698 | valgancicl | Handgrip | 0,008073 | 0,061789 | NA |
| 3 | abundance | X00702 | N-[(2S)-2-I | Tibia_length | 0,019616 | 0,052127 | NA |
| 3 | abundance | X00702 | N-[(2S)-2-I | Radius_length | 0,092032 | 0,070769 | NA |
| 3 | abundance | X00702 | N-[(2S)-2-I | Radius_SOS | -0,03769 | 0,050058 | NA |
| 3 | abundance | X00702 | N-[(2S)-2-I | Tibia_SOS | 0,080957 | 0,053372 | NA |
| 3 | abundance | X00702 | N-[(2S)-2-I | Handgrip | -0,01046 | 0,061102 | NA |
| 3 | abundance | X00722 | (3S,5R,6E) | Tibia_length | -0,02703 | 0,055521 | NA |
| 3 | abundance | X00722 | (3S,5R,6E) | Radius_length | 0,072124 | 0,07412 | NA |
| 3 | abundance | X00722 | (3S,5R,6E) | Radius_SOS | -0,03996 | 0,052816 | NA |
| 3 | abundance | X00722 | (3S,5R,6E) | Tibia_SOS | 0,042769 | 0,056549 | NA |
| 3 | abundance | X00722 | (3S,5R,6E) | Handgrip | 0,071321 | 0,06004 | NA |
| 3 | abundance | X00723 | Aspartyl-L- | Tibia_length | 0,032235 | 0,052154 | NA |
| 3 | abundance | X00723 | Aspartyl-L- | Radius_length | 0,044171 | 0,071213 | NA |
| 3 | abundance | X00723 | Aspartyl-L- | Radius_SOS | 0,032187 | 0,050121 | NA |
| 3 | abundance | X00723 | Aspartyl-L- | Tibia_SOS | 0,149875 | 0,053005 | NA |
| 3 | abundance | X00723 | Aspartyl-L- | Handgrip | 0,067048 | 0,061585 | NA |
| 3 | abundance | X00742 | MFCD1869 | Tibia_length | -0,10032 | 0,054448 | NA |
| 3 | abundance | X00742 | MFCD1869 | Radius_length | -0,16347 | 0,072445 | NA |
| 3 | abundance | X00742 | MFCD1869 | Radius_SOS | -0,07481 | 0,051485 | NA |
| 3 | abundance | X00742 | MFCD1869 | Tibia_SOS | -0,06463 | 0,056486 | NA |
| 3 | abundance | X00742 | MFCD1869 | Handgrip | -0,09229 | 0,062115 | NA |

| | | | | | | | |
|---|---|---|---|---|---|---|---|
| 3 | abundance | X00743 | 11beta,13- | Tibia_length | 0,030463 | 0,057521 | NA |
| 3 | abundance | X00743 | 11beta,13- | Radius_length | 0,062608 | 0,074774 | NA |
| 3 | abundance | X00743 | 11beta,13- | Radius_SOS | -0,10168 | 0,054542 | NA |
| 3 | abundance | X00743 | 11beta,13- | Tibia_SOS | 0,055698 | 0,056951 | NA |
| 3 | abundance | X00743 | 11beta,13- | Handgrip | -0,0198 | 0,060181 | NA |
| 3 | abundance | X00744 | (1R,2S)-1-( | Tibia_length | -0,13375 | 0,055332 | NA |
| 3 | abundance | X00744 | (1R,2S)-1-( | Radius_length | -0,17614 | 0,072553 | NA |
| 3 | abundance | X00744 | (1R,2S)-1-( | Radius_SOS | 0,052234 | 0,052379 | NA |
| 3 | abundance | X00744 | (1R,2S)-1-( | Tibia_SOS | -0,10341 | 0,054794 | NA |
| 3 | abundance | X00744 | (1R,2S)-1-( | Handgrip | -0,06975 | 0,061996 | NA |
| 3 | abundance | X00748 | 3-[3-Metho | Tibia_length | -0,17031 | 0,054537 | NA |
| 3 | abundance | X00748 | 3-[3-Metho | Radius_length | -0,1382 | 0,072745 | NA |
| 3 | abundance | X00748 | 3-[3-Metho | Radius_SOS | -0,03839 | 0,052575 | NA |
| 3 | abundance | X00748 | 3-[3-Metho | Tibia_SOS | 0,017934 | 0,055536 | NA |
| 3 | abundance | X00748 | 3-[3-Metho | Handgrip | -0,13125 | 0,061544 | NA |
| 3 | abundance | X00764 | 5-Allyl-5-se | Tibia_length | -0,0047 | 0,051111 | NA |
| 3 | abundance | X00764 | 5-Allyl-5-se | Radius_length | 0,106688 | 0,069691 | NA |
| 3 | abundance | X00764 | 5-Allyl-5-se | Radius_SOS | 0,034191 | 0,049508 | NA |
| 3 | abundance | X00764 | 5-Allyl-5-se | Tibia_SOS | -0,06466 | 0,052632 | NA |
| 3 | abundance | X00764 | 5-Allyl-5-se | Handgrip | -0,01438 | 0,060843 | NA |
| 3 | abundance | X00807 | Midodrine | Tibia_length | -0,13997 | 0,053331 | NA |
| 3 | abundance | X00807 | Midodrine | Radius_length | -0,16351 | 0,072262 | NA |
| 3 | abundance | X00807 | Midodrine | Radius_SOS | -0,03994 | 0,051485 | NA |
| 3 | abundance | X00807 | Midodrine | Tibia_SOS | -0,04332 | 0,055591 | NA |
| 3 | abundance | X00807 | Midodrine | Handgrip | -0,18596 | 0,061356 | NA |
| 3 | abundance | X00828 | Fenoterol | Tibia_length | -0,00526 | 0,052341 | NA |
| 3 | abundance | X00828 | Fenoterol | Radius_length | 0,058463 | 0,071354 | NA |
| 3 | abundance | X00828 | Fenoterol | Radius_SOS | 0,061679 | 0,050331 | NA |
| 3 | abundance | X00828 | Fenoterol | Tibia_SOS | 0,034643 | 0,053602 | NA |
| 3 | abundance | X00828 | Fenoterol | Handgrip | -5,99E-04 | 0,061999 | NA |
| 3 | abundance | X00836 | 3-Mercapto | Tibia_length | 0,076263 | 0,050727 | NA |
| 3 | abundance | X00836 | 3-Mercapto | Radius_length | 0,090852 | 0,069619 | NA |
| 3 | abundance | X00836 | 3-Mercapto | Radius_SOS | 0,073474 | 0,049272 | NA |
| 3 | abundance | X00836 | 3-Mercapto | Tibia_SOS | 0,059365 | 0,052873 | NA |
| 3 | abundance | X00836 | 3-Mercapto | Handgrip | -0,03154 | 0,060413 | NA |
| 3 | abundance | X00853 | 4-O-beta-D | Tibia_length | -0,07612 | 0,048828 | NA |
| 3 | abundance | X00853 | 4-O-beta-D | Radius_length | -0,06024 | 0,048881 | NA |
| 3 | abundance | X00853 | 4-O-beta-D | Radius_SOS | -0,03093 | 0,049006 | NA |
| 3 | abundance | X00853 | 4-O-beta-D | Tibia_SOS | -0,04093 | 0,059374 | NA |
| 3 | abundance | X00853 | 4-O-beta-D | Handgrip | -0,06338 | 0,060072 | NA |
| 3 | abundance | X00889 | LW800000 | Tibia_length | -0,0504 | 0,05557 | NA |
| 3 | abundance | X00889 | LW800000 | Radius_length | -0,02855 | 0,073794 | NA |
| 3 | abundance | X00889 | LW800000 | Radius_SOS | 0,107515 | 0,05234 | NA |
| 3 | abundance | X00889 | LW800000 | Tibia_SOS | 0,101271 | 0,055787 | NA |
| 3 | abundance | X00889 | LW800000 | Handgrip | 0,074882 | 0,062895 | NA |
| 3 | abundance | X00899 | N-(2,3,4-Tr | Tibia_length | -0,04371 | 0,054455 | NA |
| 3 | abundance | X00899 | N-(2,3,4-Tr | Radius_length | -0,09967 | 0,072699 | NA |
| 3 | abundance | X00899 | N-(2,3,4-Tr | Radius_SOS | -0,11822 | 0,051883 | NA |

| | | | | | | | |
|---|---|---|---|---|---|---|---|
| 3 | abundance | X00899 | N-(2,3,4-Tr | Tibia_SOS | -0,02906 | 0,055376 | NA |
| 3 | abundance | X00899 | N-(2,3,4-Tr | Handgrip | -0,04102 | 0,061752 | NA |
| 3 | abundance | X00907 | 1,2,3,4-Tet | Tibia_length | -0,02053 | 0,053741 | NA |
| 3 | abundance | X00907 | 1,2,3,4-Tet | Radius_length | -0,12564 | 0,072389 | NA |
| 3 | abundance | X00907 | 1,2,3,4-Tet | Radius_SOS | -0,00902 | 0,05119 | NA |
| 3 | abundance | X00907 | 1,2,3,4-Tet | Tibia_SOS | 0,050042 | 0,054344 | NA |
| 3 | abundance | X00907 | 1,2,3,4-Tet | Handgrip | 0,008022 | 0,061678 | NA |
| 3 | abundance | X00928 | (7R)-7-(5-c | Tibia_length | 0,089581 | 0,05362 | NA |
| 3 | abundance | X00928 | (7R)-7-(5-c | Radius_length | 0,033622 | 0,072276 | NA |
| 3 | abundance | X00928 | (7R)-7-(5-c | Radius_SOS | -0,00747 | 0,051109 | NA |
| 3 | abundance | X00928 | (7R)-7-(5-c | Tibia_SOS | 0,095309 | 0,054534 | NA |
| 3 | abundance | X00928 | (7R)-7-(5-c | Handgrip | 0,056067 | 0,062002 | NA |
| 3 | abundance | X00948 | Linamarin | Tibia_length | -0,07275 | 0,051495 | NA |
| 3 | abundance | X00948 | Linamarin | Radius_length | -0,09689 | 0,070269 | NA |
| 3 | abundance | X00948 | Linamarin | Radius_SOS | -0,0133 | 0,04969 | NA |
| 3 | abundance | X00948 | Linamarin | Tibia_SOS | -0,15769 | 0,052376 | NA |
| 3 | abundance | X00948 | Linamarin | Handgrip | -0,17887 | 0,059897 | NA |
| 3 | abundance | X00949 | primidone | Tibia_length | 0,16541 | 0,052738 | NA |
| 3 | abundance | X00949 | primidone | Radius_length | 0,095181 | 0,072273 | NA |
| 3 | abundance | X00949 | primidone | Radius_SOS | 0,048426 | 0,050862 | NA |
| 3 | abundance | X00949 | primidone | Tibia_SOS | -0,01145 | 0,054738 | NA |
| 3 | abundance | X00949 | primidone | Handgrip | 0,02346 | 0,062197 | NA |
| 3 | abundance | X00950 | 2-Phenylet | Tibia_length | -0,02198 | 0,055997 | NA |
| 3 | abundance | X00950 | 2-Phenylet | Radius_length | -0,02798 | 0,074909 | NA |
| 3 | abundance | X00950 | 2-Phenylet | Radius_SOS | -0,05001 | 0,053104 | NA |
| 3 | abundance | X00950 | 2-Phenylet | Tibia_SOS | 0,077726 | 0,057982 | NA |
| 3 | abundance | X00950 | 2-Phenylet | Handgrip | -0,19653 | 0,06251 | NA |
| 3 | abundance | X00963 | (4S)-4-[(2E | Tibia_length | 0,147797 | 0,05221 | NA |
| 3 | abundance | X00963 | (4S)-4-[(2E | Radius_length | 0,206846 | 0,070845 | NA |
| 3 | abundance | X00963 | (4S)-4-[(2E | Radius_SOS | 0,0838 | 0,050544 | NA |
| 3 | abundance | X00963 | (4S)-4-[(2E | Tibia_SOS | 0,087385 | 0,053936 | NA |
| 3 | abundance | X00963 | (4S)-4-[(2E | Handgrip | 0,096304 | 0,060618 | NA |
| 3 | abundance | X00985 | 1-(4-Aminc | Tibia_length | -0,0991 | 0,052912 | NA |
| 3 | abundance | X00985 | 1-(4-Aminc | Radius_length | -0,2064 | 0,070697 | NA |
| 3 | abundance | X00985 | 1-(4-Aminc | Radius_SOS | -0,05771 | 0,050582 | NA |
| 3 | abundance | X00985 | 1-(4-Aminc | Tibia_SOS | -0,20903 | 0,052927 | NA |
| 3 | abundance | X00985 | 1-(4-Aminc | Handgrip | -0,23293 | 0,0602 | NA |
| 3 | abundance | X00998 | epsilon-(ga | Tibia_length | 0,043734 | 0,053375 | NA |
| 3 | abundance | X00998 | epsilon-(ga | Radius_length | 0,078196 | 0,072519 | NA |
| 3 | abundance | X00998 | epsilon-(ga | Radius_SOS | -0,00113 | 0,051021 | NA |
| 3 | abundance | X00998 | epsilon-(ga | Tibia_SOS | 0,059686 | 0,054934 | NA |
| 3 | abundance | X00998 | epsilon-(ga | Handgrip | 0,031554 | 0,061905 | NA |
| 3 | abundance | X01007 | pentobarbi | Tibia_length | -0,10395 | 0,053804 | NA |
| 3 | abundance | X01007 | pentobarbi | Radius_length | -0,20229 | 0,071104 | NA |
| 3 | abundance | X01007 | pentobarbi | Radius_SOS | -0,05482 | 0,051079 | NA |
| 3 | abundance | X01007 | pentobarbi | Tibia_SOS | -0,13688 | 0,053438 | NA |
| 3 | abundance | X01007 | pentobarbi | Handgrip | -0,01706 | 0,061355 | NA |
| 3 | abundance | X01011 | DIBEHENIN | Tibia_length | -0,01065 | 0,054208 | NA |

| | | | | | | | |
|---|---|---|---|---|---|---|---|
| 3 | abundance | X01011 | DIBEHENII | Radius_length | -0,07879 | 0,073147 | NA |
| 3 | abundance | X01011 | DIBEHENII | Radius_SOS | -0,0922 | 0,051418 | NA |
| 3 | abundance | X01011 | DIBEHENII | Tibia_SOS | -0,06323 | 0,056213 | NA |
| 3 | abundance | X01011 | DIBEHENII | Handgrip | -0,1141 | 0,06247 | NA |
| 3 | abundance | X01017 | hexobarbit | Tibia_length | -0,14337 | 0,059203 | NA |
| 3 | abundance | X01017 | hexobarbit | Radius_length | -0,04494 | 0,076268 | NA |
| 3 | abundance | X01017 | hexobarbit | Radius_SOS | 0,046936 | 0,056382 | NA |
| 3 | abundance | X01017 | hexobarbit | Tibia_SOS | 0,050154 | 0,05997 | NA |
| 3 | abundance | X01017 | hexobarbit | Handgrip | 0,11009 | 0,063379 | NA |
| 3 | abundance | X01031 | 3-(14-Ethy | Tibia_length | 0,066944 | 0,05815 | NA |
| 3 | abundance | X01031 | 3-(14-Ethy | Radius_length | 0,249939 | 0,073148 | NA |
| 3 | abundance | X01031 | 3-(14-Ethy | Radius_SOS | -0,08124 | 0,05519 | NA |
| 3 | abundance | X01031 | 3-(14-Ethy | Tibia_SOS | 0,03813 | 0,058042 | NA |
| 3 | abundance | X01031 | 3-(14-Ethy | Handgrip | 0,119493 | 0,059762 | NA |
| 3 | abundance | X01045 | Ethyl maltc | Tibia_length | -0,01939 | 0,053843 | NA |
| 3 | abundance | X01045 | Ethyl maltc | Radius_length | 0,042631 | 0,07257 | NA |
| 3 | abundance | X01045 | Ethyl maltc | Radius_SOS | 0,014729 | 0,051348 | NA |
| 3 | abundance | X01045 | Ethyl maltc | Tibia_SOS | 0,056677 | 0,055004 | NA |
| 3 | abundance | X01045 | Ethyl maltc | Handgrip | 0,042608 | 0,06286 | NA |
| 3 | abundance | X01059 | coronatine | Tibia_length | -0,19851 | 0,054915 | NA |
| 3 | abundance | X01059 | coronatine | Radius_length | -0,12179 | 0,073159 | NA |
| 3 | abundance | X01059 | coronatine | Radius_SOS | 0,002917 | 0,052611 | NA |
| 3 | abundance | X01059 | coronatine | Tibia_SOS | -0,0543 | 0,056088 | NA |
| 3 | abundance | X01059 | coronatine | Handgrip | -0,05004 | 0,062056 | NA |
| 3 | abundance | X01067 | g-Aminobu | Tibia_length | 0,040805 | 0,055635 | NA |
| 3 | abundance | X01067 | g-Aminobu | Radius_length | -0,03386 | 0,073998 | NA |
| 3 | abundance | X01067 | g-Aminobu | Radius_SOS | -0,04501 | 0,053016 | NA |
| 3 | abundance | X01067 | g-Aminobu | Tibia_SOS | 0,082827 | 0,05544 | NA |
| 3 | abundance | X01067 | g-Aminobu | Handgrip | 0,042921 | 0,062017 | NA |
| 3 | abundance | X01078 | Dihydroura | Tibia_length | -0,09479 | 0,051747 | NA |
| 3 | abundance | X01078 | Dihydroura | Radius_length | -0,11915 | 0,070596 | NA |
| 3 | abundance | X01078 | Dihydroura | Radius_SOS | 0,063418 | 0,049927 | NA |
| 3 | abundance | X01078 | Dihydroura | Tibia_SOS | -0,08013 | 0,053477 | NA |
| 3 | abundance | X01078 | Dihydroura | Handgrip | -0,16438 | 0,060101 | NA |
| 3 | abundance | X01081 | Methyl alpl | Tibia_length | -0,07469 | 0,052168 | NA |
| 3 | abundance | X01081 | Methyl alpl | Radius_length | 0,021994 | 0,071158 | NA |
| 3 | abundance | X01081 | Methyl alpl | Radius_SOS | -0,01118 | 0,050113 | NA |
| 3 | abundance | X01081 | Methyl alpl | Tibia_SOS | -0,06995 | 0,053723 | NA |
| 3 | abundance | X01081 | Methyl alpl | Handgrip | -0,05023 | 0,061129 | NA |
| 3 | abundance | X01092 | 17,21-Dihy | Tibia_length | 0,074914 | 0,050747 | NA |
| 3 | abundance | X01092 | 17,21-Dihy | Radius_length | 0,088345 | 0,069203 | NA |
| 3 | abundance | X01092 | 17,21-Dihy | Radius_SOS | 0,007663 | 0,049379 | NA |
| 3 | abundance | X01092 | 17,21-Dihy | Tibia_SOS | 0,017681 | 0,05266 | NA |
| 3 | abundance | X01092 | 17,21-Dihy | Handgrip | 0,001617 | 0,060554 | NA |
| 3 | abundance | X01098 | (-)-Physost | Tibia_length | -0,04128 | 0,057906 | NA |
| 3 | abundance | X01098 | (-)-Physost | Radius_length | -0,11764 | 0,075152 | NA |
| 3 | abundance | X01098 | (-)-Physost | Radius_SOS | 0,029231 | 0,054957 | NA |
| 3 | abundance | X01098 | (-)-Physost | Tibia_SOS | -0,15918 | 0,056099 | NA |

| | | | | | | | |
|---|---|---|---|---|---|---|---|
| 3 | abundance | X01098 | (-)-Physost | Handgrip | -0,11407 | 0,062682 | NA |
| 3 | abundance | X01100 | (betaS)-be | Tibia_length | -0,15937 | 0,056142 | NA |
| 3 | abundance | X01100 | (betaS)-be | Radius_length | -0,13381 | 0,074494 | NA |
| 3 | abundance | X01100 | (betaS)-be | Radius_SOS | 0,017202 | 0,054061 | NA |
| 3 | abundance | X01100 | (betaS)-be | Tibia_SOS | -0,13971 | 0,056616 | NA |
| 3 | abundance | X01100 | (betaS)-be | Handgrip | -0,05068 | 0,063319 | NA |
| 3 | abundance | X01154 | N-(2,3,4-Tr | Tibia_length | -0,03868 | 0,056177 | NA |
| 3 | abundance | X01154 | N-(2,3,4-Tr | Radius_length | -0,07317 | 0,074016 | NA |
| 3 | abundance | X01154 | N-(2,3,4-Tr | Radius_SOS | -0,0679 | 0,053285 | NA |
| 3 | abundance | X01154 | N-(2,3,4-Tr | Tibia_SOS | -0,09403 | 0,057089 | NA |
| 3 | abundance | X01154 | N-(2,3,4-Tr | Handgrip | -0,00732 | 0,062333 | NA |
| 3 | abundance | X01163 | Prephenic | Tibia_length | -0,11995 | 0,054427 | NA |
| 3 | abundance | X01163 | Prephenic | Radius_length | -0,25776 | 0,070826 | NA |
| 3 | abundance | X01163 | Prephenic | Radius_SOS | -0,04159 | 0,051816 | NA |
| 3 | abundance | X01163 | Prephenic | Tibia_SOS | -0,10661 | 0,054549 | NA |
| 3 | abundance | X01163 | Prephenic | Handgrip | -0,13438 | 0,061959 | NA |
| 3 | abundance | X01164 | DL-Carboc | Tibia_length | -0,05385 | 0,056666 | NA |
| 3 | abundance | X01164 | DL-Carboc | Radius_length | -0,02939 | 0,075091 | NA |
| 3 | abundance | X01164 | DL-Carboc | Radius_SOS | 0,022607 | 0,053934 | NA |
| 3 | abundance | X01164 | DL-Carboc | Tibia_SOS | 0,021383 | 0,056857 | NA |
| 3 | abundance | X01164 | DL-Carboc | Handgrip | -0,05853 | 0,063678 | NA |
| 3 | abundance | X01181 | 2-(3,4-Dim | Tibia_length | -0,05938 | 0,051931 | NA |
| 3 | abundance | X01181 | 2-(3,4-Dim | Radius_length | -0,11302 | 0,07067 | NA |
| 3 | abundance | X01181 | 2-(3,4-Dim | Radius_SOS | 0,001284 | 0,05007 | NA |
| 3 | abundance | X01181 | 2-(3,4-Dim | Tibia_SOS | -0,12921 | 0,052618 | NA |
| 3 | abundance | X01181 | 2-(3,4-Dim | Handgrip | -0,1129 | 0,060314 | NA |
| 3 | abundance | X01186 | asn-val | Tibia_length | -0,0786 | 0,05479 | NA |
| 3 | abundance | X01186 | asn-val | Radius_length | -0,0291 | 0,073894 | NA |
| 3 | abundance | X01186 | asn-val | Radius_SOS | -0,02546 | 0,052426 | NA |
| 3 | abundance | X01186 | asn-val | Tibia_SOS | -0,02454 | 0,057019 | NA |
| 3 | abundance | X01186 | asn-val | Handgrip | -0,01575 | 0,062022 | NA |
| 3 | abundance | X01208 | Lys-Pro | Tibia_length | 0,094728 | 0,05122 | NA |
| 3 | abundance | X01208 | Lys-Pro | Radius_length | 0,093903 | 0,070453 | NA |
| 3 | abundance | X01208 | Lys-Pro | Radius_SOS | 0,084621 | 0,049455 | NA |
| 3 | abundance | X01208 | Lys-Pro | Tibia_SOS | 0,00377 | 0,053236 | NA |
| 3 | abundance | X01208 | Lys-Pro | Handgrip | -0,03662 | 0,061132 | NA |
| 3 | abundance | X01223 | DL-Mevalo | Tibia_length | -0,03461 | 0,053506 | NA |
| 3 | abundance | X01223 | DL-Mevalo | Radius_length | -0,10063 | 0,072358 | NA |
| 3 | abundance | X01223 | DL-Mevalo | Radius_SOS | -0,05034 | 0,051079 | NA |
| 3 | abundance | X01223 | DL-Mevalo | Tibia_SOS | -0,00497 | 0,054904 | NA |
| 3 | abundance | X01223 | DL-Mevalo | Handgrip | 0,021654 | 0,061531 | NA |
| 3 | abundance | X01235 | 3-[(3-Hydro | Tibia_length | 0,17846 | 0,056519 | NA |
| 3 | abundance | X01235 | 3-[(3-Hydro | Radius_length | 0,28961 | 0,072646 | NA |
| 3 | abundance | X01235 | 3-[(3-Hydro | Radius_SOS | 0,098401 | 0,054019 | NA |
| 3 | abundance | X01235 | 3-[(3-Hydro | Tibia_SOS | 0,088518 | 0,058835 | NA |
| 3 | abundance | X01235 | 3-[(3-Hydro | Handgrip | 0,143953 | 0,059566 | NA |
| 3 | abundance | X01236 | bis(4-isoth | Tibia_length | 0,070204 | 0,056166 | NA |
| 3 | abundance | X01236 | bis(4-isoth | Radius_length | 0,193557 | 0,0743 | NA |

| | | | | | | | | |
|---|---|---|---|---|---|---|---|---|
| | 3 | abundance | X01236 | bis(4-isoth | Radius_SOS | 0,101831 | 0,052944 | NA |
| | 3 | abundance | X01236 | bis(4-isoth | Tibia_SOS | 0,050492 | 0,057475 | NA |
| | 3 | abundance | X01236 | bis(4-isoth | Handgrip | 0,161715 | 0,062542 | NA |
| 2b | | abundance | X01242 | 3-Morpholi | Tibia_length | -0,0201 | 0,057968 | NA |
| 2b | | abundance | X01242 | 3-Morpholi | Radius_length | 0,042537 | 0,075308 | NA |
| 2b | | abundance | X01242 | 3-Morpholi | Radius_SOS | 0,001627 | 0,055146 | NA |
| 2b | | abundance | X01242 | 3-Morpholi | Tibia_SOS | 0,040567 | 0,058908 | NA |
| 2b | | abundance | X01242 | 3-Morpholi | Handgrip | 0,008453 | 0,063641 | NA |
| | 3 | abundance | X01246 | (2R)-1-{[(2 | Tibia_length | 0,042204 | 0,054276 | NA |
| | 3 | abundance | X01246 | (2R)-1-{[(2 | Radius_length | 0,107704 | 0,072723 | NA |
| | 3 | abundance | X01246 | (2R)-1-{[(2 | Radius_SOS | -0,03369 | 0,051807 | NA |
| | 3 | abundance | X01246 | (2R)-1-{[(2 | Tibia_SOS | 0,019857 | 0,05546 | NA |
| | 3 | abundance | X01246 | (2R)-1-{[(2 | Handgrip | 0,056156 | 0,062825 | NA |
| | 3 | abundance | X01252 | (âˆ’)-nabil | Tibia_length | -0,00643 | 0,052942 | NA |
| | 3 | abundance | X01252 | (âˆ’)-nabil | Radius_length | -0,03525 | 0,071809 | NA |
| | 3 | abundance | X01252 | (âˆ’)-nabil | Radius_SOS | 0,035319 | 0,050553 | NA |
| | 3 | abundance | X01252 | (âˆ’)-nabil | Tibia_SOS | 0,006564 | 0,053969 | NA |
| | 3 | abundance | X01252 | (âˆ’)-nabil | Handgrip | -0,02432 | 0,062432 | NA |
| | 3 | abundance | X01285 | FB950000 | Tibia_length | -0,11127 | 0,054129 | NA |
| | 3 | abundance | X01285 | FB950000 | Radius_length | -0,09951 | 0,072561 | NA |
| | 3 | abundance | X01285 | FB950000 | Radius_SOS | 0,016106 | 0,051573 | NA |
| | 3 | abundance | X01285 | FB950000 | Tibia_SOS | -0,06417 | 0,055664 | NA |
| | 3 | abundance | X01285 | FB950000 | Handgrip | -0,08813 | 0,062091 | NA |
| | 3 | abundance | X01286 | (2,4-Dihyd | Tibia_length | -0,10244 | 0,05014 | NA |
| | 3 | abundance | X01286 | (2,4-Dihyd | Radius_length | -0,17865 | 0,067827 | NA |
| | 3 | abundance | X01286 | (2,4-Dihyd | Radius_SOS | 0,011386 | 0,049145 | NA |
| | 3 | abundance | X01286 | (2,4-Dihyd | Tibia_SOS | -0,19684 | 0,051469 | NA |
| | 3 | abundance | X01286 | (2,4-Dihyd | Handgrip | -0,21369 | 0,058819 | NA |
| | 3 | abundance | X01288 | L-gamma-( | Tibia_length | -0,07763 | 0,051998 | NA |
| | 3 | abundance | X01288 | L-gamma-( | Radius_length | -0,12451 | 0,070531 | NA |
| | 3 | abundance | X01288 | L-gamma-( | Radius_SOS | 0,009216 | 0,049885 | NA |
| | 3 | abundance | X01288 | L-gamma-( | Tibia_SOS | -0,13169 | 0,052651 | NA |
| | 3 | abundance | X01288 | L-gamma-( | Handgrip | -0,0991 | 0,060194 | NA |
| | 3 | abundance | X01316 | 5-Hydanto | Tibia_length | 0,002372 | 0,054771 | NA |
| | 3 | abundance | X01316 | 5-Hydanto | Radius_length | -0,08569 | 0,073617 | NA |
| | 3 | abundance | X01316 | 5-Hydanto | Radius_SOS | -0,03564 | 0,052012 | NA |
| | 3 | abundance | X01316 | 5-Hydanto | Tibia_SOS | 0,10599 | 0,055268 | NA |
| | 3 | abundance | X01316 | 5-Hydanto | Handgrip | -0,01105 | 0,061043 | NA |
| | 3 | abundance | X01327 | threonylph | Tibia_length | -0,15021 | 0,053071 | NA |
| | 3 | abundance | X01327 | threonylph | Radius_length | -0,21427 | 0,070675 | NA |
| | 3 | abundance | X01327 | threonylph | Radius_SOS | -0,07543 | 0,050756 | NA |
| | 3 | abundance | X01327 | threonylph | Tibia_SOS | -0,14049 | 0,054014 | NA |
| | 3 | abundance | X01327 | threonylph | Handgrip | -0,10511 | 0,060784 | NA |
| | 3 | abundance | X01340 | Cadralazin | Tibia_length | 0,004105 | 0,0532 | NA |
| | 3 | abundance | X01340 | Cadralazin | Radius_length | 0,188322 | 0,070775 | NA |
| | 3 | abundance | X01340 | Cadralazin | Radius_SOS | -0,04548 | 0,050659 | NA |
| | 3 | abundance | X01340 | Cadralazin | Tibia_SOS | -0,054 | 0,054856 | NA |
| | 3 | abundance | X01340 | Cadralazin | Handgrip | -0,01938 | 0,061606 | NA |

| | | | | | | | |
|---|---|---|---|---|---|---|---|
| 3 | abundance | X01341 | metixene | Tibia_length | 0,130118 | 0,051514 | NA |
| 3 | abundance | X01341 | metixene | Radius_length | 0,153746 | 0,070172 | NA |
| 3 | abundance | X01341 | metixene | Radius_SOS | -0,01164 | 0,049805 | NA |
| 3 | abundance | X01341 | metixene | Tibia_SOS | 0,076126 | 0,053566 | NA |
| 3 | abundance | X01341 | metixene | Handgrip | 0,033096 | 0,061522 | NA |
| 3 | abundance | X01346 | IN00260 | Tibia_length | -0,06848 | 0,050071 | NA |
| 3 | abundance | X01346 | IN00260 | Radius_length | -0,07711 | 0,068376 | NA |
| 3 | abundance | X01346 | IN00260 | Radius_SOS | -0,05841 | 0,049058 | NA |
| 3 | abundance | X01346 | IN00260 | Tibia_SOS | -0,02999 | 0,052571 | NA |
| 3 | abundance | X01346 | IN00260 | Handgrip | -0,00768 | 0,060536 | NA |
| 3 | abundance | X01363 | Seryltyrosi | Tibia_length | -0,11629 | 0,053501 | NA |
| 3 | abundance | X01363 | Seryltyrosi | Radius_length | -0,14981 | 0,071505 | NA |
| 3 | abundance | X01363 | Seryltyrosi | Radius_SOS | 0,004969 | 0,050949 | NA |
| 3 | abundance | X01363 | Seryltyrosi | Tibia_SOS | -0,10358 | 0,053698 | NA |
| 3 | abundance | X01363 | Seryltyrosi | Handgrip | -0,06999 | 0,06091 | NA |
| 3 | abundance | X01364 | Oleuropeir | Tibia_length | -0,07947 | 0,056772 | NA |
| 3 | abundance | X01364 | Oleuropeir | Radius_length | 0,041928 | 0,074736 | NA |
| 3 | abundance | X01364 | Oleuropeir | Radius_SOS | 0,055862 | 0,053904 | NA |
| 3 | abundance | X01364 | Oleuropeir | Tibia_SOS | 0,093015 | 0,058012 | NA |
| 3 | abundance | X01364 | Oleuropeir | Handgrip | 0,053521 | 0,061612 | NA |
| 3 | abundance | X01367 | MFCD1869 | Tibia_length | 0,026653 | 0,054053 | NA |
| 3 | abundance | X01367 | MFCD1869 | Radius_length | 0,143937 | 0,07202 | NA |
| 3 | abundance | X01367 | MFCD1869 | Radius_SOS | -0,01044 | 0,051249 | NA |
| 3 | abundance | X01367 | MFCD1869 | Tibia_SOS | 0,025865 | 0,054668 | NA |
| 3 | abundance | X01367 | MFCD1869 | Handgrip | 0,028587 | 0,062969 | NA |
| 3 | abundance | X01380 | Spermic ac | Tibia_length | -0,05897 | 0,057715 | NA |
| 3 | abundance | X01380 | Spermic ac | Radius_length | 0,010028 | 0,074946 | NA |
| 3 | abundance | X01380 | Spermic ac | Radius_SOS | -0,03397 | 0,054871 | NA |
| 3 | abundance | X01380 | Spermic ac | Tibia_SOS | 0,010718 | 0,05751 | NA |
| 3 | abundance | X01380 | Spermic ac | Handgrip | -0,05014 | 0,062828 | NA |
| 3 | abundance | X01387 | Methyl 4-(4 | Tibia_length | -0,12664 | 0,05386 | NA |
| 3 | abundance | X01387 | Methyl 4-(4 | Radius_length | -0,15789 | 0,072791 | NA |
| 3 | abundance | X01387 | Methyl 4-(4 | Radius_SOS | -0,08734 | 0,051638 | NA |
| 3 | abundance | X01387 | Methyl 4-(4 | Tibia_SOS | -0,02685 | 0,055908 | NA |
| 3 | abundance | X01387 | Methyl 4-(4 | Handgrip | -0,12776 | 0,062504 | NA |
| 3 | abundance | X01441 | 1-{3-Carbo | Tibia_length | 0,009343 | 0,052441 | NA |
| 3 | abundance | X01441 | 1-{3-Carbo | Radius_length | -0,05353 | 0,071267 | NA |
| 3 | abundance | X01441 | 1-{3-Carbo | Radius_SOS | 0,032015 | 0,050293 | NA |
| 3 | abundance | X01441 | 1-{3-Carbo | Tibia_SOS | -0,11577 | 0,053544 | NA |
| 3 | abundance | X01441 | 1-{3-Carbo | Handgrip | -0,11084 | 0,061543 | NA |
| 3 | abundance | X01463 | 4-Thiapent | Tibia_length | 0,124044 | 0,051898 | NA |
| 3 | abundance | X01463 | 4-Thiapent | Radius_length | 0,143703 | 0,071095 | NA |
| 3 | abundance | X01463 | 4-Thiapent | Radius_SOS | 0,051839 | 0,050024 | NA |
| 3 | abundance | X01463 | 4-Thiapent | Tibia_SOS | 0,103133 | 0,053572 | NA |
| 3 | abundance | X01463 | 4-Thiapent | Handgrip | 0,174349 | 0,05968 | NA |
| 3 | abundance | X01474 | Glyceropho | Tibia_length | -0,10666 | 0,05519 | NA |
| 3 | abundance | X01474 | Glyceropho | Radius_length | 0,034048 | 0,073574 | NA |
| 3 | abundance | X01474 | Glyceropho | Radius_SOS | 0,009822 | 0,052797 | NA |

| | | | | | | | |
|---|---|---|---|---|---|---|---|
| 3 | abundance | X01474 | Glycerophc | Tibia_SOS | 0,023239 | 0,056178 | NA |
| 3 | abundance | X01474 | Glycerophc | Handgrip | -0,03199 | 0,060761 | NA |
| 3 | abundance | X01518 | L-gamma-( | Tibia_length | -0,04338 | 0,053276 | NA |
| 3 | abundance | X01518 | L-gamma-( | Radius_length | -0,1062 | 0,072147 | NA |
| 3 | abundance | X01518 | L-gamma-( | Radius_SOS | -0,07821 | 0,05076 | NA |
| 3 | abundance | X01518 | L-gamma-( | Tibia_SOS | -0,10354 | 0,053896 | NA |
| 3 | abundance | X01518 | L-gamma-( | Handgrip | -0,03965 | 0,061829 | NA |
| 3 | abundance | X01519 | Leucyltrypt | Tibia_length | -0,09888 | 0,053879 | NA |
| 3 | abundance | X01519 | Leucyltrypt | Radius_length | -0,11946 | 0,072446 | NA |
| 3 | abundance | X01519 | Leucyltrypt | Radius_SOS | 0,016754 | 0,051585 | NA |
| 3 | abundance | X01519 | Leucyltrypt | Tibia_SOS | 0,00904 | 0,054533 | NA |
| 3 | abundance | X01519 | Leucyltrypt | Handgrip | -0,16792 | 0,062418 | NA |
| 3 | abundance | X01528 | butalbital | Tibia_length | -0,07404 | 0,054025 | NA |
| 3 | abundance | X01528 | butalbital | Radius_length | 0,012401 | 0,072938 | NA |
| 3 | abundance | X01528 | butalbital | Radius_SOS | -7,69E-04 | 0,051586 | NA |
| 3 | abundance | X01528 | butalbital | Tibia_SOS | -0,03468 | 0,055483 | NA |
| 3 | abundance | X01528 | butalbital | Handgrip | 0,011923 | 0,061361 | NA |
| 2b | abundance | X01530 | INK (Peptic | Tibia_length | -0,03325 | 0,054998 | NA |
| 2b | abundance | X01530 | INK (Peptic | Radius_length | 0,045609 | 0,073677 | NA |
| 2b | abundance | X01530 | INK (Peptic | Radius_SOS | 0,064445 | 0,052375 | NA |
| 2b | abundance | X01530 | INK (Peptic | Tibia_SOS | 0,051006 | 0,055907 | NA |
| 2b | abundance | X01530 | INK (Peptic | Handgrip | 0,046725 | 0,061577 | NA |
| 3 | abundance | X01549 | 3-(Sulfooxy | Tibia_length | 0,063169 | 0,050847 | NA |
| 3 | abundance | X01549 | 3-(Sulfooxy | Radius_length | 0,08732 | 0,069542 | NA |
| 3 | abundance | X01549 | 3-(Sulfooxy | Radius_SOS | 0,016713 | 0,04938 | NA |
| 3 | abundance | X01549 | 3-(Sulfooxy | Tibia_SOS | 0,132145 | 0,05281 | NA |
| 3 | abundance | X01549 | 3-(Sulfooxy | Handgrip | 0,014147 | 0,060687 | NA |
| 3 | abundance | X01553 | uridine 5â€ | Tibia_length | 0,137448 | 0,050815 | NA |
| 3 | abundance | X01553 | uridine 5â€ | Radius_length | 0,175188 | 0,068892 | NA |
| 3 | abundance | X01553 | uridine 5â€ | Radius_SOS | -0,05553 | 0,049271 | NA |
| 3 | abundance | X01553 | uridine 5â€ | Tibia_SOS | 0,074429 | 0,052866 | NA |
| 3 | abundance | X01553 | uridine 5â€ | Handgrip | 0,040102 | 0,060442 | NA |
| 2b | abundance | X01558 | 2-Furoylgly | Tibia_length | -0,11467 | 0,051937 | NA |
| 2b | abundance | X01558 | 2-Furoylgly | Radius_length | -0,09776 | 0,071224 | NA |
| 2b | abundance | X01558 | 2-Furoylgly | Radius_SOS | -0,01483 | 0,050182 | NA |
| 2b | abundance | X01558 | 2-Furoylgly | Tibia_SOS | -0,05127 | 0,053666 | NA |
| 2b | abundance | X01558 | 2-Furoylgly | Handgrip | -0,02656 | 0,062111 | NA |
| 3 | abundance | X01561 | 2-Ammoni | Tibia_length | 0,026999 | 0,055335 | NA |
| 3 | abundance | X01561 | 2-Ammoni | Radius_length | -0,01424 | 0,073665 | NA |
| 3 | abundance | X01561 | 2-Ammoni | Radius_SOS | -0,01145 | 0,052631 | NA |
| 3 | abundance | X01561 | 2-Ammoni | Tibia_SOS | -0,07799 | 0,055944 | NA |
| 3 | abundance | X01561 | 2-Ammoni | Handgrip | -0,10376 | 0,062608 | NA |
| 3 | abundance | X01570 | carglumic a | Tibia_length | -0,08509 | 0,054904 | NA |
| 3 | abundance | X01570 | carglumic a | Radius_length | -0,11397 | 0,072683 | NA |
| 3 | abundance | X01570 | carglumic a | Radius_SOS | -0,091 | 0,051695 | NA |
| 3 | abundance | X01570 | carglumic a | Tibia_SOS | -0,03726 | 0,055726 | NA |
| 3 | abundance | X01570 | carglumic a | Handgrip | -0,00278 | 0,061298 | NA |
| 3 | abundance | X01574 | N-Acetylpr | Tibia_length | -0,08511 | 0,053093 | NA |

| | | | | | | | |
|---|---|---|---|---|---|---|---|
| 3 | abundance | X01574 | N-Acetylpr | Radius_length | -0,12248 | 0,071442 | NA |
| 3 | abundance | X01574 | N-Acetylpr | Radius_SOS | -0,04913 | 0,050765 | NA |
| 3 | abundance | X01574 | N-Acetylpr | Tibia_SOS | -0,11235 | 0,054419 | NA |
| 3 | abundance | X01574 | N-Acetylpr | Handgrip | -0,01369 | 0,062212 | NA |
| 3 | abundance | X01577 | Prunasin | Tibia_length | -0,04182 | 0,051882 | NA |
| 3 | abundance | X01577 | Prunasin | Radius_length | -0,04705 | 0,070834 | NA |
| 3 | abundance | X01577 | Prunasin | Radius_SOS | 0,023705 | 0,049852 | NA |
| 3 | abundance | X01577 | Prunasin | Tibia_SOS | 0,00679 | 0,053443 | NA |
| 3 | abundance | X01577 | Prunasin | Handgrip | -0,01824 | 0,061564 | NA |
| 3 | abundance | X01621 | 2-(2,4-Dihy | Tibia_length | 0,052312 | 0,049559 | NA |
| 3 | abundance | X01621 | 2-(2,4-Dihy | Radius_length | 0,082751 | 0,066965 | NA |
| 3 | abundance | X01621 | 2-(2,4-Dihy | Radius_SOS | -0,04287 | 0,048984 | NA |
| 3 | abundance | X01621 | 2-(2,4-Dihy | Tibia_SOS | 0,023421 | 0,052478 | NA |
| 3 | abundance | X01621 | 2-(2,4-Dihy | Handgrip | 0,115619 | 0,060075 | NA |
| 3 | abundance | X01640 | Ro 20-172 | Tibia_length | -0,12599 | 0,055069 | NA |
| 3 | abundance | X01640 | Ro 20-172 | Radius_length | -0,11441 | 0,073081 | NA |
| 3 | abundance | X01640 | Ro 20-172 | Radius_SOS | -0,02531 | 0,052122 | NA |
| 3 | abundance | X01640 | Ro 20-172 | Tibia_SOS | -0,11716 | 0,055114 | NA |
| 3 | abundance | X01640 | Ro 20-172 | Handgrip | -0,08174 | 0,06215 | NA |
| 3 | abundance | X01656 | Guanadrel | Tibia_length | -0,01112 | 0,051128 | NA |
| 3 | abundance | X01656 | Guanadrel | Radius_length | 0,011884 | 0,070013 | NA |
| 3 | abundance | X01656 | Guanadrel | Radius_SOS | -0,02161 | 0,04953 | NA |
| 3 | abundance | X01656 | Guanadrel | Tibia_SOS | -0,03312 | 0,052913 | NA |
| 3 | abundance | X01656 | Guanadrel | Handgrip | 0,053039 | 0,060771 | NA |
| 3 | abundance | X01664 | 5-methylth | Tibia_length | -0,07678 | 0,051795 | NA |
| 3 | abundance | X01664 | 5-methylth | Radius_length | -0,07958 | 0,070764 | NA |
| 3 | abundance | X01664 | 5-methylth | Radius_SOS | -0,03399 | 0,049877 | NA |
| 3 | abundance | X01664 | 5-methylth | Tibia_SOS | -0,09247 | 0,053705 | NA |
| 3 | abundance | X01664 | 5-methylth | Handgrip | 0,047864 | 0,061738 | NA |
| 3 | abundance | X01671 | N-(3,5-Dim | Tibia_length | -0,08812 | 0,054645 | NA |
| 3 | abundance | X01671 | N-(3,5-Dim | Radius_length | -0,08365 | 0,073509 | NA |
| 3 | abundance | X01671 | N-(3,5-Dim | Radius_SOS | -0,0489 | 0,052026 | NA |
| 3 | abundance | X01671 | N-(3,5-Dim | Tibia_SOS | -0,03375 | 0,055551 | NA |
| 3 | abundance | X01671 | N-(3,5-Dim | Handgrip | -0,06157 | 0,062813 | NA |
| 3 | abundance | X01672 | Triethyl citı | Tibia_length | -0,14076 | 0,054585 | NA |
| 3 | abundance | X01672 | Triethyl citı | Radius_length | -0,12636 | 0,072966 | NA |
| 3 | abundance | X01672 | Triethyl citı | Radius_SOS | -0,07835 | 0,051946 | NA |
| 3 | abundance | X01672 | Triethyl citı | Tibia_SOS | -0,11008 | 0,055898 | NA |
| 3 | abundance | X01672 | Triethyl citı | Handgrip | -0,07004 | 0,061485 | NA |
| 3 | abundance | X01679 | meprobam | Tibia_length | 0,017934 | 0,057298 | NA |
| 3 | abundance | X01679 | meprobam | Radius_length | 0,087596 | 0,074435 | NA |
| 3 | abundance | X01679 | meprobam | Radius_SOS | 0,018521 | 0,054393 | NA |
| 3 | abundance | X01679 | meprobam | Tibia_SOS | -0,02577 | 0,057646 | NA |
| 3 | abundance | X01679 | meprobam | Handgrip | 0,006239 | 0,060192 | NA |
| 3 | abundance | X01689 | Ethyl mala | Tibia_length | 0,056396 | 0,052362 | NA |
| 3 | abundance | X01689 | Ethyl mala | Radius_length | 0,021868 | 0,071396 | NA |
| 3 | abundance | X01689 | Ethyl mala | Radius_SOS | 0,01668 | 0,050326 | NA |
| 3 | abundance | X01689 | Ethyl mala | Tibia_SOS | -0,02917 | 0,054353 | NA |

| | | | | | | | |
|---|---|---|---|---|---|---|---|
| 3 | abundance | X01689 | Ethyl mala | Handgrip | 0,039278 | 0,061128 | NA |
| 3 | abundance | X01732 | 2-Hydroxy- | Tibia_length | -0,11606 | 0,052571 | NA |
| 3 | abundance | X01732 | 2-Hydroxy- | Radius_length | -0,17007 | 0,070815 | NA |
| 3 | abundance | X01732 | 2-Hydroxy- | Radius_SOS | -0,07297 | 0,050511 | NA |
| 3 | abundance | X01732 | 2-Hydroxy- | Tibia_SOS | -0,12418 | 0,053245 | NA |
| 3 | abundance | X01732 | 2-Hydroxy- | Handgrip | -0,07387 | 0,06068 | NA |
| 3 | abundance | X01738 | 2-Acetami | Tibia_length | -0,18372 | 0,054203 | NA |
| 3 | abundance | X01738 | 2-Acetami | Radius_length | -0,05984 | 0,073398 | NA |
| 3 | abundance | X01738 | 2-Acetami | Radius_SOS | -0,0427 | 0,05253 | NA |
| 3 | abundance | X01738 | 2-Acetami | Tibia_SOS | -0,01956 | 0,055707 | NA |
| 3 | abundance | X01738 | 2-Acetami | Handgrip | -0,06475 | 0,061576 | NA |
| 3 | abundance | X01746 | Astemizole | Tibia_length | -0,00959 | 0,057851 | NA |
| 3 | abundance | X01746 | Astemizole | Radius_length | -0,01186 | 0,074936 | NA |
| 3 | abundance | X01746 | Astemizole | Radius_SOS | -0,01936 | 0,055273 | NA |
| 3 | abundance | X01746 | Astemizole | Tibia_SOS | -0,00601 | 0,058094 | NA |
| 3 | abundance | X01746 | Astemizole | Handgrip | -0,04056 | 0,062368 | NA |
| 3 | abundance | X01768 | Butabarbit | Tibia_length | 0,013917 | 0,05131 | NA |
| 3 | abundance | X01768 | Butabarbit | Radius_length | 0,082526 | 0,069915 | NA |
| 3 | abundance | X01768 | Butabarbit | Radius_SOS | 0,056203 | 0,049493 | NA |
| 3 | abundance | X01768 | Butabarbit | Tibia_SOS | 0,068311 | 0,053731 | NA |
| 3 | abundance | X01768 | Butabarbit | Handgrip | -0,05468 | 0,060858 | NA |
| 3 | abundance | X01776 | N-(Carboxy | Tibia_length | -0,14683 | 0,055435 | NA |
| 3 | abundance | X01776 | N-(Carboxy | Radius_length | -0,0691 | 0,074407 | NA |
| 3 | abundance | X01776 | N-(Carboxy | Radius_SOS | -0,07437 | 0,05363 | NA |
| 3 | abundance | X01776 | N-(Carboxy | Tibia_SOS | -0,08411 | 0,054699 | NA |
| 3 | abundance | X01776 | N-(Carboxy | Handgrip | -0,01434 | 0,060989 | NA |
| 3 | abundance | X01793 | Lys-phe | Tibia_length | -0,08272 | 0,054968 | NA |
| 3 | abundance | X01793 | Lys-phe | Radius_length | 0,003764 | 0,073708 | NA |
| 3 | abundance | X01793 | Lys-phe | Radius_SOS | -0,00966 | 0,052228 | NA |
| 3 | abundance | X01793 | Lys-phe | Tibia_SOS | -0,07075 | 0,055832 | NA |
| 3 | abundance | X01793 | Lys-phe | Handgrip | -0,00449 | 0,062955 | NA |
| 3 | abundance | X01813 | Erythorbic | Tibia_length | -0,15959 | 0,056616 | NA |
| 3 | abundance | X01813 | Erythorbic | Radius_length | -0,06688 | 0,075382 | NA |
| 3 | abundance | X01813 | Erythorbic | Radius_SOS | -0,07022 | 0,054704 | NA |
| 3 | abundance | X01813 | Erythorbic | Tibia_SOS | -0,0564 | 0,058677 | NA |
| 3 | abundance | X01813 | Erythorbic | Handgrip | -0,04174 | 0,06014 | NA |
| 2b | abundance | X01833 | 2,3,4,9-Tet | Tibia_length | -0,08836 | 0,052698 | NA |
| 2b | abundance | X01833 | 2,3,4,9-Tet | Radius_length | -0,08456 | 0,071809 | NA |
| 2b | abundance | X01833 | 2,3,4,9-Tet | Radius_SOS | -0,04843 | 0,050535 | NA |
| 2b | abundance | X01833 | 2,3,4,9-Tet | Tibia_SOS | -0,08154 | 0,053673 | NA |
| 2b | abundance | X01833 | 2,3,4,9-Tet | Handgrip | -0,09167 | 0,06083 | NA |
| 3 | abundance | X01873 | Methyl 2,3- | Tibia_length | 0,110015 | 0,053316 | NA |
| 3 | abundance | X01873 | Methyl 2,3- | Radius_length | 0,259503 | 0,070176 | NA |
| 3 | abundance | X01873 | Methyl 2,3- | Radius_SOS | 0,006729 | 0,051117 | NA |
| 3 | abundance | X01873 | Methyl 2,3- | Tibia_SOS | 0,004914 | 0,055321 | NA |
| 3 | abundance | X01873 | Methyl 2,3- | Handgrip | 0,014346 | 0,062362 | NA |
| 2b | abundance | X01879 | Tetramethy | Tibia_length | -0,01311 | 0,053256 | NA |
| 2b | abundance | X01879 | Tetramethy | Radius_length | -0,00189 | 0,072258 | NA |

| | | | | | | | |
|---|---|---|---|---|---|---|---|
| 2b | abundance | X01879 | Tetramethy | Radius_SOS | 0,017281 | 0,050887 | NA |
| 2b | abundance | X01879 | Tetramethy | Tibia_SOS | 0,084343 | 0,053737 | NA |
| 2b | abundance | X01879 | Tetramethy | Handgrip | -0,01617 | 0,061793 | NA |
| 3 | abundance | X01881 | (2S)-3-Met | Tibia_length | -0,14622 | 0,056426 | NA |
| 3 | abundance | X01881 | (2S)-3-Met | Radius_length | -0,06198 | 0,075507 | NA |
| 3 | abundance | X01881 | (2S)-3-Met | Radius_SOS | 0,043216 | 0,054359 | NA |
| 3 | abundance | X01881 | (2S)-3-Met | Tibia_SOS | 0,03017 | 0,059393 | NA |
| 3 | abundance | X01881 | (2S)-3-Met | Handgrip | -0,12546 | 0,062616 | NA |
| 3 | abundance | X01884 | Roxane | Tibia_length | 0,018481 | 0,050295 | NA |
| 3 | abundance | X01884 | Roxane | Radius_length | 0,070468 | 0,068699 | NA |
| 3 | abundance | X01884 | Roxane | Radius_SOS | 0,038314 | 0,049111 | NA |
| 3 | abundance | X01884 | Roxane | Tibia_SOS | 0,119016 | 0,052544 | NA |
| 3 | abundance | X01884 | Roxane | Handgrip | 0,044138 | 0,060251 | NA |
| 3 | abundance | X01887 | lys-tyr | Tibia_length | -0,05453 | 0,054317 | NA |
| 3 | abundance | X01887 | lys-tyr | Radius_length | -0,08801 | 0,073922 | NA |
| 3 | abundance | X01887 | lys-tyr | Radius_SOS | 0,028591 | 0,051542 | NA |
| 3 | abundance | X01887 | lys-tyr | Tibia_SOS | -0,02719 | 0,055034 | NA |
| 3 | abundance | X01887 | lys-tyr | Handgrip | 0,094317 | 0,061248 | NA |
| 3 | abundance | X01893 | 4-(METHYL | Tibia_length | -0,08688 | 0,056806 | NA |
| 3 | abundance | X01893 | 4-(METHYL | Radius_length | -0,18771 | 0,073121 | NA |
| 3 | abundance | X01893 | 4-(METHYL | Radius_SOS | -0,06295 | 0,053674 | NA |
| 3 | abundance | X01893 | 4-(METHYL | Tibia_SOS | -0,12145 | 0,057024 | NA |
| 3 | abundance | X01893 | 4-(METHYL | Handgrip | -0,09029 | 0,061926 | NA |
| 3 | abundance | X01911 | 3-(2,3-Dihy | Tibia_length | -0,12214 | 0,05197 | NA |
| 3 | abundance | X01911 | 3-(2,3-Dihy | Radius_length | -0,17433 | 0,070348 | NA |
| 3 | abundance | X01911 | 3-(2,3-Dihy | Radius_SOS | -0,10667 | 0,049848 | NA |
| 3 | abundance | X01911 | 3-(2,3-Dihy | Tibia_SOS | -0,07604 | 0,053453 | NA |
| 3 | abundance | X01911 | 3-(2,3-Dihy | Handgrip | -0,04161 | 0,060671 | NA |
| 3 | abundance | X01920 | N-(Carboxy | Tibia_length | 0,031708 | 0,053537 | NA |
| 3 | abundance | X01920 | N-(Carboxy | Radius_length | 0,109102 | 0,072193 | NA |
| 3 | abundance | X01920 | N-(Carboxy | Radius_SOS | 0,035366 | 0,051154 | NA |
| 3 | abundance | X01920 | N-(Carboxy | Tibia_SOS | 0,012424 | 0,054182 | NA |
| 3 | abundance | X01920 | N-(Carboxy | Handgrip | -0,04827 | 0,062885 | NA |
| 3 | abundance | X01932 | asn-val | Tibia_length | 0,167148 | 0,054909 | NA |
| 3 | abundance | X01932 | asn-val | Radius_length | 0,163207 | 0,073133 | NA |
| 3 | abundance | X01932 | asn-val | Radius_SOS | 0,081928 | 0,052962 | NA |
| 3 | abundance | X01932 | asn-val | Tibia_SOS | 0,101887 | 0,05825 | NA |
| 3 | abundance | X01932 | asn-val | Handgrip | 0,064987 | 0,062567 | NA |
| 3 | abundance | X01943 | Tocainide | Tibia_length | 0,08615 | 0,049208 | NA |
| 3 | abundance | X01943 | Tocainide | Radius_length | 0,118993 | 0,065955 | NA |
| 3 | abundance | X01943 | Tocainide | Radius_SOS | 0,005509 | 0,049048 | NA |
| 3 | abundance | X01943 | Tocainide | Tibia_SOS | 0,077389 | 0,052355 | NA |
| 3 | abundance | X01943 | Tocainide | Handgrip | 0,010736 | 0,060234 | NA |
| 3 | abundance | X01950 | Histidylgly | Tibia_length | -0,06136 | 0,052937 | NA |
| 3 | abundance | X01950 | Histidylgly | Radius_length | -0,06685 | 0,071705 | NA |
| 3 | abundance | X01950 | Histidylgly | Radius_SOS | 0,072025 | 0,050592 | NA |
| 3 | abundance | X01950 | Histidylgly | Tibia_SOS | -0,03607 | 0,054673 | NA |
| 3 | abundance | X01950 | Histidylgly | Handgrip | -0,05083 | 0,061816 | NA |

| | | | | | | | |
|---|---|---|---|---|---|---|---|
| 3 | abundance | X01968 | 7-Chloro-5 | Tibia_length | -0,06924 | 0,057515 | NA |
| 3 | abundance | X01968 | 7-Chloro-5 | Radius_length | -0,03628 | 0,075365 | NA |
| 3 | abundance | X01968 | 7-Chloro-5 | Radius_SOS | -0,07539 | 0,054927 | NA |
| 3 | abundance | X01968 | 7-Chloro-5 | Tibia_SOS | 0,034944 | 0,058681 | NA |
| 3 | abundance | X01968 | 7-Chloro-5 | Handgrip | 0,009074 | 0,0634 | NA |
| 3 | abundance | X01996 | 3-Hydroxy- | Tibia_length | 0,121803 | 0,054772 | NA |
| 3 | abundance | X01996 | 3-Hydroxy- | Radius_length | 0,14509 | 0,072935 | NA |
| 3 | abundance | X01996 | 3-Hydroxy- | Radius_SOS | 0,043899 | 0,052462 | NA |
| 3 | abundance | X01996 | 3-Hydroxy- | Tibia_SOS | 0,071474 | 0,055592 | NA |
| 3 | abundance | X01996 | 3-Hydroxy- | Handgrip | 0,018997 | 0,062948 | NA |
| 3 | abundance | X02000 | 11-(4-Hydr | Tibia_length | 0,025582 | 0,053667 | NA |
| 3 | abundance | X02000 | 11-(4-Hydr | Radius_length | 0,105237 | 0,072028 | NA |
| 3 | abundance | X02000 | 11-(4-Hydr | Radius_SOS | 0,040772 | 0,051164 | NA |
| 3 | abundance | X02000 | 11-(4-Hydr | Tibia_SOS | 0,0473 | 0,054041 | NA |
| 3 | abundance | X02000 | 11-(4-Hydr | Handgrip | 0,064741 | 0,062129 | NA |
| 3 | abundance | X02009 | Arctiopicri | Tibia_length | -0,01382 | 0,054493 | NA |
| 3 | abundance | X02009 | Arctiopicri | Radius_length | -0,06599 | 0,073504 | NA |
| 3 | abundance | X02009 | Arctiopicri | Radius_SOS | -0,0142 | 0,051926 | NA |
| 3 | abundance | X02009 | Arctiopicri | Tibia_SOS | 0,068871 | 0,054301 | NA |
| 3 | abundance | X02009 | Arctiopicri | Handgrip | 0,074915 | 0,061034 | NA |
| 3 | abundance | X02013 | Ile-cys | Tibia_length | 0,078473 | 0,055803 | NA |
| 3 | abundance | X02013 | Ile-cys | Radius_length | 0,139657 | 0,07361 | NA |
| 3 | abundance | X02013 | Ile-cys | Radius_SOS | 0,0192 | 0,053295 | NA |
| 3 | abundance | X02013 | Ile-cys | Tibia_SOS | 0,081101 | 0,056153 | NA |
| 3 | abundance | X02013 | Ile-cys | Handgrip | 0,104265 | 0,061327 | NA |
| 3 | abundance | X02020 | Val-Ser | Tibia_length | -0,07217 | 0,056216 | NA |
| 3 | abundance | X02020 | Val-Ser | Radius_length | -0,0126 | 0,074629 | NA |
| 3 | abundance | X02020 | Val-Ser | Radius_SOS | -0,02578 | 0,053619 | NA |
| 3 | abundance | X02020 | Val-Ser | Tibia_SOS | 0,004616 | 0,058602 | NA |
| 3 | abundance | X02020 | Val-Ser | Handgrip | 0,01408 | 0,063281 | NA |
| 3 | abundance | X02023 | beta-D-Eth | Tibia_length | -0,15486 | 0,053845 | NA |
| 3 | abundance | X02023 | beta-D-Eth | Radius_length | -0,11068 | 0,073458 | NA |
| 3 | abundance | X02023 | beta-D-Eth | Radius_SOS | -0,06666 | 0,051731 | NA |
| 3 | abundance | X02023 | beta-D-Eth | Tibia_SOS | -0,01098 | 0,056356 | NA |
| 3 | abundance | X02023 | beta-D-Eth | Handgrip | -0,13196 | 0,061754 | NA |
| 3 | abundance | X02073 | N-acetyl-9 | Tibia_length | -0,14716 | 0,055966 | NA |
| 3 | abundance | X02073 | N-acetyl-9 | Radius_length | -0,02048 | 0,074895 | NA |
| 3 | abundance | X02073 | N-acetyl-9 | Radius_SOS | -0,07341 | 0,053995 | NA |
| 3 | abundance | X02073 | N-acetyl-9 | Tibia_SOS | -0,02239 | 0,056757 | NA |
| 3 | abundance | X02073 | N-acetyl-9 | Handgrip | 0,09984 | 0,062587 | NA |
| 3 | abundance | X02080 | (-)-Aspidos | Tibia_length | 0,080444 | 0,054789 | NA |
| 3 | abundance | X02080 | (-)-Aspidos | Radius_length | 0,143989 | 0,072634 | NA |
| 3 | abundance | X02080 | (-)-Aspidos | Radius_SOS | -0,05983 | 0,052182 | NA |
| 3 | abundance | X02080 | (-)-Aspidos | Tibia_SOS | 0,014734 | 0,055026 | NA |
| 3 | abundance | X02080 | (-)-Aspidos | Handgrip | 0,006147 | 0,061795 | NA |
| 3 | abundance | X02082 | 6-Hydroxyr | Tibia_length | 0,009107 | 0,052499 | NA |
| 3 | abundance | X02082 | 6-Hydroxyr | Radius_length | -0,04453 | 0,071417 | NA |
| 3 | abundance | X02082 | 6-Hydroxyr | Radius_SOS | -0,04103 | 0,050368 | NA |

| | | | | | | | |
|---|---|---|---|---|---|---|---|
| 3 | abundance | X02082 | 6-Hydroxyr | Tibia_SOS | -0,05974 | 0,053077 | NA |
| 3 | abundance | X02082 | 6-Hydroxyr | Handgrip | 0,009284 | 0,060462 | NA |
| 3 | abundance | X02101 | Succinic a | Tibia_length | 0,024177 | 0,056143 | NA |
| 3 | abundance | X02101 | Succinic a | Radius_length | 0,038431 | 0,074398 | NA |
| 3 | abundance | X02101 | Succinic a | Radius_SOS | 0,058141 | 0,053093 | NA |
| 3 | abundance | X02101 | Succinic a | Tibia_SOS | -0,01872 | 0,056521 | NA |
| 3 | abundance | X02101 | Succinic a | Handgrip | 0,139487 | 0,061669 | NA |
| 3 | abundance | X02108 | Homovanil | Tibia_length | -0,00819 | 0,053219 | NA |
| 3 | abundance | X02108 | Homovanil | Radius_length | -0,05175 | 0,072452 | NA |
| 3 | abundance | X02108 | Homovanil | Radius_SOS | -0,04183 | 0,051104 | NA |
| 3 | abundance | X02108 | Homovanil | Tibia_SOS | -0,05205 | 0,054602 | NA |
| 3 | abundance | X02108 | Homovanil | Handgrip | 0,014549 | 0,061314 | NA |
| 3 | abundance | X02121 | 2-Acetami | Tibia_length | -0,03059 | 0,053029 | NA |
| 3 | abundance | X02121 | 2-Acetami | Radius_length | 0,015614 | 0,072109 | NA |
| 3 | abundance | X02121 | 2-Acetami | Radius_SOS | -0,04236 | 0,050664 | NA |
| 3 | abundance | X02121 | 2-Acetami | Tibia_SOS | -0,06091 | 0,053706 | NA |
| 3 | abundance | X02121 | 2-Acetami | Handgrip | 0,025119 | 0,06113 | NA |
| 3 | abundance | X02139 | SECONAL | Tibia_length | -0,03225 | 0,053406 | NA |
| 3 | abundance | X02139 | SECONAL | Radius_length | -0,07004 | 0,072028 | NA |
| 3 | abundance | X02139 | SECONAL | Radius_SOS | 0,060401 | 0,050882 | NA |
| 3 | abundance | X02139 | SECONAL | Tibia_SOS | 0,010041 | 0,053995 | NA |
| 3 | abundance | X02139 | SECONAL | Handgrip | -0,07488 | 0,061328 | NA |
| 3 | abundance | X02145 | 3'-Hydroxy | Tibia_length | 0,062849 | 0,051437 | NA |
| 3 | abundance | X02145 | 3'-Hydroxy | Radius_length | 0,055526 | 0,070052 | NA |
| 3 | abundance | X02145 | 3'-Hydroxy | Radius_SOS | 0,012226 | 0,049665 | NA |
| 3 | abundance | X02145 | 3'-Hydroxy | Tibia_SOS | 0,04347 | 0,053649 | NA |
| 3 | abundance | X02145 | 3'-Hydroxy | Handgrip | -0,09567 | 0,060241 | NA |
| 3 | abundance | X02153 | Asarone | Tibia_length | -0,08039 | 0,053656 | NA |
| 3 | abundance | X02153 | Asarone | Radius_length | -0,20268 | 0,071397 | NA |
| 3 | abundance | X02153 | Asarone | Radius_SOS | -0,0153 | 0,051221 | NA |
| 3 | abundance | X02153 | Asarone | Tibia_SOS | -0,15314 | 0,053742 | NA |
| 3 | abundance | X02153 | Asarone | Handgrip | -0,09254 | 0,061697 | NA |
| 3 | abundance | X02181 | YWA1 | Tibia_length | 0,112554 | 0,049688 | NA |
| 3 | abundance | X02181 | YWA1 | Radius_length | 0,24793 | 0,066019 | NA |
| 3 | abundance | X02181 | YWA1 | Radius_SOS | 0,04695 | 0,048984 | NA |
| 3 | abundance | X02181 | YWA1 | Tibia_SOS | 0,119649 | 0,052111 | NA |
| 3 | abundance | X02181 | YWA1 | Handgrip | 0,138754 | 0,059747 | NA |
| 2b | abundance | X02184 | Piperonylo | Tibia_length | 0,051869 | 0,053314 | NA |
| 2b | abundance | X02184 | Piperonylo | Radius_length | -0,00403 | 0,072736 | NA |
| 2b | abundance | X02184 | Piperonylo | Radius_SOS | -0,05133 | 0,050896 | NA |
| 2b | abundance | X02184 | Piperonylo | Tibia_SOS | 0,09819 | 0,05411 | NA |
| 2b | abundance | X02184 | Piperonylo | Handgrip | -0,01987 | 0,061442 | NA |
| 3 | abundance | X02195 | alliin | Tibia_length | -0,04466 | 0,053803 | NA |
| 3 | abundance | X02195 | alliin | Radius_length | -0,02589 | 0,072677 | NA |
| 3 | abundance | X02195 | alliin | Radius_SOS | 0,052694 | 0,051269 | NA |
| 3 | abundance | X02195 | alliin | Tibia_SOS | 0,01807 | 0,054705 | NA |
| 3 | abundance | X02195 | alliin | Handgrip | -0,04438 | 0,061102 | NA |
| 3 | abundance | X02202 | D-2-Amino | Tibia_length | 0,051411 | 0,056147 | NA |

| | | | | | | | |
|---|---|---|---|---|---|---|---|
| 3 | abundance | X02202 | D-2-Amino | Radius_length | -0,0446 | 0,074331 | NA |
| 3 | abundance | X02202 | D-2-Amino | Radius_SOS | 0,020755 | 0,053372 | NA |
| 3 | abundance | X02202 | D-2-Amino | Tibia_SOS | 0,053971 | 0,057329 | NA |
| 3 | abundance | X02202 | D-2-Amino | Handgrip | 0,086933 | 0,06233 | NA |
| 3 | abundance | X02208 | SECONAL | Tibia_length | -0,04648 | 0,05843 | NA |
| 3 | abundance | X02208 | SECONAL | Radius_length | -0,08612 | 0,075492 | NA |
| 3 | abundance | X02208 | SECONAL | Radius_SOS | 0,053221 | 0,055595 | NA |
| 3 | abundance | X02208 | SECONAL | Tibia_SOS | -0,04937 | 0,058605 | NA |
| 3 | abundance | X02208 | SECONAL | Handgrip | 0,012048 | 0,060189 | NA |
| 3 | abundance | X02214 | (E)-4-Meth | Tibia_length | -0,06352 | 0,055463 | NA |
| 3 | abundance | X02214 | (E)-4-Meth | Radius_length | -0,0095 | 0,073713 | NA |
| 3 | abundance | X02214 | (E)-4-Meth | Radius_SOS | 1,82E-04 | 0,052915 | NA |
| 3 | abundance | X02214 | (E)-4-Meth | Tibia_SOS | 0,029821 | 0,055204 | NA |
| 3 | abundance | X02214 | (E)-4-Meth | Handgrip | -0,01635 | 0,061594 | NA |
| 3 | abundance | X02219 | Homoanse | Tibia_length | -0,09508 | 0,0538 | NA |
| 3 | abundance | X02219 | Homoanse | Radius_length | -0,14728 | 0,072133 | NA |
| 3 | abundance | X02219 | Homoanse | Radius_SOS | -0,04199 | 0,051443 | NA |
| 3 | abundance | X02219 | Homoanse | Tibia_SOS | -0,11015 | 0,053746 | NA |
| 3 | abundance | X02219 | Homoanse | Handgrip | -0,07574 | 0,061506 | NA |
| 3 | abundance | X02231 | Ala-Tyr | Tibia_length | -0,12612 | 0,052192 | NA |
| 3 | abundance | X02231 | Ala-Tyr | Radius_length | -0,18055 | 0,070678 | NA |
| 3 | abundance | X02231 | Ala-Tyr | Radius_SOS | -0,05059 | 0,050179 | NA |
| 3 | abundance | X02231 | Ala-Tyr | Tibia_SOS | -0,07477 | 0,054157 | NA |
| 3 | abundance | X02231 | Ala-Tyr | Handgrip | -0,01853 | 0,061596 | NA |
| 3 | abundance | X02238 | 6-Myoporo | Tibia_length | 0,023302 | 0,053292 | NA |
| 3 | abundance | X02238 | 6-Myoporo | Radius_length | 0,050495 | 0,071953 | NA |
| 3 | abundance | X02238 | 6-Myoporo | Radius_SOS | -0,0367 | 0,050915 | NA |
| 3 | abundance | X02238 | 6-Myoporo | Tibia_SOS | 0,020407 | 0,053701 | NA |
| 3 | abundance | X02238 | 6-Myoporo | Handgrip | -0,03848 | 0,06175 | NA |
| 3 | abundance | X02251 | Hostmania | Tibia_length | -0,06904 | 0,052451 | NA |
| 3 | abundance | X02251 | Hostmania | Radius_length | -0,09689 | 0,071472 | NA |
| 3 | abundance | X02251 | Hostmania | Radius_SOS | -0,00717 | 0,050241 | NA |
| 3 | abundance | X02251 | Hostmania | Tibia_SOS | -0,13804 | 0,053742 | NA |
| 3 | abundance | X02251 | Hostmania | Handgrip | -0,12963 | 0,060585 | NA |
| 3 | abundance | X02256 | hexobarbit | Tibia_length | 0,015655 | 0,049747 | NA |
| 3 | abundance | X02256 | hexobarbit | Radius_length | 0,15493 | 0,066713 | NA |
| 3 | abundance | X02256 | hexobarbit | Radius_SOS | 0,031481 | 0,049021 | NA |
| 3 | abundance | X02256 | hexobarbit | Tibia_SOS | 0,005521 | 0,052489 | NA |
| 3 | abundance | X02256 | hexobarbit | Handgrip | 0,043801 | 0,060633 | NA |
| 3 | abundance | X02265 | ophthalmic | Tibia_length | -0,17397 | 0,053639 | NA |
| 3 | abundance | X02265 | ophthalmic | Radius_length | -0,08823 | 0,07288 | NA |
| 3 | abundance | X02265 | ophthalmic | Radius_SOS | -0,02568 | 0,051511 | NA |
| 3 | abundance | X02265 | ophthalmic | Tibia_SOS | -5,76E-04 | 0,055468 | NA |
| 3 | abundance | X02265 | ophthalmic | Handgrip | -0,04084 | 0,061901 | NA |
| 3 | abundance | X02268 | 5-Hydroxy- | Tibia_length | -0,05708 | 0,051551 | NA |
| 3 | abundance | X02268 | 5-Hydroxy- | Radius_length | -0,01356 | 0,070451 | NA |
| 3 | abundance | X02268 | 5-Hydroxy- | Radius_SOS | 0,079973 | 0,049571 | NA |
| 3 | abundance | X02268 | 5-Hydroxy- | Tibia_SOS | -0,10435 | 0,052759 | NA |

| | | | | | | | | |
|---|---|---|---|---|---|---|---|---|
| | 3 | abundance | X02268 | 5-Hydroxy- | Handgrip | -0,10543 | 0,060778 | NA |
| | 3 | abundance | X02277 | tert-Butyl 3 | Tibia_length | -0,02862 | 0,053253 | NA |
| | 3 | abundance | X02277 | tert-Butyl 3 | Radius_length | 0,025641 | 0,072765 | NA |
| | 3 | abundance | X02277 | tert-Butyl 3 | Radius_SOS | -0,01314 | 0,050893 | NA |
| | 3 | abundance | X02277 | tert-Butyl 3 | Tibia_SOS | -0,00507 | 0,054783 | NA |
| | 3 | abundance | X02277 | tert-Butyl 3 | Handgrip | 0,012367 | 0,062003 | NA |
| | 3 | abundance | X02281 | tert-Butyl 3 | Tibia_length | 0,030353 | 0,052089 | NA |
| | 3 | abundance | X02281 | tert-Butyl 3 | Radius_length | 0,031728 | 0,071053 | NA |
| | 3 | abundance | X02281 | tert-Butyl 3 | Radius_SOS | -0,01533 | 0,050062 | NA |
| | 3 | abundance | X02281 | tert-Butyl 3 | Tibia_SOS | -0,02954 | 0,053542 | NA |
| | 3 | abundance | X02281 | tert-Butyl 3 | Handgrip | -0,01153 | 0,061071 | NA |
| 2b | | abundance | X02288 | 9-Methylur | Tibia_length | -0,14671 | 0,056462 | NA |
| 2b | | abundance | X02288 | 9-Methylur | Radius_length | -0,10514 | 0,074072 | NA |
| 2b | | abundance | X02288 | 9-Methylur | Radius_SOS | 0,01269 | 0,053995 | NA |
| 2b | | abundance | X02288 | 9-Methylur | Tibia_SOS | 0,076223 | 0,057148 | NA |
| 2b | | abundance | X02288 | 9-Methylur | Handgrip | -0,10031 | 0,062717 | NA |
| 2a | | abundance | X02289 | Kynurenic a | Tibia_length | -0,10557 | 0,04888 | NA |
| 2a | | abundance | X02289 | Kynurenic a | Radius_length | -0,10922 | 0,065245 | NA |
| 2a | | abundance | X02289 | Kynurenic a | Radius_SOS | -0,0783 | 0,048961 | NA |
| 2a | | abundance | X02289 | Kynurenic a | Tibia_SOS | 0,012957 | 0,0526 | NA |
| 2a | | abundance | X02289 | Kynurenic a | Handgrip | 0,02109 | 0,060252 | NA |
| 2b | | abundance | X02317 | 2-Aminooc | Tibia_length | -0,01079 | 0,055935 | NA |
| 2b | | abundance | X02317 | 2-Aminooc | Radius_length | 0,066563 | 0,073788 | NA |
| 2b | | abundance | X02317 | 2-Aminooc | Radius_SOS | -0,06465 | 0,052994 | NA |
| 2b | | abundance | X02317 | 2-Aminooc | Tibia_SOS | -0,06216 | 0,056634 | NA |
| 2b | | abundance | X02317 | 2-Aminooc | Handgrip | -0,05407 | 0,063482 | NA |
| | 3 | abundance | X02327 | 1H-Pyrazol | Tibia_length | -0,0143 | 0,058277 | NA |
| | 3 | abundance | X02327 | 1H-Pyrazol | Radius_length | -0,10351 | 0,075531 | NA |
| | 3 | abundance | X02327 | 1H-Pyrazol | Radius_SOS | 0,047007 | 0,055454 | NA |
| | 3 | abundance | X02327 | 1H-Pyrazol | Tibia_SOS | 0,014322 | 0,060023 | NA |
| | 3 | abundance | X02327 | 1H-Pyrazol | Handgrip | 0,028476 | 0,062937 | NA |
| | 3 | abundance | X02333 | N-Propiony | Tibia_length | 0,017904 | 0,053921 | NA |
| | 3 | abundance | X02333 | N-Propiony | Radius_length | 0,130536 | 0,072014 | NA |
| | 3 | abundance | X02333 | N-Propiony | Radius_SOS | 0,052428 | 0,051265 | NA |
| | 3 | abundance | X02333 | N-Propiony | Tibia_SOS | 0,07829 | 0,054702 | NA |
| | 3 | abundance | X02333 | N-Propiony | Handgrip | -0,00515 | 0,061316 | NA |
| | 3 | abundance | X02337 | Methyl 1-hy | Tibia_length | -0,11224 | 0,050628 | NA |
| | 3 | abundance | X02337 | Methyl 1-hy | Radius_length | -0,06615 | 0,06984 | NA |
| | 3 | abundance | X02337 | Methyl 1-hy | Radius_SOS | -0,01416 | 0,04943 | NA |
| | 3 | abundance | X02337 | Methyl 1-hy | Tibia_SOS | 0,067664 | 0,052964 | NA |
| | 3 | abundance | X02337 | Methyl 1-hy | Handgrip | -0,04072 | 0,061341 | NA |
| | 3 | abundance | X02348 | N-Benzoyla | Tibia_length | 0,093142 | 0,054694 | NA |
| | 3 | abundance | X02348 | N-Benzoyla | Radius_length | 0,059293 | 0,07406 | NA |
| | 3 | abundance | X02348 | N-Benzoyla | Radius_SOS | 0,006154 | 0,052023 | NA |
| | 3 | abundance | X02348 | N-Benzoyla | Tibia_SOS | 0,088237 | 0,057373 | NA |
| | 3 | abundance | X02348 | N-Benzoyla | Handgrip | 0,027193 | 0,061823 | NA |
| | 3 | abundance | X02352 | Indole-3-ca | Tibia_length | 0,001395 | 0,055199 | NA |
| | 3 | abundance | X02352 | Indole-3-ca | Radius_length | -0,03666 | 0,073275 | NA |

| | | | | | | | | |
|---|---|---|---|---|---|---|---|---|
| | 3 | abundance | X02352 | Indole-3-ca | Radius_SOS | 0,045118 | 0,052352 | NA |
| | 3 | abundance | X02352 | Indole-3-ca | Tibia_SOS | 0,040251 | 0,056113 | NA |
| | 3 | abundance | X02352 | Indole-3-ca | Handgrip | -0,01446 | 0,063191 | NA |
| | 3 | abundance | X02380 | Propafenon | Tibia_length | 0,124264 | 0,051538 | NA |
| | 3 | abundance | X02380 | Propafenon | Radius_length | 0,177323 | 0,070085 | NA |
| | 3 | abundance | X02380 | Propafenon | Radius_SOS | 0,039074 | 0,049778 | NA |
| | 3 | abundance | X02380 | Propafenon | Tibia_SOS | 0,050736 | 0,053615 | NA |
| | 3 | abundance | X02380 | Propafenon | Handgrip | 0,103024 | 0,060923 | NA |
| | 3 | abundance | X02419 | Glu-Glu | Tibia_length | -0,00791 | 0,053471 | NA |
| | 3 | abundance | X02419 | Glu-Glu | Radius_length | -0,01022 | 0,073002 | NA |
| | 3 | abundance | X02419 | Glu-Glu | Radius_SOS | -0,04692 | 0,050886 | NA |
| | 3 | abundance | X02419 | Glu-Glu | Tibia_SOS | -0,06634 | 0,054734 | NA |
| | 3 | abundance | X02419 | Glu-Glu | Handgrip | -0,12076 | 0,061556 | NA |
| | 3 | abundance | X02426 | Raltitrexed | Tibia_length | -0,15965 | 0,051939 | NA |
| | 3 | abundance | X02426 | Raltitrexed | Radius_length | -0,06655 | 0,071658 | NA |
| | 3 | abundance | X02426 | Raltitrexed | Radius_SOS | 0,022428 | 0,050516 | NA |
| | 3 | abundance | X02426 | Raltitrexed | Tibia_SOS | 0,027542 | 0,053171 | NA |
| | 3 | abundance | X02426 | Raltitrexed | Handgrip | -0,04309 | 0,060719 | NA |
| 2b | | abundance | X02427 | 2-(3,5-dim | Tibia_length | 0,092884 | 0,056403 | NA |
| 2b | | abundance | X02427 | 2-(3,5-dim | Radius_length | 0,007768 | 0,074353 | NA |
| 2b | | abundance | X02427 | 2-(3,5-dim | Radius_SOS | -0,01103 | 0,053829 | NA |
| 2b | | abundance | X02427 | 2-(3,5-dim | Tibia_SOS | 0,004483 | 0,058162 | NA |
| 2b | | abundance | X02427 | 2-(3,5-dim | Handgrip | -0,11532 | 0,059791 | NA |
| | 3 | abundance | X02442 | 3-(Sulfooxy | Tibia_length | 0,044114 | 0,052294 | NA |
| | 3 | abundance | X02442 | 3-(Sulfooxy | Radius_length | 0,104683 | 0,071019 | NA |
| | 3 | abundance | X02442 | 3-(Sulfooxy | Radius_SOS | 0,010619 | 0,050202 | NA |
| | 3 | abundance | X02442 | 3-(Sulfooxy | Tibia_SOS | 0,150246 | 0,05333 | NA |
| | 3 | abundance | X02442 | 3-(Sulfooxy | Handgrip | 0,005698 | 0,061036 | NA |
| | 3 | abundance | X02466 | Validamyci | Tibia_length | 0,047329 | 0,056491 | NA |
| | 3 | abundance | X02466 | Validamyci | Radius_length | 0,128316 | 0,074246 | NA |
| | 3 | abundance | X02466 | Validamyci | Radius_SOS | 0,035619 | 0,053617 | NA |
| | 3 | abundance | X02466 | Validamyci | Tibia_SOS | 0,022109 | 0,057757 | NA |
| | 3 | abundance | X02466 | Validamyci | Handgrip | -0,05195 | 0,061854 | NA |
| | 3 | abundance | X02468 | N,N-Diethy | Tibia_length | -0,04671 | 0,054255 | NA |
| | 3 | abundance | X02468 | N,N-Diethy | Radius_length | -0,03309 | 0,072858 | NA |
| | 3 | abundance | X02468 | N,N-Diethy | Radius_SOS | -0,0385 | 0,051621 | NA |
| | 3 | abundance | X02468 | N,N-Diethy | Tibia_SOS | 0,053645 | 0,055113 | NA |
| | 3 | abundance | X02468 | N,N-Diethy | Handgrip | -0,0407 | 0,061805 | NA |
| | 3 | abundance | X02471 | n-Propyl Ga | Tibia_length | -0,0144 | 0,051552 | NA |
| | 3 | abundance | X02471 | n-Propyl Ga | Radius_length | -0,18216 | 0,069508 | NA |
| | 3 | abundance | X02471 | n-Propyl Ga | Radius_SOS | -0,00479 | 0,049751 | NA |
| | 3 | abundance | X02471 | n-Propyl Ga | Tibia_SOS | -0,07744 | 0,054286 | NA |
| | 3 | abundance | X02471 | n-Propyl Ga | Handgrip | -0,06583 | 0,060905 | NA |
| | 3 | abundance | X02492 | N-(3,5-Dim | Tibia_length | -0,14187 | 0,054316 | NA |
| | 3 | abundance | X02492 | N-(3,5-Dim | Radius_length | -0,08766 | 0,073227 | NA |
| | 3 | abundance | X02492 | N-(3,5-Dim | Radius_SOS | -0,03602 | 0,052032 | NA |
| | 3 | abundance | X02492 | N-(3,5-Dim | Tibia_SOS | -0,07963 | 0,055201 | NA |
| | 3 | abundance | X02492 | N-(3,5-Dim | Handgrip | -0,12844 | 0,061556 | NA |

| | | | | | | | | |
|---|---|---|---|---|---|---|---|---|
| | 3 | abundance | X02494 | Butylphtha | Tibia_length | -0,07335 | 0,052971 | NA |
| | 3 | abundance | X02494 | Butylphtha | Radius_length | -0,18732 | 0,070962 | NA |
| | 3 | abundance | X02494 | Butylphtha | Radius_SOS | -0,00811 | 0,050708 | NA |
| | 3 | abundance | X02494 | Butylphtha | Tibia_SOS | -0,15785 | 0,053248 | NA |
| | 3 | abundance | X02494 | Butylphtha | Handgrip | -0,09127 | 0,061313 | NA |
| 2b | | abundance | X02504 | DLK (Pepti | Tibia_length | -0,07466 | 0,052597 | NA |
| 2b | | abundance | X02504 | DLK (Pepti | Radius_length | 0,046727 | 0,072082 | NA |
| 2b | | abundance | X02504 | DLK (Pepti | Radius_SOS | 0,066062 | 0,050578 | NA |
| 2b | | abundance | X02504 | DLK (Pepti | Tibia_SOS | 0,119891 | 0,053898 | NA |
| 2b | | abundance | X02504 | DLK (Pepti | Handgrip | 0,080129 | 0,061929 | NA |
| 2b | | abundance | X02513 | 7-Methylac | Tibia_length | -0,02498 | 0,052393 | NA |
| 2b | | abundance | X02513 | 7-Methylac | Radius_length | 0,018212 | 0,071481 | NA |
| 2b | | abundance | X02513 | 7-Methylac | Radius_SOS | 0,012584 | 0,050223 | NA |
| 2b | | abundance | X02513 | 7-Methylac | Tibia_SOS | 0,029887 | 0,054 | NA |
| 2b | | abundance | X02513 | 7-Methylac | Handgrip | 0,026987 | 0,061669 | NA |
| | 1 | abundance | X02514 | 3-Ureidopr | Tibia_length | -0,015 | 0,054201 | NA |
| | 1 | abundance | X02514 | 3-Ureidopr | Radius_length | -0,06923 | 0,072818 | NA |
| | 1 | abundance | X02514 | 3-Ureidopr | Radius_SOS | -0,06993 | 0,052006 | NA |
| | 1 | abundance | X02514 | 3-Ureidopr | Tibia_SOS | -0,00672 | 0,055436 | NA |
| | 1 | abundance | X02514 | 3-Ureidopr | Handgrip | 0,049732 | 0,062687 | NA |
| | 3 | abundance | X02521 | 6-(1-Hydro | Tibia_length | -0,12475 | 0,054706 | NA |
| | 3 | abundance | X02521 | 6-(1-Hydro | Radius_length | -0,1505 | 0,072863 | NA |
| | 3 | abundance | X02521 | 6-(1-Hydro | Radius_SOS | -0,01699 | 0,052055 | NA |
| | 3 | abundance | X02521 | 6-(1-Hydro | Tibia_SOS | -0,02797 | 0,054746 | NA |
| | 3 | abundance | X02521 | 6-(1-Hydro | Handgrip | -0,16119 | 0,060542 | NA |
| | 3 | abundance | X02529 | tenivastati | Tibia_length | 0,098563 | 0,05171 | NA |
| | 3 | abundance | X02529 | tenivastati | Radius_length | 0,047686 | 0,070823 | NA |
| | 3 | abundance | X02529 | tenivastati | Radius_SOS | -0,00633 | 0,049776 | NA |
| | 3 | abundance | X02529 | tenivastati | Tibia_SOS | 0,027179 | 0,053267 | NA |
| | 3 | abundance | X02529 | tenivastati | Handgrip | -0,07728 | 0,060993 | NA |
| | 3 | abundance | X02537 | Leu-pro | Tibia_length | -0,19564 | 0,053089 | NA |
| | 3 | abundance | X02537 | Leu-pro | Radius_length | -0,2295 | 0,0714 | NA |
| | 3 | abundance | X02537 | Leu-pro | Radius_SOS | -0,00644 | 0,051217 | NA |
| | 3 | abundance | X02537 | Leu-pro | Tibia_SOS | -0,03867 | 0,054344 | NA |
| | 3 | abundance | X02537 | Leu-pro | Handgrip | -0,19467 | 0,060664 | NA |
| | 3 | abundance | X02565 | Dopamine | Tibia_length | -0,09645 | 0,059184 | NA |
| | 3 | abundance | X02565 | Dopamine | Radius_length | 0,019037 | 0,076919 | NA |
| | 3 | abundance | X02565 | Dopamine | Radius_SOS | -0,03767 | 0,056866 | NA |
| | 3 | abundance | X02565 | Dopamine | Tibia_SOS | -0,13218 | 0,059107 | NA |
| | 3 | abundance | X02565 | Dopamine | Handgrip | -0,06153 | 0,060079 | NA |
| | 3 | abundance | X02583 | Ethylvanilli | Tibia_length | -0,02993 | 0,054739 | NA |
| | 3 | abundance | X02583 | Ethylvanilli | Radius_length | 0,029025 | 0,073274 | NA |
| | 3 | abundance | X02583 | Ethylvanilli | Radius_SOS | -0,05308 | 0,052011 | NA |
| | 3 | abundance | X02583 | Ethylvanilli | Tibia_SOS | 9,37E-04 | 0,054857 | NA |
| | 3 | abundance | X02583 | Ethylvanilli | Handgrip | -0,04157 | 0,061747 | NA |
| | 3 | abundance | X02599 | Dihydrothy | Tibia_length | 0,100245 | 0,054922 | NA |
| | 3 | abundance | X02599 | Dihydrothy | Radius_length | -0,05236 | 0,073925 | NA |
| | 3 | abundance | X02599 | Dihydrothy | Radius_SOS | -0,01774 | 0,052514 | NA |

| | | | | | | | |
|---|---|---|---|---|---|---|---|
| 3 | abundance | X02599 | Dihydrothy | Tibia_SOS | 0,112013 | 0,055555 | NA |
| 3 | abundance | X02599 | Dihydrothy | Handgrip | -0,02453 | 0,061423 | NA |
| 3 | abundance | X02601 | (7E,7'E)-5, | Tibia_length | 0,017123 | 0,054236 | NA |
| 3 | abundance | X02601 | (7E,7'E)-5, | Radius_length | 0,145194 | 0,07242 | NA |
| 3 | abundance | X02601 | (7E,7'E)-5, | Radius_SOS | 0,01257 | 0,051632 | NA |
| 3 | abundance | X02601 | (7E,7'E)-5, | Tibia_SOS | -0,00816 | 0,054453 | NA |
| 3 | abundance | X02601 | (7E,7'E)-5, | Handgrip | 0,067601 | 0,061725 | NA |
| 3 | abundance | X02606 | 4-Amino-1 | Tibia_length | 0,039518 | 0,04979 | NA |
| 3 | abundance | X02606 | 4-Amino-1 | Radius_length | 0,099891 | 0,067367 | NA |
| 3 | abundance | X02606 | 4-Amino-1 | Radius_SOS | 0,014285 | 0,049047 | NA |
| 3 | abundance | X02606 | 4-Amino-1 | Tibia_SOS | 0,062899 | 0,052433 | NA |
| 3 | abundance | X02606 | 4-Amino-1 | Handgrip | 0,075191 | 0,060286 | NA |
| 3 | abundance | X02615 | 6-APA | Tibia_length | -0,02579 | 0,053226 | NA |
| 3 | abundance | X02615 | 6-APA | Radius_length | -0,07598 | 0,07188 | NA |
| 3 | abundance | X02615 | 6-APA | Radius_SOS | -0,06532 | 0,050793 | NA |
| 3 | abundance | X02615 | 6-APA | Tibia_SOS | -0,07341 | 0,054143 | NA |
| 3 | abundance | X02615 | 6-APA | Handgrip | -0,01582 | 0,061352 | NA |
| 3 | abundance | X02640 | thyronine | Tibia_length | -0,07836 | 0,052424 | NA |
| 3 | abundance | X02640 | thyronine | Radius_length | -0,08933 | 0,07115 | NA |
| 3 | abundance | X02640 | thyronine | Radius_SOS | 0,014475 | 0,050375 | NA |
| 3 | abundance | X02640 | thyronine | Tibia_SOS | -0,08835 | 0,053127 | NA |
| 3 | abundance | X02640 | thyronine | Handgrip | -0,10632 | 0,060958 | NA |
| 3 | abundance | X02651 | L-gamma-( | Tibia_length | -0,07453 | 0,053203 | NA |
| 3 | abundance | X02651 | L-gamma-( | Radius_length | -0,19615 | 0,07156 | NA |
| 3 | abundance | X02651 | L-gamma-( | Radius_SOS | -0,06071 | 0,05097 | NA |
| 3 | abundance | X02651 | L-gamma-( | Tibia_SOS | -0,07139 | 0,055236 | NA |
| 3 | abundance | X02651 | L-gamma-( | Handgrip | -0,01735 | 0,062608 | NA |
| 3 | abundance | X02652 | Leucyltyros | Tibia_length | -0,12852 | 0,052753 | NA |
| 3 | abundance | X02652 | Leucyltyros | Radius_length | -0,07792 | 0,071638 | NA |
| 3 | abundance | X02652 | Leucyltyros | Radius_SOS | -0,00204 | 0,050604 | NA |
| 3 | abundance | X02652 | Leucyltyros | Tibia_SOS | -0,0874 | 0,053749 | NA |
| 3 | abundance | X02652 | Leucyltyros | Handgrip | -0,04446 | 0,06171 | NA |
| 3 | abundance | X02661 | 4-(3-Oxope | Tibia_length | -0,12534 | 0,052715 | NA |
| 3 | abundance | X02661 | 4-(3-Oxope | Radius_length | -0,12079 | 0,071786 | NA |
| 3 | abundance | X02661 | 4-(3-Oxope | Radius_SOS | -0,05813 | 0,051057 | NA |
| 3 | abundance | X02661 | 4-(3-Oxope | Tibia_SOS | 0,072024 | 0,053199 | NA |
| 3 | abundance | X02661 | 4-(3-Oxope | Handgrip | 0,043838 | 0,061101 | NA |
| 3 | abundance | X02676 | Val-Trp | Tibia_length | -0,14409 | 0,054296 | NA |
| 3 | abundance | X02676 | Val-Trp | Radius_length | -0,19325 | 0,07212 | NA |
| 3 | abundance | X02676 | Val-Trp | Radius_SOS | -0,00327 | 0,051706 | NA |
| 3 | abundance | X02676 | Val-Trp | Tibia_SOS | -0,11498 | 0,054863 | NA |
| 3 | abundance | X02676 | Val-Trp | Handgrip | -0,06815 | 0,061517 | NA |
| 3 | abundance | X02687 | S-Propylcy | Tibia_length | 0,13871 | 0,054154 | NA |
| 3 | abundance | X02687 | S-Propylcy | Radius_length | 0,126411 | 0,072937 | NA |
| 3 | abundance | X02687 | S-Propylcy | Radius_SOS | 0,211698 | 0,051401 | NA |
| 3 | abundance | X02687 | S-Propylcy | Tibia_SOS | 0,089681 | 0,05486 | NA |
| 3 | abundance | X02687 | S-Propylcy | Handgrip | 0,042178 | 0,062169 | NA |
| 3 | abundance | X02689 | piscidic ac | Tibia_length | -0,14607 | 0,051674 | NA |

| | | | | | | | |
|---|---|---|---|---|---|---|---|
| 3 | abundance | X02689 | piscidic ac | Radius_length | -0,10821 | 0,070474 | NA |
| 3 | abundance | X02689 | piscidic ac | Radius_SOS | -0,00761 | 0,050001 | NA |
| 3 | abundance | X02689 | piscidic ac | Tibia_SOS | -6,89E-04 | 0,053043 | NA |
| 3 | abundance | X02689 | piscidic ac | Handgrip | -0,11083 | 0,06015 | NA |
| 3 | abundance | X02702 | R-(+)-Etira | Tibia_length | 0,025673 | 0,054084 | NA |
| 3 | abundance | X02702 | R-(+)-Etira | Radius_length | -0,05132 | 0,072708 | NA |
| 3 | abundance | X02702 | R-(+)-Etira | Radius_SOS | 0,032996 | 0,051508 | NA |
| 3 | abundance | X02702 | R-(+)-Etira | Tibia_SOS | -0,01726 | 0,055275 | NA |
| 3 | abundance | X02702 | R-(+)-Etira | Handgrip | -0,17876 | 0,060399 | NA |
| 3 | abundance | X02720 | ala-ser | Tibia_length | 0,064492 | 0,051888 | NA |
| 3 | abundance | X02720 | ala-ser | Radius_length | 0,1313 | 0,07052 | NA |
| 3 | abundance | X02720 | ala-ser | Radius_SOS | 0,021854 | 0,049962 | NA |
| 3 | abundance | X02720 | ala-ser | Tibia_SOS | 0,064062 | 0,053306 | NA |
| 3 | abundance | X02720 | ala-ser | Handgrip | 0,068419 | 0,061189 | NA |
| 3 | abundance | X02730 | N-Phenyla | Tibia_length | -0,12157 | 0,05544 | NA |
| 3 | abundance | X02730 | N-Phenyla | Radius_length | -0,08704 | 0,073477 | NA |
| 3 | abundance | X02730 | N-Phenyla | Radius_SOS | 0,079871 | 0,052993 | NA |
| 3 | abundance | X02730 | N-Phenyla | Tibia_SOS | 0,001907 | 0,056349 | NA |
| 3 | abundance | X02730 | N-Phenyla | Handgrip | -0,01814 | 0,062023 | NA |
| 3 | abundance | X02733 | Calcitriol | Tibia_length | -0,01527 | 0,052862 | NA |
| 3 | abundance | X02733 | Calcitriol | Radius_length | -0,08921 | 0,072059 | NA |
| 3 | abundance | X02733 | Calcitriol | Radius_SOS | -0,01674 | 0,050588 | NA |
| 3 | abundance | X02733 | Calcitriol | Tibia_SOS | 0,14098 | 0,053041 | NA |
| 3 | abundance | X02733 | Calcitriol | Handgrip | -0,14746 | 0,061072 | NA |
| 3 | abundance | X02746 | N,N-Diethy | Tibia_length | -0,07832 | 0,054514 | NA |
| 3 | abundance | X02746 | N,N-Diethy | Radius_length | 0,007261 | 0,072869 | NA |
| 3 | abundance | X02746 | N,N-Diethy | Radius_SOS | 0,04419 | 0,051532 | NA |
| 3 | abundance | X02746 | N,N-Diethy | Tibia_SOS | 0,083217 | 0,054938 | NA |
| 3 | abundance | X02746 | N,N-Diethy | Handgrip | 0,078456 | 0,061717 | NA |
| 3 | abundance | X02760 | 6-hydroxyp | Tibia_length | 0,007868 | 0,055869 | NA |
| 3 | abundance | X02760 | 6-hydroxyp | Radius_length | 0,081 | 0,073924 | NA |
| 3 | abundance | X02760 | 6-hydroxyp | Radius_SOS | 0,055861 | 0,052904 | NA |
| 3 | abundance | X02760 | 6-hydroxyp | Tibia_SOS | 0,114694 | 0,058325 | NA |
| 3 | abundance | X02760 | 6-hydroxyp | Handgrip | 0,058821 | 0,062992 | NA |
| 3 | abundance | X02765 | L-Pyrrolysi | Tibia_length | 0,070793 | 0,053092 | NA |
| 3 | abundance | X02765 | L-Pyrrolysi | Radius_length | 0,034526 | 0,072001 | NA |
| 3 | abundance | X02765 | L-Pyrrolysi | Radius_SOS | -0,06402 | 0,050606 | NA |
| 3 | abundance | X02765 | L-Pyrrolysi | Tibia_SOS | 0,02575 | 0,054302 | NA |
| 3 | abundance | X02765 | L-Pyrrolysi | Handgrip | -0,09023 | 0,061278 | NA |
| 3 | abundance | X02767 | 6-APA | Tibia_length | -0,04431 | 0,058824 | NA |
| 3 | abundance | X02767 | 6-APA | Radius_length | -0,04373 | 0,075421 | NA |
| 3 | abundance | X02767 | 6-APA | Radius_SOS | -0,03568 | 0,056083 | NA |
| 3 | abundance | X02767 | 6-APA | Tibia_SOS | -0,09102 | 0,058507 | NA |
| 3 | abundance | X02767 | 6-APA | Handgrip | -0,03854 | 0,060148 | NA |
| 3 | abundance | X02769 | Tetraacety | Tibia_length | -0,01263 | 0,052604 | NA |
| 3 | abundance | X02769 | Tetraacety | Radius_length | -0,04534 | 0,071807 | NA |
| 3 | abundance | X02769 | Tetraacety | Radius_SOS | -0,05994 | 0,050331 | NA |
| 3 | abundance | X02769 | Tetraacety | Tibia_SOS | 0,086997 | 0,054033 | NA |

| | | | | | | | |
|---|---|---|---|---|---|---|---|
| 3 | abundance | X02769 | Tetraacety | Handgrip | -0,12967 | 0,061667 | NA |
| 3 | abundance | X02774 | 9-ribosylze | Tibia_length | -0,17938 | 0,055749 | NA |
| 3 | abundance | X02774 | 9-ribosylze | Radius_length | -0,18906 | 0,073092 | NA |
| 3 | abundance | X02774 | 9-ribosylze | Radius_SOS | -0,10651 | 0,052705 | NA |
| 3 | abundance | X02774 | 9-ribosylze | Tibia_SOS | -0,04848 | 0,05672 | NA |
| 3 | abundance | X02774 | 9-ribosylze | Handgrip | -0,01334 | 0,062321 | NA |
| 3 | abundance | X02793 | 3-Methoxy | Tibia_length | -0,14847 | 0,04996 | NA |
| 3 | abundance | X02793 | 3-Methoxy | Radius_length | -0,17911 | 0,067804 | NA |
| 3 | abundance | X02793 | 3-Methoxy | Radius_SOS | -0,01192 | 0,049151 | NA |
| 3 | abundance | X02793 | 3-Methoxy | Tibia_SOS | -0,10036 | 0,052427 | NA |
| 3 | abundance | X02793 | 3-Methoxy | Handgrip | -0,12884 | 0,059897 | NA |
| 3 | abundance | X02820 | FC250500 | Tibia_length | -0,13519 | 0,053718 | NA |
| 3 | abundance | X02820 | FC250500 | Radius_length | -0,1809 | 0,072009 | NA |
| 3 | abundance | X02820 | FC250500 | Radius_SOS | 0,037194 | 0,051585 | NA |
| 3 | abundance | X02820 | FC250500 | Tibia_SOS | -0,12126 | 0,055137 | NA |
| 3 | abundance | X02820 | FC250500 | Handgrip | -0,05901 | 0,062316 | NA |
| 3 | abundance | X02823 | 6,8-Dimetl | Tibia_length | -0,04267 | 0,053113 | NA |
| 3 | abundance | X02823 | 6,8-Dimetl | Radius_length | -0,11679 | 0,07176 | NA |
| 3 | abundance | X02823 | 6,8-Dimetl | Radius_SOS | -0,04745 | 0,051112 | NA |
| 3 | abundance | X02823 | 6,8-Dimetl | Tibia_SOS | -0,08383 | 0,053857 | NA |
| 3 | abundance | X02823 | 6,8-Dimetl | Handgrip | -0,01301 | 0,061554 | NA |
| 3 | abundance | X02824 | Eslicarbaz | Tibia_length | -0,06422 | 0,053719 | NA |
| 3 | abundance | X02824 | Eslicarbaz | Radius_length | -0,1019 | 0,0726 | NA |
| 3 | abundance | X02824 | Eslicarbaz | Radius_SOS | 0,057413 | 0,051068 | NA |
| 3 | abundance | X02824 | Eslicarbaz | Tibia_SOS | -0,11185 | 0,054044 | NA |
| 3 | abundance | X02824 | Eslicarbaz | Handgrip | -0,02609 | 0,06142 | NA |
| 3 | abundance | X02832 | (+/-)-2-Hyc | Tibia_length | -0,03649 | 0,055465 | NA |
| 3 | abundance | X02832 | (+/-)-2-Hyc | Radius_length | 0,068539 | 0,074634 | NA |
| 3 | abundance | X02832 | (+/-)-2-Hyc | Radius_SOS | -0,01641 | 0,052709 | NA |
| 3 | abundance | X02832 | (+/-)-2-Hyc | Tibia_SOS | 0,086285 | 0,055887 | NA |
| 3 | abundance | X02832 | (+/-)-2-Hyc | Handgrip | 0,093497 | 0,06161 | NA |
| 3 | abundance | X02841 | Kyotorphin | Tibia_length | -0,0656 | 0,053773 | NA |
| 3 | abundance | X02841 | Kyotorphin | Radius_length | 0,043161 | 0,073113 | NA |
| 3 | abundance | X02841 | Kyotorphin | Radius_SOS | 0,006374 | 0,051438 | NA |
| 3 | abundance | X02841 | Kyotorphin | Tibia_SOS | 0,014966 | 0,056188 | NA |
| 3 | abundance | X02841 | Kyotorphin | Handgrip | -0,11534 | 0,063012 | NA |
| 3 | abundance | X02843 | 4-(1-Hydro | Tibia_length | 0,060789 | 0,051837 | NA |
| 3 | abundance | X02843 | 4-(1-Hydro | Radius_length | 0,083055 | 0,070943 | NA |
| 3 | abundance | X02843 | 4-(1-Hydro | Radius_SOS | -0,01513 | 0,049802 | NA |
| 3 | abundance | X02843 | 4-(1-Hydro | Tibia_SOS | 0,139819 | 0,053978 | NA |
| 3 | abundance | X02843 | 4-(1-Hydro | Handgrip | 0,138506 | 0,06074 | NA |
| 3 | abundance | X02853 | 7alpha-Hy | Tibia_length | 0,071107 | 0,051711 | NA |
| 3 | abundance | X02853 | 7alpha-Hy | Radius_length | 0,112121 | 0,070791 | NA |
| 3 | abundance | X02853 | 7alpha-Hy | Radius_SOS | 0,021076 | 0,049798 | NA |
| 3 | abundance | X02853 | 7alpha-Hy | Tibia_SOS | 0,073659 | 0,053595 | NA |
| 3 | abundance | X02853 | 7alpha-Hy | Handgrip | 0,015418 | 0,061313 | NA |
| 3 | abundance | X02854 | Marimasta | Tibia_length | 0,082213 | 0,055374 | NA |
| 3 | abundance | X02854 | Marimasta | Radius_length | 0,129554 | 0,073222 | NA |

| | | | | | | | |
|---|---|---|---|---|---|---|---|
| 3 | abundance | X02854 | Marimasta | Radius_SOS | -0,05856 | 0,052665 | NA |
| 3 | abundance | X02854 | Marimasta | Tibia_SOS | 0,050345 | 0,055883 | NA |
| 3 | abundance | X02854 | Marimasta | Handgrip | -0,01228 | 0,062287 | NA |
| 3 | abundance | X02868 | 2-(3-Hydro | Tibia_length | -0,07597 | 0,056619 | NA |
| 3 | abundance | X02868 | 2-(3-Hydro | Radius_length | -0,25473 | 0,072137 | NA |
| 3 | abundance | X02868 | 2-(3-Hydro | Radius_SOS | -0,1041 | 0,053688 | NA |
| 3 | abundance | X02868 | 2-(3-Hydro | Tibia_SOS | -0,15704 | 0,05675 | NA |
| 3 | abundance | X02868 | 2-(3-Hydro | Handgrip | -0,14935 | 0,062151 | NA |
| 3 | abundance | X02872 | folinic acid | Tibia_length | -0,12082 | 0,052964 | NA |
| 3 | abundance | X02872 | folinic acid | Radius_length | -0,05053 | 0,072803 | NA |
| 3 | abundance | X02872 | folinic acid | Radius_SOS | -0,05349 | 0,050921 | NA |
| 3 | abundance | X02872 | folinic acid | Tibia_SOS | -0,09189 | 0,054496 | NA |
| 3 | abundance | X02872 | folinic acid | Handgrip | 0,006458 | 0,061541 | NA |
| 3 | abundance | X02878 | N~6~-[5-(1 | Tibia_length | -0,09259 | 0,050812 | NA |
| 3 | abundance | X02878 | N~6~-[5-(1 | Radius_length | -0,06516 | 0,06985 | NA |
| 3 | abundance | X02878 | N~6~-[5-(1 | Radius_SOS | -0,01211 | 0,049467 | NA |
| 3 | abundance | X02878 | N~6~-[5-(1 | Tibia_SOS | -0,04293 | 0,053849 | NA |
| 3 | abundance | X02878 | N~6~-[5-(1 | Handgrip | 0,012858 | 0,062343 | NA |
| 3 | abundance | X02881 | N-(2,3,4-Tr | Tibia_length | -0,05739 | 0,054177 | NA |
| 3 | abundance | X02881 | N-(2,3,4-Tr | Radius_length | -0,11344 | 0,072362 | NA |
| 3 | abundance | X02881 | N-(2,3,4-Tr | Radius_SOS | -0,10311 | 0,05151 | NA |
| 3 | abundance | X02881 | N-(2,3,4-Tr | Tibia_SOS | -0,08029 | 0,055448 | NA |
| 3 | abundance | X02881 | N-(2,3,4-Tr | Handgrip | -0,04048 | 0,061717 | NA |
| 3 | abundance | X02894 | Toluene | Tibia_length | -0,08387 | 0,048953 | NA |
| 3 | abundance | X02894 | Toluene | Radius_length | -0,11858 | 0,064616 | NA |
| 3 | abundance | X02894 | Toluene | Radius_SOS | -0,12544 | 0,048784 | NA |
| 3 | abundance | X02894 | Toluene | Tibia_SOS | -0,06744 | 0,052444 | NA |
| 3 | abundance | X02894 | Toluene | Handgrip | 0,034206 | 0,060322 | NA |
| 3 | abundance | X02905 | MFCD0005 | Tibia_length | -0,07477 | 0,057209 | NA |
| 3 | abundance | X02905 | MFCD0005 | Radius_length | -0,02891 | 0,075018 | NA |
| 3 | abundance | X02905 | MFCD0005 | Radius_SOS | 0,00732 | 0,054595 | NA |
| 3 | abundance | X02905 | MFCD0005 | Tibia_SOS | -0,02744 | 0,058478 | NA |
| 3 | abundance | X02905 | MFCD0005 | Handgrip | -0,01031 | 0,063122 | NA |
| 3 | abundance | X02920 | L-gamma-( | Tibia_length | 0,01488 | 0,051149 | NA |
| 3 | abundance | X02920 | L-gamma-( | Radius_length | 0,128038 | 0,069606 | NA |
| 3 | abundance | X02920 | L-gamma-( | Radius_SOS | -0,00939 | 0,049531 | NA |
| 3 | abundance | X02920 | L-gamma-( | Tibia_SOS | 0,118299 | 0,052934 | NA |
| 3 | abundance | X02920 | L-gamma-( | Handgrip | -0,0983 | 0,060933 | NA |
| 3 | abundance | X02922 | (2E)-3-Met | Tibia_length | -0,0448 | 0,057058 | NA |
| 3 | abundance | X02922 | (2E)-3-Met | Radius_length | -0,12579 | 0,074375 | NA |
| 3 | abundance | X02922 | (2E)-3-Met | Radius_SOS | -0,1752 | 0,053835 | NA |
| 3 | abundance | X02922 | (2E)-3-Met | Tibia_SOS | -0,118 | 0,058825 | NA |
| 3 | abundance | X02922 | (2E)-3-Met | Handgrip | -0,07642 | 0,062877 | NA |
| 3 | abundance | X02928 | N-Acetylpr | Tibia_length | 0,057297 | 0,055211 | NA |
| 3 | abundance | X02928 | N-Acetylpr | Radius_length | 0,162973 | 0,073184 | NA |
| 3 | abundance | X02928 | N-Acetylpr | Radius_SOS | 0,08621 | 0,052464 | NA |
| 3 | abundance | X02928 | N-Acetylpr | Tibia_SOS | 0,134813 | 0,055826 | NA |
| 3 | abundance | X02928 | N-Acetylpr | Handgrip | 0,100599 | 0,062456 | NA |

| | | | | | | | | |
|---|---|---|---|---|---|---|---|---|
| | 3 | abundance | X02943 | 3-Methoxy· | Tibia_length | -0,04187 | 0,050973 | NA |
| | 3 | abundance | X02943 | 3-Methoxy· | Radius_length | -0,07183 | 0,069545 | NA |
| | 3 | abundance | X02943 | 3-Methoxy· | Radius_SOS | -2,96E-04 | 0,049437 | NA |
| | 3 | abundance | X02943 | 3-Methoxy· | Tibia_SOS | -0,08254 | 0,052659 | NA |
| | 3 | abundance | X02943 | 3-Methoxy· | Handgrip | -0,066 | 0,060594 | NA |
| | 3 | abundance | X02944 | Valylvaline | Tibia_length | -0,04001 | 0,049797 | NA |
| | 3 | abundance | X02944 | Valylvaline | Radius_length | 0,057807 | 0,067994 | NA |
| | 3 | abundance | X02944 | Valylvaline | Radius_SOS | -0,05188 | 0,048972 | NA |
| | 3 | abundance | X02944 | Valylvaline | Tibia_SOS | 0,062885 | 0,05248 | NA |
| | 3 | abundance | X02944 | Valylvaline | Handgrip | -0,05783 | 0,060241 | NA |
| | 3 | abundance | X02948 | N-(2-Cyan | Tibia_length | -0,1379 | 0,0565 | NA |
| | 3 | abundance | X02948 | N-(2-Cyan | Radius_length | -0,11796 | 0,074583 | NA |
| | 3 | abundance | X02948 | N-(2-Cyan | Radius_SOS | -0,03001 | 0,054136 | NA |
| | 3 | abundance | X02948 | N-(2-Cyan | Tibia_SOS | -0,14302 | 0,056802 | NA |
| | 3 | abundance | X02948 | N-(2-Cyan | Handgrip | -0,13929 | 0,060925 | NA |
| | 3 | abundance | X02952 | Vorinostat | Tibia_length | -0,06217 | 0,053929 | NA |
| | 3 | abundance | X02952 | Vorinostat | Radius_length | -0,1149 | 0,072328 | NA |
| | 3 | abundance | X02952 | Vorinostat | Radius_SOS | -0,08611 | 0,051432 | NA |
| | 3 | abundance | X02952 | Vorinostat | Tibia_SOS | 0,032735 | 0,054008 | NA |
| | 3 | abundance | X02952 | Vorinostat | Handgrip | 0,01486 | 0,061064 | NA |
| | 3 | abundance | X02968 | Ro 20-172 | Tibia_length | -0,05416 | 0,052502 | NA |
| | 3 | abundance | X02968 | Ro 20-172 | Radius_length | -0,13037 | 0,07127 | NA |
| | 3 | abundance | X02968 | Ro 20-172 | Radius_SOS | -0,02495 | 0,050361 | NA |
| | 3 | abundance | X02968 | Ro 20-172 | Tibia_SOS | -0,0655 | 0,053018 | NA |
| | 3 | abundance | X02968 | Ro 20-172 | Handgrip | -0,05611 | 0,061603 | NA |
| | 3 | abundance | X02972 | 3-(Sulfoox | Tibia_length | -0,01518 | 0,051381 | NA |
| | 3 | abundance | X02972 | 3-(Sulfoox | Radius_length | -0,07552 | 0,070386 | NA |
| | 3 | abundance | X02972 | 3-(Sulfoox | Radius_SOS | 0,02812 | 0,049636 | NA |
| | 3 | abundance | X02972 | 3-(Sulfoox | Tibia_SOS | -0,01154 | 0,053128 | NA |
| | 3 | abundance | X02972 | 3-(Sulfoox | Handgrip | -0,0368 | 0,061363 | NA |
| | 3 | abundance | X02986 | Diacetin | Tibia_length | -0,01675 | 0,049694 | NA |
| | 3 | abundance | X02986 | Diacetin | Radius_length | -0,04094 | 0,067352 | NA |
| | 3 | abundance | X02986 | Diacetin | Radius_SOS | -0,10748 | 0,048749 | NA |
| | 3 | abundance | X02986 | Diacetin | Tibia_SOS | -0,10738 | 0,052184 | NA |
| | 3 | abundance | X02986 | Diacetin | Handgrip | -0,06084 | 0,060281 | NA |
| | 3 | abundance | X02990 | Pentoxifylli | Tibia_length | 0,044516 | 0,053854 | NA |
| | 3 | abundance | X02990 | Pentoxifylli | Radius_length | 0,058584 | 0,072853 | NA |
| | 3 | abundance | X02990 | Pentoxifylli | Radius_SOS | -0,01522 | 0,051476 | NA |
| | 3 | abundance | X02990 | Pentoxifylli | Tibia_SOS | 0,022525 | 0,055061 | NA |
| | 3 | abundance | X02990 | Pentoxifylli | Handgrip | 0,046716 | 0,06256 | NA |
| 2b | | abundance | X03001 | Pseudouri | Tibia_length | 0,039263 | 0,051287 | NA |
| 2b | | abundance | X03001 | Pseudouri | Radius_length | 0,021576 | 0,071073 | NA |
| 2b | | abundance | X03001 | Pseudouri | Radius_SOS | 4,49E-04 | 0,049684 | NA |
| 2b | | abundance | X03001 | Pseudouri | Tibia_SOS | 0,122125 | 0,052666 | NA |
| 2b | | abundance | X03001 | Pseudouri | Handgrip | 0,132103 | 0,060636 | NA |
| | 3 | abundance | X03004 | Bicine | Tibia_length | 0,039915 | 0,054713 | NA |
| | 3 | abundance | X03004 | Bicine | Radius_length | 0,153733 | 0,072481 | NA |
| | 3 | abundance | X03004 | Bicine | Radius_SOS | 0,08817 | 0,051942 | NA |

| | | | | | | | |
|---|---|---|---|---|---|---|---|
| 3 | abundance | X03004 | Bicine | Tibia_SOS | 0,07295 | 0,056385 | NA |
| 3 | abundance | X03004 | Bicine | Handgrip | -0,00636 | 0,06166 | NA |
| 3 | abundance | X03017 | Pro-tyr | Tibia_length | -0,0506 | 0,058091 | NA |
| 3 | abundance | X03017 | Pro-tyr | Radius_length | -0,07245 | 0,075231 | NA |
| 3 | abundance | X03017 | Pro-tyr | Radius_SOS | -0,00826 | 0,055455 | NA |
| 3 | abundance | X03017 | Pro-tyr | Tibia_SOS | -0,04476 | 0,057663 | NA |
| 3 | abundance | X03017 | Pro-tyr | Handgrip | -0,08283 | 0,063509 | NA |
| 2b | abundance | X03023 | NPK (Pepti | Tibia_length | 0,043231 | 0,053541 | NA |
| 2b | abundance | X03023 | NPK (Pepti | Radius_length | 0,059308 | 0,072177 | NA |
| 2b | abundance | X03023 | NPK (Pepti | Radius_SOS | 0,031447 | 0,051048 | NA |
| 2b | abundance | X03023 | NPK (Pepti | Tibia_SOS | 0,037228 | 0,054002 | NA |
| 2b | abundance | X03023 | NPK (Pepti | Handgrip | -0,00513 | 0,062457 | NA |
| 3 | abundance | X03064 | Formimino | Tibia_length | 0,116355 | 0,052808 | NA |
| 3 | abundance | X03064 | Formimino | Radius_length | -0,11559 | 0,071726 | NA |
| 3 | abundance | X03064 | Formimino | Radius_SOS | -0,03444 | 0,050742 | NA |
| 3 | abundance | X03064 | Formimino | Tibia_SOS | 0,017874 | 0,054048 | NA |
| 3 | abundance | X03064 | Formimino | Handgrip | -0,15815 | 0,060522 | NA |
| 3 | abundance | X03069 | N-COUMAI | Tibia_length | -0,02129 | 0,052686 | NA |
| 3 | abundance | X03069 | N-COUMAI | Radius_length | -0,05528 | 0,071695 | NA |
| 3 | abundance | X03069 | N-COUMAI | Radius_SOS | 0,011591 | 0,050574 | NA |
| 3 | abundance | X03069 | N-COUMAI | Tibia_SOS | 0,015624 | 0,054354 | NA |
| 3 | abundance | X03069 | N-COUMAI | Handgrip | -0,05037 | 0,061392 | NA |
| 3 | abundance | X03070 | 2-Methoxy- | Tibia_length | 0,007125 | 0,053238 | NA |
| 3 | abundance | X03070 | 2-Methoxy- | Radius_length | 0,039055 | 0,072036 | NA |
| 3 | abundance | X03070 | 2-Methoxy- | Radius_SOS | -0,00981 | 0,050902 | NA |
| 3 | abundance | X03070 | 2-Methoxy- | Tibia_SOS | 0,0039 | 0,053943 | NA |
| 3 | abundance | X03070 | 2-Methoxy- | Handgrip | -0,08896 | 0,062068 | NA |
| 3 | abundance | X03077 | nicotianan | Tibia_length | -0,01713 | 0,050768 | NA |
| 3 | abundance | X03077 | nicotianan | Radius_length | 0,037802 | 0,069477 | NA |
| 3 | abundance | X03077 | nicotianan | Radius_SOS | 0,054333 | 0,049259 | NA |
| 3 | abundance | X03077 | nicotianan | Tibia_SOS | 0,00484 | 0,052784 | NA |
| 3 | abundance | X03077 | nicotianan | Handgrip | 0,022965 | 0,06036 | NA |
| 3 | abundance | X03097 | 6-hydroxyn | Tibia_length | -0,14302 | 0,051777 | NA |
| 3 | abundance | X03097 | 6-hydroxyn | Radius_length | 0,006428 | 0,071257 | NA |
| 3 | abundance | X03097 | 6-hydroxyn | Radius_SOS | -0,00925 | 0,050331 | NA |
| 3 | abundance | X03097 | 6-hydroxyn | Tibia_SOS | 0,044399 | 0,054206 | NA |
| 3 | abundance | X03097 | 6-hydroxyn | Handgrip | 0,031868 | 0,062218 | NA |
| 3 | abundance | X03109 | Guanidino | Tibia_length | -0,08292 | 0,051227 | NA |
| 3 | abundance | X03109 | Guanidino | Radius_length | -0,11544 | 0,069917 | NA |
| 3 | abundance | X03109 | Guanidino | Radius_SOS | 0,013297 | 0,04968 | NA |
| 3 | abundance | X03109 | Guanidino | Tibia_SOS | -0,05579 | 0,052834 | NA |
| 3 | abundance | X03109 | Guanidino | Handgrip | -0,01659 | 0,06111 | NA |
| 3 | abundance | X03132 | (3S,4S)-7,: | Tibia_length | 0,017459 | 0,052642 | NA |
| 3 | abundance | X03132 | (3S,4S)-7,: | Radius_length | 0,112748 | 0,071249 | NA |
| 3 | abundance | X03132 | (3S,4S)-7,: | Radius_SOS | 0,023906 | 0,050425 | NA |
| 3 | abundance | X03132 | (3S,4S)-7,: | Tibia_SOS | 0,165976 | 0,05256 | NA |
| 3 | abundance | X03132 | (3S,4S)-7,: | Handgrip | 0,133811 | 0,061002 | NA |
| 3 | abundance | X03134 | Tetrahydro | Tibia_length | -0,0644 | 0,055028 | NA |

| | | | | | | | |
|---|---|---|---|---|---|---|---|
| 3 | abundance | X03134 | Tetrahydro | Radius_length | -0,11173 | 0,073201 | NA |
| 3 | abundance | X03134 | Tetrahydro | Radius_SOS | -0,1466 | 0,051792 | NA |
| 3 | abundance | X03134 | Tetrahydro | Tibia_SOS | -0,06177 | 0,055705 | NA |
| 3 | abundance | X03134 | Tetrahydro | Handgrip | -0,1775 | 0,060799 | NA |
| 3 | abundance | X03158 | 5-Hydroxy- | Tibia_length | -0,04274 | 0,052786 | NA |
| 3 | abundance | X03158 | 5-Hydroxy- | Radius_length | -0,03472 | 0,071561 | NA |
| 3 | abundance | X03158 | 5-Hydroxy- | Radius_SOS | 0,033876 | 0,050479 | NA |
| 3 | abundance | X03158 | 5-Hydroxy- | Tibia_SOS | 0,010522 | 0,055302 | NA |
| 3 | abundance | X03158 | 5-Hydroxy- | Handgrip | 0,026982 | 0,061648 | NA |
| 3 | abundance | X03223 | 1-(2-Carbo | Tibia_length | -0,11138 | 0,052656 | NA |
| 3 | abundance | X03223 | 1-(2-Carbo | Radius_length | -0,14198 | 0,071315 | NA |
| 3 | abundance | X03223 | 1-(2-Carbo | Radius_SOS | 0,048738 | 0,050428 | NA |
| 3 | abundance | X03223 | 1-(2-Carbo | Tibia_SOS | -0,10595 | 0,053299 | NA |
| 3 | abundance | X03223 | 1-(2-Carbo | Handgrip | -0,20011 | 0,060375 | NA |
| 3 | abundance | X03254 | Gly-Trp | Tibia_length | -0,08887 | 0,054673 | NA |
| 3 | abundance | X03254 | Gly-Trp | Radius_length | -0,04694 | 0,073683 | NA |
| 3 | abundance | X03254 | Gly-Trp | Radius_SOS | 0,044984 | 0,051899 | NA |
| 3 | abundance | X03254 | Gly-Trp | Tibia_SOS | 0,001037 | 0,055087 | NA |
| 3 | abundance | X03254 | Gly-Trp | Handgrip | 0,02866 | 0,061457 | NA |
| 3 | abundance | X03268 | Dihydrouri | Tibia_length | 0,007021 | 0,053203 | NA |
| 3 | abundance | X03268 | Dihydrouri | Radius_length | -0,04496 | 0,072328 | NA |
| 3 | abundance | X03268 | Dihydrouri | Radius_SOS | -0,04825 | 0,050817 | NA |
| 3 | abundance | X03268 | Dihydrouri | Tibia_SOS | 0,129193 | 0,053103 | NA |
| 3 | abundance | X03268 | Dihydrouri | Handgrip | 0,004615 | 0,062327 | NA |
| 3 | abundance | X03271 | 1ÃŽÂ²-Hyd | Tibia_length | 0,0933 | 0,050218 | NA |
| 3 | abundance | X03271 | 1ÃŽÂ²-Hyd | Radius_length | 0,142952 | 0,068266 | NA |
| 3 | abundance | X03271 | 1ÃŽÂ²-Hyd | Radius_SOS | 0,038581 | 0,049138 | NA |
| 3 | abundance | X03271 | 1ÃŽÂ²-Hyd | Tibia_SOS | -0,01473 | 0,052752 | NA |
| 3 | abundance | X03271 | 1ÃŽÂ²-Hyd | Handgrip | 0,031775 | 0,060434 | NA |
| 3 | abundance | X03275 | Methdilazi | Tibia_length | -0,11661 | 0,051746 | NA |
| 3 | abundance | X03275 | Methdilazi | Radius_length | -0,05372 | 0,071304 | NA |
| 3 | abundance | X03275 | Methdilazi | Radius_SOS | 0,017544 | 0,050102 | NA |
| 3 | abundance | X03275 | Methdilazi | Tibia_SOS | -0,01067 | 0,053222 | NA |
| 3 | abundance | X03275 | Methdilazi | Handgrip | -0,06393 | 0,06071 | NA |
| 3 | abundance | X03276 | 2'-Deoxyac | Tibia_length | -0,05226 | 0,058934 | NA |
| 3 | abundance | X03276 | 2'-Deoxyac | Radius_length | 0,012493 | 0,075316 | NA |
| 3 | abundance | X03276 | 2'-Deoxyac | Radius_SOS | -0,00807 | 0,055904 | NA |
| 3 | abundance | X03276 | 2'-Deoxyac | Tibia_SOS | -0,00922 | 0,058841 | NA |
| 3 | abundance | X03276 | 2'-Deoxyac | Handgrip | 0,170344 | 0,062638 | NA |
| 3 | abundance | X03278 | Hydroxyph | Tibia_length | -0,05165 | 0,055355 | NA |
| 3 | abundance | X03278 | Hydroxyph | Radius_length | -0,09177 | 0,073386 | NA |
| 3 | abundance | X03278 | Hydroxyph | Radius_SOS | -0,1247 | 0,052801 | NA |
| 3 | abundance | X03278 | Hydroxyph | Tibia_SOS | -0,01552 | 0,056977 | NA |
| 3 | abundance | X03278 | Hydroxyph | Handgrip | -0,04873 | 0,062667 | NA |
| 3 | abundance | X03294 | 4,4'-Thiobi | Tibia_length | -0,01852 | 0,053217 | NA |
| 3 | abundance | X03294 | 4,4'-Thiobi | Radius_length | -0,09553 | 0,071858 | NA |
| 3 | abundance | X03294 | 4,4'-Thiobi | Radius_SOS | -0,05383 | 0,050919 | NA |
| 3 | abundance | X03294 | 4,4'-Thiobi | Tibia_SOS | 0,023104 | 0,054004 | NA |

| | | | | | | | |
|---|---|---|---|---|---|---|---|
| 3 | abundance | X03294 | 4,4'-Thiobi | Handgrip | 0,111382 | 0,061312 | NA |
| 3 | abundance | X03340 | His-pro | Tibia_length | 0,007433 | 0,052603 | NA |
| 3 | abundance | X03340 | His-pro | Radius_length | 0,042916 | 0,071844 | NA |
| 3 | abundance | X03340 | His-pro | Radius_SOS | -0,0051 | 0,050416 | NA |
| 3 | abundance | X03340 | His-pro | Tibia_SOS | 0,020505 | 0,054633 | NA |
| 3 | abundance | X03340 | His-pro | Handgrip | 0,04857 | 0,061184 | NA |
| 3 | abundance | X03343 | L-alpha-As | Tibia_length | -0,03719 | 0,051565 | NA |
| 3 | abundance | X03343 | L-alpha-As | Radius_length | -0,00794 | 0,070805 | NA |
| 3 | abundance | X03343 | L-alpha-As | Radius_SOS | -0,02192 | 0,049776 | NA |
| 3 | abundance | X03343 | L-alpha-As | Tibia_SOS | -0,0217 | 0,05398 | NA |
| 3 | abundance | X03343 | L-alpha-As | Handgrip | -0,11423 | 0,061007 | NA |
| 3 | abundance | X03357 | 2-Hydroxyl | Tibia_length | -0,05717 | 0,050501 | NA |
| 3 | abundance | X03357 | 2-Hydroxyl | Radius_length | -0,13059 | 0,068447 | NA |
| 3 | abundance | X03357 | 2-Hydroxyl | Radius_SOS | -0,03753 | 0,049212 | NA |
| 3 | abundance | X03357 | 2-Hydroxyl | Tibia_SOS | -0,13757 | 0,052228 | NA |
| 3 | abundance | X03357 | 2-Hydroxyl | Handgrip | -0,13312 | 0,059918 | NA |
| 3 | abundance | X03363 | Choline Alf | Tibia_length | -0,01092 | 0,054602 | NA |
| 3 | abundance | X03363 | Choline Alf | Radius_length | 0,133123 | 0,072507 | NA |
| 3 | abundance | X03363 | Choline Alf | Radius_SOS | 0,016234 | 0,051938 | NA |
| 3 | abundance | X03363 | Choline Alf | Tibia_SOS | 0,128949 | 0,055074 | NA |
| 3 | abundance | X03363 | Choline Alf | Handgrip | 0,144786 | 0,061902 | NA |
| 3 | abundance | X03376 | 4-(9H-beta | Tibia_length | -0,14392 | 0,05332 | NA |
| 3 | abundance | X03376 | 4-(9H-beta | Radius_length | -0,13386 | 0,071772 | NA |
| 3 | abundance | X03376 | 4-(9H-beta | Radius_SOS | 0,039687 | 0,051054 | NA |
| 3 | abundance | X03376 | 4-(9H-beta | Tibia_SOS | -0,12605 | 0,053237 | NA |
| 3 | abundance | X03376 | 4-(9H-beta | Handgrip | -0,12313 | 0,06098 | NA |
| 3 | abundance | X03413 | Val-Trp | Tibia_length | 0,043372 | 0,059298 | NA |
| 3 | abundance | X03413 | Val-Trp | Radius_length | 0,086057 | 0,075806 | NA |
| 3 | abundance | X03413 | Val-Trp | Radius_SOS | 0,051426 | 0,056871 | NA |
| 3 | abundance | X03413 | Val-Trp | Tibia_SOS | 0,04018 | 0,059818 | NA |
| 3 | abundance | X03413 | Val-Trp | Handgrip | 0,078132 | 0,060009 | NA |
| 3 | abundance | X03416 | 9-(alpha-D | Tibia_length | -0,02682 | 0,053368 | NA |
| 3 | abundance | X03416 | 9-(alpha-D | Radius_length | 0,120262 | 0,072093 | NA |
| 3 | abundance | X03416 | 9-(alpha-D | Radius_SOS | 0,055874 | 0,050849 | NA |
| 3 | abundance | X03416 | 9-(alpha-D | Tibia_SOS | 0,041845 | 0,054664 | NA |
| 3 | abundance | X03416 | 9-(alpha-D | Handgrip | 0,076136 | 0,0611 | NA |
| 3 | abundance | X03434 | SECONAL | Tibia_length | -0,04593 | 0,056111 | NA |
| 3 | abundance | X03434 | SECONAL | Radius_length | 0,076681 | 0,074052 | NA |
| 3 | abundance | X03434 | SECONAL | Radius_SOS | 0,106691 | 0,053264 | NA |
| 3 | abundance | X03434 | SECONAL | Tibia_SOS | -0,011 | 0,057749 | NA |
| 3 | abundance | X03434 | SECONAL | Handgrip | 0,003869 | 0,063046 | NA |
| 3 | abundance | X03480 | epsilon-(ga | Tibia_length | -0,00222 | 0,051878 | NA |
| 3 | abundance | X03480 | epsilon-(ga | Radius_length | 0,029004 | 0,071015 | NA |
| 3 | abundance | X03480 | epsilon-(ga | Radius_SOS | 0,015726 | 0,049972 | NA |
| 3 | abundance | X03480 | epsilon-(ga | Tibia_SOS | -0,02096 | 0,053385 | NA |
| 3 | abundance | X03480 | epsilon-(ga | Handgrip | 0,039985 | 0,060663 | NA |
| 3 | abundance | X03488 | 3,4-Dihydr | Tibia_length | -0,12168 | 0,054 | NA |
| 3 | abundance | X03488 | 3,4-Dihydr | Radius_length | -0,23132 | 0,070827 | NA |

| | | | | | | | |
|---|---|---|---|---|---|---|---|
| 3 | abundance | X03488 | 3,4-Dihydr | Radius_SOS | -0,02092 | 0,051411 | NA |
| 3 | abundance | X03488 | 3,4-Dihydr | Tibia_SOS | -0,1845 | 0,054578 | NA |
| 3 | abundance | X03488 | 3,4-Dihydr | Handgrip | -0,13385 | 0,060651 | NA |
| 3 | abundance | X03535 | (1S,3R,4s) | Tibia_length | -0,0253 | 0,05371 | NA |
| 3 | abundance | X03535 | (1S,3R,4s) | Radius_length | -0,12363 | 0,072433 | NA |
| 3 | abundance | X03535 | (1S,3R,4s) | Radius_SOS | -0,03584 | 0,051206 | NA |
| 3 | abundance | X03535 | (1S,3R,4s) | Tibia_SOS | -0,07014 | 0,055322 | NA |
| 3 | abundance | X03535 | (1S,3R,4s) | Handgrip | -0,15102 | 0,061801 | NA |
| 3 | abundance | X03552 | 4-Hydroxya | Tibia_length | -0,07486 | 0,052728 | NA |
| 3 | abundance | X03552 | 4-Hydroxya | Radius_length | -0,07471 | 0,071792 | NA |
| 3 | abundance | X03552 | 4-Hydroxya | Radius_SOS | -0,00176 | 0,050521 | NA |
| 3 | abundance | X03552 | 4-Hydroxya | Tibia_SOS | -0,0046 | 0,054572 | NA |
| 3 | abundance | X03552 | 4-Hydroxya | Handgrip | -0,10986 | 0,062034 | NA |
| 3 | abundance | X03563 | 6-imino-5- | Tibia_length | -0,04483 | 0,053051 | NA |
| 3 | abundance | X03563 | 6-imino-5- | Radius_length | -0,02158 | 0,072222 | NA |
| 3 | abundance | X03563 | 6-imino-5- | Radius_SOS | -0,00796 | 0,050822 | NA |
| 3 | abundance | X03563 | 6-imino-5- | Tibia_SOS | -0,02465 | 0,054769 | NA |
| 3 | abundance | X03563 | 6-imino-5- | Handgrip | -0,10538 | 0,061462 | NA |
| 3 | abundance | X03568 | 3-[(2Z)-1-C | Tibia_length | -0,07234 | 0,052062 | NA |
| 3 | abundance | X03568 | 3-[(2Z)-1-C | Radius_length | -0,1335 | 0,070662 | NA |
| 3 | abundance | X03568 | 3-[(2Z)-1-C | Radius_SOS | -0,00225 | 0,050081 | NA |
| 3 | abundance | X03568 | 3-[(2Z)-1-C | Tibia_SOS | -0,10102 | 0,053548 | NA |
| 3 | abundance | X03568 | 3-[(2Z)-1-C | Handgrip | -0,08211 | 0,061225 | NA |
| 3 | abundance | X03592 | 2-glyceryl | Tibia_length | 0,013164 | 0,05026 | NA |
| 3 | abundance | X03592 | 2-glyceryl | Radius_length | 0,052204 | 0,068405 | NA |
| 3 | abundance | X03592 | 2-glyceryl | Radius_SOS | 0,073306 | 0,049014 | NA |
| 3 | abundance | X03592 | 2-glyceryl | Tibia_SOS | -0,02034 | 0,05274 | NA |
| 3 | abundance | X03592 | 2-glyceryl | Handgrip | -0,0568 | 0,060504 | NA |
| 3 | abundance | X03595 | Methylol D | Tibia_length | -0,02016 | 0,056366 | NA |
| 3 | abundance | X03595 | Methylol D | Radius_length | -0,03923 | 0,073993 | NA |
| 3 | abundance | X03595 | Methylol D | Radius_SOS | -0,02826 | 0,053502 | NA |
| 3 | abundance | X03595 | Methylol D | Tibia_SOS | 0,048507 | 0,056581 | NA |
| 3 | abundance | X03595 | Methylol D | Handgrip | 0,060123 | 0,060084 | NA |
| 3 | abundance | X03602 | o-Succinyl | Tibia_length | -0,04779 | 0,052008 | NA |
| 3 | abundance | X03602 | o-Succinyl | Radius_length | -0,01044 | 0,071064 | NA |
| 3 | abundance | X03602 | o-Succinyl | Radius_SOS | 0,04711 | 0,050031 | NA |
| 3 | abundance | X03602 | o-Succinyl | Tibia_SOS | 0,049058 | 0,053924 | NA |
| 3 | abundance | X03602 | o-Succinyl | Handgrip | -0,01564 | 0,061399 | NA |
| 3 | abundance | X03604 | Gly-Trp | Tibia_length | 0,022743 | 0,055543 | NA |
| 3 | abundance | X03604 | Gly-Trp | Radius_length | 0,005689 | 0,074138 | NA |
| 3 | abundance | X03604 | Gly-Trp | Radius_SOS | 0,092591 | 0,052536 | NA |
| 3 | abundance | X03604 | Gly-Trp | Tibia_SOS | 0,02654 | 0,05743 | NA |
| 3 | abundance | X03604 | Gly-Trp | Handgrip | 0,015348 | 0,063034 | NA |
| 3 | abundance | X03629 | Glycylglycy | Tibia_length | 0,06162 | 0,054525 | NA |
| 3 | abundance | X03629 | Glycylglycy | Radius_length | 0,114224 | 0,072635 | NA |
| 3 | abundance | X03629 | Glycylglycy | Radius_SOS | 0,009879 | 0,051958 | NA |
| 3 | abundance | X03629 | Glycylglycy | Tibia_SOS | 0,052256 | 0,055867 | NA |
| 3 | abundance | X03629 | Glycylglycy | Handgrip | 0,064944 | 0,062434 | NA |

| | | | | | | | |
|---|---|---|---|---|---|---|---|
| 3 | abundance | X03660 | trp-ser | Tibia_length | 0,029356 | 0,052204 | NA |
| 3 | abundance | X03660 | trp-ser | Radius_length | 0,010238 | 0,071122 | NA |
| 3 | abundance | X03660 | trp-ser | Radius_SOS | 0,038461 | 0,050147 | NA |
| 3 | abundance | X03660 | trp-ser | Tibia_SOS | -0,10515 | 0,053387 | NA |
| 3 | abundance | X03660 | trp-ser | Handgrip | -0,03378 | 0,062113 | NA |
| 3 | abundance | X03675 | asp-gln | Tibia_length | -0,05648 | 0,054118 | NA |
| 3 | abundance | X03675 | asp-gln | Radius_length | -0,00845 | 0,073224 | NA |
| 3 | abundance | X03675 | asp-gln | Radius_SOS | 0,051211 | 0,05172 | NA |
| 3 | abundance | X03675 | asp-gln | Tibia_SOS | 0,06026 | 0,055358 | NA |
| 3 | abundance | X03675 | asp-gln | Handgrip | -0,06042 | 0,061866 | NA |
| 3 | abundance | X03704 | N,N-Dimet | Tibia_length | -0,21773 | 0,053811 | NA |
| 3 | abundance | X03704 | N,N-Dimet | Radius_length | -0,18214 | 0,072059 | NA |
| 3 | abundance | X03704 | N,N-Dimet | Radius_SOS | -0,08814 | 0,051428 | NA |
| 3 | abundance | X03704 | N,N-Dimet | Tibia_SOS | -0,15215 | 0,053804 | NA |
| 3 | abundance | X03704 | N,N-Dimet | Handgrip | -0,15587 | 0,060367 | NA |
| 3 | abundance | X03707 | 9-[(5R)-5-E | Tibia_length | -0,03678 | 0,054554 | NA |
| 3 | abundance | X03707 | 9-[(5R)-5-E | Radius_length | -0,1303 | 0,072797 | NA |
| 3 | abundance | X03707 | 9-[(5R)-5-E | Radius_SOS | -0,03587 | 0,05199 | NA |
| 3 | abundance | X03707 | 9-[(5R)-5-E | Tibia_SOS | -0,12124 | 0,055396 | NA |
| 3 | abundance | X03707 | 9-[(5R)-5-E | Handgrip | -0,13976 | 0,062401 | NA |
| 3 | abundance | X03709 | Guanfacin | Tibia_length | 0,022809 | 0,055033 | NA |
| 3 | abundance | X03709 | Guanfacin | Radius_length | 0,01744 | 0,073718 | NA |
| 3 | abundance | X03709 | Guanfacin | Radius_SOS | -0,00531 | 0,052346 | NA |
| 3 | abundance | X03709 | Guanfacin | Tibia_SOS | 0,031365 | 0,056163 | NA |
| 3 | abundance | X03709 | Guanfacin | Handgrip | -0,07705 | 0,06111 | NA |
| 3 | abundance | X03714 | Sinapinic a | Tibia_length | -0,10491 | 0,053119 | NA |
| 3 | abundance | X03714 | Sinapinic a | Radius_length | -0,17783 | 0,071093 | NA |
| 3 | abundance | X03714 | Sinapinic a | Radius_SOS | -0,06201 | 0,050698 | NA |
| 3 | abundance | X03714 | Sinapinic a | Tibia_SOS | -0,09291 | 0,053761 | NA |
| 3 | abundance | X03714 | Sinapinic a | Handgrip | -0,09987 | 0,060561 | NA |
| 3 | abundance | X03718 | Propanthel | Tibia_length | -0,0386 | 0,056806 | NA |
| 3 | abundance | X03718 | Propanthel | Radius_length | -0,00999 | 0,074743 | NA |
| 3 | abundance | X03718 | Propanthel | Radius_SOS | -0,01982 | 0,054129 | NA |
| 3 | abundance | X03718 | Propanthel | Tibia_SOS | -0,04473 | 0,055564 | NA |
| 3 | abundance | X03718 | Propanthel | Handgrip | -0,02374 | 0,062447 | NA |
| 3 | abundance | X03719 | Losalen | Tibia_length | -0,09263 | 0,056318 | NA |
| 3 | abundance | X03719 | Losalen | Radius_length | -0,13902 | 0,073556 | NA |
| 3 | abundance | X03719 | Losalen | Radius_SOS | -0,03325 | 0,053621 | NA |
| 3 | abundance | X03719 | Losalen | Tibia_SOS | -0,11629 | 0,057717 | NA |
| 3 | abundance | X03719 | Losalen | Handgrip | -0,15614 | 0,062065 | NA |
| 3 | abundance | X03760 | 2-Methoxy | Tibia_length | 0,031844 | 0,052862 | NA |
| 3 | abundance | X03760 | 2-Methoxy | Radius_length | -0,09028 | 0,071908 | NA |
| 3 | abundance | X03760 | 2-Methoxy | Radius_SOS | 0,010177 | 0,050638 | NA |
| 3 | abundance | X03760 | 2-Methoxy | Tibia_SOS | 0,050687 | 0,054047 | NA |
| 3 | abundance | X03760 | 2-Methoxy | Handgrip | 0,0866 | 0,061171 | NA |
| 3 | abundance | X03793 | Desonide | Tibia_length | -0,02743 | 0,05337 | NA |
| 3 | abundance | X03793 | Desonide | Radius_length | -0,03632 | 0,072349 | NA |
| 3 | abundance | X03793 | Desonide | Radius_SOS | -0,04135 | 0,050998 | NA |

| | | | | | | | |
|---|---|---|---|---|---|---|---|
| 3 | abundance | X03793 | Desonide | Tibia_SOS | -0,00675 | 0,054334 | NA |
| 3 | abundance | X03793 | Desonide | Handgrip | -0,01389 | 0,061489 | NA |
| 3 | abundance | X03837 | 2-Methoxy· | Tibia_length | 0,098608 | 0,055569 | NA |
| 3 | abundance | X03837 | 2-Methoxy· | Radius_length | 0,024555 | 0,074131 | NA |
| 3 | abundance | X03837 | 2-Methoxy· | Radius_SOS | -0,05075 | 0,052967 | NA |
| 3 | abundance | X03837 | 2-Methoxy· | Tibia_SOS | 0,090654 | 0,056937 | NA |
| 3 | abundance | X03837 | 2-Methoxy· | Handgrip | 0,046847 | 0,062462 | NA |
| 3 | abundance | X03855 | 2-{(2S,4S)- | Tibia_length | 0,114374 | 0,051318 | NA |
| 3 | abundance | X03855 | 2-{(2S,4S)- | Radius_length | 0,092577 | 0,070369 | NA |
| 3 | abundance | X03855 | 2-{(2S,4S)- | Radius_SOS | 0,010003 | 0,049764 | NA |
| 3 | abundance | X03855 | 2-{(2S,4S)- | Tibia_SOS | 0,122544 | 0,052689 | NA |
| 3 | abundance | X03855 | 2-{(2S,4S)- | Handgrip | 0,063169 | 0,060566 | NA |
| 3 | abundance | X03860 | 2-BUTYL Pl | Tibia_length | 0,121148 | 0,053354 | NA |
| 3 | abundance | X03860 | 2-BUTYL Pl | Radius_length | 0,121199 | 0,072908 | NA |
| 3 | abundance | X03860 | 2-BUTYL Pl | Radius_SOS | 0,04737 | 0,051304 | NA |
| 3 | abundance | X03860 | 2-BUTYL Pl | Tibia_SOS | 0,135362 | 0,054428 | NA |
| 3 | abundance | X03860 | 2-BUTYL Pl | Handgrip | 0,086317 | 0,061322 | NA |
| 3 | abundance | X03892 | shinorine | Tibia_length | -0,06935 | 0,054642 | NA |
| 3 | abundance | X03892 | shinorine | Radius_length | -0,12684 | 0,073083 | NA |
| 3 | abundance | X03892 | shinorine | Radius_SOS | -0,1415 | 0,052029 | NA |
| 3 | abundance | X03892 | shinorine | Tibia_SOS | -0,12515 | 0,056132 | NA |
| 3 | abundance | X03892 | shinorine | Handgrip | -0,0343 | 0,062694 | NA |
| 3 | abundance | X03901 | ferrileghen | Tibia_length | -0,00442 | 0,049493 | NA |
| 3 | abundance | X03901 | ferrileghen | Radius_length | -0,07509 | 0,066963 | NA |
| 3 | abundance | X03901 | ferrileghen | Radius_SOS | 0,031687 | 0,049015 | NA |
| 3 | abundance | X03901 | ferrileghen | Tibia_SOS | 0,053069 | 0,052438 | NA |
| 3 | abundance | X03901 | ferrileghen | Handgrip | 0,081899 | 0,060162 | NA |
| 3 | abundance | X03916 | 2-(1-Ethox | Tibia_length | -0,04166 | 0,052182 | NA |
| 3 | abundance | X03916 | 2-(1-Ethox | Radius_length | -0,13215 | 0,071185 | NA |
| 3 | abundance | X03916 | 2-(1-Ethox | Radius_SOS | -0,02492 | 0,049895 | NA |
| 3 | abundance | X03916 | 2-(1-Ethox | Tibia_SOS | -0,05164 | 0,053574 | NA |
| 3 | abundance | X03916 | 2-(1-Ethox | Handgrip | 0,00338 | 0,061639 | NA |
| 3 | abundance | X03917 | Uramustin | Tibia_length | 0,072076 | 0,054258 | NA |
| 3 | abundance | X03917 | Uramustin | Radius_length | 0,119143 | 0,072493 | NA |
| 3 | abundance | X03917 | Uramustin | Radius_SOS | -0,00317 | 0,051844 | NA |
| 3 | abundance | X03917 | Uramustin | Tibia_SOS | 0,055331 | 0,055536 | NA |
| 3 | abundance | X03917 | Uramustin | Handgrip | -0,00893 | 0,062552 | NA |
| 3 | abundance | X03934 | 2-Aminom | Tibia_length | 0,098551 | 0,054292 | NA |
| 3 | abundance | X03934 | 2-Aminom | Radius_length | -0,07441 | 0,073142 | NA |
| 3 | abundance | X03934 | 2-Aminom | Radius_SOS | -0,03086 | 0,052045 | NA |
| 3 | abundance | X03934 | 2-Aminom | Tibia_SOS | 0,054179 | 0,054896 | NA |
| 3 | abundance | X03934 | 2-Aminom | Handgrip | 0,038435 | 0,06182 | NA |
| 3 | abundance | X03936 | (3R)-2-(3,4 | Tibia_length | -0,07006 | 0,05295 | NA |
| 3 | abundance | X03936 | (3R)-2-(3,4 | Radius_length | -0,12281 | 0,071423 | NA |
| 3 | abundance | X03936 | (3R)-2-(3,4 | Radius_SOS | 0,079663 | 0,050485 | NA |
| 3 | abundance | X03936 | (3R)-2-(3,4 | Tibia_SOS | -0,05387 | 0,054092 | NA |
| 3 | abundance | X03936 | (3R)-2-(3,4 | Handgrip | -0,00793 | 0,061327 | NA |
| 3 | abundance | X03961 | Artesunate | Tibia_length | -0,04258 | 0,052039 | NA |

| | | | | | | | | |
|---|---|---|---|---|---|---|---|---|
| | 3 | abundance | X03961 | Artesunate | Radius_length | 0,033613 | 0,071144 | NA |
| | 3 | abundance | X03961 | Artesunate | Radius_SOS | -0,00389 | 0,050092 | NA |
| | 3 | abundance | X03961 | Artesunate | Tibia_SOS | 0,09763 | 0,053903 | NA |
| | 3 | abundance | X03961 | Artesunate | Handgrip | 0,055527 | 0,061692 | NA |
| 2b | | abundance | X04021 | Bentazone | Tibia_length | 0,052097 | 0,054844 | NA |
| 2b | | abundance | X04021 | Bentazone | Radius_length | -0,02316 | 0,073481 | NA |
| 2b | | abundance | X04021 | Bentazone | Radius_SOS | 0,043325 | 0,052323 | NA |
| 2b | | abundance | X04021 | Bentazone | Tibia_SOS | -0,04142 | 0,055901 | NA |
| 2b | | abundance | X04021 | Bentazone | Handgrip | -0,0215 | 0,060179 | NA |
| | 3 | abundance | X04040 | 3-(3,4-dihy | Tibia_length | 0,003176 | 0,054265 | NA |
| | 3 | abundance | X04040 | 3-(3,4-dihy | Radius_length | 0,006687 | 0,073346 | NA |
| | 3 | abundance | X04040 | 3-(3,4-dihy | Radius_SOS | -0,01902 | 0,051622 | NA |
| | 3 | abundance | X04040 | 3-(3,4-dihy | Tibia_SOS | 0,073648 | 0,056976 | NA |
| | 3 | abundance | X04040 | 3-(3,4-dihy | Handgrip | -0,06119 | 0,062228 | NA |
| | 3 | abundance | X04117 | 2,3,4,5-tet | Tibia_length | 0,06057 | 0,05403 | NA |
| | 3 | abundance | X04117 | 2,3,4,5-tet | Radius_length | -0,01231 | 0,072906 | NA |
| | 3 | abundance | X04117 | 2,3,4,5-tet | Radius_SOS | -0,08698 | 0,051278 | NA |
| | 3 | abundance | X04117 | 2,3,4,5-tet | Tibia_SOS | -0,08971 | 0,05518 | NA |
| | 3 | abundance | X04117 | 2,3,4,5-tet | Handgrip | 0,009642 | 0,061967 | NA |
| | 3 | abundance | X04119 | Tetraacety | Tibia_length | -0,01926 | 0,051206 | NA |
| | 3 | abundance | X04119 | Tetraacety | Radius_length | 0,009732 | 0,070471 | NA |
| | 3 | abundance | X04119 | Tetraacety | Radius_SOS | -0,02322 | 0,049577 | NA |
| | 3 | abundance | X04119 | Tetraacety | Tibia_SOS | 0,057602 | 0,052793 | NA |
| | 3 | abundance | X04119 | Tetraacety | Handgrip | -0,06681 | 0,060859 | NA |
| | 3 | abundance | X04146 | L-fucopyra | Tibia_length | 0,003521 | 0,052077 | NA |
| | 3 | abundance | X04146 | L-fucopyra | Radius_length | 0,135575 | 0,07052 | NA |
| | 3 | abundance | X04146 | L-fucopyra | Radius_SOS | 0,063822 | 0,050066 | NA |
| | 3 | abundance | X04146 | L-fucopyra | Tibia_SOS | 0,110526 | 0,053886 | NA |
| | 3 | abundance | X04146 | L-fucopyra | Handgrip | 0,028714 | 0,061288 | NA |
| | 3 | abundance | X04150 | 4-(METHYL | Tibia_length | -0,11767 | 0,056307 | NA |
| | 3 | abundance | X04150 | 4-(METHYL | Radius_length | -0,04685 | 0,074472 | NA |
| | 3 | abundance | X04150 | 4-(METHYL | Radius_SOS | 0,027588 | 0,053642 | NA |
| | 3 | abundance | X04150 | 4-(METHYL | Tibia_SOS | -0,08171 | 0,055665 | NA |
| | 3 | abundance | X04150 | 4-(METHYL | Handgrip | -0,03818 | 0,063203 | NA |
| | 3 | abundance | X04157 | 3-Methoxy- | Tibia_length | -0,13822 | 0,054337 | NA |
| | 3 | abundance | X04157 | 3-Methoxy- | Radius_length | -0,14929 | 0,072694 | NA |
| | 3 | abundance | X04157 | 3-Methoxy- | Radius_SOS | -0,01693 | 0,051895 | NA |
| | 3 | abundance | X04157 | 3-Methoxy- | Tibia_SOS | -0,09629 | 0,055164 | NA |
| | 3 | abundance | X04157 | 3-Methoxy- | Handgrip | -0,16744 | 0,060508 | NA |
| | 3 | abundance | X04181 | 3-Methoxy- | Tibia_length | -0,1309 | 0,052618 | NA |
| | 3 | abundance | X04181 | 3-Methoxy- | Radius_length | -0,1617 | 0,071667 | NA |
| | 3 | abundance | X04181 | 3-Methoxy- | Radius_SOS | -0,08228 | 0,050981 | NA |
| | 3 | abundance | X04181 | 3-Methoxy- | Tibia_SOS | -0,2057 | 0,053721 | NA |
| | 3 | abundance | X04181 | 3-Methoxy- | Handgrip | -0,13192 | 0,060331 | NA |
| | 3 | abundance | X04183 | Spaglumic | Tibia_length | -0,00423 | 0,053883 | NA |
| | 3 | abundance | X04183 | Spaglumic | Radius_length | -0,03916 | 0,073184 | NA |
| | 3 | abundance | X04183 | Spaglumic | Radius_SOS | -0,05793 | 0,051585 | NA |
| | 3 | abundance | X04183 | Spaglumic | Tibia_SOS | 0,156053 | 0,052873 | NA |

| | | | | | | | |
|---|---|---|---|---|---|---|---|
| 3 | abundance | X04183 | Spaglumic | Handgrip | 0,151153 | 0,06136 | NA |
| 3 | abundance | X04229 | gamma-L-g | Tibia_length | 0,069597 | 0,053999 | NA |
| 3 | abundance | X04229 | gamma-L-g | Radius_length | 0,024724 | 0,07303 | NA |
| 3 | abundance | X04229 | gamma-L-g | Radius_SOS | -0,07498 | 0,051342 | NA |
| 3 | abundance | X04229 | gamma-L-g | Tibia_SOS | 0,090512 | 0,055551 | NA |
| 3 | abundance | X04229 | gamma-L-g | Handgrip | -0,01666 | 0,062514 | NA |
| 3 | abundance | X04259 | 3-Hydroxyt | Tibia_length | 0,041649 | 0,052266 | NA |
| 3 | abundance | X04259 | 3-Hydroxyt | Radius_length | 0,017609 | 0,071339 | NA |
| 3 | abundance | X04259 | 3-Hydroxyt | Radius_SOS | 0,00306 | 0,050295 | NA |
| 3 | abundance | X04259 | 3-Hydroxyt | Tibia_SOS | -0,00765 | 0,054012 | NA |
| 3 | abundance | X04259 | 3-Hydroxyt | Handgrip | -0,06301 | 0,061721 | NA |
| 3 | abundance | X04274 | 6-[(Z)-2-(3, | Tibia_length | 0,034708 | 0,050163 | NA |
| 3 | abundance | X04274 | 6-[(Z)-2-(3, | Radius_length | 0,145594 | 0,067785 | NA |
| 3 | abundance | X04274 | 6-[(Z)-2-(3, | Radius_SOS | 0,059909 | 0,049086 | NA |
| 3 | abundance | X04274 | 6-[(Z)-2-(3, | Tibia_SOS | 0,02155 | 0,052536 | NA |
| 3 | abundance | X04274 | 6-[(Z)-2-(3, | Handgrip | 0,112458 | 0,060421 | NA |
| 3 | abundance | X04286 | Zinecard | Tibia_length | -0,0355 | 0,054975 | NA |
| 3 | abundance | X04286 | Zinecard | Radius_length | -6,50E-04 | 0,073489 | NA |
| 3 | abundance | X04286 | Zinecard | Radius_SOS | -0,02027 | 0,052441 | NA |
| 3 | abundance | X04286 | Zinecard | Tibia_SOS | -0,00321 | 0,055851 | NA |
| 3 | abundance | X04286 | Zinecard | Handgrip | 0,001784 | 0,062284 | NA |
| 3 | abundance | X04310 | Aminohipp | Tibia_length | -0,0314 | 0,053192 | NA |
| 3 | abundance | X04310 | Aminohipp | Radius_length | 0,047388 | 0,072314 | NA |
| 3 | abundance | X04310 | Aminohipp | Radius_SOS | 0,008898 | 0,050859 | NA |
| 3 | abundance | X04310 | Aminohipp | Tibia_SOS | 0,059864 | 0,054535 | NA |
| 3 | abundance | X04310 | Aminohipp | Handgrip | 0,096257 | 0,061129 | NA |
| 3 | abundance | X04315 | Benzamide | Tibia_length | -0,01365 | 0,053567 | NA |
| 3 | abundance | X04315 | Benzamide | Radius_length | -0,11025 | 0,071952 | NA |
| 3 | abundance | X04315 | Benzamide | Radius_SOS | -0,03729 | 0,051057 | NA |
| 3 | abundance | X04315 | Benzamide | Tibia_SOS | 0,030388 | 0,054203 | NA |
| 3 | abundance | X04315 | Benzamide | Handgrip | -0,09302 | 0,060609 | NA |
| 3 | abundance | X04334 | 3-Benzyl-6 | Tibia_length | 0,093965 | 0,050662 | NA |
| 3 | abundance | X04334 | 3-Benzyl-6 | Radius_length | 0,139571 | 0,0693 | NA |
| 3 | abundance | X04334 | 3-Benzyl-6 | Radius_SOS | 0,053436 | 0,049372 | NA |
| 3 | abundance | X04334 | 3-Benzyl-6 | Tibia_SOS | 0,020399 | 0,052684 | NA |
| 3 | abundance | X04334 | 3-Benzyl-6 | Handgrip | -0,05554 | 0,060359 | NA |
| 3 | abundance | X04339 | L-Glutamic | Tibia_length | -0,19211 | 0,053343 | NA |
| 3 | abundance | X04339 | L-Glutamic | Radius_length | -0,05876 | 0,072997 | NA |
| 3 | abundance | X04339 | L-Glutamic | Radius_SOS | 0,01528 | 0,05171 | NA |
| 3 | abundance | X04339 | L-Glutamic | Tibia_SOS | -0,08043 | 0,056105 | NA |
| 3 | abundance | X04339 | L-Glutamic | Handgrip | -0,06934 | 0,061972 | NA |
| 3 | abundance | X04351 | Pyrimidine | Tibia_length | -0,09897 | 0,053816 | NA |
| 3 | abundance | X04351 | Pyrimidine | Radius_length | -0,09654 | 0,072736 | NA |
| 3 | abundance | X04351 | Pyrimidine | Radius_SOS | -0,09014 | 0,051531 | NA |
| 3 | abundance | X04351 | Pyrimidine | Tibia_SOS | 0,148846 | 0,052993 | NA |
| 3 | abundance | X04351 | Pyrimidine | Handgrip | 0,089136 | 0,061751 | NA |
| 3 | abundance | X04377 | Dihydroura | Tibia_length | -0,05375 | 0,053322 | NA |
| 3 | abundance | X04377 | Dihydroura | Radius_length | -0,01681 | 0,072449 | NA |

| | | | | | | | |
|---|---|---|---|---|---|---|---|
| 3 | abundance | X04377 | Dihydroura | Radius_SOS | -0,03491 | 0,051066 | NA |
| 3 | abundance | X04377 | Dihydroura | Tibia_SOS | 0,110452 | 0,053181 | NA |
| 3 | abundance | X04377 | Dihydroura | Handgrip | 0,055587 | 0,061741 | NA |
| 3 | abundance | X04378 | 9,11-Dihyc | Tibia_length | -0,10803 | 0,053348 | NA |
| 3 | abundance | X04378 | 9,11-Dihyc | Radius_length | -0,19534 | 0,070959 | NA |
| 3 | abundance | X04378 | 9,11-Dihyc | Radius_SOS | -0,0256 | 0,051008 | NA |
| 3 | abundance | X04378 | 9,11-Dihyc | Tibia_SOS | 0,029294 | 0,053794 | NA |
| 3 | abundance | X04378 | 9,11-Dihyc | Handgrip | -0,01113 | 0,061212 | NA |
| 3 | abundance | X04379 | 2-Amino-6 | Tibia_length | -0,0129 | 0,053585 | NA |
| 3 | abundance | X04379 | 2-Amino-6 | Radius_length | 0,077202 | 0,0725 | NA |
| 3 | abundance | X04379 | 2-Amino-6 | Radius_SOS | -5,56E-04 | 0,05118 | NA |
| 3 | abundance | X04379 | 2-Amino-6 | Tibia_SOS | -0,06895 | 0,054985 | NA |
| 3 | abundance | X04379 | 2-Amino-6 | Handgrip | 0,055832 | 0,061812 | NA |
| 3 | abundance | X04440 | Mono(3-ca | Tibia_length | -0,06483 | 0,056577 | NA |
| 3 | abundance | X04440 | Mono(3-ca | Radius_length | -0,07247 | 0,074616 | NA |
| 3 | abundance | X04440 | Mono(3-ca | Radius_SOS | -0,02511 | 0,053635 | NA |
| 3 | abundance | X04440 | Mono(3-ca | Tibia_SOS | -0,06839 | 0,056351 | NA |
| 3 | abundance | X04440 | Mono(3-ca | Handgrip | -0,13461 | 0,06293 | NA |
| 3 | abundance | X04450 | 3-Methylac | Tibia_length | -0,03943 | 0,048933 | NA |
| 3 | abundance | X04450 | 3-Methylac | Radius_length | -0,0999 | 0,062796 | NA |
| 3 | abundance | X04450 | 3-Methylac | Radius_SOS | 0,006484 | 0,049643 | NA |
| 3 | abundance | X04450 | 3-Methylac | Tibia_SOS | 0,009815 | 0,052768 | NA |
| 3 | abundance | X04450 | 3-Methylac | Handgrip | 0,041087 | 0,060153 | NA |
| 3 | abundance | X04483 | (2E)-5-Hyd | Tibia_length | -0,11936 | 0,052373 | NA |
| 3 | abundance | X04483 | (2E)-5-Hyd | Radius_length | -0,21178 | 0,070109 | NA |
| 3 | abundance | X04483 | (2E)-5-Hyd | Radius_SOS | -0,00457 | 0,050285 | NA |
| 3 | abundance | X04483 | (2E)-5-Hyd | Tibia_SOS | -0,11991 | 0,053907 | NA |
| 3 | abundance | X04483 | (2E)-5-Hyd | Handgrip | -0,08083 | 0,061125 | NA |
| 3 | abundance | X04493 | Tyrosol | Tibia_length | -0,10841 | 0,05304 | NA |
| 3 | abundance | X04493 | Tyrosol | Radius_length | -0,13896 | 0,071565 | NA |
| 3 | abundance | X04493 | Tyrosol | Radius_SOS | -0,02109 | 0,051043 | NA |
| 3 | abundance | X04493 | Tyrosol | Tibia_SOS | -0,03185 | 0,053665 | NA |
| 3 | abundance | X04493 | Tyrosol | Handgrip | -0,0732 | 0,060843 | NA |
| 3 | abundance | X04526 | N-Nonanoy | Tibia_length | 0,024383 | 0,05469 | NA |
| 3 | abundance | X04526 | N-Nonanoy | Radius_length | 0,119188 | 0,072883 | NA |
| 3 | abundance | X04526 | N-Nonanoy | Radius_SOS | -0,04639 | 0,051968 | NA |
| 3 | abundance | X04526 | N-Nonanoy | Tibia_SOS | 0,074725 | 0,055821 | NA |
| 3 | abundance | X04526 | N-Nonanoy | Handgrip | 0,028474 | 0,061966 | NA |
| 3 | abundance | X04538 | Valylvaline | Tibia_length | 0,154381 | 0,054328 | NA |
| 3 | abundance | X04538 | Valylvaline | Radius_length | 0,103311 | 0,073546 | NA |
| 3 | abundance | X04538 | Valylvaline | Radius_SOS | 0,072244 | 0,052265 | NA |
| 3 | abundance | X04538 | Valylvaline | Tibia_SOS | 0,088308 | 0,056615 | NA |
| 3 | abundance | X04538 | Valylvaline | Handgrip | -0,00802 | 0,061912 | NA |
| 3 | abundance | X04543 | N-Acetyl-S | Tibia_length | -0,1728 | 0,053381 | NA |
| 3 | abundance | X04543 | N-Acetyl-S | Radius_length | -0,16234 | 0,071906 | NA |
| 3 | abundance | X04543 | N-Acetyl-S | Radius_SOS | -0,0364 | 0,05136 | NA |
| 3 | abundance | X04543 | N-Acetyl-S | Tibia_SOS | -0,05753 | 0,054388 | NA |
| 3 | abundance | X04543 | N-Acetyl-S | Handgrip | -0,00916 | 0,061954 | NA |

| | | | | | | | |
|---|---|---|---|---|---|---|---|
| 3 | abundance | X04544 | L-gamma-( | Tibia_length | -0,02932 | 0,055092 | NA |
| 3 | abundance | X04544 | L-gamma-( | Radius_length | -0,10594 | 0,073181 | NA |
| 3 | abundance | X04544 | L-gamma-( | Radius_SOS | 0,011775 | 0,052396 | NA |
| 3 | abundance | X04544 | L-gamma-( | Tibia_SOS | -0,10166 | 0,055239 | NA |
| 3 | abundance | X04544 | L-gamma-( | Handgrip | -0,25093 | 0,060995 | NA |
| 3 | abundance | X04553 | N-(4-Amin | Tibia_length | 0,027701 | 0,054781 | NA |
| 3 | abundance | X04553 | N-(4-Amin | Radius_length | -0,04549 | 0,074387 | NA |
| 3 | abundance | X04553 | N-(4-Amin | Radius_SOS | 0,009745 | 0,052189 | NA |
| 3 | abundance | X04553 | N-(4-Amin | Tibia_SOS | 0,042402 | 0,056042 | NA |
| 3 | abundance | X04553 | N-(4-Amin | Handgrip | 0,012018 | 0,062992 | NA |
| 3 | abundance | X04557 | 1,4-Napht | Tibia_length | 0,060851 | 0,05098 | NA |
| 3 | abundance | X04557 | 1,4-Napht | Radius_length | 0,172622 | 0,068967 | NA |
| 3 | abundance | X04557 | 1,4-Napht | Radius_SOS | 0,103933 | 0,049342 | NA |
| 3 | abundance | X04557 | 1,4-Napht | Tibia_SOS | 0,174925 | 0,052776 | NA |
| 3 | abundance | X04557 | 1,4-Napht | Handgrip | 0,116019 | 0,06041 | NA |
| 3 | abundance | X04562 | Lisdexamf | Tibia_length | -0,00806 | 0,05299 | NA |
| 3 | abundance | X04562 | Lisdexamf | Radius_length | -0,00715 | 0,072019 | NA |
| 3 | abundance | X04562 | Lisdexamf | Radius_SOS | -0,06261 | 0,050733 | NA |
| 3 | abundance | X04562 | Lisdexamf | Tibia_SOS | -0,05219 | 0,054634 | NA |
| 3 | abundance | X04562 | Lisdexamf | Handgrip | 0,051557 | 0,061665 | NA |
| 3 | abundance | X04564 | asp-gln | Tibia_length | -0,05758 | 0,053542 | NA |
| 3 | abundance | X04564 | asp-gln | Radius_length | 0,054167 | 0,072428 | NA |
| 3 | abundance | X04564 | asp-gln | Radius_SOS | 0,013695 | 0,050935 | NA |
| 3 | abundance | X04564 | asp-gln | Tibia_SOS | 0,082092 | 0,054334 | NA |
| 3 | abundance | X04564 | asp-gln | Handgrip | 0,026374 | 0,061988 | NA |
| 3 | abundance | X04566 | pterin | Tibia_length | -0,05306 | 0,053836 | NA |
| 3 | abundance | X04566 | pterin | Radius_length | 0,018692 | 0,07277 | NA |
| 3 | abundance | X04566 | pterin | Radius_SOS | 0,032307 | 0,051401 | NA |
| 3 | abundance | X04566 | pterin | Tibia_SOS | 0,042306 | 0,055947 | NA |
| 3 | abundance | X04566 | pterin | Handgrip | -0,02564 | 0,062116 | NA |
| 3 | abundance | X04571 | 13a-Hydro | Tibia_length | -0,04686 | 0,058307 | NA |
| 3 | abundance | X04571 | 13a-Hydro | Radius_length | -0,09657 | 0,074699 | NA |
| 3 | abundance | X04571 | 13a-Hydro | Radius_SOS | -0,05779 | 0,055644 | NA |
| 3 | abundance | X04571 | 13a-Hydro | Tibia_SOS | -0,00681 | 0,058309 | NA |
| 3 | abundance | X04571 | 13a-Hydro | Handgrip | -0,12384 | 0,05973 | NA |
| 3 | abundance | X04579 | 6-(alpha-D | Tibia_length | -0,12174 | 0,054171 | NA |
| 3 | abundance | X04579 | 6-(alpha-D | Radius_length | -0,1076 | 0,072926 | NA |
| 3 | abundance | X04579 | 6-(alpha-D | Radius_SOS | -0,0702 | 0,051971 | NA |
| 3 | abundance | X04579 | 6-(alpha-D | Tibia_SOS | -0,03657 | 0,054871 | NA |
| 3 | abundance | X04579 | 6-(alpha-D | Handgrip | -0,02313 | 0,061592 | NA |
| 3 | abundance | X04582 | Dinoseb | Tibia_length | -0,07468 | 0,053884 | NA |
| 3 | abundance | X04582 | Dinoseb | Radius_length | -0,07891 | 0,072427 | NA |
| 3 | abundance | X04582 | Dinoseb | Radius_SOS | -0,06597 | 0,050985 | NA |
| 3 | abundance | X04582 | Dinoseb | Tibia_SOS | -0,06061 | 0,054064 | NA |
| 3 | abundance | X04582 | Dinoseb | Handgrip | -0,0593 | 0,061369 | NA |
| 3 | abundance | X04593 | Methional | Tibia_length | 0,108659 | 0,053878 | NA |
| 3 | abundance | X04593 | Methional | Radius_length | 0,189573 | 0,072151 | NA |
| 3 | abundance | X04593 | Methional | Radius_SOS | 0,084529 | 0,051055 | NA |

| | | | | | | | |
|---|---|---|---|---|---|---|---|
| 3 | abundance | X04593 | Methional | Tibia_SOS | 0,132233 | 0,056204 | NA |
| 3 | abundance | X04593 | Methional | Handgrip | 0,166006 | 0,060606 | NA |
| 3 | abundance | X04636 | epsilon-(ga | Tibia_length | -0,02348 | 0,050936 | NA |
| 3 | abundance | X04636 | epsilon-(ga | Radius_length | 0,033932 | 0,069813 | NA |
| 3 | abundance | X04636 | epsilon-(ga | Radius_SOS | 0,035282 | 0,049357 | NA |
| 3 | abundance | X04636 | epsilon-(ga | Tibia_SOS | -0,01753 | 0,052762 | NA |
| 3 | abundance | X04636 | epsilon-(ga | Handgrip | 0,071736 | 0,060206 | NA |
| 3 | abundance | X04639 | hydroxyhe› | Tibia_length | -0,06273 | 0,051289 | NA |
| 3 | abundance | X04639 | hydroxyhe› | Radius_length | -0,02781 | 0,070449 | NA |
| 3 | abundance | X04639 | hydroxyhe› | Radius_SOS | 0,020037 | 0,049566 | NA |
| 3 | abundance | X04639 | hydroxyhe› | Tibia_SOS | -0,01021 | 0,052779 | NA |
| 3 | abundance | X04639 | hydroxyhe› | Handgrip | -8,40E-04 | 0,061209 | NA |
| 3 | abundance | X04682 | 1-Methylin | Tibia_length | 0,045623 | 0,050908 | NA |
| 3 | abundance | X04682 | 1-Methylin | Radius_length | 0,015192 | 0,069582 | NA |
| 3 | abundance | X04682 | 1-Methylin | Radius_SOS | -0,04858 | 0,049198 | NA |
| 3 | abundance | X04682 | 1-Methylin | Tibia_SOS | -0,01289 | 0,052894 | NA |
| 3 | abundance | X04682 | 1-Methylin | Handgrip | 0,031538 | 0,060899 | NA |
| 3 | abundance | X04684 | AAMU | Tibia_length | -0,13334 | 0,05137 | NA |
| 3 | abundance | X04684 | AAMU | Radius_length | -0,20644 | 0,069402 | NA |
| 3 | abundance | X04684 | AAMU | Radius_SOS | -0,04256 | 0,049823 | NA |
| 3 | abundance | X04684 | AAMU | Tibia_SOS | -0,0834 | 0,052873 | NA |
| 3 | abundance | X04684 | AAMU | Handgrip | -0,11549 | 0,060414 | NA |
| 3 | abundance | X04688 | Minoxidil | Tibia_length | -0,09607 | 0,055939 | NA |
| 3 | abundance | X04688 | Minoxidil | Radius_length | -0,20095 | 0,072946 | NA |
| 3 | abundance | X04688 | Minoxidil | Radius_SOS | 0,01793 | 0,053158 | NA |
| 3 | abundance | X04688 | Minoxidil | Tibia_SOS | -8,75E-04 | 0,05729 | NA |
| 3 | abundance | X04688 | Minoxidil | Handgrip | 0,053966 | 0,06231 | NA |
| 3 | abundance | X04695 | N-[(4-Meth | Tibia_length | -0,04819 | 0,048913 | NA |
| 3 | abundance | X04695 | N-[(4-Meth | Radius_length | -0,04525 | 0,04892 | NA |
| 3 | abundance | X04695 | N-[(4-Meth | Radius_SOS | -0,05768 | 0,048947 | NA |
| 3 | abundance | X04695 | N-[(4-Meth | Tibia_SOS | -0,00614 | 0,052485 | NA |
| 3 | abundance | X04695 | N-[(4-Meth | Handgrip | -0,02588 | 0,060173 | NA |
| 3 | abundance | X04697 | Ethosuxim | Tibia_length | 0,007677 | 0,056433 | NA |
| 3 | abundance | X04697 | Ethosuxim | Radius_length | 0,025033 | 0,074599 | NA |
| 3 | abundance | X04697 | Ethosuxim | Radius_SOS | 0,05822 | 0,053921 | NA |
| 3 | abundance | X04697 | Ethosuxim | Tibia_SOS | -0,01639 | 0,057506 | NA |
| 3 | abundance | X04697 | Ethosuxim | Handgrip | -0,06552 | 0,062946 | NA |
| 3 | abundance | X04743 | 5-Methoxy- | Tibia_length | -0,12753 | 0,052456 | NA |
| 3 | abundance | X04743 | 5-Methoxy- | Radius_length | -0,18912 | 0,070635 | NA |
| 3 | abundance | X04743 | 5-Methoxy- | Radius_SOS | -0,01346 | 0,05015 | NA |
| 3 | abundance | X04743 | 5-Methoxy- | Tibia_SOS | -0,03861 | 0,053797 | NA |
| 3 | abundance | X04743 | 5-Methoxy- | Handgrip | 0,010782 | 0,061153 | NA |
| 3 | abundance | X04744 | 16alpha-hy | Tibia_length | -0,0626 | 0,054764 | NA |
| 3 | abundance | X04744 | 16alpha-hy | Radius_length | 0,065337 | 0,072942 | NA |
| 3 | abundance | X04744 | 16alpha-hy | Radius_SOS | -0,01124 | 0,052173 | NA |
| 3 | abundance | X04744 | 16alpha-hy | Tibia_SOS | 0,014056 | 0,056905 | NA |
| 3 | abundance | X04744 | 16alpha-hy | Handgrip | 0,047513 | 0,06306 | NA |
| 3 | abundance | X04759 | 7-Aminom | Tibia_length | -0,02595 | 0,051044 | NA |

| | | | | | | | |
|---|---|---|---|---|---|---|---|
| 3 | abundance | X04759 | 7-Aminom | Radius_length | 0,002652 | 0,070021 | NA |
| 3 | abundance | X04759 | 7-Aminom | Radius_SOS | -0,01507 | 0,049484 | NA |
| 3 | abundance | X04759 | 7-Aminom | Tibia_SOS | 0,011796 | 0,053014 | NA |
| 3 | abundance | X04759 | 7-Aminom | Handgrip | 0,034785 | 0,060788 | NA |
| 3 | abundance | X04788 | 2-(4-Isopro | Tibia_length | 0,115183 | 0,0529 | NA |
| 3 | abundance | X04788 | 2-(4-Isopro | Radius_length | 0,127804 | 0,071963 | NA |
| 3 | abundance | X04788 | 2-(4-Isopro | Radius_SOS | 0,011732 | 0,050609 | NA |
| 3 | abundance | X04788 | 2-(4-Isopro | Tibia_SOS | 0,128503 | 0,05459 | NA |
| 3 | abundance | X04788 | 2-(4-Isopro | Handgrip | 0,100959 | 0,061003 | NA |
| 3 | abundance | X04792 | his-asn | Tibia_length | 0,034947 | 0,054975 | NA |
| 3 | abundance | X04792 | his-asn | Radius_length | 0,096484 | 0,07352 | NA |
| 3 | abundance | X04792 | his-asn | Radius_SOS | 0,022083 | 0,052251 | NA |
| 3 | abundance | X04792 | his-asn | Tibia_SOS | 0,089294 | 0,055822 | NA |
| 3 | abundance | X04792 | his-asn | Handgrip | 0,140905 | 0,059592 | NA |
| 3 | abundance | X04814 | meprobam | Tibia_length | -0,07862 | 0,053226 | NA |
| 3 | abundance | X04814 | meprobam | Radius_length | -0,10195 | 0,071621 | NA |
| 3 | abundance | X04814 | meprobam | Radius_SOS | -0,02773 | 0,050865 | NA |
| 3 | abundance | X04814 | meprobam | Tibia_SOS | -0,06547 | 0,05408 | NA |
| 3 | abundance | X04814 | meprobam | Handgrip | 0,044943 | 0,061671 | NA |
| 3 | abundance | X04822 | 6-Sulfatoxy | Tibia_length | 0,146474 | 0,052547 | NA |
| 3 | abundance | X04822 | 6-Sulfatoxy | Radius_length | 0,224373 | 0,06999 | NA |
| 3 | abundance | X04822 | 6-Sulfatoxy | Radius_SOS | 0,080234 | 0,050361 | NA |
| 3 | abundance | X04822 | 6-Sulfatoxy | Tibia_SOS | 0,142676 | 0,053398 | NA |
| 3 | abundance | X04822 | 6-Sulfatoxy | Handgrip | 0,161954 | 0,060426 | NA |
| 3 | abundance | X04852 | Toluene | Tibia_length | 0,062891 | 0,056708 | NA |
| 3 | abundance | X04852 | Toluene | Radius_length | -0,06967 | 0,074682 | NA |
| 3 | abundance | X04852 | Toluene | Radius_SOS | -0,03105 | 0,054136 | NA |
| 3 | abundance | X04852 | Toluene | Tibia_SOS | 0,074163 | 0,058401 | NA |
| 3 | abundance | X04852 | Toluene | Handgrip | 0,006529 | 0,063829 | NA |
| 3 | abundance | X04877 | Trifluorom | Tibia_length | -0,03269 | 0,052843 | NA |
| 3 | abundance | X04877 | Trifluorom | Radius_length | -4,25E-04 | 0,071921 | NA |
| 3 | abundance | X04877 | Trifluorom | Radius_SOS | -0,04781 | 0,050621 | NA |
| 3 | abundance | X04877 | Trifluorom | Tibia_SOS | 0,033659 | 0,053834 | NA |
| 3 | abundance | X04877 | Trifluorom | Handgrip | 0,085664 | 0,061233 | NA |
| 3 | abundance | X04897 | Taxifolin | Tibia_length | 0,091633 | 0,051572 | NA |
| 3 | abundance | X04897 | Taxifolin | Radius_length | 0,104839 | 0,070672 | NA |
| 3 | abundance | X04897 | Taxifolin | Radius_SOS | -0,02522 | 0,049832 | NA |
| 3 | abundance | X04897 | Taxifolin | Tibia_SOS | 0,090502 | 0,052892 | NA |
| 3 | abundance | X04897 | Taxifolin | Handgrip | 0,121333 | 0,061152 | NA |
| 3 | abundance | X04898 | 17-Hydroxy | Tibia_length | 0,019404 | 0,050616 | NA |
| 3 | abundance | X04898 | 17-Hydroxy | Radius_length | 0,014218 | 0,069157 | NA |
| 3 | abundance | X04898 | 17-Hydroxy | Radius_SOS | 0,012545 | 0,049244 | NA |
| 3 | abundance | X04898 | 17-Hydroxy | Tibia_SOS | 0,024923 | 0,05314 | NA |
| 3 | abundance | X04898 | 17-Hydroxy | Handgrip | -0,01257 | 0,061323 | NA |
| 3 | abundance | X04919 | 5-O-alpha- | Tibia_length | -0,12821 | 0,057073 | NA |
| 3 | abundance | X04919 | 5-O-alpha- | Radius_length | 0,004503 | 0,075525 | NA |
| 3 | abundance | X04919 | 5-O-alpha- | Radius_SOS | -0,06918 | 0,054482 | NA |
| 3 | abundance | X04919 | 5-O-alpha- | Tibia_SOS | -0,01828 | 0,059268 | NA |

| | | | | | | | |
|---|---|---|---|---|---|---|---|
| 3 | abundance | X04919 | 5-O-alpha- | Handgrip | -0,03088 | 0,062695 | NA |
| 3 | abundance | X04922 | Sinapinic a | Tibia_length | -0,07997 | 0,053735 | NA |
| 3 | abundance | X04922 | Sinapinic a | Radius_length | -0,17822 | 0,071657 | NA |
| 3 | abundance | X04922 | Sinapinic a | Radius_SOS | 0,00221 | 0,051386 | NA |
| 3 | abundance | X04922 | Sinapinic a | Tibia_SOS | -0,06121 | 0,054801 | NA |
| 3 | abundance | X04922 | Sinapinic a | Handgrip | -0,11052 | 0,061186 | NA |
| 3 | abundance | X04939 | N-Desalkyl | Tibia_length | -0,11399 | 0,053833 | NA |
| 3 | abundance | X04939 | N-Desalkyl | Radius_length | -0,17704 | 0,071924 | NA |
| 3 | abundance | X04939 | N-Desalkyl | Radius_SOS | -0,11289 | 0,052083 | NA |
| 3 | abundance | X04939 | N-Desalkyl | Tibia_SOS | 0,004675 | 0,05554 | NA |
| 3 | abundance | X04939 | N-Desalkyl | Handgrip | -0,05878 | 0,061925 | NA |
| 3 | abundance | X04975 | MFCD0995 | Tibia_length | 0,119025 | 0,054022 | NA |
| 3 | abundance | X04975 | MFCD0995 | Radius_length | 0,105006 | 0,073041 | NA |
| 3 | abundance | X04975 | MFCD0995 | Radius_SOS | 0,11586 | 0,05156 | NA |
| 3 | abundance | X04975 | MFCD0995 | Tibia_SOS | 0,110767 | 0,055592 | NA |
| 3 | abundance | X04975 | MFCD0995 | Handgrip | 0,06349 | 0,061945 | NA |
| 3 | abundance | X04986 | L-Proline, 4 | Tibia_length | -0,08087 | 0,057625 | NA |
| 3 | abundance | X04986 | L-Proline, 4 | Radius_length | -0,14865 | 0,074703 | NA |
| 3 | abundance | X04986 | L-Proline, 4 | Radius_SOS | 0,055089 | 0,054754 | NA |
| 3 | abundance | X04986 | L-Proline, 4 | Tibia_SOS | 0,006251 | 0,059203 | NA |
| 3 | abundance | X04986 | L-Proline, 4 | Handgrip | -0,02857 | 0,062494 | NA |
| 3 | abundance | X04990 | Cilazapril | Tibia_length | 0,090416 | 0,052095 | NA |
| 3 | abundance | X04990 | Cilazapril | Radius_length | 0,105078 | 0,070776 | NA |
| 3 | abundance | X04990 | Cilazapril | Radius_SOS | 0,037388 | 0,050002 | NA |
| 3 | abundance | X04990 | Cilazapril | Tibia_SOS | 0,104487 | 0,05383 | NA |
| 3 | abundance | X04990 | Cilazapril | Handgrip | -0,0159 | 0,061843 | NA |
| 3 | abundance | X04996 | MFCD0272 | Tibia_length | -0,11349 | 0,054244 | NA |
| 3 | abundance | X04996 | MFCD0272 | Radius_length | -0,19976 | 0,071612 | NA |
| 3 | abundance | X04996 | MFCD0272 | Radius_SOS | -0,00979 | 0,051351 | NA |
| 3 | abundance | X04996 | MFCD0272 | Tibia_SOS | -0,10754 | 0,054567 | NA |
| 3 | abundance | X04996 | MFCD0272 | Handgrip | -0,08433 | 0,061106 | NA |
| 1 | abundance | X04999 | 2-Hydroxyp | Tibia_length | -0,14287 | 0,052941 | NA |
| 1 | abundance | X04999 | 2-Hydroxyp | Radius_length | -0,08564 | 0,071949 | NA |
| 1 | abundance | X04999 | 2-Hydroxyp | Radius_SOS | -0,04954 | 0,050865 | NA |
| 1 | abundance | X04999 | 2-Hydroxyp | Tibia_SOS | -0,12464 | 0,053607 | NA |
| 1 | abundance | X04999 | 2-Hydroxyp | Handgrip | -0,03095 | 0,061296 | NA |
| 3 | abundance | X05017 | N-(1-{[Metl | Tibia_length | -0,07801 | 0,053994 | NA |
| 3 | abundance | X05017 | N-(1-{[Metl | Radius_length | 0,01554 | 0,073591 | NA |
| 3 | abundance | X05017 | N-(1-{[Metl | Radius_SOS | -0,10193 | 0,051677 | NA |
| 3 | abundance | X05017 | N-(1-{[Metl | Tibia_SOS | 0,015959 | 0,054708 | NA |
| 3 | abundance | X05017 | N-(1-{[Metl | Handgrip | 0,073357 | 0,061875 | NA |
| 3 | abundance | X05029 | Hypericin | Tibia_length | 0,159747 | 0,05145 | NA |
| 3 | abundance | X05029 | Hypericin | Radius_length | 0,198155 | 0,069424 | NA |
| 3 | abundance | X05029 | Hypericin | Radius_SOS | 0,083323 | 0,049575 | NA |
| 3 | abundance | X05029 | Hypericin | Tibia_SOS | 0,12815 | 0,052887 | NA |
| 3 | abundance | X05029 | Hypericin | Handgrip | 0,131787 | 0,060134 | NA |
| 3 | abundance | X05033 | (2E)-N-3,7- | Tibia_length | 0,077261 | 0,052782 | NA |
| 3 | abundance | X05033 | (2E)-N-3,7- | Radius_length | 0,127394 | 0,07159 | NA |

| | | | | | | | | |
|---|---|---|---|---|---|---|---|---|
| | 3 | abundance | X05033 | (2E)-N-3,7· | Radius_SOS | 0,050618 | 0,050591 | NA |
| | 3 | abundance | X05033 | (2E)-N-3,7· | Tibia_SOS | 0,114958 | 0,053919 | NA |
| | 3 | abundance | X05033 | (2E)-N-3,7· | Handgrip | 0,166641 | 0,060307 | NA |
| | 3 | abundance | X05046 | Casimiroin | Tibia_length | -0,12026 | 0,057503 | NA |
| | 3 | abundance | X05046 | Casimiroin | Radius_length | -0,04843 | 0,074893 | NA |
| | 3 | abundance | X05046 | Casimiroin | Radius_SOS | -0,03487 | 0,055208 | NA |
| | 3 | abundance | X05046 | Casimiroin | Tibia_SOS | 0,015293 | 0,058124 | NA |
| | 3 | abundance | X05046 | Casimiroin | Handgrip | 0,012681 | 0,061763 | NA |
| | 3 | abundance | X05068 | pro-gln | Tibia_length | -0,11495 | 0,054444 | NA |
| | 3 | abundance | X05068 | pro-gln | Radius_length | -0,09099 | 0,072805 | NA |
| | 3 | abundance | X05068 | pro-gln | Radius_SOS | -0,04142 | 0,051945 | NA |
| | 3 | abundance | X05068 | pro-gln | Tibia_SOS | -0,15194 | 0,055248 | NA |
| | 3 | abundance | X05068 | pro-gln | Handgrip | -0,07971 | 0,061406 | NA |
| | 3 | abundance | X05071 | Histidylphe | Tibia_length | -3,36E-04 | 0,0583 | NA |
| | 3 | abundance | X05071 | Histidylphe | Radius_length | 0,001006 | 0,075843 | NA |
| | 3 | abundance | X05071 | Histidylphe | Radius_SOS | -0,01884 | 0,055481 | NA |
| | 3 | abundance | X05071 | Histidylphe | Tibia_SOS | 0,016123 | 0,058091 | NA |
| | 3 | abundance | X05071 | Histidylphe | Handgrip | -0,07397 | 0,060028 | NA |
| | 3 | abundance | X05072 | L-gamma-( | Tibia_length | -0,07384 | 0,052074 | NA |
| | 3 | abundance | X05072 | L-gamma-( | Radius_length | -0,06852 | 0,07097 | NA |
| | 3 | abundance | X05072 | L-gamma-( | Radius_SOS | 2,91E-04 | 0,04998 | NA |
| | 3 | abundance | X05072 | L-gamma-( | Tibia_SOS | -0,08289 | 0,053632 | NA |
| | 3 | abundance | X05072 | L-gamma-( | Handgrip | -0,0088 | 0,061256 | NA |
| 2b | | abundance | X05081 | ELK (Peptic | Tibia_length | -0,05935 | 0,055499 | NA |
| 2b | | abundance | X05081 | ELK (Peptic | Radius_length | 0,005442 | 0,073838 | NA |
| 2b | | abundance | X05081 | ELK (Peptic | Radius_SOS | 0,034576 | 0,052863 | NA |
| 2b | | abundance | X05081 | ELK (Peptic | Tibia_SOS | 0,013283 | 0,055824 | NA |
| 2b | | abundance | X05081 | ELK (Peptic | Handgrip | 0,031272 | 0,062132 | NA |
| | 3 | abundance | X05126 | (3aS,5S,6F | Tibia_length | 0,012505 | 0,050531 | NA |
| | 3 | abundance | X05126 | (3aS,5S,6F | Radius_length | 0,085293 | 0,068954 | NA |
| | 3 | abundance | X05126 | (3aS,5S,6F | Radius_SOS | 0,029314 | 0,049204 | NA |
| | 3 | abundance | X05126 | (3aS,5S,6F | Tibia_SOS | -0,05533 | 0,052474 | NA |
| | 3 | abundance | X05126 | (3aS,5S,6F | Handgrip | 0,009086 | 0,060298 | NA |
| | 3 | abundance | X05146 | UQ367500 | Tibia_length | 0,007026 | 0,053454 | NA |
| | 3 | abundance | X05146 | UQ367500 | Radius_length | -0,11075 | 0,072431 | NA |
| | 3 | abundance | X05146 | UQ367500 | Radius_SOS | 0,002076 | 0,051054 | NA |
| | 3 | abundance | X05146 | UQ367500 | Tibia_SOS | 0,099097 | 0,055224 | NA |
| | 3 | abundance | X05146 | UQ367500 | Handgrip | 0,0607 | 0,063002 | NA |
| | 3 | abundance | X05149 | S-Sulfocys | Tibia_length | 0,107982 | 0,05027 | NA |
| | 3 | abundance | X05149 | S-Sulfocys | Radius_length | 0,123272 | 0,068527 | NA |
| | 3 | abundance | X05149 | S-Sulfocys | Radius_SOS | 0,007156 | 0,049188 | NA |
| | 3 | abundance | X05149 | S-Sulfocys | Tibia_SOS | 0,08751 | 0,052566 | NA |
| | 3 | abundance | X05149 | S-Sulfocys | Handgrip | 0,083899 | 0,060046 | NA |
| | 3 | abundance | X05177 | cis-3-Hexe | Tibia_length | -0,1006 | 0,05563 | NA |
| | 3 | abundance | X05177 | cis-3-Hexe | Radius_length | -0,03237 | 0,073876 | NA |
| | 3 | abundance | X05177 | cis-3-Hexe | Radius_SOS | 0,002912 | 0,053071 | NA |
| | 3 | abundance | X05177 | cis-3-Hexe | Tibia_SOS | 0,054571 | 0,055645 | NA |
| | 3 | abundance | X05177 | cis-3-Hexe | Handgrip | -0,00723 | 0,061627 | NA |

| | | | | | | | |
|---|---|---|---|---|---|---|---|
| 3 | abundance | X05183 | 3,6-Dichlo | Tibia_length | 0,006434 | 0,054261 | NA |
| 3 | abundance | X05183 | 3,6-Dichlo | Radius_length | -0,00169 | 0,073108 | NA |
| 3 | abundance | X05183 | 3,6-Dichlo | Radius_SOS | 0,028467 | 0,051726 | NA |
| 3 | abundance | X05183 | 3,6-Dichlo | Tibia_SOS | -0,01799 | 0,055278 | NA |
| 3 | abundance | X05183 | 3,6-Dichlo | Handgrip | 0,024267 | 0,062534 | NA |
| 3 | abundance | X05195 | Lys-phe | Tibia_length | -0,02046 | 0,05644 | NA |
| 3 | abundance | X05195 | Lys-phe | Radius_length | -0,06424 | 0,074227 | NA |
| 3 | abundance | X05195 | Lys-phe | Radius_SOS | 0,033792 | 0,053619 | NA |
| 3 | abundance | X05195 | Lys-phe | Tibia_SOS | -0,0878 | 0,055837 | NA |
| 3 | abundance | X05195 | Lys-phe | Handgrip | 0,032923 | 0,062726 | NA |
| 3 | abundance | X05203 | felbamate | Tibia_length | -0,15821 | 0,057004 | NA |
| 3 | abundance | X05203 | felbamate | Radius_length | -0,02083 | 0,075056 | NA |
| 3 | abundance | X05203 | felbamate | Radius_SOS | -0,01099 | 0,054926 | NA |
| 3 | abundance | X05203 | felbamate | Tibia_SOS | -0,10441 | 0,057806 | NA |
| 3 | abundance | X05203 | felbamate | Handgrip | -0,08607 | 0,06337 | NA |
| 3 | abundance | X05204 | Urothion | Tibia_length | -0,08126 | 0,050622 | NA |
| 3 | abundance | X05204 | Urothion | Radius_length | -0,0058 | 0,069711 | NA |
| 3 | abundance | X05204 | Urothion | Radius_SOS | -0,02128 | 0,049466 | NA |
| 3 | abundance | X05204 | Urothion | Tibia_SOS | -0,0409 | 0,052754 | NA |
| 3 | abundance | X05204 | Urothion | Handgrip | 0,015653 | 0,061032 | NA |
| 3 | abundance | X05214 | [7-Hydroxy | Tibia_length | 0,102512 | 0,051164 | NA |
| 3 | abundance | X05214 | [7-Hydroxy | Radius_length | 0,071701 | 0,070242 | NA |
| 3 | abundance | X05214 | [7-Hydroxy | Radius_SOS | -0,02571 | 0,049608 | NA |
| 3 | abundance | X05214 | [7-Hydroxy | Tibia_SOS | 0,080337 | 0,053368 | NA |
| 3 | abundance | X05214 | [7-Hydroxy | Handgrip | -4,80E-04 | 0,061125 | NA |
| 3 | abundance | X05225 | 4-(METHYL | Tibia_length | -0,02798 | 0,056395 | NA |
| 3 | abundance | X05225 | 4-(METHYL | Radius_length | -0,00264 | 0,074873 | NA |
| 3 | abundance | X05225 | 4-(METHYL | Radius_SOS | 0,026211 | 0,053425 | NA |
| 3 | abundance | X05225 | 4-(METHYL | Tibia_SOS | -0,04596 | 0,057027 | NA |
| 3 | abundance | X05225 | 4-(METHYL | Handgrip | 0,126249 | 0,059711 | NA |
| 3 | abundance | X05234 | 3,4-dihydro | Tibia_length | -0,16498 | 0,052238 | NA |
| 3 | abundance | X05234 | 3,4-dihydro | Radius_length | -0,15619 | 0,071045 | NA |
| 3 | abundance | X05234 | 3,4-dihydro | Radius_SOS | -0,02739 | 0,0503 | NA |
| 3 | abundance | X05234 | 3,4-dihydro | Tibia_SOS | -0,11151 | 0,053953 | NA |
| 3 | abundance | X05234 | 3,4-dihydro | Handgrip | -0,04684 | 0,060964 | NA |
| 3 | abundance | X05237 | imazameth | Tibia_length | -0,11546 | 0,057313 | NA |
| 3 | abundance | X05237 | imazameth | Radius_length | -0,17681 | 0,074551 | NA |
| 3 | abundance | X05237 | imazameth | Radius_SOS | -0,04992 | 0,054975 | NA |
| 3 | abundance | X05237 | imazameth | Tibia_SOS | -0,13088 | 0,057909 | NA |
| 3 | abundance | X05237 | imazameth | Handgrip | -0,01214 | 0,062174 | NA |
| 3 | abundance | X05240 | Dimeric mo | Tibia_length | -0,05444 | 0,053759 | NA |
| 3 | abundance | X05240 | Dimeric mo | Radius_length | -0,09609 | 0,072319 | NA |
| 3 | abundance | X05240 | Dimeric mo | Radius_SOS | -0,01349 | 0,051248 | NA |
| 3 | abundance | X05240 | Dimeric mo | Tibia_SOS | 0,037548 | 0,054502 | NA |
| 3 | abundance | X05240 | Dimeric mo | Handgrip | -0,08016 | 0,061798 | NA |
| 3 | abundance | X05243 | N-(4-Hydro | Tibia_length | -0,16276 | 0,052882 | NA |
| 3 | abundance | X05243 | N-(4-Hydro | Radius_length | -0,13841 | 0,071725 | NA |
| 3 | abundance | X05243 | N-(4-Hydro | Radius_SOS | -0,05274 | 0,050765 | NA |

| | | | | | | | |
|---|---|---|---|---|---|---|---|
| 3 | abundance | X05243 | N-(4-Hydrc | Tibia_SOS | -0,10624 | 0,053647 | NA |
| 3 | abundance | X05243 | N-(4-Hydrc | Handgrip | -0,06683 | 0,061255 | NA |
| 3 | abundance | X05275 | H-DL-MET- | Tibia_length | 0,052213 | 0,056406 | NA |
| 3 | abundance | X05275 | H-DL-MET- | Radius_length | 0,169372 | 0,074346 | NA |
| 3 | abundance | X05275 | H-DL-MET- | Radius_SOS | 0,006341 | 0,053629 | NA |
| 3 | abundance | X05275 | H-DL-MET- | Tibia_SOS | 0,03579 | 0,057555 | NA |
| 3 | abundance | X05275 | H-DL-MET- | Handgrip | 0,085015 | 0,063503 | NA |
| 3 | abundance | X05295 | 3-(4,7-Dim | Tibia_length | -0,08569 | 0,055151 | NA |
| 3 | abundance | X05295 | 3-(4,7-Dim | Radius_length | -0,18008 | 0,07231 | NA |
| 3 | abundance | X05295 | 3-(4,7-Dim | Radius_SOS | -0,05981 | 0,052201 | NA |
| 3 | abundance | X05295 | 3-(4,7-Dim | Tibia_SOS | -0,1151 | 0,055593 | NA |
| 3 | abundance | X05295 | 3-(4,7-Dim | Handgrip | -0,04404 | 0,061922 | NA |
| 3 | abundance | X05306 | 2-methoxy | Tibia_length | 0,062459 | 0,049707 | NA |
| 3 | abundance | X05306 | 2-methoxy | Radius_length | 0,183707 | 0,066655 | NA |
| 3 | abundance | X05306 | 2-methoxy | Radius_SOS | -0,00957 | 0,049035 | NA |
| 3 | abundance | X05306 | 2-methoxy | Tibia_SOS | 0,037985 | 0,052472 | NA |
| 3 | abundance | X05306 | 2-methoxy | Handgrip | 0,043055 | 0,060442 | NA |
| 3 | abundance | X05357 | NPYR | Tibia_length | 0,096384 | 0,05461 | NA |
| 3 | abundance | X05357 | NPYR | Radius_length | -0,06291 | 0,073614 | NA |
| 3 | abundance | X05357 | NPYR | Radius_SOS | -0,03305 | 0,052176 | NA |
| 3 | abundance | X05357 | NPYR | Tibia_SOS | 0,102885 | 0,055284 | NA |
| 3 | abundance | X05357 | NPYR | Handgrip | -0,02004 | 0,061511 | NA |
| 3 | abundance | X05391 | 2-Acetami | Tibia_length | -0,0144 | 0,056771 | NA |
| 3 | abundance | X05391 | 2-Acetami | Radius_length | 0,106734 | 0,074093 | NA |
| 3 | abundance | X05391 | 2-Acetami | Radius_SOS | -0,02713 | 0,054029 | NA |
| 3 | abundance | X05391 | 2-Acetami | Tibia_SOS | 0,005619 | 0,057719 | NA |
| 3 | abundance | X05391 | 2-Acetami | Handgrip | 0,021026 | 0,061555 | NA |
| 3 | abundance | X05398 | DIBOA | Tibia_length | -0,11838 | 0,05495 | NA |
| 3 | abundance | X05398 | DIBOA | Radius_length | -0,09145 | 0,073934 | NA |
| 3 | abundance | X05398 | DIBOA | Radius_SOS | -0,02162 | 0,05239 | NA |
| 3 | abundance | X05398 | DIBOA | Tibia_SOS | -0,07671 | 0,055385 | NA |
| 3 | abundance | X05398 | DIBOA | Handgrip | -0,17129 | 0,062051 | NA |
| 3 | abundance | X05411 | 3-(Sulfoox | Tibia_length | -0,07929 | 0,057482 | NA |
| 3 | abundance | X05411 | 3-(Sulfoox | Radius_length | 0,021068 | 0,075079 | NA |
| 3 | abundance | X05411 | 3-(Sulfoox | Radius_SOS | 0,103198 | 0,054752 | NA |
| 3 | abundance | X05411 | 3-(Sulfoox | Tibia_SOS | 0,121111 | 0,058274 | NA |
| 3 | abundance | X05411 | 3-(Sulfoox | Handgrip | -0,05177 | 0,062641 | NA |
| 3 | abundance | X05415 | Redul | Tibia_length | -0,00806 | 0,051629 | NA |
| 3 | abundance | X05415 | Redul | Radius_length | 0,164021 | 0,069676 | NA |
| 3 | abundance | X05415 | Redul | Radius_SOS | 0,056063 | 0,049817 | NA |
| 3 | abundance | X05415 | Redul | Tibia_SOS | 0,054274 | 0,05289 | NA |
| 3 | abundance | X05415 | Redul | Handgrip | 0,019186 | 0,060487 | NA |
| 3 | abundance | X05417 | (S)-2-hydra | Tibia_length | -0,03656 | 0,053385 | NA |
| 3 | abundance | X05417 | (S)-2-hydra | Radius_length | -0,03855 | 0,072634 | NA |
| 3 | abundance | X05417 | (S)-2-hydra | Radius_SOS | 0,036638 | 0,051071 | NA |
| 3 | abundance | X05417 | (S)-2-hydra | Tibia_SOS | -0,05801 | 0,054105 | NA |
| 3 | abundance | X05417 | (S)-2-hydra | Handgrip | -0,02425 | 0,061804 | NA |
| 3 | abundance | X05421 | S-Allylcyst | Tibia_length | -0,04054 | 0,054644 | NA |

| | | | | | | | |
|---|---|---|---|---|---|---|---|
| 3 | abundance | X05421 | S-Allylcyst | Radius_length | 0,057137 | 0,073071 | NA |
| 3 | abundance | X05421 | S-Allylcyst | Radius_SOS | 0,00523 | 0,051851 | NA |
| 3 | abundance | X05421 | S-Allylcyst | Tibia_SOS | -0,02598 | 0,055223 | NA |
| 3 | abundance | X05421 | S-Allylcyst | Handgrip | -0,05044 | 0,062544 | NA |
| 3 | abundance | X05459 | Scopoletin | Tibia_length | 0,114587 | 0,05277 | NA |
| 3 | abundance | X05459 | Scopoletin | Radius_length | 0,154163 | 0,071194 | NA |
| 3 | abundance | X05459 | Scopoletin | Radius_SOS | 0,063131 | 0,050474 | NA |
| 3 | abundance | X05459 | Scopoletin | Tibia_SOS | 0,131143 | 0,053192 | NA |
| 3 | abundance | X05459 | Scopoletin | Handgrip | 0,146135 | 0,060541 | NA |
| 3 | abundance | X05467 | Azulfidine | Tibia_length | -0,12265 | 0,053254 | NA |
| 3 | abundance | X05467 | Azulfidine | Radius_length | -0,13575 | 0,071824 | NA |
| 3 | abundance | X05467 | Azulfidine | Radius_SOS | -0,08405 | 0,050985 | NA |
| 3 | abundance | X05467 | Azulfidine | Tibia_SOS | 0,066422 | 0,053782 | NA |
| 3 | abundance | X05467 | Azulfidine | Handgrip | 0,029479 | 0,061555 | NA |
| 3 | abundance | X05478 | S(8)-aminc | Tibia_length | 0,042555 | 0,052151 | NA |
| 3 | abundance | X05478 | S(8)-aminc | Radius_length | 0,095765 | 0,071392 | NA |
| 3 | abundance | X05478 | S(8)-aminc | Radius_SOS | 0,029436 | 0,05006 | NA |
| 3 | abundance | X05478 | S(8)-aminc | Tibia_SOS | 0,045698 | 0,053382 | NA |
| 3 | abundance | X05478 | S(8)-aminc | Handgrip | 0,019181 | 0,061769 | NA |
| 3 | abundance | X05499 | GLY-MET | Tibia_length | -0,09102 | 0,055609 | NA |
| 3 | abundance | X05499 | GLY-MET | Radius_length | 0,026006 | 0,074386 | NA |
| 3 | abundance | X05499 | GLY-MET | Radius_SOS | 0,017759 | 0,052726 | NA |
| 3 | abundance | X05499 | GLY-MET | Tibia_SOS | 0,012694 | 0,056688 | NA |
| 3 | abundance | X05499 | GLY-MET | Handgrip | 0,056327 | 0,062839 | NA |
| 3 | abundance | X05510 | 3-Benzyl-6 | Tibia_length | 0,072653 | 0,050843 | NA |
| 3 | abundance | X05510 | 3-Benzyl-6 | Radius_length | 0,060784 | 0,069626 | NA |
| 3 | abundance | X05510 | 3-Benzyl-6 | Radius_SOS | 0,053232 | 0,049429 | NA |
| 3 | abundance | X05510 | 3-Benzyl-6 | Tibia_SOS | 0,05811 | 0,052943 | NA |
| 3 | abundance | X05510 | 3-Benzyl-6 | Handgrip | -0,0241 | 0,061016 | NA |
| 3 | abundance | X05514 | (3aS,5S,6F | Tibia_length | -0,02366 | 0,050601 | NA |
| 3 | abundance | X05514 | (3aS,5S,6F | Radius_length | -0,11134 | 0,068795 | NA |
| 3 | abundance | X05514 | (3aS,5S,6F | Radius_SOS | 0,03792 | 0,049217 | NA |
| 3 | abundance | X05514 | (3aS,5S,6F | Tibia_SOS | -0,02015 | 0,052861 | NA |
| 3 | abundance | X05514 | (3aS,5S,6F | Handgrip | -0,04604 | 0,060575 | NA |
| 3 | abundance | X05551 | Trolox | Tibia_length | -0,07299 | 0,052029 | NA |
| 3 | abundance | X05551 | Trolox | Radius_length | -0,07521 | 0,070909 | NA |
| 3 | abundance | X05551 | Trolox | Radius_SOS | 0,039625 | 0,050087 | NA |
| 3 | abundance | X05551 | Trolox | Tibia_SOS | 0,002699 | 0,053213 | NA |
| 3 | abundance | X05551 | Trolox | Handgrip | -0,15207 | 0,059681 | NA |
| 3 | abundance | X05561 | 1ÃŽÂ²-Hyd | Tibia_length | -0,03043 | 0,051651 | NA |
| 3 | abundance | X05561 | 1ÃŽÂ²-Hyd | Radius_length | -0,10629 | 0,070389 | NA |
| 3 | abundance | X05561 | 1ÃŽÂ²-Hyd | Radius_SOS | 0,036089 | 0,04977 | NA |
| 3 | abundance | X05561 | 1ÃŽÂ²-Hyd | Tibia_SOS | -0,05592 | 0,053318 | NA |
| 3 | abundance | X05561 | 1ÃŽÂ²-Hyd | Handgrip | -0,02626 | 0,061065 | NA |
| 3 | abundance | X05580 | Xanthureni | Tibia_length | -0,06045 | 0,055531 | NA |
| 3 | abundance | X05580 | Xanthureni | Radius_length | -0,03276 | 0,073372 | NA |
| 3 | abundance | X05580 | Xanthureni | Radius_SOS | -0,09718 | 0,052327 | NA |
| 3 | abundance | X05580 | Xanthureni | Tibia_SOS | 0,018266 | 0,055381 | NA |

| | | | | | | | |
|---|---|---|---|---|---|---|---|
| 3 | abundance | X05580 | Xanthureni | Handgrip | 0,022769 | 0,062273 | NA |
| 3 | abundance | X05581 | N-[(10Z)-7 | Tibia_length | -0,04964 | 0,049312 | NA |
| 3 | abundance | X05581 | N-[(10Z)-7 | Radius_length | -0,03355 | 0,066552 | NA |
| 3 | abundance | X05581 | N-[(10Z)-7 | Radius_SOS | -0,02866 | 0,049018 | NA |
| 3 | abundance | X05581 | N-[(10Z)-7 | Tibia_SOS | -0,05756 | 0,052404 | NA |
| 3 | abundance | X05581 | N-[(10Z)-7 | Handgrip | -0,03494 | 0,060349 | NA |
| 1 | abundance | X05584 | Serotonin | Tibia_length | 0,044454 | 0,050355 | NA |
| 1 | abundance | X05584 | Serotonin | Radius_length | -0,0383 | 0,068748 | NA |
| 1 | abundance | X05584 | Serotonin | Radius_SOS | -0,05696 | 0,049095 | NA |
| 1 | abundance | X05584 | Serotonin | Tibia_SOS | -0,00914 | 0,05283 | NA |
| 1 | abundance | X05584 | Serotonin | Handgrip | -0,09428 | 0,060127 | NA |
| 3 | abundance | X05656 | mesifurane | Tibia_length | -0,04134 | 0,053123 | NA |
| 3 | abundance | X05656 | mesifurane | Radius_length | -0,15511 | 0,071041 | NA |
| 3 | abundance | X05656 | mesifurane | Radius_SOS | -0,0311 | 0,050775 | NA |
| 3 | abundance | X05656 | mesifurane | Tibia_SOS | -0,09787 | 0,053624 | NA |
| 3 | abundance | X05656 | mesifurane | Handgrip | -0,03982 | 0,061055 | NA |
| 3 | abundance | X05709 | 2,3-Dihydr | Tibia_length | -0,06542 | 0,052438 | NA |
| 3 | abundance | X05709 | 2,3-Dihydr | Radius_length | -0,14541 | 0,070847 | NA |
| 3 | abundance | X05709 | 2,3-Dihydr | Radius_SOS | -0,00733 | 0,050207 | NA |
| 3 | abundance | X05709 | 2,3-Dihydr | Tibia_SOS | -0,1288 | 0,053069 | NA |
| 3 | abundance | X05709 | 2,3-Dihydr | Handgrip | -0,22035 | 0,059634 | NA |
| 3 | abundance | X05726 | methocarb | Tibia_length | -0,11235 | 0,051362 | NA |
| 3 | abundance | X05726 | methocarb | Radius_length | -0,14246 | 0,069734 | NA |
| 3 | abundance | X05726 | methocarb | Radius_SOS | -0,03188 | 0,049649 | NA |
| 3 | abundance | X05726 | methocarb | Tibia_SOS | -0,09827 | 0,052726 | NA |
| 3 | abundance | X05726 | methocarb | Handgrip | -0,1043 | 0,060434 | NA |
| 3 | abundance | X05734 | N-[(2S)-2-I | Tibia_length | -0,08726 | 0,054691 | NA |
| 3 | abundance | X05734 | N-[(2S)-2-I | Radius_length | 0,008213 | 0,073651 | NA |
| 3 | abundance | X05734 | N-[(2S)-2-I | Radius_SOS | 0,018569 | 0,052373 | NA |
| 3 | abundance | X05734 | N-[(2S)-2-I | Tibia_SOS | -0,00141 | 0,05506 | NA |
| 3 | abundance | X05734 | N-[(2S)-2-I | Handgrip | -0,05445 | 0,062102 | NA |
| 3 | abundance | X05741 | g-Aminobu | Tibia_length | 0,048084 | 0,0526 | NA |
| 3 | abundance | X05741 | g-Aminobu | Radius_length | 0,018081 | 0,071537 | NA |
| 3 | abundance | X05741 | g-Aminobu | Radius_SOS | -0,02317 | 0,050448 | NA |
| 3 | abundance | X05741 | g-Aminobu | Tibia_SOS | -0,04953 | 0,054357 | NA |
| 3 | abundance | X05741 | g-Aminobu | Handgrip | 0,029539 | 0,061861 | NA |
| 3 | abundance | X05758 | quinol sulf | Tibia_length | -0,0533 | 0,054778 | NA |
| 3 | abundance | X05758 | quinol sulf | Radius_length | -0,11793 | 0,073093 | NA |
| 3 | abundance | X05758 | quinol sulf | Radius_SOS | -0,16278 | 0,052645 | NA |
| 3 | abundance | X05758 | quinol sulf | Tibia_SOS | -0,02935 | 0,055855 | NA |
| 3 | abundance | X05758 | quinol sulf | Handgrip | -0,05739 | 0,062447 | NA |
| 3 | abundance | X05786 | Nitrendipir | Tibia_length | 0,027985 | 0,051604 | NA |
| 3 | abundance | X05786 | Nitrendipir | Radius_length | 0,032212 | 0,070944 | NA |
| 3 | abundance | X05786 | Nitrendipir | Radius_SOS | 0,039469 | 0,0497 | NA |
| 3 | abundance | X05786 | Nitrendipir | Tibia_SOS | 0,053342 | 0,052817 | NA |
| 3 | abundance | X05786 | Nitrendipir | Handgrip | -0,06186 | 0,06062 | NA |
| 3 | abundance | X05812 | (2S)-3-(1H | Tibia_length | -0,01635 | 0,057204 | NA |
| 3 | abundance | X05812 | (2S)-3-(1H | Radius_length | 0,078454 | 0,0746 | NA |

| | | | | | | | |
|---|---|---|---|---|---|---|---|
| 3 | abundance | X05812 | (2S)-3-(1H | Radius_SOS | 0,100206 | 0,054617 | NA |
| 3 | abundance | X05812 | (2S)-3-(1H | Tibia_SOS | -0,02215 | 0,059488 | NA |
| 3 | abundance | X05812 | (2S)-3-(1H | Handgrip | 0,027733 | 0,063339 | NA |
| 3 | abundance | X05857 | N-D-Gluco | Tibia_length | -0,22338 | 0,053523 | NA |
| 3 | abundance | X05857 | N-D-Gluco | Radius_length | -0,07587 | 0,073167 | NA |
| 3 | abundance | X05857 | N-D-Gluco | Radius_SOS | -0,03423 | 0,05208 | NA |
| 3 | abundance | X05857 | N-D-Gluco | Tibia_SOS | -0,07561 | 0,055829 | NA |
| 3 | abundance | X05857 | N-D-Gluco | Handgrip | 0,049048 | 0,062607 | NA |
| 3 | abundance | X05859 | 3,7,12,17-T | Tibia_length | -0,03676 | 0,05046 | NA |
| 3 | abundance | X05859 | 3,7,12,17-T | Radius_length | -0,11659 | 0,06864 | NA |
| 3 | abundance | X05859 | 3,7,12,17-T | Radius_SOS | 0,032444 | 0,049165 | NA |
| 3 | abundance | X05859 | 3,7,12,17-T | Tibia_SOS | -0,00357 | 0,052659 | NA |
| 3 | abundance | X05859 | 3,7,12,17-T | Handgrip | 0,019612 | 0,060624 | NA |
| 3 | abundance | X05878 | 1,2-dihydro | Tibia_length | 0,00703 | 0,053187 | NA |
| 3 | abundance | X05878 | 1,2-dihydro | Radius_length | 0,044215 | 0,072121 | NA |
| 3 | abundance | X05878 | 1,2-dihydro | Radius_SOS | -0,08547 | 0,050928 | NA |
| 3 | abundance | X05878 | 1,2-dihydro | Tibia_SOS | 0,026126 | 0,054789 | NA |
| 3 | abundance | X05878 | 1,2-dihydro | Handgrip | -0,02583 | 0,061384 | NA |
| 3 | abundance | X05892 | (5Z)-2-Ami | Tibia_length | -0,04695 | 0,053029 | NA |
| 3 | abundance | X05892 | (5Z)-2-Ami | Radius_length | -0,0865 | 0,071731 | NA |
| 3 | abundance | X05892 | (5Z)-2-Ami | Radius_SOS | -0,02436 | 0,050693 | NA |
| 3 | abundance | X05892 | (5Z)-2-Ami | Tibia_SOS | -0,07948 | 0,054461 | NA |
| 3 | abundance | X05892 | (5Z)-2-Ami | Handgrip | -0,00816 | 0,062068 | NA |
| 3 | abundance | X05919 | 2-(2-Amino | Tibia_length | -0,04415 | 0,054083 | NA |
| 3 | abundance | X05919 | 2-(2-Amino | Radius_length | -0,16457 | 0,071678 | NA |
| 3 | abundance | X05919 | 2-(2-Amino | Radius_SOS | -0,02935 | 0,051348 | NA |
| 3 | abundance | X05919 | 2-(2-Amino | Tibia_SOS | -0,07899 | 0,055449 | NA |
| 3 | abundance | X05919 | 2-(2-Amino | Handgrip | -0,0436 | 0,061478 | NA |
| 3 | abundance | X05968 | Tetraacety | Tibia_length | -0,10168 | 0,054087 | NA |
| 3 | abundance | X05968 | Tetraacety | Radius_length | -0,21319 | 0,072369 | NA |
| 3 | abundance | X05968 | Tetraacety | Radius_SOS | -0,08344 | 0,0513 | NA |
| 3 | abundance | X05968 | Tetraacety | Tibia_SOS | -0,10764 | 0,054086 | NA |
| 3 | abundance | X05968 | Tetraacety | Handgrip | -0,05923 | 0,062103 | NA |
| 3 | abundance | X05969 | 4-(METHYL | Tibia_length | -0,17285 | 0,055857 | NA |
| 3 | abundance | X05969 | 4-(METHYL | Radius_length | -0,20319 | 0,073371 | NA |
| 3 | abundance | X05969 | 4-(METHYL | Radius_SOS | -0,01159 | 0,053434 | NA |
| 3 | abundance | X05969 | 4-(METHYL | Tibia_SOS | 0,009908 | 0,057084 | NA |
| 3 | abundance | X05969 | 4-(METHYL | Handgrip | -0,06322 | 0,063519 | NA |
| 3 | abundance | X05971 | L-gamma-( | Tibia_length | 0,022567 | 0,050688 | NA |
| 3 | abundance | X05971 | L-gamma-( | Radius_length | -0,05788 | 0,069518 | NA |
| 3 | abundance | X05971 | L-gamma-( | Radius_SOS | 0,093733 | 0,049079 | NA |
| 3 | abundance | X05971 | L-gamma-( | Tibia_SOS | -0,00345 | 0,052879 | NA |
| 3 | abundance | X05971 | L-gamma-( | Handgrip | 0,024284 | 0,06122 | NA |
| 3 | abundance | X05982 | (19R,25S)- | Tibia_length | 0,044066 | 0,054753 | NA |
| 3 | abundance | X05982 | (19R,25S)- | Radius_length | 0,185906 | 0,072199 | NA |
| 3 | abundance | X05982 | (19R,25S)- | Radius_SOS | -0,02122 | 0,051923 | NA |
| 3 | abundance | X05982 | (19R,25S)- | Tibia_SOS | 0,037 | 0,054568 | NA |
| 3 | abundance | X05982 | (19R,25S)- | Handgrip | -0,00513 | 0,062475 | NA |

| | | | | | | | |
|---|---|---|---|---|---|---|---|
| 2b | abundance | X05984 | 1-(4-Methy | Tibia_length | 0,0936 | 0,050629 | NA |
| 2b | abundance | X05984 | 1-(4-Methy | Radius_length | 0,090154 | 0,069391 | NA |
| 2b | abundance | X05984 | 1-(4-Methy | Radius_SOS | 0,02293 | 0,049395 | NA |
| 2b | abundance | X05984 | 1-(4-Methy | Tibia_SOS | 0,087611 | 0,052624 | NA |
| 2b | abundance | X05984 | 1-(4-Methy | Handgrip | 0,033179 | 0,060404 | NA |
| 3 | abundance | X05988 | Selsun | Tibia_length | -0,01441 | 0,053163 | NA |
| 3 | abundance | X05988 | Selsun | Radius_length | 0,055911 | 0,072278 | NA |
| 3 | abundance | X05988 | Selsun | Radius_SOS | -0,06651 | 0,050901 | NA |
| 3 | abundance | X05988 | Selsun | Tibia_SOS | 0,070994 | 0,054239 | NA |
| 3 | abundance | X05988 | Selsun | Handgrip | 0,016128 | 0,060938 | NA |
| 3 | abundance | X05995 | Diacetin | Tibia_length | -0,06771 | 0,054764 | NA |
| 3 | abundance | X05995 | Diacetin | Radius_length | 0,008377 | 0,073672 | NA |
| 3 | abundance | X05995 | Diacetin | Radius_SOS | 0,030281 | 0,052036 | NA |
| 3 | abundance | X05995 | Diacetin | Tibia_SOS | 0,008525 | 0,056664 | NA |
| 3 | abundance | X05995 | Diacetin | Handgrip | 0,06998 | 0,060536 | NA |
| 3 | abundance | X05997 | FB950000( | Tibia_length | 0,005926 | 0,054597 | NA |
| 3 | abundance | X05997 | FB950000( | Radius_length | -0,05752 | 0,073519 | NA |
| 3 | abundance | X05997 | FB950000( | Radius_SOS | 0,024438 | 0,051985 | NA |
| 3 | abundance | X05997 | FB950000( | Tibia_SOS | 0,03422 | 0,056314 | NA |
| 3 | abundance | X05997 | FB950000( | Handgrip | -0,03613 | 0,060154 | NA |
| 3 | abundance | X06011 | Coprine | Tibia_length | -0,07289 | 0,056247 | NA |
| 3 | abundance | X06011 | Coprine | Radius_length | -0,12803 | 0,07359 | NA |
| 3 | abundance | X06011 | Coprine | Radius_SOS | -0,0348 | 0,053584 | NA |
| 3 | abundance | X06011 | Coprine | Tibia_SOS | -0,05068 | 0,056386 | NA |
| 3 | abundance | X06011 | Coprine | Handgrip | -0,01839 | 0,061924 | NA |
| 3 | abundance | X06016 | N,N-Dimet | Tibia_length | 0,029741 | 0,055103 | NA |
| 3 | abundance | X06016 | N,N-Dimet | Radius_length | 0,059434 | 0,073666 | NA |
| 3 | abundance | X06016 | N,N-Dimet | Radius_SOS | 0,161791 | 0,051565 | NA |
| 3 | abundance | X06016 | N,N-Dimet | Tibia_SOS | -0,02781 | 0,055191 | NA |
| 3 | abundance | X06016 | N,N-Dimet | Handgrip | 0,064673 | 0,062073 | NA |
| 3 | abundance | X06034 | N-[(2S)-2-I | Tibia_length | 0,111917 | 0,052607 | NA |
| 3 | abundance | X06034 | N-[(2S)-2-I | Radius_length | 0,042594 | 0,071641 | NA |
| 3 | abundance | X06034 | N-[(2S)-2-I | Radius_SOS | -0,02349 | 0,050648 | NA |
| 3 | abundance | X06034 | N-[(2S)-2-I | Tibia_SOS | 0,18852 | 0,053311 | NA |
| 3 | abundance | X06034 | N-[(2S)-2-I | Handgrip | 0,010699 | 0,061141 | NA |
| 3 | abundance | X06039 | gamma-Gl | Tibia_length | -0,01647 | 0,053378 | NA |
| 3 | abundance | X06039 | gamma-Gl | Radius_length | 0,189705 | 0,070965 | NA |
| 3 | abundance | X06039 | gamma-Gl | Radius_SOS | 0,0897 | 0,050849 | NA |
| 3 | abundance | X06039 | gamma-Gl | Tibia_SOS | 0,161061 | 0,053984 | NA |
| 3 | abundance | X06039 | gamma-Gl | Handgrip | 0,095695 | 0,062868 | NA |
| 3 | abundance | X06041 | (DL)-3-O-M | Tibia_length | -0,10502 | 0,052675 | NA |
| 3 | abundance | X06041 | (DL)-3-O-M | Radius_length | -0,10188 | 0,071675 | NA |
| 3 | abundance | X06041 | (DL)-3-O-M | Radius_SOS | -0,05796 | 0,050476 | NA |
| 3 | abundance | X06041 | (DL)-3-O-M | Tibia_SOS | -0,11443 | 0,054396 | NA |
| 3 | abundance | X06041 | (DL)-3-O-M | Handgrip | -0,13361 | 0,061644 | NA |
| 3 | abundance | X06043 | vinyl sulfid | Tibia_length | 0,090124 | 0,051078 | NA |
| 3 | abundance | X06043 | vinyl sulfid | Radius_length | 0,167208 | 0,068944 | NA |
| 3 | abundance | X06043 | vinyl sulfid | Radius_SOS | 0,02503 | 0,049449 | NA |

| | | | | | | | |
|---|---|---|---|---|---|---|---|
| 3 | abundance | X06043 | vinyl sulfid | Tibia_SOS | 0,132958 | 0,052524 | NA |
| 3 | abundance | X06043 | vinyl sulfid | Handgrip | 0,105814 | 0,060213 | NA |
| 3 | abundance | X06046 | Valylvaline | Tibia_length | 0,078074 | 0,052892 | NA |
| 3 | abundance | X06046 | Valylvaline | Radius_length | 0,191752 | 0,070619 | NA |
| 3 | abundance | X06046 | Valylvaline | Radius_SOS | 0,05588 | 0,050393 | NA |
| 3 | abundance | X06046 | Valylvaline | Tibia_SOS | 0,14771 | 0,053614 | NA |
| 3 | abundance | X06046 | Valylvaline | Handgrip | 0,090151 | 0,061721 | NA |
| 3 | abundance | X06062 | 1-(2,3-Dihy | Tibia_length | -0,05173 | 0,052052 | NA |
| 3 | abundance | X06062 | 1-(2,3-Dihy | Radius_length | -0,10325 | 0,070673 | NA |
| 3 | abundance | X06062 | 1-(2,3-Dihy | Radius_SOS | -0,07852 | 0,0497 | NA |
| 3 | abundance | X06062 | 1-(2,3-Dihy | Tibia_SOS | 0,093025 | 0,052933 | NA |
| 3 | abundance | X06062 | 1-(2,3-Dihy | Handgrip | -0,11887 | 0,060219 | NA |
| 1 | abundance | X06079 | Glycylleuci | Tibia_length | -0,07355 | 0,054723 | NA |
| 1 | abundance | X06079 | Glycylleuci | Radius_length | -0,02101 | 0,074062 | NA |
| 1 | abundance | X06079 | Glycylleuci | Radius_SOS | -0,01957 | 0,052214 | NA |
| 1 | abundance | X06079 | Glycylleuci | Tibia_SOS | -0,02076 | 0,056536 | NA |
| 1 | abundance | X06079 | Glycylleuci | Handgrip | 0,014957 | 0,061489 | NA |
| 3 | abundance | X06082 | O-heptano | Tibia_length | -0,04609 | 0,055592 | NA |
| 3 | abundance | X06082 | O-heptano | Radius_length | -0,10724 | 0,073722 | NA |
| 3 | abundance | X06082 | O-heptano | Radius_SOS | -0,05777 | 0,052667 | NA |
| 3 | abundance | X06082 | O-heptano | Tibia_SOS | -0,12541 | 0,05739 | NA |
| 3 | abundance | X06082 | O-heptano | Handgrip | -0,18729 | 0,061113 | NA |
| 3 | abundance | X06085 | Tetraacety | Tibia_length | -0,10727 | 0,051204 | NA |
| 3 | abundance | X06085 | Tetraacety | Radius_length | -0,09219 | 0,069936 | NA |
| 3 | abundance | X06085 | Tetraacety | Radius_SOS | -0,02889 | 0,049542 | NA |
| 3 | abundance | X06085 | Tetraacety | Tibia_SOS | -0,10632 | 0,052846 | NA |
| 3 | abundance | X06085 | Tetraacety | Handgrip | -0,12067 | 0,060354 | NA |
| 3 | abundance | X06089 | riboprine | Tibia_length | -0,06428 | 0,055545 | NA |
| 3 | abundance | X06089 | riboprine | Radius_length | 0,005182 | 0,074244 | NA |
| 3 | abundance | X06089 | riboprine | Radius_SOS | -0,05389 | 0,053116 | NA |
| 3 | abundance | X06089 | riboprine | Tibia_SOS | -0,01535 | 0,056275 | NA |
| 3 | abundance | X06089 | riboprine | Handgrip | 0,111784 | 0,062566 | NA |
| 3 | abundance | X06100 | Midodrine | Tibia_length | -0,18109 | 0,052353 | NA |
| 3 | abundance | X06100 | Midodrine | Radius_length | -0,10907 | 0,071532 | NA |
| 3 | abundance | X06100 | Midodrine | Radius_SOS | -0,07342 | 0,050503 | NA |
| 3 | abundance | X06100 | Midodrine | Tibia_SOS | -0,04055 | 0,054098 | NA |
| 3 | abundance | X06100 | Midodrine | Handgrip | 0,063902 | 0,061533 | NA |
| 3 | abundance | X06107 | Hydroxycai | Tibia_length | 0,068185 | 0,049585 | NA |
| 3 | abundance | X06107 | Hydroxycai | Radius_length | 0,097444 | 0,067284 | NA |
| 3 | abundance | X06107 | Hydroxycai | Radius_SOS | 0,012584 | 0,049026 | NA |
| 3 | abundance | X06107 | Hydroxycai | Tibia_SOS | 0,069971 | 0,052382 | NA |
| 3 | abundance | X06107 | Hydroxycai | Handgrip | 0,080614 | 0,060191 | NA |
| 3 | abundance | X06112 | Val-Ser | Tibia_length | 0,071109 | 0,049405 | NA |
| 3 | abundance | X06112 | Val-Ser | Radius_length | 0,074063 | 0,067011 | NA |
| 3 | abundance | X06112 | Val-Ser | Radius_SOS | -0,05864 | 0,048945 | NA |
| 3 | abundance | X06112 | Val-Ser | Tibia_SOS | -0,03476 | 0,052615 | NA |
| 3 | abundance | X06112 | Val-Ser | Handgrip | -0,11045 | 0,06001 | NA |
| 3 | abundance | X06124 | MFCD099! | Tibia_length | -0,07456 | 0,055394 | NA |

| | | | | | | | | |
|---|---|---|---|---|---|---|---|---|
| | 3 | abundance | X06124 | MFCD0995 | Radius_length | 0,009873 | 0,073736 | NA |
| | 3 | abundance | X06124 | MFCD0995 | Radius_SOS | 0,004475 | 0,052676 | NA |
| | 3 | abundance | X06124 | MFCD0995 | Tibia_SOS | 0,011462 | 0,055978 | NA |
| | 3 | abundance | X06124 | MFCD0995 | Handgrip | -0,08015 | 0,062837 | NA |
| | 3 | abundance | X06127 | 2-Methoxy- | Tibia_length | -0,09205 | 0,052677 | NA |
| | 3 | abundance | X06127 | 2-Methoxy- | Radius_length | -0,00739 | 0,071836 | NA |
| | 3 | abundance | X06127 | 2-Methoxy- | Radius_SOS | -0,03529 | 0,050591 | NA |
| | 3 | abundance | X06127 | 2-Methoxy- | Tibia_SOS | 0,028723 | 0,054076 | NA |
| | 3 | abundance | X06127 | 2-Methoxy- | Handgrip | 0,003804 | 0,0612 | NA |
| 2a | | abundance | X06143 | 2-Methylbu | Tibia_length | -0,00549 | 0,056193 | NA |
| 2a | | abundance | X06143 | 2-Methylbu | Radius_length | 0,068655 | 0,07411 | NA |
| 2a | | abundance | X06143 | 2-Methylbu | Radius_SOS | 0,006015 | 0,05344 | NA |
| 2a | | abundance | X06143 | 2-Methylbu | Tibia_SOS | 0,005753 | 0,056855 | NA |
| 2a | | abundance | X06143 | 2-Methylbu | Handgrip | -0,00905 | 0,062658 | NA |
| | 3 | abundance | X06146 | S-Methyl-1 | Tibia_length | 0,038654 | 0,052256 | NA |
| | 3 | abundance | X06146 | S-Methyl-1 | Radius_length | -0,00899 | 0,071276 | NA |
| | 3 | abundance | X06146 | S-Methyl-1 | Radius_SOS | 0,008988 | 0,050235 | NA |
| | 3 | abundance | X06146 | S-Methyl-1 | Tibia_SOS | 0,056029 | 0,053634 | NA |
| | 3 | abundance | X06146 | S-Methyl-1 | Handgrip | -0,14453 | 0,059999 | NA |
| | 3 | abundance | X06149 | Yangonin | Tibia_length | -0,02286 | 0,052602 | NA |
| | 3 | abundance | X06149 | Yangonin | Radius_length | 0,04252 | 0,071791 | NA |
| | 3 | abundance | X06149 | Yangonin | Radius_SOS | 0,006035 | 0,050496 | NA |
| | 3 | abundance | X06149 | Yangonin | Tibia_SOS | 0,014138 | 0,053721 | NA |
| | 3 | abundance | X06149 | Yangonin | Handgrip | -0,03491 | 0,062215 | NA |
| | 3 | abundance | X06150 | Zalcitabine | Tibia_length | 0,012754 | 0,052309 | NA |
| | 3 | abundance | X06150 | Zalcitabine | Radius_length | -5,97E-04 | 0,071603 | NA |
| | 3 | abundance | X06150 | Zalcitabine | Radius_SOS | 0,012733 | 0,050238 | NA |
| | 3 | abundance | X06150 | Zalcitabine | Tibia_SOS | 0,097833 | 0,05358 | NA |
| | 3 | abundance | X06150 | Zalcitabine | Handgrip | 0,03959 | 0,061783 | NA |
| | 3 | abundance | X06152 | alpha-keto | Tibia_length | -0,05837 | 0,055568 | NA |
| | 3 | abundance | X06152 | alpha-keto | Radius_length | 0,021281 | 0,074414 | NA |
| | 3 | abundance | X06152 | alpha-keto | Radius_SOS | 0,062542 | 0,052552 | NA |
| | 3 | abundance | X06152 | alpha-keto | Tibia_SOS | 0,057128 | 0,057517 | NA |
| | 3 | abundance | X06152 | alpha-keto | Handgrip | 0,067046 | 0,062086 | NA |
| 2a | | abundance | X06167 | N-Acetylas | Tibia_length | -0,07438 | 0,054151 | NA |
| 2a | | abundance | X06167 | N-Acetylas | Radius_length | 0,015486 | 0,07353 | NA |
| 2a | | abundance | X06167 | N-Acetylas | Radius_SOS | -0,05619 | 0,051826 | NA |
| 2a | | abundance | X06167 | N-Acetylas | Tibia_SOS | 0,021957 | 0,056566 | NA |
| 2a | | abundance | X06167 | N-Acetylas | Handgrip | -0,02495 | 0,061848 | NA |
| | 3 | abundance | X06168 | leu-gln | Tibia_length | -0,04108 | 0,053138 | NA |
| | 3 | abundance | X06168 | leu-gln | Radius_length | 0,031587 | 0,072296 | NA |
| | 3 | abundance | X06168 | leu-gln | Radius_SOS | 0,002424 | 0,050823 | NA |
| | 3 | abundance | X06168 | leu-gln | Tibia_SOS | -0,02657 | 0,054585 | NA |
| | 3 | abundance | X06168 | leu-gln | Handgrip | 0,001045 | 0,061112 | NA |
| | 3 | abundance | X06183 | Nisinic aci | Tibia_length | 0,014739 | 0,049657 | NA |
| | 3 | abundance | X06183 | Nisinic aci | Radius_length | 0,082715 | 0,066952 | NA |
| | 3 | abundance | X06183 | Nisinic aci | Radius_SOS | 0,029689 | 0,049007 | NA |
| | 3 | abundance | X06183 | Nisinic aci | Tibia_SOS | 0,025946 | 0,052483 | NA |

| | | | | | | | | |
|---|---|---|---|---|---|---|---|---|
| | 3 | abundance | X06183 | Nisinic aci | Handgrip | 0,041522 | 0,060616 | NA |
| | 3 | abundance | X06189 | delta-Guar | Tibia_length | -0,06835 | 0,054262 | NA |
| | 3 | abundance | X06189 | delta-Guar | Radius_length | -0,0291 | 0,07283 | NA |
| | 3 | abundance | X06189 | delta-Guar | Radius_SOS | 0,010987 | 0,051822 | NA |
| | 3 | abundance | X06189 | delta-Guar | Tibia_SOS | 0,006866 | 0,055908 | NA |
| | 3 | abundance | X06189 | delta-Guar | Handgrip | -0,02197 | 0,062821 | NA |
| | 3 | abundance | X06197 | 4-(Nitrosoa | Tibia_length | -0,00241 | 0,05692 | NA |
| | 3 | abundance | X06197 | 4-(Nitrosoa | Radius_length | 0,064446 | 0,07499 | NA |
| | 3 | abundance | X06197 | 4-(Nitrosoa | Radius_SOS | 0,054649 | 0,054029 | NA |
| | 3 | abundance | X06197 | 4-(Nitrosoa | Tibia_SOS | -0,12386 | 0,056175 | NA |
| | 3 | abundance | X06197 | 4-(Nitrosoa | Handgrip | 0,06178 | 0,061815 | NA |
| | 3 | abundance | X06220 | Tetrahydro | Tibia_length | -0,08561 | 0,053618 | NA |
| | 3 | abundance | X06220 | Tetrahydro | Radius_length | -0,09851 | 0,072373 | NA |
| | 3 | abundance | X06220 | Tetrahydro | Radius_SOS | 0,014926 | 0,051222 | NA |
| | 3 | abundance | X06220 | Tetrahydro | Tibia_SOS | -0,11757 | 0,054559 | NA |
| | 3 | abundance | X06220 | Tetrahydro | Handgrip | -0,02403 | 0,061508 | NA |
| | 3 | abundance | X06222 | Leucylaspa | Tibia_length | -0,08619 | 0,05391 | NA |
| | 3 | abundance | X06222 | Leucylaspa | Radius_length | -0,01682 | 0,073067 | NA |
| | 3 | abundance | X06222 | Leucylaspa | Radius_SOS | 0,009799 | 0,051495 | NA |
| | 3 | abundance | X06222 | Leucylaspa | Tibia_SOS | -0,02822 | 0,055069 | NA |
| | 3 | abundance | X06222 | Leucylaspa | Handgrip | 0,054702 | 0,061129 | NA |
| | 3 | abundance | X06227 | lys-tyr | Tibia_length | -0,13701 | 0,054863 | NA |
| | 3 | abundance | X06227 | lys-tyr | Radius_length | -0,11013 | 0,07343 | NA |
| | 3 | abundance | X06227 | lys-tyr | Radius_SOS | -0,04874 | 0,052234 | NA |
| | 3 | abundance | X06227 | lys-tyr | Tibia_SOS | -0,0691 | 0,055167 | NA |
| | 3 | abundance | X06227 | lys-tyr | Handgrip | -0,05089 | 0,060394 | NA |
| | 3 | abundance | X06230 | Glu-Gly | Tibia_length | -0,10444 | 0,053131 | NA |
| | 3 | abundance | X06230 | Glu-Gly | Radius_length | -0,01436 | 0,072573 | NA |
| | 3 | abundance | X06230 | Glu-Gly | Radius_SOS | -0,06942 | 0,051276 | NA |
| | 3 | abundance | X06230 | Glu-Gly | Tibia_SOS | -0,05161 | 0,055899 | NA |
| | 3 | abundance | X06230 | Glu-Gly | Handgrip | -0,00399 | 0,062985 | NA |
| | 3 | abundance | X06240 | His-pro | Tibia_length | 0,00527 | 0,051744 | NA |
| | 3 | abundance | X06240 | His-pro | Radius_length | 0,014607 | 0,07088 | NA |
| | 3 | abundance | X06240 | His-pro | Radius_SOS | -0,00381 | 0,049859 | NA |
| | 3 | abundance | X06240 | His-pro | Tibia_SOS | 0,060949 | 0,053832 | NA |
| | 3 | abundance | X06240 | His-pro | Handgrip | 0,061535 | 0,060792 | NA |
| 2b | | abundance | X06250 | [3-({3-[(Cy | Tibia_length | -0,13802 | 0,054848 | NA |
| 2b | | abundance | X06250 | [3-({3-[(Cy | Radius_length | -0,32189 | 0,069814 | NA |
| 2b | | abundance | X06250 | [3-({3-[(Cy | Radius_SOS | -0,14715 | 0,051927 | NA |
| 2b | | abundance | X06250 | [3-({3-[(Cy | Tibia_SOS | -0,18609 | 0,054707 | NA |
| 2b | | abundance | X06250 | [3-({3-[(Cy | Handgrip | -0,20076 | 0,061585 | NA |
| | 3 | abundance | X06253 | N-(4-Amin | Tibia_length | -0,06751 | 0,054503 | NA |
| | 3 | abundance | X06253 | N-(4-Amin | Radius_length | -0,11649 | 0,072572 | NA |
| | 3 | abundance | X06253 | N-(4-Amin | Radius_SOS | -0,06298 | 0,051875 | NA |
| | 3 | abundance | X06253 | N-(4-Amin | Tibia_SOS | -0,02607 | 0,056193 | NA |
| | 3 | abundance | X06253 | N-(4-Amin | Handgrip | -0,07082 | 0,062275 | NA |
| | 3 | abundance | X06254 | D-Alanyl-D | Tibia_length | -0,05277 | 0,055473 | NA |
| | 3 | abundance | X06254 | D-Alanyl-D | Radius_length | -0,06977 | 0,074101 | NA |

| | | | | | | | | |
|---|---|---|---|---|---|---|---|---|
| | 3 | abundance | X06254 | D-Alanyl-D | Radius_SOS | -0,05579 | 0,053028 | NA |
| | 3 | abundance | X06254 | D-Alanyl-D | Tibia_SOS | -0,01378 | 0,056824 | NA |
| | 3 | abundance | X06254 | D-Alanyl-D | Handgrip | 0,035791 | 0,063468 | NA |
| | 3 | abundance | X06259 | (S)-?-glyce | Tibia_length | 0,019149 | 0,055032 | NA |
| | 3 | abundance | X06259 | (S)-?-glyce | Radius_length | 0,026943 | 0,073814 | NA |
| | 3 | abundance | X06259 | (S)-?-glyce | Radius_SOS | 0,043583 | 0,052337 | NA |
| | 3 | abundance | X06259 | (S)-?-glyce | Tibia_SOS | 0,052428 | 0,055705 | NA |
| | 3 | abundance | X06259 | (S)-?-glyce | Handgrip | 0,061106 | 0,06206 | NA |
| | 3 | abundance | X06268 | mesifurane | Tibia_length | 0,011122 | 0,053762 | NA |
| | 3 | abundance | X06268 | mesifurane | Radius_length | -0,01353 | 0,07281 | NA |
| | 3 | abundance | X06268 | mesifurane | Radius_SOS | 0,007958 | 0,051354 | NA |
| | 3 | abundance | X06268 | mesifurane | Tibia_SOS | -0,01818 | 0,055859 | NA |
| | 3 | abundance | X06268 | mesifurane | Handgrip | 0,034033 | 0,063405 | NA |
| 2b | | abundance | X06270 | 9-Methylur | Tibia_length | -0,02417 | 0,054684 | NA |
| 2b | | abundance | X06270 | 9-Methylur | Radius_length | -0,1069 | 0,073237 | NA |
| 2b | | abundance | X06270 | 9-Methylur | Radius_SOS | -0,03399 | 0,051879 | NA |
| 2b | | abundance | X06270 | 9-Methylur | Tibia_SOS | -0,09685 | 0,055496 | NA |
| 2b | | abundance | X06270 | 9-Methylur | Handgrip | -0,13781 | 0,061911 | NA |
| | 3 | abundance | X06276 | asn-pro | Tibia_length | -0,04759 | 0,056281 | NA |
| | 3 | abundance | X06276 | asn-pro | Radius_length | -0,06384 | 0,074691 | NA |
| | 3 | abundance | X06276 | asn-pro | Radius_SOS | -0,06485 | 0,053375 | NA |
| | 3 | abundance | X06276 | asn-pro | Tibia_SOS | -0,11471 | 0,054685 | NA |
| | 3 | abundance | X06276 | asn-pro | Handgrip | -0,14474 | 0,061113 | NA |
| | 3 | abundance | X06278 | 3-Hydroxy- | Tibia_length | 0,005425 | 0,049926 | NA |
| | 3 | abundance | X06278 | 3-Hydroxy- | Radius_length | 0,059115 | 0,067989 | NA |
| | 3 | abundance | X06278 | 3-Hydroxy- | Radius_SOS | -0,0423 | 0,049033 | NA |
| | 3 | abundance | X06278 | 3-Hydroxy- | Tibia_SOS | 0,002828 | 0,05259 | NA |
| | 3 | abundance | X06278 | 3-Hydroxy- | Handgrip | 0,011361 | 0,060323 | NA |
| | 3 | abundance | X06289 | Nicotinatel | Tibia_length | 0,034658 | 0,053987 | NA |
| | 3 | abundance | X06289 | Nicotinatel | Radius_length | -0,00477 | 0,073421 | NA |
| | 3 | abundance | X06289 | Nicotinatel | Radius_SOS | 0,052567 | 0,051573 | NA |
| | 3 | abundance | X06289 | Nicotinatel | Tibia_SOS | -0,01355 | 0,055268 | NA |
| | 3 | abundance | X06289 | Nicotinatel | Handgrip | 0,076065 | 0,063531 | NA |
| 2b | | abundance | X06291 | Corticoste | Tibia_length | 0,019422 | 0,050012 | NA |
| 2b | | abundance | X06291 | Corticoste | Radius_length | 0,062755 | 0,068044 | NA |
| 2b | | abundance | X06291 | Corticoste | Radius_SOS | -0,02968 | 0,049042 | NA |
| 2b | | abundance | X06291 | Corticoste | Tibia_SOS | 0,050399 | 0,052422 | NA |
| 2b | | abundance | X06291 | Corticoste | Handgrip | 0,062533 | 0,0601 | NA |
| | 3 | abundance | X06292 | mesifurane | Tibia_length | 0,005974 | 0,057711 | NA |
| | 3 | abundance | X06292 | mesifurane | Radius_length | 0,020427 | 0,075574 | NA |
| | 3 | abundance | X06292 | mesifurane | Radius_SOS | -0,02763 | 0,054885 | NA |
| | 3 | abundance | X06292 | mesifurane | Tibia_SOS | -0,02393 | 0,059902 | NA |
| | 3 | abundance | X06292 | mesifurane | Handgrip | 0,026939 | 0,060171 | NA |
| | 3 | abundance | X06293 | (4R)-4-{[3- | Tibia_length | -0,09859 | 0,053438 | NA |
| | 3 | abundance | X06293 | (4R)-4-{[3- | Radius_length | -0,11437 | 0,071998 | NA |
| | 3 | abundance | X06293 | (4R)-4-{[3- | Radius_SOS | -0,02794 | 0,051071 | NA |
| | 3 | abundance | X06293 | (4R)-4-{[3- | Tibia_SOS | -0,07517 | 0,053638 | NA |
| | 3 | abundance | X06293 | (4R)-4-{[3- | Handgrip | -0,02393 | 0,061153 | NA |

| | | | | | | | |
|---|---|---|---|---|---|---|---|
| 2b | abundance | X06313 | Isoquinolir | Tibia_length | 0,044929 | 0,054168 | NA |
| 2b | abundance | X06313 | Isoquinolir | Radius_length | -0,08393 | 0,072726 | NA |
| 2b | abundance | X06313 | Isoquinolir | Radius_SOS | -0,13437 | 0,051183 | NA |
| 2b | abundance | X06313 | Isoquinolir | Tibia_SOS | 0,057328 | 0,054729 | NA |
| 2b | abundance | X06313 | Isoquinolir | Handgrip | -0,01431 | 0,06249 | NA |
| 2b | abundance | X06318 | Propamoca | Tibia_length | 0,060736 | 0,053625 | NA |
| 2b | abundance | X06318 | Propamoca | Radius_length | 0,072961 | 0,072505 | NA |
| 2b | abundance | X06318 | Propamoca | Radius_SOS | 0,023715 | 0,051363 | NA |
| 2b | abundance | X06318 | Propamoca | Tibia_SOS | -0,0522 | 0,054825 | NA |
| 2b | abundance | X06318 | Propamoca | Handgrip | 0,019658 | 0,06241 | NA |
| 3 | abundance | X06320 | 7alpha-Hy | Tibia_length | 0,040547 | 0,051857 | NA |
| 3 | abundance | X06320 | 7alpha-Hy | Radius_length | -0,00957 | 0,070781 | NA |
| 3 | abundance | X06320 | 7alpha-Hy | Radius_SOS | 0,020558 | 0,049928 | NA |
| 3 | abundance | X06320 | 7alpha-Hy | Tibia_SOS | -0,07736 | 0,053788 | NA |
| 3 | abundance | X06320 | 7alpha-Hy | Handgrip | -0,02516 | 0,061657 | NA |
| 3 | abundance | X06329 | Leu-Leu | Tibia_length | -0,07605 | 0,055098 | NA |
| 3 | abundance | X06329 | Leu-Leu | Radius_length | -0,05216 | 0,073348 | NA |
| 3 | abundance | X06329 | Leu-Leu | Radius_SOS | -0,02566 | 0,052379 | NA |
| 3 | abundance | X06329 | Leu-Leu | Tibia_SOS | -0,05281 | 0,056859 | NA |
| 3 | abundance | X06329 | Leu-Leu | Handgrip | -0,04359 | 0,062131 | NA |
| 3 | abundance | X06334 | Bis-D-fruct | Tibia_length | -0,0262 | 0,052361 | NA |
| 3 | abundance | X06334 | Bis-D-fruct | Radius_length | -0,01936 | 0,071783 | NA |
| 3 | abundance | X06334 | Bis-D-fruct | Radius_SOS | -0,03009 | 0,050236 | NA |
| 3 | abundance | X06334 | Bis-D-fruct | Tibia_SOS | 0,072936 | 0,053501 | NA |
| 3 | abundance | X06334 | Bis-D-fruct | Handgrip | -0,02648 | 0,062344 | NA |
| 3 | abundance | X06337 | Zalcitabine | Tibia_length | -0,05029 | 0,053576 | NA |
| 3 | abundance | X06337 | Zalcitabine | Radius_length | -0,01442 | 0,072772 | NA |
| 3 | abundance | X06337 | Zalcitabine | Radius_SOS | 0,006677 | 0,051127 | NA |
| 3 | abundance | X06337 | Zalcitabine | Tibia_SOS | -0,08015 | 0,053419 | NA |
| 3 | abundance | X06337 | Zalcitabine | Handgrip | -0,08022 | 0,062384 | NA |
| 2b | abundance | X06354 | trans-Zeati | Tibia_length | -0,11675 | 0,056285 | NA |
| 2b | abundance | X06354 | trans-Zeati | Radius_length | -0,12675 | 0,073327 | NA |
| 2b | abundance | X06354 | trans-Zeati | Radius_SOS | -0,04904 | 0,052882 | NA |
| 2b | abundance | X06354 | trans-Zeati | Tibia_SOS | -0,01896 | 0,056078 | NA |
| 2b | abundance | X06354 | trans-Zeati | Handgrip | 0,00192 | 0,062505 | NA |
| 3 | abundance | X06361 | Oxprenolol | Tibia_length | 0,191295 | 0,049028 | NA |
| 3 | abundance | X06361 | Oxprenolol | Radius_length | 0,200661 | 0,066894 | NA |
| 3 | abundance | X06361 | Oxprenolol | Radius_SOS | 0,065064 | 0,048957 | NA |
| 3 | abundance | X06361 | Oxprenolol | Tibia_SOS | 0,117601 | 0,052263 | NA |
| 3 | abundance | X06361 | Oxprenolol | Handgrip | 0,107707 | 0,059958 | NA |
| 3 | abundance | X06368 | TDP-2 | Tibia_length | 0,053941 | 0,050864 | NA |
| 3 | abundance | X06368 | TDP-2 | Radius_length | 0,05697 | 0,069687 | NA |
| 3 | abundance | X06368 | TDP-2 | Radius_SOS | 0,019009 | 0,04945 | NA |
| 3 | abundance | X06368 | TDP-2 | Tibia_SOS | 0,092788 | 0,052489 | NA |
| 3 | abundance | X06368 | TDP-2 | Handgrip | -0,03304 | 0,060332 | NA |
| 3 | abundance | X06371 | L-gamma-( | Tibia_length | -0,09136 | 0,055851 | NA |
| 3 | abundance | X06371 | L-gamma-( | Radius_length | -0,06769 | 0,074259 | NA |
| 3 | abundance | X06371 | L-gamma-( | Radius_SOS | -0,00788 | 0,053082 | NA |

| | | | | | | | | |
|---|---|---|---|---|---|---|---|---|
| | 3 | abundance | X06371 | L-gamma-( | Tibia_SOS | 0,007457 | 0,057575 | NA |
| | 3 | abundance | X06371 | L-gamma-( | Handgrip | -0,02258 | 0,062355 | NA |
| 2b | | abundance | X06372 | N,N-dimetl | Tibia_length | 0,072157 | 0,051159 | NA |
| 2b | | abundance | X06372 | N,N-dimetl | Radius_length | 0,109093 | 0,069705 | NA |
| 2b | | abundance | X06372 | N,N-dimetl | Radius_SOS | -0,01136 | 0,049548 | NA |
| 2b | | abundance | X06372 | N,N-dimetl | Tibia_SOS | 0,127747 | 0,052424 | NA |
| 2b | | abundance | X06372 | N,N-dimetl | Handgrip | 0,080396 | 0,060963 | NA |
| | 3 | abundance | X06381 | MFCD1869 | Tibia_length | -0,18008 | 0,04817 | NA |
| | 3 | abundance | X06381 | MFCD1869 | Radius_length | -0,16987 | 0,048259 | NA |
| | 3 | abundance | X06381 | MFCD1869 | Radius_SOS | -0,01823 | 0,049021 | NA |
| | 3 | abundance | X06381 | MFCD1869 | Tibia_SOS | -0,09552 | 0,052246 | NA |
| | 3 | abundance | X06381 | MFCD1869 | Handgrip | -0,08537 | 0,059973 | NA |
| | 1 | abundance | X06388 | Caffeic aci | Tibia_length | -0,13063 | 0,05366 | NA |
| | 1 | abundance | X06388 | Caffeic aci | Radius_length | -0,10431 | 0,072493 | NA |
| | 1 | abundance | X06388 | Caffeic aci | Radius_SOS | -0,11649 | 0,051073 | NA |
| | 1 | abundance | X06388 | Caffeic aci | Tibia_SOS | -0,1475 | 0,054394 | NA |
| | 1 | abundance | X06388 | Caffeic aci | Handgrip | 0,036944 | 0,061844 | NA |
| | 3 | abundance | X06400 | Phloionolic | Tibia_length | 0,049637 | 0,049977 | NA |
| | 3 | abundance | X06400 | Phloionolic | Radius_length | -0,01737 | 0,068068 | NA |
| | 3 | abundance | X06400 | Phloionolic | Radius_SOS | -0,03446 | 0,049047 | NA |
| | 3 | abundance | X06400 | Phloionolic | Tibia_SOS | -0,0578 | 0,052557 | NA |
| | 3 | abundance | X06400 | Phloionolic | Handgrip | -0,04204 | 0,06074 | NA |
| 2b | | abundance | X06401 | Isophthalic | Tibia_length | -0,15365 | 0,050098 | NA |
| 2b | | abundance | X06401 | Isophthalic | Radius_length | -0,12551 | 0,068797 | NA |
| 2b | | abundance | X06401 | Isophthalic | Radius_SOS | -0,01582 | 0,049248 | NA |
| 2b | | abundance | X06401 | Isophthalic | Tibia_SOS | -0,08404 | 0,052416 | NA |
| 2b | | abundance | X06401 | Isophthalic | Handgrip | -0,11225 | 0,059921 | NA |
| | 3 | abundance | X06404 | Flemichap | Tibia_length | 0,024902 | 0,053044 | NA |
| | 3 | abundance | X06404 | Flemichap | Radius_length | 0,035236 | 0,071741 | NA |
| | 3 | abundance | X06404 | Flemichap | Radius_SOS | 0,030225 | 0,050727 | NA |
| | 3 | abundance | X06404 | Flemichap | Tibia_SOS | 0,066691 | 0,05493 | NA |
| | 3 | abundance | X06404 | Flemichap | Handgrip | -0,04765 | 0,060866 | NA |
| | 3 | abundance | X06405 | Asparaginy | Tibia_length | -0,07442 | 0,052787 | NA |
| | 3 | abundance | X06405 | Asparaginy | Radius_length | -0,07572 | 0,071946 | NA |
| | 3 | abundance | X06405 | Asparaginy | Radius_SOS | -0,04239 | 0,050503 | NA |
| | 3 | abundance | X06405 | Asparaginy | Tibia_SOS | -0,07288 | 0,053132 | NA |
| | 3 | abundance | X06405 | Asparaginy | Handgrip | -0,07977 | 0,061049 | NA |
| 2b | | abundance | X06409 | Glycylproli | Tibia_length | 0,069406 | 0,052731 | NA |
| 2b | | abundance | X06409 | Glycylproli | Radius_length | 0,009603 | 0,071587 | NA |
| 2b | | abundance | X06409 | Glycylproli | Radius_SOS | -0,03709 | 0,050466 | NA |
| 2b | | abundance | X06409 | Glycylproli | Tibia_SOS | 0,10099 | 0,055239 | NA |
| 2b | | abundance | X06409 | Glycylproli | Handgrip | 0,07444 | 0,061937 | NA |
| | 3 | abundance | X06412 | Butenylcar | Tibia_length | 0,087623 | 0,056446 | NA |
| | 3 | abundance | X06412 | Butenylcar | Radius_length | 0,049298 | 0,074404 | NA |
| | 3 | abundance | X06412 | Butenylcar | Radius_SOS | -0,03367 | 0,053716 | NA |
| | 3 | abundance | X06412 | Butenylcar | Tibia_SOS | 0,06437 | 0,056819 | NA |
| | 3 | abundance | X06412 | Butenylcar | Handgrip | -0,07966 | 0,062488 | NA |
| | 3 | abundance | X06416 | KYNURAMI | Tibia_length | 0,118456 | 0,049635 | NA |

| | | | | | | | |
|---|---|---|---|---|---|---|---|
| 3 | abundance | X06416 | KYNURAMI | Radius_length | 0,097884 | 0,067721 | NA |
| 3 | abundance | X06416 | KYNURAMI | Radius_SOS | 0,022409 | 0,049064 | NA |
| 3 | abundance | X06416 | KYNURAMI | Tibia_SOS | 0,093638 | 0,052329 | NA |
| 3 | abundance | X06416 | KYNURAMI | Handgrip | 0,039492 | 0,060271 | NA |
| 3 | abundance | X06419 | Spermic ac | Tibia_length | -0,07428 | 0,054135 | NA |
| 3 | abundance | X06419 | Spermic ac | Radius_length | 0,067321 | 0,073066 | NA |
| 3 | abundance | X06419 | Spermic ac | Radius_SOS | -0,01975 | 0,051716 | NA |
| 3 | abundance | X06419 | Spermic ac | Tibia_SOS | -0,01119 | 0,056219 | NA |
| 3 | abundance | X06419 | Spermic ac | Handgrip | 0,019181 | 0,062404 | NA |
| 2b | abundance | X06433 | 4-(2,5-Diflu | Tibia_length | -0,07759 | 0,050959 | NA |
| 2b | abundance | X06433 | 4-(2,5-Diflu | Radius_length | -0,0638 | 0,070049 | NA |
| 2b | abundance | X06433 | 4-(2,5-Diflu | Radius_SOS | -0,00424 | 0,049496 | NA |
| 2b | abundance | X06433 | 4-(2,5-Diflu | Tibia_SOS | 0,082756 | 0,052709 | NA |
| 2b | abundance | X06433 | 4-(2,5-Diflu | Handgrip | 0,002398 | 0,061637 | NA |
| 3 | abundance | X06434 | butyrin | Tibia_length | 0,058961 | 0,051606 | NA |
| 3 | abundance | X06434 | butyrin | Radius_length | 0,058292 | 0,070468 | NA |
| 3 | abundance | X06434 | butyrin | Radius_SOS | 0,055945 | 0,049651 | NA |
| 3 | abundance | X06434 | butyrin | Tibia_SOS | 0,021276 | 0,053463 | NA |
| 3 | abundance | X06434 | butyrin | Handgrip | -0,01428 | 0,061079 | NA |
| 3 | abundance | X06437 | 2-Acetami | Tibia_length | 0,091956 | 0,055857 | NA |
| 3 | abundance | X06437 | 2-Acetami | Radius_length | 0,086354 | 0,074512 | NA |
| 3 | abundance | X06437 | 2-Acetami | Radius_SOS | -0,05022 | 0,053428 | NA |
| 3 | abundance | X06437 | 2-Acetami | Tibia_SOS | 0,001714 | 0,057231 | NA |
| 3 | abundance | X06437 | 2-Acetami | Handgrip | 0,084135 | 0,063306 | NA |
| 2b | abundance | X06448 | 3,8,9-trihy | Tibia_length | -0,02774 | 0,054296 | NA |
| 2b | abundance | X06448 | 3,8,9-trihy | Radius_length | 0,060451 | 0,073067 | NA |
| 2b | abundance | X06448 | 3,8,9-trihy | Radius_SOS | 0,062404 | 0,051755 | NA |
| 2b | abundance | X06448 | 3,8,9-trihy | Tibia_SOS | -0,02379 | 0,055143 | NA |
| 2b | abundance | X06448 | 3,8,9-trihy | Handgrip | -0,09117 | 0,061803 | NA |
| 3 | abundance | X06454 | 8-Amino-7- | Tibia_length | -0,1283 | 0,05255 | NA |
| 3 | abundance | X06454 | 8-Amino-7- | Radius_length | 0,010186 | 0,071921 | NA |
| 3 | abundance | X06454 | 8-Amino-7- | Radius_SOS | 0,013555 | 0,050669 | NA |
| 3 | abundance | X06454 | 8-Amino-7- | Tibia_SOS | 0,001984 | 0,054159 | NA |
| 3 | abundance | X06454 | 8-Amino-7- | Handgrip | -0,10008 | 0,061226 | NA |
| 3 | abundance | X06455 | 4-Hydroxyp | Tibia_length | 0,041174 | 0,054336 | NA |
| 3 | abundance | X06455 | 4-Hydroxyp | Radius_length | 0,077081 | 0,072998 | NA |
| 3 | abundance | X06455 | 4-Hydroxyp | Radius_SOS | 0,045603 | 0,05182 | NA |
| 3 | abundance | X06455 | 4-Hydroxyp | Tibia_SOS | -0,07963 | 0,057052 | NA |
| 3 | abundance | X06455 | 4-Hydroxyp | Handgrip | 0,013532 | 0,061714 | NA |
| 3 | abundance | X06462 | DNOP | Tibia_length | 0,036803 | 0,050273 | NA |
| 3 | abundance | X06462 | DNOP | Radius_length | 0,104732 | 0,06828 | NA |
| 3 | abundance | X06462 | DNOP | Radius_SOS | 0,058215 | 0,049047 | NA |
| 3 | abundance | X06462 | DNOP | Tibia_SOS | -0,04316 | 0,052729 | NA |
| 3 | abundance | X06462 | DNOP | Handgrip | -1,87E-04 | 0,060881 | NA |
| 3 | abundance | X06463 | 8-Amino-7- | Tibia_length | -0,1102 | 0,059354 | NA |
| 3 | abundance | X06463 | 8-Amino-7- | Radius_length | -0,11097 | 0,075516 | NA |
| 3 | abundance | X06463 | 8-Amino-7- | Radius_SOS | -0,05053 | 0,056734 | NA |
| 3 | abundance | X06463 | 8-Amino-7- | Tibia_SOS | -0,00459 | 0,052486 | NA |

| | | | | | | | |
|---|---|---|---|---|---|---|---|
| 3 | abundance | X06463 | 8-Amino-7- | Handgrip | 0,01064 | 0,06019 | NA |
| 3 | abundance | X06472 | N(alpha)-B | Tibia_length | 0,095061 | 0,053743 | NA |
| 3 | abundance | X06472 | N(alpha)-B | Radius_length | 0,097071 | 0,072773 | NA |
| 3 | abundance | X06472 | N(alpha)-B | Radius_SOS | 0,095307 | 0,051346 | NA |
| 3 | abundance | X06472 | N(alpha)-B | Tibia_SOS | 0,088685 | 0,055244 | NA |
| 3 | abundance | X06472 | N(alpha)-B | Handgrip | 0,054292 | 0,061755 | NA |
| 3 | abundance | X06473 | MFCD0002 | Tibia_length | 0,112181 | 0,055165 | NA |
| 3 | abundance | X06473 | MFCD0002 | Radius_length | 0,125161 | 0,073269 | NA |
| 3 | abundance | X06473 | MFCD0002 | Radius_SOS | 0,080069 | 0,052333 | NA |
| 3 | abundance | X06473 | MFCD0002 | Tibia_SOS | 0,090441 | 0,055366 | NA |
| 3 | abundance | X06473 | MFCD0002 | Handgrip | 0,01004 | 0,062499 | NA |
| 3 | abundance | X06477 | APM | Tibia_length | -0,0017 | 0,056695 | NA |
| 3 | abundance | X06477 | APM | Radius_length | 0,228024 | 0,07275 | NA |
| 3 | abundance | X06477 | APM | Radius_SOS | 0,021026 | 0,053717 | NA |
| 3 | abundance | X06477 | APM | Tibia_SOS | 0,025153 | 0,055685 | NA |
| 3 | abundance | X06477 | APM | Handgrip | 0,003888 | 0,062059 | NA |
| 3 | abundance | X06478 | Caffeic aci | Tibia_length | -0,04645 | 0,056036 | NA |
| 3 | abundance | X06478 | Caffeic aci | Radius_length | -0,123 | 0,073477 | NA |
| 3 | abundance | X06478 | Caffeic aci | Radius_SOS | -0,06974 | 0,053168 | NA |
| 3 | abundance | X06478 | Caffeic aci | Tibia_SOS | -0,05996 | 0,056361 | NA |
| 3 | abundance | X06478 | Caffeic aci | Handgrip | -0,09454 | 0,062827 | NA |
| 3 | abundance | X06482 | glu-pro | Tibia_length | -0,06765 | 0,053292 | NA |
| 3 | abundance | X06482 | glu-pro | Radius_length | -0,13311 | 0,071572 | NA |
| 3 | abundance | X06482 | glu-pro | Radius_SOS | -0,08093 | 0,050744 | NA |
| 3 | abundance | X06482 | glu-pro | Tibia_SOS | -0,06234 | 0,054218 | NA |
| 3 | abundance | X06482 | glu-pro | Handgrip | -0,0848 | 0,061366 | NA |
| 3 | abundance | X06486 | Leu-Val | Tibia_length | -0,01324 | 0,053182 | NA |
| 3 | abundance | X06486 | Leu-Val | Radius_length | -0,07705 | 0,072042 | NA |
| 3 | abundance | X06486 | Leu-Val | Radius_SOS | 0,021845 | 0,050773 | NA |
| 3 | abundance | X06486 | Leu-Val | Tibia_SOS | 0,041334 | 0,055247 | NA |
| 3 | abundance | X06486 | Leu-Val | Handgrip | -0,01612 | 0,06248 | NA |
| 3 | abundance | X06495 | 2-Acetamid | Tibia_length | -0,0193 | 0,056134 | NA |
| 3 | abundance | X06495 | 2-Acetamid | Radius_length | 1,89E-05 | 0,074378 | NA |
| 3 | abundance | X06495 | 2-Acetamid | Radius_SOS | 0,035743 | 0,053637 | NA |
| 3 | abundance | X06495 | 2-Acetamid | Tibia_SOS | -0,04111 | 0,057275 | NA |
| 3 | abundance | X06495 | 2-Acetamid | Handgrip | -0,0788 | 0,062652 | NA |
| 3 | abundance | X06502 | MFCD2836 | Tibia_length | -0,06256 | 0,051566 | NA |
| 3 | abundance | X06502 | MFCD2836 | Radius_length | -0,07658 | 0,070584 | NA |
| 3 | abundance | X06502 | MFCD2836 | Radius_SOS | -0,00964 | 0,049629 | NA |
| 3 | abundance | X06502 | MFCD2836 | Tibia_SOS | -0,01539 | 0,053262 | NA |
| 3 | abundance | X06502 | MFCD2836 | Handgrip | 0,026111 | 0,061344 | NA |
| 3 | abundance | X06506 | L-Homocys | Tibia_length | 0,055354 | 0,054099 | NA |
| 3 | abundance | X06506 | L-Homocys | Radius_length | 0,072092 | 0,072551 | NA |
| 3 | abundance | X06506 | L-Homocys | Radius_SOS | 0,063445 | 0,051474 | NA |
| 3 | abundance | X06506 | L-Homocys | Tibia_SOS | 1,10E-04 | 0,055116 | NA |
| 3 | abundance | X06506 | L-Homocys | Handgrip | 0,012217 | 0,063125 | NA |
| 3 | abundance | X06507 | Gly-Lys | Tibia_length | -0,00309 | 0,055844 | NA |
| 3 | abundance | X06507 | Gly-Lys | Radius_length | -0,02209 | 0,074097 | NA |

| | | | | | | | | |
|---|---|---|---|---|---|---|---|---|
| | 3 | abundance | X06507 | Gly-Lys | Radius_SOS | 0,069145 | 0,052917 | NA |
| | 3 | abundance | X06507 | Gly-Lys | Tibia_SOS | 0,04169 | 0,057081 | NA |
| | 3 | abundance | X06507 | Gly-Lys | Handgrip | 0,039007 | 0,06356 | NA |
| | 3 | abundance | X06508 | Asparaginy | Tibia_length | -0,08607 | 0,05509 | NA |
| | 3 | abundance | X06508 | Asparaginy | Radius_length | -0,12188 | 0,073385 | NA |
| | 3 | abundance | X06508 | Asparaginy | Radius_SOS | -0,03701 | 0,052438 | NA |
| | 3 | abundance | X06508 | Asparaginy | Tibia_SOS | -0,08406 | 0,054066 | NA |
| | 3 | abundance | X06508 | Asparaginy | Handgrip | -0,07456 | 0,061585 | NA |
| 2b | | abundance | X06510 | N-{4-[(2R,3 | Tibia_length | -0,04655 | 0,053173 | NA |
| 2b | | abundance | X06510 | N-{4-[(2R,3 | Radius_length | 0,027708 | 0,072668 | NA |
| 2b | | abundance | X06510 | N-{4-[(2R,3 | Radius_SOS | 0,003778 | 0,050859 | NA |
| 2b | | abundance | X06510 | N-{4-[(2R,3 | Tibia_SOS | 0,015784 | 0,054478 | NA |
| 2b | | abundance | X06510 | N-{4-[(2R,3 | Handgrip | 0,095043 | 0,061003 | NA |
| 2a | | abundance | X06529 | Methanesu | Tibia_length | -0,00534 | 0,055554 | NA |
| 2a | | abundance | X06529 | Methanesu | Radius_length | 0,070077 | 0,073366 | NA |
| 2a | | abundance | X06529 | Methanesu | Radius_SOS | 0,03024 | 0,052799 | NA |
| 2a | | abundance | X06529 | Methanesu | Tibia_SOS | 0,116933 | 0,055832 | NA |
| 2a | | abundance | X06529 | Methanesu | Handgrip | 0,055134 | 0,06173 | NA |
| 2b | | abundance | X06530 | 3-(1-hydro: | Tibia_length | -0,04252 | 0,051674 | NA |
| 2b | | abundance | X06530 | 3-(1-hydro: | Radius_length | -0,06248 | 0,070604 | NA |
| 2b | | abundance | X06530 | 3-(1-hydro: | Radius_SOS | -0,03073 | 0,049769 | NA |
| 2b | | abundance | X06530 | 3-(1-hydro: | Tibia_SOS | -0,0177 | 0,053124 | NA |
| 2b | | abundance | X06530 | 3-(1-hydro: | Handgrip | -0,0808 | 0,060237 | NA |
| | 3 | abundance | X06532 | Leupeptin | Tibia_length | -6,36E-04 | 0,052485 | NA |
| | 3 | abundance | X06532 | Leupeptin | Radius_length | 0,030462 | 0,071392 | NA |
| | 3 | abundance | X06532 | Leupeptin | Radius_SOS | -0,01795 | 0,050318 | NA |
| | 3 | abundance | X06532 | Leupeptin | Tibia_SOS | -0,02827 | 0,054194 | NA |
| | 3 | abundance | X06532 | Leupeptin | Handgrip | 0,041097 | 0,063223 | NA |
| 2b | | abundance | X06546 | 4-Acetami | Tibia_length | -0,07817 | 0,052361 | NA |
| 2b | | abundance | X06546 | 4-Acetami | Radius_length | 0,001242 | 0,071485 | NA |
| 2b | | abundance | X06546 | 4-Acetami | Radius_SOS | -0,00304 | 0,050334 | NA |
| 2b | | abundance | X06546 | 4-Acetami | Tibia_SOS | -0,00929 | 0,05352 | NA |
| 2b | | abundance | X06546 | 4-Acetami | Handgrip | 0,060077 | 0,061484 | NA |
| | 3 | abundance | X06549 | Panthenol | Tibia_length | 0,025992 | 0,056796 | NA |
| | 3 | abundance | X06549 | Panthenol | Radius_length | 0,058043 | 0,074702 | NA |
| | 3 | abundance | X06549 | Panthenol | Radius_SOS | -0,00359 | 0,053964 | NA |
| | 3 | abundance | X06549 | Panthenol | Tibia_SOS | -0,0039 | 0,056695 | NA |
| | 3 | abundance | X06549 | Panthenol | Handgrip | 0,01808 | 0,062824 | NA |
| | 3 | abundance | X06551 | Methyl 1-hy | Tibia_length | -0,17448 | 0,053912 | NA |
| | 3 | abundance | X06551 | Methyl 1-hy | Radius_length | -0,1325 | 0,072452 | NA |
| | 3 | abundance | X06551 | Methyl 1-hy | Radius_SOS | -0,09407 | 0,051672 | NA |
| | 3 | abundance | X06551 | Methyl 1-hy | Tibia_SOS | -0,14404 | 0,054813 | NA |
| | 3 | abundance | X06551 | Methyl 1-hy | Handgrip | -0,12509 | 0,061957 | NA |
| 2b | | abundance | X06560 | 2,6-Dimeth | Tibia_length | -0,13895 | 0,048762 | NA |
| 2b | | abundance | X06560 | 2,6-Dimeth | Radius_length | -0,19891 | 0,064282 | NA |
| 2b | | abundance | X06560 | 2,6-Dimeth | Radius_SOS | -0,04648 | 0,049121 | NA |
| 2b | | abundance | X06560 | 2,6-Dimeth | Tibia_SOS | -0,07618 | 0,052666 | NA |
| 2b | | abundance | X06560 | 2,6-Dimeth | Handgrip | -0,06395 | 0,060081 | NA |

| | | | | | | | |
|---|---|---|---|---|---|---|---|
| 3 | abundance | X06590 | 3-hydroxy-: | Tibia_length | -0,07176 | 0,056861 | NA |
| 3 | abundance | X06590 | 3-hydroxy-: | Radius_length | -0,09218 | 0,074156 | NA |
| 3 | abundance | X06590 | 3-hydroxy-: | Radius_SOS | -0,04382 | 0,054112 | NA |
| 3 | abundance | X06590 | 3-hydroxy-: | Tibia_SOS | 0,003062 | 0,057719 | NA |
| 3 | abundance | X06590 | 3-hydroxy-: | Handgrip | -0,10993 | 0,063298 | NA |
| 3 | abundance | X06600 | MFCD001 | Tibia_length | 0,034702 | 0,053978 | NA |
| 3 | abundance | X06600 | MFCD001 | Radius_length | -0,00478 | 0,073413 | NA |
| 3 | abundance | X06600 | MFCD001 | Radius_SOS | 0,052592 | 0,051567 | NA |
| 3 | abundance | X06600 | MFCD001 | Tibia_SOS | -0,01328 | 0,055262 | NA |
| 3 | abundance | X06600 | MFCD001 | Handgrip | 0,076002 | 0,063528 | NA |
| 3 | abundance | X06606 | Leu-Val | Tibia_length | 0,083745 | 0,054077 | NA |
| 3 | abundance | X06606 | Leu-Val | Radius_length | 0,037899 | 0,073488 | NA |
| 3 | abundance | X06606 | Leu-Val | Radius_SOS | 0,093234 | 0,051888 | NA |
| 3 | abundance | X06606 | Leu-Val | Tibia_SOS | 0,045525 | 0,055634 | NA |
| 3 | abundance | X06606 | Leu-Val | Handgrip | -0,10517 | 0,06315 | NA |
| 3 | abundance | X06612 | Spermic ac | Tibia_length | -0,11243 | 0,053635 | NA |
| 3 | abundance | X06612 | Spermic ac | Radius_length | 0,040578 | 0,072905 | NA |
| 3 | abundance | X06612 | Spermic ac | Radius_SOS | -0,00546 | 0,051334 | NA |
| 3 | abundance | X06612 | Spermic ac | Tibia_SOS | -0,00334 | 0,055054 | NA |
| 3 | abundance | X06612 | Spermic ac | Handgrip | 0,056369 | 0,060823 | NA |
| 3 | abundance | X06622 | Lys-Pro | Tibia_length | -0,01674 | 0,053387 | NA |
| 3 | abundance | X06622 | Lys-Pro | Radius_length | 0,013556 | 0,072486 | NA |
| 3 | abundance | X06622 | Lys-Pro | Radius_SOS | -0,00924 | 0,051031 | NA |
| 3 | abundance | X06622 | Lys-Pro | Tibia_SOS | 0,07917 | 0,054245 | NA |
| 3 | abundance | X06622 | Lys-Pro | Handgrip | 0,089221 | 0,060697 | NA |
| 3 | abundance | X06623 | quinol sulf | Tibia_length | -0,09121 | 0,054187 | NA |
| 3 | abundance | X06623 | quinol sulf | Radius_length | -0,09329 | 0,073039 | NA |
| 3 | abundance | X06623 | quinol sulf | Radius_SOS | 0,018626 | 0,051751 | NA |
| 3 | abundance | X06623 | quinol sulf | Tibia_SOS | -0,05295 | 0,05455 | NA |
| 3 | abundance | X06623 | quinol sulf | Handgrip | -0,03626 | 0,061874 | NA |
| 3 | abundance | X06626 | N-Butyryl-l | Tibia_length | 0,083905 | 0,052171 | NA |
| 3 | abundance | X06626 | N-Butyryl-l | Radius_length | 0,108349 | 0,070868 | NA |
| 3 | abundance | X06626 | N-Butyryl-l | Radius_SOS | 0,026337 | 0,050151 | NA |
| 3 | abundance | X06626 | N-Butyryl-l | Tibia_SOS | 0,117375 | 0,053354 | NA |
| 3 | abundance | X06626 | N-Butyryl-l | Handgrip | 0,041711 | 0,060858 | NA |
| 3 | abundance | X06630 | 4-(METHYL | Tibia_length | -0,02789 | 0,053959 | NA |
| 3 | abundance | X06630 | 4-(METHYL | Radius_length | 0,041762 | 0,072929 | NA |
| 3 | abundance | X06630 | 4-(METHYL | Radius_SOS | 0,043886 | 0,051434 | NA |
| 3 | abundance | X06630 | 4-(METHYL | Tibia_SOS | 0,091947 | 0,056172 | NA |
| 3 | abundance | X06630 | 4-(METHYL | Handgrip | 0,053172 | 0,063144 | NA |
| 2b | abundance | X06631 | 2-Isopropy | Tibia_length | -0,07738 | 0,054009 | NA |
| 2b | abundance | X06631 | 2-Isopropy | Radius_length | -0,09126 | 0,072521 | NA |
| 2b | abundance | X06631 | 2-Isopropy | Radius_SOS | -0,03017 | 0,051475 | NA |
| 2b | abundance | X06631 | 2-Isopropy | Tibia_SOS | 0,002804 | 0,055211 | NA |
| 2b | abundance | X06631 | 2-Isopropy | Handgrip | -0,0028 | 0,0623 | NA |
| 3 | abundance | X06634 | Glycitein | Tibia_length | -0,00774 | 0,058922 | NA |
| 3 | abundance | X06634 | Glycitein | Radius_length | 0,047477 | 0,075938 | NA |
| 3 | abundance | X06634 | Glycitein | Radius_SOS | -0,0787 | 0,056281 | NA |

| | | | | | | | |
|---|---|---|---|---|---|---|---|
| 3 | abundance | X06634 | Glycitein | Tibia_SOS | 0,038897 | 0,05946 | NA |
| 3 | abundance | X06634 | Glycitein | Handgrip | 0,103494 | 0,05987 | NA |
| 2a | abundance | X06642 | Homocitru | Tibia_length | 0,028947 | 0,049508 | NA |
| 2a | abundance | X06642 | Homocitru | Radius_length | 0,098153 | 0,066607 | NA |
| 2a | abundance | X06642 | Homocitru | Radius_SOS | -0,03886 | 0,049004 | NA |
| 2a | abundance | X06642 | Homocitru | Tibia_SOS | 0,045314 | 0,052433 | NA |
| 2a | abundance | X06642 | Homocitru | Handgrip | -0,00863 | 0,060382 | NA |
| 3 | abundance | X06646 | (S)-3-sulfo | Tibia_length | 0,037842 | 0,058133 | NA |
| 3 | abundance | X06646 | (S)-3-sulfo | Radius_length | -0,11991 | 0,074671 | NA |
| 3 | abundance | X06646 | (S)-3-sulfo | Radius_SOS | -0,05032 | 0,055447 | NA |
| 3 | abundance | X06646 | (S)-3-sulfo | Tibia_SOS | 0,011386 | 0,052483 | NA |
| 3 | abundance | X06646 | (S)-3-sulfo | Handgrip | -0,02312 | 0,060177 | NA |
| 3 | abundance | X06655 | Hydroxyca | Tibia_length | 0,024806 | 0,052269 | NA |
| 3 | abundance | X06655 | Hydroxyca | Radius_length | 0,01916 | 0,071731 | NA |
| 3 | abundance | X06655 | Hydroxyca | Radius_SOS | 0,060133 | 0,050345 | NA |
| 3 | abundance | X06655 | Hydroxyca | Tibia_SOS | -0,01027 | 0,054244 | NA |
| 3 | abundance | X06655 | Hydroxyca | Handgrip | -0,08892 | 0,061912 | NA |
| 3 | abundance | X06656 | Valylvaline | Tibia_length | -0,0317 | 0,055095 | NA |
| 3 | abundance | X06656 | Valylvaline | Radius_length | -0,02478 | 0,073437 | NA |
| 3 | abundance | X06656 | Valylvaline | Radius_SOS | 0,036989 | 0,052365 | NA |
| 3 | abundance | X06656 | Valylvaline | Tibia_SOS | -0,00816 | 0,056005 | NA |
| 3 | abundance | X06656 | Valylvaline | Handgrip | -0,05226 | 0,060111 | NA |
| 3 | abundance | X06663 | Leucyltyros | Tibia_length | -0,0709 | 0,051855 | NA |
| 3 | abundance | X06663 | Leucyltyros | Radius_length | 0,010166 | 0,07104 | NA |
| 3 | abundance | X06663 | Leucyltyros | Radius_SOS | 0,099586 | 0,049551 | NA |
| 3 | abundance | X06663 | Leucyltyros | Tibia_SOS | -0,05339 | 0,053558 | NA |
| 3 | abundance | X06663 | Leucyltyros | Handgrip | -0,01579 | 0,060849 | NA |
| 3 | abundance | X06675 | L-gamma-( | Tibia_length | 0,036356 | 0,055886 | NA |
| 3 | abundance | X06675 | L-gamma-( | Radius_length | 0,04521 | 0,074187 | NA |
| 3 | abundance | X06675 | L-gamma-( | Radius_SOS | 0,101688 | 0,052696 | NA |
| 3 | abundance | X06675 | L-gamma-( | Tibia_SOS | 3,96E-04 | 0,056606 | NA |
| 3 | abundance | X06675 | L-gamma-( | Handgrip | 0,087877 | 0,062511 | NA |
| 2b | abundance | X06680 | 7-Hydroxy- | Tibia_length | -0,08058 | 0,053952 | NA |
| 2b | abundance | X06680 | 7-Hydroxy- | Radius_length | -0,14723 | 0,072008 | NA |
| 2b | abundance | X06680 | 7-Hydroxy- | Radius_SOS | 0,034995 | 0,051514 | NA |
| 2b | abundance | X06680 | 7-Hydroxy- | Tibia_SOS | -0,06741 | 0,054174 | NA |
| 2b | abundance | X06680 | 7-Hydroxy- | Handgrip | -0,04877 | 0,061759 | NA |
| 3 | abundance | X06681 | Scymnol | Tibia_length | -0,06738 | 0,049478 | NA |
| 3 | abundance | X06681 | Scymnol | Radius_length | -0,10062 | 0,066608 | NA |
| 3 | abundance | X06681 | Scymnol | Radius_SOS | 0,009776 | 0,049027 | NA |
| 3 | abundance | X06681 | Scymnol | Tibia_SOS | 0,073858 | 0,052353 | NA |
| 3 | abundance | X06681 | Scymnol | Handgrip | -0,11106 | 0,060032 | NA |
| 3 | abundance | X06684 | Bromazine | Tibia_length | 0,014308 | 0,057047 | NA |
| 3 | abundance | X06684 | Bromazine | Radius_length | 0,100909 | 0,073986 | NA |
| 3 | abundance | X06684 | Bromazine | Radius_SOS | -0,01711 | 0,054296 | NA |
| 3 | abundance | X06684 | Bromazine | Tibia_SOS | 0,109592 | 0,057288 | NA |
| 3 | abundance | X06684 | Bromazine | Handgrip | -0,01926 | 0,060182 | NA |
| 2b | abundance | X06690 | 4-Methylca | Tibia_length | -0,13185 | 0,056951 | NA |

| | | | | | | | |
|---|---|---|---|---|---|---|---|
| 2b | abundance | X06690 | 4-Methylca | Radius_length | -0,08969 | 0,075065 | NA |
| 2b | abundance | X06690 | 4-Methylca | Radius_SOS | -0,13296 | 0,054364 | NA |
| 2b | abundance | X06690 | 4-Methylca | Tibia_SOS | -0,06872 | 0,060499 | NA |
| 2b | abundance | X06690 | 4-Methylca | Handgrip | -0,01645 | 0,063435 | NA |
| 3 | abundance | X06698 | entecavir | Tibia_length | 0,101358 | 0,053603 | NA |
| 3 | abundance | X06698 | entecavir | Radius_length | 0,125455 | 0,073105 | NA |
| 3 | abundance | X06698 | entecavir | Radius_SOS | 0,047574 | 0,051412 | NA |
| 3 | abundance | X06698 | entecavir | Tibia_SOS | 0,146076 | 0,054626 | NA |
| 3 | abundance | X06698 | entecavir | Handgrip | 0,103758 | 0,061238 | NA |
| 3 | abundance | X06703 | N6-METHY | Tibia_length | -0,01833 | 0,055883 | NA |
| 3 | abundance | X06703 | N6-METHY | Radius_length | -0,0327 | 0,073711 | NA |
| 3 | abundance | X06703 | N6-METHY | Radius_SOS | -0,03553 | 0,052985 | NA |
| 3 | abundance | X06703 | N6-METHY | Tibia_SOS | -0,14313 | 0,057121 | NA |
| 3 | abundance | X06703 | N6-METHY | Handgrip | -0,03742 | 0,060151 | NA |
| 3 | abundance | X06704 | feruloylagr | Tibia_length | -0,03691 | 0,051345 | NA |
| 3 | abundance | X06704 | feruloylagr | Radius_length | 0,021049 | 0,070145 | NA |
| 3 | abundance | X06704 | feruloylagr | Radius_SOS | -0,02052 | 0,049663 | NA |
| 3 | abundance | X06704 | feruloylagr | Tibia_SOS | 0,050821 | 0,052941 | NA |
| 3 | abundance | X06704 | feruloylagr | Handgrip | 0,017247 | 0,061357 | NA |
| 3 | abundance | X06709 | Tetraacety | Tibia_length | -0,10496 | 0,052876 | NA |
| 3 | abundance | X06709 | Tetraacety | Radius_length | -0,02949 | 0,072277 | NA |
| 3 | abundance | X06709 | Tetraacety | Radius_SOS | 0,010049 | 0,050712 | NA |
| 3 | abundance | X06709 | Tetraacety | Tibia_SOS | -0,09949 | 0,053939 | NA |
| 3 | abundance | X06709 | Tetraacety | Handgrip | -0,00132 | 0,061115 | NA |
| 3 | abundance | X06722 | LW800000 | Tibia_length | -0,00675 | 0,053179 | NA |
| 3 | abundance | X06722 | LW800000 | Radius_length | 0,015904 | 0,071926 | NA |
| 3 | abundance | X06722 | LW800000 | Radius_SOS | 0,003364 | 0,050807 | NA |
| 3 | abundance | X06722 | LW800000 | Tibia_SOS | 0,060353 | 0,053865 | NA |
| 3 | abundance | X06722 | LW800000 | Handgrip | 0,040105 | 0,061282 | NA |
| 3 | abundance | X06723 | 3-Hydroxy- | Tibia_length | -0,09098 | 0,054999 | NA |
| 3 | abundance | X06723 | 3-Hydroxy- | Radius_length | -0,11703 | 0,072912 | NA |
| 3 | abundance | X06723 | 3-Hydroxy- | Radius_SOS | -0,08827 | 0,052287 | NA |
| 3 | abundance | X06723 | 3-Hydroxy- | Tibia_SOS | -0,05767 | 0,055261 | NA |
| 3 | abundance | X06723 | 3-Hydroxy- | Handgrip | -0,14083 | 0,061739 | NA |
| 3 | abundance | X06727 | MFCD0087 | Tibia_length | 0,021897 | 0,054925 | NA |
| 3 | abundance | X06727 | MFCD0087 | Radius_length | 0,075461 | 0,073314 | NA |
| 3 | abundance | X06727 | MFCD0087 | Radius_SOS | 0,089648 | 0,052571 | NA |
| 3 | abundance | X06727 | MFCD0087 | Tibia_SOS | 0,027418 | 0,053738 | NA |
| 3 | abundance | X06727 | MFCD0087 | Handgrip | -0,02834 | 0,061094 | NA |
| 2b | abundance | X06735 | 7-Methylxa | Tibia_length | -0,04275 | 0,054748 | NA |
| 2b | abundance | X06735 | 7-Methylxa | Radius_length | -0,0288 | 0,073545 | NA |
| 2b | abundance | X06735 | 7-Methylxa | Radius_SOS | 0,068979 | 0,051909 | NA |
| 2b | abundance | X06735 | 7-Methylxa | Tibia_SOS | -0,01125 | 0,05434 | NA |
| 2b | abundance | X06735 | 7-Methylxa | Handgrip | 0,021141 | 0,061705 | NA |
| 3 | abundance | X06744 | Diethylpyro | Tibia_length | -0,08562 | 0,053141 | NA |
| 3 | abundance | X06744 | Diethylpyro | Radius_length | -0,17959 | 0,072013 | NA |
| 3 | abundance | X06744 | Diethylpyro | Radius_SOS | -0,08754 | 0,050655 | NA |
| 3 | abundance | X06744 | Diethylpyro | Tibia_SOS | -0,09135 | 0,055114 | NA |

| | | | | | | | |
|---|---|---|---|---|---|---|---|
| 3 | abundance | X06744 | Diethylpyr‹ | Handgrip | 0,01781 | 0,063007 | NA |
| 3 | abundance | X06750 | 1-PYRENYl | Tibia_length | -0,05846 | 0,054532 | NA |
| 3 | abundance | X06750 | 1-PYRENYl | Radius_length | -0,0132 | 0,073379 | NA |
| 3 | abundance | X06750 | 1-PYRENYl | Radius_SOS | 0,091798 | 0,051646 | NA |
| 3 | abundance | X06750 | 1-PYRENYl | Tibia_SOS | 0,053935 | 0,054281 | NA |
| 3 | abundance | X06750 | 1-PYRENYl | Handgrip | 0,055804 | 0,061677 | NA |
| 3 | abundance | X06764 | N-(4-Hydrc | Tibia_length | -0,16915 | 0,056521 | NA |
| 3 | abundance | X06764 | N-(4-Hydrc | Radius_length | -0,14114 | 0,074985 | NA |
| 3 | abundance | X06764 | N-(4-Hydrc | Radius_SOS | -0,05961 | 0,054297 | NA |
| 3 | abundance | X06764 | N-(4-Hydrc | Tibia_SOS | -0,12962 | 0,056727 | NA |
| 3 | abundance | X06764 | N-(4-Hydrc | Handgrip | -0,07431 | 0,061727 | NA |
| 1 | abundance | X06767 | 2-Hydroxyl | Tibia_length | -0,04938 | 0,053012 | NA |
| 1 | abundance | X06767 | 2-Hydroxyl | Radius_length | -0,05323 | 0,072316 | NA |
| 1 | abundance | X06767 | 2-Hydroxyl | Radius_SOS | 0,060691 | 0,050511 | NA |
| 1 | abundance | X06767 | 2-Hydroxyl | Tibia_SOS | -0,02594 | 0,054107 | NA |
| 1 | abundance | X06767 | 2-Hydroxyl | Handgrip | 0,149222 | 0,060532 | NA |
| 3 | abundance | X06768 | Sinapyl alc | Tibia_length | 0,043564 | 0,054993 | NA |
| 3 | abundance | X06768 | Sinapyl alc | Radius_length | 0,076407 | 0,073781 | NA |
| 3 | abundance | X06768 | Sinapyl alc | Radius_SOS | 0,07818 | 0,052158 | NA |
| 3 | abundance | X06768 | Sinapyl alc | Tibia_SOS | -0,11395 | 0,056292 | NA |
| 3 | abundance | X06768 | Sinapyl alc | Handgrip | 0,149412 | 0,062158 | NA |
| 3 | abundance | X06771 | MFCD0015 | Tibia_length | 0,17044 | 0,053559 | NA |
| 3 | abundance | X06771 | MFCD0015 | Radius_length | 0,187557 | 0,071867 | NA |
| 3 | abundance | X06771 | MFCD0015 | Radius_SOS | -0,00312 | 0,051758 | NA |
| 3 | abundance | X06771 | MFCD0015 | Tibia_SOS | 0,029434 | 0,055303 | NA |
| 3 | abundance | X06771 | MFCD0015 | Handgrip | 0,0503 | 0,062068 | NA |
| 3 | abundance | X06783 | N~6~-Octa | Tibia_length | -0,02314 | 0,053407 | NA |
| 3 | abundance | X06783 | N~6~-Octa | Radius_length | -8,32E-04 | 0,072051 | NA |
| 3 | abundance | X06783 | N~6~-Octa | Radius_SOS | 0,001686 | 0,050984 | NA |
| 3 | abundance | X06783 | N~6~-Octa | Tibia_SOS | 0,018038 | 0,054885 | NA |
| 3 | abundance | X06783 | N~6~-Octa | Handgrip | 0,046904 | 0,061478 | NA |
| 3 | abundance | X06797 | pro-gln | Tibia_length | -0,0966 | 0,05381 | NA |
| 3 | abundance | X06797 | pro-gln | Radius_length | -0,04364 | 0,072664 | NA |
| 3 | abundance | X06797 | pro-gln | Radius_SOS | -0,04105 | 0,051192 | NA |
| 3 | abundance | X06797 | pro-gln | Tibia_SOS | -0,11199 | 0,054305 | NA |
| 3 | abundance | X06797 | pro-gln | Handgrip | -0,08485 | 0,061934 | NA |
| 3 | abundance | X06805 | (9cis)-O~1 | Tibia_length | -0,00244 | 0,053064 | NA |
| 3 | abundance | X06805 | (9cis)-O~1 | Radius_length | 0,026608 | 0,072132 | NA |
| 3 | abundance | X06805 | (9cis)-O~1 | Radius_SOS | 0,018717 | 0,050656 | NA |
| 3 | abundance | X06805 | (9cis)-O~1 | Tibia_SOS | 0,056511 | 0,053544 | NA |
| 3 | abundance | X06805 | (9cis)-O~1 | Handgrip | 0,038791 | 0,061391 | NA |
| 3 | abundance | X06807 | 1,1'-[1,12-I | Tibia_length | 0,049484 | 0,050797 | NA |
| 3 | abundance | X06807 | 1,1'-[1,12-I | Radius_length | 0,062061 | 0,0694 | NA |
| 3 | abundance | X06807 | 1,1'-[1,12-I | Radius_SOS | 0,039404 | 0,049322 | NA |
| 3 | abundance | X06807 | 1,1'-[1,12-I | Tibia_SOS | -0,06482 | 0,05295 | NA |
| 3 | abundance | X06807 | 1,1'-[1,12-I | Handgrip | 0,017726 | 0,061034 | NA |
| 3 | abundance | X06812 | 2-Hydroxy- | Tibia_length | 0,065471 | 0,051536 | NA |
| 3 | abundance | X06812 | 2-Hydroxy- | Radius_length | 0,121866 | 0,070334 | NA |

| | | | | | | | |
|---|---|---|---|---|---|---|---|
| 3 | abundance | X06812 | 2-Hydroxy- | Radius_SOS | 0,003955 | 0,049757 | NA |
| 3 | abundance | X06812 | 2-Hydroxy- | Tibia_SOS | 0,092413 | 0,05281 | NA |
| 3 | abundance | X06812 | 2-Hydroxy- | Handgrip | 0,022469 | 0,06093 | NA |
| 3 | abundance | X06814 | N-Benzoyla | Tibia_length | -0,13136 | 0,051254 | NA |
| 3 | abundance | X06814 | N-Benzoyla | Radius_length | -0,15255 | 0,069824 | NA |
| 3 | abundance | X06814 | N-Benzoyla | Radius_SOS | -0,02911 | 0,049639 | NA |
| 3 | abundance | X06814 | N-Benzoyla | Tibia_SOS | -0,13185 | 0,052896 | NA |
| 3 | abundance | X06814 | N-Benzoyla | Handgrip | -0,09298 | 0,060525 | NA |
| 3 | abundance | X06818 | Homoanse | Tibia_length | 0,032353 | 0,052677 | NA |
| 3 | abundance | X06818 | Homoanse | Radius_length | 0,08255 | 0,071432 | NA |
| 3 | abundance | X06818 | Homoanse | Radius_SOS | 0,046599 | 0,050538 | NA |
| 3 | abundance | X06818 | Homoanse | Tibia_SOS | 0,009381 | 0,053944 | NA |
| 3 | abundance | X06818 | Homoanse | Handgrip | -0,11111 | 0,060978 | NA |
| 3 | abundance | X06819 | Primaquine | Tibia_length | 0,069175 | 0,050275 | NA |
| 3 | abundance | X06819 | Primaquine | Radius_length | -0,02637 | 0,068934 | NA |
| 3 | abundance | X06819 | Primaquine | Radius_SOS | -0,04668 | 0,049187 | NA |
| 3 | abundance | X06819 | Primaquine | Tibia_SOS | -0,0263 | 0,052827 | NA |
| 3 | abundance | X06819 | Primaquine | Handgrip | 0,10708 | 0,060206 | NA |
| 2b | abundance | X06820 | 1-(3,4-dim | Tibia_length | -0,10595 | 0,050716 | NA |
| 2b | abundance | X06820 | 1-(3,4-dim | Radius_length | -0,1193 | 0,069003 | NA |
| 2b | abundance | X06820 | 1-(3,4-dim | Radius_SOS | 0,002599 | 0,049341 | NA |
| 2b | abundance | X06820 | 1-(3,4-dim | Tibia_SOS | -0,0508 | 0,052711 | NA |
| 2b | abundance | X06820 | 1-(3,4-dim | Handgrip | -0,05757 | 0,060129 | NA |
| 3 | abundance | X06836 | asn-lys | Tibia_length | -0,11015 | 0,057788 | NA |
| 3 | abundance | X06836 | asn-lys | Radius_length | -0,05364 | 0,07541 | NA |
| 3 | abundance | X06836 | asn-lys | Radius_SOS | -0,13845 | 0,055073 | NA |
| 3 | abundance | X06836 | asn-lys | Tibia_SOS | 0,070933 | 0,057623 | NA |
| 3 | abundance | X06836 | asn-lys | Handgrip | 0,059631 | 0,063449 | NA |
| 3 | abundance | X06850 | Leu-Val | Tibia_length | 0,00905 | 0,05286 | NA |
| 3 | abundance | X06850 | Leu-Val | Radius_length | -0,02058 | 0,072052 | NA |
| 3 | abundance | X06850 | Leu-Val | Radius_SOS | -0,08239 | 0,050239 | NA |
| 3 | abundance | X06850 | Leu-Val | Tibia_SOS | -0,0039 | 0,054078 | NA |
| 3 | abundance | X06850 | Leu-Val | Handgrip | 0,005546 | 0,061803 | NA |
| 2b | abundance | X06856 | Tiglic acid | Tibia_length | -0,02767 | 0,054759 | NA |
| 2b | abundance | X06856 | Tiglic acid | Radius_length | -0,00171 | 0,07342 | NA |
| 2b | abundance | X06856 | Tiglic acid | Radius_SOS | -0,05072 | 0,052061 | NA |
| 2b | abundance | X06856 | Tiglic acid | Tibia_SOS | 0,024569 | 0,056093 | NA |
| 2b | abundance | X06856 | Tiglic acid | Handgrip | 0,029463 | 0,062545 | NA |
| 3 | abundance | X06858 | tert-Butyl 3 | Tibia_length | -0,00153 | 0,052093 | NA |
| 3 | abundance | X06858 | tert-Butyl 3 | Radius_length | 0,094369 | 0,071222 | NA |
| 3 | abundance | X06858 | tert-Butyl 3 | Radius_SOS | -0,00341 | 0,050089 | NA |
| 3 | abundance | X06858 | tert-Butyl 3 | Tibia_SOS | 0,072174 | 0,053817 | NA |
| 3 | abundance | X06858 | tert-Butyl 3 | Handgrip | 0,07778 | 0,060849 | NA |
| 3 | abundance | X06859 | Methylol D | Tibia_length | 0,013763 | 0,052407 | NA |
| 3 | abundance | X06859 | Methylol D | Radius_length | 0,147736 | 0,070831 | NA |
| 3 | abundance | X06859 | Methylol D | Radius_SOS | 0,033363 | 0,050311 | NA |
| 3 | abundance | X06859 | Methylol D | Tibia_SOS | -0,02246 | 0,054224 | NA |
| 3 | abundance | X06859 | Methylol D | Handgrip | -0,09627 | 0,061282 | NA |

| | | | | | | | |
|---|---|---|---|---|---|---|---|
| 3 | abundance | X06861 | 4-(4-Deoxy | Tibia_length | 0,089862 | 0,051573 | NA |
| 3 | abundance | X06861 | 4-(4-Deoxy | Radius_length | 0,161718 | 0,069556 | NA |
| 3 | abundance | X06861 | 4-(4-Deoxy | Radius_SOS | -0,0367 | 0,049691 | NA |
| 3 | abundance | X06861 | 4-(4-Deoxy | Tibia_SOS | -0,02612 | 0,053247 | NA |
| 3 | abundance | X06861 | 4-(4-Deoxy | Handgrip | 0,108632 | 0,061093 | NA |
| 3 | abundance | X06866 | tert-Butyl 3 | Tibia_length | -0,04579 | 0,052775 | NA |
| 3 | abundance | X06866 | tert-Butyl 3 | Radius_length | 0,026096 | 0,07201 | NA |
| 3 | abundance | X06866 | tert-Butyl 3 | Radius_SOS | 0,020269 | 0,050582 | NA |
| 3 | abundance | X06866 | tert-Butyl 3 | Tibia_SOS | -0,02372 | 0,054468 | NA |
| 3 | abundance | X06866 | tert-Butyl 3 | Handgrip | 0,026373 | 0,062104 | NA |
| 3 | abundance | X06870 | Menadiol | Tibia_length | -0,02258 | 0,056304 | NA |
| 3 | abundance | X06870 | Menadiol | Radius_length | 0,037226 | 0,074458 | NA |
| 3 | abundance | X06870 | Menadiol | Radius_SOS | -0,03109 | 0,05371 | NA |
| 3 | abundance | X06870 | Menadiol | Tibia_SOS | -0,09844 | 0,057728 | NA |
| 3 | abundance | X06870 | Menadiol | Handgrip | -0,02768 | 0,062667 | NA |
| 3 | abundance | X06874 | N-Acetyl-5 | Tibia_length | 0,038152 | 0,055535 | NA |
| 3 | abundance | X06874 | N-Acetyl-5 | Radius_length | 0,061546 | 0,073755 | NA |
| 3 | abundance | X06874 | N-Acetyl-5 | Radius_SOS | -0,08377 | 0,052462 | NA |
| 3 | abundance | X06874 | N-Acetyl-5 | Tibia_SOS | -0,07474 | 0,057796 | NA |
| 3 | abundance | X06874 | N-Acetyl-5 | Handgrip | 0,011704 | 0,063472 | NA |
| 3 | abundance | X06875 | Nicotine gl | Tibia_length | 0,043837 | 0,053253 | NA |
| 3 | abundance | X06875 | Nicotine gl | Radius_length | 0,043331 | 0,072177 | NA |
| 3 | abundance | X06875 | Nicotine gl | Radius_SOS | 0,030089 | 0,050966 | NA |
| 3 | abundance | X06875 | Nicotine gl | Tibia_SOS | 0,076538 | 0,054199 | NA |
| 3 | abundance | X06875 | Nicotine gl | Handgrip | -0,05668 | 0,061503 | NA |
| 3 | abundance | X06880 | pentobarbi | Tibia_length | 0,056551 | 0,051559 | NA |
| 3 | abundance | X06880 | pentobarbi | Radius_length | 0,070223 | 0,070739 | NA |
| 3 | abundance | X06880 | pentobarbi | Radius_SOS | -0,0039 | 0,049824 | NA |
| 3 | abundance | X06880 | pentobarbi | Tibia_SOS | -0,01054 | 0,053448 | NA |
| 3 | abundance | X06880 | pentobarbi | Handgrip | 0,048896 | 0,061211 | NA |
| 3 | abundance | X06883 | Agomelatir | Tibia_length | -0,09117 | 0,053593 | NA |
| 3 | abundance | X06883 | Agomelatir | Radius_length | -0,10354 | 0,072102 | NA |
| 3 | abundance | X06883 | Agomelatir | Radius_SOS | -0,01737 | 0,051079 | NA |
| 3 | abundance | X06883 | Agomelatir | Tibia_SOS | -0,09055 | 0,054464 | NA |
| 3 | abundance | X06883 | Agomelatir | Handgrip | 0,005076 | 0,061538 | NA |
| 2a | abundance | X06889 | 3,4-Dimetl | Tibia_length | 0,076816 | 0,052617 | NA |
| 2a | abundance | X06889 | 3,4-Dimetl | Radius_length | 0,122478 | 0,071219 | NA |
| 2a | abundance | X06889 | 3,4-Dimetl | Radius_SOS | 0,033918 | 0,050469 | NA |
| 2a | abundance | X06889 | 3,4-Dimetl | Tibia_SOS | -0,00998 | 0,053909 | NA |
| 2a | abundance | X06889 | 3,4-Dimetl | Handgrip | 0,120551 | 0,061114 | NA |
| 3 | abundance | X06896 | Pirbuterol | Tibia_length | -0,11454 | 0,055738 | NA |
| 3 | abundance | X06896 | Pirbuterol | Radius_length | -0,12466 | 0,073643 | NA |
| 3 | abundance | X06896 | Pirbuterol | Radius_SOS | -0,00712 | 0,053037 | NA |
| 3 | abundance | X06896 | Pirbuterol | Tibia_SOS | -0,00527 | 0,056137 | NA |
| 3 | abundance | X06896 | Pirbuterol | Handgrip | 0,001672 | 0,062864 | NA |
| 3 | abundance | X06902 | Homocyste | Tibia_length | 0,081324 | 0,055105 | NA |
| 3 | abundance | X06902 | Homocyste | Radius_length | 0,104777 | 0,074082 | NA |
| 3 | abundance | X06902 | Homocyste | Radius_SOS | 0,089676 | 0,052168 | NA |

| | | | | | | | |
|---|---|---|---|---|---|---|---|
| 3 | abundance | X06902 | Homocyst | Tibia_SOS | 0,102947 | 0,05573 | NA |
| 3 | abundance | X06902 | Homocyst | Handgrip | 0,045326 | 0,063152 | NA |
| 2b | abundance | X06903 | Esculin | Tibia_length | -0,05818 | 0,055993 | NA |
| 2b | abundance | X06903 | Esculin | Radius_length | -0,08847 | 0,073603 | NA |
| 2b | abundance | X06903 | Esculin | Radius_SOS | -0,09378 | 0,053585 | NA |
| 2b | abundance | X06903 | Esculin | Tibia_SOS | -0,04748 | 0,056707 | NA |
| 2b | abundance | X06903 | Esculin | Handgrip | -0,07039 | 0,062551 | NA |
| 3 | abundance | X06905 | Ectoine | Tibia_length | 0,001601 | 0,055002 | NA |
| 3 | abundance | X06905 | Ectoine | Radius_length | -0,02741 | 0,073509 | NA |
| 3 | abundance | X06905 | Ectoine | Radius_SOS | -0,03201 | 0,052243 | NA |
| 3 | abundance | X06905 | Ectoine | Tibia_SOS | -0,06419 | 0,055673 | NA |
| 3 | abundance | X06905 | Ectoine | Handgrip | -0,05445 | 0,062132 | NA |
| 3 | abundance | X06906 | Arg-pro | Tibia_length | 0,025879 | 0,051776 | NA |
| 3 | abundance | X06906 | Arg-pro | Radius_length | 0,072553 | 0,070589 | NA |
| 3 | abundance | X06906 | Arg-pro | Radius_SOS | 0,012145 | 0,049839 | NA |
| 3 | abundance | X06906 | Arg-pro | Tibia_SOS | 0,06442 | 0,053373 | NA |
| 3 | abundance | X06906 | Arg-pro | Handgrip | 0,042568 | 0,060565 | NA |
| 3 | abundance | X06917 | Alanyltrypt | Tibia_length | -0,10399 | 0,055014 | NA |
| 3 | abundance | X06917 | Alanyltrypt | Radius_length | -0,04648 | 0,073905 | NA |
| 3 | abundance | X06917 | Alanyltrypt | Radius_SOS | 0,024889 | 0,05233 | NA |
| 3 | abundance | X06917 | Alanyltrypt | Tibia_SOS | 0,009431 | 0,055804 | NA |
| 3 | abundance | X06917 | Alanyltrypt | Handgrip | 0,061714 | 0,061015 | NA |
| 3 | abundance | X06933 | 3-Methoxy- | Tibia_length | -0,09044 | 0,052294 | NA |
| 3 | abundance | X06933 | 3-Methoxy- | Radius_length | -0,08699 | 0,071566 | NA |
| 3 | abundance | X06933 | 3-Methoxy- | Radius_SOS | 0,018143 | 0,050061 | NA |
| 3 | abundance | X06933 | 3-Methoxy- | Tibia_SOS | -0,09548 | 0,053282 | NA |
| 3 | abundance | X06933 | 3-Methoxy- | Handgrip | -0,13456 | 0,060573 | NA |
| 3 | abundance | X06943 | NSC 9277 | Tibia_length | -0,11298 | 0,054299 | NA |
| 3 | abundance | X06943 | NSC 9277 | Radius_length | 0,024543 | 0,073394 | NA |
| 3 | abundance | X06943 | NSC 9277 | Radius_SOS | 0,068491 | 0,051798 | NA |
| 3 | abundance | X06943 | NSC 9277 | Tibia_SOS | 0,080848 | 0,055383 | NA |
| 3 | abundance | X06943 | NSC 9277 | Handgrip | 0,0717 | 0,061844 | NA |
| 3 | abundance | X06946 | SECONAL | Tibia_length | -0,11713 | 0,054175 | NA |
| 3 | abundance | X06946 | SECONAL | Radius_length | -0,12585 | 0,072506 | NA |
| 3 | abundance | X06946 | SECONAL | Radius_SOS | -0,04603 | 0,051903 | NA |
| 3 | abundance | X06946 | SECONAL | Tibia_SOS | -0,09318 | 0,055212 | NA |
| 3 | abundance | X06946 | SECONAL | Handgrip | -0,15414 | 0,061152 | NA |
| 3 | abundance | X06948 | (1R,3R,5R) | Tibia_length | 0,023472 | 0,054973 | NA |
| 3 | abundance | X06948 | (1R,3R,5R) | Radius_length | 0,119594 | 0,072886 | NA |
| 3 | abundance | X06948 | (1R,3R,5R) | Radius_SOS | 0,024257 | 0,052007 | NA |
| 3 | abundance | X06948 | (1R,3R,5R) | Tibia_SOS | 0,060276 | 0,05574 | NA |
| 3 | abundance | X06948 | (1R,3R,5R) | Handgrip | -0,07682 | 0,063576 | NA |
| 2b | abundance | X06958 | 2-[(carboxy | Tibia_length | -0,02865 | 0,051944 | NA |
| 2b | abundance | X06958 | 2-[(carboxy | Radius_length | 0,114578 | 0,070477 | NA |
| 2b | abundance | X06958 | 2-[(carboxy | Radius_SOS | 0,083662 | 0,049763 | NA |
| 2b | abundance | X06958 | 2-[(carboxy | Tibia_SOS | 0,114901 | 0,053475 | NA |
| 2b | abundance | X06958 | 2-[(carboxy | Handgrip | -0,04664 | 0,061802 | NA |
| 3 | abundance | X06961 | Zalcitabine | Tibia_length | -0,15419 | 0,056592 | NA |

| | | | | | | | | |
|---|---|---|---|---|---|---|---|---|
| | 3 | abundance | X06961 | Zalcitabine | Radius_length | -0,17899 | 0,074107 | NA |
| | 3 | abundance | X06961 | Zalcitabine | Radius_SOS | -0,03748 | 0,053851 | NA |
| | 3 | abundance | X06961 | Zalcitabine | Tibia_SOS | -0,24567 | 0,056158 | NA |
| | 3 | abundance | X06961 | Zalcitabine | Handgrip | -0,14281 | 0,06285 | NA |
| | 3 | abundance | X06977 | 4-Hydroxyp | Tibia_length | -0,0686 | 0,055717 | NA |
| | 3 | abundance | X06977 | 4-Hydroxyp | Radius_length | -0,03907 | 0,074514 | NA |
| | 3 | abundance | X06977 | 4-Hydroxyp | Radius_SOS | -0,02258 | 0,052958 | NA |
| | 3 | abundance | X06977 | 4-Hydroxyp | Tibia_SOS | 0,104063 | 0,055155 | NA |
| | 3 | abundance | X06977 | 4-Hydroxyp | Handgrip | 0,026248 | 0,062609 | NA |
| | 3 | abundance | X06985 | N-Ethylpro | Tibia_length | -0,03885 | 0,053908 | NA |
| | 3 | abundance | X06985 | N-Ethylpro | Radius_length | -0,01268 | 0,07311 | NA |
| | 3 | abundance | X06985 | N-Ethylpro | Radius_SOS | -0,03012 | 0,051307 | NA |
| | 3 | abundance | X06985 | N-Ethylpro | Tibia_SOS | 0,065901 | 0,055055 | NA |
| | 3 | abundance | X06985 | N-Ethylpro | Handgrip | 0,03299 | 0,061223 | NA |
| | 3 | abundance | X07002 | Paraldehyc | Tibia_length | 0,021049 | 0,05586 | NA |
| | 3 | abundance | X07002 | Paraldehyc | Radius_length | 0,060662 | 0,073961 | NA |
| | 3 | abundance | X07002 | Paraldehyc | Radius_SOS | -0,00641 | 0,053165 | NA |
| | 3 | abundance | X07002 | Paraldehyc | Tibia_SOS | 0,05237 | 0,056911 | NA |
| | 3 | abundance | X07002 | Paraldehyc | Handgrip | 0,006827 | 0,063346 | NA |
| | 3 | abundance | X07013 | pimethixer | Tibia_length | 0,052023 | 0,051502 | NA |
| | 3 | abundance | X07013 | pimethixer | Radius_length | 0,022907 | 0,07041 | NA |
| | 3 | abundance | X07013 | pimethixer | Radius_SOS | -4,30E-04 | 0,049738 | NA |
| | 3 | abundance | X07013 | pimethixer | Tibia_SOS | -0,00665 | 0,053202 | NA |
| | 3 | abundance | X07013 | pimethixer | Handgrip | -0,00722 | 0,061181 | NA |
| | 3 | abundance | X07014 | Octyl benz | Tibia_length | 0,050226 | 0,053022 | NA |
| | 3 | abundance | X07014 | Octyl benz | Radius_length | 0,08625 | 0,071601 | NA |
| | 3 | abundance | X07014 | Octyl benz | Radius_SOS | -0,03123 | 0,050708 | NA |
| | 3 | abundance | X07014 | Octyl benz | Tibia_SOS | 0,018126 | 0,053493 | NA |
| | 3 | abundance | X07014 | Octyl benz | Handgrip | -0,01663 | 0,061499 | NA |
| 2b | | abundance | X07026 | 7Î±-Hydrox | Tibia_length | 0,029098 | 0,052006 | NA |
| 2b | | abundance | X07026 | 7Î±-Hydrox | Radius_length | 0,107971 | 0,070796 | NA |
| 2b | | abundance | X07026 | 7Î±-Hydrox | Radius_SOS | 0,097288 | 0,049809 | NA |
| 2b | | abundance | X07026 | 7Î±-Hydrox | Tibia_SOS | 0,13538 | 0,05389 | NA |
| 2b | | abundance | X07026 | 7Î±-Hydrox | Handgrip | 0,063795 | 0,061052 | NA |
| | 3 | abundance | X07027 | Allyl merca | Tibia_length | -0,11209 | 0,055583 | NA |
| | 3 | abundance | X07027 | Allyl merca | Radius_length | -0,06685 | 0,073822 | NA |
| | 3 | abundance | X07027 | Allyl merca | Radius_SOS | -0,02035 | 0,052721 | NA |
| | 3 | abundance | X07027 | Allyl merca | Tibia_SOS | 0,05716 | 0,055942 | NA |
| | 3 | abundance | X07027 | Allyl merca | Handgrip | 0,051205 | 0,060114 | NA |
| | 3 | abundance | X07028 | N-Pentano | Tibia_length | 0,193979 | 0,053041 | NA |
| | 3 | abundance | X07028 | N-Pentano | Radius_length | 0,145861 | 0,07263 | NA |
| | 3 | abundance | X07028 | N-Pentano | Radius_SOS | 0,095882 | 0,051335 | NA |
| | 3 | abundance | X07028 | N-Pentano | Tibia_SOS | 0,156631 | 0,054944 | NA |
| | 3 | abundance | X07028 | N-Pentano | Handgrip | 0,070124 | 0,061043 | NA |
| | 3 | abundance | X07038 | Piperidine | Tibia_length | -0,01698 | 0,05044 | NA |
| | 3 | abundance | X07038 | Piperidine | Radius_length | -0,02204 | 0,068842 | NA |
| | 3 | abundance | X07038 | Piperidine | Radius_SOS | -0,03273 | 0,049187 | NA |
| | 3 | abundance | X07038 | Piperidine | Tibia_SOS | -0,02928 | 0,052777 | NA |

| | | | | | | | | |
|---|---|---|---|---|---|---|---|---|
| | 3 | abundance | X07038 | Piperidine | Handgrip | 0,044143 | 0,060733 | NA |
| | 3 | abundance | X07040 | 6-Hydroxyp | Tibia_length | 0,083608 | 0,050118 | NA |
| | 3 | abundance | X07040 | 6-Hydroxyp | Radius_length | 0,141239 | 0,068021 | NA |
| | 3 | abundance | X07040 | 6-Hydroxyp | Radius_SOS | 0,036518 | 0,049112 | NA |
| | 3 | abundance | X07040 | 6-Hydroxyp | Tibia_SOS | -0,00307 | 0,052568 | NA |
| | 3 | abundance | X07040 | 6-Hydroxyp | Handgrip | 0,045804 | 0,060312 | NA |
| | 3 | abundance | X07050 | 2-Methylth | Tibia_length | 0,090124 | 0,051078 | NA |
| | 3 | abundance | X07050 | 2-Methylth | Radius_length | 0,167208 | 0,068944 | NA |
| | 3 | abundance | X07050 | 2-Methylth | Radius_SOS | 0,02503 | 0,049449 | NA |
| | 3 | abundance | X07050 | 2-Methylth | Tibia_SOS | 0,132958 | 0,052524 | NA |
| | 3 | abundance | X07050 | 2-Methylth | Handgrip | 0,105814 | 0,060213 | NA |
| | 3 | abundance | X07051 | O-succinyl | Tibia_length | 0,012573 | 0,049856 | NA |
| | 3 | abundance | X07051 | O-succinyl | Radius_length | 0,119294 | 0,06732 | NA |
| | 3 | abundance | X07051 | O-succinyl | Radius_SOS | 1,45E-04 | 0,049048 | NA |
| | 3 | abundance | X07051 | O-succinyl | Tibia_SOS | 0,010989 | 0,052879 | NA |
| | 3 | abundance | X07051 | O-succinyl | Handgrip | -0,053 | 0,060217 | NA |
| 2b | | abundance | X07054 | Valylprolin | Tibia_length | -0,02519 | 0,053275 | NA |
| 2b | | abundance | X07054 | Valylprolin | Radius_length | 0,008665 | 0,072453 | NA |
| 2b | | abundance | X07054 | Valylprolin | Radius_SOS | -0,01655 | 0,050965 | NA |
| 2b | | abundance | X07054 | Valylprolin | Tibia_SOS | 0,06465 | 0,054966 | NA |
| 2b | | abundance | X07054 | Valylprolin | Handgrip | 0,101705 | 0,061576 | NA |
| | 3 | abundance | X07057 | Leu-Leu | Tibia_length | -0,00783 | 0,053728 | NA |
| | 3 | abundance | X07057 | Leu-Leu | Radius_length | 0,0819 | 0,072631 | NA |
| | 3 | abundance | X07057 | Leu-Leu | Radius_SOS | -0,01967 | 0,05121 | NA |
| | 3 | abundance | X07057 | Leu-Leu | Tibia_SOS | 0,043505 | 0,056063 | NA |
| | 3 | abundance | X07057 | Leu-Leu | Handgrip | -0,00326 | 0,062931 | NA |
| | 3 | abundance | X07073 | asn-val | Tibia_length | -0,01297 | 0,052478 | NA |
| | 3 | abundance | X07073 | asn-val | Radius_length | 0,125858 | 0,071428 | NA |
| | 3 | abundance | X07073 | asn-val | Radius_SOS | 0,031459 | 0,050435 | NA |
| | 3 | abundance | X07073 | asn-val | Tibia_SOS | 0,041468 | 0,053417 | NA |
| | 3 | abundance | X07073 | asn-val | Handgrip | -0,08298 | 0,06115 | NA |
| | 1 | abundance | X07077 | Hyodeoxyc | Tibia_length | -0,0031 | 0,051544 | NA |
| | 1 | abundance | X07077 | Hyodeoxyc | Radius_length | -0,04374 | 0,070861 | NA |
| | 1 | abundance | X07077 | Hyodeoxyc | Radius_SOS | 0,094373 | 0,04963 | NA |
| | 1 | abundance | X07077 | Hyodeoxyc | Tibia_SOS | 0,021679 | 0,053424 | NA |
| | 1 | abundance | X07077 | Hyodeoxyc | Handgrip | 0,048467 | 0,060696 | NA |
| | 3 | abundance | X07079 | N-Pentano | Tibia_length | 0,187092 | 0,052438 | NA |
| | 3 | abundance | X07079 | N-Pentano | Radius_length | 0,130749 | 0,072105 | NA |
| | 3 | abundance | X07079 | N-Pentano | Radius_SOS | 0,082246 | 0,050879 | NA |
| | 3 | abundance | X07079 | N-Pentano | Tibia_SOS | 0,157963 | 0,054173 | NA |
| | 3 | abundance | X07079 | N-Pentano | Handgrip | 0,073031 | 0,060752 | NA |
| | 3 | abundance | X07081 | N-LACTOYI | Tibia_length | -0,03171 | 0,053697 | NA |
| | 3 | abundance | X07081 | N-LACTOYI | Radius_length | -0,07256 | 0,072201 | NA |
| | 3 | abundance | X07081 | N-LACTOYI | Radius_SOS | 0,012966 | 0,051154 | NA |
| | 3 | abundance | X07081 | N-LACTOYI | Tibia_SOS | -0,0183 | 0,054973 | NA |
| | 3 | abundance | X07081 | N-LACTOYI | Handgrip | -0,07126 | 0,062295 | NA |
| | 3 | abundance | X07084 | heptabarbi | Tibia_length | 0,029443 | 0,052254 | NA |
| | 3 | abundance | X07084 | heptabarbi | Radius_length | 0,071104 | 0,07128 | NA |

| | | | | | | | |
|---|---|---|---|---|---|---|---|
| | 3 | abundance | X07084 | heptabarbi | Radius_SOS | 0,034692 | 0,050223 | NA |
| | 3 | abundance | X07084 | heptabarbi | Tibia_SOS | 0,019146 | 0,053361 | NA |
| | 3 | abundance | X07084 | heptabarbi | Handgrip | -0,02888 | 0,061371 | NA |
| | 3 | abundance | X07089 | Pregabalin | Tibia_length | -0,14134 | 0,052381 | NA |
| | 3 | abundance | X07089 | Pregabalin | Radius_length | -0,16872 | 0,070773 | NA |
| | 3 | abundance | X07089 | Pregabalin | Radius_SOS | -0,11396 | 0,050174 | NA |
| | 3 | abundance | X07089 | Pregabalin | Tibia_SOS | -0,0663 | 0,054416 | NA |
| | 3 | abundance | X07089 | Pregabalin | Handgrip | -0,10563 | 0,0605 | NA |
| | 3 | abundance | X07092 | Dibutyl ma | Tibia_length | 0,095306 | 0,050728 | NA |
| | 3 | abundance | X07092 | Dibutyl ma | Radius_length | 0,05183 | 0,06981 | NA |
| | 3 | abundance | X07092 | Dibutyl ma | Radius_SOS | 0,073963 | 0,049361 | NA |
| | 3 | abundance | X07092 | Dibutyl ma | Tibia_SOS | 0,008564 | 0,053103 | NA |
| | 3 | abundance | X07092 | Dibutyl ma | Handgrip | 0,002374 | 0,061292 | NA |
| | 3 | abundance | X07107 | Lysylvaline | Tibia_length | 0,062336 | 0,051776 | NA |
| | 3 | abundance | X07107 | Lysylvaline | Radius_length | 0,033788 | 0,070941 | NA |
| | 3 | abundance | X07107 | Lysylvaline | Radius_SOS | -0,00102 | 0,050008 | NA |
| | 3 | abundance | X07107 | Lysylvaline | Tibia_SOS | -0,03893 | 0,054016 | NA |
| | 3 | abundance | X07107 | Lysylvaline | Handgrip | 0,068033 | 0,062356 | NA |
| | 3 | abundance | X07112 | MFCD0272 | Tibia_length | -0,08099 | 0,053333 | NA |
| | 3 | abundance | X07112 | MFCD0272 | Radius_length | -0,08935 | 0,072374 | NA |
| | 3 | abundance | X07112 | MFCD0272 | Radius_SOS | 0,014578 | 0,051134 | NA |
| | 3 | abundance | X07112 | MFCD0272 | Tibia_SOS | -0,06979 | 0,054402 | NA |
| | 3 | abundance | X07112 | MFCD0272 | Handgrip | -0,10836 | 0,062826 | NA |
| | 3 | abundance | X07113 | Sulfoaceta | Tibia_length | 0,021406 | 0,054894 | NA |
| | 3 | abundance | X07113 | Sulfoaceta | Radius_length | -0,16059 | 0,072979 | NA |
| | 3 | abundance | X07113 | Sulfoaceta | Radius_SOS | -0,11149 | 0,052573 | NA |
| | 3 | abundance | X07113 | Sulfoaceta | Tibia_SOS | -0,01553 | 0,056807 | NA |
| | 3 | abundance | X07113 | Sulfoaceta | Handgrip | -0,01974 | 0,062705 | NA |
| | 3 | abundance | X07125 | Zalcitabine | Tibia_length | -0,05401 | 0,057689 | NA |
| | 3 | abundance | X07125 | Zalcitabine | Radius_length | -0,0093 | 0,075344 | NA |
| | 3 | abundance | X07125 | Zalcitabine | Radius_SOS | 0,048086 | 0,054877 | NA |
| | 3 | abundance | X07125 | Zalcitabine | Tibia_SOS | -0,06863 | 0,057305 | NA |
| | 3 | abundance | X07125 | Zalcitabine | Handgrip | -0,06472 | 0,063179 | NA |
| | 3 | abundance | X07126 | Leu-Val | Tibia_length | -0,03713 | 0,050849 | NA |
| | 3 | abundance | X07126 | Leu-Val | Radius_length | -0,09651 | 0,069209 | NA |
| | 3 | abundance | X07126 | Leu-Val | Radius_SOS | 0,006319 | 0,049411 | NA |
| | 3 | abundance | X07126 | Leu-Val | Tibia_SOS | -0,00444 | 0,052768 | NA |
| | 3 | abundance | X07126 | Leu-Val | Handgrip | -0,07313 | 0,060875 | NA |
| | 3 | abundance | X07127 | N~5~-[P-Ar | Tibia_length | 0,033681 | 0,056274 | NA |
| | 3 | abundance | X07127 | N~5~-[P-Ar | Radius_length | 0,08383 | 0,074322 | NA |
| | 3 | abundance | X07127 | N~5~-[P-Ar | Radius_SOS | -0,02498 | 0,053473 | NA |
| | 3 | abundance | X07127 | N~5~-[P-Ar | Tibia_SOS | 0,005774 | 0,056538 | NA |
| | 3 | abundance | X07127 | N~5~-[P-Ar | Handgrip | 0,121057 | 0,062138 | NA |
| 2b | | abundance | X07134 | 4-Phenolsu | Tibia_length | -0,09771 | 0,051685 | NA |
| 2b | | abundance | X07134 | 4-Phenolsu | Radius_length | -0,06719 | 0,070723 | NA |
| 2b | | abundance | X07134 | 4-Phenolsu | Radius_SOS | -0,09754 | 0,049722 | NA |
| 2b | | abundance | X07134 | 4-Phenolsu | Tibia_SOS | 0,035396 | 0,053604 | NA |
| 2b | | abundance | X07134 | 4-Phenolsu | Handgrip | -0,05085 | 0,061344 | NA |

| | | | | | | | |
|---|---|---|---|---|---|---|---|
| 3 | abundance | X07136 | hexobarbit | Tibia_length | -0,19506 | 0,053667 | NA |
| 3 | abundance | X07136 | hexobarbit | Radius_length | -0,10116 | 0,072686 | NA |
| 3 | abundance | X07136 | hexobarbit | Radius_SOS | -0,06975 | 0,051604 | NA |
| 3 | abundance | X07136 | hexobarbit | Tibia_SOS | -0,06928 | 0,055688 | NA |
| 3 | abundance | X07136 | hexobarbit | Handgrip | 0,043566 | 0,061805 | NA |
| 3 | abundance | X07139 | N,N-Diethy | Tibia_length | -0,00163 | 0,052542 | NA |
| 3 | abundance | X07139 | N,N-Diethy | Radius_length | -0,05084 | 0,071296 | NA |
| 3 | abundance | X07139 | N,N-Diethy | Radius_SOS | 0,009261 | 0,050304 | NA |
| 3 | abundance | X07139 | N,N-Diethy | Tibia_SOS | -0,02563 | 0,053711 | NA |
| 3 | abundance | X07139 | N,N-Diethy | Handgrip | 0,08475 | 0,060803 | NA |
| 3 | abundance | X07146 | meprobam | Tibia_length | -0,07175 | 0,056424 | NA |
| 3 | abundance | X07146 | meprobam | Radius_length | 0,041712 | 0,074657 | NA |
| 3 | abundance | X07146 | meprobam | Radius_SOS | -0,03676 | 0,053743 | NA |
| 3 | abundance | X07146 | meprobam | Tibia_SOS | -0,00664 | 0,058833 | NA |
| 3 | abundance | X07146 | meprobam | Handgrip | 0,007537 | 0,063789 | NA |
| 3 | abundance | X07155 | Naphthale | Tibia_length | 0,049107 | 0,051611 | NA |
| 3 | abundance | X07155 | Naphthale | Radius_length | 0,10071 | 0,069797 | NA |
| 3 | abundance | X07155 | Naphthale | Radius_SOS | -0,03361 | 0,049561 | NA |
| 3 | abundance | X07155 | Naphthale | Tibia_SOS | -0,00703 | 0,053064 | NA |
| 3 | abundance | X07155 | Naphthale | Handgrip | -0,05313 | 0,060803 | NA |
| 3 | abundance | X07164 | N-Acetyl-5 | Tibia_length | -0,02619 | 0,052612 | NA |
| 3 | abundance | X07164 | N-Acetyl-5 | Radius_length | 0,068938 | 0,071458 | NA |
| 3 | abundance | X07164 | N-Acetyl-5 | Radius_SOS | 0,030908 | 0,050222 | NA |
| 3 | abundance | X07164 | N-Acetyl-5 | Tibia_SOS | 0,026429 | 0,054934 | NA |
| 3 | abundance | X07164 | N-Acetyl-5 | Handgrip | 0,058975 | 0,061378 | NA |
| 3 | abundance | X07165 | pro-gln | Tibia_length | 0,062673 | 0,051595 | NA |
| 3 | abundance | X07165 | pro-gln | Radius_length | -0,01495 | 0,070923 | NA |
| 3 | abundance | X07165 | pro-gln | Radius_SOS | 0,031768 | 0,049883 | NA |
| 3 | abundance | X07165 | pro-gln | Tibia_SOS | 0,01233 | 0,053225 | NA |
| 3 | abundance | X07165 | pro-gln | Handgrip | -0,09777 | 0,060721 | NA |
| 3 | abundance | X07168 | L-(+)-Eryth | Tibia_length | 0,043361 | 0,05447 | NA |
| 3 | abundance | X07168 | L-(+)-Eryth | Radius_length | 0,073138 | 0,073465 | NA |
| 3 | abundance | X07168 | L-(+)-Eryth | Radius_SOS | -0,02762 | 0,051971 | NA |
| 3 | abundance | X07168 | L-(+)-Eryth | Tibia_SOS | 0,037731 | 0,055703 | NA |
| 3 | abundance | X07168 | L-(+)-Eryth | Handgrip | 0,109726 | 0,062916 | NA |
| 3 | abundance | X07193 | N-(1-Methy | Tibia_length | -0,02535 | 0,053437 | NA |
| 3 | abundance | X07193 | N-(1-Methy | Radius_length | 0,007394 | 0,072298 | NA |
| 3 | abundance | X07193 | N-(1-Methy | Radius_SOS | -0,02202 | 0,051028 | NA |
| 3 | abundance | X07193 | N-(1-Methy | Tibia_SOS | -0,00385 | 0,054115 | NA |
| 3 | abundance | X07193 | N-(1-Methy | Handgrip | -0,08663 | 0,061651 | NA |
| 2b | abundance | X07201 | 2,4-Quinol | Tibia_length | 0,012319 | 0,053162 | NA |
| 2b | abundance | X07201 | 2,4-Quinol | Radius_length | -0,03458 | 0,072374 | NA |
| 2b | abundance | X07201 | 2,4-Quinol | Radius_SOS | 0,006306 | 0,050926 | NA |
| 2b | abundance | X07201 | 2,4-Quinol | Tibia_SOS | -0,02751 | 0,05431 | NA |
| 2b | abundance | X07201 | 2,4-Quinol | Handgrip | 0,07342 | 0,062223 | NA |
| 3 | abundance | X07216 | pentobarbi | Tibia_length | 0,074128 | 0,051686 | NA |
| 3 | abundance | X07216 | pentobarbi | Radius_length | 0,104118 | 0,070325 | NA |
| 3 | abundance | X07216 | pentobarbi | Radius_SOS | 0,01832 | 0,04984 | NA |

| | | | | | | | | |
|---|---|---|---|---|---|---|---|---|
| | 3 | abundance | X07216 | pentobarbi | Tibia_SOS | 0,034764 | 0,053311 | NA |
| | 3 | abundance | X07216 | pentobarbi | Handgrip | -0,1044 | 0,060999 | NA |
| 2b | | abundance | X07219 | 3,3-Dimetl | Tibia_length | 0,024001 | 0,054467 | NA |
| 2b | | abundance | X07219 | 3,3-Dimetl | Radius_length | -0,04157 | 0,073401 | NA |
| 2b | | abundance | X07219 | 3,3-Dimetl | Radius_SOS | -0,00463 | 0,052007 | NA |
| 2b | | abundance | X07219 | 3,3-Dimetl | Tibia_SOS | 7,53E-04 | 0,055928 | NA |
| 2b | | abundance | X07219 | 3,3-Dimetl | Handgrip | 0,141138 | 0,061746 | NA |
| | 3 | abundance | X07220 | Glycylproly | Tibia_length | -0,06881 | 0,052534 | NA |
| | 3 | abundance | X07220 | Glycylproly | Radius_length | -0,06508 | 0,071465 | NA |
| | 3 | abundance | X07220 | Glycylproly | Radius_SOS | -0,05053 | 0,050348 | NA |
| | 3 | abundance | X07220 | Glycylproly | Tibia_SOS | -0,04178 | 0,053994 | NA |
| | 3 | abundance | X07220 | Glycylproly | Handgrip | -0,09286 | 0,060541 | NA |
| | 3 | abundance | X07226 | threonylph | Tibia_length | -0,06391 | 0,054466 | NA |
| | 3 | abundance | X07226 | threonylph | Radius_length | -0,10517 | 0,072854 | NA |
| | 3 | abundance | X07226 | threonylph | Radius_SOS | -0,04593 | 0,051901 | NA |
| | 3 | abundance | X07226 | threonylph | Tibia_SOS | -0,03235 | 0,055616 | NA |
| | 3 | abundance | X07226 | threonylph | Handgrip | -0,17228 | 0,062061 | NA |
| | 3 | abundance | X07236 | 2-methylci | Tibia_length | -0,09624 | 0,053628 | NA |
| | 3 | abundance | X07236 | 2-methylci | Radius_length | -0,05269 | 0,072974 | NA |
| | 3 | abundance | X07236 | 2-methylci | Radius_SOS | 0,036082 | 0,051342 | NA |
| | 3 | abundance | X07236 | 2-methylci | Tibia_SOS | -0,00887 | 0,054608 | NA |
| | 3 | abundance | X07236 | 2-methylci | Handgrip | -0,04404 | 0,0612 | NA |
| | 3 | abundance | X07250 | Leucylaspa | Tibia_length | -3,34E-04 | 0,054511 | NA |
| | 3 | abundance | X07250 | Leucylaspa | Radius_length | 0,073979 | 0,073518 | NA |
| | 3 | abundance | X07250 | Leucylaspa | Radius_SOS | -0,00485 | 0,051865 | NA |
| | 3 | abundance | X07250 | Leucylaspa | Tibia_SOS | -0,01181 | 0,056172 | NA |
| | 3 | abundance | X07250 | Leucylaspa | Handgrip | -0,00689 | 0,062196 | NA |
| | 3 | abundance | X07260 | (7E,7'E)-5, | Tibia_length | -0,09821 | 0,056169 | NA |
| | 3 | abundance | X07260 | (7E,7'E)-5, | Radius_length | -0,0853 | 0,074181 | NA |
| | 3 | abundance | X07260 | (7E,7'E)-5, | Radius_SOS | -0,15478 | 0,053552 | NA |
| | 3 | abundance | X07260 | (7E,7'E)-5, | Tibia_SOS | -0,06095 | 0,057946 | NA |
| | 3 | abundance | X07260 | (7E,7'E)-5, | Handgrip | -0,11503 | 0,059793 | NA |
| 2b | | abundance | X07263 | 3,3-Dimetl | Tibia_length | 0,004582 | 0,055249 | NA |
| 2b | | abundance | X07263 | 3,3-Dimetl | Radius_length | 0,014736 | 0,073308 | NA |
| 2b | | abundance | X07263 | 3,3-Dimetl | Radius_SOS | 0,011365 | 0,052584 | NA |
| 2b | | abundance | X07263 | 3,3-Dimetl | Tibia_SOS | 0,033287 | 0,056536 | NA |
| 2b | | abundance | X07263 | 3,3-Dimetl | Handgrip | 0,136461 | 0,061024 | NA |
| | 3 | abundance | X07269 | (3aR,4R,5I | Tibia_length | 0,033666 | 0,050381 | NA |
| | 3 | abundance | X07269 | (3aR,4R,5I | Radius_length | -0,00308 | 0,068819 | NA |
| | 3 | abundance | X07269 | (3aR,4R,5I | Radius_SOS | 0,008457 | 0,049229 | NA |
| | 3 | abundance | X07269 | (3aR,4R,5I | Tibia_SOS | -0,011 | 0,052833 | NA |
| | 3 | abundance | X07269 | (3aR,4R,5I | Handgrip | -0,00759 | 0,061178 | NA |
| | 3 | abundance | X07270 | 5-(2-Carbo | Tibia_length | -0,00159 | 0,055706 | NA |
| | 3 | abundance | X07270 | 5-(2-Carbo | Radius_length | 0,057529 | 0,073787 | NA |
| | 3 | abundance | X07270 | 5-(2-Carbo | Radius_SOS | 0,148391 | 0,052624 | NA |
| | 3 | abundance | X07270 | 5-(2-Carbo | Tibia_SOS | 0,126037 | 0,056299 | NA |
| | 3 | abundance | X07270 | 5-(2-Carbo | Handgrip | 0,120646 | 0,062758 | NA |
| | 3 | abundance | X07278 | 5-Phospho | Tibia_length | -0,02866 | 0,051177 | NA |

| | | | | | | | | |
|---|---|---|---|---|---|---|---|---|
| | 3 | abundance | X07278 | 5-Phospho | Radius_length | -0,09963 | 0,06979 | NA |
| | 3 | abundance | X07278 | 5-Phospho | Radius_SOS | 0,070246 | 0,049395 | NA |
| | 3 | abundance | X07278 | 5-Phospho | Tibia_SOS | -0,05923 | 0,053042 | NA |
| | 3 | abundance | X07278 | 5-Phospho | Handgrip | -0,00151 | 0,061163 | NA |
| | 3 | abundance | X07283 | 8-Hydroxyl | Tibia_length | 0,060744 | 0,054079 | NA |
| | 3 | abundance | X07283 | 8-Hydroxyl | Radius_length | 0,055447 | 0,072865 | NA |
| | 3 | abundance | X07283 | 8-Hydroxyl | Radius_SOS | 0,021132 | 0,051621 | NA |
| | 3 | abundance | X07283 | 8-Hydroxyl | Tibia_SOS | -0,03535 | 0,054907 | NA |
| | 3 | abundance | X07283 | 8-Hydroxyl | Handgrip | -0,0526 | 0,061994 | NA |
| | 3 | abundance | X07285 | 2-(Carboxy | Tibia_length | -0,01644 | 0,058426 | NA |
| | 3 | abundance | X07285 | 2-(Carboxy | Radius_length | -0,03809 | 0,075218 | NA |
| | 3 | abundance | X07285 | 2-(Carboxy | Radius_SOS | -0,05306 | 0,055669 | NA |
| | 3 | abundance | X07285 | 2-(Carboxy | Tibia_SOS | 0,020046 | 0,059538 | NA |
| | 3 | abundance | X07285 | 2-(Carboxy | Handgrip | -0,07295 | 0,060033 | NA |
| | 3 | abundance | X07294 | 6-Acetami | Tibia_length | -0,13891 | 0,056614 | NA |
| | 3 | abundance | X07294 | 6-Acetami | Radius_length | -0,07191 | 0,074958 | NA |
| | 3 | abundance | X07294 | 6-Acetami | Radius_SOS | -0,0677 | 0,054176 | NA |
| | 3 | abundance | X07294 | 6-Acetami | Tibia_SOS | -0,01523 | 0,057384 | NA |
| | 3 | abundance | X07294 | 6-Acetami | Handgrip | -0,08632 | 0,062543 | NA |
| | 3 | abundance | X07303 | Triacetin | Tibia_length | -0,10421 | 0,053858 | NA |
| | 3 | abundance | X07303 | Triacetin | Radius_length | -0,1342 | 0,072507 | NA |
| | 3 | abundance | X07303 | Triacetin | Radius_SOS | -0,02548 | 0,051401 | NA |
| | 3 | abundance | X07303 | Triacetin | Tibia_SOS | -7,66E-05 | 0,054894 | NA |
| | 3 | abundance | X07303 | Triacetin | Handgrip | -0,09232 | 0,061381 | NA |
| 2a | | abundance | X07308 | Nicotinic a | Tibia_length | -0,07122 | 0,053639 | NA |
| 2a | | abundance | X07308 | Nicotinic a | Radius_length | -0,02484 | 0,073068 | NA |
| 2a | | abundance | X07308 | Nicotinic a | Radius_SOS | -0,13017 | 0,051365 | NA |
| 2a | | abundance | X07308 | Nicotinic a | Tibia_SOS | -0,13351 | 0,055965 | NA |
| 2a | | abundance | X07308 | Nicotinic a | Handgrip | -0,03495 | 0,062574 | NA |
| | 3 | abundance | X07309 | 3,4,15-Trih | Tibia_length | -0,04257 | 0,054518 | NA |
| | 3 | abundance | X07309 | 3,4,15-Trih | Radius_length | -0,04779 | 0,07292 | NA |
| | 3 | abundance | X07309 | 3,4,15-Trih | Radius_SOS | 0,02824 | 0,051999 | NA |
| | 3 | abundance | X07309 | 3,4,15-Trih | Tibia_SOS | -0,0067 | 0,055285 | NA |
| | 3 | abundance | X07309 | 3,4,15-Trih | Handgrip | -0,00228 | 0,062056 | NA |
| | 3 | abundance | X07315 | Rivastigmi | Tibia_length | -0,08064 | 0,052693 | NA |
| | 3 | abundance | X07315 | Rivastigmi | Radius_length | -0,11685 | 0,071424 | NA |
| | 3 | abundance | X07315 | Rivastigmi | Radius_SOS | -0,03098 | 0,050575 | NA |
| | 3 | abundance | X07315 | Rivastigmi | Tibia_SOS | -0,13228 | 0,054427 | NA |
| | 3 | abundance | X07315 | Rivastigmi | Handgrip | -0,01874 | 0,06189 | NA |
| | 3 | abundance | X07320 | asn-pro | Tibia_length | -0,16499 | 0,053347 | NA |
| | 3 | abundance | X07320 | asn-pro | Radius_length | -0,14969 | 0,071856 | NA |
| | 3 | abundance | X07320 | asn-pro | Radius_SOS | -0,04432 | 0,051119 | NA |
| | 3 | abundance | X07320 | asn-pro | Tibia_SOS | -0,09762 | 0,05417 | NA |
| | 3 | abundance | X07320 | asn-pro | Handgrip | -0,13636 | 0,060222 | NA |
| | 3 | abundance | X07327 | Aspartyl-L- | Tibia_length | -0,09795 | 0,056207 | NA |
| | 3 | abundance | X07327 | Aspartyl-L- | Radius_length | -0,07506 | 0,074316 | NA |
| | 3 | abundance | X07327 | Aspartyl-L- | Radius_SOS | 0,030902 | 0,053579 | NA |
| | 3 | abundance | X07327 | Aspartyl-L- | Tibia_SOS | -0,03313 | 0,05824 | NA |

| | | | | | | | | |
|---|---|---|---|---|---|---|---|---|
| | 3 | abundance | X07327 | Aspartyl-L- | Handgrip | -0,0027 | 0,06283 | NA |
| | 3 | abundance | X07334 | 2,2-Bis(hy | Tibia_length | -0,02963 | 0,055111 | NA |
| | 3 | abundance | X07334 | 2,2-Bis(hy | Radius_length | -0,05214 | 0,074267 | NA |
| | 3 | abundance | X07334 | 2,2-Bis(hy | Radius_SOS | -0,10606 | 0,052668 | NA |
| | 3 | abundance | X07334 | 2,2-Bis(hy | Tibia_SOS | -0,00682 | 0,056498 | NA |
| | 3 | abundance | X07334 | 2,2-Bis(hy | Handgrip | 0,133614 | 0,061762 | NA |
| | 3 | abundance | X07336 | Threonylse | Tibia_length | -0,04995 | 0,055804 | NA |
| | 3 | abundance | X07336 | Threonylse | Radius_length | 0,05304 | 0,074246 | NA |
| | 3 | abundance | X07336 | Threonylse | Radius_SOS | 0,012411 | 0,053052 | NA |
| | 3 | abundance | X07336 | Threonylse | Tibia_SOS | 0,002745 | 0,058112 | NA |
| | 3 | abundance | X07336 | Threonylse | Handgrip | 0,004495 | 0,063176 | NA |
| | 3 | abundance | X07344 | (âˆ’)-nabil | Tibia_length | -0,02115 | 0,053851 | NA |
| | 3 | abundance | X07344 | (âˆ’)-nabil | Radius_length | -0,04912 | 0,072744 | NA |
| | 3 | abundance | X07344 | (âˆ’)-nabil | Radius_SOS | 0,083069 | 0,051079 | NA |
| | 3 | abundance | X07344 | (âˆ’)-nabil | Tibia_SOS | 0,008266 | 0,054675 | NA |
| | 3 | abundance | X07344 | (âˆ’)-nabil | Handgrip | 0,06473 | 0,060861 | NA |
| | 3 | abundance | X07345 | Phenyl D-g | Tibia_length | -0,00572 | 0,055382 | NA |
| | 3 | abundance | X07345 | Phenyl D-g | Radius_length | -0,13536 | 0,072941 | NA |
| | 3 | abundance | X07345 | Phenyl D-g | Radius_SOS | -0,06256 | 0,052534 | NA |
| | 3 | abundance | X07345 | Phenyl D-g | Tibia_SOS | -0,05937 | 0,055919 | NA |
| | 3 | abundance | X07345 | Phenyl D-g | Handgrip | -0,0791 | 0,062799 | NA |
| | 3 | abundance | X07375 | porphobili | Tibia_length | -0,02991 | 0,050341 | NA |
| | 3 | abundance | X07375 | porphobili | Radius_length | -0,04556 | 0,068777 | NA |
| | 3 | abundance | X07375 | porphobili | Radius_SOS | -0,06814 | 0,049044 | NA |
| | 3 | abundance | X07375 | porphobili | Tibia_SOS | -0,01425 | 0,052658 | NA |
| | 3 | abundance | X07375 | porphobili | Handgrip | -0,08516 | 0,0601 | NA |
| | 3 | abundance | X07381 | Bis-D-fruct | Tibia_length | -0,10447 | 0,05486 | NA |
| | 3 | abundance | X07381 | Bis-D-fruct | Radius_length | -0,07034 | 0,07377 | NA |
| | 3 | abundance | X07381 | Bis-D-fruct | Radius_SOS | -0,05361 | 0,052291 | NA |
| | 3 | abundance | X07381 | Bis-D-fruct | Tibia_SOS | 0,066094 | 0,055991 | NA |
| | 3 | abundance | X07381 | Bis-D-fruct | Handgrip | -0,01883 | 0,062417 | NA |
| | 3 | abundance | X07394 | Hydroxypr | Tibia_length | 0,109409 | 0,056658 | NA |
| | 3 | abundance | X07394 | Hydroxypr | Radius_length | 0,216922 | 0,07316 | NA |
| | 3 | abundance | X07394 | Hydroxypr | Radius_SOS | 0,062831 | 0,053934 | NA |
| | 3 | abundance | X07394 | Hydroxypr | Tibia_SOS | 0,039003 | 0,059304 | NA |
| | 3 | abundance | X07394 | Hydroxypr | Handgrip | 0,177365 | 0,062821 | NA |
| | 3 | abundance | X07399 | Homoanse | Tibia_length | -0,03375 | 0,054761 | NA |
| | 3 | abundance | X07399 | Homoanse | Radius_length | -0,0021 | 0,073531 | NA |
| | 3 | abundance | X07399 | Homoanse | Radius_SOS | -0,09373 | 0,051837 | NA |
| | 3 | abundance | X07399 | Homoanse | Tibia_SOS | 0,059337 | 0,055391 | NA |
| | 3 | abundance | X07399 | Homoanse | Handgrip | -0,01181 | 0,062513 | NA |
| 2b | | abundance | X07400 | Tranexami | Tibia_length | 0,006156 | 0,049686 | NA |
| 2b | | abundance | X07400 | Tranexami | Radius_length | 0,108297 | 0,067363 | NA |
| 2b | | abundance | X07400 | Tranexami | Radius_SOS | -0,01192 | 0,049028 | NA |
| 2b | | abundance | X07400 | Tranexami | Tibia_SOS | 0,087307 | 0,052313 | NA |
| 2b | | abundance | X07400 | Tranexami | Handgrip | 0,079096 | 0,060196 | NA |
| | 1 | abundance | X07413 | Chenodeo: | Tibia_length | -0,02875 | 0,050302 | NA |
| | 1 | abundance | X07413 | Chenodeo: | Radius_length | 0,051037 | 0,06869 | NA |

| | | | | | | | | |
|---|---|---|---|---|---|---|---|---|
| | 1 | abundance | X07413 | Chenodeo: | Radius_SOS | -0,03698 | 0,049185 | NA |
| | 1 | abundance | X07413 | Chenodeo: | Tibia_SOS | -0,05206 | 0,052631 | NA |
| | 1 | abundance | X07413 | Chenodeo: | Handgrip | 0,012682 | 0,060809 | NA |
| | 3 | abundance | X07414 | 2,2-Bis(hy | Tibia_length | -0,03858 | 0,053783 | NA |
| | 3 | abundance | X07414 | 2,2-Bis(hy | Radius_length | 0,031799 | 0,073337 | NA |
| | 3 | abundance | X07414 | 2,2-Bis(hy | Radius_SOS | 0,013659 | 0,051182 | NA |
| | 3 | abundance | X07414 | 2,2-Bis(hy | Tibia_SOS | 0,013899 | 0,055892 | NA |
| | 3 | abundance | X07414 | 2,2-Bis(hy | Handgrip | 0,089777 | 0,06125 | NA |
| 2b | | abundance | X07417 | Î´-Valerola | Tibia_length | 0,027512 | 0,054467 | NA |
| 2b | | abundance | X07417 | Î´-Valerola | Radius_length | -0,00213 | 0,073212 | NA |
| 2b | | abundance | X07417 | Î´-Valerola | Radius_SOS | -0,03751 | 0,051933 | NA |
| 2b | | abundance | X07417 | Î´-Valerola | Tibia_SOS | -0,01274 | 0,054755 | NA |
| 2b | | abundance | X07417 | Î´-Valerola | Handgrip | 0,030278 | 0,061712 | NA |
| | 3 | abundance | X07420 | DNOP | Tibia_length | 0,057327 | 0,052776 | NA |
| | 3 | abundance | X07420 | DNOP | Radius_length | 0,079998 | 0,072035 | NA |
| | 3 | abundance | X07420 | DNOP | Radius_SOS | 0,009899 | 0,050448 | NA |
| | 3 | abundance | X07420 | DNOP | Tibia_SOS | 0,009909 | 0,053878 | NA |
| | 3 | abundance | X07420 | DNOP | Handgrip | 0,033313 | 0,061197 | NA |
| | 3 | abundance | X07424 | Rutinose (I | Tibia_length | 0,06221 | 0,057132 | NA |
| | 3 | abundance | X07424 | Rutinose (I | Radius_length | 0,13216 | 0,074508 | NA |
| | 3 | abundance | X07424 | Rutinose (I | Radius_SOS | 0,020259 | 0,054419 | NA |
| | 3 | abundance | X07424 | Rutinose (I | Tibia_SOS | 0,0644 | 0,057913 | NA |
| | 3 | abundance | X07424 | Rutinose (I | Handgrip | 0,0548 | 0,062901 | NA |
| 2a | | abundance | X07425 | N-Acetylas | Tibia_length | 0,164971 | 0,0526 | NA |
| 2a | | abundance | X07425 | N-Acetylas | Radius_length | 0,067226 | 0,072696 | NA |
| 2a | | abundance | X07425 | N-Acetylas | Radius_SOS | 0,033216 | 0,051014 | NA |
| 2a | | abundance | X07425 | N-Acetylas | Tibia_SOS | 0,098075 | 0,055218 | NA |
| 2a | | abundance | X07425 | N-Acetylas | Handgrip | -0,03629 | 0,061877 | NA |
| | 3 | abundance | X07428 | N6-METHY | Tibia_length | -0,02026 | 0,049214 | NA |
| | 3 | abundance | X07428 | N6-METHY | Radius_length | -0,03843 | 0,06548 | NA |
| | 3 | abundance | X07428 | N6-METHY | Radius_SOS | 0,019345 | 0,049118 | NA |
| | 3 | abundance | X07428 | N6-METHY | Tibia_SOS | -0,07451 | 0,05239 | NA |
| | 3 | abundance | X07428 | N6-METHY | Handgrip | -0,01044 | 0,060191 | NA |
| | 3 | abundance | X07429 | DNOP | Tibia_length | 0,019673 | 0,049922 | NA |
| | 3 | abundance | X07429 | DNOP | Radius_length | -0,01385 | 0,067886 | NA |
| | 3 | abundance | X07429 | DNOP | Radius_SOS | 0,044535 | 0,049021 | NA |
| | 3 | abundance | X07429 | DNOP | Tibia_SOS | -0,08315 | 0,052475 | NA |
| | 3 | abundance | X07429 | DNOP | Handgrip | 0,017707 | 0,061235 | NA |
| 2b | | abundance | X07434 | N'-Hydroxy | Tibia_length | -0,05275 | 0,052203 | NA |
| 2b | | abundance | X07434 | N'-Hydroxy | Radius_length | -0,12844 | 0,070667 | NA |
| 2b | | abundance | X07434 | N'-Hydroxy | Radius_SOS | -0,06424 | 0,050241 | NA |
| 2b | | abundance | X07434 | N'-Hydroxy | Tibia_SOS | 0,013957 | 0,053971 | NA |
| 2b | | abundance | X07434 | N'-Hydroxy | Handgrip | -0,04044 | 0,061284 | NA |
| | 1 | abundance | X07439 | 4-pyridoxic | Tibia_length | -0,02306 | 0,049463 | NA |
| | 1 | abundance | X07439 | 4-pyridoxic | Radius_length | 0,10742 | 0,066694 | NA |
| | 1 | abundance | X07439 | 4-pyridoxic | Radius_SOS | 0,04012 | 0,048991 | NA |
| | 1 | abundance | X07439 | 4-pyridoxic | Tibia_SOS | 0,076675 | 0,052353 | NA |
| | 1 | abundance | X07439 | 4-pyridoxic | Handgrip | 0,060765 | 0,060097 | NA |

| | | | | | | | |
|---|---|---|---|---|---|---|---|
| 3 | abundance | X07442 | coenzyme | Tibia_length | 0,021261 | 0,049326 | NA |
| 3 | abundance | X07442 | coenzyme | Radius_length | -0,01147 | 0,066216 | NA |
| 3 | abundance | X07442 | coenzyme | Radius_SOS | 0,009681 | 0,049059 | NA |
| 3 | abundance | X07442 | coenzyme | Tibia_SOS | -5,71E-04 | 0,052711 | NA |
| 3 | abundance | X07442 | coenzyme | Handgrip | 0,084291 | 0,060015 | NA |
| 3 | abundance | X07443 | Butylparab | Tibia_length | -0,01092 | 0,052098 | NA |
| 3 | abundance | X07443 | Butylparab | Radius_length | 0,016982 | 0,071145 | NA |
| 3 | abundance | X07443 | Butylparab | Radius_SOS | 0,071953 | 0,050183 | NA |
| 3 | abundance | X07443 | Butylparab | Tibia_SOS | -0,04233 | 0,053398 | NA |
| 3 | abundance | X07443 | Butylparab | Handgrip | -0,10195 | 0,060874 | NA |
| 3 | abundance | X07445 | 3-(Sulfooxy | Tibia_length | 0,007346 | 0,053667 | NA |
| 3 | abundance | X07445 | 3-(Sulfooxy | Radius_length | -0,16235 | 0,071541 | NA |
| 3 | abundance | X07445 | 3-(Sulfooxy | Radius_SOS | -0,08105 | 0,050872 | NA |
| 3 | abundance | X07445 | 3-(Sulfooxy | Tibia_SOS | -0,06188 | 0,055151 | NA |
| 3 | abundance | X07445 | 3-(Sulfooxy | Handgrip | -0,11909 | 0,06143 | NA |
| 3 | abundance | X07451 | Valylvaline | Tibia_length | 0,068765 | 0,052839 | NA |
| 3 | abundance | X07451 | Valylvaline | Radius_length | 0,085293 | 0,071974 | NA |
| 3 | abundance | X07451 | Valylvaline | Radius_SOS | 0,071869 | 0,050431 | NA |
| 3 | abundance | X07451 | Valylvaline | Tibia_SOS | 0,113124 | 0,053911 | NA |
| 3 | abundance | X07451 | Valylvaline | Handgrip | 0,040499 | 0,061041 | NA |
| 3 | abundance | X07452 | 4-Amino-1 | Tibia_length | -0,10505 | 0,0539 | NA |
| 3 | abundance | X07452 | 4-Amino-1 | Radius_length | -0,12597 | 0,072031 | NA |
| 3 | abundance | X07452 | 4-Amino-1 | Radius_SOS | -0,00753 | 0,051071 | NA |
| 3 | abundance | X07452 | 4-Amino-1 | Tibia_SOS | 0,02315 | 0,0553 | NA |
| 3 | abundance | X07452 | 4-Amino-1 | Handgrip | -0,07792 | 0,061803 | NA |
| 2b | abundance | X07454 | 2,4-Quinol | Tibia_length | -0,13414 | 0,051138 | NA |
| 2b | abundance | X07454 | 2,4-Quinol | Radius_length | -0,00302 | 0,071023 | NA |
| 2b | abundance | X07454 | 2,4-Quinol | Radius_SOS | -0,02422 | 0,04997 | NA |
| 2b | abundance | X07454 | 2,4-Quinol | Tibia_SOS | 0,100593 | 0,053046 | NA |
| 2b | abundance | X07454 | 2,4-Quinol | Handgrip | -0,05915 | 0,061272 | NA |
| 1 | abundance | X07457 | Glucosami | Tibia_length | -0,03176 | 0,056103 | NA |
| 1 | abundance | X07457 | Glucosami | Radius_length | 0,01984 | 0,074996 | NA |
| 1 | abundance | X07457 | Glucosami | Radius_SOS | -0,03791 | 0,053334 | NA |
| 1 | abundance | X07457 | Glucosami | Tibia_SOS | -0,02765 | 0,057629 | NA |
| 1 | abundance | X07457 | Glucosami | Handgrip | 0,093792 | 0,063536 | NA |
| 3 | abundance | X07460 | 1-pyrroline | Tibia_length | 0,06433 | 0,052463 | NA |
| 3 | abundance | X07460 | 1-pyrroline | Radius_length | 0,080005 | 0,071889 | NA |
| 3 | abundance | X07460 | 1-pyrroline | Radius_SOS | -0,00522 | 0,050291 | NA |
| 3 | abundance | X07460 | 1-pyrroline | Tibia_SOS | 0,144975 | 0,052954 | NA |
| 3 | abundance | X07460 | 1-pyrroline | Handgrip | 0,072097 | 0,060421 | NA |
| 3 | abundance | X07465 | TO012790 | Tibia_length | -0,05961 | 0,051863 | NA |
| 3 | abundance | X07465 | TO012790 | Radius_length | -0,09961 | 0,070328 | NA |
| 3 | abundance | X07465 | TO012790 | Radius_SOS | -0,07033 | 0,049845 | NA |
| 3 | abundance | X07465 | TO012790 | Tibia_SOS | -0,05583 | 0,05361 | NA |
| 3 | abundance | X07465 | TO012790 | Handgrip | -0,05997 | 0,061451 | NA |
| 3 | abundance | X07469 | 1,1'-[1,12- | Tibia_length | 0,02059 | 0,050088 | NA |
| 3 | abundance | X07469 | 1,1'-[1,12- | Radius_length | 0,06552 | 0,06823 | NA |
| 3 | abundance | X07469 | 1,1'-[1,12- | Radius_SOS | -0,00101 | 0,049114 | NA |

| | | | | | | | | |
|---|---|---|---|---|---|---|---|---|
| | 3 | abundance | X07469 | 1,1'-[1,12-I | Tibia_SOS | -0,07406 | 0,052454 | NA |
| | 3 | abundance | X07469 | 1,1'-[1,12-I | Handgrip | 0,049803 | 0,060684 | NA |
| | 3 | abundance | X07475 | 2'-Deoxy-5 | Tibia_length | -4,68E-04 | 0,053589 | NA |
| | 3 | abundance | X07475 | 2'-Deoxy-5 | Radius_length | -0,01699 | 0,072445 | NA |
| | 3 | abundance | X07475 | 2'-Deoxy-5 | Radius_SOS | 0,045056 | 0,051176 | NA |
| | 3 | abundance | X07475 | 2'-Deoxy-5 | Tibia_SOS | -0,00836 | 0,054775 | NA |
| | 3 | abundance | X07475 | 2'-Deoxy-5 | Handgrip | 0,022658 | 0,062278 | NA |
| | 1 | abundance | X07476 | 4-Hydroxyl | Tibia_length | 0,183258 | 0,052349 | NA |
| | 1 | abundance | X07476 | 4-Hydroxyl | Radius_length | 0,139486 | 0,071645 | NA |
| | 1 | abundance | X07476 | 4-Hydroxyl | Radius_SOS | 0,018813 | 0,050741 | NA |
| | 1 | abundance | X07476 | 4-Hydroxyl | Tibia_SOS | 0,126187 | 0,053634 | NA |
| | 1 | abundance | X07476 | 4-Hydroxyl | Handgrip | 0,093218 | 0,060526 | NA |
| 2a | | abundance | X07480 | Naringenin | Tibia_length | -0,01142 | 0,055726 | NA |
| 2a | | abundance | X07480 | Naringenin | Radius_length | 0,045111 | 0,074188 | NA |
| 2a | | abundance | X07480 | Naringenin | Radius_SOS | -0,05585 | 0,052786 | NA |
| 2a | | abundance | X07480 | Naringenin | Tibia_SOS | -0,03558 | 0,056421 | NA |
| 2a | | abundance | X07480 | Naringenin | Handgrip | 0,046752 | 0,061681 | NA |
| | 3 | abundance | X07484 | MFCD0002 | Tibia_length | 0,013285 | 0,05087 | NA |
| | 3 | abundance | X07484 | MFCD0002 | Radius_length | -0,07857 | 0,069269 | NA |
| | 3 | abundance | X07484 | MFCD0002 | Radius_SOS | -0,02073 | 0,049375 | NA |
| | 3 | abundance | X07484 | MFCD0002 | Tibia_SOS | 0,049416 | 0,052748 | NA |
| | 3 | abundance | X07484 | MFCD0002 | Handgrip | -0,02123 | 0,060325 | NA |
| | 3 | abundance | X07487 | Oxypeucec | Tibia_length | 0,001246 | 0,05503 | NA |
| | 3 | abundance | X07487 | Oxypeucec | Radius_length | 0,004789 | 0,073591 | NA |
| | 3 | abundance | X07487 | Oxypeucec | Radius_SOS | -0,04462 | 0,05236 | NA |
| | 3 | abundance | X07487 | Oxypeucec | Tibia_SOS | 0,016047 | 0,055442 | NA |
| | 3 | abundance | X07487 | Oxypeucec | Handgrip | 0,032968 | 0,061837 | NA |
| 2b | | abundance | X07491 | 1-Vinylimic | Tibia_length | -0,09577 | 0,050267 | NA |
| 2b | | abundance | X07491 | 1-Vinylimic | Radius_length | -0,13613 | 0,068523 | NA |
| 2b | | abundance | X07491 | 1-Vinylimic | Radius_SOS | -0,04581 | 0,04914 | NA |
| 2b | | abundance | X07491 | 1-Vinylimic | Tibia_SOS | -0,08339 | 0,052499 | NA |
| 2b | | abundance | X07491 | 1-Vinylimic | Handgrip | 0,007301 | 0,060623 | NA |
| 2b | | abundance | X07504 | 4-Acetamic | Tibia_length | 0,03474 | 0,0506 | NA |
| 2b | | abundance | X07504 | 4-Acetamic | Radius_length | 0,056238 | 0,069626 | NA |
| 2b | | abundance | X07504 | 4-Acetamic | Radius_SOS | 0,042019 | 0,049259 | NA |
| 2b | | abundance | X07504 | 4-Acetamic | Tibia_SOS | 0,110632 | 0,052734 | NA |
| 2b | | abundance | X07504 | 4-Acetamic | Handgrip | 0,031441 | 0,061011 | NA |
| 2b | | abundance | X07508 | Procaine | Tibia_length | -0,12238 | 0,051995 | NA |
| 2b | | abundance | X07508 | Procaine | Radius_length | -0,08601 | 0,070867 | NA |
| 2b | | abundance | X07508 | Procaine | Radius_SOS | 0,035632 | 0,050102 | NA |
| 2b | | abundance | X07508 | Procaine | Tibia_SOS | -0,03999 | 0,053981 | NA |
| 2b | | abundance | X07508 | Procaine | Handgrip | 0,006215 | 0,061378 | NA |
| | 3 | abundance | X07511 | Piperidine | Tibia_length | -0,05128 | 0,049841 | NA |
| | 3 | abundance | X07511 | Piperidine | Radius_length | -0,0322 | 0,06781 | NA |
| | 3 | abundance | X07511 | Piperidine | Radius_SOS | 0,008508 | 0,049065 | NA |
| | 3 | abundance | X07511 | Piperidine | Tibia_SOS | 0,044032 | 0,052543 | NA |
| | 3 | abundance | X07511 | Piperidine | Handgrip | -0,00571 | 0,060269 | NA |
| | 3 | abundance | X07512 | 4-{[(3-Hydr | Tibia_length | -0,10488 | 0,055778 | NA |

| | | | | | | | | |
|---|---|---|---|---|---|---|---|---|
| | 3 | abundance | X07512 | 4-{[(3-Hydr | Radius_length | 0,06503 | 0,074429 | NA |
| | 3 | abundance | X07512 | 4-{[(3-Hydr | Radius_SOS | 0,018602 | 0,053086 | NA |
| | 3 | abundance | X07512 | 4-{[(3-Hydr | Tibia_SOS | 0,101849 | 0,057316 | NA |
| | 3 | abundance | X07512 | 4-{[(3-Hydr | Handgrip | 0,047789 | 0,06314 | NA |
| | 3 | abundance | X07513 | (âˆ')-nabil | Tibia_length | 0,009467 | 0,051798 | NA |
| | 3 | abundance | X07513 | (âˆ')-nabil | Radius_length | 0,082794 | 0,070732 | NA |
| | 3 | abundance | X07513 | (âˆ')-nabil | Radius_SOS | 0,032348 | 0,049891 | NA |
| | 3 | abundance | X07513 | (âˆ')-nabil | Tibia_SOS | -0,06681 | 0,053567 | NA |
| | 3 | abundance | X07513 | (âˆ')-nabil | Handgrip | -0,0237 | 0,061387 | NA |
| | 3 | abundance | X07514 | L-(+)-Eryth | Tibia_length | -0,03831 | 0,055745 | NA |
| | 3 | abundance | X07514 | L-(+)-Eryth | Radius_length | 0,019335 | 0,074647 | NA |
| | 3 | abundance | X07514 | L-(+)-Eryth | Radius_SOS | -0,03769 | 0,053031 | NA |
| | 3 | abundance | X07514 | L-(+)-Eryth | Tibia_SOS | 0,004369 | 0,056982 | NA |
| | 3 | abundance | X07514 | L-(+)-Eryth | Handgrip | 0,099953 | 0,06309 | NA |
| | 3 | abundance | X07518 | (+/-)-2-Hyc | Tibia_length | -0,09824 | 0,053713 | NA |
| | 3 | abundance | X07518 | (+/-)-2-Hyc | Radius_length | -0,0266 | 0,073408 | NA |
| | 3 | abundance | X07518 | (+/-)-2-Hyc | Radius_SOS | -0,01623 | 0,051242 | NA |
| | 3 | abundance | X07518 | (+/-)-2-Hyc | Tibia_SOS | 0,032752 | 0,054626 | NA |
| | 3 | abundance | X07518 | (+/-)-2-Hyc | Handgrip | -0,02391 | 0,061791 | NA |
| | 3 | abundance | X07519 | Pyrrolidine | Tibia_length | -0,12554 | 0,051539 | NA |
| | 3 | abundance | X07519 | Pyrrolidine | Radius_length | -0,1061 | 0,070376 | NA |
| | 3 | abundance | X07519 | Pyrrolidine | Radius_SOS | 0,010469 | 0,049788 | NA |
| | 3 | abundance | X07519 | Pyrrolidine | Tibia_SOS | 0,03124 | 0,053453 | NA |
| | 3 | abundance | X07519 | Pyrrolidine | Handgrip | 0,012938 | 0,061032 | NA |
| | 1 | abundance | X07523 | Ursodeoxy | Tibia_length | 0,030724 | 0,049207 | NA |
| | 1 | abundance | X07523 | Ursodeoxy | Radius_length | 0,060665 | 0,065524 | NA |
| | 1 | abundance | X07523 | Ursodeoxy | Radius_SOS | 0,017946 | 0,049096 | NA |
| | 1 | abundance | X07523 | Ursodeoxy | Tibia_SOS | -0,00882 | 0,052554 | NA |
| | 1 | abundance | X07523 | Ursodeoxy | Handgrip | 0,025276 | 0,060305 | NA |
| 2b | | abundance | X07524 | Prolinamid | Tibia_length | -0,06045 | 0,051613 | NA |
| 2b | | abundance | X07524 | Prolinamid | Radius_length | -0,01289 | 0,070786 | NA |
| 2b | | abundance | X07524 | Prolinamid | Radius_SOS | 0,018226 | 0,04972 | NA |
| 2b | | abundance | X07524 | Prolinamid | Tibia_SOS | 0,017635 | 0,052984 | NA |
| 2b | | abundance | X07524 | Prolinamid | Handgrip | 0,032048 | 0,060625 | NA |
| 2b | | abundance | X07527 | 1,7-Dimetl | Tibia_length | -0,07081 | 0,051452 | NA |
| 2b | | abundance | X07527 | 1,7-Dimetl | Radius_length | -0,11553 | 0,070108 | NA |
| 2b | | abundance | X07527 | 1,7-Dimetl | Radius_SOS | 0,007157 | 0,049766 | NA |
| 2b | | abundance | X07527 | 1,7-Dimetl | Tibia_SOS | -0,06318 | 0,052851 | NA |
| 2b | | abundance | X07527 | 1,7-Dimetl | Handgrip | -0,02104 | 0,061139 | NA |
| | 1 | abundance | X07530 | Alanine | Tibia_length | 0,143892 | 0,053473 | NA |
| | 1 | abundance | X07530 | Alanine | Radius_length | 0,184118 | 0,07201 | NA |
| | 1 | abundance | X07530 | Alanine | Radius_SOS | 0,032143 | 0,05138 | NA |
| | 1 | abundance | X07530 | Alanine | Tibia_SOS | 0,145468 | 0,054344 | NA |
| | 1 | abundance | X07530 | Alanine | Handgrip | 0,166331 | 0,059862 | NA |
| | 3 | abundance | X07537 | alpha-Cha | Tibia_length | 0,026371 | 0,05629 | NA |
| | 3 | abundance | X07537 | alpha-Cha | Radius_length | 0,210586 | 0,072524 | NA |
| | 3 | abundance | X07537 | alpha-Cha | Radius_SOS | -6,38E-04 | 0,05328 | NA |
| | 3 | abundance | X07537 | alpha-Cha | Tibia_SOS | 0,017049 | 0,05593 | NA |

| | | | | | | | | |
|---|---|---|---|---|---|---|---|---|
| | 3 | abundance | X07537 | alpha-Cha | Handgrip | 0,100769 | 0,062753 | NA |
| | 3 | abundance | X07538 | Homoanse | Tibia_length | 0,014205 | 0,049723 | NA |
| | 3 | abundance | X07538 | Homoanse | Radius_length | 0,160203 | 0,066597 | NA |
| | 3 | abundance | X07538 | Homoanse | Radius_SOS | 0,048674 | 0,048985 | NA |
| | 3 | abundance | X07538 | Homoanse | Tibia_SOS | 0,004652 | 0,052488 | NA |
| | 3 | abundance | X07538 | Homoanse | Handgrip | 0,048039 | 0,060754 | NA |
| | 1 | abundance | X07541 | Saccharin | Tibia_length | -0,01314 | 0,054871 | NA |
| | 1 | abundance | X07541 | Saccharin | Radius_length | 0,119113 | 0,072913 | NA |
| | 1 | abundance | X07541 | Saccharin | Radius_SOS | 0,010357 | 0,052286 | NA |
| | 1 | abundance | X07541 | Saccharin | Tibia_SOS | 0,061427 | 0,055635 | NA |
| | 1 | abundance | X07541 | Saccharin | Handgrip | 0,057012 | 0,061181 | NA |
| | 3 | abundance | X07544 | Ethyl sulfat | Tibia_length | -0,0172 | 0,054959 | NA |
| | 3 | abundance | X07544 | Ethyl sulfat | Radius_length | -0,0587 | 0,073363 | NA |
| | 3 | abundance | X07544 | Ethyl sulfat | Radius_SOS | -0,02788 | 0,052368 | NA |
| | 3 | abundance | X07544 | Ethyl sulfat | Tibia_SOS | 0,049725 | 0,055571 | NA |
| | 3 | abundance | X07544 | Ethyl sulfat | Handgrip | 0,063832 | 0,062363 | NA |
| 2b | | abundance | X07558 | Leucylproli | Tibia_length | -0,03427 | 0,051824 | NA |
| 2b | | abundance | X07558 | Leucylproli | Radius_length | 0,017519 | 0,071082 | NA |
| 2b | | abundance | X07558 | Leucylproli | Radius_SOS | -0,01007 | 0,049968 | NA |
| 2b | | abundance | X07558 | Leucylproli | Tibia_SOS | 0,063969 | 0,053955 | NA |
| 2b | | abundance | X07558 | Leucylproli | Handgrip | 0,05206 | 0,06038 | NA |
| | 1 | abundance | X07563 | Xanthine | Tibia_length | 0,071987 | 0,051585 | NA |
| | 1 | abundance | X07563 | Xanthine | Radius_length | 0,106026 | 0,071412 | NA |
| | 1 | abundance | X07563 | Xanthine | Radius_SOS | 0,023315 | 0,049821 | NA |
| | 1 | abundance | X07563 | Xanthine | Tibia_SOS | 0,138707 | 0,052851 | NA |
| | 1 | abundance | X07563 | Xanthine | Handgrip | 0,131496 | 0,059798 | NA |
| 2b | | abundance | X07565 | 5-Hydroxyi | Tibia_length | -0,03121 | 0,049103 | NA |
| 2b | | abundance | X07565 | 5-Hydroxyi | Radius_length | -0,00647 | 0,065351 | NA |
| 2b | | abundance | X07565 | 5-Hydroxyi | Radius_SOS | -0,01395 | 0,049184 | NA |
| 2b | | abundance | X07565 | 5-Hydroxyi | Tibia_SOS | 0,059275 | 0,052438 | NA |
| 2b | | abundance | X07565 | 5-Hydroxyi | Handgrip | -0,00956 | 0,06037 | NA |
| | 3 | abundance | X07566 | 5-Allyl-5-se | Tibia_length | -0,12387 | 0,055157 | NA |
| | 3 | abundance | X07566 | 5-Allyl-5-se | Radius_length | -0,19045 | 0,072298 | NA |
| | 3 | abundance | X07566 | 5-Allyl-5-se | Radius_SOS | 0,030126 | 0,052787 | NA |
| | 3 | abundance | X07566 | 5-Allyl-5-se | Tibia_SOS | -0,00865 | 0,055103 | NA |
| | 3 | abundance | X07566 | 5-Allyl-5-se | Handgrip | -0,06986 | 0,062441 | NA |
| | 3 | abundance | X07572 | 8-(3-Furyl)- | Tibia_length | -0,02294 | 0,056769 | NA |
| | 3 | abundance | X07572 | 8-(3-Furyl)- | Radius_length | 0,062042 | 0,074688 | NA |
| | 3 | abundance | X07572 | 8-(3-Furyl)- | Radius_SOS | -0,00328 | 0,053926 | NA |
| | 3 | abundance | X07572 | 8-(3-Furyl)- | Tibia_SOS | 0,050518 | 0,057465 | NA |
| | 3 | abundance | X07572 | 8-(3-Furyl)- | Handgrip | 0,053135 | 0,063524 | NA |
| | 3 | abundance | X07574 | 7-ketodeox | Tibia_length | 0,035202 | 0,050924 | NA |
| | 3 | abundance | X07574 | 7-ketodeox | Radius_length | 0,030711 | 0,069549 | NA |
| | 3 | abundance | X07574 | 7-ketodeox | Radius_SOS | 0,044063 | 0,049357 | NA |
| | 3 | abundance | X07574 | 7-ketodeox | Tibia_SOS | -0,04687 | 0,05333 | NA |
| | 3 | abundance | X07574 | 7-ketodeox | Handgrip | -0,04052 | 0,061247 | NA |
| | 3 | abundance | X07577 | 2-[(Sulfoox | Tibia_length | -0,06084 | 0,054836 | NA |
| | 3 | abundance | X07577 | 2-[(Sulfoox | Radius_length | -0,19313 | 0,071953 | NA |

| | | | | | | | |
|---|---|---|---|---|---|---|---|
| 3 | abundance | X07577 | 2-[(Sulfoox | Radius_SOS | -0,05653 | 0,051854 | NA |
| 3 | abundance | X07577 | 2-[(Sulfoox | Tibia_SOS | -0,07707 | 0,056116 | NA |
| 3 | abundance | X07577 | 2-[(Sulfoox | Handgrip | -0,13484 | 0,062371 | NA |
| 3 | abundance | X07578 | 5-Allyl-5-se | Tibia_length | -0,15716 | 0,053919 | NA |
| 3 | abundance | X07578 | 5-Allyl-5-se | Radius_length | -0,0692 | 0,072856 | NA |
| 3 | abundance | X07578 | 5-Allyl-5-se | Radius_SOS | 0,023987 | 0,051651 | NA |
| 3 | abundance | X07578 | 5-Allyl-5-se | Tibia_SOS | -0,07424 | 0,054634 | NA |
| 3 | abundance | X07578 | 5-Allyl-5-se | Handgrip | 0,010047 | 0,061869 | NA |
| 3 | abundance | X07580 | N-Pentano | Tibia_length | 0,099508 | 0,053614 | NA |
| 3 | abundance | X07580 | N-Pentano | Radius_length | 0,100006 | 0,072693 | NA |
| 3 | abundance | X07580 | N-Pentano | Radius_SOS | 0,06809 | 0,051473 | NA |
| 3 | abundance | X07580 | N-Pentano | Tibia_SOS | 0,037046 | 0,055154 | NA |
| 3 | abundance | X07580 | N-Pentano | Handgrip | -0,04805 | 0,06239 | NA |
| 3 | abundance | X07581 | N-Acetylva | Tibia_length | -0,0075 | 0,054011 | NA |
| 3 | abundance | X07581 | N-Acetylva | Radius_length | 0,014107 | 0,072885 | NA |
| 3 | abundance | X07581 | N-Acetylva | Radius_SOS | -0,02851 | 0,051404 | NA |
| 3 | abundance | X07581 | N-Acetylva | Tibia_SOS | -8,29E-04 | 0,055257 | NA |
| 3 | abundance | X07581 | N-Acetylva | Handgrip | 0,044225 | 0,062285 | NA |
| 2b | abundance | X07582 | 4-methylpy | Tibia_length | -0,1037 | 0,055169 | NA |
| 2b | abundance | X07582 | 4-methylpy | Radius_length | -0,05071 | 0,073782 | NA |
| 2b | abundance | X07582 | 4-methylpy | Radius_SOS | 0,108528 | 0,052087 | NA |
| 2b | abundance | X07582 | 4-methylpy | Tibia_SOS | 0,036407 | 0,054558 | NA |
| 2b | abundance | X07582 | 4-methylpy | Handgrip | 0,066724 | 0,061572 | NA |
| 2b | abundance | X07596 | 5-amino-2- | Tibia_length | 0,017124 | 0,051352 | NA |
| 2b | abundance | X07596 | 5-amino-2- | Radius_length | 0,076829 | 0,070271 | NA |
| 2b | abundance | X07596 | 5-amino-2- | Radius_SOS | -0,03027 | 0,049605 | NA |
| 2b | abundance | X07596 | 5-amino-2- | Tibia_SOS | 0,045954 | 0,053464 | NA |
| 2b | abundance | X07596 | 5-amino-2- | Handgrip | 0,014391 | 0,060605 | NA |
| 3 | abundance | X07597 | p-Cresylsu | Tibia_length | -0,00953 | 0,054926 | NA |
| 3 | abundance | X07597 | p-Cresylsu | Radius_length | -0,04657 | 0,073468 | NA |
| 3 | abundance | X07597 | p-Cresylsu | Radius_SOS | 0,001611 | 0,052292 | NA |
| 3 | abundance | X07597 | p-Cresylsu | Tibia_SOS | 0,018762 | 0,056116 | NA |
| 3 | abundance | X07597 | p-Cresylsu | Handgrip | -0,20319 | 0,06105 | NA |
| 3 | abundance | X07601 | Asp-lys | Tibia_length | -0,03641 | 0,051775 | NA |
| 3 | abundance | X07601 | Asp-lys | Radius_length | 0,00275 | 0,071087 | NA |
| 3 | abundance | X07601 | Asp-lys | Radius_SOS | 0,047715 | 0,04968 | NA |
| 3 | abundance | X07601 | Asp-lys | Tibia_SOS | 0,1437 | 0,052482 | NA |
| 3 | abundance | X07601 | Asp-lys | Handgrip | -0,05065 | 0,061229 | NA |
| 3 | abundance | X07613 | 1-(4-Aminc | Tibia_length | -0,08107 | 0,052174 | NA |
| 3 | abundance | X07613 | 1-(4-Aminc | Radius_length | -0,03026 | 0,071169 | NA |
| 3 | abundance | X07613 | 1-(4-Aminc | Radius_SOS | 0,003101 | 0,050062 | NA |
| 3 | abundance | X07613 | 1-(4-Aminc | Tibia_SOS | -0,00915 | 0,053345 | NA |
| 3 | abundance | X07613 | 1-(4-Aminc | Handgrip | 0,041526 | 0,061067 | NA |
| 3 | abundance | X07618 | N-(3-aceta | Tibia_length | 0,042082 | 0,049628 | NA |
| 3 | abundance | X07618 | N-(3-aceta | Radius_length | 0,123032 | 0,067112 | NA |
| 3 | abundance | X07618 | N-(3-aceta | Radius_SOS | 0,012801 | 0,049026 | NA |
| 3 | abundance | X07618 | N-(3-aceta | Tibia_SOS | 0,07934 | 0,052429 | NA |
| 3 | abundance | X07618 | N-(3-aceta | Handgrip | 0,100117 | 0,060267 | NA |

| | | | | | | | | |
|---|---|---|---|---|---|---|---|---|
| | 1 | abundance | X07630 | Methionine | Tibia_length | 0,049281 | 0,056188 | NA |
| | 1 | abundance | X07630 | Methionine | Radius_length | 0,090601 | 0,073718 | NA |
| | 1 | abundance | X07630 | Methionine | Radius_SOS | -0,02008 | 0,053401 | NA |
| | 1 | abundance | X07630 | Methionine | Tibia_SOS | 0,101076 | 0,055453 | NA |
| | 1 | abundance | X07630 | Methionine | Handgrip | 0,076788 | 0,061668 | NA |
| 2b | | abundance | X07634 | 3,8,9-trihy | Tibia_length | -0,02141 | 0,051023 | NA |
| 2b | | abundance | X07634 | 3,8,9-trihy | Radius_length | 0,036262 | 0,069883 | NA |
| 2b | | abundance | X07634 | 3,8,9-trihy | Radius_SOS | 0,055101 | 0,049494 | NA |
| 2b | | abundance | X07634 | 3,8,9-trihy | Tibia_SOS | -0,00641 | 0,052866 | NA |
| 2b | | abundance | X07634 | 3,8,9-trihy | Handgrip | -0,03531 | 0,060787 | NA |
| | 3 | abundance | X07638 | 7-ketodeo | Tibia_length | 0,073864 | 0,050429 | NA |
| | 3 | abundance | X07638 | 7-ketodeo | Radius_length | 0,099891 | 0,068602 | NA |
| | 3 | abundance | X07638 | 7-ketodeo | Radius_SOS | 0,017825 | 0,049213 | NA |
| | 3 | abundance | X07638 | 7-ketodeo | Tibia_SOS | -0,04804 | 0,052771 | NA |
| | 3 | abundance | X07638 | 7-ketodeo | Handgrip | 0,00999 | 0,060632 | NA |
| | 3 | abundance | X07645 | O-propeno | Tibia_length | -0,02676 | 0,058818 | NA |
| | 3 | abundance | X07645 | O-propeno | Radius_length | 0,109392 | 0,075328 | NA |
| | 3 | abundance | X07645 | O-propeno | Radius_SOS | 0,077666 | 0,056191 | NA |
| | 3 | abundance | X07645 | O-propeno | Tibia_SOS | 0,006664 | 0,057979 | NA |
| | 3 | abundance | X07645 | O-propeno | Handgrip | -0,09383 | 0,062997 | NA |
| | 3 | abundance | X07646 | (2Z)-2-({[(6 | Tibia_length | -0,05021 | 0,05902 | NA |
| | 3 | abundance | X07646 | (2Z)-2-({[(6 | Radius_length | 0,033899 | 0,075926 | NA |
| | 3 | abundance | X07646 | (2Z)-2-({[(6 | Radius_SOS | 0,028459 | 0,056215 | NA |
| | 3 | abundance | X07646 | (2Z)-2-({[(6 | Tibia_SOS | 0,142858 | 0,058733 | NA |
| | 3 | abundance | X07646 | (2Z)-2-({[(6 | Handgrip | 0,050734 | 0,060115 | NA |
| | 1 | abundance | X07647 | Genistein | Tibia_length | -0,02173 | 0,052035 | NA |
| | 1 | abundance | X07647 | Genistein | Radius_length | -0,01071 | 0,071285 | NA |
| | 1 | abundance | X07647 | Genistein | Radius_SOS | 0,004726 | 0,050063 | NA |
| | 1 | abundance | X07647 | Genistein | Tibia_SOS | 0,071723 | 0,054428 | NA |
| | 1 | abundance | X07647 | Genistein | Handgrip | 0,003108 | 0,062105 | NA |
| | 3 | abundance | X07648 | NL851300 | Tibia_length | 0,098866 | 0,049884 | NA |
| | 3 | abundance | X07648 | NL851300 | Radius_length | 0,1343 | 0,067735 | NA |
| | 3 | abundance | X07648 | NL851300 | Radius_SOS | 0,055516 | 0,049044 | NA |
| | 3 | abundance | X07648 | NL851300 | Tibia_SOS | 0,088653 | 0,052377 | NA |
| | 3 | abundance | X07648 | NL851300 | Handgrip | 0,0383 | 0,060297 | NA |
| 2b | | abundance | X07657 | Docosahe | Tibia_length | 0,023293 | 0,051566 | NA |
| 2b | | abundance | X07657 | Docosahe | Radius_length | -0,03085 | 0,070911 | NA |
| 2b | | abundance | X07657 | Docosahe | Radius_SOS | 0,103904 | 0,049609 | NA |
| 2b | | abundance | X07657 | Docosahe | Tibia_SOS | -0,00716 | 0,05362 | NA |
| 2b | | abundance | X07657 | Docosahe | Handgrip | 0,053012 | 0,060937 | NA |
| | 1 | abundance | X07658 | Î´-Glucono | Tibia_length | -0,03725 | 0,056502 | NA |
| | 1 | abundance | X07658 | Î´-Glucono | Radius_length | -0,00848 | 0,075058 | NA |
| | 1 | abundance | X07658 | Î´-Glucono | Radius_SOS | 0,021002 | 0,05368 | NA |
| | 1 | abundance | X07658 | Î´-Glucono | Tibia_SOS | 0,019914 | 0,058023 | NA |
| | 1 | abundance | X07658 | Î´-Glucono | Handgrip | 0,053244 | 0,062783 | NA |
| | 1 | abundance | X07674 | Lysine | Tibia_length | 0,144023 | 0,049801 | NA |
| | 1 | abundance | X07674 | Lysine | Radius_length | 0,208921 | 0,066996 | NA |
| | 1 | abundance | X07674 | Lysine | Radius_SOS | 0,035407 | 0,04906 | NA |

| | | | | | | | | |
|---|---|---|---|---|---|---|---|---|
| | 1 | abundance | X07674 | Lysine | Tibia_SOS | 0,141824 | 0,052233 | NA |
| | 1 | abundance | X07674 | Lysine | Handgrip | 0,089096 | 0,060446 | NA |
| 2b | | abundance | X07678 | (2E)-3-(3,4 | Tibia_length | -0,16359 | 0,054599 | NA |
| 2b | | abundance | X07678 | (2E)-3-(3,4 | Radius_length | -0,14041 | 0,072891 | NA |
| 2b | | abundance | X07678 | (2E)-3-(3,4 | Radius_SOS | -0,04674 | 0,05264 | NA |
| 2b | | abundance | X07678 | (2E)-3-(3,4 | Tibia_SOS | -0,12755 | 0,05485 | NA |
| 2b | | abundance | X07678 | (2E)-3-(3,4 | Handgrip | -0,07404 | 0,062075 | NA |
| | 1 | abundance | X07680 | Indole-3-a | Tibia_length | 0,029684 | 0,053994 | NA |
| | 1 | abundance | X07680 | Indole-3-a | Radius_length | -0,09566 | 0,072534 | NA |
| | 1 | abundance | X07680 | Indole-3-a | Radius_SOS | -0,15769 | 0,050784 | NA |
| | 1 | abundance | X07680 | Indole-3-a | Tibia_SOS | 0,064672 | 0,0544 | NA |
| | 1 | abundance | X07680 | Indole-3-a | Handgrip | -0,0112 | 0,061873 | NA |
| | 3 | abundance | X07694 | DNOP | Tibia_length | 0,027987 | 0,052985 | NA |
| | 3 | abundance | X07694 | DNOP | Radius_length | -0,06337 | 0,071656 | NA |
| | 3 | abundance | X07694 | DNOP | Radius_SOS | 0,097625 | 0,050277 | NA |
| | 3 | abundance | X07694 | DNOP | Tibia_SOS | -0,0333 | 0,055035 | NA |
| | 3 | abundance | X07694 | DNOP | Handgrip | 0,042618 | 0,061444 | NA |
| | 3 | abundance | X07699 | N-(1-Methy | Tibia_length | -0,0735 | 0,054815 | NA |
| | 3 | abundance | X07699 | N-(1-Methy | Radius_length | -0,05673 | 0,07339 | NA |
| | 3 | abundance | X07699 | N-(1-Methy | Radius_SOS | -0,00409 | 0,052279 | NA |
| | 3 | abundance | X07699 | N-(1-Methy | Tibia_SOS | 0,020856 | 0,054751 | NA |
| | 3 | abundance | X07699 | N-(1-Methy | Handgrip | -0,07158 | 0,061763 | NA |
| | 3 | abundance | X07701 | 3-Benzyl-6 | Tibia_length | -0,10104 | 0,054205 | NA |
| | 3 | abundance | X07701 | 3-Benzyl-6 | Radius_length | -0,07226 | 0,072799 | NA |
| | 3 | abundance | X07701 | 3-Benzyl-6 | Radius_SOS | 0,006438 | 0,051632 | NA |
| | 3 | abundance | X07701 | 3-Benzyl-6 | Tibia_SOS | 0,058094 | 0,055211 | NA |
| | 3 | abundance | X07701 | 3-Benzyl-6 | Handgrip | 0,09431 | 0,061885 | NA |
| | 3 | abundance | X07710 | 3-O-beta-D | Tibia_length | 0,0578 | 0,053809 | NA |
| | 3 | abundance | X07710 | 3-O-beta-D | Radius_length | 0,065841 | 0,073246 | NA |
| | 3 | abundance | X07710 | 3-O-beta-D | Radius_SOS | 0,002371 | 0,051415 | NA |
| | 3 | abundance | X07710 | 3-O-beta-D | Tibia_SOS | 0,061044 | 0,054752 | NA |
| | 3 | abundance | X07710 | 3-O-beta-D | Handgrip | 0,026279 | 0,061472 | NA |
| | 3 | abundance | X07712 | paracetam | Tibia_length | -0,07069 | 0,054783 | NA |
| | 3 | abundance | X07712 | paracetam | Radius_length | -0,08983 | 0,073176 | NA |
| | 3 | abundance | X07712 | paracetam | Radius_SOS | -0,00446 | 0,052202 | NA |
| | 3 | abundance | X07712 | paracetam | Tibia_SOS | -0,07439 | 0,055036 | NA |
| | 3 | abundance | X07712 | paracetam | Handgrip | -0,01548 | 0,061746 | NA |
| 2b | | abundance | X07731 | 3-[4-methy | Tibia_length | -0,08961 | 0,053252 | NA |
| 2b | | abundance | X07731 | 3-[4-methy | Radius_length | -0,1516 | 0,071494 | NA |
| 2b | | abundance | X07731 | 3-[4-methy | Radius_SOS | 0,053957 | 0,051045 | NA |
| 2b | | abundance | X07731 | 3-[4-methy | Tibia_SOS | -0,04324 | 0,054048 | NA |
| 2b | | abundance | X07731 | 3-[4-methy | Handgrip | -0,03255 | 0,061598 | NA |
| 2b | | abundance | X07732 | N-Acetylva | Tibia_length | 0,034365 | 0,05398 | NA |
| 2b | | abundance | X07732 | N-Acetylva | Radius_length | 0,086872 | 0,072508 | NA |
| 2b | | abundance | X07732 | N-Acetylva | Radius_SOS | 0,023053 | 0,051357 | NA |
| 2b | | abundance | X07732 | N-Acetylva | Tibia_SOS | 0,126213 | 0,054626 | NA |
| 2b | | abundance | X07732 | N-Acetylva | Handgrip | 0,00865 | 0,061599 | NA |
| 2b | | abundance | X07743 | D-Alanine | Tibia_length | -0,01959 | 0,049591 | NA |

| | | | | | | | |
|---|---|---|---|---|---|---|---|
| 2b | abundance | X07743 | D-Alanine | Radius_length | -0,04137 | 0,067684 | NA |
| 2b | abundance | X07743 | D-Alanine | Radius_SOS | 0,003469 | 0,049029 | NA |
| 2b | abundance | X07743 | D-Alanine | Tibia_SOS | -0,02225 | 0,05248 | NA |
| 2b | abundance | X07743 | D-Alanine | Handgrip | 0,126175 | 0,06002 | NA |
| 3 | abundance | X07745 | APM | Tibia_length | -0,09503 | 0,05007 | NA |
| 3 | abundance | X07745 | APM | Radius_length | -0,12331 | 0,068075 | NA |
| 3 | abundance | X07745 | APM | Radius_SOS | -0,02999 | 0,049133 | NA |
| 3 | abundance | X07745 | APM | Tibia_SOS | -0,05713 | 0,052723 | NA |
| 3 | abundance | X07745 | APM | Handgrip | -0,04308 | 0,06045 | NA |
| 2b | abundance | X07746 | 3-(2-Oxo-2 | Tibia_length | -0,03166 | 0,05117 | NA |
| 2b | abundance | X07746 | 3-(2-Oxo-2 | Radius_length | -0,09216 | 0,069801 | NA |
| 2b | abundance | X07746 | 3-(2-Oxo-2 | Radius_SOS | 0,078653 | 0,049347 | NA |
| 2b | abundance | X07746 | 3-(2-Oxo-2 | Tibia_SOS | -0,05417 | 0,053052 | NA |
| 2b | abundance | X07746 | 3-(2-Oxo-2 | Handgrip | 0,016932 | 0,060915 | NA |
| 3 | abundance | X07747 | Diethylpyr | Tibia_length | -0,02123 | 0,055614 | NA |
| 3 | abundance | X07747 | Diethylpyr | Radius_length | 0,041428 | 0,074433 | NA |
| 3 | abundance | X07747 | Diethylpyr | Radius_SOS | -0,02222 | 0,05289 | NA |
| 3 | abundance | X07747 | Diethylpyr | Tibia_SOS | -0,0168 | 0,056882 | NA |
| 3 | abundance | X07747 | Diethylpyr | Handgrip | 0,104346 | 0,063468 | NA |
| 3 | abundance | X07749 | (DL)-3-O-M | Tibia_length | -0,11832 | 0,056045 | NA |
| 3 | abundance | X07749 | (DL)-3-O-M | Radius_length | -0,20034 | 0,073456 | NA |
| 3 | abundance | X07749 | (DL)-3-O-M | Radius_SOS | -0,06357 | 0,053412 | NA |
| 3 | abundance | X07749 | (DL)-3-O-M | Tibia_SOS | -0,14097 | 0,056261 | NA |
| 3 | abundance | X07749 | (DL)-3-O-M | Handgrip | -0,13088 | 0,061163 | NA |
| 3 | abundance | X07753 | Lovastatin | Tibia_length | 0,052226 | 0,052936 | NA |
| 3 | abundance | X07753 | Lovastatin | Radius_length | 0,071328 | 0,071513 | NA |
| 3 | abundance | X07753 | Lovastatin | Radius_SOS | 0,019761 | 0,050601 | NA |
| 3 | abundance | X07753 | Lovastatin | Tibia_SOS | 0,018003 | 0,055041 | NA |
| 3 | abundance | X07753 | Lovastatin | Handgrip | -0,08845 | 0,061769 | NA |
| 3 | abundance | X07754 | hexobarbit | Tibia_length | 0,025421 | 0,056431 | NA |
| 3 | abundance | X07754 | hexobarbit | Radius_length | 0,091417 | 0,074169 | NA |
| 3 | abundance | X07754 | hexobarbit | Radius_SOS | 0,055307 | 0,053521 | NA |
| 3 | abundance | X07754 | hexobarbit | Tibia_SOS | -0,05278 | 0,057532 | NA |
| 3 | abundance | X07754 | hexobarbit | Handgrip | 0,061091 | 0,063013 | NA |
| 3 | abundance | X07764 | (S)-2-meth | Tibia_length | 0,101078 | 0,054404 | NA |
| 3 | abundance | X07764 | (S)-2-meth | Radius_length | 0,138237 | 0,073259 | NA |
| 3 | abundance | X07764 | (S)-2-meth | Radius_SOS | 0,072107 | 0,051534 | NA |
| 3 | abundance | X07764 | (S)-2-meth | Tibia_SOS | 0,06622 | 0,055853 | NA |
| 3 | abundance | X07764 | (S)-2-meth | Handgrip | 0,183512 | 0,060648 | NA |
| 1 | abundance | X07770 | Piperine | Tibia_length | -0,03421 | 0,056851 | NA |
| 1 | abundance | X07770 | Piperine | Radius_length | 0,123031 | 0,073999 | NA |
| 1 | abundance | X07770 | Piperine | Radius_SOS | 0,036006 | 0,053879 | NA |
| 1 | abundance | X07770 | Piperine | Tibia_SOS | 0,00638 | 0,057426 | NA |
| 1 | abundance | X07770 | Piperine | Handgrip | 0,060569 | 0,063447 | NA |
| 2b | abundance | X07771 | Caprolacta | Tibia_length | -0,08884 | 0,051143 | NA |
| 2b | abundance | X07771 | Caprolacta | Radius_length | -0,10722 | 0,069904 | NA |
| 2b | abundance | X07771 | Caprolacta | Radius_SOS | -0,1099 | 0,049483 | NA |
| 2b | abundance | X07771 | Caprolacta | Tibia_SOS | -0,03366 | 0,052887 | NA |

| | | | | | | | | |
|---|---|---|---|---|---|---|---|---|
| 2b | | abundance | X07771 | Caprolacta | Handgrip | 0,017919 | 0,06094 | NA |
| | 3 | abundance | X07774 | 7alpha-Hy | Tibia_length | 0,039275 | 0,054984 | NA |
| | 3 | abundance | X07774 | 7alpha-Hy | Radius_length | 0,026382 | 0,073505 | NA |
| | 3 | abundance | X07774 | 7alpha-Hy | Radius_SOS | 0,001584 | 0,052212 | NA |
| | 3 | abundance | X07774 | 7alpha-Hy | Tibia_SOS | -0,01003 | 0,056185 | NA |
| | 3 | abundance | X07774 | 7alpha-Hy | Handgrip | -0,00559 | 0,061933 | NA |
| | 1 | abundance | X07777 | Arginine | Tibia_length | -0,0082 | 0,052496 | NA |
| | 1 | abundance | X07777 | Arginine | Radius_length | 0,10463 | 0,071004 | NA |
| | 1 | abundance | X07777 | Arginine | Radius_SOS | -0,00415 | 0,050346 | NA |
| | 1 | abundance | X07777 | Arginine | Tibia_SOS | 0,050424 | 0,0536 | NA |
| | 1 | abundance | X07777 | Arginine | Handgrip | 0,026025 | 0,061871 | NA |
| | 1 | abundance | X07782 | Cytosine | Tibia_length | -0,01612 | 0,053325 | NA |
| | 1 | abundance | X07782 | Cytosine | Radius_length | 0,083246 | 0,072283 | NA |
| | 1 | abundance | X07782 | Cytosine | Radius_SOS | 0,029968 | 0,050883 | NA |
| | 1 | abundance | X07782 | Cytosine | Tibia_SOS | 0,061425 | 0,05452 | NA |
| | 1 | abundance | X07782 | Cytosine | Handgrip | 0,089316 | 0,061724 | NA |
| | 3 | abundance | X07783 | 1,3-dimeth | Tibia_length | -0,06265 | 0,054093 | NA |
| | 3 | abundance | X07783 | 1,3-dimeth | Radius_length | 0,040697 | 0,072841 | NA |
| | 3 | abundance | X07783 | 1,3-dimeth | Radius_SOS | 0,047506 | 0,051469 | NA |
| | 3 | abundance | X07783 | 1,3-dimeth | Tibia_SOS | 0,010687 | 0,054898 | NA |
| | 3 | abundance | X07783 | 1,3-dimeth | Handgrip | -0,01109 | 0,06181 | NA |
| 2b | | abundance | X07792 | N-Acetylhi: | Tibia_length | -0,07059 | 0,051037 | NA |
| 2b | | abundance | X07792 | N-Acetylhi: | Radius_length | -0,18816 | 0,068797 | NA |
| 2b | | abundance | X07792 | N-Acetylhi: | Radius_SOS | -0,09751 | 0,049295 | NA |
| 2b | | abundance | X07792 | N-Acetylhi: | Tibia_SOS | -0,09188 | 0,053142 | NA |
| 2b | | abundance | X07792 | N-Acetylhi: | Handgrip | 0,039776 | 0,060822 | NA |
| 2b | | abundance | X07794 | Prolylleuci | Tibia_length | -0,06367 | 0,052555 | NA |
| 2b | | abundance | X07794 | Prolylleuci | Radius_length | -0,11277 | 0,071348 | NA |
| 2b | | abundance | X07794 | Prolylleuci | Radius_SOS | -0,08908 | 0,050242 | NA |
| 2b | | abundance | X07794 | Prolylleuci | Tibia_SOS | 0,028622 | 0,054341 | NA |
| 2b | | abundance | X07794 | Prolylleuci | Handgrip | 0,01926 | 0,062004 | NA |
| | 3 | abundance | X07798 | N-lauroylgl | Tibia_length | -0,06543 | 0,053664 | NA |
| | 3 | abundance | X07798 | N-lauroylgl | Radius_length | -0,0779 | 0,072217 | NA |
| | 3 | abundance | X07798 | N-lauroylgl | Radius_SOS | -0,04995 | 0,051119 | NA |
| | 3 | abundance | X07798 | N-lauroylgl | Tibia_SOS | -0,08959 | 0,055371 | NA |
| | 3 | abundance | X07798 | N-lauroylgl | Handgrip | -0,13977 | 0,060758 | NA |
| 2b | | abundance | X07799 | Capryloylg | Tibia_length | 0,096043 | 0,051877 | NA |
| 2b | | abundance | X07799 | Capryloylg | Radius_length | 0,098338 | 0,071017 | NA |
| 2b | | abundance | X07799 | Capryloylg | Radius_SOS | 0,052412 | 0,049938 | NA |
| 2b | | abundance | X07799 | Capryloylg | Tibia_SOS | 0,048435 | 0,053343 | NA |
| 2b | | abundance | X07799 | Capryloylg | Handgrip | -0,00411 | 0,061303 | NA |
| | 3 | abundance | X07811 | Metirosine | Tibia_length | -0,07846 | 0,051586 | NA |
| | 3 | abundance | X07811 | Metirosine | Radius_length | -0,02434 | 0,07071 | NA |
| | 3 | abundance | X07811 | Metirosine | Radius_SOS | -0,00483 | 0,049788 | NA |
| | 3 | abundance | X07811 | Metirosine | Tibia_SOS | 0,006147 | 0,053729 | NA |
| | 3 | abundance | X07811 | Metirosine | Handgrip | -0,17638 | 0,06022 | NA |
| | 3 | abundance | X07812 | Triethyl citı | Tibia_length | -0,11385 | 0,057643 | NA |
| | 3 | abundance | X07812 | Triethyl citı | Radius_length | 0,001445 | 0,075244 | NA |

| | | | | | | | |
|---|---|---|---|---|---|---|---|
| 3 | abundance | X07812 | Triethyl citı | Radius_SOS | -0,11366 | 0,055313 | NA |
| 3 | abundance | X07812 | Triethyl citı | Tibia_SOS | 0,007905 | 0,057768 | NA |
| 3 | abundance | X07812 | Triethyl citı | Handgrip | 0,067781 | 0,063421 | NA |
| 3 | abundance | X07815 | 3-[2-[(Z)-[3 | Tibia_length | -0,01148 | 0,051443 | NA |
| 3 | abundance | X07815 | 3-[2-[(Z)-[3 | Radius_length | 0,068352 | 0,070591 | NA |
| 3 | abundance | X07815 | 3-[2-[(Z)-[3 | Radius_SOS | 0,03638 | 0,049632 | NA |
| 3 | abundance | X07815 | 3-[2-[(Z)-[3 | Tibia_SOS | -0,00331 | 0,053089 | NA |
| 3 | abundance | X07815 | 3-[2-[(Z)-[3 | Handgrip | -0,00873 | 0,061033 | NA |
| 1 | abundance | X07820 | Creatinine | Tibia_length | -0,12657 | 0,053633 | NA |
| 1 | abundance | X07820 | Creatinine | Radius_length | -0,12764 | 0,072771 | NA |
| 1 | abundance | X07820 | Creatinine | Radius_SOS | -0,12413 | 0,050986 | NA |
| 1 | abundance | X07820 | Creatinine | Tibia_SOS | -0,0939 | 0,05468 | NA |
| 1 | abundance | X07820 | Creatinine | Handgrip | 0,040661 | 0,061327 | NA |
| 1 | abundance | X07825 | Maltotriose | Tibia_length | -0,06289 | 0,055038 | NA |
| 1 | abundance | X07825 | Maltotriose | Radius_length | 0,063697 | 0,073798 | NA |
| 1 | abundance | X07825 | Maltotriose | Radius_SOS | -0,00425 | 0,052399 | NA |
| 1 | abundance | X07825 | Maltotriose | Tibia_SOS | -0,016 | 0,05696 | NA |
| 1 | abundance | X07825 | Maltotriose | Handgrip | 0,032563 | 0,060161 | NA |
| 2b | abundance | X07830 | 1,5-Isoquir | Tibia_length | -0,05407 | 0,057577 | NA |
| 2b | abundance | X07830 | 1,5-Isoquir | Radius_length | -0,05937 | 0,074697 | NA |
| 2b | abundance | X07830 | 1,5-Isoquir | Radius_SOS | -0,05678 | 0,054778 | NA |
| 2b | abundance | X07830 | 1,5-Isoquir | Tibia_SOS | 0,008627 | 0,058477 | NA |
| 2b | abundance | X07830 | 1,5-Isoquir | Handgrip | -0,06868 | 0,060051 | NA |
| 3 | abundance | X07834 | Choline su | Tibia_length | 0,005045 | 0,05274 | NA |
| 3 | abundance | X07834 | Choline su | Radius_length | -0,07703 | 0,07163 | NA |
| 3 | abundance | X07834 | Choline su | Radius_SOS | -0,00982 | 0,050678 | NA |
| 3 | abundance | X07834 | Choline su | Tibia_SOS | -0,05752 | 0,054798 | NA |
| 3 | abundance | X07834 | Choline su | Handgrip | 0,027126 | 0,061858 | NA |
| 3 | abundance | X07835 | 7-ketodeo› | Tibia_length | 0,010405 | 0,050301 | NA |
| 3 | abundance | X07835 | 7-ketodeo› | Radius_length | 0,062094 | 0,068687 | NA |
| 3 | abundance | X07835 | 7-ketodeo› | Radius_SOS | -0,01452 | 0,049174 | NA |
| 3 | abundance | X07835 | 7-ketodeo› | Tibia_SOS | -0,06937 | 0,052563 | NA |
| 3 | abundance | X07835 | 7-ketodeo› | Handgrip | -5,32E-04 | 0,060765 | NA |
| 3 | abundance | X07841 | 2-(5-Benzy | Tibia_length | -0,06786 | 0,053771 | NA |
| 3 | abundance | X07841 | 2-(5-Benzy | Radius_length | -0,08999 | 0,07311 | NA |
| 3 | abundance | X07841 | 2-(5-Benzy | Radius_SOS | -0,00107 | 0,051259 | NA |
| 3 | abundance | X07841 | 2-(5-Benzy | Tibia_SOS | 7,25E-04 | 0,05429 | NA |
| 3 | abundance | X07841 | 2-(5-Benzy | Handgrip | -0,11288 | 0,061396 | NA |
| 3 | abundance | X07843 | Piperidine | Tibia_length | -0,05491 | 0,049581 | NA |
| 3 | abundance | X07843 | Piperidine | Radius_length | -0,04331 | 0,067069 | NA |
| 3 | abundance | X07843 | Piperidine | Radius_SOS | -0,03165 | 0,049008 | NA |
| 3 | abundance | X07843 | Piperidine | Tibia_SOS | -0,10448 | 0,052201 | NA |
| 3 | abundance | X07843 | Piperidine | Handgrip | 0,035734 | 0,060195 | NA |
| 3 | abundance | X07846 | Carbofurar | Tibia_length | 0,054662 | 0,053533 | NA |
| 3 | abundance | X07846 | Carbofurar | Radius_length | 0,006792 | 0,072815 | NA |
| 3 | abundance | X07846 | Carbofurar | Radius_SOS | -0,00653 | 0,051175 | NA |
| 3 | abundance | X07846 | Carbofurar | Tibia_SOS | 0,098382 | 0,0551 | NA |
| 3 | abundance | X07846 | Carbofurar | Handgrip | -0,02182 | 0,06346 | NA |

| | | | | | | | |
|---|---|---|---|---|---|---|---|
| 3 | abundance | X07848 | (âˆ’)-nabil | Tibia_length | 0,010463 | 0,049541 | NA |
| 3 | abundance | X07848 | (âˆ’)-nabil | Radius_length | -0,02487 | 0,066858 | NA |
| 3 | abundance | X07848 | (âˆ’)-nabil | Radius_SOS | 0,030418 | 0,049007 | NA |
| 3 | abundance | X07848 | (âˆ’)-nabil | Tibia_SOS | -0,09229 | 0,052351 | NA |
| 3 | abundance | X07848 | (âˆ’)-nabil | Handgrip | 0,007796 | 0,061017 | NA |
| 3 | abundance | X07849 | Crotamitor | Tibia_length | 0,014823 | 0,050701 | NA |
| 3 | abundance | X07849 | Crotamitor | Radius_length | 0,003259 | 0,069471 | NA |
| 3 | abundance | X07849 | Crotamitor | Radius_SOS | -0,02841 | 0,049323 | NA |
| 3 | abundance | X07849 | Crotamitor | Tibia_SOS | 0,025134 | 0,052972 | NA |
| 3 | abundance | X07849 | Crotamitor | Handgrip | -0,00751 | 0,060987 | NA |
| 3 | abundance | X07852 | N6,N6,N6- | Tibia_length | -0,06029 | 0,05155 | NA |
| 3 | abundance | X07852 | N6,N6,N6- | Radius_length | -0,12041 | 0,06979 | NA |
| 3 | abundance | X07852 | N6,N6,N6- | Radius_SOS | -0,06969 | 0,049645 | NA |
| 3 | abundance | X07852 | N6,N6,N6- | Tibia_SOS | -0,05821 | 0,053371 | NA |
| 3 | abundance | X07852 | N6,N6,N6- | Handgrip | -0,05672 | 0,061224 | NA |
| 3 | abundance | X07854 | TO0127900 | Tibia_length | -0,01418 | 0,05041 | NA |
| 3 | abundance | X07854 | TO0127900 | Radius_length | -0,01292 | 0,069178 | NA |
| 3 | abundance | X07854 | TO0127900 | Radius_SOS | -0,07672 | 0,049039 | NA |
| 3 | abundance | X07854 | TO0127900 | Tibia_SOS | -0,03018 | 0,052751 | NA |
| 3 | abundance | X07854 | TO0127900 | Handgrip | 0,0189 | 0,06069 | NA |

| | | | | | | | |
|---|---|---|---|---|---|---|---|
| 3 | abundance | X07856 | [illegible],6R,7R,9R,11R,12R,13S,14R)-6-{[(2S,3R,4S,6R)-4-(Dimethylamino)-3-hydroxy-6-methyltetrahydro-2H-pyran-2-yl]oxy}-12,13-dihydroxy-14-[(1S)-1-hydroxyethyl]-4-{[(2R,4R,5S,6S)-5-hydroxy-4-methoxy-4,6-dimethyltetrahydro-2H-pyran-2-yl]oxy}-7[illegible] | Tibia_length | 0,015311 | 0,058575 | NA |

| | | | | | | | |
|---|---|---|---|---|---|---|---|
| 3 | abundance | X07856 | ,6R,7R,9R,11R,12R,13S,14R)-6-{[(2S,3R,4S,6R)-4-(Dimethylamino)-3-hydroxy-6-methyltetrahydro-2H-pyran-2-yl]oxy}-12,13-dihydroxy-14-[(1S)-1-hydroxyethyl]-4-{[(2R,4R,5S,6S)-5-hydroxy-4-methoxy-4,6-dimethyltetrahydro-2H-pyran-2-yl]oxy}-7 | Radius_length | 0,182576 | 0,073966 | NA |

| | | | | | | | |
|---|---|---|---|---|---|---|---|
| 3 | abundance | X07856 | [illegible],6R,7R,9R,11R,12R,13S,14R)-6-{[(2S,3R,4S,6R)-4-(Dimethylamino)-3-hydroxy-6-methyltetrahydro-2H-pyran-2-yl]oxy}-12,13-dihydroxy-14-[(1S)-1-hydroxyethyl]-4-{[(2R,4R,5S,6S)-5-hydroxy-4-methoxy-4,6-dimethyltetrahydro-2H-pyran-2-yl]oxy}-7 | Radius_SOS | 0,013029 | 0,055562 | NA |

| | | | | | | | |
|---|---|---|---|---|---|---|---|
| 3 | abundance | X07856 | ,6R,7R,9R ,11R,12R, 13S,14R)- 6- {[(2S,3R,4 S,6R)-4- (Dimethyl amino)-3- hydroxy-6- methyltetr ahydro- 2H-pyran- 2-yl]oxy}- 12,13- dihydroxy- 14-[(1S)-1- hydroxyet hyl]-4- {[(2R,4R,5 S,6S)-5- hydroxy-4- methoxy- 4 ,6- dimethylt etrahydro- 2H-pyran- 2-yl]oxy}-7 | Tibia_SOS | 0,089392 | 0,058444 | NA |

| | | | | | | | | |
|---|---|---|---|---|---|---|---|---|
| | 3 | abundance | X07856 | [illegible],6R,7R,9R,11R,12R,13S,14R)-6-{[(2S,3R,4S,6R)-4-(Dimethylamino)-3-hydroxy-6-methyltetrahydro-2H-pyran-2-yl]oxy}-12,13-dihydroxy-14-[(1S)-1-hydroxyethyl]-4-{[(2R,4R,5S,6S)-5-hydroxy-4-methoxy-4,6-dimethyltetrahydro-2H-pyran-2-yl]oxy}-7 | Handgrip | 0,090305 | 0,059947 | NA |
| 2b | | abundance | X07866 | 3,3,5,5-Tet | Tibia_length | -0,0115 | 0,051055 | NA |
| 2b | | abundance | X07866 | 3,3,5,5-Tet | Radius_length | -0,05724 | 0,069643 | NA |
| 2b | | abundance | X07866 | 3,3,5,5-Tet | Radius_SOS | -0,0417 | 0,049405 | NA |
| 2b | | abundance | X07866 | 3,3,5,5-Tet | Tibia_SOS | -0,02183 | 0,05342 | NA |
| 2b | | abundance | X07866 | 3,3,5,5-Tet | Handgrip | 0,018504 | 0,061341 | NA |
| | 3 | abundance | X07867 | N-Acetylva | Tibia_length | -0,04367 | 0,057069 | NA |
| | 3 | abundance | X07867 | N-Acetylva | Radius_length | -0,12125 | 0,074199 | NA |
| | 3 | abundance | X07867 | N-Acetylva | Radius_SOS | -0,02087 | 0,05396 | NA |
| | 3 | abundance | X07867 | N-Acetylva | Tibia_SOS | -0,00616 | 0,057578 | NA |
| | 3 | abundance | X07867 | N-Acetylva | Handgrip | -0,04629 | 0,062294 | NA |
| 2b | | abundance | X07868 | Biotin | Tibia_length | 0,053104 | 0,054089 | NA |
| 2b | | abundance | X07868 | Biotin | Radius_length | 0,019804 | 0,072976 | NA |
| 2b | | abundance | X07868 | Biotin | Radius_SOS | 0,022732 | 0,051705 | NA |
| 2b | | abundance | X07868 | Biotin | Tibia_SOS | -0,03283 | 0,054604 | NA |
| 2b | | abundance | X07868 | Biotin | Handgrip | -0,15687 | 0,061445 | NA |
| 2a | | abundance | X07869 | Hexamethy | Tibia_length | 0,054785 | 0,052011 | NA |
| 2a | | abundance | X07869 | Hexamethy | Radius_length | 0,032519 | 0,071196 | NA |
| 2a | | abundance | X07869 | Hexamethy | Radius_SOS | 0,012406 | 0,050129 | NA |
| 2a | | abundance | X07869 | Hexamethy | Tibia_SOS | -0,00824 | 0,053913 | NA |

| | | | | | | | | |
|---|---|---|---|---|---|---|---|---|
| 2a | | abundance | X07869 | Hexamethy | Handgrip | 0,063074 | 0,061539 | NA |
| | 3 | abundance | X07873 | (3Z,6Z,9Z,: | Tibia_length | 0,081502 | 0,054887 | NA |
| | 3 | abundance | X07873 | (3Z,6Z,9Z,: | Radius_length | 0,178263 | 0,07243 | NA |
| | 3 | abundance | X07873 | (3Z,6Z,9Z,: | Radius_SOS | -0,05146 | 0,052033 | NA |
| | 3 | abundance | X07873 | (3Z,6Z,9Z,: | Tibia_SOS | 0,049316 | 0,055146 | NA |
| | 3 | abundance | X07873 | (3Z,6Z,9Z,: | Handgrip | 0,00541 | 0,061955 | NA |
| 2b | | abundance | X07874 | Î´-Valerola | Tibia_length | -0,06967 | 0,053512 | NA |
| 2b | | abundance | X07874 | Î´-Valerola | Radius_length | -0,0747 | 0,072546 | NA |
| 2b | | abundance | X07874 | Î´-Valerola | Radius_SOS | -0,06919 | 0,051173 | NA |
| 2b | | abundance | X07874 | Î´-Valerola | Tibia_SOS | 0,024973 | 0,054579 | NA |
| 2b | | abundance | X07874 | Î´-Valerola | Handgrip | 0,005603 | 0,061503 | NA |
| | 3 | abundance | X07886 | L-Urobilin | Tibia_length | -0,00851 | 0,049539 | NA |
| | 3 | abundance | X07886 | L-Urobilin | Radius_length | 0,060657 | 0,067216 | NA |
| | 3 | abundance | X07886 | L-Urobilin | Radius_SOS | 0,012218 | 0,049025 | NA |
| | 3 | abundance | X07886 | L-Urobilin | Tibia_SOS | 0,012529 | 0,052499 | NA |
| | 3 | abundance | X07886 | L-Urobilin | Handgrip | 0,064326 | 0,0603 | NA |
| | 3 | abundance | X07889 | UROBILIN, | Tibia_length | 0,001385 | 0,050836 | NA |
| | 3 | abundance | X07889 | UROBILIN, | Radius_length | 0,063977 | 0,069607 | NA |
| | 3 | abundance | X07889 | UROBILIN, | Radius_SOS | 0,041284 | 0,049389 | NA |
| | 3 | abundance | X07889 | UROBILIN, | Tibia_SOS | 0,06172 | 0,052693 | NA |
| | 3 | abundance | X07889 | UROBILIN, | Handgrip | -0,07384 | 0,060702 | NA |
| 2b | | abundance | X07890 | N-(5-aceta | Tibia_length | 0,060732 | 0,050581 | NA |
| 2b | | abundance | X07890 | N-(5-aceta | Radius_length | 0,072368 | 0,069251 | NA |
| 2b | | abundance | X07890 | N-(5-aceta | Radius_SOS | 0,016611 | 0,049335 | NA |
| 2b | | abundance | X07890 | N-(5-aceta | Tibia_SOS | -0,05008 | 0,052766 | NA |
| 2b | | abundance | X07890 | N-(5-aceta | Handgrip | 0,054902 | 0,060998 | NA |
| 2b | | abundance | X07891 | Cyclamic a | Tibia_length | -0,04836 | 0,056007 | NA |
| 2b | | abundance | X07891 | Cyclamic a | Radius_length | -0,06668 | 0,074116 | NA |
| 2b | | abundance | X07891 | Cyclamic a | Radius_SOS | -0,00232 | 0,0534 | NA |
| 2b | | abundance | X07891 | Cyclamic a | Tibia_SOS | -0,03135 | 0,055491 | NA |
| 2b | | abundance | X07891 | Cyclamic a | Handgrip | -0,06045 | 0,061825 | NA |
| | 3 | abundance | X07892 | urobilinoge | Tibia_length | 0,001871 | 0,051819 | NA |
| | 3 | abundance | X07892 | urobilinoge | Radius_length | 0,052779 | 0,071636 | NA |
| | 3 | abundance | X07892 | urobilinoge | Radius_SOS | 0,051592 | 0,049982 | NA |
| | 3 | abundance | X07892 | urobilinoge | Tibia_SOS | 0,131447 | 0,053844 | NA |
| | 3 | abundance | X07892 | urobilinoge | Handgrip | -0,0179 | 0,061364 | NA |
| 2b | | abundance | X07899 | N-Acetylpu | Tibia_length | -0,09754 | 0,050307 | NA |
| 2b | | abundance | X07899 | N-Acetylpu | Radius_length | -0,11089 | 0,068686 | NA |
| 2b | | abundance | X07899 | N-Acetylpu | Radius_SOS | -0,08572 | 0,049127 | NA |
| 2b | | abundance | X07899 | N-Acetylpu | Tibia_SOS | -0,01433 | 0,052638 | NA |
| 2b | | abundance | X07899 | N-Acetylpu | Handgrip | 0,021191 | 0,060944 | NA |
| | 3 | abundance | X07900 | presqualer | Tibia_length | 0,032448 | 0,057039 | NA |
| | 3 | abundance | X07900 | presqualer | Radius_length | 0,159619 | 0,073869 | NA |
| | 3 | abundance | X07900 | presqualer | Radius_SOS | 0,031137 | 0,054094 | NA |
| | 3 | abundance | X07900 | presqualer | Tibia_SOS | 0,090225 | 0,05646 | NA |
| | 3 | abundance | X07900 | presqualer | Handgrip | 2,14E-04 | 0,063294 | NA |
| 2b | | abundance | X07903 | Acetophen | Tibia_length | -0,12125 | 0,049417 | NA |
| 2b | | abundance | X07903 | Acetophen | Radius_length | -0,15505 | 0,066799 | NA |

| | | | | | | | |
|---|---|---|---|---|---|---|---|
| 2b | abundance | X07903 | Acetophen | Radius_SOS | -0,15903 | 0,048426 | NA |
| 2b | abundance | X07903 | Acetophen | Tibia_SOS | -0,07237 | 0,052371 | NA |
| 2b | abundance | X07903 | Acetophen | Handgrip | -0,03693 | 0,060655 | NA |
| 2b | abundance | X07905 | Prilocaine | Tibia_length | 0,049833 | 0,050538 | NA |
| 2b | abundance | X07905 | Prilocaine | Radius_length | 0,003622 | 0,069423 | NA |
| 2b | abundance | X07905 | Prilocaine | Radius_SOS | -0,01322 | 0,049291 | NA |
| 2b | abundance | X07905 | Prilocaine | Tibia_SOS | 0,008649 | 0,053007 | NA |
| 2b | abundance | X07905 | Prilocaine | Handgrip | -0,02725 | 0,060869 | NA |
| 1 | abundance | X07907 | 4-Acetami | Tibia_length | -0,02026 | 0,04896 | NA |
| 1 | abundance | X07907 | 4-Acetami | Radius_length | 0,100265 | 0,048723 | NA |
| 1 | abundance | X07907 | 4-Acetami | Radius_SOS | -0,04723 | 0,048974 | NA |
| 1 | abundance | X07907 | 4-Acetami | Tibia_SOS | 0,064564 | 0,059731 | NA |
| 1 | abundance | X07907 | 4-Acetami | Handgrip | 0,105677 | 0,059856 | NA |
| 2b | abundance | X07909 | (Â±)-Albute | Tibia_length | -0,01962 | 0,057889 | NA |
| 2b | abundance | X07909 | (Â±)-Albute | Radius_length | -1,32E-04 | 0,075551 | NA |
| 2b | abundance | X07909 | (Â±)-Albute | Radius_SOS | -0,07314 | 0,054402 | NA |
| 2b | abundance | X07909 | (Â±)-Albute | Tibia_SOS | -0,04093 | 0,060059 | NA |
| 2b | abundance | X07909 | (Â±)-Albute | Handgrip | 0,112937 | 0,062621 | NA |
| 3 | abundance | X07914 | (-)-Erythror | Tibia_length | -0,03898 | 0,055847 | NA |
| 3 | abundance | X07914 | (-)-Erythror | Radius_length | 0,149645 | 0,073403 | NA |
| 3 | abundance | X07914 | (-)-Erythror | Radius_SOS | 0,03203 | 0,053029 | NA |
| 3 | abundance | X07914 | (-)-Erythror | Tibia_SOS | 0,119384 | 0,055666 | NA |
| 3 | abundance | X07914 | (-)-Erythror | Handgrip | 0,126326 | 0,061687 | NA |
| 3 | abundance | X07916 | N6,N6,N6- | Tibia_length | -0,02816 | 0,050432 | NA |
| 3 | abundance | X07916 | N6,N6,N6- | Radius_length | -0,0357 | 0,069068 | NA |
| 3 | abundance | X07916 | N6,N6,N6- | Radius_SOS | -0,0693 | 0,049081 | NA |
| 3 | abundance | X07916 | N6,N6,N6- | Tibia_SOS | -0,04373 | 0,052773 | NA |
| 3 | abundance | X07916 | N6,N6,N6- | Handgrip | -0,00803 | 0,060777 | NA |
| 1 | abundance | X07921 | Pipecolinic | Tibia_length | 0,054872 | 0,051332 | NA |
| 1 | abundance | X07921 | Pipecolinic | Radius_length | 0,203248 | 0,06902 | NA |
| 1 | abundance | X07921 | Pipecolinic | Radius_SOS | 0,13199 | 0,049246 | NA |
| 1 | abundance | X07921 | Pipecolinic | Tibia_SOS | 0,077302 | 0,053117 | NA |
| 1 | abundance | X07921 | Pipecolinic | Handgrip | 0,027809 | 0,061031 | NA |
| 3 | abundance | X07925 | DNOP | Tibia_length | 0,020695 | 0,052845 | NA |
| 3 | abundance | X07925 | DNOP | Radius_length | -0,00199 | 0,072123 | NA |
| 3 | abundance | X07925 | DNOP | Radius_SOS | 0,002203 | 0,050532 | NA |
| 3 | abundance | X07925 | DNOP | Tibia_SOS | 0,002927 | 0,054357 | NA |
| 3 | abundance | X07925 | DNOP | Handgrip | 0,014263 | 0,061303 | NA |
| 2b | abundance | X07930 | N-Methylca | Tibia_length | -0,00359 | 0,050141 | NA |
| 2b | abundance | X07930 | N-Methylca | Radius_length | -0,03671 | 0,068252 | NA |
| 2b | abundance | X07930 | N-Methylca | Radius_SOS | -0,04212 | 0,049071 | NA |
| 2b | abundance | X07930 | N-Methylca | Tibia_SOS | -0,0636 | 0,052707 | NA |
| 2b | abundance | X07930 | N-Methylca | Handgrip | 0,011812 | 0,060848 | NA |
| 3 | abundance | X07935 | Limonin | Tibia_length | 0,017767 | 0,05816 | NA |
| 3 | abundance | X07935 | Limonin | Radius_length | 0,08559 | 0,074802 | NA |
| 3 | abundance | X07935 | Limonin | Radius_SOS | -0,01328 | 0,055372 | NA |
| 3 | abundance | X07935 | Limonin | Tibia_SOS | -0,02916 | 0,058934 | NA |
| 3 | abundance | X07935 | Limonin | Handgrip | 0,105448 | 0,059857 | NA |

| | | | | | | | | |
|---|---|---|---|---|---|---|---|---|
| | 1 | abundance | X07937 | Atenolol | Tibia_length | -0,06213 | 0,051585 | NA |
| | 1 | abundance | X07937 | Atenolol | Radius_length | -0,09534 | 0,070475 | NA |
| | 1 | abundance | X07937 | Atenolol | Radius_SOS | 4,62E-04 | 0,04984 | NA |
| | 1 | abundance | X07937 | Atenolol | Tibia_SOS | -0,02303 | 0,053009 | NA |
| | 1 | abundance | X07937 | Atenolol | Handgrip | 0,038839 | 0,06105 | NA |
| | 3 | abundance | X07944 | Styrene | Tibia_length | -0,0162 | 0,050897 | NA |
| | 3 | abundance | X07944 | Styrene | Radius_length | -0,15548 | 0,068925 | NA |
| | 3 | abundance | X07944 | Styrene | Radius_SOS | -0,19338 | 0,048441 | NA |
| | 3 | abundance | X07944 | Styrene | Tibia_SOS | -0,04855 | 0,052973 | NA |
| | 3 | abundance | X07944 | Styrene | Handgrip | -0,15638 | 0,06047 | NA |
| | 3 | abundance | X07955 | Valylvaline | Tibia_length | -0,10289 | 0,054945 | NA |
| | 3 | abundance | X07955 | Valylvaline | Radius_length | -0,03377 | 0,073842 | NA |
| | 3 | abundance | X07955 | Valylvaline | Radius_SOS | -0,05754 | 0,052641 | NA |
| | 3 | abundance | X07955 | Valylvaline | Tibia_SOS | 0,00482 | 0,056956 | NA |
| | 3 | abundance | X07955 | Valylvaline | Handgrip | -0,07991 | 0,062236 | NA |
| | 3 | abundance | X07961 | 2-(Hydroxy | Tibia_length | 0,076885 | 0,049801 | NA |
| | 3 | abundance | X07961 | 2-(Hydroxy | Radius_length | 0,158449 | 0,067268 | NA |
| | 3 | abundance | X07961 | 2-(Hydroxy | Radius_SOS | 0,014108 | 0,04905 | NA |
| | 3 | abundance | X07961 | 2-(Hydroxy | Tibia_SOS | 0,100084 | 0,052298 | NA |
| | 3 | abundance | X07961 | 2-(Hydroxy | Handgrip | 0,009772 | 0,06045 | NA |
| | 1 | abundance | X07963 | Theobromi | Tibia_length | -0,07634 | 0,055423 | NA |
| | 1 | abundance | X07963 | Theobromi | Radius_length | -0,01924 | 0,073997 | NA |
| | 1 | abundance | X07963 | Theobromi | Radius_SOS | 0,033707 | 0,05279 | NA |
| | 1 | abundance | X07963 | Theobromi | Tibia_SOS | -0,07457 | 0,055741 | NA |
| | 1 | abundance | X07963 | Theobromi | Handgrip | -0,02957 | 0,063627 | NA |
| | 3 | abundance | X07969 | 6-hydroxyp | Tibia_length | 0,015869 | 0,049659 | NA |
| | 3 | abundance | X07969 | 6-hydroxyp | Radius_length | 0,04797 | 0,067164 | NA |
| | 3 | abundance | X07969 | 6-hydroxyp | Radius_SOS | -0,03278 | 0,049005 | NA |
| | 3 | abundance | X07969 | 6-hydroxyp | Tibia_SOS | -0,01121 | 0,05253 | NA |
| | 3 | abundance | X07969 | 6-hydroxyp | Handgrip | 0,007499 | 0,060913 | NA |
| | 3 | abundance | X07974 | Isoprene | Tibia_length | 0,083101 | 0,050614 | NA |
| | 3 | abundance | X07974 | Isoprene | Radius_length | 0,138028 | 0,06878 | NA |
| | 3 | abundance | X07974 | Isoprene | Radius_SOS | 0,020789 | 0,049277 | NA |
| | 3 | abundance | X07974 | Isoprene | Tibia_SOS | 0,146559 | 0,052199 | NA |
| | 3 | abundance | X07974 | Isoprene | Handgrip | 0,066748 | 0,060235 | NA |
| 2b | | abundance | X07977 | Tyramine | Tibia_length | -0,13145 | 0,049771 | NA |
| 2b | | abundance | X07977 | Tyramine | Radius_length | -0,1574 | 0,067714 | NA |
| 2b | | abundance | X07977 | Tyramine | Radius_SOS | -0,16095 | 0,048516 | NA |
| 2b | | abundance | X07977 | Tyramine | Tibia_SOS | -0,0739 | 0,052473 | NA |
| 2b | | abundance | X07977 | Tyramine | Handgrip | -0,04435 | 0,060901 | NA |
| | 3 | abundance | X07978 | 1-[(4E)-4-( | Tibia_length | 0,02052 | 0,054118 | NA |
| | 3 | abundance | X07978 | 1-[(4E)-4-( | Radius_length | 0,020312 | 0,073111 | NA |
| | 3 | abundance | X07978 | 1-[(4E)-4-( | Radius_SOS | 0,045207 | 0,051503 | NA |
| | 3 | abundance | X07978 | 1-[(4E)-4-( | Tibia_SOS | 0,063832 | 0,055475 | NA |
| | 3 | abundance | X07978 | 1-[(4E)-4-( | Handgrip | 0,002478 | 0,062851 | NA |
| 2b | | abundance | X07981 | Methylimic | Tibia_length | 0,03825 | 0,053222 | NA |
| 2b | | abundance | X07981 | Methylimic | Radius_length | 0,067335 | 0,072175 | NA |
| 2b | | abundance | X07981 | Methylimic | Radius_SOS | 0,115759 | 0,050676 | NA |

| | | | | | | | |
|---|---|---|---|---|---|---|---|
| 2b | abundance | X07981 | Methylimic | Tibia_SOS | 0,045006 | 0,054243 | NA |
| 2b | abundance | X07981 | Methylimic | Handgrip | 0,071448 | 0,061845 | NA |
| 3 | abundance | X07983 | butalbital | Tibia_length | -0,12521 | 0,052393 | NA |
| 3 | abundance | X07983 | butalbital | Radius_length | -0,10612 | 0,071829 | NA |
| 3 | abundance | X07983 | butalbital | Radius_SOS | -0,00494 | 0,050404 | NA |
| 3 | abundance | X07983 | butalbital | Tibia_SOS | -0,05652 | 0,054624 | NA |
| 3 | abundance | X07983 | butalbital | Handgrip | 0,040035 | 0,062194 | NA |
| 3 | abundance | X07989 | Piperidine | Tibia_length | 0,094505 | 0,050348 | NA |
| 3 | abundance | X07989 | Piperidine | Radius_length | 0,141343 | 0,068485 | NA |
| 3 | abundance | X07989 | Piperidine | Radius_SOS | 0,004185 | 0,049224 | NA |
| 3 | abundance | X07989 | Piperidine | Tibia_SOS | 0,124752 | 0,052323 | NA |
| 3 | abundance | X07989 | Piperidine | Handgrip | 0,088217 | 0,060079 | NA |
| 3 | abundance | X07993 | UROBILIN, | Tibia_length | 0,039627 | 0,052289 | NA |
| 3 | abundance | X07993 | UROBILIN, | Radius_length | 0,108659 | 0,071662 | NA |
| 3 | abundance | X07993 | UROBILIN, | Radius_SOS | 0,099417 | 0,050047 | NA |
| 3 | abundance | X07993 | UROBILIN, | Tibia_SOS | 0,093939 | 0,054427 | NA |
| 3 | abundance | X07993 | UROBILIN, | Handgrip | -0,02083 | 0,062674 | NA |
| 3 | abundance | X08002 | (2S)-6-Ami | Tibia_length | 0,033618 | 0,053605 | NA |
| 3 | abundance | X08002 | (2S)-6-Ami | Radius_length | 0,108596 | 0,072786 | NA |
| 3 | abundance | X08002 | (2S)-6-Ami | Radius_SOS | 0,095517 | 0,051441 | NA |
| 3 | abundance | X08002 | (2S)-6-Ami | Tibia_SOS | 0,009348 | 0,055902 | NA |
| 3 | abundance | X08002 | (2S)-6-Ami | Handgrip | -0,02625 | 0,062777 | NA |
| 2b | abundance | X08004 | Î´-Valerola | Tibia_length | -0,02357 | 0,053294 | NA |
| 2b | abundance | X08004 | Î´-Valerola | Radius_length | -0,05455 | 0,072289 | NA |
| 2b | abundance | X08004 | Î´-Valerola | Radius_SOS | -0,05305 | 0,050978 | NA |
| 2b | abundance | X08004 | Î´-Valerola | Tibia_SOS | -0,00882 | 0,054282 | NA |
| 2b | abundance | X08004 | Î´-Valerola | Handgrip | -0,00677 | 0,060906 | NA |
| 3 | abundance | X08007 | MFCD0002 | Tibia_length | 0,010915 | 0,051021 | NA |
| 3 | abundance | X08007 | MFCD0002 | Radius_length | -0,03588 | 0,069847 | NA |
| 3 | abundance | X08007 | MFCD0002 | Radius_SOS | -0,07342 | 0,049316 | NA |
| 3 | abundance | X08007 | MFCD0002 | Tibia_SOS | -0,05581 | 0,052972 | NA |
| 3 | abundance | X08007 | MFCD0002 | Handgrip | 0,04028 | 0,061083 | NA |
| 1 | abundance | X08008 | N-Acetylne | Tibia_length | 0,005488 | 0,053453 | NA |
| 1 | abundance | X08008 | N-Acetylne | Radius_length | 0,064249 | 0,072828 | NA |
| 1 | abundance | X08008 | N-Acetylne | Radius_SOS | -0,03463 | 0,051076 | NA |
| 1 | abundance | X08008 | N-Acetylne | Tibia_SOS | 0,010702 | 0,054264 | NA |
| 1 | abundance | X08008 | N-Acetylne | Handgrip | 0,090081 | 0,062474 | NA |
| 3 | abundance | X08012 | Methylol D | Tibia_length | 0,151391 | 0,049991 | NA |
| 3 | abundance | X08012 | Methylol D | Radius_length | 0,28193 | 0,066276 | NA |
| 3 | abundance | X08012 | Methylol D | Radius_SOS | 0,056176 | 0,049103 | NA |
| 3 | abundance | X08012 | Methylol D | Tibia_SOS | 0,156098 | 0,052071 | NA |
| 3 | abundance | X08012 | Methylol D | Handgrip | 0,109775 | 0,059871 | NA |
| 1 | abundance | X08013 | Deoxysuga | Tibia_length | -0,07156 | 0,054368 | NA |
| 1 | abundance | X08013 | Deoxysuga | Radius_length | -0,04444 | 0,073608 | NA |
| 1 | abundance | X08013 | Deoxysuga | Radius_SOS | -0,03742 | 0,052063 | NA |
| 1 | abundance | X08013 | Deoxysuga | Tibia_SOS | -0,03504 | 0,055391 | NA |
| 1 | abundance | X08013 | Deoxysuga | Handgrip | 0,049928 | 0,062065 | NA |
| 3 | abundance | X08014 | DNOP | Tibia_length | 0,027763 | 0,052965 | NA |

| | | | | | | | | |
|---|---|---|---|---|---|---|---|---|
| | 3 | abundance | X08014 | DNOP | Radius_length | 0,015316 | 0,072287 | NA |
| | 3 | abundance | X08014 | DNOP | Radius_SOS | 0,018523 | 0,050675 | NA |
| | 3 | abundance | X08014 | DNOP | Tibia_SOS | 0,017422 | 0,054422 | NA |
| | 3 | abundance | X08014 | DNOP | Handgrip | -0,01667 | 0,061435 | NA |
| 2b | | abundance | X08017 | Tropinone | Tibia_length | 0,053118 | 0,052838 | NA |
| 2b | | abundance | X08017 | Tropinone | Radius_length | 0,055309 | 0,071998 | NA |
| 2b | | abundance | X08017 | Tropinone | Radius_SOS | 0,011145 | 0,050613 | NA |
| 2b | | abundance | X08017 | Tropinone | Tibia_SOS | 0,041884 | 0,054098 | NA |
| 2b | | abundance | X08017 | Tropinone | Handgrip | -0,02755 | 0,061482 | NA |
| | 1 | abundance | X08019 | Thymidine | Tibia_length | 0,012586 | 0,050637 | NA |
| | 1 | abundance | X08019 | Thymidine | Radius_length | 0,073271 | 0,069077 | NA |
| | 1 | abundance | X08019 | Thymidine | Radius_SOS | -0,08013 | 0,049126 | NA |
| | 1 | abundance | X08019 | Thymidine | Tibia_SOS | 0,059926 | 0,052619 | NA |
| | 1 | abundance | X08019 | Thymidine | Handgrip | 0,043257 | 0,060832 | NA |
| | 3 | abundance | X08024 | 8-Methyl-8 | Tibia_length | -0,08384 | 0,055201 | NA |
| | 3 | abundance | X08024 | 8-Methyl-8 | Radius_length | 0,003353 | 0,074301 | NA |
| | 3 | abundance | X08024 | 8-Methyl-8 | Radius_SOS | -0,06499 | 0,052809 | NA |
| | 3 | abundance | X08024 | 8-Methyl-8 | Tibia_SOS | -0,00237 | 0,056374 | NA |
| | 3 | abundance | X08024 | 8-Methyl-8 | Handgrip | -0,00487 | 0,062615 | NA |
| | 3 | abundance | X08028 | Solanidine | Tibia_length | -0,00924 | 0,054843 | NA |
| | 3 | abundance | X08028 | Solanidine | Radius_length | 0,155234 | 0,072574 | NA |
| | 3 | abundance | X08028 | Solanidine | Radius_SOS | 0,017579 | 0,052139 | NA |
| | 3 | abundance | X08028 | Solanidine | Tibia_SOS | -0,01124 | 0,055502 | NA |
| | 3 | abundance | X08028 | Solanidine | Handgrip | 0,119661 | 0,06245 | NA |
| | 3 | abundance | X08035 | Tetrahydro | Tibia_length | 0,092631 | 0,053142 | NA |
| | 3 | abundance | X08035 | Tetrahydro | Radius_length | 0,147901 | 0,07205 | NA |
| | 3 | abundance | X08035 | Tetrahydro | Radius_SOS | 0,069843 | 0,050566 | NA |
| | 3 | abundance | X08035 | Tetrahydro | Tibia_SOS | 0,066032 | 0,054551 | NA |
| | 3 | abundance | X08035 | Tetrahydro | Handgrip | 0,179606 | 0,059914 | NA |
| 2b | | abundance | X08037 | Crotonic a | Tibia_length | -0,16031 | 0,050598 | NA |
| 2b | | abundance | X08037 | Crotonic a | Radius_length | -0,10625 | 0,069895 | NA |
| 2b | | abundance | X08037 | Crotonic a | Radius_SOS | -0,04215 | 0,049331 | NA |
| 2b | | abundance | X08037 | Crotonic a | Tibia_SOS | -0,03246 | 0,052803 | NA |
| 2b | | abundance | X08037 | Crotonic a | Handgrip | -0,02522 | 0,060399 | NA |
| 2b | | abundance | X08038 | N-{6-[(7-Cl | Tibia_length | -5,15E-04 | 0,054161 | NA |
| 2b | | abundance | X08038 | N-{6-[(7-Cl | Radius_length | -0,00622 | 0,072749 | NA |
| 2b | | abundance | X08038 | N-{6-[(7-Cl | Radius_SOS | 0,051797 | 0,05162 | NA |
| 2b | | abundance | X08038 | N-{6-[(7-Cl | Tibia_SOS | 0,014529 | 0,055364 | NA |
| 2b | | abundance | X08038 | N-{6-[(7-Cl | Handgrip | 0,039477 | 0,062795 | NA |
| 2b | | abundance | X08040 | Prolylleuci | Tibia_length | -0,06209 | 0,053351 | NA |
| 2b | | abundance | X08040 | Prolylleuci | Radius_length | -0,09382 | 0,072143 | NA |
| 2b | | abundance | X08040 | Prolylleuci | Radius_SOS | -0,06668 | 0,050837 | NA |
| 2b | | abundance | X08040 | Prolylleuci | Tibia_SOS | 0,031689 | 0,055002 | NA |
| 2b | | abundance | X08040 | Prolylleuci | Handgrip | 0,046009 | 0,061882 | NA |
| | 3 | abundance | X08044 | 3-[2-[(Z)-[3 | Tibia_length | 0,007606 | 0,050235 | NA |
| | 3 | abundance | X08044 | 3-[2-[(Z)-[3 | Radius_length | 0,06867 | 0,06866 | NA |
| | 3 | abundance | X08044 | 3-[2-[(Z)-[3 | Radius_SOS | 0,060385 | 0,049099 | NA |
| | 3 | abundance | X08044 | 3-[2-[(Z)-[3 | Tibia_SOS | 0,029321 | 0,052525 | NA |

| | | | | | | | | |
|---|---|---|---|---|---|---|---|---|
| | 3 | abundance | X08044 | 3-[2-[(Z)-[3 | Handgrip | -0,04912 | 0,060381 | NA |
| | 3 | abundance | X08046 | 3-Hydroxy- | Tibia_length | -3,39E-04 | 0,056525 | NA |
| | 3 | abundance | X08046 | 3-Hydroxy- | Radius_length | -0,05195 | 0,074216 | NA |
| | 3 | abundance | X08046 | 3-Hydroxy- | Radius_SOS | -0,10584 | 0,053416 | NA |
| | 3 | abundance | X08046 | 3-Hydroxy- | Tibia_SOS | -0,02336 | 0,058018 | NA |
| | 3 | abundance | X08046 | 3-Hydroxy- | Handgrip | -0,10396 | 0,062355 | NA |
| 2b | | abundance | X08054 | Docosahe› | Tibia_length | 0,043228 | 0,052621 | NA |
| 2b | | abundance | X08054 | Docosahe› | Radius_length | 0,035214 | 0,071942 | NA |
| 2b | | abundance | X08054 | Docosahe› | Radius_SOS | 0,115473 | 0,050268 | NA |
| 2b | | abundance | X08054 | Docosahe› | Tibia_SOS | -0,01595 | 0,053862 | NA |
| 2b | | abundance | X08054 | Docosahe› | Handgrip | 0,034277 | 0,060759 | NA |
| 2b | | abundance | X08056 | 3-(2-Oxo-2 | Tibia_length | -0,05403 | 0,056664 | NA |
| 2b | | abundance | X08056 | 3-(2-Oxo-2 | Radius_length | -0,04941 | 0,074242 | NA |
| 2b | | abundance | X08056 | 3-(2-Oxo-2 | Radius_SOS | -0,0452 | 0,053871 | NA |
| 2b | | abundance | X08056 | 3-(2-Oxo-2 | Tibia_SOS | 0,024144 | 0,057619 | NA |
| 2b | | abundance | X08056 | 3-(2-Oxo-2 | Handgrip | -0,05051 | 0,060116 | NA |
| | 3 | abundance | X08057 | N-{3-Carbo | Tibia_length | -0,08005 | 0,053583 | NA |
| | 3 | abundance | X08057 | N-{3-Carbo | Radius_length | -0,07852 | 0,072584 | NA |
| | 3 | abundance | X08057 | N-{3-Carbo | Radius_SOS | -0,09244 | 0,051233 | NA |
| | 3 | abundance | X08057 | N-{3-Carbo | Tibia_SOS | -0,03424 | 0,054999 | NA |
| | 3 | abundance | X08057 | N-{3-Carbo | Handgrip | -0,03315 | 0,06259 | NA |
| | 3 | abundance | X08058 | 2-Hydroxy- | Tibia_length | -0,01284 | 0,056783 | NA |
| | 3 | abundance | X08058 | 2-Hydroxy- | Radius_length | 0,055006 | 0,074562 | NA |
| | 3 | abundance | X08058 | 2-Hydroxy- | Radius_SOS | -0,02271 | 0,05398 | NA |
| | 3 | abundance | X08058 | 2-Hydroxy- | Tibia_SOS | -0,02647 | 0,05704 | NA |
| | 3 | abundance | X08058 | 2-Hydroxy- | Handgrip | 0,041845 | 0,062604 | NA |
| | 3 | abundance | X08059 | Cadaverin‹ | Tibia_length | -0,05192 | 0,049605 | NA |
| | 3 | abundance | X08059 | Cadaverin‹ | Radius_length | -0,04483 | 0,067097 | NA |
| | 3 | abundance | X08059 | Cadaverin‹ | Radius_SOS | -0,03426 | 0,049005 | NA |
| | 3 | abundance | X08059 | Cadaverin‹ | Tibia_SOS | -0,10757 | 0,052182 | NA |
| | 3 | abundance | X08059 | Cadaverin‹ | Handgrip | 0,032556 | 0,060213 | NA |
| | 1 | abundance | X08062 | Indole-3-ca | Tibia_length | 0,178147 | 0,050241 | NA |
| | 1 | abundance | X08062 | Indole-3-ca | Radius_length | 0,124814 | 0,069358 | NA |
| | 1 | abundance | X08062 | Indole-3-ca | Radius_SOS | -0,04193 | 0,049334 | NA |
| | 1 | abundance | X08062 | Indole-3-ca | Tibia_SOS | 0,127775 | 0,052431 | NA |
| | 1 | abundance | X08062 | Indole-3-ca | Handgrip | 0,112211 | 0,059936 | NA |
| | 3 | abundance | X08072 | 4-Hydroxyp | Tibia_length | 0,062423 | 0,051159 | NA |
| | 3 | abundance | X08072 | 4-Hydroxyp | Radius_length | 0,074762 | 0,070361 | NA |
| | 3 | abundance | X08072 | 4-Hydroxyp | Radius_SOS | -0,01775 | 0,049597 | NA |
| | 3 | abundance | X08072 | 4-Hydroxyp | Tibia_SOS | -0,02114 | 0,053423 | NA |
| | 3 | abundance | X08072 | 4-Hydroxyp | Handgrip | 0,050254 | 0,06153 | NA |
| 2b | | abundance | X08076 | 3',5,7-Trihy | Tibia_length | -0,07474 | 0,057627 | NA |
| 2b | | abundance | X08076 | 3',5,7-Trihy | Radius_length | -0,01745 | 0,075493 | NA |
| 2b | | abundance | X08076 | 3',5,7-Trihy | Radius_SOS | -0,05843 | 0,055055 | NA |
| 2b | | abundance | X08076 | 3',5,7-Trihy | Tibia_SOS | 0,013226 | 0,058912 | NA |
| 2b | | abundance | X08076 | 3',5,7-Trihy | Handgrip | 0,068207 | 0,061597 | NA |
| | 3 | abundance | X08077 | 6-(alpha-D | Tibia_length | -0,15738 | 0,054861 | NA |
| | 3 | abundance | X08077 | 6-(alpha-D | Radius_length | -0,11328 | 0,073345 | NA |

| | | | | | | | |
|---|---|---|---|---|---|---|---|
| 3 | abundance | X08077 | 6-(alpha-D | Radius_SOS | -0,04479 | 0,052886 | NA |
| 3 | abundance | X08077 | 6-(alpha-D | Tibia_SOS | -0,00442 | 0,055736 | NA |
| 3 | abundance | X08077 | 6-(alpha-D | Handgrip | -0,00131 | 0,062416 | NA |
| 3 | abundance | X08078 | Ethyl mala | Tibia_length | -0,06439 | 0,056866 | NA |
| 3 | abundance | X08078 | Ethyl mala | Radius_length | 0,005186 | 0,074482 | NA |
| 3 | abundance | X08078 | Ethyl mala | Radius_SOS | -0,06559 | 0,054379 | NA |
| 3 | abundance | X08078 | Ethyl mala | Tibia_SOS | 0,072512 | 0,057832 | NA |
| 3 | abundance | X08078 | Ethyl mala | Handgrip | -0,02742 | 0,062983 | NA |
| 3 | abundance | X08092 | Safrole | Tibia_length | -0,05582 | 0,0571 | NA |
| 3 | abundance | X08092 | Safrole | Radius_length | 0,005262 | 0,075596 | NA |
| 3 | abundance | X08092 | Safrole | Radius_SOS | -0,02605 | 0,054457 | NA |
| 3 | abundance | X08092 | Safrole | Tibia_SOS | 0,018042 | 0,05731 | NA |
| 3 | abundance | X08092 | Safrole | Handgrip | 0,057867 | 0,062464 | NA |
| 3 | abundance | X08094 | 1,1'-[1,12-I | Tibia_length | 0,040189 | 0,049385 | NA |
| 3 | abundance | X08094 | 1,1'-[1,12-I | Radius_length | 0,01234 | 0,066443 | NA |
| 3 | abundance | X08094 | 1,1'-[1,12-I | Radius_SOS | 0,021171 | 0,049031 | NA |
| 3 | abundance | X08094 | 1,1'-[1,12-I | Tibia_SOS | -0,06605 | 0,05238 | NA |
| 3 | abundance | X08094 | 1,1'-[1,12-I | Handgrip | 0,068819 | 0,060398 | NA |
| 3 | abundance | X08095 | DNOP | Tibia_length | -0,0024 | 0,050493 | NA |
| 3 | abundance | X08095 | DNOP | Radius_length | 0,109271 | 0,068679 | NA |
| 3 | abundance | X08095 | DNOP | Radius_SOS | 0,001067 | 0,049247 | NA |
| 3 | abundance | X08095 | DNOP | Tibia_SOS | -0,01033 | 0,052756 | NA |
| 3 | abundance | X08095 | DNOP | Handgrip | 0,055974 | 0,060533 | NA |
| 3 | abundance | X08096 | (2E)-3-Met | Tibia_length | -0,08032 | 0,057747 | NA |
| 3 | abundance | X08096 | (2E)-3-Met | Radius_length | -0,08933 | 0,075273 | NA |
| 3 | abundance | X08096 | (2E)-3-Met | Radius_SOS | -0,12773 | 0,05486 | NA |
| 3 | abundance | X08096 | (2E)-3-Met | Tibia_SOS | -0,08869 | 0,060057 | NA |
| 3 | abundance | X08096 | (2E)-3-Met | Handgrip | -0,06278 | 0,060074 | NA |
| 1 | abundance | X08098 | Citrulline | Tibia_length | 0,173684 | 0,049491 | NA |
| 1 | abundance | X08098 | Citrulline | Radius_length | 0,232945 | 0,06668 | NA |
| 1 | abundance | X08098 | Citrulline | Radius_SOS | 0,025595 | 0,049091 | NA |
| 1 | abundance | X08098 | Citrulline | Tibia_SOS | 0,148024 | 0,052125 | NA |
| 1 | abundance | X08098 | Citrulline | Handgrip | 0,05298 | 0,06024 | NA |
| 3 | abundance | X08099 | g-Butyrobe | Tibia_length | 0,010104 | 0,051802 | NA |
| 3 | abundance | X08099 | g-Butyrobe | Radius_length | 0,066637 | 0,070943 | NA |
| 3 | abundance | X08099 | g-Butyrobe | Radius_SOS | 0,011406 | 0,049927 | NA |
| 3 | abundance | X08099 | g-Butyrobe | Tibia_SOS | -0,05505 | 0,053168 | NA |
| 3 | abundance | X08099 | g-Butyrobe | Handgrip | -0,10231 | 0,060674 | NA |
| 3 | abundance | X08100 | Piperidine | Tibia_length | -0,00278 | 0,051312 | NA |
| 3 | abundance | X08100 | Piperidine | Radius_length | -0,05694 | 0,070048 | NA |
| 3 | abundance | X08100 | Piperidine | Radius_SOS | -0,04154 | 0,049532 | NA |
| 3 | abundance | X08100 | Piperidine | Tibia_SOS | -0,01394 | 0,053544 | NA |
| 3 | abundance | X08100 | Piperidine | Handgrip | 0,03772 | 0,061549 | NA |
| 3 | abundance | X08103 | Isopelletie | Tibia_length | -0,0343 | 0,051604 | NA |
| 3 | abundance | X08103 | Isopelletie | Radius_length | -0,13674 | 0,069827 | NA |
| 3 | abundance | X08103 | Isopelletie | Radius_SOS | -0,11548 | 0,049523 | NA |
| 3 | abundance | X08103 | Isopelletie | Tibia_SOS | -0,08121 | 0,05348 | NA |
| 3 | abundance | X08103 | Isopelletie | Handgrip | -0,0078 | 0,062063 | NA |

| | | | | | | | |
|---|---|---|---|---|---|---|---|
| | 3 | abundance | X08105 | 2-Acetamid | Tibia_length | 0,013215 | 0,053831 | NA |
| | 3 | abundance | X08105 | 2-Acetamid | Radius_length | 0,11495 | 0,072169 | NA |
| | 3 | abundance | X08105 | 2-Acetamid | Radius_SOS | 0,054813 | 0,051446 | NA |
| | 3 | abundance | X08105 | 2-Acetamid | Tibia_SOS | -0,02644 | 0,055204 | NA |
| | 3 | abundance | X08105 | 2-Acetamid | Handgrip | -0,06031 | 0,062824 | NA |
| | 1 | abundance | X08106 | 4-Vinylphe | Tibia_length | 4,47E-04 | 0,051333 | NA |
| | 1 | abundance | X08106 | 4-Vinylphe | Radius_length | -0,15825 | 0,069636 | NA |
| | 1 | abundance | X08106 | 4-Vinylphe | Radius_SOS | 0,017349 | 0,049633 | NA |
| | 1 | abundance | X08106 | 4-Vinylphe | Tibia_SOS | -0,03519 | 0,05414 | NA |
| | 1 | abundance | X08106 | 4-Vinylphe | Handgrip | -0,01429 | 0,061315 | NA |
| 2b | | abundance | X08111 | 2-Hydroxyc | Tibia_length | 0,142868 | 0,049517 | NA |
| 2b | | abundance | X08111 | 2-Hydroxyc | Radius_length | 0,136073 | 0,067227 | NA |
| 2b | | abundance | X08111 | 2-Hydroxyc | Radius_SOS | 0,036791 | 0,049032 | NA |
| 2b | | abundance | X08111 | 2-Hydroxyc | Tibia_SOS | 0,096249 | 0,052251 | NA |
| 2b | | abundance | X08111 | 2-Hydroxyc | Handgrip | 0,059261 | 0,06013 | NA |
| | 3 | abundance | X08116 | Methyl (2Z, | Tibia_length | 0,047458 | 0,05711 | NA |
| | 3 | abundance | X08116 | Methyl (2Z, | Radius_length | 0,290935 | 0,072176 | NA |
| | 3 | abundance | X08116 | Methyl (2Z, | Radius_SOS | 0,063529 | 0,054142 | NA |
| | 3 | abundance | X08116 | Methyl (2Z, | Tibia_SOS | 0,13439 | 0,056402 | NA |
| | 3 | abundance | X08116 | Methyl (2Z, | Handgrip | 0,125533 | 0,063158 | NA |
| 2b | | abundance | X08123 | Tiglic acid | Tibia_length | -0,10287 | 0,054242 | NA |
| 2b | | abundance | X08123 | Tiglic acid | Radius_length | -0,07271 | 0,073593 | NA |
| 2b | | abundance | X08123 | Tiglic acid | Radius_SOS | -0,08381 | 0,051962 | NA |
| 2b | | abundance | X08123 | Tiglic acid | Tibia_SOS | 0,027944 | 0,055935 | NA |
| 2b | | abundance | X08123 | Tiglic acid | Handgrip | 0,016682 | 0,062013 | NA |
| | 3 | abundance | X08146 | O-Ethyl (4- | Tibia_length | 0,07138 | 0,054262 | NA |
| | 3 | abundance | X08146 | O-Ethyl (4- | Radius_length | 0,028869 | 0,073125 | NA |
| | 3 | abundance | X08146 | O-Ethyl (4- | Radius_SOS | -0,03969 | 0,051681 | NA |
| | 3 | abundance | X08146 | O-Ethyl (4- | Tibia_SOS | 0,051507 | 0,05489 | NA |
| | 3 | abundance | X08146 | O-Ethyl (4- | Handgrip | -0,05716 | 0,062585 | NA |
| | 3 | abundance | X08147 | porphobilir | Tibia_length | -0,14218 | 0,056419 | NA |
| | 3 | abundance | X08147 | porphobilir | Radius_length | -0,14644 | 0,07411 | NA |
| | 3 | abundance | X08147 | porphobilir | Radius_SOS | -0,05853 | 0,053456 | NA |
| | 3 | abundance | X08147 | porphobilir | Tibia_SOS | -0,10019 | 0,055826 | NA |
| | 3 | abundance | X08147 | porphobilir | Handgrip | -0,09824 | 0,06042 | NA |
| 2b | | abundance | X08150 | DL-4-Hydro | Tibia_length | 0,133515 | 0,052014 | NA |
| 2b | | abundance | X08150 | DL-4-Hydro | Radius_length | 0,22174 | 0,070295 | NA |
| 2b | | abundance | X08150 | DL-4-Hydro | Radius_SOS | 0,17869 | 0,048993 | NA |
| 2b | | abundance | X08150 | DL-4-Hydro | Tibia_SOS | 0,100249 | 0,053351 | NA |
| 2b | | abundance | X08150 | DL-4-Hydro | Handgrip | 0,190159 | 0,059685 | NA |
| | 3 | abundance | X08154 | 4-(METHYL | Tibia_length | -0,03205 | 0,052878 | NA |
| | 3 | abundance | X08154 | 4-(METHYL | Radius_length | 0,012608 | 0,071727 | NA |
| | 3 | abundance | X08154 | 4-(METHYL | Radius_SOS | 0,020472 | 0,050676 | NA |
| | 3 | abundance | X08154 | 4-(METHYL | Tibia_SOS | -0,02793 | 0,054184 | NA |
| | 3 | abundance | X08154 | 4-(METHYL | Handgrip | -0,06836 | 0,06175 | NA |
| | 3 | abundance | X08157 | Leu-Val | Tibia_length | 0,079543 | 0,053801 | NA |
| | 3 | abundance | X08157 | Leu-Val | Radius_length | 0,099715 | 0,072419 | NA |
| | 3 | abundance | X08157 | Leu-Val | Radius_SOS | 0,082724 | 0,051617 | NA |

| | | | | | | | |
|---|---|---|---|---|---|---|---|
| 3 | abundance | X08157 | Leu-Val | Tibia_SOS | 0,026612 | 0,054906 | NA |
| 3 | abundance | X08157 | Leu-Val | Handgrip | -0,03994 | 0,061765 | NA |
| 3 | abundance | X08162 | Valylvaline | Tibia_length | 0,028177 | 0,052496 | NA |
| 3 | abundance | X08162 | Valylvaline | Radius_length | 0,101111 | 0,071287 | NA |
| 3 | abundance | X08162 | Valylvaline | Radius_SOS | -0,02423 | 0,05034 | NA |
| 3 | abundance | X08162 | Valylvaline | Tibia_SOS | 0,008968 | 0,054458 | NA |
| 3 | abundance | X08162 | Valylvaline | Handgrip | -0,00206 | 0,061131 | NA |
| 3 | abundance | X08163 | Phenyl D-g | Tibia_length | -0,13879 | 0,052032 | NA |
| 3 | abundance | X08163 | Phenyl D-g | Radius_length | -0,11946 | 0,070958 | NA |
| 3 | abundance | X08163 | Phenyl D-g | Radius_SOS | -0,06064 | 0,050363 | NA |
| 3 | abundance | X08163 | Phenyl D-g | Tibia_SOS | -0,0378 | 0,053649 | NA |
| 3 | abundance | X08163 | Phenyl D-g | Handgrip | -0,06591 | 0,061032 | NA |
| 3 | abundance | X08167 | Lanthionin | Tibia_length | -0,04868 | 0,056008 | NA |
| 3 | abundance | X08167 | Lanthionin | Radius_length | -0,06812 | 0,074178 | NA |
| 3 | abundance | X08167 | Lanthionin | Radius_SOS | -0,09205 | 0,053105 | NA |
| 3 | abundance | X08167 | Lanthionin | Tibia_SOS | -0,05797 | 0,057582 | NA |
| 3 | abundance | X08167 | Lanthionin | Handgrip | -0,08434 | 0,062198 | NA |
| 2b | abundance | X08173 | 9-Methylur | Tibia_length | -0,07874 | 0,054016 | NA |
| 2b | abundance | X08173 | 9-Methylur | Radius_length | -0,11465 | 0,072582 | NA |
| 2b | abundance | X08173 | 9-Methylur | Radius_SOS | 4,19E-04 | 0,051663 | NA |
| 2b | abundance | X08173 | 9-Methylur | Tibia_SOS | -0,05068 | 0,054818 | NA |
| 2b | abundance | X08173 | 9-Methylur | Handgrip | -0,13649 | 0,062226 | NA |
| 2b | abundance | X08188 | 2-Hydroxy- | Tibia_length | 0,121372 | 0,054696 | NA |
| 2b | abundance | X08188 | 2-Hydroxy- | Radius_length | 0,17188 | 0,073077 | NA |
| 2b | abundance | X08188 | 2-Hydroxy- | Radius_SOS | 0,088412 | 0,051731 | NA |
| 2b | abundance | X08188 | 2-Hydroxy- | Tibia_SOS | 0,1031 | 0,056616 | NA |
| 2b | abundance | X08188 | 2-Hydroxy- | Handgrip | 0,147217 | 0,061033 | NA |
| 3 | abundance | X08193 | Seryltyrosi | Tibia_length | -0,14022 | 0,051431 | NA |
| 3 | abundance | X08193 | Seryltyrosi | Radius_length | -0,16715 | 0,069521 | NA |
| 3 | abundance | X08193 | Seryltyrosi | Radius_SOS | -0,10794 | 0,049398 | NA |
| 3 | abundance | X08193 | Seryltyrosi | Tibia_SOS | -0,09366 | 0,05333 | NA |
| 3 | abundance | X08193 | Seryltyrosi | Handgrip | -0,06576 | 0,060683 | NA |
| 2b | abundance | X08204 | 2-Amino-4 | Tibia_length | 0,048487 | 0,049489 | NA |
| 2b | abundance | X08204 | 2-Amino-4 | Radius_length | 0,242443 | 0,064975 | NA |
| 2b | abundance | X08204 | 2-Amino-4 | Radius_SOS | 0,024904 | 0,049017 | NA |
| 2b | abundance | X08204 | 2-Amino-4 | Tibia_SOS | 0,110306 | 0,052166 | NA |
| 2b | abundance | X08204 | 2-Amino-4 | Handgrip | 0,103103 | 0,059873 | NA |
| 3 | abundance | X08212 | LU345300 | Tibia_length | 0,018436 | 0,055477 | NA |
| 3 | abundance | X08212 | LU345300 | Radius_length | 0,068327 | 0,074609 | NA |
| 3 | abundance | X08212 | LU345300 | Radius_SOS | -0,00218 | 0,052782 | NA |
| 3 | abundance | X08212 | LU345300 | Tibia_SOS | 0,026175 | 0,057022 | NA |
| 3 | abundance | X08212 | LU345300 | Handgrip | 0,124035 | 0,063414 | NA |
| 2b | abundance | X08221 | Brilliant blu | Tibia_length | -0,10183 | 0,054113 | NA |
| 2b | abundance | X08221 | Brilliant blu | Radius_length | -0,0729 | 0,073024 | NA |
| 2b | abundance | X08221 | Brilliant blu | Radius_SOS | -0,02722 | 0,051973 | NA |
| 2b | abundance | X08221 | Brilliant blu | Tibia_SOS | 3,64E-04 | 0,054782 | NA |
| 2b | abundance | X08221 | Brilliant blu | Handgrip | -0,12142 | 0,061922 | NA |
| 3 | abundance | X08230 | Coumaron | Tibia_length | 0,122435 | 0,049228 | NA |

| | | | | | | | | |
|---|---|---|---|---|---|---|---|---|
| | 3 | abundance | X08230 | Coumaron | Radius_length | 0,174115 | 0,065867 | NA |
| | 3 | abundance | X08230 | Coumaron | Radius_SOS | 0,072636 | 0,048901 | NA |
| | 3 | abundance | X08230 | Coumaron | Tibia_SOS | 0,13205 | 0,052065 | NA |
| | 3 | abundance | X08230 | Coumaron | Handgrip | 0,068961 | 0,060058 | NA |
| 2b | | abundance | X08235 | 1-Methylhi | Tibia_length | -0,07755 | 0,054589 | NA |
| 2b | | abundance | X08235 | 1-Methylhi | Radius_length | -0,04677 | 0,073723 | NA |
| 2b | | abundance | X08235 | 1-Methylhi | Radius_SOS | -0,02919 | 0,052192 | NA |
| 2b | | abundance | X08235 | 1-Methylhi | Tibia_SOS | -0,0752 | 0,05546 | NA |
| 2b | | abundance | X08235 | 1-Methylhi | Handgrip | 0,04851 | 0,063654 | NA |
| | 3 | abundance | X08244 | Setoclavin | Tibia_length | -0,00506 | 0,050202 | NA |
| | 3 | abundance | X08244 | Setoclavin | Radius_length | 0,087587 | 0,068815 | NA |
| | 3 | abundance | X08244 | Setoclavin | Radius_SOS | 0,026542 | 0,049133 | NA |
| | 3 | abundance | X08244 | Setoclavin | Tibia_SOS | 0,048187 | 0,052507 | NA |
| | 3 | abundance | X08244 | Setoclavin | Handgrip | 0,045502 | 0,06077 | NA |
| | 3 | abundance | X08270 | Butabarbit | Tibia_length | -0,11266 | 0,052964 | NA |
| | 3 | abundance | X08270 | Butabarbit | Radius_length | -0,15804 | 0,071438 | NA |
| | 3 | abundance | X08270 | Butabarbit | Radius_SOS | -0,02852 | 0,050633 | NA |
| | 3 | abundance | X08270 | Butabarbit | Tibia_SOS | -0,04872 | 0,054672 | NA |
| | 3 | abundance | X08270 | Butabarbit | Handgrip | -0,01779 | 0,061745 | NA |
| 2b | | abundance | X08274 | Mevalonic | Tibia_length | -0,06393 | 0,05403 | NA |
| 2b | | abundance | X08274 | Mevalonic | Radius_length | -6,00E-04 | 0,073239 | NA |
| 2b | | abundance | X08274 | Mevalonic | Radius_SOS | 0,015856 | 0,051418 | NA |
| 2b | | abundance | X08274 | Mevalonic | Tibia_SOS | 0,02605 | 0,056307 | NA |
| 2b | | abundance | X08274 | Mevalonic | Handgrip | 0,064517 | 0,061424 | NA |
| | 3 | abundance | X08277 | adrenaline | Tibia_length | 0,049735 | 0,056359 | NA |
| | 3 | abundance | X08277 | adrenaline | Radius_length | 0,096629 | 0,073917 | NA |
| | 3 | abundance | X08277 | adrenaline | Radius_SOS | -0,03784 | 0,053406 | NA |
| | 3 | abundance | X08277 | adrenaline | Tibia_SOS | -0,04926 | 0,057378 | NA |
| | 3 | abundance | X08277 | adrenaline | Handgrip | -0,01728 | 0,062098 | NA |
| | 3 | abundance | X08279 | DNOP | Tibia_length | 0,042791 | 0,052578 | NA |
| | 3 | abundance | X08279 | DNOP | Radius_length | 0,172601 | 0,070401 | NA |
| | 3 | abundance | X08279 | DNOP | Radius_SOS | 0,055804 | 0,050246 | NA |
| | 3 | abundance | X08279 | DNOP | Tibia_SOS | -0,01718 | 0,05376 | NA |
| | 3 | abundance | X08279 | DNOP | Handgrip | 0,055043 | 0,062092 | NA |
| | 3 | abundance | X08288 | Capryloylg | Tibia_length | -0,12484 | 0,05771 | NA |
| | 3 | abundance | X08288 | Capryloylg | Radius_length | -0,03818 | 0,075222 | NA |
| | 3 | abundance | X08288 | Capryloylg | Radius_SOS | -0,08278 | 0,054739 | NA |
| | 3 | abundance | X08288 | Capryloylg | Tibia_SOS | 0,044086 | 0,06052 | NA |
| | 3 | abundance | X08288 | Capryloylg | Handgrip | -0,04358 | 0,060136 | NA |
| 2b | | abundance | X08292 | Lidocaine | Tibia_length | -0,05811 | 0,051232 | NA |
| 2b | | abundance | X08292 | Lidocaine | Radius_length | 0,033499 | 0,070092 | NA |
| 2b | | abundance | X08292 | Lidocaine | Radius_SOS | -0,03218 | 0,04957 | NA |
| 2b | | abundance | X08292 | Lidocaine | Tibia_SOS | 0,040178 | 0,053515 | NA |
| 2b | | abundance | X08292 | Lidocaine | Handgrip | 0,013338 | 0,060837 | NA |
| 2b | | abundance | X08298 | N6,N6,N6- | Tibia_length | -0,05541 | 0,050295 | NA |
| 2b | | abundance | X08298 | N6,N6,N6- | Radius_length | -0,03348 | 0,068608 | NA |
| 2b | | abundance | X08298 | N6,N6,N6- | Radius_SOS | 0,010431 | 0,049151 | NA |
| 2b | | abundance | X08298 | N6,N6,N6- | Tibia_SOS | -0,07537 | 0,052353 | NA |

| | | | | | | | |
|---|---|---|---|---|---|---|---|
| 2b | abundance | X08298 | N6,N6,N6- | Handgrip | -0,02558 | 0,060288 | NA |
| 3 | abundance | X08300 | Indole-3-ac | Tibia_length | 0,044443 | 0,050343 | NA |
| 3 | abundance | X08300 | Indole-3-ac | Radius_length | -0,03738 | 0,068725 | NA |
| 3 | abundance | X08300 | Indole-3-ac | Radius_SOS | -0,05459 | 0,049099 | NA |
| 3 | abundance | X08300 | Indole-3-ac | Tibia_SOS | -0,00714 | 0,052838 | NA |
| 3 | abundance | X08300 | Indole-3-ac | Handgrip | -0,09283 | 0,060127 | NA |
| 2a | abundance | X08305 | Prolylhydrc | Tibia_length | -0,07696 | 0,055202 | NA |
| 2a | abundance | X08305 | Prolylhydrc | Radius_length | -0,15321 | 0,073273 | NA |
| 2a | abundance | X08305 | Prolylhydrc | Radius_SOS | -0,09079 | 0,052356 | NA |
| 2a | abundance | X08305 | Prolylhydrc | Tibia_SOS | -0,09629 | 0,055258 | NA |
| 2a | abundance | X08305 | Prolylhydrc | Handgrip | -0,07414 | 0,061585 | NA |
| 2b | abundance | X08306 | 1-Methylxa | Tibia_length | -0,03353 | 0,054302 | NA |
| 2b | abundance | X08306 | 1-Methylxa | Radius_length | -0,14563 | 0,072435 | NA |
| 2b | abundance | X08306 | 1-Methylxa | Radius_SOS | 0,008471 | 0,051427 | NA |
| 2b | abundance | X08306 | 1-Methylxa | Tibia_SOS | -0,01217 | 0,05471 | NA |
| 2b | abundance | X08306 | 1-Methylxa | Handgrip | -0,00907 | 0,061973 | NA |
| 3 | abundance | X08311 | Methyl 3-fc | Tibia_length | -0,16881 | 0,054349 | NA |
| 3 | abundance | X08311 | Methyl 3-fc | Radius_length | -0,08297 | 0,073049 | NA |
| 3 | abundance | X08311 | Methyl 3-fc | Radius_SOS | 0,04738 | 0,05185 | NA |
| 3 | abundance | X08311 | Methyl 3-fc | Tibia_SOS | 0,010144 | 0,054502 | NA |
| 3 | abundance | X08311 | Methyl 3-fc | Handgrip | -0,01905 | 0,061306 | NA |
| 2b | abundance | X08317 | 8-Hydroxyc | Tibia_length | -2,42E-04 | 0,05573 | NA |
| 2b | abundance | X08317 | 8-Hydroxyc | Radius_length | -0,01227 | 0,073825 | NA |
| 2b | abundance | X08317 | 8-Hydroxyc | Radius_SOS | 0,007313 | 0,052913 | NA |
| 2b | abundance | X08317 | 8-Hydroxyc | Tibia_SOS | 0,050635 | 0,055926 | NA |
| 2b | abundance | X08317 | 8-Hydroxyc | Handgrip | -0,02061 | 0,06295 | NA |
| 3 | abundance | X08322 | Glycylproly | Tibia_length | -0,10256 | 0,052131 | NA |
| 3 | abundance | X08322 | Glycylproly | Radius_length | -0,11943 | 0,070735 | NA |
| 3 | abundance | X08322 | Glycylproly | Radius_SOS | 0,030865 | 0,050057 | NA |
| 3 | abundance | X08322 | Glycylproly | Tibia_SOS | -0,04206 | 0,053357 | NA |
| 3 | abundance | X08322 | Glycylproly | Handgrip | 0,060435 | 0,060792 | NA |
| 2b | abundance | X08323 | (15Z)-9,12 | Tibia_length | 0,102356 | 0,049525 | NA |
| 2b | abundance | X08323 | (15Z)-9,12 | Radius_length | 0,148534 | 0,067078 | NA |
| 2b | abundance | X08323 | (15Z)-9,12 | Radius_SOS | 0,016637 | 0,049035 | NA |
| 2b | abundance | X08323 | (15Z)-9,12 | Tibia_SOS | -0,02269 | 0,05256 | NA |
| 2b | abundance | X08323 | (15Z)-9,12 | Handgrip | 0,066436 | 0,060119 | NA |
| 3 | abundance | X08326 | 2-(1,3-Ben | Tibia_length | -0,06568 | 0,051452 | NA |
| 3 | abundance | X08326 | 2-(1,3-Ben | Radius_length | -3,35E-04 | 0,070532 | NA |
| 3 | abundance | X08326 | 2-(1,3-Ben | Radius_SOS | 0,015755 | 0,049815 | NA |
| 3 | abundance | X08326 | 2-(1,3-Ben | Tibia_SOS | -0,05687 | 0,052938 | NA |
| 3 | abundance | X08326 | 2-(1,3-Ben | Handgrip | -0,0192 | 0,061573 | NA |
| 3 | abundance | X08338 | Methyl [9-( | Tibia_length | 0,012541 | 0,059461 | NA |
| 3 | abundance | X08338 | Methyl [9-( | Radius_length | 0,084289 | 0,075456 | NA |
| 3 | abundance | X08338 | Methyl [9-( | Radius_SOS | -0,02255 | 0,056628 | NA |
| 3 | abundance | X08338 | Methyl [9-( | Tibia_SOS | 0,129346 | 0,058851 | NA |
| 3 | abundance | X08338 | Methyl [9-( | Handgrip | 0,019951 | 0,062927 | NA |
| 3 | abundance | X08341 | Midodrine | Tibia_length | -0,06762 | 0,055373 | NA |
| 3 | abundance | X08341 | Midodrine | Radius_length | -0,08928 | 0,074007 | NA |

| | | | | | | | | |
|---|---|---|---|---|---|---|---|---|
| | 3 | abundance | X08341 | Midodrine | Radius_SOS | 0,053699 | 0,052698 | NA |
| | 3 | abundance | X08341 | Midodrine | Tibia_SOS | -0,09865 | 0,055512 | NA |
| | 3 | abundance | X08341 | Midodrine | Handgrip | -0,10156 | 0,062973 | NA |
| 2b | | abundance | X08352 | 7-[4-(tert-b | Tibia_length | 0,05658 | 0,056834 | NA |
| 2b | | abundance | X08352 | 7-[4-(tert-b | Radius_length | 0,034342 | 0,074735 | NA |
| 2b | | abundance | X08352 | 7-[4-(tert-b | Radius_SOS | -0,01964 | 0,054273 | NA |
| 2b | | abundance | X08352 | 7-[4-(tert-b | Tibia_SOS | -0,01872 | 0,058151 | NA |
| 2b | | abundance | X08352 | 7-[4-(tert-b | Handgrip | -0,0152 | 0,063566 | NA |
| | 3 | abundance | X08365 | Lysylvaline | Tibia_length | -0,03871 | 0,051214 | NA |
| | 3 | abundance | X08365 | Lysylvaline | Radius_length | -0,07481 | 0,070006 | NA |
| | 3 | abundance | X08365 | Lysylvaline | Radius_SOS | -0,00329 | 0,049586 | NA |
| | 3 | abundance | X08365 | Lysylvaline | Tibia_SOS | -0,07525 | 0,053233 | NA |
| | 3 | abundance | X08365 | Lysylvaline | Handgrip | 0,011568 | 0,06076 | NA |
| 2b | | abundance | X08367 | 11-dehydro | Tibia_length | -0,03525 | 0,057438 | NA |
| 2b | | abundance | X08367 | 11-dehydro | Radius_length | 0,053907 | 0,074711 | NA |
| 2b | | abundance | X08367 | 11-dehydro | Radius_SOS | 0,072632 | 0,054237 | NA |
| 2b | | abundance | X08367 | 11-dehydro | Tibia_SOS | -0,05084 | 0,057899 | NA |
| 2b | | abundance | X08367 | 11-dehydro | Handgrip | -0,03451 | 0,063629 | NA |
| | 3 | abundance | X08373 | 2-Hydroxy- | Tibia_length | -0,00621 | 0,056722 | NA |
| | 3 | abundance | X08373 | 2-Hydroxy- | Radius_length | 0,068745 | 0,074253 | NA |
| | 3 | abundance | X08373 | 2-Hydroxy- | Radius_SOS | 0,033771 | 0,053842 | NA |
| | 3 | abundance | X08373 | 2-Hydroxy- | Tibia_SOS | 0,098109 | 0,056104 | NA |
| | 3 | abundance | X08373 | 2-Hydroxy- | Handgrip | 0,038367 | 0,063442 | NA |
| | 3 | abundance | X08386 | 3b-Hydroxy | Tibia_length | -0,02409 | 0,051484 | NA |
| | 3 | abundance | X08386 | 3b-Hydroxy | Radius_length | -0,03328 | 0,070861 | NA |
| | 3 | abundance | X08386 | 3b-Hydroxy | Radius_SOS | 0,114054 | 0,049557 | NA |
| | 3 | abundance | X08386 | 3b-Hydroxy | Tibia_SOS | 0,022454 | 0,053425 | NA |
| | 3 | abundance | X08386 | 3b-Hydroxy | Handgrip | 0,053688 | 0,060896 | NA |
| | 3 | abundance | X08387 | 4-[(2E,4Z)- | Tibia_length | 0,005654 | 0,05721 | NA |
| | 3 | abundance | X08387 | 4-[(2E,4Z)- | Radius_length | -0,00407 | 0,074683 | NA |
| | 3 | abundance | X08387 | 4-[(2E,4Z)- | Radius_SOS | 0,02952 | 0,054418 | NA |
| | 3 | abundance | X08387 | 4-[(2E,4Z)- | Tibia_SOS | 0,10648 | 0,057283 | NA |
| | 3 | abundance | X08387 | 4-[(2E,4Z)- | Handgrip | -0,01543 | 0,060186 | NA |
| 2b | | abundance | X08412 | 2-Hydroxyp | Tibia_length | 0,053205 | 0,051049 | NA |
| 2b | | abundance | X08412 | 2-Hydroxyp | Radius_length | 0,054675 | 0,07036 | NA |
| 2b | | abundance | X08412 | 2-Hydroxyp | Radius_SOS | 0,061382 | 0,04944 | NA |
| 2b | | abundance | X08412 | 2-Hydroxyp | Tibia_SOS | 0,11376 | 0,052907 | NA |
| 2b | | abundance | X08412 | 2-Hydroxyp | Handgrip | 0,023222 | 0,061626 | NA |
| | 3 | abundance | X08417 | asn-pro | Tibia_length | -0,05651 | 0,054612 | NA |
| | 3 | abundance | X08417 | asn-pro | Radius_length | -0,01856 | 0,073274 | NA |
| | 3 | abundance | X08417 | asn-pro | Radius_SOS | 0,017093 | 0,052088 | NA |
| | 3 | abundance | X08417 | asn-pro | Tibia_SOS | 0,010722 | 0,055199 | NA |
| | 3 | abundance | X08417 | asn-pro | Handgrip | -0,08079 | 0,063 | NA |
| | 1 | abundance | X08420 | Dodecane | Tibia_length | 0,086289 | 0,05856 | NA |
| | 1 | abundance | X08420 | Dodecane | Radius_length | 0,08195 | 0,075326 | NA |
| | 1 | abundance | X08420 | Dodecane | Radius_SOS | 0,105434 | 0,055697 | NA |
| | 1 | abundance | X08420 | Dodecane | Tibia_SOS | 0,038174 | 0,058303 | NA |
| | 1 | abundance | X08420 | Dodecane | Handgrip | 0,02143 | 0,062871 | NA |

| | | | | | | | |
|---|---|---|---|---|---|---|---|
| 3 | abundance | X08421 | Cys-tyr | Tibia_length | -0,06938 | 0,058293 | NA |
| 3 | abundance | X08421 | Cys-tyr | Radius_length | 0,034139 | 0,075425 | NA |
| 3 | abundance | X08421 | Cys-tyr | Radius_SOS | -0,16071 | 0,055613 | NA |
| 3 | abundance | X08421 | Cys-tyr | Tibia_SOS | -0,05501 | 0,058722 | NA |
| 3 | abundance | X08421 | Cys-tyr | Handgrip | 0,073757 | 0,063418 | NA |
| 3 | abundance | X08422 | N-(3-aceta | Tibia_length | -0,1235 | 0,051033 | NA |
| 3 | abundance | X08422 | N-(3-aceta | Radius_length | -0,08204 | 0,070524 | NA |
| 3 | abundance | X08422 | N-(3-aceta | Radius_SOS | -0,05349 | 0,049798 | NA |
| 3 | abundance | X08422 | N-(3-aceta | Tibia_SOS | 0,073877 | 0,052635 | NA |
| 3 | abundance | X08422 | N-(3-aceta | Handgrip | 0,055945 | 0,061442 | NA |
| 3 | abundance | X08425 | MFCD1297 | Tibia_length | 0,042021 | 0,052629 | NA |
| 3 | abundance | X08425 | MFCD1297 | Radius_length | 0,008149 | 0,071705 | NA |
| 3 | abundance | X08425 | MFCD1297 | Radius_SOS | 0,027764 | 0,050475 | NA |
| 3 | abundance | X08425 | MFCD1297 | Tibia_SOS | -0,00482 | 0,054026 | NA |
| 3 | abundance | X08425 | MFCD1297 | Handgrip | -0,07118 | 0,061908 | NA |
| 3 | abundance | X08436 | N,N-Dimet | Tibia_length | 0,080413 | 0,054827 | NA |
| 3 | abundance | X08436 | N,N-Dimet | Radius_length | 0,095793 | 0,073082 | NA |
| 3 | abundance | X08436 | N,N-Dimet | Radius_SOS | 0,032614 | 0,05187 | NA |
| 3 | abundance | X08436 | N,N-Dimet | Tibia_SOS | 0,026321 | 0,055864 | NA |
| 3 | abundance | X08436 | N,N-Dimet | Handgrip | 0,116785 | 0,062483 | NA |
| 3 | abundance | X08439 | Leucyltyros | Tibia_length | -0,0441 | 0,050684 | NA |
| 3 | abundance | X08439 | Leucyltyros | Radius_length | -0,14423 | 0,068769 | NA |
| 3 | abundance | X08439 | Leucyltyros | Radius_SOS | 9,73E-04 | 0,049321 | NA |
| 3 | abundance | X08439 | Leucyltyros | Tibia_SOS | -0,07756 | 0,052612 | NA |
| 3 | abundance | X08439 | Leucyltyros | Handgrip | -0,09822 | 0,060888 | NA |
| 3 | abundance | X08451 | L-gamma-( | Tibia_length | -0,08127 | 0,053098 | NA |
| 3 | abundance | X08451 | L-gamma-( | Radius_length | -0,00382 | 0,072147 | NA |
| 3 | abundance | X08451 | L-gamma-( | Radius_SOS | -0,01026 | 0,050896 | NA |
| 3 | abundance | X08451 | L-gamma-( | Tibia_SOS | -0,02356 | 0,054521 | NA |
| 3 | abundance | X08451 | L-gamma-( | Handgrip | -0,02657 | 0,061225 | NA |
| 1 | abundance | X08460 | Î±-Muricho | Tibia_length | 0,002963 | 0,052972 | NA |
| 1 | abundance | X08460 | Î±-Muricho | Radius_length | 0,023307 | 0,072136 | NA |
| 1 | abundance | X08460 | Î±-Muricho | Radius_SOS | 0,088061 | 0,05052 | NA |
| 1 | abundance | X08460 | Î±-Muricho | Tibia_SOS | -0,03573 | 0,054545 | NA |
| 1 | abundance | X08460 | Î±-Muricho | Handgrip | 0,011276 | 0,061313 | NA |
| 3 | abundance | X08461 | Dihydrouri | Tibia_length | -0,13522 | 0,056541 | NA |
| 3 | abundance | X08461 | Dihydrouri | Radius_length | -0,04837 | 0,075387 | NA |
| 3 | abundance | X08461 | Dihydrouri | Radius_SOS | -0,09602 | 0,054209 | NA |
| 3 | abundance | X08461 | Dihydrouri | Tibia_SOS | -0,0618 | 0,057454 | NA |
| 3 | abundance | X08461 | Dihydrouri | Handgrip | -0,02031 | 0,063398 | NA |
| 3 | abundance | X08475 | 1-(4-Aminc | Tibia_length | -0,11308 | 0,053297 | NA |
| 3 | abundance | X08475 | 1-(4-Aminc | Radius_length | -0,15509 | 0,071536 | NA |
| 3 | abundance | X08475 | 1-(4-Aminc | Radius_SOS | -0,0849 | 0,050754 | NA |
| 3 | abundance | X08475 | 1-(4-Aminc | Tibia_SOS | -0,08929 | 0,053909 | NA |
| 3 | abundance | X08475 | 1-(4-Aminc | Handgrip | -0,02835 | 0,060428 | NA |
| 3 | abundance | X08476 | Coprine | Tibia_length | 0,070297 | 0,053869 | NA |
| 3 | abundance | X08476 | Coprine | Radius_length | 0,07389 | 0,072496 | NA |
| 3 | abundance | X08476 | Coprine | Radius_SOS | 0,071998 | 0,05141 | NA |

| | | | | | | | |
|---|---|---|---|---|---|---|---|
| 3 | abundance | X08476 | Coprine | Tibia_SOS | 0,042757 | 0,054555 | NA |
| 3 | abundance | X08476 | Coprine | Handgrip | 0,1153 | 0,061216 | NA |
| 3 | abundance | X08478 | 1-(beta-D-I | Tibia_length | -0,1019 | 0,05641 | NA |
| 3 | abundance | X08478 | 1-(beta-D-I | Radius_length | -0,10432 | 0,073979 | NA |
| 3 | abundance | X08478 | 1-(beta-D-I | Radius_SOS | -0,07437 | 0,053683 | NA |
| 3 | abundance | X08478 | 1-(beta-D-I | Tibia_SOS | -0,09458 | 0,055549 | NA |
| 3 | abundance | X08478 | 1-(beta-D-I | Handgrip | -0,10999 | 0,063257 | NA |
| 3 | abundance | X08485 | Isoprenalir | Tibia_length | 0,105656 | 0,052613 | NA |
| 3 | abundance | X08485 | Isoprenalir | Radius_length | 0,082483 | 0,071457 | NA |
| 3 | abundance | X08485 | Isoprenalir | Radius_SOS | 0,005511 | 0,050504 | NA |
| 3 | abundance | X08485 | Isoprenalir | Tibia_SOS | 0,043541 | 0,054141 | NA |
| 3 | abundance | X08485 | Isoprenalir | Handgrip | 0,021274 | 0,06178 | NA |
| 3 | abundance | X08486 | (2E,6E)-9-[ | Tibia_length | 0,100846 | 0,05286 | NA |
| 3 | abundance | X08486 | (2E,6E)-9-[ | Radius_length | 0,066597 | 0,072106 | NA |
| 3 | abundance | X08486 | (2E,6E)-9-[ | Radius_SOS | 0,007055 | 0,050552 | NA |
| 3 | abundance | X08486 | (2E,6E)-9-[ | Tibia_SOS | 0,06931 | 0,0543 | NA |
| 3 | abundance | X08486 | (2E,6E)-9-[ | Handgrip | -0,06029 | 0,060797 | NA |
| 2a | abundance | X08504 | Indole-3-ca | Tibia_length | -0,09774 | 0,051542 | NA |
| 2a | abundance | X08504 | Indole-3-ca | Radius_length | 0,033397 | 0,071118 | NA |
| 2a | abundance | X08504 | Indole-3-ca | Radius_SOS | -0,02305 | 0,050003 | NA |
| 2a | abundance | X08504 | Indole-3-ca | Tibia_SOS | 0,106862 | 0,052899 | NA |
| 2a | abundance | X08504 | Indole-3-ca | Handgrip | -0,02879 | 0,061603 | NA |
| 3 | abundance | X08509 | Piceid | Tibia_length | -0,01764 | 0,050602 | NA |
| 3 | abundance | X08509 | Piceid | Radius_length | -0,07442 | 0,068996 | NA |
| 3 | abundance | X08509 | Piceid | Radius_SOS | 0,022225 | 0,049279 | NA |
| 3 | abundance | X08509 | Piceid | Tibia_SOS | 0,016067 | 0,053037 | NA |
| 3 | abundance | X08509 | Piceid | Handgrip | 0,002182 | 0,061003 | NA |
| 2b | abundance | X08512 | Bile acid I ( | Tibia_length | 0,026762 | 0,052313 | NA |
| 2b | abundance | X08512 | Bile acid I ( | Radius_length | 1,74E-04 | 0,071691 | NA |
| 2b | abundance | X08512 | Bile acid I ( | Radius_SOS | 0,114951 | 0,050089 | NA |
| 2b | abundance | X08512 | Bile acid I ( | Tibia_SOS | -0,00187 | 0,053625 | NA |
| 2b | abundance | X08512 | Bile acid I ( | Handgrip | 0,039189 | 0,061097 | NA |
| 3 | abundance | X08514 | N-Ethylpro | Tibia_length | -0,05126 | 0,049541 | NA |
| 3 | abundance | X08514 | N-Ethylpro | Radius_length | 0,015882 | 0,066989 | NA |
| 3 | abundance | X08514 | N-Ethylpro | Radius_SOS | 0,023494 | 0,049016 | NA |
| 3 | abundance | X08514 | N-Ethylpro | Tibia_SOS | -0,08299 | 0,052311 | NA |
| 3 | abundance | X08514 | N-Ethylpro | Handgrip | 0,067024 | 0,060058 | NA |
| 3 | abundance | X08525 | MFCD1869 | Tibia_length | -0,08698 | 0,051972 | NA |
| 3 | abundance | X08525 | MFCD1869 | Radius_length | 0,007672 | 0,071123 | NA |
| 3 | abundance | X08525 | MFCD1869 | Radius_SOS | 0,035193 | 0,050126 | NA |
| 3 | abundance | X08525 | MFCD1869 | Tibia_SOS | 0,016873 | 0,053242 | NA |
| 3 | abundance | X08525 | MFCD1869 | Handgrip | 0,038093 | 0,061281 | NA |
| 3 | abundance | X08529 | 7,8-Diamir | Tibia_length | -0,04388 | 0,049768 | NA |
| 3 | abundance | X08529 | 7,8-Diamir | Radius_length | -0,03323 | 0,067502 | NA |
| 3 | abundance | X08529 | 7,8-Diamir | Radius_SOS | -0,01618 | 0,049037 | NA |
| 3 | abundance | X08529 | 7,8-Diamir | Tibia_SOS | -0,07977 | 0,05232 | NA |
| 3 | abundance | X08529 | 7,8-Diamir | Handgrip | -0,02716 | 0,060293 | NA |
| 3 | abundance | X08533 | tyramine si | Tibia_length | -0,11943 | 0,055846 | NA |

| | | | | | | | | |
|---|---|---|---|---|---|---|---|---|
| | 3 | abundance | X08533 | tyramine sı | Radius_length | -0,15901 | 0,073522 | NA |
| | 3 | abundance | X08533 | tyramine sı | Radius_SOS | -0,01407 | 0,053488 | NA |
| | 3 | abundance | X08533 | tyramine sı | Tibia_SOS | -0,03323 | 0,056566 | NA |
| | 3 | abundance | X08533 | tyramine sı | Handgrip | -0,094 | 0,061636 | NA |
| 2b | | abundance | X08535 | Nitrosohep | Tibia_length | -0,00909 | 0,049124 | NA |
| 2b | | abundance | X08535 | Nitrosohep | Radius_length | 0,037569 | 0,065028 | NA |
| 2b | | abundance | X08535 | Nitrosohep | Radius_SOS | 0,03593 | 0,04916 | NA |
| 2b | | abundance | X08535 | Nitrosohep | Tibia_SOS | -0,04315 | 0,052645 | NA |
| 2b | | abundance | X08535 | Nitrosohep | Handgrip | 0,019273 | 0,060268 | NA |
| 2b | | abundance | X08546 | 1,3,7-Trim | Tibia_length | -0,09986 | 0,051618 | NA |
| 2b | | abundance | X08546 | 1,3,7-Trim | Radius_length | -0,06316 | 0,070617 | NA |
| 2b | | abundance | X08546 | 1,3,7-Trim | Radius_SOS | 0,013394 | 0,049947 | NA |
| 2b | | abundance | X08546 | 1,3,7-Trim | Tibia_SOS | -0,04789 | 0,053142 | NA |
| 2b | | abundance | X08546 | 1,3,7-Trim | Handgrip | -0,05072 | 0,061014 | NA |
| | 1 | abundance | X08548 | N2-Acetyll | Tibia_length | 0,097855 | 0,050775 | NA |
| | 1 | abundance | X08548 | N2-Acetyll | Radius_length | 0,142857 | 0,069007 | NA |
| | 1 | abundance | X08548 | N2-Acetyll | Radius_SOS | 0,021342 | 0,049362 | NA |
| | 1 | abundance | X08548 | N2-Acetyll | Tibia_SOS | 0,098081 | 0,052408 | NA |
| | 1 | abundance | X08548 | N2-Acetyll | Handgrip | 0,053602 | 0,060123 | NA |
| | 3 | abundance | X08549 | 3-Methylsu | Tibia_length | 0,08809 | 0,051064 | NA |
| | 3 | abundance | X08549 | 3-Methylsu | Radius_length | 0,166508 | 0,068927 | NA |
| | 3 | abundance | X08549 | 3-Methylsu | Radius_SOS | 0,025681 | 0,049438 | NA |
| | 3 | abundance | X08549 | 3-Methylsu | Tibia_SOS | 0,131949 | 0,052548 | NA |
| | 3 | abundance | X08549 | 3-Methylsu | Handgrip | 0,108729 | 0,060195 | NA |
| | 3 | abundance | X08561 | N-(Carboxy | Tibia_length | -0,06835 | 0,052997 | NA |
| | 3 | abundance | X08561 | N-(Carboxy | Radius_length | 0,122568 | 0,071905 | NA |
| | 3 | abundance | X08561 | N-(Carboxy | Radius_SOS | 0,081871 | 0,050662 | NA |
| | 3 | abundance | X08561 | N-(Carboxy | Tibia_SOS | 0,078007 | 0,054776 | NA |
| | 3 | abundance | X08561 | N-(Carboxy | Handgrip | 0,060315 | 0,062232 | NA |
| | 3 | abundance | X08578 | tert-Butyl 3 | Tibia_length | 0,020583 | 0,054508 | NA |
| | 3 | abundance | X08578 | tert-Butyl 3 | Radius_length | 0,066164 | 0,072951 | NA |
| | 3 | abundance | X08578 | tert-Butyl 3 | Radius_SOS | 0,006159 | 0,05177 | NA |
| | 3 | abundance | X08578 | tert-Butyl 3 | Tibia_SOS | -0,01966 | 0,055236 | NA |
| | 3 | abundance | X08578 | tert-Butyl 3 | Handgrip | 0,057245 | 0,061797 | NA |
| | 3 | abundance | X08584 | meticillin | Tibia_length | -0,04979 | 0,056601 | NA |
| | 3 | abundance | X08584 | meticillin | Radius_length | -0,03805 | 0,074587 | NA |
| | 3 | abundance | X08584 | meticillin | Radius_SOS | -0,03673 | 0,054154 | NA |
| | 3 | abundance | X08584 | meticillin | Tibia_SOS | -0,02036 | 0,055754 | NA |
| | 3 | abundance | X08584 | meticillin | Handgrip | -0,03735 | 0,062185 | NA |
| | 3 | abundance | X08588 | 2-Acetami | Tibia_length | 0,06508 | 0,053645 | NA |
| | 3 | abundance | X08588 | 2-Acetami | Radius_length | 0,111555 | 0,072632 | NA |
| | 3 | abundance | X08588 | 2-Acetami | Radius_SOS | -0,01325 | 0,051293 | NA |
| | 3 | abundance | X08588 | 2-Acetami | Tibia_SOS | 0,055023 | 0,054585 | NA |
| | 3 | abundance | X08588 | 2-Acetami | Handgrip | 0,103506 | 0,062641 | NA |
| | 1 | abundance | X08593 | Uric acid | Tibia_length | 0,095544 | 0,052993 | NA |
| | 1 | abundance | X08593 | Uric acid | Radius_length | 0,065408 | 0,072175 | NA |
| | 1 | abundance | X08593 | Uric acid | Radius_SOS | 0,029681 | 0,050865 | NA |
| | 1 | abundance | X08593 | Uric acid | Tibia_SOS | 0,089129 | 0,054408 | NA |

| | | | | | | | | |
|---|---|---|---|---|---|---|---|---|
| | 1 | abundance | X08593 | Uric acid | Handgrip | -0,03096 | 0,061278 | NA |
| | 3 | abundance | X08594 | 7-ketodeo | Tibia_length | 0,007638 | 0,054571 | NA |
| | 3 | abundance | X08594 | 7-ketodeo | Radius_length | -0,044 | 0,073011 | NA |
| | 3 | abundance | X08594 | 7-ketodeo | Radius_SOS | 0,044125 | 0,051893 | NA |
| | 3 | abundance | X08594 | 7-ketodeo | Tibia_SOS | -0,06891 | 0,05513 | NA |
| | 3 | abundance | X08594 | 7-ketodeo | Handgrip | -0,04881 | 0,062423 | NA |
| | 3 | abundance | X08600 | N-Propiony | Tibia_length | 0,104381 | 0,051681 | NA |
| | 3 | abundance | X08600 | N-Propiony | Radius_length | 0,032975 | 0,070747 | NA |
| | 3 | abundance | X08600 | N-Propiony | Radius_SOS | -0,01462 | 0,049947 | NA |
| | 3 | abundance | X08600 | N-Propiony | Tibia_SOS | 0,070451 | 0,053319 | NA |
| | 3 | abundance | X08600 | N-Propiony | Handgrip | -0,05108 | 0,061508 | NA |
| | 3 | abundance | X08606 | Butabarbit | Tibia_length | -0,03719 | 0,051246 | NA |
| | 3 | abundance | X08606 | Butabarbit | Radius_length | -0,0693 | 0,070183 | NA |
| | 3 | abundance | X08606 | Butabarbit | Radius_SOS | -0,05221 | 0,049585 | NA |
| | 3 | abundance | X08606 | Butabarbit | Tibia_SOS | -0,02708 | 0,053671 | NA |
| | 3 | abundance | X08606 | Butabarbit | Handgrip | -0,00988 | 0,060664 | NA |
| | 3 | abundance | X08610 | Leucyltyros | Tibia_length | -0,12827 | 0,05042 | NA |
| | 3 | abundance | X08610 | Leucyltyros | Radius_length | -0,0675 | 0,069047 | NA |
| | 3 | abundance | X08610 | Leucyltyros | Radius_SOS | 0,022208 | 0,049295 | NA |
| | 3 | abundance | X08610 | Leucyltyros | Tibia_SOS | -0,09084 | 0,052632 | NA |
| | 3 | abundance | X08610 | Leucyltyros | Handgrip | -0,01089 | 0,060395 | NA |
| | 3 | abundance | X08616 | MFCD0002 | Tibia_length | 0,004225 | 0,05044 | NA |
| | 3 | abundance | X08616 | MFCD0002 | Radius_length | -0,03195 | 0,068972 | NA |
| | 3 | abundance | X08616 | MFCD0002 | Radius_SOS | 0,005928 | 0,049217 | NA |
| | 3 | abundance | X08616 | MFCD0002 | Tibia_SOS | 0,076103 | 0,052572 | NA |
| | 3 | abundance | X08616 | MFCD0002 | Handgrip | 0,150695 | 0,060157 | NA |
| | 3 | abundance | X08625 | Hept-2-ulo | Tibia_length | -0,02249 | 0,054553 | NA |
| | 3 | abundance | X08625 | Hept-2-ulo | Radius_length | 0,099938 | 0,073273 | NA |
| | 3 | abundance | X08625 | Hept-2-ulo | Radius_SOS | 0,038171 | 0,051843 | NA |
| | 3 | abundance | X08625 | Hept-2-ulo | Tibia_SOS | 0,004525 | 0,055811 | NA |
| | 3 | abundance | X08625 | Hept-2-ulo | Handgrip | 0,007046 | 0,061731 | NA |
| 2b | | abundance | X08634 | 1,5-Isoquir | Tibia_length | -0,03348 | 0,051237 | NA |
| 2b | | abundance | X08634 | 1,5-Isoquir | Radius_length | -0,08654 | 0,069907 | NA |
| 2b | | abundance | X08634 | 1,5-Isoquir | Radius_SOS | 0,078391 | 0,049386 | NA |
| 2b | | abundance | X08634 | 1,5-Isoquir | Tibia_SOS | -0,05234 | 0,05315 | NA |
| 2b | | abundance | X08634 | 1,5-Isoquir | Handgrip | 0,010771 | 0,060926 | NA |
| | 3 | abundance | X08639 | hypaphorir | Tibia_length | -0,13346 | 0,048768 | NA |
| | 3 | abundance | X08639 | hypaphorir | Radius_length | -0,151 | 0,064992 | NA |
| | 3 | abundance | X08639 | hypaphorir | Radius_SOS | -0,03314 | 0,049091 | NA |
| | 3 | abundance | X08639 | hypaphorir | Tibia_SOS | 0,059356 | 0,052567 | NA |
| | 3 | abundance | X08639 | hypaphorir | Handgrip | 0,049493 | 0,060138 | NA |
| | 3 | abundance | X08641 | meprobam | Tibia_length | -0,07479 | 0,055189 | NA |
| | 3 | abundance | X08641 | meprobam | Radius_length | -0,00709 | 0,07407 | NA |
| | 3 | abundance | X08641 | meprobam | Radius_SOS | -0,00521 | 0,052761 | NA |
| | 3 | abundance | X08641 | meprobam | Tibia_SOS | -0,00203 | 0,057322 | NA |
| | 3 | abundance | X08641 | meprobam | Handgrip | -0,07826 | 0,063269 | NA |
| 2b | | abundance | X08644 | 2,5-Dimetl | Tibia_length | 0,006138 | 0,051283 | NA |
| 2b | | abundance | X08644 | 2,5-Dimetl | Radius_length | -0,17277 | 0,069305 | NA |

| | | | | | | | | |
|---|---|---|---|---|---|---|---|---|
| 2b | | abundance | X08644 | 2,5-Dimetl | Radius_SOS | -0,0122 | 0,049611 | NA |
| 2b | | abundance | X08644 | 2,5-Dimetl | Tibia_SOS | -0,07197 | 0,053611 | NA |
| 2b | | abundance | X08644 | 2,5-Dimetl | Handgrip | 0,003841 | 0,060887 | NA |
| | 3 | abundance | X08646 | N-Stearoyl | Tibia_length | -0,00476 | 0,04897 | NA |
| | 3 | abundance | X08646 | N-Stearoyl | Radius_length | 0,067476 | 0,048859 | NA |
| | 3 | abundance | X08646 | N-Stearoyl | Radius_SOS | -0,01532 | 0,049023 | NA |
| | 3 | abundance | X08646 | N-Stearoyl | Tibia_SOS | 0,003852 | 0,052486 | NA |
| | 3 | abundance | X08646 | N-Stearoyl | Handgrip | -0,01639 | 0,060185 | NA |
| | 3 | abundance | X08657 | 1-(4-Aminc | Tibia_length | -0,06285 | 0,049178 | NA |
| | 3 | abundance | X08657 | 1-(4-Aminc | Radius_length | 0,027631 | 0,065953 | NA |
| | 3 | abundance | X08657 | 1-(4-Aminc | Radius_SOS | -0,04473 | 0,049022 | NA |
| | 3 | abundance | X08657 | 1-(4-Aminc | Tibia_SOS | -0,09281 | 0,052269 | NA |
| | 3 | abundance | X08657 | 1-(4-Aminc | Handgrip | 0,055599 | 0,060141 | NA |
| 2b | | abundance | X08659 | 3,3-Dimetl | Tibia_length | -0,04302 | 0,049152 | NA |
| 2b | | abundance | X08659 | 3,3-Dimetl | Radius_length | 0,008955 | 0,065459 | NA |
| 2b | | abundance | X08659 | 3,3-Dimetl | Radius_SOS | 0,044301 | 0,049065 | NA |
| 2b | | abundance | X08659 | 3,3-Dimetl | Tibia_SOS | 0,04621 | 0,052557 | NA |
| 2b | | abundance | X08659 | 3,3-Dimetl | Handgrip | -0,05597 | 0,060117 | NA |
| 2b | | abundance | X08663 | Pilocarpine | Tibia_length | 0,004847 | 0,050267 | NA |
| 2b | | abundance | X08663 | Pilocarpine | Radius_length | -0,08905 | 0,068528 | NA |
| 2b | | abundance | X08663 | Pilocarpine | Radius_SOS | -0,0705 | 0,049018 | NA |
| 2b | | abundance | X08663 | Pilocarpine | Tibia_SOS | -0,08878 | 0,052677 | NA |
| 2b | | abundance | X08663 | Pilocarpine | Handgrip | -0,00115 | 0,060998 | NA |
| | 3 | abundance | X08670 | Hydroxypro | Tibia_length | -0,00213 | 0,054294 | NA |
| | 3 | abundance | X08670 | Hydroxypro | Radius_length | 0,046983 | 0,073303 | NA |
| | 3 | abundance | X08670 | Hydroxypro | Radius_SOS | -0,03422 | 0,051867 | NA |
| | 3 | abundance | X08670 | Hydroxypro | Tibia_SOS | -3,17E-04 | 0,054394 | NA |
| | 3 | abundance | X08670 | Hydroxypro | Handgrip | -0,01488 | 0,061604 | NA |
| | 3 | abundance | X08673 | NPC | Tibia_length | 0,013646 | 0,054438 | NA |
| | 3 | abundance | X08673 | NPC | Radius_length | 0,084763 | 0,073221 | NA |
| | 3 | abundance | X08673 | NPC | Radius_SOS | -0,02542 | 0,051709 | NA |
| | 3 | abundance | X08673 | NPC | Tibia_SOS | 0,072424 | 0,055114 | NA |
| | 3 | abundance | X08673 | NPC | Handgrip | 0,06369 | 0,061766 | NA |
| | 3 | abundance | X08677 | 1-methylhy | Tibia_length | -0,00827 | 0,053627 | NA |
| | 3 | abundance | X08677 | 1-methylhy | Radius_length | 0,077453 | 0,072641 | NA |
| | 3 | abundance | X08677 | 1-methylhy | Radius_SOS | 0,09538 | 0,05062 | NA |
| | 3 | abundance | X08677 | 1-methylhy | Tibia_SOS | 0,098411 | 0,053962 | NA |
| | 3 | abundance | X08677 | 1-methylhy | Handgrip | 0,045441 | 0,061345 | NA |
| | 3 | abundance | X08690 | Ethanoic a | Tibia_length | -0,02787 | 0,055952 | NA |
| | 3 | abundance | X08690 | Ethanoic a | Radius_length | 0,034562 | 0,074718 | NA |
| | 3 | abundance | X08690 | Ethanoic a | Radius_SOS | -0,04866 | 0,053188 | NA |
| | 3 | abundance | X08690 | Ethanoic a | Tibia_SOS | 0,018138 | 0,056652 | NA |
| | 3 | abundance | X08690 | Ethanoic a | Handgrip | 0,097074 | 0,063294 | NA |
| | 3 | abundance | X08693 | 7-Sulfocho | Tibia_length | -0,06003 | 0,050303 | NA |
| | 3 | abundance | X08693 | 7-Sulfocho | Radius_length | -0,12802 | 0,068307 | NA |
| | 3 | abundance | X08693 | 7-Sulfocho | Radius_SOS | 0,021287 | 0,049146 | NA |
| | 3 | abundance | X08693 | 7-Sulfocho | Tibia_SOS | -0,00883 | 0,052636 | NA |
| | 3 | abundance | X08693 | 7-Sulfocho | Handgrip | 0,003903 | 0,060681 | NA |

| | | | | | | | | |
|---|---|---|---|---|---|---|---|---|
| | 3 | abundance | X08695 | 3b-Hydroxy | Tibia_length | 0,023133 | 0,050348 | NA |
| | 3 | abundance | X08695 | 3b-Hydroxy | Radius_length | 0,080285 | 0,068619 | NA |
| | 3 | abundance | X08695 | 3b-Hydroxy | Radius_SOS | -0,02308 | 0,049201 | NA |
| | 3 | abundance | X08695 | 3b-Hydroxy | Tibia_SOS | -0,03838 | 0,052675 | NA |
| | 3 | abundance | X08695 | 3b-Hydroxy | Handgrip | 0,063025 | 0,060887 | NA |
| | 3 | abundance | X08699 | Aurorix | Tibia_length | -0,00486 | 0,05643 | NA |
| | 3 | abundance | X08699 | Aurorix | Radius_length | -0,00592 | 0,07436 | NA |
| | 3 | abundance | X08699 | Aurorix | Radius_SOS | -0,06823 | 0,05352 | NA |
| | 3 | abundance | X08699 | Aurorix | Tibia_SOS | -0,02244 | 0,05638 | NA |
| | 3 | abundance | X08699 | Aurorix | Handgrip | 0,00802 | 0,062236 | NA |
| | 3 | abundance | X08707 | 2-(1-Ethoxy | Tibia_length | -0,04683 | 0,052097 | NA |
| | 3 | abundance | X08707 | 2-(1-Ethoxy | Radius_length | -0,11993 | 0,070825 | NA |
| | 3 | abundance | X08707 | 2-(1-Ethoxy | Radius_SOS | -0,07561 | 0,05 | NA |
| | 3 | abundance | X08707 | 2-(1-Ethoxy | Tibia_SOS | 0,008055 | 0,053992 | NA |
| | 3 | abundance | X08707 | 2-(1-Ethoxy | Handgrip | 0,020816 | 0,061641 | NA |
| | 3 | abundance | X08711 | 2-Acetami | Tibia_length | 0,165222 | 0,055213 | NA |
| | 3 | abundance | X08711 | 2-Acetami | Radius_length | 0,196857 | 0,072706 | NA |
| | 3 | abundance | X08711 | 2-Acetami | Radius_SOS | 0,047479 | 0,053208 | NA |
| | 3 | abundance | X08711 | 2-Acetami | Tibia_SOS | 0,125135 | 0,055586 | NA |
| | 3 | abundance | X08711 | 2-Acetami | Handgrip | -0,03051 | 0,062931 | NA |
| | 3 | abundance | X08712 | 4-Iodoanis | Tibia_length | 0,076453 | 0,050953 | NA |
| | 3 | abundance | X08712 | 4-Iodoanis | Radius_length | 0,210508 | 0,068449 | NA |
| | 3 | abundance | X08712 | 4-Iodoanis | Radius_SOS | 0,056156 | 0,049355 | NA |
| | 3 | abundance | X08712 | 4-Iodoanis | Tibia_SOS | 0,084398 | 0,052604 | NA |
| | 3 | abundance | X08712 | 4-Iodoanis | Handgrip | 0,155118 | 0,059918 | NA |
| 2b | | abundance | X08713 | Phenethyla | Tibia_length | -0,03502 | 0,050664 | NA |
| 2b | | abundance | X08713 | Phenethyla | Radius_length | -0,15883 | 0,068576 | NA |
| 2b | | abundance | X08713 | Phenethyla | Radius_SOS | -0,18975 | 0,048374 | NA |
| 2b | | abundance | X08713 | Phenethyla | Tibia_SOS | -0,03903 | 0,052816 | NA |
| 2b | | abundance | X08713 | Phenethyla | Handgrip | -0,16303 | 0,060045 | NA |
| | 3 | abundance | X08714 | Arabic acic | Tibia_length | -0,08174 | 0,05581 | NA |
| | 3 | abundance | X08714 | Arabic acic | Radius_length | -0,03061 | 0,075095 | NA |
| | 3 | abundance | X08714 | Arabic acic | Radius_SOS | -0,04959 | 0,053177 | NA |
| | 3 | abundance | X08714 | Arabic acic | Tibia_SOS | -0,0147 | 0,056301 | NA |
| | 3 | abundance | X08714 | Arabic acic | Handgrip | 0,038692 | 0,063221 | NA |
| 2b | | abundance | X08719 | 6-Methylqu | Tibia_length | 0,054349 | 0,049302 | NA |
| 2b | | abundance | X08719 | 6-Methylqu | Radius_length | -0,00369 | 0,066372 | NA |
| 2b | | abundance | X08719 | 6-Methylqu | Radius_SOS | -0,0575 | 0,048977 | NA |
| 2b | | abundance | X08719 | 6-Methylqu | Tibia_SOS | -0,00961 | 0,052489 | NA |
| 2b | | abundance | X08719 | 6-Methylqu | Handgrip | -0,08956 | 0,059966 | NA |
| | 3 | abundance | X08723 | S-Allylcyst | Tibia_length | -0,02273 | 0,053547 | NA |
| | 3 | abundance | X08723 | S-Allylcyst | Radius_length | -0,04185 | 0,072786 | NA |
| | 3 | abundance | X08723 | S-Allylcyst | Radius_SOS | -0,07394 | 0,051179 | NA |
| | 3 | abundance | X08723 | S-Allylcyst | Tibia_SOS | -0,01336 | 0,054575 | NA |
| | 3 | abundance | X08723 | S-Allylcyst | Handgrip | 0,071257 | 0,061615 | NA |
| | 3 | abundance | X08726 | YV8195000 | Tibia_length | 0,022069 | 0,051977 | NA |
| | 3 | abundance | X08726 | YV8195000 | Radius_length | 0,195819 | 0,069748 | NA |
| | 3 | abundance | X08726 | YV8195000 | Radius_SOS | 0,063843 | 0,049899 | NA |

| | | | | | | | |
|---|---|---|---|---|---|---|---|
| 3 | abundance | X08726 | YV819500( | Tibia_SOS | 0,100613 | 0,053414 | NA |
| 3 | abundance | X08726 | YV819500( | Handgrip | 0,036541 | 0,061638 | NA |
| 3 | abundance | X08733 | N-Methyl-1 | Tibia_length | 0,041542 | 0,050781 | NA |
| 3 | abundance | X08733 | N-Methyl-1 | Radius_length | 0,114327 | 0,068625 | NA |
| 3 | abundance | X08733 | N-Methyl-1 | Radius_SOS | -0,01544 | 0,049236 | NA |
| 3 | abundance | X08733 | N-Methyl-1 | Tibia_SOS | -0,0062 | 0,052764 | NA |
| 3 | abundance | X08733 | N-Methyl-1 | Handgrip | -0,04266 | 0,060667 | NA |
| 3 | abundance | X08739 | 1,1'-[1,12-I | Tibia_length | 0,025925 | 0,049905 | NA |
| 3 | abundance | X08739 | 1,1'-[1,12-I | Radius_length | 0,007226 | 0,067823 | NA |
| 3 | abundance | X08739 | 1,1'-[1,12-I | Radius_SOS | 0,00216 | 0,049075 | NA |
| 3 | abundance | X08739 | 1,1'-[1,12-I | Tibia_SOS | -0,04288 | 0,052654 | NA |
| 3 | abundance | X08739 | 1,1'-[1,12-I | Handgrip | 0,072572 | 0,060401 | NA |
| 3 | abundance | X08758 | Methohexit | Tibia_length | -0,04856 | 0,050953 | NA |
| 3 | abundance | X08758 | Methohexit | Radius_length | 0,023059 | 0,070082 | NA |
| 3 | abundance | X08758 | Methohexit | Radius_SOS | -0,02336 | 0,049505 | NA |
| 3 | abundance | X08758 | Methohexit | Tibia_SOS | 0,040455 | 0,05295 | NA |
| 3 | abundance | X08758 | Methohexit | Handgrip | 0,079454 | 0,060273 | NA |
| 2b | abundance | X08777 | 3-Aminosa | Tibia_length | -0,02776 | 0,049111 | NA |
| 2b | abundance | X08777 | 3-Aminosa | Radius_length | 0,061043 | 0,065245 | NA |
| 2b | abundance | X08777 | 3-Aminosa | Radius_SOS | -0,04588 | 0,049109 | NA |
| 2b | abundance | X08777 | 3-Aminosa | Tibia_SOS | 0,123172 | 0,052114 | NA |
| 2b | abundance | X08777 | 3-Aminosa | Handgrip | -0,08612 | 0,060109 | NA |
| 2b | abundance | X08779 | 3-(1-hydro: | Tibia_length | -0,1039 | 0,054218 | NA |
| 2b | abundance | X08779 | 3-(1-hydro: | Radius_length | -0,22238 | 0,07215 | NA |
| 2b | abundance | X08779 | 3-(1-hydro: | Radius_SOS | -0,07405 | 0,051568 | NA |
| 2b | abundance | X08779 | 3-(1-hydro: | Tibia_SOS | -0,10464 | 0,054312 | NA |
| 2b | abundance | X08779 | 3-(1-hydro: | Handgrip | -0,03226 | 0,062169 | NA |
| 3 | abundance | X08781 | Procaine | Tibia_length | -0,0265 | 0,051618 | NA |
| 3 | abundance | X08781 | Procaine | Radius_length | -0,02269 | 0,070484 | NA |
| 3 | abundance | X08781 | Procaine | Radius_SOS | -0,01492 | 0,049717 | NA |
| 3 | abundance | X08781 | Procaine | Tibia_SOS | 0,003045 | 0,05376 | NA |
| 3 | abundance | X08781 | Procaine | Handgrip | -0,04272 | 0,061558 | NA |
| 2b | abundance | X08789 | Acrylic acid | Tibia_length | -0,01858 | 0,056008 | NA |
| 2b | abundance | X08789 | Acrylic acid | Radius_length | 0,031532 | 0,074842 | NA |
| 2b | abundance | X08789 | Acrylic acid | Radius_SOS | -0,03472 | 0,053225 | NA |
| 2b | abundance | X08789 | Acrylic acid | Tibia_SOS | 0,010509 | 0,05684 | NA |
| 2b | abundance | X08789 | Acrylic acid | Handgrip | 0,112628 | 0,063152 | NA |
| 3 | abundance | X08792 | 3,4-Methyl | Tibia_length | 0,05785 | 0,059142 | NA |
| 3 | abundance | X08792 | 3,4-Methyl | Radius_length | 0,162772 | 0,075082 | NA |
| 3 | abundance | X08792 | 3,4-Methyl | Radius_SOS | 0,060053 | 0,056187 | NA |
| 3 | abundance | X08792 | 3,4-Methyl | Tibia_SOS | -0,03731 | 0,058941 | NA |
| 3 | abundance | X08792 | 3,4-Methyl | Handgrip | 0,043709 | 0,063724 | NA |
| 3 | abundance | X08795 | N-Acetylva | Tibia_length | 0,045009 | 0,054602 | NA |
| 3 | abundance | X08795 | N-Acetylva | Radius_length | 0,028171 | 0,073175 | NA |
| 3 | abundance | X08795 | N-Acetylva | Radius_SOS | 0,04974 | 0,05206 | NA |
| 3 | abundance | X08795 | N-Acetylva | Tibia_SOS | -0,01346 | 0,056093 | NA |
| 3 | abundance | X08795 | N-Acetylva | Handgrip | 0,001905 | 0,062067 | NA |
| 2b | abundance | X08801 | 3,7-Dimetl | Tibia_length | -0,06149 | 0,051311 | NA |

| | | | | | | | |
|---|---|---|---|---|---|---|---|
| 2b | abundance | X08801 | 3,7-Dimetl | Radius_length | -0,0936 | 0,070158 | NA |
| 2b | abundance | X08801 | 3,7-Dimetl | Radius_SOS | 0,042512 | 0,049612 | NA |
| 2b | abundance | X08801 | 3,7-Dimetl | Tibia_SOS | -0,15674 | 0,052662 | NA |
| 2b | abundance | X08801 | 3,7-Dimetl | Handgrip | -0,07427 | 0,060885 | NA |
| 3 | abundance | X08810 | N~6~,N~6 | Tibia_length | -0,02811 | 0,049102 | NA |
| 3 | abundance | X08810 | N~6~,N~6 | Radius_length | -0,05543 | 0,064922 | NA |
| 3 | abundance | X08810 | N~6~,N~6 | Radius_SOS | -0,01286 | 0,04919 | NA |
| 3 | abundance | X08810 | N~6~,N~6 | Tibia_SOS | -0,07475 | 0,052529 | NA |
| 3 | abundance | X08810 | N~6~,N~6 | Handgrip | -0,02882 | 0,060185 | NA |
| 3 | abundance | X08822 | CYS-ASP | Tibia_length | -0,08206 | 0,05706 | NA |
| 3 | abundance | X08822 | CYS-ASP | Radius_length | -0,06831 | 0,074648 | NA |
| 3 | abundance | X08822 | CYS-ASP | Radius_SOS | -0,11136 | 0,054203 | NA |
| 3 | abundance | X08822 | CYS-ASP | Tibia_SOS | -0,00865 | 0,058184 | NA |
| 3 | abundance | X08822 | CYS-ASP | Handgrip | -0,15072 | 0,062704 | NA |
| 3 | abundance | X08825 | Chenodeo: | Tibia_length | 0,091741 | 0,052253 | NA |
| 3 | abundance | X08825 | Chenodeo: | Radius_length | 0,094265 | 0,071086 | NA |
| 3 | abundance | X08825 | Chenodeo: | Radius_SOS | 0,047064 | 0,050233 | NA |
| 3 | abundance | X08825 | Chenodeo: | Tibia_SOS | -0,05111 | 0,053679 | NA |
| 3 | abundance | X08825 | Chenodeo: | Handgrip | 0,097845 | 0,061432 | NA |
| 3 | abundance | X08842 | Spermic ac | Tibia_length | -0,09026 | 0,054478 | NA |
| 3 | abundance | X08842 | Spermic ac | Radius_length | 0,042207 | 0,073833 | NA |
| 3 | abundance | X08842 | Spermic ac | Radius_SOS | -0,02152 | 0,05218 | NA |
| 3 | abundance | X08842 | Spermic ac | Tibia_SOS | 0,006641 | 0,05703 | NA |
| 3 | abundance | X08842 | Spermic ac | Handgrip | -0,10503 | 0,062577 | NA |
| 3 | abundance | X08843 | (2R,3S)-3-I | Tibia_length | 0,006324 | 0,050976 | NA |
| 3 | abundance | X08843 | (2R,3S)-3-I | Radius_length | 0,117945 | 0,069379 | NA |
| 3 | abundance | X08843 | (2R,3S)-3-I | Radius_SOS | 0,032254 | 0,049463 | NA |
| 3 | abundance | X08843 | (2R,3S)-3-I | Tibia_SOS | 0,157639 | 0,052555 | NA |
| 3 | abundance | X08843 | (2R,3S)-3-I | Handgrip | 0,035096 | 0,060737 | NA |
| 3 | abundance | X08844 | 4-Hydroxy- | Tibia_length | -0,0188 | 0,05632 | NA |
| 3 | abundance | X08844 | 4-Hydroxy- | Radius_length | 0,025766 | 0,074869 | NA |
| 3 | abundance | X08844 | 4-Hydroxy- | Radius_SOS | -0,04569 | 0,053508 | NA |
| 3 | abundance | X08844 | 4-Hydroxy- | Tibia_SOS | 0,019409 | 0,05684 | NA |
| 3 | abundance | X08844 | 4-Hydroxy- | Handgrip | 0,136446 | 0,063032 | NA |
| 3 | abundance | X08845 | 14-Hydroxy | Tibia_length | -0,02646 | 0,054533 | NA |
| 3 | abundance | X08845 | 14-Hydroxy | Radius_length | -0,09553 | 0,072988 | NA |
| 3 | abundance | X08845 | 14-Hydroxy | Radius_SOS | -0,02076 | 0,051771 | NA |
| 3 | abundance | X08845 | 14-Hydroxy | Tibia_SOS | 0,00602 | 0,054882 | NA |
| 3 | abundance | X08845 | 14-Hydroxy | Handgrip | -0,06082 | 0,061407 | NA |
| 2b | abundance | X08847 | 1-Vinylimic | Tibia_length | -0,09546 | 0,051969 | NA |
| 2b | abundance | X08847 | 1-Vinylimic | Radius_length | -0,18639 | 0,069914 | NA |
| 2b | abundance | X08847 | 1-Vinylimic | Radius_SOS | -0,12138 | 0,049764 | NA |
| 2b | abundance | X08847 | 1-Vinylimic | Tibia_SOS | -0,10612 | 0,053416 | NA |
| 2b | abundance | X08847 | 1-Vinylimic | Handgrip | 0,042198 | 0,060851 | NA |
| 3 | abundance | X08869 | Lanthionin | Tibia_length | -0,05365 | 0,055366 | NA |
| 3 | abundance | X08869 | Lanthionin | Radius_length | -0,06518 | 0,073708 | NA |
| 3 | abundance | X08869 | Lanthionin | Radius_SOS | -0,08191 | 0,052851 | NA |
| 3 | abundance | X08869 | Lanthionin | Tibia_SOS | -0,06136 | 0,056532 | NA |

| | | | | | | | | |
|---|---|---|---|---|---|---|---|---|
| | 3 | abundance | X08869 | Lanthionin | Handgrip | -0,05779 | 0,062361 | NA |
| | 3 | abundance | X08870 | Tetraacety | Tibia_length | -0,0571 | 0,058432 | NA |
| | 3 | abundance | X08870 | Tetraacety | Radius_length | 0,008388 | 0,075711 | NA |
| | 3 | abundance | X08870 | Tetraacety | Radius_SOS | -0,03028 | 0,055857 | NA |
| | 3 | abundance | X08870 | Tetraacety | Tibia_SOS | -0,05001 | 0,058601 | NA |
| | 3 | abundance | X08870 | Tetraacety | Handgrip | 0,042452 | 0,063326 | NA |
| | 3 | abundance | X08883 | 3-Phenylpr | Tibia_length | 0,006676 | 0,054607 | NA |
| | 3 | abundance | X08883 | 3-Phenylpr | Radius_length | -0,01075 | 0,072972 | NA |
| | 3 | abundance | X08883 | 3-Phenylpr | Radius_SOS | -0,03068 | 0,051936 | NA |
| | 3 | abundance | X08883 | 3-Phenylpr | Tibia_SOS | -0,04544 | 0,055021 | NA |
| | 3 | abundance | X08883 | 3-Phenylpr | Handgrip | -0,00515 | 0,061234 | NA |
| | 3 | abundance | X08893 | 5beta-Cho | Tibia_length | -0,17351 | 0,052632 | NA |
| | 3 | abundance | X08893 | 5beta-Cho | Radius_length | -0,15215 | 0,072163 | NA |
| | 3 | abundance | X08893 | 5beta-Cho | Radius_SOS | -0,02894 | 0,051072 | NA |
| | 3 | abundance | X08893 | 5beta-Cho | Tibia_SOS | -0,07613 | 0,054544 | NA |
| | 3 | abundance | X08893 | 5beta-Cho | Handgrip | -0,18992 | 0,060926 | NA |
| | 3 | abundance | X08895 | pentoxyl | Tibia_length | -0,15378 | 0,051761 | NA |
| | 3 | abundance | X08895 | pentoxyl | Radius_length | -0,01062 | 0,071552 | NA |
| | 3 | abundance | X08895 | pentoxyl | Radius_SOS | -0,03398 | 0,050303 | NA |
| | 3 | abundance | X08895 | pentoxyl | Tibia_SOS | -0,0603 | 0,053965 | NA |
| | 3 | abundance | X08895 | pentoxyl | Handgrip | 0,064942 | 0,062131 | NA |
| | 3 | abundance | X08908 | Vorinostat | Tibia_length | -0,00106 | 0,053053 | NA |
| | 3 | abundance | X08908 | Vorinostat | Radius_length | -0,00703 | 0,072306 | NA |
| | 3 | abundance | X08908 | Vorinostat | Radius_SOS | 0,032611 | 0,050837 | NA |
| | 3 | abundance | X08908 | Vorinostat | Tibia_SOS | -0,0053 | 0,054466 | NA |
| | 3 | abundance | X08908 | Vorinostat | Handgrip | -0,00177 | 0,062179 | NA |
| 2b | | abundance | X08909 | GLK (Pepti | Tibia_length | -0,05759 | 0,054948 | NA |
| 2b | | abundance | X08909 | GLK (Pepti | Radius_length | 0,005018 | 0,074014 | NA |
| 2b | | abundance | X08909 | GLK (Pepti | Radius_SOS | 0,032855 | 0,052162 | NA |
| 2b | | abundance | X08909 | GLK (Pepti | Tibia_SOS | -0,0023 | 0,054965 | NA |
| 2b | | abundance | X08909 | GLK (Pepti | Handgrip | 0,09963 | 0,060417 | NA |
| | 3 | abundance | X08923 | Semilicois | Tibia_length | -0,00225 | 0,055437 | NA |
| | 3 | abundance | X08923 | Semilicois | Radius_length | -0,12814 | 0,073259 | NA |
| | 3 | abundance | X08923 | Semilicois | Radius_SOS | -0,11687 | 0,052794 | NA |
| | 3 | abundance | X08923 | Semilicois | Tibia_SOS | -0,08433 | 0,056647 | NA |
| | 3 | abundance | X08923 | Semilicois | Handgrip | -0,10636 | 0,060971 | NA |
| | 3 | abundance | X08938 | Nonivamid | Tibia_length | 0,093284 | 0,050684 | NA |
| | 3 | abundance | X08938 | Nonivamid | Radius_length | 0,14699 | 0,069082 | NA |
| | 3 | abundance | X08938 | Nonivamid | Radius_SOS | 0,081355 | 0,049201 | NA |
| | 3 | abundance | X08938 | Nonivamid | Tibia_SOS | 0,142421 | 0,052613 | NA |
| | 3 | abundance | X08938 | Nonivamid | Handgrip | 0,092842 | 0,060287 | NA |
| | 3 | abundance | X08944 | Dipivefrin | Tibia_length | 0,153598 | 0,053922 | NA |
| | 3 | abundance | X08944 | Dipivefrin | Radius_length | 0,210937 | 0,072185 | NA |
| | 3 | abundance | X08944 | Dipivefrin | Radius_SOS | 0,096094 | 0,051822 | NA |
| | 3 | abundance | X08944 | Dipivefrin | Tibia_SOS | 0,164985 | 0,055135 | NA |
| | 3 | abundance | X08944 | Dipivefrin | Handgrip | 0,102028 | 0,061934 | NA |
| | 3 | abundance | X08965 | 1,9-Nonan | Tibia_length | -0,09059 | 0,055059 | NA |
| | 3 | abundance | X08965 | 1,9-Nonan | Radius_length | -0,01841 | 0,074199 | NA |

| | | | | | | | |
|---|---|---|---|---|---|---|---|
| 3 | abundance | X08965 | 1,9-Nonan | Radius_SOS | 0,054925 | 0,052208 | NA |
| 3 | abundance | X08965 | 1,9-Nonan | Tibia_SOS | 0,111736 | 0,054684 | NA |
| 3 | abundance | X08965 | 1,9-Nonan | Handgrip | 0,151883 | 0,061106 | NA |
| 3 | abundance | X08973 | Aminohipp | Tibia_length | 0,080595 | 0,054533 | NA |
| 3 | abundance | X08973 | Aminohipp | Radius_length | -0,00388 | 0,07313 | NA |
| 3 | abundance | X08973 | Aminohipp | Radius_SOS | -0,02284 | 0,052111 | NA |
| 3 | abundance | X08973 | Aminohipp | Tibia_SOS | -0,01088 | 0,057049 | NA |
| 3 | abundance | X08973 | Aminohipp | Handgrip | -0,02156 | 0,060179 | NA |
| 3 | abundance | X08983 | IN00150 | Tibia_length | -0,05538 | 0,051893 | NA |
| 3 | abundance | X08983 | IN00150 | Radius_length | -0,06949 | 0,071257 | NA |
| 3 | abundance | X08983 | IN00150 | Radius_SOS | 0,106715 | 0,049829 | NA |
| 3 | abundance | X08983 | IN00150 | Tibia_SOS | 0,045312 | 0,053537 | NA |
| 3 | abundance | X08983 | IN00150 | Handgrip | -0,01214 | 0,061863 | NA |
| 3 | abundance | X08987 | feruloylser | Tibia_length | -0,10547 | 0,055395 | NA |
| 3 | abundance | X08987 | feruloylser | Radius_length | 0,008676 | 0,073896 | NA |
| 3 | abundance | X08987 | feruloylser | Radius_SOS | -0,05471 | 0,052935 | NA |
| 3 | abundance | X08987 | feruloylser | Tibia_SOS | 0,011178 | 0,056354 | NA |
| 3 | abundance | X08987 | feruloylser | Handgrip | -0,01821 | 0,062205 | NA |
| 3 | abundance | X08990 | Procaine | Tibia_length | -0,14917 | 0,052915 | NA |
| 3 | abundance | X08990 | Procaine | Radius_length | -0,12071 | 0,071672 | NA |
| 3 | abundance | X08990 | Procaine | Radius_SOS | -0,02578 | 0,050893 | NA |
| 3 | abundance | X08990 | Procaine | Tibia_SOS | -0,07543 | 0,054209 | NA |
| 3 | abundance | X08990 | Procaine | Handgrip | -0,18563 | 0,060615 | NA |
| 3 | abundance | X08992 | N(2)-succi | Tibia_length | -0,17791 | 0,051354 | NA |
| 3 | abundance | X08992 | N(2)-succi | Radius_length | -0,05434 | 0,071053 | NA |
| 3 | abundance | X08992 | N(2)-succi | Radius_SOS | -0,02348 | 0,049948 | NA |
| 3 | abundance | X08992 | N(2)-succi | Tibia_SOS | 0,016387 | 0,053962 | NA |
| 3 | abundance | X08992 | N(2)-succi | Handgrip | 0,012516 | 0,061205 | NA |
| 3 | abundance | X09001 | Varanic ac | Tibia_length | -0,00164 | 0,052121 | NA |
| 3 | abundance | X09001 | Varanic ac | Radius_length | -0,01188 | 0,071126 | NA |
| 3 | abundance | X09001 | Varanic ac | Radius_SOS | -0,06526 | 0,04993 | NA |
| 3 | abundance | X09001 | Varanic ac | Tibia_SOS | -0,06027 | 0,053673 | NA |
| 3 | abundance | X09001 | Varanic ac | Handgrip | -0,11252 | 0,060589 | NA |
| 3 | abundance | X09004 | Arenaine | Tibia_length | 0,010785 | 0,05028 | NA |
| 3 | abundance | X09004 | Arenaine | Radius_length | 0,03267 | 0,068489 | NA |
| 3 | abundance | X09004 | Arenaine | Radius_SOS | -0,06931 | 0,049007 | NA |
| 3 | abundance | X09004 | Arenaine | Tibia_SOS | 0,064287 | 0,052509 | NA |
| 3 | abundance | X09004 | Arenaine | Handgrip | 0,0775 | 0,060215 | NA |
| 3 | abundance | X09008 | 5-guanidin | Tibia_length | 0,125014 | 0,050463 | NA |
| 3 | abundance | X09008 | 5-guanidin | Radius_length | 0,131771 | 0,068999 | NA |
| 3 | abundance | X09008 | 5-guanidin | Radius_SOS | 0,074697 | 0,049209 | NA |
| 3 | abundance | X09008 | 5-guanidin | Tibia_SOS | 0,104171 | 0,052651 | NA |
| 3 | abundance | X09008 | 5-guanidin | Handgrip | 0,157024 | 0,060219 | NA |
| 1 | abundance | X09010 | 4-Hydroxyp | Tibia_length | 0,019335 | 0,053752 | NA |
| 1 | abundance | X09010 | 4-Hydroxyp | Radius_length | 0,1638 | 0,072475 | NA |
| 1 | abundance | X09010 | 4-Hydroxyp | Radius_SOS | 0,018221 | 0,051203 | NA |
| 1 | abundance | X09010 | 4-Hydroxyp | Tibia_SOS | 0,085339 | 0,053884 | NA |
| 1 | abundance | X09010 | 4-Hydroxyp | Handgrip | 0,232762 | 0,059999 | NA |

| | | | | | | | |
|---|---|---|---|---|---|---|---|
| 3 | abundance | X09055 | N-[(1R,2S,( | Tibia_length | 0,032809 | 0,052749 | NA |
| 3 | abundance | X09055 | N-[(1R,2S,( | Radius_length | 0,124025 | 0,071114 | NA |
| 3 | abundance | X09055 | N-[(1R,2S,( | Radius_SOS | -0,02043 | 0,050463 | NA |
| 3 | abundance | X09055 | N-[(1R,2S,( | Tibia_SOS | 0,109438 | 0,05362 | NA |
| 3 | abundance | X09055 | N-[(1R,2S,( | Handgrip | 0,1121 | 0,061078 | NA |
| 3 | abundance | X09064 | Tetrahydro | Tibia_length | -0,1504 | 0,053156 | NA |
| 3 | abundance | X09064 | Tetrahydro | Radius_length | -0,14616 | 0,071629 | NA |
| 3 | abundance | X09064 | Tetrahydro | Radius_SOS | -0,07948 | 0,050868 | NA |
| 3 | abundance | X09064 | Tetrahydro | Tibia_SOS | -0,07914 | 0,054435 | NA |
| 3 | abundance | X09064 | Tetrahydro | Handgrip | -0,02844 | 0,061494 | NA |
| 3 | abundance | X09068 | N,N-Diethy | Tibia_length | 0,04333 | 0,053282 | NA |
| 3 | abundance | X09068 | N,N-Diethy | Radius_length | 0,013416 | 0,072437 | NA |
| 3 | abundance | X09068 | N,N-Diethy | Radius_SOS | -0,01365 | 0,050951 | NA |
| 3 | abundance | X09068 | N,N-Diethy | Tibia_SOS | -0,01189 | 0,055695 | NA |
| 3 | abundance | X09068 | N,N-Diethy | Handgrip | 0,062932 | 0,063076 | NA |
| 3 | abundance | X09082 | 1_2-Dihydr | Tibia_length | -0,09956 | 0,055877 | NA |
| 3 | abundance | X09082 | 1_2-Dihydr | Radius_length | -0,05037 | 0,074708 | NA |
| 3 | abundance | X09082 | 1_2-Dihydr | Radius_SOS | -0,10081 | 0,05336 | NA |
| 3 | abundance | X09082 | 1_2-Dihydr | Tibia_SOS | 0,046593 | 0,055854 | NA |
| 3 | abundance | X09082 | 1_2-Dihydr | Handgrip | 0,139681 | 0,063241 | NA |
| 3 | abundance | X09091 | N-(3-aceta | Tibia_length | -0,01706 | 0,055433 | NA |
| 3 | abundance | X09091 | N-(3-aceta | Radius_length | 0,055711 | 0,073703 | NA |
| 3 | abundance | X09091 | N-(3-aceta | Radius_SOS | -0,02055 | 0,052616 | NA |
| 3 | abundance | X09091 | N-(3-aceta | Tibia_SOS | 0,033633 | 0,056606 | NA |
| 3 | abundance | X09091 | N-(3-aceta | Handgrip | -0,04342 | 0,062297 | NA |
| 3 | abundance | X09092 | LysoSM(d1 | Tibia_length | 0,032399 | 0,052334 | NA |
| 3 | abundance | X09092 | LysoSM(d1 | Radius_length | 0,047097 | 0,071723 | NA |
| 3 | abundance | X09092 | LysoSM(d1 | Radius_SOS | 0,032005 | 0,050264 | NA |
| 3 | abundance | X09092 | LysoSM(d1 | Tibia_SOS | 0,059106 | 0,053614 | NA |
| 3 | abundance | X09092 | LysoSM(d1 | Handgrip | -0,10647 | 0,060478 | NA |
| 3 | abundance | X09099 | 1-(2-Hydro | Tibia_length | -0,00389 | 0,049462 | NA |
| 3 | abundance | X09099 | 1-(2-Hydro | Radius_length | 0,009815 | 0,066785 | NA |
| 3 | abundance | X09099 | 1-(2-Hydro | Radius_SOS | 0,067797 | 0,048916 | NA |
| 3 | abundance | X09099 | 1-(2-Hydro | Tibia_SOS | 0,001563 | 0,0525 | NA |
| 3 | abundance | X09099 | 1-(2-Hydro | Handgrip | -0,05713 | 0,060194 | NA |
| 3 | abundance | X09100 | Ethyl aceta | Tibia_length | 0,05081 | 0,050971 | NA |
| 3 | abundance | X09100 | Ethyl aceta | Radius_length | 0,017473 | 0,06968 | NA |
| 3 | abundance | X09100 | Ethyl aceta | Radius_SOS | 0,055077 | 0,04934 | NA |
| 3 | abundance | X09100 | Ethyl aceta | Tibia_SOS | 0,065691 | 0,052677 | NA |
| 3 | abundance | X09100 | Ethyl aceta | Handgrip | 0,038334 | 0,060482 | NA |
| 3 | abundance | X09110 | Valylvaline | Tibia_length | -0,07295 | 0,051097 | NA |
| 3 | abundance | X09110 | Valylvaline | Radius_length | -0,10202 | 0,069589 | NA |
| 3 | abundance | X09110 | Valylvaline | Radius_SOS | -0,0489 | 0,049582 | NA |
| 3 | abundance | X09110 | Valylvaline | Tibia_SOS | -0,01317 | 0,052884 | NA |
| 3 | abundance | X09110 | Valylvaline | Handgrip | -0,03796 | 0,061104 | NA |
| 3 | abundance | X09122 | Valyl-4-hyc | Tibia_length | -0,05255 | 0,053157 | NA |
| 3 | abundance | X09122 | Valyl-4-hyc | Radius_length | -0,06173 | 0,071933 | NA |
| 3 | abundance | X09122 | Valyl-4-hyc | Radius_SOS | 0,057459 | 0,05086 | NA |

| | | | | | | | |
|---|---|---|---|---|---|---|---|
| 3 | abundance | X09122 | Valyl-4-hyc | Tibia_SOS | 0,037745 | 0,055062 | NA |
| 3 | abundance | X09122 | Valyl-4-hyc | Handgrip | -0,03158 | 0,062588 | NA |
| 3 | abundance | X09129 | Coprine | Tibia_length | -0,01123 | 0,05763 | NA |
| 3 | abundance | X09129 | Coprine | Radius_length | -0,00522 | 0,075388 | NA |
| 3 | abundance | X09129 | Coprine | Radius_SOS | -0,0232 | 0,054753 | NA |
| 3 | abundance | X09129 | Coprine | Tibia_SOS | 0,056568 | 0,05748 | NA |
| 3 | abundance | X09129 | Coprine | Handgrip | -0,02927 | 0,062575 | NA |
| 1 | abundance | X09152 | Ferulic aci | Tibia_length | 0,016195 | 0,053974 | NA |
| 1 | abundance | X09152 | Ferulic aci | Radius_length | 0,099672 | 0,072966 | NA |
| 1 | abundance | X09152 | Ferulic aci | Radius_SOS | -0,02513 | 0,051337 | NA |
| 1 | abundance | X09152 | Ferulic aci | Tibia_SOS | -0,03024 | 0,05457 | NA |
| 1 | abundance | X09152 | Ferulic aci | Handgrip | 0,068673 | 0,062816 | NA |
| 3 | abundance | X09163 | Prenisteine | Tibia_length | -0,00523 | 0,05106 | NA |
| 3 | abundance | X09163 | Prenisteine | Radius_length | -0,0235 | 0,069784 | NA |
| 3 | abundance | X09163 | Prenisteine | Radius_SOS | 0,049264 | 0,049365 | NA |
| 3 | abundance | X09163 | Prenisteine | Tibia_SOS | 0,058154 | 0,05263 | NA |
| 3 | abundance | X09163 | Prenisteine | Handgrip | -0,09294 | 0,060802 | NA |
| 3 | abundance | X09165 | N-[1-Carbc | Tibia_length | 0,023196 | 0,051924 | NA |
| 3 | abundance | X09165 | N-[1-Carbc | Radius_length | 0,076971 | 0,070676 | NA |
| 3 | abundance | X09165 | N-[1-Carbc | Radius_SOS | -0,05251 | 0,049928 | NA |
| 3 | abundance | X09165 | N-[1-Carbc | Tibia_SOS | 0,090101 | 0,05332 | NA |
| 3 | abundance | X09165 | N-[1-Carbc | Handgrip | 0,043483 | 0,061432 | NA |
| 1 | abundance | X09187 | Salicylic ac | Tibia_length | -0,03499 | 0,054637 | NA |
| 1 | abundance | X09187 | Salicylic ac | Radius_length | -0,06202 | 0,074124 | NA |
| 1 | abundance | X09187 | Salicylic ac | Radius_SOS | 0,028093 | 0,051999 | NA |
| 1 | abundance | X09187 | Salicylic ac | Tibia_SOS | -0,03304 | 0,055777 | NA |
| 1 | abundance | X09187 | Salicylic ac | Handgrip | 0,119471 | 0,063071 | NA |
| 3 | abundance | X09189 | 4-[(E)-2-(3, | Tibia_length | -0,02417 | 0,051564 | NA |
| 3 | abundance | X09189 | 4-[(E)-2-(3, | Radius_length | 0,035037 | 0,070932 | NA |
| 3 | abundance | X09189 | 4-[(E)-2-(3, | Radius_SOS | -0,00237 | 0,04982 | NA |
| 3 | abundance | X09189 | 4-[(E)-2-(3, | Tibia_SOS | 0,043818 | 0,053261 | NA |
| 3 | abundance | X09189 | 4-[(E)-2-(3, | Handgrip | -0,04949 | 0,061682 | NA |
| 3 | abundance | X09207 | AAMU | Tibia_length | -0,12997 | 0,051601 | NA |
| 3 | abundance | X09207 | AAMU | Radius_length | -0,1368 | 0,070464 | NA |
| 3 | abundance | X09207 | AAMU | Radius_SOS | 0,016058 | 0,04994 | NA |
| 3 | abundance | X09207 | AAMU | Tibia_SOS | -0,13905 | 0,053386 | NA |
| 3 | abundance | X09207 | AAMU | Handgrip | -0,19252 | 0,060076 | NA |
| 3 | abundance | X09208 | IN00260 | Tibia_length | 0,04151 | 0,054946 | NA |
| 3 | abundance | X09208 | IN00260 | Radius_length | 0,04349 | 0,073386 | NA |
| 3 | abundance | X09208 | IN00260 | Radius_SOS | -0,13431 | 0,051849 | NA |
| 3 | abundance | X09208 | IN00260 | Tibia_SOS | -0,03154 | 0,055544 | NA |
| 3 | abundance | X09208 | IN00260 | Handgrip | -0,05846 | 0,061867 | NA |
| 3 | abundance | X09222 | L-gamma-( | Tibia_length | 0,001097 | 0,049889 | NA |
| 3 | abundance | X09222 | L-gamma-( | Radius_length | 0,051996 | 0,068052 | NA |
| 3 | abundance | X09222 | L-gamma-( | Radius_SOS | -0,03126 | 0,049038 | NA |
| 3 | abundance | X09222 | L-gamma-( | Tibia_SOS | 0,025226 | 0,052511 | NA |
| 3 | abundance | X09222 | L-gamma-( | Handgrip | -0,06808 | 0,060344 | NA |
| 3 | abundance | X09231 | 2-Keto-glut | Tibia_length | 0,120889 | 0,049899 | NA |

| | | | | | | | |
|---|---|---|---|---|---|---|---|
| 3 | abundance | X09231 | 2-Keto-glut | Radius_length | 0,155821 | 0,067633 | NA |
| 3 | abundance | X09231 | 2-Keto-glut | Radius_SOS | -0,06299 | 0,048965 | NA |
| 3 | abundance | X09231 | 2-Keto-glut | Tibia_SOS | 0,06173 | 0,052583 | NA |
| 3 | abundance | X09231 | 2-Keto-glut | Handgrip | -5,90E-04 | 0,060499 | NA |
| 3 | abundance | X09244 | gamma-Gl | Tibia_length | -1,16E-04 | 0,056372 | NA |
| 3 | abundance | X09244 | gamma-Gl | Radius_length | 0,091002 | 0,074635 | NA |
| 3 | abundance | X09244 | gamma-Gl | Radius_SOS | 0,006474 | 0,053527 | NA |
| 3 | abundance | X09244 | gamma-Gl | Tibia_SOS | 0,024224 | 0,056804 | NA |
| 3 | abundance | X09244 | gamma-Gl | Handgrip | 0,079191 | 0,062961 | NA |
| 3 | abundance | X09245 | L-gamma-( | Tibia_length | -0,11806 | 0,0546 | NA |
| 3 | abundance | X09245 | L-gamma-( | Radius_length | -0,01379 | 0,073548 | NA |
| 3 | abundance | X09245 | L-gamma-( | Radius_SOS | -0,0295 | 0,052061 | NA |
| 3 | abundance | X09245 | L-gamma-( | Tibia_SOS | 0,021986 | 0,055222 | NA |
| 3 | abundance | X09245 | L-gamma-( | Handgrip | 0,004639 | 0,060997 | NA |
| 3 | abundance | X09256 | Aspartyl-L- | Tibia_length | 0,114101 | 0,049696 | NA |
| 3 | abundance | X09256 | Aspartyl-L- | Radius_length | 0,113718 | 0,067407 | NA |
| 3 | abundance | X09256 | Aspartyl-L- | Radius_SOS | -0,05563 | 0,048975 | NA |
| 3 | abundance | X09256 | Aspartyl-L- | Tibia_SOS | 0,10126 | 0,052412 | NA |
| 3 | abundance | X09256 | Aspartyl-L- | Handgrip | 0,089089 | 0,060893 | NA |
| 3 | abundance | X09260 | (4S)-4-[(2E | Tibia_length | 0,22391 | 0,053544 | NA |
| 3 | abundance | X09260 | (4S)-4-[(2E | Radius_length | 0,291636 | 0,071435 | NA |
| 3 | abundance | X09260 | (4S)-4-[(2E | Radius_SOS | 0,14566 | 0,052031 | NA |
| 3 | abundance | X09260 | (4S)-4-[(2E | Tibia_SOS | 0,144969 | 0,054893 | NA |
| 3 | abundance | X09260 | (4S)-4-[(2E | Handgrip | 0,189808 | 0,060209 | NA |
| 3 | abundance | X09284 | 4-(Nitrosoa | Tibia_length | -0,03791 | 0,055657 | NA |
| 3 | abundance | X09284 | 4-(Nitrosoa | Radius_length | -0,03807 | 0,073912 | NA |
| 3 | abundance | X09284 | 4-(Nitrosoa | Radius_SOS | 0,079449 | 0,052851 | NA |
| 3 | abundance | X09284 | 4-(Nitrosoa | Tibia_SOS | 0,020502 | 0,05572 | NA |
| 3 | abundance | X09284 | 4-(Nitrosoa | Handgrip | 0,035479 | 0,062683 | NA |
| 3 | abundance | X09285 | MFCD0272 | Tibia_length | -0,06365 | 0,052772 | NA |
| 3 | abundance | X09285 | MFCD0272 | Radius_length | -0,17256 | 0,070936 | NA |
| 3 | abundance | X09285 | MFCD0272 | Radius_SOS | 0,003296 | 0,050577 | NA |
| 3 | abundance | X09285 | MFCD0272 | Tibia_SOS | -0,09389 | 0,053381 | NA |
| 3 | abundance | X09285 | MFCD0272 | Handgrip | -0,13094 | 0,061001 | NA |
| 3 | abundance | X09302 | Sular | Tibia_length | 0,023048 | 0,053652 | NA |
| 3 | abundance | X09302 | Sular | Radius_length | 0,071358 | 0,072228 | NA |
| 3 | abundance | X09302 | Sular | Radius_SOS | 0,057669 | 0,051134 | NA |
| 3 | abundance | X09302 | Sular | Tibia_SOS | 0,125785 | 0,055429 | NA |
| 3 | abundance | X09302 | Sular | Handgrip | 0,045917 | 0,061578 | NA |
| 3 | abundance | X09318 | butalbital | Tibia_length | -0,03481 | 0,051974 | NA |
| 3 | abundance | X09318 | butalbital | Radius_length | -0,02059 | 0,071195 | NA |
| 3 | abundance | X09318 | butalbital | Radius_SOS | 0,081311 | 0,049883 | NA |
| 3 | abundance | X09318 | butalbital | Tibia_SOS | -0,0651 | 0,053284 | NA |
| 3 | abundance | X09318 | butalbital | Handgrip | -0,16799 | 0,060424 | NA |
| 3 | abundance | X09336 | 1,1'-[1,12-I | Tibia_length | 0,054987 | 0,05447 | NA |
| 3 | abundance | X09336 | 1,1'-[1,12-I | Radius_length | 0,056651 | 0,073172 | NA |
| 3 | abundance | X09336 | 1,1'-[1,12-I | Radius_SOS | 0,056608 | 0,051787 | NA |
| 3 | abundance | X09336 | 1,1'-[1,12-I | Tibia_SOS | 0,012368 | 0,056029 | NA |

| | | | | | | | |
|---|---|---|---|---|---|---|---|
| 3 | abundance | X09336 | 1,1'-[1,12-I | Handgrip | 0,027993 | 0,062084 | NA |
| 3 | abundance | X09357 | 5-(5-Methy | Tibia_length | -0,06289 | 0,05421 | NA |
| 3 | abundance | X09357 | 5-(5-Methy | Radius_length | -0,09318 | 0,072819 | NA |
| 3 | abundance | X09357 | 5-(5-Methy | Radius_SOS | -0,06997 | 0,051322 | NA |
| 3 | abundance | X09357 | 5-(5-Methy | Tibia_SOS | -0,05033 | 0,055521 | NA |
| 3 | abundance | X09357 | 5-(5-Methy | Handgrip | 0,020731 | 0,062383 | NA |
| 3 | abundance | X09374 | 3-Deoxy-D | Tibia_length | -0,15367 | 0,054473 | NA |
| 3 | abundance | X09374 | 3-Deoxy-D | Radius_length | -0,13516 | 0,073218 | NA |
| 3 | abundance | X09374 | 3-Deoxy-D | Radius_SOS | -0,08846 | 0,052378 | NA |
| 3 | abundance | X09374 | 3-Deoxy-D | Tibia_SOS | 0,010256 | 0,056555 | NA |
| 3 | abundance | X09374 | 3-Deoxy-D | Handgrip | -0,08406 | 0,061961 | NA |
| 3 | abundance | X09385 | 2-(1-Napht | Tibia_length | -0,02321 | 0,048957 | NA |
| 3 | abundance | X09385 | 2-(1-Napht | Radius_length | 0,079437 | 0,048815 | NA |
| 3 | abundance | X09385 | 2-(1-Napht | Radius_SOS | 0,015975 | 0,049023 | NA |
| 3 | abundance | X09385 | 2-(1-Napht | Tibia_SOS | 0,05937 | 0,052394 | NA |
| 3 | abundance | X09385 | 2-(1-Napht | Handgrip | 0,049917 | 0,060118 | NA |
| 3 | abundance | X09390 | asn-pro | Tibia_length | -0,03318 | 0,052923 | NA |
| 3 | abundance | X09390 | asn-pro | Radius_length | 0,066753 | 0,071994 | NA |
| 3 | abundance | X09390 | asn-pro | Radius_SOS | 0,013789 | 0,050672 | NA |
| 3 | abundance | X09390 | asn-pro | Tibia_SOS | 0,061727 | 0,054244 | NA |
| 3 | abundance | X09390 | asn-pro | Handgrip | 0,100582 | 0,060898 | NA |
| 3 | abundance | X09391 | ophthalmic | Tibia_length | -0,12184 | 0,052075 | NA |
| 3 | abundance | X09391 | ophthalmic | Radius_length | -0,00216 | 0,071524 | NA |
| 3 | abundance | X09391 | ophthalmic | Radius_SOS | 0,040285 | 0,050067 | NA |
| 3 | abundance | X09391 | ophthalmic | Tibia_SOS | 0,069454 | 0,0538 | NA |
| 3 | abundance | X09391 | ophthalmic | Handgrip | 0,117022 | 0,061133 | NA |
| 3 | abundance | X09393 | 2-Isopropy | Tibia_length | 0,091471 | 0,055562 | NA |
| 3 | abundance | X09393 | 2-Isopropy | Radius_length | 0,046083 | 0,073934 | NA |
| 3 | abundance | X09393 | 2-Isopropy | Radius_SOS | -0,01992 | 0,053143 | NA |
| 3 | abundance | X09393 | 2-Isopropy | Tibia_SOS | 0,033524 | 0,056881 | NA |
| 3 | abundance | X09393 | 2-Isopropy | Handgrip | 0,117566 | 0,062678 | NA |
| 3 | abundance | X09399 | L-gamma-( | Tibia_length | -0,11651 | 0,053089 | NA |
| 3 | abundance | X09399 | L-gamma-( | Radius_length | -0,06751 | 0,072048 | NA |
| 3 | abundance | X09399 | L-gamma-( | Radius_SOS | 0,00477 | 0,050854 | NA |
| 3 | abundance | X09399 | L-gamma-( | Tibia_SOS | 0,019591 | 0,054596 | NA |
| 3 | abundance | X09399 | L-gamma-( | Handgrip | 0,050466 | 0,06141 | NA |
| 3 | abundance | X09454 | 2-(2-Hydro | Tibia_length | -0,09633 | 0,053026 | NA |
| 3 | abundance | X09454 | 2-(2-Hydro | Radius_length | -0,16088 | 0,071253 | NA |
| 3 | abundance | X09454 | 2-(2-Hydro | Radius_SOS | 0,012904 | 0,050746 | NA |
| 3 | abundance | X09454 | 2-(2-Hydro | Tibia_SOS | -0,03631 | 0,054184 | NA |
| 3 | abundance | X09454 | 2-(2-Hydro | Handgrip | -0,08002 | 0,061512 | NA |
| 3 | abundance | X09476 | N(1),N(8)-I | Tibia_length | 0,013932 | 0,051418 | NA |
| 3 | abundance | X09476 | N(1),N(8)-I | Radius_length | 0,089192 | 0,070631 | NA |
| 3 | abundance | X09476 | N(1),N(8)-I | Radius_SOS | 0,060564 | 0,049577 | NA |
| 3 | abundance | X09476 | N(1),N(8)-I | Tibia_SOS | 0,085512 | 0,053091 | NA |
| 3 | abundance | X09476 | N(1),N(8)-I | Handgrip | 0,131477 | 0,061439 | NA |
| 3 | abundance | X09480 | Spermic ac | Tibia_length | 0,037957 | 0,051973 | NA |
| 3 | abundance | X09480 | Spermic ac | Radius_length | 0,032369 | 0,071013 | NA |

| | | | | | | | |
|---|---|---|---|---|---|---|---|
| 3 | abundance | X09480 | Spermic ac | Radius_SOS | 0,01524 | 0,050086 | NA |
| 3 | abundance | X09480 | Spermic ac | Tibia_SOS | -0,05839 | 0,053498 | NA |
| 3 | abundance | X09480 | Spermic ac | Handgrip | 0,047416 | 0,061792 | NA |
| 3 | abundance | X09490 | 2-Hydroxy- | Tibia_length | -0,08906 | 0,056099 | NA |
| 3 | abundance | X09490 | 2-Hydroxy- | Radius_length | -0,11697 | 0,07398 | NA |
| 3 | abundance | X09490 | 2-Hydroxy- | Radius_SOS | -0,09002 | 0,054076 | NA |
| 3 | abundance | X09490 | 2-Hydroxy- | Tibia_SOS | -0,11517 | 0,056856 | NA |
| 3 | abundance | X09490 | 2-Hydroxy- | Handgrip | -0,06445 | 0,062227 | NA |
| 3 | abundance | X09503 | 4-(METHYL | Tibia_length | -0,10324 | 0,052917 | NA |
| 3 | abundance | X09503 | 4-(METHYL | Radius_length | 0,017356 | 0,072326 | NA |
| 3 | abundance | X09503 | 4-(METHYL | Radius_SOS | -0,00228 | 0,050872 | NA |
| 3 | abundance | X09503 | 4-(METHYL | Tibia_SOS | 0,036867 | 0,054131 | NA |
| 3 | abundance | X09503 | 4-(METHYL | Handgrip | -0,05164 | 0,062635 | NA |
| 3 | abundance | X09505 | (-)-Physost | Tibia_length | -0,02631 | 0,054803 | NA |
| 3 | abundance | X09505 | (-)-Physost | Radius_length | -0,04862 | 0,073543 | NA |
| 3 | abundance | X09505 | (-)-Physost | Radius_SOS | -0,02538 | 0,052229 | NA |
| 3 | abundance | X09505 | (-)-Physost | Tibia_SOS | 0,02696 | 0,054465 | NA |
| 3 | abundance | X09505 | (-)-Physost | Handgrip | -0,10846 | 0,061634 | NA |
| 3 | abundance | X09523 | N-(3,5-Dim | Tibia_length | -0,16996 | 0,056196 | NA |
| 3 | abundance | X09523 | N-(3,5-Dim | Radius_length | -0,11048 | 0,074461 | NA |
| 3 | abundance | X09523 | N-(3,5-Dim | Radius_SOS | -0,00894 | 0,054149 | NA |
| 3 | abundance | X09523 | N-(3,5-Dim | Tibia_SOS | -0,05174 | 0,057451 | NA |
| 3 | abundance | X09523 | N-(3,5-Dim | Handgrip | 0,051117 | 0,062687 | NA |
| 3 | abundance | X09525 | Homoanse | Tibia_length | -0,04461 | 0,055883 | NA |
| 3 | abundance | X09525 | Homoanse | Radius_length | 0,047628 | 0,074708 | NA |
| 3 | abundance | X09525 | Homoanse | Radius_SOS | -0,05342 | 0,05317 | NA |
| 3 | abundance | X09525 | Homoanse | Tibia_SOS | -0,01402 | 0,056184 | NA |
| 3 | abundance | X09525 | Homoanse | Handgrip | 0,072824 | 0,062361 | NA |
| 3 | abundance | X09533 | S(6)-acetyl | Tibia_length | 0,012014 | 0,052325 | NA |
| 3 | abundance | X09533 | S(6)-acetyl | Radius_length | 0,042381 | 0,071455 | NA |
| 3 | abundance | X09533 | S(6)-acetyl | Radius_SOS | 0,054809 | 0,050173 | NA |
| 3 | abundance | X09533 | S(6)-acetyl | Tibia_SOS | 0,076803 | 0,054056 | NA |
| 3 | abundance | X09533 | S(6)-acetyl | Handgrip | -0,14206 | 0,061505 | NA |
| 3 | abundance | X09545 | 4-Methyler | Tibia_length | 0,057747 | 0,052541 | NA |
| 3 | abundance | X09545 | 4-Methyler | Radius_length | 0,00568 | 0,07158 | NA |
| 3 | abundance | X09545 | 4-Methyler | Radius_SOS | 0,074496 | 0,050405 | NA |
| 3 | abundance | X09545 | 4-Methyler | Tibia_SOS | -0,02317 | 0,053882 | NA |
| 3 | abundance | X09545 | 4-Methyler | Handgrip | -0,14228 | 0,06021 | NA |
| 3 | abundance | X09546 | asn-phe | Tibia_length | -0,03136 | 0,053712 | NA |
| 3 | abundance | X09546 | asn-phe | Radius_length | -0,04323 | 0,072319 | NA |
| 3 | abundance | X09546 | asn-phe | Radius_SOS | -0,07883 | 0,050985 | NA |
| 3 | abundance | X09546 | asn-phe | Tibia_SOS | -0,12011 | 0,054511 | NA |
| 3 | abundance | X09546 | asn-phe | Handgrip | -0,05289 | 0,062018 | NA |
| 3 | abundance | X09548 | 2-O-beta-D | Tibia_length | -0,07417 | 0,056306 | NA |
| 3 | abundance | X09548 | 2-O-beta-D | Radius_length | 0,017916 | 0,07518 | NA |
| 3 | abundance | X09548 | 2-O-beta-D | Radius_SOS | -0,0222 | 0,053606 | NA |
| 3 | abundance | X09548 | 2-O-beta-D | Tibia_SOS | -0,00854 | 0,05731 | NA |
| 3 | abundance | X09548 | 2-O-beta-D | Handgrip | 0,00403 | 0,062877 | NA |

| | | | | | | | |
|---|---|---|---|---|---|---|---|
| 3 | abundance | X09554 | N-Propiony | Tibia_length | 0,052106 | 0,053563 | NA |
| 3 | abundance | X09554 | N-Propiony | Radius_length | 0,01105 | 0,072239 | NA |
| 3 | abundance | X09554 | N-Propiony | Radius_SOS | 0,102164 | 0,051013 | NA |
| 3 | abundance | X09554 | N-Propiony | Tibia_SOS | -0,06708 | 0,053692 | NA |
| 3 | abundance | X09554 | N-Propiony | Handgrip | -0,07217 | 0,061185 | NA |
| 3 | abundance | X09555 | N4-(beta-N | Tibia_length | 0,058077 | 0,051727 | NA |
| 3 | abundance | X09555 | N4-(beta-N | Radius_length | 0,011539 | 0,070824 | NA |
| 3 | abundance | X09555 | N4-(beta-N | Radius_SOS | -0,10539 | 0,049565 | NA |
| 3 | abundance | X09555 | N4-(beta-N | Tibia_SOS | 0,003575 | 0,053467 | NA |
| 3 | abundance | X09555 | N4-(beta-N | Handgrip | 0,043789 | 0,061197 | NA |
| 3 | abundance | X09565 | 3-Methoxy- | Tibia_length | -0,07543 | 0,051416 | NA |
| 3 | abundance | X09565 | 3-Methoxy- | Radius_length | -0,11119 | 0,069911 | NA |
| 3 | abundance | X09565 | 3-Methoxy- | Radius_SOS | -0,02268 | 0,049656 | NA |
| 3 | abundance | X09565 | 3-Methoxy- | Tibia_SOS | -0,09218 | 0,052867 | NA |
| 3 | abundance | X09565 | 3-Methoxy- | Handgrip | -0,08141 | 0,060431 | NA |
| 3 | abundance | X09577 | Spermic ac | Tibia_length | -0,09471 | 0,052897 | NA |
| 3 | abundance | X09577 | Spermic ac | Radius_length | -0,05197 | 0,071829 | NA |
| 3 | abundance | X09577 | Spermic ac | Radius_SOS | -0,0189 | 0,050786 | NA |
| 3 | abundance | X09577 | Spermic ac | Tibia_SOS | -0,06152 | 0,053867 | NA |
| 3 | abundance | X09577 | Spermic ac | Handgrip | 0,010362 | 0,061443 | NA |
| 3 | abundance | X09581 | L-gamma-( | Tibia_length | -0,03065 | 0,04969 | NA |
| 3 | abundance | X09581 | L-gamma-( | Radius_length | 0,048777 | 0,067801 | NA |
| 3 | abundance | X09581 | L-gamma-( | Radius_SOS | -8,03E-04 | 0,049038 | NA |
| 3 | abundance | X09581 | L-gamma-( | Tibia_SOS | 0,03377 | 0,052513 | NA |
| 3 | abundance | X09581 | L-gamma-( | Handgrip | -0,05586 | 0,060609 | NA |
| 3 | abundance | X09607 | N-(1H-Pyrr | Tibia_length | -0,05013 | 0,053151 | NA |
| 3 | abundance | X09607 | N-(1H-Pyrr | Radius_length | -0,02657 | 0,072323 | NA |
| 3 | abundance | X09607 | N-(1H-Pyrr | Radius_SOS | -0,01416 | 0,050879 | NA |
| 3 | abundance | X09607 | N-(1H-Pyrr | Tibia_SOS | -0,03182 | 0,054808 | NA |
| 3 | abundance | X09607 | N-(1H-Pyrr | Handgrip | -0,122 | 0,061417 | NA |
| 3 | abundance | X09608 | N-(3,5-Dim | Tibia_length | -0,10328 | 0,056088 | NA |
| 3 | abundance | X09608 | N-(3,5-Dim | Radius_length | -0,15028 | 0,074177 | NA |
| 3 | abundance | X09608 | N-(3,5-Dim | Radius_SOS | -0,06 | 0,053487 | NA |
| 3 | abundance | X09608 | N-(3,5-Dim | Tibia_SOS | -0,15154 | 0,055174 | NA |
| 3 | abundance | X09608 | N-(3,5-Dim | Handgrip | -0,04706 | 0,061301 | NA |
| 3 | abundance | X09609 | N-{3-[(4-Ac | Tibia_length | -0,07881 | 0,053549 | NA |
| 3 | abundance | X09609 | N-{3-[(4-Ac | Radius_length | -0,119 | 0,071804 | NA |
| 3 | abundance | X09609 | N-{3-[(4-Ac | Radius_SOS | -0,04475 | 0,05133 | NA |
| 3 | abundance | X09609 | N-{3-[(4-Ac | Tibia_SOS | 0,017759 | 0,05438 | NA |
| 3 | abundance | X09609 | N-{3-[(4-Ac | Handgrip | -0,00761 | 0,061118 | NA |
| 3 | abundance | X09612 | 3-hydroxy- | Tibia_length | -0,0333 | 0,051325 | NA |
| 3 | abundance | X09612 | 3-hydroxy- | Radius_length | -0,10562 | 0,06998 | NA |
| 3 | abundance | X09612 | 3-hydroxy- | Radius_SOS | 0,077214 | 0,049421 | NA |
| 3 | abundance | X09612 | 3-hydroxy- | Tibia_SOS | -0,05733 | 0,053207 | NA |
| 3 | abundance | X09612 | 3-hydroxy- | Handgrip | 0,019681 | 0,061218 | NA |
| 3 | abundance | X09615 | tyramine s | Tibia_length | -0,11941 | 0,051315 | NA |
| 3 | abundance | X09615 | tyramine s | Radius_length | -0,14484 | 0,070097 | NA |
| 3 | abundance | X09615 | tyramine s | Radius_SOS | -0,12958 | 0,049565 | NA |

| | | | | | | | | |
|---|---|---|---|---|---|---|---|---|
| | 3 | abundance | X09615 | tyramine sı | Tibia_SOS | -0,06182 | 0,053178 | NA |
| | 3 | abundance | X09615 | tyramine sı | Handgrip | -0,03043 | 0,061275 | NA |
| | 3 | abundance | X09622 | N-Hydroxy- | Tibia_length | -0,00748 | 0,049049 | NA |
| | 3 | abundance | X09622 | N-Hydroxy- | Radius_length | 0,023812 | 0,061829 | NA |
| | 3 | abundance | X09622 | N-Hydroxy- | Radius_SOS | -0,04725 | 0,050009 | NA |
| | 3 | abundance | X09622 | N-Hydroxy- | Tibia_SOS | 0,126369 | 0,053019 | NA |
| | 3 | abundance | X09622 | N-Hydroxy- | Handgrip | 0,012226 | 0,060405 | NA |
| 2b | | abundance | X09623 | Endothal | Tibia_length | -0,17515 | 0,053373 | NA |
| 2b | | abundance | X09623 | Endothal | Radius_length | -0,23975 | 0,070523 | NA |
| 2b | | abundance | X09623 | Endothal | Radius_SOS | -0,02113 | 0,051072 | NA |
| 2b | | abundance | X09623 | Endothal | Tibia_SOS | -0,16373 | 0,054881 | NA |
| 2b | | abundance | X09623 | Endothal | Handgrip | -0,15493 | 0,060564 | NA |
| | 3 | abundance | X09635 | 2,4,6-Triisc | Tibia_length | 0,063804 | 0,054969 | NA |
| | 3 | abundance | X09635 | 2,4,6-Triisc | Radius_length | 0,11365 | 0,07304 | NA |
| | 3 | abundance | X09635 | 2,4,6-Triisc | Radius_SOS | 0,075456 | 0,052315 | NA |
| | 3 | abundance | X09635 | 2,4,6-Triisc | Tibia_SOS | 0,033625 | 0,056943 | NA |
| | 3 | abundance | X09635 | 2,4,6-Triisc | Handgrip | 0,02946 | 0,06274 | NA |
| | 3 | abundance | X09643 | N-(3,5-Dim | Tibia_length | -0,10778 | 0,05579 | NA |
| | 3 | abundance | X09643 | N-(3,5-Dim | Radius_length | -0,13317 | 0,073612 | NA |
| | 3 | abundance | X09643 | N-(3,5-Dim | Radius_SOS | 0,042264 | 0,053099 | NA |
| | 3 | abundance | X09643 | N-(3,5-Dim | Tibia_SOS | -0,10677 | 0,055991 | NA |
| | 3 | abundance | X09643 | N-(3,5-Dim | Handgrip | -0,15283 | 0,061809 | NA |
| | 3 | abundance | X09645 | TOLMETIN | Tibia_length | -0,14668 | 0,053524 | NA |
| | 3 | abundance | X09645 | TOLMETIN | Radius_length | -0,11528 | 0,072179 | NA |
| | 3 | abundance | X09645 | TOLMETIN | Radius_SOS | -0,04365 | 0,051132 | NA |
| | 3 | abundance | X09645 | TOLMETIN | Tibia_SOS | -0,06112 | 0,054299 | NA |
| | 3 | abundance | X09645 | TOLMETIN | Handgrip | -0,00758 | 0,061228 | NA |
| | 3 | abundance | X09654 | linatine | Tibia_length | 0,056995 | 0,051815 | NA |
| | 3 | abundance | X09654 | linatine | Radius_length | 0,110159 | 0,070439 | NA |
| | 3 | abundance | X09654 | linatine | Radius_SOS | 0,042926 | 0,049962 | NA |
| | 3 | abundance | X09654 | linatine | Tibia_SOS | 0,029559 | 0,053015 | NA |
| | 3 | abundance | X09654 | linatine | Handgrip | -0,03676 | 0,061317 | NA |
| | 3 | abundance | X09657 | 2,3,4,5,6-F | Tibia_length | 0,082339 | 0,051033 | NA |
| | 3 | abundance | X09657 | 2,3,4,5,6-F | Radius_length | 0,150403 | 0,069233 | NA |
| | 3 | abundance | X09657 | 2,3,4,5,6-F | Radius_SOS | 0,068291 | 0,04943 | NA |
| | 3 | abundance | X09657 | 2,3,4,5,6-F | Tibia_SOS | 0,031701 | 0,053078 | NA |
| | 3 | abundance | X09657 | 2,3,4,5,6-F | Handgrip | 0,056335 | 0,060764 | NA |
| | 3 | abundance | X09675 | UK387000 | Tibia_length | 0,07886 | 0,052647 | NA |
| | 3 | abundance | X09675 | UK387000 | Radius_length | 0,08252 | 0,072073 | NA |
| | 3 | abundance | X09675 | UK387000 | Radius_SOS | 0,04474 | 0,050516 | NA |
| | 3 | abundance | X09675 | UK387000 | Tibia_SOS | 0,076291 | 0,055319 | NA |
| | 3 | abundance | X09675 | UK387000 | Handgrip | 0,093847 | 0,060851 | NA |
| | 3 | abundance | X09684 | (+)-Etomid | Tibia_length | -0,0415 | 0,049737 | NA |
| | 3 | abundance | X09684 | (+)-Etomid | Radius_length | 0,059324 | 0,067937 | NA |
| | 3 | abundance | X09684 | (+)-Etomid | Radius_SOS | 0,004625 | 0,049047 | NA |
| | 3 | abundance | X09684 | (+)-Etomid | Tibia_SOS | 0,042586 | 0,052499 | NA |
| | 3 | abundance | X09684 | (+)-Etomid | Handgrip | -0,07374 | 0,060492 | NA |
| | 3 | abundance | X09703 | 7,8-Didehy | Tibia_length | -0,14044 | 0,052953 | NA |

| | | | | | | | | |
|---|---|---|---|---|---|---|---|---|
| | 3 | abundance | X09703 | 7,8-Didehy | Radius_length | -0,12126 | 0,071684 | NA |
| | 3 | abundance | X09703 | 7,8-Didehy | Radius_SOS | 0,042648 | 0,050718 | NA |
| | 3 | abundance | X09703 | 7,8-Didehy | Tibia_SOS | -0,10022 | 0,053747 | NA |
| | 3 | abundance | X09703 | 7,8-Didehy | Handgrip | -0,13211 | 0,061305 | NA |
| | 3 | abundance | X09710 | LW800000 | Tibia_length | -0,07642 | 0,054709 | NA |
| | 3 | abundance | X09710 | LW800000 | Radius_length | -0,04915 | 0,07351 | NA |
| | 3 | abundance | X09710 | LW800000 | Radius_SOS | 0,033147 | 0,05212 | NA |
| | 3 | abundance | X09710 | LW800000 | Tibia_SOS | 0,017519 | 0,055337 | NA |
| | 3 | abundance | X09710 | LW800000 | Handgrip | 0,07235 | 0,062483 | NA |
| | 3 | abundance | X09713 | N(2)-succi | Tibia_length | 0,025192 | 0,050978 | NA |
| | 3 | abundance | X09713 | N(2)-succi | Radius_length | 0,018359 | 0,069968 | NA |
| | 3 | abundance | X09713 | N(2)-succi | Radius_SOS | 0,001834 | 0,049452 | NA |
| | 3 | abundance | X09713 | N(2)-succi | Tibia_SOS | 0,048009 | 0,052909 | NA |
| | 3 | abundance | X09713 | N(2)-succi | Handgrip | -0,06953 | 0,061184 | NA |
| | 3 | abundance | X09721 | 3-(2,3-Dihy | Tibia_length | -0,09914 | 0,056523 | NA |
| | 3 | abundance | X09721 | 3-(2,3-Dihy | Radius_length | -0,16646 | 0,073559 | NA |
| | 3 | abundance | X09721 | 3-(2,3-Dihy | Radius_SOS | -0,0019 | 0,053479 | NA |
| | 3 | abundance | X09721 | 3-(2,3-Dihy | Tibia_SOS | 0,009787 | 0,057603 | NA |
| | 3 | abundance | X09721 | 3-(2,3-Dihy | Handgrip | 0,023364 | 0,063407 | NA |
| | 3 | abundance | X09724 | Formylkynı | Tibia_length | 0,14262 | 0,052547 | NA |
| | 3 | abundance | X09724 | Formylkynı | Radius_length | 0,104316 | 0,071817 | NA |
| | 3 | abundance | X09724 | Formylkynı | Radius_SOS | -0,0616 | 0,05083 | NA |
| | 3 | abundance | X09724 | Formylkynı | Tibia_SOS | 0,036885 | 0,053869 | NA |
| | 3 | abundance | X09724 | Formylkynı | Handgrip | 0,010562 | 0,061093 | NA |
| 2b | | abundance | X09726 | indoline-2- | Tibia_length | 0,053272 | 0,052429 | NA |
| 2b | | abundance | X09726 | indoline-2- | Radius_length | 0,095185 | 0,07152 | NA |
| 2b | | abundance | X09726 | indoline-2- | Radius_SOS | -0,05069 | 0,05028 | NA |
| 2b | | abundance | X09726 | indoline-2- | Tibia_SOS | 0,08365 | 0,053365 | NA |
| 2b | | abundance | X09726 | indoline-2- | Handgrip | 0,045384 | 0,060598 | NA |
| | 3 | abundance | X09728 | {[(15-Hydro | Tibia_length | -0,03691 | 0,050741 | NA |
| | 3 | abundance | X09728 | {[(15-Hydro | Radius_length | -0,09145 | 0,069485 | NA |
| | 3 | abundance | X09728 | {[(15-Hydro | Radius_SOS | -0,00874 | 0,049328 | NA |
| | 3 | abundance | X09728 | {[(15-Hydro | Tibia_SOS | 0,079847 | 0,053296 | NA |
| | 3 | abundance | X09728 | {[(15-Hydro | Handgrip | 0,032694 | 0,060999 | NA |
| | 3 | abundance | X09738 | (7E,7'E)-5, | Tibia_length | -0,09714 | 0,051877 | NA |
| | 3 | abundance | X09738 | (7E,7'E)-5, | Radius_length | -0,20403 | 0,069825 | NA |
| | 3 | abundance | X09738 | (7E,7'E)-5, | Radius_SOS | -0,0622 | 0,050189 | NA |
| | 3 | abundance | X09738 | (7E,7'E)-5, | Tibia_SOS | -0,12973 | 0,053255 | NA |
| | 3 | abundance | X09738 | (7E,7'E)-5, | Handgrip | -0,06307 | 0,061205 | NA |
| | 3 | abundance | X09740 | MFCD0166 | Tibia_length | -0,05057 | 0,056308 | NA |
| | 3 | abundance | X09740 | MFCD0166 | Radius_length | -0,00958 | 0,075055 | NA |
| | 3 | abundance | X09740 | MFCD0166 | Radius_SOS | 0,020661 | 0,053519 | NA |
| | 3 | abundance | X09740 | MFCD0166 | Tibia_SOS | 0,045421 | 0,057661 | NA |
| | 3 | abundance | X09740 | MFCD0166 | Handgrip | 0,055839 | 0,062541 | NA |
| | 3 | abundance | X09741 | Estrone glu | Tibia_length | 0,106268 | 0,051299 | NA |
| | 3 | abundance | X09741 | Estrone glu | Radius_length | 0,082117 | 0,070347 | NA |
| | 3 | abundance | X09741 | Estrone glu | Radius_SOS | -0,01356 | 0,049701 | NA |
| | 3 | abundance | X09741 | Estrone glu | Tibia_SOS | 0,08029 | 0,053529 | NA |

| | | | | | | | | |
|---|---|---|---|---|---|---|---|---|
| | 3 | abundance | X09741 | Estrone glu | Handgrip | 0,014015 | 0,061108 | NA |
| 2b | | abundance | X09746 | ELK (Peptic | Tibia_length | -0,03112 | 0,053008 | NA |
| 2b | | abundance | X09746 | ELK (Peptic | Radius_length | 0,081738 | 0,072026 | NA |
| 2b | | abundance | X09746 | ELK (Peptic | Radius_SOS | 0,041496 | 0,050715 | NA |
| 2b | | abundance | X09746 | ELK (Peptic | Tibia_SOS | -0,00372 | 0,054589 | NA |
| 2b | | abundance | X09746 | ELK (Peptic | Handgrip | 0,065917 | 0,060553 | NA |
| | 3 | abundance | X09765 | Tetrahydro | Tibia_length | 0,017826 | 0,052768 | NA |
| | 3 | abundance | X09765 | Tetrahydro | Radius_length | 0,036745 | 0,071961 | NA |
| | 3 | abundance | X09765 | Tetrahydro | Radius_SOS | -0,01215 | 0,050548 | NA |
| | 3 | abundance | X09765 | Tetrahydro | Tibia_SOS | -0,05471 | 0,053299 | NA |
| | 3 | abundance | X09765 | Tetrahydro | Handgrip | -0,15451 | 0,06102 | NA |
| | 3 | abundance | X09771 | 2-(3-CARB | Tibia_length | -0,12049 | 0,052887 | NA |
| | 3 | abundance | X09771 | 2-(3-CARB | Radius_length | -0,11901 | 0,071553 | NA |
| | 3 | abundance | X09771 | 2-(3-CARB | Radius_SOS | -0,04373 | 0,050975 | NA |
| | 3 | abundance | X09771 | 2-(3-CARB | Tibia_SOS | -0,05583 | 0,053949 | NA |
| | 3 | abundance | X09771 | 2-(3-CARB | Handgrip | -0,04683 | 0,061112 | NA |
| | 3 | abundance | X09787 | Queuosine | Tibia_length | -0,12219 | 0,053289 | NA |
| | 3 | abundance | X09787 | Queuosine | Radius_length | -0,1596 | 0,071832 | NA |
| | 3 | abundance | X09787 | Queuosine | Radius_SOS | -0,20754 | 0,050106 | NA |
| | 3 | abundance | X09787 | Queuosine | Tibia_SOS | 0,02477 | 0,055194 | NA |
| | 3 | abundance | X09787 | Queuosine | Handgrip | 0,023877 | 0,062421 | NA |
| | 3 | abundance | X09805 | Methyl alpl | Tibia_length | 0,038465 | 0,055047 | NA |
| | 3 | abundance | X09805 | Methyl alpl | Radius_length | -0,00978 | 0,073723 | NA |
| | 3 | abundance | X09805 | Methyl alpl | Radius_SOS | -0,09859 | 0,052127 | NA |
| | 3 | abundance | X09805 | Methyl alpl | Tibia_SOS | 0,123584 | 0,056581 | NA |
| | 3 | abundance | X09805 | Methyl alpl | Handgrip | 0,018596 | 0,062297 | NA |
| | 3 | abundance | X09810 | Zalcitabine | Tibia_length | -0,11719 | 0,056275 | NA |
| | 3 | abundance | X09810 | Zalcitabine | Radius_length | 0,034666 | 0,074336 | NA |
| | 3 | abundance | X09810 | Zalcitabine | Radius_SOS | 0,006894 | 0,054004 | NA |
| | 3 | abundance | X09810 | Zalcitabine | Tibia_SOS | -0,07127 | 0,058724 | NA |
| | 3 | abundance | X09810 | Zalcitabine | Handgrip | -0,02774 | 0,06017 | NA |
| | 3 | abundance | X09825 | (2S)-6-Ami | Tibia_length | -0,03425 | 0,053792 | NA |
| | 3 | abundance | X09825 | (2S)-6-Ami | Radius_length | -0,05791 | 0,072594 | NA |
| | 3 | abundance | X09825 | (2S)-6-Ami | Radius_SOS | -0,00944 | 0,051327 | NA |
| | 3 | abundance | X09825 | (2S)-6-Ami | Tibia_SOS | -0,00856 | 0,055386 | NA |
| | 3 | abundance | X09825 | (2S)-6-Ami | Handgrip | 0,130958 | 0,061754 | NA |
| | 3 | abundance | X09849 | 3-Methylac | Tibia_length | -0,01873 | 0,051241 | NA |
| | 3 | abundance | X09849 | 3-Methylac | Radius_length | 0,025421 | 0,070322 | NA |
| | 3 | abundance | X09849 | 3-Methylac | Radius_SOS | -0,08903 | 0,049466 | NA |
| | 3 | abundance | X09849 | 3-Methylac | Tibia_SOS | 0,052152 | 0,053395 | NA |
| | 3 | abundance | X09849 | 3-Methylac | Handgrip | 0,008331 | 0,061591 | NA |
| 2b | | abundance | X09854 | N-Benzyl-3 | Tibia_length | 0,040976 | 0,05582 | NA |
| 2b | | abundance | X09854 | N-Benzyl-3 | Radius_length | 0,037878 | 0,074317 | NA |
| 2b | | abundance | X09854 | N-Benzyl-3 | Radius_SOS | -0,01559 | 0,052746 | NA |
| 2b | | abundance | X09854 | N-Benzyl-3 | Tibia_SOS | 0,010895 | 0,055953 | NA |
| 2b | | abundance | X09854 | N-Benzyl-3 | Handgrip | -0,02859 | 0,062235 | NA |
| | 3 | abundance | X09875 | 4-[(E)-2-(3, | Tibia_length | -0,03949 | 0,053 | NA |
| | 3 | abundance | X09875 | 4-[(E)-2-(3, | Radius_length | 0,053591 | 0,071787 | NA |

| | | | | | | | |
|---|---|---|---|---|---|---|---|
| 3 | abundance | X09875 | 4-[(E)-2-(3, | Radius_SOS | 0,047118 | 0,050807 | NA |
| 3 | abundance | X09875 | 4-[(E)-2-(3, | Tibia_SOS | -0,01369 | 0,054012 | NA |
| 3 | abundance | X09875 | 4-[(E)-2-(3, | Handgrip | 0,007423 | 0,0626 | NA |
| 3 | abundance | X09882 | Harmane | Tibia_length | 0,079326 | 0,049754 | NA |
| 3 | abundance | X09882 | Harmane | Radius_length | 0,096156 | 0,067561 | NA |
| 3 | abundance | X09882 | Harmane | Radius_SOS | 0,018642 | 0,049055 | NA |
| 3 | abundance | X09882 | Harmane | Tibia_SOS | 0,070531 | 0,052385 | NA |
| 3 | abundance | X09882 | Harmane | Handgrip | 0,005329 | 0,060517 | NA |
| 3 | abundance | X09895 | 1-(beta-D-r | Tibia_length | 0,061678 | 0,055282 | NA |
| 3 | abundance | X09895 | 1-(beta-D-r | Radius_length | 0,054948 | 0,07432 | NA |
| 3 | abundance | X09895 | 1-(beta-D-r | Radius_SOS | -0,07189 | 0,053154 | NA |
| 3 | abundance | X09895 | 1-(beta-D-r | Tibia_SOS | 0,081836 | 0,056845 | NA |
| 3 | abundance | X09895 | 1-(beta-D-r | Handgrip | 0,082202 | 0,061418 | NA |
| 3 | abundance | X09898 | N(6),N(6)-I | Tibia_length | -0,02736 | 0,053694 | NA |
| 3 | abundance | X09898 | N(6),N(6)-I | Radius_length | 0,057382 | 0,072405 | NA |
| 3 | abundance | X09898 | N(6),N(6)-I | Radius_SOS | 0,023504 | 0,051159 | NA |
| 3 | abundance | X09898 | N(6),N(6)-I | Tibia_SOS | 0,080266 | 0,054252 | NA |
| 3 | abundance | X09898 | N(6),N(6)-I | Handgrip | -0,01973 | 0,061638 | NA |
| 3 | abundance | X09912 | MC055530 | Tibia_length | -0,10637 | 0,053384 | NA |
| 3 | abundance | X09912 | MC055530 | Radius_length | -0,11019 | 0,072053 | NA |
| 3 | abundance | X09912 | MC055530 | Radius_SOS | -0,03573 | 0,051074 | NA |
| 3 | abundance | X09912 | MC055530 | Tibia_SOS | -0,07495 | 0,054832 | NA |
| 3 | abundance | X09912 | MC055530 | Handgrip | -0,02442 | 0,061485 | NA |
| 3 | abundance | X09919 | 4-ethylphe | Tibia_length | -0,14342 | 0,054385 | NA |
| 3 | abundance | X09919 | 4-ethylphe | Radius_length | -0,13605 | 0,072706 | NA |
| 3 | abundance | X09919 | 4-ethylphe | Radius_SOS | -0,01519 | 0,05219 | NA |
| 3 | abundance | X09919 | 4-ethylphe | Tibia_SOS | 0,050688 | 0,055085 | NA |
| 3 | abundance | X09919 | 4-ethylphe | Handgrip | -0,05092 | 0,061786 | NA |
| 3 | abundance | X09921 | 5-Hydroxy- | Tibia_length | 0,057506 | 0,05029 | NA |
| 3 | abundance | X09921 | 5-Hydroxy- | Radius_length | 0,101428 | 0,068519 | NA |
| 3 | abundance | X09921 | 5-Hydroxy- | Radius_SOS | 0,025798 | 0,049171 | NA |
| 3 | abundance | X09921 | 5-Hydroxy- | Tibia_SOS | -0,0527 | 0,052538 | NA |
| 3 | abundance | X09921 | 5-Hydroxy- | Handgrip | 0,013386 | 0,060265 | NA |
| 3 | abundance | X09932 | g-Aminobu | Tibia_length | 0,050897 | 0,051846 | NA |
| 3 | abundance | X09932 | g-Aminobu | Radius_length | 0,012208 | 0,070822 | NA |
| 3 | abundance | X09932 | g-Aminobu | Radius_SOS | -3,23E-04 | 0,049978 | NA |
| 3 | abundance | X09932 | g-Aminobu | Tibia_SOS | -0,03792 | 0,053798 | NA |
| 3 | abundance | X09932 | g-Aminobu | Handgrip | 0,037076 | 0,061389 | NA |
| 3 | abundance | X09940 | pentobarbi | Tibia_length | 0,011887 | 0,057789 | NA |
| 3 | abundance | X09940 | pentobarbi | Radius_length | 0,134566 | 0,075011 | NA |
| 3 | abundance | X09940 | pentobarbi | Radius_SOS | 0,095833 | 0,05476 | NA |
| 3 | abundance | X09940 | pentobarbi | Tibia_SOS | -0,0448 | 0,055834 | NA |
| 3 | abundance | X09940 | pentobarbi | Handgrip | -0,10466 | 0,062345 | NA |
| 3 | abundance | X09942 | Spermic ac | Tibia_length | -0,04508 | 0,054967 | NA |
| 3 | abundance | X09942 | Spermic ac | Radius_length | -0,0434 | 0,07333 | NA |
| 3 | abundance | X09942 | Spermic ac | Radius_SOS | -0,05746 | 0,052105 | NA |
| 3 | abundance | X09942 | Spermic ac | Tibia_SOS | -0,06799 | 0,055301 | NA |
| 3 | abundance | X09942 | Spermic ac | Handgrip | -0,07975 | 0,062242 | NA |

| | | | | | | | | |
|---|---|---|---|---|---|---|---|---|
| | 3 | abundance | X09950 | 3-Hydroxyh | Tibia_length | 0,003243 | 0,050373 | NA |
| | 3 | abundance | X09950 | 3-Hydroxyh | Radius_length | 0,047149 | 0,068605 | NA |
| | 3 | abundance | X09950 | 3-Hydroxyh | Radius_SOS | 0,069859 | 0,04906 | NA |
| | 3 | abundance | X09950 | 3-Hydroxyh | Tibia_SOS | -0,02221 | 0,052785 | NA |
| | 3 | abundance | X09950 | 3-Hydroxyh | Handgrip | -0,04647 | 0,060442 | NA |
| | 3 | abundance | X09951 | 5beta-Cypr | Tibia_length | -0,05758 | 0,050821 | NA |
| | 3 | abundance | X09951 | 5beta-Cypr | Radius_length | -0,05984 | 0,06951 | NA |
| | 3 | abundance | X09951 | 5beta-Cypr | Radius_SOS | -0,01319 | 0,04947 | NA |
| | 3 | abundance | X09951 | 5beta-Cypr | Tibia_SOS | -0,02615 | 0,053442 | NA |
| | 3 | abundance | X09951 | 5beta-Cypr | Handgrip | -0,16171 | 0,060447 | NA |
| | 3 | abundance | X09963 | Iminoglycin | Tibia_length | 0,09566 | 0,049707 | NA |
| | 3 | abundance | X09963 | Iminoglycin | Radius_length | 0,199129 | 0,06643 | NA |
| | 3 | abundance | X09963 | Iminoglycin | Radius_SOS | 0,017495 | 0,049044 | NA |
| | 3 | abundance | X09963 | Iminoglycin | Tibia_SOS | 0,117733 | 0,052221 | NA |
| | 3 | abundance | X09963 | Iminoglycin | Handgrip | 0,074615 | 0,060269 | NA |
| 2b | | abundance | X09965 | Biocytin | Tibia_length | 0,087623 | 0,051692 | NA |
| 2b | | abundance | X09965 | Biocytin | Radius_length | -0,06527 | 0,070809 | NA |
| 2b | | abundance | X09965 | Biocytin | Radius_SOS | -0,04714 | 0,049921 | NA |
| 2b | | abundance | X09965 | Biocytin | Tibia_SOS | 0,01204 | 0,054054 | NA |
| 2b | | abundance | X09965 | Biocytin | Handgrip | -0,12002 | 0,062086 | NA |
| | 3 | abundance | X09969 | 3-Methylac | Tibia_length | 0,049117 | 0,05345 | NA |
| | 3 | abundance | X09969 | 3-Methylac | Radius_length | 0,127273 | 0,072513 | NA |
| | 3 | abundance | X09969 | 3-Methylac | Radius_SOS | 0,016399 | 0,050973 | NA |
| | 3 | abundance | X09969 | 3-Methylac | Tibia_SOS | 0,144866 | 0,05383 | NA |
| | 3 | abundance | X09969 | 3-Methylac | Handgrip | 0,101968 | 0,061722 | NA |
| 2b | | abundance | X09977 | TLK (Peptic | Tibia_length | 0,018372 | 0,055264 | NA |
| 2b | | abundance | X09977 | TLK (Peptic | Radius_length | 0,104415 | 0,073456 | NA |
| 2b | | abundance | X09977 | TLK (Peptic | Radius_SOS | -0,00283 | 0,052516 | NA |
| 2b | | abundance | X09977 | TLK (Peptic | Tibia_SOS | 0,074474 | 0,056214 | NA |
| 2b | | abundance | X09977 | TLK (Peptic | Handgrip | 0,130429 | 0,061735 | NA |
| | 3 | abundance | X09980 | N-(4-Hepta | Tibia_length | 0,093606 | 0,050513 | NA |
| | 3 | abundance | X09980 | N-(4-Hepta | Radius_length | 0,138913 | 0,069006 | NA |
| | 3 | abundance | X09980 | N-(4-Hepta | Radius_SOS | 0,05949 | 0,049226 | NA |
| | 3 | abundance | X09980 | N-(4-Hepta | Tibia_SOS | 0,121126 | 0,052666 | NA |
| | 3 | abundance | X09980 | N-(4-Hepta | Handgrip | 0,062811 | 0,060668 | NA |
| | 3 | abundance | X09981 | N-Acetylva | Tibia_length | 0,106002 | 0,049654 | NA |
| | 3 | abundance | X09981 | N-Acetylva | Radius_length | 0,130934 | 0,067437 | NA |
| | 3 | abundance | X09981 | N-Acetylva | Radius_SOS | 0,029732 | 0,049028 | NA |
| | 3 | abundance | X09981 | N-Acetylva | Tibia_SOS | 0,125416 | 0,052192 | NA |
| | 3 | abundance | X09981 | N-Acetylva | Handgrip | 0,020334 | 0,060518 | NA |
| | 3 | abundance | X09998 | N-[(10Z)-7- | Tibia_length | 0,029392 | 0,049888 | NA |
| | 3 | abundance | X09998 | N-[(10Z)-7- | Radius_length | -0,0105 | 0,068117 | NA |
| | 3 | abundance | X09998 | N-[(10Z)-7- | Radius_SOS | 0,027418 | 0,049056 | NA |
| | 3 | abundance | X09998 | N-[(10Z)-7- | Tibia_SOS | -0,06236 | 0,052531 | NA |
| | 3 | abundance | X09998 | N-[(10Z)-7- | Handgrip | -0,06794 | 0,06057 | NA |
| | 3 | abundance | X10011 | Midodrine | Tibia_length | -0,191 | 0,056455 | NA |
| | 3 | abundance | X10011 | Midodrine | Radius_length | -0,14845 | 0,075068 | NA |
| | 3 | abundance | X10011 | Midodrine | Radius_SOS | -0,06253 | 0,054364 | NA |

| | | | | | | | |
|---|---|---|---|---|---|---|---|
| 3 | abundance | X10011 | Midodrine | Tibia_SOS | -0,11074 | 0,058012 | NA |
| 3 | abundance | X10011 | Midodrine | Handgrip | -0,14464 | 0,062403 | NA |
| 3 | abundance | X10025 | 2-[4-(3-Hy | Tibia_length | -0,01103 | 0,051462 | NA |
| 3 | abundance | X10025 | 2-[4-(3-Hy | Radius_length | 0,012686 | 0,070443 | NA |
| 3 | abundance | X10025 | 2-[4-(3-Hy | Radius_SOS | 0,035632 | 0,049681 | NA |
| 3 | abundance | X10025 | 2-[4-(3-Hy | Tibia_SOS | -0,03627 | 0,053838 | NA |
| 3 | abundance | X10025 | 2-[4-(3-Hy | Handgrip | 0,120878 | 0,061198 | NA |
| 3 | abundance | X10032 | 4,9a-Dime | Tibia_length | -0,02941 | 0,058571 | NA |
| 3 | abundance | X10032 | 4,9a-Dime | Radius_length | -0,04128 | 0,075603 | NA |
| 3 | abundance | X10032 | 4,9a-Dime | Radius_SOS | -0,1239 | 0,055862 | NA |
| 3 | abundance | X10032 | 4,9a-Dime | Tibia_SOS | -0,02472 | 0,05247 | NA |
| 3 | abundance | X10032 | 4,9a-Dime | Handgrip | 0,032883 | 0,06016 | NA |
| 3 | abundance | X10033 | epsilon-(g | Tibia_length | -0,04098 | 0,053716 | NA |
| 3 | abundance | X10033 | epsilon-(g | Radius_length | -0,09118 | 0,072228 | NA |
| 3 | abundance | X10033 | epsilon-(g | Radius_SOS | -0,02517 | 0,051144 | NA |
| 3 | abundance | X10033 | epsilon-(g | Tibia_SOS | -0,03811 | 0,054473 | NA |
| 3 | abundance | X10033 | epsilon-(g | Handgrip | -0,05871 | 0,061681 | NA |
| 3 | abundance | X10036 | Nimodipin | Tibia_length | -0,05748 | 0,055397 | NA |
| 3 | abundance | X10036 | Nimodipin | Radius_length | -0,01665 | 0,073729 | NA |
| 3 | abundance | X10036 | Nimodipin | Radius_SOS | -0,00886 | 0,052704 | NA |
| 3 | abundance | X10036 | Nimodipin | Tibia_SOS | 0,099531 | 0,056188 | NA |
| 3 | abundance | X10036 | Nimodipin | Handgrip | -0,05835 | 0,061673 | NA |
| 3 | abundance | X10037 | his-gln | Tibia_length | -0,09452 | 0,051956 | NA |
| 3 | abundance | X10037 | his-gln | Radius_length | -0,10742 | 0,070637 | NA |
| 3 | abundance | X10037 | his-gln | Radius_SOS | 0,030657 | 0,049951 | NA |
| 3 | abundance | X10037 | his-gln | Tibia_SOS | -0,04252 | 0,053275 | NA |
| 3 | abundance | X10037 | his-gln | Handgrip | 0,087995 | 0,060459 | NA |
| 2b | abundance | X10046 | Dehydroac | Tibia_length | -0,07816 | 0,05681 | NA |
| 2b | abundance | X10046 | Dehydroac | Radius_length | -0,01319 | 0,075527 | NA |
| 2b | abundance | X10046 | Dehydroac | Radius_SOS | -0,05365 | 0,054121 | NA |
| 2b | abundance | X10046 | Dehydroac | Tibia_SOS | -0,05608 | 0,057977 | NA |
| 2b | abundance | X10046 | Dehydroac | Handgrip | 0,011659 | 0,060189 | NA |
| 3 | abundance | X10076 | trimethadi | Tibia_length | -8,80E-04 | 0,049592 | NA |
| 3 | abundance | X10076 | trimethadi | Radius_length | -0,03036 | 0,067512 | NA |
| 3 | abundance | X10076 | trimethadi | Radius_SOS | 0,028374 | 0,049012 | NA |
| 3 | abundance | X10076 | trimethadi | Tibia_SOS | 0,065704 | 0,052424 | NA |
| 3 | abundance | X10076 | trimethadi | Handgrip | 0,078664 | 0,060125 | NA |
| 3 | abundance | X10091 | (2E,6E)-9-[ | Tibia_length | -0,00858 | 0,052253 | NA |
| 3 | abundance | X10091 | (2E,6E)-9-[ | Radius_length | 0,065677 | 0,071111 | NA |
| 3 | abundance | X10091 | (2E,6E)-9-[ | Radius_SOS | -0,0301 | 0,050207 | NA |
| 3 | abundance | X10091 | (2E,6E)-9-[ | Tibia_SOS | 0,021258 | 0,054192 | NA |
| 3 | abundance | X10091 | (2E,6E)-9-[ | Handgrip | -0,0271 | 0,061348 | NA |
| 3 | abundance | X10097 | Coixol | Tibia_length | -0,00938 | 0,054503 | NA |
| 3 | abundance | X10097 | Coixol | Radius_length | -0,11519 | 0,072472 | NA |
| 3 | abundance | X10097 | Coixol | Radius_SOS | -0,04026 | 0,051783 | NA |
| 3 | abundance | X10097 | Coixol | Tibia_SOS | 0,026316 | 0,054938 | NA |
| 3 | abundance | X10097 | Coixol | Handgrip | -0,07837 | 0,06131 | NA |
| 3 | abundance | X10098 | Methyl 2,3- | Tibia_length | -0,12073 | 0,055423 | NA |

| | | | | | | | |
|---|---|---|---|---|---|---|---|
| 3 | abundance | X10098 | Methyl 2,3- | Radius_length | -0,05617 | 0,07373 | NA |
| 3 | abundance | X10098 | Methyl 2,3- | Radius_SOS | -0,0424 | 0,052918 | NA |
| 3 | abundance | X10098 | Methyl 2,3- | Tibia_SOS | -0,0189 | 0,057397 | NA |
| 3 | abundance | X10098 | Methyl 2,3- | Handgrip | -0,05782 | 0,062591 | NA |
| 3 | abundance | X10099 | ala-met | Tibia_length | 0,15334 | 0,05631 | NA |
| 3 | abundance | X10099 | ala-met | Radius_length | 0,197652 | 0,07357 | NA |
| 3 | abundance | X10099 | ala-met | Radius_SOS | 0,071959 | 0,0538 | NA |
| 3 | abundance | X10099 | ala-met | Tibia_SOS | 0,126119 | 0,058209 | NA |
| 3 | abundance | X10099 | ala-met | Handgrip | 0,038361 | 0,062978 | NA |
| 3 | abundance | X10121 | Leu-pro | Tibia_length | -0,17913 | 0,053933 | NA |
| 3 | abundance | X10121 | Leu-pro | Radius_length | -0,28939 | 0,069625 | NA |
| 3 | abundance | X10121 | Leu-pro | Radius_SOS | -0,02526 | 0,051081 | NA |
| 3 | abundance | X10121 | Leu-pro | Tibia_SOS | -0,13494 | 0,053404 | NA |
| 3 | abundance | X10121 | Leu-pro | Handgrip | -0,23066 | 0,060063 | NA |
| 3 | abundance | X10129 | SECONAL | Tibia_length | -0,05783 | 0,054075 | NA |
| 3 | abundance | X10129 | SECONAL | Radius_length | -0,05934 | 0,072812 | NA |
| 3 | abundance | X10129 | SECONAL | Radius_SOS | 0,043489 | 0,051451 | NA |
| 3 | abundance | X10129 | SECONAL | Tibia_SOS | -0,05072 | 0,054467 | NA |
| 3 | abundance | X10129 | SECONAL | Handgrip | -0,11527 | 0,061593 | NA |
| 3 | abundance | X10130 | N-(4-Hydro | Tibia_length | -0,15984 | 0,054111 | NA |
| 3 | abundance | X10130 | N-(4-Hydro | Radius_length | -0,1305 | 0,0729 | NA |
| 3 | abundance | X10130 | N-(4-Hydro | Radius_SOS | -0,04283 | 0,0519 | NA |
| 3 | abundance | X10130 | N-(4-Hydro | Tibia_SOS | -0,09418 | 0,054984 | NA |
| 3 | abundance | X10130 | N-(4-Hydro | Handgrip | -0,085 | 0,061895 | NA |
| 3 | abundance | X10137 | 1-Methyl-1 | Tibia_length | -0,0112 | 0,055082 | NA |
| 3 | abundance | X10137 | 1-Methyl-1 | Radius_length | 0,092025 | 0,073061 | NA |
| 3 | abundance | X10137 | 1-Methyl-1 | Radius_SOS | 0,02183 | 0,052452 | NA |
| 3 | abundance | X10137 | 1-Methyl-1 | Tibia_SOS | 0,134014 | 0,056539 | NA |
| 3 | abundance | X10137 | 1-Methyl-1 | Handgrip | -0,00551 | 0,063011 | NA |
| 3 | abundance | X10144 | imazameth | Tibia_length | 0,128195 | 0,053041 | NA |
| 3 | abundance | X10144 | imazameth | Radius_length | 0,135141 | 0,072082 | NA |
| 3 | abundance | X10144 | imazameth | Radius_SOS | 0,113969 | 0,050917 | NA |
| 3 | abundance | X10144 | imazameth | Tibia_SOS | 0,087394 | 0,054805 | NA |
| 3 | abundance | X10144 | imazameth | Handgrip | 0,105309 | 0,060635 | NA |
| 3 | abundance | X10148 | cys-met | Tibia_length | -0,05984 | 0,054967 | NA |
| 3 | abundance | X10148 | cys-met | Radius_length | -0,03052 | 0,073764 | NA |
| 3 | abundance | X10148 | cys-met | Radius_SOS | -0,01989 | 0,052465 | NA |
| 3 | abundance | X10148 | cys-met | Tibia_SOS | 0,052811 | 0,05584 | NA |
| 3 | abundance | X10148 | cys-met | Handgrip | 0,102324 | 0,062038 | NA |
| 3 | abundance | X10151 | 2-(3-CARB | Tibia_length | -0,16704 | 0,053486 | NA |
| 3 | abundance | X10151 | 2-(3-CARB | Radius_length | -0,16255 | 0,071944 | NA |
| 3 | abundance | X10151 | 2-(3-CARB | Radius_SOS | -0,03753 | 0,051402 | NA |
| 3 | abundance | X10151 | 2-(3-CARB | Tibia_SOS | -0,05614 | 0,054432 | NA |
| 3 | abundance | X10151 | 2-(3-CARB | Handgrip | -0,00857 | 0,062027 | NA |
| 2a | abundance | X10162 | Quinaldic a | Tibia_length | 0,105264 | 0,05229 | NA |
| 2a | abundance | X10162 | Quinaldic a | Radius_length | 0,108218 | 0,070861 | NA |
| 2a | abundance | X10162 | Quinaldic a | Radius_SOS | 0,101926 | 0,049934 | NA |
| 2a | abundance | X10162 | Quinaldic a | Tibia_SOS | -0,01328 | 0,053025 | NA |

| | | | | | | | |
|---|---|---|---|---|---|---|---|
| 2a | abundance | X10162 | Quinaldic a | Handgrip | 0,041163 | 0,060196 | NA |
| 3 | abundance | X10177 | Gly-DL-Phe | Tibia_length | -0,16268 | 0,05653 | NA |
| 3 | abundance | X10177 | Gly-DL-Phe | Radius_length | -0,28107 | 0,071815 | NA |
| 3 | abundance | X10177 | Gly-DL-Phe | Radius_SOS | -0,0898 | 0,053812 | NA |
| 3 | abundance | X10177 | Gly-DL-Phe | Tibia_SOS | -0,02548 | 0,057844 | NA |
| 3 | abundance | X10177 | Gly-DL-Phe | Handgrip | -0,10767 | 0,062559 | NA |
| 3 | abundance | X10180 | 4-Formyl-2 | Tibia_length | -0,07941 | 0,053448 | NA |
| 3 | abundance | X10180 | 4-Formyl-2 | Radius_length | -0,11344 | 0,072323 | NA |
| 3 | abundance | X10180 | 4-Formyl-2 | Radius_SOS | -0,09591 | 0,051181 | NA |
| 3 | abundance | X10180 | 4-Formyl-2 | Tibia_SOS | -0,04384 | 0,05449 | NA |
| 3 | abundance | X10180 | 4-Formyl-2 | Handgrip | -0,04385 | 0,062431 | NA |
| 3 | abundance | X10203 | L-gamma-( | Tibia_length | -0,06057 | 0,049904 | NA |
| 3 | abundance | X10203 | L-gamma-( | Radius_length | -0,0801 | 0,06765 | NA |
| 3 | abundance | X10203 | L-gamma-( | Radius_SOS | 0,020111 | 0,04902 | NA |
| 3 | abundance | X10203 | L-gamma-( | Tibia_SOS | -0,05334 | 0,052422 | NA |
| 3 | abundance | X10203 | L-gamma-( | Handgrip | 0,006506 | 0,060503 | NA |
| 2b | abundance | X10224 | Imidazolela | Tibia_length | 0,097378 | 0,052147 | NA |
| 2b | abundance | X10224 | Imidazolela | Radius_length | 0,136559 | 0,070874 | NA |
| 2b | abundance | X10224 | Imidazolela | Radius_SOS | 0,054781 | 0,050089 | NA |
| 2b | abundance | X10224 | Imidazolela | Tibia_SOS | 0,096909 | 0,054258 | NA |
| 2b | abundance | X10224 | Imidazolela | Handgrip | 0,024936 | 0,060552 | NA |
| 3 | abundance | X10246 | Ethynodiol | Tibia_length | -0,00572 | 0,0556 | NA |
| 3 | abundance | X10246 | Ethynodiol | Radius_length | -0,02426 | 0,073912 | NA |
| 3 | abundance | X10246 | Ethynodiol | Radius_SOS | -0,0445 | 0,052781 | NA |
| 3 | abundance | X10246 | Ethynodiol | Tibia_SOS | 0,040606 | 0,056783 | NA |
| 3 | abundance | X10246 | Ethynodiol | Handgrip | -0,05323 | 0,061891 | NA |
| 3 | abundance | X10248 | (+)-Etomid | Tibia_length | 0,053662 | 0,054665 | NA |
| 3 | abundance | X10248 | (+)-Etomid | Radius_length | 0,025848 | 0,07336 | NA |
| 3 | abundance | X10248 | (+)-Etomid | Radius_SOS | -0,11431 | 0,051785 | NA |
| 3 | abundance | X10248 | (+)-Etomid | Tibia_SOS | 0,170121 | 0,056204 | NA |
| 3 | abundance | X10248 | (+)-Etomid | Handgrip | 0,048821 | 0,062272 | NA |
| 3 | abundance | X10265 | 3-methyl-4 | Tibia_length | 0,093829 | 0,056245 | NA |
| 3 | abundance | X10265 | 3-methyl-4 | Radius_length | 0,231717 | 0,07288 | NA |
| 3 | abundance | X10265 | 3-methyl-4 | Radius_SOS | 0,067941 | 0,05321 | NA |
| 3 | abundance | X10265 | 3-methyl-4 | Tibia_SOS | 0,08131 | 0,056662 | NA |
| 3 | abundance | X10265 | 3-methyl-4 | Handgrip | 0,141105 | 0,062218 | NA |
| 3 | abundance | X10276 | Maleamate | Tibia_length | 0,099216 | 0,049533 | NA |
| 3 | abundance | X10276 | Maleamate | Radius_length | 0,209397 | 0,065932 | NA |
| 3 | abundance | X10276 | Maleamate | Radius_SOS | 0,015183 | 0,049028 | NA |
| 3 | abundance | X10276 | Maleamate | Tibia_SOS | 0,110757 | 0,052229 | NA |
| 3 | abundance | X10276 | Maleamate | Handgrip | 0,068693 | 0,060189 | NA |
| 3 | abundance | X10279 | Lys-Pro | Tibia_length | 0,068173 | 0,05165 | NA |
| 3 | abundance | X10279 | Lys-Pro | Radius_length | 0,061977 | 0,070759 | NA |
| 3 | abundance | X10279 | Lys-Pro | Radius_SOS | 0,050258 | 0,049954 | NA |
| 3 | abundance | X10279 | Lys-Pro | Tibia_SOS | -0,11218 | 0,053388 | NA |
| 3 | abundance | X10279 | Lys-Pro | Handgrip | 0,010282 | 0,062535 | NA |
| 3 | abundance | X10304 | 2,3,8,9-Tet | Tibia_length | 0,017223 | 0,05322 | NA |
| 3 | abundance | X10304 | 2,3,8,9-Tet | Radius_length | 0,053191 | 0,072358 | NA |

| | | | | | | | |
|---|---|---|---|---|---|---|---|
| 3 | abundance | X10304 | 2,3,8,9-Tet | Radius_SOS | -0,02524 | 0,050748 | NA |
| 3 | abundance | X10304 | 2,3,8,9-Tet | Tibia_SOS | 0,082596 | 0,053318 | NA |
| 3 | abundance | X10304 | 2,3,8,9-Tet | Handgrip | -0,03372 | 0,060877 | NA |
| 2b | abundance | X10314 | 4-morphol | Tibia_length | -0,08555 | 0,052946 | NA |
| 2b | abundance | X10314 | 4-morphol | Radius_length | -0,14646 | 0,071511 | NA |
| 2b | abundance | X10314 | 4-morphol | Radius_SOS | 0,029972 | 0,050669 | NA |
| 2b | abundance | X10314 | 4-morphol | Tibia_SOS | 0,027701 | 0,053745 | NA |
| 2b | abundance | X10314 | 4-morphol | Handgrip | 0,00429 | 0,060885 | NA |
| 3 | abundance | X10319 | 1-Methyl-1 | Tibia_length | -0,04898 | 0,052849 | NA |
| 3 | abundance | X10319 | 1-Methyl-1 | Radius_length | 0,029399 | 0,071895 | NA |
| 3 | abundance | X10319 | 1-Methyl-1 | Radius_SOS | -0,02592 | 0,050662 | NA |
| 3 | abundance | X10319 | 1-Methyl-1 | Tibia_SOS | -0,07461 | 0,053792 | NA |
| 3 | abundance | X10319 | 1-Methyl-1 | Handgrip | -0,11527 | 0,061891 | NA |
| 3 | abundance | X10325 | Lovastatin | Tibia_length | 0,044831 | 0,051413 | NA |
| 3 | abundance | X10325 | Lovastatin | Radius_length | -0,0094 | 0,070892 | NA |
| 3 | abundance | X10325 | Lovastatin | Radius_SOS | -0,01154 | 0,049752 | NA |
| 3 | abundance | X10325 | Lovastatin | Tibia_SOS | 0,016759 | 0,053289 | NA |
| 3 | abundance | X10325 | Lovastatin | Handgrip | -0,07421 | 0,06208 | NA |
| 3 | abundance | X10330 | Sulfurous a | Tibia_length | -0,07706 | 0,052603 | NA |
| 3 | abundance | X10330 | Sulfurous a | Radius_length | -0,18794 | 0,070371 | NA |
| 3 | abundance | X10330 | Sulfurous a | Radius_SOS | -0,0797 | 0,050418 | NA |
| 3 | abundance | X10330 | Sulfurous a | Tibia_SOS | -0,04767 | 0,054185 | NA |
| 3 | abundance | X10330 | Sulfurous a | Handgrip | -0,03842 | 0,061614 | NA |
| 2b | abundance | X10339 | 4H-1-Benz | Tibia_length | 0,009642 | 0,055517 | NA |
| 2b | abundance | X10339 | 4H-1-Benz | Radius_length | -0,05303 | 0,074428 | NA |
| 2b | abundance | X10339 | 4H-1-Benz | Radius_SOS | 0,043082 | 0,052623 | NA |
| 2b | abundance | X10339 | 4H-1-Benz | Tibia_SOS | 0,030176 | 0,056535 | NA |
| 2b | abundance | X10339 | 4H-1-Benz | Handgrip | 0,044027 | 0,062501 | NA |
| 3 | abundance | X10345 | S-Allylcyst | Tibia_length | -0,02148 | 0,051249 | NA |
| 3 | abundance | X10345 | S-Allylcyst | Radius_length | 0,047166 | 0,069974 | NA |
| 3 | abundance | X10345 | S-Allylcyst | Radius_SOS | -0,10033 | 0,049542 | NA |
| 3 | abundance | X10345 | S-Allylcyst | Tibia_SOS | -0,01031 | 0,052969 | NA |
| 3 | abundance | X10345 | S-Allylcyst | Handgrip | 0,015073 | 0,060479 | NA |
| 3 | abundance | X10353 | L-gamma-( | Tibia_length | -0,13451 | 0,051286 | NA |
| 3 | abundance | X10353 | L-gamma-( | Radius_length | 0,012845 | 0,07057 | NA |
| 3 | abundance | X10353 | L-gamma-( | Radius_SOS | 0,01902 | 0,049714 | NA |
| 3 | abundance | X10353 | L-gamma-( | Tibia_SOS | -0,02432 | 0,053021 | NA |
| 3 | abundance | X10353 | L-gamma-( | Handgrip | -0,02644 | 0,060172 | NA |
| 3 | abundance | X10367 | 2-(1-Napht | Tibia_length | -0,07371 | 0,056755 | NA |
| 3 | abundance | X10367 | 2-(1-Napht | Radius_length | 0,010875 | 0,074491 | NA |
| 3 | abundance | X10367 | 2-(1-Napht | Radius_SOS | 0,015312 | 0,053823 | NA |
| 3 | abundance | X10367 | 2-(1-Napht | Tibia_SOS | 0,038281 | 0,057067 | NA |
| 3 | abundance | X10367 | 2-(1-Napht | Handgrip | 0,046293 | 0,063195 | NA |
| 3 | abundance | X10370 | L-Arogenat | Tibia_length | -0,01487 | 0,056491 | NA |
| 3 | abundance | X10370 | L-Arogenat | Radius_length | 0,023988 | 0,075024 | NA |
| 3 | abundance | X10370 | L-Arogenat | Radius_SOS | -0,03363 | 0,053643 | NA |
| 3 | abundance | X10370 | L-Arogenat | Tibia_SOS | -0,02154 | 0,055882 | NA |
| 3 | abundance | X10370 | L-Arogenat | Handgrip | -0,08589 | 0,062869 | NA |

| | | | | | | | | |
|---|---|---|---|---|---|---|---|---|
| 2b | | abundance | X10375 | 4-Indoleca | Tibia_length | 0,053272 | 0,052429 | NA |
| 2b | | abundance | X10375 | 4-Indoleca | Radius_length | 0,095185 | 0,07152 | NA |
| 2b | | abundance | X10375 | 4-Indoleca | Radius_SOS | -0,05069 | 0,05028 | NA |
| 2b | | abundance | X10375 | 4-Indoleca | Tibia_SOS | 0,08365 | 0,053365 | NA |
| 2b | | abundance | X10375 | 4-Indoleca | Handgrip | 0,045384 | 0,060598 | NA |
| | 3 | abundance | X10379 | Phenyl D-g | Tibia_length | -0,06079 | 0,053844 | NA |
| | 3 | abundance | X10379 | Phenyl D-g | Radius_length | -0,16079 | 0,071758 | NA |
| | 3 | abundance | X10379 | Phenyl D-g | Radius_SOS | -0,10268 | 0,051169 | NA |
| | 3 | abundance | X10379 | Phenyl D-g | Tibia_SOS | -0,05134 | 0,055774 | NA |
| | 3 | abundance | X10379 | Phenyl D-g | Handgrip | -0,09801 | 0,062303 | NA |
| | 3 | abundance | X10382 | Zalcitabine | Tibia_length | -0,08393 | 0,05116 | NA |
| | 3 | abundance | X10382 | Zalcitabine | Radius_length | -0,13333 | 0,069331 | NA |
| | 3 | abundance | X10382 | Zalcitabine | Radius_SOS | 0,014543 | 0,049421 | NA |
| | 3 | abundance | X10382 | Zalcitabine | Tibia_SOS | -0,06058 | 0,052823 | NA |
| | 3 | abundance | X10382 | Zalcitabine | Handgrip | 0,058339 | 0,060515 | NA |
| | 3 | abundance | X10384 | L-N2-(2-Ca | Tibia_length | -0,0014 | 0,059206 | NA |
| | 3 | abundance | X10384 | L-N2-(2-Ca | Radius_length | 0,105062 | 0,075741 | NA |
| | 3 | abundance | X10384 | L-N2-(2-Ca | Radius_SOS | -0,02547 | 0,056479 | NA |
| | 3 | abundance | X10384 | L-N2-(2-Ca | Tibia_SOS | 0,179291 | 0,057935 | NA |
| | 3 | abundance | X10384 | L-N2-(2-Ca | Handgrip | 0,078421 | 0,060008 | NA |
| | 3 | abundance | X10387 | Coenzyme | Tibia_length | 0,089777 | 0,05019 | NA |
| | 3 | abundance | X10387 | Coenzyme | Radius_length | 0,197456 | 0,068161 | NA |
| | 3 | abundance | X10387 | Coenzyme | Radius_SOS | 0,050677 | 0,04914 | NA |
| | 3 | abundance | X10387 | Coenzyme | Tibia_SOS | -0,01225 | 0,052705 | NA |
| | 3 | abundance | X10387 | Coenzyme | Handgrip | -0,00607 | 0,060372 | NA |
| | 3 | abundance | X10389 | Leucylaspa | Tibia_length | -0,06426 | 0,054758 | NA |
| | 3 | abundance | X10389 | Leucylaspa | Radius_length | 0,033346 | 0,073368 | NA |
| | 3 | abundance | X10389 | Leucylaspa | Radius_SOS | -0,00538 | 0,052377 | NA |
| | 3 | abundance | X10389 | Leucylaspa | Tibia_SOS | -9,50E-04 | 0,05652 | NA |
| | 3 | abundance | X10389 | Leucylaspa | Handgrip | -0,10897 | 0,062004 | NA |
| | 3 | abundance | X10393 | Ro 20-1724 | Tibia_length | -0,05413 | 0,051886 | NA |
| | 3 | abundance | X10393 | Ro 20-1724 | Radius_length | -0,04712 | 0,070816 | NA |
| | 3 | abundance | X10393 | Ro 20-1724 | Radius_SOS | -0,01191 | 0,050061 | NA |
| | 3 | abundance | X10393 | Ro 20-1724 | Tibia_SOS | 0,005242 | 0,053424 | NA |
| | 3 | abundance | X10393 | Ro 20-1724 | Handgrip | -0,00721 | 0,061103 | NA |
| | 1 | abundance | X10401 | 3-Methylxa | Tibia_length | -0,1079 | 0,0516 | NA |
| | 1 | abundance | X10401 | 3-Methylxa | Radius_length | -0,09416 | 0,070689 | NA |
| | 1 | abundance | X10401 | 3-Methylxa | Radius_SOS | 0,017363 | 0,049896 | NA |
| | 1 | abundance | X10401 | 3-Methylxa | Tibia_SOS | -0,07214 | 0,053348 | NA |
| | 1 | abundance | X10401 | 3-Methylxa | Handgrip | -0,0024 | 0,061597 | NA |
| | 3 | abundance | X10415 | beta-D-Glc | Tibia_length | 0,004523 | 0,054215 | NA |
| | 3 | abundance | X10415 | beta-D-Glc | Radius_length | 0,050039 | 0,072972 | NA |
| | 3 | abundance | X10415 | beta-D-Glc | Radius_SOS | 0,034281 | 0,051533 | NA |
| | 3 | abundance | X10415 | beta-D-Glc | Tibia_SOS | 0,112194 | 0,055152 | NA |
| | 3 | abundance | X10415 | beta-D-Glc | Handgrip | 0,059216 | 0,061296 | NA |
| 2b | | abundance | X10417 | L-Sacchar | Tibia_length | -0,09847 | 0,056041 | NA |
| 2b | | abundance | X10417 | L-Sacchar | Radius_length | 0,045416 | 0,075369 | NA |
| 2b | | abundance | X10417 | L-Sacchar | Radius_SOS | -0,01841 | 0,053618 | NA |

| | | | | | | | |
|---|---|---|---|---|---|---|---|
| 2b | abundance | X10417 | L-Sacchar( | Tibia_SOS | -0,06303 | 0,056695 | NA |
| 2b | abundance | X10417 | L-Sacchar( | Handgrip | -0,17289 | 0,060191 | NA |
| 2b | abundance | X10429 | 1,5-Isoquir | Tibia_length | -0,01101 | 0,059516 | NA |
| 2b | abundance | X10429 | 1,5-Isoquir | Radius_length | -0,00706 | 0,075892 | NA |
| 2b | abundance | X10429 | 1,5-Isoquir | Radius_SOS | -0,03733 | 0,056858 | NA |
| 2b | abundance | X10429 | 1,5-Isoquir | Tibia_SOS | 0,036982 | 0,05245 | NA |
| 2b | abundance | X10429 | 1,5-Isoquir | Handgrip | -0,02678 | 0,060171 | NA |
| 3 | abundance | X10442 | (6alpha,11 | Tibia_length | -0,03154 | 0,051056 | NA |
| 3 | abundance | X10442 | (6alpha,11 | Radius_length | 0,030258 | 0,070131 | NA |
| 3 | abundance | X10442 | (6alpha,11 | Radius_SOS | -0,04916 | 0,049467 | NA |
| 3 | abundance | X10442 | (6alpha,11 | Tibia_SOS | 0,041885 | 0,05358 | NA |
| 3 | abundance | X10442 | (6alpha,11 | Handgrip | 0,07838 | 0,060962 | NA |
| 3 | abundance | X10445 | 2-(Carboxy | Tibia_length | 0,030718 | 0,053434 | NA |
| 3 | abundance | X10445 | 2-(Carboxy | Radius_length | -0,04759 | 0,072521 | NA |
| 3 | abundance | X10445 | 2-(Carboxy | Radius_SOS | 0,025369 | 0,051051 | NA |
| 3 | abundance | X10445 | 2-(Carboxy | Tibia_SOS | -0,0429 | 0,054817 | NA |
| 3 | abundance | X10445 | 2-(Carboxy | Handgrip | 0,073507 | 0,062759 | NA |
| 3 | abundance | X10451 | Ro 20-172 | Tibia_length | 0,025152 | 0,053592 | NA |
| 3 | abundance | X10451 | Ro 20-172 | Radius_length | 0,101712 | 0,072471 | NA |
| 3 | abundance | X10451 | Ro 20-172 | Radius_SOS | 0,069148 | 0,051008 | NA |
| 3 | abundance | X10451 | Ro 20-172 | Tibia_SOS | 0,050442 | 0,055241 | NA |
| 3 | abundance | X10451 | Ro 20-172 | Handgrip | 1,16E-04 | 0,062184 | NA |
| 3 | abundance | X10454 | 8-Amino-7 | Tibia_length | -0,15379 | 0,053033 | NA |
| 3 | abundance | X10454 | 8-Amino-7 | Radius_length | -0,04634 | 0,072644 | NA |
| 3 | abundance | X10454 | 8-Amino-7 | Radius_SOS | 0,007134 | 0,050833 | NA |
| 3 | abundance | X10454 | 8-Amino-7 | Tibia_SOS | -0,07555 | 0,05473 | NA |
| 3 | abundance | X10454 | 8-Amino-7 | Handgrip | -0,14514 | 0,060624 | NA |
| 3 | abundance | X10457 | glu-ser | Tibia_length | -0,07871 | 0,05516 | NA |
| 3 | abundance | X10457 | glu-ser | Radius_length | -0,04788 | 0,073937 | NA |
| 3 | abundance | X10457 | glu-ser | Radius_SOS | -0,01213 | 0,052319 | NA |
| 3 | abundance | X10457 | glu-ser | Tibia_SOS | 0,045789 | 0,056751 | NA |
| 3 | abundance | X10457 | glu-ser | Handgrip | -0,03381 | 0,062247 | NA |
| 3 | abundance | X10476 | GAMMA-H\ | Tibia_length | -0,05669 | 0,057587 | NA |
| 3 | abundance | X10476 | GAMMA-H\ | Radius_length | 0,029901 | 0,075606 | NA |
| 3 | abundance | X10476 | GAMMA-H\ | Radius_SOS | -0,03352 | 0,05517 | NA |
| 3 | abundance | X10476 | GAMMA-H\ | Tibia_SOS | 0,095762 | 0,057459 | NA |
| 3 | abundance | X10476 | GAMMA-H\ | Handgrip | 0,083393 | 0,059983 | NA |
| 2b | abundance | X10486 | Coumarin | Tibia_length | 0,128856 | 0,049179 | NA |
| 2b | abundance | X10486 | Coumarin | Radius_length | 0,165877 | 0,065935 | NA |
| 2b | abundance | X10486 | Coumarin | Radius_SOS | 0,08044 | 0,048872 | NA |
| 2b | abundance | X10486 | Coumarin | Tibia_SOS | 0,134742 | 0,052036 | NA |
| 2b | abundance | X10486 | Coumarin | Handgrip | 0,078694 | 0,060013 | NA |
| 1 | abundance | X10488 | 4-Guanidir | Tibia_length | -0,09908 | 0,049896 | NA |
| 1 | abundance | X10488 | 4-Guanidir | Radius_length | -0,09338 | 0,068167 | NA |
| 1 | abundance | X10488 | 4-Guanidir | Radius_SOS | -0,01845 | 0,0491 | NA |
| 1 | abundance | X10488 | 4-Guanidir | Tibia_SOS | -0,01441 | 0,052548 | NA |
| 1 | abundance | X10488 | 4-Guanidir | Handgrip | 0,006416 | 0,060527 | NA |
| 3 | abundance | X10494 | 26Q0EO75 | Tibia_length | -0,05805 | 0,054548 | NA |

| | | | | | | | |
|---|---|---|---|---|---|---|---|
| 3 | abundance | X10494 | 26Q0EO75 | Radius_length | -0,01284 | 0,073389 | NA |
| 3 | abundance | X10494 | 26Q0EO75 | Radius_SOS | 0,091345 | 0,051659 | NA |
| 3 | abundance | X10494 | 26Q0EO75 | Tibia_SOS | 0,053464 | 0,054289 | NA |
| 3 | abundance | X10494 | 26Q0EO75 | Handgrip | 0,056562 | 0,061672 | NA |
| 3 | abundance | X10497 | 4-Guanidir | Tibia_length | -0,10038 | 0,048723 | NA |
| 3 | abundance | X10497 | 4-Guanidir | Radius_length | -0,01293 | 0,048966 | NA |
| 3 | abundance | X10497 | 4-Guanidir | Radius_SOS | -0,00935 | 0,049027 | NA |
| 3 | abundance | X10497 | 4-Guanidir | Tibia_SOS | -0,04522 | 0,052433 | NA |
| 3 | abundance | X10497 | 4-Guanidir | Handgrip | 0,053975 | 0,060105 | NA |
| 3 | abundance | X10503 | APM | Tibia_length | -0,17016 | 0,05907 | NA |
| 3 | abundance | X10503 | APM | Radius_length | -0,13001 | 0,075729 | NA |
| 3 | abundance | X10503 | APM | Radius_SOS | -0,08946 | 0,056259 | NA |
| 3 | abundance | X10503 | APM | Tibia_SOS | -0,15184 | 0,058495 | NA |
| 3 | abundance | X10503 | APM | Handgrip | -0,01218 | 0,060188 | NA |
| 3 | abundance | X10511 | dopaquino | Tibia_length | -0,02922 | 0,050748 | NA |
| 3 | abundance | X10511 | dopaquino | Radius_length | -0,06309 | 0,06934 | NA |
| 3 | abundance | X10511 | dopaquino | Radius_SOS | 0,032571 | 0,049307 | NA |
| 3 | abundance | X10511 | dopaquino | Tibia_SOS | -0,10259 | 0,05231 | NA |
| 3 | abundance | X10511 | dopaquino | Handgrip | -0,10363 | 0,060331 | NA |
| 3 | abundance | X10513 | ophthalmi | Tibia_length | 6,24E-04 | 0,054121 | NA |
| 3 | abundance | X10513 | ophthalmi | Radius_length | 0,022431 | 0,073246 | NA |
| 3 | abundance | X10513 | ophthalmi | Radius_SOS | 0,01673 | 0,051605 | NA |
| 3 | abundance | X10513 | ophthalmi | Tibia_SOS | 0,030784 | 0,056034 | NA |
| 3 | abundance | X10513 | ophthalmi | Handgrip | 0,125514 | 0,062753 | NA |
| 3 | abundance | X10518 | MFCD0002 | Tibia_length | -0,11893 | 0,057908 | NA |
| 3 | abundance | X10518 | MFCD0002 | Radius_length | 0,02933 | 0,075097 | NA |
| 3 | abundance | X10518 | MFCD0002 | Radius_SOS | -0,02112 | 0,05524 | NA |
| 3 | abundance | X10518 | MFCD0002 | Tibia_SOS | -0,00342 | 0,057342 | NA |
| 3 | abundance | X10518 | MFCD0002 | Handgrip | 0,025594 | 0,060173 | NA |
| 3 | abundance | X10534 | L-N2-(2-Ca | Tibia_length | 9,25E-04 | 0,050151 | NA |
| 3 | abundance | X10534 | L-N2-(2-Ca | Radius_length | -0,04488 | 0,06853 | NA |
| 3 | abundance | X10534 | L-N2-(2-Ca | Radius_SOS | -0,08561 | 0,048933 | NA |
| 3 | abundance | X10534 | L-N2-(2-Ca | Tibia_SOS | -0,05926 | 0,052682 | NA |
| 3 | abundance | X10534 | L-N2-(2-Ca | Handgrip | -0,06491 | 0,060205 | NA |
| 3 | abundance | X10536 | CMPF | Tibia_length | -0,06267 | 0,050801 | NA |
| 3 | abundance | X10536 | CMPF | Radius_length | -0,0237 | 0,069951 | NA |
| 3 | abundance | X10536 | CMPF | Radius_SOS | 0,058201 | 0,04928 | NA |
| 3 | abundance | X10536 | CMPF | Tibia_SOS | -0,09314 | 0,052828 | NA |
| 3 | abundance | X10536 | CMPF | Handgrip | -0,02771 | 0,060973 | NA |
| 2b | abundance | X10549 | Glaucine | Tibia_length | -0,06753 | 0,056005 | NA |
| 2b | abundance | X10549 | Glaucine | Radius_length | -0,0061 | 0,074098 | NA |
| 2b | abundance | X10549 | Glaucine | Radius_SOS | 0,009982 | 0,053083 | NA |
| 2b | abundance | X10549 | Glaucine | Tibia_SOS | -0,03206 | 0,05628 | NA |
| 2b | abundance | X10549 | Glaucine | Handgrip | -0,0245 | 0,063102 | NA |
| 3 | abundance | X10550 | pentobarbi | Tibia_length | -0,09755 | 0,053255 | NA |
| 3 | abundance | X10550 | pentobarbi | Radius_length | -0,15985 | 0,071562 | NA |
| 3 | abundance | X10550 | pentobarbi | Radius_SOS | 0,010467 | 0,050859 | NA |
| 3 | abundance | X10550 | pentobarbi | Tibia_SOS | -0,14903 | 0,053994 | NA |

| | | | | | | | | |
|---|---|---|---|---|---|---|---|---|
| | 3 | abundance | X10550 | pentobarbi | Handgrip | -0,07611 | 0,06169 | NA |
| | 1 | abundance | X10554 | Glycine | Tibia_length | 0,088119 | 0,05012 | NA |
| | 1 | abundance | X10554 | Glycine | Radius_length | 0,092077 | 0,068395 | NA |
| | 1 | abundance | X10554 | Glycine | Radius_SOS | -0,01614 | 0,049138 | NA |
| | 1 | abundance | X10554 | Glycine | Tibia_SOS | 0,079014 | 0,05251 | NA |
| | 1 | abundance | X10554 | Glycine | Handgrip | 0,08703 | 0,060138 | NA |
| | 3 | abundance | X10564 | 2-(1-Ethoxy | Tibia_length | 4,29E-04 | 0,051713 | NA |
| | 3 | abundance | X10564 | 2-(1-Ethoxy | Radius_length | 0,124221 | 0,070313 | NA |
| | 3 | abundance | X10564 | 2-(1-Ethoxy | Radius_SOS | 0,050264 | 0,049814 | NA |
| | 3 | abundance | X10564 | 2-(1-Ethoxy | Tibia_SOS | 0,058835 | 0,053609 | NA |
| | 3 | abundance | X10564 | 2-(1-Ethoxy | Handgrip | 0,062087 | 0,061427 | NA |
| | 3 | abundance | X10580 | Leu-Leu | Tibia_length | 0,041398 | 0,054631 | NA |
| | 3 | abundance | X10580 | Leu-Leu | Radius_length | 0,058291 | 0,07306 | NA |
| | 3 | abundance | X10580 | Leu-Leu | Radius_SOS | 0,058362 | 0,052309 | NA |
| | 3 | abundance | X10580 | Leu-Leu | Tibia_SOS | 0,033963 | 0,05501 | NA |
| | 3 | abundance | X10580 | Leu-Leu | Handgrip | -0,04665 | 0,062227 | NA |
| | 3 | abundance | X10588 | (8)-Ginger( | Tibia_length | 0,044372 | 0,051331 | NA |
| | 3 | abundance | X10588 | (8)-Ginger( | Radius_length | 0,093985 | 0,070089 | NA |
| | 3 | abundance | X10588 | (8)-Ginger( | Radius_SOS | 0,100839 | 0,049488 | NA |
| | 3 | abundance | X10588 | (8)-Ginger( | Tibia_SOS | 0,123579 | 0,053329 | NA |
| | 3 | abundance | X10588 | (8)-Ginger( | Handgrip | 0,057646 | 0,060875 | NA |
| | 3 | abundance | X10593 | Vorinostat | Tibia_length | -0,17411 | 0,053162 | NA |
| | 3 | abundance | X10593 | Vorinostat | Radius_length | -0,1249 | 0,072388 | NA |
| | 3 | abundance | X10593 | Vorinostat | Radius_SOS | 0,027436 | 0,051104 | NA |
| | 3 | abundance | X10593 | Vorinostat | Tibia_SOS | 0,001475 | 0,055781 | NA |
| | 3 | abundance | X10593 | Vorinostat | Handgrip | -0,00711 | 0,06203 | NA |
| | 3 | abundance | X10600 | Glycocyam | Tibia_length | -0,00129 | 0,05456 | NA |
| | 3 | abundance | X10600 | Glycocyam | Radius_length | 0,083033 | 0,073224 | NA |
| | 3 | abundance | X10600 | Glycocyam | Radius_SOS | 0,022204 | 0,051949 | NA |
| | 3 | abundance | X10600 | Glycocyam | Tibia_SOS | -0,06994 | 0,055378 | NA |
| | 3 | abundance | X10600 | Glycocyam | Handgrip | 0,191995 | 0,061366 | NA |
| | 3 | abundance | X10611 | 2-Acrylami | Tibia_length | 0,155823 | 0,052199 | NA |
| | 3 | abundance | X10611 | 2-Acrylami | Radius_length | 0,105456 | 0,071249 | NA |
| | 3 | abundance | X10611 | 2-Acrylami | Radius_SOS | 0,011709 | 0,050408 | NA |
| | 3 | abundance | X10611 | 2-Acrylami | Tibia_SOS | 0,138738 | 0,053222 | NA |
| | 3 | abundance | X10611 | 2-Acrylami | Handgrip | 0,102412 | 0,06042 | NA |
| 2a | | abundance | X10614 | Monometh | Tibia_length | -0,04283 | 0,053115 | NA |
| 2a | | abundance | X10614 | Monometh | Radius_length | -0,04734 | 0,071954 | NA |
| 2a | | abundance | X10614 | Monometh | Radius_SOS | -0,04376 | 0,050659 | NA |
| 2a | | abundance | X10614 | Monometh | Tibia_SOS | -0,1403 | 0,053638 | NA |
| 2a | | abundance | X10614 | Monometh | Handgrip | -0,09461 | 0,061393 | NA |
| 2b | | abundance | X10617 | 2-methyl-1 | Tibia_length | 0,117428 | 0,050984 | NA |
| 2b | | abundance | X10617 | 2-methyl-1 | Radius_length | 0,206438 | 0,068777 | NA |
| 2b | | abundance | X10617 | 2-methyl-1 | Radius_SOS | 0,03117 | 0,049521 | NA |
| 2b | | abundance | X10617 | 2-methyl-1 | Tibia_SOS | 0,124557 | 0,05272 | NA |
| 2b | | abundance | X10617 | 2-methyl-1 | Handgrip | 0,067617 | 0,060956 | NA |
| | 3 | abundance | X10623 | N-Nonanoy | Tibia_length | 0,114239 | 0,058213 | NA |
| | 3 | abundance | X10623 | N-Nonanoy | Radius_length | 0,122755 | 0,075569 | NA |

| | | | | | | | | |
|---|---|---|---|---|---|---|---|---|
| | 3 | abundance | X10623 | N-Nonanoy | Radius_SOS | -0,03999 | 0,055845 | NA |
| | 3 | abundance | X10623 | N-Nonanoy | Tibia_SOS | 0,021846 | 0,058851 | NA |
| | 3 | abundance | X10623 | N-Nonanoy | Handgrip | 0,008008 | 0,063794 | NA |
| 2b | | abundance | X10636 | 8-hydroxy- | Tibia_length | 0,041204 | 0,054428 | NA |
| 2b | | abundance | X10636 | 8-hydroxy- | Radius_length | 0,084032 | 0,072663 | NA |
| 2b | | abundance | X10636 | 8-hydroxy- | Radius_SOS | 0,026512 | 0,051903 | NA |
| 2b | | abundance | X10636 | 8-hydroxy- | Tibia_SOS | 0,022472 | 0,055924 | NA |
| 2b | | abundance | X10636 | 8-hydroxy- | Handgrip | 0,030428 | 0,06238 | NA |
| | 3 | abundance | X10647 | 4-Hydroxyp | Tibia_length | -0,01703 | 0,055579 | NA |
| | 3 | abundance | X10647 | 4-Hydroxyp | Radius_length | -0,08099 | 0,073287 | NA |
| | 3 | abundance | X10647 | 4-Hydroxyp | Radius_SOS | 0,029216 | 0,05262 | NA |
| | 3 | abundance | X10647 | 4-Hydroxyp | Tibia_SOS | 0,063316 | 0,056941 | NA |
| | 3 | abundance | X10647 | 4-Hydroxyp | Handgrip | 0,04325 | 0,062979 | NA |
| | 3 | abundance | X10651 | Methyl 1-h | Tibia_length | -0,11102 | 0,053096 | NA |
| | 3 | abundance | X10651 | Methyl 1-h | Radius_length | -0,17179 | 0,071081 | NA |
| | 3 | abundance | X10651 | Methyl 1-h | Radius_SOS | -0,02781 | 0,050738 | NA |
| | 3 | abundance | X10651 | Methyl 1-h | Tibia_SOS | -0,13543 | 0,053835 | NA |
| | 3 | abundance | X10651 | Methyl 1-h | Handgrip | -0,10799 | 0,061137 | NA |
| | 3 | abundance | X10660 | 2-Methylth | Tibia_length | 0,179829 | 0,050915 | NA |
| | 3 | abundance | X10660 | 2-Methylth | Radius_length | 0,132699 | 0,069745 | NA |
| | 3 | abundance | X10660 | 2-Methylth | Radius_SOS | -0,02165 | 0,049609 | NA |
| | 3 | abundance | X10660 | 2-Methylth | Tibia_SOS | 0,140965 | 0,052699 | NA |
| | 3 | abundance | X10660 | 2-Methylth | Handgrip | 0,047523 | 0,060445 | NA |
| 2b | | abundance | X10664 | N1-(5-metl | Tibia_length | -0,11855 | 0,056149 | NA |
| 2b | | abundance | X10664 | N1-(5-metl | Radius_length | -0,19907 | 0,072891 | NA |
| 2b | | abundance | X10664 | N1-(5-metl | Radius_SOS | 0,009345 | 0,05306 | NA |
| 2b | | abundance | X10664 | N1-(5-metl | Tibia_SOS | -0,00457 | 0,055554 | NA |
| 2b | | abundance | X10664 | N1-(5-metl | Handgrip | -0,09386 | 0,062056 | NA |
| | 3 | abundance | X10665 | Aprobarbit | Tibia_length | -0,04554 | 0,053021 | NA |
| | 3 | abundance | X10665 | Aprobarbit | Radius_length | 0,088063 | 0,07175 | NA |
| | 3 | abundance | X10665 | Aprobarbit | Radius_SOS | -0,02951 | 0,05076 | NA |
| | 3 | abundance | X10665 | Aprobarbit | Tibia_SOS | 0,091309 | 0,054783 | NA |
| | 3 | abundance | X10665 | Aprobarbit | Handgrip | -0,0453 | 0,061967 | NA |
| | 3 | abundance | X10667 | N-Phenyla | Tibia_length | -0,06766 | 0,057209 | NA |
| | 3 | abundance | X10667 | N-Phenyla | Radius_length | 0,109225 | 0,074367 | NA |
| | 3 | abundance | X10667 | N-Phenyla | Radius_SOS | 0,038978 | 0,054582 | NA |
| | 3 | abundance | X10667 | N-Phenyla | Tibia_SOS | 0,031039 | 0,058651 | NA |
| | 3 | abundance | X10667 | N-Phenyla | Handgrip | -0,07506 | 0,060023 | NA |
| | 3 | abundance | X10676 | 6-(1-Hydro | Tibia_length | -0,12521 | 0,054866 | NA |
| | 3 | abundance | X10676 | 6-(1-Hydro | Radius_length | -0,08425 | 0,074795 | NA |
| | 3 | abundance | X10676 | 6-(1-Hydro | Radius_SOS | -0,02035 | 0,052625 | NA |
| | 3 | abundance | X10676 | 6-(1-Hydro | Tibia_SOS | -0,06248 | 0,056025 | NA |
| | 3 | abundance | X10676 | 6-(1-Hydro | Handgrip | -0,22646 | 0,060833 | NA |
| | 3 | abundance | X10680 | 8-hydroxy- | Tibia_length | -0,00494 | 0,05068 | NA |
| | 3 | abundance | X10680 | 8-hydroxy- | Radius_length | 0,031112 | 0,069375 | NA |
| | 3 | abundance | X10680 | 8-hydroxy- | Radius_SOS | -0,00783 | 0,049324 | NA |
| | 3 | abundance | X10680 | 8-hydroxy- | Tibia_SOS | 0,010911 | 0,052878 | NA |
| | 3 | abundance | X10680 | 8-hydroxy- | Handgrip | -0,00199 | 0,061031 | NA |

| | | | | | | | |
|---|---|---|---|---|---|---|---|
| 2a | abundance | X10685 | Glycylvalin | Tibia_length | 0,059206 | 0,056986 | NA |
| 2a | abundance | X10685 | Glycylvalin | Radius_length | -0,06876 | 0,074864 | NA |
| 2a | abundance | X10685 | Glycylvalin | Radius_SOS | 0,021048 | 0,054213 | NA |
| 2a | abundance | X10685 | Glycylvalin | Tibia_SOS | 0,043454 | 0,057814 | NA |
| 2a | abundance | X10685 | Glycylvalin | Handgrip | 0,084574 | 0,06238 | NA |
| 3 | abundance | X10687 | Gln-Gln | Tibia_length | -0,02574 | 0,053195 | NA |
| 3 | abundance | X10687 | Gln-Gln | Radius_length | -0,01691 | 0,072106 | NA |
| 3 | abundance | X10687 | Gln-Gln | Radius_SOS | -0,02965 | 0,050873 | NA |
| 3 | abundance | X10687 | Gln-Gln | Tibia_SOS | 0,017694 | 0,054146 | NA |
| 3 | abundance | X10687 | Gln-Gln | Handgrip | 0,135337 | 0,060791 | NA |
| 3 | abundance | X10691 | (âˆ’)-nabilo | Tibia_length | 0,03596 | 0,05305 | NA |
| 3 | abundance | X10691 | (âˆ’)-nabilo | Radius_length | 0,048387 | 0,072151 | NA |
| 3 | abundance | X10691 | (âˆ’)-nabilo | Radius_SOS | 0,048786 | 0,050729 | NA |
| 3 | abundance | X10691 | (âˆ’)-nabilo | Tibia_SOS | 0,05104 | 0,054473 | NA |
| 3 | abundance | X10691 | (âˆ’)-nabilo | Handgrip | 0,018515 | 0,06153 | NA |
| 3 | abundance | X10706 | Furaneol | Tibia_length | 0,062299 | 0,051773 | NA |
| 3 | abundance | X10706 | Furaneol | Radius_length | 0,157073 | 0,069902 | NA |
| 3 | abundance | X10706 | Furaneol | Radius_SOS | 0,007466 | 0,04982 | NA |
| 3 | abundance | X10706 | Furaneol | Tibia_SOS | 0,198195 | 0,052305 | NA |
| 3 | abundance | X10706 | Furaneol | Handgrip | 0,075002 | 0,060748 | NA |
| 3 | abundance | X10713 | norhaman | Tibia_length | -0,06412 | 0,056636 | NA |
| 3 | abundance | X10713 | norhaman | Radius_length | -0,05613 | 0,075 | NA |
| 3 | abundance | X10713 | norhaman | Radius_SOS | -0,12552 | 0,053759 | NA |
| 3 | abundance | X10713 | norhaman | Tibia_SOS | 0,008521 | 0,05686 | NA |
| 3 | abundance | X10713 | norhaman | Handgrip | -0,04504 | 0,063131 | NA |
| 2b | abundance | X10722 | Acetanilide | Tibia_length | 0,128947 | 0,049257 | NA |
| 2b | abundance | X10722 | Acetanilide | Radius_length | 0,175492 | 0,066015 | NA |
| 2b | abundance | X10722 | Acetanilide | Radius_SOS | 0,076415 | 0,048886 | NA |
| 2b | abundance | X10722 | Acetanilide | Tibia_SOS | 0,143786 | 0,051963 | NA |
| 2b | abundance | X10722 | Acetanilide | Handgrip | 0,069061 | 0,060055 | NA |
| 3 | abundance | X10725 | Ethyl mala | Tibia_length | -0,05379 | 0,051701 | NA |
| 3 | abundance | X10725 | Ethyl mala | Radius_length | -0,0165 | 0,071009 | NA |
| 3 | abundance | X10725 | Ethyl mala | Radius_SOS | 0,019486 | 0,049837 | NA |
| 3 | abundance | X10725 | Ethyl mala | Tibia_SOS | 0,046422 | 0,05334 | NA |
| 3 | abundance | X10725 | Ethyl mala | Handgrip | 0,119242 | 0,060927 | NA |
| 3 | abundance | X10736 | Toxopyrimi | Tibia_length | -0,05254 | 0,05209 | NA |
| 3 | abundance | X10736 | Toxopyrimi | Radius_length | -0,11492 | 0,070947 | NA |
| 3 | abundance | X10736 | Toxopyrimi | Radius_SOS | 0,049583 | 0,04989 | NA |
| 3 | abundance | X10736 | Toxopyrimi | Tibia_SOS | 0,040079 | 0,053586 | NA |
| 3 | abundance | X10736 | Toxopyrimi | Handgrip | 0,075057 | 0,060849 | NA |
| 3 | abundance | X10741 | 5-Nitro-2-p | Tibia_length | 0,002812 | 0,056544 | NA |
| 3 | abundance | X10741 | 5-Nitro-2-p | Radius_length | -0,07672 | 0,074805 | NA |
| 3 | abundance | X10741 | 5-Nitro-2-p | Radius_SOS | -0,0287 | 0,053534 | NA |
| 3 | abundance | X10741 | 5-Nitro-2-p | Tibia_SOS | -0,00959 | 0,059506 | NA |
| 3 | abundance | X10741 | 5-Nitro-2-p | Handgrip | -0,10399 | 0,061231 | NA |
| 3 | abundance | X10757 | Glycyrin | Tibia_length | -0,00565 | 0,051732 | NA |
| 3 | abundance | X10757 | Glycyrin | Radius_length | -0,12583 | 0,070149 | NA |
| 3 | abundance | X10757 | Glycyrin | Radius_SOS | -0,00941 | 0,049835 | NA |

| | | | | | | | | |
|---|---|---|---|---|---|---|---|---|
| | 3 | abundance | X10757 | Glycyrin | Tibia_SOS | -0,05387 | 0,054123 | NA |
| | 3 | abundance | X10757 | Glycyrin | Handgrip | -0,02368 | 0,061502 | NA |
| | 1 | abundance | X10769 | Cystine | Tibia_length | 0,062309 | 0,049407 | NA |
| | 1 | abundance | X10769 | Cystine | Radius_length | 0,051246 | 0,06679 | NA |
| | 1 | abundance | X10769 | Cystine | Radius_SOS | 0,025553 | 0,049016 | NA |
| | 1 | abundance | X10769 | Cystine | Tibia_SOS | 0,053068 | 0,052413 | NA |
| | 1 | abundance | X10769 | Cystine | Handgrip | 0,082813 | 0,059998 | NA |
| | 3 | abundance | X10770 | 2-(4-Isoprc | Tibia_length | 0,006987 | 0,056164 | NA |
| | 3 | abundance | X10770 | 2-(4-Isoprc | Radius_length | 0,027084 | 0,074345 | NA |
| | 3 | abundance | X10770 | 2-(4-Isoprc | Radius_SOS | 0,103392 | 0,053336 | NA |
| | 3 | abundance | X10770 | 2-(4-Isoprc | Tibia_SOS | 0,009668 | 0,057353 | NA |
| | 3 | abundance | X10770 | 2-(4-Isoprc | Handgrip | -0,01218 | 0,060188 | NA |
| | 3 | abundance | X10772 | Bicine | Tibia_length | 0,032505 | 0,056634 | NA |
| | 3 | abundance | X10772 | Bicine | Radius_length | 0,029193 | 0,074769 | NA |
| | 3 | abundance | X10772 | Bicine | Radius_SOS | 0,020097 | 0,053814 | NA |
| | 3 | abundance | X10772 | Bicine | Tibia_SOS | 0,024178 | 0,057133 | NA |
| | 3 | abundance | X10772 | Bicine | Handgrip | 0,098589 | 0,062753 | NA |
| | 3 | abundance | X10776 | Histidylgly | Tibia_length | -0,03711 | 0,05514 | NA |
| | 3 | abundance | X10776 | Histidylgly | Radius_length | 0,085366 | 0,0739 | NA |
| | 3 | abundance | X10776 | Histidylgly | Radius_SOS | 0,001646 | 0,052389 | NA |
| | 3 | abundance | X10776 | Histidylgly | Tibia_SOS | 0,111027 | 0,055519 | NA |
| | 3 | abundance | X10776 | Histidylgly | Handgrip | 0,116692 | 0,062416 | NA |
| | 3 | abundance | X10793 | Zalcitabine | Tibia_length | -0,09575 | 0,055985 | NA |
| | 3 | abundance | X10793 | Zalcitabine | Radius_length | 0,025594 | 0,074459 | NA |
| | 3 | abundance | X10793 | Zalcitabine | Radius_SOS | 0,037743 | 0,053532 | NA |
| | 3 | abundance | X10793 | Zalcitabine | Tibia_SOS | 0,081892 | 0,056274 | NA |
| | 3 | abundance | X10793 | Zalcitabine | Handgrip | 0,081323 | 0,061287 | NA |
| | 3 | abundance | X10818 | butalbital | Tibia_length | 0,106893 | 0,053869 | NA |
| | 3 | abundance | X10818 | butalbital | Radius_length | 0,076143 | 0,073081 | NA |
| | 3 | abundance | X10818 | butalbital | Radius_SOS | 0,108841 | 0,051581 | NA |
| | 3 | abundance | X10818 | butalbital | Tibia_SOS | 0,156525 | 0,054767 | NA |
| | 3 | abundance | X10818 | butalbital | Handgrip | -0,02949 | 0,062254 | NA |
| 2a | | abundance | X10819 | Hyocholic | Tibia_length | 0,038143 | 0,049433 | NA |
| 2a | | abundance | X10819 | Hyocholic | Radius_length | 0,060945 | 0,066462 | NA |
| 2a | | abundance | X10819 | Hyocholic | Radius_SOS | -0,00398 | 0,049033 | NA |
| 2a | | abundance | X10819 | Hyocholic | Tibia_SOS | -0,04949 | 0,052434 | NA |
| 2a | | abundance | X10819 | Hyocholic | Handgrip | -0,03289 | 0,060519 | NA |
| 2b | | abundance | X10864 | 5-amino-2 | Tibia_length | -0,07987 | 0,055059 | NA |
| 2b | | abundance | X10864 | 5-amino-2 | Radius_length | -0,07844 | 0,073284 | NA |
| 2b | | abundance | X10864 | 5-amino-2 | Radius_SOS | 0,038973 | 0,052458 | NA |
| 2b | | abundance | X10864 | 5-amino-2 | Tibia_SOS | -0,01342 | 0,054744 | NA |
| 2b | | abundance | X10864 | 5-amino-2 | Handgrip | -0,15264 | 0,061085 | NA |
| | 3 | abundance | X10865 | Melatonin | Tibia_length | -0,04615 | 0,054208 | NA |
| | 3 | abundance | X10865 | Melatonin | Radius_length | 0,005964 | 0,072881 | NA |
| | 3 | abundance | X10865 | Melatonin | Radius_SOS | -0,0184 | 0,051581 | NA |
| | 3 | abundance | X10865 | Melatonin | Tibia_SOS | 0,022843 | 0,055187 | NA |
| | 3 | abundance | X10865 | Melatonin | Handgrip | 0,051998 | 0,061947 | NA |
| | 3 | abundance | X10881 | L-gamma-( | Tibia_length | -0,02533 | 0,053342 | NA |

| | | | | | | | | |
|---|---|---|---|---|---|---|---|---|
| | 3 | abundance | X10881 | L-gamma-( | Radius_length | -0,02522 | 0,072749 | NA |
| | 3 | abundance | X10881 | L-gamma-( | Radius_SOS | 0,016428 | 0,050973 | NA |
| | 3 | abundance | X10881 | L-gamma-( | Tibia_SOS | -0,01295 | 0,054953 | NA |
| | 3 | abundance | X10881 | L-gamma-( | Handgrip | -0,0218 | 0,060712 | NA |
| | 3 | abundance | X10886 | L-gamma-( | Tibia_length | -0,05113 | 0,05099 | NA |
| | 3 | abundance | X10886 | L-gamma-( | Radius_length | -0,13952 | 0,068901 | NA |
| | 3 | abundance | X10886 | L-gamma-( | Radius_SOS | 0,004486 | 0,049336 | NA |
| | 3 | abundance | X10886 | L-gamma-( | Tibia_SOS | -0,03701 | 0,052809 | NA |
| | 3 | abundance | X10886 | L-gamma-( | Handgrip | -0,08308 | 0,061035 | NA |
| | 3 | abundance | X10889 | N-acetyl-b | Tibia_length | -0,09487 | 0,056135 | NA |
| | 3 | abundance | X10889 | N-acetyl-b | Radius_length | 0,014562 | 0,074753 | NA |
| | 3 | abundance | X10889 | N-acetyl-b | Radius_SOS | -0,0155 | 0,053699 | NA |
| | 3 | abundance | X10889 | N-acetyl-b | Tibia_SOS | 0,006646 | 0,058374 | NA |
| | 3 | abundance | X10889 | N-acetyl-b | Handgrip | -0,0266 | 0,063209 | NA |
| 2b | | abundance | X10896 | 6-Hydroxyr | Tibia_length | 0,014688 | 0,054222 | NA |
| 2b | | abundance | X10896 | 6-Hydroxyr | Radius_length | 0,030253 | 0,073073 | NA |
| 2b | | abundance | X10896 | 6-Hydroxyr | Radius_SOS | 0,042061 | 0,051616 | NA |
| 2b | | abundance | X10896 | 6-Hydroxyr | Tibia_SOS | 0,009368 | 0,054838 | NA |
| 2b | | abundance | X10896 | 6-Hydroxyr | Handgrip | -0,04056 | 0,06143 | NA |
| | 3 | abundance | X10901 | bis-noryan | Tibia_length | 0,105883 | 0,049847 | NA |
| | 3 | abundance | X10901 | bis-noryan | Radius_length | 0,242208 | 0,06637 | NA |
| | 3 | abundance | X10901 | bis-noryan | Radius_SOS | 0,062125 | 0,04896 | NA |
| | 3 | abundance | X10901 | bis-noryan | Tibia_SOS | 0,135274 | 0,052029 | NA |
| | 3 | abundance | X10901 | bis-noryan | Handgrip | 0,155631 | 0,05966 | NA |
| 2b | | abundance | X10902 | Isoquinolir | Tibia_length | 0,094877 | 0,056281 | NA |
| 2b | | abundance | X10902 | Isoquinolir | Radius_length | 0,072258 | 0,074296 | NA |
| 2b | | abundance | X10902 | Isoquinolir | Radius_SOS | -0,0854 | 0,053906 | NA |
| 2b | | abundance | X10902 | Isoquinolir | Tibia_SOS | 0,118345 | 0,056815 | NA |
| 2b | | abundance | X10902 | Isoquinolir | Handgrip | 0,145014 | 0,062382 | NA |
| | 3 | abundance | X10918 | meprobam | Tibia_length | 0,038575 | 0,050403 | NA |
| | 3 | abundance | X10918 | meprobam | Radius_length | 0,091911 | 0,068864 | NA |
| | 3 | abundance | X10918 | meprobam | Radius_SOS | -5,96E-04 | 0,049206 | NA |
| | 3 | abundance | X10918 | meprobam | Tibia_SOS | -0,06668 | 0,052572 | NA |
| | 3 | abundance | X10918 | meprobam | Handgrip | -0,19735 | 0,059587 | NA |
| | 3 | abundance | X10929 | Temozolon | Tibia_length | -0,04592 | 0,05636 | NA |
| | 3 | abundance | X10929 | Temozolon | Radius_length | 0,047465 | 0,074883 | NA |
| | 3 | abundance | X10929 | Temozolon | Radius_SOS | 0,014445 | 0,053551 | NA |
| | 3 | abundance | X10929 | Temozolon | Tibia_SOS | 0,093054 | 0,057321 | NA |
| | 3 | abundance | X10929 | Temozolon | Handgrip | 0,094927 | 0,062323 | NA |
| | 3 | abundance | X10930 | MFCD0995 | Tibia_length | 0,03017 | 0,054567 | NA |
| | 3 | abundance | X10930 | MFCD0995 | Radius_length | 0,049861 | 0,073123 | NA |
| | 3 | abundance | X10930 | MFCD0995 | Radius_SOS | 0,068189 | 0,052157 | NA |
| | 3 | abundance | X10930 | MFCD0995 | Tibia_SOS | -0,04696 | 0,054807 | NA |
| | 3 | abundance | X10930 | MFCD0995 | Handgrip | -0,1036 | 0,062953 | NA |
| | 3 | abundance | X10936 | clavulanic | Tibia_length | 0,003886 | 0,05182 | NA |
| | 3 | abundance | X10936 | clavulanic | Radius_length | -0,04436 | 0,070941 | NA |
| | 3 | abundance | X10936 | clavulanic | Radius_SOS | 0,040443 | 0,049844 | NA |
| | 3 | abundance | X10936 | clavulanic | Tibia_SOS | -0,0234 | 0,053408 | NA |

| | | | | | | | | |
|---|---|---|---|---|---|---|---|---|
| | 3 | abundance | X10936 | clavulanic | Handgrip | -0,07214 | 0,060639 | NA |
| | 3 | abundance | X10947 | 1,1'-[1,12-| | Tibia_length | 0,07165 | 0,050332 | NA |
| | 3 | abundance | X10947 | 1,1'-[1,12-| | Radius_length | 0,093059 | 0,068396 | NA |
| | 3 | abundance | X10947 | 1,1'-[1,12-| | Radius_SOS | 0,005253 | 0,049176 | NA |
| | 3 | abundance | X10947 | 1,1'-[1,12-| | Tibia_SOS | -0,01685 | 0,052742 | NA |
| | 3 | abundance | X10947 | 1,1'-[1,12-| | Handgrip | 0,008886 | 0,060584 | NA |
| | 3 | abundance | X10951 | 8-Methyl-8 | Tibia_length | -0,03872 | 0,049372 | NA |
| | 3 | abundance | X10951 | 8-Methyl-8 | Radius_length | 0,030798 | 0,066519 | NA |
| | 3 | abundance | X10951 | 8-Methyl-8 | Radius_SOS | -0,05605 | 0,048968 | NA |
| | 3 | abundance | X10951 | 8-Methyl-8 | Tibia_SOS | -0,03112 | 0,052593 | NA |
| | 3 | abundance | X10951 | 8-Methyl-8 | Handgrip | -0,02624 | 0,06018 | NA |
| | 3 | abundance | X10953 | Vorinostat | Tibia_length | -0,03339 | 0,051647 | NA |
| | 3 | abundance | X10953 | Vorinostat | Radius_length | -0,04915 | 0,070354 | NA |
| | 3 | abundance | X10953 | Vorinostat | Radius_SOS | -5,24E-04 | 0,04976 | NA |
| | 3 | abundance | X10953 | Vorinostat | Tibia_SOS | 0,026273 | 0,053178 | NA |
| | 3 | abundance | X10953 | Vorinostat | Handgrip | 0,030962 | 0,061654 | NA |
| | 3 | abundance | X10974 | Threonylglı | Tibia_length | -0,04208 | 0,056882 | NA |
| | 3 | abundance | X10974 | Threonylglı | Radius_length | 0,147572 | 0,074226 | NA |
| | 3 | abundance | X10974 | Threonylglı | Radius_SOS | -0,02341 | 0,054196 | NA |
| | 3 | abundance | X10974 | Threonylglı | Tibia_SOS | 0,006775 | 0,057345 | NA |
| | 3 | abundance | X10974 | Threonylglı | Handgrip | -0,03421 | 0,062758 | NA |
| | 3 | abundance | X10977 | pentobarbi | Tibia_length | -0,08498 | 0,055257 | NA |
| | 3 | abundance | X10977 | pentobarbi | Radius_length | -0,17664 | 0,072591 | NA |
| | 3 | abundance | X10977 | pentobarbi | Radius_SOS | -7,15E-04 | 0,05233 | NA |
| | 3 | abundance | X10977 | pentobarbi | Tibia_SOS | -0,0589 | 0,056745 | NA |
| | 3 | abundance | X10977 | pentobarbi | Handgrip | -0,04888 | 0,063438 | NA |
| | 3 | abundance | X10979 | acetyltauri | Tibia_length | -0,02733 | 0,053843 | NA |
| | 3 | abundance | X10979 | acetyltauri | Radius_length | -0,1109 | 0,072916 | NA |
| | 3 | abundance | X10979 | acetyltauri | Radius_SOS | -0,0386 | 0,051644 | NA |
| | 3 | abundance | X10979 | acetyltauri | Tibia_SOS | -0,01852 | 0,055653 | NA |
| | 3 | abundance | X10979 | acetyltauri | Handgrip | -0,00162 | 0,062145 | NA |
| | 3 | abundance | X10982 | 8-Methyl-8 | Tibia_length | -0,04096 | 0,049941 | NA |
| | 3 | abundance | X10982 | 8-Methyl-8 | Radius_length | 0,004285 | 0,068592 | NA |
| | 3 | abundance | X10982 | 8-Methyl-8 | Radius_SOS | -0,05465 | 0,048989 | NA |
| | 3 | abundance | X10982 | 8-Methyl-8 | Tibia_SOS | -0,01959 | 0,052593 | NA |
| | 3 | abundance | X10982 | 8-Methyl-8 | Handgrip | -0,14553 | 0,060037 | NA |
| | 3 | abundance | X10987 | 2-BUTYL Pl | Tibia_length | 0,010237 | 0,051698 | NA |
| | 3 | abundance | X10987 | 2-BUTYL Pl | Radius_length | 0,023515 | 0,070827 | NA |
| | 3 | abundance | X10987 | 2-BUTYL Pl | Radius_SOS | -0,00316 | 0,049827 | NA |
| | 3 | abundance | X10987 | 2-BUTYL Pl | Tibia_SOS | 0,109739 | 0,053079 | NA |
| | 3 | abundance | X10987 | 2-BUTYL Pl | Handgrip | 0,014175 | 0,061001 | NA |
| 2b | | abundance | X11001 | 2-[(carboxy | Tibia_length | -0,01399 | 0,053498 | NA |
| 2b | | abundance | X11001 | 2-[(carboxy | Radius_length | 0,084887 | 0,072483 | NA |
| 2b | | abundance | X11001 | 2-[(carboxy | Radius_SOS | 0,065543 | 0,050977 | NA |
| 2b | | abundance | X11001 | 2-[(carboxy | Tibia_SOS | 0,072588 | 0,054805 | NA |
| 2b | | abundance | X11001 | 2-[(carboxy | Handgrip | -0,05231 | 0,062091 | NA |
| | 3 | abundance | X11012 | Furan | Tibia_length | 0,005922 | 0,054961 | NA |
| | 3 | abundance | X11012 | Furan | Radius_length | -0,01515 | 0,073789 | NA |

| | | | | | | | |
|---|---|---|---|---|---|---|---|
| 3 | abundance | X11012 | Furan | Radius_SOS | -0,07174 | 0,052307 | NA |
| 3 | abundance | X11012 | Furan | Tibia_SOS | 0,107196 | 0,056423 | NA |
| 3 | abundance | X11012 | Furan | Handgrip | 0,060533 | 0,062823 | NA |
| 3 | abundance | X11028 | butalbital | Tibia_length | -0,11738 | 0,054105 | NA |
| 3 | abundance | X11028 | butalbital | Radius_length | -0,09058 | 0,072769 | NA |
| 3 | abundance | X11028 | butalbital | Radius_SOS | 0,041577 | 0,051958 | NA |
| 3 | abundance | X11028 | butalbital | Tibia_SOS | -0,02498 | 0,055062 | NA |
| 3 | abundance | X11028 | butalbital | Handgrip | -0,07645 | 0,061585 | NA |
| 3 | abundance | X11032 | 2,4-Bis(3-r | Tibia_length | 0,05747 | 0,050319 | NA |
| 3 | abundance | X11032 | 2,4-Bis(3-r | Radius_length | 0,201492 | 0,067469 | NA |
| 3 | abundance | X11032 | 2,4-Bis(3-r | Radius_SOS | 0,116883 | 0,048836 | NA |
| 3 | abundance | X11032 | 2,4-Bis(3-r | Tibia_SOS | 0,048286 | 0,052499 | NA |
| 3 | abundance | X11032 | 2,4-Bis(3-r | Handgrip | 0,039523 | 0,060431 | NA |
| 3 | abundance | X11033 | N-Acetyl-5 | Tibia_length | -6,69E-05 | 0,052785 | NA |
| 3 | abundance | X11033 | N-Acetyl-5 | Radius_length | 0,079828 | 0,072253 | NA |
| 3 | abundance | X11033 | N-Acetyl-5 | Radius_SOS | 0,036202 | 0,050683 | NA |
| 3 | abundance | X11033 | N-Acetyl-5 | Tibia_SOS | -0,05233 | 0,053333 | NA |
| 3 | abundance | X11033 | N-Acetyl-5 | Handgrip | -0,11752 | 0,060882 | NA |
| 3 | abundance | X11034 | 1-(3-Aminc | Tibia_length | -0,12681 | 0,053672 | NA |
| 3 | abundance | X11034 | 1-(3-Aminc | Radius_length | -0,05969 | 0,072663 | NA |
| 3 | abundance | X11034 | 1-(3-Aminc | Radius_SOS | -0,02466 | 0,051459 | NA |
| 3 | abundance | X11034 | 1-(3-Aminc | Tibia_SOS | -0,06057 | 0,055662 | NA |
| 3 | abundance | X11034 | 1-(3-Aminc | Handgrip | -0,04432 | 0,062609 | NA |
| 2b | abundance | X11036 | 3-Hydroxyp | Tibia_length | 0,154582 | 0,049942 | NA |
| 2b | abundance | X11036 | 3-Hydroxyp | Radius_length | 0,283469 | 0,066193 | NA |
| 2b | abundance | X11036 | 3-Hydroxyp | Radius_SOS | 0,044317 | 0,049114 | NA |
| 2b | abundance | X11036 | 3-Hydroxyp | Tibia_SOS | 0,162141 | 0,051953 | NA |
| 2b | abundance | X11036 | 3-Hydroxyp | Handgrip | 0,093731 | 0,060032 | NA |
| 3 | abundance | X11038 | (2E)-3-(3,4 | Tibia_length | 0,021091 | 0,055342 | NA |
| 3 | abundance | X11038 | (2E)-3-(3,4 | Radius_length | 0,010551 | 0,073882 | NA |
| 3 | abundance | X11038 | (2E)-3-(3,4 | Radius_SOS | 0,130081 | 0,052342 | NA |
| 3 | abundance | X11038 | (2E)-3-(3,4 | Tibia_SOS | -0,0317 | 0,055508 | NA |
| 3 | abundance | X11038 | (2E)-3-(3,4 | Handgrip | 0,052698 | 0,062053 | NA |
| 3 | abundance | X11042 | Leu-arg | Tibia_length | -0,15314 | 0,055276 | NA |
| 3 | abundance | X11042 | Leu-arg | Radius_length | -0,04031 | 0,073802 | NA |
| 3 | abundance | X11042 | Leu-arg | Radius_SOS | -0,05549 | 0,05233 | NA |
| 3 | abundance | X11042 | Leu-arg | Tibia_SOS | -0,06943 | 0,056619 | NA |
| 3 | abundance | X11042 | Leu-arg | Handgrip | 0,032876 | 0,062707 | NA |
| 3 | abundance | X11059 | Dihydrocoı | Tibia_length | 0,021791 | 0,050081 | NA |
| 3 | abundance | X11059 | Dihydrocoı | Radius_length | 0,058686 | 0,068159 | NA |
| 3 | abundance | X11059 | Dihydrocoı | Radius_SOS | -0,01914 | 0,049071 | NA |
| 3 | abundance | X11059 | Dihydrocoı | Tibia_SOS | 0,041161 | 0,052453 | NA |
| 3 | abundance | X11059 | Dihydrocoı | Handgrip | 0,088915 | 0,059976 | NA |
| 3 | abundance | X11067 | Resveratro | Tibia_length | 0,123911 | 0,051632 | NA |
| 3 | abundance | X11067 | Resveratro | Radius_length | 0,17658 | 0,070064 | NA |
| 3 | abundance | X11067 | Resveratro | Radius_SOS | 0,031288 | 0,049892 | NA |
| 3 | abundance | X11067 | Resveratro | Tibia_SOS | 0,14165 | 0,052632 | NA |
| 3 | abundance | X11067 | Resveratro | Handgrip | 0,004884 | 0,060766 | NA |

| | | | | | | | |
|---|---|---|---|---|---|---|---|
| 3 | abundance | X11068 | N,N'-Bis[4- | Tibia_length | -0,01527 | 0,052744 | NA |
| 3 | abundance | X11068 | N,N'-Bis[4- | Radius_length | -0,02427 | 0,071838 | NA |
| 3 | abundance | X11068 | N,N'-Bis[4- | Radius_SOS | 0,030367 | 0,050734 | NA |
| 3 | abundance | X11068 | N,N'-Bis[4- | Tibia_SOS | 0,078151 | 0,053998 | NA |
| 3 | abundance | X11068 | N,N'-Bis[4- | Handgrip | -0,01289 | 0,061604 | NA |
| 3 | abundance | X11110 | Tauropine | Tibia_length | -0,1275 | 0,056303 | NA |
| 3 | abundance | X11110 | Tauropine | Radius_length | -0,11531 | 0,074153 | NA |
| 3 | abundance | X11110 | Tauropine | Radius_SOS | 0,04279 | 0,05323 | NA |
| 3 | abundance | X11110 | Tauropine | Tibia_SOS | -0,03995 | 0,0571 | NA |
| 3 | abundance | X11110 | Tauropine | Handgrip | 0,040631 | 0,061847 | NA |
| 3 | abundance | X11115 | Hydroxyca | Tibia_length | -0,15206 | 0,054479 | NA |
| 3 | abundance | X11115 | Hydroxyca | Radius_length | -0,05765 | 0,073295 | NA |
| 3 | abundance | X11115 | Hydroxyca | Radius_SOS | -0,00158 | 0,052031 | NA |
| 3 | abundance | X11115 | Hydroxyca | Tibia_SOS | -0,03997 | 0,055322 | NA |
| 3 | abundance | X11115 | Hydroxyca | Handgrip | 0,049654 | 0,06181 | NA |
| 3 | abundance | X11117 | Coprine | Tibia_length | 7,70E-04 | 0,050897 | NA |
| 3 | abundance | X11117 | Coprine | Radius_length | 0,110585 | 0,069489 | NA |
| 3 | abundance | X11117 | Coprine | Radius_SOS | -0,00908 | 0,049409 | NA |
| 3 | abundance | X11117 | Coprine | Tibia_SOS | 0,03715 | 0,053233 | NA |
| 3 | abundance | X11117 | Coprine | Handgrip | 0,033942 | 0,060448 | NA |
| 3 | abundance | X11125 | 5,8,12-Trih | Tibia_length | -0,08103 | 0,050469 | NA |
| 3 | abundance | X11125 | 5,8,12-Trih | Radius_length | -0,09259 | 0,068834 | NA |
| 3 | abundance | X11125 | 5,8,12-Trih | Radius_SOS | 0,044237 | 0,049182 | NA |
| 3 | abundance | X11125 | 5,8,12-Trih | Tibia_SOS | -0,01126 | 0,052663 | NA |
| 3 | abundance | X11125 | 5,8,12-Trih | Handgrip | 0,001447 | 0,060754 | NA |
| 3 | abundance | X11140 | Leu-Leu | Tibia_length | -0,02843 | 0,053445 | NA |
| 3 | abundance | X11140 | Leu-Leu | Radius_length | -0,02712 | 0,072422 | NA |
| 3 | abundance | X11140 | Leu-Leu | Radius_SOS | -0,05183 | 0,051209 | NA |
| 3 | abundance | X11140 | Leu-Leu | Tibia_SOS | -0,07024 | 0,053968 | NA |
| 3 | abundance | X11140 | Leu-Leu | Handgrip | -0,05668 | 0,06143 | NA |
| 3 | abundance | X11163 | Lys-Pro | Tibia_length | 0,055175 | 0,049348 | NA |
| 3 | abundance | X11163 | Lys-Pro | Radius_length | 0,126984 | 0,06626 | NA |
| 3 | abundance | X11163 | Lys-Pro | Radius_SOS | 0,015498 | 0,049033 | NA |
| 3 | abundance | X11163 | Lys-Pro | Tibia_SOS | -0,06483 | 0,052485 | NA |
| 3 | abundance | X11163 | Lys-Pro | Handgrip | -0,03495 | 0,060175 | NA |
| 3 | abundance | X11170 | QJ972000( | Tibia_length | 0,164479 | 0,051997 | NA |
| 3 | abundance | X11170 | QJ972000( | Radius_length | 0,115252 | 0,071309 | NA |
| 3 | abundance | X11170 | QJ972000( | Radius_SOS | -0,05064 | 0,050367 | NA |
| 3 | abundance | X11170 | QJ972000( | Tibia_SOS | 0,112584 | 0,053292 | NA |
| 3 | abundance | X11170 | QJ972000( | Handgrip | 0,120497 | 0,06039 | NA |
| 2b | abundance | X11186 | Fluocinolo | Tibia_length | -0,00747 | 0,055504 | NA |
| 2b | abundance | X11186 | Fluocinolo | Radius_length | -0,0312 | 0,073521 | NA |
| 2b | abundance | X11186 | Fluocinolo | Radius_SOS | 0,005775 | 0,052835 | NA |
| 2b | abundance | X11186 | Fluocinolo | Tibia_SOS | -0,02304 | 0,056185 | NA |
| 2b | abundance | X11186 | Fluocinolo | Handgrip | 0,019418 | 0,063137 | NA |
| 3 | abundance | X11215 | Leu-Leu | Tibia_length | -0,02857 | 0,052224 | NA |
| 3 | abundance | X11215 | Leu-Leu | Radius_length | -0,13999 | 0,070634 | NA |
| 3 | abundance | X11215 | Leu-Leu | Radius_SOS | -0,03474 | 0,050065 | NA |

| | | | | | | | |
|---|---|---|---|---|---|---|---|
| 3 | abundance | X11215 | Leu-Leu | Tibia_SOS | -0,00671 | 0,054141 | NA |
| 3 | abundance | X11215 | Leu-Leu | Handgrip | -0,07461 | 0,061256 | NA |
| 3 | abundance | X11223 | Ethyl mala | Tibia_length | -0,0855 | 0,058623 | NA |
| 3 | abundance | X11223 | Ethyl mala | Radius_length | -0,00473 | 0,075831 | NA |
| 3 | abundance | X11223 | Ethyl mala | Radius_SOS | 0,045072 | 0,056106 | NA |
| 3 | abundance | X11223 | Ethyl mala | Tibia_SOS | -0,13618 | 0,057606 | NA |
| 3 | abundance | X11223 | Ethyl mala | Handgrip | 0,00638 | 0,063408 | NA |
| 3 | abundance | X11245 | MFCD0003 | Tibia_length | -0,13827 | 0,055616 | NA |
| 3 | abundance | X11245 | MFCD0003 | Radius_length | -0,0312 | 0,07455 | NA |
| 3 | abundance | X11245 | MFCD0003 | Radius_SOS | -0,02675 | 0,053522 | NA |
| 3 | abundance | X11245 | MFCD0003 | Tibia_SOS | -0,06403 | 0,057654 | NA |
| 3 | abundance | X11245 | MFCD0003 | Handgrip | 0,025606 | 0,062971 | NA |
| 3 | abundance | X11251 | N-Nonanoy | Tibia_length | -0,04286 | 0,052558 | NA |
| 3 | abundance | X11251 | N-Nonanoy | Radius_length | -0,01734 | 0,071696 | NA |
| 3 | abundance | X11251 | N-Nonanoy | Radius_SOS | -0,02683 | 0,050399 | NA |
| 3 | abundance | X11251 | N-Nonanoy | Tibia_SOS | 0,096789 | 0,054557 | NA |
| 3 | abundance | X11251 | N-Nonanoy | Handgrip | 0,039805 | 0,061807 | NA |
| 3 | abundance | X11254 | (4R,5S,9S, | Tibia_length | -0,03232 | 0,049804 | NA |
| 3 | abundance | X11254 | (4R,5S,9S, | Radius_length | -0,03125 | 0,067627 | NA |
| 3 | abundance | X11254 | (4R,5S,9S, | Radius_SOS | 0,079106 | 0,048885 | NA |
| 3 | abundance | X11254 | (4R,5S,9S, | Tibia_SOS | 0,034428 | 0,052462 | NA |
| 3 | abundance | X11254 | (4R,5S,9S, | Handgrip | -0,02885 | 0,06018 | NA |
| 2b | abundance | X11257 | 1-Methylgu | Tibia_length | -0,05844 | 0,052625 | NA |
| 2b | abundance | X11257 | 1-Methylgu | Radius_length | 0,007934 | 0,071654 | NA |
| 2b | abundance | X11257 | 1-Methylgu | Radius_SOS | -0,1174 | 0,050527 | NA |
| 2b | abundance | X11257 | 1-Methylgu | Tibia_SOS | 0,052529 | 0,053639 | NA |
| 2b | abundance | X11257 | 1-Methylgu | Handgrip | 0,03968 | 0,061718 | NA |
| 3 | abundance | X11258 | Valyl-4-hyc | Tibia_length | 0,049789 | 0,053947 | NA |
| 3 | abundance | X11258 | Valyl-4-hyc | Radius_length | 0,028295 | 0,07267 | NA |
| 3 | abundance | X11258 | Valyl-4-hyc | Radius_SOS | 0,04049 | 0,05153 | NA |
| 3 | abundance | X11258 | Valyl-4-hyc | Tibia_SOS | 0,022851 | 0,054707 | NA |
| 3 | abundance | X11258 | Valyl-4-hyc | Handgrip | -0,03503 | 0,061812 | NA |
| 3 | abundance | X11265 | Sulfurol | Tibia_length | -0,06036 | 0,055286 | NA |
| 3 | abundance | X11265 | Sulfurol | Radius_length | -0,0806 | 0,073367 | NA |
| 3 | abundance | X11265 | Sulfurol | Radius_SOS | -0,03619 | 0,05258 | NA |
| 3 | abundance | X11265 | Sulfurol | Tibia_SOS | 0,076923 | 0,055348 | NA |
| 3 | abundance | X11265 | Sulfurol | Handgrip | -0,06465 | 0,062023 | NA |
| 3 | abundance | X11273 | trp-pro | Tibia_length | -0,01046 | 0,051215 | NA |
| 3 | abundance | X11273 | trp-pro | Radius_length | 0,05933 | 0,070151 | NA |
| 3 | abundance | X11273 | trp-pro | Radius_SOS | -0,05671 | 0,049468 | NA |
| 3 | abundance | X11273 | trp-pro | Tibia_SOS | 0,024148 | 0,053354 | NA |
| 3 | abundance | X11273 | trp-pro | Handgrip | 0,047567 | 0,06069 | NA |
| 3 | abundance | X11277 | lys-leu | Tibia_length | 0,082731 | 0,053552 | NA |
| 3 | abundance | X11277 | lys-leu | Radius_length | 0,042809 | 0,072576 | NA |
| 3 | abundance | X11277 | lys-leu | Radius_SOS | 0,01048 | 0,051372 | NA |
| 3 | abundance | X11277 | lys-leu | Tibia_SOS | 0,035179 | 0,055896 | NA |
| 3 | abundance | X11277 | lys-leu | Handgrip | 0,122517 | 0,062819 | NA |
| 3 | abundance | X11281 | Penbutolol | Tibia_length | 0,050149 | 0,057084 | NA |

| | | | | | | | |
|---|---|---|---|---|---|---|---|
| 3 | abundance | X11281 | Penbutolol | Radius_length | 0,135145 | 0,074786 | NA |
| 3 | abundance | X11281 | Penbutolol | Radius_SOS | 0,003582 | 0,054273 | NA |
| 3 | abundance | X11281 | Penbutolol | Tibia_SOS | -0,04431 | 0,057266 | NA |
| 3 | abundance | X11281 | Penbutolol | Handgrip | 0,082006 | 0,062885 | NA |
| 3 | abundance | X11301 | (6S)-2-Ami | Tibia_length | -0,04977 | 0,053886 | NA |
| 3 | abundance | X11301 | (6S)-2-Ami | Radius_length | -0,06583 | 0,072815 | NA |
| 3 | abundance | X11301 | (6S)-2-Ami | Radius_SOS | -0,07793 | 0,0511 | NA |
| 3 | abundance | X11301 | (6S)-2-Ami | Tibia_SOS | -0,04428 | 0,053923 | NA |
| 3 | abundance | X11301 | (6S)-2-Ami | Handgrip | 0,019104 | 0,061905 | NA |
| 3 | abundance | X11310 | Leu-Val | Tibia_length | 0,15967 | 0,05192 | NA |
| 3 | abundance | X11310 | Leu-Val | Radius_length | 0,103905 | 0,071555 | NA |
| 3 | abundance | X11310 | Leu-Val | Radius_SOS | 0,079713 | 0,050278 | NA |
| 3 | abundance | X11310 | Leu-Val | Tibia_SOS | 0,090358 | 0,053965 | NA |
| 3 | abundance | X11310 | Leu-Val | Handgrip | 0,075329 | 0,061548 | NA |
| 3 | abundance | X11319 | Methohexit | Tibia_length | 0,037731 | 0,054617 | NA |
| 3 | abundance | X11319 | Methohexit | Radius_length | 0,041965 | 0,074187 | NA |
| 3 | abundance | X11319 | Methohexit | Radius_SOS | -0,02423 | 0,051814 | NA |
| 3 | abundance | X11319 | Methohexit | Tibia_SOS | -0,02155 | 0,054849 | NA |
| 3 | abundance | X11319 | Methohexit | Handgrip | 0,01476 | 0,061295 | NA |
| 3 | abundance | X11323 | dihydroxyb | Tibia_length | -0,06516 | 0,049668 | NA |
| 3 | abundance | X11323 | dihydroxyb | Radius_length | 0,073088 | 0,068072 | NA |
| 3 | abundance | X11323 | dihydroxyb | Radius_SOS | -1,77E-04 | 0,049064 | NA |
| 3 | abundance | X11323 | dihydroxyb | Tibia_SOS | 0,01664 | 0,052522 | NA |
| 3 | abundance | X11323 | dihydroxyb | Handgrip | -0,00423 | 0,0606 | NA |
| 3 | abundance | X11331 | N~6~-Octa | Tibia_length | -0,01528 | 0,049689 | NA |
| 3 | abundance | X11331 | N~6~-Octa | Radius_length | -0,06255 | 0,067199 | NA |
| 3 | abundance | X11331 | N~6~-Octa | Radius_SOS | 0,003637 | 0,049033 | NA |
| 3 | abundance | X11331 | N~6~-Octa | Tibia_SOS | -0,0907 | 0,052282 | NA |
| 3 | abundance | X11331 | N~6~-Octa | Handgrip | -0,07715 | 0,060069 | NA |
| 3 | abundance | X11339 | L-gamma-( | Tibia_length | -0,03176 | 0,051125 | NA |
| 3 | abundance | X11339 | L-gamma-( | Radius_length | -0,0691 | 0,070062 | NA |
| 3 | abundance | X11339 | L-gamma-( | Radius_SOS | -0,01232 | 0,049565 | NA |
| 3 | abundance | X11339 | L-gamma-( | Tibia_SOS | 0,00387 | 0,05336 | NA |
| 3 | abundance | X11339 | L-gamma-( | Handgrip | -0,04464 | 0,061119 | NA |
| 3 | abundance | X11347 | Voglibose | Tibia_length | -0,0196 | 0,052181 | NA |
| 3 | abundance | X11347 | Voglibose | Radius_length | -0,08421 | 0,071951 | NA |
| 3 | abundance | X11347 | Voglibose | Radius_SOS | -0,04241 | 0,049997 | NA |
| 3 | abundance | X11347 | Voglibose | Tibia_SOS | -0,07152 | 0,053378 | NA |
| 3 | abundance | X11347 | Voglibose | Handgrip | -0,17173 | 0,060593 | NA |
| 3 | abundance | X11349 | Butabarbit | Tibia_length | -0,08249 | 0,053622 | NA |
| 3 | abundance | X11349 | Butabarbit | Radius_length | -0,11782 | 0,071922 | NA |
| 3 | abundance | X11349 | Butabarbit | Radius_SOS | -0,01501 | 0,051275 | NA |
| 3 | abundance | X11349 | Butabarbit | Tibia_SOS | -0,07096 | 0,053109 | NA |
| 3 | abundance | X11349 | Butabarbit | Handgrip | -0,17799 | 0,060289 | NA |
| 3 | abundance | X11354 | 3-Oxo-4,6- | Tibia_length | 0,045547 | 0,050951 | NA |
| 3 | abundance | X11354 | 3-Oxo-4,6- | Radius_length | 0,070868 | 0,069616 | NA |
| 3 | abundance | X11354 | 3-Oxo-4,6- | Radius_SOS | 0,09886 | 0,049199 | NA |
| 3 | abundance | X11354 | 3-Oxo-4,6- | Tibia_SOS | -0,02946 | 0,0531 | NA |

| | | | | | | | |
|---|---|---|---|---|---|---|---|
| 3 | abundance | X11354 | 3-Oxo-4,6- | Handgrip | -0,04925 | 0,061154 | NA |
| 3 | abundance | X11361 | 1-Methylhi | Tibia_length | -0,138 | 0,048774 | NA |
| 3 | abundance | X11361 | 1-Methylhi | Radius_length | -0,19979 | 0,064294 | NA |
| 3 | abundance | X11361 | 1-Methylhi | Radius_SOS | -0,04799 | 0,049113 | NA |
| 3 | abundance | X11361 | 1-Methylhi | Tibia_SOS | -0,0758 | 0,052655 | NA |
| 3 | abundance | X11361 | 1-Methylhi | Handgrip | -0,06356 | 0,060083 | NA |
| 3 | abundance | X11366 | 3-Sulfino-L | Tibia_length | -0,05369 | 0,054639 | NA |
| 3 | abundance | X11366 | 3-Sulfino-L | Radius_length | -0,11181 | 0,073327 | NA |
| 3 | abundance | X11366 | 3-Sulfino-L | Radius_SOS | -0,06479 | 0,052338 | NA |
| 3 | abundance | X11366 | 3-Sulfino-L | Tibia_SOS | -0,0365 | 0,056494 | NA |
| 3 | abundance | X11366 | 3-Sulfino-L | Handgrip | 0,016662 | 0,062185 | NA |
| 2b | abundance | X11373 | 3-Succinoy | Tibia_length | -0,08815 | 0,052017 | NA |
| 2b | abundance | X11373 | 3-Succinoy | Radius_length | -0,14358 | 0,070384 | NA |
| 2b | abundance | X11373 | 3-Succinoy | Radius_SOS | -0,04443 | 0,049868 | NA |
| 2b | abundance | X11373 | 3-Succinoy | Tibia_SOS | -0,03881 | 0,053791 | NA |
| 2b | abundance | X11373 | 3-Succinoy | Handgrip | -0,05022 | 0,060964 | NA |
| 3 | abundance | X11375 | L-gamma-( | Tibia_length | 0,063048 | 0,054491 | NA |
| 3 | abundance | X11375 | L-gamma-( | Radius_length | 0,041386 | 0,07344 | NA |
| 3 | abundance | X11375 | L-gamma-( | Radius_SOS | -0,08991 | 0,051708 | NA |
| 3 | abundance | X11375 | L-gamma-( | Tibia_SOS | 0,178839 | 0,055505 | NA |
| 3 | abundance | X11375 | L-gamma-( | Handgrip | 0,053698 | 0,062104 | NA |
| 3 | abundance | X11383 | N-{3-[(4-Ac | Tibia_length | 0,00298 | 0,055386 | NA |
| 3 | abundance | X11383 | N-{3-[(4-Ac | Radius_length | -0,04597 | 0,073342 | NA |
| 3 | abundance | X11383 | N-{3-[(4-Ac | Radius_SOS | -0,05043 | 0,052701 | NA |
| 3 | abundance | X11383 | N-{3-[(4-Ac | Tibia_SOS | -0,04596 | 0,057415 | NA |
| 3 | abundance | X11383 | N-{3-[(4-Ac | Handgrip | -0,06556 | 0,060063 | NA |
| 3 | abundance | X11408 | 5-Amino-6- | Tibia_length | -0,12883 | 0,053115 | NA |
| 3 | abundance | X11408 | 5-Amino-6- | Radius_length | -0,13989 | 0,071446 | NA |
| 3 | abundance | X11408 | 5-Amino-6- | Radius_SOS | -0,11937 | 0,050485 | NA |
| 3 | abundance | X11408 | 5-Amino-6- | Tibia_SOS | -0,07694 | 0,053388 | NA |
| 3 | abundance | X11408 | 5-Amino-6- | Handgrip | -0,04392 | 0,06085 | NA |
| 3 | abundance | X11421 | 8-Amino-7- | Tibia_length | -0,03937 | 0,053097 | NA |
| 3 | abundance | X11421 | 8-Amino-7- | Radius_length | 0,028334 | 0,072032 | NA |
| 3 | abundance | X11421 | 8-Amino-7- | Radius_SOS | 0,053583 | 0,050742 | NA |
| 3 | abundance | X11421 | 8-Amino-7- | Tibia_SOS | -0,03173 | 0,054387 | NA |
| 3 | abundance | X11421 | 8-Amino-7- | Handgrip | -0,105 | 0,060766 | NA |
| 3 | abundance | X11438 | pro-met | Tibia_length | -0,02797 | 0,054013 | NA |
| 3 | abundance | X11438 | pro-met | Radius_length | 0,025082 | 0,072991 | NA |
| 3 | abundance | X11438 | pro-met | Radius_SOS | -0,02456 | 0,051506 | NA |
| 3 | abundance | X11438 | pro-met | Tibia_SOS | -0,05996 | 0,055043 | NA |
| 3 | abundance | X11438 | pro-met | Handgrip | 0,02725 | 0,061204 | NA |
| 3 | abundance | X11441 | beta-D-Eth | Tibia_length | -0,10641 | 0,053195 | NA |
| 3 | abundance | X11441 | beta-D-Eth | Radius_length | -0,1006 | 0,07253 | NA |
| 3 | abundance | X11441 | beta-D-Eth | Radius_SOS | -0,05544 | 0,050873 | NA |
| 3 | abundance | X11441 | beta-D-Eth | Tibia_SOS | 0,017786 | 0,054969 | NA |
| 3 | abundance | X11441 | beta-D-Eth | Handgrip | -0,15857 | 0,061543 | NA |
| 3 | abundance | X11477 | gamma-Gl | Tibia_length | -0,07708 | 0,055681 | NA |
| 3 | abundance | X11477 | gamma-Gl | Radius_length | -0,09242 | 0,073939 | NA |

| | | | | | | | | |
|---|---|---|---|---|---|---|---|---|
| | 3 | abundance | X11477 | gamma-Gl | Radius_SOS | -0,0478 | 0,053228 | NA |
| | 3 | abundance | X11477 | gamma-Gl | Tibia_SOS | -0,03769 | 0,056207 | NA |
| | 3 | abundance | X11477 | gamma-Gl | Handgrip | -0,05539 | 0,060101 | NA |
| | 3 | abundance | X11481 | his-gln | Tibia_length | -0,07784 | 0,052138 | NA |
| | 3 | abundance | X11481 | his-gln | Radius_length | -0,06042 | 0,070812 | NA |
| | 3 | abundance | X11481 | his-gln | Radius_SOS | -0,04276 | 0,049924 | NA |
| | 3 | abundance | X11481 | his-gln | Tibia_SOS | -0,05205 | 0,054098 | NA |
| | 3 | abundance | X11481 | his-gln | Handgrip | 0,056883 | 0,062025 | NA |
| | 3 | abundance | X11494 | 3-Hydroxys | Tibia_length | -0,09187 | 0,053335 | NA |
| | 3 | abundance | X11494 | 3-Hydroxys | Radius_length | -0,11454 | 0,07192 | NA |
| | 3 | abundance | X11494 | 3-Hydroxys | Radius_SOS | 0,0612 | 0,050779 | NA |
| | 3 | abundance | X11494 | 3-Hydroxys | Tibia_SOS | -0,14401 | 0,053197 | NA |
| | 3 | abundance | X11494 | 3-Hydroxys | Handgrip | -0,12054 | 0,061196 | NA |
| | 3 | abundance | X11498 | Valylvaline | Tibia_length | -0,06828 | 0,054397 | NA |
| | 3 | abundance | X11498 | Valylvaline | Radius_length | -0,13512 | 0,072368 | NA |
| | 3 | abundance | X11498 | Valylvaline | Radius_SOS | 0,014875 | 0,051842 | NA |
| | 3 | abundance | X11498 | Valylvaline | Tibia_SOS | 0,051739 | 0,057604 | NA |
| | 3 | abundance | X11498 | Valylvaline | Handgrip | -0,00372 | 0,062693 | NA |
| | 3 | abundance | X11505 | FB950000( | Tibia_length | -0,006 | 0,055718 | NA |
| | 3 | abundance | X11505 | FB950000( | Radius_length | 0,002042 | 0,073806 | NA |
| | 3 | abundance | X11505 | FB950000( | Radius_SOS | 0,037275 | 0,05309 | NA |
| | 3 | abundance | X11505 | FB950000( | Tibia_SOS | -0,01138 | 0,055064 | NA |
| | 3 | abundance | X11505 | FB950000( | Handgrip | -0,02719 | 0,061936 | NA |
| 2b | | abundance | X11515 | 13(S)-HOT | Tibia_length | 0,078608 | 0,054567 | NA |
| 2b | | abundance | X11515 | 13(S)-HOT | Radius_length | 0,110711 | 0,073306 | NA |
| 2b | | abundance | X11515 | 13(S)-HOT | Radius_SOS | 0,024304 | 0,051932 | NA |
| 2b | | abundance | X11515 | 13(S)-HOT | Tibia_SOS | 0,024247 | 0,056115 | NA |
| 2b | | abundance | X11515 | 13(S)-HOT | Handgrip | -0,04113 | 0,062716 | NA |
| | 3 | abundance | X11522 | thr-trp | Tibia_length | -0,09404 | 0,054379 | NA |
| | 3 | abundance | X11522 | thr-trp | Radius_length | -0,13636 | 0,072342 | NA |
| | 3 | abundance | X11522 | thr-trp | Radius_SOS | -0,10162 | 0,05128 | NA |
| | 3 | abundance | X11522 | thr-trp | Tibia_SOS | -0,03325 | 0,056232 | NA |
| | 3 | abundance | X11522 | thr-trp | Handgrip | 0,041312 | 0,062715 | NA |
| | 3 | abundance | X11524 | 6-hydroxyp | Tibia_length | -0,09517 | 0,057523 | NA |
| | 3 | abundance | X11524 | 6-hydroxyp | Radius_length | -0,06374 | 0,074603 | NA |
| | 3 | abundance | X11524 | 6-hydroxyp | Radius_SOS | -0,02021 | 0,055202 | NA |
| | 3 | abundance | X11524 | 6-hydroxyp | Tibia_SOS | -0,06879 | 0,057552 | NA |
| | 3 | abundance | X11524 | 6-hydroxyp | Handgrip | -0,08084 | 0,059996 | NA |
| 2b | | abundance | X11529 | 2-Aminooc | Tibia_length | -0,04954 | 0,054866 | NA |
| 2b | | abundance | X11529 | 2-Aminooc | Radius_length | -0,01834 | 0,073182 | NA |
| 2b | | abundance | X11529 | 2-Aminooc | Radius_SOS | -0,03451 | 0,052209 | NA |
| 2b | | abundance | X11529 | 2-Aminooc | Tibia_SOS | 0,038232 | 0,055951 | NA |
| 2b | | abundance | X11529 | 2-Aminooc | Handgrip | -0,07999 | 0,061521 | NA |
| | 3 | abundance | X11549 | 1,1'-[1,12-l | Tibia_length | -0,00803 | 0,051711 | NA |
| | 3 | abundance | X11549 | 1,1'-[1,12-l | Radius_length | -0,03138 | 0,070721 | NA |
| | 3 | abundance | X11549 | 1,1'-[1,12-l | Radius_SOS | 0,113276 | 0,049445 | NA |
| | 3 | abundance | X11549 | 1,1'-[1,12-l | Tibia_SOS | -7,27E-04 | 0,053353 | NA |
| | 3 | abundance | X11549 | 1,1'-[1,12-l | Handgrip | 0,118909 | 0,060228 | NA |

| | | | | | | | |
|---|---|---|---|---|---|---|---|
| 3 | abundance | X11556 | N-[(2E)-3-( | Tibia_length | 0,102347 | 0,050752 | NA |
| 3 | abundance | X11556 | N-[(2E)-3-( | Radius_length | 0,115514 | 0,069545 | NA |
| 3 | abundance | X11556 | N-[(2E)-3-( | Radius_SOS | 0,010461 | 0,049411 | NA |
| 3 | abundance | X11556 | N-[(2E)-3-( | Tibia_SOS | 0,123794 | 0,052364 | NA |
| 3 | abundance | X11556 | N-[(2E)-3-( | Handgrip | 0,059561 | 0,060136 | NA |
| 3 | abundance | X11562 | ALA-PRO | Tibia_length | 0,018269 | 0,056659 | NA |
| 3 | abundance | X11562 | ALA-PRO | Radius_length | 0,002278 | 0,074568 | NA |
| 3 | abundance | X11562 | ALA-PRO | Radius_SOS | 0,083006 | 0,053723 | NA |
| 3 | abundance | X11562 | ALA-PRO | Tibia_SOS | 0,01613 | 0,05714 | NA |
| 3 | abundance | X11562 | ALA-PRO | Handgrip | -0,01191 | 0,060189 | NA |
| 3 | abundance | X11563 | threonylph | Tibia_length | -0,08928 | 0,051104 | NA |
| 3 | abundance | X11563 | threonylph | Radius_length | -0,14826 | 0,069605 | NA |
| 3 | abundance | X11563 | threonylph | Radius_SOS | -0,00359 | 0,049621 | NA |
| 3 | abundance | X11563 | threonylph | Tibia_SOS | -0,06253 | 0,052621 | NA |
| 3 | abundance | X11563 | threonylph | Handgrip | -0,1745 | 0,060415 | NA |
| 3 | abundance | X11576 | Phenyl D-g | Tibia_length | -0,0362 | 0,055726 | NA |
| 3 | abundance | X11576 | Phenyl D-g | Radius_length | -0,20399 | 0,072257 | NA |
| 3 | abundance | X11576 | Phenyl D-g | Radius_SOS | -0,10155 | 0,052928 | NA |
| 3 | abundance | X11576 | Phenyl D-g | Tibia_SOS | -0,0862 | 0,056726 | NA |
| 3 | abundance | X11576 | Phenyl D-g | Handgrip | -0,07188 | 0,063099 | NA |
| 3 | abundance | X11605 | 3-Methyl-2 | Tibia_length | 0,12315 | 0,052268 | NA |
| 3 | abundance | X11605 | 3-Methyl-2 | Radius_length | 0,148259 | 0,071401 | NA |
| 3 | abundance | X11605 | 3-Methyl-2 | Radius_SOS | 0,066732 | 0,05014 | NA |
| 3 | abundance | X11605 | 3-Methyl-2 | Tibia_SOS | 0,093133 | 0,05395 | NA |
| 3 | abundance | X11605 | 3-Methyl-2 | Handgrip | 0,181612 | 0,05966 | NA |
| 3 | abundance | X11614 | 2-Methylth | Tibia_length | 0,170678 | 0,050023 | NA |
| 3 | abundance | X11614 | 2-Methylth | Radius_length | 0,176449 | 0,067733 | NA |
| 3 | abundance | X11614 | 2-Methylth | Radius_SOS | 0,058042 | 0,049085 | NA |
| 3 | abundance | X11614 | 2-Methylth | Tibia_SOS | 0,145524 | 0,052229 | NA |
| 3 | abundance | X11614 | 2-Methylth | Handgrip | 0,120413 | 0,059757 | NA |
| 3 | abundance | X11627 | N-(3-aceta | Tibia_length | 0,017375 | 0,050788 | NA |
| 3 | abundance | X11627 | N-(3-aceta | Radius_length | 0,096769 | 0,069074 | NA |
| 3 | abundance | X11627 | N-(3-aceta | Radius_SOS | 0,068749 | 0,049236 | NA |
| 3 | abundance | X11627 | N-(3-aceta | Tibia_SOS | -0,03225 | 0,052902 | NA |
| 3 | abundance | X11627 | N-(3-aceta | Handgrip | 0,035361 | 0,061098 | NA |
| 3 | abundance | X11639 | Midodrine | Tibia_length | -0,14048 | 0,057077 | NA |
| 3 | abundance | X11639 | Midodrine | Radius_length | -0,08227 | 0,075202 | NA |
| 3 | abundance | X11639 | Midodrine | Radius_SOS | -0,03211 | 0,054011 | NA |
| 3 | abundance | X11639 | Midodrine | Tibia_SOS | -0,17998 | 0,056494 | NA |
| 3 | abundance | X11639 | Midodrine | Handgrip | -0,13347 | 0,06216 | NA |
| 3 | abundance | X11645 | glu-thr | Tibia_length | -0,02041 | 0,055833 | NA |
| 3 | abundance | X11645 | glu-thr | Radius_length | 0,021987 | 0,074674 | NA |
| 3 | abundance | X11645 | glu-thr | Radius_SOS | -0,00421 | 0,053039 | NA |
| 3 | abundance | X11645 | glu-thr | Tibia_SOS | 0,0327 | 0,056886 | NA |
| 3 | abundance | X11645 | glu-thr | Handgrip | -0,00615 | 0,062272 | NA |
| 3 | abundance | X11666 | N-(4-Hydrc | Tibia_length | -0,15223 | 0,050254 | NA |
| 3 | abundance | X11666 | N-(4-Hydrc | Radius_length | -0,12625 | 0,069036 | NA |
| 3 | abundance | X11666 | N-(4-Hydrc | Radius_SOS | -0,03871 | 0,049291 | NA |

| | | | | | | | |
|---|---|---|---|---|---|---|---|
| 3 | abundance | X11666 | N-(4-Hydrc | Tibia_SOS | -0,09268 | 0,052491 | NA |
| 3 | abundance | X11666 | N-(4-Hydrc | Handgrip | -0,11245 | 0,05999 | NA |
| 3 | abundance | X11667 | Tetraacety | Tibia_length | -0,0303 | 0,05296 | NA |
| 3 | abundance | X11667 | Tetraacety | Radius_length | -0,03054 | 0,072531 | NA |
| 3 | abundance | X11667 | Tetraacety | Radius_SOS | -0,01102 | 0,050721 | NA |
| 3 | abundance | X11667 | Tetraacety | Tibia_SOS | 0,02071 | 0,054975 | NA |
| 3 | abundance | X11667 | Tetraacety | Handgrip | -0,05568 | 0,061777 | NA |
| 3 | abundance | X11692 | 2_7-Anhyd | Tibia_length | -0,00575 | 0,053459 | NA |
| 3 | abundance | X11692 | 2_7-Anhyd | Radius_length | 0,056315 | 0,072835 | NA |
| 3 | abundance | X11692 | 2_7-Anhyd | Radius_SOS | -0,01301 | 0,051093 | NA |
| 3 | abundance | X11692 | 2_7-Anhyd | Tibia_SOS | 0,022063 | 0,054457 | NA |
| 3 | abundance | X11692 | 2_7-Anhyd | Handgrip | 0,084921 | 0,062466 | NA |
| 3 | abundance | X11698 | 1-(beta-D-I | Tibia_length | -0,00738 | 0,053302 | NA |
| 3 | abundance | X11698 | 1-(beta-D-I | Radius_length | 0,187096 | 0,070924 | NA |
| 3 | abundance | X11698 | 1-(beta-D-I | Radius_SOS | 0,090466 | 0,050779 | NA |
| 3 | abundance | X11698 | 1-(beta-D-I | Tibia_SOS | 0,163723 | 0,053974 | NA |
| 3 | abundance | X11698 | 1-(beta-D-I | Handgrip | 0,092677 | 0,062939 | NA |
| 3 | abundance | X11699 | {2-[2-(Isob | Tibia_length | -0,006 | 0,050013 | NA |
| 3 | abundance | X11699 | {2-[2-(Isob | Radius_length | 0,218077 | 0,066862 | NA |
| 3 | abundance | X11699 | {2-[2-(Isob | Radius_SOS | 0,039483 | 0,049055 | NA |
| 3 | abundance | X11699 | {2-[2-(Isob | Tibia_SOS | 0,066048 | 0,052429 | NA |
| 3 | abundance | X11699 | {2-[2-(Isob | Handgrip | -0,01913 | 0,060486 | NA |
| 3 | abundance | X11716 | ala-ser | Tibia_length | 0,03907 | 0,055341 | NA |
| 3 | abundance | X11716 | ala-ser | Radius_length | 0,029627 | 0,074162 | NA |
| 3 | abundance | X11716 | ala-ser | Radius_SOS | -0,01958 | 0,052696 | NA |
| 3 | abundance | X11716 | ala-ser | Tibia_SOS | 0,037359 | 0,055811 | NA |
| 3 | abundance | X11716 | ala-ser | Handgrip | 0,044096 | 0,062573 | NA |
| 3 | abundance | X11717 | nicotianan | Tibia_length | -0,00639 | 0,055566 | NA |
| 3 | abundance | X11717 | nicotianan | Radius_length | 0,147162 | 0,073129 | NA |
| 3 | abundance | X11717 | nicotianan | Radius_SOS | 0,050297 | 0,052768 | NA |
| 3 | abundance | X11717 | nicotianan | Tibia_SOS | 0,015448 | 0,056333 | NA |
| 3 | abundance | X11717 | nicotianan | Handgrip | 0,021625 | 0,062949 | NA |
| 3 | abundance | X11733 | 1,3-Dihydr | Tibia_length | 7,41E-04 | 0,052794 | NA |
| 3 | abundance | X11733 | 1,3-Dihydr | Radius_length | 0,00134 | 0,071987 | NA |
| 3 | abundance | X11733 | 1,3-Dihydr | Radius_SOS | -0,02389 | 0,050455 | NA |
| 3 | abundance | X11733 | 1,3-Dihydr | Tibia_SOS | -0,01644 | 0,054268 | NA |
| 3 | abundance | X11733 | 1,3-Dihydr | Handgrip | 6,04E-04 | 0,061814 | NA |
| 3 | abundance | X11745 | 2-Despipei | Tibia_length | -0,08914 | 0,054487 | NA |
| 3 | abundance | X11745 | 2-Despipei | Radius_length | -0,06255 | 0,072948 | NA |
| 3 | abundance | X11745 | 2-Despipei | Radius_SOS | -0,06091 | 0,051936 | NA |
| 3 | abundance | X11745 | 2-Despipei | Tibia_SOS | -0,07181 | 0,055348 | NA |
| 3 | abundance | X11745 | 2-Despipei | Handgrip | 0,022906 | 0,062301 | NA |
| 3 | abundance | X11755 | Furfuranol | Tibia_length | -0,00726 | 0,055152 | NA |
| 3 | abundance | X11755 | Furfuranol | Radius_length | -0,00384 | 0,074024 | NA |
| 3 | abundance | X11755 | Furfuranol | Radius_SOS | -0,04027 | 0,052448 | NA |
| 3 | abundance | X11755 | Furfuranol | Tibia_SOS | 0,123617 | 0,056687 | NA |
| 3 | abundance | X11755 | Furfuranol | Handgrip | 0,044696 | 0,062896 | NA |
| 1 | abundance | p_cresol | p-Cresol | Tibia_length | 0,078042 | 0,054031 | NA |

| | | | | | | | |
|---|---|---|---|---|---|---|---|
| 1 | abundance | p_cresol | p-Cresol | Radius_length | 0,019396 | 0,073198 | NA |
| 1 | abundance | p_cresol | p-Cresol | Radius_SOS | 0,063676 | 0,051886 | NA |
| 1 | abundance | p_cresol | p-Cresol | Tibia_SOS | 0,012656 | 0,055272 | NA |
| 1 | abundance | p_cresol | p-Cresol | Handgrip | -0,15135 | 0,061875 | NA |

| Test statist | Sample siz | Degrees of | P-value | FDR | FDR featur |
|---|---|---|---|---|---|
| 0,940161 | 417 | 340,954 | 0,347801 | 1 | 0,637437 |
| -0,53498 | 417 | 189,7765 | 0,593288 | 1 | 0,808395 |
| 0,478916 | 416 | 372,5932 | 0,632279 | 1 | 0,964878 |
| -0,70316 | 364 | 336,4325 | 0,482441 | 1 | 0,785988 |
| -2,06712 | 278 | 254,7753 | 0,039733 | 0,238397 | 0,360707 |
| 0,598382 | 409 | 336,793 | 0,549987 | 1 | 0,786367 |
| -0,46823 | 409 | 197,1044 | 0,640138 | 1 | 0,838647 |
| 0,795436 | 408 | 368,0154 | 0,426873 | 1 | 0,95743 |
| -0,5744 | 358 | 327,3629 | 0,566089 | 1 | 0,837838 |
| -2,68769 | 274 | 250,6664 | 0,007677 | 0,053737 | 0,199202 |
| -0,21408 | 421 | 325,5488 | 0,830615 | 1 | 0,936611 |
| 0,006491 | 421 | 183,3901 | 0,994828 | 1 | 0,997935 |
| 0,583166 | 420 | 361,0078 | 0,560146 | 1 | 0,960096 |
| 0,363202 | 367 | 316,4288 | 0,716697 | 1 | 0,913008 |
| 0,483672 | 280 | 257,2008 | 0,62903 | 1 | 0,901091 |
| -1,25644 | 420 | 354,6647 | 0,209785 | 1 | 0,498652 |
| -0,61736 | 420 | 193,7982 | 0,537725 | 1 | 0,771054 |
| 0,50878 | 419 | 386,6837 | 0,611197 | 1 | 0,964878 |
| 0,23152 | 367 | 334,1406 | 0,817052 | 1 | 0,95095 |
| -0,15297 | 280 | 262,5589 | 0,878539 | 1 | 0,973498 |
| -0,26556 | 242 | 221,8572 | 0,790822 | 1 | 0,919136 |
| -0,08735 | 242 | 145,0154 | 0,930513 | 1 | 0,97433 |
| 1,283233 | 241 | 221,3849 | 0,200752 | 1 | 0,928589 |
| -1,10253 | 214 | 207,0998 | 0,271509 | 1 | 0,609149 |
| 0,720072 | 156 | 150,7491 | 0,472596 | 1 | 0,824256 |
| -1,22492 | 421 | 351,1787 | 0,221427 | 1 | 0,51655 |
| -0,35925 | 421 | 187,4574 | 0,719812 | 1 | 0,875826 |
| -1,07603 | 420 | 382,4549 | 0,282592 | 1 | 0,943897 |
| 0,006183 | 367 | 335,4267 | 0,995071 | 1 | 0,997335 |
| 1,252247 | 280 | 257,7 | 0,211615 | 1 | 0,644251 |
| -0,23342 | 421 | 372,5824 | 0,815563 | 1 | 0,928288 |
| -0,4537 | 421 | 195,8242 | 0,650544 | 1 | 0,839358 |
| -0,77418 | 420 | 400,5356 | 0,439282 | 1 | 0,960096 |
| 2,206809 | 367 | 353,1883 | 0,02797 | 0,811136 | 0,257802 |
| 1,86746 | 280 | 264,5884 | 0,062943 | 1 | 0,422605 |
| -1,96618 | 421 | 385,4299 | 0,049995 | 1 | 0,246877 |
| -1,65893 | 421 | 206,0306 | 0,09865 | 1 | 0,379251 |
| 0,795725 | 420 | 410,4416 | 0,426652 | 1 | 0,95743 |
| -2,80953 | 367 | 360,0738 | 0,005232 | 0,172665 | 0,178183 |
| -3,05846 | 280 | 270,8291 | 0,002448 | 0,083218 | 0,135787 |
| 2,366297 | 421 | 402,303 | 0,01844 | 0,516322 | 0,158072 |
| 2,808307 | 421 | 215,5692 | 0,005438 | 0,174001 | 0,1403 |
| 1,407476 | 420 | 415,7315 | 0,160034 | 1 | 0,914005 |
| 1,707782 | 367 | 362,4631 | 0,088533 | 1 | 0,394732 |
| 1,432094 | 280 | 275,2263 | 0,153252 | 1 | 0,565026 |
| 3,424119 | 421 | 359,6481 | 6,88E-04 | 0,02339 | 0,051145 |
| 1,05236 | 421 | 192,1179 | 0,293956 | 1 | 0,590656 |

| | | | | | |
|---|---|---|---|---|---|
| 0,155587 | 420 | 392,2919 | 0,876438 | 1 | 0,982463 |
| 2,54507 | 367 | 331,5024 | 0,011378 | 0,364105 | 0,189546 |
| 1,007772 | 280 | 268,99 | 0,314469 | 1 | 0,734737 |
| 0,483361 | 421 | 371,3837 | 0,629124 | 1 | 0,833119 |
| 0,240464 | 421 | 196,9791 | 0,81022 | 1 | 0,926063 |
| 0,934101 | 420 | 400,4943 | 0,350815 | 1 | 0,947373 |
| 0,189168 | 367 | 348,1538 | 0,850071 | 1 | 0,956935 |
| 0,023022 | 280 | 269,5658 | 0,981649 | 1 | 0,99236 |
| -1,70657 | 421 | 352,8029 | 0,088782 | 1 | 0,319007 |
| 0,253945 | 421 | 191,287 | 0,799811 | 1 | 0,921861 |
| 0,23943 | 420 | 388,2938 | 0,810898 | 1 | 0,969427 |
| -0,39089 | 367 | 338,221 | 0,696124 | 1 | 0,905141 |
| 0,589659 | 280 | 271,5839 | 0,55591 | 1 | 0,856601 |
| 2,04899 | 421 | 349,6849 | 0,041209 | 0,989015 | 0,22748 |
| 2,343653 | 421 | 187,7855 | 0,020142 | 0,552641 | 0,198392 |
| 1,806514 | 420 | 389,22 | 0,07161 | 1 | 0,848246 |
| 1,434686 | 367 | 332,1031 | 0,152318 | 1 | 0,500093 |
| 3,033948 | 280 | 270,3887 | 0,002649 | 0,087406 | 0,135787 |
| -1,06506 | 421 | 315,9777 | 0,287664 | 1 | 0,584852 |
| 0,171008 | 421 | 178,7707 | 0,864411 | 1 | 0,947215 |
| -0,33363 | 420 | 349,3103 | 0,738862 | 1 | 0,969427 |
| 0,063846 | 367 | 296,4374 | 0,949136 | 1 | 0,979539 |
| -0,00323 | 280 | 253,2259 | 0,997425 | 1 | 0,998213 |
| -2,16886 | 421 | 362,1765 | 0,030743 | 0,79933 | 0,200565 |
| -2,7401 | 421 | 195,8886 | 0,00671 | 0,201289 | 0,141161 |
| -1,50798 | 420 | 395,1664 | 0,132359 | 1 | 0,888479 |
| -1,74521 | 367 | 343,5403 | 0,081843 | 1 | 0,381841 |
| -2,20158 | 280 | 264,5103 | 0,028559 | 0,913879 | 0,326641 |
| 0,263973 | 421 | 410,526 | 0,791933 | 1 | 0,919136 |
| 0,862549 | 421 | 224,4208 | 0,389306 | 1 | 0,664422 |
| 0,813359 | 420 | 415,754 | 0,416478 | 1 | 0,95743 |
| 1,250407 | 367 | 362,9911 | 0,211956 | 1 | 0,563532 |
| -0,12742 | 280 | 274,4961 | 0,898704 | 1 | 0,975243 |
| 2,516964 | 421 | 407,9843 | 0,01222 | 0,366585 | 0,131857 |
| 2,953631 | 421 | 225,9277 | 0,003473 | 0,118069 | 0,118336 |
| 0,448991 | 420 | 415,7268 | 0,653672 | 1 | 0,966867 |
| 2,948846 | 367 | 362,7826 | 0,003396 | 0,115475 | 0,176492 |
| 1,621911 | 280 | 275,9916 | 0,105964 | 1 | 0,492865 |
| -0,41496 | 421 | 415,4472 | 0,678388 | 1 | 0,861268 |
| 1,688314 | 421 | 237,3741 | 0,092664 | 1 | 0,373113 |
| -0,26217 | 420 | 412,3401 | 0,793324 | 1 | 0,969427 |
| 0,902331 | 367 | 359,1509 | 0,367486 | 1 | 0,678946 |
| 0,329149 | 280 | 275,9935 | 0,742293 | 1 | 0,94096 |
| -0,99344 | 421 | 322,885 | 0,321241 | 1 | 0,615134 |
| -1,64204 | 421 | 183,136 | 0,102298 | 1 | 0,383322 |
| -2,63786 | 420 | 352,5412 | 0,008713 | 0,296237 | 0,582436 |
| -1,29375 | 367 | 299,1008 | 0,196749 | 1 | 0,549567 |
| -1,56517 | 280 | 255,9602 | 0,118779 | 1 | 0,521866 |

| | | | | | |
|---|---|---|---|---|---|
| -0,60232 | 421 | 317,3493 | 0,54739 | 1 | 0,784719 |
| 1,294711 | 421 | 179,9817 | 0,197078 | 1 | 0,486881 |
| -0,52033 | 420 | 351,0539 | 0,60316 | 1 | 0,964878 |
| 1,603312 | 367 | 314,9177 | 0,109868 | 1 | 0,435936 |
| 1,527912 | 280 | 252,3714 | 0,127787 | 1 | 0,536498 |
| 1,655188 | 421 | 406,2959 | 0,098658 | 1 | 0,339344 |
| 1,443921 | 421 | 221,1176 | 0,150177 | 1 | 0,430627 |
| 0,150628 | 420 | 415,9924 | 0,880343 | 1 | 0,982463 |
| 1,262222 | 367 | 362,9999 | 0,207679 | 1 | 0,560883 |
| -0,24283 | 280 | 275,2554 | 0,808318 | 1 | 0,95747 |
| -1,70436 | 421 | 385,8626 | 0,089119 | 1 | 0,319007 |
| -1,49657 | 421 | 206,2703 | 0,136032 | 1 | 0,419225 |
| -1,7427 | 420 | 408,6207 | 0,082138 | 1 | 0,848246 |
| -0,79118 | 367 | 359,5352 | 0,42936 | 1 | 0,737794 |
| 0,068765 | 280 | 270,1818 | 0,945228 | 1 | 0,982385 |
| 1,060343 | 421 | 336,9783 | 0,289748 | 1 | 0,586895 |
| 2,614072 | 421 | 187,7901 | 0,009673 | 0,28051 | 0,149771 |
| -0,27911 | 420 | 376,9109 | 0,780312 | 1 | 0,969427 |
| 1,052621 | 367 | 332,1272 | 0,29328 | 1 | 0,627013 |
| 0,403816 | 280 | 253,6556 | 0,686688 | 1 | 0,915426 |
| -1,62472 | 421 | 397,1563 | 0,105016 | 1 | 0,347821 |
| -1,97115 | 421 | 214,2528 | 0,049994 | 1 | 0,289774 |
| -0,94915 | 420 | 414,9125 | 0,343098 | 1 | 0,947373 |
| -1,9049 | 367 | 362,1968 | 0,057585 | 1 | 0,338985 |
| -1,48843 | 280 | 275,2255 | 0,137783 | 1 | 0,54931 |
| -0,96941 | 421 | 390,2172 | 0,33294 | 1 | 0,62159 |
| -1,38648 | 421 | 208,0464 | 0,167085 | 1 | 0,453987 |
| 1,512837 | 420 | 412,2967 | 0,131087 | 1 | 0,888479 |
| -0,85037 | 367 | 357,8837 | 0,395689 | 1 | 0,707784 |
| -0,18545 | 280 | 269,6694 | 0,853014 | 1 | 0,966655 |
| -2,68048 | 421 | 416,8588 | 0,007643 | 0,244585 | 0,108664 |
| -2,82547 | 421 | 247,0705 | 0,005107 | 0,168528 | 0,137431 |
| -1,74236 | 420 | 407,8797 | 0,082199 | 1 | 0,848246 |
| -1,89804 | 367 | 354,409 | 0,058503 | 1 | 0,339097 |
| 0,020324 | 280 | 274,5196 | 0,9838 | 1 | 0,99236 |
| -0,34777 | 421 | 302,5929 | 0,728251 | 1 | 0,890807 |
| -0,46954 | 421 | 178,9269 | 0,639256 | 1 | 0,838432 |
| -0,45581 | 420 | 334,6956 | 0,648824 | 1 | 0,964878 |
| 0,363203 | 367 | 293,6671 | 0,716715 | 1 | 0,913008 |
| -0,78317 | 280 | 276 | 0,434201 | 1 | 0,803161 |
| -2,44696 | 421 | 372,3889 | 0,014868 | 0,431181 | 0,147897 |
| -2,07109 | 421 | 199,5111 | 0,039637 | 1 | 0,26998 |
| -0,5178 | 420 | 402,972 | 0,604879 | 1 | 0,964878 |
| -2,505 | 367 | 353,5256 | 0,012694 | 0,393522 | 0,189546 |
| -2,13344 | 280 | 268,3371 | 0,033796 | 1 | 0,329568 |
| -1,44548 | 421 | 406,113 | 0,149096 | 1 | 0,422053 |
| -1,66412 | 421 | 223,449 | 0,09749 | 1 | 0,379251 |
| 0,033953 | 420 | 415,9847 | 0,972931 | 1 | 0,997342 |

| | | | | | |
|---|---|---|---|---|---|
| -0,91906 | 367 | 362,8223 | 0,358677 | 1 | 0,674813 |
| -0,71913 | 280 | 275,3646 | 0,472672 | 1 | 0,824256 |
| -1,04844 | 421 | 355,155 | 0,295151 | 1 | 0,593552 |
| 0,288637 | 421 | 191,804 | 0,773171 | 1 | 0,909577 |
| 0,010961 | 420 | 386,7903 | 0,99126 | 1 | 0,997342 |
| -0,06582 | 367 | 353,1487 | 0,947561 | 1 | 0,979539 |
| -1,27541 | 280 | 266,0574 | 0,203276 | 1 | 0,632429 |
| -2,2182 | 421 | 351,1607 | 0,027181 | 0,733878 | 0,192509 |
| -2,3514 | 421 | 188,2281 | 0,019737 | 0,552641 | 0,195731 |
| 0,142906 | 420 | 387,8317 | 0,886439 | 1 | 0,982463 |
| -2,34106 | 367 | 344,5821 | 0,019799 | 0,593956 | 0,221237 |
| -1,85548 | 280 | 259,8525 | 0,06466 | 1 | 0,423666 |
| -0,58791 | 421 | 414,462 | 0,556912 | 1 | 0,791015 |
| 0,013211 | 421 | 236,8894 | 0,98947 | 1 | 0,997412 |
| 0,498721 | 420 | 413,2641 | 0,618241 | 1 | 0,964878 |
| -1,30821 | 367 | 360,0974 | 0,191637 | 1 | 0,542851 |
| -0,10244 | 280 | 275,5809 | 0,918479 | 1 | 0,977051 |
| 0,016815 | 421 | 354,8043 | 0,986594 | 1 | 0,998043 |
| -0,02817 | 421 | 192,4406 | 0,977556 | 1 | 0,991789 |
| 1,134841 | 420 | 386,8471 | 0,257145 | 1 | 0,943897 |
| -0,11695 | 367 | 336,7968 | 0,906971 | 1 | 0,970486 |
| 0,565797 | 280 | 260,2059 | 0,572019 | 1 | 0,869862 |
| 0,268707 | 421 | 416,653 | 0,788288 | 1 | 0,918755 |
| 0,879811 | 421 | 257,6631 | 0,379782 | 1 | 0,657096 |
| 0,310836 | 420 | 403,8049 | 0,756086 | 1 | 0,969427 |
| -0,6704 | 367 | 356,1869 | 0,503039 | 1 | 0,793601 |
| 0,210913 | 280 | 275,4307 | 0,833111 | 1 | 0,964667 |
| 0,528765 | 421 | 386,8889 | 0,597272 | 1 | 0,815122 |
| 0,090983 | 421 | 205,8635 | 0,927594 | 1 | 0,972749 |
| -0,48389 | 420 | 410,0513 | 0,628725 | 1 | 0,964878 |
| -0,414 | 367 | 353,8331 | 0,679123 | 1 | 0,899694 |
| 0,676458 | 280 | 266,6874 | 0,499336 | 1 | 0,831491 |
| -2,6456 | 421 | 365,7279 | 0,008507 | 0,263703 | 0,112451 |
| -2,75665 | 421 | 198,2909 | 0,006385 | 0,197924 | 0,141161 |
| -1,09554 | 420 | 401,7213 | 0,273936 | 1 | 0,943897 |
| -2,16331 | 367 | 348,7886 | 0,031196 | 0,873493 | 0,266905 |
| -1,46462 | 280 | 271,6023 | 0,144181 | 1 | 0,555466 |
| 2,10593 | 421 | 360,3432 | 0,035902 | 0,897543 | 0,216416 |
| 1,322744 | 421 | 192,6863 | 0,187488 | 1 | 0,475664 |
| 1,421111 | 420 | 390,0283 | 0,156083 | 1 | 0,914005 |
| 0,839448 | 367 | 341,739 | 0,401805 | 1 | 0,715745 |
| -0,95587 | 280 | 260,0617 | 0,340025 | 1 | 0,756894 |
| -3,19769 | 421 | 377,6591 | 0,001502 | 0,049572 | 0,069718 |
| -1,53722 | 421 | 200,3647 | 0,125818 | 1 | 0,410224 |
| -0,48511 | 420 | 406,6734 | 0,627857 | 1 | 0,964878 |
| -1,36002 | 367 | 349,9599 | 0,174699 | 1 | 0,523018 |
| 0,131238 | 280 | 270,0609 | 0,895684 | 1 | 0,975243 |
| -0,54607 | 421 | 391,3521 | 0,585329 | 1 | 0,807036 |

| | | | | | |
|---|---|---|---|---|---|
| 0,799957 | 421 | 207,0572 | 0,424652 | 1 | 0,693378 |
| -0,27468 | 420 | 411,7471 | 0,783699 | 1 | 0,969427 |
| 1,106627 | 367 | 355,1849 | 0,269204 | 1 | 0,609149 |
| 0,737109 | 280 | 271,634 | 0,461692 | 1 | 0,821452 |
| -0,31533 | 405 | 349,9665 | 0,752702 | 1 | 0,898125 |
| -0,12882 | 405 | 198,5898 | 0,897627 | 1 | 0,960059 |
| 0,522575 | 404 | 381,8159 | 0,601573 | 1 | 0,964878 |
| -1,31522 | 355 | 335,5575 | 0,189334 | 1 | 0,53937 |
| -1,09358 | 270 | 251,2068 | 0,275188 | 1 | 0,701156 |
| 0,105238 | 421 | 378,0441 | 0,916243 | 1 | 0,966828 |
| -0,81101 | 421 | 202,0244 | 0,418313 | 1 | 0,688959 |
| -0,49563 | 420 | 405,1165 | 0,620424 | 1 | 0,964878 |
| 0,506918 | 367 | 352,4692 | 0,612529 | 1 | 0,871158 |
| -0,36104 | 280 | 266,1066 | 0,718352 | 1 | 0,931633 |
| -2,09567 | 421 | 332,5915 | 0,036867 | 1 | 0,217944 |
| -1,00441 | 421 | 184,2914 | 0,3165 | 1 | 0,614646 |
| -1,12922 | 420 | 370,6449 | 0,259535 | 1 | 0,943897 |
| -0,75407 | 367 | 336,5668 | 0,451336 | 1 | 0,756578 |
| -2,10247 | 280 | 258,1964 | 0,036481 | 1 | 0,34044 |
| 0,219369 | 421 | 343,0603 | 0,826493 | 1 | 0,936485 |
| 0,61945 | 421 | 187,1775 | 0,536373 | 1 | 0,770962 |
| 0,061319 | 420 | 377,6697 | 0,951138 | 1 | 0,992811 |
| 1,011641 | 367 | 320,9083 | 0,312472 | 1 | 0,646834 |
| 1,256006 | 280 | 260,1753 | 0,210241 | 1 | 0,641361 |
| 2,18004 | 421 | 399,4892 | 0,029838 | 1 | 0,200565 |
| 3,222267 | 421 | 216,3532 | 0,001468 | 0,145341 | 0,093671 |
| 0,297922 | 420 | 415,3334 | 0,765912 | 1 | 0,969427 |
| 2,249468 | 367 | 360,3696 | 0,025086 | 1 | 0,247087 |
| 1,265257 | 280 | 273,9733 | 0,206854 | 1 | 0,63879 |
| 0,50522 | 421 | 365,9535 | 0,613708 | 1 | 0,825172 |
| -0,17879 | 421 | 196,814 | 0,858289 | 1 | 0,946189 |
| -1,22958 | 420 | 395,6222 | 0,219586 | 1 | 0,943055 |
| -1,27026 | 367 | 346,1827 | 0,204847 | 1 | 0,558434 |
| 0,085236 | 280 | 263,371 | 0,932138 | 1 | 0,979504 |
| -1,26388 | 421 | 339,1022 | 0,207142 | 1 | 0,496728 |
| -1,12648 | 421 | 185,7002 | 0,261416 | 1 | 0,557979 |
| -1,39515 | 420 | 373,0307 | 0,1638 | 1 | 0,914005 |
| -0,09345 | 367 | 320,5106 | 0,925603 | 1 | 0,976733 |
| 0,431779 | 280 | 260,773 | 0,666259 | 1 | 0,908644 |
| 0,520947 | 421 | 335,7481 | 0,602747 | 1 | 0,820542 |
| 0,624449 | 421 | 184,572 | 0,533104 | 1 | 0,769894 |
| -0,19816 | 420 | 370,2978 | 0,843026 | 1 | 0,973894 |
| 0,997844 | 367 | 324,3788 | 0,319099 | 1 | 0,653184 |
| 0,508948 | 280 | 256,6814 | 0,611226 | 1 | 0,888252 |
| -0,77874 | 421 | 388,6902 | 0,436607 | 1 | 0,707193 |
| -1,43008 | 421 | 206,8234 | 0,154202 | 1 | 0,435989 |
| -1,53467 | 420 | 412,0431 | 0,125631 | 1 | 0,880587 |
| -1,41461 | 367 | 358,3053 | 0,158051 | 1 | 0,504558 |

| | | | | | |
|---|---|---|---|---|---|
| -1,62283 | 280 | 273,192 | 0,105779 | 1 | 0,492865 |
| 1,293491 | 421 | 359,2499 | 0,196673 | 1 | 0,482712 |
| 1,135606 | 421 | 195,4154 | 0,257512 | 1 | 0,556025 |
| 0,037741 | 420 | 398,3943 | 0,969913 | 1 | 0,997342 |
| 1,210457 | 367 | 345,4793 | 0,226931 | 1 | 0,576775 |
| 1,134934 | 280 | 270,7141 | 0,257407 | 1 | 0,680548 |
| 1,823692 | 421 | 324,469 | 0,069119 | 1 | 0,282663 |
| 2,778855 | 421 | 182,6351 | 0,006026 | 0,584682 | 0,141161 |
| 1,186346 | 420 | 358,8557 | 0,23627 | 1 | 0,943055 |
| 0,904294 | 367 | 316,5429 | 0,366527 | 1 | 0,678571 |
| -0,20468 | 280 | 247,2618 | 0,837987 | 1 | 0,964667 |
| 2,023809 | 421 | 362,364 | 0,043723 | 1 | 0,235324 |
| 1,431568 | 421 | 192,2246 | 0,153891 | 1 | 0,435929 |
| 2,096976 | 420 | 398,7585 | 0,036625 | 1 | 0,828713 |
| 1,818242 | 367 | 343,4417 | 0,069898 | 1 | 0,364488 |
| -0,26949 | 280 | 265,4449 | 0,787761 | 1 | 0,953235 |
| 2,174047 | 421 | 368,069 | 0,030338 | 1 | 0,200565 |
| 0,986 | 421 | 194,3918 | 0,325359 | 1 | 0,619085 |
| -0,11214 | 420 | 399,73 | 0,910765 | 1 | 0,987181 |
| 2,18841 | 367 | 345,4108 | 0,029309 | 1 | 0,260082 |
| 1,488737 | 280 | 273,0983 | 0,137711 | 1 | 0,54931 |
| 0,112108 | 421 | 369,4192 | 0,910799 | 1 | 0,966828 |
| 1,224323 | 421 | 197,099 | 0,222292 | 1 | 0,518566 |
| -0,66682 | 420 | 399,9273 | 0,505269 | 1 | 0,960096 |
| 1,635519 | 367 | 349,746 | 0,10284 | 1 | 0,426647 |
| -0,41529 | 280 | 263,5783 | 0,678267 | 1 | 0,909385 |
| 0,34572 | 421 | 351,7017 | 0,72976 | 1 | 0,891206 |
| 0,678194 | 421 | 189,2662 | 0,498477 | 1 | 0,749329 |
| -1,40236 | 420 | 383,0811 | 0,161617 | 1 | 0,914005 |
| 2,126863 | 367 | 339,7339 | 0,034152 | 1 | 0,279197 |
| 1,626715 | 280 | 267,4116 | 0,104975 | 1 | 0,492014 |
| -1,63291 | 421 | 371,9155 | 0,103334 | 1 | 0,345287 |
| 0,505877 | 421 | 197,1577 | 0,613508 | 1 | 0,824026 |
| -0,32299 | 420 | 400,0619 | 0,74687 | 1 | 0,969427 |
| -0,02707 | 367 | 352,4483 | 0,978421 | 1 | 0,99209 |
| 0,531264 | 280 | 269,4822 | 0,595674 | 1 | 0,880079 |
| 2,040286 | 421 | 305,7404 | 0,042181 | 1 | 0,229482 |
| 2,297106 | 421 | 171,053 | 0,022824 | 1 | 0,212324 |
| 3,044897 | 420 | 345,1777 | 0,002506 | 0,333362 | 0,314772 |
| 1,481493 | 367 | 297,7674 | 0,139533 | 1 | 0,487879 |
| 2,26903 | 280 | 255,2685 | 0,024102 | 1 | 0,313396 |
| 1,29524 | 421 | 371,2048 | 0,196042 | 1 | 0,481951 |
| 1,915094 | 421 | 195,3424 | 0,056941 | 1 | 0,312039 |
| -0,6769 | 420 | 401,7833 | 0,498858 | 1 | 0,960096 |
| -0,26311 | 367 | 351,8518 | 0,792617 | 1 | 0,944997 |
| 0,074212 | 280 | 268,9521 | 0,940897 | 1 | 0,982175 |
| -2,86587 | 421 | 328,1325 | 0,004427 | 0,562235 | 0,087964 |
| -0,9942 | 421 | 179,3376 | 0,321464 | 1 | 0,617772 |

| | | | | | |
|---|---|---|---|---|---|
| -2,22225 | 420 | 357,0618 | 0,026893 | 1 | 0,767829 |
| -1,41067 | 367 | 318,5754 | 0,159319 | 1 | 0,504558 |
| -2,70978 | 280 | 256,9615 | 0,007186 | 0,919788 | 0,195931 |
| -0,19793 | 421 | 322,8182 | 0,843224 | 1 | 0,939735 |
| 0,958829 | 421 | 180,0878 | 0,33893 | 1 | 0,632922 |
| -0,41404 | 420 | 357,6658 | 0,679092 | 1 | 0,96781 |
| 0,361626 | 367 | 313,6702 | 0,717875 | 1 | 0,913715 |
| 1,520321 | 280 | 254,9178 | 0,12967 | 1 | 0,536847 |
| 0,77257 | 421 | 347,9701 | 0,440301 | 1 | 0,710422 |
| 0,567056 | 421 | 187,719 | 0,571354 | 1 | 0,792729 |
| 0,3025 | 420 | 381,7471 | 0,762436 | 1 | 0,969427 |
| 0,831038 | 367 | 337,99 | 0,406539 | 1 | 0,719923 |
| 1,699857 | 280 | 263,8898 | 0,090336 | 1 | 0,468147 |
| -0,46402 | 421 | 317,5423 | 0,642949 | 1 | 0,836 |
| 0,208644 | 421 | 178,5696 | 0,834964 | 1 | 0,937624 |
| -0,82744 | 420 | 350,8033 | 0,40855 | 1 | 0,95743 |
| -0,03876 | 367 | 304,1579 | 0,96911 | 1 | 0,988125 |
| 1,437854 | 280 | 247,0248 | 0,151741 | 1 | 0,565026 |
| -2,53589 | 421 | 348,8497 | 0,011652 | 1 | 0,130072 |
| -0,49933 | 421 | 190,2061 | 0,618121 | 1 | 0,825805 |
| -1,64392 | 420 | 378,2835 | 0,101023 | 1 | 0,848246 |
| -0,23136 | 367 | 346,0079 | 0,817173 | 1 | 0,95095 |
| 1,39943 | 280 | 263,5318 | 0,16286 | 1 | 0,584818 |
| -2,34227 | 421 | 329,2059 | 0,019762 | 1 | 0,162476 |
| -0,32307 | 421 | 182,3012 | 0,747011 | 1 | 0,889215 |
| -0,5942 | 420 | 366,0168 | 0,552742 | 1 | 0,960096 |
| 0,243801 | 367 | 321,2 | 0,80754 | 1 | 0,950149 |
| 0,611048 | 280 | 252,2009 | 0,541718 | 1 | 0,849499 |
| 0,02837 | 421 | 361,9689 | 0,977383 | 1 | 0,995982 |
| 0,677763 | 421 | 195,3553 | 0,498724 | 1 | 0,749329 |
| 0,240685 | 420 | 393,4231 | 0,809925 | 1 | 0,969427 |
| 1,902688 | 367 | 336,9528 | 0,057934 | 1 | 0,339097 |
| -0,20986 | 280 | 262,8698 | 0,83394 | 1 | 0,964667 |
| 1,555678 | 421 | 355,0834 | 0,120675 | 1 | 0,382054 |
| 0,402085 | 421 | 194,3528 | 0,688063 | 1 | 0,863373 |
| -0,93801 | 420 | 391,3075 | 0,348818 | 1 | 0,947373 |
| -0,77921 | 367 | 343,5162 | 0,436394 | 1 | 0,742744 |
| 0,259119 | 280 | 264,65 | 0,795745 | 1 | 0,954269 |
| -0,14568 | 421 | 318,7034 | 0,884267 | 1 | 0,961484 |
| -1,04272 | 421 | 181,2629 | 0,298466 | 1 | 0,597214 |
| 0,975057 | 420 | 354,0481 | 0,330198 | 1 | 0,947373 |
| -1,43598 | 367 | 317,5324 | 0,151994 | 1 | 0,500093 |
| -0,05208 | 280 | 252,6431 | 0,958508 | 1 | 0,986271 |
| 1,890656 | 421 | 366,2263 | 0,059459 | 1 | 0,268966 |
| 2,726289 | 421 | 193,9287 | 0,006992 | 0,887979 | 0,141161 |
| 1,112543 | 420 | 398,1068 | 0,266576 | 1 | 0,943897 |
| 2,681418 | 367 | 349,7048 | 0,007679 | 0,975186 | 0,180403 |
| 1,083103 | 280 | 274,9071 | 0,279712 | 1 | 0,703716 |

| | | | | | |
|---|---|---|---|---|---|
| 0,238024 | 421 | 353,9064 | 0,812 | 1 | 0,924931 |
| 1,125186 | 421 | 189,8061 | 0,261931 | 1 | 0,557979 |
| -0,48754 | 420 | 387,8689 | 0,626148 | 1 | 0,964878 |
| -0,35129 | 367 | 336,6047 | 0,725591 | 1 | 0,919068 |
| 0,113835 | 280 | 262,685 | 0,909456 | 1 | 0,976008 |
| -0,68396 | 421 | 320,6954 | 0,494496 | 1 | 0,74916 |
| 0,611718 | 421 | 180,2178 | 0,541495 | 1 | 0,773814 |
| 0,316911 | 420 | 354,6829 | 0,751498 | 1 | 0,969427 |
| 0,216698 | 367 | 312,0687 | 0,828585 | 1 | 0,956109 |
| 1,51589 | 280 | 252,323 | 0,130799 | 1 | 0,538563 |
| 0,481095 | 421 | 367,6657 | 0,630735 | 1 | 0,834519 |
| 1,021089 | 421 | 190,8293 | 0,308505 | 1 | 0,60631 |
| 0,169237 | 420 | 398,4033 | 0,865696 | 1 | 0,979433 |
| 2,538125 | 367 | 354,4637 | 0,011572 | 1 | 0,189546 |
| 2,505076 | 280 | 269,603 | 0,012833 | 1 | 0,2448 |
| 0,154051 | 421 | 364,5277 | 0,877655 | 1 | 0,95861 |
| 1,117277 | 421 | 194,7009 | 0,265253 | 1 | 0,562747 |
| -1,42696 | 420 | 395,3386 | 0,15438 | 1 | 0,914005 |
| -0,06239 | 367 | 346,0356 | 0,950289 | 1 | 0,979539 |
| -0,01861 | 280 | 266,1745 | 0,985167 | 1 | 0,992411 |
| 0,246459 | 421 | 304,0021 | 0,805493 | 1 | 0,922616 |
| -2,06574 | 421 | 177,1111 | 0,040309 | 1 | 0,26998 |
| -1,59098 | 420 | 332,4871 | 0,112564 | 1 | 0,848246 |
| 0,443184 | 367 | 295,5441 | 0,657957 | 1 | 0,892476 |
| -0,0618 | 280 | 247,7695 | 0,950774 | 1 | 0,982409 |
| -0,43251 | 421 | 327,3567 | 0,665656 | 1 | 0,853014 |
| -0,84954 | 421 | 184,0767 | 0,396686 | 1 | 0,667939 |
| -0,5136 | 420 | 365,3298 | 0,607843 | 1 | 0,964878 |
| -0,95182 | 367 | 323,1393 | 0,3419 | 1 | 0,664831 |
| 0,333058 | 280 | 249,885 | 0,73937 | 1 | 0,940183 |
| 2,677691 | 421 | 315,6563 | 0,007801 | 0,741091 | 0,108852 |
| 0,278583 | 421 | 178,4785 | 0,780888 | 1 | 0,91393 |
| -0,84939 | 420 | 349,8294 | 0,396245 | 1 | 0,95743 |
| 2,05148 | 367 | 300,0478 | 0,041088 | 1 | 0,289956 |
| 1,117873 | 280 | 255,6261 | 0,264671 | 1 | 0,688875 |
| -1,36727 | 421 | 366,794 | 0,172379 | 1 | 0,444059 |
| -0,43612 | 421 | 195,7272 | 0,663234 | 1 | 0,845595 |
| -1,96851 | 420 | 396,743 | 0,049705 | 1 | 0,846627 |
| -0,19845 | 367 | 344,7757 | 0,842811 | 1 | 0,956935 |
| 1,373416 | 280 | 263,1554 | 0,170792 | 1 | 0,59737 |
| 0,681756 | 421 | 361,3531 | 0,49583 | 1 | 0,750117 |
| 0,755298 | 421 | 192,2886 | 0,450994 | 1 | 0,713861 |
| 0,306096 | 420 | 392,2657 | 0,759694 | 1 | 0,969427 |
| 0,648816 | 367 | 333,2407 | 0,516904 | 1 | 0,80059 |
| 0,75648 | 280 | 257,0096 | 0,450054 | 1 | 0,813228 |
| 0,156795 | 421 | 368,2376 | 0,875492 | 1 | 0,95861 |
| 1,938991 | 421 | 193,8709 | 0,053954 | 1 | 0,303391 |
| 0,349756 | 420 | 399,6188 | 0,726706 | 1 | 0,969427 |

| | | | | | |
|---|---|---|---|---|---|
| 1,5967 | 367 | 354,5759 | 0,111224 | 1 | 0,437634 |
| 2,704292 | 280 | 269,432 | 0,007281 | 0,698956 | 0,195931 |
| -0,75069 | 421 | 307,8076 | 0,453413 | 1 | 0,717744 |
| -0,26704 | 421 | 176,9498 | 0,78975 | 1 | 0,918329 |
| -1,36137 | 420 | 337,6271 | 0,174304 | 1 | 0,9199 |
| -1,27814 | 367 | 306,7889 | 0,202167 | 1 | 0,556171 |
| 0,242903 | 280 | 253,1775 | 0,808278 | 1 | 0,95747 |
| -0,91076 | 421 | 313,5594 | 0,363119 | 1 | 0,653008 |
| -0,6968 | 421 | 178,0402 | 0,486836 | 1 | 0,741822 |
| -2,68019 | 420 | 334,7233 | 0,007722 | 0,764482 | 0,554149 |
| -0,47676 | 367 | 309,7686 | 0,633869 | 1 | 0,880707 |
| 1,68325 | 280 | 244,6705 | 0,093602 | 1 | 0,47335 |
| -0,71907 | 421 | 378,7233 | 0,472544 | 1 | 0,731548 |
| -2,74148 | 421 | 202,5446 | 0,006664 | 0,639698 | 0,141161 |
| -0,41615 | 420 | 406,3692 | 0,677523 | 1 | 0,96781 |
| -2,88708 | 367 | 353,5432 | 0,004127 | 0,404487 | 0,178183 |
| -2,56863 | 280 | 271,0657 | 0,010746 | 1 | 0,231345 |
| 1,383947 | 421 | 324,8093 | 0,167325 | 1 | 0,441901 |
| 2,553414 | 421 | 182,462 | 0,011485 | 1 | 0,163285 |
| 0,767577 | 420 | 363,6186 | 0,443237 | 1 | 0,960096 |
| 2,544432 | 367 | 309,008 | 0,011432 | 1 | 0,189546 |
| 1,019199 | 280 | 257,7083 | 0,309064 | 1 | 0,730032 |
| -0,87694 | 421 | 350,3942 | 0,38112 | 1 | 0,671614 |
| -0,75966 | 421 | 187,8477 | 0,448409 | 1 | 0,711318 |
| -0,7328 | 420 | 382,3271 | 0,46413 | 1 | 0,960096 |
| 1,104689 | 367 | 331,1227 | 0,270096 | 1 | 0,609149 |
| 2,03762 | 280 | 257,62 | 0,04261 | 1 | 0,377723 |
| -0,55492 | 421 | 321,4842 | 0,579335 | 1 | 0,803921 |
| 0,04868 | 421 | 181,6032 | 0,961228 | 1 | 0,985228 |
| -0,43067 | 420 | 351,8058 | 0,666975 | 1 | 0,96781 |
| -0,238 | 367 | 312,5972 | 0,812039 | 1 | 0,950149 |
| 2,531838 | 280 | 261,5847 | 0,011933 | 1 | 0,239766 |
| 1,796802 | 421 | 340,9179 | 0,073252 | 1 | 0,292681 |
| 2,16823 | 421 | 188,3647 | 0,031395 | 1 | 0,241423 |
| -0,11555 | 420 | 376,7942 | 0,908068 | 1 | 0,986632 |
| 1,754209 | 367 | 317,0658 | 0,080361 | 1 | 0,381841 |
| 0,93858 | 280 | 259,6001 | 0,348819 | 1 | 0,761841 |
| -1,74966 | 421 | 326,7572 | 0,081115 | 1 | 0,30376 |
| -0,5031 | 421 | 184,7705 | 0,615492 | 1 | 0,825219 |
| 0,469305 | 420 | 364,3207 | 0,639132 | 1 | 0,964878 |
| -1,15713 | 367 | 313,8578 | 0,2481 | 1 | 0,602072 |
| -0,95744 | 280 | 247,8113 | 0,33928 | 1 | 0,756354 |
| 1,413861 | 421 | 323,6046 | 0,158364 | 1 | 0,431264 |
| 0,402797 | 421 | 180,7602 | 0,687574 | 1 | 0,863373 |
| -1,12978 | 420 | 352,3242 | 0,259336 | 1 | 0,943897 |
| 1,222055 | 367 | 316,5402 | 0,222596 | 1 | 0,574455 |
| 3,479524 | 280 | 259,6851 | 5,89E-04 | 0,077738 | 0,093926 |
| 0,102612 | 421 | 335,8349 | 0,918332 | 1 | 0,966828 |

| | | | | | |
|---|---|---|---|---|---|
| 1,160249 | 421 | 181,8157 | 0,247469 | 1 | 0,548314 |
| 1,03394 | 420 | 372,4143 | 0,301835 | 1 | 0,943897 |
| 0,104571 | 367 | 330,7437 | 0,91678 | 1 | 0,973634 |
| 2,274834 | 280 | 262,4567 | 0,023723 | 1 | 0,313396 |
| 0,182518 | 421 | 300,1993 | 0,8553 | 1 | 0,947051 |
| 1,074721 | 421 | 171,8375 | 0,284006 | 1 | 0,582654 |
| 1,445104 | 420 | 335,5285 | 0,149362 | 1 | 0,914005 |
| 0,169278 | 367 | 297,304 | 0,865693 | 1 | 0,959636 |
| 2,768545 | 280 | 260,0171 | 0,006036 | 0,585479 | 0,186692 |
| -0,20358 | 421 | 363,6472 | 0,838796 | 1 | 0,93721 |
| 0,327029 | 421 | 191,5071 | 0,744003 | 1 | 0,888441 |
| 0,125969 | 420 | 395,6891 | 0,89982 | 1 | 0,98503 |
| 0,677286 | 367 | 345,219 | 0,498678 | 1 | 0,793601 |
| 1,753243 | 280 | 263,2 | 0,080724 | 1 | 0,455624 |
| 0,211007 | 421 | 289,6284 | 0,83303 | 1 | 0,936611 |
| -0,88783 | 421 | 175,0932 | 0,375851 | 1 | 0,654686 |
| -1,01517 | 420 | 315,1101 | 0,310803 | 1 | 0,943897 |
| 1,089042 | 367 | 274,553 | 0,27709 | 1 | 0,611884 |
| 0,161264 | 280 | 254,6609 | 0,872014 | 1 | 0,969656 |
| 0,177738 | 421 | 347,3956 | 0,859032 | 1 | 0,949789 |
| 1,914007 | 421 | 185,84 | 0,057156 | 1 | 0,312079 |
| -0,0065 | 420 | 382,7002 | 0,994821 | 1 | 0,997342 |
| 1,290757 | 367 | 344,0779 | 0,197654 | 1 | 0,549567 |
| 1,84142 | 280 | 260,5525 | 0,066697 | 1 | 0,425749 |
| -0,3814 | 421 | 330,1591 | 0,703152 | 1 | 0,874567 |
| 0,690847 | 421 | 182,4941 | 0,49054 | 1 | 0,744768 |
| -0,77927 | 420 | 364,2023 | 0,436327 | 1 | 0,960096 |
| 0,05129 | 367 | 322,5202 | 0,959126 | 1 | 0,981063 |
| 1,837023 | 280 | 251,6783 | 0,067386 | 1 | 0,425749 |
| -2,04279 | 421 | 331,8556 | 0,041863 | 1 | 0,228575 |
| -0,38767 | 421 | 184,6645 | 0,698704 | 1 | 0,865528 |
| -1,0508 | 420 | 366,3947 | 0,294042 | 1 | 0,943897 |
| -2,23248 | 367 | 311,1579 | 0,026295 | 1 | 0,252225 |
| -0,49152 | 280 | 254,6783 | 0,623484 | 1 | 0,89827 |
| 2,062161 | 421 | 355,776 | 0,039918 | 1 | 0,224527 |
| 1,814842 | 421 | 193,5021 | 0,071097 | 1 | 0,339864 |
| 0,554638 | 420 | 392,7085 | 0,579458 | 1 | 0,964878 |
| 1,807911 | 367 | 338,3942 | 0,071508 | 1 | 0,367791 |
| -0,48805 | 280 | 263,6945 | 0,625918 | 1 | 0,899494 |
| 0,326905 | 421 | 333,0427 | 0,743945 | 1 | 0,895468 |
| 1,463385 | 421 | 183,958 | 0,145069 | 1 | 0,426157 |
| 0,635791 | 420 | 370,0009 | 0,525306 | 1 | 0,960096 |
| 0,677798 | 367 | 319,014 | 0,498391 | 1 | 0,793601 |
| 1,933197 | 280 | 261,2428 | 0,054292 | 1 | 0,40654 |
| 0,303503 | 421 | 342,0107 | 0,761691 | 1 | 0,902412 |
| -0,18778 | 421 | 185,8799 | 0,851252 | 1 | 0,944105 |
| -0,45902 | 420 | 376,2929 | 0,646488 | 1 | 0,964878 |
| 2,78676 | 367 | 308,8542 | 0,005653 | 0,740582 | 0,178183 |

| | | | | | |
|---|---|---|---|---|---|
| 0,338204 | 280 | 246,5634 | 0,735498 | 1 | 0,938189 |
| -1,48201 | 421 | 353,856 | 0,139228 | 1 | 0,4106 |
| 0,352512 | 421 | 193,145 | 0,724839 | 1 | 0,878959 |
| 0,741833 | 420 | 386,5738 | 0,458639 | 1 | 0,960096 |
| -0,36649 | 367 | 350,4557 | 0,714219 | 1 | 0,912652 |
| 0,296825 | 280 | 269,1122 | 0,766829 | 1 | 0,945436 |
| -2,92438 | 421 | 340,5357 | 0,003683 | 0,3609 | 0,087964 |
| -1,81759 | 421 | 190,8139 | 0,070695 | 1 | 0,339864 |
| -1,79882 | 420 | 379,9496 | 0,07284 | 1 | 0,848246 |
| -1,67132 | 367 | 336,7584 | 0,095587 | 1 | 0,412696 |
| -0,54628 | 280 | 263,9551 | 0,585336 | 1 | 0,87561 |
| -2,61141 | 421 | 286,9843 | 0,009491 | 0,88267 | 0,114424 |
| -2,23936 | 421 | 173,5228 | 0,026403 | 1 | 0,222284 |
| -0,88798 | 420 | 320,9434 | 0,375216 | 1 | 0,95743 |
| -2,64297 | 367 | 363 | 0,008574 | 0,823129 | 0,183886 |
| -3,01429 | 280 | 258,1043 | 0,002832 | 0,280405 | 0,135787 |
| -0,64993 | 421 | 357,9475 | 0,516153 | 1 | 0,767364 |
| 0,486269 | 421 | 194,1613 | 0,627325 | 1 | 0,832379 |
| -0,75682 | 420 | 389,5051 | 0,449614 | 1 | 0,960096 |
| -2,20705 | 367 | 331,5724 | 0,027995 | 1 | 0,257802 |
| 2,000972 | 280 | 252,8934 | 0,046464 | 1 | 0,387749 |
| -0,82586 | 421 | 383,0917 | 0,409398 | 1 | 0,695339 |
| -1,0649 | 421 | 205,5979 | 0,28817 | 1 | 0,585544 |
| -0,04269 | 420 | 409,8801 | 0,965972 | 1 | 0,996924 |
| -1,89791 | 367 | 361,483 | 0,058504 | 1 | 0,339097 |
| -1,69315 | 280 | 271,9759 | 0,091572 | 1 | 0,468147 |
| -0,24683 | 421 | 289,3452 | 0,805215 | 1 | 0,922616 |
| 0,239893 | 421 | 169,4393 | 0,810703 | 1 | 0,926063 |
| 1,524263 | 420 | 321,0578 | 0,128427 | 1 | 0,887798 |
| 0,146229 | 367 | 298,142 | 0,883839 | 1 | 0,962871 |
| 0,020349 | 280 | 276 | 0,98378 | 1 | 0,99236 |
| 0,315144 | 421 | 336,3509 | 0,752847 | 1 | 0,898125 |
| 0,828026 | 421 | 182,9941 | 0,408734 | 1 | 0,681984 |
| -0,6286 | 420 | 369,4799 | 0,53 | 1 | 0,960096 |
| 0,411411 | 367 | 325,608 | 0,681042 | 1 | 0,900574 |
| 1,456168 | 280 | 252,9048 | 0,146587 | 1 | 0,555466 |
| -0,53449 | 421 | 299,1761 | 0,5934 | 1 | 0,812958 |
| 1,290825 | 421 | 176,8942 | 0,198449 | 1 | 0,488664 |
| 0,42461 | 420 | 329,8427 | 0,671398 | 1 | 0,96781 |
| 1,502617 | 367 | 301,058 | 0,133986 | 1 | 0,477542 |
| -0,17874 | 280 | 254,7183 | 0,858281 | 1 | 0,966655 |
| 0,757373 | 421 | 341,9906 | 0,449348 | 1 | 0,714643 |
| -0,09703 | 421 | 186,386 | 0,922808 | 1 | 0,971139 |
| -1,84198 | 420 | 378,32 | 0,066261 | 1 | 0,848246 |
| 1,395137 | 367 | 338,1779 | 0,16389 | 1 | 0,510294 |
| 0,072315 | 280 | 261,8677 | 0,942407 | 1 | 0,982175 |
| -0,00593 | 421 | 408,5344 | 0,995275 | 1 | 0,998054 |
| -1,17188 | 421 | 222,7089 | 0,242497 | 1 | 0,542422 |

| | | | | | |
|---|---|---|---|---|---|
| -0,1558 | 420 | 415,8361 | 0,876263 | 1 | 0,982463 |
| 1,048421 | 367 | 361,4775 | 0,295145 | 1 | 0,62734 |
| 0,554411 | 280 | 274,663 | 0,579749 | 1 | 0,874212 |
| 0,733434 | 421 | 388,989 | 0,463736 | 1 | 0,727964 |
| 1,636918 | 421 | 207,8753 | 0,103161 | 1 | 0,383322 |
| 1,470739 | 420 | 412,2279 | 0,142125 | 1 | 0,90372 |
| -0,66841 | 367 | 352,6374 | 0,504312 | 1 | 0,793601 |
| 0,507514 | 280 | 264,0923 | 0,612218 | 1 | 0,888837 |
| 1,740403 | 421 | 383,4825 | 0,08259 | 1 | 0,305058 |
| 2,503938 | 421 | 205,6 | 0,013059 | 1 | 0,171643 |
| 0,54747 | 420 | 409,7699 | 0,584354 | 1 | 0,964878 |
| 3,09924 | 367 | 356,6416 | 0,002094 | 0,207339 | 0,176492 |
| 1,692793 | 280 | 271,6763 | 0,091641 | 1 | 0,468147 |
| -1,77644 | 421 | 345,9665 | 0,07654 | 1 | 0,300378 |
| -0,80249 | 421 | 188,8458 | 0,423279 | 1 | 0,693349 |
| 0,10971 | 420 | 381,398 | 0,912697 | 1 | 0,987181 |
| 0,234481 | 367 | 335,0449 | 0,814754 | 1 | 0,95095 |
| -0,11096 | 280 | 257,7267 | 0,911736 | 1 | 0,976008 |
| -1,91066 | 421 | 327,417 | 0,056922 | 1 | 0,266382 |
| -0,70276 | 421 | 181,7787 | 0,483103 | 1 | 0,739236 |
| -1,26295 | 420 | 365,4105 | 0,207414 | 1 | 0,930277 |
| -1,67323 | 367 | 310,9175 | 0,095287 | 1 | 0,412696 |
| -1,14134 | 280 | 269,7576 | 0,254739 | 1 | 0,679673 |
| 1,507622 | 421 | 402,5399 | 0,132436 | 1 | 0,404829 |
| 1,900044 | 421 | 218,6299 | 0,058743 | 1 | 0,314262 |
| 0,266809 | 420 | 415,6339 | 0,789749 | 1 | 0,969427 |
| 0,615437 | 367 | 362,9876 | 0,538652 | 1 | 0,819758 |
| 2,148593 | 280 | 275,9522 | 0,032537 | 1 | 0,329568 |
| -2,84416 | 421 | 346,772 | 0,004717 | 0,594351 | 0,087964 |
| -3,19874 | 421 | 189,1967 | 0,001618 | 0,212013 | 0,093671 |
| -0,26119 | 420 | 387,2982 | 0,794081 | 1 | 0,969427 |
| -1,05438 | 367 | 335,7133 | 0,292468 | 1 | 0,627013 |
| -1,12437 | 280 | 261,8719 | 0,261888 | 1 | 0,687569 |
| -2,3674 | 421 | 347,3745 | 0,018461 | 1 | 0,158072 |
| -1,23575 | 421 | 190,6404 | 0,21807 | 1 | 0,514565 |
| 0,275231 | 420 | 384,2739 | 0,783287 | 1 | 0,969427 |
| -1,85939 | 367 | 340,3772 | 0,063834 | 1 | 0,350852 |
| -1,2144 | 280 | 265,6953 | 0,225672 | 1 | 0,654014 |
| -0,23824 | 421 | 313,2693 | 0,811851 | 1 | 0,924931 |
| 0,631625 | 421 | 181,043 | 0,528429 | 1 | 0,768085 |
| 0,544819 | 420 | 347,1096 | 0,586228 | 1 | 0,964878 |
| 0,926256 | 367 | 307,426 | 0,35504 | 1 | 0,674813 |
| -0,853 | 280 | 252,5056 | 0,394467 | 1 | 0,782194 |
| 1,415104 | 421 | 379,9243 | 0,157857 | 1 | 0,430962 |
| 0,019373 | 421 | 201,1462 | 0,984563 | 1 | 0,995074 |
| 0,379744 | 420 | 406,2992 | 0,704334 | 1 | 0,968866 |
| -0,84995 | 367 | 352,7214 | 0,395927 | 1 | 0,707784 |
| -0,28649 | 280 | 266,9731 | 0,774722 | 1 | 0,947651 |

| | | | | | |
|---|---|---|---|---|---|
| 1,761045 | 421 | 344,3668 | 0,079118 | 1 | 0,302838 |
| 1,90802 | 421 | 186,4937 | 0,057925 | 1 | 0,312877 |
| 1,282994 | 420 | 383,0896 | 0,20027 | 1 | 0,928589 |
| 1,053424 | 367 | 331,4512 | 0,292914 | 1 | 0,627013 |
| 2,998861 | 280 | 267,8608 | 0,002965 | 0,290544 | 0,135787 |
| 0,479246 | 421 | 316,4877 | 0,632095 | 1 | 0,83455 |
| 1,057293 | 421 | 181,7838 | 0,291781 | 1 | 0,588663 |
| 1,62149 | 420 | 351,7227 | 0,105809 | 1 | 0,848246 |
| 0,469665 | 367 | 308,4848 | 0,638926 | 1 | 0,880707 |
| 1,187079 | 280 | 246,0053 | 0,236342 | 1 | 0,659852 |
| -1,36471 | 421 | 330,3627 | 0,173273 | 1 | 0,445601 |
| -2,23987 | 421 | 183,6588 | 0,026299 | 1 | 0,222284 |
| -1,38305 | 420 | 370,6997 | 0,167481 | 1 | 0,914005 |
| -1,02686 | 367 | 335,3703 | 0,305225 | 1 | 0,637666 |
| -0,4522 | 280 | 269,2347 | 0,651488 | 1 | 0,908644 |
| 0,537377 | 421 | 377,0488 | 0,591324 | 1 | 0,81147 |
| -0,7933 | 421 | 201,4207 | 0,428539 | 1 | 0,694836 |
| 0,164169 | 420 | 403,7117 | 0,869681 | 1 | 0,980994 |
| 0,179495 | 367 | 355,9029 | 0,857651 | 1 | 0,958813 |
| -0,31878 | 280 | 261,2172 | 0,750149 | 1 | 0,942071 |
| -1,06914 | 421 | 373,9378 | 0,285695 | 1 | 0,583534 |
| -0,45669 | 421 | 200,4989 | 0,648389 | 1 | 0,839358 |
| -1,24739 | 420 | 402,3766 | 0,212978 | 1 | 0,943055 |
| -0,98111 | 367 | 349,2488 | 0,327217 | 1 | 0,655788 |
| 0,89123 | 280 | 272,2392 | 0,373593 | 1 | 0,763914 |
| 0,132662 | 421 | 382,9723 | 0,894531 | 1 | 0,962 |
| 1,372465 | 421 | 203,4823 | 0,17143 | 1 | 0,456365 |
| -0,17314 | 420 | 407,9182 | 0,862631 | 1 | 0,977432 |
| -0,69888 | 367 | 355,4422 | 0,485083 | 1 | 0,786044 |
| -1,01419 | 280 | 266,6888 | 0,311413 | 1 | 0,733729 |
| -0,09935 | 421 | 397,9423 | 0,920909 | 1 | 0,966828 |
| -0,5517 | 421 | 214,5748 | 0,581727 | 1 | 0,798417 |
| -0,85584 | 420 | 414,6128 | 0,392583 | 1 | 0,95743 |
| -1,20728 | 367 | 358,5152 | 0,22812 | 1 | 0,577307 |
| 0,162446 | 280 | 270,0312 | 0,871076 | 1 | 0,969656 |
| -0,66885 | 421 | 297,9486 | 0,504109 | 1 | 0,75707 |
| -0,57207 | 421 | 178,1536 | 0,567994 | 1 | 0,78927 |
| -0,26157 | 420 | 328,848 | 0,793813 | 1 | 0,969427 |
| 1,256805 | 367 | 293,3996 | 0,209824 | 1 | 0,563436 |
| 0,273906 | 280 | 276 | 0,784362 | 1 | 0,953235 |
| 0,77003 | 421 | 416,6381 | 0,441718 | 1 | 0,711185 |
| 2,42621 | 421 | 244,2952 | 0,015982 | 1 | 0,179127 |
| 1,068849 | 420 | 409,4399 | 0,285767 | 1 | 0,943897 |
| -0,00879 | 367 | 359,7972 | 0,992988 | 1 | 0,997335 |
| 2,210226 | 280 | 275,5855 | 0,027911 | 1 | 0,326641 |
| -2,6185 | 421 | 307,4641 | 0,009269 | 0,871303 | 0,113566 |
| -0,87955 | 421 | 180,7994 | 0,380269 | 1 | 0,657185 |
| -1,75005 | 420 | 348,757 | 0,080989 | 1 | 0,848246 |

| | | | | | |
|---|---|---|---|---|---|
| -1,81872 | 367 | 316,8158 | 0,069898 | 1 | 0,364488 |
| -1,71567 | 280 | 256,1882 | 0,087431 | 1 | 0,460069 |
| 0,641437 | 421 | 376,9035 | 0,521629 | 1 | 0,769172 |
| 2,061612 | 421 | 198,9196 | 0,040545 | 1 | 0,270359 |
| 0,314452 | 420 | 404,0615 | 0,75334 | 1 | 0,969427 |
| 0,365956 | 367 | 351,0396 | 0,714618 | 1 | 0,912652 |
| -0,23619 | 280 | 262,6273 | 0,813467 | 1 | 0,95747 |
| 0,755926 | 421 | 346,4492 | 0,450207 | 1 | 0,714643 |
| 0,369028 | 421 | 189,5066 | 0,712519 | 1 | 0,872396 |
| 1,363773 | 420 | 381,7732 | 0,173443 | 1 | 0,9199 |
| 1,934065 | 367 | 321,1918 | 0,053983 | 1 | 0,334402 |
| 0,026921 | 280 | 258,5065 | 0,978543 | 1 | 0,99236 |
| 1,089249 | 421 | 361,2544 | 0,27677 | 1 | 0,57291 |
| -0,7926 | 421 | 195,0737 | 0,428972 | 1 | 0,694836 |
| -1,36536 | 420 | 394,2329 | 0,172918 | 1 | 0,9199 |
| -0,66717 | 367 | 339,1857 | 0,505116 | 1 | 0,793601 |
| -0,38982 | 280 | 253,1574 | 0,696997 | 1 | 0,92034 |
| -2,67978 | 421 | 349,7835 | 0,007715 | 0,740677 | 0,108664 |
| -1,23679 | 421 | 190,1101 | 0,217692 | 1 | 0,514565 |
| -0,73399 | 420 | 386,0862 | 0,463401 | 1 | 0,960096 |
| -1,89235 | 367 | 340,6708 | 0,059292 | 1 | 0,340409 |
| -0,91016 | 280 | 263,2884 | 0,36357 | 1 | 0,762031 |
| 2,259586 | 421 | 364,4617 | 0,024437 | 1 | 0,182309 |
| 1,47822 | 421 | 195,8642 | 0,140955 | 1 | 0,423499 |
| 1,038336 | 420 | 397,0482 | 0,299746 | 1 | 0,943897 |
| 1,241084 | 367 | 340,9545 | 0,215428 | 1 | 0,56658 |
| -0,11218 | 280 | 261,8314 | 0,910765 | 1 | 0,976008 |
| -1,14691 | 421 | 332,2245 | 0,252245 | 1 | 0,546261 |
| -1,48335 | 421 | 185,333 | 0,13968 | 1 | 0,423351 |
| -0,15214 | 420 | 365,6514 | 0,879162 | 1 | 0,982463 |
| -0,36997 | 367 | 331,8533 | 0,711644 | 1 | 0,912652 |
| -0,90117 | 280 | 262,6047 | 0,368322 | 1 | 0,763914 |
| -1,02783 | 421 | 376,161 | 0,304692 | 1 | 0,601009 |
| -1,75388 | 421 | 200,1319 | 0,080981 | 1 | 0,360416 |
| -0,87652 | 420 | 404,2818 | 0,381268 | 1 | 0,95743 |
| -1,36521 | 367 | 348,4898 | 0,173069 | 1 | 0,522676 |
| -0,34471 | 280 | 272,9637 | 0,730577 | 1 | 0,936637 |
| -0,96124 | 421 | 334,1762 | 0,337124 | 1 | 0,627993 |
| -2,40358 | 421 | 184,0963 | 0,017228 | 1 | 0,181988 |
| -1,40128 | 420 | 369,87 | 0,161968 | 1 | 0,914005 |
| -2,04286 | 367 | 326,5031 | 0,041869 | 1 | 0,291798 |
| -2,25647 | 280 | 258,1614 | 0,024878 | 1 | 0,315049 |
| -2,71607 | 421 | 409,5691 | 0,006886 | 0,667938 | 0,10482 |
| -2,42689 | 421 | 224,4612 | 0,016018 | 1 | 0,179127 |
| -1,4337 | 420 | 415,9468 | 0,15241 | 1 | 0,914005 |
| -2,16101 | 367 | 362,6483 | 0,031348 | 1 | 0,266905 |
| -0,34292 | 280 | 274,2284 | 0,731922 | 1 | 0,936637 |
| -1,06487 | 421 | 349,1616 | 0,287669 | 1 | 0,584852 |

| | | | | | |
|---|---|---|---|---|---|
| -0,23547 | 421 | 190,7353 | 0,814094 | 1 | 0,926063 |
| 0,484273 | 420 | 383,4703 | 0,628468 | 1 | 0,964878 |
| 0,240255 | 367 | 336,9295 | 0,810279 | 1 | 0,950149 |
| -0,21962 | 280 | 258,6026 | 0,826339 | 1 | 0,96121 |
| -1,27213 | 421 | 349,6439 | 0,204173 | 1 | 0,494095 |
| -2,39569 | 421 | 189,4028 | 0,017563 | 1 | 0,182536 |
| -0,14913 | 420 | 384,2939 | 0,881528 | 1 | 0,982463 |
| -2,87197 | 367 | 339,5721 | 0,004335 | 0,420541 | 0,178183 |
| -1,88633 | 280 | 258,1757 | 0,060373 | 1 | 0,422605 |
| -1,5576 | 421 | 346,5078 | 0,12024 | 1 | 0,381479 |
| 1,683495 | 421 | 186,7694 | 0,09395 | 1 | 0,373113 |
| -0,44466 | 420 | 379,8227 | 0,656816 | 1 | 0,96781 |
| -0,42933 | 367 | 335,8906 | 0,667957 | 1 | 0,894765 |
| 0,239561 | 280 | 260,1924 | 0,81086 | 1 | 0,95747 |
| 0,316286 | 421 | 346,1685 | 0,751976 | 1 | 0,898125 |
| 1,242218 | 421 | 190,3798 | 0,215684 | 1 | 0,514565 |
| 1,628376 | 420 | 381,2408 | 0,104271 | 1 | 0,848246 |
| 1,215799 | 367 | 341,5284 | 0,224901 | 1 | 0,576775 |
| 0,51618 | 280 | 268,0465 | 0,606154 | 1 | 0,88515 |
| -0,15188 | 421 | 341,6175 | 0,87937 | 1 | 0,95861 |
| -0,28367 | 421 | 184,5822 | 0,776981 | 1 | 0,911265 |
| 0,213856 | 420 | 374,9309 | 0,830775 | 1 | 0,973894 |
| 0,797509 | 367 | 326,6733 | 0,425735 | 1 | 0,733464 |
| 0,264908 | 280 | 248,9713 | 0,7913 | 1 | 0,953435 |
| 0,329055 | 421 | 358,7989 | 0,742306 | 1 | 0,894925 |
| 2,780287 | 421 | 193,9782 | 0,005966 | 0,584682 | 0,141161 |
| 0,223391 | 420 | 391,755 | 0,823348 | 1 | 0,972623 |
| 1,59096 | 367 | 337,3743 | 0,112555 | 1 | 0,438296 |
| 2,005076 | 280 | 268,4246 | 0,045959 | 1 | 0,387749 |
| -1,55896 | 421 | 317,3176 | 0,120003 | 1 | 0,381479 |
| -0,87481 | 421 | 180,7585 | 0,382839 | 1 | 0,659083 |
| -1,84507 | 420 | 345,405 | 0,065883 | 1 | 0,848246 |
| -0,44048 | 367 | 313,7942 | 0,659892 | 1 | 0,893492 |
| 0,024636 | 280 | 252,7957 | 0,980365 | 1 | 0,99236 |
| 0,406695 | 421 | 343,2761 | 0,684485 | 1 | 0,863919 |
| -0,04497 | 421 | 185,9522 | 0,964177 | 1 | 0,985228 |
| -1,03722 | 420 | 375,315 | 0,300301 | 1 | 0,943897 |
| -0,39001 | 367 | 313,5799 | 0,696796 | 1 | 0,905235 |
| 1,446168 | 280 | 252,0211 | 0,149373 | 1 | 0,561358 |
| 2,060189 | 421 | 370,6119 | 0,040078 | 1 | 0,224527 |
| 2,518889 | 421 | 199,9442 | 0,012556 | 1 | 0,169272 |
| 0,811017 | 420 | 403,5289 | 0,417834 | 1 | 0,95743 |
| 0,27603 | 367 | 354,1688 | 0,782686 | 1 | 0,94337 |
| 0,346726 | 280 | 270,6415 | 0,729066 | 1 | 0,936637 |
| -0,27672 | 421 | 346,3147 | 0,782158 | 1 | 0,915149 |
| -0,38554 | 421 | 189,8947 | 0,700267 | 1 | 0,865528 |
| 0,94199 | 420 | 379,233 | 0,346798 | 1 | 0,947373 |
| -0,0265 | 367 | 332,8107 | 0,978876 | 1 | 0,99209 |

| | | | | | |
|---|---|---|---|---|---|
| 0,653619 | 280 | 257,1646 | 0,513941 | 1 | 0,831912 |
| 0,761286 | 421 | 373,7025 | 0,446966 | 1 | 0,714643 |
| -0,73584 | 421 | 198,4296 | 0,4627 | 1 | 0,721079 |
| 0,401186 | 420 | 402,024 | 0,688496 | 1 | 0,968866 |
| 1,186262 | 367 | 340,4124 | 0,236346 | 1 | 0,585627 |
| 0,651635 | 280 | 267,7769 | 0,515195 | 1 | 0,83211 |
| 1,596003 | 421 | 364,7889 | 0,111354 | 1 | 0,358728 |
| 1,56664 | 421 | 197,3491 | 0,118801 | 1 | 0,396436 |
| -0,15115 | 420 | 399,1967 | 0,879934 | 1 | 0,982463 |
| 2,565891 | 367 | 350,6723 | 0,010706 | 1 | 0,189546 |
| 0,516182 | 280 | 271,2229 | 0,606148 | 1 | 0,88515 |
| 0,683739 | 421 | 320,4866 | 0,494634 | 1 | 0,74916 |
| 0,447004 | 421 | 182,3297 | 0,655403 | 1 | 0,841107 |
| 0,662791 | 420 | 355,68 | 0,507894 | 1 | 0,960096 |
| -0,67382 | 367 | 318,5946 | 0,500917 | 1 | 0,793601 |
| 2,096842 | 280 | 260,2183 | 0,036973 | 1 | 0,34183 |
| -1,16646 | 421 | 298,8357 | 0,244357 | 1 | 0,541538 |
| -0,87276 | 421 | 175,029 | 0,383991 | 1 | 0,659083 |
| 0,594196 | 420 | 337,2803 | 0,552779 | 1 | 0,960096 |
| -0,34634 | 367 | 299,3205 | 0,729329 | 1 | 0,922837 |
| 0,913383 | 280 | 251,9268 | 0,361914 | 1 | 0,762031 |
| 2,282877 | 421 | 375,1051 | 0,022997 | 1 | 0,177419 |
| 3,231 | 421 | 200,8891 | 0,001441 | 0,190274 | 0,093671 |
| 0,988044 | 420 | 406,8304 | 0,323718 | 1 | 0,945573 |
| 3,369421 | 367 | 355,2434 | 8,36E-04 | 0,112016 | 0,139974 |
| 1,226673 | 280 | 272,3915 | 0,221005 | 1 | 0,653047 |
| -1,26509 | 421 | 386,5772 | 0,2066 | 1 | 0,496566 |
| -1,4215 | 421 | 202,5397 | 0,156708 | 1 | 0,438625 |
| -0,70213 | 420 | 409,6853 | 0,482995 | 1 | 0,960096 |
| -0,92302 | 367 | 357,7803 | 0,356621 | 1 | 0,674813 |
| -2,45016 | 280 | 274,1624 | 0,014905 | 1 | 0,263522 |
| -0,95319 | 421 | 358,9711 | 0,341136 | 1 | 0,63164 |
| -1,96442 | 421 | 191,5368 | 0,050927 | 1 | 0,294052 |
| 0,681747 | 420 | 393,3773 | 0,4958 | 1 | 0,960096 |
| -0,20799 | 367 | 332,335 | 0,835362 | 1 | 0,956935 |
| -0,78959 | 280 | 263,7572 | 0,430475 | 1 | 0,801886 |
| -0,57102 | 421 | 320,164 | 0,568383 | 1 | 0,796637 |
| -0,87142 | 421 | 181,8357 | 0,384674 | 1 | 0,659504 |
| -2,16828 | 420 | 353,221 | 0,030805 | 1 | 0,80141 |
| 0,988245 | 367 | 316,515 | 0,323788 | 1 | 0,655597 |
| 0,461243 | 280 | 256,6757 | 0,645015 | 1 | 0,907804 |
| 2,834002 | 421 | 359,8424 | 0,004856 | 0,607022 | 0,087964 |
| 2,78222 | 421 | 195,9348 | 0,005927 | 0,758613 | 0,141161 |
| 0,596319 | 420 | 396,9399 | 0,551302 | 1 | 0,960096 |
| 1,346105 | 367 | 355,9509 | 0,179125 | 1 | 0,526202 |
| 0,144776 | 280 | 272,9712 | 0,884995 | 1 | 0,973784 |
| 0,122606 | 421 | 349,8235 | 0,90249 | 1 | 0,965517 |
| -1,50292 | 421 | 188,1665 | 0,134535 | 1 | 0,417169 |

| | | | | | |
|---|---|---|---|---|---|
| -0,00853 | 420 | 383,5007 | 0,993197 | 1 | 0,997342 |
| 2,080609 | 367 | 325,8065 | 0,038251 | 1 | 0,28956 |
| 0,934912 | 280 | 250,5371 | 0,350734 | 1 | 0,761846 |
| -3,02766 | 421 | 345,7989 | 0,00265 | 0,34445 | 0,073944 |
| -0,88713 | 421 | 190,1779 | 0,376128 | 1 | 0,654686 |
| 0,131268 | 420 | 384,3475 | 0,895632 | 1 | 0,98503 |
| -0,96414 | 367 | 339,5607 | 0,335664 | 1 | 0,662363 |
| 0,675014 | 280 | 255,4503 | 0,500277 | 1 | 0,831912 |
| 0,412868 | 421 | 335,2299 | 0,679967 | 1 | 0,861268 |
| 0,974545 | 421 | 186,0173 | 0,331051 | 1 | 0,626331 |
| 0,77193 | 420 | 370,8254 | 0,440647 | 1 | 0,960096 |
| 1,109686 | 367 | 325,7066 | 0,267953 | 1 | 0,609149 |
| 1,944799 | 280 | 251,1354 | 0,052915 | 1 | 0,402746 |
| 0,499732 | 421 | 367,058 | 0,617564 | 1 | 0,826526 |
| 0,252733 | 421 | 195,6396 | 0,80074 | 1 | 0,921861 |
| 1,95664 | 420 | 395,4648 | 0,051093 | 1 | 0,846627 |
| -0,0931 | 367 | 347,9994 | 0,925875 | 1 | 0,976733 |
| 0,461561 | 280 | 270,5513 | 0,644767 | 1 | 0,907804 |
| 2,076724 | 421 | 350,3698 | 0,038555 | 1 | 0,221767 |
| 1,625023 | 421 | 190,12 | 0,105814 | 1 | 0,385174 |
| 0,831027 | 420 | 385,6183 | 0,406472 | 1 | 0,95743 |
| 1,398657 | 367 | 331,3182 | 0,162851 | 1 | 0,510222 |
| 1,386404 | 280 | 272,5164 | 0,166757 | 1 | 0,5913 |
| -1,27946 | 421 | 340,6521 | 0,201607 | 1 | 0,490826 |
| -1,47116 | 421 | 187,4123 | 0,142925 | 1 | 0,423993 |
| -1,77772 | 420 | 366,5003 | 0,076278 | 1 | 0,848246 |
| -0,57359 | 367 | 327,9016 | 0,566636 | 1 | 0,837838 |
| -0,07228 | 280 | 258,542 | 0,942438 | 1 | 0,982175 |
| 0,493649 | 421 | 374,8659 | 0,621843 | 1 | 0,828413 |
| 0,940548 | 421 | 200,82 | 0,348066 | 1 | 0,642814 |
| -0,69383 | 420 | 402,7754 | 0,488188 | 1 | 0,960096 |
| 1,35665 | 367 | 351,703 | 0,175763 | 1 | 0,523018 |
| 1,852253 | 280 | 262,1248 | 0,065114 | 1 | 0,424733 |
| -2,1958 | 421 | 414,7276 | 0,028659 | 1 | 0,197211 |
| -2,48118 | 421 | 237,6112 | 0,013788 | 1 | 0,17172 |
| -0,47479 | 420 | 411,597 | 0,635188 | 1 | 0,964878 |
| -2,55936 | 367 | 360,4131 | 0,010894 | 1 | 0,189546 |
| 0,435822 | 280 | 274,655 | 0,663308 | 1 | 0,908644 |
| 1,758954 | 421 | 363,7784 | 0,079426 | 1 | 0,302838 |
| 1,373401 | 421 | 195,7659 | 0,171199 | 1 | 0,456365 |
| 1,752774 | 420 | 396,643 | 0,080413 | 1 | 0,848246 |
| 1,577505 | 367 | 350,2264 | 0,115582 | 1 | 0,443225 |
| -0,60524 | 280 | 268,991 | 0,545533 | 1 | 0,851507 |
| -0,2169 | 421 | 315,9284 | 0,828426 | 1 | 0,936611 |
| -0,21978 | 421 | 177,4269 | 0,826299 | 1 | 0,932758 |
| -0,90875 | 420 | 351,1132 | 0,364106 | 1 | 0,956033 |
| 0,589492 | 367 | 314,432 | 0,555955 | 1 | 0,830257 |
| 2,060237 | 280 | 246,6695 | 0,040425 | 1 | 0,364793 |

| | | | | | |
|---|---|---|---|---|---|
| 1,435106 | 421 | 377,9844 | 0,152084 | 1 | 0,42445 |
| 1,656072 | 421 | 201,9085 | 0,09926 | 1 | 0,379659 |
| -0,45653 | 420 | 405,972 | 0,648251 | 1 | 0,964878 |
| 1,472753 | 367 | 354,264 | 0,141706 | 1 | 0,489794 |
| 1,76418 | 280 | 271,4847 | 0,078826 | 1 | 0,453386 |
| -1,15474 | 421 | 353,7791 | 0,248977 | 1 | 0,545362 |
| 0,006725 | 421 | 192,1043 | 0,994641 | 1 | 0,997935 |
| -1,06176 | 420 | 386,2803 | 0,28901 | 1 | 0,943897 |
| 0,497143 | 367 | 340,1701 | 0,619409 | 1 | 0,875495 |
| 0,7167 | 280 | 261,8411 | 0,474198 | 1 | 0,824256 |
| 0,262689 | 421 | 306,463 | 0,792967 | 1 | 0,919136 |
| 0,189944 | 421 | 175,6259 | 0,849572 | 1 | 0,944105 |
| -0,11119 | 420 | 340,1732 | 0,91153 | 1 | 0,987181 |
| 1,007129 | 367 | 292,3943 | 0,314706 | 1 | 0,647568 |
| 2,200574 | 280 | 276 | 0,028595 | 1 | 0,326641 |
| 0,602088 | 421 | 347,5032 | 0,547508 | 1 | 0,784719 |
| -0,64957 | 421 | 188,5686 | 0,516761 | 1 | 0,76125 |
| 1,222422 | 420 | 383,6817 | 0,222299 | 1 | 0,943055 |
| 0,134372 | 367 | 318,6362 | 0,893193 | 1 | 0,969073 |
| 0,851167 | 280 | 258,7232 | 0,395464 | 1 | 0,782194 |
| 0,012854 | 421 | 340,3181 | 0,989752 | 1 | 0,998054 |
| 0,685944 | 421 | 185,8364 | 0,493603 | 1 | 0,746291 |
| 0,011601 | 420 | 374,9416 | 0,99075 | 1 | 0,997342 |
| 0,729 | 367 | 328,1466 | 0,466522 | 1 | 0,771623 |
| -0,66912 | 280 | 263,9313 | 0,504007 | 1 | 0,831912 |
| -2,64757 | 421 | 330,376 | 0,008496 | 1 | 0,112451 |
| -0,55862 | 421 | 183,7565 | 0,577103 | 1 | 0,794719 |
| -1,08055 | 420 | 367,2509 | 0,280608 | 1 | 0,943897 |
| -1,92471 | 367 | 325,2589 | 0,055138 | 1 | 0,335053 |
| 1,789363 | 280 | 267,5238 | 0,074688 | 1 | 0,447755 |
| -0,06239 | 421 | 334,0884 | 0,950289 | 1 | 0,984251 |
| 0,479458 | 421 | 186,1747 | 0,632175 | 1 | 0,835691 |
| 1,324237 | 420 | 369,8185 | 0,186242 | 1 | 0,923408 |
| -1,15952 | 367 | 330,9593 | 0,247081 | 1 | 0,600675 |
| -0,91421 | 280 | 255,4472 | 0,361469 | 1 | 0,762031 |
| -0,02977 | 421 | 398,3421 | 0,976264 | 1 | 0,995982 |
| -0,60979 | 421 | 214,2749 | 0,542645 | 1 | 0,774002 |
| -1,36486 | 420 | 414,3595 | 0,173039 | 1 | 0,9199 |
| 1,111662 | 367 | 357,7998 | 0,26703 | 1 | 0,609149 |
| -0,8861 | 280 | 269,6596 | 0,376352 | 1 | 0,767473 |
| 1,92845 | 421 | 339,255 | 0,054633 | 1 | 0,259162 |
| 1,900368 | 421 | 184,618 | 0,058943 | 1 | 0,314262 |
| 1,337016 | 420 | 378,6396 | 0,18202 | 1 | 0,923408 |
| 1,287005 | 367 | 321,0334 | 0,19902 | 1 | 0,551573 |
| 3,1135 | 280 | 264,0337 | 0,002052 | 0,266792 | 0,135787 |
| 1,227599 | 421 | 343,4434 | 0,220438 | 1 | 0,515839 |
| 1,028797 | 421 | 187,3841 | 0,304901 | 1 | 0,602952 |
| 1,324899 | 420 | 377,3163 | 0,186006 | 1 | 0,923408 |

| | | | | | |
|---|---|---|---|---|---|
| 1,443458 | 367 | 340,2888 | 0,149811 | 1 | 0,499998 |
| -0,05655 | 280 | 262,8967 | 0,95495 | 1 | 0,984343 |
| 0,757787 | 421 | 408,521 | 0,449016 | 1 | 0,714643 |
| -0,067 | 421 | 224,4329 | 0,94664 | 1 | 0,980846 |
| -1,0104 | 420 | 415,8334 | 0,312893 | 1 | 0,943897 |
| -0,47138 | 367 | 362,6481 | 0,637655 | 1 | 0,880707 |
| -1,52928 | 280 | 275,7835 | 0,127342 | 1 | 0,536498 |
| -1,76382 | 421 | 326,4084 | 0,078697 | 1 | 0,302838 |
| -0,12963 | 421 | 180,4628 | 0,897007 | 1 | 0,960059 |
| -1,33856 | 420 | 359,0741 | 0,181559 | 1 | 0,923408 |
| 1,493816 | 367 | 320,578 | 0,136207 | 1 | 0,479589 |
| 1,470449 | 280 | 251,305 | 0,142691 | 1 | 0,554427 |
| 2,821922 | 421 | 387,5174 | 0,00502 | 0,622425 | 0,087964 |
| 2,357531 | 421 | 210,1825 | 0,019315 | 1 | 0,195731 |
| 0,663607 | 420 | 412,7806 | 0,507312 | 1 | 0,960096 |
| 2,055167 | 367 | 358,6988 | 0,040587 | 1 | 0,289956 |
| 1,143702 | 280 | 275,3496 | 0,25374 | 1 | 0,679673 |
| -2,91662 | 421 | 363,5551 | 0,003758 | 0,484802 | 0,087964 |
| -1,68717 | 421 | 194,0786 | 0,093178 | 1 | 0,373113 |
| -0,7458 | 420 | 399,3941 | 0,456229 | 1 | 0,960096 |
| -1,17163 | 367 | 349,6307 | 0,242143 | 1 | 0,59405 |
| 0,092755 | 280 | 274,0546 | 0,926166 | 1 | 0,979504 |
| 3,532367 | 421 | 409,1716 | 4,59E-04 | 0,061017 | 0,04642 |
| 2,214153 | 421 | 227,149 | 0,027812 | 1 | 0,226812 |
| 1,210112 | 420 | 415,7238 | 0,226924 | 1 | 0,943055 |
| 1,974572 | 367 | 362,3363 | 0,049076 | 1 | 0,314711 |
| 0,595531 | 280 | 275,9185 | 0,551976 | 1 | 0,854033 |
| -1,88597 | 421 | 329,0351 | 0,06018 | 1 | 0,270611 |
| -0,68273 | 421 | 184,2899 | 0,495635 | 1 | 0,747712 |
| -1,77191 | 420 | 360,9004 | 0,077253 | 1 | 0,848246 |
| -0,67889 | 367 | 328,4753 | 0,497685 | 1 | 0,793601 |
| -0,16181 | 280 | 254,024 | 0,871582 | 1 | 0,969656 |
| 1,866546 | 421 | 356,3081 | 0,062786 | 1 | 0,273465 |
| 2,439122 | 421 | 191,2629 | 0,015636 | 1 | 0,179127 |
| 1,289924 | 420 | 395,2622 | 0,197831 | 1 | 0,928589 |
| 1,759116 | 367 | 337,6802 | 0,079464 | 1 | 0,381841 |
| 3,968968 | 280 | 268,4838 | 9,27E-05 | 0,012424 | 0,046455 |
| 0,849953 | 421 | 383,4022 | 0,395882 | 1 | 0,686065 |
| 0,086678 | 421 | 205,4958 | 0,931012 | 1 | 0,97433 |
| 1,059055 | 420 | 409,1698 | 0,2902 | 1 | 0,943897 |
| 1,131329 | 367 | 358,1613 | 0,258674 | 1 | 0,609149 |
| 0,593773 | 280 | 272,3697 | 0,553157 | 1 | 0,854225 |
| 2,380993 | 421 | 391,6578 | 0,017743 | 1 | 0,15546 |
| 2,15866 | 421 | 210,3719 | 0,032009 | 1 | 0,241423 |
| 0,233176 | 420 | 413,337 | 0,81574 | 1 | 0,969427 |
| 2,777008 | 367 | 360,1829 | 0,005773 | 0,750497 | 0,178183 |
| 1,303998 | 280 | 275,28 | 0,193324 | 1 | 0,62543 |
| 0,016574 | 421 | 392,1099 | 0,986785 | 1 | 0,998043 |

| | | | | | |
|---|---|---|---|---|---|
| -0,93092 | 421 | 208,1287 | 0,352971 | 1 | 0,643758 |
| -1,63568 | 420 | 412,1823 | 0,102669 | 1 | 0,848246 |
| -0,97986 | 367 | 356,7202 | 0,32782 | 1 | 0,655788 |
| 0,186827 | 280 | 271,3233 | 0,851936 | 1 | 0,966655 |
| -8,80E-04 | 421 | 399,495 | 0,999298 | 1 | 0,999298 |
| 0,887466 | 421 | 214,7017 | 0,375821 | 1 | 0,654686 |
| -0,13849 | 420 | 414,7987 | 0,889921 | 1 | 0,983483 |
| -1,31677 | 367 | 361,5123 | 0,188749 | 1 | 0,538722 |
| 0,437764 | 280 | 270,7268 | 0,661906 | 1 | 0,908644 |
| -1,21373 | 421 | 390,2789 | 0,225583 | 1 | 0,518147 |
| -1,15202 | 421 | 208,6058 | 0,250633 | 1 | 0,548794 |
| 0,559135 | 420 | 412,0877 | 0,576373 | 1 | 0,964878 |
| -0,99195 | 367 | 362,7919 | 0,321885 | 1 | 0,653748 |
| 0,087227 | 280 | 274,5375 | 0,930554 | 1 | 0,979504 |
| -0,64252 | 421 | 352,9978 | 0,520952 | 1 | 0,769172 |
| -1,18318 | 421 | 192,5727 | 0,238196 | 1 | 0,538172 |
| -0,52507 | 420 | 386,9614 | 0,599836 | 1 | 0,964878 |
| -1,59245 | 367 | 346,1897 | 0,112197 | 1 | 0,438296 |
| -0,37789 | 280 | 262,3846 | 0,705817 | 1 | 0,922762 |
| -0,93373 | 421 | 320,8366 | 0,351144 | 1 | 0,640439 |
| -1,49876 | 421 | 182,5074 | 0,135664 | 1 | 0,418945 |
| -1,46835 | 420 | 347,6517 | 0,142913 | 1 | 0,904916 |
| -0,14172 | 367 | 316,0889 | 0,887389 | 1 | 0,964898 |
| -0,38643 | 280 | 258,5706 | 0,699494 | 1 | 0,92034 |
| -0,57034 | 421 | 363,4435 | 0,5688 | 1 | 0,796637 |
| -0,37979 | 421 | 194,4411 | 0,704514 | 1 | 0,867404 |
| -1,31467 | 420 | 394,3205 | 0,189383 | 1 | 0,925729 |
| 0,550713 | 367 | 355,1226 | 0,582177 | 1 | 0,845222 |
| 3,462242 | 280 | 268,1811 | 6,23E-04 | 0,081647 | 0,093926 |
| -0,55707 | 421 | 351,9632 | 0,577836 | 1 | 0,802579 |
| -1,59608 | 421 | 188,8067 | 0,112144 | 1 | 0,391524 |
| -1,10563 | 420 | 384,2693 | 0,269577 | 1 | 0,943897 |
| 0,569782 | 367 | 335,0732 | 0,569208 | 1 | 0,837838 |
| 1,523638 | 280 | 252,4748 | 0,128851 | 1 | 0,536498 |
| -2,00596 | 421 | 376,732 | 0,045575 | 1 | 0,237403 |
| -0,26832 | 421 | 200,9375 | 0,78873 | 1 | 0,918329 |
| 0,009638 | 420 | 403,9462 | 0,992315 | 1 | 0,997342 |
| 0,670983 | 367 | 351,6451 | 0,502672 | 1 | 0,793601 |
| -0,03549 | 280 | 266,1456 | 0,971715 | 1 | 0,992124 |
| 0,313466 | 421 | 372,1573 | 0,754103 | 1 | 0,898125 |
| 0,736199 | 421 | 199,4363 | 0,462475 | 1 | 0,721079 |
| -1,39762 | 420 | 401,8831 | 0,162997 | 1 | 0,914005 |
| 0,767889 | 367 | 343,9403 | 0,44308 | 1 | 0,747728 |
| 1,636696 | 280 | 263,3375 | 0,102888 | 1 | 0,486059 |
| 1,99343 | 421 | 385,2465 | 0,046919 | 1 | 0,239762 |
| 2,623871 | 421 | 207,6318 | 0,009339 | 1 | 0,14815 |
| 0,481569 | 420 | 411,0817 | 0,630368 | 1 | 0,964878 |
| 2,590987 | 367 | 357,7162 | 0,009962 | 1 | 0,183886 |

| | | | | | |
|---|---|---|---|---|---|
| 1,864553 | 280 | 274,0769 | 0,063313 | 1 | 0,422605 |
| -2,2262 | 421 | 380,0743 | 0,026586 | 1 | 0,1917 |
| -1,37763 | 421 | 200,5801 | 0,169852 | 1 | 0,456365 |
| -0,66331 | 420 | 406,1307 | 0,507508 | 1 | 0,960096 |
| -1,35838 | 367 | 352,7254 | 0,175213 | 1 | 0,523018 |
| 0,058076 | 280 | 271,5206 | 0,953731 | 1 | 0,983759 |
| -1,99516 | 421 | 371,1883 | 0,046756 | 1 | 0,239762 |
| -1,62866 | 421 | 197,3732 | 0,104981 | 1 | 0,38493 |
| -0,93935 | 420 | 398,2066 | 0,348122 | 1 | 0,947373 |
| -0,0637 | 367 | 347,05 | 0,949246 | 1 | 0,979539 |
| -0,44743 | 280 | 256,4882 | 0,65494 | 1 | 0,908644 |
| 0,375108 | 421 | 366,2215 | 0,707798 | 1 | 0,875021 |
| 1,008285 | 421 | 194,0451 | 0,314573 | 1 | 0,61407 |
| 1,021226 | 420 | 397,9055 | 0,307768 | 1 | 0,943897 |
| 0,825235 | 367 | 345,9097 | 0,409807 | 1 | 0,722315 |
| 1,602627 | 280 | 270,712 | 0,110183 | 1 | 0,50165 |
| 0,911571 | 421 | 356,7666 | 0,36261 | 1 | 0,652872 |
| 1,541786 | 421 | 190,5442 | 0,124785 | 1 | 0,409697 |
| -0,64945 | 420 | 389,4323 | 0,516433 | 1 | 0,960096 |
| 1,81869 | 367 | 343,3177 | 0,06983 | 1 | 0,364488 |
| 0,484826 | 280 | 271,5365 | 0,628191 | 1 | 0,900874 |
| -0,35601 | 421 | 306,5804 | 0,72208 | 1 | 0,886136 |
| -0,69541 | 421 | 176,3063 | 0,487714 | 1 | 0,742408 |
| -0,51934 | 420 | 339,0085 | 0,603863 | 1 | 0,964878 |
| 0,270179 | 367 | 307,0635 | 0,787204 | 1 | 0,943768 |
| -0,20474 | 280 | 249,1058 | 0,837946 | 1 | 0,964667 |
| 2,148605 | 421 | 367,8403 | 0,032318 | 1 | 0,20468 |
| 2,644373 | 421 | 198,0905 | 0,00884 | 1 | 0,14815 |
| 1,940205 | 420 | 399,5724 | 0,053058 | 1 | 0,846627 |
| 2,095809 | 367 | 347,9021 | 0,036821 | 1 | 0,28464 |
| 0,974942 | 280 | 269,0204 | 0,330465 | 1 | 0,752282 |
| 0,141743 | 421 | 348,7469 | 0,887365 | 1 | 0,962 |
| -0,99975 | 421 | 188,7404 | 0,318711 | 1 | 0,617081 |
| -1,18078 | 420 | 382,9329 | 0,238423 | 1 | 0,943055 |
| 1,718107 | 367 | 338,1913 | 0,086692 | 1 | 0,391959 |
| -0,08373 | 280 | 255,7537 | 0,933333 | 1 | 0,979504 |
| 0,132986 | 421 | 324,5744 | 0,894287 | 1 | 0,962 |
| -0,68218 | 421 | 183,0403 | 0,49599 | 1 | 0,747712 |
| 0,734122 | 420 | 358,9402 | 0,463353 | 1 | 0,960096 |
| 1,17111 | 367 | 312,5532 | 0,242447 | 1 | 0,59405 |
| -0,54648 | 280 | 247,4966 | 0,585231 | 1 | 0,87561 |
| 1,806316 | 421 | 309,1336 | 0,071841 | 1 | 0,288707 |
| 1,196105 | 421 | 179,5539 | 0,233233 | 1 | 0,533704 |
| -1,33288 | 420 | 339,2819 | 0,183467 | 1 | 0,923408 |
| 2,436245 | 367 | 307,3588 | 0,015408 | 1 | 0,204372 |
| 2,392986 | 280 | 251,4394 | 0,017446 | 1 | 0,26828 |
| 1,016579 | 421 | 407,6866 | 0,309957 | 1 | 0,605668 |
| 3,690145 | 421 | 223,4056 | 2,82E-04 | 0,037736 | 0,035752 |

| | | | | | |
|---|---|---|---|---|---|
| 0,455165 | 420 | 415,9149 | 0,649228 | 1 | 0,964878 |
| 2,069324 | 367 | 363 | 0,039223 | 1 | 0,28956 |
| 1,735315 | 280 | 275,9977 | 0,083801 | 1 | 0,456738 |
| -0,79218 | 421 | 331,1414 | 0,428824 | 1 | 0,700922 |
| 0,592712 | 421 | 185,8598 | 0,554094 | 1 | 0,778237 |
| -1,70282 | 420 | 362,6613 | 0,089458 | 1 | 0,848246 |
| 0,195511 | 367 | 318,2352 | 0,845117 | 1 | 0,956935 |
| -0,80533 | 280 | 254,4483 | 0,421379 | 1 | 0,796701 |
| -0,63983 | 421 | 377,0304 | 0,522669 | 1 | 0,769202 |
| 0,383523 | 421 | 202,5454 | 0,701734 | 1 | 0,866104 |
| 0,590918 | 420 | 405,7996 | 0,554904 | 1 | 0,960096 |
| -1,87002 | 367 | 355,2656 | 0,062303 | 1 | 0,347744 |
| 0,23337 | 280 | 271,8122 | 0,81565 | 1 | 0,95747 |
| 2,296635 | 421 | 391,4916 | 0,022167 | 1 | 0,176042 |
| 1,64528 | 421 | 210,0306 | 0,101408 | 1 | 0,383011 |
| -0,14694 | 420 | 412,6549 | 0,883253 | 1 | 0,982463 |
| 2,053866 | 367 | 360,4578 | 0,040709 | 1 | 0,289956 |
| 0,850998 | 280 | 275,7039 | 0,395509 | 1 | 0,782194 |
| 2,005333 | 421 | 389,4155 | 0,045619 | 1 | 0,237403 |
| 2,128965 | 421 | 208,3466 | 0,034431 | 1 | 0,252759 |
| 0,051653 | 420 | 412,0718 | 0,95883 | 1 | 0,996522 |
| 2,407072 | 367 | 358,8386 | 0,016585 | 1 | 0,210035 |
| 1,457139 | 280 | 274,9854 | 0,146219 | 1 | 0,555466 |
| -0,69784 | 421 | 343,9624 | 0,485751 | 1 | 0,745445 |
| -1,37096 | 421 | 185,208 | 0,172047 | 1 | 0,456365 |
| -0,94649 | 420 | 372,6044 | 0,344511 | 1 | 0,947373 |
| -0,27226 | 367 | 324,5151 | 0,785593 | 1 | 0,94337 |
| 0,579383 | 280 | 254,6474 | 0,562843 | 1 | 0,861728 |
| -0,27938 | 421 | 317,7411 | 0,780134 | 1 | 0,9142 |
| 0,602313 | 421 | 179,7743 | 0,547725 | 1 | 0,776424 |
| -0,41993 | 420 | 347,759 | 0,674798 | 1 | 0,96781 |
| 1,408394 | 367 | 314,5385 | 0,160002 | 1 | 0,504558 |
| 1,295506 | 280 | 250,3155 | 0,196339 | 1 | 0,626871 |
| -0,6964 | 421 | 360,2955 | 0,486627 | 1 | 0,74603 |
| -1,13238 | 421 | 194,1037 | 0,258871 | 1 | 0,556516 |
| -2,10898 | 420 | 391,4758 | 0,035581 | 1 | 0,828713 |
| -3,11274 | 367 | 323,801 | 0,002019 | 0,268504 | 0,176492 |
| -0,87701 | 280 | 268,5395 | 0,381263 | 1 | 0,770948 |
| 0,54639 | 421 | 384,8449 | 0,585115 | 1 | 0,807036 |
| 1,194291 | 421 | 205,8623 | 0,233738 | 1 | 0,533704 |
| -1,48475 | 420 | 409,5319 | 0,138378 | 1 | 0,891179 |
| 1,169882 | 367 | 359,3835 | 0,242824 | 1 | 0,59405 |
| 0,685823 | 280 | 270,8531 | 0,493411 | 1 | 0,827962 |
| 0,386603 | 421 | 361,7141 | 0,699277 | 1 | 0,873083 |
| 0,952991 | 421 | 193,0779 | 0,341786 | 1 | 0,635891 |
| 0,016149 | 420 | 394,4707 | 0,987124 | 1 | 0,997342 |
| 0,620927 | 367 | 346,6334 | 0,535055 | 1 | 0,816122 |
| 0,617856 | 280 | 266,8046 | 0,537197 | 1 | 0,845931 |

| | | | | | |
|---|---|---|---|---|---|
| 0,21005 | 421 | 360,3783 | 0,833747 | 1 | 0,936611 |
| 0,690447 | 421 | 192,0819 | 0,490747 | 1 | 0,744768 |
| -0,48561 | 420 | 391,5192 | 0,627513 | 1 | 0,964878 |
| 0,599507 | 367 | 341,8858 | 0,549232 | 1 | 0,828521 |
| 1,17947 | 280 | 255,3491 | 0,239309 | 1 | 0,662939 |
| 4,209855 | 421 | 335,9773 | 3,28E-05 | 0,035439 | 0,019267 |
| 1,803898 | 421 | 185,0828 | 0,072873 | 1 | 0,342119 |
| 1,66888 | 420 | 370,0816 | 0,095987 | 1 | 0,848246 |
| 1,614232 | 367 | 320,9227 | 0,10746 | 1 | 0,43416 |
| 1,365828 | 280 | 257,8501 | 0,173183 | 1 | 0,598625 |
| 0,603881 | 421 | 343,2753 | 0,546321 | 1 | 0,784719 |
| -0,06305 | 421 | 184,7521 | 0,949793 | 1 | 0,980953 |
| -1,82658 | 420 | 378,4254 | 0,068551 | 1 | 0,848246 |
| 1,162811 | 367 | 325,2963 | 0,245758 | 1 | 0,599343 |
| 0,278598 | 280 | 259,7905 | 0,780775 | 1 | 0,951006 |
| -2,17097 | 421 | 318,3981 | 0,030672 | 1 | 0,200565 |
| -1,54986 | 421 | 179,3698 | 0,122939 | 1 | 0,405402 |
| -1,18507 | 420 | 354,9007 | 0,236783 | 1 | 0,943055 |
| -1,51116 | 367 | 317,56 | 0,131742 | 1 | 0,475026 |
| -2,22797 | 280 | 259,9821 | 0,026739 | 1 | 0,324814 |
| 1,680079 | 421 | 368,6619 | 0,093789 | 1 | 0,328698 |
| 0,714594 | 421 | 194,4679 | 0,475717 | 1 | 0,734136 |
| 0,683316 | 420 | 397,9333 | 0,494805 | 1 | 0,960096 |
| -0,36661 | 367 | 351,5161 | 0,714128 | 1 | 0,912652 |
| -3,05591 | 280 | 266,4202 | 0,002472 | 1 | 0,135787 |
| -1,48988 | 421 | 331,2379 | 0,137208 | 1 | 0,409449 |
| -1,71079 | 421 | 185,1226 | 0,088794 | 1 | 0,370674 |
| -1,30045 | 420 | 365,4765 | 0,194267 | 1 | 0,928589 |
| -1,35935 | 367 | 304,9009 | 0,175039 | 1 | 0,523018 |
| -1,45712 | 280 | 254,9576 | 0,146315 | 1 | 0,555466 |
| -0,34045 | 421 | 389,5407 | 0,733705 | 1 | 0,893273 |
| 1,729944 | 421 | 207,594 | 0,085127 | 1 | 0,365475 |
| 0,929591 | 420 | 411,3676 | 0,353128 | 1 | 0,947373 |
| 1,661508 | 367 | 356,0661 | 0,097492 | 1 | 0,412696 |
| 1,312479 | 280 | 268,8923 | 0,190478 | 1 | 0,625249 |
| 1,152477 | 421 | 405,5012 | 0,249804 | 1 | 0,545586 |
| 2,241231 | 421 | 222,1854 | 0,026 | 1 | 0,222284 |
| -0,77546 | 420 | 415,9538 | 0,438506 | 1 | 0,960096 |
| 1,123594 | 367 | 362,8397 | 0,261928 | 1 | 0,609149 |
| 0,562918 | 280 | 274,1923 | 0,573951 | 1 | 0,870165 |
| 0,028254 | 421 | 341,6188 | 0,977476 | 1 | 0,995982 |
| -0,20081 | 421 | 186,4356 | 0,841065 | 1 | 0,940271 |
| -0,50773 | 420 | 374,5741 | 0,611941 | 1 | 0,964878 |
| 0,068016 | 367 | 339,1871 | 0,945813 | 1 | 0,979539 |
| -1,3619 | 280 | 257,4537 | 0,174419 | 1 | 0,598625 |
| 0,787873 | 421 | 343,8283 | 0,431314 | 1 | 0,704214 |
| 0,722057 | 421 | 188,0622 | 0,471156 | 1 | 0,731044 |
| -0,02652 | 420 | 376,5545 | 0,978857 | 1 | 0,997342 |

| | | | | | |
|---|---|---|---|---|---|
| -0,95333 | 367 | 329,9666 | 0,341122 | 1 | 0,664831 |
| 1,24191 | 280 | 252,7421 | 0,215421 | 1 | 0,648472 |
| 0,092829 | 421 | 378,9364 | 0,926088 | 1 | 0,968505 |
| 0,224185 | 421 | 202,6447 | 0,82284 | 1 | 0,930247 |
| 0,963537 | 420 | 406,1514 | 0,335851 | 1 | 0,947373 |
| 0,663308 | 367 | 354,4386 | 0,507565 | 1 | 0,795942 |
| -0,89322 | 280 | 267,825 | 0,372544 | 1 | 0,763914 |
| -1,98795 | 421 | 314,0213 | 0,047685 | 1 | 0,241202 |
| 0,37477 | 421 | 178,7183 | 0,708276 | 1 | 0,870812 |
| -1,3022 | 420 | 344,4671 | 0,193717 | 1 | 0,928589 |
| 1,10226 | 367 | 302,2262 | 0,271226 | 1 | 0,609149 |
| 0,698695 | 280 | 251,3969 | 0,485389 | 1 | 0,826532 |
| -1,01839 | 421 | 377,6997 | 0,309143 | 1 | 0,605668 |
| 0,007282 | 421 | 200,3375 | 0,994197 | 1 | 0,997935 |
| 0,829221 | 420 | 401,9973 | 0,407471 | 1 | 0,95743 |
| 0,324896 | 367 | 355,5222 | 0,745451 | 1 | 0,923669 |
| -2,02784 | 280 | 270,5668 | 0,043557 | 1 | 0,383858 |
| -1,20409 | 421 | 320,2351 | 0,229444 | 1 | 0,520357 |
| -0,68052 | 421 | 182,2466 | 0,49704 | 1 | 0,748291 |
| -0,25048 | 420 | 357,6387 | 0,802361 | 1 | 0,969427 |
| -0,06079 | 367 | 309,9695 | 0,951565 | 1 | 0,979686 |
| 1,359281 | 280 | 257,4192 | 0,175247 | 1 | 0,598625 |
| 0,78656 | 421 | 316,4432 | 0,432128 | 1 | 0,70478 |
| 1,894022 | 421 | 181,7821 | 0,05981 | 1 | 0,314727 |
| -1,26405 | 420 | 348,4078 | 0,207056 | 1 | 0,930277 |
| 0,262319 | 367 | 316,2982 | 0,793246 | 1 | 0,944997 |
| -0,14311 | 280 | 249,2967 | 0,886317 | 1 | 0,973784 |
| 1,586009 | 421 | 332,3537 | 0,113688 | 1 | 0,363754 |
| 2,027039 | 421 | 186,0862 | 0,044085 | 1 | 0,277922 |
| 0,619715 | 420 | 367,6693 | 0,53583 | 1 | 0,960096 |
| 0,129713 | 367 | 315,8309 | 0,896876 | 1 | 0,970443 |
| 0,331362 | 280 | 248,7716 | 0,74065 | 1 | 0,94032 |
| 1,756342 | 421 | 350,9696 | 0,079902 | 1 | 0,302838 |
| 0,79216 | 421 | 189,4407 | 0,429258 | 1 | 0,694836 |
| -1,09403 | 420 | 383,0714 | 0,274627 | 1 | 0,943897 |
| -0,46822 | 367 | 335,3083 | 0,63993 | 1 | 0,880707 |
| -0,51254 | 280 | 254,6599 | 0,608716 | 1 | 0,886655 |
| -0,94868 | 421 | 379,2157 | 0,343388 | 1 | 0,63185 |
| -1,0893 | 421 | 202,3555 | 0,277316 | 1 | 0,578094 |
| 1,242551 | 420 | 406,6358 | 0,214749 | 1 | 0,943055 |
| -0,00478 | 367 | 355,7578 | 0,996192 | 1 | 0,997516 |
| -0,94077 | 280 | 267,9901 | 0,347671 | 1 | 0,761841 |
| -1,76172 | 421 | 380,0784 | 0,07892 | 1 | 0,302838 |
| -2,43475 | 421 | 201,5687 | 0,015772 | 1 | 0,179127 |
| -0,64598 | 420 | 405,4417 | 0,518656 | 1 | 0,960096 |
| -0,70122 | 367 | 355,8835 | 0,483625 | 1 | 0,786044 |
| -0,55903 | 280 | 268,4008 | 0,57661 | 1 | 0,871183 |
| -1,46608 | 421 | 321,944 | 0,143603 | 1 | 0,412154 |

| | | | | | |
|---|---|---|---|---|---|
| -0,51185 | 421 | 182,4439 | 0,609376 | 1 | 0,82272 |
| 1,033199 | 420 | 356,3226 | 0,302212 | 1 | 0,943897 |
| -1,11625 | 367 | 319,5807 | 0,265154 | 1 | 0,609149 |
| -1,91559 | 280 | 251,5664 | 0,056551 | 1 | 0,406703 |
| 0,959016 | 421 | 339,3175 | 0,338233 | 1 | 0,629281 |
| 2,135883 | 421 | 186,7939 | 0,033991 | 1 | 0,252338 |
| 0,435029 | 420 | 380,2635 | 0,663788 | 1 | 0,96781 |
| 1,379469 | 367 | 332,8091 | 0,168677 | 1 | 0,514971 |
| 1,664512 | 280 | 251,6949 | 0,097254 | 1 | 0,480557 |
| -0,95294 | 421 | 378,8882 | 0,341227 | 1 | 0,63164 |
| -0,04686 | 421 | 201,9939 | 0,962671 | 1 | 0,985228 |
| 0,702573 | 420 | 404,7767 | 0,482726 | 1 | 0,960096 |
| -0,56394 | 367 | 342,9856 | 0,573164 | 1 | 0,840232 |
| -1,83645 | 280 | 265,457 | 0,067409 | 1 | 0,425749 |
| -1,70624 | 421 | 321,0485 | 0,08893 | 1 | 0,319007 |
| -1,64082 | 421 | 185,1371 | 0,102532 | 1 | 0,383322 |
| -0,28002 | 420 | 359,8406 | 0,779623 | 1 | 0,969427 |
| -1,62405 | 367 | 312,1372 | 0,105374 | 1 | 0,429907 |
| -1,16341 | 280 | 248,1266 | 0,24578 | 1 | 0,676332 |
| 0,578688 | 421 | 323,3547 | 0,563203 | 1 | 0,796197 |
| 2,08945 | 421 | 182,3577 | 0,038055 | 1 | 0,267984 |
| -0,33465 | 420 | 358,499 | 0,738086 | 1 | 0,969427 |
| 1,486307 | 367 | 308,6549 | 0,138219 | 1 | 0,484408 |
| 1,788445 | 280 | 258,8964 | 0,074873 | 1 | 0,447755 |
| -0,84968 | 421 | 360,004 | 0,396069 | 1 | 0,686065 |
| -1,24564 | 421 | 195,5003 | 0,214389 | 1 | 0,514565 |
| 0,346272 | 420 | 394,5267 | 0,729323 | 1 | 0,969427 |
| -1,06599 | 367 | 351,4225 | 0,287161 | 1 | 0,623561 |
| -0,68543 | 280 | 258,2894 | 0,493688 | 1 | 0,827962 |
| -1,6727 | 421 | 318,4072 | 0,095368 | 1 | 0,331916 |
| -1,89473 | 421 | 182,0867 | 0,059713 | 1 | 0,314727 |
| -1,90265 | 420 | 348,5499 | 0,057909 | 1 | 0,848246 |
| -0,72932 | 367 | 320,5617 | 0,466338 | 1 | 0,771623 |
| -1,41243 | 280 | 259,5882 | 0,159022 | 1 | 0,578854 |
| 1,022258 | 421 | 340,9489 | 0,307384 | 1 | 0,604218 |
| 0,983511 | 421 | 188,8085 | 0,326614 | 1 | 0,620691 |
| -0,23161 | 420 | 377,0743 | 0,816969 | 1 | 0,969427 |
| -0,39994 | 367 | 323,5064 | 0,689465 | 1 | 0,903032 |
| 0,428494 | 280 | 266,5982 | 0,668638 | 1 | 0,908644 |
| -2,49486 | 421 | 321,1813 | 0,013102 | 1 | 0,138079 |
| -2,43274 | 421 | 183,4668 | 0,015945 | 1 | 0,179127 |
| -0,80887 | 420 | 357,545 | 0,419127 | 1 | 0,95743 |
| -1,88802 | 367 | 326,0117 | 0,059911 | 1 | 0,340409 |
| -1,96901 | 280 | 266,1366 | 0,04999 | 1 | 0,39442 |
| -1,63236 | 421 | 318,8708 | 0,103591 | 1 | 0,345379 |
| -1,58781 | 421 | 183,0965 | 0,114054 | 1 | 0,391524 |
| -0,03853 | 420 | 354,7722 | 0,96929 | 1 | 0,997342 |
| -0,94971 | 367 | 313,29 | 0,342992 | 1 | 0,665683 |

| | | | | | |
|---|---|---|---|---|---|
| -2,31822 | 280 | 253,7659 | 0,021233 | 1 | 0,30229 |
| -1,71172 | 421 | 338,3697 | 0,087864 | 1 | 0,317534 |
| -0,15672 | 421 | 186,59 | 0,875632 | 1 | 0,953452 |
| -0,51056 | 420 | 373,2939 | 0,60996 | 1 | 0,964878 |
| -0,32812 | 367 | 328,8679 | 0,743028 | 1 | 0,923669 |
| 1,059226 | 280 | 253,2045 | 0,290506 | 1 | 0,720054 |
| 0,313388 | 421 | 324,7982 | 0,754187 | 1 | 0,898125 |
| 0,230964 | 421 | 181,7099 | 0,817603 | 1 | 0,926377 |
| -1,03421 | 420 | 355,1533 | 0,301743 | 1 | 0,943897 |
| -1,00645 | 367 | 313,5493 | 0,314975 | 1 | 0,647568 |
| -0,96053 | 280 | 259,5011 | 0,337681 | 1 | 0,755731 |
| 2,375303 | 421 | 359,5553 | 0,018058 | 1 | 0,156401 |
| 1,647058 | 421 | 194,3969 | 0,101162 | 1 | 0,383011 |
| 0,87544 | 420 | 392,5764 | 0,38187 | 1 | 0,95743 |
| -1,15933 | 367 | 343,5731 | 0,247126 | 1 | 0,600675 |
| 2,101036 | 280 | 261,0489 | 0,036597 | 1 | 0,34044 |
| 0,626488 | 421 | 402,4946 | 0,53135 | 1 | 0,776668 |
| -0,32582 | 421 | 218,9282 | 0,744868 | 1 | 0,888542 |
| -0,70323 | 420 | 415,6989 | 0,482304 | 1 | 0,960096 |
| -0,43534 | 367 | 362,8329 | 0,663574 | 1 | 0,894765 |
| -0,41553 | 280 | 272,0988 | 0,678079 | 1 | 0,909385 |
| -0,99222 | 421 | 332,2909 | 0,32181 | 1 | 0,615441 |
| -1,46406 | 421 | 186,096 | 0,144864 | 1 | 0,426157 |
| 0,70045 | 420 | 366,843 | 0,48409 | 1 | 0,960096 |
| -0,96787 | 367 | 331,0009 | 0,333816 | 1 | 0,661796 |
| -0,41619 | 280 | 261,0038 | 0,677616 | 1 | 0,909385 |
| -1,31962 | 421 | 373,1427 | 0,187772 | 1 | 0,472953 |
| -2,39594 | 421 | 198,8033 | 0,017505 | 1 | 0,182536 |
| -1,07329 | 420 | 402,1359 | 0,283786 | 1 | 0,943897 |
| -2,74337 | 367 | 348,2813 | 0,006396 | 1 | 0,178183 |
| -1,86028 | 280 | 268,733 | 0,063938 | 1 | 0,422605 |
| -0,84208 | 421 | 366,9098 | 0,400291 | 1 | 0,689415 |
| -1,34242 | 421 | 196,2058 | 0,181013 | 1 | 0,470641 |
| 0,405935 | 420 | 399,3875 | 0,685007 | 1 | 0,968613 |
| -1,65386 | 367 | 339,9432 | 0,099079 | 1 | 0,415911 |
| -1,18588 | 280 | 258,8139 | 0,236756 | 1 | 0,659852 |
| -0,40215 | 421 | 323,9427 | 0,687842 | 1 | 0,865979 |
| 0,407585 | 421 | 183,9853 | 0,684053 | 1 | 0,859821 |
| -0,01355 | 420 | 356,8221 | 0,989193 | 1 | 0,997342 |
| -1,71074 | 367 | 305,6033 | 0,088144 | 1 | 0,394165 |
| 1,481193 | 280 | 253,7702 | 0,139796 | 1 | 0,552945 |
| 0,211478 | 421 | 355,852 | 0,832635 | 1 | 0,936611 |
| 1,218693 | 421 | 192,2684 | 0,224454 | 1 | 0,521994 |
| 0,462802 | 420 | 388,7593 | 0,643765 | 1 | 0,964878 |
| 2,303223 | 367 | 338,9246 | 0,021871 | 1 | 0,230489 |
| -0,38557 | 280 | 264,8673 | 0,700126 | 1 | 0,92034 |
| -0,46512 | 421 | 409,4036 | 0,642096 | 1 | 0,835611 |
| -0,2949 | 421 | 226,7995 | 0,768339 | 1 | 0,905801 |

| | | | | | |
|---|---|---|---|---|---|
| 0,284916 | 420 | 415,682 | 0,77585 | 1 | 0,969427 |
| -0,89292 | 367 | 362,4629 | 0,37249 | 1 | 0,682069 |
| -0,62873 | 280 | 275,9965 | 0,530044 | 1 | 0,840949 |
| -0,19652 | 421 | 357,3876 | 0,844317 | 1 | 0,939735 |
| -0,75207 | 421 | 194,245 | 0,452919 | 1 | 0,71471 |
| 0,721604 | 420 | 390,1659 | 0,47097 | 1 | 0,960096 |
| -2,66461 | 367 | 348,7368 | 0,008066 | 1 | 0,181422 |
| -2,18308 | 280 | 269,8707 | 0,029892 | 1 | 0,328807 |
| -0,36535 | 421 | 313,5694 | 0,715093 | 1 | 0,881149 |
| 0,445062 | 421 | 181,0622 | 0,656807 | 1 | 0,841515 |
| -0,42673 | 420 | 348,7287 | 0,669836 | 1 | 0,96781 |
| -0,46484 | 367 | 305,8107 | 0,642378 | 1 | 0,882539 |
| 2,010098 | 280 | 249,0282 | 0,045499 | 1 | 0,387749 |
| -2,44553 | 421 | 330,5268 | 0,014985 | 1 | 0,147897 |
| -1,41803 | 421 | 184,9643 | 0,157863 | 1 | 0,43893 |
| -0,41727 | 420 | 368,4116 | 0,676724 | 1 | 0,96781 |
| 0,313076 | 367 | 322,1756 | 0,754426 | 1 | 0,92504 |
| -0,91251 | 280 | 246,984 | 0,362389 | 1 | 0,762031 |
| 0,720655 | 421 | 310,012 | 0,471665 | 1 | 0,731548 |
| 1,08517 | 421 | 178,9461 | 0,279306 | 1 | 0,578974 |
| -0,32786 | 420 | 343,0449 | 0,743215 | 1 | 0,969427 |
| -2,2033 | 367 | 308,9442 | 0,028312 | 1 | 0,257802 |
| 1,500053 | 280 | 248,7848 | 0,134869 | 1 | 0,544898 |
| -0,68782 | 421 | 331,1258 | 0,492051 | 1 | 0,748579 |
| -1,69961 | 421 | 185,3088 | 0,090882 | 1 | 0,373113 |
| 0,834571 | 420 | 364,0105 | 0,404507 | 1 | 0,95743 |
| -1,12769 | 367 | 334,1037 | 0,260258 | 1 | 0,609149 |
| 0,13066 | 280 | 261,9096 | 0,896144 | 1 | 0,975243 |
| 0,376313 | 421 | 367,8797 | 0,706901 | 1 | 0,875021 |
| 1,300459 | 421 | 197,9817 | 0,194956 | 1 | 0,483337 |
| -0,753 | 420 | 398,5073 | 0,451893 | 1 | 0,960096 |
| 1,51686 | 367 | 348,7579 | 0,130208 | 1 | 0,47397 |
| -0,17121 | 280 | 267,8193 | 0,864186 | 1 | 0,968274 |
| -0,48687 | 421 | 324,1637 | 0,626678 | 1 | 0,832808 |
| 0,973078 | 421 | 181,0788 | 0,331813 | 1 | 0,626331 |
| -0,75662 | 420 | 357,9055 | 0,449774 | 1 | 0,960096 |
| 0,756317 | 367 | 312,1434 | 0,45003 | 1 | 0,755345 |
| 1,187894 | 280 | 276 | 0,235896 | 1 | 0,659852 |
| 0,618086 | 421 | 367,2654 | 0,536902 | 1 | 0,778195 |
| 0,620274 | 421 | 196,8047 | 0,535795 | 1 | 0,770962 |
| 0,642188 | 420 | 397,6632 | 0,521121 | 1 | 0,960096 |
| 2,827545 | 367 | 347,931 | 0,004962 | 1 | 0,178183 |
| 1,088708 | 280 | 262,4767 | 0,277281 | 1 | 0,701156 |
| -1,84252 | 421 | 333,9214 | 0,066286 | 1 | 0,278382 |
| -2,25643 | 421 | 185,4448 | 0,025211 | 1 | 0,218351 |
| -1,45312 | 420 | 375,1536 | 0,147025 | 1 | 0,914005 |
| -1,14426 | 367 | 312,1015 | 0,253393 | 1 | 0,609031 |
| -1,48587 | 280 | 256,9764 | 0,138539 | 1 | 0,550865 |

| | | | | | |
|---|---|---|---|---|---|
| 0,529596 | 421 | 301,9574 | 0,596781 | 1 | 0,815122 |
| 0,837295 | 421 | 178,1524 | 0,403549 | 1 | 0,677774 |
| -1,86416 | 420 | 332,673 | 0,06318 | 1 | 0,848246 |
| 0,978006 | 367 | 307,3658 | 0,32884 | 1 | 0,656374 |
| -0,329 | 280 | 276 | 0,742403 | 1 | 0,94096 |
| -2,41731 | 421 | 320,7851 | 0,016193 | 1 | 0,147897 |
| -2,42771 | 421 | 184,0791 | 0,016157 | 1 | 0,179127 |
| 0,997229 | 420 | 363,4896 | 0,319317 | 1 | 0,945573 |
| -1,88722 | 367 | 329,5118 | 0,060009 | 1 | 0,340409 |
| -1,12508 | 280 | 258,9151 | 0,261598 | 1 | 0,687569 |
| -3,12277 | 421 | 326,4639 | 0,001952 | 1 | 0,069718 |
| -1,89977 | 421 | 185,36 | 0,059015 | 1 | 0,314262 |
| -0,73018 | 420 | 361,2446 | 0,465755 | 1 | 0,960096 |
| 0,322932 | 367 | 324,1215 | 0,746955 | 1 | 0,923669 |
| -2,13259 | 280 | 259,4661 | 0,033897 | 1 | 0,329568 |
| -0,09187 | 421 | 382,7967 | 0,92685 | 1 | 0,968629 |
| 1,530861 | 421 | 203,55 | 0,127356 | 1 | 0,410224 |
| 0,690607 | 420 | 407,5104 | 0,490206 | 1 | 0,960096 |
| -1,22858 | 367 | 359,4888 | 0,220034 | 1 | 0,572696 |
| -0,23638 | 280 | 270,0793 | 0,813315 | 1 | 0,95747 |
| -2,62452 | 421 | 344,7036 | 0,009063 | 1 | 0,113397 |
| -2,2628 | 421 | 186,3847 | 0,024802 | 1 | 0,218351 |
| -0,77577 | 420 | 376,662 | 0,43837 | 1 | 0,960096 |
| -0,77932 | 367 | 322,9821 | 0,436365 | 1 | 0,742744 |
| -3,03086 | 280 | 256,4497 | 0,002688 | 1 | 0,135787 |
| -0,1005 | 421 | 365,0038 | 0,920001 | 1 | 0,966828 |
| 0,819328 | 421 | 195,7369 | 0,413596 | 1 | 0,684933 |
| 1,225456 | 420 | 393,2492 | 0,221137 | 1 | 0,943055 |
| 0,646297 | 367 | 347,6258 | 0,518513 | 1 | 0,801766 |
| -0,00966 | 280 | 260,1511 | 0,992303 | 1 | 0,996269 |
| 1,503396 | 421 | 386,3545 | 0,133554 | 1 | 0,406597 |
| 1,304988 | 421 | 204,6189 | 0,193363 | 1 | 0,481649 |
| 1,491172 | 420 | 409,6761 | 0,136686 | 1 | 0,891179 |
| 1,122793 | 367 | 356,4507 | 0,262281 | 1 | 0,609149 |
| -0,52207 | 280 | 273,7215 | 0,602044 | 1 | 0,882569 |
| -1,55896 | 421 | 417 | 0,119765 | 1 | 0,381479 |
| -1,23244 | 421 | 417 | 0,218479 | 1 | 0,514565 |
| -0,63112 | 420 | 416 | 0,528308 | 1 | 0,960096 |
| -0,68944 | 367 | 283,1911 | 0,491111 | 1 | 0,792348 |
| -1,05505 | 280 | 276 | 0,292327 | 1 | 0,722191 |
| -0,90693 | 421 | 323,0142 | 0,365121 | 1 | 0,655826 |
| -0,38694 | 421 | 183,4845 | 0,69925 | 1 | 0,865528 |
| 2,054155 | 420 | 360,807 | 0,040681 | 1 | 0,846132 |
| 1,815338 | 367 | 318,0271 | 0,070414 | 1 | 0,36493 |
| 1,190582 | 280 | 251,377 | 0,234941 | 1 | 0,659852 |
| -0,8027 | 421 | 336,5846 | 0,422715 | 1 | 0,697735 |
| -1,37104 | 421 | 187,328 | 0,172002 | 1 | 0,456365 |
| -2,27848 | 420 | 366,2969 | 0,023273 | 1 | 0,75904 |

| | | | | | |
|---|---|---|---|---|---|
| -0,52477 | 367 | 325,8241 | 0,600097 | 1 | 0,862104 |
| -0,66422 | 280 | 261,7969 | 0,507135 | 1 | 0,831912 |
| -0,38201 | 421 | 346,1037 | 0,702685 | 1 | 0,874567 |
| -1,73564 | 421 | 187,8224 | 0,084267 | 1 | 0,365475 |
| -0,17624 | 420 | 381,5928 | 0,860201 | 1 | 0,976988 |
| 0,920822 | 367 | 337,7545 | 0,357801 | 1 | 0,674813 |
| 0,130065 | 280 | 262,8495 | 0,896614 | 1 | 0,975243 |
| 1,670649 | 421 | 345,0164 | 0,095698 | 1 | 0,332296 |
| 0,465191 | 421 | 191,2167 | 0,642324 | 1 | 0,838647 |
| -0,14622 | 420 | 382,804 | 0,883827 | 1 | 0,982463 |
| 1,747694 | 367 | 333,1962 | 0,081438 | 1 | 0,381841 |
| 0,904271 | 280 | 259,3109 | 0,366691 | 1 | 0,763914 |
| -1,41281 | 421 | 375,1203 | 0,15854 | 1 | 0,431264 |
| -1,37881 | 421 | 200,6188 | 0,169489 | 1 | 0,456365 |
| -0,26761 | 420 | 404,9385 | 0,789134 | 1 | 0,969427 |
| -3,01075 | 367 | 355,4673 | 0,002792 | 1 | 0,176492 |
| -2,98635 | 280 | 269,8125 | 0,003083 | 1 | 0,136636 |
| 3,13647 | 421 | 349,7123 | 0,001855 | 1 | 0,069718 |
| 1,316958 | 421 | 189,7112 | 0,189441 | 1 | 0,476746 |
| 0,9521 | 420 | 385,652 | 0,341643 | 1 | 0,947373 |
| -0,20925 | 367 | 333,7097 | 0,834379 | 1 | 0,956935 |
| 0,377181 | 280 | 258,3543 | 0,706349 | 1 | 0,922762 |
| -0,39245 | 421 | 318,7575 | 0,694985 | 1 | 0,870609 |
| -0,3735 | 421 | 178,072 | 0,709222 | 1 | 0,870869 |
| -0,9418 | 420 | 353,7145 | 0,346938 | 1 | 0,947373 |
| 1,34051 | 367 | 295,6478 | 0,181109 | 1 | 0,527769 |
| -3,14388 | 280 | 246,0309 | 0,001872 | 1 | 0,135787 |
| 2,830827 | 421 | 358,8424 | 0,004904 | 1 | 0,087964 |
| 2,9197 | 421 | 190,7183 | 0,003926 | 1 | 0,118336 |
| 1,657965 | 420 | 388,6881 | 0,098131 | 1 | 0,848246 |
| 1,62017 | 367 | 341,1268 | 0,10612 | 1 | 0,431058 |
| 1,588718 | 280 | 269,6216 | 0,113296 | 1 | 0,508687 |
| -1,87296 | 421 | 353,671 | 0,061898 | 1 | 0,271708 |
| -2,9195 | 421 | 191,5542 | 0,003926 | 1 | 0,118336 |
| -1,14094 | 420 | 389,5524 | 0,254594 | 1 | 0,943897 |
| -3,94941 | 367 | 341,3783 | 9,52E-05 | 0,102617 | 0,053579 |
| -3,86937 | 280 | 260,9652 | 1,38E-04 | 0,148664 | 0,046455 |
| 0,819369 | 421 | 350,3365 | 0,413133 | 1 | 0,695339 |
| 1,078272 | 421 | 188,9859 | 0,282287 | 1 | 0,581951 |
| -0,02208 | 420 | 384,1508 | 0,982397 | 1 | 0,997342 |
| 1,086491 | 367 | 330,1871 | 0,278054 | 1 | 0,612614 |
| 0,509712 | 280 | 260,6841 | 0,610684 | 1 | 0,888252 |
| -1,93204 | 421 | 341,7002 | 0,054181 | 1 | 0,259162 |
| -2,84503 | 421 | 189,7 | 0,004928 | 1 | 0,137431 |
| -1,07326 | 420 | 382,1213 | 0,283833 | 1 | 0,943897 |
| -2,5615 | 367 | 343,6309 | 0,010848 | 1 | 0,189546 |
| -0,27811 | 280 | 265,5697 | 0,781142 | 1 | 0,951006 |
| -0,19644 | 421 | 340,2744 | 0,844381 | 1 | 0,939735 |

| | | | | | |
|---|---|---|---|---|---|
| -1,07712 | 421 | 185,7399 | 0,282824 | 1 | 0,582262 |
| -1,79315 | 420 | 375,0219 | 0,073755 | 1 | 0,848246 |
| -1,12487 | 367 | 315,1952 | 0,261499 | 1 | 0,609149 |
| -1,82641 | 280 | 252,9107 | 0,068968 | 1 | 0,429481 |
| -2,42158 | 421 | 279,4383 | 0,01609 | 1 | 0,147897 |
| -0,58919 | 421 | 171,5692 | 0,556507 | 1 | 0,778696 |
| 0,832472 | 420 | 313,8818 | 0,405775 | 1 | 0,95743 |
| 0,836324 | 367 | 277,3579 | 0,403692 | 1 | 0,716566 |
| 1,737018 | 280 | 245,9324 | 0,083636 | 1 | 0,456738 |
| 1,151216 | 421 | 294,4045 | 0,250577 | 1 | 0,546261 |
| 3,416893 | 421 | 175,2191 | 7,87E-04 | 0,840194 | 0,065272 |
| -1,47194 | 420 | 326,1456 | 0,142 | 1 | 0,90372 |
| 0,656943 | 367 | 296,401 | 0,511727 | 1 | 0,797829 |
| 1,999487 | 280 | 276 | 0,046536 | 1 | 0,387749 |
| -0,36015 | 421 | 344,8103 | 0,718952 | 1 | 0,884157 |
| 0,587443 | 421 | 189,5389 | 0,557605 | 1 | 0,779138 |
| 0,286841 | 420 | 379,1988 | 0,774391 | 1 | 0,969427 |
| 1,030418 | 367 | 329,4693 | 0,30357 | 1 | 0,636272 |
| 0,677818 | 280 | 252,6134 | 0,498508 | 1 | 0,83103 |
| -3,61478 | 421 | 318,5376 | 3,49E-04 | 0,373728 | 0,04642 |
| -1,66475 | 421 | 184,0682 | 0,097664 | 1 | 0,379251 |
| 0,055439 | 420 | 361,283 | 0,955819 | 1 | 0,995923 |
| -0,96816 | 367 | 316,9403 | 0,333703 | 1 | 0,661796 |
| -0,80637 | 280 | 259,0303 | 0,420767 | 1 | 0,796701 |
| 0,733449 | 421 | 322,5398 | 0,463817 | 1 | 0,727964 |
| -0,45756 | 421 | 182,4174 | 0,647814 | 1 | 0,839358 |
| -0,84899 | 420 | 355,0611 | 0,396459 | 1 | 0,95743 |
| 1,493995 | 367 | 323,1178 | 0,136153 | 1 | 0,479589 |
| 0,692082 | 280 | 259,5249 | 0,489505 | 1 | 0,826999 |
| -1,83188 | 421 | 370,0954 | 0,067773 | 1 | 0,280051 |
| -1,6877 | 421 | 197,7994 | 0,093046 | 1 | 0,373113 |
| 1,270197 | 420 | 399,5515 | 0,204754 | 1 | 0,928589 |
| -1,49848 | 367 | 347,4335 | 0,134917 | 1 | 0,477542 |
| -2,73515 | 280 | 269,3672 | 0,006649 | 1 | 0,192695 |
| -1,43182 | 421 | 365,4 | 0,153049 | 1 | 0,42476 |
| 0,309091 | 421 | 197,3956 | 0,757578 | 1 | 0,898246 |
| -0,22315 | 420 | 398,1452 | 0,823534 | 1 | 0,972623 |
| -1,30206 | 367 | 344,7814 | 0,193766 | 1 | 0,547402 |
| -0,82177 | 280 | 266,9322 | 0,411943 | 1 | 0,788151 |
| 1,476219 | 421 | 386,1244 | 0,1407 | 1 | 0,410878 |
| 1,276603 | 421 | 207,1804 | 0,203172 | 1 | 0,497853 |
| 0,155193 | 420 | 410,1018 | 0,876745 | 1 | 0,982463 |
| 0,335749 | 367 | 360,4973 | 0,737255 | 1 | 0,923669 |
| 0,02671 | 280 | 272,7211 | 0,978711 | 1 | 0,99236 |
| -0,7129 | 421 | 297,7232 | 0,476468 | 1 | 0,734941 |
| -1,56543 | 421 | 174,6099 | 0,119294 | 1 | 0,396436 |
| 0,531895 | 420 | 330,8111 | 0,595156 | 1 | 0,964878 |
| -2,83744 | 367 | 309,7026 | 0,004848 | 1 | 0,178183 |

| | | | | | |
|---|---|---|---|---|---|
| -1,81974 | 280 | 251,2013 | 0,069989 | 1 | 0,432271 |
| -2,83866 | 421 | 309,206 | 0,00483 | 1 | 0,087964 |
| -1,79625 | 421 | 176,9731 | 0,074161 | 1 | 0,344631 |
| 0,318201 | 420 | 342,0608 | 0,750526 | 1 | 0,969427 |
| -2,46768 | 367 | 305,8839 | 0,014146 | 1 | 0,201413 |
| -0,80041 | 280 | 248,779 | 0,424235 | 1 | 0,797636 |
| -0,68845 | 421 | 316,3922 | 0,491676 | 1 | 0,748579 |
| -0,98851 | 421 | 181,5591 | 0,32422 | 1 | 0,619085 |
| -1,27431 | 420 | 350,5728 | 0,203397 | 1 | 0,928589 |
| -1,64704 | 367 | 304,1144 | 0,100583 | 1 | 0,42105 |
| -0,11746 | 280 | 257,3637 | 0,906588 | 1 | 0,975308 |
| -2,20387 | 421 | 332,7127 | 0,028218 | 1 | 0,19529 |
| -3,63941 | 421 | 186,1064 | 3,54E-04 | 0,378683 | 0,038096 |
| -0,80269 | 420 | 371,8112 | 0,422664 | 1 | 0,95743 |
| -1,95445 | 367 | 332,2506 | 0,051486 | 1 | 0,32195 |
| -2,16883 | 280 | 255,7904 | 0,031017 | 1 | 0,329568 |
| -0,95025 | 421 | 310,5259 | 0,342723 | 1 | 0,63164 |
| -0,39133 | 421 | 177,1927 | 0,696023 | 1 | 0,865526 |
| 0,41916 | 420 | 343,5978 | 0,675361 | 1 | 0,96781 |
| 0,376087 | 367 | 309,1963 | 0,70711 | 1 | 0,91071 |
| -0,91923 | 280 | 245,7744 | 0,358878 | 1 | 0,761846 |
| -1,14336 | 421 | 369,4957 | 0,253628 | 1 | 0,546261 |
| -1,59923 | 421 | 197,6717 | 0,111367 | 1 | 0,391524 |
| 0,025644 | 420 | 398,8749 | 0,979554 | 1 | 0,997342 |
| -2,45559 | 367 | 355,1585 | 0,014544 | 1 | 0,204372 |
| -1,8718 | 280 | 271,39 | 0,06231 | 1 | 0,422605 |
| -1,4345 | 421 | 331,0548 | 0,152374 | 1 | 0,42445 |
| -0,39375 | 421 | 182,9819 | 0,694223 | 1 | 0,865526 |
| -0,48556 | 420 | 363,6066 | 0,627573 | 1 | 0,964878 |
| -0,43032 | 367 | 307,4014 | 0,667264 | 1 | 0,894765 |
| -0,25396 | 280 | 259,8997 | 0,799728 | 1 | 0,954985 |
| 1,849456 | 421 | 377,7584 | 0,065173 | 1 | 0,276236 |
| 1,332861 | 421 | 199,6916 | 0,184096 | 1 | 0,473593 |
| 1,711076 | 420 | 405,9387 | 0,087831 | 1 | 0,848246 |
| 0,070812 | 367 | 352,8454 | 0,943587 | 1 | 0,979539 |
| -0,59902 | 280 | 267,2306 | 0,549665 | 1 | 0,852208 |
| -0,64691 | 421 | 348,8799 | 0,518115 | 1 | 0,768119 |
| -1,3907 | 421 | 189,0606 | 0,16595 | 1 | 0,453055 |
| -0,98546 | 420 | 382,3141 | 0,325021 | 1 | 0,945573 |
| -0,0906 | 367 | 331,7287 | 0,927865 | 1 | 0,976733 |
| 0,351922 | 280 | 264,0023 | 0,725177 | 1 | 0,936493 |
| 3,157493 | 421 | 303,0739 | 0,001752 | 1 | 0,069718 |
| 3,986601 | 421 | 173,5941 | 9,86E-05 | 0,105979 | 0,021224 |
| 1,82159 | 420 | 339,3723 | 0,069397 | 1 | 0,848246 |
| 1,504513 | 367 | 286,6263 | 0,133551 | 1 | 0,477542 |
| 2,416704 | 280 | 276 | 0,016311 | 1 | 0,266517 |
| 1,24994 | 421 | 315,4375 | 0,212248 | 1 | 0,502921 |
| 2,605084 | 421 | 174,3588 | 0,009979 | 1 | 0,150377 |

| | | | | | |
|---|---|---|---|---|---|
| 1,923362 | 420 | 353,0493 | 0,055239 | 1 | 0,848246 |
| 0,878495 | 367 | 301,9481 | 0,380374 | 1 | 0,691076 |
| 2,585688 | 280 | 248,9666 | 0,010288 | 1 | 0,229419 |
| -0,34679 | 421 | 297,478 | 0,728993 | 1 | 0,890992 |
| 0,564848 | 421 | 176,0082 | 0,572897 | 1 | 0,792796 |
| 0,029504 | 420 | 328,8258 | 0,97648 | 1 | 0,997342 |
| 0,688644 | 367 | 287,696 | 0,491603 | 1 | 0,792348 |
| 0,132824 | 280 | 246,8869 | 0,894441 | 1 | 0,975243 |
| 0,777576 | 421 | 338,8497 | 0,437362 | 1 | 0,707193 |
| 1,481011 | 421 | 186,8909 | 0,140288 | 1 | 0,423351 |
| -0,65022 | 420 | 372,1548 | 0,515948 | 1 | 0,960096 |
| 0,358039 | 367 | 324,9923 | 0,720547 | 1 | 0,914797 |
| 0,893851 | 280 | 252,5624 | 0,372253 | 1 | 0,763914 |
| -0,12149 | 421 | 356,7697 | 0,90337 | 1 | 0,965517 |
| -0,4909 | 421 | 193,687 | 0,624053 | 1 | 0,830785 |
| 0,698649 | 420 | 390,8116 | 0,485187 | 1 | 0,960096 |
| 0,121632 | 367 | 343,3179 | 0,903262 | 1 | 0,970486 |
| -0,38952 | 280 | 256,4036 | 0,697216 | 1 | 0,92034 |
| -2,05563 | 421 | 337,072 | 0,040588 | 1 | 0,226301 |
| -1,37139 | 421 | 188,047 | 0,171886 | 1 | 0,456365 |
| 0,312302 | 420 | 375,8693 | 0,754984 | 1 | 0,969427 |
| -1,15276 | 367 | 321,4095 | 0,249865 | 1 | 0,604409 |
| -1,41945 | 280 | 257,3723 | 0,156979 | 1 | 0,575588 |
| -2,04311 | 421 | 393,6028 | 0,041707 | 1 | 0,228553 |
| -2,63391 | 421 | 210,433 | 0,009068 | 1 | 0,14815 |
| 0,231676 | 420 | 413,9775 | 0,816904 | 1 | 0,969427 |
| -3,82453 | 367 | 362,867 | 1,54E-04 | 0,165916 | 0,053579 |
| -3,6329 | 280 | 275,8427 | 3,34E-04 | 0,358847 | 0,06294 |
| -1,49294 | 421 | 367,6288 | 0,13631 | 1 | 0,407576 |
| -1,76534 | 421 | 197,9025 | 0,079048 | 1 | 0,355599 |
| 0,184743 | 420 | 401,8197 | 0,853524 | 1 | 0,973894 |
| -2,50116 | 367 | 354,4778 | 0,012829 | 1 | 0,189546 |
| -1,64635 | 280 | 273,2807 | 0,10084 | 1 | 0,480557 |
| 0,043306 | 421 | 333,3528 | 0,965483 | 1 | 0,992572 |
| -1,16405 | 421 | 183,1662 | 0,24592 | 1 | 0,546224 |
| -0,6852 | 420 | 369,1802 | 0,493645 | 1 | 0,960096 |
| 1,917739 | 367 | 323,7002 | 0,056024 | 1 | 0,337523 |
| -0,18097 | 280 | 268,3374 | 0,856531 | 1 | 0,966655 |
| -2,83032 | 421 | 347,0363 | 0,004921 | 1 | 0,087964 |
| -3,03172 | 421 | 191,0084 | 0,002769 | 1 | 0,114447 |
| -1,48614 | 420 | 385,9621 | 0,138059 | 1 | 0,891179 |
| -2,60101 | 367 | 335,9949 | 0,009706 | 1 | 0,183886 |
| -1,72921 | 280 | 267,6715 | 0,084924 | 1 | 0,45831 |
| 0,077166 | 421 | 353,322 | 0,938535 | 1 | 0,97543 |
| 2,660864 | 421 | 192,5582 | 0,008452 | 1 | 0,148102 |
| -0,89782 | 420 | 388,8481 | 0,369838 | 1 | 0,95743 |
| -0,98445 | 367 | 331,3493 | 0,325611 | 1 | 0,655597 |
| -0,31455 | 280 | 263,3819 | 0,753353 | 1 | 0,942071 |

| | | | | | |
|---|---|---|---|---|---|
| 2,52588 | 421 | 370,4558 | 0,011957 | 1 | 0,131857 |
| 2,190993 | 421 | 198,2817 | 0,029618 | 1 | 0,236159 |
| -0,23365 | 420 | 403,0765 | 0,815371 | 1 | 0,969427 |
| 1,421168 | 367 | 346,4986 | 0,156167 | 1 | 0,502197 |
| 0,537957 | 280 | 263,9155 | 0,59106 | 1 | 0,8767 |
| -1,36772 | 421 | 397,0033 | 0,172174 | 1 | 0,444059 |
| -1,1277 | 421 | 212,6171 | 0,260719 | 1 | 0,557979 |
| -1,19055 | 420 | 414,0841 | 0,234511 | 1 | 0,943055 |
| -0,57051 | 367 | 361,5091 | 0,568684 | 1 | 0,837838 |
| -0,12688 | 280 | 272,8679 | 0,899131 | 1 | 0,975243 |
| -2,17353 | 421 | 344,6354 | 0,030421 | 1 | 0,200565 |
| -2,0951 | 421 | 191,1915 | 0,037479 | 1 | 0,266419 |
| 0,097539 | 420 | 385,2307 | 0,922349 | 1 | 0,989204 |
| -1,92893 | 367 | 343,0807 | 0,054564 | 1 | 0,335053 |
| -1,14904 | 280 | 268,2209 | 0,251565 | 1 | 0,679007 |
| -1,39989 | 421 | 308,3024 | 0,162553 | 1 | 0,436662 |
| 0,561022 | 421 | 178,7227 | 0,575485 | 1 | 0,794719 |
| 1,036318 | 420 | 343,0789 | 0,300784 | 1 | 0,943897 |
| 1,603372 | 367 | 294,5728 | 0,109924 | 1 | 0,435936 |
| 0,868683 | 280 | 262,6821 | 0,385813 | 1 | 0,773166 |
| 0,493081 | 421 | 342,0149 | 0,622272 | 1 | 0,828413 |
| 1,998569 | 421 | 188,7997 | 0,04709 | 1 | 0,284997 |
| -0,20377 | 420 | 380,7065 | 0,838645 | 1 | 0,973894 |
| 0,473138 | 367 | 334,3866 | 0,636424 | 1 | 0,880707 |
| 0,453982 | 280 | 251,9935 | 0,650233 | 1 | 0,908644 |
| -1,02176 | 421 | 299,1653 | 0,307719 | 1 | 0,604218 |
| 0,133803 | 421 | 178,0153 | 0,893709 | 1 | 0,958882 |
| -0,61909 | 420 | 331,7532 | 0,536279 | 1 | 0,960096 |
| 0,186374 | 367 | 302,3193 | 0,852277 | 1 | 0,956935 |
| -0,79813 | 280 | 252,6953 | 0,425547 | 1 | 0,797636 |
| -2,35125 | 421 | 339,1889 | 0,019282 | 1 | 0,159771 |
| -2,16907 | 421 | 184,0264 | 0,03136 | 1 | 0,241423 |
| -1,69132 | 420 | 372,1633 | 0,091614 | 1 | 0,848246 |
| -0,48031 | 367 | 319,693 | 0,631338 | 1 | 0,880707 |
| -2,0441 | 280 | 251,7894 | 0,041984 | 1 | 0,374382 |
| 0,178164 | 421 | 363,5919 | 0,858693 | 1 | 0,949789 |
| -0,75111 | 421 | 196,3262 | 0,453486 | 1 | 0,714856 |
| 0,636561 | 420 | 394,944 | 0,524779 | 1 | 0,960096 |
| -2,1621 | 367 | 344,1308 | 0,031299 | 1 | 0,266905 |
| -1,80109 | 280 | 260,7785 | 0,072844 | 1 | 0,44087 |
| 2,390153 | 421 | 365,5677 | 0,017347 | 1 | 0,153976 |
| 2,021274 | 421 | 193,7573 | 0,044627 | 1 | 0,279059 |
| 1,03627 | 420 | 398,5347 | 0,300705 | 1 | 0,943897 |
| 1,92514 | 367 | 344,733 | 0,055034 | 1 | 0,335053 |
| 2,921402 | 280 | 272,2313 | 0,003777 | 1 | 0,147267 |
| -1,93258 | 421 | 324,5681 | 0,054158 | 1 | 0,259162 |
| 0,462774 | 421 | 184,5241 | 0,644072 | 1 | 0,838647 |
| 0,186027 | 420 | 358,7072 | 0,852529 | 1 | 0,973894 |

| | | | | | |
|---|---|---|---|---|---|
| 0,413668 | 367 | 316,6939 | 0,679397 | 1 | 0,899694 |
| -0,52646 | 280 | 270,5825 | 0,598997 | 1 | 0,881231 |
| -0,81417 | 421 | 351,6625 | 0,416099 | 1 | 0,695339 |
| -1,47193 | 421 | 189,9476 | 0,142695 | 1 | 0,423993 |
| -1,54075 | 420 | 385,7334 | 0,124198 | 1 | 0,87461 |
| -1,92116 | 367 | 340,5723 | 0,055546 | 1 | 0,336177 |
| -0,64124 | 280 | 261,1722 | 0,521931 | 1 | 0,833721 |
| -1,83526 | 421 | 341,1048 | 0,067337 | 1 | 0,279551 |
| -1,64896 | 421 | 187,8157 | 0,100828 | 1 | 0,383011 |
| 0,324786 | 420 | 375,697 | 0,745524 | 1 | 0,969427 |
| 0,165765 | 367 | 336,2388 | 0,868441 | 1 | 0,959978 |
| -2,69026 | 280 | 249,435 | 0,007622 | 1 | 0,199202 |
| -1,37044 | 421 | 340,7435 | 0,171452 | 1 | 0,443949 |
| 0,170025 | 421 | 187,9446 | 0,865173 | 1 | 0,947215 |
| -0,01491 | 420 | 375,7791 | 0,988115 | 1 | 0,997342 |
| -0,6251 | 367 | 324,4585 | 0,532343 | 1 | 0,814457 |
| 0,194304 | 280 | 265,5515 | 0,846086 | 1 | 0,965181 |
| -0,60462 | 421 | 330,2331 | 0,545847 | 1 | 0,784719 |
| 0,619042 | 421 | 183,8363 | 0,536655 | 1 | 0,770962 |
| 1,230447 | 420 | 363,0291 | 0,219326 | 1 | 0,943055 |
| 0,912338 | 367 | 319,112 | 0,36228 | 1 | 0,677287 |
| 0,758812 | 280 | 263,159 | 0,448644 | 1 | 0,813228 |
| 1,242346 | 421 | 385,2484 | 0,214865 | 1 | 0,506731 |
| 1,255644 | 421 | 205,2037 | 0,210673 | 1 | 0,509968 |
| 0,338467 | 420 | 410,0009 | 0,735185 | 1 | 0,969427 |
| 2,502283 | 367 | 352,3091 | 0,012792 | 1 | 0,189546 |
| 0,233111 | 280 | 271,4693 | 0,815851 | 1 | 0,95747 |
| 2,704879 | 421 | 379,9553 | 0,00714 | 1 | 0,105763 |
| 2,542918 | 421 | 204,2296 | 0,011734 | 1 | 0,165261 |
| -1,12701 | 420 | 410,6461 | 0,260398 | 1 | 0,943897 |
| 1,407882 | 367 | 355,8247 | 0,160039 | 1 | 0,504558 |
| 0,663477 | 280 | 273,2936 | 0,507584 | 1 | 0,831912 |
| -2,20796 | 421 | 365,8455 | 0,027867 | 1 | 0,19529 |
| -1,37254 | 421 | 195,246 | 0,17147 | 1 | 0,456365 |
| -0,2955 | 420 | 397,009 | 0,767764 | 1 | 0,969427 |
| -0,9554 | 367 | 346,2988 | 0,340042 | 1 | 0,664831 |
| -0,42765 | 280 | 259,0374 | 0,669263 | 1 | 0,908644 |
| 0,487919 | 421 | 326,3473 | 0,625935 | 1 | 0,832554 |
| -0,19335 | 421 | 184,2414 | 0,846898 | 1 | 0,943293 |
| -0,21759 | 420 | 360,9658 | 0,82787 | 1 | 0,973894 |
| -1,39401 | 367 | 317,5757 | 0,164289 | 1 | 0,510483 |
| -1,65737 | 280 | 252,3712 | 0,098688 | 1 | 0,480557 |
| -1,54976 | 421 | 329,3391 | 0,122159 | 1 | 0,383489 |
| -1,5681 | 421 | 186,8358 | 0,118551 | 1 | 0,396436 |
| -1,76034 | 420 | 371,0984 | 0,079173 | 1 | 0,848246 |
| -0,66863 | 367 | 321,5754 | 0,504214 | 1 | 0,793601 |
| -0,0453 | 280 | 266,1343 | 0,963899 | 1 | 0,989749 |
| -1,60308 | 421 | 352,1827 | 0,109814 | 1 | 0,35666 |

| | | | | | |
|---|---|---|---|---|---|
| -1,71434 | 421 | 192,9852 | 0,088071 | 1 | 0,368676 |
| -0,9678 | 420 | 387,0991 | 0,333748 | 1 | 0,947373 |
| -2,06454 | 367 | 333,4073 | 0,03974 | 1 | 0,28956 |
| -0,22001 | 280 | 258,3268 | 0,826039 | 1 | 0,96121 |
| -0,80606 | 421 | 370,856 | 0,420724 | 1 | 0,697735 |
| -0,66426 | 421 | 198,8657 | 0,507292 | 1 | 0,754678 |
| 0,475521 | 420 | 402,1602 | 0,634674 | 1 | 0,964878 |
| 0,127059 | 367 | 350,1052 | 0,898967 | 1 | 0,970486 |
| -0,29626 | 280 | 263,7521 | 0,767265 | 1 | 0,945436 |
| 1,055552 | 421 | 406,0371 | 0,291801 | 1 | 0,589468 |
| 1,235726 | 421 | 221,4694 | 0,21787 | 1 | 0,514565 |
| -0,87508 | 420 | 415,9932 | 0,382035 | 1 | 0,95743 |
| 0,446296 | 367 | 362,9171 | 0,655649 | 1 | 0,890147 |
| 1,924581 | 280 | 273,3819 | 0,055319 | 1 | 0,40654 |
| -2,28795 | 421 | 324,5218 | 0,022783 | 1 | 0,177419 |
| -1,56553 | 421 | 184,7853 | 0,11917 | 1 | 0,396436 |
| -0,48558 | 420 | 367,8543 | 0,627551 | 1 | 0,964878 |
| -2,1258 | 367 | 324,6972 | 0,034274 | 1 | 0,279197 |
| -1,31522 | 280 | 257,1582 | 0,189608 | 1 | 0,625017 |
| -0,2174 | 421 | 382,5044 | 0,828014 | 1 | 0,936611 |
| 0,169745 | 421 | 203,9768 | 0,865379 | 1 | 0,947215 |
| -0,43625 | 420 | 407,4436 | 0,662886 | 1 | 0,96781 |
| -0,626 | 367 | 356,7739 | 0,531712 | 1 | 0,814457 |
| 0,872762 | 280 | 270,0102 | 0,383568 | 1 | 0,772778 |
| -1,48232 | 421 | 370,5601 | 0,139106 | 1 | 0,4106 |
| -1,12454 | 421 | 198,4352 | 0,262143 | 1 | 0,557979 |
| -0,68156 | 420 | 401,5117 | 0,49591 | 1 | 0,960096 |
| -1,72174 | 367 | 343,7475 | 0,086016 | 1 | 0,390442 |
| 0,775282 | 280 | 261,7586 | 0,438873 | 1 | 0,808534 |
| -1,61251 | 421 | 332,2817 | 0,1078 | 1 | 0,353931 |
| -1,13792 | 421 | 183,7655 | 0,256634 | 1 | 0,556025 |
| -0,93996 | 420 | 368,5727 | 0,347851 | 1 | 0,947373 |
| -0,60754 | 367 | 323,6895 | 0,543915 | 1 | 0,824628 |
| -0,98027 | 280 | 252,4947 | 0,327892 | 1 | 0,752105 |
| -2,57868 | 421 | 328,9734 | 0,010352 | 1 | 0,120936 |
| -1,73178 | 421 | 184,8265 | 0,084982 | 1 | 0,365475 |
| -1,5083 | 420 | 368,317 | 0,132335 | 1 | 0,888479 |
| -1,96929 | 367 | 316,1629 | 0,049793 | 1 | 0,315285 |
| -1,13917 | 280 | 263,2284 | 0,25567 | 1 | 0,679673 |
| 0,312998 | 421 | 304,4939 | 0,754497 | 1 | 0,898125 |
| 1,176804 | 421 | 179,1003 | 0,240835 | 1 | 0,5417 |
| 0,340504 | 420 | 337,8843 | 0,733688 | 1 | 0,969427 |
| -0,44708 | 367 | 300,7289 | 0,655142 | 1 | 0,890147 |
| 0,103658 | 280 | 276 | 0,917516 | 1 | 0,977051 |
| 1,077046 | 421 | 363,571 | 0,282174 | 1 | 0,581718 |
| 0,306291 | 421 | 196,0871 | 0,759708 | 1 | 0,899424 |
| 0,331435 | 420 | 394,7194 | 0,740492 | 1 | 0,969427 |
| -0,53668 | 367 | 338,2129 | 0,591844 | 1 | 0,855959 |

| | | | | | |
|---|---|---|---|---|---|
| 0,642557 | 280 | 267,2102 | 0,521062 | 1 | 0,833721 |
| -2,2076 | 421 | 356,9542 | 0,027908 | 1 | 0,19529 |
| -2,4016 | 421 | 193,6414 | 0,017269 | 1 | 0,181988 |
| -1,44462 | 420 | 389,8563 | 0,149367 | 1 | 0,914005 |
| -2,3323 | 367 | 347,2841 | 0,020256 | 1 | 0,224016 |
| -1,21733 | 280 | 270,1011 | 0,224543 | 1 | 0,654014 |
| -3,38948 | 421 | 328,8874 | 7,85E-04 | 0,834891 | 0,053127 |
| -0,81523 | 421 | 184,958 | 0,415988 | 1 | 0,686888 |
| -0,81295 | 420 | 361,7333 | 0,41678 | 1 | 0,95743 |
| -0,3511 | 367 | 322,1168 | 0,725741 | 1 | 0,919068 |
| -1,0516 | 280 | 262,6315 | 0,293948 | 1 | 0,723324 |
| -0,16581 | 421 | 298,7658 | 0,868423 | 1 | 0,954568 |
| -0,15829 | 421 | 178,0556 | 0,874404 | 1 | 0,952803 |
| -0,35033 | 420 | 327,2023 | 0,726313 | 1 | 0,969427 |
| -0,10343 | 367 | 296,2925 | 0,91769 | 1 | 0,973915 |
| -0,65037 | 280 | 256,6589 | 0,516038 | 1 | 0,83211 |
| 0,271233 | 421 | 379,7695 | 0,786359 | 1 | 0,917552 |
| 1,18037 | 421 | 203,1832 | 0,239233 | 1 | 0,539707 |
| 1,135573 | 420 | 406,9419 | 0,256804 | 1 | 0,943897 |
| 1,271348 | 367 | 344,7605 | 0,204462 | 1 | 0,558434 |
| -0,89843 | 280 | 269,1909 | 0,369758 | 1 | 0,763914 |
| -2,64879 | 421 | 318,3992 | 0,008481 | 1 | 0,112451 |
| -0,92862 | 421 | 179,7624 | 0,354329 | 1 | 0,643758 |
| -1,38669 | 420 | 345,7572 | 0,16643 | 1 | 0,914005 |
| -1,53776 | 367 | 331,8663 | 0,125059 | 1 | 0,462104 |
| -0,23507 | 280 | 268,7875 | 0,814337 | 1 | 0,95747 |
| -1,50487 | 421 | 328,6952 | 0,133319 | 1 | 0,406597 |
| 0,051071 | 421 | 184,0634 | 0,959324 | 1 | 0,98414 |
| -0,18505 | 420 | 366,5594 | 0,853293 | 1 | 0,973894 |
| -1,26713 | 367 | 319,1893 | 0,206031 | 1 | 0,558434 |
| -0,07126 | 280 | 252,3108 | 0,943246 | 1 | 0,982175 |
| -2,8188 | 421 | 304,0327 | 0,005137 | 1 | 0,087964 |
| -0,88715 | 421 | 175,1928 | 0,376216 | 1 | 0,654686 |
| -1,28362 | 420 | 332,524 | 0,200168 | 1 | 0,928589 |
| -0,96122 | 367 | 289,5257 | 0,337244 | 1 | 0,662616 |
| -0,69411 | 280 | 276 | 0,488196 | 1 | 0,826999 |
| -1,67666 | 421 | 357,278 | 0,094483 | 1 | 0,330363 |
| -1,17763 | 421 | 192,5397 | 0,240397 | 1 | 0,541523 |
| -0,95832 | 420 | 390,6535 | 0,338496 | 1 | 0,947373 |
| -1,51914 | 367 | 344,8206 | 0,129644 | 1 | 0,473621 |
| -1,50696 | 280 | 267,9769 | 0,132998 | 1 | 0,543276 |
| 2,063454 | 421 | 347,5333 | 0,039812 | 1 | 0,224527 |
| 3,697873 | 421 | 189,3841 | 2,85E-04 | 0,305187 | 0,035752 |
| 0,131632 | 420 | 382,6881 | 0,895345 | 1 | 0,98503 |
| 0,088827 | 367 | 326,7499 | 0,929274 | 1 | 0,976733 |
| 0,230043 | 280 | 257,0787 | 0,818241 | 1 | 0,95778 |
| -0,24622 | 421 | 352,5275 | 0,805656 | 1 | 0,922616 |
| -0,02614 | 421 | 191,5255 | 0,979174 | 1 | 0,992344 |

| | | | | | |
|---|---|---|---|---|---|
| 0,339584 | 420 | 386,0545 | 0,734354 | 1 | 0,969427 |
| 1,569531 | 367 | 343,8313 | 0,117444 | 1 | 0,445308 |
| -0,26173 | 280 | 261,8226 | 0,793736 | 1 | 0,954269 |
| -2,59143 | 421 | 307,3621 | 0,010014 | 1 | 0,117898 |
| -0,82081 | 421 | 174,7247 | 0,412874 | 1 | 0,684933 |
| 0,795007 | 420 | 337,7918 | 0,427167 | 1 | 0,95743 |
| 0,507974 | 367 | 283,2266 | 0,611867 | 1 | 0,871158 |
| -2,00369 | 280 | 251,0358 | 0,046178 | 1 | 0,387749 |
| 0,367442 | 421 | 395,1869 | 0,713486 | 1 | 0,881149 |
| 1,025759 | 421 | 210,8341 | 0,30618 | 1 | 0,603743 |
| 0,780164 | 420 | 414,0113 | 0,43574 | 1 | 0,960096 |
| 2,265093 | 367 | 357,0774 | 0,024106 | 1 | 0,245454 |
| 0,732566 | 280 | 274,9309 | 0,464447 | 1 | 0,824256 |
| -1,00398 | 421 | 337,9406 | 0,316107 | 1 | 0,609953 |
| -1,19064 | 421 | 181,584 | 0,235348 | 1 | 0,535889 |
| 0,554702 | 420 | 376,1105 | 0,579428 | 1 | 0,964878 |
| -0,49402 | 367 | 329,9276 | 0,621619 | 1 | 0,875495 |
| 1,539913 | 280 | 264,2012 | 0,124778 | 1 | 0,535729 |
| -1,52943 | 421 | 307,5539 | 0,127186 | 1 | 0,392764 |
| -2,56713 | 421 | 180,4428 | 0,011065 | 1 | 0,161895 |
| -1,17274 | 420 | 345,7413 | 0,241707 | 1 | 0,943659 |
| -2,12972 | 367 | 302,9926 | 0,034 | 1 | 0,279197 |
| -1,45795 | 280 | 258,6428 | 0,146066 | 1 | 0,555466 |
| -2,3502 | 421 | 364,7326 | 0,019295 | 1 | 0,159771 |
| -2,47808 | 421 | 195,9251 | 0,014055 | 1 | 0,173612 |
| -2,13995 | 420 | 397,864 | 0,032966 | 1 | 0,828006 |
| -1,42264 | 367 | 347,9718 | 0,155735 | 1 | 0,502197 |
| -0,68588 | 280 | 271,1955 | 0,493374 | 1 | 0,827962 |
| 0,59227 | 421 | 348,5409 | 0,554054 | 1 | 0,790572 |
| 1,511267 | 421 | 189,5888 | 0,132385 | 1 | 0,414338 |
| 0,69136 | 420 | 381,6753 | 0,48976 | 1 | 0,960096 |
| 0,229297 | 367 | 340,5827 | 0,818776 | 1 | 0,951644 |
| -0,7676 | 280 | 252,286 | 0,44344 | 1 | 0,810247 |
| 3,044085 | 421 | 322,4072 | 0,002526 | 1 | 0,073944 |
| 2,231628 | 421 | 181,9878 | 0,02686 | 1 | 0,223635 |
| 1,546927 | 420 | 354,1193 | 0,122774 | 1 | 0,87201 |
| 1,749128 | 367 | 291,6567 | 0,081321 | 1 | 0,381841 |
| 1,038691 | 280 | 254,3762 | 0,299935 | 1 | 0,723324 |
| 1,75072 | 421 | 409,9064 | 0,080742 | 1 | 0,30376 |
| 1,804162 | 421 | 226,6301 | 0,072534 | 1 | 0,341588 |
| 0,112327 | 420 | 415,6615 | 0,910619 | 1 | 0,987181 |
| 1,478144 | 367 | 362,6358 | 0,140237 | 1 | 0,488078 |
| 0,178244 | 280 | 275,589 | 0,858663 | 1 | 0,966655 |
| -1,15907 | 421 | 355,5068 | 0,247205 | 1 | 0,544195 |
| -0,93228 | 421 | 193,6202 | 0,352354 | 1 | 0,643758 |
| 1,42364 | 420 | 388,6646 | 0,155353 | 1 | 0,914005 |
| -0,65973 | 367 | 334,1106 | 0,509885 | 1 | 0,797092 |
| -0,82221 | 280 | 261,0238 | 0,411709 | 1 | 0,788151 |

| | | | | | |
|---|---|---|---|---|---|
| -1,20392 | 421 | 300,8466 | 0,229567 | 1 | 0,520357 |
| -0,48142 | 421 | 175,8267 | 0,630815 | 1 | 0,835359 |
| -1,37261 | 420 | 329,5722 | 0,170805 | 1 | 0,9199 |
| 0,595491 | 367 | 290,0498 | 0,55198 | 1 | 0,830257 |
| 0,143129 | 280 | 248,7619 | 0,886304 | 1 | 0,973784 |
| 2,223839 | 421 | 328,3967 | 0,026839 | 1 | 0,192509 |
| 1,989322 | 421 | 184,0322 | 0,048147 | 1 | 0,286754 |
| 0,836765 | 420 | 362,6318 | 0,403276 | 1 | 0,95743 |
| 1,285694 | 367 | 321,9245 | 0,199474 | 1 | 0,551573 |
| 0,301785 | 280 | 252,2805 | 0,763065 | 1 | 0,944296 |
| 0,476684 | 421 | 346,9768 | 0,633887 | 1 | 0,83455 |
| 1,461055 | 421 | 190,6178 | 0,145647 | 1 | 0,426194 |
| 0,7969 | 420 | 381,3761 | 0,426005 | 1 | 0,95743 |
| 0,875269 | 367 | 341,649 | 0,382042 | 1 | 0,692825 |
| 1,042046 | 280 | 257,9818 | 0,298366 | 1 | 0,723324 |
| -0,25368 | 421 | 336,6902 | 0,799901 | 1 | 0,922616 |
| -0,89773 | 421 | 184,283 | 0,370504 | 1 | 0,649999 |
| -0,27344 | 420 | 370,802 | 0,784667 | 1 | 0,969427 |
| 1,268315 | 367 | 337,5372 | 0,20556 | 1 | 0,558434 |
| 1,227428 | 280 | 266,9377 | 0,220743 | 1 | 0,653047 |
| 1,406248 | 421 | 319,1545 | 0,160623 | 1 | 0,434577 |
| 1,897255 | 421 | 180,9546 | 0,059387 | 1 | 0,314727 |
| 0,360255 | 420 | 351,9323 | 0,718873 | 1 | 0,969427 |
| 1,444284 | 367 | 315,0529 | 0,149653 | 1 | 0,499998 |
| 1,700154 | 280 | 262,9977 | 0,090284 | 1 | 0,468147 |
| -1,28382 | 421 | 314,7783 | 0,200151 | 1 | 0,488069 |
| -0,16881 | 421 | 179,5217 | 0,866133 | 1 | 0,947215 |
| -0,48086 | 420 | 347,5893 | 0,630921 | 1 | 0,964878 |
| 0,078763 | 367 | 291,1824 | 0,937275 | 1 | 0,978845 |
| 0,222499 | 280 | 249,6737 | 0,824108 | 1 | 0,96121 |
| -2,87614 | 421 | 336,6441 | 0,004282 | 1 | 0,087964 |
| -1,50666 | 421 | 183,0495 | 0,133621 | 1 | 0,415189 |
| -1,28865 | 420 | 372,0179 | 0,198322 | 1 | 0,928589 |
| -0,19483 | 367 | 314,8202 | 0,845653 | 1 | 0,956935 |
| -2,13686 | 280 | 257,6591 | 0,033551 | 1 | 0,329568 |
| -2,6294 | 421 | 312,3496 | 0,008977 | 1 | 0,113397 |
| -0,27345 | 421 | 178,2001 | 0,784826 | 1 | 0,916847 |
| -1,35948 | 420 | 341,15 | 0,174894 | 1 | 0,9199 |
| -0,39443 | 367 | 310,2691 | 0,693537 | 1 | 0,904597 |
| 1,595205 | 280 | 252,7417 | 0,111916 | 1 | 0,506479 |
| 1,468264 | 421 | 330,9759 | 0,142982 | 1 | 0,412154 |
| 1,982386 | 421 | 185,6181 | 0,048911 | 1 | 0,286808 |
| -1,14665 | 420 | 365,9345 | 0,252275 | 1 | 0,943897 |
| 0,267765 | 367 | 330,2005 | 0,789048 | 1 | 0,944997 |
| 0,099483 | 280 | 261,8677 | 0,920831 | 1 | 0,977136 |
| 0,173471 | 421 | 362,7947 | 0,862378 | 1 | 0,951261 |
| -0,62355 | 421 | 195,6754 | 0,53365 | 1 | 0,769894 |
| -0,81455 | 420 | 393,5188 | 0,415822 | 1 | 0,95743 |

| | | | | | |
|---|---|---|---|---|---|
| -1,12544 | 367 | 353,6949 | 0,261164 | 1 | 0,609149 |
| 0,153554 | 280 | 273,522 | 0,878074 | 1 | 0,973498 |
| 0,43063 | 421 | 317,0663 | 0,66703 | 1 | 0,853323 |
| 0,516554 | 421 | 180,3992 | 0,6061 | 1 | 0,821338 |
| 1,095079 | 420 | 353,5535 | 0,274228 | 1 | 0,943897 |
| -0,33128 | 367 | 312,9173 | 0,740655 | 1 | 0,923669 |
| 2,261858 | 280 | 257,8279 | 0,024539 | 1 | 0,313396 |
| -0,15382 | 421 | 353,0516 | 0,877837 | 1 | 0,95861 |
| -0,71425 | 421 | 189,9915 | 0,475946 | 1 | 0,734136 |
| -0,81848 | 420 | 382,2276 | 0,413594 | 1 | 0,95743 |
| -0,95333 | 367 | 334,5098 | 0,341109 | 1 | 0,664831 |
| 0,237278 | 280 | 265,9415 | 0,812624 | 1 | 0,95747 |
| -0,57691 | 421 | 355,2722 | 0,564364 | 1 | 0,796637 |
| 0,21654 | 421 | 192,2738 | 0,828796 | 1 | 0,934178 |
| -0,83605 | 420 | 388,8824 | 0,403643 | 1 | 0,95743 |
| -1,13407 | 367 | 345,417 | 0,257551 | 1 | 0,609149 |
| 0,410908 | 280 | 267,436 | 0,681469 | 1 | 0,911245 |
| -0,60389 | 421 | 350,2468 | 0,546306 | 1 | 0,784719 |
| -0,97242 | 421 | 191,8077 | 0,332067 | 1 | 0,626331 |
| 1,187071 | 420 | 384,8418 | 0,235932 | 1 | 0,943055 |
| 0,185955 | 367 | 342,9693 | 0,85259 | 1 | 0,956935 |
| -1,22096 | 280 | 264,3868 | 0,223188 | 1 | 0,654014 |
| 1,221865 | 421 | 376,474 | 0,222523 | 1 | 0,51676 |
| 0,792637 | 421 | 203,1517 | 0,428914 | 1 | 0,694836 |
| 0,246164 | 420 | 405,3608 | 0,80568 | 1 | 0,969427 |
| 0,81027 | 367 | 346,7845 | 0,418341 | 1 | 0,729676 |
| -1,58812 | 280 | 273,0388 | 0,113417 | 1 | 0,508687 |
| -1,49833 | 421 | 345,1067 | 0,134961 | 1 | 0,407576 |
| -2,8388 | 421 | 188,1138 | 0,005026 | 1 | 0,137431 |
| -0,29867 | 420 | 381,0731 | 0,765357 | 1 | 0,969427 |
| -2,84956 | 367 | 338,1146 | 0,004646 | 1 | 0,178183 |
| -1,49992 | 280 | 260,4532 | 0,134847 | 1 | 0,544898 |
| 2,265197 | 421 | 399,903 | 0,024035 | 1 | 0,181103 |
| 3,755445 | 421 | 215,3338 | 2,23E-04 | 0,238948 | 0,035752 |
| 0,95849 | 420 | 415,8525 | 0,338372 | 1 | 0,947373 |
| 2,296039 | 367 | 362,9771 | 0,022243 | 1 | 0,231173 |
| 2,322369 | 280 | 274,7461 | 0,020944 | 1 | 0,30229 |
| 0,972896 | 421 | 350,8692 | 0,331275 | 1 | 0,619505 |
| -0,05547 | 421 | 189,0128 | 0,955822 | 1 | 0,98414 |
| -1,00857 | 420 | 385,0306 | 0,313815 | 1 | 0,943897 |
| 1,814623 | 367 | 338,2463 | 0,070468 | 1 | 0,36493 |
| -0,32335 | 280 | 264,7844 | 0,746684 | 1 | 0,942071 |
| -0,83001 | 421 | 344,7569 | 0,40711 | 1 | 0,695339 |
| -0,35623 | 421 | 189,1993 | 0,722066 | 1 | 0,877543 |
| 1,027802 | 420 | 379,3862 | 0,304698 | 1 | 0,943897 |
| 0,33031 | 367 | 334,0406 | 0,741373 | 1 | 0,923669 |
| -0,7264 | 280 | 267,318 | 0,468232 | 1 | 0,824256 |
| 0,915644 | 421 | 316,3673 | 0,360551 | 1 | 0,650719 |

| | | | | | |
|---|---|---|---|---|---|
| -0,60006 | 421 | 180,634 | 0,549216 | 1 | 0,776424 |
| 0,388867 | 420 | 350,8987 | 0,69761 | 1 | 0,968866 |
| 0,941422 | 367 | 303,3749 | 0,347238 | 1 | 0,668671 |
| 1,394722 | 280 | 255,4563 | 0,164311 | 1 | 0,586115 |
| -0,79551 | 421 | 292,2703 | 0,426963 | 1 | 0,700145 |
| -1,14079 | 421 | 174,1687 | 0,255526 | 1 | 0,555667 |
| 0,957303 | 420 | 322,6233 | 0,339131 | 1 | 0,947373 |
| -0,84249 | 367 | 290,4507 | 0,400209 | 1 | 0,713745 |
| 0,200164 | 280 | 276 | 0,8415 | 1 | 0,965181 |
| -1,14525 | 421 | 323,7758 | 0,252949 | 1 | 0,546261 |
| -0,12886 | 421 | 184,0211 | 0,897609 | 1 | 0,960059 |
| 0,003438 | 420 | 357,149 | 0,997259 | 1 | 0,997643 |
| 0,540192 | 367 | 327,8438 | 0,589431 | 1 | 0,853288 |
| -0,26552 | 280 | 263,5144 | 0,790817 | 1 | 0,953435 |
| -1,76731 | 421 | 342,3672 | 0,078068 | 1 | 0,302838 |
| -2,04183 | 421 | 188,0219 | 0,042566 | 1 | 0,273775 |
| -0,81619 | 420 | 377,2068 | 0,414909 | 1 | 0,95743 |
| -2,04951 | 367 | 341,9874 | 0,041175 | 1 | 0,289956 |
| -1,23135 | 280 | 262,8233 | 0,219293 | 1 | 0,650539 |
| -2,41653 | 421 | 361,2736 | 0,016164 | 1 | 0,147897 |
| -2,55457 | 421 | 193,6597 | 0,011401 | 1 | 0,163285 |
| -1,00814 | 420 | 396,1296 | 0,314 | 1 | 0,943897 |
| -1,38057 | 367 | 339,0433 | 0,168321 | 1 | 0,514971 |
| -0,30076 | 280 | 263,4825 | 0,763833 | 1 | 0,944296 |
| 0,437255 | 421 | 351,9221 | 0,662195 | 1 | 0,851474 |
| 0,701773 | 421 | 192,6609 | 0,483667 | 1 | 0,739236 |
| -0,7208 | 420 | 385,2341 | 0,471468 | 1 | 0,960096 |
| 0,38001 | 367 | 346,6155 | 0,704171 | 1 | 0,908549 |
| -0,62321 | 280 | 261,8718 | 0,533686 | 1 | 0,84326 |
| -1,31628 | 421 | 361,7638 | 0,188913 | 1 | 0,472953 |
| -1,35563 | 421 | 193,9234 | 0,176792 | 1 | 0,46335 |
| -0,14266 | 420 | 396,1548 | 0,886629 | 1 | 0,982463 |
| -2,56854 | 367 | 339,6392 | 0,01064 | 1 | 0,189546 |
| -2,13966 | 280 | 267,865 | 0,033286 | 1 | 0,329568 |
| 0,314686 | 421 | 403,9728 | 0,753162 | 1 | 0,898125 |
| 2,322353 | 421 | 219,2961 | 0,021131 | 1 | 0,2022 |
| 0,642201 | 420 | 415,7287 | 0,521096 | 1 | 0,960096 |
| 0,10519 | 367 | 362,9487 | 0,916283 | 1 | 0,973634 |
| 0,722389 | 280 | 271,485 | 0,470677 | 1 | 0,824256 |
| -3,24328 | 421 | 337,0501 | 0,0013 | 1 | 0,066922 |
| -1,21056 | 421 | 186,8036 | 0,227593 | 1 | 0,526855 |
| -0,49857 | 420 | 376,6277 | 0,618372 | 1 | 0,964878 |
| -0,01039 | 367 | 325,0225 | 0,991714 | 1 | 0,997007 |
| -0,65978 | 280 | 260,5449 | 0,509975 | 1 | 0,831912 |
| -1,10734 | 421 | 375,0698 | 0,268857 | 1 | 0,563516 |
| -0,19251 | 421 | 201,4401 | 0,84754 | 1 | 0,94331 |
| 1,613309 | 420 | 404,3494 | 0,107457 | 1 | 0,848246 |
| -1,9778 | 367 | 355,3456 | 0,048723 | 1 | 0,313787 |

| | | | | | |
|---|---|---|---|---|---|
| -1,73466 | 280 | 267,7038 | 0,083953 | 1 | 0,456738 |
| -0,53738 | 421 | 352,3344 | 0,591347 | 1 | 0,81147 |
| 0,352377 | 421 | 188,7412 | 0,724949 | 1 | 0,878959 |
| -0,25823 | 420 | 386,0151 | 0,796369 | 1 | 0,969427 |
| -0,09249 | 367 | 333,1906 | 0,926362 | 1 | 0,976733 |
| 0,199451 | 280 | 260,08 | 0,842065 | 1 | 0,965181 |
| 0,58271 | 421 | 368,2211 | 0,560445 | 1 | 0,793043 |
| 0,446546 | 421 | 197,8807 | 0,655692 | 1 | 0,841107 |
| -0,30628 | 420 | 398,9172 | 0,759552 | 1 | 0,969427 |
| -0,55165 | 367 | 348,5255 | 0,581543 | 1 | 0,845115 |
| -0,18874 | 280 | 268,081 | 0,850438 | 1 | 0,966155 |
| -2,59845 | 421 | 306,9295 | 0,009816 | 1 | 0,116484 |
| -1,41948 | 421 | 180,2446 | 0,157485 | 1 | 0,43893 |
| 0,235028 | 420 | 342,9493 | 0,814327 | 1 | 0,969427 |
| 1,333782 | 367 | 304,4131 | 0,183272 | 1 | 0,529103 |
| -1,59941 | 280 | 251,6728 | 0,110984 | 1 | 0,503775 |
| -2,15974 | 421 | 413,8844 | 0,031366 | 1 | 0,201145 |
| -1,67395 | 421 | 232,1096 | 0,095487 | 1 | 0,377685 |
| -1,5993 | 420 | 414,606 | 0,110516 | 1 | 0,848246 |
| 0,246336 | 367 | 361,3695 | 0,805562 | 1 | 0,950149 |
| 0,350038 | 280 | 275,3377 | 0,726578 | 1 | 0,936637 |
| -0,19297 | 421 | 319,582 | 0,847101 | 1 | 0,940738 |
| 0,902091 | 421 | 182,8545 | 0,368195 | 1 | 0,648213 |
| -1,21988 | 420 | 354,59 | 0,223319 | 1 | 0,943055 |
| -1,09756 | 367 | 310,5712 | 0,273248 | 1 | 0,609149 |
| -0,85166 | 280 | 247,4151 | 0,395224 | 1 | 0,782194 |
| -0,24543 | 421 | 294,3827 | 0,806294 | 1 | 0,922616 |
| -1,37047 | 421 | 173,4082 | 0,17231 | 1 | 0,456365 |
| 0,84768 | 420 | 324,4674 | 0,397241 | 1 | 0,95743 |
| 0,238615 | 367 | 277,5116 | 0,811581 | 1 | 0,950149 |
| 0,452458 | 280 | 252,25 | 0,651328 | 1 | 0,908644 |
| 0,332037 | 421 | 343,8283 | 0,740064 | 1 | 0,893273 |
| 1,812636 | 421 | 189,538 | 0,07147 | 1 | 0,339864 |
| 1,022683 | 420 | 379,4566 | 0,307109 | 1 | 0,943897 |
| 1,431208 | 367 | 332,1369 | 0,153311 | 1 | 0,501171 |
| -0,08406 | 280 | 265,9715 | 0,933069 | 1 | 0,979504 |
| -2,21696 | 421 | 385,2208 | 0,027209 | 1 | 0,192509 |
| -0,94716 | 421 | 204,1221 | 0,344677 | 1 | 0,638117 |
| -0,2865 | 420 | 409,1951 | 0,774637 | 1 | 0,969427 |
| 1,277548 | 367 | 354,8555 | 0,202244 | 1 | 0,556171 |
| -0,66375 | 280 | 265,3222 | 0,507428 | 1 | 0,831912 |
| 1,702968 | 421 | 331,3874 | 0,089512 | 1 | 0,319654 |
| 0,800614 | 421 | 181,6787 | 0,424401 | 1 | 0,693378 |
| 0,118295 | 420 | 369,4845 | 0,905898 | 1 | 0,98503 |
| 1,537956 | 367 | 301,4333 | 0,125108 | 1 | 0,462104 |
| 0,439853 | 280 | 261,4471 | 0,660407 | 1 | 0,908644 |
| 0,025268 | 421 | 328,1935 | 0,979857 | 1 | 0,997025 |
| -0,50038 | 421 | 185,9982 | 0,617402 | 1 | 0,825805 |

| | | | | | |
|---|---|---|---|---|---|
| 0,861823 | 420 | 364,1205 | 0,389352 | 1 | 0,95743 |
| 0,717318 | 367 | 317,0785 | 0,473706 | 1 | 0,776795 |
| -0,22878 | 280 | 250,3821 | 0,819229 | 1 | 0,95778 |
| 2,41113 | 421 | 370,6734 | 0,016389 | 1 | 0,148783 |
| 2,530106 | 421 | 197,1841 | 0,012185 | 1 | 0,169272 |
| 0,784971 | 420 | 402,961 | 0,432932 | 1 | 0,960096 |
| 0,946293 | 367 | 346,9818 | 0,344658 | 1 | 0,667378 |
| 1,691056 | 280 | 266,5653 | 0,091995 | 1 | 0,468366 |
| -0,14785 | 421 | 349,7315 | 0,882544 | 1 | 0,960978 |
| -0,14 | 421 | 187,6219 | 0,888812 | 1 | 0,958798 |
| -0,92213 | 420 | 385,3348 | 0,357037 | 1 | 0,947373 |
| -1,21208 | 367 | 332,3278 | 0,226344 | 1 | 0,576775 |
| -1,96171 | 280 | 260,0599 | 0,050863 | 1 | 0,398004 |
| -3,07373 | 421 | 361,2447 | 0,002275 | 1 | 0,069718 |
| -0,92879 | 421 | 193,8867 | 0,354154 | 1 | 0,643758 |
| 0,44398 | 420 | 391,6803 | 0,657302 | 1 | 0,96781 |
| 0,517999 | 367 | 353,4452 | 0,604783 | 1 | 0,867182 |
| -0,7097 | 280 | 270,7356 | 0,478502 | 1 | 0,824256 |
| 1,64678 | 421 | 311,6226 | 0,100611 | 1 | 0,342121 |
| 0,104473 | 421 | 180,8732 | 0,91691 | 1 | 0,969673 |
| -0,20482 | 420 | 345,0799 | 0,837836 | 1 | 0,973894 |
| 0,077081 | 367 | 295,6017 | 0,938611 | 1 | 0,979214 |
| -1,92865 | 280 | 276 | 0,054799 | 1 | 0,40654 |
| 0,843585 | 421 | 364,9668 | 0,399454 | 1 | 0,688761 |
| 1,474017 | 421 | 196,0952 | 0,142081 | 1 | 0,423499 |
| 0,211517 | 420 | 396,7369 | 0,832592 | 1 | 0,973894 |
| 2,817298 | 367 | 343,6727 | 0,005123 | 1 | 0,178183 |
| 0,093361 | 280 | 268,421 | 0,925687 | 1 | 0,979504 |
| 0,837817 | 421 | 312,6603 | 0,402774 | 1 | 0,692899 |
| 1,728248 | 421 | 178,419 | 0,085674 | 1 | 0,365475 |
| 0,66433 | 420 | 347,4099 | 0,50692 | 1 | 0,960096 |
| 0,382791 | 367 | 299,625 | 0,702146 | 1 | 0,908269 |
| -0,83984 | 280 | 260,6694 | 0,401767 | 1 | 0,784757 |
| -0,86094 | 421 | 338,9724 | 0,389882 | 1 | 0,679251 |
| -0,45411 | 421 | 188,1759 | 0,65027 | 1 | 0,839358 |
| -0,7458 | 420 | 374,7178 | 0,456259 | 1 | 0,960096 |
| 0,973373 | 367 | 328,2792 | 0,331085 | 1 | 0,659108 |
| -0,65855 | 280 | 261,3574 | 0,510765 | 1 | 0,831912 |
| -0,27939 | 421 | 376,207 | 0,780101 | 1 | 0,9142 |
| -2,62076 | 421 | 200,1097 | 0,009447 | 1 | 0,148297 |
| -0,09631 | 420 | 403,9972 | 0,923322 | 1 | 0,989204 |
| -1,42653 | 367 | 337,2908 | 0,154639 | 1 | 0,502197 |
| -1,08091 | 280 | 268,4171 | 0,280706 | 1 | 0,703928 |
| -2,61201 | 421 | 332,1377 | 0,00941 | 1 | 0,114362 |
| -1,19714 | 421 | 185,0569 | 0,232784 | 1 | 0,533704 |
| -0,69229 | 420 | 368,883 | 0,489193 | 1 | 0,960096 |
| -1,44249 | 367 | 326,0981 | 0,150125 | 1 | 0,499998 |
| -2,08657 | 280 | 259,5624 | 0,037903 | 1 | 0,348289 |

| | | | | | |
|---|---|---|---|---|---|
| -1,38478 | 421 | 354,4678 | 0,16699 | 1 | 0,441901 |
| -2,63974 | 421 | 191,6176 | 0,00898 | 1 | 0,14815 |
| -0,1599 | 420 | 388,8855 | 0,873041 | 1 | 0,982463 |
| -2,96436 | 367 | 343,9035 | 0,003245 | 1 | 0,176492 |
| -1,48865 | 280 | 263,788 | 0,137775 | 1 | 0,54931 |
| -1,41941 | 421 | 359,4645 | 0,156647 | 1 | 0,429212 |
| 0,648244 | 421 | 192,042 | 0,517602 | 1 | 0,761675 |
| 1,306154 | 420 | 389,2108 | 0,192272 | 1 | 0,925729 |
| 2,2244 | 367 | 339,2876 | 0,026779 | 1 | 0,252225 |
| 1,29389 | 280 | 259,0694 | 0,196856 | 1 | 0,627192 |
| -0,47677 | 421 | 364,0687 | 0,633812 | 1 | 0,83455 |
| 0,25478 | 421 | 195,6456 | 0,799161 | 1 | 0,921861 |
| 0,250566 | 420 | 396,3991 | 0,80228 | 1 | 0,969427 |
| 0,55346 | 367 | 342,6254 | 0,580309 | 1 | 0,844137 |
| 0,43761 | 280 | 262,7544 | 0,662029 | 1 | 0,908644 |
| -0,27683 | 421 | 340,3201 | 0,782077 | 1 | 0,915149 |
| -0,95071 | 421 | 187,6892 | 0,342976 | 1 | 0,636552 |
| -1,34457 | 420 | 367,9278 | 0,179594 | 1 | 0,923408 |
| -0,12126 | 367 | 325,3888 | 0,903559 | 1 | 0,970486 |
| 0,793348 | 280 | 253,8487 | 0,428316 | 1 | 0,800834 |
| -2,28044 | 421 | 328,9448 | 0,023221 | 1 | 0,177419 |
| -2,06556 | 421 | 184,0897 | 0,040271 | 1 | 0,26998 |
| -0,32646 | 420 | 368,937 | 0,744258 | 1 | 0,969427 |
| -0,51098 | 367 | 333,3934 | 0,6097 | 1 | 0,870021 |
| -2,66253 | 280 | 265,741 | 0,008229 | 1 | 0,203287 |
| 1,906079 | 421 | 370,3544 | 0,057414 | 1 | 0,266382 |
| 0,673313 | 421 | 198,9124 | 0,50153 | 1 | 0,750551 |
| -0,12723 | 420 | 403,5943 | 0,898821 | 1 | 0,98503 |
| 0,510241 | 367 | 352,1793 | 0,610202 | 1 | 0,870021 |
| -1,26711 | 280 | 267,1992 | 0,20622 | 1 | 0,638139 |
| -3,68508 | 421 | 341,2196 | 2,66E-04 | 0,285222 | 0,04642 |
| -3,21426 | 421 | 185,8272 | 0,001542 | 1 | 0,093671 |
| -0,12578 | 420 | 381,2044 | 0,899972 | 1 | 0,98503 |
| -0,71163 | 367 | 338,1057 | 0,477186 | 1 | 0,780803 |
| -3,20901 | 280 | 261,4328 | 0,001498 | 1 | 0,135787 |
| -1,62974 | 421 | 282,838 | 0,10427 | 1 | 0,346875 |
| 0,247491 | 421 | 168,9577 | 0,804828 | 1 | 0,923205 |
| -0,66248 | 420 | 308,8002 | 0,508158 | 1 | 0,960096 |
| -2,23628 | 367 | 281,2352 | 0,026118 | 1 | 0,252225 |
| -1,02423 | 280 | 276 | 0,306625 | 1 | 0,72861 |
| -0,54676 | 421 | 333,4453 | 0,584908 | 1 | 0,807036 |
| 0,396114 | 421 | 186,0925 | 0,692475 | 1 | 0,865526 |
| -1,02047 | 420 | 368,6294 | 0,308177 | 1 | 0,943897 |
| 0,017076 | 367 | 332,2993 | 0,986386 | 1 | 0,996248 |
| -0,67327 | 280 | 261,829 | 0,501372 | 1 | 0,831912 |
| 1,825221 | 421 | 328,1839 | 0,068876 | 1 | 0,282663 |
| -0,70835 | 421 | 182,4859 | 0,479634 | 1 | 0,736057 |
| -0,33783 | 420 | 362,5049 | 0,735686 | 1 | 0,969427 |

| | | | | | |
|---|---|---|---|---|---|
| 2,016276 | 367 | 319,9469 | 0,044606 | 1 | 0,302719 |
| -0,39928 | 280 | 264,8949 | 0,690009 | 1 | 0,91859 |
| 0,315713 | 421 | 339,8613 | 0,752414 | 1 | 0,898125 |
| 2,004891 | 421 | 186,652 | 0,04642 | 1 | 0,282075 |
| 0,243454 | 420 | 375,0562 | 0,807787 | 1 | 0,969427 |
| -0,14978 | 367 | 337,2362 | 0,881025 | 1 | 0,962453 |
| 1,095201 | 280 | 261,2703 | 0,274437 | 1 | 0,701156 |
| 0,79369 | 421 | 402,7467 | 0,427844 | 1 | 0,700826 |
| 1,482785 | 421 | 218,1445 | 0,139575 | 1 | 0,423351 |
| 0,291248 | 420 | 415,608 | 0,771007 | 1 | 0,969427 |
| 1,199619 | 367 | 362,3068 | 0,231071 | 1 | 0,582149 |
| 1,247227 | 280 | 273,5889 | 0,213381 | 1 | 0,646827 |
| -0,4845 | 421 | 352,7497 | 0,628329 | 1 | 0,833119 |
| -1,0571 | 421 | 192,4276 | 0,291792 | 1 | 0,588663 |
| -1,28592 | 420 | 385,9596 | 0,199241 | 1 | 0,928589 |
| -1,35587 | 367 | 339,2905 | 0,176042 | 1 | 0,523018 |
| -0,25781 | 280 | 265,6046 | 0,796755 | 1 | 0,954269 |
| -1,4947 | 421 | 361,6268 | 0,135865 | 1 | 0,407576 |
| -1,25559 | 421 | 195,9639 | 0,210761 | 1 | 0,509968 |
| 0,287344 | 420 | 393,9912 | 0,774 | 1 | 0,969427 |
| -1,66297 | 367 | 351,5307 | 0,09721 | 1 | 0,412696 |
| -1,74412 | 280 | 266,0778 | 0,082293 | 1 | 0,456738 |
| -1,40095 | 421 | 351,3243 | 0,162113 | 1 | 0,436257 |
| -2,74102 | 421 | 187,7683 | 0,006717 | 1 | 0,141161 |
| -1,19102 | 420 | 383,4993 | 0,234383 | 1 | 0,943055 |
| -1,2925 | 367 | 326,0875 | 0,197098 | 1 | 0,549567 |
| -0,27716 | 280 | 255,0389 | 0,781882 | 1 | 0,951006 |
| -2,43618 | 421 | 353,4075 | 0,015337 | 1 | 0,147897 |
| -1,08774 | 421 | 193,6705 | 0,278063 | 1 | 0,578094 |
| -0,04037 | 420 | 390,4993 | 0,967817 | 1 | 0,996924 |
| -1,62609 | 367 | 343,5077 | 0,104848 | 1 | 0,429852 |
| -0,72053 | 280 | 262,0792 | 0,471839 | 1 | 0,824256 |
| -2,37765 | 421 | 354,1991 | 0,017953 | 1 | 0,156391 |
| -1,68261 | 421 | 191,2206 | 0,094083 | 1 | 0,373113 |
| -1,13848 | 420 | 382,3083 | 0,255634 | 1 | 0,943897 |
| 1,353854 | 367 | 351,5076 | 0,176653 | 1 | 0,523018 |
| 0,717463 | 280 | 267,3417 | 0,473715 | 1 | 0,824256 |
| -2,65385 | 421 | 332,1598 | 0,008341 | 1 | 0,112451 |
| -2,6795 | 421 | 185,0808 | 0,008038 | 1 | 0,148102 |
| -0,0632 | 420 | 374,0378 | 0,949637 | 1 | 0,992811 |
| -2,09568 | 367 | 327,838 | 0,036877 | 1 | 0,28464 |
| -1,10782 | 280 | 263,0191 | 0,268951 | 1 | 0,693794 |
| 2,561384 | 421 | 334,4227 | 0,010864 | 1 | 0,124028 |
| 1,733155 | 421 | 184,9741 | 0,084735 | 1 | 0,365475 |
| 4,118544 | 420 | 361,5284 | 4,73E-05 | 0,050981 | 0,035635 |
| 1,634724 | 367 | 329,5951 | 0,103062 | 1 | 0,426647 |
| 0,67844 | 280 | 258,2769 | 0,4981 | 1 | 0,83103 |
| -2,82673 | 421 | 366,5124 | 0,004961 | 1 | 0,087964 |

| | | | | | |
|---|---|---|---|---|---|
| -1,53553 | 421 | 198,9888 | 0,126242 | 1 | 0,410224 |
| -0,15222 | 420 | 399,9683 | 0,879087 | 1 | 0,982463 |
| -0,013 | 367 | 355,4261 | 0,989638 | 1 | 0,996248 |
| -1,84257 | 280 | 272,9969 | 0,066476 | 1 | 0,425749 |
| 0,474696 | 421 | 341,65 | 0,635307 | 1 | 0,83455 |
| -0,70582 | 421 | 188,6666 | 0,481169 | 1 | 0,737662 |
| 0,640589 | 420 | 376,5051 | 0,52218 | 1 | 0,960096 |
| -0,3123 | 367 | 327,2007 | 0,75501 | 1 | 0,92504 |
| -2,95961 | 280 | 265,3607 | 0,003359 | 1 | 0,143818 |
| 1,242899 | 421 | 369,8708 | 0,214692 | 1 | 0,506731 |
| 1,861885 | 421 | 197,6156 | 0,064104 | 1 | 0,324177 |
| 0,437411 | 420 | 400,4108 | 0,662049 | 1 | 0,96781 |
| 1,201782 | 367 | 350,4851 | 0,23026 | 1 | 0,581242 |
| 1,11817 | 280 | 265,8394 | 0,264504 | 1 | 0,688875 |
| -2,1929 | 421 | 320,5476 | 0,029033 | 1 | 0,197976 |
| -1,18456 | 421 | 183,8233 | 0,237719 | 1 | 0,537903 |
| 1,507196 | 420 | 353,8233 | 0,132653 | 1 | 0,888479 |
| 0,033845 | 367 | 314,9432 | 0,973022 | 1 | 0,989436 |
| -0,29244 | 280 | 259,8642 | 0,770185 | 1 | 0,946609 |
| -0,28893 | 421 | 357,7741 | 0,772799 | 1 | 0,90985 |
| -1,23808 | 421 | 191,055 | 0,217206 | 1 | 0,514565 |
| -0,33094 | 420 | 390,6472 | 0,740868 | 1 | 0,969427 |
| 2,657932 | 367 | 348,3803 | 0,008225 | 1 | 0,182271 |
| -2,41455 | 280 | 262,2774 | 0,01644 | 1 | 0,266517 |
| -1,43665 | 421 | 334,4396 | 0,151752 | 1 | 0,42445 |
| 0,09964 | 421 | 188,3188 | 0,920736 | 1 | 0,970834 |
| 0,857526 | 420 | 375,8376 | 0,391701 | 1 | 0,95743 |
| 1,51475 | 367 | 329,0314 | 0,130796 | 1 | 0,47485 |
| 1,271219 | 280 | 260,9225 | 0,204783 | 1 | 0,634995 |
| 0,140836 | 421 | 320,3564 | 0,888088 | 1 | 0,962 |
| 1,095717 | 421 | 181,7904 | 0,274652 | 1 | 0,576463 |
| 1,055878 | 420 | 356,1726 | 0,29174 | 1 | 0,943897 |
| 1,966449 | 367 | 290,0909 | 0,0502 | 1 | 0,316533 |
| 0,933786 | 280 | 251,1464 | 0,351311 | 1 | 0,761846 |
| 1,333402 | 421 | 352,9918 | 0,18326 | 1 | 0,465721 |
| 0,479519 | 421 | 192,664 | 0,632113 | 1 | 0,835691 |
| -1,26511 | 420 | 388,8842 | 0,206591 | 1 | 0,930277 |
| 0,47419 | 367 | 338,9025 | 0,63567 | 1 | 0,880707 |
| -1,4725 | 280 | 264,1437 | 0,142078 | 1 | 0,554427 |
| -0,75326 | 421 | 288,4321 | 0,451909 | 1 | 0,716117 |
| -0,57988 | 421 | 175,4626 | 0,562742 | 1 | 0,785233 |
| -0,63622 | 420 | 317,5329 | 0,525091 | 1 | 0,960096 |
| -1,55564 | 367 | 289,7173 | 0,120885 | 1 | 0,452158 |
| -0,6407 | 280 | 276 | 0,522251 | 1 | 0,833721 |
| -0,24009 | 421 | 361,3212 | 0,810396 | 1 | 0,924931 |
| -0,63138 | 421 | 193,5405 | 0,528536 | 1 | 0,768085 |
| -1,19093 | 420 | 393,3309 | 0,234399 | 1 | 0,943055 |
| 1,610086 | 367 | 339,9283 | 0,108307 | 1 | 0,435251 |

| | | | | | |
|---|---|---|---|---|---|
| -2,10269 | 280 | 258,5387 | 0,03646 | 1 | 0,34044 |
| -3,21771 | 421 | 311,4031 | 0,001428 | 1 | 0,069432 |
| -2,58662 | 421 | 180,4913 | 0,01048 | 1 | 0,156367 |
| -2,02091 | 420 | 355,9149 | 0,044037 | 1 | 0,846132 |
| -0,85468 | 367 | 310,1065 | 0,393386 | 1 | 0,705753 |
| -0,21409 | 280 | 257,4297 | 0,830647 | 1 | 0,964667 |
| -2,97173 | 421 | 391,8044 | 0,003144 | 1 | 0,078203 |
| -2,64159 | 421 | 210,5343 | 0,008871 | 1 | 0,14815 |
| -0,24242 | 420 | 413,8821 | 0,808575 | 1 | 0,969427 |
| -1,91434 | 367 | 360,1598 | 0,056368 | 1 | 0,337523 |
| -2,151 | 280 | 274,1069 | 0,03235 | 1 | 0,329568 |
| -2,5166 | 421 | 340,2185 | 0,012309 | 1 | 0,131857 |
| -2,51222 | 421 | 186,5418 | 0,012845 | 1 | 0,171309 |
| 0,721021 | 420 | 375,2842 | 0,471345 | 1 | 0,960096 |
| -2,19921 | 367 | 324,0974 | 0,028569 | 1 | 0,257802 |
| -0,94693 | 280 | 256,6152 | 0,344566 | 1 | 0,758934 |
| -0,80334 | 421 | 353,8382 | 0,422316 | 1 | 0,697735 |
| -1,62751 | 421 | 191,5444 | 0,105273 | 1 | 0,385063 |
| -0,92837 | 420 | 381,928 | 0,353804 | 1 | 0,947373 |
| -1,55662 | 367 | 342,3388 | 0,120486 | 1 | 0,452158 |
| -0,2114 | 280 | 263,8818 | 0,832736 | 1 | 0,964667 |
| -1,19542 | 421 | 345,1052 | 0,232743 | 1 | 0,525066 |
| -1,40359 | 421 | 187,757 | 0,162094 | 1 | 0,445827 |
| 1,124254 | 420 | 382,1834 | 0,261611 | 1 | 0,943897 |
| -2,06963 | 367 | 338,0905 | 0,039246 | 1 | 0,28956 |
| -0,42477 | 280 | 264,9047 | 0,67135 | 1 | 0,908644 |
| -0,65793 | 421 | 324,6232 | 0,511047 | 1 | 0,764035 |
| 0,91833 | 421 | 178,6801 | 0,359684 | 1 | 0,643758 |
| -0,31142 | 420 | 359,8456 | 0,755661 | 1 | 0,969427 |
| 1,543917 | 367 | 317,7866 | 0,123604 | 1 | 0,458795 |
| 1,517571 | 280 | 261,1476 | 0,130332 | 1 | 0,538112 |
| -1,22002 | 421 | 344,3522 | 0,223292 | 1 | 0,51676 |
| 0,59034 | 421 | 186,7254 | 0,555676 | 1 | 0,778696 |
| 0,123919 | 420 | 377,9368 | 0,901445 | 1 | 0,98503 |
| 0,266359 | 367 | 316,6727 | 0,790136 | 1 | 0,944997 |
| -1,83047 | 280 | 248,5072 | 0,068377 | 1 | 0,427568 |
| 1,17268 | 421 | 370,7724 | 0,241677 | 1 | 0,538768 |
| 1,170726 | 421 | 197,3204 | 0,243121 | 1 | 0,542422 |
| -0,30386 | 420 | 403,0993 | 0,761389 | 1 | 0,969427 |
| 2,590317 | 367 | 336,5089 | 0,010006 | 1 | 0,183886 |
| 2,280324 | 280 | 265,8545 | 0,02338 | 1 | 0,313396 |
| 1,375093 | 421 | 372,0781 | 0,16993 | 1 | 0,442288 |
| 1,58384 | 421 | 197,0385 | 0,114834 | 1 | 0,392379 |
| 0,423229 | 420 | 403,073 | 0,672354 | 1 | 0,96781 |
| 1,374361 | 367 | 346,2447 | 0,170219 | 1 | 0,518221 |
| 0,251469 | 280 | 265,9479 | 0,801646 | 1 | 0,956473 |
| 1,484697 | 421 | 323,9288 | 0,138597 | 1 | 0,410344 |
| 1,769321 | 421 | 183,3842 | 0,078502 | 1 | 0,355264 |

| | | | | | |
|---|---|---|---|---|---|
| -1,11198 | 420 | 359,3012 | 0,266891 | 1 | 0,943897 |
| 0,900899 | 367 | 319,3974 | 0,368321 | 1 | 0,678946 |
| -0,19721 | 280 | 257,7169 | 0,84382 | 1 | 0,965181 |
| -1,3418 | 421 | 310,1417 | 0,180641 | 1 | 0,46049 |
| -3,53117 | 421 | 179,702 | 5,26E-04 | 0,561433 | 0,049513 |
| -1,93894 | 420 | 343,1743 | 0,053328 | 1 | 0,846627 |
| -2,76726 | 367 | 302,8416 | 0,006001 | 1 | 0,178183 |
| -2,40308 | 280 | 253,1115 | 0,016978 | 1 | 0,266517 |
| -2,28112 | 421 | 351,2858 | 0,02314 | 1 | 0,177419 |
| -0,69403 | 421 | 188,188 | 0,488522 | 1 | 0,742889 |
| -1,0504 | 420 | 384,5554 | 0,294194 | 1 | 0,943897 |
| -1,68615 | 367 | 333,8723 | 0,0927 | 1 | 0,40529 |
| 0,104941 | 280 | 264,0295 | 0,916503 | 1 | 0,977051 |
| -1,82214 | 421 | 383,9974 | 0,069212 | 1 | 0,282663 |
| -0,9329 | 421 | 204,0917 | 0,351975 | 1 | 0,643758 |
| -0,24472 | 420 | 408,5996 | 0,806796 | 1 | 0,969427 |
| -0,79723 | 367 | 344,2293 | 0,425867 | 1 | 0,733464 |
| 0,206255 | 280 | 257,2509 | 0,836755 | 1 | 0,964667 |
| -1,05929 | 421 | 339,5734 | 0,290219 | 1 | 0,587061 |
| -1,56765 | 421 | 188,5195 | 0,118639 | 1 | 0,396436 |
| -2,00171 | 420 | 372,8781 | 0,04604 | 1 | 0,846132 |
| -1,44808 | 367 | 323,1604 | 0,148565 | 1 | 0,499998 |
| -0,65589 | 280 | 262,107 | 0,512471 | 1 | 0,831912 |
| -1,7132 | 421 | 414,3569 | 0,087423 | 1 | 0,316698 |
| -1,83519 | 421 | 236,1399 | 0,067735 | 1 | 0,334428 |
| -2,57122 | 420 | 413,5714 | 0,010483 | 1 | 0,607592 |
| -1,28596 | 367 | 361,9373 | 0,19928 | 1 | 0,551573 |
| 0,567064 | 280 | 274,4987 | 0,571134 | 1 | 0,869862 |
| -1,30692 | 421 | 303,8349 | 0,192229 | 1 | 0,478035 |
| -0,38533 | 421 | 177,5437 | 0,700458 | 1 | 0,865528 |
| 0,134077 | 420 | 335,4814 | 0,893422 | 1 | 0,98503 |
| -0,4692 | 367 | 292,2101 | 0,639275 | 1 | 0,880707 |
| -0,16336 | 280 | 250,9552 | 0,870363 | 1 | 0,969656 |
| 0,290923 | 421 | 382,1449 | 0,771268 | 1 | 0,909469 |
| 1,83947 | 421 | 203,0166 | 0,067306 | 1 | 0,333654 |
| -0,18967 | 420 | 407,573 | 0,849664 | 1 | 0,973894 |
| 2,234847 | 367 | 351,892 | 0,026055 | 1 | 0,252225 |
| -1,6133 | 280 | 266,7363 | 0,107862 | 1 | 0,497151 |
| -0,78511 | 421 | 306,5442 | 0,432993 | 1 | 0,705289 |
| -1,69129 | 421 | 177,9168 | 0,092532 | 1 | 0,373113 |
| -3,2544 | 420 | 334,4514 | 0,001252 | 1 | 0,235929 |
| -2,00594 | 367 | 284,9618 | 0,045808 | 1 | 0,306809 |
| -1,21538 | 280 | 251,4584 | 0,225362 | 1 | 0,654014 |
| 1,037783 | 421 | 326,9855 | 0,300138 | 1 | 0,596172 |
| 2,226885 | 421 | 181,7493 | 0,027183 | 1 | 0,22508 |
| 1,643208 | 420 | 360,6037 | 0,101212 | 1 | 0,848246 |
| 2,414894 | 367 | 315,0385 | 0,016309 | 1 | 0,208286 |
| 1,610713 | 280 | 253,7666 | 0,108485 | 1 | 0,497151 |

| | | | | | |
|---|---|---|---|---|---|
| -0,8214 | 421 | 384,2025 | 0,41193 | 1 | 0,695339 |
| -1,03289 | 421 | 205,6945 | 0,302867 | 1 | 0,601345 |
| -0,00598 | 420 | 409,1586 | 0,995233 | 1 | 0,997342 |
| -1,56751 | 367 | 358,1625 | 0,117879 | 1 | 0,445308 |
| -1,08918 | 280 | 271,1692 | 0,277042 | 1 | 0,701156 |
| -0,80342 | 421 | 402,6236 | 0,422208 | 1 | 0,697735 |
| 0,850177 | 421 | 215,5797 | 0,39617 | 1 | 0,667817 |
| -1,05928 | 420 | 415,849 | 0,290085 | 1 | 0,943897 |
| 1,198274 | 367 | 361,6584 | 0,231595 | 1 | 0,582149 |
| -0,95993 | 280 | 274,6377 | 0,337935 | 1 | 0,755731 |
| -2,44073 | 421 | 307,2961 | 0,015222 | 1 | 0,147897 |
| -1,58155 | 421 | 177,2676 | 0,115535 | 1 | 0,392379 |
| -0,55438 | 420 | 340,9017 | 0,579685 | 1 | 0,964878 |
| -2,51779 | 367 | 303,5945 | 0,012324 | 1 | 0,189546 |
| -2,28634 | 280 | 264,1806 | 0,023026 | 1 | 0,313396 |
| -1,15281 | 421 | 342,5148 | 0,249793 | 1 | 0,545586 |
| -1,58866 | 421 | 188,6339 | 0,113813 | 1 | 0,391524 |
| -1,6743 | 420 | 375,239 | 0,094905 | 1 | 0,848246 |
| 0,606107 | 367 | 342,4683 | 0,544845 | 1 | 0,825208 |
| 0,243352 | 280 | 268,1218 | 0,807919 | 1 | 0,95747 |
| -1,03152 | 421 | 361,7155 | 0,302984 | 1 | 0,599209 |
| -1,82928 | 421 | 193,5247 | 0,068896 | 1 | 0,335504 |
| -0,49549 | 420 | 394,0363 | 0,62053 | 1 | 0,964878 |
| -1,23541 | 367 | 354,2353 | 0,217498 | 1 | 0,569492 |
| -0,91085 | 280 | 262,6828 | 0,36321 | 1 | 0,762031 |
| -0,29535 | 421 | 378,7062 | 0,767887 | 1 | 0,906901 |
| -1,07291 | 421 | 200,6995 | 0,2846 | 1 | 0,582733 |
| 0,56652 | 420 | 405,5602 | 0,571354 | 1 | 0,964199 |
| -0,21724 | 367 | 354,2365 | 0,828147 | 1 | 0,956109 |
| -0,59978 | 280 | 265,2129 | 0,549162 | 1 | 0,852208 |
| -0,33713 | 421 | 404,8228 | 0,736192 | 1 | 0,893273 |
| -0,60786 | 421 | 220,0781 | 0,543908 | 1 | 0,774002 |
| -2,20468 | 420 | 415,9345 | 0,028023 | 1 | 0,767829 |
| -2,05778 | 367 | 362,9797 | 0,040325 | 1 | 0,289956 |
| -1,00923 | 280 | 274,1726 | 0,313755 | 1 | 0,734206 |
| 0,826612 | 421 | 344,1152 | 0,40903 | 1 | 0,695339 |
| 0,804139 | 421 | 187,7657 | 0,422334 | 1 | 0,692554 |
| -0,29564 | 420 | 377,2966 | 0,767669 | 1 | 0,969427 |
| 0,409097 | 367 | 329,6813 | 0,682734 | 1 | 0,900574 |
| 0,746736 | 280 | 254,9534 | 0,455911 | 1 | 0,815983 |
| 0,765554 | 421 | 379,5849 | 0,444418 | 1 | 0,713799 |
| 0,30358 | 421 | 197,8764 | 0,761767 | 1 | 0,901085 |
| 0,009039 | 420 | 405,1122 | 0,992792 | 1 | 0,997342 |
| 2,318869 | 367 | 355,1562 | 0,020969 | 1 | 0,225192 |
| 2,178623 | 280 | 267,2342 | 0,030234 | 1 | 0,329568 |
| 0,729525 | 421 | 333,5225 | 0,466193 | 1 | 0,728034 |
| 2,121008 | 421 | 185,8498 | 0,035247 | 1 | 0,255449 |
| 1,697462 | 420 | 367,7605 | 0,090455 | 1 | 0,848246 |

| | | | | | |
|---|---|---|---|---|---|
| 1,293785 | 367 | 312,8648 | 0,196694 | 1 | 0,549567 |
| -0,1031 | 280 | 263,0148 | 0,917965 | 1 | 0,977051 |
| -0,87112 | 421 | 295,5786 | 0,384394 | 1 | 0,674247 |
| -0,96299 | 421 | 175,7601 | 0,336874 | 1 | 0,63143 |
| -0,14903 | 420 | 325,1485 | 0,881622 | 1 | 0,982463 |
| -0,77619 | 367 | 300,1448 | 0,438248 | 1 | 0,742744 |
| -1,30428 | 280 | 246,2271 | 0,193354 | 1 | 0,62543 |
| 0,807442 | 421 | 348,1858 | 0,419963 | 1 | 0,697735 |
| 0,821702 | 421 | 191,2795 | 0,412269 | 1 | 0,684933 |
| 0,616027 | 420 | 383,3723 | 0,538242 | 1 | 0,960096 |
| 0,689377 | 367 | 342,429 | 0,491053 | 1 | 0,792348 |
| -0,08207 | 280 | 256,342 | 0,934657 | 1 | 0,979504 |
| 2,203356 | 421 | 353,7368 | 0,028214 | 1 | 0,19529 |
| -1,61156 | 421 | 191,783 | 0,108702 | 1 | 0,388185 |
| -0,67875 | 420 | 387,9316 | 0,497699 | 1 | 0,960096 |
| 0,330706 | 367 | 342,2205 | 0,741069 | 1 | 0,923669 |
| -2,61302 | 280 | 266,1754 | 0,009485 | 1 | 0,226737 |
| -0,404 | 421 | 360,0912 | 0,686454 | 1 | 0,864986 |
| -0,7711 | 421 | 193,9526 | 0,441587 | 1 | 0,706446 |
| 0,22918 | 420 | 390,9227 | 0,818849 | 1 | 0,97003 |
| 0,287446 | 367 | 338,406 | 0,773947 | 1 | 0,93908 |
| -0,82055 | 280 | 264,6524 | 0,412642 | 1 | 0,788151 |
| 0,133827 | 421 | 352,8096 | 0,893616 | 1 | 0,962 |
| 0,542169 | 421 | 192,417 | 0,58833 | 1 | 0,80382 |
| -0,19265 | 420 | 385,91 | 0,847335 | 1 | 0,973894 |
| 0,072297 | 367 | 343,6499 | 0,942408 | 1 | 0,979539 |
| -1,43328 | 280 | 257,5229 | 0,152991 | 1 | 0,565026 |
| -0,33742 | 421 | 387,8776 | 0,73598 | 1 | 0,893273 |
| 0,544098 | 421 | 206,8705 | 0,58696 | 1 | 0,802811 |
| 1,102994 | 420 | 410,9008 | 0,270675 | 1 | 0,943897 |
| 0,09169 | 367 | 358,913 | 0,926995 | 1 | 0,976733 |
| 0,380465 | 280 | 274,3266 | 0,703894 | 1 | 0,921995 |
| -2,76215 | 421 | 365,3872 | 0,006032 | 1 | 0,097752 |
| 0,090211 | 421 | 196,9369 | 0,928211 | 1 | 0,972749 |
| -0,18387 | 420 | 394,718 | 0,85421 | 1 | 0,973894 |
| 0,819078 | 367 | 339,6692 | 0,413316 | 1 | 0,724248 |
| 0,512199 | 280 | 258,0596 | 0,60895 | 1 | 0,886655 |
| -1,61869 | 421 | 378,4524 | 0,106347 | 1 | 0,351457 |
| -1,65106 | 421 | 201,8408 | 0,100282 | 1 | 0,382595 |
| 0,267656 | 420 | 405,0932 | 0,789101 | 1 | 0,969427 |
| -1,05598 | 367 | 357,1198 | 0,29169 | 1 | 0,627013 |
| -0,27143 | 280 | 267,7044 | 0,786268 | 1 | 0,953235 |
| 0,331654 | 421 | 360,7466 | 0,740343 | 1 | 0,893273 |
| 1,582441 | 421 | 194,4827 | 0,115174 | 1 | 0,392379 |
| 0,474097 | 420 | 393,0618 | 0,635694 | 1 | 0,964878 |
| 3,157861 | 367 | 352,0154 | 0,001727 | 1 | 0,176492 |
| 2,193544 | 280 | 263,9148 | 0,029141 | 1 | 0,326641 |
| -1,17034 | 421 | 328,8754 | 0,242712 | 1 | 0,53918 |

| | | | | | |
|---|---|---|---|---|---|
| -1,5263 | 421 | 184,2919 | 0,12865 | 1 | 0,410224 |
| -2,83062 | 420 | 364,7921 | 0,004903 | 1 | 0,449965 |
| -1,10889 | 367 | 321,0355 | 0,268308 | 1 | 0,609149 |
| -2,91947 | 280 | 262,0047 | 0,003811 | 1 | 0,147267 |
| -0,80973 | 421 | 358,2337 | 0,418633 | 1 | 0,697735 |
| -0,48518 | 421 | 195,0391 | 0,628091 | 1 | 0,832483 |
| 0,67109 | 420 | 391,9953 | 0,502558 | 1 | 0,960096 |
| 0,190256 | 367 | 326,9406 | 0,849226 | 1 | 0,956935 |
| 0,437683 | 280 | 262,9328 | 0,661976 | 1 | 0,908644 |
| -2,11532 | 421 | 356,1883 | 0,035097 | 1 | 0,215003 |
| -1,99095 | 421 | 192,6606 | 0,047899 | 1 | 0,286754 |
| 0,966472 | 420 | 392,2993 | 0,334404 | 1 | 0,947373 |
| -1,98782 | 367 | 348,067 | 0,047615 | 1 | 0,311448 |
| -3,31455 | 280 | 263,3553 | 0,001047 | 1 | 0,121989 |
| -1,62552 | 421 | 331,8978 | 0,105001 | 1 | 0,347821 |
| -0,637 | 421 | 183,7845 | 0,52492 | 1 | 0,766525 |
| 0,866757 | 420 | 370,5091 | 0,386636 | 1 | 0,95743 |
| 0,018831 | 367 | 329,5335 | 0,984987 | 1 | 0,996248 |
| 0,466337 | 280 | 264,547 | 0,641358 | 1 | 0,907537 |
| 0,13197 | 421 | 353,2736 | 0,895083 | 1 | 0,962 |
| -0,62155 | 421 | 190,7693 | 0,534982 | 1 | 0,770763 |
| -0,94946 | 420 | 386,3412 | 0,34298 | 1 | 0,947373 |
| 2,432857 | 367 | 348,6969 | 0,015482 | 1 | 0,204372 |
| 0,074049 | 280 | 257,4162 | 0,941029 | 1 | 0,982175 |
| 1,857883 | 421 | 393,0786 | 0,063933 | 1 | 0,275801 |
| 2,094054 | 421 | 210,1981 | 0,037454 | 1 | 0,266419 |
| 0,78516 | 420 | 413,5489 | 0,43281 | 1 | 0,960096 |
| -0,27916 | 367 | 359,2735 | 0,780283 | 1 | 0,94337 |
| 0,52577 | 280 | 273,5221 | 0,599474 | 1 | 0,881231 |
| -2,25354 | 421 | 368,3868 | 0,024813 | 1 | 0,183798 |
| -0,75334 | 421 | 196,1169 | 0,452147 | 1 | 0,714241 |
| 0,350172 | 420 | 398,2541 | 0,726395 | 1 | 0,969427 |
| -0,20051 | 367 | 352,9896 | 0,841199 | 1 | 0,956935 |
| -1,05307 | 280 | 270,209 | 0,293248 | 1 | 0,723282 |
| -0,88677 | 421 | 287,1334 | 0,375947 | 1 | 0,669684 |
| 0,165881 | 421 | 176,2616 | 0,868441 | 1 | 0,947676 |
| -0,14444 | 420 | 319,9572 | 0,885247 | 1 | 0,982463 |
| -0,15673 | 367 | 288,8031 | 0,875564 | 1 | 0,962417 |
| 2,719482 | 280 | 247,4749 | 0,007002 | 1 | 0,195931 |
| -0,93311 | 421 | 325,4759 | 0,351455 | 1 | 0,640439 |
| -1,25056 | 421 | 184,1218 | 0,212682 | 1 | 0,512644 |
| -2,36161 | 420 | 353,1087 | 0,018738 | 1 | 0,75904 |
| -0,27241 | 367 | 307,9662 | 0,785487 | 1 | 0,94337 |
| -0,77767 | 280 | 254,032 | 0,43749 | 1 | 0,806974 |
| -0,34802 | 421 | 352,9749 | 0,728036 | 1 | 0,890807 |
| -1,32938 | 421 | 191,8993 | 0,1853 | 1 | 0,473945 |
| -1,05726 | 420 | 384,5791 | 0,291058 | 1 | 0,943897 |
| 0,427824 | 367 | 342,6991 | 0,669048 | 1 | 0,895431 |

| | | | | | |
|---|---|---|---|---|---|
| 1,816641 | 280 | 262,7143 | 0,070412 | 1 | 0,433103 |
| 0,141296 | 421 | 361,3676 | 0,887715 | 1 | 0,962 |
| 0,597348 | 421 | 193,3846 | 0,550974 | 1 | 0,777451 |
| -0,10113 | 420 | 393,4093 | 0,919495 | 1 | 0,989204 |
| 0,375322 | 367 | 334,8981 | 0,707658 | 1 | 0,91071 |
| 0,793843 | 280 | 266,5049 | 0,427993 | 1 | 0,800834 |
| -0,7213 | 421 | 375,5647 | 0,471171 | 1 | 0,731548 |
| -0,1122 | 421 | 199,4544 | 0,910775 | 1 | 0,96646 |
| -0,44042 | 420 | 403,4081 | 0,659868 | 1 | 0,96781 |
| -0,40195 | 367 | 343,0286 | 0,687975 | 1 | 0,903032 |
| -1,87233 | 280 | 265,1789 | 0,062261 | 1 | 0,422605 |
| -1,132 | 421 | 390,8249 | 0,258331 | 1 | 0,550898 |
| -1,90784 | 421 | 209,8052 | 0,057777 | 1 | 0,312877 |
| -0,76264 | 420 | 412,3269 | 0,446114 | 1 | 0,960096 |
| -2,63408 | 367 | 359,6659 | 0,008801 | 1 | 0,183886 |
| -2,22176 | 280 | 273,5982 | 0,027118 | 1 | 0,324814 |
| -0,19993 | 421 | 335,3791 | 0,841657 | 1 | 0,939539 |
| 1,835992 | 421 | 186,84 | 0,067948 | 1 | 0,334428 |
| 0,312559 | 420 | 370,6029 | 0,754792 | 1 | 0,969427 |
| 2,341363 | 367 | 324,207 | 0,019819 | 1 | 0,221237 |
| 2,338954 | 280 | 255,4978 | 0,020109 | 1 | 0,297103 |
| -2,69925 | 421 | 344,4485 | 0,007292 | 1 | 0,105763 |
| -1,8651 | 421 | 190,6514 | 0,063704 | 1 | 0,323237 |
| 0,777354 | 420 | 383,0462 | 0,43743 | 1 | 0,960096 |
| -2,36772 | 367 | 347,232 | 0,018446 | 1 | 0,217108 |
| -2,01922 | 280 | 264,8427 | 0,044471 | 1 | 0,387749 |
| 0,731426 | 421 | 283,8612 | 0,465123 | 1 | 0,727964 |
| 1,135226 | 421 | 172,7288 | 0,257853 | 1 | 0,556025 |
| 0,904263 | 420 | 308,3707 | 0,366562 | 1 | 0,95743 |
| 0,671699 | 367 | 279,0193 | 0,502331 | 1 | 0,793601 |
| 1,302009 | 280 | 276 | 0,193999 | 1 | 0,62543 |
| -0,50257 | 421 | 350,8591 | 0,615581 | 1 | 0,826526 |
| 1,668154 | 421 | 189,6227 | 0,096936 | 1 | 0,379251 |
| 1,098814 | 420 | 385,5456 | 0,272535 | 1 | 0,943897 |
| 0,765494 | 367 | 334,071 | 0,444518 | 1 | 0,749316 |
| 1,246095 | 280 | 266,3134 | 0,213825 | 1 | 0,646827 |
| -0,81864 | 421 | 316,9526 | 0,413607 | 1 | 0,695339 |
| 1,035505 | 421 | 181,2862 | 0,301812 | 1 | 0,600041 |
| 2,003062 | 420 | 348,4646 | 0,045945 | 1 | 0,846132 |
| -0,19055 | 367 | 299,8225 | 0,849003 | 1 | 0,956935 |
| 0,061364 | 280 | 251,5822 | 0,951118 | 1 | 0,982409 |
| -0,04278 | 421 | 371,5628 | 0,965898 | 1 | 0,992572 |
| 0,408421 | 421 | 198,1204 | 0,683406 | 1 | 0,859821 |
| 0,31469 | 420 | 400,3488 | 0,753161 | 1 | 0,969427 |
| -0,39265 | 367 | 350,7233 | 0,694815 | 1 | 0,904597 |
| 0,659131 | 280 | 271,3076 | 0,51037 | 1 | 0,831912 |
| -2,25331 | 421 | 337,8528 | 0,02488 | 1 | 0,183798 |
| -3,26601 | 421 | 188,6784 | 0,001296 | 1 | 0,092971 |

| | | | | | |
|---|---|---|---|---|---|
| -0,40683 | 420 | 378,1723 | 0,68436 | 1 | 0,968613 |
| -3,38045 | 367 | 324,2815 | 8,12E-04 | 0,871337 | 0,139974 |
| -2,20696 | 280 | 266,978 | 0,028168 | 1 | 0,326641 |
| -0,47098 | 421 | 346,4216 | 0,637952 | 1 | 0,834842 |
| -1,70687 | 421 | 187,6887 | 0,0895 | 1 | 0,37156 |
| -0,69982 | 420 | 380,8897 | 0,484466 | 1 | 0,960096 |
| -1,26788 | 367 | 325,1377 | 0,205746 | 1 | 0,558434 |
| -2,44371 | 280 | 255,8553 | 0,015213 | 1 | 0,263522 |
| -1,41966 | 421 | 357,6599 | 0,156577 | 1 | 0,429212 |
| -1,04061 | 421 | 192,9375 | 0,299358 | 1 | 0,597527 |
| -0,03483 | 420 | 391,7902 | 0,972232 | 1 | 0,997342 |
| -0,08422 | 367 | 335,774 | 0,932929 | 1 | 0,977109 |
| -1,77101 | 280 | 256,7253 | 0,077746 | 1 | 0,453386 |
| -0,84507 | 421 | 354,6055 | 0,398644 | 1 | 0,688151 |
| -0,29876 | 421 | 191,6305 | 0,765444 | 1 | 0,90393 |
| -0,15656 | 420 | 387,1451 | 0,875674 | 1 | 0,982463 |
| -0,45004 | 367 | 333,1684 | 0,652975 | 1 | 0,888919 |
| -1,71451 | 280 | 261,7792 | 0,087618 | 1 | 0,460069 |
| -1,38948 | 421 | 367,0063 | 0,165529 | 1 | 0,440727 |
| -1,88933 | 421 | 196,7056 | 0,060319 | 1 | 0,314727 |
| -0,04496 | 420 | 398,7057 | 0,964162 | 1 | 0,996924 |
| -1,88657 | 367 | 345,1932 | 0,060057 | 1 | 0,340409 |
| -1,34108 | 280 | 264,9777 | 0,181042 | 1 | 0,607861 |
| 0,261922 | 421 | 395,7989 | 0,793518 | 1 | 0,919136 |
| 0,76316 | 421 | 213,1282 | 0,446212 | 1 | 0,710076 |
| 1,495627 | 420 | 414,0236 | 0,135513 | 1 | 0,891179 |
| -0,38572 | 367 | 359,3718 | 0,69993 | 1 | 0,906491 |
| -0,93881 | 280 | 272,2895 | 0,348663 | 1 | 0,761841 |
| -0,35763 | 421 | 314,6208 | 0,720858 | 1 | 0,885357 |
| -0,53023 | 421 | 182,3682 | 0,596596 | 1 | 0,811435 |
| -0,52817 | 420 | 349,0733 | 0,597715 | 1 | 0,964878 |
| 0,857308 | 367 | 311,627 | 0,391933 | 1 | 0,705088 |
| 1,000655 | 280 | 276 | 0,31787 | 1 | 0,741533 |
| -0,91895 | 421 | 368,8713 | 0,358721 | 1 | 0,648267 |
| -0,14687 | 421 | 197,9958 | 0,883383 | 1 | 0,957051 |
| 0,941619 | 420 | 398,6145 | 0,346958 | 1 | 0,947373 |
| 0,909762 | 367 | 343,072 | 0,363587 | 1 | 0,677287 |
| -0,25474 | 280 | 265,1992 | 0,799121 | 1 | 0,954985 |
| 0,409473 | 421 | 323,9817 | 0,682463 | 1 | 0,862088 |
| 0,076739 | 421 | 181,9285 | 0,938915 | 1 | 0,977847 |
| 1,762433 | 420 | 359,2124 | 0,078847 | 1 | 0,848246 |
| 0,462136 | 367 | 302,9838 | 0,644316 | 1 | 0,882712 |
| 0,243487 | 280 | 251,6219 | 0,807827 | 1 | 0,95747 |
| 1,130128 | 421 | 335,0888 | 0,25923 | 1 | 0,55178 |
| 1,572576 | 421 | 187,07 | 0,117507 | 1 | 0,396436 |
| 0,190135 | 420 | 370,3822 | 0,849308 | 1 | 0,973894 |
| 0,935356 | 367 | 319,5188 | 0,350311 | 1 | 0,669099 |
| 1,040195 | 280 | 255,4586 | 0,299233 | 1 | 0,723324 |

| | | | | | |
|---|---|---|---|---|---|
| 0,56233 | 421 | 366,6219 | 0,574235 | 1 | 0,800529 |
| 0,143955 | 421 | 197,6738 | 0,885682 | 1 | 0,957782 |
| 0,766971 | 420 | 397,0717 | 0,443555 | 1 | 0,960096 |
| -1,96964 | 367 | 346,9822 | 0,049675 | 1 | 0,315285 |
| -0,54379 | 280 | 258,9062 | 0,587052 | 1 | 0,87561 |
| -1,04371 | 421 | 340,3536 | 0,297362 | 1 | 0,59543 |
| -0,11539 | 421 | 186,4953 | 0,908258 | 1 | 0,96646 |
| 0,990169 | 420 | 372,862 | 0,322733 | 1 | 0,945573 |
| 1,088558 | 367 | 325,1329 | 0,277155 | 1 | 0,611884 |
| -0,97657 | 280 | 260,3237 | 0,329689 | 1 | 0,752282 |
| -4,04619 | 421 | 328,9785 | 6,49E-05 | 0,069863 | 0,024462 |
| -2,52768 | 421 | 186,1954 | 0,012313 | 1 | 0,169272 |
| -1,71385 | 420 | 375,152 | 0,087382 | 1 | 0,848246 |
| -2,82787 | 367 | 337,4379 | 0,004966 | 1 | 0,178183 |
| -2,5821 | 280 | 267,7389 | 0,010352 | 1 | 0,229419 |
| -0,67424 | 421 | 335,5566 | 0,50062 | 1 | 0,754435 |
| -1,78988 | 421 | 185,4961 | 0,075104 | 1 | 0,345067 |
| -0,69 | 420 | 369,4851 | 0,490629 | 1 | 0,960096 |
| -2,18869 | 367 | 321,0782 | 0,029339 | 1 | 0,260082 |
| -2,23974 | 280 | 251,7987 | 0,025981 | 1 | 0,324814 |
| 0,414468 | 421 | 330,0091 | 0,678801 | 1 | 0,861268 |
| 0,236575 | 421 | 183,9565 | 0,81325 | 1 | 0,926063 |
| -0,10138 | 420 | 364,9369 | 0,919302 | 1 | 0,989204 |
| 0,558465 | 367 | 316,7155 | 0,576921 | 1 | 0,843313 |
| -1,26083 | 280 | 266,186 | 0,208475 | 1 | 0,64091 |
| -1,97497 | 421 | 350,5067 | 0,049056 | 1 | 0,244581 |
| -2,50129 | 421 | 191,5965 | 0,013212 | 1 | 0,171643 |
| -1,22303 | 420 | 387,5586 | 0,222063 | 1 | 0,943055 |
| -1,7282 | 367 | 342,9994 | 0,084853 | 1 | 0,388246 |
| -1,64905 | 280 | 269,9372 | 0,100301 | 1 | 0,480557 |
| -0,67948 | 421 | 309,432 | 0,49734 | 1 | 0,750993 |
| -0,13363 | 421 | 178,9866 | 0,893842 | 1 | 0,958882 |
| -0,36607 | 420 | 341,1694 | 0,714536 | 1 | 0,968866 |
| -0,80493 | 367 | 323,2478 | 0,421451 | 1 | 0,731532 |
| -0,38009 | 280 | 256,2867 | 0,704191 | 1 | 0,921995 |
| -1,64476 | 421 | 312,5773 | 0,101024 | 1 | 0,342121 |
| -1,88999 | 421 | 181,2535 | 0,060356 | 1 | 0,314727 |
| -0,62005 | 420 | 347,4214 | 0,535631 | 1 | 0,960096 |
| -2,01486 | 367 | 296,1231 | 0,044822 | 1 | 0,302719 |
| -2,51577 | 280 | 253,2756 | 0,012497 | 1 | 0,244579 |
| 0,602403 | 421 | 357,4946 | 0,547288 | 1 | 0,784719 |
| -1,2555 | 421 | 191,8172 | 0,210823 | 1 | 0,509968 |
| 0,200964 | 420 | 389,9363 | 0,840832 | 1 | 0,973894 |
| 0,937836 | 367 | 341,4636 | 0,348992 | 1 | 0,668671 |
| 1,415705 | 280 | 265,2392 | 0,158035 | 1 | 0,576656 |
| -0,51392 | 421 | 350,8104 | 0,60763 | 1 | 0,824024 |
| -0,50199 | 421 | 190,7923 | 0,616252 | 1 | 0,825504 |
| -0,81076 | 420 | 383,8373 | 0,418005 | 1 | 0,95743 |

| | | | | | |
|---|---|---|---|---|---|
| -0,12415 | 367 | 338,7143 | 0,901267 | 1 | 0,970486 |
| -0,22594 | 280 | 264,4403 | 0,821422 | 1 | 0,9596 |
| 1,774516 | 421 | 320,6921 | 0,076927 | 1 | 0,301112 |
| 0,331245 | 421 | 181,8622 | 0,740841 | 1 | 0,886773 |
| -0,9582 | 420 | 355,521 | 0,338611 | 1 | 0,947373 |
| 1,592177 | 367 | 305,9295 | 0,112378 | 1 | 0,438296 |
| 0,750009 | 280 | 255,7471 | 0,453939 | 1 | 0,814707 |
| 2,228716 | 421 | 374,7479 | 0,026425 | 1 | 0,191457 |
| 1,3156 | 421 | 200,2181 | 0,189813 | 1 | 0,476746 |
| 0,201009 | 420 | 403,7613 | 0,840793 | 1 | 0,973894 |
| 2,325808 | 367 | 354,8049 | 0,020592 | 1 | 0,224016 |
| 1,042974 | 280 | 271,524 | 0,297888 | 1 | 0,723324 |
| 2,270672 | 421 | 346,1407 | 0,023781 | 1 | 0,180089 |
| 1,66235 | 421 | 185,3619 | 0,098133 | 1 | 0,379251 |
| 0,923318 | 420 | 379,0737 | 0,356429 | 1 | 0,947373 |
| 2,486971 | 367 | 331,3743 | 0,013375 | 1 | 0,193812 |
| 1,407606 | 280 | 263,9471 | 0,160424 | 1 | 0,581516 |
| -1,26923 | 421 | 333,3075 | 0,205244 | 1 | 0,494095 |
| -1,7355 | 421 | 184,2122 | 0,084325 | 1 | 0,365475 |
| -2,71969 | 420 | 362,0129 | 0,006849 | 1 | 0,543263 |
| -2,2295 | 367 | 312,4065 | 0,026492 | 1 | 0,252225 |
| -0,54711 | 280 | 254,1161 | 0,58478 | 1 | 0,87561 |
| -0,08927 | 421 | 408,2323 | 0,928908 | 1 | 0,970107 |
| -1,12143 | 421 | 221,7567 | 0,263319 | 1 | 0,559693 |
| 0,646482 | 420 | 415,8196 | 0,518324 | 1 | 0,960096 |
| 1,012041 | 367 | 362,6459 | 0,312193 | 1 | 0,646834 |
| 1,361298 | 280 | 274,4279 | 0,174536 | 1 | 0,598625 |
| -0,79841 | 421 | 366,6066 | 0,425151 | 1 | 0,6997 |
| -1,85637 | 421 | 193,8972 | 0,064918 | 1 | 0,325826 |
| -0,49937 | 420 | 401,4341 | 0,617794 | 1 | 0,964878 |
| -0,96388 | 367 | 347,479 | 0,335776 | 1 | 0,662363 |
| 0,054842 | 280 | 263,2025 | 0,956306 | 1 | 0,985067 |
| 1,328401 | 421 | 337,9206 | 0,184942 | 1 | 0,46885 |
| 1,643514 | 421 | 187,5851 | 0,101952 | 1 | 0,383322 |
| -0,06109 | 420 | 372,0542 | 0,951317 | 1 | 0,992811 |
| 0,996303 | 367 | 323,2337 | 0,319848 | 1 | 0,653184 |
| -0,14281 | 280 | 255,5575 | 0,886551 | 1 | 0,973784 |
| 1,81519 | 421 | 335,9588 | 0,070386 | 1 | 0,28514 |
| -1,01736 | 421 | 185,8871 | 0,310302 | 1 | 0,607306 |
| -0,59288 | 420 | 368,827 | 0,553628 | 1 | 0,960096 |
| 0,986939 | 367 | 330,8602 | 0,324394 | 1 | 0,655597 |
| 0,621728 | 280 | 261,28 | 0,534663 | 1 | 0,84326 |
| -1,32312 | 421 | 354,9258 | 0,186647 | 1 | 0,471942 |
| -1,71948 | 421 | 193,0714 | 0,08713 | 1 | 0,3678 |
| 1,577949 | 420 | 389,854 | 0,115388 | 1 | 0,848246 |
| -0,99588 | 367 | 340,7807 | 0,320014 | 1 | 0,653184 |
| -0,12937 | 280 | 265,8672 | 0,897165 | 1 | 0,975243 |
| -0,81829 | 421 | 368,6026 | 0,413722 | 1 | 0,695339 |

| | | | | | |
|---|---|---|---|---|---|
| 0,472468 | 421 | 197,3467 | 0,637115 | 1 | 0,837812 |
| -0,07773 | 420 | 398,5184 | 0,938078 | 1 | 0,992059 |
| 1,811214 | 367 | 340,8888 | 0,070988 | 1 | 0,366365 |
| 0,90007 | 280 | 261,9436 | 0,36891 | 1 | 0,763914 |
| 0,949901 | 421 | 331,5542 | 0,342854 | 1 | 0,63164 |
| -0,31516 | 421 | 185,1068 | 0,752993 | 1 | 0,893512 |
| 0,828022 | 420 | 364,5783 | 0,4082 | 1 | 0,95743 |
| -0,74097 | 367 | 319,457 | 0,459258 | 1 | 0,763917 |
| -0,35733 | 280 | 276 | 0,721119 | 1 | 0,932812 |
| 0,058531 | 421 | 339,5915 | 0,95336 | 1 | 0,986076 |
| 0,09117 | 421 | 185,8756 | 0,927455 | 1 | 0,972749 |
| -0,36842 | 420 | 375,1218 | 0,712767 | 1 | 0,968866 |
| 1,292625 | 367 | 306,3794 | 0,197115 | 1 | 0,549567 |
| -0,98334 | 280 | 257,2743 | 0,326365 | 1 | 0,749743 |
| 1,121052 | 421 | 341,3032 | 0,263054 | 1 | 0,556772 |
| -0,16887 | 421 | 188,1056 | 0,86608 | 1 | 0,947215 |
| -1,69635 | 420 | 377,4387 | 0,090645 | 1 | 0,848246 |
| -1,62576 | 367 | 325,7861 | 0,104967 | 1 | 0,429852 |
| 0,155597 | 280 | 260,398 | 0,876471 | 1 | 0,972638 |
| -0,3761 | 421 | 381,2359 | 0,707054 | 1 | 0,875021 |
| 0,138103 | 421 | 201,3449 | 0,890297 | 1 | 0,958882 |
| -0,46836 | 420 | 406,6291 | 0,639778 | 1 | 0,964878 |
| 1,091093 | 367 | 357,6018 | 0,275966 | 1 | 0,611884 |
| -1,09782 | 280 | 268,7893 | 0,273267 | 1 | 0,701156 |
| 0,067608 | 421 | 368,7192 | 0,946134 | 1 | 0,981297 |
| 1,922494 | 421 | 197,3852 | 0,055983 | 1 | 0,309036 |
| 1,274761 | 420 | 397,3238 | 0,203139 | 1 | 0,928589 |
| 2,051097 | 367 | 340,1804 | 0,041023 | 1 | 0,289956 |
| 0,468518 | 280 | 266,0078 | 0,639798 | 1 | 0,907449 |
| -2,0897 | 421 | 311,0395 | 0,037457 | 1 | 0,217944 |
| -0,62915 | 421 | 179,9116 | 0,530046 | 1 | 0,768955 |
| 0,514305 | 420 | 347,2677 | 0,607366 | 1 | 0,964878 |
| -1,46784 | 367 | 320,5722 | 0,143129 | 1 | 0,493583 |
| -0,60402 | 280 | 249,9751 | 0,54638 | 1 | 0,851507 |
| -2,54382 | 421 | 332,2248 | 0,011417 | 1 | 0,128401 |
| -2,05372 | 421 | 185,0155 | 0,041409 | 1 | 0,272972 |
| -0,32632 | 420 | 371,2088 | 0,744367 | 1 | 0,969427 |
| -1,74557 | 367 | 325,5745 | 0,081828 | 1 | 0,381841 |
| -2,76723 | 280 | 265,4731 | 0,006051 | 1 | 0,186692 |
| -2,48777 | 421 | 354,9916 | 0,013313 | 1 | 0,138362 |
| -2,25622 | 421 | 189,6077 | 0,025199 | 1 | 0,218351 |
| -1,61397 | 420 | 382,154 | 0,107358 | 1 | 0,848246 |
| -3,82905 | 367 | 331,8493 | 1,54E-04 | 0,165612 | 0,053579 |
| -2,18667 | 280 | 269,9561 | 0,029625 | 1 | 0,32827 |
| -0,07855 | 421 | 344,426 | 0,937433 | 1 | 0,974957 |
| -0,53516 | 421 | 186,4229 | 0,59318 | 1 | 0,808395 |
| -1,12301 | 420 | 374,5326 | 0,262154 | 1 | 0,943897 |
| 2,951486 | 367 | 349,0045 | 0,003376 | 1 | 0,176492 |

| | | | | | |
|---|---|---|---|---|---|
| 2,463375 | 280 | 259,5321 | 0,014413 | 1 | 0,258576 |
| 1,288869 | 421 | 341,2887 | 0,198317 | 1 | 0,484381 |
| 0,338544 | 421 | 187,3842 | 0,735332 | 1 | 0,884613 |
| -1,46049 | 420 | 377,2333 | 0,144988 | 1 | 0,914005 |
| 1,629345 | 367 | 321,3932 | 0,10422 | 1 | 0,429123 |
| -0,26651 | 280 | 255,8155 | 0,790063 | 1 | 0,953435 |
| 0,79686 | 421 | 365,4331 | 0,42605 | 1 | 0,700145 |
| 0,246833 | 421 | 196,4325 | 0,805296 | 1 | 0,923205 |
| 0,060849 | 420 | 395,3108 | 0,95151 | 1 | 0,992811 |
| -0,14167 | 367 | 342,7618 | 0,887424 | 1 | 0,964898 |
| -1,02082 | 280 | 261,4639 | 0,308286 | 1 | 0,729704 |
| 0,691906 | 421 | 396,9272 | 0,489401 | 1 | 0,748388 |
| 2,14789 | 421 | 213,0248 | 0,032851 | 1 | 0,246304 |
| 1,220487 | 420 | 413,5438 | 0,222976 | 1 | 0,943055 |
| 0,410183 | 367 | 362,1412 | 0,681914 | 1 | 0,900574 |
| 1,861249 | 280 | 270,4601 | 0,063794 | 1 | 0,422605 |
| -0,64576 | 421 | 330,4593 | 0,518882 | 1 | 0,768129 |
| -0,00884 | 421 | 185,1619 | 0,992956 | 1 | 0,997935 |
| -0,38656 | 420 | 363,4756 | 0,699305 | 1 | 0,968866 |
| -0,05747 | 367 | 320,5735 | 0,95421 | 1 | 0,980228 |
| 0,028636 | 280 | 257,7767 | 0,977177 | 1 | 0,99236 |
| -0,59036 | 421 | 353,0911 | 0,555324 | 1 | 0,790572 |
| 0,655302 | 421 | 190,7982 | 0,513063 | 1 | 0,756803 |
| 0,174959 | 420 | 386,5669 | 0,861204 | 1 | 0,977284 |
| 1,097705 | 367 | 335,0321 | 0,273121 | 1 | 0,609149 |
| 1,574665 | 280 | 265,1352 | 0,116526 | 1 | 0,518958 |
| -0,25475 | 421 | 348,4399 | 0,799064 | 1 | 0,922616 |
| -1,53221 | 421 | 190,8108 | 0,127127 | 1 | 0,410224 |
| -0,7304 | 420 | 383,0725 | 0,46559 | 1 | 0,960096 |
| 0,56063 | 367 | 340,0523 | 0,575419 | 1 | 0,842717 |
| -1,53482 | 280 | 269,8638 | 0,125999 | 1 | 0,536498 |
| 1,854748 | 421 | 386,1749 | 0,064394 | 1 | 0,275801 |
| 2,014013 | 421 | 204,1704 | 0,045321 | 1 | 0,281076 |
| 1,082317 | 420 | 409,0654 | 0,279749 | 1 | 0,943897 |
| 0,3872 | 367 | 360,1351 | 0,698837 | 1 | 0,906323 |
| -0,9201 | 280 | 273,6399 | 0,358332 | 1 | 0,761846 |
| -3,60145 | 421 | 338,47 | 3,64E-04 | 0,38893 | 0,04642 |
| -0,80498 | 421 | 187,0223 | 0,421851 | 1 | 0,692515 |
| 0,295499 | 420 | 373,8959 | 0,767777 | 1 | 0,969427 |
| -1,43354 | 367 | 315,6348 | 0,152692 | 1 | 0,500233 |
| -1,11893 | 280 | 259,1323 | 0,264207 | 1 | 0,688875 |
| -1,839 | 421 | 341,9035 | 0,066782 | 1 | 0,278784 |
| -1,32722 | 421 | 187,2556 | 0,18605 | 1 | 0,474401 |
| -1,74921 | 420 | 373,5219 | 0,081075 | 1 | 0,848246 |
| 2,808777 | 367 | 348,2005 | 0,005254 | 1 | 0,178183 |
| 1,443464 | 280 | 260,1617 | 0,150093 | 1 | 0,562661 |
| -1,00797 | 421 | 350,6983 | 0,314162 | 1 | 0,608538 |
| -0,23198 | 421 | 190,4658 | 0,816803 | 1 | 0,926377 |

| | | | | | |
|---|---|---|---|---|---|
| -0,68365 | 420 | 383,0028 | 0,49461 | 1 | 0,960096 |
| 2,076894 | 367 | 349,263 | 0,038542 | 1 | 0,28956 |
| 0,900325 | 280 | 261,5252 | 0,368776 | 1 | 0,763914 |
| -2,02497 | 421 | 347,2718 | 0,043635 | 1 | 0,235324 |
| -2,75287 | 421 | 191,0249 | 0,006477 | 1 | 0,141161 |
| -0,5019 | 420 | 384,0948 | 0,616028 | 1 | 0,964878 |
| 0,544559 | 367 | 345,2714 | 0,586408 | 1 | 0,849728 |
| -0,18188 | 280 | 266,8528 | 0,855814 | 1 | 0,966655 |
| -0,24068 | 421 | 348,2149 | 0,809946 | 1 | 0,924931 |
| 1,064845 | 421 | 189,1134 | 0,288304 | 1 | 0,585544 |
| -0,01087 | 420 | 381,7734 | 0,991334 | 1 | 0,997342 |
| -1,25402 | 367 | 329,1897 | 0,210723 | 1 | 0,563436 |
| 0,903259 | 280 | 260,9181 | 0,367222 | 1 | 0,763914 |
| -1,1459 | 421 | 311,0884 | 0,252718 | 1 | 0,546261 |
| -0,97123 | 421 | 178,6671 | 0,332749 | 1 | 0,626816 |
| -0,46823 | 420 | 347,3977 | 0,639914 | 1 | 0,964878 |
| -1,21364 | 367 | 313,4474 | 0,225798 | 1 | 0,576775 |
| -2,139 | 280 | 247,9397 | 0,033413 | 1 | 0,329568 |
| -0,80571 | 421 | 416,9813 | 0,420871 | 1 | 0,697735 |
| -1,59079 | 421 | 251,0584 | 0,112916 | 1 | 0,391524 |
| 0,130622 | 420 | 405,752 | 0,896139 | 1 | 0,98503 |
| 0,185995 | 367 | 359,1042 | 0,852554 | 1 | 0,956935 |
| 0,683029 | 280 | 275,8965 | 0,495161 | 1 | 0,8282 |
| -2,27906 | 421 | 359,3757 | 0,023249 | 1 | 0,177419 |
| -3,02072 | 421 | 194,3238 | 0,002861 | 1 | 0,114447 |
| -0,09093 | 420 | 395,464 | 0,927592 | 1 | 0,990001 |
| -2,22442 | 367 | 339,1708 | 0,026778 | 1 | 0,252225 |
| -1,32245 | 280 | 265,8999 | 0,187156 | 1 | 0,621241 |
| -2,04399 | 421 | 351,2835 | 0,0417 | 1 | 0,228553 |
| -1,94168 | 421 | 191,4843 | 0,053644 | 1 | 0,302776 |
| -0,41327 | 420 | 383,6531 | 0,679641 | 1 | 0,96781 |
| -0,5934 | 367 | 346,8728 | 0,5533 | 1 | 0,830257 |
| -1,20314 | 280 | 268,6839 | 0,229982 | 1 | 0,657169 |
| 0,445833 | 421 | 334,1353 | 0,656007 | 1 | 0,846936 |
| 1,635339 | 421 | 185,5829 | 0,103673 | 1 | 0,383322 |
| -0,89268 | 420 | 369,4789 | 0,372611 | 1 | 0,95743 |
| 1,338663 | 367 | 319,1379 | 0,181633 | 1 | 0,527769 |
| 0,459503 | 280 | 260,2188 | 0,646257 | 1 | 0,907804 |
| 2,841663 | 421 | 330,7366 | 0,004766 | 1 | 0,087964 |
| 1,404726 | 421 | 182,9058 | 0,161799 | 1 | 0,445827 |
| 1,38226 | 420 | 364,1698 | 0,167739 | 1 | 0,914005 |
| 1,559802 | 367 | 309,5582 | 0,119828 | 1 | 0,451453 |
| -0,12947 | 280 | 260,8681 | 0,897089 | 1 | 0,975243 |
| -3,23705 | 421 | 340,4617 | 0,001326 | 1 | 0,066922 |
| -2,25766 | 421 | 188,3073 | 0,025115 | 1 | 0,218351 |
| -0,70875 | 420 | 378,5973 | 0,478913 | 1 | 0,960096 |
| -1,05774 | 367 | 336,9474 | 0,290931 | 1 | 0,627013 |
| -0,1479 | 280 | 260,5063 | 0,882538 | 1 | 0,973784 |

| | | | | | |
|---|---|---|---|---|---|
| -0,53218 | 421 | 329,1862 | 0,594961 | 1 | 0,813756 |
| -1,44767 | 421 | 184,6275 | 0,149406 | 1 | 0,430506 |
| 0,224736 | 420 | 364,1976 | 0,822311 | 1 | 0,972623 |
| -1,84041 | 367 | 324,3387 | 0,066621 | 1 | 0,359849 |
| -4,11389 | 280 | 251,8654 | 5,27E-05 | 0,056893 | 0,046455 |
| 0,505663 | 421 | 332,9748 | 0,613428 | 1 | 0,825172 |
| -0,61154 | 421 | 180,3442 | 0,541611 | 1 | 0,773814 |
| 0,186718 | 420 | 367,1156 | 0,851985 | 1 | 0,973894 |
| 0,756614 | 367 | 317,8236 | 0,449842 | 1 | 0,755345 |
| 0,190779 | 280 | 251,9783 | 0,848852 | 1 | 0,966155 |
| 1,193628 | 421 | 383,3443 | 0,233362 | 1 | 0,525147 |
| 2,502968 | 421 | 203,9761 | 0,0131 | 1 | 0,171643 |
| 2,106401 | 420 | 406,3069 | 0,035782 | 1 | 0,828713 |
| 3,314459 | 367 | 348,0373 | 0,001015 | 1 | 0,152933 |
| 1,920537 | 280 | 270,3348 | 0,055842 | 1 | 0,40654 |
| -0,15206 | 421 | 356,1079 | 0,879223 | 1 | 0,95861 |
| -0,09922 | 421 | 192,7918 | 0,92107 | 1 | 0,970834 |
| -1,23417 | 420 | 386,9998 | 0,21789 | 1 | 0,943055 |
| -0,95535 | 367 | 334,1067 | 0,340092 | 1 | 0,664831 |
| 0,836071 | 280 | 262,2781 | 0,403876 | 1 | 0,786359 |
| -1,0754 | 421 | 347,6763 | 0,282941 | 1 | 0,582502 |
| 0,747878 | 421 | 190,0701 | 0,455458 | 1 | 0,716467 |
| 0,26888 | 420 | 385,3763 | 0,788166 | 1 | 0,969427 |
| 1,510875 | 367 | 336,4488 | 0,131759 | 1 | 0,475026 |
| 0,425472 | 280 | 260,0613 | 0,670844 | 1 | 0,908644 |
| -0,98551 | 421 | 344,0562 | 0,325064 | 1 | 0,61749 |
| 0,256858 | 421 | 188,7735 | 0,797568 | 1 | 0,921861 |
| 0,628538 | 420 | 378,1029 | 0,530031 | 1 | 0,960096 |
| 0,75618 | 367 | 318,9055 | 0,450099 | 1 | 0,755345 |
| -0,41271 | 280 | 259,0036 | 0,680158 | 1 | 0,91111 |
| -0,80376 | 421 | 293,4985 | 0,422185 | 1 | 0,697735 |
| -1,29284 | 421 | 177,5398 | 0,197746 | 1 | 0,48773 |
| -1,03857 | 420 | 321,8928 | 0,299786 | 1 | 0,943897 |
| -0,11671 | 367 | 294,1079 | 0,907169 | 1 | 0,970486 |
| -2,07329 | 280 | 276 | 0,039073 | 1 | 0,356865 |
| -2,24739 | 421 | 335,7226 | 0,025264 | 1 | 0,184916 |
| -1,47544 | 421 | 185,8554 | 0,141785 | 1 | 0,423499 |
| -1,35067 | 420 | 368,415 | 0,177632 | 1 | 0,9199 |
| -0,66652 | 367 | 331,6858 | 0,505545 | 1 | 0,793601 |
| -0,3756 | 280 | 263,4639 | 0,70752 | 1 | 0,923145 |
| -1,38592 | 421 | 342,4892 | 0,166674 | 1 | 0,441901 |
| -1,08946 | 421 | 189,4463 | 0,277336 | 1 | 0,578094 |
| -1,29383 | 420 | 383,0216 | 0,196502 | 1 | 0,928589 |
| -1,12106 | 367 | 340,8719 | 0,26305 | 1 | 0,609149 |
| -0,96621 | 280 | 264,5862 | 0,334821 | 1 | 0,755731 |
| 2,016783 | 421 | 340,4293 | 0,044503 | 1 | 0,236315 |
| 2,627433 | 421 | 185,189 | 0,009324 | 1 | 0,14815 |
| 1,655644 | 420 | 380,9008 | 0,098617 | 1 | 0,848246 |

| | | | | | |
|---|---|---|---|---|---|
| 2,352756 | 367 | 311,036 | 0,019257 | 1 | 0,218203 |
| 2,739114 | 280 | 264,7494 | 0,006579 | 1 | 0,192695 |
| -0,46101 | 421 | 385,2281 | 0,645049 | 1 | 0,838007 |
| 0,486037 | 421 | 204,9414 | 0,62746 | 1 | 0,832379 |
| 0,714841 | 420 | 409,9869 | 0,475114 | 1 | 0,960096 |
| -0,33216 | 367 | 359,1034 | 0,73996 | 1 | 0,923669 |
| 1,191517 | 280 | 274,4615 | 0,23448 | 1 | 0,659852 |
| -1,22313 | 421 | 378,6531 | 0,222043 | 1 | 0,51676 |
| -0,39474 | 421 | 201,3339 | 0,693456 | 1 | 0,865526 |
| 0,404245 | 420 | 406,8684 | 0,686245 | 1 | 0,968613 |
| -0,19351 | 367 | 358,9423 | 0,846667 | 1 | 0,956935 |
| -0,01372 | 280 | 266,9092 | 0,989062 | 1 | 0,995004 |
| 0,896184 | 421 | 385,0552 | 0,370714 | 1 | 0,662712 |
| 0,218335 | 421 | 206,4919 | 0,827384 | 1 | 0,933284 |
| -0,98746 | 420 | 412,1729 | 0,323998 | 1 | 0,945573 |
| -0,24371 | 367 | 357,3679 | 0,807598 | 1 | 0,950149 |
| 0,517874 | 280 | 269,3695 | 0,604971 | 1 | 0,885137 |
| -2,59565 | 421 | 372,2138 | 0,009815 | 1 | 0,116484 |
| -2,9746 | 421 | 198,7648 | 0,003297 | 1 | 0,118304 |
| -0,85424 | 420 | 402,1128 | 0,393479 | 1 | 0,95743 |
| -1,57744 | 367 | 355,2218 | 0,115585 | 1 | 0,443225 |
| -1,91159 | 280 | 270,3312 | 0,056986 | 1 | 0,407001 |
| -1,71732 | 421 | 316,6202 | 0,086899 | 1 | 0,315558 |
| -2,75476 | 421 | 180,3404 | 0,006476 | 1 | 0,141161 |
| 0,337297 | 420 | 353,7674 | 0,736093 | 1 | 0,969427 |
| -0,01528 | 367 | 304,6741 | 0,987822 | 1 | 0,996248 |
| 0,866087 | 280 | 256,8101 | 0,387251 | 1 | 0,775016 |
| -0,98522 | 421 | 417 | 0,325086 | 1 | 0,61749 |
| -0,92489 | 421 | 417 | 0,355556 | 1 | 0,643758 |
| -1,17847 | 420 | 416 | 0,239283 | 1 | 0,943659 |
| -0,11704 | 367 | 363 | 0,906892 | 1 | 0,970486 |
| -0,43016 | 280 | 276 | 0,667413 | 1 | 0,908644 |
| 0,136034 | 421 | 313,9816 | 0,891882 | 1 | 0,962 |
| 0,335571 | 421 | 179,5793 | 0,737586 | 1 | 0,884986 |
| 1,079717 | 420 | 342,7709 | 0,281027 | 1 | 0,943897 |
| -0,28496 | 367 | 302,3181 | 0,775868 | 1 | 0,940654 |
| -1,04091 | 280 | 251,2998 | 0,298918 | 1 | 0,723324 |
| -2,43121 | 421 | 357,5105 | 0,015539 | 1 | 0,147897 |
| -2,6774 | 421 | 193,2622 | 0,008058 | 1 | 0,148102 |
| -0,26839 | 420 | 397,533 | 0,788542 | 1 | 0,969427 |
| -0,71777 | 367 | 345,0119 | 0,473387 | 1 | 0,776795 |
| 0,17632 | 280 | 267,3742 | 0,860176 | 1 | 0,966655 |
| -1,14309 | 421 | 332,1235 | 0,253825 | 1 | 0,546261 |
| 0,895732 | 421 | 187,1468 | 0,371546 | 1 | 0,650743 |
| -0,21543 | 420 | 367,3245 | 0,82955 | 1 | 0,973894 |
| 0,247002 | 367 | 308,7536 | 0,805071 | 1 | 0,950149 |
| 0,753467 | 280 | 250,9084 | 0,451876 | 1 | 0,814566 |
| -0,50846 | 421 | 383,5481 | 0,611426 | 1 | 0,825172 |

| | | | | | |
|---|---|---|---|---|---|
| 0,037869 | 421 | 203,9601 | 0,969829 | 1 | 0,988858 |
| -0,30446 | 420 | 408,295 | 0,760935 | 1 | 0,969427 |
| 0,222499 | 367 | 355,7606 | 0,824053 | 1 | 0,953906 |
| 0,572241 | 280 | 270,2938 | 0,567634 | 1 | 0,866692 |
| 2,177367 | 421 | 352,6047 | 0,030116 | 1 | 0,200565 |
| 1,775967 | 421 | 189,9463 | 0,077339 | 1 | 0,353183 |
| 0,231808 | 420 | 390,3719 | 0,816809 | 1 | 0,969427 |
| 2,353985 | 367 | 330,026 | 0,019159 | 1 | 0,218203 |
| 1,654996 | 280 | 265,9815 | 0,099105 | 1 | 0,480557 |
| 0,635685 | 421 | 330,4744 | 0,525422 | 1 | 0,770244 |
| 1,312348 | 421 | 183,2841 | 0,191044 | 1 | 0,47904 |
| 0,422633 | 420 | 366,0963 | 0,672811 | 1 | 0,96781 |
| 1,599618 | 367 | 318,3571 | 0,110675 | 1 | 0,437634 |
| 2,364481 | 280 | 276 | 0,018748 | 1 | 0,281002 |
| -1,47718 | 421 | 350,8023 | 0,140525 | 1 | 0,410878 |
| -1,42349 | 421 | 192,9245 | 0,15621 | 1 | 0,438625 |
| -0,5451 | 420 | 386,211 | 0,586002 | 1 | 0,964878 |
| -1,21063 | 367 | 340,454 | 0,226877 | 1 | 0,576775 |
| 0,728746 | 280 | 262,3963 | 0,466807 | 1 | 0,824256 |
| 2,787513 | 421 | 354,3984 | 0,005598 | 1 | 0,09373 |
| 3,205793 | 421 | 193,8638 | 0,001575 | 1 | 0,093671 |
| 1,593169 | 420 | 391,7459 | 0,111929 | 1 | 0,848246 |
| 2,671934 | 367 | 343,5724 | 0,007901 | 1 | 0,180403 |
| 2,680192 | 280 | 266,6882 | 0,007816 | 1 | 0,199202 |
| 1,109044 | 421 | 309,7371 | 0,268272 | 1 | 0,563516 |
| -0,93296 | 421 | 178,4268 | 0,352103 | 1 | 0,643758 |
| -0,57348 | 420 | 340,8861 | 0,5667 | 1 | 0,962539 |
| 1,269887 | 367 | 291,5845 | 0,205137 | 1 | 0,558434 |
| 0,102288 | 280 | 245,4368 | 0,918612 | 1 | 0,977051 |
| -0,6187 | 421 | 357,7327 | 0,536509 | 1 | 0,778195 |
| -0,00592 | 421 | 193,3238 | 0,995287 | 1 | 0,997935 |
| -0,9445 | 420 | 389,3517 | 0,345498 | 1 | 0,947373 |
| 0,625226 | 367 | 344,6604 | 0,532236 | 1 | 0,814457 |
| 1,398983 | 280 | 264,7469 | 0,162988 | 1 | 0,584818 |
| 1,776817 | 421 | 372,8364 | 0,076414 | 1 | 0,300378 |
| 1,483473 | 421 | 198,0207 | 0,139539 | 1 | 0,423351 |
| -0,50614 | 420 | 402,4499 | 0,613035 | 1 | 0,964878 |
| 1,711079 | 367 | 354,5261 | 0,087941 | 1 | 0,394165 |
| 1,984144 | 280 | 263,4779 | 0,048277 | 1 | 0,393265 |
| 0,383353 | 421 | 390,1686 | 0,701667 | 1 | 0,874567 |
| 0,205594 | 421 | 209,0447 | 0,837308 | 1 | 0,938419 |
| 0,254759 | 420 | 412,3054 | 0,799037 | 1 | 0,969427 |
| 0,469004 | 367 | 353,8999 | 0,639356 | 1 | 0,880707 |
| -0,20502 | 280 | 265,878 | 0,83771 | 1 | 0,964667 |
| -2,24642 | 421 | 301,9517 | 0,0254 | 1 | 0,184916 |
| 0,059622 | 421 | 175,3099 | 0,952525 | 1 | 0,982515 |
| -1,26974 | 420 | 335,2816 | 0,205059 | 1 | 0,928589 |
| -0,30835 | 367 | 284,5844 | 0,758041 | 1 | 0,928 |

| | | | | | |
|---|---|---|---|---|---|
| -0,4925 | 280 | 254,1714 | 0,622793 | 1 | 0,898132 |
| -1,48828 | 421 | 344,1144 | 0,137592 | 1 | 0,409785 |
| -2,48714 | 421 | 188,565 | 0,013746 | 1 | 0,17172 |
| 0,043002 | 420 | 378,7131 | 0,965723 | 1 | 0,996924 |
| -1,11697 | 367 | 331,7385 | 0,264815 | 1 | 0,609149 |
| -1,80626 | 280 | 263,8475 | 0,072017 | 1 | 0,439393 |
| -2,11748 | 421 | 340,5811 | 0,034943 | 1 | 0,214936 |
| -2,46154 | 421 | 187,2494 | 0,014739 | 1 | 0,178042 |
| -2,16751 | 420 | 363,9452 | 0,030844 | 1 | 0,80141 |
| 0,084178 | 367 | 324,1735 | 0,932967 | 1 | 0,977109 |
| -0,94921 | 280 | 259,8742 | 0,343395 | 1 | 0,758791 |
| 2,20327 | 421 | 337,8039 | 0,02825 | 1 | 0,19529 |
| 1,437635 | 421 | 185,3758 | 0,152224 | 1 | 0,432832 |
| 2,247102 | 420 | 371,1185 | 0,025221 | 1 | 0,767829 |
| 1,992506 | 367 | 319,6085 | 0,047166 | 1 | 0,311448 |
| 1,024937 | 280 | 259,5543 | 0,306347 | 1 | 0,72861 |
| -1,40338 | 421 | 299,1771 | 0,161541 | 1 | 0,435497 |
| -1,98984 | 421 | 175,2359 | 0,048164 | 1 | 0,286754 |
| 1,006115 | 420 | 332,5421 | 0,315092 | 1 | 0,943897 |
| 0,105592 | 367 | 285,2913 | 0,91598 | 1 | 0,973634 |
| -0,45716 | 280 | 255,8414 | 0,647945 | 1 | 0,908644 |
| 1,735607 | 421 | 365,4671 | 0,083476 | 1 | 0,306826 |
| 1,484653 | 421 | 197,427 | 0,139231 | 1 | 0,423351 |
| 0,747733 | 420 | 399,4167 | 0,455061 | 1 | 0,960096 |
| 1,941057 | 367 | 341,3394 | 0,053074 | 1 | 0,330508 |
| -0,25708 | 280 | 261,4063 | 0,79732 | 1 | 0,954269 |
| -2,09225 | 421 | 335,4755 | 0,037167 | 1 | 0,217944 |
| -2,78946 | 421 | 187,2142 | 0,005826 | 1 | 0,141161 |
| -0,19057 | 420 | 379,1877 | 0,848965 | 1 | 0,973894 |
| -1,9708 | 367 | 331,9634 | 0,049578 | 1 | 0,315285 |
| -1,38006 | 280 | 265,9072 | 0,168728 | 1 | 0,59271 |
| -2,69871 | 421 | 349,5087 | 0,007299 | 0,897761 | 0,105763 |
| -1,19028 | 421 | 191,7593 | 0,235407 | 1 | 0,535889 |
| -0,97386 | 420 | 385,5642 | 0,330739 | 1 | 0,947373 |
| -2,32517 | 367 | 342,5811 | 0,020646 | 1 | 0,224016 |
| -0,50498 | 280 | 265,9049 | 0,613992 | 1 | 0,889365 |
| -1,44473 | 421 | 340,9231 | 0,149452 | 1 | 0,422053 |
| 0,211172 | 421 | 184,6058 | 0,832986 | 1 | 0,936588 |
| -1,97246 | 420 | 370,5686 | 0,0493 | 1 | 0,846627 |
| 0,291716 | 367 | 334,0298 | 0,770685 | 1 | 0,938144 |
| 1,185555 | 280 | 259,79 | 0,236881 | 1 | 0,659852 |
| 3,104908 | 421 | 368,1334 | 0,002051 | 1 | 0,069718 |
| 2,854272 | 421 | 199,336 | 0,004769 | 1 | 0,135614 |
| 1,680731 | 420 | 404,0578 | 0,093588 | 1 | 0,848246 |
| 2,423107 | 367 | 351,6556 | 0,015893 | 1 | 0,204703 |
| 2,191546 | 280 | 271,7379 | 0,029261 | 1 | 0,326641 |
| 1,463794 | 421 | 356,8069 | 0,144131 | 1 | 0,412154 |
| 1,779492 | 421 | 191,9507 | 0,076741 | 1 | 0,351518 |

| | | | | | |
|---|---|---|---|---|---|
| 1,00055 | 420 | 389,7155 | 0,317665 | 1 | 0,945573 |
| 2,132069 | 367 | 339,4262 | 0,033719 | 1 | 0,279197 |
| 2,763225 | 280 | 267,3248 | 0,006121 | 1 | 0,186692 |
| -2,09142 | 421 | 298,0564 | 0,037337 | 1 | 0,217944 |
| -0,64664 | 421 | 177,8698 | 0,518696 | 1 | 0,761752 |
| -0,63165 | 420 | 327,6936 | 0,528053 | 1 | 0,960096 |
| 0,263114 | 367 | 295,9256 | 0,792646 | 1 | 0,944997 |
| 0,20532 | 280 | 262,105 | 0,837482 | 1 | 0,964667 |
| -2,11129 | 421 | 332,9125 | 0,035492 | 1 | 0,215166 |
| -1,24976 | 421 | 187,098 | 0,21295 | 1 | 0,512644 |
| -0,79741 | 420 | 369,9738 | 0,425723 | 1 | 0,95743 |
| -2,7501 | 367 | 320,0586 | 0,006297 | 1 | 0,178183 |
| -1,29814 | 280 | 263,5185 | 0,195375 | 1 | 0,626244 |
| -0,00576 | 421 | 294,2101 | 0,995405 | 1 | 0,998054 |
| 0,013265 | 421 | 173,8489 | 0,989431 | 1 | 0,997412 |
| -0,33954 | 420 | 324,7589 | 0,734421 | 1 | 0,969427 |
| 0,277554 | 367 | 296,2535 | 0,781549 | 1 | 0,94337 |
| -1,23225 | 280 | 276 | 0,218906 | 1 | 0,650539 |
| -1,41796 | 421 | 366,7558 | 0,157052 | 1 | 0,429541 |
| -0,96553 | 421 | 197,6061 | 0,335459 | 1 | 0,630335 |
| 0,005824 | 420 | 400,3212 | 0,995356 | 1 | 0,997342 |
| -1,54554 | 367 | 345,2693 | 0,123131 | 1 | 0,458167 |
| -0,14358 | 280 | 266,4847 | 0,885938 | 1 | 0,973784 |
| -1,06946 | 421 | 323,515 | 0,285658 | 1 | 0,583534 |
| 0,073696 | 421 | 183,4106 | 0,941333 | 1 | 0,979688 |
| 0,654063 | 420 | 357,4206 | 0,513492 | 1 | 0,960096 |
| 0,237951 | 367 | 320,8301 | 0,812071 | 1 | 0,950149 |
| 0,50332 | 280 | 258,785 | 0,615168 | 1 | 0,889691 |
| 0,247477 | 421 | 391,5689 | 0,804668 | 1 | 0,922616 |
| 1,236957 | 421 | 208,793 | 0,217493 | 1 | 0,514565 |
| 0,595766 | 420 | 412,6839 | 0,551658 | 1 | 0,960096 |
| -1,05433 | 367 | 362,0564 | 0,292435 | 1 | 0,627013 |
| 0,150694 | 280 | 275,0184 | 0,880328 | 1 | 0,973784 |
| 0,131436 | 421 | 349,9535 | 0,895506 | 1 | 0,962 |
| -1,52902 | 421 | 188,2734 | 0,127939 | 1 | 0,410224 |
| 0,040653 | 420 | 383,6573 | 0,967593 | 1 | 0,996924 |
| 1,794474 | 367 | 324,6863 | 0,073668 | 1 | 0,373795 |
| 0,963467 | 280 | 251,0118 | 0,33624 | 1 | 0,755731 |
| 2,14803 | 421 | 391,0987 | 0,032325 | 1 | 0,20468 |
| 1,798873 | 421 | 209,711 | 0,073477 | 1 | 0,342815 |
| 0,145484 | 420 | 413,2983 | 0,8844 | 1 | 0,982463 |
| 1,664766 | 367 | 359,1265 | 0,096832 | 1 | 0,412696 |
| 1,397242 | 280 | 275,3984 | 0,163465 | 1 | 0,585137 |
| -1,80839 | 421 | 319,8613 | 0,071485 | 1 | 0,288707 |
| -0,43814 | 421 | 183,0364 | 0,661801 | 1 | 0,845199 |
| 0,054872 | 420 | 355,044 | 0,956271 | 1 | 0,995923 |
| 0,980704 | 367 | 322,0019 | 0,327475 | 1 | 0,655788 |
| -0,11731 | 280 | 263,2927 | 0,906706 | 1 | 0,975308 |

| | | | | | |
|---|---|---|---|---|---|
| 0,118566 | 421 | 339,6284 | 0,905689 | 1 | 0,96594 |
| -0,02305 | 421 | 187,1001 | 0,981635 | 1 | 0,993502 |
| 0,550352 | 420 | 373,4536 | 0,582407 | 1 | 0,964878 |
| -0,32544 | 367 | 327,1511 | 0,745056 | 1 | 0,923669 |
| 0,388061 | 280 | 255,5744 | 0,698294 | 1 | 0,92034 |
| -0,36246 | 421 | 313,7936 | 0,71725 | 1 | 0,883085 |
| -0,86546 | 421 | 180,7511 | 0,387933 | 1 | 0,663581 |
| 0,630217 | 420 | 347,4243 | 0,528967 | 1 | 0,960096 |
| -1,57236 | 367 | 318,2725 | 0,116861 | 1 | 0,445308 |
| 0,524878 | 280 | 253,8829 | 0,600126 | 1 | 0,881231 |
| -2,77537 | 421 | 300,0448 | 0,00586 | 1 | 0,097046 |
| -0,2775 | 421 | 177,436 | 0,781723 | 1 | 0,91393 |
| -0,20012 | 420 | 331,4281 | 0,841508 | 1 | 0,973894 |
| -1,8063 | 367 | 296,0061 | 0,071887 | 1 | 0,36848 |
| -1,3583 | 280 | 247,1764 | 0,175607 | 1 | 0,598625 |
| -1,60514 | 421 | 387,6563 | 0,109277 | 1 | 0,355682 |
| -0,08327 | 421 | 205,7696 | 0,933721 | 1 | 0,975655 |
| -0,4303 | 420 | 408,5057 | 0,667206 | 1 | 0,96781 |
| -0,77533 | 367 | 358,7269 | 0,438658 | 1 | 0,742744 |
| 0,25647 | 280 | 268,3965 | 0,797784 | 1 | 0,954269 |
| 2,003597 | 421 | 377,9942 | 0,045827 | 1 | 0,237403 |
| 1,02077 | 421 | 201,6368 | 0,308587 | 1 | 0,60631 |
| -0,51822 | 420 | 406,0814 | 0,604589 | 1 | 0,964878 |
| 1,505348 | 367 | 348,8435 | 0,13314 | 1 | 0,477542 |
| -0,00786 | 280 | 267,6442 | 0,993737 | 1 | 0,996381 |
| -0,49608 | 421 | 314,1759 | 0,620184 | 1 | 0,827828 |
| -0,03526 | 421 | 178,3823 | 0,971909 | 1 | 0,990309 |
| 0,490624 | 420 | 350,122 | 0,624 | 1 | 0,964878 |
| -0,8059 | 367 | 306,8497 | 0,420923 | 1 | 0,731532 |
| 2,114321 | 280 | 276 | 0,035385 | 1 | 0,339407 |
| -3,15818 | 421 | 356,485 | 0,001723 | 1 | 0,069718 |
| -2,19852 | 421 | 193,288 | 0,029099 | 1 | 0,233259 |
| -0,54448 | 420 | 394,9482 | 0,586415 | 1 | 0,964878 |
| -2,06683 | 367 | 339,2602 | 0,039508 | 1 | 0,28956 |
| -0,76839 | 280 | 268,4748 | 0,442932 | 1 | 0,810247 |
| -2,01457 | 421 | 300,3767 | 0,044841 | 1 | 0,236315 |
| -2,37161 | 421 | 174,3022 | 0,018802 | 1 | 0,19275 |
| -0,90807 | 420 | 330,0592 | 0,364504 | 1 | 0,956033 |
| -2,26004 | 367 | 293,0875 | 0,024553 | 1 | 0,247087 |
| -0,19521 | 280 | 258,655 | 0,845378 | 1 | 0,965181 |
| -1,01272 | 421 | 344,9963 | 0,311904 | 1 | 0,608071 |
| -1,32868 | 421 | 189,4394 | 0,185552 | 1 | 0,473945 |
| -0,26328 | 420 | 380,6908 | 0,792474 | 1 | 0,969427 |
| 0,688925 | 367 | 336,1731 | 0,491346 | 1 | 0,792348 |
| -1,29715 | 280 | 260,164 | 0,195727 | 1 | 0,626244 |
| -3,07784 | 421 | 348,1212 | 0,00225 | 1 | 0,069718 |
| -1,92977 | 421 | 190,6577 | 0,05512 | 1 | 0,308795 |
| -1,03892 | 420 | 386,9536 | 0,299491 | 1 | 0,943897 |

| | | | | | |
|---|---|---|---|---|---|
| -1,98032 | 367 | 343,5354 | 0,048465 | 1 | 0,313787 |
| -1,09093 | 280 | 265,3202 | 0,276292 | 1 | 0,701156 |
| 0,925662 | 421 | 313,4494 | 0,355334 | 1 | 0,645166 |
| 2,278151 | 421 | 175,7284 | 0,023922 | 1 | 0,218351 |
| 0,118248 | 420 | 347,6876 | 0,90594 | 1 | 0,98503 |
| 0,621839 | 367 | 301,4892 | 0,534518 | 1 | 0,816122 |
| 1,338742 | 280 | 246,1816 | 0,18189 | 1 | 0,607861 |
| -1,55378 | 421 | 326,3537 | 0,121206 | 1 | 0,382129 |
| -2,49035 | 421 | 185,0481 | 0,013643 | 1 | 0,17172 |
| -1,14572 | 420 | 365,6615 | 0,252661 | 1 | 0,943897 |
| -2,07042 | 367 | 319,2825 | 0,039216 | 1 | 0,28956 |
| -0,71127 | 280 | 260,2967 | 0,477554 | 1 | 0,824256 |
| 1,256523 | 421 | 403,1434 | 0,209654 | 1 | 0,498652 |
| 2,756097 | 421 | 217,4838 | 0,006346 | 1 | 0,141161 |
| -0,19511 | 420 | 415,8568 | 0,845401 | 1 | 0,973894 |
| 0,723908 | 367 | 362,6772 | 0,469589 | 1 | 0,775105 |
| 0,712338 | 280 | 273,2196 | 0,476863 | 1 | 0,824256 |
| 1,764967 | 421 | 332,2078 | 0,078488 | 1 | 0,302838 |
| -0,8546 | 421 | 183,8051 | 0,393886 | 1 | 0,665792 |
| -0,63352 | 420 | 366,9345 | 0,526791 | 1 | 0,960096 |
| 1,861031 | 367 | 323,7272 | 0,063646 | 1 | 0,350852 |
| -0,32576 | 280 | 264,1936 | 0,744865 | 1 | 0,942071 |
| -0,25361 | 421 | 310,2111 | 0,799967 | 1 | 0,922616 |
| 1,440551 | 421 | 180,083 | 0,151448 | 1 | 0,432258 |
| -0,50205 | 420 | 342,3161 | 0,615953 | 1 | 0,964878 |
| 0,097349 | 367 | 300,1546 | 0,922514 | 1 | 0,975687 |
| 0,341579 | 280 | 263,8036 | 0,73294 | 1 | 0,936751 |
| -2,15429 | 421 | 326,5393 | 0,031949 | 1 | 0,204011 |
| -1,23698 | 421 | 181,4133 | 0,217693 | 1 | 0,514565 |
| -0,41276 | 420 | 364,1677 | 0,680024 | 1 | 0,96781 |
| -1,385 | 367 | 324,0824 | 0,167005 | 1 | 0,514677 |
| -2,76056 | 280 | 252,1016 | 0,006194 | 1 | 0,186692 |
| -1,37945 | 421 | 300,7472 | 0,168781 | 1 | 0,442288 |
| 0,280614 | 421 | 177,3233 | 0,779334 | 1 | 0,913263 |
| 1,88485 | 420 | 330,0334 | 0,060329 | 1 | 0,848246 |
| 2,078287 | 367 | 290,1529 | 0,038562 | 1 | 0,28956 |
| -0,82649 | 280 | 254,1675 | 0,409299 | 1 | 0,788151 |
| -0,15607 | 421 | 375,1347 | 0,876059 | 1 | 0,95861 |
| 2,354056 | 421 | 200,4432 | 0,019537 | 1 | 0,195731 |
| 1,125393 | 420 | 401,6818 | 0,261095 | 1 | 0,943897 |
| 1,026177 | 367 | 356,4303 | 0,305504 | 1 | 0,637666 |
| 0,317193 | 280 | 273,2205 | 0,751339 | 1 | 0,942071 |
| -0,68483 | 421 | 350,4071 | 0,493906 | 1 | 0,74916 |
| -0,53077 | 421 | 189,267 | 0,596202 | 1 | 0,811435 |
| 0,717397 | 420 | 382,8844 | 0,473567 | 1 | 0,960096 |
| -1,07211 | 367 | 340,4578 | 0,284431 | 1 | 0,619419 |
| -0,39236 | 280 | 261,6402 | 0,695108 | 1 | 0,92034 |
| -0,74191 | 421 | 334,3437 | 0,458665 | 1 | 0,724077 |

| | | | | | |
|---|---|---|---|---|---|
| 0,781935 | 421 | 186,6743 | 0,435243 | 1 | 0,702261 |
| 0,100871 | 420 | 371,9425 | 0,919707 | 1 | 0,989204 |
| -0,47036 | 367 | 327,6904 | 0,638408 | 1 | 0,880707 |
| -0,80654 | 280 | 254,9929 | 0,420682 | 1 | 0,796701 |
| 2,171441 | 421 | 354,3953 | 0,030561 | 1 | 0,200565 |
| 2,165402 | 421 | 192,6064 | 0,031586 | 1 | 0,241423 |
| 1,250758 | 420 | 390,9582 | 0,211771 | 1 | 0,943055 |
| 2,465447 | 367 | 347,3508 | 0,014167 | 1 | 0,201413 |
| 2,413806 | 280 | 267,0065 | 0,01646 | 1 | 0,266517 |
| -2,30311 | 421 | 347,3127 | 0,021863 | 1 | 0,175251 |
| -1,89004 | 421 | 190,2733 | 0,060273 | 1 | 0,314727 |
| -1,64861 | 420 | 381,9723 | 0,10005 | 1 | 0,848246 |
| 1,23501 | 367 | 344,1935 | 0,217669 | 1 | 0,569492 |
| 0,478897 | 280 | 263,6875 | 0,632409 | 1 | 0,903355 |
| 0,815993 | 421 | 367,017 | 0,415033 | 1 | 0,695339 |
| 1,341396 | 421 | 194,4026 | 0,181357 | 1 | 0,470641 |
| 0,588001 | 420 | 398,6902 | 0,556864 | 1 | 0,960096 |
| 0,856065 | 367 | 350,1923 | 0,392547 | 1 | 0,705088 |
| 0,310527 | 280 | 261,9976 | 0,756407 | 1 | 0,942071 |
| -1,63675 | 421 | 320,6977 | 0,102663 | 1 | 0,344557 |
| 0,349604 | 421 | 180,6034 | 0,727044 | 1 | 0,880751 |
| 0,336816 | 420 | 359,6004 | 0,736452 | 1 | 0,969427 |
| 0,223928 | 367 | 311,1297 | 0,82296 | 1 | 0,953906 |
| 0,896378 | 280 | 252,4439 | 0,370905 | 1 | 0,763914 |
| 1,428974 | 421 | 384,8082 | 0,153823 | 1 | 0,426123 |
| 0,87301 | 421 | 205,5186 | 0,383677 | 1 | 0,659083 |
| 1,076934 | 420 | 408,139 | 0,282146 | 1 | 0,943897 |
| 1,097581 | 367 | 355,556 | 0,27313 | 1 | 0,609149 |
| -0,39501 | 280 | 268,4455 | 0,693152 | 1 | 0,920335 |
| -0,46751 | 421 | 390,3317 | 0,640397 | 1 | 0,834842 |
| -1,61846 | 421 | 208,6723 | 0,107074 | 1 | 0,386957 |
| 0,770462 | 420 | 412,2367 | 0,441467 | 1 | 0,960096 |
| -0,38126 | 367 | 357,7271 | 0,703236 | 1 | 0,908463 |
| -0,7601 | 280 | 271,9526 | 0,447851 | 1 | 0,813228 |
| -1,40288 | 421 | 367,4394 | 0,161496 | 1 | 0,435497 |
| -1,06059 | 421 | 197,7566 | 0,29017 | 1 | 0,586961 |
| 0,791131 | 420 | 397,9857 | 0,429339 | 1 | 0,958538 |
| 0,050717 | 367 | 353,1519 | 0,95958 | 1 | 0,981063 |
| -2,54801 | 280 | 274,2662 | 0,01138 | 1 | 0,234919 |
| -0,58923 | 421 | 374,4911 | 0,556066 | 1 | 0,790572 |
| -1,51 | 421 | 199,5536 | 0,132624 | 1 | 0,414338 |
| 0,725128 | 420 | 403,1821 | 0,468794 | 1 | 0,960096 |
| -1,04882 | 367 | 350,6692 | 0,294985 | 1 | 0,62734 |
| -0,43 | 280 | 267,9841 | 0,667541 | 1 | 0,908644 |
| -1,0886 | 421 | 323,0973 | 0,277141 | 1 | 0,57291 |
| -0,44646 | 421 | 185,5571 | 0,655783 | 1 | 0,841107 |
| -1,85718 | 420 | 361,7638 | 0,064097 | 1 | 0,848246 |
| 0,329817 | 367 | 325,9364 | 0,74175 | 1 | 0,923669 |

| | | | | | |
|---|---|---|---|---|---|
| 0,365635 | 280 | 257,7401 | 0,714938 | 1 | 0,929604 |
| -1,00668 | 421 | 410,2239 | 0,31468 | 1 | 0,608758 |
| -0,50406 | 421 | 225,5208 | 0,614712 | 1 | 0,824908 |
| -0,58477 | 420 | 415,8466 | 0,559018 | 1 | 0,960096 |
| -1,09841 | 367 | 362,9384 | 0,272752 | 1 | 0,609149 |
| -0,57899 | 280 | 274,2406 | 0,563073 | 1 | 0,861728 |
| 0,882819 | 421 | 393,6035 | 0,377873 | 1 | 0,670163 |
| -0,55717 | 421 | 211,2744 | 0,578 | 1 | 0,794719 |
| -1,1601 | 420 | 413,529 | 0,246675 | 1 | 0,943897 |
| -0,17299 | 367 | 358,2578 | 0,862759 | 1 | 0,959636 |
| -1,5681 | 280 | 274,1506 | 0,118011 | 1 | 0,520006 |
| -0,77819 | 421 | 353,7413 | 0,43698 | 1 | 0,707193 |
| -2,18332 | 421 | 193,3777 | 0,030216 | 1 | 0,238409 |
| -0,61255 | 420 | 387,4998 | 0,540531 | 1 | 0,960096 |
| -1,82519 | 367 | 344,4365 | 0,068838 | 1 | 0,364488 |
| -0,65217 | 280 | 267,8368 | 0,514848 | 1 | 0,83211 |
| -1,24755 | 421 | 362,1201 | 0,213002 | 1 | 0,503916 |
| -2,05245 | 421 | 195,0196 | 0,041461 | 1 | 0,272972 |
| -0,14592 | 420 | 396,6855 | 0,884058 | 1 | 0,982463 |
| -2,42696 | 367 | 349,1786 | 0,015731 | 1 | 0,204372 |
| -3,69498 | 280 | 267,5443 | 2,67E-04 | 0,286655 | 0,057407 |
| -2,18743 | 421 | 374,2763 | 0,029329 | 1 | 0,199094 |
| -2,04294 | 421 | 201,4692 | 0,042361 | 1 | 0,273775 |
| -0,6421 | 420 | 405,2657 | 0,521174 | 1 | 0,960096 |
| -1,86388 | 367 | 356,2399 | 0,063161 | 1 | 0,34994 |
| -1,72586 | 280 | 270,8237 | 0,085513 | 1 | 0,458532 |
| -1,59554 | 421 | 331,7831 | 0,111543 | 1 | 0,358728 |
| 0,111514 | 421 | 184,3379 | 0,91133 | 1 | 0,96646 |
| 0,35455 | 420 | 364,4432 | 0,723132 | 1 | 0,969427 |
| -0,02553 | 367 | 329,8586 | 0,979652 | 1 | 0,992161 |
| -0,87678 | 280 | 258,5254 | 0,381419 | 1 | 0,770948 |
| 0,914151 | 421 | 360,6026 | 0,361248 | 1 | 0,651198 |
| 0,25275 | 421 | 195,3443 | 0,800727 | 1 | 0,921861 |
| -0,45927 | 420 | 392,7153 | 0,646294 | 1 | 0,964878 |
| -0,91113 | 367 | 337,6143 | 0,362876 | 1 | 0,677287 |
| 0,477505 | 280 | 261,0874 | 0,633402 | 1 | 0,903918 |
| -0,9731 | 421 | 332,3109 | 0,331214 | 1 | 0,619505 |
| -1,61338 | 421 | 184,5704 | 0,10837 | 1 | 0,388185 |
| -3,09207 | 420 | 351,2598 | 0,002147 | 1 | 0,294094 |
| -0,52551 | 367 | 320,2622 | 0,599594 | 1 | 0,862104 |
| -0,91899 | 280 | 255,5915 | 0,358966 | 1 | 0,761846 |
| 0,542308 | 421 | 375,2195 | 0,587929 | 1 | 0,80988 |
| 0,454043 | 421 | 198,4796 | 0,650294 | 1 | 0,839358 |
| 0,794155 | 420 | 404,216 | 0,427571 | 1 | 0,95743 |
| 1,009937 | 367 | 357,4538 | 0,313209 | 1 | 0,64747 |
| -1,02041 | 280 | 271,0822 | 0,308442 | 1 | 0,729704 |
| -0,2859 | 421 | 305,5089 | 0,77515 | 1 | 0,911337 |
| 1,051665 | 421 | 178,5826 | 0,294374 | 1 | 0,590708 |

| | | | | | |
|---|---|---|---|---|---|
| 1,834708 | 420 | 331,865 | 0,067444 | 1 | 0,848246 |
| -0,37238 | 367 | 282,4456 | 0,709888 | 1 | 0,912021 |
| 0,437857 | 280 | 249,0717 | 0,661869 | 1 | 0,908644 |
| -4,17356 | 421 | 331,6505 | 3,84E-05 | 0,041309 | 0,019267 |
| -1,037 | 421 | 185,7217 | 0,301085 | 1 | 0,600041 |
| -0,65721 | 420 | 368,2547 | 0,511459 | 1 | 0,960096 |
| -1,35438 | 367 | 318,995 | 0,176574 | 1 | 0,523018 |
| 0,783421 | 280 | 254,5124 | 0,434108 | 1 | 0,803161 |
| -0,72847 | 421 | 392,2095 | 0,466762 | 1 | 0,728168 |
| -1,6986 | 421 | 209,3658 | 0,09088 | 1 | 0,373113 |
| 0,659897 | 420 | 413,2626 | 0,509687 | 1 | 0,960096 |
| -0,06783 | 367 | 360,6228 | 0,945957 | 1 | 0,979539 |
| 0,323504 | 280 | 271,9814 | 0,746562 | 1 | 0,942071 |
| 0,132172 | 421 | 353,4875 | 0,894924 | 1 | 0,962 |
| 0,613068 | 421 | 191,8765 | 0,540557 | 1 | 0,773814 |
| -1,67815 | 420 | 382,7339 | 0,094133 | 1 | 0,848246 |
| 0,476848 | 367 | 332,9033 | 0,633784 | 1 | 0,880707 |
| -0,42073 | 280 | 265,216 | 0,67429 | 1 | 0,909385 |
| -0,88545 | 421 | 354,8238 | 0,376513 | 1 | 0,669899 |
| -1,20585 | 421 | 192,8986 | 0,229353 | 1 | 0,529303 |
| -0,48051 | 420 | 388,9081 | 0,631134 | 1 | 0,964878 |
| -1,4594 | 367 | 335,0252 | 0,145391 | 1 | 0,496835 |
| -0,13143 | 280 | 259,5556 | 0,895539 | 1 | 0,975243 |
| -0,81638 | 421 | 341,213 | 0,414855 | 1 | 0,695339 |
| -2,29602 | 421 | 189,3688 | 0,02277 | 1 | 0,212324 |
| -0,5716 | 420 | 378,9455 | 0,567929 | 1 | 0,962723 |
| -1,42455 | 367 | 323,2224 | 0,155252 | 1 | 0,502197 |
| -0,70919 | 280 | 264,0791 | 0,478835 | 1 | 0,824256 |
| -1,87988 | 421 | 338,3047 | 0,060984 | 1 | 0,270611 |
| -2,94593 | 421 | 182,2596 | 0,00364 | 1 | 0,118336 |
| -1,62643 | 420 | 377,3342 | 0,104693 | 1 | 0,848246 |
| -1,99022 | 367 | 337,8808 | 0,047372 | 1 | 0,311448 |
| -0,95371 | 280 | 258,3731 | 0,341121 | 1 | 0,757098 |
| -3,09451 | 421 | 310,9309 | 0,002151 | 1 | 0,069718 |
| -2,76938 | 421 | 178,0919 | 0,006211 | 1 | 0,141161 |
| -0,21682 | 420 | 350,1947 | 0,828475 | 1 | 0,973894 |
| 0,173562 | 367 | 306,8567 | 0,862324 | 1 | 0,959636 |
| -0,99525 | 280 | 246,8638 | 0,320588 | 1 | 0,744349 |
| 0,445207 | 421 | 389,0207 | 0,656418 | 1 | 0,846936 |
| -0,83264 | 421 | 206,2301 | 0,406013 | 1 | 0,679847 |
| 1,909847 | 420 | 411,512 | 0,056848 | 1 | 0,848246 |
| -0,06524 | 367 | 357,6274 | 0,948022 | 1 | 0,979539 |
| 0,396668 | 280 | 266,6567 | 0,69193 | 1 | 0,919522 |
| 0,804803 | 421 | 332,9161 | 0,421508 | 1 | 0,697735 |
| 2,574914 | 421 | 185,2094 | 0,010807 | 1 | 0,159671 |
| -0,40864 | 420 | 370,7524 | 0,683041 | 1 | 0,968338 |
| 0,678043 | 367 | 335,3696 | 0,498212 | 1 | 0,793601 |
| -0,08211 | 280 | 256,1976 | 0,934625 | 1 | 0,979504 |

| | | | | | |
|---|---|---|---|---|---|
| 1,848761 | 421 | 386,7111 | 0,065255 | 1 | 0,276236 |
| 1,299213 | 421 | 205,9914 | 0,195323 | 1 | 0,483337 |
| 0,464228 | 420 | 409,6476 | 0,642731 | 1 | 0,964878 |
| 1,664842 | 367 | 358,3256 | 0,096818 | 1 | 0,412696 |
| 0,549278 | 280 | 273,772 | 0,583262 | 1 | 0,87561 |
| -0,27114 | 421 | 353,7442 | 0,786444 | 1 | 0,917552 |
| 0,773556 | 421 | 190,8193 | 0,440151 | 1 | 0,705407 |
| -1,30659 | 420 | 384,2582 | 0,192134 | 1 | 0,925729 |
| 1,308901 | 367 | 338,2023 | 0,191456 | 1 | 0,542851 |
| 0,264663 | 280 | 269,2261 | 0,791471 | 1 | 0,953435 |
| -1,23642 | 421 | 331,8998 | 0,217178 | 1 | 0,510588 |
| 0,113702 | 421 | 184,2327 | 0,909598 | 1 | 0,96646 |
| 0,581925 | 420 | 368,9704 | 0,560973 | 1 | 0,960096 |
| 0,150454 | 367 | 311,426 | 0,880504 | 1 | 0,962453 |
| 1,155998 | 280 | 271,5414 | 0,248698 | 1 | 0,677438 |
| 0,108547 | 421 | 335,4657 | 0,913627 | 1 | 0,966828 |
| -0,78242 | 421 | 184,4023 | 0,434973 | 1 | 0,702261 |
| 0,470089 | 420 | 369,8161 | 0,638569 | 1 | 0,964878 |
| 0,607657 | 367 | 314,962 | 0,543853 | 1 | 0,824628 |
| -0,60063 | 280 | 276 | 0,548581 | 1 | 0,852208 |
| -1,29584 | 421 | 314,4034 | 0,19598 | 1 | 0,481951 |
| -1,73974 | 421 | 181,6289 | 0,083598 | 1 | 0,365475 |
| -0,64954 | 420 | 347,8617 | 0,51642 | 1 | 0,960096 |
| -0,89879 | 367 | 313,7171 | 0,369452 | 1 | 0,679634 |
| -0,29705 | 280 | 260,6987 | 0,766666 | 1 | 0,945436 |
| 0,539731 | 421 | 329,0559 | 0,589748 | 1 | 0,81147 |
| 0,806805 | 421 | 183,6249 | 0,420823 | 1 | 0,691581 |
| 3,137613 | 420 | 366,2428 | 0,001841 | 1 | 0,294094 |
| -0,50385 | 367 | 328,0369 | 0,614707 | 1 | 0,872281 |
| 1,041883 | 280 | 258,4483 | 0,298439 | 1 | 0,723324 |
| 2,127428 | 421 | 356,8182 | 0,03407 | 1 | 0,212429 |
| 0,59455 | 421 | 194,4883 | 0,552835 | 1 | 0,778237 |
| -0,46371 | 420 | 389,6139 | 0,643118 | 1 | 0,964878 |
| 3,536237 | 367 | 339,3541 | 4,62E-04 | 0,49647 | 0,116105 |
| 0,174992 | 280 | 267,4771 | 0,861219 | 1 | 0,967106 |
| -0,30853 | 421 | 350,8809 | 0,757865 | 1 | 0,900919 |
| 2,673201 | 421 | 191,4203 | 0,008162 | 1 | 0,148102 |
| 1,764046 | 420 | 383,6445 | 0,07852 | 1 | 0,848246 |
| 2,983479 | 367 | 334,2325 | 0,00306 | 1 | 0,176492 |
| 1,52216 | 280 | 250,6968 | 0,129229 | 1 | 0,536498 |
| -1,99381 | 421 | 356,429 | 0,046934 | 1 | 0,239762 |
| -1,42142 | 421 | 192,6346 | 0,156812 | 1 | 0,438625 |
| -1,14827 | 420 | 391,1713 | 0,25156 | 1 | 0,943897 |
| -2,10369 | 367 | 333,5381 | 0,036154 | 1 | 0,28464 |
| -2,16745 | 280 | 258,4584 | 0,031114 | 1 | 0,329568 |
| 1,764429 | 421 | 380,1779 | 0,078463 | 1 | 0,302838 |
| 2,425267 | 421 | 204,4994 | 0,016165 | 1 | 0,179127 |
| 0,506182 | 420 | 408,7094 | 0,613002 | 1 | 0,964878 |

| | | | | | |
|---|---|---|---|---|---|
| 2,531366 | 367 | 356,0689 | 0,011792 | 1 | 0,189546 |
| 1,757324 | 280 | 272,7278 | 0,079985 | 1 | 0,453386 |
| 1,476119 | 421 | 355,2789 | 0,140798 | 1 | 0,410878 |
| 2,715323 | 421 | 193,1493 | 0,007221 | 1 | 0,14319 |
| 1,108885 | 420 | 392,5552 | 0,268159 | 1 | 0,943897 |
| 2,755089 | 367 | 340,3055 | 0,006183 | 1 | 0,178183 |
| 1,460624 | 280 | 260,3698 | 0,145324 | 1 | 0,555466 |
| -0,99378 | 421 | 368,0918 | 0,320981 | 1 | 0,615134 |
| -1,46102 | 421 | 198,0804 | 0,145593 | 1 | 0,426194 |
| -1,57992 | 420 | 402,3492 | 0,114912 | 1 | 0,848246 |
| 1,757413 | 367 | 353,813 | 0,079712 | 1 | 0,381841 |
| -1,97397 | 280 | 271,8612 | 0,049398 | 1 | 0,39442 |
| -1,34403 | 421 | 332,133 | 0,179857 | 1 | 0,459396 |
| -0,28362 | 421 | 182,2297 | 0,777024 | 1 | 0,911265 |
| -0,3748 | 420 | 366,6533 | 0,708024 | 1 | 0,968866 |
| -0,36725 | 367 | 312,7257 | 0,713684 | 1 | 0,912652 |
| 0,243239 | 280 | 264,427 | 0,808009 | 1 | 0,95747 |
| -0,82907 | 421 | 322,889 | 0,407677 | 1 | 0,695339 |
| -1,45459 | 421 | 181,8776 | 0,147506 | 1 | 0,428784 |
| -1,09683 | 420 | 359,3092 | 0,273452 | 1 | 0,943897 |
| -2,18514 | 367 | 298,8422 | 0,029655 | 1 | 0,260259 |
| -3,06462 | 280 | 258,3638 | 0,00241 | 1 | 0,135787 |
| -2,09494 | 421 | 377,0283 | 0,036843 | 1 | 0,217944 |
| -1,31818 | 421 | 202,719 | 0,188931 | 1 | 0,476746 |
| -0,58309 | 420 | 407,0911 | 0,560157 | 1 | 0,960096 |
| -2,01182 | 367 | 354,0302 | 0,044996 | 1 | 0,302719 |
| -1,99935 | 280 | 270,5338 | 0,046571 | 1 | 0,387749 |
| -1,1572 | 421 | 322,7897 | 0,248045 | 1 | 0,544195 |
| 0,069795 | 421 | 181,4111 | 0,944434 | 1 | 0,980711 |
| -1,01466 | 420 | 353,4149 | 0,31096 | 1 | 0,943897 |
| -0,27275 | 367 | 315,6962 | 0,785227 | 1 | 0,94337 |
| 1,786653 | 280 | 252,2671 | 0,075194 | 1 | 0,447896 |
| -3,45908 | 421 | 352,8876 | 6,08E-04 | 0,647889 | 0,048251 |
| -1,5248 | 421 | 193,1103 | 0,128945 | 1 | 0,410224 |
| -1,4537 | 420 | 389,9518 | 0,146835 | 1 | 0,914005 |
| -0,74964 | 367 | 341,1346 | 0,453987 | 1 | 0,758491 |
| 1,038513 | 280 | 263,0336 | 0,299985 | 1 | 0,723324 |
| 1,375102 | 421 | 404,8294 | 0,16986 | 1 | 0,442288 |
| 1,448248 | 421 | 218,794 | 0,148979 | 1 | 0,430099 |
| 0,256685 | 420 | 415,9813 | 0,797549 | 1 | 0,969427 |
| 1,335775 | 367 | 362,661 | 0,18246 | 1 | 0,527769 |
| 1,33932 | 280 | 274,2278 | 0,181575 | 1 | 0,607861 |
| 1,439306 | 421 | 407,6197 | 0,150831 | 1 | 0,424072 |
| 1,105237 | 421 | 221,4736 | 0,270256 | 1 | 0,569587 |
| -1,19815 | 420 | 415,995 | 0,231542 | 1 | 0,943055 |
| -0,66073 | 367 | 360,7976 | 0,509206 | 1 | 0,797092 |
| -1,84051 | 280 | 274,2962 | 0,066774 | 1 | 0,425749 |
| -1,34606 | 421 | 324,0802 | 0,179222 | 1 | 0,458554 |

| | | | | | |
|---|---|---|---|---|---|
| 0,133899 | 421 | 183,9055 | 0,893629 | 1 | 0,958882 |
| 0,084959 | 420 | 360,385 | 0,932342 | 1 | 0,991559 |
| 0,204756 | 367 | 319,0842 | 0,837893 | 1 | 0,956935 |
| -1,27557 | 280 | 251,6338 | 0,203283 | 1 | 0,632429 |
| -1,74737 | 421 | 357,3226 | 0,081433 | 1 | 0,30376 |
| -0,10286 | 421 | 193,7725 | 0,918177 | 1 | 0,970154 |
| -0,69747 | 420 | 390,2242 | 0,485925 | 1 | 0,960096 |
| 0,531157 | 367 | 341,6848 | 0,595655 | 1 | 0,858998 |
| 0,062162 | 280 | 266,9863 | 0,95048 | 1 | 0,982409 |
| -0,09774 | 421 | 316,6862 | 0,9222 | 1 | 0,966828 |
| 0,926396 | 421 | 181,2171 | 0,355472 | 1 | 0,643758 |
| 0,112562 | 420 | 350,15 | 0,910442 | 1 | 0,987181 |
| 0,101182 | 367 | 309,3521 | 0,919472 | 1 | 0,974433 |
| -0,14451 | 280 | 254,6888 | 0,885215 | 1 | 0,973784 |
| 0,739718 | 421 | 365,6651 | 0,459946 | 1 | 0,724425 |
| -0,12613 | 421 | 196,8217 | 0,899758 | 1 | 0,960975 |
| 0,178927 | 420 | 396,2287 | 0,858087 | 1 | 0,976689 |
| 1,044669 | 367 | 346,545 | 0,296904 | 1 | 0,629303 |
| -2,40893 | 280 | 271,9799 | 0,016665 | 1 | 0,266517 |
| -0,43461 | 421 | 361,2108 | 0,664107 | 1 | 0,852478 |
| 0,592268 | 421 | 193,6737 | 0,554362 | 1 | 0,778237 |
| 0,119505 | 420 | 392,1639 | 0,904936 | 1 | 0,98503 |
| 0,263184 | 367 | 346,4404 | 0,792565 | 1 | 0,944997 |
| -0,5611 | 280 | 258,0386 | 0,575219 | 1 | 0,870337 |
| 0,243827 | 421 | 365,4042 | 0,807502 | 1 | 0,923297 |
| -0,00834 | 421 | 195,0437 | 0,993356 | 1 | 0,997935 |
| 0,253454 | 420 | 396,1555 | 0,800049 | 1 | 0,969427 |
| 1,825925 | 367 | 345,0026 | 0,068726 | 1 | 0,364488 |
| 0,640792 | 280 | 261,5653 | 0,522219 | 1 | 0,833721 |
| -1,05051 | 421 | 322,754 | 0,294268 | 1 | 0,592864 |
| 0,285975 | 421 | 180,5052 | 0,775225 | 1 | 0,910824 |
| 1,190094 | 420 | 360,6732 | 0,234792 | 1 | 0,943055 |
| 0,993236 | 367 | 301,289 | 0,321392 | 1 | 0,653627 |
| 1,079893 | 280 | 258,2582 | 0,281198 | 1 | 0,703928 |
| -1,37352 | 421 | 339,1428 | 0,170499 | 1 | 0,443003 |
| 0,210614 | 421 | 184,9108 | 0,833421 | 1 | 0,936588 |
| -1,08426 | 420 | 371,1376 | 0,278954 | 1 | 0,943897 |
| 0,388174 | 367 | 312,3804 | 0,698152 | 1 | 0,906214 |
| -0,40334 | 280 | 261,267 | 0,687025 | 1 | 0,915426 |
| -0,77316 | 421 | 353,5508 | 0,439945 | 1 | 0,710422 |
| 0,436904 | 421 | 191,1314 | 0,662674 | 1 | 0,845595 |
| 0,047689 | 420 | 387,1524 | 0,961989 | 1 | 0,996924 |
| -0,48678 | 367 | 335,3839 | 0,626733 | 1 | 0,88023 |
| 0,017092 | 280 | 267,7621 | 0,986376 | 1 | 0,992965 |
| 0,29681 | 421 | 405,4609 | 0,766764 | 1 | 0,906285 |
| 1,235433 | 421 | 221,5585 | 0,217978 | 1 | 0,514565 |
| 0,605813 | 420 | 415,9996 | 0,544969 | 1 | 0,960096 |
| 0,494374 | 367 | 362,7965 | 0,621341 | 1 | 0,875495 |

| | | | | | |
|---|---|---|---|---|---|
| 0,685012 | 280 | 271,6957 | 0,49392 | 1 | 0,827962 |
| -1,2597 | 421 | 338,0487 | 0,208645 | 1 | 0,497513 |
| -0,39955 | 421 | 188,3719 | 0,689943 | 1 | 0,864363 |
| 0,21202 | 420 | 372,3247 | 0,832208 | 1 | 0,973894 |
| 0,122801 | 367 | 319,9117 | 0,902342 | 1 | 0,970486 |
| -0,34976 | 280 | 253,266 | 0,726812 | 1 | 0,936637 |
| -0,04238 | 421 | 308,6554 | 0,966226 | 1 | 0,992572 |
| 0,859394 | 421 | 177,0876 | 0,391285 | 1 | 0,665538 |
| 1,011473 | 420 | 341,5403 | 0,312506 | 1 | 0,943897 |
| -2,20492 | 367 | 312,0316 | 0,028189 | 1 | 0,257802 |
| 0,999444 | 280 | 260,7078 | 0,318506 | 1 | 0,741869 |
| -1,59672 | 421 | 345,2905 | 0,111243 | 1 | 0,358728 |
| -1,36119 | 421 | 189,0631 | 0,175074 | 1 | 0,459682 |
| 0,291399 | 420 | 381,0515 | 0,770904 | 1 | 0,969427 |
| -2,15487 | 367 | 331,301 | 0,031892 | 1 | 0,27001 |
| -0,3906 | 280 | 264,172 | 0,696405 | 1 | 0,92034 |
| -1,59873 | 421 | 341,5218 | 0,110806 | 1 | 0,358728 |
| -0,2302 | 421 | 187,2567 | 0,818187 | 1 | 0,926377 |
| 0,190299 | 420 | 377,0735 | 0,849178 | 1 | 0,973894 |
| -0,51248 | 367 | 329,4915 | 0,608661 | 1 | 0,870021 |
| 0,894863 | 280 | 266,8139 | 0,371667 | 1 | 0,763914 |
| -2,49734 | 421 | 325,9916 | 0,013006 | 1 | 0,138025 |
| -1,49981 | 421 | 183,2114 | 0,135385 | 1 | 0,418943 |
| -0,9332 | 420 | 365,6511 | 0,351334 | 1 | 0,947373 |
| -1,25261 | 367 | 327,0069 | 0,211242 | 1 | 0,563436 |
| -0,8426 | 280 | 273,4527 | 0,400187 | 1 | 0,784757 |
| -1,96565 | 421 | 350,386 | 0,050129 | 1 | 0,246877 |
| -0,19783 | 421 | 189,8298 | 0,843392 | 1 | 0,941476 |
| -1,35385 | 420 | 378,5044 | 0,176593 | 1 | 0,9199 |
| -0,92331 | 367 | 319,1825 | 0,356544 | 1 | 0,674813 |
| -0,06342 | 280 | 252,0714 | 0,949478 | 1 | 0,982409 |
| 0,101841 | 421 | 373,4761 | 0,918938 | 1 | 0,966828 |
| 0,206073 | 421 | 199,0011 | 0,836945 | 1 | 0,938419 |
| -0,07641 | 420 | 402,2618 | 0,939132 | 1 | 0,992477 |
| 1,132207 | 367 | 343,7978 | 0,258336 | 1 | 0,609149 |
| 1,012232 | 280 | 269,566 | 0,312335 | 1 | 0,733729 |
| -2,51636 | 421 | 326,0781 | 0,012337 | 1 | 0,131857 |
| -4,61072 | 421 | 183,9132 | 7,49E-06 | 0,001154 | 0,011291 |
| -2,83382 | 420 | 362,8293 | 0,004857 | 0,728496 | 0,449965 |
| -3,40148 | 367 | 322,5557 | 7,55E-04 | 0,116201 | 0,139974 |
| -3,25997 | 280 | 253,0388 | 0,001267 | 0,19514 | 0,135787 |
| -1,23869 | 421 | 335,0949 | 0,216327 | 1 | 0,509382 |
| -1,6051 | 421 | 187,2957 | 0,110158 | 1 | 0,389736 |
| -1,21407 | 420 | 370,1305 | 0,225495 | 1 | 0,943055 |
| -0,46393 | 367 | 316,4809 | 0,643018 | 1 | 0,882539 |
| -1,13717 | 280 | 256,5577 | 0,256529 | 1 | 0,680087 |
| -0,95121 | 421 | 324,0651 | 0,342206 | 1 | 0,63164 |
| -0,94154 | 421 | 181,2303 | 0,34768 | 1 | 0,642814 |

| | | | | | |
|---|---|---|---|---|---|
| -1,05211 | 420 | 354,5117 | 0,293464 | 1 | 0,943897 |
| -0,24243 | 367 | 309,6337 | 0,808608 | 1 | 0,950149 |
| 0,563923 | 280 | 247,9358 | 0,573317 | 1 | 0,870079 |
| 0,347958 | 421 | 330,0697 | 0,728094 | 1 | 0,890807 |
| 0,365005 | 421 | 183,4029 | 0,715528 | 1 | 0,874476 |
| 0,832742 | 420 | 364,3883 | 0,405536 | 1 | 0,95743 |
| 0,941166 | 367 | 321,3783 | 0,347327 | 1 | 0,668671 |
| 0,984635 | 280 | 258,6741 | 0,325723 | 1 | 0,749412 |
| 0,206866 | 421 | 345,9339 | 0,836236 | 1 | 0,93721 |
| -0,18588 | 421 | 188,5988 | 0,852737 | 1 | 0,944105 |
| 0,154957 | 420 | 379,1685 | 0,876938 | 1 | 0,982463 |
| -0,32548 | 367 | 320,3805 | 0,745031 | 1 | 0,923669 |
| 0,536752 | 280 | 248,4581 | 0,591919 | 1 | 0,876993 |
| -0,44191 | 421 | 334,2153 | 0,658838 | 1 | 0,848606 |
| -1,4596 | 421 | 184,3089 | 0,146102 | 1 | 0,426698 |
| -0,6551 | 420 | 371,1214 | 0,512812 | 1 | 0,960096 |
| -1,74511 | 367 | 321,6483 | 0,081921 | 1 | 0,381841 |
| -2,22598 | 280 | 255,9396 | 0,026886 | 1 | 0,324814 |
| -0,84549 | 421 | 314,9812 | 0,398477 | 1 | 0,688151 |
| -0,85476 | 421 | 178,5192 | 0,393832 | 1 | 0,665792 |
| -1,21506 | 420 | 349,5313 | 0,225163 | 1 | 0,943055 |
| -2,09772 | 367 | 329,9933 | 0,03669 | 1 | 0,28464 |
| -2,36838 | 280 | 262,145 | 0,018592 | 1 | 0,281002 |
| 0,108661 | 421 | 401,1812 | 0,913525 | 1 | 0,966828 |
| 0,869479 | 421 | 215,5778 | 0,385552 | 1 | 0,660258 |
| -0,86277 | 420 | 415,1901 | 0,388763 | 1 | 0,95743 |
| 0,053777 | 367 | 361,5639 | 0,957142 | 1 | 0,981063 |
| 0,188333 | 280 | 274,7782 | 0,850755 | 1 | 0,966155 |
| 0,64197 | 421 | 342,6873 | 0,521322 | 1 | 0,769172 |
| -0,06493 | 421 | 185,5 | 0,948303 | 1 | 0,980846 |
| 1,019276 | 420 | 374,9285 | 0,308729 | 1 | 0,943897 |
| -0,24525 | 367 | 327,3245 | 0,806416 | 1 | 0,950149 |
| 1,197285 | 280 | 246,3251 | 0,232346 | 1 | 0,659852 |
| 0,388349 | 421 | 399,661 | 0,697965 | 1 | 0,872167 |
| 0,922262 | 421 | 215,1303 | 0,357425 | 1 | 0,643758 |
| -0,60527 | 420 | 415,415 | 0,54533 | 1 | 0,960096 |
| 0,961399 | 367 | 362,9631 | 0,336992 | 1 | 0,662616 |
| 1,040485 | 280 | 275,7698 | 0,299026 | 1 | 0,723324 |
| 0,103522 | 421 | 300,2391 | 0,917618 | 1 | 0,966828 |
| 0,270287 | 421 | 175,0153 | 0,787258 | 1 | 0,917996 |
| -0,50336 | 420 | 331,7107 | 0,615047 | 1 | 0,964878 |
| -0,39951 | 367 | 278,5258 | 0,689825 | 1 | 0,903032 |
| 0,447709 | 280 | 276 | 0,654714 | 1 | 0,908644 |
| -1,84487 | 421 | 346,7801 | 0,06591 | 1 | 0,278224 |
| -1,58845 | 421 | 190,3869 | 0,113843 | 1 | 0,391524 |
| -0,54706 | 420 | 383,0963 | 0,584658 | 1 | 0,964878 |
| -1,40143 | 367 | 345,617 | 0,161983 | 1 | 0,508558 |
| -0,39139 | 280 | 267,247 | 0,695821 | 1 | 0,92034 |

| | | | | | |
|---|---|---|---|---|---|
| 0,829432 | 421 | 340,1244 | 0,407442 | 1 | 0,695339 |
| -1,15401 | 421 | 187,737 | 0,249961 | 1 | 0,548314 |
| -2,6253 | 420 | 374,8251 | 0,009012 | 1 | 0,582436 |
| 1,047481 | 367 | 332,7619 | 0,295638 | 1 | 0,627502 |
| -0,22894 | 280 | 256,0328 | 0,819099 | 1 | 0,95778 |
| 1,132594 | 421 | 346,4609 | 0,258168 | 1 | 0,550898 |
| 1,00629 | 421 | 189,2106 | 0,315561 | 1 | 0,614499 |
| 0,461719 | 420 | 378,8452 | 0,644548 | 1 | 0,964878 |
| -0,95203 | 367 | 331,7855 | 0,341773 | 1 | 0,664831 |
| 0,314982 | 280 | 256,6433 | 0,753032 | 1 | 0,942071 |
| 0,781893 | 421 | 371,248 | 0,434776 | 1 | 0,706341 |
| -0,1352 | 421 | 199,5834 | 0,892588 | 1 | 0,958882 |
| 0,41175 | 420 | 400,9927 | 0,680743 | 1 | 0,96781 |
| -1,43818 | 367 | 343,5719 | 0,151293 | 1 | 0,499998 |
| -0,40809 | 280 | 262,8844 | 0,683538 | 1 | 0,913202 |
| -1,38026 | 421 | 327,5002 | 0,168448 | 1 | 0,442288 |
| -0,71109 | 421 | 185,3717 | 0,477925 | 1 | 0,734183 |
| -0,48982 | 420 | 364,2548 | 0,624559 | 1 | 0,964878 |
| -0,92881 | 367 | 308,457 | 0,353714 | 1 | 0,673889 |
| -0,70161 | 280 | 258,5602 | 0,483554 | 1 | 0,824339 |
| -0,50039 | 421 | 364,4945 | 0,617106 | 1 | 0,826526 |
| -0,26975 | 421 | 193,9945 | 0,787637 | 1 | 0,917996 |
| -0,59902 | 420 | 395,8895 | 0,549503 | 1 | 0,960096 |
| 1,363281 | 367 | 347,5077 | 0,173677 | 1 | 0,523018 |
| -0,42467 | 280 | 257,1062 | 0,671431 | 1 | 0,908644 |
| -0,9387 | 421 | 347,4999 | 0,348539 | 1 | 0,637437 |
| -0,19813 | 421 | 188,7908 | 0,843154 | 1 | 0,941476 |
| 0,130588 | 420 | 382,5389 | 0,89617 | 1 | 0,98503 |
| -1,50045 | 367 | 348,1882 | 0,134403 | 1 | 0,477542 |
| -1,28591 | 280 | 255,299 | 0,199639 | 1 | 0,631529 |
| -2,0742 | 421 | 311,356 | 0,038882 | 1 | 0,222577 |
| -1,72853 | 421 | 182,9966 | 0,085581 | 1 | 0,365475 |
| -0,92737 | 420 | 356,7293 | 0,354362 | 1 | 0,947373 |
| -0,33812 | 367 | 317,8817 | 0,735498 | 1 | 0,923669 |
| 0,030715 | 280 | 255,9556 | 0,975521 | 1 | 0,99236 |
| 3,901761 | 421 | 400,7965 | 1,12E-04 | 0,120378 | 0,033751 |
| 2,99969 | 421 | 214,476 | 0,003022 | 1 | 0,114447 |
| 1,328991 | 420 | 415,4531 | 0,18458 | 1 | 0,923408 |
| 2,250173 | 367 | 361,0465 | 0,025039 | 1 | 0,247087 |
| 1,796358 | 280 | 274,9372 | 0,073535 | 1 | 0,443271 |
| 1,060486 | 421 | 385,3988 | 0,289587 | 1 | 0,586895 |
| 0,81751 | 421 | 205,2503 | 0,414585 | 1 | 0,685818 |
| 0,384404 | 420 | 408,7972 | 0,700879 | 1 | 0,968866 |
| 1,767738 | 367 | 359,833 | 0,077952 | 1 | 0,381409 |
| -0,54766 | 280 | 274,4276 | 0,584368 | 1 | 0,87561 |
| -1,63572 | 421 | 317,9097 | 0,102887 | 1 | 0,344557 |
| -0,9115 | 421 | 180,5143 | 0,363249 | 1 | 0,646044 |
| -0,14839 | 420 | 354,8761 | 0,882116 | 1 | 0,982463 |

| | | | | | |
|---|---|---|---|---|---|
| 0,129521 | 367 | 301,6522 | 0,897032 | 1 | 0,970443 |
| -0,36216 | 280 | 257,064 | 0,717527 | 1 | 0,931363 |
| 1,410462 | 421 | 380,0976 | 0,159221 | 1 | 0,432185 |
| 1,565061 | 421 | 203,3604 | 0,119123 | 1 | 0,396436 |
| -0,22931 | 420 | 407,2742 | 0,818742 | 1 | 0,97003 |
| 2,436802 | 367 | 357,9244 | 0,015304 | 1 | 0,204372 |
| 1,318762 | 280 | 267,3297 | 0,188377 | 1 | 0,623041 |
| -3,73855 | 421 | 417 | 2,11E-04 | 0,226527 | 0,04642 |
| -3,51997 | 421 | 417 | 4,79E-04 | 0,51221 | 0,048139 |
| -0,37198 | 420 | 416 | 0,710097 | 1 | 0,968866 |
| -1,82824 | 367 | 363 | 0,068334 | 1 | 0,364488 |
| -1,42348 | 280 | 276 | 0,155728 | 1 | 0,572395 |
| -2,43442 | 421 | 341,3735 | 0,015428 | 1 | 0,147897 |
| -1,43895 | 421 | 188,218 | 0,151824 | 1 | 0,432513 |
| -2,28084 | 420 | 378,1639 | 0,023113 | 1 | 0,75904 |
| -2,71173 | 367 | 330,6376 | 0,007043 | 0,901534 | 0,180403 |
| 0,597378 | 280 | 261,1071 | 0,550772 | 1 | 0,853046 |
| 0,993199 | 421 | 399,3848 | 0,321214 | 1 | 0,615134 |
| -0,2552 | 421 | 215,767 | 0,798815 | 1 | 0,921861 |
| -0,70265 | 420 | 415,2057 | 0,482668 | 1 | 0,960096 |
| -1,09968 | 367 | 360,8198 | 0,272203 | 1 | 0,609149 |
| -0,69219 | 280 | 270,5742 | 0,48941 | 1 | 0,826999 |
| -3,06703 | 421 | 389,0227 | 0,002313 | 0,349283 | 0,069718 |
| -1,82436 | 421 | 207,9554 | 0,069533 | 1 | 0,336932 |
| -0,3212 | 420 | 412,2113 | 0,748221 | 1 | 0,969427 |
| -1,60329 | 367 | 361,405 | 0,109744 | 1 | 0,435936 |
| -1,87327 | 280 | 275,0017 | 0,062092 | 1 | 0,422605 |
| 0,469462 | 421 | 355,1818 | 0,639028 | 1 | 0,834842 |
| 0,49115 | 421 | 194,0539 | 0,623875 | 1 | 0,830785 |
| 0,595835 | 420 | 388,2638 | 0,551633 | 1 | 0,960096 |
| 1,21411 | 367 | 329,9456 | 0,225574 | 1 | 0,576775 |
| -0,78293 | 280 | 269,321 | 0,434357 | 1 | 0,803161 |
| -1,40986 | 421 | 356,8934 | 0,159453 | 1 | 0,432185 |
| -1,05242 | 421 | 192,0844 | 0,293931 | 1 | 0,590656 |
| -0,83925 | 420 | 391,3615 | 0,40184 | 1 | 0,95743 |
| -1,3716 | 367 | 352,3526 | 0,171061 | 1 | 0,51869 |
| -1,30659 | 280 | 266,6104 | 0,192478 | 1 | 0,62543 |
| 1,31624 | 421 | 357,9134 | 0,188936 | 1 | 0,472953 |
| 0,13414 | 421 | 195,1166 | 0,89343 | 1 | 0,958882 |
| -0,73493 | 420 | 392,1041 | 0,462824 | 1 | 0,960096 |
| 1,82822 | 367 | 324,3768 | 0,068435 | 1 | 0,364488 |
| 1,201864 | 280 | 259,2291 | 0,230513 | 1 | 0,657169 |
| 1,55234 | 421 | 311,4504 | 0,121597 | 1 | 0,382559 |
| 0,66257 | 421 | 180,1974 | 0,508452 | 1 | 0,754914 |
| -0,62679 | 420 | 346,1786 | 0,531213 | 1 | 0,960096 |
| 1,132892 | 367 | 308,4693 | 0,258139 | 1 | 0,609149 |
| -1,27482 | 280 | 254,4733 | 0,203536 | 1 | 0,632429 |
| 2,386537 | 421 | 400,2074 | 0,017472 | 1 | 0,153976 |

| | | | | | |
|---|---|---|---|---|---|
| 1,445398 | 421 | 215,9583 | 0,149796 | 1 | 0,430627 |
| 0,456734 | 420 | 415,2025 | 0,648101 | 1 | 0,964878 |
| 1,789409 | 367 | 361,9827 | 0,074385 | 1 | 0,374909 |
| 0,655244 | 280 | 274,8551 | 0,512858 | 1 | 0,831912 |
| -1,37212 | 421 | 339,3431 | 0,170932 | 1 | 0,443364 |
| 0,921374 | 421 | 186,4666 | 0,358046 | 1 | 0,643758 |
| -0,38192 | 420 | 373,7545 | 0,702736 | 1 | 0,968866 |
| -0,19896 | 367 | 316,362 | 0,84242 | 1 | 0,956935 |
| 0,307367 | 280 | 256,6904 | 0,758813 | 1 | 0,94273 |
| -1,5225 | 421 | 382,7633 | 0,128709 | 1 | 0,396656 |
| -0,91079 | 421 | 202,9694 | 0,363484 | 1 | 0,646044 |
| -0,08561 | 420 | 408,1738 | 0,931816 | 1 | 0,991559 |
| 1,57007 | 367 | 357,4785 | 0,117283 | 1 | 0,445308 |
| 0,038908 | 280 | 263,2134 | 0,968993 | 1 | 0,991502 |
| 1,142523 | 421 | 374,1848 | 0,253967 | 1 | 0,546261 |
| 0,827211 | 421 | 200,6952 | 0,4091 | 1 | 0,681984 |
| 1,126745 | 420 | 404,3656 | 0,260519 | 1 | 0,943897 |
| 0,397955 | 367 | 349,7055 | 0,690906 | 1 | 0,903032 |
| -0,23379 | 280 | 267,9997 | 0,815328 | 1 | 0,95747 |
| 1,646284 | 421 | 317,8058 | 0,100694 | 1 | 0,342121 |
| 1,158926 | 421 | 178,7691 | 0,248032 | 1 | 0,548314 |
| -0,93997 | 420 | 349,4337 | 0,347885 | 1 | 0,947373 |
| 0,02995 | 367 | 305,3103 | 0,976127 | 1 | 0,9914 |
| 1,329027 | 280 | 247,7589 | 0,185062 | 1 | 0,617008 |
| -0,51097 | 421 | 338,9512 | 0,609703 | 1 | 0,824796 |
| 0,827344 | 421 | 186,6249 | 0,409099 | 1 | 0,681984 |
| 1,205747 | 420 | 371,8745 | 0,228682 | 1 | 0,943055 |
| -0,43148 | 367 | 328,6805 | 0,666404 | 1 | 0,894765 |
| -1,47519 | 280 | 259,6337 | 0,141374 | 1 | 0,554427 |
| -2,44152 | 421 | 356,1655 | 0,015112 | 1 | 0,147897 |
| 0,141632 | 421 | 193,3078 | 0,887518 | 1 | 0,958088 |
| 0,267525 | 420 | 389,4319 | 0,789206 | 1 | 0,969427 |
| 0,036631 | 367 | 340,92 | 0,970801 | 1 | 0,98918 |
| -1,63462 | 280 | 264,0905 | 0,10332 | 1 | 0,486574 |
| 0,757773 | 421 | 338,1336 | 0,449115 | 1 | 0,714643 |
| 1,055934 | 421 | 186,5458 | 0,292364 | 1 | 0,589027 |
| 0,880021 | 420 | 371,6187 | 0,379417 | 1 | 0,95743 |
| -1,39572 | 367 | 305,2818 | 0,163812 | 1 | 0,510294 |
| 0,219263 | 280 | 262,5117 | 0,826616 | 1 | 0,96121 |
| 0,73206 | 421 | 395,133 | 0,464566 | 1 | 0,727964 |
| 1,533859 | 421 | 212,1401 | 0,126554 | 1 | 0,410224 |
| 1,186919 | 420 | 414,2784 | 0,23594 | 1 | 0,943055 |
| -0,81851 | 367 | 358,9971 | 0,41361 | 1 | 0,724248 |
| -0,00307 | 280 | 269,7969 | 0,997551 | 1 | 0,998213 |
| -1,85657 | 421 | 280,4075 | 0,064421 | 1 | 0,275801 |
| -1,46944 | 421 | 173,1969 | 0,143528 | 1 | 0,42451 |
| -0,8907 | 420 | 309,8846 | 0,373779 | 1 | 0,95743 |
| -0,0874 | 367 | 363 | 0,930402 | 1 | 0,977084 |

| | | | | | |
|---|---|---|---|---|---|
| 0,176781 | 280 | 276 | 0,85981 | 1 | 0,966655 |
| 1,768792 | 421 | 343,0908 | 0,077817 | 1 | 0,302838 |
| 1,333883 | 421 | 187,0438 | 0,183864 | 1 | 0,473593 |
| 1,856176 | 420 | 375,8585 | 0,064211 | 1 | 0,848246 |
| 1,605338 | 367 | 325,0931 | 0,10939 | 1 | 0,435936 |
| 0,879154 | 280 | 261,4382 | 0,380125 | 1 | 0,770948 |
| 2,03357 | 421 | 324,4714 | 0,042808 | 1 | 0,232058 |
| 1,708226 | 421 | 183,3573 | 0,089287 | 1 | 0,37156 |
| 1,529995 | 420 | 362,7914 | 0,126889 | 1 | 0,885289 |
| 1,633506 | 367 | 323,5534 | 0,103335 | 1 | 0,426647 |
| 0,160648 | 280 | 255,9832 | 0,872497 | 1 | 0,969656 |
| -0,03 | 421 | 311,1052 | 0,976083 | 1 | 0,995982 |
| 3,134364 | 421 | 179,1211 | 0,002013 | 1 | 0,099921 |
| 0,391431 | 420 | 346,4114 | 0,695719 | 1 | 0,968866 |
| 0,451696 | 367 | 322,2967 | 0,651792 | 1 | 0,88811 |
| 0,062657 | 280 | 259,6513 | 0,950088 | 1 | 0,982409 |
| -0,82894 | 421 | 317,7811 | 0,407763 | 1 | 0,695339 |
| -1,67399 | 421 | 182,4239 | 0,095847 | 1 | 0,378119 |
| -1,31168 | 420 | 352,0339 | 0,190484 | 1 | 0,925729 |
| -1,06381 | 367 | 313,6698 | 0,288234 | 1 | 0,624991 |
| -1,50483 | 280 | 251,0812 | 0,133625 | 1 | 0,543276 |
| -1,26936 | 421 | 350,4908 | 0,205156 | 1 | 0,494095 |
| -1,85984 | 421 | 191,7577 | 0,06444 | 1 | 0,324784 |
| -1,59492 | 420 | 385,8178 | 0,11155 | 1 | 0,848246 |
| -1,14985 | 367 | 338,8576 | 0,251016 | 1 | 0,605249 |
| -1,3819 | 280 | 263,6373 | 0,168173 | 1 | 0,59271 |
| -0,24899 | 421 | 353,5061 | 0,803511 | 1 | 0,922616 |
| -1,06955 | 421 | 191,5343 | 0,286169 | 1 | 0,584263 |
| 0,430243 | 420 | 387,728 | 0,667258 | 1 | 0,96781 |
| 0,74817 | 367 | 327,067 | 0,454895 | 1 | 0,758509 |
| -0,25801 | 280 | 256,0992 | 0,796607 | 1 | 0,954269 |
| -0,34374 | 421 | 317,2345 | 0,731271 | 1 | 0,892329 |
| 2,54E-04 | 421 | 180,765 | 0,999797 | 1 | 0,999797 |
| 0,666388 | 420 | 347,1445 | 0,505606 | 1 | 0,960096 |
| -0,71776 | 367 | 304,3181 | 0,473458 | 1 | 0,776795 |
| -1,25768 | 280 | 253,1803 | 0,209667 | 1 | 0,64091 |
| -1,21324 | 421 | 374,601 | 0,225801 | 1 | 0,518147 |
| -1,0849 | 421 | 199,5437 | 0,279274 | 1 | 0,578974 |
| -0,19418 | 420 | 405,9683 | 0,846134 | 1 | 0,973894 |
| -0,28893 | 367 | 352,4269 | 0,772806 | 1 | 0,938452 |
| 0,425654 | 280 | 265,5601 | 0,670705 | 1 | 0,908644 |
| 1,023192 | 421 | 340,636 | 0,306943 | 1 | 0,604218 |
| 0,993666 | 421 | 188,9934 | 0,321656 | 1 | 0,617772 |
| 1,232569 | 420 | 375,9061 | 0,218507 | 1 | 0,943055 |
| 0,001998 | 367 | 329,1846 | 0,998407 | 1 | 0,998887 |
| 0,193537 | 280 | 250,9208 | 0,846695 | 1 | 0,965181 |
| -0,05525 | 421 | 320,6585 | 0,955976 | 1 | 0,987258 |
| -0,29809 | 421 | 182,0485 | 0,765972 | 1 | 0,90393 |

| | | | | | |
|---|---|---|---|---|---|
| 1,306669 | 420 | 355,4055 | 0,19217 | 1 | 0,925729 |
| 0,730374 | 367 | 306,3831 | 0,46572 | 1 | 0,771623 |
| 0,613711 | 280 | 247,1576 | 0,539971 | 1 | 0,84937 |
| -1,56241 | 421 | 327,0597 | 0,119158 | 1 | 0,380448 |
| -1,66084 | 421 | 182,9281 | 0,098459 | 1 | 0,379251 |
| -0,70573 | 420 | 363,1676 | 0,48081 | 1 | 0,960096 |
| -1,55484 | 367 | 339,6815 | 0,120916 | 1 | 0,452158 |
| -1,21065 | 280 | 262,1937 | 0,227118 | 1 | 0,656943 |
| -0,87553 | 421 | 352,9236 | 0,381881 | 1 | 0,671614 |
| 0,381297 | 421 | 189,2242 | 0,70341 | 1 | 0,866753 |
| 0,074274 | 420 | 386,5936 | 0,940831 | 1 | 0,992811 |
| 0,28974 | 367 | 336,8625 | 0,772194 | 1 | 0,938452 |
| 1,557989 | 280 | 266,2874 | 0,120424 | 1 | 0,526024 |
| -0,09615 | 421 | 324,005 | 0,923459 | 1 | 0,966828 |
| 0,95518 | 421 | 184,874 | 0,340734 | 1 | 0,635503 |
| 0,572745 | 420 | 358,3917 | 0,567176 | 1 | 0,962539 |
| 2,094393 | 367 | 316,4171 | 0,03702 | 1 | 0,28464 |
| 0,89314 | 280 | 261,6259 | 0,372603 | 1 | 0,763914 |
| -0,82293 | 421 | 373,8249 | 0,411071 | 1 | 0,695339 |
| -0,88489 | 421 | 199,821 | 0,377282 | 1 | 0,655017 |
| -0,61746 | 420 | 403,3481 | 0,537282 | 1 | 0,960096 |
| -0,33316 | 367 | 354,2265 | 0,739211 | 1 | 0,923669 |
| -1,34137 | 280 | 273,7954 | 0,180911 | 1 | 0,607861 |
| -0,01213 | 421 | 363,0178 | 0,990332 | 1 | 0,998054 |
| 0,426692 | 421 | 196,0177 | 0,670072 | 1 | 0,851432 |
| -0,35669 | 420 | 394,827 | 0,721515 | 1 | 0,969427 |
| -0,52163 | 367 | 340,2119 | 0,602271 | 1 | 0,864402 |
| 0,650026 | 280 | 249,7554 | 0,516273 | 1 | 0,83211 |
| -1,493 | 421 | 362,518 | 0,136306 | 1 | 0,407576 |
| 0,017369 | 421 | 195,6885 | 0,98616 | 1 | 0,995407 |
| -0,06048 | 420 | 394,699 | 0,951801 | 1 | 0,992811 |
| -0,17366 | 367 | 349,0866 | 0,862231 | 1 | 0,959636 |
| 0,977115 | 280 | 263,5786 | 0,329408 | 1 | 0,752282 |
| 0,457642 | 421 | 309,7943 | 0,64753 | 1 | 0,839784 |
| 0,777003 | 421 | 178,5968 | 0,438185 | 1 | 0,704743 |
| -0,06646 | 420 | 343,395 | 0,947053 | 1 | 0,992811 |
| -0,0688 | 367 | 311,0989 | 0,945196 | 1 | 0,979539 |
| 0,287794 | 280 | 253,2867 | 0,77374 | 1 | 0,947219 |
| -3,23632 | 421 | 333,5868 | 0,001332 | 1 | 0,066922 |
| -1,82884 | 421 | 187,1584 | 0,069015 | 1 | 0,335504 |
| -1,82049 | 420 | 371,2255 | 0,069489 | 1 | 0,848246 |
| -2,62787 | 367 | 325,9329 | 0,008999 | 1 | 0,183886 |
| -2,01893 | 280 | 256,434 | 0,044535 | 1 | 0,387749 |
| -2,84947 | 421 | 412,4442 | 0,004599 | 0,680607 | 0,087964 |
| -3,09432 | 421 | 232,4301 | 0,002214 | 0,329944 | 0,101124 |
| -0,94628 | 420 | 413,5522 | 0,344558 | 1 | 0,947373 |
| -1,44648 | 367 | 358,4311 | 0,148917 | 1 | 0,499998 |
| -1,06444 | 280 | 275,8935 | 0,288058 | 1 | 0,71621 |

| | | | | | |
|---|---|---|---|---|---|
| -1,26197 | 421 | 307,7015 | 0,207915 | 1 | 0,496728 |
| -1,24302 | 421 | 180,3033 | 0,215474 | 1 | 0,514565 |
| -0,80977 | 420 | 340,8617 | 0,418634 | 1 | 0,95743 |
| 0,053043 | 367 | 300,1645 | 0,957733 | 1 | 0,981063 |
| -1,73668 | 280 | 246,5673 | 0,083693 | 1 | 0,456738 |
| 0,642878 | 421 | 342,7956 | 0,520733 | 1 | 0,769172 |
| -0,0651 | 421 | 185,5431 | 0,948162 | 1 | 0,980846 |
| 1,019885 | 420 | 375,0228 | 0,308441 | 1 | 0,943897 |
| -0,24022 | 367 | 327,39 | 0,81031 | 1 | 0,950149 |
| 1,19635 | 280 | 246,3481 | 0,23271 | 1 | 0,659852 |
| 1,548635 | 421 | 339,5633 | 0,122401 | 1 | 0,383489 |
| 0,515723 | 421 | 184,9021 | 0,606664 | 1 | 0,821338 |
| 1,796823 | 420 | 368,1854 | 0,073183 | 1 | 0,848246 |
| 0,818305 | 367 | 322,4226 | 0,413787 | 1 | 0,724248 |
| -1,66544 | 280 | 247,9866 | 0,097088 | 1 | 0,480557 |
| -2,09611 | 421 | 343,2201 | 0,036805 | 1 | 0,217944 |
| 0,556581 | 421 | 187,8317 | 0,578476 | 1 | 0,794719 |
| -0,10644 | 420 | 379,4677 | 0,915292 | 1 | 0,988069 |
| -0,06058 | 367 | 329,9245 | 0,951732 | 1 | 0,979686 |
| 0,926773 | 280 | 269,4529 | 0,354874 | 1 | 0,761846 |
| -0,3135 | 421 | 350,7521 | 0,754087 | 1 | 0,898125 |
| 0,187014 | 421 | 190,2869 | 0,851849 | 1 | 0,944105 |
| -0,18102 | 420 | 383,9701 | 0,856448 | 1 | 0,975561 |
| 1,459506 | 367 | 337,7185 | 0,145355 | 1 | 0,496835 |
| 1,46994 | 280 | 269,276 | 0,142746 | 1 | 0,554427 |
| -1,68325 | 421 | 337,7353 | 0,093251 | 1 | 0,327788 |
| -1,27726 | 421 | 185,8204 | 0,203103 | 1 | 0,497853 |
| 0,359925 | 420 | 373,2647 | 0,719106 | 1 | 0,969427 |
| -0,97064 | 367 | 335,1133 | 0,332428 | 1 | 0,660908 |
| -0,58598 | 280 | 260,8652 | 0,558396 | 1 | 0,858676 |
| 1,608267 | 421 | 364,8168 | 0,108642 | 1 | 0,355148 |
| 1,528897 | 421 | 196,7771 | 0,127896 | 1 | 0,410224 |
| 0,525146 | 420 | 397,3139 | 0,599775 | 1 | 0,964878 |
| 2,199953 | 367 | 346,456 | 0,02847 | 1 | 0,257802 |
| 0,685383 | 280 | 269,5274 | 0,493691 | 1 | 0,827962 |
| -0,5169 | 421 | 343,1947 | 0,605556 | 1 | 0,822137 |
| 0,572641 | 421 | 187,6892 | 0,567573 | 1 | 0,78927 |
| 0,853253 | 420 | 377,2842 | 0,39406 | 1 | 0,95743 |
| 1,636877 | 367 | 314,2438 | 0,102657 | 1 | 0,426647 |
| 0,842065 | 280 | 250,094 | 0,400556 | 1 | 0,784757 |
| -1,43279 | 421 | 340,7678 | 0,152834 | 1 | 0,42476 |
| -1,2584 | 421 | 188,5543 | 0,209801 | 1 | 0,509953 |
| -0,58606 | 420 | 377,0662 | 0,558186 | 1 | 0,960096 |
| 0,050782 | 367 | 328,0538 | 0,95953 | 1 | 0,981063 |
| -0,04501 | 280 | 257,6472 | 0,964135 | 1 | 0,989749 |
| -0,13132 | 421 | 288,0161 | 0,895612 | 1 | 0,962 |
| 0,625201 | 421 | 173,0203 | 0,532663 | 1 | 0,769894 |
| -1,39833 | 420 | 313,7467 | 0,163 | 1 | 0,914005 |

| | | | | | |
|---|---|---|---|---|---|
| 0,654173 | 367 | 282,4149 | 0,513533 | 1 | 0,797829 |
| 1,728653 | 280 | 276 | 0,084989 | 1 | 0,45831 |
| 0,584696 | 421 | 407,6488 | 0,559076 | 1 | 0,792594 |
| 1,473617 | 421 | 223,2317 | 0,141993 | 1 | 0,423499 |
| -0,79298 | 420 | 415,8011 | 0,428239 | 1 | 0,957502 |
| 0,864223 | 367 | 362,9965 | 0,388036 | 1 | 0,702006 |
| -0,14288 | 280 | 274,2517 | 0,886492 | 1 | 0,973784 |
| 0,650955 | 421 | 295,4838 | 0,515582 | 1 | 0,767364 |
| -1,60585 | 421 | 176,7681 | 0,110093 | 1 | 0,389736 |
| -0,90757 | 420 | 324,441 | 0,364777 | 1 | 0,956033 |
| 0,216944 | 367 | 363 | 0,828374 | 1 | 0,956109 |
| -0,38424 | 280 | 276 | 0,701095 | 1 | 0,92034 |
| 0,474591 | 421 | 365,7979 | 0,635362 | 1 | 0,83455 |
| 0,267114 | 421 | 194,282 | 0,789665 | 1 | 0,918329 |
| 1,194426 | 420 | 393,1131 | 0,233031 | 1 | 0,943055 |
| -0,18938 | 367 | 339,8265 | 0,84991 | 1 | 0,956935 |
| -1,43621 | 280 | 258,8195 | 0,15215 | 1 | 0,565026 |
| -0,57532 | 421 | 329,1036 | 0,565469 | 1 | 0,796637 |
| -0,33748 | 421 | 185,3144 | 0,736138 | 1 | 0,884613 |
| 0,706358 | 420 | 364,1794 | 0,480417 | 1 | 0,960096 |
| -0,14566 | 367 | 318,7942 | 0,884282 | 1 | 0,962871 |
| -0,86947 | 280 | 276 | 0,385345 | 1 | 0,773166 |
| -1,36725 | 421 | 370,0179 | 0,172377 | 1 | 0,444059 |
| 0,143105 | 421 | 198,1271 | 0,886353 | 1 | 0,957782 |
| 2,009772 | 420 | 403,2478 | 0,045121 | 1 | 0,846132 |
| -0,99687 | 367 | 347,621 | 0,31952 | 1 | 0,653184 |
| -0,25949 | 280 | 270,0145 | 0,795457 | 1 | 0,954269 |
| 0,650544 | 421 | 319,7623 | 0,515808 | 1 | 0,767364 |
| 0,609407 | 421 | 181,3252 | 0,543018 | 1 | 0,774002 |
| 1,92972 | 420 | 356,3966 | 0,054435 | 1 | 0,848246 |
| 0,006995 | 367 | 312,0876 | 0,994423 | 1 | 0,997335 |
| 1,405782 | 280 | 253,9344 | 0,161011 | 1 | 0,581516 |
| -1,49361 | 421 | 341,318 | 0,136202 | 1 | 0,407576 |
| -2,04462 | 421 | 188,6768 | 0,042281 | 1 | 0,273775 |
| 0,679328 | 420 | 376,3736 | 0,497348 | 1 | 0,960096 |
| -1,24423 | 367 | 339,1844 | 0,214274 | 1 | 0,564902 |
| -0,78975 | 280 | 261,5524 | 0,430387 | 1 | 0,801886 |
| -1,36174 | 421 | 406,6334 | 0,174033 | 1 | 0,446794 |
| -1,51066 | 421 | 223,1146 | 0,13229 | 1 | 0,414338 |
| 0,199394 | 420 | 415,9963 | 0,842052 | 1 | 0,973894 |
| 1,410763 | 367 | 362,8602 | 0,159171 | 1 | 0,504558 |
| -1,85002 | 280 | 274,0597 | 0,065387 | 1 | 0,424733 |
| 0,250808 | 421 | 307,2213 | 0,802131 | 1 | 0,922616 |
| 1,363901 | 421 | 180,8256 | 0,174294 | 1 | 0,459198 |
| -0,3151 | 420 | 339,1034 | 0,75288 | 1 | 0,969427 |
| 1,913014 | 367 | 301,0442 | 0,056695 | 1 | 0,337704 |
| -0,32002 | 280 | 276 | 0,749198 | 1 | 0,942071 |
| -2,31516 | 421 | 302,9563 | 0,021272 | 1 | 0,172351 |

| | | | | | |
|---|---|---|---|---|---|
| -1,19489 | 421 | 176,0421 | 0,233739 | 1 | 0,533704 |
| -2,44576 | 420 | 332,3802 | 0,014973 | 1 | 0,723749 |
| -1,13587 | 367 | 271,9282 | 0,257011 | 1 | 0,609149 |
| -0,2593 | 280 | 248,4418 | 0,795622 | 1 | 0,954269 |
| 1,890901 | 421 | 344,4608 | 0,059477 | 1 | 0,268966 |
| 1,716092 | 421 | 184,1687 | 0,087827 | 1 | 0,368676 |
| 0,925355 | 420 | 377,4746 | 0,355372 | 1 | 0,947373 |
| 2,674114 | 367 | 327,969 | 0,007868 | 1 | 0,180403 |
| 1,694345 | 280 | 263,7913 | 0,09138 | 1 | 0,468147 |
| -0,328 | 421 | 320,1103 | 0,743123 | 1 | 0,895193 |
| -0,44367 | 421 | 183,8531 | 0,6578 | 1 | 0,841515 |
| -0,67058 | 420 | 355,7484 | 0,502926 | 1 | 0,960096 |
| -2,50566 | 367 | 300,2041 | 0,012752 | 1 | 0,189546 |
| -0,62212 | 280 | 276 | 0,534378 | 1 | 0,84326 |
| -0,7189 | 421 | 378,8003 | 0,472647 | 1 | 0,731548 |
| 0,300073 | 421 | 203,1476 | 0,764428 | 1 | 0,903524 |
| -0,41314 | 420 | 405,274 | 0,679724 | 1 | 0,96781 |
| 0,959957 | 367 | 355,8699 | 0,337729 | 1 | 0,662705 |
| 0,281089 | 280 | 265,5483 | 0,778861 | 1 | 0,951006 |
| -1,98494 | 421 | 353,7277 | 0,047923 | 1 | 0,241202 |
| -0,40808 | 421 | 191,2606 | 0,683674 | 1 | 0,859821 |
| 0,198156 | 420 | 388,8078 | 0,843026 | 1 | 0,973894 |
| -1,84448 | 367 | 340,3121 | 0,065982 | 1 | 0,358968 |
| -0,02165 | 280 | 267,7366 | 0,982744 | 1 | 0,99236 |
| -0,12693 | 421 | 353,5926 | 0,899066 | 1 | 0,965023 |
| 0,22111 | 421 | 193,2482 | 0,825241 | 1 | 0,932262 |
| 0,066216 | 420 | 387,3938 | 0,94724 | 1 | 0,992811 |
| 1,120453 | 367 | 343,4063 | 0,263304 | 1 | 0,609149 |
| 0,654433 | 280 | 265,8496 | 0,513399 | 1 | 0,831912 |
| -1,65427 | 421 | 327,8577 | 0,099029 | 1 | 0,339344 |
| -1,60511 | 421 | 185,5298 | 0,110171 | 1 | 0,389736 |
| -1,68817 | 420 | 362,9176 | 0,092237 | 1 | 0,848246 |
| -1,04353 | 367 | 326,376 | 0,297477 | 1 | 0,629631 |
| -2,28104 | 280 | 257,1461 | 0,023364 | 1 | 0,313396 |
| 0,39868 | 421 | 331,3244 | 0,690386 | 1 | 0,867733 |
| 1,029279 | 421 | 184,9883 | 0,304692 | 1 | 0,602952 |
| 1,705259 | 420 | 358,9219 | 0,089011 | 1 | 0,848246 |
| 0,510213 | 367 | 346,0332 | 0,610227 | 1 | 0,870021 |
| -0,4639 | 280 | 267,7003 | 0,643094 | 1 | 0,907804 |
| -0,78088 | 421 | 333,0233 | 0,435428 | 1 | 0,706341 |
| -0,39153 | 421 | 184,7259 | 0,695854 | 1 | 0,865526 |
| 1,328841 | 420 | 369,355 | 0,184721 | 1 | 0,923408 |
| -0,20698 | 367 | 338,616 | 0,836151 | 1 | 0,956935 |
| 0,342624 | 280 | 262,5246 | 0,732155 | 1 | 0,936637 |
| -1,61117 | 421 | 351,5163 | 0,10804 | 1 | 0,353949 |
| -2,49391 | 421 | 186,6105 | 0,013504 | 1 | 0,17172 |
| -1,72815 | 420 | 386,7296 | 0,08476 | 1 | 0,848246 |
| -1,65756 | 367 | 326,469 | 0,098367 | 1 | 0,414924 |

| | | | | | |
|---|---|---|---|---|---|
| 0,282665 | 280 | 251,8163 | 0,777666 | 1 | 0,950481 |
| -1,0721 | 421 | 335,1256 | 0,284448 | 1 | 0,583534 |
| -0,17983 | 421 | 185,6871 | 0,857481 | 1 | 0,945992 |
| 1,777447 | 420 | 371,7523 | 0,076312 | 1 | 0,848246 |
| 0,993627 | 367 | 338,4095 | 0,321114 | 1 | 0,653627 |
| 0,904791 | 280 | 262,0627 | 0,366407 | 1 | 0,763914 |
| -2,99271 | 421 | 304,0653 | 0,002992 | 1 | 0,077359 |
| -1,88228 | 421 | 174,3063 | 0,061465 | 1 | 0,317217 |
| -1,09787 | 420 | 337,9902 | 0,273042 | 1 | 0,943897 |
| -2,2849 | 367 | 305,5373 | 0,023004 | 1 | 0,237443 |
| -1,2039 | 280 | 261,0026 | 0,229721 | 1 | 0,657169 |
| -0,93156 | 421 | 354,9687 | 0,352195 | 1 | 0,641012 |
| -0,73605 | 421 | 190,677 | 0,462606 | 1 | 0,721079 |
| 1,201544 | 420 | 390,5083 | 0,230268 | 1 | 0,943055 |
| -0,47939 | 367 | 341,3564 | 0,631967 | 1 | 0,880707 |
| 2,465167 | 280 | 266,8391 | 0,014325 | 1 | 0,258576 |
| 0,792167 | 421 | 330,0358 | 0,428832 | 1 | 0,700922 |
| 1,035602 | 421 | 182,6297 | 0,301757 | 1 | 0,600041 |
| 1,498908 | 420 | 365,342 | 0,134761 | 1 | 0,890998 |
| -2,02428 | 367 | 311,4797 | 0,043795 | 1 | 0,299994 |
| 2,403749 | 280 | 253,0488 | 0,016948 | 1 | 0,266517 |
| 3,182281 | 421 | 338,4767 | 0,001597 | 1 | 0,069718 |
| 2,609771 | 421 | 186,8045 | 0,009795 | 1 | 0,149771 |
| -0,06028 | 420 | 373,2857 | 0,951966 | 1 | 0,992811 |
| 0,532222 | 367 | 326,6779 | 0,594934 | 1 | 0,85878 |
| 0,810406 | 280 | 258,9201 | 0,418451 | 1 | 0,795216 |
| -0,43325 | 421 | 350,4119 | 0,665098 | 1 | 0,853014 |
| -0,01155 | 421 | 192,6258 | 0,990795 | 1 | 0,997935 |
| 0,033063 | 420 | 384,7007 | 0,973642 | 1 | 0,997342 |
| 0,328657 | 367 | 331,8539 | 0,742622 | 1 | 0,923669 |
| 0,76294 | 280 | 264,0028 | 0,44618 | 1 | 0,813051 |
| -1,79525 | 421 | 342,1423 | 0,073497 | 1 | 0,292681 |
| -0,60055 | 421 | 189,0311 | 0,548859 | 1 | 0,776424 |
| -0,80185 | 420 | 380,9479 | 0,42314 | 1 | 0,95743 |
| -2,06229 | 367 | 334,8353 | 0,039952 | 1 | 0,28956 |
| -1,36996 | 280 | 258,8216 | 0,171886 | 1 | 0,598625 |
| -0,04601 | 421 | 355,1398 | 0,963331 | 1 | 0,991625 |
| 0,368883 | 421 | 192,06 | 0,712621 | 1 | 0,872396 |
| 0,369499 | 420 | 389,5752 | 0,711957 | 1 | 0,968866 |
| 1,055424 | 367 | 347,6904 | 0,291964 | 1 | 0,627013 |
| 0,631872 | 280 | 264,935 | 0,528015 | 1 | 0,839366 |
| 0,974149 | 421 | 386,599 | 0,330592 | 1 | 0,619505 |
| 0,894258 | 421 | 206,8264 | 0,372223 | 1 | 0,650743 |
| 0,798914 | 420 | 410,4331 | 0,424803 | 1 | 0,95743 |
| -1,22425 | 367 | 355,1665 | 0,221669 | 1 | 0,573978 |
| 0,290436 | 280 | 268,3605 | 0,771707 | 1 | 0,947038 |
| 1,270408 | 421 | 374,904 | 0,204727 | 1 | 0,494095 |
| 1,732662 | 421 | 199,1435 | 0,084704 | 1 | 0,365475 |

| | | | | | |
|---|---|---|---|---|---|
| 0,079496 | 420 | 403,9098 | 0,936678 | 1 | 0,992059 |
| 1,749937 | 367 | 355,5087 | 0,080992 | 1 | 0,381841 |
| 0,368772 | 280 | 269,2275 | 0,712588 | 1 | 0,928432 |
| -2,56285 | 421 | 374,0967 | 0,010772 | 1 | 0,12392 |
| -2,18485 | 421 | 200,3399 | 0,03006 | 1 | 0,238409 |
| -0,58644 | 420 | 405,4949 | 0,557905 | 1 | 0,960096 |
| -2,49256 | 367 | 351,1838 | 0,013143 | 1 | 0,192294 |
| -1,53629 | 280 | 270,6232 | 0,125635 | 1 | 0,536498 |
| 0,614188 | 421 | 360,004 | 0,539479 | 1 | 0,779259 |
| 1,15564 | 421 | 194,6438 | 0,249245 | 1 | 0,548314 |
| 0,922049 | 420 | 390,6767 | 0,357072 | 1 | 0,947373 |
| 0,1739 | 367 | 343,6219 | 0,862047 | 1 | 0,959636 |
| -1,82211 | 280 | 265,6231 | 0,069563 | 1 | 0,431405 |
| 1,375922 | 421 | 393,7408 | 0,169628 | 1 | 0,442288 |
| -0,38258 | 421 | 210,293 | 0,70242 | 1 | 0,866241 |
| -0,94912 | 420 | 412,4237 | 0,343115 | 1 | 0,947373 |
| -0,49777 | 367 | 358,0859 | 0,61895 | 1 | 0,875495 |
| 1,77856 | 280 | 272,7185 | 0,076426 | 1 | 0,450485 |
| -2,08912 | 421 | 384,418 | 0,037354 | 1 | 0,217944 |
| -1,72889 | 421 | 207,0354 | 0,085319 | 1 | 0,365475 |
| 0,052675 | 420 | 410,7598 | 0,958017 | 1 | 0,996364 |
| -0,9638 | 367 | 358,9845 | 0,335796 | 1 | 0,662363 |
| -0,95743 | 280 | 275,6717 | 0,339189 | 1 | 0,756354 |
| -1,90609 | 421 | 295,8189 | 0,057608 | 1 | 0,266382 |
| -0,7113 | 421 | 175,3431 | 0,477842 | 1 | 0,734183 |
| -2,51393 | 420 | 323,3786 | 0,012425 | 1 | 0,668711 |
| 1,230984 | 367 | 299,6558 | 0,219294 | 1 | 0,572696 |
| 0,939831 | 280 | 247,5153 | 0,348221 | 1 | 0,761841 |
| 0,171215 | 421 | 357,8587 | 0,864151 | 1 | 0,951261 |
| -0,28567 | 421 | 192,5425 | 0,775439 | 1 | 0,910824 |
| -1,64002 | 420 | 393,5181 | 0,101801 | 1 | 0,848246 |
| -0,0721 | 367 | 341,9435 | 0,942565 | 1 | 0,979539 |
| 0,089733 | 280 | 261,7959 | 0,928568 | 1 | 0,979504 |
| -0,50526 | 421 | 333,2452 | 0,613709 | 1 | 0,825172 |
| -0,02332 | 421 | 185,5094 | 0,981424 | 1 | 0,993502 |
| -0,97425 | 420 | 368,0114 | 0,330571 | 1 | 0,947373 |
| 0,438009 | 367 | 317,632 | 0,661677 | 1 | 0,894303 |
| 0,471065 | 280 | 255,4135 | 0,637996 | 1 | 0,907449 |
| -0,02934 | 421 | 368,506 | 0,976611 | 1 | 0,995982 |
| 1,325009 | 421 | 195,3846 | 0,186715 | 1 | 0,474503 |
| -0,0681 | 420 | 398,5799 | 0,945737 | 1 | 0,992811 |
| 1,341111 | 367 | 343,4758 | 0,180771 | 1 | 0,527769 |
| 1,278247 | 280 | 268,4472 | 0,202266 | 1 | 0,632429 |
| 0,262623 | 421 | 364,0359 | 0,79299 | 1 | 0,919136 |
| 2,085738 | 421 | 194,9685 | 0,038304 | 1 | 0,268483 |
| 0,663133 | 420 | 394,6352 | 0,507633 | 1 | 0,960096 |
| -0,41428 | 367 | 339,9382 | 0,678931 | 1 | 0,899694 |
| -1,57093 | 280 | 263,811 | 0,117398 | 1 | 0,518958 |

| | | | | | |
|---|---|---|---|---|---|
| 1,742399 | 421 | 372,9288 | 0,082263 | 1 | 0,304997 |
| 2,325012 | 421 | 201,2914 | 0,021068 | 1 | 0,2022 |
| -0,73861 | 420 | 404,4445 | 0,460571 | 1 | 0,960096 |
| -0,49057 | 367 | 352,4651 | 0,624039 | 1 | 0,878083 |
| 1,778157 | 280 | 264,7689 | 0,076526 | 1 | 0,450485 |
| -0,86763 | 421 | 358,2867 | 0,386177 | 1 | 0,675138 |
| 0,362399 | 421 | 192,7181 | 0,717451 | 1 | 0,874811 |
| 0,40071 | 420 | 390,6935 | 0,688853 | 1 | 0,968866 |
| -0,4354 | 367 | 336,8814 | 0,663551 | 1 | 0,894765 |
| 0,424651 | 280 | 259,0931 | 0,671444 | 1 | 0,908644 |
| -0,40111 | 421 | 315,2803 | 0,688609 | 1 | 0,866222 |
| 0,499962 | 421 | 180,1274 | 0,617712 | 1 | 0,825805 |
| -0,57878 | 420 | 346,3186 | 0,563111 | 1 | 0,961052 |
| -1,70532 | 367 | 297,165 | 0,08918 | 1 | 0,396441 |
| -0,44163 | 280 | 254,444 | 0,659132 | 1 | 0,908644 |
| 0,686989 | 421 | 323,7626 | 0,492582 | 1 | 0,748579 |
| 0,83446 | 421 | 183,1326 | 0,405109 | 1 | 0,679087 |
| -1,59683 | 420 | 360,7834 | 0,111179 | 1 | 0,848246 |
| -1,29317 | 367 | 297,6946 | 0,196953 | 1 | 0,549567 |
| 0,1844 | 280 | 248,1834 | 0,85385 | 1 | 0,966655 |
| 0,823173 | 421 | 351,944 | 0,410967 | 1 | 0,695339 |
| 0,600347 | 421 | 191,598 | 0,548985 | 1 | 0,776424 |
| 0,59038 | 420 | 384,6331 | 0,555283 | 1 | 0,960096 |
| 1,412162 | 367 | 338,4243 | 0,158821 | 1 | 0,504558 |
| -0,92161 | 280 | 263,5172 | 0,357577 | 1 | 0,761846 |
| 1,096809 | 421 | 374,967 | 0,273429 | 1 | 0,56914 |
| 0,99271 | 421 | 198,8565 | 0,322057 | 1 | 0,617772 |
| -0,07819 | 420 | 402,8216 | 0,937714 | 1 | 0,992059 |
| -0,19727 | 367 | 350,0211 | 0,843731 | 1 | 0,956935 |
| 0,79881 | 280 | 266,2552 | 0,425112 | 1 | 0,797636 |
| -1,7011 | 421 | 345,2754 | 0,089825 | 1 | 0,320014 |
| -1,43601 | 421 | 190,2963 | 0,152641 | 1 | 0,433201 |
| -0,34016 | 420 | 383,1698 | 0,733925 | 1 | 0,969427 |
| -1,66262 | 367 | 334,3484 | 0,097325 | 1 | 0,412696 |
| 0,082493 | 280 | 264,0634 | 0,934318 | 1 | 0,979504 |
| 1,459889 | 421 | 359,0631 | 0,145195 | 1 | 0,41441 |
| 1,719747 | 421 | 194,1986 | 0,087072 | 1 | 0,3678 |
| 0,672058 | 420 | 392,1466 | 0,501943 | 1 | 0,960096 |
| -0,18516 | 367 | 344,0602 | 0,853211 | 1 | 0,956935 |
| 1,972555 | 280 | 263,8532 | 0,04959 | 1 | 0,39442 |
| -2,05501 | 421 | 317,6547 | 0,040695 | 1 | 0,226301 |
| -1,69273 | 421 | 181,5234 | 0,092223 | 1 | 0,373113 |
| -0,13433 | 420 | 355,4875 | 0,893219 | 1 | 0,98503 |
| -0,0938 | 367 | 317,3168 | 0,925328 | 1 | 0,976733 |
| 0,026597 | 280 | 253,043 | 0,978802 | 1 | 0,99236 |
| 1,475805 | 421 | 327,1426 | 0,140958 | 1 | 0,410878 |
| 1,414349 | 421 | 180,2123 | 0,158985 | 1 | 0,440423 |
| 1,718971 | 420 | 364,4826 | 0,086469 | 1 | 0,848246 |

| | | | | | |
|---|---|---|---|---|---|
| 1,847244 | 367 | 318,5603 | 0,065639 | 1 | 0,358398 |
| 0,717722 | 280 | 250,2226 | 0,473599 | 1 | 0,824256 |
| -1,03911 | 421 | 317,8728 | 0,299542 | 1 | 0,596172 |
| -1,20204 | 421 | 183,1454 | 0,230899 | 1 | 0,531243 |
| -1,75019 | 420 | 345,2078 | 0,080974 | 1 | 0,848246 |
| -0,8373 | 367 | 310,2765 | 0,403066 | 1 | 0,716298 |
| -1,12526 | 280 | 254,3141 | 0,261539 | 1 | 0,687569 |
| 0,029112 | 421 | 330,559 | 0,976793 | 1 | 0,995982 |
| -0,37288 | 421 | 184,9241 | 0,709666 | 1 | 0,870869 |
| -0,61277 | 420 | 366,0132 | 0,540411 | 1 | 0,960096 |
| -1,15306 | 367 | 321,3062 | 0,249741 | 1 | 0,604409 |
| -0,87638 | 280 | 258,2751 | 0,381637 | 1 | 0,770948 |
| 0,499815 | 421 | 372,7772 | 0,6175 | 1 | 0,826526 |
| 1,027822 | 421 | 199,6325 | 0,305277 | 1 | 0,602952 |
| 0,24368 | 420 | 402,5273 | 0,807603 | 1 | 0,969427 |
| 1,206984 | 367 | 349,5841 | 0,228254 | 1 | 0,577307 |
| 0,702857 | 280 | 272,1276 | 0,482746 | 1 | 0,824339 |
| -1,89023 | 421 | 326,8418 | 0,059612 | 1 | 0,268966 |
| -0,62897 | 421 | 182,6896 | 0,530155 | 1 | 0,768955 |
| 0,475617 | 420 | 364,9529 | 0,634632 | 1 | 0,964878 |
| 0,169009 | 367 | 321,0901 | 0,865896 | 1 | 0,959636 |
| 1,011451 | 280 | 267,5863 | 0,312714 | 1 | 0,733729 |
| -1,7294 | 421 | 362,6804 | 0,084589 | 1 | 0,309688 |
| -1,21549 | 421 | 193,7695 | 0,22566 | 1 | 0,523989 |
| 0,362409 | 420 | 398,8937 | 0,717238 | 1 | 0,969427 |
| -1,79205 | 367 | 349,0268 | 0,07399 | 1 | 0,374173 |
| -2,22145 | 280 | 267,6123 | 0,027158 | 1 | 0,324814 |
| -2,08064 | 421 | 334,834 | 0,038227 | 1 | 0,220722 |
| 0,334403 | 421 | 185,5297 | 0,738453 | 1 | 0,885321 |
| 1,322287 | 420 | 370,9711 | 0,186887 | 1 | 0,923408 |
| 1,459815 | 367 | 323,8962 | 0,14531 | 1 | 0,496835 |
| 1,159368 | 280 | 260,1167 | 0,247369 | 1 | 0,677438 |
| -2,16214 | 421 | 336,0445 | 0,031312 | 1 | 0,201145 |
| -1,73574 | 421 | 187,2069 | 0,084254 | 1 | 0,365475 |
| -0,88681 | 420 | 370,416 | 0,375755 | 1 | 0,95743 |
| -1,68767 | 367 | 325,2019 | 0,092434 | 1 | 0,40529 |
| -2,52055 | 280 | 261,0618 | 0,012314 | 1 | 0,244171 |
| 0,426978 | 421 | 330,7159 | 0,669673 | 1 | 0,855252 |
| 1,640834 | 421 | 185,5487 | 0,102526 | 1 | 0,383322 |
| 0,46642 | 420 | 369,5026 | 0,64119 | 1 | 0,964878 |
| 1,081379 | 367 | 320,6949 | 0,280341 | 1 | 0,614784 |
| -1,2083 | 280 | 245,9501 | 0,228094 | 1 | 0,657169 |
| -0,55154 | 421 | 370,3118 | 0,581595 | 1 | 0,805573 |
| 1,625748 | 421 | 198,6856 | 0,105588 | 1 | 0,385174 |
| 1,681223 | 420 | 400,9986 | 0,093498 | 1 | 0,848246 |
| 2,148697 | 367 | 345,0875 | 0,032354 | 1 | 0,270873 |
| -0,75466 | 280 | 261,2424 | 0,451134 | 1 | 0,814202 |
| -2,72456 | 421 | 304,8228 | 0,00681 | 1 | 0,10482 |

| | | | | | |
|---|---|---|---|---|---|
| -2,41532 | 421 | 176,2544 | 0,016743 | 1 | 0,181525 |
| -0,69606 | 420 | 344,3483 | 0,486863 | 1 | 0,960096 |
| -4,37463 | 367 | 297,9442 | 1,68E-05 | 0,018171 | 0,025378 |
| -2,2722 | 280 | 247,995 | 0,023931 | 1 | 0,313396 |
| -1,23122 | 421 | 320,6098 | 0,219142 | 1 | 0,513603 |
| -0,52434 | 421 | 179,8288 | 0,600689 | 1 | 0,816265 |
| -0,42643 | 420 | 356,3854 | 0,670054 | 1 | 0,96781 |
| 1,886745 | 367 | 325,165 | 0,060085 | 1 | 0,340409 |
| 0,419228 | 280 | 254,9317 | 0,675403 | 1 | 0,909385 |
| -0,72074 | 421 | 343,5882 | 0,471557 | 1 | 0,731548 |
| -0,17346 | 421 | 187,0588 | 0,862479 | 1 | 0,947215 |
| -0,58698 | 420 | 379,5393 | 0,557566 | 1 | 0,960096 |
| 1,197014 | 367 | 328,4904 | 0,232164 | 1 | 0,582149 |
| 0,538861 | 280 | 266,5035 | 0,590433 | 1 | 0,876633 |
| 0,376817 | 421 | 320,3405 | 0,706559 | 1 | 0,875021 |
| 0,820191 | 421 | 182,1355 | 0,413179 | 1 | 0,684933 |
| -0,12066 | 420 | 353,7744 | 0,90403 | 1 | 0,98503 |
| 0,920207 | 367 | 307,902 | 0,358185 | 1 | 0,674813 |
| 0,107771 | 280 | 249,196 | 0,914264 | 1 | 0,977051 |
| 1,010111 | 421 | 375,992 | 0,313092 | 1 | 0,608538 |
| 0,325333 | 421 | 201,6062 | 0,745267 | 1 | 0,888542 |
| -0,00865 | 420 | 404,223 | 0,993105 | 1 | 0,997342 |
| -0,12491 | 367 | 353,2795 | 0,900669 | 1 | 0,970486 |
| -0,11796 | 280 | 267,1453 | 0,906189 | 1 | 0,975308 |
| 0,947264 | 421 | 354,8071 | 0,344149 | 1 | 0,632479 |
| 1,204606 | 421 | 193,6086 | 0,229826 | 1 | 0,529584 |
| -0,61589 | 420 | 388,5275 | 0,538326 | 1 | 0,960096 |
| 0,338845 | 367 | 349,3502 | 0,73493 | 1 | 0,923669 |
| -0,27049 | 280 | 264,3323 | 0,786996 | 1 | 0,953235 |
| 0,559511 | 421 | 369,4289 | 0,576152 | 1 | 0,801719 |
| 1,525111 | 421 | 197,194 | 0,128834 | 1 | 0,410224 |
| 1,953239 | 420 | 399,262 | 0,051489 | 1 | 0,846627 |
| 2,512173 | 367 | 338,0323 | 0,012464 | 1 | 0,189546 |
| 1,044928 | 280 | 267,198 | 0,297001 | 1 | 0,723324 |
| -2,01662 | 421 | 319,6165 | 0,044571 | 1 | 0,236315 |
| -0,90559 | 421 | 182,6781 | 0,366346 | 1 | 0,647779 |
| -0,38598 | 420 | 359,6301 | 0,699741 | 1 | 0,968866 |
| 1,021763 | 367 | 318,492 | 0,307668 | 1 | 0,640947 |
| 0,851805 | 280 | 276 | 0,395061 | 1 | 0,782194 |
| 3,657168 | 421 | 342,0778 | 2,95E-04 | 0,3164 | 0,04642 |
| 2,008284 | 421 | 185,5373 | 0,046063 | 1 | 0,281076 |
| 1,867768 | 420 | 375,9797 | 0,062572 | 1 | 0,848246 |
| 2,850752 | 367 | 323,1288 | 0,004642 | 1 | 0,178183 |
| 1,148764 | 280 | 267,0478 | 0,251681 | 1 | 0,679007 |
| -0,33656 | 421 | 392,9457 | 0,736629 | 1 | 0,893273 |
| -0,32021 | 421 | 210,9018 | 0,749126 | 1 | 0,890124 |
| -0,66536 | 420 | 412,8956 | 0,506194 | 1 | 0,960096 |
| -0,55486 | 367 | 358,7001 | 0,579336 | 1 | 0,844137 |

| | | | | | |
|---|---|---|---|---|---|
| 0,726832 | 280 | 270,5849 | 0,467957 | 1 | 0,824256 |
| 1,668227 | 421 | 395,3336 | 0,096063 | 1 | 0,332744 |
| 2,076396 | 421 | 211,8173 | 0,039064 | 1 | 0,26998 |
| 0,743562 | 420 | 414,0492 | 0,457563 | 1 | 0,960096 |
| -0,05836 | 367 | 361,8688 | 0,953491 | 1 | 0,980157 |
| 0,759448 | 280 | 274,33 | 0,448237 | 1 | 0,813228 |
| 1,764429 | 421 | 380,1779 | 0,078463 | 1 | 0,302838 |
| 2,425267 | 421 | 204,4994 | 0,016165 | 1 | 0,179127 |
| 0,506182 | 420 | 408,7094 | 0,613002 | 1 | 0,964878 |
| 2,531366 | 367 | 356,0689 | 0,011792 | 1 | 0,189546 |
| 1,757324 | 280 | 272,7278 | 0,079985 | 1 | 0,453386 |
| 0,252194 | 421 | 402,2566 | 0,80102 | 1 | 0,922616 |
| 1,772041 | 421 | 217,5131 | 0,077788 | 1 | 0,353243 |
| 0,002956 | 420 | 415,6822 | 0,997643 | 1 | 0,997643 |
| 0,207812 | 367 | 357,5851 | 0,835494 | 1 | 0,956935 |
| -0,88013 | 280 | 275,0072 | 0,379557 | 1 | 0,770948 |
| -0,47275 | 421 | 352,1126 | 0,636688 | 1 | 0,834842 |
| 0,119596 | 421 | 190,4803 | 0,904929 | 1 | 0,96513 |
| -0,32474 | 420 | 384,8961 | 0,745551 | 1 | 0,969427 |
| 1,176174 | 367 | 329,6029 | 0,240374 | 1 | 0,59287 |
| 1,651686 | 280 | 261,0102 | 0,099801 | 1 | 0,480557 |
| -0,14565 | 421 | 346,3988 | 0,884285 | 1 | 0,961484 |
| 1,127619 | 421 | 188,2942 | 0,260916 | 1 | 0,557979 |
| -0,38415 | 420 | 381,1751 | 0,701081 | 1 | 0,968866 |
| 0,776005 | 367 | 317,5612 | 0,438324 | 1 | 0,742744 |
| -0,05173 | 280 | 252,5041 | 0,958784 | 1 | 0,986271 |
| -0,2471 | 421 | 363,0577 | 0,80497 | 1 | 0,922616 |
| 1,762039 | 421 | 192,901 | 0,079647 | 1 | 0,357225 |
| 0,623758 | 420 | 392,7457 | 0,533149 | 1 | 0,960096 |
| 0,776301 | 367 | 349,8565 | 0,438096 | 1 | 0,742744 |
| -1,35694 | 280 | 265,5846 | 0,175951 | 1 | 0,598625 |
| -0,06023 | 421 | 376,3953 | 0,952002 | 1 | 0,985348 |
| -0,6173 | 421 | 198,7701 | 0,537743 | 1 | 0,771054 |
| 1,901504 | 420 | 402,363 | 0,05795 | 1 | 0,848246 |
| 0,405795 | 367 | 350,204 | 0,685141 | 1 | 0,902395 |
| 0,798518 | 280 | 270,8043 | 0,42527 | 1 | 0,797636 |
| 3,567894 | 421 | 350,945 | 4,10E-04 | 0,437735 | 0,04642 |
| 1,81333 | 421 | 189,054 | 0,071367 | 1 | 0,339864 |
| 1,616497 | 420 | 383,6865 | 0,106809 | 1 | 0,848246 |
| 2,915876 | 367 | 332,2422 | 0,003788 | 1 | 0,178183 |
| 1,202108 | 280 | 269,4946 | 0,230377 | 1 | 0,657169 |
| -0,59051 | 421 | 346,4633 | 0,555235 | 1 | 0,790572 |
| -1,00497 | 421 | 190,819 | 0,316183 | 1 | 0,614646 |
| 0,253477 | 420 | 382,09 | 0,800036 | 1 | 0,969427 |
| -0,33281 | 367 | 330,7953 | 0,739486 | 1 | 0,923669 |
| -1,14397 | 280 | 256,3758 | 0,253704 | 1 | 0,679673 |
| 0,563463 | 421 | 365,919 | 0,573465 | 1 | 0,800196 |
| 0,997539 | 421 | 195,8241 | 0,319734 | 1 | 0,617081 |

| | | | | | |
|---|---|---|---|---|---|
| 0,690747 | 420 | 395,9741 | 0,490129 | 1 | 0,960096 |
| 0,358802 | 367 | 351,0641 | 0,719959 | 1 | 0,914797 |
| -0,47064 | 280 | 265,2804 | 0,638284 | 1 | 0,907449 |
| -2,69837 | 421 | 357,1825 | 0,007299 | 1 | 0,105763 |
| -2,38393 | 421 | 193,9674 | 0,018093 | 1 | 0,186759 |
| -2,27126 | 420 | 392,0644 | 0,023673 | 1 | 0,75904 |
| -1,21832 | 367 | 336,2233 | 0,223956 | 1 | 0,576775 |
| -1,74596 | 280 | 270,1559 | 0,081955 | 1 | 0,456738 |
| 1,878767 | 421 | 385,0737 | 0,061031 | 1 | 0,270611 |
| 0,742443 | 421 | 204,6428 | 0,45867 | 1 | 0,719268 |
| 1,498399 | 420 | 408,1737 | 0,134803 | 1 | 0,890998 |
| 0,161269 | 367 | 354,599 | 0,871974 | 1 | 0,959978 |
| 0,038733 | 280 | 266,1884 | 0,969132 | 1 | 0,991502 |
| 1,203944 | 421 | 371,5746 | 0,229377 | 1 | 0,520357 |
| 0,476276 | 421 | 198,4753 | 0,634402 | 1 | 0,835703 |
| -0,02044 | 420 | 399,8789 | 0,983706 | 1 | 0,997342 |
| -0,72079 | 367 | 342,2166 | 0,471531 | 1 | 0,77576 |
| 1,091054 | 280 | 255,9953 | 0,276275 | 1 | 0,701156 |
| -1,51849 | 421 | 349,2605 | 0,129795 | 1 | 0,397564 |
| -1,23455 | 421 | 189,3868 | 0,218528 | 1 | 0,514565 |
| 0,285101 | 420 | 382,3767 | 0,775721 | 1 | 0,969427 |
| -1,28286 | 367 | 336,2429 | 0,200424 | 1 | 0,553185 |
| -1,72477 | 280 | 250,3734 | 0,085804 | 1 | 0,458532 |
| 0,389944 | 421 | 331,7016 | 0,696828 | 1 | 0,871469 |
| -2,20046 | 421 | 182,9167 | 0,029026 | 1 | 0,233259 |
| -2,12074 | 420 | 357,3108 | 0,034632 | 1 | 0,828713 |
| -0,27338 | 367 | 309,8053 | 0,784741 | 1 | 0,94337 |
| -0,31477 | 280 | 254,2294 | 0,753193 | 1 | 0,942071 |
| -0,93616 | 421 | 299,6033 | 0,349947 | 1 | 0,639237 |
| -0,1235 | 421 | 176,145 | 0,901854 | 1 | 0,962531 |
| 0,876245 | 420 | 331,2963 | 0,381532 | 1 | 0,95743 |
| -1,19759 | 367 | 303,0825 | 0,232013 | 1 | 0,582149 |
| -1,02432 | 280 | 249,4785 | 0,306675 | 1 | 0,72861 |
| -0,73028 | 421 | 386,2254 | 0,465665 | 1 | 0,727964 |
| -1,39447 | 421 | 206,8313 | 0,164671 | 1 | 0,451197 |
| 0,127878 | 420 | 409,5715 | 0,898308 | 1 | 0,98503 |
| -0,08411 | 367 | 359,1235 | 0,933019 | 1 | 0,977109 |
| -1,20132 | 280 | 268,4023 | 0,230685 | 1 | 0,657169 |
| 0,59851 | 421 | 315,4176 | 0,549929 | 1 | 0,786367 |
| 1,127934 | 421 | 179,7646 | 0,260851 | 1 | 0,557979 |
| -0,46715 | 420 | 349,5044 | 0,64068 | 1 | 0,964878 |
| 0,102121 | 367 | 312,8228 | 0,918726 | 1 | 0,974328 |
| 1,948176 | 280 | 255,1928 | 0,05249 | 1 | 0,401537 |
| -1,8905 | 421 | 370,7732 | 0,059471 | 1 | 0,268966 |
| -0,9501 | 421 | 199,0273 | 0,343214 | 1 | 0,636552 |
| -1,96179 | 420 | 400,6439 | 0,050479 | 1 | 0,846627 |
| 0,660322 | 367 | 347,5813 | 0,509485 | 1 | 0,797092 |
| -0,82889 | 280 | 265,0519 | 0,407912 | 1 | 0,788151 |

| | | | | | |
|---|---|---|---|---|---|
| -3,63462 | 421 | 333,9909 | 3,22E-04 | 0,345098 | 0,04642 |
| -1,39179 | 421 | 187,3385 | 0,165635 | 1 | 0,453017 |
| -1,35161 | 420 | 373,688 | 0,177317 | 1 | 0,9199 |
| -1,24398 | 367 | 320,9077 | 0,214415 | 1 | 0,564902 |
| 0,704891 | 280 | 261,2946 | 0,481506 | 1 | 0,824339 |
| -0,03111 | 421 | 362,2349 | 0,975201 | 1 | 0,995982 |
| -0,71311 | 421 | 196,2191 | 0,476624 | 1 | 0,734183 |
| 0,184106 | 420 | 395,1493 | 0,854025 | 1 | 0,973894 |
| -0,4772 | 367 | 346,4075 | 0,633518 | 1 | 0,880707 |
| 1,393845 | 280 | 268,5456 | 0,164517 | 1 | 0,586115 |
| -1,27155 | 421 | 312,4841 | 0,204478 | 1 | 0,494095 |
| 0,558709 | 421 | 179,1029 | 0,577058 | 1 | 0,794719 |
| -0,68402 | 420 | 345,7489 | 0,494423 | 1 | 0,960096 |
| -0,11284 | 367 | 288,8904 | 0,910234 | 1 | 0,970788 |
| 0,118162 | 280 | 245,7461 | 0,906035 | 1 | 0,975308 |
| 0,951485 | 421 | 374,5093 | 0,341972 | 1 | 0,63164 |
| 1,442892 | 421 | 203,187 | 0,150591 | 1 | 0,430627 |
| -0,67806 | 420 | 406,6629 | 0,498118 | 1 | 0,960096 |
| -0,13246 | 367 | 355,1276 | 0,894696 | 1 | 0,969307 |
| -0,8738 | 280 | 269,7214 | 0,383002 | 1 | 0,772669 |
| -0,49778 | 421 | 361,0213 | 0,618944 | 1 | 0,826904 |
| 0,964737 | 421 | 194,9093 | 0,335872 | 1 | 0,630335 |
| 0,615431 | 420 | 396,0853 | 0,538624 | 1 | 0,960096 |
| 0,481111 | 367 | 331,1413 | 0,630755 | 1 | 0,880707 |
| 0,960851 | 280 | 264,5244 | 0,337505 | 1 | 0,755731 |
| 1,214711 | 421 | 374,1808 | 0,225242 | 1 | 0,518147 |
| -0,21085 | 421 | 198,7573 | 0,83322 | 1 | 0,936588 |
| 0,636842 | 420 | 401,4665 | 0,52459 | 1 | 0,960096 |
| 0,231664 | 367 | 352,9372 | 0,816933 | 1 | 0,95095 |
| -1,61017 | 280 | 268,6269 | 0,108535 | 1 | 0,497151 |
| 0,796063 | 421 | 336,4126 | 0,426557 | 1 | 0,700145 |
| 0,995547 | 421 | 184,295 | 0,320776 | 1 | 0,617772 |
| -0,53149 | 420 | 369,9564 | 0,595395 | 1 | 0,964878 |
| 0,677359 | 367 | 321,8288 | 0,498665 | 1 | 0,793601 |
| 1,743996 | 280 | 249,5827 | 0,082391 | 1 | 0,456738 |
| -0,47443 | 421 | 349,9743 | 0,635493 | 1 | 0,83455 |
| 0,102266 | 421 | 191,304 | 0,918653 | 1 | 0,970154 |
| -0,43151 | 420 | 383,8574 | 0,666337 | 1 | 0,96781 |
| -0,07119 | 367 | 341,4783 | 0,943289 | 1 | 0,979539 |
| -1,40518 | 280 | 261,1267 | 0,161156 | 1 | 0,581516 |
| 0,231725 | 421 | 353,7755 | 0,816886 | 1 | 0,929092 |
| -0,47782 | 421 | 190,685 | 0,633327 | 1 | 0,835702 |
| 0,123828 | 420 | 385,5682 | 0,901516 | 1 | 0,98503 |
| -0,50661 | 367 | 338,7776 | 0,612759 | 1 | 0,871158 |
| 1,179952 | 280 | 256,8916 | 0,239111 | 1 | 0,662939 |
| 1,434203 | 421 | 372,2729 | 0,152354 | 1 | 0,42445 |
| 1,480541 | 421 | 200,0103 | 0,140302 | 1 | 0,423351 |
| 0,367566 | 420 | 402,4326 | 0,71339 | 1 | 0,968866 |

| | | | | | |
|---|---|---|---|---|---|
| 0,652104 | 367 | 351,436 | 0,51476 | 1 | 0,798912 |
| -1,71142 | 280 | 265,824 | 0,08817 | 1 | 0,461362 |
| 0,440656 | 421 | 336,8893 | 0,659744 | 1 | 0,849048 |
| -0,5663 | 421 | 185,2886 | 0,571875 | 1 | 0,792729 |
| -0,089 | 420 | 369,7122 | 0,929131 | 1 | 0,990942 |
| 0,013462 | 367 | 319,7039 | 0,989267 | 1 | 0,996248 |
| 2,285796 | 280 | 257,069 | 0,02308 | 1 | 0,313396 |
| -1,30979 | 421 | 360,6209 | 0,191099 | 1 | 0,47601 |
| -0,91059 | 421 | 194,9707 | 0,363636 | 1 | 0,646044 |
| -1,00356 | 420 | 393,4867 | 0,31621 | 1 | 0,943897 |
| -0,77376 | 367 | 342,4088 | 0,439607 | 1 | 0,742744 |
| -1,53382 | 280 | 270,4866 | 0,126242 | 1 | 0,536498 |
| -1,17336 | 421 | 335,7163 | 0,241484 | 1 | 0,538768 |
| -1,44355 | 421 | 186,3218 | 0,150544 | 1 | 0,430627 |
| -0,88498 | 420 | 370,4524 | 0,376739 | 1 | 0,95743 |
| -0,58171 | 367 | 322,9591 | 0,561165 | 1 | 0,834 |
| -2,77604 | 280 | 251,9303 | 0,005915 | 1 | 0,186692 |
| -1,79452 | 421 | 344,4921 | 0,073607 | 1 | 0,292681 |
| -0,72197 | 421 | 187,2652 | 0,471213 | 1 | 0,731044 |
| 0,702781 | 420 | 378,8656 | 0,482624 | 1 | 0,960096 |
| -0,16249 | 367 | 335,3115 | 0,871017 | 1 | 0,959978 |
| -0,71964 | 280 | 266,472 | 0,472375 | 1 | 0,824256 |
| -0,00613 | 421 | 336,533 | 0,99511 | 1 | 0,998054 |
| 1,006267 | 421 | 184,0047 | 0,315609 | 1 | 0,614499 |
| -0,09353 | 420 | 371,7411 | 0,925533 | 1 | 0,989204 |
| -0,21024 | 367 | 316,8825 | 0,833619 | 1 | 0,956935 |
| -0,11076 | 280 | 258,4969 | 0,911891 | 1 | 0,976008 |
| -1,74851 | 421 | 313,9047 | 0,081353 | 1 | 0,30376 |
| -1,14994 | 421 | 180,4036 | 0,251691 | 1 | 0,549707 |
| -2,89026 | 420 | 340,3417 | 0,004096 | 1 | 0,443819 |
| -1,05181 | 367 | 296,7096 | 0,293743 | 1 | 0,627013 |
| -1,92376 | 280 | 276 | 0,055413 | 1 | 0,40654 |
| 0,08293 | 421 | 327,6037 | 0,933958 | 1 | 0,972684 |
| 0,201015 | 421 | 186,0393 | 0,840907 | 1 | 0,940271 |
| 0,21613 | 420 | 361,6084 | 0,829009 | 1 | 0,973894 |
| 0,588767 | 367 | 312,5094 | 0,556443 | 1 | 0,830257 |
| 2,236184 | 280 | 263,5309 | 0,026177 | 1 | 0,324814 |
| 0,66823 | 421 | 393,5269 | 0,504378 | 1 | 0,75707 |
| -0,0448 | 421 | 211,1424 | 0,964308 | 1 | 0,985228 |
| 0,171788 | 420 | 412,5987 | 0,863688 | 1 | 0,977895 |
| -0,20814 | 367 | 358,2115 | 0,835237 | 1 | 0,956935 |
| -0,12406 | 280 | 267,1709 | 0,901358 | 1 | 0,975243 |
| -0,02853 | 421 | 322,2548 | 0,97726 | 1 | 0,995982 |
| 0,779664 | 421 | 183,064 | 0,436595 | 1 | 0,703689 |
| 2,819839 | 420 | 353,1527 | 0,005076 | 1 | 0,449965 |
| 2,238706 | 367 | 310,4892 | 0,025883 | 1 | 0,252225 |
| 1,922397 | 280 | 250,205 | 0,055691 | 1 | 0,40654 |
| -0,55997 | 421 | 381,5023 | 0,575828 | 1 | 0,801719 |

| | | | | | |
|---|---|---|---|---|---|
| -1,42764 | 421 | 203,2732 | 0,15493 | 1 | 0,437229 |
| 1,42213 | 420 | 407,8369 | 0,155753 | 1 | 0,914005 |
| -1,11663 | 367 | 354,1847 | 0,264911 | 1 | 0,609149 |
| -0,0247 | 280 | 267,3132 | 0,980314 | 1 | 0,99236 |
| 1,123239 | 421 | 340,6675 | 0,262127 | 1 | 0,555878 |
| 0,760962 | 421 | 187,7712 | 0,447634 | 1 | 0,710837 |
| 0,409374 | 420 | 375,0993 | 0,682499 | 1 | 0,968338 |
| -0,64384 | 367 | 331,287 | 0,520122 | 1 | 0,802988 |
| -0,84851 | 280 | 259,474 | 0,396935 | 1 | 0,782458 |
| -0,28135 | 421 | 292,8697 | 0,77864 | 1 | 0,913934 |
| -0,50637 | 421 | 176,4939 | 0,61323 | 1 | 0,824026 |
| -0,95311 | 420 | 321,7744 | 0,341248 | 1 | 0,947373 |
| 0,336688 | 367 | 281,9933 | 0,736602 | 1 | 0,923669 |
| -1,21512 | 280 | 276 | 0,225358 | 1 | 0,654014 |
| -2,45368 | 421 | 305,9826 | 0,014698 | 1 | 0,147663 |
| -0,95929 | 421 | 177,0555 | 0,338718 | 1 | 0,632922 |
| -1,24964 | 420 | 339,1497 | 0,212292 | 1 | 0,943055 |
| -0,26533 | 367 | 303,6077 | 0,790937 | 1 | 0,944997 |
| -1,38024 | 280 | 253,744 | 0,168727 | 1 | 0,59271 |
| -1,93487 | 421 | 341,0001 | 0,053833 | 1 | 0,259162 |
| -1,85084 | 421 | 186,7895 | 0,065771 | 1 | 0,328203 |
| -0,4957 | 420 | 378,2412 | 0,620394 | 1 | 0,964878 |
| -0,0014 | 367 | 331,8519 | 0,998887 | 1 | 0,998887 |
| -1,50413 | 280 | 263,159 | 0,133746 | 1 | 0,543276 |
| -1,32782 | 421 | 345,8028 | 0,185113 | 1 | 0,46885 |
| -0,34003 | 421 | 187,19 | 0,734218 | 1 | 0,884613 |
| -2,53426 | 420 | 372,6023 | 0,011677 | 1 | 0,651769 |
| -2,38566 | 367 | 313,5884 | 0,017642 | 1 | 0,216225 |
| -0,55859 | 280 | 255,0801 | 0,576935 | 1 | 0,871183 |
| -0,78088 | 421 | 335,8358 | 0,435425 | 1 | 0,706341 |
| -0,65542 | 421 | 187,6372 | 0,512997 | 1 | 0,756803 |
| 0,543079 | 420 | 369,5434 | 0,587402 | 1 | 0,964878 |
| -0,1212 | 367 | 327,1632 | 0,90361 | 1 | 0,970486 |
| -0,0368 | 280 | 259,6716 | 0,970674 | 1 | 0,992124 |
| -1,53033 | 421 | 357,8201 | 0,126817 | 1 | 0,392431 |
| -1,63597 | 421 | 193,348 | 0,103472 | 1 | 0,383322 |
| -0,61264 | 420 | 390,5886 | 0,54047 | 1 | 0,960096 |
| -2,43047 | 367 | 331,6669 | 0,015609 | 1 | 0,204372 |
| -0,30283 | 280 | 260,9803 | 0,762263 | 1 | 0,944296 |
| -3,09276 | 421 | 341,8152 | 0,002146 | 1 | 0,069718 |
| -2,08322 | 421 | 189,3348 | 0,038574 | 1 | 0,269128 |
| -0,867 | 420 | 381,9326 | 0,386485 | 1 | 0,95743 |
| -1,80206 | 367 | 337,5367 | 0,072428 | 1 | 0,369998 |
| -2,26421 | 280 | 270,6078 | 0,024353 | 1 | 0,313396 |
| -1,74267 | 421 | 313,4994 | 0,082371 | 1 | 0,304997 |
| -1,01 | 421 | 180,0432 | 0,31385 | 1 | 0,613453 |
| 0,576759 | 420 | 348,009 | 0,564475 | 1 | 0,961448 |
| -0,56881 | 367 | 294,4921 | 0,569916 | 1 | 0,837915 |

| | | | | | |
|---|---|---|---|---|---|
| -0,04293 | 280 | 253,3187 | 0,965788 | 1 | 0,990771 |
| -0,5376 | 421 | 328,9598 | 0,591215 | 1 | 0,81147 |
| -0,7021 | 421 | 180,8114 | 0,48352 | 1 | 0,739236 |
| -2,01367 | 420 | 356,4401 | 0,044795 | 1 | 0,846132 |
| -0,12067 | 367 | 313,2687 | 0,904029 | 1 | 0,970486 |
| 2,16336 | 280 | 257,4719 | 0,031434 | 1 | 0,329568 |
| -0,89503 | 421 | 320,3257 | 0,371446 | 1 | 0,663233 |
| 0,714381 | 421 | 180,8945 | 0,475912 | 1 | 0,734136 |
| 0,233944 | 420 | 355,2492 | 0,815163 | 1 | 0,969427 |
| 0,04724 | 367 | 296,1207 | 0,962354 | 1 | 0,983232 |
| 0,071149 | 280 | 250,5472 | 0,943336 | 1 | 0,982175 |
| -0,39281 | 421 | 344,6844 | 0,694702 | 1 | 0,870609 |
| -0,6752 | 421 | 188,5175 | 0,500378 | 1 | 0,750551 |
| 1,6263 | 420 | 380,6367 | 0,104714 | 1 | 0,848246 |
| 0,151189 | 367 | 334,4954 | 0,879918 | 1 | 0,962453 |
| 1,063565 | 280 | 268,8416 | 0,28848 | 1 | 0,71621 |
| -0,10334 | 421 | 326,0262 | 0,917755 | 1 | 0,966828 |
| -1,85579 | 421 | 184,5104 | 0,065079 | 1 | 0,325826 |
| -1,19088 | 420 | 360,9288 | 0,234485 | 1 | 0,943055 |
| -1,06167 | 367 | 318,6758 | 0,289191 | 1 | 0,626164 |
| -1,25959 | 280 | 251,9792 | 0,208982 | 1 | 0,64091 |
| -0,59407 | 421 | 394,2501 | 0,552807 | 1 | 0,78965 |
| -0,66243 | 421 | 210,9649 | 0,508421 | 1 | 0,754914 |
| -1,38926 | 420 | 413,813 | 0,1655 | 1 | 0,914005 |
| -0,27063 | 367 | 360,5593 | 0,786827 | 1 | 0,943768 |
| -1,41695 | 280 | 274,8464 | 0,157631 | 1 | 0,576579 |
| -1,90423 | 421 | 328,638 | 0,057753 | 1 | 0,266382 |
| -0,95353 | 421 | 182,8482 | 0,341579 | 1 | 0,635891 |
| -1,02514 | 420 | 364,6701 | 0,305979 | 1 | 0,943897 |
| 1,180437 | 367 | 317,5824 | 0,23871 | 1 | 0,589731 |
| -0,30166 | 280 | 256,5904 | 0,763153 | 1 | 0,944296 |
| 1,931031 | 421 | 307,7807 | 0,054397 | 1 | 0,259162 |
| 2,965019 | 421 | 178,0393 | 0,003442 | 1 | 0,118336 |
| 1,164958 | 420 | 342,417 | 0,244847 | 1 | 0,943897 |
| 0,657667 | 367 | 283,8996 | 0,511285 | 1 | 0,797829 |
| 2,823341 | 280 | 245,4197 | 0,005143 | 1 | 0,180228 |
| -0,61636 | 421 | 333,0931 | 0,538075 | 1 | 0,778195 |
| -0,02853 | 421 | 184,9489 | 0,977269 | 1 | 0,991789 |
| -1,80813 | 420 | 368,8824 | 0,071401 | 1 | 0,848246 |
| 1,071239 | 367 | 324,7762 | 0,284857 | 1 | 0,619451 |
| -0,18899 | 280 | 255,8609 | 0,850248 | 1 | 0,966155 |
| 0,123888 | 421 | 405,0566 | 0,901465 | 1 | 0,965517 |
| 1,607667 | 421 | 217,789 | 0,109357 | 1 | 0,3896 |
| -0,24312 | 420 | 415,9567 | 0,808029 | 1 | 0,969427 |
| 1,668917 | 367 | 362,619 | 0,095997 | 1 | 0,412696 |
| 1,313976 | 280 | 274,2432 | 0,189952 | 1 | 0,625017 |
| -0,57155 | 421 | 394,8918 | 0,567954 | 1 | 0,796637 |
| 0,743 | 421 | 211,3887 | 0,458307 | 1 | 0,719268 |

| | | | | | |
|---|---|---|---|---|---|
| -0,75178 | 420 | 412,8043 | 0,452611 | 1 | 0,960096 |
| -0,98917 | 367 | 360,0261 | 0,323243 | 1 | 0,655597 |
| 0,208555 | 280 | 270,3955 | 0,834953 | 1 | 0,964667 |
| -0,71741 | 421 | 345,1912 | 0,473607 | 1 | 0,732026 |
| 0,433604 | 421 | 185,7417 | 0,665079 | 1 | 0,846515 |
| 0,266868 | 420 | 381,6678 | 0,789715 | 1 | 0,969427 |
| 0,248683 | 367 | 320,0521 | 0,803766 | 1 | 0,950149 |
| 1,465763 | 280 | 264,411 | 0,143901 | 1 | 0,555466 |
| 0,505107 | 421 | 336,8254 | 0,613814 | 1 | 0,825172 |
| -0,02909 | 421 | 186,5674 | 0,976821 | 1 | 0,991789 |
| -0,72233 | 420 | 370,2538 | 0,470548 | 1 | 0,960096 |
| -0,23273 | 367 | 333,4949 | 0,816112 | 1 | 0,95095 |
| 0,490627 | 280 | 262,3369 | 0,624101 | 1 | 0,898299 |
| 1,086233 | 421 | 357,8508 | 0,278107 | 1 | 0,57412 |
| 1,110541 | 421 | 191,4802 | 0,268159 | 1 | 0,566781 |
| 0,196228 | 420 | 392,895 | 0,844533 | 1 | 0,973894 |
| 0,18392 | 367 | 344,4588 | 0,854184 | 1 | 0,957067 |
| 0,544366 | 280 | 266,7237 | 0,586645 | 1 | 0,87561 |
| 1,088896 | 421 | 305,1849 | 0,277059 | 1 | 0,57291 |
| 1,773766 | 421 | 176,987 | 0,077821 | 1 | 0,353243 |
| 0,372279 | 420 | 337,5377 | 0,709919 | 1 | 0,968866 |
| 1,112019 | 367 | 296,9256 | 0,267029 | 1 | 0,609149 |
| 0,871212 | 280 | 251,9845 | 0,384468 | 1 | 0,773166 |
| 3,136308 | 421 | 351,5939 | 0,001855 | 0,183661 | 0,069718 |
| 0,924748 | 421 | 188,3698 | 0,35628 | 1 | 0,643758 |
| 0,651116 | 420 | 383,8379 | 0,515361 | 1 | 0,960096 |
| 1,776127 | 367 | 324,8138 | 0,076648 | 1 | 0,378716 |
| -0,58656 | 280 | 260,8385 | 0,558008 | 1 | 0,858676 |
| -0,41175 | 421 | 412,71 | 0,680733 | 1 | 0,861268 |
| -0,58687 | 421 | 232,8864 | 0,557857 | 1 | 0,779138 |
| 0,393849 | 420 | 414,3397 | 0,693895 | 1 | 0,968866 |
| -1,4223 | 367 | 362,3139 | 0,155801 | 1 | 0,502197 |
| -0,17338 | 280 | 275,9886 | 0,862479 | 1 | 0,967527 |
| 0,394076 | 421 | 401,0993 | 0,693734 | 1 | 0,870489 |
| -0,20409 | 421 | 216,9477 | 0,838473 | 1 | 0,938766 |
| 0,90849 | 420 | 415,3158 | 0,364146 | 1 | 0,956033 |
| -1,58461 | 367 | 360,6405 | 0,113931 | 1 | 0,440054 |
| 0,289161 | 280 | 266,5999 | 0,772683 | 1 | 0,947219 |
| -1,01051 | 421 | 365,936 | 0,312918 | 1 | 0,608538 |
| -1,81748 | 421 | 196,9419 | 0,070663 | 1 | 0,339864 |
| -1,27866 | 420 | 394,5399 | 0,201768 | 1 | 0,928589 |
| 0,258601 | 367 | 343,2328 | 0,796098 | 1 | 0,946151 |
| -0,65991 | 280 | 265,8256 | 0,509882 | 1 | 0,831912 |
| -0,46629 | 421 | 408,5203 | 0,641258 | 1 | 0,835242 |
| 1,610641 | 421 | 222,2229 | 0,108678 | 1 | 0,388185 |
| 0,818929 | 420 | 415,9696 | 0,413296 | 1 | 0,95743 |
| 1,464593 | 367 | 362,7114 | 0,143898 | 1 | 0,495102 |
| 1,011114 | 280 | 275,8561 | 0,312848 | 1 | 0,733729 |

| | | | | | |
|---|---|---|---|---|---|
| 0,431028 | 421 | 410,8255 | 0,666674 | 1 | 0,853323 |
| -0,17318 | 421 | 228,0461 | 0,862663 | 1 | 0,947215 |
| 0,197335 | 420 | 415,4462 | 0,843662 | 1 | 0,973894 |
| -0,01084 | 367 | 359,9192 | 0,991357 | 1 | 0,997007 |
| 1,404497 | 280 | 275,6635 | 0,161297 | 1 | 0,581516 |
| -0,20964 | 421 | 368,3881 | 0,834062 | 1 | 0,936611 |
| 0,238691 | 421 | 197,508 | 0,811593 | 1 | 0,926063 |
| 1,433812 | 420 | 395,0291 | 0,152417 | 1 | 0,914005 |
| -0,79277 | 367 | 350,0884 | 0,428451 | 1 | 0,737072 |
| -1,67474 | 280 | 267,0513 | 0,095156 | 1 | 0,476412 |
| 0,136873 | 421 | 347,187 | 0,89121 | 1 | 0,962 |
| -2,26932 | 421 | 190,2324 | 0,024371 | 1 | 0,218351 |
| -1,59327 | 420 | 383,867 | 0,111923 | 1 | 0,848246 |
| -1,12205 | 367 | 327,5109 | 0,262663 | 1 | 0,609149 |
| -1,93866 | 280 | 261,2396 | 0,05362 | 1 | 0,406056 |
| 1,301413 | 421 | 356,4785 | 0,193958 | 1 | 0,479958 |
| 1,185057 | 421 | 191,6386 | 0,237462 | 1 | 0,537903 |
| 1,425091 | 420 | 391,1562 | 0,154928 | 1 | 0,914005 |
| 2,098353 | 367 | 339,6705 | 0,036612 | 1 | 0,28464 |
| 0,663467 | 280 | 267,9418 | 0,507602 | 1 | 0,831912 |
| -1,94894 | 421 | 340,4102 | 0,052124 | 1 | 0,254208 |
| -1,74886 | 421 | 189,6744 | 0,081933 | 1 | 0,362173 |
| -0,14746 | 420 | 383,3842 | 0,88285 | 1 | 0,982463 |
| 0,418624 | 367 | 326,8245 | 0,675766 | 1 | 0,898834 |
| -1,26085 | 280 | 260,2209 | 0,208493 | 1 | 0,64091 |
| -2,62305 | 421 | 375,5129 | 0,00907 | 1 | 0,113397 |
| -0,04254 | 421 | 198,2413 | 0,966107 | 1 | 0,986398 |
| -0,48474 | 420 | 400,2456 | 0,628127 | 1 | 0,964878 |
| 1,896335 | 367 | 351,7866 | 0,058735 | 1 | 0,339132 |
| -0,96537 | 280 | 265,4288 | 0,335238 | 1 | 0,755731 |
| -0,56604 | 421 | 317,3923 | 0,571764 | 1 | 0,79928 |
| 0,264547 | 421 | 177,7291 | 0,791665 | 1 | 0,918572 |
| -0,71088 | 420 | 351,0529 | 0,477632 | 1 | 0,960096 |
| -0,47982 | 367 | 300,8707 | 0,631703 | 1 | 0,880707 |
| 1,476215 | 280 | 245,5421 | 0,141168 | 1 | 0,554427 |
| 1,226187 | 421 | 361,8154 | 0,220926 | 1 | 0,516179 |
| 1,112883 | 421 | 192,2564 | 0,267148 | 1 | 0,565438 |
| -0,10376 | 420 | 395,3774 | 0,917409 | 1 | 0,989204 |
| 2,737749 | 367 | 349,1204 | 0,006503 | 1 | 0,178183 |
| 1,193239 | 280 | 272,4927 | 0,233814 | 1 | 0,659852 |
| -1,14946 | 421 | 370,4547 | 0,251108 | 1 | 0,546261 |
| -1,41636 | 421 | 200,1758 | 0,158226 | 1 | 0,439127 |
| -1,41104 | 420 | 400,5014 | 0,159008 | 1 | 0,914005 |
| -1,04147 | 367 | 346,8529 | 0,298382 | 1 | 0,630662 |
| -0,97597 | 280 | 263,8663 | 0,329973 | 1 | 0,752282 |
| 0,41107 | 421 | 398,4215 | 0,681242 | 1 | 0,861268 |
| 0,960274 | 421 | 213,8849 | 0,338001 | 1 | 0,632756 |
| -0,02063 | 420 | 414,5592 | 0,983555 | 1 | 0,997342 |

| | | | | | |
|---|---|---|---|---|---|
| -1,4119 | 367 | 361,4611 | 0,158839 | 1 | 0,504558 |
| 0,820705 | 280 | 270,8814 | 0,412536 | 1 | 0,788151 |
| -0,00873 | 421 | 348,2107 | 0,99304 | 1 | 0,998054 |
| -0,23454 | 421 | 190,4812 | 0,814818 | 1 | 0,926063 |
| 0,880398 | 420 | 381,0458 | 0,379199 | 1 | 0,95743 |
| -0,15259 | 367 | 333,274 | 0,878814 | 1 | 0,962453 |
| 0,363824 | 280 | 257,6988 | 0,716288 | 1 | 0,930557 |
| 3,500663 | 421 | 352,6467 | 5,24E-04 | 0,068598 | 0,04642 |
| 1,946898 | 421 | 191,0247 | 0,053013 | 1 | 0,302617 |
| 0,370758 | 420 | 388,2622 | 0,71102 | 1 | 0,968866 |
| 2,352714 | 367 | 342,0903 | 0,019203 | 1 | 0,218203 |
| 1,540145 | 280 | 270,6019 | 0,124694 | 1 | 0,535729 |
| -0,20487 | 421 | 321,9835 | 0,837806 | 1 | 0,93721 |
| 0,608072 | 421 | 181,3233 | 0,5439 | 1 | 0,774002 |
| -1,05797 | 420 | 357,7683 | 0,290784 | 1 | 0,943897 |
| -0,63061 | 367 | 313,7425 | 0,528755 | 1 | 0,812268 |
| 0,757959 | 280 | 262,2655 | 0,449156 | 1 | 0,813228 |
| 0,261165 | 421 | 386,3657 | 0,794104 | 1 | 0,919136 |
| -1,13433 | 421 | 207,1278 | 0,257968 | 1 | 0,556025 |
| -0,41976 | 420 | 410,0164 | 0,67488 | 1 | 0,96781 |
| 0,936837 | 367 | 358,5345 | 0,349473 | 1 | 0,668671 |
| -0,35187 | 280 | 274,6652 | 0,725207 | 1 | 0,936493 |
| 0,022648 | 421 | 330,2203 | 0,981945 | 1 | 0,997025 |
| 0,065079 | 421 | 184,6457 | 0,948182 | 1 | 0,980846 |
| -0,85223 | 420 | 364,0253 | 0,394649 | 1 | 0,95743 |
| 0,289442 | 367 | 325,2496 | 0,772427 | 1 | 0,938452 |
| 0,533141 | 280 | 261,2364 | 0,594389 | 1 | 0,879042 |
| -1,90517 | 421 | 392,1294 | 0,057489 | 1 | 0,266382 |
| -1,98655 | 421 | 209,0257 | 0,048278 | 1 | 0,286754 |
| -0,9323 | 420 | 413,2587 | 0,351725 | 1 | 0,947373 |
| -1,58835 | 367 | 360,2988 | 0,113085 | 1 | 0,439223 |
| 0,120433 | 280 | 272,087 | 0,904229 | 1 | 0,975308 |
| 0,686571 | 421 | 390,0994 | 0,492761 | 1 | 0,748579 |
| 0,807713 | 421 | 205,6247 | 0,420189 | 1 | 0,691294 |
| 0,853028 | 420 | 411,4056 | 0,39414 | 1 | 0,95743 |
| 2,097903 | 367 | 355,1923 | 0,03662 | 1 | 0,28464 |
| 0,515337 | 280 | 268,3817 | 0,606742 | 1 | 0,88515 |
| -2,35376 | 421 | 364,3576 | 0,019115 | 1 | 0,159771 |
| -1,21364 | 421 | 197,6429 | 0,226332 | 1 | 0,524743 |
| 0,711196 | 420 | 397,8642 | 0,477379 | 1 | 0,960096 |
| -0,7409 | 367 | 342,6329 | 0,459263 | 1 | 0,763917 |
| 0,101256 | 280 | 265,4341 | 0,919423 | 1 | 0,977051 |
| -1,02887 | 421 | 401,4932 | 0,30416 | 1 | 0,600745 |
| -0,4748 | 421 | 217,251 | 0,635404 | 1 | 0,836291 |
| 0,173409 | 420 | 415,3675 | 0,862415 | 1 | 0,977432 |
| 0,838019 | 367 | 361,5132 | 0,402574 | 1 | 0,716268 |
| -0,09469 | 280 | 275,2942 | 0,924634 | 1 | 0,979504 |
| -1,8804 | 421 | 317,8854 | 0,060968 | 1 | 0,270611 |

| | | | | | |
|---|---|---|---|---|---|
| 0,873707 | 421 | 179,7506 | 0,383443 | 1 | 0,659083 |
| 0,350411 | 420 | 354,728 | 0,726238 | 1 | 0,969427 |
| 1,776954 | 367 | 301,2407 | 0,076585 | 1 | 0,378716 |
| 0,756872 | 280 | 250,2639 | 0,449839 | 1 | 0,813228 |
| 0,182764 | 421 | 372,6793 | 0,855083 | 1 | 0,947051 |
| 1,170541 | 421 | 198,5105 | 0,243186 | 1 | 0,542422 |
| 0,648385 | 420 | 401,3336 | 0,517107 | 1 | 0,960096 |
| -1,24729 | 367 | 346,9433 | 0,213131 | 1 | 0,563645 |
| -0,38602 | 280 | 265,2199 | 0,69979 | 1 | 0,92034 |
| -0,68723 | 421 | 321,3329 | 0,492436 | 1 | 0,748579 |
| 0,259025 | 421 | 179,3958 | 0,795913 | 1 | 0,92123 |
| -0,7107 | 420 | 355,0785 | 0,477736 | 1 | 0,960096 |
| 0,07668 | 367 | 307,9784 | 0,938928 | 1 | 0,979214 |
| 1,584287 | 280 | 248,7221 | 0,114399 | 1 | 0,51157 |
| -1,82899 | 421 | 343,2689 | 0,068269 | 1 | 0,281097 |
| -0,36236 | 421 | 185,4395 | 0,717496 | 1 | 0,874811 |
| -0,31674 | 420 | 380,7448 | 0,751611 | 1 | 0,969427 |
| 0,599573 | 367 | 334,7574 | 0,549196 | 1 | 0,828521 |
| -0,38696 | 280 | 261,7585 | 0,699101 | 1 | 0,92034 |
| -2,43574 | 421 | 370,5323 | 0,015332 | 1 | 0,147897 |
| -1,50756 | 421 | 199,6353 | 0,133247 | 1 | 0,414884 |
| 0,210266 | 420 | 403,3709 | 0,833567 | 1 | 0,973894 |
| 0,58445 | 367 | 349,6542 | 0,559294 | 1 | 0,832862 |
| 0,211992 | 280 | 268,4182 | 0,832274 | 1 | 0,964667 |
| 0,624397 | 421 | 412,6149 | 0,532712 | 1 | 0,777904 |
| 0,925843 | 421 | 232,0588 | 0,35549 | 1 | 0,643758 |
| 0,365523 | 420 | 414,7357 | 0,714907 | 1 | 0,968866 |
| -0,16779 | 367 | 362,0322 | 0,866845 | 1 | 0,959636 |
| 0,419143 | 280 | 274,799 | 0,675439 | 1 | 0,909385 |
| -1,17129 | 421 | 374,0104 | 0,242227 | 1 | 0,53918 |
| -0,18206 | 421 | 199,539 | 0,855718 | 1 | 0,945471 |
| 0,366584 | 420 | 404,3884 | 0,714121 | 1 | 0,968866 |
| 0,332836 | 367 | 356,1043 | 0,739454 | 1 | 0,923669 |
| 0,528617 | 280 | 271,7961 | 0,597503 | 1 | 0,880334 |
| -1,37618 | 421 | 375,8516 | 0,169584 | 1 | 0,442288 |
| -1,64794 | 421 | 200,7389 | 0,100929 | 1 | 0,383011 |
| 0,143821 | 420 | 403,7468 | 0,885714 | 1 | 0,982463 |
| -1,19542 | 367 | 356,5785 | 0,232719 | 1 | 0,582158 |
| -0,34421 | 280 | 267,4034 | 0,730958 | 1 | 0,936637 |
| 2,690951 | 421 | 342,4931 | 0,007474 | 0,911863 | 0,107274 |
| 2,556827 | 421 | 186,3078 | 0,01136 | 1 | 0,163285 |
| 0,625596 | 420 | 378,4146 | 0,531957 | 1 | 0,960096 |
| 2,676773 | 367 | 331,4374 | 0,007803 | 0,983202 | 0,180403 |
| 2,77857 | 280 | 271,3374 | 0,005841 | 0,753505 | 0,186692 |
| 0,46849 | 421 | 315,38 | 0,639758 | 1 | 0,834842 |
| 2,903664 | 421 | 181,6914 | 0,004146 | 1 | 0,120964 |
| -0,01198 | 420 | 352,2675 | 0,990447 | 1 | 0,997342 |
| 0,304833 | 367 | 319,5874 | 0,760691 | 1 | 0,929966 |

| | | | | | |
|---|---|---|---|---|---|
| 1,605806 | 280 | 251,3611 | 0,109572 | 1 | 0,500379 |
| 0,285677 | 421 | 404,3918 | 0,775271 | 1 | 0,911337 |
| 2,405566 | 421 | 219,6866 | 0,016976 | 1 | 0,181988 |
| 0,993658 | 420 | 415,7624 | 0,320968 | 1 | 0,945573 |
| 0,088637 | 367 | 362,9668 | 0,929419 | 1 | 0,976733 |
| 0,790709 | 280 | 270,2979 | 0,429807 | 1 | 0,801886 |
| -0,23955 | 421 | 332,0782 | 0,810827 | 1 | 0,924931 |
| 1,633628 | 421 | 185,4318 | 0,104034 | 1 | 0,383322 |
| 0,198086 | 420 | 365,7437 | 0,843088 | 1 | 0,973894 |
| 1,1041 | 367 | 321,853 | 0,270375 | 1 | 0,609149 |
| 0,931865 | 280 | 266,2887 | 0,35225 | 1 | 0,761846 |
| -0,313 | 421 | 330,9683 | 0,754479 | 1 | 0,898125 |
| -0,8001 | 421 | 185,1617 | 0,424677 | 1 | 0,693378 |
| -0,53235 | 420 | 364,3543 | 0,59481 | 1 | 0,964878 |
| 0,894798 | 367 | 323,0218 | 0,371561 | 1 | 0,68168 |
| 1,023553 | 280 | 256,0784 | 0,307012 | 1 | 0,72861 |
| -0,66135 | 421 | 371,9074 | 0,508797 | 1 | 0,762184 |
| 0,246458 | 421 | 197,8571 | 0,805583 | 1 | 0,923205 |
| -0,20147 | 420 | 400,4687 | 0,840436 | 1 | 0,973894 |
| 1,185613 | 367 | 342,1052 | 0,236598 | 1 | 0,585627 |
| 0,862219 | 280 | 273,5532 | 0,389322 | 1 | 0,777098 |
| 1,395483 | 421 | 373,8429 | 0,163699 | 1 | 0,438177 |
| 1,484709 | 421 | 193,8851 | 0,139245 | 1 | 0,423351 |
| 0,46798 | 420 | 402,6647 | 0,640052 | 1 | 0,964878 |
| 2,62447 | 367 | 351,1159 | 0,009057 | 1 | 0,183886 |
| 2,198986 | 280 | 274,8183 | 0,028712 | 1 | 0,326641 |
| -0,63561 | 421 | 414,3484 | 0,525382 | 1 | 0,770244 |
| -0,09899 | 421 | 234,1414 | 0,921229 | 1 | 0,970834 |
| -0,28366 | 420 | 413,3083 | 0,776809 | 1 | 0,969427 |
| 1,130387 | 367 | 362,3904 | 0,259061 | 1 | 0,609149 |
| -0,15828 | 280 | 274,3616 | 0,874351 | 1 | 0,971 |
| -2,24576 | 421 | 323,6558 | 0,025394 | 1 | 0,184916 |
| -2,63423 | 421 | 184,3765 | 0,00915 | 1 | 0,14815 |
| 0,570698 | 420 | 358,5467 | 0,568562 | 1 | 0,962723 |
| -0,15702 | 367 | 329,3156 | 0,875325 | 1 | 0,962417 |
| -1,11876 | 280 | 255,2349 | 0,264296 | 1 | 0,688875 |
| -0,40402 | 421 | 310,1323 | 0,686479 | 1 | 0,864986 |
| 0,830679 | 421 | 178,5766 | 0,407264 | 1 | 0,681185 |
| -0,06089 | 420 | 343,8757 | 0,951482 | 1 | 0,992811 |
| 0,879105 | 367 | 302,0558 | 0,380043 | 1 | 0,691076 |
| 0,836456 | 280 | 247,1131 | 0,403706 | 1 | 0,786359 |
| 0,691255 | 421 | 385,1319 | 0,489822 | 1 | 0,748388 |
| 0,441574 | 421 | 206,5433 | 0,65926 | 1 | 0,842667 |
| 0,89274 | 420 | 409,6856 | 0,37252 | 1 | 0,95743 |
| -0,87895 | 367 | 350,83 | 0,380031 | 1 | 0,691076 |
| -0,66165 | 280 | 266,1438 | 0,508767 | 1 | 0,831912 |
| -1,10947 | 421 | 331,3281 | 0,268034 | 1 | 0,563516 |
| -2,68415 | 421 | 185,9491 | 0,007928 | 1 | 0,148102 |

| | | | | | |
|---|---|---|---|---|---|
| -1,09013 | 420 | 370,7165 | 0,276363 | 1 | 0,943897 |
| -1,37333 | 367 | 315,68 | 0,170625 | 1 | 0,51841 |
| -2,16186 | 280 | 252,3862 | 0,031569 | 1 | 0,329568 |
| -2,9147 | 421 | 335,4656 | 0,0038 | 1 | 0,087964 |
| -0,94986 | 421 | 187,4911 | 0,343408 | 1 | 0,636552 |
| 0,4644 | 420 | 374,6156 | 0,642631 | 1 | 0,964878 |
| -1,35883 | 367 | 333,1804 | 0,175119 | 1 | 0,523018 |
| 0,162399 | 280 | 261,2224 | 0,871117 | 1 | 0,969656 |
| 1,856013 | 421 | 344,4514 | 0,064306 | 1 | 0,275801 |
| 1,375729 | 421 | 187,3477 | 0,170548 | 1 | 0,456365 |
| 1,322835 | 420 | 375,6871 | 0,186695 | 1 | 0,923408 |
| 0,67168 | 367 | 328,2808 | 0,50226 | 1 | 0,793601 |
| -0,77021 | 280 | 256,3085 | 0,441883 | 1 | 0,810247 |
| -0,13894 | 421 | 342,7796 | 0,889582 | 1 | 0,962 |
| 0,193557 | 421 | 188,2083 | 0,846731 | 1 | 0,943293 |
| -0,55459 | 420 | 378,1352 | 0,579503 | 1 | 0,964878 |
| -0,015 | 367 | 327,51 | 0,988038 | 1 | 0,996248 |
| 0,710041 | 280 | 257,2632 | 0,478322 | 1 | 0,824256 |
| -1,87963 | 421 | 325,0274 | 0,061054 | 1 | 0,270611 |
| -0,68735 | 421 | 183,2253 | 0,492729 | 1 | 0,746275 |
| 2,083572 | 420 | 364,242 | 0,037896 | 1 | 0,83984 |
| 0,667308 | 367 | 335,5129 | 0,505034 | 1 | 0,793601 |
| 1,083664 | 280 | 262,598 | 0,279508 | 1 | 0,703716 |
| 0,333467 | 421 | 379,1098 | 0,738966 | 1 | 0,893273 |
| 1,093324 | 421 | 201,3152 | 0,275558 | 1 | 0,576759 |
| -0,61027 | 420 | 406,0298 | 0,542024 | 1 | 0,960096 |
| 0,859528 | 367 | 349,1095 | 0,390639 | 1 | 0,705022 |
| 0,237455 | 280 | 272,2022 | 0,812483 | 1 | 0,95747 |
| -0,17359 | 421 | 331,4373 | 0,862296 | 1 | 0,951261 |
| -0,63385 | 421 | 184,8652 | 0,526963 | 1 | 0,767321 |
| 0,030801 | 420 | 365,6975 | 0,975445 | 1 | 0,997342 |
| 0,33434 | 367 | 317,4446 | 0,738344 | 1 | 0,923669 |
| -3,32828 | 280 | 257,2297 | 0,001002 | 1 | 0,121989 |
| -0,70322 | 421 | 372,5466 | 0,48236 | 1 | 0,74251 |
| 0,038687 | 421 | 197,8867 | 0,969179 | 1 | 0,988858 |
| 0,960441 | 420 | 404,2441 | 0,337407 | 1 | 0,947373 |
| 2,738083 | 367 | 355,5653 | 0,006491 | 1 | 0,178183 |
| -0,82721 | 280 | 266,0567 | 0,408858 | 1 | 0,788151 |
| -1,55378 | 421 | 364,9421 | 0,121103 | 1 | 0,382129 |
| -0,42516 | 421 | 197,2531 | 0,67118 | 1 | 0,852122 |
| 0,061942 | 420 | 399,0087 | 0,95064 | 1 | 0,992811 |
| -0,1715 | 367 | 351,3819 | 0,863929 | 1 | 0,959636 |
| 0,680014 | 280 | 267,695 | 0,497083 | 1 | 0,830492 |
| 0,847958 | 421 | 405,3011 | 0,396962 | 1 | 0,686821 |
| 1,833227 | 421 | 218,6602 | 0,068128 | 1 | 0,334428 |
| 0,261099 | 420 | 415,9756 | 0,794146 | 1 | 0,969427 |
| 1,513289 | 367 | 361,5083 | 0,13108 | 1 | 0,47485 |
| 1,661235 | 280 | 272,5664 | 0,097816 | 1 | 0,480557 |

| | | | | | |
|---|---|---|---|---|---|
| 0,877074 | 421 | 315,9817 | 0,381113 | 1 | 0,671614 |
| 1,229017 | 421 | 182,5045 | 0,220648 | 1 | 0,516786 |
| -0,37606 | 420 | 350,5334 | 0,707098 | 1 | 0,968866 |
| 1,822729 | 367 | 321,877 | 0,069272 | 1 | 0,364488 |
| 1,245187 | 280 | 261,4069 | 0,214178 | 1 | 0,646827 |
| -0,41962 | 421 | 383,9488 | 0,674997 | 1 | 0,859543 |
| 0,518896 | 421 | 204,4937 | 0,604394 | 1 | 0,820561 |
| 1,113288 | 420 | 406,9765 | 0,266242 | 1 | 0,943897 |
| -0,12126 | 367 | 357,7856 | 0,903552 | 1 | 0,970486 |
| -0,58085 | 280 | 270,2957 | 0,561828 | 1 | 0,861728 |
| 1,464725 | 421 | 391,0827 | 0,143799 | 1 | 0,412154 |
| 1,45609 | 421 | 210,3614 | 0,146859 | 1 | 0,428077 |
| 0,362201 | 420 | 412,7593 | 0,717387 | 1 | 0,969427 |
| -0,91044 | 367 | 358,2706 | 0,363203 | 1 | 0,677287 |
| 0,164772 | 280 | 271,989 | 0,869246 | 1 | 0,969656 |
| -0,45496 | 421 | 288,8439 | 0,649477 | 1 | 0,841583 |
| 1,452222 | 421 | 174,1257 | 0,148239 | 1 | 0,428784 |
| 1,38218 | 420 | 314,8003 | 0,167896 | 1 | 0,914005 |
| 0,114938 | 367 | 297,4677 | 0,908572 | 1 | 0,970624 |
| -1,4895 | 280 | 249,7545 | 0,137618 | 1 | 0,54931 |
| -0,85075 | 421 | 286,3578 | 0,395618 | 1 | 0,686065 |
| 0,446482 | 421 | 173,2704 | 0,655806 | 1 | 0,841107 |
| 0,506261 | 420 | 316,1905 | 0,613026 | 1 | 0,964878 |
| 2,432326 | 367 | 283,9737 | 0,01562 | 1 | 0,204372 |
| 0,84395 | 280 | 276 | 0,399428 | 1 | 0,784757 |
| -0,41756 | 421 | 369,1451 | 0,676512 | 1 | 0,860341 |
| -0,15024 | 421 | 196,7671 | 0,880732 | 1 | 0,956931 |
| 0,094398 | 420 | 398,9898 | 0,924841 | 1 | 0,989204 |
| 1,317779 | 367 | 335,8324 | 0,188476 | 1 | 0,538722 |
| 0,050049 | 280 | 259,2614 | 0,960122 | 1 | 0,986974 |
| 1,981915 | 421 | 397,9303 | 0,048176 | 1 | 0,241202 |
| 1,982732 | 421 | 214,0287 | 0,048676 | 1 | 0,286808 |
| 1,131957 | 420 | 414,4592 | 0,258307 | 1 | 0,943897 |
| 1,692593 | 367 | 361,651 | 0,091394 | 1 | 0,403903 |
| 0,635189 | 280 | 274,6457 | 0,525834 | 1 | 0,837665 |
| 0,451715 | 421 | 375,8685 | 0,651735 | 1 | 0,843783 |
| -0,43503 | 421 | 198,6833 | 0,664015 | 1 | 0,845876 |
| 2,094468 | 420 | 401,9486 | 0,036844 | 1 | 0,828713 |
| -0,13352 | 367 | 347,796 | 0,893857 | 1 | 0,969095 |
| 0,869949 | 280 | 268,5469 | 0,385105 | 1 | 0,773166 |
| -0,65935 | 421 | 312,8036 | 0,510156 | 1 | 0,763461 |
| -0,11298 | 421 | 177,4882 | 0,910176 | 1 | 0,96646 |
| 0,391235 | 420 | 346,8796 | 0,695864 | 1 | 0,968866 |
| 0,343206 | 367 | 296,9155 | 0,731687 | 1 | 0,923669 |
| 0,848071 | 280 | 252,9802 | 0,3972 | 1 | 0,782458 |
| 2,891958 | 421 | 394,8344 | 0,00404 | 0,517157 | 0,087964 |
| 3,118397 | 421 | 213,0665 | 0,00207 | 0,26904 | 0,099921 |
| 0,721708 | 420 | 414,9563 | 0,47088 | 1 | 0,960096 |

| | | | | | |
|---|---|---|---|---|---|
| 2,715232 | 367 | 359,1638 | 0,006943 | 0,895625 | 0,180403 |
| 1,473967 | 280 | 271,5174 | 0,141649 | 1 | 0,554427 |
| -2,99615 | 421 | 326,4726 | 0,002943 | 0,441515 | 0,077359 |
| -1,9263 | 421 | 184,5061 | 0,055604 | 1 | 0,309036 |
| -0,8879 | 420 | 360,0954 | 0,375185 | 1 | 0,95743 |
| -2,32539 | 367 | 326,978 | 0,020662 | 1 | 0,224016 |
| -1,1928 | 280 | 258,0987 | 0,234042 | 1 | 0,659852 |
| 0,549767 | 421 | 342,7123 | 0,582837 | 1 | 0,806553 |
| -1,31886 | 421 | 188,3332 | 0,188816 | 1 | 0,476746 |
| -3,10504 | 420 | 378,1066 | 0,002046 | 0,274155 | 0,294094 |
| 1,188808 | 367 | 336,4936 | 0,235353 | 1 | 0,585627 |
| -0,18103 | 280 | 261,1816 | 0,856487 | 1 | 0,966655 |
| 0,528209 | 421 | 355,9146 | 0,597684 | 1 | 0,815122 |
| -0,88437 | 421 | 193,9782 | 0,377593 | 1 | 0,655017 |
| 1,941739 | 420 | 391,8351 | 0,052885 | 1 | 0,846627 |
| -0,60498 | 367 | 329,7922 | 0,545605 | 1 | 0,825529 |
| 0,693602 | 280 | 264,3906 | 0,488541 | 1 | 0,826999 |
| -1,34083 | 421 | 331,019 | 0,180896 | 1 | 0,46049 |
| -0,77307 | 421 | 185,0679 | 0,44047 | 1 | 0,705407 |
| -0,07814 | 420 | 365,8752 | 0,937757 | 1 | 0,992059 |
| 0,380925 | 367 | 333,4454 | 0,703501 | 1 | 0,908463 |
| -1,15887 | 280 | 260,8006 | 0,24757 | 1 | 0,677438 |
| -1,86395 | 421 | 336,8716 | 0,063199 | 1 | 0,274468 |
| -0,99255 | 421 | 187,7066 | 0,322209 | 1 | 0,617772 |
| 0,12469 | 420 | 375,0966 | 0,900836 | 1 | 0,98503 |
| 1,052233 | 367 | 326,9544 | 0,29347 | 1 | 0,627013 |
| 1,523959 | 280 | 258,7944 | 0,12874 | 1 | 0,536498 |
| 1,074185 | 421 | 344,2251 | 0,283492 | 1 | 0,582841 |
| 0,898905 | 421 | 185,5869 | 0,369868 | 1 | 0,64964 |
| 0,046109 | 420 | 378,2789 | 0,963247 | 1 | 0,996924 |
| 1,114921 | 367 | 332,3354 | 0,26569 | 1 | 0,609149 |
| 0,427499 | 280 | 264,447 | 0,669364 | 1 | 0,908644 |
| -1,2904 | 421 | 331,5352 | 0,197811 | 1 | 0,484381 |
| -1,22763 | 421 | 185,2451 | 0,221142 | 1 | 0,516786 |
| -0,0855 | 420 | 366,9638 | 0,931907 | 1 | 0,991559 |
| -1,35175 | 367 | 328,3201 | 0,177385 | 1 | 0,523131 |
| -0,2507 | 280 | 262,23 | 0,802244 | 1 | 0,956473 |
| -1,68277 | 421 | 349,8119 | 0,093312 | 1 | 0,327788 |
| -2,12045 | 421 | 191,1459 | 0,035258 | 1 | 0,255449 |
| 1,057054 | 420 | 382,6747 | 0,291154 | 1 | 0,943897 |
| -0,79996 | 367 | 341,6848 | 0,424292 | 1 | 0,733267 |
| -0,5285 | 280 | 263,2729 | 0,597599 | 1 | 0,880334 |
| 0,636628 | 421 | 342,7812 | 0,524792 | 1 | 0,770244 |
| 1,198095 | 421 | 188,7721 | 0,232382 | 1 | 0,533704 |
| 0,448881 | 420 | 378,943 | 0,653774 | 1 | 0,966867 |
| 2,310479 | 367 | 329,7775 | 0,021478 | 1 | 0,227943 |
| 0,140418 | 280 | 263,5209 | 0,888437 | 1 | 0,975145 |
| -0,39508 | 421 | 406,4711 | 0,692994 | 1 | 0,870285 |

| | | | | | |
|---|---|---|---|---|---|
| -0,61117 | 421 | 217,9119 | 0,541721 | 1 | 0,773814 |
| 0,070762 | 420 | 415,9991 | 0,943621 | 1 | 0,992811 |
| -0,42406 | 367 | 362,9048 | 0,671772 | 1 | 0,897482 |
| 2,102209 | 280 | 273,1731 | 0,03645 | 1 | 0,34044 |
| -1,89803 | 421 | 395,2871 | 0,05842 | 1 | 0,268334 |
| -1,8114 | 421 | 212,5055 | 0,071491 | 1 | 0,339864 |
| -0,61045 | 420 | 413,8736 | 0,541897 | 1 | 0,960096 |
| -1,08365 | 367 | 358,5795 | 0,279246 | 1 | 0,613446 |
| -0,71274 | 280 | 273,1506 | 0,476618 | 1 | 0,824256 |
| -0,61872 | 421 | 381,5365 | 0,536469 | 1 | 0,778195 |
| -1,32036 | 421 | 203,5046 | 0,188198 | 1 | 0,476663 |
| 1,593883 | 420 | 408,1208 | 0,111736 | 1 | 0,848246 |
| -1,02105 | 367 | 354,2529 | 0,307927 | 1 | 0,640947 |
| 0,27796 | 280 | 269,4141 | 0,781257 | 1 | 0,951006 |
| -0,38165 | 421 | 323,1705 | 0,702971 | 1 | 0,874567 |
| 0,55658 | 421 | 180,1846 | 0,578505 | 1 | 0,794719 |
| -0,42012 | 420 | 357,3044 | 0,674653 | 1 | 0,96781 |
| -0,29526 | 367 | 308,9778 | 0,767992 | 1 | 0,935621 |
| 1,644059 | 280 | 245,5458 | 0,101444 | 1 | 0,480741 |
| -2,11113 | 421 | 313,9074 | 0,035552 | 1 | 0,215166 |
| -2,7273 | 421 | 177,8901 | 0,007025 | 1 | 0,141161 |
| -1,19014 | 420 | 349,1058 | 0,234801 | 1 | 0,943055 |
| -2,5056 | 367 | 309,6481 | 0,012737 | 1 | 0,189546 |
| -2,13983 | 280 | 262,7331 | 0,03329 | 1 | 0,329568 |
| 0,986581 | 421 | 355,8835 | 0,324518 | 1 | 0,61749 |
| 0,997417 | 421 | 194,5439 | 0,319801 | 1 | 0,617081 |
| 0,390526 | 420 | 390,4039 | 0,696361 | 1 | 0,968866 |
| 0,327085 | 367 | 329,9738 | 0,743811 | 1 | 0,923669 |
| -1,43199 | 280 | 260,0426 | 0,153348 | 1 | 0,565026 |
| 0,450482 | 421 | 313,8198 | 0,652674 | 1 | 0,844274 |
| 1,232546 | 421 | 180,265 | 0,21935 | 1 | 0,515696 |
| 1,033357 | 420 | 348,0289 | 0,302154 | 1 | 0,943897 |
| -0,91749 | 367 | 301,2833 | 0,359622 | 1 | 0,674813 |
| 0,969503 | 280 | 250,9081 | 0,333228 | 1 | 0,755731 |
| 1,85793 | 421 | 334,4123 | 0,064058 | 1 | 0,275801 |
| 1,886957 | 421 | 182,7652 | 0,060751 | 1 | 0,315698 |
| 1,399198 | 420 | 374,5787 | 0,162581 | 1 | 0,914005 |
| 1,185606 | 367 | 319,1515 | 0,23666 | 1 | 0,585627 |
| 3,025839 | 280 | 262,7156 | 0,002725 | 1 | 0,135787 |
| -0,60173 | 421 | 309,0412 | 0,547793 | 1 | 0,784719 |
| 1,662595 | 421 | 179,8539 | 0,098135 | 1 | 0,379251 |
| 0,668271 | 420 | 344,0325 | 0,504409 | 1 | 0,960096 |
| 0,111102 | 367 | 303,2259 | 0,911609 | 1 | 0,971566 |
| 0,954635 | 280 | 247,5013 | 0,340694 | 1 | 0,757098 |
| -1,73717 | 421 | 379,3001 | 0,08317 | 1 | 0,306447 |
| -1,53384 | 421 | 202,2884 | 0,126632 | 1 | 0,410224 |
| -2,22101 | 420 | 403,4673 | 0,026905 | 1 | 0,767829 |
| -0,63641 | 367 | 357,119 | 0,524916 | 1 | 0,808843 |

| | | | | | |
|---|---|---|---|---|---|
| 0,294036 | 280 | 269,1857 | 0,768957 | 1 | 0,946609 |
| 0,714285 | 421 | 330,2562 | 0,475556 | 1 | 0,734285 |
| 0,358909 | 421 | 184,9523 | 0,720072 | 1 | 0,875826 |
| 0,030339 | 420 | 366,8282 | 0,975813 | 1 | 0,997342 |
| -0,17853 | 367 | 316,7513 | 0,858423 | 1 | 0,958964 |
| -0,09025 | 280 | 260,7022 | 0,92816 | 1 | 0,979504 |
| -0,15627 | 421 | 362,8407 | 0,875905 | 1 | 0,95861 |
| 1,473583 | 421 | 196,1804 | 0,142197 | 1 | 0,423499 |
| -0,08235 | 420 | 394,522 | 0,934406 | 1 | 0,992059 |
| 0,940739 | 367 | 347,1827 | 0,347493 | 1 | 0,668671 |
| 0,420639 | 280 | 261,0522 | 0,674365 | 1 | 0,909385 |
| -0,30224 | 421 | 351,5805 | 0,762645 | 1 | 0,902833 |
| 1,151658 | 421 | 190,0652 | 0,250908 | 1 | 0,548794 |
| 0,588959 | 420 | 385,8864 | 0,556233 | 1 | 0,960096 |
| 1,126654 | 367 | 335,1554 | 0,260695 | 1 | 0,609149 |
| 1,447031 | 280 | 260,387 | 0,149091 | 1 | 0,561358 |
| -1,15821 | 421 | 340,4153 | 0,247591 | 1 | 0,544195 |
| 0,558711 | 421 | 188,1593 | 0,577023 | 1 | 0,794719 |
| 0,923003 | 420 | 376,6377 | 0,356596 | 1 | 0,947373 |
| 0,194663 | 367 | 331,7747 | 0,845776 | 1 | 0,956935 |
| -0,17946 | 280 | 261,7192 | 0,857719 | 1 | 0,966655 |
| -1,38312 | 421 | 382,0044 | 0,167435 | 1 | 0,441901 |
| -2,73504 | 421 | 203,7986 | 0,006787 | 0,990894 | 0,141161 |
| -1,97806 | 420 | 407,6153 | 0,048595 | 1 | 0,846627 |
| -1,729 | 367 | 351,1092 | 0,084689 | 1 | 0,388246 |
| 0,653971 | 280 | 269,8911 | 0,513687 | 1 | 0,831912 |
| -1,21147 | 421 | 360,589 | 0,226509 | 1 | 0,518767 |
| -1,58061 | 421 | 193,9419 | 0,115597 | 1 | 0,392379 |
| -1,77305 | 420 | 393,0112 | 0,076996 | 1 | 0,848246 |
| 0,526704 | 367 | 338,3657 | 0,598744 | 1 | 0,861803 |
| 0,310621 | 280 | 260,012 | 0,756338 | 1 | 0,942071 |
| -1,21926 | 421 | 345,7544 | 0,223575 | 1 | 0,51676 |
| -1,0787 | 421 | 190,5779 | 0,282085 | 1 | 0,581951 |
| -0,97708 | 420 | 381,724 | 0,329151 | 1 | 0,947373 |
| -1,61793 | 367 | 323,5469 | 0,106653 | 1 | 0,432057 |
| -2,30043 | 280 | 265,6023 | 0,022199 | 1 | 0,309763 |
| 1,851372 | 421 | 368,1565 | 0,064917 | 1 | 0,276236 |
| 1,384711 | 421 | 196,3607 | 0,167712 | 1 | 0,45454 |
| 1,049549 | 420 | 399,8968 | 0,294559 | 1 | 0,943897 |
| 0,908003 | 367 | 350,6145 | 0,3645 | 1 | 0,677906 |
| -0,06699 | 280 | 266,0934 | 0,946637 | 1 | 0,982409 |
| -1,521 | 421 | 373,4741 | 0,129105 | 1 | 0,397064 |
| -0,34427 | 421 | 199,8846 | 0,731004 | 1 | 0,882616 |
| -0,09699 | 420 | 403,3985 | 0,922781 | 1 | 0,989204 |
| 0,114409 | 367 | 346,3845 | 0,90898 | 1 | 0,970624 |
| -2,92896 | 280 | 267,1698 | 0,003694 | 1 | 0,147267 |
| -1,97516 | 421 | 297,0574 | 0,049176 | 1 | 0,244581 |
| 0,019207 | 421 | 176,6269 | 0,984698 | 1 | 0,995074 |

| | | | | | |
|---|---|---|---|---|---|
| -2,05477 | 420 | 322,6269 | 0,040706 | 1 | 0,846132 |
| 0,136848 | 367 | 299,644 | 0,891243 | 1 | 0,968351 |
| 1,068752 | 280 | 247,4777 | 0,286223 | 1 | 0,714135 |
| -0,22307 | 421 | 377,8307 | 0,823603 | 1 | 0,933913 |
| 0,968289 | 421 | 199,7419 | 0,334071 | 1 | 0,62852 |
| 0,733 | 420 | 405,4129 | 0,463982 | 1 | 0,960096 |
| -0,06242 | 367 | 354,8035 | 0,950262 | 1 | 0,979539 |
| -0,143 | 280 | 268,4367 | 0,886398 | 1 | 0,973784 |
| -2,35998 | 421 | 342,0787 | 0,018837 | 1 | 0,15948 |
| -1,75402 | 421 | 185,7604 | 0,081076 | 1 | 0,360416 |
| -2,43463 | 420 | 378,7515 | 0,015368 | 1 | 0,723749 |
| -1,71723 | 367 | 331,5114 | 0,086871 | 1 | 0,391959 |
| 0,663031 | 280 | 265,4504 | 0,507886 | 1 | 0,831912 |
| -1,14259 | 421 | 328,8185 | 0,25404 | 1 | 0,546261 |
| 0,863129 | 421 | 182,8703 | 0,389197 | 1 | 0,664422 |
| -0,08119 | 420 | 364,2012 | 0,935337 | 1 | 0,992059 |
| -0,28086 | 367 | 308,1366 | 0,779007 | 1 | 0,942943 |
| 0,541267 | 280 | 276 | 0,58876 | 1 | 0,87561 |
| -0,93905 | 421 | 300,7722 | 0,348458 | 1 | 0,637437 |
| -0,79487 | 421 | 178,5929 | 0,427747 | 1 | 0,694836 |
| -1,0365 | 420 | 332,1866 | 0,300723 | 1 | 0,943897 |
| 0,147527 | 367 | 292,4143 | 0,882818 | 1 | 0,962667 |
| -1,14367 | 280 | 276 | 0,253751 | 1 | 0,679673 |
| 0,09566 | 421 | 359,5069 | 0,923844 | 1 | 0,966828 |
| -1,0754 | 421 | 193,7452 | 0,283531 | 1 | 0,582654 |
| -0,19378 | 420 | 389,3374 | 0,846451 | 1 | 0,973894 |
| -1,04971 | 367 | 331,9234 | 0,294614 | 1 | 0,62734 |
| 0,438521 | 280 | 261,1523 | 0,661372 | 1 | 0,908644 |
| 0,206847 | 421 | 395,1825 | 0,836236 | 1 | 0,93721 |
| 0,904012 | 421 | 211,143 | 0,367019 | 1 | 0,647779 |
| -0,29531 | 420 | 413,4716 | 0,767903 | 1 | 0,969427 |
| -1,31966 | 367 | 360,1996 | 0,187787 | 1 | 0,538014 |
| -0,00875 | 280 | 270,8259 | 0,993025 | 1 | 0,996331 |
| -1,262 | 421 | 344,2699 | 0,207802 | 1 | 0,496728 |
| -1,23087 | 421 | 185,5748 | 0,219931 | 1 | 0,516256 |
| -0,02084 | 420 | 380,5925 | 0,983383 | 1 | 0,997342 |
| 0,013363 | 367 | 339,2774 | 0,989346 | 1 | 0,996248 |
| -1,83855 | 280 | 261,9063 | 0,067113 | 1 | 0,425749 |
| -1,10745 | 421 | 405,5686 | 0,268756 | 1 | 0,563516 |
| -0,64573 | 421 | 221,8911 | 0,519124 | 1 | 0,761752 |
| -0,6458 | 420 | 415,9446 | 0,518763 | 1 | 0,960096 |
| -2,00143 | 367 | 362,968 | 0,04609 | 1 | 0,307337 |
| 0,593642 | 280 | 275,6302 | 0,553238 | 1 | 0,854225 |
| 1,021087 | 421 | 347,9017 | 0,307923 | 1 | 0,604218 |
| 0,09328 | 421 | 188,5995 | 0,92578 | 1 | 0,972483 |
| -0,12752 | 420 | 381,8207 | 0,898599 | 1 | 0,98503 |
| 1,785532 | 367 | 326,1943 | 0,075104 | 1 | 0,376019 |
| -0,34378 | 280 | 248,1955 | 0,731306 | 1 | 0,936637 |

| | | | | | |
|---|---|---|---|---|---|
| 0,211196 | 421 | 407,4088 | 0,83284 | 1 | 0,936611 |
| -0,37204 | 421 | 223,5729 | 0,710218 | 1 | 0,870869 |
| 0,620685 | 420 | 415,9933 | 0,535147 | 1 | 0,960096 |
| -1,763 | 367 | 361,7733 | 0,078745 | 1 | 0,381638 |
| 0,127765 | 280 | 268,5809 | 0,898431 | 1 | 0,975243 |
| 0,292369 | 421 | 388,9313 | 0,77016 | 1 | 0,908874 |
| 0,046912 | 421 | 207,1991 | 0,962628 | 1 | 0,985228 |
| -0,57599 | 420 | 410,7217 | 0,56494 | 1 | 0,961448 |
| 0,474481 | 367 | 356,1453 | 0,635448 | 1 | 0,880707 |
| -0,12315 | 280 | 268,8432 | 0,90208 | 1 | 0,975243 |
| -1,16952 | 421 | 374,9388 | 0,242935 | 1 | 0,53918 |
| -1,72528 | 421 | 202,3339 | 0,086005 | 1 | 0,365475 |
| -1,40368 | 420 | 403,7631 | 0,161184 | 1 | 0,914005 |
| -1,0907 | 367 | 349,8801 | 0,276155 | 1 | 0,611884 |
| -0,92651 | 280 | 265,9239 | 0,355021 | 1 | 0,761846 |
| -0,28122 | 421 | 393,4367 | 0,778693 | 1 | 0,913934 |
| -0,18671 | 421 | 208,9251 | 0,852072 | 1 | 0,944105 |
| -1,56444 | 420 | 413,3765 | 0,11848 | 1 | 0,858144 |
| -0,57207 | 367 | 359,0357 | 0,567635 | 1 | 0,837838 |
| 0,31141 | 280 | 271,3999 | 0,755728 | 1 | 0,942071 |

| | | | | | |
|---|---|---|---|---|---|
| 0,261387 | 421 | 291,3901 | 0,793979 | 1 | 0,919136 |

| | | | | | | |
|---|---|---|---|---|---|---|
| 2,468389 | 421 | 176,6911 | 0,014524 | | 1 | 0,177943 |

| | | | | | |
|---|---|---|---|---|---|
| 0,234489 | 420 | 323,8752 | 0,814753 | 1 | 0,969427 |

| | | | | | |
|---|---|---|---|---|---|
| 1,529538 | 367 | 290,4254 | 0,12722 | 1 | 0,468753 |

| | | | | | |
|---|---|---|---|---|---|
| 1,506406 | 280 | 276 | 0,133107 | 1 | 0,543276 |
| -0,22518 | 421 | 383,5916 | 0,821962 | 1 | 0,932754 |
| -0,82185 | 421 | 205,5047 | 0,412118 | 1 | 0,684933 |
| -0,84402 | 420 | 408,9753 | 0,399154 | 1 | 0,95743 |
| -0,40865 | 367 | 350,2533 | 0,68305 | 1 | 0,900574 |
| 0,301654 | 280 | 265,6756 | 0,763152 | 1 | 0,944296 |
| -0,76516 | 421 | 306,4555 | 0,444763 | 1 | 0,713799 |
| -1,63416 | 421 | 178,965 | 0,103984 | 1 | 0,383322 |
| -0,38674 | 420 | 343,3008 | 0,69919 | 1 | 0,968866 |
| -0,10693 | 367 | 301,6272 | 0,914918 | 1 | 0,973634 |
| -0,74309 | 280 | 257,1426 | 0,458109 | 1 | 0,817005 |
| 0,981789 | 421 | 340,8426 | 0,3269 | 1 | 0,61749 |
| 0,271377 | 421 | 187,7001 | 0,786399 | 1 | 0,917973 |
| 0,439635 | 420 | 373,8547 | 0,660455 | 1 | 0,96781 |
| -0,60126 | 367 | 335,0321 | 0,548076 | 1 | 0,828435 |
| -2,55307 | 280 | 258,3456 | 0,011253 | 1 | 0,234919 |
| 1,053335 | 421 | 368,5601 | 0,292878 | 1 | 0,590852 |
| 0,456747 | 421 | 197,0735 | 0,648356 | 1 | 0,839358 |
| 0,247483 | 420 | 397,8762 | 0,804662 | 1 | 0,969427 |
| -0,15276 | 367 | 344,0178 | 0,878675 | 1 | 0,962453 |

| | | | | | |
|---|---|---|---|---|---|
| 1,024944 | 280 | 263,0076 | 0,306332 | 1 | 0,72861 |
| 1,484916 | 421 | 329,7369 | 0,138522 | 1 | 0,410344 |
| 2,461161 | 421 | 184,5584 | 0,014768 | 1 | 0,178042 |
| -0,98898 | 420 | 368,3697 | 0,323323 | 1 | 0,945573 |
| 0,894284 | 367 | 328,0314 | 0,371826 | 1 | 0,68168 |
| 0,087328 | 280 | 260,5172 | 0,930478 | 1 | 0,979504 |
| -1,30203 | 421 | 347,5191 | 0,193768 | 1 | 0,479958 |
| -1,0297 | 421 | 188,9502 | 0,304467 | 1 | 0,602952 |
| -1,35203 | 420 | 380,0379 | 0,177169 | 1 | 0,9199 |
| 0,457548 | 367 | 335,4884 | 0,647573 | 1 | 0,884762 |
| 0,091101 | 280 | 264,363 | 0,927481 | 1 | 0,979504 |
| -0,17183 | 421 | 407,4554 | 0,863654 | 1 | 0,951261 |
| 0,902428 | 421 | 220,524 | 0,367814 | 1 | 0,648213 |
| 0,249209 | 420 | 415,9997 | 0,803322 | 1 | 0,969427 |
| 0,238646 | 367 | 362,7635 | 0,811515 | 1 | 0,950149 |
| 1,066772 | 280 | 273,8836 | 0,287014 | 1 | 0,714926 |
| 0,02724 | 421 | 386,948 | 0,978283 | 1 | 0,99613 |
| 0,919114 | 421 | 205,546 | 0,359113 | 1 | 0,643758 |
| 0,835882 | 420 | 409,2553 | 0,403709 | 1 | 0,95743 |
| 1,171314 | 367 | 358,7931 | 0,24225 | 1 | 0,59405 |
| -1,21638 | 280 | 269,9085 | 0,224901 | 1 | 0,654014 |
| 1,200699 | 421 | 389,4238 | 0,230598 | 1 | 0,521788 |
| 1,045011 | 421 | 207,4269 | 0,297234 | 1 | 0,595654 |
| 0,336691 | 420 | 410,7426 | 0,736522 | 1 | 0,969427 |
| -0,94907 | 367 | 358,2581 | 0,343222 | 1 | 0,665683 |
| 0,900072 | 280 | 267,9561 | 0,36889 | 1 | 0,763914 |
| -0,86343 | 421 | 318,0471 | 0,388551 | 1 | 0,677715 |
| -0,89969 | 421 | 181,236 | 0,369479 | 1 | 0,64964 |
| -0,04341 | 420 | 350,6882 | 0,965403 | 1 | 0,996924 |
| -0,56488 | 367 | 324,4311 | 0,572547 | 1 | 0,840144 |
| -0,97771 | 280 | 260,6618 | 0,329122 | 1 | 0,752282 |
| 0,036102 | 421 | 372,4119 | 0,97122 | 1 | 0,995666 |
| 0,736772 | 421 | 194,3263 | 0,46215 | 1 | 0,721079 |
| 1,032202 | 420 | 399,2189 | 0,302602 | 1 | 0,943897 |
| 2,441265 | 367 | 338,9704 | 0,015147 | 1 | 0,204372 |
| -0,29172 | 280 | 265,4827 | 0,770729 | 1 | 0,946609 |
| -1,93881 | 421 | 391,3723 | 0,053242 | 1 | 0,258827 |
| -1,61449 | 421 | 209,3561 | 0,107928 | 1 | 0,388181 |
| -1,74491 | 420 | 411,3007 | 0,081748 | 1 | 0,848246 |
| -0,27221 | 367 | 360,8366 | 0,78562 | 1 | 0,94337 |
| 0,347723 | 280 | 269,1211 | 0,72832 | 1 | 0,936637 |
| 0,568884 | 421 | 307,0467 | 0,569851 | 1 | 0,797368 |
| 2,160832 | 421 | 178,5938 | 0,03204 | 1 | 0,241423 |
| 0,575611 | 420 | 341,4071 | 0,565257 | 1 | 0,961448 |
| 1,598041 | 367 | 311,15 | 0,111049 | 1 | 0,437634 |
| 0,003387 | 280 | 249,6167 | 0,9973 | 1 | 0,998213 |
| -2,45357 | 421 | 403,4711 | 0,014567 | 1 | 0,14733 |
| -2,32115 | 421 | 218,723 | 0,021199 | 1 | 0,2022 |

| | | | | | |
|---|---|---|---|---|---|
| -3,28401 | 420 | 415,6324 | 0,00111 | 0,167613 | 0,235929 |
| -1,3819 | 367 | 362,6856 | 0,167854 | 1 | 0,514971 |
| -0,60891 | 280 | 271,4431 | 0,543092 | 1 | 0,849885 |
| 0,986053 | 421 | 390,5526 | 0,324718 | 1 | 0,61749 |
| 0,05217 | 421 | 207,4835 | 0,958444 | 1 | 0,98414 |
| -0,26814 | 420 | 411,5228 | 0,788722 | 1 | 0,969427 |
| 0,163165 | 367 | 355,8754 | 0,870481 | 1 | 0,959978 |
| -0,44763 | 280 | 269,7018 | 0,654777 | 1 | 0,908644 |
| -0,41385 | 421 | 417 | 0,679195 | 1 | 0,861268 |
| 2,057842 | 421 | 417 | 0,040226 | 1 | 0,26998 |
| -0,96448 | 420 | 416 | 0,335364 | 1 | 0,947373 |
| 1,080908 | 367 | 279,1165 | 0,280671 | 1 | 0,614784 |
| 1,765519 | 280 | 276 | 0,078582 | 1 | 0,453386 |
| -0,33884 | 421 | 298,2922 | 0,734969 | 1 | 0,893273 |
| -0,00175 | 421 | 175,1964 | 0,998603 | 1 | 0,999267 |
| -1,34441 | 420 | 336,0726 | 0,179724 | 1 | 0,923408 |
| -0,68157 | 367 | 276,7699 | 0,496082 | 1 | 0,793601 |
| 1,803485 | 280 | 251,756 | 0,072508 | 1 | 0,4406 |
| -0,69796 | 421 | 320,1413 | 0,485712 | 1 | 0,745445 |
| 2,038677 | 421 | 181,4417 | 0,042933 | 1 | 0,274154 |
| 0,604012 | 420 | 355,2417 | 0,546221 | 1 | 0,960096 |
| 2,144633 | 367 | 318,1115 | 0,032739 | 1 | 0,272587 |
| 2,047839 | 280 | 258,596 | 0,041586 | 1 | 0,373036 |
| -0,55829 | 421 | 392,8603 | 0,576962 | 1 | 0,802105 |
| -0,51689 | 421 | 209,3615 | 0,605779 | 1 | 0,821338 |
| -1,41197 | 420 | 413,1194 | 0,15871 | 1 | 0,914005 |
| -0,82874 | 367 | 358,3838 | 0,407804 | 1 | 0,72047 |
| -0,13212 | 280 | 270,6998 | 0,89499 | 1 | 0,975243 |
| 1,068969 | 421 | 378,3651 | 0,285765 | 1 | 0,583534 |
| 2,944786 | 421 | 201,2488 | 0,003612 | 0,465976 | 0,118336 |
| 2,680185 | 420 | 405,1516 | 0,007658 | 1 | 0,554149 |
| 1,455316 | 367 | 352,3125 | 0,146472 | 1 | 0,498729 |
| 0,45566 | 280 | 268,2637 | 0,649003 | 1 | 0,908644 |
| 0,39162 | 421 | 357,9396 | 0,695572 | 1 | 0,870621 |
| -0,02765 | 421 | 192,2457 | 0,977969 | 1 | 0,991789 |
| 0,043597 | 420 | 391,6265 | 0,965248 | 1 | 0,996924 |
| 0,053852 | 367 | 338,4467 | 0,957085 | 1 | 0,981063 |
| 0,232663 | 280 | 266,0413 | 0,816202 | 1 | 0,95747 |
| -0,07155 | 421 | 397,7491 | 0,942999 | 1 | 0,978719 |
| -0,5378 | 421 | 214,3812 | 0,591275 | 1 | 0,807111 |
| -0,85839 | 420 | 414,5491 | 0,391171 | 1 | 0,95743 |
| -1,20677 | 367 | 358,514 | 0,228318 | 1 | 0,577307 |
| 0,194122 | 280 | 270,0533 | 0,846226 | 1 | 0,965181 |
| 0,30548 | 421 | 295,5399 | 0,760215 | 1 | 0,901373 |
| 1,144227 | 421 | 177,4109 | 0,254071 | 1 | 0,553302 |
| -0,23978 | 420 | 326,0899 | 0,81065 | 1 | 0,969427 |
| -0,4948 | 367 | 287,6717 | 0,621117 | 1 | 0,875495 |
| 1,761658 | 280 | 276 | 0,079234 | 1 | 0,453386 |

| | | | | | |
|---|---|---|---|---|---|
| -1,20451 | 421 | 374,344 | 0,229152 | 1 | 0,520357 |
| -1,35284 | 421 | 199,511 | 0,177637 | 1 | 0,464755 |
| 0,009274 | 420 | 402,5761 | 0,992605 | 1 | 0,997342 |
| -0,43449 | 367 | 355,6837 | 0,664199 | 1 | 0,894765 |
| 0,636188 | 280 | 267,9003 | 0,525197 | 1 | 0,837536 |
| -0,31827 | 421 | 385,9218 | 0,750454 | 1 | 0,898125 |
| -2,25585 | 421 | 205,4097 | 0,025134 | 1 | 0,218351 |
| -3,99208 | 420 | 410,2187 | 7,76E-05 | 0,083572 | 0,038681 |
| -0,91651 | 367 | 355,5264 | 0,36002 | 1 | 0,674813 |
| -2,58608 | 280 | 266,7874 | 0,010238 | 1 | 0,229419 |
| -1,87254 | 421 | 327,7354 | 0,062022 | 1 | 0,271708 |
| -0,4573 | 421 | 183,1878 | 0,648001 | 1 | 0,839358 |
| -1,09308 | 420 | 359,6744 | 0,275091 | 1 | 0,943897 |
| 0,084636 | 367 | 308,2607 | 0,932606 | 1 | 0,977109 |
| -1,28397 | 280 | 256,5265 | 0,200312 | 1 | 0,631529 |
| 1,543822 | 421 | 400,8131 | 0,123421 | 1 | 0,385082 |
| 2,355496 | 421 | 215,4486 | 0,019396 | 1 | 0,195731 |
| 0,287633 | 420 | 415,5648 | 0,773771 | 1 | 0,969427 |
| 1,91375 | 367 | 361,9636 | 0,056441 | 1 | 0,337523 |
| 0,161652 | 280 | 273,6316 | 0,871699 | 1 | 0,969656 |
| -1,3774 | 421 | 323,651 | 0,16934 | 1 | 0,442288 |
| -0,26004 | 421 | 182,5619 | 0,79513 | 1 | 0,92103 |
| 0,638508 | 420 | 358,434 | 0,523551 | 1 | 0,960096 |
| -1,33784 | 367 | 320,0573 | 0,181899 | 1 | 0,527769 |
| -0,4647 | 280 | 246,7956 | 0,642558 | 1 | 0,907804 |
| 0,319559 | 421 | 405,4088 | 0,749468 | 1 | 0,898125 |
| 0,714218 | 421 | 221,1722 | 0,475846 | 1 | 0,734136 |
| -0,66889 | 420 | 415,9536 | 0,503938 | 1 | 0,960096 |
| -0,21341 | 367 | 362,348 | 0,831126 | 1 | 0,956843 |
| 0,123105 | 280 | 269,5017 | 0,902116 | 1 | 0,975243 |
| 1,641852 | 421 | 387,6569 | 0,101432 | 1 | 0,34273 |
| 2,006807 | 421 | 207,3597 | 0,046069 | 1 | 0,281076 |
| 0,42187 | 420 | 411,6427 | 0,67334 | 1 | 0,96781 |
| 2,807719 | 367 | 359,1298 | 0,005262 | 1 | 0,178183 |
| 1,108121 | 280 | 274,3846 | 0,26878 | 1 | 0,693794 |
| -2,64104 | 421 | 396,7151 | 0,008592 | 1 | 0,112587 |
| -2,32453 | 421 | 212,6916 | 0,02104 | 1 | 0,2022 |
| -3,31741 | 420 | 413,8376 | 9,89E-04 | 0,150303 | 0,235929 |
| -1,40836 | 367 | 361,2028 | 0,159886 | 1 | 0,504558 |
| -0,72824 | 280 | 269,0903 | 0,467102 | 1 | 0,824256 |
| 0,379169 | 421 | 341,2996 | 0,704798 | 1 | 0,875021 |
| 0,277827 | 421 | 187,0082 | 0,781452 | 1 | 0,91393 |
| 0,877742 | 420 | 376,2167 | 0,380644 | 1 | 0,95743 |
| 1,150634 | 367 | 323,617 | 0,250732 | 1 | 0,605249 |
| 0,039433 | 280 | 253,1467 | 0,968576 | 1 | 0,991502 |
| 0,718684 | 421 | 352,5149 | 0,472812 | 1 | 0,731548 |
| 0,932942 | 421 | 191,0964 | 0,352027 | 1 | 0,643758 |
| 2,284305 | 420 | 384,185 | 0,022898 | 1 | 0,75904 |

| | | | | | |
|---|---|---|---|---|---|
| 0,829703 | 367 | 339,1757 | 0,40729 | 1 | 0,720407 |
| 1,155274 | 280 | 260,1165 | 0,249038 | 1 | 0,677438 |
| -2,38988 | 421 | 358,5878 | 0,017369 | 1 | 0,153976 |
| -1,47733 | 421 | 191,6376 | 0,14123 | 1 | 0,423499 |
| -0,09809 | 420 | 393,6064 | 0,921908 | 1 | 0,989204 |
| -1,03469 | 367 | 334,076 | 0,301561 | 1 | 0,633826 |
| 0,643702 | 280 | 258,1095 | 0,52034 | 1 | 0,833721 |
| 1,877061 | 421 | 390,9727 | 0,061254 | 1 | 0,270704 |
| 2,063846 | 421 | 208,9483 | 0,040268 | 1 | 0,26998 |
| 0,085025 | 420 | 412,6984 | 0,932283 | 1 | 0,991559 |
| 2,384285 | 367 | 359,5889 | 0,01763 | 1 | 0,216225 |
| 1,468356 | 280 | 274,894 | 0,143151 | 1 | 0,554571 |
| 0,757829 | 421 | 365,1647 | 0,449042 | 1 | 0,714643 |
| 1,516282 | 421 | 192,4284 | 0,131089 | 1 | 0,413287 |
| 1,986494 | 420 | 395,3095 | 0,047669 | 1 | 0,846627 |
| 1,725976 | 367 | 334,6011 | 0,085275 | 1 | 0,388246 |
| -0,33234 | 280 | 254,4738 | 0,739903 | 1 | 0,940183 |
| 0,627153 | 421 | 347,6161 | 0,53097 | 1 | 0,776668 |
| 1,491998 | 421 | 186,534 | 0,137389 | 1 | 0,422541 |
| 1,856821 | 420 | 374,4558 | 0,064122 | 1 | 0,848246 |
| 0,167221 | 367 | 319,9663 | 0,867302 | 1 | 0,959636 |
| -0,41814 | 280 | 253,5674 | 0,676202 | 1 | 0,909385 |
| -0,44234 | 421 | 351,8812 | 0,658513 | 1 | 0,848606 |
| -0,75458 | 421 | 190,7904 | 0,451433 | 1 | 0,713861 |
| -1,0407 | 420 | 383,719 | 0,29867 | 1 | 0,943897 |
| -0,1625 | 367 | 339,3567 | 0,871011 | 1 | 0,959978 |
| -0,11114 | 280 | 269,5606 | 0,911588 | 1 | 0,976008 |
| 0,213922 | 421 | 384,1044 | 0,830721 | 1 | 0,936611 |
| -0,51374 | 421 | 204,7112 | 0,607988 | 1 | 0,821738 |
| -1,48869 | 420 | 408,9587 | 0,137341 | 1 | 0,891179 |
| -1,05354 | 367 | 355,2693 | 0,29281 | 1 | 0,627013 |
| 0,659423 | 280 | 267,5771 | 0,510191 | 1 | 0,831912 |
| 0,102664 | 421 | 349,9846 | 0,918288 | 1 | 0,966828 |
| 0,882204 | 421 | 187,7631 | 0,378794 | 1 | 0,656141 |
| -0,67794 | 420 | 382,8639 | 0,498221 | 1 | 0,960096 |
| 0,197227 | 367 | 339,562 | 0,843768 | 1 | 0,956935 |
| 1,441899 | 280 | 254,1342 | 0,150562 | 1 | 0,56302 |
| 3,028396 | 421 | 390,9804 | 0,002622 | 1 | 0,073944 |
| 4,253844 | 421 | 209,562 | 3,17E-05 | 0,034167 | 0,015907 |
| 1,144032 | 420 | 413,436 | 0,253272 | 1 | 0,943897 |
| 2,997789 | 367 | 359,8282 | 0,002908 | 1 | 0,176492 |
| 1,833521 | 280 | 275,6126 | 0,067803 | 1 | 0,425749 |
| -1,31621 | 421 | 336,5821 | 0,188999 | 1 | 0,472953 |
| -0,60372 | 421 | 184,1998 | 0,546772 | 1 | 0,776424 |
| -0,71867 | 420 | 368,414 | 0,472803 | 1 | 0,960096 |
| -0,63259 | 367 | 325,5314 | 0,527447 | 1 | 0,811085 |
| 0,804448 | 280 | 258,9554 | 0,421876 | 1 | 0,796701 |
| 0,524183 | 421 | 356,1993 | 0,600477 | 1 | 0,818191 |

| | | | | | |
|---|---|---|---|---|---|
| 0,211878 | 421 | 191,3281 | 0,832428 | 1 | 0,936588 |
| 0,365528 | 420 | 389,2739 | 0,714916 | 1 | 0,968866 |
| 0,320123 | 367 | 337,5396 | 0,749073 | 1 | 0,923669 |
| -0,27134 | 280 | 264,876 | 0,786338 | 1 | 0,953235 |
| 1,005292 | 421 | 357,1757 | 0,315437 | 1 | 0,60944 |
| 0,768197 | 421 | 192,3221 | 0,443312 | 1 | 0,708362 |
| 0,220208 | 420 | 390,317 | 0,825825 | 1 | 0,973894 |
| 0,774219 | 367 | 341,0887 | 0,439338 | 1 | 0,742744 |
| -0,44808 | 280 | 264,3438 | 0,654464 | 1 | 0,908644 |
| 0,248551 | 421 | 389,9447 | 0,803839 | 1 | 0,922616 |
| 1,060718 | 421 | 208,4461 | 0,290045 | 1 | 0,586961 |
| -1,6311 | 420 | 411,7016 | 0,103635 | 1 | 0,848246 |
| 1,138858 | 367 | 359,8762 | 0,25552 | 1 | 0,609149 |
| 0,711099 | 280 | 269,7281 | 0,477637 | 1 | 0,824256 |
| -1,51888 | 421 | 325,8689 | 0,129762 | 1 | 0,397564 |
| 0,045122 | 421 | 181,1358 | 0,964059 | 1 | 0,985228 |
| -1,23071 | 420 | 357,0648 | 0,219242 | 1 | 0,943055 |
| -0,04201 | 367 | 314,6606 | 0,96652 | 1 | 0,986276 |
| -0,0777 | 280 | 255,0582 | 0,938129 | 1 | 0,981778 |
| -0,16857 | 421 | 332,446 | 0,866237 | 1 | 0,95286 |
| 2,138968 | 421 | 185,286 | 0,033747 | 1 | 0,251769 |
| 0,337166 | 420 | 367,7441 | 0,736184 | 1 | 0,969427 |
| -0,20246 | 367 | 324,5857 | 0,839688 | 1 | 0,956935 |
| 1,916111 | 280 | 252,7381 | 0,056479 | 1 | 0,406703 |
| 1,743078 | 421 | 351,0605 | 0,082196 | 1 | 0,304997 |
| 2,05274 | 421 | 188,4177 | 0,04148 | 1 | 0,272972 |
| 1,381225 | 420 | 389,1861 | 0,168002 | 1 | 0,914005 |
| 1,210453 | 367 | 334,5755 | 0,226959 | 1 | 0,576775 |
| 2,997761 | 280 | 269,5935 | 0,002973 | 1 | 0,135787 |
| -3,1684 | 421 | 380,566 | 0,001657 | 0,253473 | 0,069718 |
| -1,52009 | 421 | 202,3843 | 0,130049 | 1 | 0,410868 |
| -0,85445 | 420 | 410,1934 | 0,393356 | 1 | 0,95743 |
| -0,61481 | 367 | 358,2799 | 0,539071 | 1 | 0,819758 |
| -0,41752 | 280 | 273,9461 | 0,676627 | 1 | 0,909385 |
| -0,0095 | 421 | 340,9034 | 0,992422 | 1 | 0,998054 |
| -0,08555 | 421 | 188,9444 | 0,931916 | 1 | 0,974599 |
| 1,003428 | 420 | 374,2851 | 0,316302 | 1 | 0,943897 |
| 0,26242 | 367 | 326,1826 | 0,793163 | 1 | 0,944997 |
| 0,628677 | 280 | 253,2086 | 0,530127 | 1 | 0,840949 |
| -1,16376 | 421 | 349,9736 | 0,245315 | 1 | 0,542863 |
| -1,30041 | 421 | 190,4479 | 0,195032 | 1 | 0,483337 |
| -1,31167 | 420 | 385,2089 | 0,190413 | 1 | 0,925729 |
| 0,576152 | 367 | 330,2263 | 0,564905 | 1 | 0,837838 |
| 0,743493 | 280 | 260,5862 | 0,457853 | 1 | 0,817005 |
| 0,151405 | 421 | 396,2405 | 0,879733 | 1 | 0,95861 |
| 1,000152 | 421 | 211,1251 | 0,318382 | 1 | 0,617081 |
| 1,22988 | 420 | 413,3089 | 0,219442 | 1 | 0,943055 |
| 0,558224 | 367 | 362,1551 | 0,577036 | 1 | 0,843313 |

| | | | | | |
|---|---|---|---|---|---|
| -0,81354 | 280 | 273,6199 | 0,416613 | 1 | 0,792723 |
| -0,00599 | 421 | 312,9761 | 0,995224 | 1 | 0,998054 |
| -0,69995 | 421 | 181,0641 | 0,484859 | 1 | 0,740307 |
| -1,98144 | 420 | 346,5524 | 0,048332 | 1 | 0,846627 |
| -0,40261 | 367 | 296,9141 | 0,687525 | 1 | 0,903032 |
| -1,66723 | 280 | 254,4096 | 0,096698 | 1 | 0,480557 |
| 0,821506 | 421 | 360,4764 | 0,411901 | 1 | 0,695339 |
| 0,489487 | 421 | 192,975 | 0,625053 | 1 | 0,831381 |
| 2,297159 | 420 | 390,4707 | 0,022138 | 1 | 0,75904 |
| -0,2961 | 367 | 344,6023 | 0,767329 | 1 | 0,93557 |
| 0,564152 | 280 | 270,5633 | 0,573118 | 1 | 0,870079 |
| -0,95348 | 421 | 310,5382 | 0,341087 | 1 | 0,63164 |
| -0,66551 | 421 | 180,9834 | 0,506572 | 1 | 0,754351 |
| -0,83913 | 420 | 343,8698 | 0,401981 | 1 | 0,95743 |
| 0,419038 | 367 | 301,0386 | 0,675487 | 1 | 0,898834 |
| -0,84014 | 280 | 276 | 0,401556 | 1 | 0,784757 |
| -1,4939 | 421 | 346,0635 | 0,136112 | 1 | 0,407576 |
| -1,08175 | 421 | 188,6381 | 0,280743 | 1 | 0,581153 |
| -1,80426 | 420 | 377,7193 | 0,071986 | 1 | 0,848246 |
| -0,62257 | 367 | 330,2062 | 0,533998 | 1 | 0,816122 |
| -0,52967 | 280 | 254,9807 | 0,596798 | 1 | 0,880334 |
| -0,22618 | 421 | 310,0929 | 0,821208 | 1 | 0,932601 |
| 0,737725 | 421 | 179,3271 | 0,461646 | 1 | 0,721079 |
| -0,42066 | 420 | 343,0103 | 0,674269 | 1 | 0,96781 |
| -0,46402 | 367 | 307,1363 | 0,642962 | 1 | 0,882539 |
| 0,668412 | 280 | 254,7067 | 0,504476 | 1 | 0,831912 |
| -1,04661 | 421 | 405,2993 | 0,295902 | 1 | 0,593774 |
| -0,6681 | 421 | 221,6749 | 0,504762 | 1 | 0,753145 |
| -0,69918 | 420 | 415,9228 | 0,484831 | 1 | 0,960096 |
| -2,06151 | 367 | 362,9919 | 0,039966 | 1 | 0,28956 |
| 0,540683 | 280 | 275,5251 | 0,589163 | 1 | 0,87561 |
| 3,54586 | 421 | 383,6023 | 4,40E-04 | 0,058922 | 0,04642 |
| 1,799562 | 421 | 204,639 | 0,073403 | 1 | 0,342815 |
| -0,84983 | 420 | 410,151 | 0,395917 | 1 | 0,95743 |
| 2,437019 | 367 | 357,8285 | 0,015296 | 1 | 0,204372 |
| 1,872165 | 280 | 274,8617 | 0,062246 | 1 | 0,422605 |
| 1,220187 | 421 | 380,6 | 0,22315 | 1 | 0,51676 |
| 1,062562 | 421 | 200,8663 | 0,289256 | 1 | 0,586688 |
| -0,35796 | 420 | 406,398 | 0,720559 | 1 | 0,969427 |
| -0,39577 | 367 | 350,2247 | 0,692516 | 1 | 0,904352 |
| 0,816737 | 280 | 263,4705 | 0,414817 | 1 | 0,791302 |
| -1,29701 | 421 | 299,4393 | 0,195627 | 1 | 0,481951 |
| -0,23121 | 421 | 175,4113 | 0,81742 | 1 | 0,926377 |
| -1,06121 | 420 | 328,7896 | 0,289373 | 1 | 0,943897 |
| 0,224505 | 367 | 288,0811 | 0,822524 | 1 | 0,953906 |
| 1,107317 | 280 | 262,3382 | 0,269171 | 1 | 0,693794 |
| -2,86861 | 421 | 324,0245 | 0,004393 | 1 | 0,087964 |
| -1,54453 | 421 | 183,5082 | 0,124182 | 1 | 0,408607 |

| | | | | | |
|---|---|---|---|---|---|
| -0,847 | 420 | 356,8136 | 0,397561 | 1 | 0,95743 |
| -0,07928 | 367 | 321,8976 | 0,936856 | 1 | 0,978845 |
| -0,02102 | 280 | 256,6897 | 0,983248 | 1 | 0,99236 |
| -1,13229 | 421 | 307,9593 | 0,258395 | 1 | 0,550898 |
| 0,069627 | 421 | 180,2539 | 0,944568 | 1 | 0,980711 |
| -1,20623 | 420 | 336,716 | 0,228576 | 1 | 0,943055 |
| 1,253844 | 367 | 297,4254 | 0,210883 | 1 | 0,563436 |
| -0,43534 | 280 | 251,898 | 0,663687 | 1 | 0,908644 |
| -0,97762 | 421 | 305,7499 | 0,329037 | 1 | 0,618807 |
| 0,069605 | 421 | 174,9801 | 0,944588 | 1 | 0,980711 |
| -0,47844 | 420 | 336,9751 | 0,632644 | 1 | 0,964878 |
| 0,314814 | 367 | 304,3636 | 0,753119 | 1 | 0,924494 |
| 0,926408 | 280 | 255,4391 | 0,355108 | 1 | 0,761846 |
| 0,813793 | 421 | 409,3638 | 0,416236 | 1 | 0,695339 |
| 0,185715 | 421 | 226,4821 | 0,852834 | 1 | 0,944105 |
| 0,431789 | 420 | 415,7797 | 0,666118 | 1 | 0,96781 |
| -1,26105 | 367 | 362,8911 | 0,2081 | 1 | 0,561015 |
| 1,139435 | 280 | 272,8319 | 0,255521 | 1 | 0,679673 |
| -0,04746 | 421 | 392,2278 | 0,962167 | 1 | 0,991104 |
| 1,591034 | 421 | 209,4748 | 0,11311 | 1 | 0,391524 |
| 0,021671 | 420 | 412,3311 | 0,982721 | 1 | 0,997342 |
| -0,19579 | 367 | 359,2615 | 0,844887 | 1 | 0,956935 |
| 0,924688 | 280 | 272,052 | 0,355948 | 1 | 0,761846 |
| -1,39096 | 421 | 297,9361 | 0,165276 | 1 | 0,440727 |
| -1,18676 | 421 | 175,0805 | 0,236931 | 1 | 0,537903 |
| -2,32827 | 420 | 326,8464 | 0,020507 | 1 | 0,75904 |
| -1,47677 | 367 | 275,0715 | 0,140882 | 1 | 0,48919 |
| -1,0451 | 280 | 276 | 0,296892 | 1 | 0,723324 |
| 3,509408 | 421 | 395,9528 | 5,01E-04 | 0,066094 | 0,04642 |
| 3,49346 | 421 | 212,7027 | 5,80E-04 | 0,077134 | 0,051411 |
| 0,521372 | 420 | 414,6819 | 0,602386 | 1 | 0,964878 |
| 2,839781 | 367 | 359,9863 | 0,004771 | 0,629756 | 0,178183 |
| 0,879478 | 280 | 274,7973 | 0,379911 | 1 | 0,770948 |
| 0,195054 | 421 | 372,6211 | 0,845457 | 1 | 0,939735 |
| 0,939294 | 421 | 197,8084 | 0,348725 | 1 | 0,643243 |
| 0,228456 | 420 | 401,1166 | 0,819408 | 1 | 0,97003 |
| -1,03536 | 367 | 352,6744 | 0,301212 | 1 | 0,633826 |
| -1,68627 | 280 | 268,7947 | 0,092904 | 1 | 0,471402 |
| -0,05409 | 421 | 379,8092 | 0,956889 | 1 | 0,987258 |
| -0,81287 | 421 | 203,1417 | 0,417247 | 1 | 0,687955 |
| -0,83857 | 420 | 406,889 | 0,402201 | 1 | 0,95743 |
| -0,26035 | 367 | 348,7312 | 0,79475 | 1 | 0,945294 |
| 0,612844 | 280 | 263,598 | 0,540508 | 1 | 0,84937 |
| -0,66459 | 421 | 375,0804 | 0,506721 | 1 | 0,75983 |
| -1,95827 | 421 | 201,2586 | 0,051581 | 1 | 0,296106 |
| -2,33192 | 420 | 402,3088 | 0,020198 | 1 | 0,75904 |
| -1,5185 | 367 | 347,3267 | 0,129798 | 1 | 0,473621 |
| -0,12572 | 280 | 259,6033 | 0,900054 | 1 | 0,975243 |

| | | | | | |
|---|---|---|---|---|---|
| 0,245494 | 421 | 345,0306 | 0,80622 | 1 | 0,922616 |
| 1,592796 | 421 | 189,4635 | 0,112873 | 1 | 0,391524 |
| 1,065449 | 420 | 376,6948 | 0,287355 | 1 | 0,943897 |
| -0,47899 | 367 | 327,9068 | 0,632262 | 1 | 0,880707 |
| -0,95995 | 280 | 252,4485 | 0,337998 | 1 | 0,755731 |
| 0,008714 | 421 | 379,4923 | 0,993052 | 1 | 0,998054 |
| -2,27251 | 421 | 201,0573 | 0,024114 | 1 | 0,218351 |
| 0,349547 | 420 | 405,8121 | 0,72686 | 1 | 0,969427 |
| -0,65 | 367 | 340,7467 | 0,516131 | 1 | 0,800216 |
| -0,23304 | 280 | 265,9341 | 0,81591 | 1 | 0,95747 |
| 2,885226 | 421 | 399,5137 | 0,004123 | 0,614373 | 0,087964 |
| 2,024083 | 421 | 217,1692 | 0,044186 | 1 | 0,277922 |
| 0,750363 | 420 | 415,3932 | 0,453461 | 1 | 0,960096 |
| 1,84204 | 367 | 362,8789 | 0,066285 | 1 | 0,359322 |
| 0,985549 | 280 | 275,6088 | 0,325219 | 1 | 0,749395 |
| 0,830989 | 421 | 305,9147 | 0,406628 | 1 | 0,695339 |
| 4,030909 | 421 | 175,7127 | 8,27E-05 | 0,088941 | 0,020761 |
| 1,173375 | 420 | 339,7661 | 0,241467 | 1 | 0,943659 |
| 2,382694 | 367 | 308,6664 | 0,017792 | 1 | 0,216225 |
| 1,987623 | 280 | 246,7465 | 0,047959 | 1 | 0,393265 |
| -1,89644 | 421 | 336,2814 | 0,058759 | 1 | 0,268334 |
| -0,98798 | 421 | 183,6667 | 0,324464 | 1 | 0,619085 |
| -1,61292 | 420 | 367,7599 | 0,10762 | 1 | 0,848246 |
| 0,499582 | 367 | 319,3752 | 0,617714 | 1 | 0,875495 |
| 0,269001 | 280 | 259,961 | 0,788142 | 1 | 0,953235 |
| 1,31547 | 421 | 337,9061 | 0,189244 | 1 | 0,472953 |
| 0,394785 | 421 | 186,8545 | 0,693452 | 1 | 0,865526 |
| -0,76793 | 420 | 373,8101 | 0,443015 | 1 | 0,960096 |
| 0,938369 | 367 | 331,0257 | 0,348739 | 1 | 0,668671 |
| -0,91331 | 280 | 254,471 | 0,361942 | 1 | 0,762031 |
| -2,52002 | 421 | 307,8054 | 0,012241 | 1 | 0,131857 |
| -1,97596 | 421 | 178,1669 | 0,049703 | 1 | 0,289197 |
| -1,09499 | 420 | 348,753 | 0,274279 | 1 | 0,943897 |
| -1,79476 | 367 | 317,6477 | 0,073642 | 1 | 0,373795 |
| -1,62592 | 280 | 271,2856 | 0,105128 | 1 | 0,492014 |
| 2,566926 | 421 | 363,041 | 0,01066 | 1 | 0,12358 |
| 3,154432 | 421 | 192,4239 | 0,001866 | 0,279939 | 0,096981 |
| 3,647284 | 420 | 403,3152 | 3,00E-04 | 0,045865 | 0,09035 |
| 1,879057 | 367 | 347,8042 | 0,061073 | 1 | 0,34342 |
| 3,186029 | 280 | 270,5652 | 0,001611 | 0,246546 | 0,135787 |
| -0,60608 | 421 | 357,2729 | 0,544849 | 1 | 0,784719 |
| 0,175784 | 421 | 194,3411 | 0,860647 | 1 | 0,947215 |
| 0,40398 | 420 | 389,2325 | 0,686449 | 1 | 0,968613 |
| -0,51547 | 367 | 340,349 | 0,606559 | 1 | 0,868127 |
| -1,10698 | 280 | 261,0312 | 0,269323 | 1 | 0,693794 |
| 1,47846 | 421 | 343,2893 | 0,140202 | 1 | 0,410878 |
| 1,376929 | 421 | 188,7815 | 0,170165 | 1 | 0,456365 |
| 1,602655 | 420 | 372,7658 | 0,109858 | 1 | 0,848246 |

| | | | | | |
|---|---|---|---|---|---|
| 0,484678 | 367 | 331,4785 | 0,628225 | 1 | 0,880707 |
| -0,64671 | 280 | 261,7128 | 0,518387 | 1 | 0,833721 |
| 0,536741 | 421 | 362,5745 | 0,591776 | 1 | 0,81147 |
| 1,418365 | 421 | 194,7665 | 0,157682 | 1 | 0,43893 |
| -0,48133 | 420 | 394,3769 | 0,630548 | 1 | 0,964878 |
| 0,164686 | 367 | 337,1633 | 0,86929 | 1 | 0,959978 |
| -0,03374 | 280 | 267,5907 | 0,973111 | 1 | 0,99236 |
| -2,66741 | 421 | 362,252 | 0,007987 | 1 | 0,110425 |
| -1,68347 | 421 | 195,7724 | 0,093878 | 1 | 0,373113 |
| -1,20414 | 420 | 392,8096 | 0,22926 | 1 | 0,943055 |
| -0,70454 | 367 | 346,9449 | 0,481568 | 1 | 0,785415 |
| -1,07998 | 280 | 267,2926 | 0,281126 | 1 | 0,703928 |
| -0,86919 | 421 | 318,027 | 0,385396 | 1 | 0,674555 |
| -0,9184 | 421 | 180,8952 | 0,359635 | 1 | 0,643758 |
| -1,73335 | 420 | 351,5893 | 0,08391 | 1 | 0,848246 |
| -1,00679 | 367 | 300,5845 | 0,314847 | 1 | 0,647568 |
| -1,35591 | 280 | 256,6522 | 0,176319 | 1 | 0,598625 |
| -1,45768 | 421 | 340,6136 | 0,145851 | 1 | 0,414711 |
| -1,57954 | 421 | 187,3225 | 0,115899 | 1 | 0,392493 |
| 0,00812 | 420 | 374,6698 | 0,993526 | 1 | 0,997342 |
| -0,92446 | 367 | 331,9262 | 0,355919 | 1 | 0,674813 |
| -2,19339 | 280 | 253,4477 | 0,029188 | 1 | 0,326641 |
| 2,219006 | 421 | 329,3339 | 0,027168 | 1 | 0,192509 |
| 2,352028 | 421 | 181,7229 | 0,019742 | 1 | 0,195731 |
| 1,709059 | 420 | 370,755 | 0,088277 | 1 | 0,848246 |
| 1,821036 | 367 | 308,6597 | 0,069569 | 1 | 0,364488 |
| 2,4121 | 280 | 262,6387 | 0,016547 | 1 | 0,266517 |
| -2,72628 | 421 | 370,6193 | 0,006709 | 1 | 0,10482 |
| -2,40439 | 421 | 201,1242 | 0,017106 | 1 | 0,181988 |
| -2,18507 | 420 | 405,0367 | 0,029456 | 1 | 0,792693 |
| -1,75628 | 367 | 348,52 | 0,079919 | 1 | 0,381841 |
| -1,08369 | 280 | 270,3853 | 0,279466 | 1 | 0,703716 |
| 0,979748 | 421 | 407,3453 | 0,327792 | 1 | 0,618251 |
| 3,731326 | 421 | 222,9463 | 2,42E-04 | 0,036743 | 0,035752 |
| 0,508068 | 420 | 415,9424 | 0,611675 | 1 | 0,964878 |
| 2,114511 | 367 | 362,9999 | 0,035153 | 1 | 0,283291 |
| 1,722044 | 280 | 275,9961 | 0,086182 | 1 | 0,458713 |
| 0,332315 | 421 | 324,8042 | 0,739866 | 1 | 0,893273 |
| 0,915802 | 421 | 178,807 | 0,361004 | 1 | 0,64452 |
| -0,04128 | 420 | 358,9438 | 0,967099 | 1 | 0,996924 |
| 0,459044 | 367 | 307,3436 | 0,646527 | 1 | 0,884339 |
| 1,955964 | 280 | 244,8488 | 0,051607 | 1 | 0,398826 |
| -1,88187 | 421 | 337,9586 | 0,060713 | 1 | 0,270611 |
| -0,99833 | 421 | 186,5324 | 0,319411 | 1 | 0,617081 |
| -0,52376 | 420 | 369,9299 | 0,600757 | 1 | 0,964878 |
| 0,006645 | 367 | 333,2143 | 0,994702 | 1 | 0,997335 |
| -1,96089 | 280 | 256,959 | 0,050972 | 1 | 0,398004 |
| 2,487107 | 421 | 406,4633 | 0,013279 | 1 | 0,138362 |

| | | | | | |
|---|---|---|---|---|---|
| 2,64343 | 421 | 223,5071 | 0,008788 | 1 | 0,14815 |
| 1,485364 | 420 | 415,9745 | 0,138205 | 1 | 0,891179 |
| 2,536243 | 367 | 362,4622 | 0,011624 | 1 | 0,189546 |
| 1,148231 | 280 | 275,9216 | 0,251868 | 1 | 0,679007 |
| -1,42061 | 421 | 333,5531 | 0,156364 | 1 | 0,429212 |
| -0,63443 | 421 | 183,5899 | 0,526587 | 1 | 0,767321 |
| -0,5593 | 420 | 366,7885 | 0,5763 | 1 | 0,964878 |
| -1,35591 | 367 | 323,277 | 0,176073 | 1 | 0,523018 |
| 0,762081 | 280 | 246,2202 | 0,446741 | 1 | 0,81309 |
| -0,10083 | 421 | 396,7702 | 0,919739 | 1 | 0,966828 |
| 1,2728 | 421 | 209,5537 | 0,204499 | 1 | 0,500293 |
| 0,540207 | 420 | 413,9528 | 0,589344 | 1 | 0,964878 |
| 0,917737 | 367 | 361,8776 | 0,359368 | 1 | 0,674813 |
| 0,748753 | 280 | 270,2197 | 0,454657 | 1 | 0,814707 |
| -2,12703 | 421 | 351,9591 | 0,034113 | 1 | 0,212429 |
| -2,21228 | 421 | 191,0521 | 0,028132 | 1 | 0,227932 |
| -0,5633 | 420 | 389,74 | 0,573555 | 1 | 0,964878 |
| -0,89118 | 367 | 333,7622 | 0,373472 | 1 | 0,683037 |
| -0,28805 | 280 | 262,2157 | 0,773539 | 1 | 0,947219 |
| -1,1833 | 421 | 341,1559 | 0,237513 | 1 | 0,532577 |
| -0,00819 | 421 | 186,4307 | 0,993474 | 1 | 0,997935 |
| 0,308363 | 420 | 378,1432 | 0,757976 | 1 | 0,969427 |
| 0,462637 | 367 | 315,1921 | 0,643944 | 1 | 0,882712 |
| 1,050352 | 280 | 263,9401 | 0,294517 | 1 | 0,723324 |
| 0,882459 | 421 | 314,049 | 0,378204 | 1 | 0,670163 |
| 1,307266 | 421 | 181,318 | 0,192778 | 1 | 0,480987 |
| -0,70855 | 420 | 350,1026 | 0,479076 | 1 | 0,960096 |
| -0,85849 | 367 | 303,0081 | 0,391298 | 1 | 0,705088 |
| -0,27828 | 280 | 259,2493 | 0,781021 | 1 | 0,951006 |
| 0,813855 | 421 | 361,0768 | 0,416264 | 1 | 0,695339 |
| 2,4517 | 421 | 195,7548 | 0,015095 | 1 | 0,179127 |
| 1,110631 | 420 | 394,865 | 0,267403 | 1 | 0,943897 |
| -0,3195 | 367 | 345,8988 | 0,749537 | 1 | 0,923669 |
| 0,88647 | 280 | 258,5875 | 0,376188 | 1 | 0,767473 |
| -2,16321 | 421 | 295,5778 | 0,031326 | 1 | 0,201145 |
| -0,50759 | 421 | 176,4738 | 0,612376 | 1 | 0,823974 |
| -1,51229 | 420 | 331,4502 | 0,131414 | 1 | 0,888479 |
| 0,728444 | 367 | 272,4924 | 0,466967 | 1 | 0,771623 |
| -0,72465 | 280 | 276 | 0,469281 | 1 | 0,824256 |
| -1,1342 | 421 | 379,7111 | 0,257427 | 1 | 0,550898 |
| 0,477931 | 421 | 203,3197 | 0,633213 | 1 | 0,835702 |
| -0,64927 | 420 | 406,5461 | 0,516528 | 1 | 0,960096 |
| 0,750795 | 367 | 348,622 | 0,453282 | 1 | 0,758154 |
| 0,219241 | 280 | 270,1373 | 0,826628 | 1 | 0,96121 |
| -1,10176 | 421 | 394,1102 | 0,271238 | 1 | 0,56693 |
| -0,48805 | 421 | 212,2084 | 0,626015 | 1 | 0,831927 |
| 0,212218 | 420 | 413,8968 | 0,832041 | 1 | 0,973894 |
| -1,43969 | 367 | 362,7802 | 0,150818 | 1 | 0,499998 |

| | | | | | |
|---|---|---|---|---|---|
| -0,42429 | 280 | 274,9512 | 0,671685 | 1 | 0,908644 |
| 0,882815 | 421 | 393,7944 | 0,377875 | 1 | 0,670163 |
| -0,54394 | 421 | 211,4274 | 0,587059 | 1 | 0,802811 |
| -1,11193 | 420 | 413,5817 | 0,266813 | 1 | 0,943897 |
| -0,13521 | 367 | 358,173 | 0,892522 | 1 | 0,969042 |
| -1,54396 | 280 | 274,2203 | 0,12375 | 1 | 0,535729 |
| -1,39423 | 421 | 326,2192 | 0,164196 | 1 | 0,438729 |
| -2,091 | 421 | 181,8849 | 0,037918 | 1 | 0,267984 |
| -1,73405 | 420 | 361,7979 | 0,08376 | 1 | 0,848246 |
| -1,7426 | 367 | 324,4584 | 0,082351 | 1 | 0,381856 |
| -1,20378 | 280 | 262,2113 | 0,229759 | 1 | 0,657169 |
| -0,61746 | 421 | 338,747 | 0,537346 | 1 | 0,778195 |
| -2,01046 | 421 | 186,5478 | 0,045823 | 1 | 0,281076 |
| 0,164716 | 420 | 378,088 | 0,869256 | 1 | 0,980994 |
| -0,22239 | 367 | 334,0429 | 0,824144 | 1 | 0,953906 |
| -0,14637 | 280 | 260,352 | 0,883744 | 1 | 0,973784 |
| -3,10603 | 421 | 328,9033 | 0,002061 | 1 | 0,069718 |
| -1,13582 | 421 | 186,1122 | 0,257494 | 1 | 0,556025 |
| 0,913789 | 420 | 371,1248 | 0,361421 | 1 | 0,955547 |
| 0,186114 | 367 | 336,6115 | 0,852467 | 1 | 0,956935 |
| -0,31075 | 280 | 265,9721 | 0,756232 | 1 | 0,942071 |
| -0,00434 | 421 | 321,9759 | 0,996538 | 1 | 0,998526 |
| -0,1662 | 421 | 183,4552 | 0,868185 | 1 | 0,947676 |
| 0,1382 | 420 | 357,1451 | 0,89016 | 1 | 0,983483 |
| 0,90539 | 367 | 318,903 | 0,365942 | 1 | 0,678321 |
| -0,32746 | 280 | 252,2478 | 0,743591 | 1 | 0,941674 |
| -1,96735 | 421 | 364,0957 | 0,049902 | 1 | 0,246877 |
| -1,68839 | 421 | 197,0104 | 0,092919 | 1 | 0,373113 |
| 0,616596 | 420 | 398,7091 | 0,537853 | 1 | 0,960096 |
| -0,78822 | 367 | 350,6341 | 0,431102 | 1 | 0,73975 |
| 0,994126 | 280 | 269,6008 | 0,321053 | 1 | 0,744349 |
| 2,066749 | 421 | 403,4324 | 0,039395 | 1 | 0,224033 |
| 2,214349 | 421 | 217,3464 | 0,027844 | 1 | 0,226812 |
| 0,339286 | 420 | 415,7851 | 0,734565 | 1 | 0,969427 |
| -0,43177 | 367 | 361,7941 | 0,666163 | 1 | 0,894765 |
| 1,105078 | 280 | 275,4601 | 0,27009 | 1 | 0,694583 |
| -1,27656 | 421 | 376,1125 | 0,202545 | 1 | 0,491523 |
| -0,00475 | 421 | 201,0136 | 0,996212 | 1 | 0,998199 |
| 0,316271 | 420 | 402,8773 | 0,751961 | 1 | 0,969427 |
| -1,07425 | 367 | 355,6843 | 0,283438 | 1 | 0,61815 |
| -0,31191 | 280 | 263,6728 | 0,755357 | 1 | 0,942071 |
| 0,210908 | 421 | 282,7875 | 0,833111 | 1 | 0,936611 |
| 1,117066 | 421 | 174,3888 | 0,265503 | 1 | 0,562747 |
| -0,3982 | 420 | 311,6847 | 0,690758 | 1 | 0,968866 |
| 2,197866 | 367 | 283,9018 | 0,028766 | 1 | 0,258038 |
| 0,317049 | 280 | 252,441 | 0,751468 | 1 | 0,942071 |
| -1,22112 | 421 | 324,6545 | 0,222925 | 1 | 0,51676 |
| -1,20636 | 421 | 181,1243 | 0,229252 | 1 | 0,529303 |

| | | | | | |
|---|---|---|---|---|---|
| 1,018992 | 420 | 359,0568 | 0,308893 | 1 | 0,943897 |
| -1,77714 | 367 | 321,3469 | 0,076491 | 1 | 0,378716 |
| -1,61282 | 280 | 249,5713 | 0,108048 | 1 | 0,497151 |
| 0,995521 | 421 | 308,5914 | 0,320263 | 1 | 0,615134 |
| 0,459521 | 421 | 178,8293 | 0,646418 | 1 | 0,839358 |
| -0,36196 | 420 | 339,3613 | 0,717608 | 1 | 0,969427 |
| -0,32195 | 367 | 295,6209 | 0,74772 | 1 | 0,923669 |
| -0,23912 | 280 | 247,4281 | 0,811212 | 1 | 0,95747 |
| -0,75593 | 421 | 380,6889 | 0,450161 | 1 | 0,714643 |
| -1,06862 | 421 | 202,9021 | 0,28651 | 1 | 0,584263 |
| -0,06644 | 420 | 406,7078 | 0,947059 | 1 | 0,992811 |
| -1,41354 | 367 | 350,8966 | 0,158384 | 1 | 0,504558 |
| 0,190381 | 280 | 270,8356 | 0,849153 | 1 | 0,966155 |
| -0,61374 | 421 | 302,7342 | 0,539845 | 1 | 0,779259 |
| 0,721547 | 421 | 178,6353 | 0,471516 | 1 | 0,731044 |
| 1,339159 | 420 | 338,1481 | 0,181418 | 1 | 0,923408 |
| -0,87806 | 367 | 297,5303 | 0,380619 | 1 | 0,691076 |
| -0,54244 | 280 | 246,7019 | 0,588007 | 1 | 0,87561 |
| -0,10942 | 421 | 310,8023 | 0,912939 | 1 | 0,966828 |
| 0,925821 | 421 | 180,5162 | 0,355775 | 1 | 0,643758 |
| 0,627229 | 420 | 344,5587 | 0,530924 | 1 | 0,960096 |
| 1,748707 | 367 | 314,6422 | 0,081317 | 1 | 0,381841 |
| 0,604762 | 280 | 248,0912 | 0,54589 | 1 | 0,851507 |
| -0,46795 | 421 | 377,0561 | 0,640092 | 1 | 0,834842 |
| -0,4697 | 421 | 198,9307 | 0,639082 | 1 | 0,838432 |
| 2,30146 | 420 | 401,8864 | 0,021876 | 1 | 0,75904 |
| 0,420289 | 367 | 350,1798 | 0,674532 | 1 | 0,898779 |
| 0,881645 | 280 | 268,8894 | 0,378756 | 1 | 0,770948 |
| 0,098837 | 421 | 305,5179 | 0,921333 | 1 | 0,966828 |
| -0,0545 | 421 | 179,2872 | 0,956601 | 1 | 0,98414 |
| 0,542465 | 420 | 337,3918 | 0,587857 | 1 | 0,964878 |
| 1,858845 | 367 | 301,3004 | 0,064024 | 1 | 0,350852 |
| -0,25636 | 280 | 276 | 0,797863 | 1 | 0,954269 |
| 1,042232 | 421 | 382,6393 | 0,297962 | 1 | 0,595529 |
| 0,777071 | 421 | 201,3922 | 0,438029 | 1 | 0,704743 |
| 1,241535 | 420 | 407,5687 | 0,215122 | 1 | 0,943055 |
| 2,150203 | 367 | 352,634 | 0,032219 | 1 | 0,270873 |
| 0,376815 | 280 | 263,1696 | 0,706614 | 1 | 0,922762 |
| -1,03477 | 421 | 334,2196 | 0,301525 | 1 | 0,597107 |
| -0,25332 | 421 | 186,1856 | 0,800297 | 1 | 0,921861 |
| 0,32816 | 420 | 368,4632 | 0,742977 | 1 | 0,969427 |
| 0,194247 | 367 | 328,1625 | 0,846103 | 1 | 0,956935 |
| -1,2824 | 280 | 250,3066 | 0,200888 | 1 | 0,631761 |
| 1,473508 | 421 | 289,4359 | 0,141701 | 1 | 0,412154 |
| 1,087946 | 421 | 175,0608 | 0,278114 | 1 | 0,578094 |
| 1,892998 | 420 | 318,7735 | 0,059264 | 1 | 0,848246 |
| 0,654758 | 367 | 293,7578 | 0,513136 | 1 | 0,797829 |
| 0,340865 | 280 | 252,8739 | 0,733488 | 1 | 0,936751 |

| | | | | | |
|---|---|---|---|---|---|
| -1,19027 | 421 | 292,8686 | 0,234905 | 1 | 0,527573 |
| 0,452621 | 421 | 175,5733 | 0,65138 | 1 | 0,839718 |
| -2,88974 | 420 | 314,9845 | 0,004123 | 1 | 0,443819 |
| -0,9368 | 367 | 289,1259 | 0,349644 | 1 | 0,668671 |
| 1,163029 | 280 | 247,2901 | 0,245939 | 1 | 0,676332 |
| -2,42003 | 421 | 378,1189 | 0,01599 | 1 | 0,147897 |
| -1,16327 | 421 | 199,7098 | 0,246109 | 1 | 0,546224 |
| -1,0741 | 420 | 402,102 | 0,28342 | 1 | 0,943897 |
| 1,403579 | 367 | 358,9895 | 0,161309 | 1 | 0,5075 |
| 0,910538 | 280 | 264,0638 | 0,363369 | 1 | 0,762031 |
| 0,798442 | 421 | 360,396 | 0,42514 | 1 | 0,6997 |
| 0,113653 | 421 | 194,4798 | 0,90963 | 1 | 0,96646 |
| 0,550056 | 420 | 392,2053 | 0,582594 | 1 | 0,964878 |
| -0,08915 | 367 | 342,5975 | 0,929011 | 1 | 0,976733 |
| -1,14979 | 280 | 259,5988 | 0,251289 | 1 | 0,679007 |
| 1,466671 | 421 | 330,5182 | 0,143416 | 1 | 0,412154 |
| 1,310765 | 421 | 185,5137 | 0,191558 | 1 | 0,47953 |
| 0,628765 | 420 | 371,2824 | 0,529889 | 1 | 0,960096 |
| 0,471171 | 367 | 320,2125 | 0,63784 | 1 | 0,880707 |
| 1,869089 | 280 | 252,6494 | 0,062767 | 1 | 0,422605 |
| -0,87012 | 421 | 388,5162 | 0,384773 | 1 | 0,674247 |
| -2,0973 | 421 | 207,0535 | 0,037181 | 1 | 0,266419 |
| 0,019732 | 420 | 411,0897 | 0,984267 | 1 | 0,997342 |
| -1,47411 | 367 | 359,102 | 0,141329 | 1 | 0,489614 |
| -1,6132 | 280 | 267,1303 | 0,107883 | 1 | 0,497151 |
| -1,53059 | 421 | 352,3386 | 0,126768 | 1 | 0,392431 |
| -0,05301 | 421 | 192,1116 | 0,957783 | 1 | 0,98414 |
| -0,20153 | 420 | 386,0007 | 0,840389 | 1 | 0,973894 |
| -0,43213 | 367 | 336,2254 | 0,665921 | 1 | 0,894765 |
| -0,43404 | 280 | 266,5869 | 0,664612 | 1 | 0,908644 |
| 0,05594 | 421 | 356,3778 | 0,955421 | 1 | 0,987258 |
| 0,323102 | 421 | 192,0701 | 0,74697 | 1 | 0,889215 |
| 1,743096 | 420 | 388,7691 | 0,082107 | 1 | 0,848246 |
| -0,65512 | 367 | 335,6835 | 0,51284 | 1 | 0,797829 |
| 0,183905 | 280 | 265,9766 | 0,854228 | 1 | 0,966655 |
| -2,3915 | 421 | 307,0811 | 0,017381 | 1 | 0,153976 |
| -0,64165 | 421 | 175,5478 | 0,521938 | 1 | 0,765001 |
| -1,77124 | 420 | 337,1581 | 0,077424 | 1 | 0,848246 |
| -1,07561 | 367 | 301,7824 | 0,282963 | 1 | 0,618008 |
| -0,32028 | 280 | 248,6943 | 0,749023 | 1 | 0,942071 |
| -2,12171 | 421 | 347,5352 | 0,034569 | 1 | 0,213506 |
| -2,16797 | 421 | 190,7107 | 0,0314 | 1 | 0,241423 |
| -1,67267 | 420 | 385,403 | 0,095203 | 1 | 0,848246 |
| -1,65636 | 367 | 341,352 | 0,098569 | 1 | 0,414924 |
| -0,46912 | 280 | 273,6323 | 0,63936 | 1 | 0,907449 |
| 1,304958 | 421 | 342,9018 | 0,192782 | 1 | 0,478621 |
| 1,019239 | 421 | 189,234 | 0,309391 | 1 | 0,607099 |
| 1,400466 | 420 | 376,3928 | 0,162198 | 1 | 0,914005 |

| | | | | | |
|---|---|---|---|---|---|
| 0,783736 | 367 | 335,3732 | 0,433748 | 1 | 0,74195 |
| 1,88348 | 280 | 263,3022 | 0,060737 | 1 | 0,422605 |
| -1,80648 | 421 | 310,9924 | 0,07181 | 1 | 0,288707 |
| -1,41019 | 421 | 180,7308 | 0,160203 | 1 | 0,442985 |
| -1,3853 | 420 | 345,0766 | 0,166855 | 1 | 0,914005 |
| -1,70263 | 367 | 321,1735 | 0,089605 | 1 | 0,397163 |
| -1,7387 | 280 | 246,8828 | 0,083335 | 1 | 0,456738 |
| 2,008169 | 421 | 357,2193 | 0,045378 | 1 | 0,237403 |
| 1,154308 | 421 | 194,5124 | 0,24979 | 1 | 0,548314 |
| 0,109125 | 420 | 392,0386 | 0,913159 | 1 | 0,987181 |
| 0,804218 | 367 | 340,5037 | 0,421832 | 1 | 0,731532 |
| 0,344357 | 280 | 261,8799 | 0,730854 | 1 | 0,936637 |
| 1,907792 | 421 | 354,2446 | 0,057226 | 1 | 0,266382 |
| 0,9236 | 421 | 191,4797 | 0,356857 | 1 | 0,643758 |
| 0,139552 | 420 | 391,292 | 0,889086 | 1 | 0,983483 |
| 1,276448 | 367 | 337,5333 | 0,202675 | 1 | 0,556341 |
| -0,99171 | 280 | 269,5601 | 0,322227 | 1 | 0,745172 |
| -1,89629 | 421 | 372,8233 | 0,058695 | 1 | 0,268334 |
| 0,469599 | 421 | 197,4951 | 0,639159 | 1 | 0,838432 |
| -0,46107 | 420 | 399,7383 | 0,645 | 1 | 0,964878 |
| 2,020108 | 367 | 353,2732 | 0,044127 | 1 | 0,300899 |
| -0,46727 | 280 | 263,2893 | 0,640694 | 1 | 0,907449 |
| -0,34867 | 421 | 390,42 | 0,727526 | 1 | 0,890807 |
| -1,07858 | 421 | 208,9035 | 0,28202 | 1 | 0,581951 |
| 0,450995 | 420 | 411,5854 | 0,652231 | 1 | 0,966867 |
| 0,302936 | 367 | 355,4145 | 0,762116 | 1 | 0,929966 |
| 0,035767 | 280 | 268,7134 | 0,971495 | 1 | 0,992124 |
| 0,51158 | 421 | 365,1446 | 0,609255 | 1 | 0,824796 |
| 0,002427 | 421 | 194,567 | 0,998066 | 1 | 0,999267 |
| 2,29493 | 420 | 393,3081 | 0,022263 | 1 | 0,75904 |
| -0,03492 | 367 | 347,7534 | 0,972162 | 1 | 0,989229 |
| 0,641425 | 280 | 267,4801 | 0,521796 | 1 | 0,833721 |
| -1,03467 | 421 | 406,3705 | 0,301436 | 1 | 0,597107 |
| 0,237089 | 421 | 222,7826 | 0,812806 | 1 | 0,926063 |
| 0,479318 | 420 | 415,9938 | 0,631964 | 1 | 0,964878 |
| -1,58645 | 367 | 362,9189 | 0,113508 | 1 | 0,439732 |
| 1,115989 | 280 | 275,9997 | 0,265397 | 1 | 0,689576 |
| -1,67358 | 421 | 367,4248 | 0,095065 | 1 | 0,331627 |
| 0,107868 | 421 | 197,6753 | 0,91421 | 1 | 0,968176 |
| 0,702088 | 420 | 397,5062 | 0,483035 | 1 | 0,960096 |
| 0,316907 | 367 | 352,6718 | 0,751502 | 1 | 0,924056 |
| 0,621619 | 280 | 265,9026 | 0,534725 | 1 | 0,84326 |
| -0,88175 | 421 | 402,9676 | 0,37844 | 1 | 0,670163 |
| -0,49236 | 421 | 219,2249 | 0,62296 | 1 | 0,830785 |
| -0,32996 | 420 | 415,7575 | 0,741598 | 1 | 0,969427 |
| -1,52463 | 367 | 362,9943 | 0,128222 | 1 | 0,471294 |
| -0,45046 | 280 | 274,8785 | 0,652732 | 1 | 0,908644 |
| -2,13861 | 421 | 316,0605 | 0,033233 | 1 | 0,209551 |

| | | | | | |
|---|---|---|---|---|---|
| -2,16275 | 421 | 180,321 | 0,031877 | 1 | 0,241423 |
| -0,26312 | 420 | 349,4618 | 0,792612 | 1 | 0,969427 |
| -0,58749 | 367 | 312,1822 | 0,557298 | 1 | 0,83071 |
| -1,52504 | 280 | 260,9048 | 0,12846 | 1 | 0,536498 |
| -0,18498 | 421 | 414,3569 | 0,853337 | 1 | 0,946355 |
| 0,57774 | 421 | 236,1468 | 0,56399 | 1 | 0,786247 |
| 0,730886 | 420 | 413,2583 | 0,465263 | 1 | 0,960096 |
| -0,81969 | 367 | 360,1392 | 0,412937 | 1 | 0,724248 |
| 0,319792 | 280 | 275,2126 | 0,749369 | 1 | 0,942071 |
| -1,9346 | 421 | 371,5745 | 0,053798 | 1 | 0,259162 |
| -0,89442 | 421 | 199,7327 | 0,372173 | 1 | 0,650743 |
| 0,268176 | 420 | 400,7849 | 0,788702 | 1 | 0,969427 |
| -0,90114 | 367 | 353,2878 | 0,368126 | 1 | 0,678946 |
| -0,83131 | 280 | 267,9322 | 0,406538 | 1 | 0,788151 |
| 1,927238 | 421 | 384,1736 | 0,054687 | 1 | 0,259162 |
| 2,07018 | 421 | 205,712 | 0,039684 | 1 | 0,26998 |
| 0,432348 | 420 | 410,2208 | 0,665716 | 1 | 0,96781 |
| 1,871484 | 367 | 360,582 | 0,062087 | 1 | 0,347744 |
| 0,891537 | 280 | 275,8465 | 0,373418 | 1 | 0,763914 |
| 1,725094 | 421 | 380,5282 | 0,085322 | 1 | 0,311334 |
| 2,415715 | 421 | 204,6496 | 0,016583 | 1 | 0,181207 |
| 0,519447 | 420 | 408,8722 | 0,60373 | 1 | 0,964878 |
| 2,511035 | 367 | 355,8507 | 0,01248 | 1 | 0,189546 |
| 1,806268 | 280 | 272,7164 | 0,071979 | 1 | 0,439393 |
| -1,28968 | 421 | 354,378 | 0,198004 | 1 | 0,484381 |
| 1,704589 | 421 | 190,5058 | 0,089901 | 1 | 0,372201 |
| 1,616025 | 420 | 387,001 | 0,106904 | 1 | 0,848246 |
| 1,424106 | 367 | 331,2547 | 0,155357 | 1 | 0,502197 |
| 0,969194 | 280 | 257,2669 | 0,333359 | 1 | 0,755731 |
| 0,377612 | 421 | 336,4324 | 0,705957 | 1 | 0,875021 |
| 0,906961 | 421 | 187,0831 | 0,365594 | 1 | 0,647779 |
| 0,118977 | 420 | 373,1014 | 0,905358 | 1 | 0,98503 |
| -0,35597 | 367 | 327,6356 | 0,722095 | 1 | 0,91599 |
| 0,926337 | 280 | 260,9998 | 0,355126 | 1 | 0,761846 |
| -0,87966 | 421 | 311,3713 | 0,37972 | 1 | 0,671614 |
| -0,51011 | 421 | 179,4903 | 0,610601 | 1 | 0,82272 |
| -0,67833 | 420 | 340,532 | 0,498021 | 1 | 0,960096 |
| -0,36511 | 367 | 321,5677 | 0,715266 | 1 | 0,912706 |
| -0,60065 | 280 | 258,2362 | 0,5486 | 1 | 0,852208 |
| 1,213165 | 421 | 346,0224 | 0,225894 | 1 | 0,518147 |
| 1,535897 | 421 | 187,2004 | 0,126252 | 1 | 0,410224 |
| -0,25825 | 420 | 380,0149 | 0,796352 | 1 | 0,969427 |
| 1,008016 | 367 | 334,6069 | 0,314175 | 1 | 0,647568 |
| 1,652362 | 280 | 252,1158 | 0,099706 | 1 | 0,480557 |
| 1,802947 | 421 | 352,8395 | 0,07225 | 1 | 0,289575 |
| 0,906243 | 421 | 191,148 | 0,365948 | 1 | 0,647779 |
| 0,583518 | 420 | 386,1709 | 0,559885 | 1 | 0,960096 |
| 1,638155 | 367 | 335,1298 | 0,102328 | 1 | 0,426647 |

| | | | | | |
|---|---|---|---|---|---|
| -0,50522 | 280 | 266,0531 | 0,61382 | 1 | 0,889365 |
| 0,139971 | 421 | 335,7723 | 0,888767 | 1 | 0,962 |
| -0,60269 | 421 | 187,2342 | 0,547447 | 1 | 0,776424 |
| 0,850309 | 420 | 370,6216 | 0,395702 | 1 | 0,95743 |
| -1,25001 | 367 | 327,459 | 0,212188 | 1 | 0,563532 |
| -0,782 | 280 | 256,0227 | 0,434939 | 1 | 0,803251 |
| 2,019707 | 421 | 370,32 | 0,044134 | 1 | 0,236315 |
| 0,466091 | 421 | 199,5763 | 0,641658 | 1 | 0,838647 |
| -0,29275 | 420 | 400,7636 | 0,769863 | 1 | 0,969427 |
| 1,321325 | 367 | 350,0102 | 0,187256 | 1 | 0,537514 |
| -0,83047 | 280 | 263,6337 | 0,407025 | 1 | 0,788151 |
| -0,72566 | 421 | 380,2603 | 0,468493 | 1 | 0,729358 |
| -0,9874 | 421 | 202,0416 | 0,324627 | 1 | 0,619085 |
| -1,05301 | 420 | 405,6128 | 0,292963 | 1 | 0,943897 |
| -0,50454 | 367 | 346,9015 | 0,614205 | 1 | 0,872281 |
| -0,1628 | 280 | 271,7066 | 0,870798 | 1 | 0,969656 |
| -2,54395 | 421 | 386,8959 | 0,011349 | 1 | 0,128401 |
| -0,97765 | 421 | 208,7987 | 0,329379 | 1 | 0,625156 |
| 0,450504 | 420 | 411,3172 | 0,652585 | 1 | 0,966867 |
| -1,72602 | 367 | 358,0146 | 0,085207 | 1 | 0,388246 |
| -0,18034 | 280 | 274,1267 | 0,857017 | 1 | 0,966655 |
| 0,083753 | 421 | 393,0394 | 0,933295 | 1 | 0,972684 |
| -0,4632 | 421 | 209,9942 | 0,643698 | 1 | 0,838647 |
| 0,120449 | 420 | 412,8067 | 0,904186 | 1 | 0,98503 |
| 1,447597 | 367 | 359,7225 | 0,148601 | 1 | 0,499998 |
| 2,505044 | 280 | 270,0583 | 0,012833 | 1 | 0,2448 |
| -0,41226 | 421 | 335,8438 | 0,680411 | 1 | 0,861268 |
| 1,363905 | 421 | 184,395 | 0,17426 | 1 | 0,459198 |
| 0,736273 | 420 | 371,5189 | 0,462029 | 1 | 0,960096 |
| 0,081074 | 367 | 321,0303 | 0,935434 | 1 | 0,978278 |
| 0,114145 | 280 | 262,4054 | 0,90921 | 1 | 0,976008 |
| -0,65337 | 421 | 380,4995 | 0,513914 | 1 | 0,767364 |
| -1,23795 | 421 | 203,0941 | 0,217162 | 1 | 0,514565 |
| 1,587312 | 420 | 407,4932 | 0,113218 | 1 | 0,848246 |
| -0,98477 | 367 | 353,0264 | 0,32541 | 1 | 0,655597 |
| 0,176786 | 280 | 269,3633 | 0,85981 | 1 | 0,966655 |
| -2,7366 | 421 | 412,9718 | 0,006476 | 1 | 0,102724 |
| -2,32341 | 421 | 231,343 | 0,021025 | 1 | 0,2022 |
| -0,67507 | 420 | 414,4896 | 0,500009 | 1 | 0,960096 |
| 1,129159 | 367 | 360,6162 | 0,259581 | 1 | 0,609149 |
| 0,822985 | 280 | 275,8235 | 0,411228 | 1 | 0,788151 |
| -1,35522 | 421 | 326,4788 | 0,176284 | 1 | 0,451804 |
| -0,09578 | 421 | 182,2599 | 0,923804 | 1 | 0,971509 |
| -0,09884 | 420 | 359,2158 | 0,921323 | 1 | 0,989204 |
| -0,03536 | 367 | 304,3403 | 0,971816 | 1 | 0,989229 |
| -1,23701 | 280 | 248,2874 | 0,217251 | 1 | 0,650539 |
| 0,119683 | 421 | 380,2272 | 0,904798 | 1 | 0,965673 |
| -2,4929 | 421 | 201,9798 | 0,013474 | 1 | 0,17172 |

| | | | | | |
|---|---|---|---|---|---|
| -0,24583 | 420 | 406,2342 | 0,805938 | 1 | 0,969427 |
| -1,34252 | 367 | 346,1269 | 0,180307 | 1 | 0,527769 |
| 0,063076 | 280 | 269,743 | 0,949752 | 1 | 0,982409 |
| -0,09717 | 421 | 417 | 0,922635 | 1 | 0,966828 |
| 1,381054 | 421 | 417 | 0,168002 | 1 | 0,45454 |
| -0,31254 | 420 | 416 | 0,754785 | 1 | 0,969427 |
| 0,073382 | 367 | 363 | 0,941542 | 1 | 0,979539 |
| -0,27237 | 280 | 276 | 0,785541 | 1 | 0,953235 |
| -1,27801 | 421 | 411,8544 | 0,201965 | 1 | 0,490904 |
| 0,418944 | 421 | 229,7203 | 0,675649 | 1 | 0,854676 |
| -0,91239 | 420 | 415,2821 | 0,36209 | 1 | 0,955639 |
| -1,77564 | 367 | 362,8715 | 0,07663 | 1 | 0,378716 |
| 0,924486 | 280 | 275,6225 | 0,356042 | 1 | 0,761846 |
| -0,87527 | 421 | 413,1565 | 0,381933 | 1 | 0,671614 |
| 0,136809 | 421 | 233,3615 | 0,8913 | 1 | 0,958882 |
| 0,902906 | 420 | 414,5719 | 0,3671 | 1 | 0,95743 |
| 0,879235 | 367 | 361,2532 | 0,379858 | 1 | 0,691076 |
| -0,93096 | 280 | 275,829 | 0,35269 | 1 | 0,761846 |
| 0,096429 | 421 | 395,7551 | 0,923229 | 1 | 0,966828 |
| -1,29952 | 421 | 211,2546 | 0,195182 | 1 | 0,483337 |
| -1,43818 | 420 | 414,1136 | 0,151139 | 1 | 0,914005 |
| -1,6854 | 367 | 357,543 | 0,092784 | 1 | 0,40529 |
| -0,0189 | 280 | 268,7598 | 0,984931 | 1 | 0,992411 |
| -0,03919 | 421 | 339,2272 | 0,968759 | 1 | 0,993819 |
| 0,640939 | 421 | 185,6914 | 0,522353 | 1 | 0,765001 |
| -0,65969 | 420 | 371,2818 | 0,509863 | 1 | 0,960096 |
| -0,00583 | 367 | 337,9864 | 0,99535 | 1 | 0,997335 |
| -0,24155 | 280 | 263,443 | 0,80932 | 1 | 0,95747 |
| 0,250671 | 421 | 337,3721 | 0,802221 | 1 | 0,922616 |
| 1,157628 | 421 | 185,1788 | 0,248507 | 1 | 0,548314 |
| -0,49152 | 420 | 373,7519 | 0,623345 | 1 | 0,964878 |
| 1,314068 | 367 | 327,485 | 0,189743 | 1 | 0,539515 |
| 1,031157 | 280 | 261,0599 | 0,303421 | 1 | 0,726446 |
| -0,15425 | 421 | 347,7046 | 0,877502 | 1 | 0,95861 |
| 1,066234 | 421 | 188,3727 | 0,287683 | 1 | 0,585544 |
| 1,884235 | 420 | 386,7104 | 0,060283 | 1 | 0,848246 |
| 1,823715 | 367 | 340,0921 | 0,069073 | 1 | 0,364488 |
| 0,740736 | 280 | 265,1779 | 0,459509 | 1 | 0,818534 |
| -0,49805 | 421 | 319,1808 | 0,61879 | 1 | 0,826904 |
| 0,462571 | 421 | 178,9106 | 0,644234 | 1 | 0,838647 |
| -0,91485 | 420 | 352,6521 | 0,360893 | 1 | 0,955547 |
| 0,320159 | 367 | 311,481 | 0,749062 | 1 | 0,923669 |
| 1,533685 | 280 | 247,2629 | 0,126386 | 1 | 0,536498 |
| -1,19328 | 421 | 393,7655 | 0,233476 | 1 | 0,525147 |
| -1,87427 | 421 | 210,8132 | 0,062277 | 1 | 0,319638 |
| 0,433149 | 420 | 413,8421 | 0,665132 | 1 | 0,96781 |
| -0,16767 | 367 | 360,9151 | 0,866937 | 1 | 0,959636 |
| 0,064315 | 280 | 271,5771 | 0,948767 | 1 | 0,982409 |

| | | | | | |
|---|---|---|---|---|---|
| 0,459466 | 421 | 394,2861 | 0,646153 | 1 | 0,838719 |
| 1,170012 | 421 | 211,0092 | 0,243316 | 1 | 0,542422 |
| -0,46911 | 420 | 412,8733 | 0,639236 | 1 | 0,964878 |
| -0,72855 | 367 | 359,8699 | 0,466753 | 1 | 0,771623 |
| 1,035113 | 280 | 268,6732 | 0,301547 | 1 | 0,723617 |
| -0,08612 | 421 | 314,0301 | 0,931426 | 1 | 0,972063 |
| -0,07967 | 421 | 180,844 | 0,936592 | 1 | 0,97636 |
| -1,27484 | 420 | 347,4931 | 0,203216 | 1 | 0,928589 |
| -0,39804 | 367 | 314,434 | 0,69087 | 1 | 0,903032 |
| 0,128866 | 280 | 258,164 | 0,897564 | 1 | 0,975243 |
| -0,89894 | 421 | 367,6415 | 0,369272 | 1 | 0,661703 |
| -1,6934 | 421 | 196,4882 | 0,091964 | 1 | 0,373113 |
| -1,51213 | 420 | 397,7111 | 0,131296 | 1 | 0,888479 |
| 0,14919 | 367 | 343,0198 | 0,881491 | 1 | 0,962453 |
| 0,3377 | 280 | 263,0734 | 0,735859 | 1 | 0,938189 |
| 2,992439 | 421 | 319,0757 | 0,002983 | 1 | 0,077359 |
| 2,707564 | 421 | 181,8404 | 0,007424 | 1 | 0,145306 |
| 0,892338 | 420 | 352,4297 | 0,37282 | 1 | 0,95743 |
| 2,251196 | 367 | 318,5761 | 0,025054 | 1 | 0,247087 |
| -0,48474 | 280 | 252,2678 | 0,62828 | 1 | 0,900874 |
| 1,500458 | 421 | 382,92 | 0,13432 | 1 | 0,407576 |
| 3,075385 | 421 | 203,9758 | 0,00239 | 1 | 0,10292 |
| 1,137801 | 420 | 409,2253 | 0,255869 | 1 | 0,943897 |
| 1,604412 | 367 | 358,8058 | 0,109503 | 1 | 0,435936 |
| 2,588832 | 280 | 271,8358 | 0,010149 | 1 | 0,229419 |
| -0,69126 | 421 | 389,1036 | 0,489816 | 1 | 0,748388 |
| -2,31607 | 421 | 207,2808 | 0,021532 | 1 | 0,204081 |
| -3,92244 | 420 | 411,9522 | 1,03E-04 | 0,015811 | 0,038681 |
| -0,73889 | 367 | 357,9356 | 0,460457 | 1 | 0,765059 |
| -2,71519 | 280 | 269,991 | 0,007051 | 1 | 0,195931 |
| -1,46461 | 421 | 318,9094 | 0,144013 | 1 | 0,412154 |
| -0,40756 | 421 | 177,1622 | 0,684092 | 1 | 0,859821 |
| -0,93252 | 420 | 352,7583 | 0,351706 | 1 | 0,947373 |
| -0,26116 | 367 | 315,4071 | 0,794142 | 1 | 0,945294 |
| 0,612012 | 280 | 249,8235 | 0,541087 | 1 | 0,849393 |
| 1,102367 | 421 | 410,1829 | 0,270949 | 1 | 0,56693 |
| -0,05554 | 421 | 226,9993 | 0,95576 | 1 | 0,98414 |
| -1,17393 | 420 | 415,514 | 0,241096 | 1 | 0,943659 |
| -0,183 | 367 | 362,9295 | 0,854899 | 1 | 0,957157 |
| -1,49358 | 280 | 275,8614 | 0,136428 | 1 | 0,54931 |
| -0,42443 | 421 | 348,5768 | 0,671517 | 1 | 0,856155 |
| -0,5749 | 421 | 188,4259 | 0,566043 | 1 | 0,788379 |
| -1,44467 | 420 | 379,6967 | 0,149377 | 1 | 0,914005 |
| -0,24473 | 367 | 335,6903 | 0,806817 | 1 | 0,950149 |
| 1,156474 | 280 | 262,0665 | 0,24854 | 1 | 0,677438 |
| 0,424598 | 421 | 369,9654 | 0,671376 | 1 | 0,856155 |
| 2,807515 | 421 | 197,6762 | 0,005493 | 1 | 0,1403 |
| 1,279435 | 420 | 399,9807 | 0,201486 | 1 | 0,928589 |

| | | | | | |
|---|---|---|---|---|---|
| 1,883631 | 367 | 346,9476 | 0,060451 | 1 | 0,341198 |
| 0,592836 | 280 | 262,8606 | 0,553801 | 1 | 0,854225 |
| 0,818056 | 421 | 387,1151 | 0,413828 | 1 | 0,695339 |
| 1,665971 | 421 | 209,5689 | 0,097213 | 1 | 0,379251 |
| -0,31353 | 420 | 412,4163 | 0,754034 | 1 | 0,969427 |
| -0,11755 | 367 | 359,1707 | 0,906492 | 1 | 0,970486 |
| -0,70326 | 280 | 271,2127 | 0,4825 | 1 | 0,824339 |
| 0,519487 | 421 | 401,259 | 0,603708 | 1 | 0,821108 |
| 0,106535 | 421 | 217,3806 | 0,915256 | 1 | 0,968604 |
| 0,044014 | 420 | 415,2273 | 0,964914 | 1 | 0,996924 |
| -0,81441 | 367 | 360,0284 | 0,41595 | 1 | 0,727188 |
| 1,201512 | 280 | 272,6588 | 0,230595 | 1 | 0,657169 |
| -0,95303 | 421 | 384,2729 | 0,341176 | 1 | 0,63164 |
| 0,329027 | 421 | 203,4946 | 0,742474 | 1 | 0,888022 |
| -0,47193 | 420 | 407,8185 | 0,63723 | 1 | 0,964878 |
| 0,764031 | 367 | 356,0909 | 0,445355 | 1 | 0,749888 |
| 1,318245 | 280 | 273,5321 | 0,188525 | 1 | 0,623041 |
| -0,56515 | 421 | 414,2937 | 0,572278 | 1 | 0,79928 |
| 0,935605 | 421 | 234,0395 | 0,350441 | 1 | 0,643758 |
| -0,93417 | 420 | 413,7813 | 0,35076 | 1 | 0,947373 |
| 2,363507 | 367 | 362,6169 | 0,01863 | 1 | 0,217108 |
| -1,43271 | 280 | 274,7186 | 0,153079 | 1 | 0,565026 |
| -1,91637 | 421 | 336,5146 | 0,056165 | 1 | 0,2645 |
| -3,08218 | 421 | 182,6014 | 0,002373 | 0,351256 | 0,10292 |
| -1,43604 | 420 | 373,9794 | 0,151825 | 1 | 0,914005 |
| -1,92657 | 367 | 335,2943 | 0,054879 | 1 | 0,335053 |
| -0,51892 | 280 | 258,4616 | 0,60426 | 1 | 0,884956 |
| -0,51331 | 421 | 375,056 | 0,608039 | 1 | 0,824024 |
| -0,32188 | 421 | 201,1877 | 0,747876 | 1 | 0,889542 |
| -0,30012 | 420 | 404,4742 | 0,764242 | 1 | 0,969427 |
| 0,056636 | 367 | 346,0006 | 0,954867 | 1 | 0,980235 |
| -0,694 | 280 | 263,4167 | 0,488296 | 1 | 0,826999 |
| -0,33179 | 421 | 318,6714 | 0,740269 | 1 | 0,893273 |
| 0,421313 | 421 | 178,3538 | 0,674034 | 1 | 0,853746 |
| -0,65225 | 420 | 352,5703 | 0,514664 | 1 | 0,960096 |
| 0,184897 | 367 | 309,4924 | 0,853431 | 1 | 0,956935 |
| 1,78346 | 280 | 247,564 | 0,075736 | 1 | 0,449349 |
| 0,978156 | 421 | 284,9411 | 0,328827 | 1 | 0,618807 |
| 2,167925 | 421 | 172,691 | 0,031533 | 1 | 0,241423 |
| 1,068806 | 420 | 315,6195 | 0,285974 | 1 | 0,943897 |
| -0,63297 | 367 | 287,4459 | 0,527258 | 1 | 0,811085 |
| 0,685908 | 280 | 245,7876 | 0,493417 | 1 | 0,827962 |
| 0,824307 | 421 | 334,7309 | 0,410352 | 1 | 0,695339 |
| 0,384977 | 421 | 186,6071 | 0,700693 | 1 | 0,865528 |
| 0,955426 | 420 | 368,0531 | 0,339989 | 1 | 0,947373 |
| -0,24003 | 367 | 317,7672 | 0,810465 | 1 | 0,950149 |
| 0,0307 | 280 | 259,5813 | 0,975532 | 1 | 0,99236 |
| -1,1984 | 421 | 378,384 | 0,231511 | 1 | 0,523069 |

| | | | | | |
|---|---|---|---|---|---|
| -1,3341 | 421 | 201,3864 | 0,183679 | 1 | 0,473593 |
| 0,856885 | 420 | 405,5458 | 0,392014 | 1 | 0,95743 |
| -2,97624 | 367 | 351,7199 | 0,00312 | 0,471125 | 0,176492 |
| -1,2199 | 280 | 268,2771 | 0,223573 | 1 | 0,654014 |
| -0,57248 | 421 | 414,4287 | 0,56731 | 1 | 0,796637 |
| -0,85379 | 421 | 236,5252 | 0,394085 | 1 | 0,665792 |
| -0,26135 | 420 | 413,2087 | 0,793949 | 1 | 0,969427 |
| -1,42304 | 367 | 360,391 | 0,15559 | 1 | 0,502197 |
| -0,47892 | 280 | 275,8408 | 0,632374 | 1 | 0,903355 |
| -1,43822 | 421 | 305,0721 | 0,151397 | 1 | 0,42445 |
| -0,91506 | 421 | 178,6192 | 0,361393 | 1 | 0,64452 |
| -2,05455 | 420 | 336,1562 | 0,040695 | 1 | 0,846132 |
| -0,14859 | 367 | 295,3664 | 0,881982 | 1 | 0,962453 |
| -2,40365 | 280 | 248,5584 | 0,016965 | 1 | 0,266517 |
| 1,755717 | 421 | 363,1739 | 0,07998 | 1 | 0,302838 |
| 1,326064 | 421 | 196,1346 | 0,186361 | 1 | 0,474401 |
| 0,936926 | 420 | 395,4276 | 0,349369 | 1 | 0,947373 |
| -0,95211 | 367 | 346,1415 | 0,341707 | 1 | 0,664831 |
| 1,592727 | 280 | 262,4411 | 0,112425 | 1 | 0,50726 |
| -1,65675 | 421 | 334,1975 | 0,098508 | 1 | 0,339344 |
| 0,57166 | 421 | 183,1155 | 0,568254 | 1 | 0,78927 |
| -0,41234 | 420 | 367,1048 | 0,680331 | 1 | 0,96781 |
| 0,116446 | 367 | 307,4477 | 0,907375 | 1 | 0,970486 |
| -1,67836 | 280 | 252,5562 | 0,094514 | 1 | 0,474774 |
| 0,124058 | 421 | 384,8157 | 0,901334 | 1 | 0,965517 |
| 1,700011 | 421 | 204,8602 | 0,090646 | 1 | 0,373113 |
| 0,652078 | 420 | 408,308 | 0,514718 | 1 | 0,960096 |
| 2,999476 | 367 | 353,0503 | 0,002896 | 1 | 0,176492 |
| 0,577843 | 280 | 270,7444 | 0,56385 | 1 | 0,861787 |
| -0,33375 | 421 | 315,1521 | 0,738793 | 1 | 0,893273 |
| 0,34415 | 421 | 178,2797 | 0,731139 | 1 | 0,882616 |
| -0,85389 | 420 | 348,5427 | 0,393751 | 1 | 0,95743 |
| 0,341465 | 367 | 309,4003 | 0,732985 | 1 | 0,923669 |
| 2,164711 | 280 | 247,0127 | 0,031368 | 1 | 0,329568 |
| -0,48517 | 421 | 336,0256 | 0,627875 | 1 | 0,833119 |
| -1,30892 | 421 | 186,0032 | 0,192176 | 1 | 0,48028 |
| -0,40102 | 420 | 372,935 | 0,688638 | 1 | 0,968866 |
| 0,109696 | 367 | 331,99 | 0,912717 | 1 | 0,972059 |
| -0,99039 | 280 | 264,2102 | 0,322891 | 1 | 0,745172 |
| -1,83682 | 421 | 366,8837 | 0,067046 | 1 | 0,279111 |
| -2,66601 | 421 | 197,4745 | 0,008312 | 1 | 0,148102 |
| -2,43915 | 420 | 397,8489 | 0,015158 | 1 | 0,723749 |
| -1,98672 | 367 | 346,5264 | 0,04774 | 1 | 0,311448 |
| 0,693474 | 280 | 269,5858 | 0,488609 | 1 | 0,826999 |
| -0,96898 | 421 | 325,286 | 0,333275 | 1 | 0,62159 |
| -0,88427 | 421 | 183,2823 | 0,377711 | 1 | 0,655017 |
| -1,54984 | 420 | 355,6058 | 0,122069 | 1 | 0,87201 |
| -1,08545 | 367 | 311,7266 | 0,278563 | 1 | 0,612838 |

| | | | | | |
|---|---|---|---|---|---|
| -0,92677 | 280 | 256,2846 | 0,354919 | 1 | 0,761846 |
| -0,97712 | 421 | 291,9268 | 0,329319 | 1 | 0,618807 |
| 0,110787 | 421 | 174,4399 | 0,911913 | 1 | 0,96646 |
| -0,5421 | 420 | 320,2197 | 0,588126 | 1 | 0,964878 |
| -0,85342 | 367 | 290,4671 | 0,39413 | 1 | 0,706248 |
| 0,670369 | 280 | 248,9153 | 0,503244 | 1 | 0,831912 |
| 0,12225 | 421 | 335,3351 | 0,902774 | 1 | 0,965517 |
| -0,14735 | 421 | 187,7766 | 0,883016 | 1 | 0,957051 |
| -0,59068 | 420 | 370,3786 | 0,555093 | 1 | 0,960096 |
| -0,82578 | 367 | 329,6482 | 0,409524 | 1 | 0,722315 |
| -0,08404 | 280 | 266,6837 | 0,933092 | 1 | 0,979504 |
| -3,29672 | 421 | 350,1292 | 0,001078 | 1 | 0,064989 |
| -2,10838 | 421 | 187,5832 | 0,036326 | 1 | 0,261928 |
| -0,56673 | 420 | 383,0667 | 0,571232 | 1 | 0,964199 |
| -1,39579 | 367 | 334,1796 | 0,163703 | 1 | 0,510294 |
| -3,11726 | 280 | 259,6821 | 0,002031 | 1 | 0,135787 |
| -2,97097 | 421 | 364,4191 | 0,003165 | 1 | 0,078203 |
| -0,14838 | 421 | 195,3024 | 0,882195 | 1 | 0,957051 |
| -0,67548 | 420 | 394,745 | 0,499764 | 1 | 0,960096 |
| -1,11736 | 367 | 342,1266 | 0,264625 | 1 | 0,609149 |
| 1,045235 | 280 | 257,9539 | 0,296893 | 1 | 0,723324 |
| -0,01992 | 421 | 355,2843 | 0,984121 | 1 | 0,998028 |
| -0,09721 | 421 | 191,2627 | 0,922658 | 1 | 0,971139 |
| 0,641486 | 420 | 386,5271 | 0,521587 | 1 | 0,960096 |
| -0,09724 | 367 | 337,0796 | 0,922597 | 1 | 0,975687 |
| -0,02855 | 280 | 258,648 | 0,977249 | 1 | 0,99236 |
| -1,04802 | 421 | 330,1023 | 0,295397 | 1 | 0,593552 |
| 0,067802 | 421 | 182,5403 | 0,946018 | 1 | 0,980846 |
| 0,629857 | 420 | 367,1333 | 0,52918 | 1 | 0,960096 |
| -0,04185 | 367 | 331,0031 | 0,966642 | 1 | 0,986276 |
| 1,649038 | 280 | 271,2348 | 0,100298 | 1 | 0,480557 |
| -0,0405 | 421 | 325,3836 | 0,967717 | 1 | 0,993427 |
| -1,74908 | 421 | 183,2684 | 0,081952 | 1 | 0,362173 |
| -2,21372 | 420 | 353,8769 | 0,027485 | 1 | 0,767829 |
| -1,4886 | 367 | 309,4171 | 0,13761 | 1 | 0,4834 |
| -1,74442 | 280 | 265,9595 | 0,082242 | 1 | 0,456738 |
| 1,84051 | 421 | 385,8965 | 0,066461 | 1 | 0,278382 |
| 2,127765 | 421 | 205,0155 | 0,034551 | 1 | 0,252759 |
| 1,653506 | 420 | 410,3586 | 0,098993 | 1 | 0,848246 |
| 2,706974 | 367 | 353,9312 | 0,007119 | 1 | 0,180403 |
| 1,539998 | 280 | 272,7661 | 0,12472 | 1 | 0,535729 |
| 2,848518 | 421 | 335,8126 | 0,004663 | 1 | 0,087964 |
| 2,922194 | 421 | 183,3767 | 0,003913 | 1 | 0,118336 |
| 1,85433 | 420 | 368,934 | 0,06449 | 1 | 0,848246 |
| 2,992389 | 367 | 320,0069 | 0,002983 | 1 | 0,176492 |
| 1,647364 | 280 | 257,9854 | 0,100701 | 1 | 0,480557 |
| -1,64528 | 421 | 327,1648 | 0,100873 | 1 | 0,342121 |
| -0,24808 | 421 | 181,576 | 0,804353 | 1 | 0,923205 |

| | | | | | |
|---|---|---|---|---|---|
| 1,052041 | 420 | 365,7809 | 0,293475 | 1 | 0,943897 |
| 2,043324 | 367 | 330,2396 | 0,041814 | 1 | 0,291798 |
| 2,485578 | 280 | 261,6365 | 0,013559 | 1 | 0,252268 |
| 1,477914 | 421 | 334,0784 | 0,140373 | 1 | 0,410878 |
| -0,05312 | 421 | 186,9832 | 0,95769 | 1 | 0,98414 |
| -0,43828 | 420 | 368,0537 | 0,66144 | 1 | 0,96781 |
| -0,1907 | 367 | 307,219 | 0,848886 | 1 | 0,956935 |
| -0,35823 | 280 | 276 | 0,720448 | 1 | 0,932745 |
| -1,06721 | 421 | 370,2052 | 0,286571 | 1 | 0,584387 |
| -0,97525 | 421 | 195,9932 | 0,330637 | 1 | 0,626331 |
| 2,141617 | 420 | 398,1637 | 0,032831 | 1 | 0,828006 |
| 0,846375 | 367 | 348,1776 | 0,397925 | 1 | 0,710512 |
| -0,19624 | 280 | 261,2605 | 0,844577 | 1 | 0,965181 |
| -1,90399 | 421 | 322,2538 | 0,057802 | 1 | 0,266382 |
| 0,117403 | 421 | 183,1154 | 0,906669 | 1 | 0,966302 |
| -1,03345 | 420 | 355,8029 | 0,302093 | 1 | 0,943897 |
| 0,19836 | 367 | 314,8494 | 0,842891 | 1 | 0,956935 |
| -0,2928 | 280 | 258,3483 | 0,76991 | 1 | 0,946609 |
| -2,81899 | 421 | 349,2007 | 0,005092 | 1 | 0,087964 |
| -1,68421 | 421 | 191,8326 | 0,093768 | 1 | 0,373113 |
| -0,50659 | 420 | 385,8319 | 0,612735 | 1 | 0,964878 |
| -1,39147 | 367 | 338,3561 | 0,164997 | 1 | 0,511628 |
| -3,06244 | 280 | 262,7949 | 0,002423 | 1 | 0,135787 |
| -3,4644 | 421 | 367,1807 | 5,94E-04 | 0,633427 | 0,048251 |
| -0,76484 | 421 | 197,4901 | 0,445282 | 1 | 0,709954 |
| -0,47016 | 420 | 400,615 | 0,638495 | 1 | 0,964878 |
| 0,303677 | 367 | 343,3279 | 0,761558 | 1 | 0,929966 |
| 0,204493 | 280 | 266,9101 | 0,838124 | 1 | 0,964667 |
| -0,03149 | 421 | 368,104 | 0,974893 | 1 | 0,995982 |
| -0,16707 | 421 | 197,6424 | 0,867486 | 1 | 0,947676 |
| -1,30695 | 420 | 399,4146 | 0,191983 | 1 | 0,925729 |
| -1,12293 | 367 | 345,8713 | 0,262247 | 1 | 0,609149 |
| -1,85711 | 280 | 268,9587 | 0,064389 | 1 | 0,423666 |
| 0,214507 | 421 | 395,5173 | 0,830262 | 1 | 0,936611 |
| 0,477007 | 421 | 212,9607 | 0,633847 | 1 | 0,835702 |
| -1,41439 | 420 | 414,376 | 0,158 | 1 | 0,914005 |
| 1,224302 | 367 | 361,1857 | 0,221636 | 1 | 0,573978 |
| 1,287053 | 280 | 274,1416 | 0,199161 | 1 | 0,631529 |
| 2,47734 | 421 | 386,559 | 0,013663 | 1 | 0,140071 |
| 1,909756 | 421 | 206,3998 | 0,057551 | 1 | 0,312877 |
| 1,517966 | 420 | 410,6613 | 0,129792 | 1 | 0,888479 |
| 1,978496 | 367 | 356,8142 | 0,048641 | 1 | 0,313787 |
| 2,607528 | 280 | 268,9589 | 0,009629 | 1 | 0,226737 |
| 0,359696 | 421 | 345,9731 | 0,719294 | 1 | 0,884157 |
| 2,260095 | 421 | 185,2741 | 0,024979 | 1 | 0,218351 |
| 0,355868 | 420 | 381,302 | 0,722136 | 1 | 0,969427 |
| 1,583752 | 367 | 341,9041 | 0,114175 | 1 | 0,440054 |
| 3,879423 | 280 | 262,7357 | 1,32E-04 | 0,017612 | 0,046455 |

| | | | | | |
|---|---|---|---|---|---|
| 0,621981 | 421 | 359,0061 | 0,534349 | 1 | 0,778195 |
| 1,744021 | 421 | 194,6942 | 0,082734 | 1 | 0,36456 |
| -0,40486 | 420 | 392,5231 | 0,6858 | 1 | 0,968613 |
| 2,040971 | 367 | 343,6427 | 0,042017 | 1 | 0,291798 |
| 1,83535 | 280 | 264,6864 | 0,067576 | 1 | 0,425749 |
| -2,82935 | 421 | 345,9053 | 0,004936 | 1 | 0,087964 |
| -2,0405 | 421 | 190,7387 | 0,042678 | 1 | 0,273775 |
| -1,56242 | 420 | 384,022 | 0,119013 | 1 | 0,858144 |
| -1,45379 | 367 | 335,3668 | 0,14694 | 1 | 0,498736 |
| -0,46254 | 280 | 264,2323 | 0,644074 | 1 | 0,907804 |
| 0,813207 | 421 | 351,5726 | 0,41665 | 1 | 0,695339 |
| 0,185204 | 421 | 190,5475 | 0,853266 | 1 | 0,944105 |
| -0,26793 | 420 | 385,1346 | 0,788898 | 1 | 0,969427 |
| -0,21353 | 367 | 322,3311 | 0,831052 | 1 | 0,956843 |
| 0,997724 | 280 | 250,3507 | 0,319377 | 1 | 0,742748 |
| -1,78179 | 421 | 317,1114 | 0,075741 | 1 | 0,2988 |
| -0,6742 | 421 | 178,7173 | 0,501056 | 1 | 0,750551 |
| -1,8892 | 420 | 347,6485 | 0,059697 | 1 | 0,848246 |
| 0,834203 | 367 | 319,8536 | 0,404789 | 1 | 0,717667 |
| 2,208721 | 280 | 245,1604 | 0,02812 | 1 | 0,326641 |
| -0,30768 | 421 | 325,3374 | 0,758526 | 1 | 0,900919 |
| 0,755883 | 421 | 183,5188 | 0,450688 | 1 | 0,713861 |
| -0,39058 | 420 | 361,0623 | 0,696339 | 1 | 0,968866 |
| 0,594164 | 367 | 311,7336 | 0,552833 | 1 | 0,830257 |
| -0,69703 | 280 | 257,183 | 0,486413 | 1 | 0,826999 |
| 0,619087 | 421 | 364,738 | 0,536245 | 1 | 0,778195 |
| 0,656643 | 421 | 193,9603 | 0,512189 | 1 | 0,756803 |
| 0,636734 | 420 | 395,4068 | 0,524667 | 1 | 0,960096 |
| 1,102426 | 367 | 346,6711 | 0,271041 | 1 | 0,609149 |
| -1,76051 | 280 | 270,3014 | 0,079452 | 1 | 0,453386 |
| -0,07857 | 421 | 408,7492 | 0,937414 | 1 | 0,974957 |
| 0,146962 | 421 | 224,1845 | 0,883294 | 1 | 0,957051 |
| 1,385988 | 420 | 415,9996 | 0,166493 | 1 | 0,914005 |
| 0,029768 | 367 | 362,8119 | 0,976269 | 1 | 0,9914 |
| -0,94916 | 280 | 275,0885 | 0,343372 | 1 | 0,758791 |
| 0,996848 | 421 | 383,9121 | 0,319466 | 1 | 0,61486 |
| 0,250764 | 421 | 205,8988 | 0,802246 | 1 | 0,922323 |
| 1,116285 | 420 | 409,5273 | 0,264955 | 1 | 0,943897 |
| 1,247056 | 367 | 358,8285 | 0,21319 | 1 | 0,563645 |
| 0,633805 | 280 | 272,9639 | 0,526739 | 1 | 0,838222 |
| -1,42772 | 421 | 380,9707 | 0,154192 | 1 | 0,426363 |
| -1,46606 | 421 | 204,3498 | 0,144169 | 1 | 0,425172 |
| -0,98617 | 420 | 405,7957 | 0,324636 | 1 | 0,945573 |
| -0,24904 | 367 | 357,4966 | 0,803477 | 1 | 0,950149 |
| -0,62128 | 280 | 267,4477 | 0,534942 | 1 | 0,84326 |
| -0,98852 | 421 | 352,9246 | 0,323576 | 1 | 0,61749 |
| -0,85811 | 421 | 192,5222 | 0,391901 | 1 | 0,665733 |
| 1,129759 | 420 | 385,3133 | 0,259281 | 1 | 0,943897 |

| | | | | | |
|---|---|---|---|---|---|
| 0,685497 | 367 | 329,3661 | 0,493512 | 1 | 0,793601 |
| -0,50449 | 280 | 255,022 | 0,614353 | 1 | 0,889365 |
| -0,19494 | 421 | 301,058 | 0,845574 | 1 | 0,939735 |
| -0,06919 | 421 | 175,9478 | 0,944918 | 1 | 0,980711 |
| -0,42378 | 420 | 333,3917 | 0,671998 | 1 | 0,96781 |
| 0,984128 | 367 | 301,6995 | 0,325841 | 1 | 0,655597 |
| -0,46776 | 280 | 255,1679 | 0,640358 | 1 | 0,907449 |
| 0,300054 | 421 | 343,1802 | 0,764317 | 1 | 0,904102 |
| 1,366007 | 421 | 185,9598 | 0,173586 | 1 | 0,458938 |
| -0,48955 | 420 | 379,1952 | 0,624735 | 1 | 0,964878 |
| -0,55411 | 367 | 335,4992 | 0,579872 | 1 | 0,844137 |
| 1,093233 | 280 | 252,2335 | 0,275335 | 1 | 0,701156 |
| -0,10235 | 421 | 383,5601 | 0,91853 | 1 | 0,966828 |
| -0,3367 | 421 | 205,2325 | 0,736688 | 1 | 0,884613 |
| 0,997952 | 420 | 409,3639 | 0,318892 | 1 | 0,945573 |
| 1,104959 | 367 | 359,7997 | 0,269916 | 1 | 0,609149 |
| -1,52849 | 280 | 268,1604 | 0,12757 | 1 | 0,536498 |
| 0,446718 | 421 | 370,6997 | 0,655339 | 1 | 0,846936 |
| 1,089064 | 421 | 199,0082 | 0,277443 | 1 | 0,578094 |
| -1,05175 | 420 | 400,0411 | 0,293551 | 1 | 0,943897 |
| 1,689807 | 367 | 348,88 | 0,091958 | 1 | 0,405208 |
| 0,707826 | 280 | 264,4742 | 0,479676 | 1 | 0,824256 |
| -0,64045 | 421 | 334,581 | 0,522316 | 1 | 0,769202 |
| -0,83668 | 421 | 181,3066 | 0,403876 | 1 | 0,677774 |
| 0,540266 | 420 | 369,5408 | 0,589339 | 1 | 0,964878 |
| -0,59244 | 367 | 321,0848 | 0,553975 | 1 | 0,830257 |
| 1,894228 | 280 | 247,7957 | 0,059359 | 1 | 0,421955 |
| -0,46871 | 421 | 375,8798 | 0,639549 | 1 | 0,834842 |
| 0,493956 | 421 | 198,5118 | 0,621883 | 1 | 0,830096 |
| -0,04758 | 420 | 402,8887 | 0,962076 | 1 | 0,996924 |
| 0,82271 | 367 | 351,8395 | 0,41123 | 1 | 0,723976 |
| -0,80227 | 280 | 262,1887 | 0,423122 | 1 | 0,797055 |
| -2,51882 | 421 | 369,2126 | 0,012197 | 1 | 0,131857 |
| -1,94146 | 421 | 197,6358 | 0,053624 | 1 | 0,302776 |
| 0,321551 | 420 | 400,8507 | 0,747961 | 1 | 0,969427 |
| -2,60465 | 367 | 344,0899 | 0,009596 | 1 | 0,183886 |
| -3,20453 | 280 | 266,8068 | 0,001517 | 1 | 0,135787 |
| 0,755469 | 421 | 330,6574 | 0,450505 | 1 | 0,714643 |
| 0,592623 | 421 | 185,3343 | 0,554156 | 1 | 0,778237 |
| -2,59041 | 420 | 365,2649 | 0,00997 | 1 | 0,600995 |
| -0,5679 | 367 | 323,8067 | 0,570493 | 1 | 0,837947 |
| -0,9449 | 280 | 260,3684 | 0,345587 | 1 | 0,759183 |
| 0,021986 | 421 | 401,7804 | 0,98247 | 1 | 0,997025 |
| 0,764065 | 421 | 215,3475 | 0,445665 | 1 | 0,709954 |
| -0,63754 | 420 | 415,4345 | 0,524125 | 1 | 0,960096 |
| 0,480395 | 367 | 362,4251 | 0,631236 | 1 | 0,880707 |
| -1,12816 | 280 | 273,3512 | 0,260243 | 1 | 0,686841 |
| 2,42267 | 421 | 395,7527 | 0,015855 | 1 | 0,147897 |

| | | | | | |
|---|---|---|---|---|---|
| 2,303897 | 421 | 213,3054 | 0,022191 | 1 | 0,209014 |
| -1,28642 | 420 | 415,441 | 0,199012 | 1 | 0,928589 |
| 1,173957 | 367 | 360,2875 | 0,241188 | 1 | 0,593905 |
| -0,00975 | 280 | 273,2123 | 0,992226 | 1 | 0,996269 |
| -0,00206 | 421 | 314,6823 | 0,998355 | 1 | 0,999298 |
| 1,2193 | 421 | 178,036 | 0,224343 | 1 | 0,521994 |
| 0,120944 | 420 | 349,0044 | 0,903805 | 1 | 0,98503 |
| 0,426453 | 367 | 309,7382 | 0,670074 | 1 | 0,896008 |
| 1,257791 | 280 | 250,6857 | 0,209638 | 1 | 0,64091 |
| -2,16233 | 421 | 330,7598 | 0,031309 | 1 | 0,201145 |
| -0,18749 | 421 | 184,8335 | 0,851481 | 1 | 0,944105 |
| -0,56671 | 420 | 368,6302 | 0,571259 | 1 | 0,964199 |
| 0,398141 | 367 | 327,7669 | 0,690785 | 1 | 0,903032 |
| 0,076045 | 280 | 268,7624 | 0,93944 | 1 | 0,982175 |
| 2,29596 | 421 | 399,6308 | 0,022195 | 1 | 0,176042 |
| 1,687042 | 421 | 217,2391 | 0,093031 | 1 | 0,373113 |
| -1,13581 | 420 | 415,6295 | 0,25669 | 1 | 0,943897 |
| 1,931999 | 367 | 360,2983 | 0,054143 | 1 | 0,334402 |
| 1,463045 | 280 | 267,5481 | 0,144629 | 1 | 0,555466 |
| 4,181797 | 421 | 331,3137 | 3,71E-05 | 0,039958 | 0,019267 |
| 4,082518 | 421 | 179,2959 | 6,70E-05 | 0,072182 | 0,0202 |
| 2,799487 | 420 | 361,5461 | 0,005393 | 1 | 0,451515 |
| 2,640948 | 367 | 324,8985 | 0,008667 | 1 | 0,183886 |
| 3,152494 | 280 | 265,9154 | 0,001804 | 1 | 0,135787 |
| -0,68116 | 421 | 322,3574 | 0,496262 | 1 | 0,750117 |
| -0,51504 | 421 | 182,7857 | 0,607147 | 1 | 0,821338 |
| 1,503257 | 420 | 355,7423 | 0,13366 | 1 | 0,890998 |
| 0,367948 | 367 | 321,9551 | 0,713154 | 1 | 0,912652 |
| 0,566006 | 280 | 254,1897 | 0,571889 | 1 | 0,869862 |
| -1,20608 | 421 | 357,6289 | 0,228586 | 1 | 0,520357 |
| -2,43263 | 421 | 192,8162 | 0,015902 | 1 | 0,179127 |
| 0,06516 | 420 | 390,9262 | 0,94808 | 1 | 0,992811 |
| -1,75894 | 367 | 347,8348 | 0,079467 | 1 | 0,381841 |
| -2,14653 | 280 | 264,1258 | 0,032741 | 1 | 0,329568 |
| 0,429584 | 421 | 347,2162 | 0,667765 | 1 | 0,853538 |
| 0,987964 | 421 | 190,711 | 0,324422 | 1 | 0,619085 |
| 1,127803 | 420 | 381,1829 | 0,260113 | 1 | 0,943897 |
| 2,269306 | 367 | 320,3353 | 0,023914 | 1 | 0,245162 |
| 0,745671 | 280 | 263,1695 | 0,456532 | 1 | 0,816125 |
| -0,66977 | 421 | 369,7436 | 0,50342 | 1 | 0,75707 |
| -0,28914 | 421 | 197,206 | 0,772774 | 1 | 0,909577 |
| 1,630028 | 420 | 399,2188 | 0,103884 | 1 | 0,848246 |
| -1,22177 | 367 | 350,7193 | 0,222616 | 1 | 0,574455 |
| -2,78016 | 280 | 266,1668 | 0,005821 | 1 | 0,186692 |
| 1,009491 | 421 | 336,0295 | 0,313466 | 1 | 0,608538 |
| 0,774211 | 421 | 186,1702 | 0,439788 | 1 | 0,705407 |
| 1,093104 | 420 | 371,6823 | 0,275056 | 1 | 0,943897 |
| 0,220749 | 367 | 318,4978 | 0,825429 | 1 | 0,95466 |

| | | | | | |
|---|---|---|---|---|---|
| 0,450894 | 280 | 259,238 | 0,652443 | 1 | 0,908644 |
| -1,16003 | 421 | 338,9363 | 0,246852 | 1 | 0,544195 |
| -1,27954 | 421 | 186,9494 | 0,202292 | 1 | 0,497314 |
| -1,36332 | 420 | 377,7971 | 0,173593 | 1 | 0,9199 |
| -0,9065 | 367 | 323,5761 | 0,365346 | 1 | 0,678049 |
| 0,332321 | 280 | 256,8531 | 0,739918 | 1 | 0,940183 |
| -2,82097 | 421 | 329,0503 | 0,005079 | 1 | 0,087964 |
| -1,84595 | 421 | 183,1302 | 0,066513 | 1 | 0,330811 |
| -1,68889 | 420 | 361,6466 | 0,092103 | 1 | 0,848246 |
| 0,18134 | 367 | 312,6213 | 0,856218 | 1 | 0,957922 |
| -1,35663 | 280 | 258,63 | 0,176083 | 1 | 0,598625 |
| -0,47401 | 421 | 417 | 0,635742 | 1 | 0,83455 |
| 1,627285 | 421 | 417 | 0,104432 | 1 | 0,383851 |
| 0,325876 | 420 | 416 | 0,744682 | 1 | 0,969427 |
| 1,133141 | 367 | 363 | 0,257903 | 1 | 0,609149 |
| 0,830326 | 280 | 276 | 0,407072 | 1 | 0,788151 |
| -0,62694 | 421 | 356,643 | 0,531102 | 1 | 0,776668 |
| 0,927192 | 421 | 192,0711 | 0,354991 | 1 | 0,643758 |
| 0,272119 | 420 | 389,3929 | 0,785674 | 1 | 0,969427 |
| 1,137938 | 367 | 338,5565 | 0,255951 | 1 | 0,609149 |
| 1,651643 | 280 | 266,9158 | 0,099783 | 1 | 0,480557 |
| -2,33975 | 421 | 363,2789 | 0,019838 | 1 | 0,162476 |
| -0,03025 | 421 | 195,4758 | 0,975896 | 1 | 0,991789 |
| 0,804634 | 420 | 398,2893 | 0,421511 | 1 | 0,95743 |
| 1,290983 | 367 | 343,8297 | 0,197577 | 1 | 0,549567 |
| 1,914216 | 280 | 263,9112 | 0,056674 | 1 | 0,406703 |
| 1,646289 | 421 | 321,2157 | 0,100682 | 1 | 0,342121 |
| 0,623296 | 421 | 182,5525 | 0,533868 | 1 | 0,769894 |
| -0,37475 | 420 | 353,9479 | 0,708067 | 1 | 0,968866 |
| 0,589373 | 367 | 308,7314 | 0,556042 | 1 | 0,830257 |
| 1,875705 | 280 | 251,0287 | 0,061856 | 1 | 0,422605 |
| -2,19463 | 421 | 349,9881 | 0,028846 | 1 | 0,197598 |
| -0,93698 | 421 | 191,7648 | 0,349949 | 1 | 0,643758 |
| 0,093804 | 420 | 386,6665 | 0,925313 | 1 | 0,989204 |
| 0,358844 | 367 | 335,3655 | 0,719938 | 1 | 0,914797 |
| 0,821791 | 280 | 264,4961 | 0,411936 | 1 | 0,788151 |
| -1,81672 | 421 | 352,3461 | 0,07011 | 1 | 0,284787 |
| -2,25783 | 421 | 191,8717 | 0,025083 | 1 | 0,218351 |
| 0,254294 | 420 | 388,2592 | 0,799403 | 1 | 0,969427 |
| -0,67014 | 367 | 340,1654 | 0,503223 | 1 | 0,793601 |
| -1,30083 | 280 | 262,5982 | 0,194458 | 1 | 0,62543 |
| 0,27096 | 421 | 378,1717 | 0,786569 | 1 | 0,917552 |
| 1,262787 | 421 | 198,8577 | 0,208145 | 1 | 0,508069 |
| 1,221622 | 420 | 405,3664 | 0,222561 | 1 | 0,943055 |
| 1,610653 | 367 | 352,1818 | 0,108151 | 1 | 0,435251 |
| 2,139969 | 280 | 260,341 | 0,033287 | 1 | 0,329568 |
| 0,730309 | 421 | 369,6667 | 0,465664 | 1 | 0,727964 |
| 0,455823 | 421 | 198,0936 | 0,649017 | 1 | 0,839358 |

| | | | | | |
|---|---|---|---|---|---|
| 0,304282 | 420 | 398,5312 | 0,761072 | 1 | 0,969427 |
| -1,0914 | 367 | 348,2116 | 0,275852 | 1 | 0,611884 |
| 0,76735 | 280 | 261,3146 | 0,443566 | 1 | 0,810247 |
| -1,58763 | 421 | 315,2332 | 0,113373 | 1 | 0,363517 |
| -1,58112 | 421 | 180,213 | 0,115605 | 1 | 0,392379 |
| -1,66474 | 420 | 339,1952 | 0,096888 | 1 | 0,848246 |
| -2,02569 | 367 | 305,2445 | 0,043666 | 1 | 0,299994 |
| -1,03573 | 280 | 257,1793 | 0,3013 | 1 | 0,723617 |
| -1,95092 | 421 | 353,3063 | 0,051857 | 1 | 0,253727 |
| 0,239969 | 421 | 191,1091 | 0,810611 | 1 | 0,926063 |
| -0,04486 | 420 | 386,3956 | 0,96424 | 1 | 0,996924 |
| 0,681067 | 367 | 340,8079 | 0,496291 | 1 | 0,793601 |
| -0,82446 | 280 | 254,2183 | 0,410449 | 1 | 0,788151 |
| -0,48015 | 421 | 332,7248 | 0,631436 | 1 | 0,83455 |
| -0,66118 | 421 | 184,4558 | 0,509324 | 1 | 0,755464 |
| -0,486 | 420 | 366,3563 | 0,627254 | 1 | 0,964878 |
| 0,494987 | 367 | 336,8544 | 0,620932 | 1 | 0,875495 |
| -1,75981 | 280 | 260,1482 | 0,079616 | 1 | 0,453386 |
| -3,02448 | 421 | 307,5152 | 0,002701 | 1 | 0,073997 |
| -1,48378 | 421 | 178,1607 | 0,139634 | 1 | 0,423351 |
| -0,16519 | 420 | 341,0265 | 0,868894 | 1 | 0,980994 |
| -0,90057 | 367 | 302,1581 | 0,368532 | 1 | 0,678946 |
| 0,815442 | 280 | 253,8128 | 0,415584 | 1 | 0,791764 |
| -0,79828 | 421 | 319,5741 | 0,425298 | 1 | 0,6997 |
| 0,637518 | 421 | 178,7637 | 0,524603 | 1 | 0,766525 |
| -1,00467 | 420 | 352,7199 | 0,315743 | 1 | 0,943897 |
| -0,2495 | 367 | 316,7267 | 0,803139 | 1 | 0,950149 |
| 1,16777 | 280 | 255,7753 | 0,243987 | 1 | 0,673421 |
| 0,229613 | 421 | 365,1928 | 0,818521 | 1 | 0,93025 |
| 0,593121 | 421 | 195,503 | 0,553786 | 1 | 0,778237 |
| 1,092409 | 420 | 396,058 | 0,275317 | 1 | 0,943897 |
| 1,420799 | 367 | 340,2012 | 0,156291 | 1 | 0,502197 |
| -2,30972 | 280 | 259,0165 | 0,02169 | 1 | 0,305489 |
| 1,099099 | 421 | 361,0444 | 0,272457 | 1 | 0,567902 |
| 0,079346 | 421 | 195,1642 | 0,936839 | 1 | 0,97636 |
| 1,477949 | 420 | 391,41 | 0,140226 | 1 | 0,899233 |
| -0,43003 | 367 | 344,2559 | 0,667445 | 1 | 0,894765 |
| -2,36306 | 280 | 270,2607 | 0,018833 | 1 | 0,281002 |
| -0,5839 | 421 | 346,2844 | 0,559669 | 1 | 0,79269 |
| -0,59781 | 421 | 190,8458 | 0,550673 | 1 | 0,777451 |
| -1,54606 | 420 | 382,3073 | 0,122918 | 1 | 0,87201 |
| -2,20349 | 367 | 331,6803 | 0,028247 | 1 | 0,257802 |
| -0,85277 | 280 | 259,2637 | 0,394573 | 1 | 0,782194 |
| -1,31727 | 421 | 313,6879 | 0,188709 | 1 | 0,472953 |
| 0,238304 | 421 | 176,8705 | 0,811921 | 1 | 0,926063 |
| -0,4142 | 420 | 347,8237 | 0,678985 | 1 | 0,96781 |
| -0,1491 | 367 | 304,4482 | 0,881573 | 1 | 0,962453 |
| 0,0641 | 280 | 252,9358 | 0,948941 | 1 | 0,982409 |

| | | | | | |
|---|---|---|---|---|---|
| 0,972789 | 421 | 347,6045 | 0,331335 | 1 | 0,619505 |
| 0,152964 | 421 | 191,6013 | 0,878587 | 1 | 0,955289 |
| 2,002702 | 420 | 380,2604 | 0,045919 | 1 | 0,846132 |
| -1,24931 | 367 | 345,3198 | 0,212399 | 1 | 0,563532 |
| -1,17961 | 280 | 265,7326 | 0,239209 | 1 | 0,662939 |
| 1,122761 | 421 | 372,4808 | 0,262262 | 1 | 0,555878 |
| 0,162918 | 421 | 199,333 | 0,870748 | 1 | 0,949506 |
| -2,1264 | 420 | 402,5338 | 0,034078 | 1 | 0,828713 |
| 0,066857 | 367 | 349,7993 | 0,946734 | 1 | 0,979539 |
| 0,715536 | 280 | 266,5051 | 0,474904 | 1 | 0,824256 |
| -1,46712 | 421 | 376,1211 | 0,143179 | 1 | 0,412154 |
| -1,59048 | 421 | 202,0726 | 0,11329 | 1 | 0,391524 |
| -0,45664 | 420 | 405,346 | 0,648174 | 1 | 0,964878 |
| -1,7436 | 367 | 354,7459 | 0,082094 | 1 | 0,381841 |
| -1,34708 | 280 | 272,0142 | 0,179075 | 1 | 0,605081 |
| -1,7904 | 421 | 354,1745 | 0,074244 | 1 | 0,294383 |
| -0,7235 | 421 | 193,2991 | 0,470249 | 1 | 0,731044 |
| -0,37224 | 420 | 387,5692 | 0,709916 | 1 | 0,968866 |
| -1,14199 | 367 | 343,3225 | 0,254255 | 1 | 0,609149 |
| 0,168636 | 280 | 264,8536 | 0,866212 | 1 | 0,969656 |
| -0,61678 | 421 | 404,6322 | 0,537724 | 1 | 0,778195 |
| 0,719411 | 421 | 217,0148 | 0,472662 | 1 | 0,732067 |
| -0,01637 | 420 | 415,8555 | 0,98695 | 1 | 0,997342 |
| 0,643073 | 367 | 362,2158 | 0,520584 | 1 | 0,802988 |
| -0,92173 | 280 | 271,3743 | 0,35749 | 1 | 0,761846 |
| -0,94312 | 421 | 353,0844 | 0,346265 | 1 | 0,635592 |
| -0,36732 | 421 | 191,0447 | 0,713787 | 1 | 0,873114 |
| -0,2784 | 420 | 386,2254 | 0,780852 | 1 | 0,969427 |
| -0,58062 | 367 | 332,5641 | 0,561889 | 1 | 0,834252 |
| -1,98643 | 280 | 261,159 | 0,048031 | 1 | 0,393265 |
| -1,84142 | 421 | 314,4863 | 0,066501 | 1 | 0,278382 |
| -2,026 | 421 | 177,6396 | 0,044261 | 1 | 0,277922 |
| -1,12171 | 420 | 348,2901 | 0,262758 | 1 | 0,943897 |
| -2,74663 | 367 | 320,9487 | 0,006361 | 1 | 0,178183 |
| -0,76765 | 280 | 265,5215 | 0,443375 | 1 | 0,810247 |
| -1,47178 | 421 | 346,5676 | 0,141989 | 1 | 0,412154 |
| -1,65731 | 421 | 191,2069 | 0,099097 | 1 | 0,379659 |
| -0,87178 | 420 | 378,7759 | 0,38388 | 1 | 0,95743 |
| 0,326565 | 367 | 338,0557 | 0,744199 | 1 | 0,923669 |
| -0,12453 | 280 | 267,6921 | 0,90099 | 1 | 0,975243 |
| -0,64883 | 421 | 379,2001 | 0,516838 | 1 | 0,767364 |
| -1,50925 | 421 | 201,9176 | 0,132797 | 1 | 0,414338 |
| 1,562348 | 420 | 406,9784 | 0,118983 | 1 | 0,858144 |
| -1,07754 | 367 | 352,0664 | 0,281978 | 1 | 0,61675 |
| 0,321498 | 280 | 266,7351 | 0,748085 | 1 | 0,942071 |
| -2,32704 | 421 | 374,352 | 0,020496 | 1 | 0,16696 |
| -2,06635 | 421 | 199,2497 | 0,040088 | 1 | 0,26998 |
| -2,61441 | 420 | 400,2246 | 0,009276 | 1 | 0,582436 |

| | | | | | |
|---|---|---|---|---|---|
| -1,16259 | 367 | 352,2621 | 0,245782 | 1 | 0,599343 |
| -0,49668 | 280 | 266,0922 | 0,619826 | 1 | 0,894711 |
| -0,15257 | 421 | 415,6387 | 0,878813 | 1 | 0,95861 |
| 0,38512 | 421 | 261,4344 | 0,700462 | 1 | 0,865528 |
| -0,94479 | 420 | 398,9652 | 0,345336 | 1 | 0,947373 |
| 2,383476 | 367 | 350,0643 | 0,017683 | 1 | 0,216225 |
| 0,202392 | 280 | 274,021 | 0,83976 | 1 | 0,964667 |
| -3,28165 | 421 | 340,276 | 0,001139 | 0,1754 | 0,064989 |
| -3,39954 | 421 | 189,5102 | 8,23E-04 | 0,124263 | 0,065272 |
| -0,41378 | 420 | 383,2107 | 0,679269 | 1 | 0,96781 |
| -2,98335 | 367 | 323,1125 | 0,003068 | 0,466364 | 0,176492 |
| -2,55813 | 280 | 266,0883 | 0,011078 | 1 | 0,234919 |
| 1,160722 | 421 | 329,6004 | 0,246595 | 1 | 0,544195 |
| 1,55599 | 421 | 185,0252 | 0,121419 | 1 | 0,402151 |
| 1,442342 | 420 | 363,3061 | 0,150067 | 1 | 0,914005 |
| 0,590506 | 367 | 308,0536 | 0,555284 | 1 | 0,830257 |
| 0,469567 | 280 | 253,8273 | 0,639068 | 1 | 0,907449 |
| -1,93196 | 421 | 317,5547 | 0,054254 | 1 | 0,259162 |
| -1,80913 | 421 | 181,2702 | 0,072087 | 1 | 0,341588 |
| 0,795946 | 420 | 354,0451 | 0,426597 | 1 | 0,95743 |
| -1,90695 | 367 | 315,3404 | 0,057435 | 1 | 0,338985 |
| -2,47258 | 280 | 255,6438 | 0,014065 | 1 | 0,258495 |
| -2,74038 | 421 | 341,5526 | 0,006459 | 1 | 0,102724 |
| -1,59712 | 421 | 189,3938 | 0,111905 | 1 | 0,391524 |
| -0,85369 | 420 | 381,7627 | 0,39381 | 1 | 0,95743 |
| -1,12562 | 367 | 337,8966 | 0,261124 | 1 | 0,609149 |
| -0,12375 | 280 | 266,7344 | 0,901604 | 1 | 0,975243 |
| 1,099963 | 421 | 371,2548 | 0,272061 | 1 | 0,567861 |
| 1,563893 | 421 | 199,0991 | 0,119431 | 1 | 0,396436 |
| 0,859176 | 420 | 399,8722 | 0,390758 | 1 | 0,95743 |
| 0,557547 | 367 | 355,4802 | 0,577504 | 1 | 0,843313 |
| -0,59949 | 280 | 265,618 | 0,549359 | 1 | 0,852208 |
| 1,613448 | 421 | 381,372 | 0,107474 | 1 | 0,353632 |
| 2,17241 | 421 | 203,9067 | 0,030978 | 1 | 0,241423 |
| 1,381552 | 420 | 407,364 | 0,167867 | 1 | 0,914005 |
| 0,597262 | 367 | 354,6015 | 0,550713 | 1 | 0,829925 |
| 0,927101 | 280 | 269,9736 | 0,354702 | 1 | 0,761846 |
| 1,497911 | 421 | 358,5509 | 0,135036 | 1 | 0,407576 |
| 1,144961 | 421 | 191,2021 | 0,253656 | 1 | 0,553199 |
| 0,885668 | 420 | 391,0911 | 0,376341 | 1 | 0,95743 |
| 1,37911 | 367 | 324,8728 | 0,168809 | 1 | 0,514971 |
| 1,542249 | 280 | 267,6845 | 0,124194 | 1 | 0,535729 |
| -0,83445 | 421 | 403,5479 | 0,404518 | 1 | 0,695107 |
| 0,873218 | 421 | 215,8994 | 0,383514 | 1 | 0,659083 |
| 0,094293 | 420 | 415,6888 | 0,924922 | 1 | 0,989204 |
| 0,811175 | 367 | 362,1643 | 0,417798 | 1 | 0,729573 |
| -1,21903 | 280 | 271,7878 | 0,223889 | 1 | 0,654014 |
| -2,65214 | 421 | 349,6009 | 0,008363 | 1 | 0,112451 |

| | | | | | |
|---|---|---|---|---|---|
| -1,69163 | 421 | 191,7417 | 0,092341 | 1 | 0,373113 |
| 0,840876 | 420 | 388,0467 | 0,400935 | 1 | 0,95743 |
| -1,86467 | 367 | 342,7001 | 0,063083 | 1 | 0,34994 |
| -2,15499 | 280 | 261,4301 | 0,032076 | 1 | 0,329568 |
| -1,39693 | 421 | 332,1548 | 0,163366 | 1 | 0,438064 |
| -0,66862 | 421 | 184,61 | 0,504575 | 1 | 0,753145 |
| 0,63597 | 420 | 367,7207 | 0,525191 | 1 | 0,960096 |
| 0,316595 | 367 | 326,4676 | 0,751754 | 1 | 0,924056 |
| 1,157922 | 280 | 254,8011 | 0,24798 | 1 | 0,677438 |
| 0,494183 | 421 | 384,5589 | 0,621459 | 1 | 0,828413 |
| 0,262392 | 421 | 204,2007 | 0,793284 | 1 | 0,919599 |
| 0,037084 | 420 | 408,9146 | 0,970436 | 1 | 0,997342 |
| 0,907381 | 367 | 356,4008 | 0,364819 | 1 | 0,677906 |
| -1,13647 | 280 | 265,8415 | 0,256781 | 1 | 0,680087 |
| -1,75403 | 421 | 309,9246 | 0,080414 | 1 | 0,303719 |
| -2,26289 | 421 | 179,6919 | 0,024839 | 1 | 0,218351 |
| -0,03545 | 420 | 349,6466 | 0,971743 | 1 | 0,997342 |
| 0,169905 | 367 | 301,3502 | 0,865199 | 1 | 0,959636 |
| 0,368474 | 280 | 248,5892 | 0,712833 | 1 | 0,928432 |
| 2,714147 | 421 | 354,7978 | 0,006969 | 1 | 0,105025 |
| 1,452537 | 421 | 191,7772 | 0,147986 | 1 | 0,428784 |
| -1,21191 | 420 | 385,582 | 0,226288 | 1 | 0,943055 |
| 0,684721 | 367 | 344,1366 | 0,493981 | 1 | 0,793601 |
| 0,172879 | 280 | 267,8989 | 0,862877 | 1 | 0,967527 |
| 1,016078 | 421 | 362,7644 | 0,310269 | 1 | 0,605668 |
| 1,3309 | 421 | 193,7298 | 0,184786 | 1 | 0,473593 |
| -1,00811 | 420 | 394,5444 | 0,314018 | 1 | 0,943897 |
| 1,567514 | 367 | 348,6905 | 0,117902 | 1 | 0,445308 |
| 0,748935 | 280 | 271,7611 | 0,454544 | 1 | 0,814707 |
| -0,72744 | 421 | 387,8656 | 0,467395 | 1 | 0,728402 |
| -1,31611 | 421 | 205,3847 | 0,189603 | 1 | 0,476746 |
| -0,17728 | 420 | 410,9483 | 0,859377 | 1 | 0,976988 |
| 1,498166 | 367 | 349,807 | 0,134992 | 1 | 0,477542 |
| 0,535972 | 280 | 268,4681 | 0,592421 | 1 | 0,876993 |
| -1,87243 | 421 | 368,0659 | 0,06194 | 1 | 0,271708 |
| -2,92195 | 421 | 196,5668 | 0,003886 | 1 | 0,118336 |
| -1,23929 | 420 | 395,4533 | 0,215971 | 1 | 0,943055 |
| -2,43606 | 367 | 346,6591 | 0,015351 | 1 | 0,204372 |
| -1,03055 | 280 | 265,8875 | 0,30369 | 1 | 0,726446 |
| -0,8981 | 421 | 314,5915 | 0,369819 | 1 | 0,661896 |
| -0,12768 | 421 | 177,5032 | 0,898545 | 1 | 0,960359 |
| 0,386054 | 420 | 348,9735 | 0,699692 | 1 | 0,968866 |
| 0,787723 | 367 | 300,1543 | 0,43148 | 1 | 0,73975 |
| 0,892836 | 280 | 254,865 | 0,372787 | 1 | 0,763914 |
| 2,071528 | 421 | 375,7018 | 0,038992 | 1 | 0,222577 |
| 1,16732 | 421 | 200,7128 | 0,244466 | 1 | 0,54418 |
| -0,27274 | 420 | 404,7535 | 0,785192 | 1 | 0,969427 |
| 1,499933 | 367 | 346,7505 | 0,134542 | 1 | 0,477542 |

| | | | | | |
|---|---|---|---|---|---|
| 0,229347 | 280 | 267,747 | 0,818774 | 1 | 0,95778 |
| -0,58702 | 421 | 355,5486 | 0,557564 | 1 | 0,791195 |
| 1,134843 | 421 | 191,4729 | 0,25786 | 1 | 0,556025 |
| 0,818212 | 420 | 388,1265 | 0,413738 | 1 | 0,95743 |
| -0,06821 | 367 | 335,5662 | 0,945663 | 1 | 0,979539 |
| 1,088593 | 280 | 271,5438 | 0,277299 | 1 | 0,701156 |
| 0,337818 | 421 | 359,0232 | 0,735698 | 1 | 0,893273 |
| 0,510615 | 421 | 192,8472 | 0,610205 | 1 | 0,82272 |
| -0,2404 | 420 | 391,3205 | 0,810147 | 1 | 0,969427 |
| -1,02657 | 367 | 350,962 | 0,305332 | 1 | 0,637666 |
| -2,53213 | 280 | 262,1566 | 0,011922 | 1 | 0,239766 |
| -2,27824 | 421 | 352,3344 | 0,02331 | 1 | 0,177419 |
| -1,66332 | 421 | 192,555 | 0,097875 | 1 | 0,379251 |
| -0,85795 | 420 | 384,1122 | 0,391455 | 1 | 0,95743 |
| -1,03494 | 367 | 342,512 | 0,301426 | 1 | 0,633826 |
| -0,76631 | 280 | 267,1718 | 0,444167 | 1 | 0,810362 |
| -2,29288 | 421 | 346,8862 | 0,022453 | 1 | 0,177154 |
| -2,22189 | 421 | 188,8651 | 0,027478 | 1 | 0,22628 |
| -4,14205 | 420 | 381,1527 | 4,24E-05 | 0,045775 | 0,035635 |
| 0,448782 | 367 | 328,0538 | 0,653885 | 1 | 0,889355 |
| 0,38252 | 280 | 256,5007 | 0,702392 | 1 | 0,92124 |
| 0,698778 | 421 | 329,5283 | 0,485184 | 1 | 0,745445 |
| -0,1327 | 421 | 183,9718 | 0,894579 | 1 | 0,958882 |
| -1,8914 | 420 | 364,445 | 0,059364 | 1 | 0,848246 |
| 2,184174 | 367 | 307,5871 | 0,029704 | 1 | 0,260259 |
| 0,298507 | 280 | 257,5821 | 0,765557 | 1 | 0,945436 |
| -2,08248 | 421 | 311,4314 | 0,038114 | 1 | 0,220722 |
| 0,46634 | 421 | 180,7495 | 0,641534 | 1 | 0,838647 |
| 0,127657 | 420 | 342,8673 | 0,898495 | 1 | 0,98503 |
| -1,21367 | 367 | 288,5071 | 0,225867 | 1 | 0,576775 |
| -0,46099 | 280 | 276 | 0,64517 | 1 | 0,907804 |
| -0,63665 | 421 | 345,1906 | 0,524772 | 1 | 0,770244 |
| -0,79767 | 421 | 189,1186 | 0,42606 | 1 | 0,694133 |
| -0,184 | 420 | 379,5538 | 0,854114 | 1 | 0,973894 |
| -0,15455 | 367 | 325,9635 | 0,87727 | 1 | 0,962453 |
| 2,120631 | 280 | 257,7218 | 0,034908 | 1 | 0,337221 |
| -0,36554 | 421 | 380,7206 | 0,714911 | 1 | 0,881149 |
| 0,361491 | 421 | 202,0871 | 0,71811 | 1 | 0,874852 |
| -1,79991 | 420 | 405,4388 | 0,072618 | 1 | 0,848246 |
| 0,976722 | 367 | 349,8028 | 0,329382 | 1 | 0,656585 |
| 0,135269 | 280 | 263,5934 | 0,892502 | 1 | 0,975243 |
| 0,734081 | 421 | 320,3989 | 0,463436 | 1 | 0,727964 |
| 0,509678 | 421 | 180,8014 | 0,610898 | 1 | 0,82272 |
| -0,29565 | 420 | 359,3506 | 0,767664 | 1 | 0,969427 |
| 0,194709 | 367 | 319,3758 | 0,845745 | 1 | 0,956935 |
| -0,45935 | 280 | 257,975 | 0,646366 | 1 | 0,907804 |
| -0,74517 | 421 | 355,4466 | 0,456661 | 1 | 0,722128 |
| 0,746518 | 421 | 193,4883 | 0,456261 | 1 | 0,716981 |

| | | | | | |
|---|---|---|---|---|---|
| 0,927387 | 420 | 386,5321 | 0,354304 | 1 | 0,947373 |
| -0,25338 | 367 | 342,7167 | 0,800126 | 1 | 0,950149 |
| 0,118571 | 280 | 255,1671 | 0,905709 | 1 | 0,975308 |
| 1,594365 | 421 | 401,4231 | 0,111641 | 1 | 0,358728 |
| 1,423244 | 421 | 217,054 | 0,156101 | 1 | 0,438625 |
| 0,380026 | 420 | 415,4158 | 0,70412 | 1 | 0,968866 |
| 1,346384 | 367 | 362,5874 | 0,17902 | 1 | 0,526202 |
| 0,088059 | 280 | 273,046 | 0,929894 | 1 | 0,979504 |
| 1,11571 | 421 | 325,9717 | 0,265368 | 1 | 0,560098 |
| 0,739333 | 421 | 180,4969 | 0,460665 | 1 | 0,721079 |
| -1,35242 | 420 | 352,1151 | 0,177108 | 1 | 0,9199 |
| 1,439621 | 367 | 307,3909 | 0,150992 | 1 | 0,499998 |
| 1,338419 | 280 | 263,3115 | 0,181915 | 1 | 0,607861 |
| -0,50953 | 421 | 346,5986 | 0,610708 | 1 | 0,825172 |
| 0,79251 | 421 | 190,1199 | 0,429051 | 1 | 0,694836 |
| 0,459433 | 420 | 381,8699 | 0,646185 | 1 | 0,964878 |
| 1,479507 | 367 | 337,57 | 0,139937 | 1 | 0,488078 |
| -0,32012 | 280 | 263,1112 | 0,749134 | 1 | 0,942071 |
| -1,99246 | 421 | 346,9233 | 0,047104 | 1 | 0,239816 |
| -1,52922 | 421 | 190,2785 | 0,127869 | 1 | 0,410224 |
| -0,69949 | 420 | 382,8631 | 0,484673 | 1 | 0,960096 |
| -1,36691 | 367 | 330,7436 | 0,172582 | 1 | 0,522252 |
| -0,39715 | 280 | 264,3599 | 0,691576 | 1 | 0,919522 |
| -2,63715 | 421 | 331,1404 | 0,008755 | 1 | 0,113397 |
| -1,87126 | 421 | 185,6709 | 0,062882 | 1 | 0,321231 |
| -0,29101 | 420 | 367,0561 | 0,771205 | 1 | 0,969427 |
| 0,920181 | 367 | 328,717 | 0,358153 | 1 | 0,674813 |
| -0,82416 | 280 | 261,2734 | 0,410602 | 1 | 0,788151 |
| 1,143492 | 421 | 394,0953 | 0,253528 | 1 | 0,546261 |
| 1,480288 | 421 | 210,8077 | 0,140289 | 1 | 0,423351 |
| 0,524664 | 420 | 413,3342 | 0,600098 | 1 | 0,964878 |
| -1,00308 | 367 | 361,2801 | 0,316491 | 1 | 0,649799 |
| 0,222112 | 280 | 275,2924 | 0,824392 | 1 | 0,96121 |
| 0,981687 | 421 | 371,0562 | 0,326894 | 1 | 0,61749 |
| 0,172379 | 421 | 199,343 | 0,863315 | 1 | 0,947215 |
| -0,00646 | 420 | 400,3528 | 0,994846 | 1 | 0,997342 |
| -0,70487 | 367 | 345,0138 | 0,481366 | 1 | 0,785415 |
| 0,603958 | 280 | 264,9879 | 0,546388 | 1 | 0,851507 |
| 0,205692 | 421 | 299,4013 | 0,837171 | 1 | 0,93721 |
| 1,793954 | 421 | 174,5084 | 0,074552 | 1 | 0,344631 |
| 1,750046 | 420 | 330,4159 | 0,081039 | 1 | 0,848246 |
| -0,80242 | 367 | 320,1358 | 0,422903 | 1 | 0,732546 |
| -1,67875 | 280 | 254,4562 | 0,094428 | 1 | 0,474774 |
| -0,82014 | 421 | 330,3024 | 0,412726 | 1 | 0,695339 |
| -0,59191 | 421 | 185,6175 | 0,554629 | 1 | 0,778237 |
| -1,10268 | 420 | 367,1121 | 0,270889 | 1 | 0,943897 |
| -1,22952 | 367 | 325,4803 | 0,219765 | 1 | 0,572696 |
| -1,28136 | 280 | 256,4836 | 0,201224 | 1 | 0,631761 |

| | | | | | |
|---|---|---|---|---|---|
| 0,064371 | 421 | 394,0978 | 0,948707 | 1 | 0,983289 |
| 0,68725 | 421 | 211,9926 | 0,492676 | 1 | 0,746275 |
| 1,423952 | 420 | 413,4498 | 0,155215 | 1 | 0,914005 |
| -0,4208 | 367 | 358,7325 | 0,674151 | 1 | 0,898779 |
| -0,7689 | 280 | 273,1425 | 0,442617 | 1 | 0,810247 |
| -1,13292 | 421 | 385,8972 | 0,257951 | 1 | 0,550898 |
| -0,86084 | 421 | 206,2284 | 0,390323 | 1 | 0,664652 |
| -0,26663 | 420 | 408,5456 | 0,789892 | 1 | 0,969427 |
| -0,48931 | 367 | 349,8927 | 0,624928 | 1 | 0,878514 |
| -2,67518 | 280 | 266,5249 | 0,007931 | 1 | 0,199202 |
| 1,92449 | 421 | 401,0312 | 0,055 | 1 | 0,259829 |
| 2,997557 | 421 | 217,6185 | 0,003038 | 1 | 0,114447 |
| 0,356727 | 420 | 415,6213 | 0,721477 | 1 | 0,969427 |
| 2,254509 | 367 | 361,6155 | 0,024762 | 1 | 0,247087 |
| 1,238043 | 280 | 273,772 | 0,216761 | 1 | 0,650539 |
| 1,695075 | 421 | 371,3632 | 0,090899 | 1 | 0,32265 |
| -0,92185 | 421 | 198,5972 | 0,357725 | 1 | 0,643758 |
| -0,94438 | 420 | 400,3706 | 0,345543 | 1 | 0,947373 |
| 0,222736 | 367 | 342,2036 | 0,823874 | 1 | 0,953906 |
| -1,93314 | 280 | 255,6907 | 0,054322 | 1 | 0,40654 |
| 0,918938 | 421 | 349,1898 | 0,358762 | 1 | 0,648267 |
| 1,755169 | 421 | 187,0995 | 0,080867 | 1 | 0,360416 |
| 0,321723 | 420 | 384,7728 | 0,747837 | 1 | 0,969427 |
| 2,691169 | 367 | 337,8622 | 0,007474 | 1 | 0,180403 |
| 1,652043 | 280 | 259,7634 | 0,099734 | 1 | 0,480557 |
| 0,332436 | 421 | 327,3152 | 0,739773 | 1 | 0,893273 |
| 1,421466 | 421 | 183,3112 | 0,15688 | 1 | 0,438625 |
| -0,05386 | 420 | 362,5821 | 0,957077 | 1 | 0,996074 |
| 1,324826 | 367 | 314,6971 | 0,18619 | 1 | 0,535663 |
| 2,112732 | 280 | 257,923 | 0,035585 | 1 | 0,339407 |
| 1,85311 | 421 | 388,4864 | 0,064625 | 1 | 0,27589 |
| 2,013058 | 421 | 205,9498 | 0,04541 | 1 | 0,281076 |
| 1,208527 | 420 | 411,2248 | 0,227539 | 1 | 0,943055 |
| 2,299863 | 367 | 355,2335 | 0,022035 | 1 | 0,230605 |
| 1,035334 | 280 | 270,6263 | 0,301437 | 1 | 0,723617 |
| 2,134795 | 421 | 401,0287 | 0,033383 | 1 | 0,20962 |
| 1,941579 | 421 | 216,1203 | 0,053488 | 1 | 0,302776 |
| 0,606428 | 420 | 415,6551 | 0,544562 | 1 | 0,960096 |
| 2,402973 | 367 | 361,3296 | 0,016766 | 1 | 0,210552 |
| 0,336005 | 280 | 272,9266 | 0,737125 | 1 | 0,939009 |
| 0,589174 | 421 | 401,4582 | 0,556076 | 1 | 0,790572 |
| -0,15414 | 421 | 215,4966 | 0,877643 | 1 | 0,954952 |
| 0,558898 | 420 | 415,2225 | 0,576533 | 1 | 0,964878 |
| -1,18715 | 367 | 360,9753 | 0,235949 | 1 | 0,585627 |
| -1,12168 | 280 | 271,3139 | 0,262992 | 1 | 0,688875 |
| -3,38322 | 421 | 302,3081 | 8,11E-04 | 0,861102 | 0,053127 |
| -1,9775 | 421 | 173,5462 | 0,049569 | 1 | 0,289197 |
| -1,1503 | 420 | 337,0392 | 0,250834 | 1 | 0,943897 |

| | | | | | |
|---|---|---|---|---|---|
| -1,90897 | 367 | 293,4957 | 0,057241 | 1 | 0,338985 |
| -2,31782 | 280 | 251,4259 | 0,021263 | 1 | 0,30229 |
| -0,21439 | 421 | 377,5464 | 0,830362 | 1 | 0,936611 |
| 0,180092 | 421 | 201,4922 | 0,857261 | 1 | 0,945992 |
| 0,717212 | 420 | 404,6395 | 0,473657 | 1 | 0,960096 |
| -0,67373 | 367 | 344,5443 | 0,500935 | 1 | 0,793601 |
| 1,975203 | 280 | 263,1081 | 0,04929 | 1 | 0,39442 |
| -0,50207 | 421 | 291,2411 | 0,615996 | 1 | 0,826526 |
| -0,546 | 421 | 174,6541 | 0,585763 | 1 | 0,802496 |
| -2,21804 | 420 | 315,5389 | 0,027264 | 1 | 0,767829 |
| -0,47107 | 367 | 363 | 0,637872 | 1 | 0,880707 |
| 0,546597 | 280 | 276 | 0,585097 | 1 | 0,87561 |
| -0,76291 | 421 | 345,9956 | 0,446037 | 1 | 0,714643 |
| -1,2624 | 421 | 190,0914 | 0,208352 | 1 | 0,508069 |
| -0,49219 | 420 | 382,0686 | 0,622866 | 1 | 0,964878 |
| -0,6996 | 367 | 336,5119 | 0,484661 | 1 | 0,786044 |
| -0,95178 | 280 | 261,9341 | 0,342084 | 1 | 0,75812 |
| -1,03753 | 421 | 324,7844 | 0,300262 | 1 | 0,596172 |
| -0,22584 | 421 | 183,9069 | 0,821573 | 1 | 0,929512 |
| -0,16804 | 420 | 359,9738 | 0,866649 | 1 | 0,979775 |
| 1,771398 | 367 | 313,6095 | 0,077466 | 1 | 0,380263 |
| -0,9461 | 280 | 262,0198 | 0,34497 | 1 | 0,758934 |
| -1,8193 | 421 | 367,1458 | 0,069681 | 1 | 0,283808 |
| -1,52072 | 421 | 198,1063 | 0,129924 | 1 | 0,410868 |
| 0,613731 | 420 | 400,4036 | 0,539742 | 1 | 0,960096 |
| -0,79811 | 367 | 351,6991 | 0,425347 | 1 | 0,733464 |
| 1,455454 | 280 | 271,4612 | 0,146699 | 1 | 0,555466 |
| -1,37575 | 421 | 307,9567 | 0,169898 | 1 | 0,442288 |
| -0,1746 | 421 | 175,2748 | 0,861599 | 1 | 0,947215 |
| -0,99132 | 420 | 340,4193 | 0,322233 | 1 | 0,945573 |
| -0,96728 | 367 | 296,5628 | 0,334192 | 1 | 0,661796 |
| 0,193714 | 280 | 276 | 0,846542 | 1 | 0,965181 |
| -0,01774 | 421 | 406,6098 | 0,985857 | 1 | 0,998043 |
| -0,44963 | 421 | 219,199 | 0,653419 | 1 | 0,841107 |
| 0,578928 | 420 | 415,9553 | 0,562951 | 1 | 0,961052 |
| 1,253318 | 367 | 362,2971 | 0,210898 | 1 | 0,563436 |
| 1,30834 | 280 | 274,9114 | 0,191851 | 1 | 0,62543 |
| -0,16423 | 421 | 366,223 | 0,869643 | 1 | 0,955213 |
| 0,923578 | 421 | 196,9014 | 0,356836 | 1 | 0,643758 |
| -0,59951 | 420 | 396,3547 | 0,549176 | 1 | 0,960096 |
| 0,392269 | 367 | 340,3608 | 0,695105 | 1 | 0,904597 |
| -0,4417 | 280 | 265,5092 | 0,659066 | 1 | 0,908644 |
| -0,17217 | 421 | 336,603 | 0,863407 | 1 | 0,951261 |
| -1,58942 | 421 | 187,868 | 0,113646 | 1 | 0,391524 |
| -0,77753 | 420 | 372,3283 | 0,43734 | 1 | 0,960096 |
| 0,479009 | 367 | 331,1014 | 0,632249 | 1 | 0,880707 |
| -1,27821 | 280 | 264,4032 | 0,202295 | 1 | 0,632429 |
| -2,17828 | 421 | 320,8056 | 0,030113 | 1 | 0,200565 |

| | | | | | |
|---|---|---|---|---|---|
| -0,76177 | 421 | 183,3768 | 0,447175 | 1 | 0,710837 |
| -0,8012 | 420 | 356,4557 | 0,423548 | 1 | 0,95743 |
| -0,32924 | 367 | 303,4355 | 0,742205 | 1 | 0,923669 |
| -0,92381 | 280 | 254,4028 | 0,356458 | 1 | 0,761846 |
| 2,723148 | 421 | 307,9625 | 0,006835 | 1 | 0,10482 |
| 2,686584 | 421 | 177,5377 | 0,007904 | 1 | 0,148102 |
| 1,337537 | 420 | 343,7054 | 0,181932 | 1 | 0,923408 |
| 2,16665 | 367 | 290,4388 | 0,031075 | 1 | 0,266905 |
| 0,609109 | 280 | 251,7549 | 0,543001 | 1 | 0,849885 |
| -3,32139 | 421 | 332,7553 | 9,95E-04 | 1 | 0,062491 |
| -4,1564 | 421 | 189,0096 | 4,90E-05 | 0,052783 | 0,018447 |
| -0,49448 | 420 | 383,0098 | 0,621254 | 1 | 0,964878 |
| -2,52674 | 367 | 344,2422 | 0,01196 | 1 | 0,189546 |
| -3,84027 | 280 | 262,4497 | 1,54E-04 | 0,165998 | 0,046455 |
| -1,06946 | 421 | 340,8429 | 0,285621 | 1 | 0,583534 |
| -0,81493 | 421 | 187,9591 | 0,416144 | 1 | 0,686888 |
| 0,845242 | 420 | 377,0386 | 0,398512 | 1 | 0,95743 |
| -0,93113 | 367 | 336,2102 | 0,352454 | 1 | 0,67234 |
| -1,87146 | 280 | 260,094 | 0,062405 | 1 | 0,422605 |
| -2,95403 | 421 | 332,8081 | 0,00336 | 1 | 0,081668 |
| -1,79019 | 421 | 184,9633 | 0,07506 | 1 | 0,345067 |
| -0,82532 | 420 | 370,5713 | 0,409721 | 1 | 0,95743 |
| -1,71287 | 367 | 327,8354 | 0,087681 | 1 | 0,394165 |
| -1,3733 | 280 | 259,1397 | 0,170847 | 1 | 0,59737 |
| -0,2034 | 421 | 329,5572 | 0,838949 | 1 | 0,93721 |
| 1,259562 | 421 | 185,753 | 0,209408 | 1 | 0,509818 |
| 0,416194 | 420 | 363,3091 | 0,677514 | 1 | 0,96781 |
| 2,370316 | 367 | 307,2135 | 0,01839 | 1 | 0,217108 |
| -0,08746 | 280 | 251,854 | 0,930373 | 1 | 0,979504 |
| 2,416907 | 421 | 349,6098 | 0,016164 | 1 | 0,147897 |
| 1,874837 | 421 | 188,9497 | 0,062358 | 1 | 0,319638 |
| 2,238326 | 420 | 380,7107 | 0,025777 | 1 | 0,767829 |
| 1,594632 | 367 | 330,3913 | 0,111751 | 1 | 0,438296 |
| 1,736774 | 280 | 268,9747 | 0,083572 | 1 | 0,456738 |
| -1,0886 | 421 | 329,7888 | 0,277125 | 1 | 0,57291 |
| -0,41379 | 421 | 183,6144 | 0,679509 | 1 | 0,857638 |
| -0,37903 | 420 | 363,1531 | 0,704887 | 1 | 0,968866 |
| 0,945766 | 367 | 319,8187 | 0,344982 | 1 | 0,667378 |
| 1,64937 | 280 | 257,1045 | 0,100293 | 1 | 0,480557 |
| -3,12306 | 421 | 339,8107 | 0,001944 | 1 | 0,069718 |
| -2,25939 | 421 | 188,0962 | 0,025006 | 1 | 0,218351 |
| -0,73018 | 420 | 377,9418 | 0,465732 | 1 | 0,960096 |
| -1,03139 | 367 | 336,4474 | 0,3031 | 1 | 0,636172 |
| -0,13812 | 280 | 259,9025 | 0,890256 | 1 | 0,975243 |
| 2,013068 | 421 | 361,6752 | 0,044848 | 1 | 0,236315 |
| 1,527187 | 421 | 196,8188 | 0,12832 | 1 | 0,410224 |
| 2,041224 | 420 | 396,8918 | 0,041889 | 1 | 0,846132 |
| -0,25039 | 367 | 355,6048 | 0,802434 | 1 | 0,950149 |

| | | | | | |
|---|---|---|---|---|---|
| 0,683814 | 280 | 275,5021 | 0,494668 | 1 | 0,8282 |
| -2,87777 | 421 | 304,6496 | 0,004288 | 1 | 0,087964 |
| -3,91382 | 421 | 178,5792 | 1,29E-04 | 0,138609 | 0,024311 |
| -1,6688 | 420 | 342,5535 | 0,09607 | 1 | 0,848246 |
| -0,44056 | 367 | 298,678 | 0,659849 | 1 | 0,893492 |
| -1,72116 | 280 | 252,5533 | 0,086446 | 1 | 0,458713 |
| -1,48566 | 421 | 347,8447 | 0,138274 | 1 | 0,410344 |
| -1,56854 | 421 | 188,7222 | 0,11843 | 1 | 0,396436 |
| -1,87394 | 420 | 378,2358 | 0,061709 | 1 | 0,848246 |
| -0,80451 | 367 | 336,1424 | 0,42167 | 1 | 0,731532 |
| -0,70231 | 280 | 256,0761 | 0,483124 | 1 | 0,824339 |
| -1,21371 | 421 | 400,0734 | 0,225574 | 1 | 0,518147 |
| -1,18403 | 421 | 217,1047 | 0,237696 | 1 | 0,537903 |
| 0,410251 | 420 | 415,9799 | 0,681833 | 1 | 0,968338 |
| -1,01757 | 367 | 362,8528 | 0,309559 | 1 | 0,643456 |
| 0,107536 | 280 | 273,1645 | 0,914443 | 1 | 0,977051 |
| 1,867384 | 421 | 364,2545 | 0,062651 | 1 | 0,273465 |
| 1,926779 | 421 | 195,3661 | 0,055458 | 1 | 0,309036 |
| 1,093673 | 420 | 397,3763 | 0,274761 | 1 | 0,943897 |
| 1,78608 | 367 | 336,4938 | 0,074987 | 1 | 0,376019 |
| 0,411819 | 280 | 272,5694 | 0,680796 | 1 | 0,911154 |
| -0,10295 | 421 | 323,4754 | 0,918068 | 1 | 0,966828 |
| -0,32819 | 421 | 182,9434 | 0,743143 | 1 | 0,888118 |
| -0,84312 | 420 | 358,2434 | 0,399727 | 1 | 0,95743 |
| 0,715112 | 367 | 309,6287 | 0,475079 | 1 | 0,7782 |
| -0,86004 | 280 | 260,323 | 0,390556 | 1 | 0,778529 |
| 0,981659 | 421 | 333,6805 | 0,326979 | 1 | 0,61749 |
| 0,352343 | 421 | 185,6913 | 0,724981 | 1 | 0,878959 |
| -2,2075 | 420 | 368,0317 | 0,027895 | 1 | 0,767829 |
| 3,026825 | 367 | 307,4008 | 0,00268 | 1 | 0,176492 |
| 0,784004 | 280 | 257,2645 | 0,433759 | 1 | 0,803161 |
| 1,668234 | 421 | 313,3276 | 0,096268 | 1 | 0,332744 |
| 3,179432 | 421 | 178,1614 | 0,00174 | 1 | 0,093671 |
| 1,276857 | 420 | 351,566 | 0,202495 | 1 | 0,928589 |
| 1,434997 | 367 | 309,411 | 0,152298 | 1 | 0,500093 |
| 2,267908 | 280 | 253,1815 | 0,024178 | 1 | 0,313396 |
| 2,003032 | 421 | 403,571 | 0,045842 | 1 | 0,237403 |
| 3,175973 | 421 | 219,9582 | 0,001708 | 1 | 0,093671 |
| 0,309673 | 420 | 415,9273 | 0,756965 | 1 | 0,969427 |
| 2,120593 | 367 | 362,0875 | 0,034635 | 1 | 0,280622 |
| 1,141304 | 280 | 274,7378 | 0,254737 | 1 | 0,679673 |
| 1,31989 | 421 | 373,107 | 0,187681 | 1 | 0,472953 |
| 0,875898 | 421 | 198,9616 | 0,382142 | 1 | 0,659083 |
| 1,006091 | 420 | 399,7217 | 0,31498 | 1 | 0,943897 |
| -2,10128 | 367 | 346,4253 | 0,036338 | 1 | 0,28464 |
| 0,164413 | 280 | 255,6848 | 0,869536 | 1 | 0,969656 |
| 0,32362 | 421 | 352,9531 | 0,746417 | 1 | 0,897726 |
| 0,735113 | 421 | 190,4565 | 0,463175 | 1 | 0,721079 |

| | | | | | |
|---|---|---|---|---|---|
| -0,49742 | 420 | 388,0439 | 0,619174 | 1 | 0,964878 |
| 1,549112 | 367 | 349,359 | 0,12226 | 1 | 0,456054 |
| -0,55391 | 280 | 269,5249 | 0,580101 | 1 | 0,874212 |
| -1,61574 | 421 | 354,1206 | 0,107041 | 1 | 0,352976 |
| -2,0481 | 421 | 191,3557 | 0,041916 | 1 | 0,273775 |
| 0,591525 | 420 | 389,1596 | 0,554512 | 1 | 0,960096 |
| 0,515412 | 367 | 345,9292 | 0,606594 | 1 | 0,868127 |
| 0,070468 | 280 | 269,757 | 0,943873 | 1 | 0,982175 |
| -0,92671 | 421 | 357,1822 | 0,354704 | 1 | 0,644799 |
| 0,408918 | 421 | 193,2975 | 0,683052 | 1 | 0,859821 |
| -0,51164 | 420 | 389,3511 | 0,609195 | 1 | 0,964878 |
| -1,3871 | 367 | 343,6699 | 0,16631 | 1 | 0,513583 |
| -1,8624 | 280 | 257,5908 | 0,063685 | 1 | 0,422605 |
| 0,871978 | 421 | 377,5489 | 0,383775 | 1 | 0,674066 |
| -0,13263 | 421 | 198,9619 | 0,894617 | 1 | 0,958882 |
| -0,23195 | 420 | 403,9464 | 0,816691 | 1 | 0,969427 |
| 0,314488 | 367 | 352,0458 | 0,753337 | 1 | 0,924494 |
| -1,19544 | 280 | 258,0438 | 0,233011 | 1 | 0,659852 |
| -1,46484 | 421 | 359,2408 | 0,143838 | 1 | 0,412154 |
| -2,67068 | 421 | 194,8003 | 0,00821 | 1 | 0,148102 |
| -1,58076 | 420 | 390,9006 | 0,114741 | 1 | 0,848246 |
| -0,8798 | 367 | 339,8245 | 0,379589 | 1 | 0,691076 |
| -0,62361 | 280 | 263,0257 | 0,533426 | 1 | 0,84326 |
| 0,173685 | 421 | 324,422 | 0,862221 | 1 | 0,951261 |
| -0,71251 | 421 | 180,0148 | 0,477071 | 1 | 0,734183 |
| 0,818694 | 420 | 360,4515 | 0,413502 | 1 | 0,95743 |
| 0,533762 | 367 | 312,5857 | 0,593885 | 1 | 0,858088 |
| 0,704413 | 280 | 255,4924 | 0,481817 | 1 | 0,824339 |
| -0,41919 | 421 | 380,5626 | 0,675315 | 1 | 0,859543 |
| 0,674045 | 421 | 203,7788 | 0,501047 | 1 | 0,750551 |
| -2,02518 | 420 | 403,3258 | 0,043507 | 1 | 0,846132 |
| -0,19466 | 367 | 356,3754 | 0,845771 | 1 | 0,956935 |
| 0,249228 | 280 | 273,3318 | 0,803371 | 1 | 0,95706 |
| -2,62279 | 421 | 373,3134 | 0,009079 | 1 | 0,113397 |
| 0,182014 | 421 | 200,7633 | 0,855755 | 1 | 0,945471 |
| 0,382598 | 420 | 404,4702 | 0,702219 | 1 | 0,968866 |
| -0,45877 | 367 | 355,5084 | 0,646676 | 1 | 0,884339 |
| -0,43934 | 280 | 275,9994 | 0,660762 | 1 | 0,908644 |
| -1,2988 | 421 | 308,7635 | 0,194983 | 1 | 0,481703 |
| 0,145994 | 421 | 180,196 | 0,884089 | 1 | 0,957128 |
| 0,284489 | 420 | 345,1151 | 0,776206 | 1 | 0,969427 |
| 0,670807 | 367 | 306,6172 | 0,502848 | 1 | 0,793601 |
| 0,73254 | 280 | 249,8626 | 0,464525 | 1 | 0,824256 |
| -0,26326 | 421 | 313,2854 | 0,79252 | 1 | 0,919136 |
| 0,319732 | 421 | 177,5624 | 0,749547 | 1 | 0,890124 |
| -0,62688 | 420 | 347,1274 | 0,531153 | 1 | 0,960096 |
| -0,38544 | 367 | 320,0758 | 0,700169 | 1 | 0,906491 |
| -1,36617 | 280 | 251,1344 | 0,173108 | 1 | 0,598625 |

| | | | | | |
|---|---|---|---|---|---|
| 1,016078 | 421 | 362,7644 | 0,310269 | 1 | 0,605668 |
| 1,3309 | 421 | 193,7298 | 0,184786 | 1 | 0,473593 |
| -1,00811 | 420 | 394,5444 | 0,314018 | 1 | 0,943897 |
| 1,567514 | 367 | 348,6905 | 0,117902 | 1 | 0,445308 |
| 0,748935 | 280 | 271,7611 | 0,454544 | 1 | 0,814707 |
| -1,12899 | 421 | 343,6453 | 0,25969 | 1 | 0,551979 |
| -2,24069 | 421 | 189,1831 | 0,026209 | 1 | 0,222284 |
| -2,00673 | 420 | 377,9106 | 0,045491 | 1 | 0,846132 |
| -0,92054 | 367 | 320,6143 | 0,357981 | 1 | 0,674813 |
| -1,57311 | 280 | 255,1481 | 0,116933 | 1 | 0,518958 |
| -1,64051 | 421 | 379,3701 | 0,101728 | 1 | 0,342962 |
| -1,92314 | 421 | 204,3405 | 0,055852 | 1 | 0,309036 |
| 0,294264 | 420 | 409,3468 | 0,768706 | 1 | 0,969427 |
| -1,1468 | 367 | 357,0693 | 0,252231 | 1 | 0,607208 |
| 0,964037 | 280 | 272,1374 | 0,335883 | 1 | 0,755731 |
| -0,02365 | 421 | 285,2787 | 0,981148 | 1 | 0,997025 |
| 1,387123 | 421 | 172,3926 | 0,167195 | 1 | 0,453987 |
| -0,45099 | 420 | 313,291 | 0,652306 | 1 | 0,966867 |
| 3,094708 | 367 | 288,36 | 0,002164 | 1 | 0,176492 |
| 1,306846 | 280 | 276 | 0,192353 | 1 | 0,62543 |
| 1,788736 | 421 | 393,7764 | 0,074426 | 1 | 0,294383 |
| 2,896915 | 421 | 206,8525 | 0,004174 | 1 | 0,120964 |
| 1,031289 | 420 | 413,0626 | 0,303009 | 1 | 0,943897 |
| -0,23245 | 367 | 359,9392 | 0,816323 | 1 | 0,95095 |
| -0,10053 | 280 | 274,351 | 0,919997 | 1 | 0,977051 |
| -1,17346 | 421 | 332,1243 | 0,241454 | 1 | 0,538768 |
| 0,454509 | 421 | 185,5693 | 0,649994 | 1 | 0,839358 |
| -0,10267 | 420 | 364,5006 | 0,918283 | 1 | 0,989204 |
| -0,01682 | 367 | 313,0363 | 0,986594 | 1 | 0,996248 |
| -1,75748 | 280 | 257,0206 | 0,080027 | 1 | 0,453386 |
| -1,04324 | 421 | 370,3571 | 0,297518 | 1 | 0,59543 |
| -0,6654 | 421 | 198,9601 | 0,506566 | 1 | 0,754351 |
| -0,23789 | 420 | 398,9712 | 0,812086 | 1 | 0,969427 |
| 0,09812 | 367 | 350,3663 | 0,921893 | 1 | 0,975687 |
| -0,11806 | 280 | 267,8253 | 0,906108 | 1 | 0,975308 |
| -2,09119 | 421 | 371,2105 | 0,037191 | 1 | 0,217944 |
| -1,33208 | 421 | 198,3502 | 0,184364 | 1 | 0,473593 |
| 0,347981 | 420 | 401,5561 | 0,728037 | 1 | 0,969427 |
| -1,35228 | 367 | 349,5394 | 0,17716 | 1 | 0,523131 |
| -0,03899 | 280 | 263,5582 | 0,968931 | 1 | 0,991502 |
| 0,083435 | 421 | 340,2112 | 0,933554 | 1 | 0,972684 |
| 0,685731 | 421 | 187,326 | 0,493731 | 1 | 0,746291 |
| 0,665221 | 420 | 376,1094 | 0,506317 | 1 | 0,960096 |
| 2,034271 | 367 | 324,6208 | 0,042737 | 1 | 0,295433 |
| 0,966063 | 280 | 265,2183 | 0,334893 | 1 | 0,755731 |
| -1,75712 | 421 | 315,3212 | 0,079868 | 1 | 0,302838 |
| 0,602585 | 421 | 175,6794 | 0,547562 | 1 | 0,776424 |
| -0,34333 | 420 | 347,7171 | 0,731557 | 1 | 0,969427 |

| | | | | | |
|---|---|---|---|---|---|
| -1,11172 | 367 | 309,8666 | 0,267121 | 1 | 0,609149 |
| -2,87228 | 280 | 267,7646 | 0,004401 | 0,669004 | 0,161776 |
| -0,18493 | 421 | 282,2774 | 0,853415 | 1 | 0,946355 |
| -0,09299 | 421 | 173,6144 | 0,926021 | 1 | 0,972483 |
| -0,65657 | 420 | 308,8926 | 0,511949 | 1 | 0,960096 |
| 0,705087 | 367 | 363 | 0,481208 | 1 | 0,785415 |
| -0,44499 | 280 | 276 | 0,656676 | 1 | 0,908644 |
| -0,61777 | 421 | 383,2461 | 0,537096 | 1 | 0,778195 |
| 0,431454 | 421 | 203,1355 | 0,666596 | 1 | 0,84773 |
| -0,99382 | 420 | 407,6765 | 0,320898 | 1 | 0,945573 |
| 0,781729 | 367 | 347,7221 | 0,434906 | 1 | 0,742246 |
| 1,285723 | 280 | 267,4258 | 0,199652 | 1 | 0,631529 |
| 0,574883 | 421 | 349,9134 | 0,56574 | 1 | 0,796637 |
| -0,6562 | 421 | 189,7115 | 0,512488 | 1 | 0,756803 |
| 0,49694 | 420 | 383,4588 | 0,619516 | 1 | 0,964878 |
| -0,78262 | 367 | 332,1749 | 0,434409 | 1 | 0,742238 |
| 1,171262 | 280 | 252,5193 | 0,242597 | 1 | 0,670815 |
| 0,469317 | 421 | 347,9504 | 0,639137 | 1 | 0,834842 |
| 1,403482 | 421 | 188,4297 | 0,162119 | 1 | 0,445827 |
| 1,355623 | 420 | 382,5092 | 0,176019 | 1 | 0,9199 |
| 0,913122 | 367 | 326,8698 | 0,361851 | 1 | 0,677287 |
| 0,001865 | 280 | 258,6067 | 0,998514 | 1 | 0,998514 |
| -2,89986 | 421 | 347,1431 | 0,003971 | 1 | 0,087964 |
| -0,63795 | 421 | 189,0895 | 0,524278 | 1 | 0,766525 |
| 0,140335 | 420 | 386,971 | 0,888468 | 1 | 0,983483 |
| -1,38048 | 367 | 331,9461 | 0,168369 | 1 | 0,514971 |
| -2,39417 | 280 | 266,3586 | 0,01735 | 1 | 0,26828 |
| -1,42702 | 421 | 326,6326 | 0,154531 | 1 | 0,426516 |
| -0,6476 | 421 | 182,5051 | 0,518061 | 1 | 0,761675 |
| -0,23179 | 420 | 365,2731 | 0,816832 | 1 | 0,969427 |
| 0,806849 | 367 | 309,8468 | 0,420372 | 1 | 0,731532 |
| -0,54321 | 280 | 257,7876 | 0,587454 | 1 | 0,87561 |
| -0,98441 | 421 | 300,5785 | 0,325708 | 1 | 0,61749 |
| 0,39549 | 421 | 174,7822 | 0,692963 | 1 | 0,865526 |
| -0,60767 | 420 | 328,1797 | 0,543826 | 1 | 0,960096 |
| 1,666609 | 367 | 300,1084 | 0,096635 | 1 | 0,412696 |
| 1,390275 | 280 | 276 | 0,165566 | 1 | 0,588461 |
| 2,620137 | 421 | 406,6027 | 0,009119 | 1 | 0,113397 |
| 2,515746 | 421 | 223,6889 | 0,01258 | 1 | 0,169272 |
| 1,64592 | 420 | 415,9667 | 0,100536 | 1 | 0,848246 |
| 2,589374 | 367 | 362,6008 | 0,010002 | 1 | 0,183886 |
| 1,311282 | 280 | 275,9407 | 0,190852 | 1 | 0,625249 |
| -1,9857 | 421 | 397,731 | 0,047753 | 1 | 0,241202 |
| -1,36984 | 421 | 213,3285 | 0,172178 | 1 | 0,456365 |
| -0,37575 | 420 | 414,6632 | 0,707293 | 1 | 0,968866 |
| -0,27417 | 367 | 362,0687 | 0,784114 | 1 | 0,94337 |
| 0,105998 | 280 | 272,9487 | 0,915662 | 1 | 0,977051 |
| -1,06429 | 421 | 334,9518 | 0,287963 | 1 | 0,584852 |

| | | | | | |
|---|---|---|---|---|---|
| -0,17493 | 421 | 185,6366 | 0,861328 | 1 | 0,947215 |
| 1,768217 | 420 | 371,5872 | 0,077845 | 1 | 0,848246 |
| 0,984812 | 367 | 338,3289 | 0,32542 | 1 | 0,655597 |
| 0,917145 | 280 | 262,0799 | 0,35991 | 1 | 0,761846 |
| -2,06019 | 421 | 417 | 0,040001 | 1 | 0,224527 |
| -0,26416 | 421 | 417 | 0,791788 | 1 | 0,918572 |
| -0,19073 | 420 | 416 | 0,848832 | 1 | 0,973894 |
| -0,86237 | 367 | 363 | 0,389055 | 1 | 0,703005 |
| 0,89801 | 280 | 276 | 0,369963 | 1 | 0,763914 |
| -2,88065 | 421 | 278,2977 | 0,004277 | 1 | 0,087964 |
| -1,7168 | 421 | 171,422 | 0,087821 | 1 | 0,368676 |
| -1,59009 | 420 | 313,4189 | 0,112822 | 1 | 0,848246 |
| -2,59579 | 367 | 285,5201 | 0,009925 | 1 | 0,183886 |
| -0,20229 | 280 | 276 | 0,839842 | 1 | 0,964667 |
| -0,57573 | 421 | 387,959 | 0,565134 | 1 | 0,796637 |
| -0,90984 | 421 | 207,1596 | 0,363962 | 1 | 0,646044 |
| 0,660586 | 420 | 410,8894 | 0,509248 | 1 | 0,960096 |
| -1,96121 | 367 | 361,6017 | 0,050622 | 1 | 0,317862 |
| -1,71765 | 280 | 271,7894 | 0,087 | 1 | 0,460033 |
| 0,01153 | 421 | 341,4078 | 0,990807 | 1 | 0,998054 |
| 0,306238 | 421 | 186,2987 | 0,759765 | 1 | 0,899424 |
| 0,324188 | 420 | 375,4016 | 0,745976 | 1 | 0,969427 |
| 0,549386 | 367 | 318,1933 | 0,583126 | 1 | 0,845786 |
| 2,000122 | 280 | 249,9399 | 0,046569 | 1 | 0,387749 |
| -2,05385 | 421 | 293,9954 | 0,040875 | 1 | 0,226465 |
| 0,390555 | 421 | 177,1656 | 0,696596 | 1 | 0,865526 |
| -0,38242 | 420 | 327,5654 | 0,702399 | 1 | 0,968866 |
| -0,05966 | 367 | 304,1177 | 0,952465 | 1 | 0,979771 |
| 0,425343 | 280 | 276 | 0,670918 | 1 | 0,908644 |
| 0,018438 | 421 | 397,5987 | 0,985298 | 1 | 0,998043 |
| -0,6549 | 421 | 212,5027 | 0,51324 | 1 | 0,756803 |
| -1,74948 | 420 | 414,5734 | 0,080948 | 1 | 0,848246 |
| -1,12484 | 367 | 359,0384 | 0,261406 | 1 | 0,609149 |
| -1,07817 | 280 | 274,7254 | 0,281905 | 1 | 0,70453 |
| -1,23359 | 421 | 385,9616 | 0,218107 | 1 | 0,511974 |
| -0,33887 | 421 | 204,2532 | 0,735054 | 1 | 0,884613 |
| 1,18102 | 420 | 410,3779 | 0,238279 | 1 | 0,943055 |
| -1,76301 | 367 | 355,2118 | 0,078759 | 1 | 0,381638 |
| -0,45445 | 280 | 268,7791 | 0,649875 | 1 | 0,908644 |
| -1,20582 | 421 | 317,3667 | 0,228784 | 1 | 0,520357 |
| -0,08238 | 421 | 182,1247 | 0,934435 | 1 | 0,975655 |
| 0,188053 | 420 | 354,853 | 0,850942 | 1 | 0,973894 |
| -0,56967 | 367 | 315,3903 | 0,569307 | 1 | 0,837838 |
| -0,38834 | 280 | 250,9889 | 0,698094 | 1 | 0,92034 |
| -1,83182 | 421 | 349,2376 | 0,067829 | 1 | 0,280051 |
| -2,23371 | 421 | 190,282 | 0,026667 | 1 | 0,223261 |
| 0,2058 | 420 | 386,5606 | 0,837056 | 1 | 0,973894 |
| -2,76018 | 367 | 335,3899 | 0,006094 | 1 | 0,178183 |

| | | | | | |
|---|---|---|---|---|---|
| -1,23371 | 280 | 261,245 | 0,218418 | 1 | 0,650539 |
| 1,758154 | 421 | 394,9928 | 0,079496 | 1 | 0,302838 |
| 1,346257 | 421 | 211,9613 | 0,179658 | 1 | 0,469227 |
| -0,32845 | 420 | 414,0429 | 0,742738 | 1 | 0,969427 |
| 1,504746 | 367 | 360,4147 | 0,133265 | 1 | 0,477542 |
| 1,447158 | 280 | 274,4071 | 0,148994 | 1 | 0,561358 |
| 0,008293 | 421 | 373,9399 | 0,993387 | 1 | 0,998054 |
| 1,766687 | 421 | 199,1463 | 0,078813 | 1 | 0,355599 |
| 1,009037 | 420 | 401,9757 | 0,313564 | 1 | 0,943897 |
| 1,09748 | 367 | 346,7496 | 0,273193 | 1 | 0,609149 |
| 1,010744 | 280 | 263,9966 | 0,313064 | 1 | 0,733729 |
| 0,757776 | 421 | 334,4863 | 0,449118 | 1 | 0,714643 |
| 0,797855 | 421 | 186,7079 | 0,425968 | 1 | 0,694133 |
| 1,115715 | 420 | 364,2158 | 0,26528 | 1 | 0,943897 |
| 0,617399 | 367 | 330,0824 | 0,537397 | 1 | 0,818865 |
| -0,74961 | 280 | 257,6896 | 0,454171 | 1 | 0,814707 |
| 0,864428 | 421 | 378,7754 | 0,3879 | 1 | 0,677364 |
| 1,340938 | 421 | 201,764 | 0,181448 | 1 | 0,470641 |
| 2,037659 | 420 | 404,1736 | 0,042235 | 1 | 0,846132 |
| 2,317307 | 367 | 346,2531 | 0,02107 | 1 | 0,225192 |
| 0,946964 | 280 | 268,9526 | 0,344507 | 1 | 0,758934 |
| -3,275 | 421 | 343,1078 | 0,001164 | 1 | 0,064989 |
| -1,72543 | 421 | 187,8591 | 0,086094 | 1 | 0,365475 |
| 0,536854 | 420 | 382,6113 | 0,591681 | 1 | 0,964878 |
| 0,026439 | 367 | 321,3857 | 0,978923 | 1 | 0,99209 |
| -0,1147 | 280 | 259,8847 | 0,90877 | 1 | 0,976008 |
| -0,02373 | 421 | 335,9261 | 0,981084 | 1 | 0,997025 |
| 1,133969 | 421 | 185,2216 | 0,258273 | 1 | 0,556025 |
| 0,427416 | 420 | 370,3629 | 0,669325 | 1 | 0,96781 |
| -1,2629 | 367 | 324,4849 | 0,207532 | 1 | 0,560883 |
| 3,128716 | 280 | 255,764 | 0,001959 | 1 | 0,135787 |
| 2,985205 | 421 | 358,103 | 0,003029 | 1 | 0,077359 |
| 1,480102 | 421 | 194,7995 | 0,140461 | 1 | 0,423351 |
| 0,232294 | 420 | 393,4951 | 0,816431 | 1 | 0,969427 |
| 2,606769 | 367 | 346,2388 | 0,009535 | 1 | 0,183886 |
| 1,695005 | 280 | 271,055 | 0,091223 | 1 | 0,468147 |
| -0,80645 | 421 | 353,8044 | 0,420525 | 1 | 0,697735 |
| -0,65792 | 421 | 192,7128 | 0,511376 | 1 | 0,756803 |
| -0,86381 | 420 | 388,9131 | 0,388227 | 1 | 0,95743 |
| -2,61577 | 367 | 340,7399 | 0,009299 | 0,883377 | 0,183886 |
| -1,5411 | 280 | 262,9409 | 0,124494 | 1 | 0,535729 |
| 2,303236 | 421 | 379,4055 | 0,021806 | 1 | 0,175251 |
| 3,001541 | 421 | 202,3938 | 0,003024 | 0,444555 | 0,114447 |
| 0,629418 | 420 | 407,3718 | 0,529428 | 1 | 0,960096 |
| 2,362603 | 367 | 354,2021 | 0,018687 | 1 | 0,217108 |
| 1,109279 | 280 | 267,9047 | 0,268304 | 1 | 0,693794 |
| 1,962421 | 421 | 291,2412 | 0,050665 | 1 | 0,248706 |
| 1,624413 | 421 | 172,473 | 0,106114 | 1 | 0,385334 |

| | | | | | |
|---|---|---|---|---|---|
| -0,71605 | 420 | 320,134 | 0,474482 | 1 | 0,960096 |
| 0,371198 | 367 | 288,5893 | 0,710762 | 1 | 0,912367 |
| 0,125525 | 280 | 245,7077 | 0,90021 | 1 | 0,975243 |
| 0,757032 | 421 | 336,9901 | 0,44956 | 1 | 0,714643 |
| 1,156455 | 421 | 188,0579 | 0,248962 | 1 | 0,548314 |
| 0,510793 | 420 | 370,9503 | 0,6098 | 1 | 0,964878 |
| 0,401837 | 367 | 319,5815 | 0,688073 | 1 | 0,903032 |
| 0,487777 | 280 | 256,7452 | 0,626124 | 1 | 0,899494 |
| -0,30638 | 421 | 323,6296 | 0,759515 | 1 | 0,901251 |
| -1,10511 | 421 | 184,9624 | 0,270548 | 1 | 0,569587 |
| 0,555228 | 420 | 360,8503 | 0,579083 | 1 | 0,964878 |
| 1,111962 | 367 | 307,1866 | 0,267024 | 1 | 0,609149 |
| 0,686729 | 280 | 251,6479 | 0,492886 | 1 | 0,827962 |
| -2,09089 | 421 | 350,339 | 0,037258 | 1 | 0,217944 |
| -2,41678 | 421 | 192,0827 | 0,016594 | 1 | 0,181207 |
| -0,54809 | 420 | 388,1487 | 0,583944 | 1 | 0,964878 |
| -2,5156 | 367 | 338,7137 | 0,012345 | 1 | 0,189546 |
| -1,76644 | 280 | 264,4217 | 0,078476 | 1 | 0,453386 |
| 3,531914 | 421 | 373,2717 | 4,64E-04 | 0,495334 | 0,04642 |
| 1,902644 | 421 | 201,9585 | 0,05851 | 1 | 0,314262 |
| -0,43644 | 420 | 406,1375 | 0,66275 | 1 | 0,96781 |
| 2,674924 | 367 | 352,9248 | 0,007823 | 1 | 0,180403 |
| 0,786215 | 280 | 273,0843 | 0,432423 | 1 | 0,803161 |
| -2,11135 | 421 | 312,7291 | 0,035535 | 1 | 0,215166 |
| -2,73103 | 421 | 180,7543 | 0,006939 | 1 | 0,141161 |
| 0,176127 | 420 | 355,1658 | 0,860294 | 1 | 0,976988 |
| -0,08223 | 367 | 324,0058 | 0,934517 | 1 | 0,977998 |
| -1,51246 | 280 | 257,3867 | 0,131643 | 1 | 0,540563 |
| -0,85888 | 421 | 354,9827 | 0,390987 | 1 | 0,68039 |
| 1,227356 | 421 | 192,741 | 0,221186 | 1 | 0,516786 |
| -0,58145 | 420 | 387,7802 | 0,561277 | 1 | 0,960096 |
| 1,666734 | 367 | 330,4253 | 0,096515 | 1 | 0,412696 |
| -0,73101 | 280 | 259,8915 | 0,465434 | 1 | 0,824256 |
| -1,18273 | 421 | 304,1407 | 0,237839 | 1 | 0,532577 |
| 1,468736 | 421 | 178,6605 | 0,143663 | 1 | 0,42451 |
| 0,714124 | 420 | 335,1542 | 0,475647 | 1 | 0,960096 |
| 0,529207 | 367 | 290,4182 | 0,597066 | 1 | 0,860209 |
| -1,25045 | 280 | 276 | 0,212193 | 1 | 0,644706 |
| -2,28216 | 421 | 326,9886 | 0,023122 | 1 | 0,177419 |
| -1,12642 | 421 | 177,4842 | 0,261507 | 1 | 0,557979 |
| -0,38665 | 420 | 360,9378 | 0,699244 | 1 | 0,968866 |
| -1,11516 | 367 | 317,3483 | 0,265625 | 1 | 0,609149 |
| -3,72258 | 280 | 256,3634 | 2,42E-04 | 0,260853 | 0,057407 |
| -0,09753 | 421 | 389,3311 | 0,922355 | 1 | 0,966828 |
| 0,448468 | 421 | 207,5758 | 0,654284 | 1 | 0,841107 |
| -0,15877 | 420 | 411,0151 | 0,873927 | 1 | 0,982463 |
| 0,206334 | 367 | 357,5965 | 0,836648 | 1 | 0,956935 |
| -0,0326 | 280 | 268,4728 | 0,974021 | 1 | 0,99236 |

| | | | | | |
|---|---|---|---|---|---|
| 1,038949 | 421 | 306,8549 | 0,299646 | 1 | 0,596172 |
| -0,91851 | 421 | 177,5783 | 0,359598 | 1 | 0,643758 |
| 0,388251 | 420 | 340,0944 | 0,698073 | 1 | 0,968866 |
| 0,75161 | 367 | 298,6131 | 0,452877 | 1 | 0,758154 |
| 1,355774 | 280 | 255,1447 | 0,17637 | 1 | 0,598625 |
| -0,48389 | 421 | 353,1639 | 0,628767 | 1 | 0,833119 |
| -0,23451 | 421 | 192,2805 | 0,814837 | 1 | 0,926063 |
| -0,58282 | 420 | 386,0532 | 0,560358 | 1 | 0,960096 |
| 0,32678 | 367 | 340,9857 | 0,744034 | 1 | 0,923669 |
| 2,226245 | 280 | 265,636 | 0,026837 | 1 | 0,324814 |
| 0,677848 | 421 | 354,8659 | 0,49831 | 1 | 0,751705 |
| 0,670639 | 421 | 191,6469 | 0,503258 | 1 | 0,75239 |
| 0,961684 | 420 | 387,6559 | 0,336808 | 1 | 0,947373 |
| 0,936971 | 367 | 336,1274 | 0,349446 | 1 | 0,668671 |
| 0,300908 | 280 | 264,046 | 0,763722 | 1 | 0,944296 |
| 1,203316 | 421 | 371,6277 | 0,22962 | 1 | 0,520357 |
| 2,247047 | 421 | 199,6042 | 0,025732 | 1 | 0,221586 |
| 0,149857 | 420 | 402,8782 | 0,880952 | 1 | 0,982463 |
| 3,78924 | 367 | 351,1683 | 1,78E-04 | 0,191099 | 0,053579 |
| 1,234639 | 280 | 269,4521 | 0,21804 | 1 | 0,650539 |
| -1,13214 | 421 | 310,4769 | 0,258451 | 1 | 0,550898 |
| -0,74836 | 421 | 177,22 | 0,455233 | 1 | 0,716467 |
| -2,33488 | 420 | 340,5717 | 0,02013 | 1 | 0,75904 |
| 0,149859 | 367 | 309,2791 | 0,880973 | 1 | 0,962453 |
| -0,7134 | 280 | 250,3957 | 0,476263 | 1 | 0,824256 |
| 2,617844 | 421 | 405,3053 | 0,00918 | 1 | 0,113397 |
| 2,65837 | 421 | 222,397 | 0,008422 | 1 | 0,148102 |
| 1,563141 | 420 | 415,9996 | 0,11878 | 1 | 0,858144 |
| 2,767067 | 367 | 362,6869 | 0,005946 | 0,891928 | 0,178183 |
| 1,14996 | 280 | 275,9482 | 0,251156 | 1 | 0,679007 |
| -1,04051 | 421 | 373,0368 | 0,298776 | 1 | 0,596172 |
| -0,23236 | 421 | 198,2694 | 0,816501 | 1 | 0,926377 |
| 0,390982 | 420 | 402,46 | 0,696018 | 1 | 0,968866 |
| 0,870296 | 367 | 350,714 | 0,384734 | 1 | 0,696868 |
| 1,957127 | 280 | 265,5605 | 0,05138 | 1 | 0,398826 |
| -1,00855 | 421 | 367,5238 | 0,313856 | 1 | 0,608538 |
| -1,61982 | 421 | 196,0447 | 0,106878 | 1 | 0,386957 |
| 0,993852 | 420 | 400,7841 | 0,320895 | 1 | 0,945573 |
| 0,747935 | 367 | 347,6894 | 0,455005 | 1 | 0,758509 |
| 1,233509 | 280 | 268,5619 | 0,218465 | 1 | 0,650539 |
| 0,049739 | 421 | 312,7677 | 0,960362 | 1 | 0,989922 |
| -1,02559 | 421 | 177,6542 | 0,306479 | 1 | 0,603743 |
| -0,53607 | 420 | 348,645 | 0,59225 | 1 | 0,964878 |
| -0,16118 | 367 | 282,3824 | 0,87207 | 1 | 0,959978 |
| -1,6983 | 280 | 263,8357 | 0,09063 | 1 | 0,468147 |
| -0,10912 | 421 | 373,6465 | 0,913166 | 1 | 0,966828 |
| -1,79379 | 421 | 199,9963 | 0,074357 | 1 | 0,344631 |
| -0,18884 | 420 | 402,6104 | 0,850316 | 1 | 0,973894 |

| | | | | | |
|---|---|---|---|---|---|
| -0,99528 | 367 | 340,3847 | 0,320307 | 1 | 0,653184 |
| -0,38496 | 280 | 264,2318 | 0,700575 | 1 | 0,92034 |
| 1,26112 | 421 | 408,0635 | 0,207986 | 1 | 0,496728 |
| 0,767279 | 421 | 223,5844 | 0,443725 | 1 | 0,708362 |
| 0,521315 | 420 | 415,9492 | 0,602425 | 1 | 0,964878 |
| 1,012493 | 367 | 362,9897 | 0,311977 | 1 | 0,646834 |
| 1,380274 | 280 | 275,8943 | 0,168619 | 1 | 0,59271 |
| 0,124398 | 421 | 316,9992 | 0,901079 | 1 | 0,965517 |
| 0,364298 | 421 | 180,7908 | 0,716061 | 1 | 0,874476 |
| 1,938504 | 420 | 347,7671 | 0,053371 | 1 | 0,846627 |
| 0,168565 | 367 | 303,9793 | 0,866251 | 1 | 0,959636 |
| -0,20242 | 280 | 276 | 0,839735 | 1 | 0,964667 |
| 0,573957 | 421 | 311,4499 | 0,566411 | 1 | 0,796637 |
| 0,390447 | 421 | 178,7275 | 0,696671 | 1 | 0,865526 |
| 0,37345 | 420 | 345,1683 | 0,709043 | 1 | 0,968866 |
| 0,423193 | 367 | 306,1815 | 0,672451 | 1 | 0,897595 |
| 1,571049 | 280 | 251,4695 | 0,117429 | 1 | 0,518958 |
| -0,673 | 421 | 328,4477 | 0,501422 | 1 | 0,754888 |
| 1,155156 | 421 | 181,7753 | 0,249543 | 1 | 0,548314 |
| 0,031416 | 420 | 364,3497 | 0,974955 | 1 | 0,997342 |
| 1,999807 | 367 | 320,429 | 0,046365 | 1 | 0,307806 |
| 1,869604 | 280 | 253,1978 | 0,062693 | 1 | 0,422605 |
| -1,71038 | 421 | 316,1281 | 0,088177 | 1 | 0,3179 |
| 0,343736 | 421 | 180,2503 | 0,731445 | 1 | 0,882616 |
| 0,705051 | 420 | 348,4586 | 0,481249 | 1 | 0,960096 |
| 1,455224 | 367 | 313,658 | 0,146607 | 1 | 0,498729 |
| 1,326916 | 280 | 264,472 | 0,185681 | 1 | 0,617707 |
| 1,984295 | 421 | 340,6618 | 0,048025 | 1 | 0,241202 |
| 1,041909 | 421 | 186,1531 | 0,298805 | 1 | 0,597214 |
| 2,110098 | 420 | 371,4043 | 0,035518 | 1 | 0,828713 |
| 2,858008 | 367 | 325,2263 | 0,004538 | 1 | 0,178183 |
| -0,47363 | 280 | 257,8011 | 0,636161 | 1 | 0,906996 |
| 0,771626 | 421 | 408,64 | 0,440782 | 1 | 0,710437 |
| 0,917003 | 421 | 225,5502 | 0,360121 | 1 | 0,643774 |
| -0,08109 | 420 | 415,9306 | 0,935411 | 1 | 0,992059 |
| -0,94381 | 367 | 362,8344 | 0,345897 | 1 | 0,66829 |
| -0,54352 | 280 | 272,7383 | 0,587215 | 1 | 0,87561 |
| -1,45058 | 421 | 327,7656 | 0,147853 | 1 | 0,419613 |
| -1,07037 | 421 | 185,0551 | 0,285846 | 1 | 0,584263 |
| 0,742937 | 420 | 362,8403 | 0,458001 | 1 | 0,960096 |
| -0,2452 | 367 | 333,6156 | 0,80645 | 1 | 0,950149 |
| -2,49877 | 280 | 261,7576 | 0,013076 | 1 | 0,246323 |
| -0,85129 | 421 | 339,5798 | 0,39521 | 1 | 0,686065 |
| 0,081831 | 421 | 188,2587 | 0,934868 | 1 | 0,975655 |
| -0,35668 | 420 | 375,7325 | 0,721535 | 1 | 0,969427 |
| 0,413918 | 367 | 328,1653 | 0,679205 | 1 | 0,899694 |
| 0,839407 | 280 | 259,8894 | 0,402012 | 1 | 0,784757 |
| -0,47492 | 421 | 351,2221 | 0,635138 | 1 | 0,83455 |

| | | | | | |
|---|---|---|---|---|---|
| -0,34673 | 421 | 188,8305 | 0,729181 | 1 | 0,882616 |
| 0,322295 | 420 | 384,7745 | 0,747404 | 1 | 0,969427 |
| -0,23562 | 367 | 331,0932 | 0,813877 | 1 | 0,95095 |
| -0,35913 | 280 | 271,1695 | 0,719774 | 1 | 0,932673 |
| -1,00271 | 421 | 383,6152 | 0,316633 | 1 | 0,610186 |
| -2,02501 | 421 | 206,545 | 0,044153 | 1 | 0,277922 |
| 0,090936 | 420 | 410,8395 | 0,927588 | 1 | 0,990001 |
| -0,70085 | 367 | 358,0886 | 0,48385 | 1 | 0,786044 |
| -1,36127 | 280 | 266,5857 | 0,174579 | 1 | 0,598625 |
| -1,68997 | 421 | 314,4926 | 0,092024 | 1 | 0,325541 |
| 0,194797 | 421 | 178,9155 | 0,845773 | 1 | 0,943293 |
| -0,28862 | 420 | 346,7018 | 0,773042 | 1 | 0,969427 |
| 0,113847 | 367 | 293,4507 | 0,909437 | 1 | 0,970624 |
| -0,42085 | 280 | 250,1124 | 0,674229 | 1 | 0,909385 |
| 0,27088 | 421 | 340,0564 | 0,786647 | 1 | 0,917552 |
| 0,414018 | 421 | 187,1087 | 0,679335 | 1 | 0,857638 |
| 0,814868 | 420 | 374,6764 | 0,415666 | 1 | 0,95743 |
| 0,17083 | 367 | 332,5113 | 0,864462 | 1 | 0,959636 |
| -0,66029 | 280 | 264,5588 | 0,509642 | 1 | 0,831912 |
| 2,124161 | 421 | 397,9495 | 0,034273 | 1 | 0,212548 |
| 3,649374 | 421 | 213,6998 | 3,30E-04 | 0,353916 | 0,038096 |
| 1,268893 | 420 | 415,5606 | 0,205189 | 1 | 0,928589 |
| 2,600003 | 367 | 362,6563 | 0,009703 | 1 | 0,183886 |
| 2,608654 | 280 | 274,1512 | 0,009589 | 1 | 0,226737 |
| 1,685779 | 421 | 312,862 | 0,092835 | 1 | 0,327642 |
| 0,97256 | 421 | 180,2151 | 0,332076 | 1 | 0,626331 |
| -1,58427 | 420 | 341,6218 | 0,114057 | 1 | 0,848246 |
| 2,082992 | 367 | 305,4559 | 0,038084 | 1 | 0,28956 |
| 2,324614 | 280 | 251,5674 | 0,020889 | 1 | 0,30229 |
| 0,765341 | 421 | 393,0499 | 0,444528 | 1 | 0,713799 |
| 1,334679 | 421 | 209,0897 | 0,183433 | 1 | 0,473593 |
| -0,01212 | 420 | 413,0092 | 0,990339 | 1 | 0,997342 |
| -1,26833 | 367 | 360,2167 | 0,205501 | 1 | 0,558434 |
| -3,31201 | 280 | 270,669 | 0,001052 | 1 | 0,121989 |
| -0,81484 | 421 | 314,1524 | 0,415784 | 1 | 0,695339 |
| 0,63385 | 421 | 177,9319 | 0,526992 | 1 | 0,767321 |
| 0,269746 | 420 | 348,6396 | 0,787515 | 1 | 0,969427 |
| 1,623386 | 367 | 301,7135 | 0,105551 | 1 | 0,429907 |
| 1,523149 | 280 | 255,1368 | 0,12896 | 1 | 0,536498 |
| 0,552896 | 421 | 335,5372 | 0,580703 | 1 | 0,805078 |
| 0,681882 | 421 | 186,5567 | 0,496159 | 1 | 0,747712 |
| 1,307386 | 420 | 365,8961 | 0,191903 | 1 | 0,925729 |
| -0,85676 | 367 | 332,1748 | 0,392193 | 1 | 0,705088 |
| -1,64569 | 280 | 249,6176 | 0,101086 | 1 | 0,480557 |
| 0,074991 | 421 | 372,3887 | 0,940262 | 1 | 0,97655 |
| -0,62532 | 421 | 198,3139 | 0,532482 | 1 | 0,769894 |
| 0,811393 | 420 | 401,8454 | 0,41762 | 1 | 0,95743 |
| -0,43821 | 367 | 350,3945 | 0,661503 | 1 | 0,894303 |

| | | | | | |
|---|---|---|---|---|---|
| -1,18964 | 280 | 270,5414 | 0,235232 | 1 | 0,659852 |
| 1,423561 | 421 | 392,7209 | 0,155367 | 1 | 0,428042 |
| 1,360592 | 421 | 211,9163 | 0,175088 | 1 | 0,459682 |
| 0,106825 | 420 | 413,5082 | 0,914979 | 1 | 0,988069 |
| -0,31941 | 367 | 359,3819 | 0,7496 | 1 | 0,923669 |
| 0,146671 | 280 | 272,4257 | 0,8835 | 1 | 0,973784 |
| -0,7842 | 421 | 409,6316 | 0,433376 | 1 | 0,705289 |
| 0,463003 | 421 | 225,787 | 0,643808 | 1 | 0,838647 |
| -1,14463 | 420 | 415,7317 | 0,253021 | 1 | 0,943897 |
| -0,59163 | 367 | 361,1787 | 0,554468 | 1 | 0,830257 |
| -0,43595 | 280 | 275,9257 | 0,663214 | 1 | 0,908644 |
| -0,64648 | 421 | 374,482 | 0,518366 | 1 | 0,768119 |
| -0,69859 | 421 | 201,5468 | 0,485616 | 1 | 0,740711 |
| -0,01052 | 420 | 403,8725 | 0,991608 | 1 | 0,997342 |
| 0,494058 | 367 | 353,3777 | 0,621572 | 1 | 0,875495 |
| 0,502187 | 280 | 262,8239 | 0,615956 | 1 | 0,889977 |
| -0,73971 | 421 | 308,5217 | 0,460036 | 1 | 0,724425 |
| 1,988153 | 421 | 177,5532 | 0,048331 | 1 | 0,286754 |
| -0,43192 | 420 | 340,2697 | 0,666072 | 1 | 0,96781 |
| 0,11815 | 367 | 304,0849 | 0,906027 | 1 | 0,970486 |
| -0,54513 | 280 | 253,6061 | 0,58614 | 1 | 0,87561 |
| -1,53786 | 421 | 325,144 | 0,125054 | 1 | 0,38857 |
| -2,43332 | 421 | 183,8519 | 0,015918 | 1 | 0,179127 |
| -0,01365 | 420 | 365,1663 | 0,989113 | 1 | 0,997342 |
| -1,03797 | 367 | 309,4796 | 0,300094 | 1 | 0,633391 |
| -0,77048 | 280 | 247,892 | 0,441751 | 1 | 0,810247 |
| -0,50761 | 421 | 344,6782 | 0,61205 | 1 | 0,825172 |
| -1,52088 | 421 | 185,7719 | 0,129989 | 1 | 0,410868 |
| -0,74749 | 420 | 374,3751 | 0,455236 | 1 | 0,960096 |
| -0,33276 | 367 | 322,751 | 0,739535 | 1 | 0,923669 |
| -0,02607 | 280 | 258,9337 | 0,979225 | 1 | 0,99236 |
| -0,82023 | 421 | 400,2724 | 0,412571 | 1 | 0,695339 |
| 0,062477 | 421 | 212,5393 | 0,950242 | 1 | 0,980953 |
| -1,11556 | 420 | 415,4417 | 0,265255 | 1 | 0,943897 |
| -0,37257 | 367 | 361,3899 | 0,709683 | 1 | 0,912021 |
| -2,42401 | 280 | 271,5589 | 0,016004 | 1 | 0,266517 |
| 0,198019 | 421 | 374,1179 | 0,843138 | 1 | 0,939735 |
| 0,332002 | 421 | 199,2335 | 0,740237 | 1 | 0,886754 |
| -0,06338 | 420 | 402,7728 | 0,949494 | 1 | 0,992811 |
| 2,067488 | 367 | 350,6706 | 0,039421 | 1 | 0,28956 |
| 0,232376 | 280 | 268,6849 | 0,816423 | 1 | 0,95747 |
| -0,26146 | 421 | 349,3311 | 0,793889 | 1 | 0,919136 |
| 1,17113 | 421 | 188,9668 | 0,243021 | 1 | 0,542422 |
| 1,285755 | 420 | 383,1674 | 0,199305 | 1 | 0,928589 |
| 1,324483 | 367 | 331,1853 | 0,186256 | 1 | 0,535663 |
| -0,84247 | 280 | 258,6771 | 0,400302 | 1 | 0,784757 |
| 0,107756 | 421 | 331,0368 | 0,914255 | 1 | 0,966828 |
| -0,20533 | 421 | 183,6212 | 0,837541 | 1 | 0,938419 |

| | | | | | |
|---|---|---|---|---|---|
| -1,37147 | 420 | 363,6153 | 0,171073 | 1 | 0,9199 |
| 1,899866 | 367 | 310,5056 | 0,058377 | 1 | 0,339097 |
| 0,963557 | 280 | 252,4494 | 0,33619 | 1 | 0,755731 |
| -2,1695 | 421 | 336,9031 | 0,030744 | 1 | 0,200565 |
| -1,24482 | 421 | 187,2978 | 0,214754 | 1 | 0,514565 |
| 0,800207 | 420 | 369,7844 | 0,424105 | 1 | 0,95743 |
| -0,45367 | 367 | 329,6274 | 0,650365 | 1 | 0,887772 |
| -1,24136 | 280 | 262,1244 | 0,215584 | 1 | 0,648472 |
| 1,142115 | 421 | 393,6404 | 0,2541 | 1 | 0,546261 |
| 2,986443 | 421 | 210,763 | 0,003157 | 1 | 0,116024 |
| 2,393397 | 420 | 413,5717 | 0,017138 | 1 | 0,75904 |
| 0,919739 | 367 | 361,977 | 0,358322 | 1 | 0,674813 |
| 0,654008 | 280 | 273,399 | 0,513657 | 1 | 0,831912 |
| -0,00127 | 421 | 358,9014 | 0,998989 | 1 | 0,999298 |
| 1,104849 | 421 | 190,3343 | 0,27062 | 1 | 0,569587 |
| 0,71429 | 420 | 388,7824 | 0,475477 | 1 | 0,960096 |
| -0,9811 | 367 | 350,6026 | 0,32722 | 1 | 0,655788 |
| -1,93023 | 280 | 266,0587 | 0,054641 | 1 | 0,40654 |
| -2,36273 | 421 | 341,5615 | 0,018701 | 1 | 0,159223 |
| -0,82141 | 421 | 188,722 | 0,412449 | 1 | 0,684933 |
| -0,47922 | 420 | 377,4115 | 0,632062 | 1 | 0,964878 |
| -1,08821 | 367 | 321,5757 | 0,277317 | 1 | 0,611884 |
| -0,70787 | 280 | 254,6096 | 0,479673 | 1 | 0,824256 |
| 3,095249 | 421 | 391,3564 | 0,002108 | 0,320402 | 0,069718 |
| 4,282465 | 421 | 209,8918 | 2,81E-05 | 0,004303 | 0,015907 |
| 0,902333 | 420 | 413,7445 | 0,367405 | 1 | 0,95743 |
| 3,120889 | 367 | 360,7462 | 0,001948 | 0,298109 | 0,176492 |
| 1,561356 | 280 | 275,0465 | 0,11959 | 1 | 0,5239 |
| 0,381113 | 421 | 326,362 | 0,703368 | 1 | 0,874567 |
| 0,142806 | 421 | 183,1809 | 0,8866 | 1 | 0,957782 |
| 2,485227 | 420 | 358,8325 | 0,013402 | 1 | 0,69643 |
| -0,57102 | 367 | 324,2271 | 0,568384 | 1 | 0,837838 |
| 0,849233 | 280 | 258,9782 | 0,396536 | 1 | 0,782458 |
| -2,77049 | 421 | 319,611 | 0,005925 | 1 | 0,09705 |
| -0,54624 | 421 | 183,2975 | 0,585568 | 1 | 0,802496 |
| -1,0603 | 420 | 364,0426 | 0,28971 | 1 | 0,943897 |
| -1,22625 | 367 | 310,4442 | 0,221033 | 1 | 0,573978 |
| 0,524271 | 280 | 254,0357 | 0,600547 | 1 | 0,881231 |
| 0,435114 | 421 | 398,5157 | 0,663716 | 1 | 0,852478 |
| 0,861007 | 421 | 214,511 | 0,390196 | 1 | 0,664652 |
| -0,38998 | 420 | 415,1436 | 0,696753 | 1 | 0,968866 |
| 0,784719 | 367 | 362,8528 | 0,43313 | 1 | 0,741735 |
| 1,482501 | 280 | 275,7997 | 0,139349 | 1 | 0,552627 |
| 2,399886 | 421 | 369,3532 | 0,016895 | 1 | 0,152456 |
| 2,520281 | 421 | 197,3595 | 0,012518 | 1 | 0,169272 |
| 0,627125 | 420 | 401,344 | 0,530934 | 1 | 0,960096 |
| 2,691347 | 367 | 353,7537 | 0,007454 | 1 | 0,180403 |
| 0,080379 | 280 | 270,8129 | 0,935995 | 1 | 0,980226 |

| | | | | | |
|---|---|---|---|---|---|
| -0,28951 | 421 | 359,377 | 0,772356 | 1 | 0,90985 |
| -0,33789 | 421 | 193,6585 | 0,735811 | 1 | 0,884613 |
| 0,598547 | 420 | 388,145 | 0,549824 | 1 | 0,960096 |
| 1,447284 | 367 | 340,8629 | 0,148737 | 1 | 0,499998 |
| -0,2093 | 280 | 263,4609 | 0,834379 | 1 | 0,964667 |
| -2,26445 | 421 | 310,3259 | 0,024236 | 1 | 0,181711 |
| -1,555 | 421 | 179,4424 | 0,121707 | 1 | 0,402221 |
| 0,803883 | 420 | 352,2882 | 0,422007 | 1 | 0,95743 |
| -0,6997 | 367 | 306,2249 | 0,484644 | 1 | 0,786044 |
| 0,656965 | 280 | 261,0067 | 0,511782 | 1 | 0,831912 |
| -2,79114 | 421 | 329,1384 | 0,005559 | 1 | 0,09373 |
| -0,7866 | 421 | 185,5281 | 0,432517 | 1 | 0,69936 |
| -0,03043 | 420 | 369,3743 | 0,975743 | 1 | 0,997342 |
| -0,72257 | 367 | 326,2228 | 0,470459 | 1 | 0,775692 |
| 0,803332 | 280 | 261,0983 | 0,422514 | 1 | 0,796907 |
| 0,015129 | 421 | 386,0305 | 0,987937 | 1 | 0,998054 |
| 1,591394 | 421 | 204,5589 | 0,113065 | 1 | 0,391524 |
| -0,1837 | 420 | 409,5978 | 0,854338 | 1 | 0,973894 |
| 0,697874 | 367 | 352,4001 | 0,485716 | 1 | 0,786224 |
| 0,561497 | 280 | 273,3581 | 0,574919 | 1 | 0,870337 |
| -1,60553 | 421 | 390,0292 | 0,109186 | 1 | 0,355682 |
| -1,34509 | 421 | 209,2436 | 0,180052 | 1 | 0,469443 |
| 0,89944 | 420 | 412,6017 | 0,368943 | 1 | 0,95743 |
| -0,21388 | 367 | 360,5264 | 0,830761 | 1 | 0,956843 |
| 0,023815 | 280 | 270,9258 | 0,981017 | 1 | 0,99236 |
| -0,532 | 421 | 349,812 | 0,595062 | 1 | 0,813756 |
| -0,37451 | 421 | 190,5176 | 0,708438 | 1 | 0,870812 |
| -1,01203 | 420 | 380,3103 | 0,312165 | 1 | 0,943897 |
| -1,30148 | 367 | 341,646 | 0,19397 | 1 | 0,547402 |
| -0,92266 | 280 | 264,1439 | 0,357026 | 1 | 0,761846 |
| 1,11807 | 421 | 409,3852 | 0,264193 | 1 | 0,558399 |
| 1,916457 | 421 | 224,0978 | 0,056579 | 1 | 0,311185 |
| 0,316079 | 420 | 415,8333 | 0,752101 | 1 | 0,969427 |
| -1,23521 | 367 | 361,4998 | 0,217555 | 1 | 0,569492 |
| -0,58089 | 280 | 275,8305 | 0,561787 | 1 | 0,861728 |
| 3,163233 | 421 | 359,8592 | 0,001693 | 1 | 0,069718 |
| 1,616235 | 421 | 194,0476 | 0,107668 | 1 | 0,388172 |
| -1,00534 | 420 | 393,1771 | 0,315352 | 1 | 0,943897 |
| 2,112594 | 367 | 347,6468 | 0,035348 | 1 | 0,283349 |
| 1,995314 | 280 | 270,219 | 0,047013 | 1 | 0,389275 |
| -0,13456 | 421 | 324,58 | 0,893044 | 1 | 0,962 |
| -0,42439 | 421 | 184,8218 | 0,671772 | 1 | 0,852155 |
| 0,109303 | 420 | 358,2178 | 0,913023 | 1 | 0,987181 |
| -0,41011 | 367 | 316,612 | 0,682002 | 1 | 0,900574 |
| 0,30755 | 280 | 250,7635 | 0,75868 | 1 | 0,94273 |
| -0,54703 | 421 | 366,3515 | 0,584691 | 1 | 0,807036 |
| -1,98186 | 421 | 196,5041 | 0,048889 | 1 | 0,286808 |
| -0,69387 | 420 | 398,4784 | 0,488168 | 1 | 0,960096 |

| | | | | | |
|---|---|---|---|---|---|
| -0,1239 | 367 | 341,1398 | 0,901465 | 1 | 0,970486 |
| -1,21794 | 280 | 265,0216 | 0,224331 | 1 | 0,654014 |
| -1,45849 | 421 | 288,8507 | 0,145792 | 1 | 0,414711 |
| -0,06235 | 421 | 173,8982 | 0,950359 | 1 | 0,980953 |
| 0,803342 | 420 | 317,0324 | 0,422379 | 1 | 0,95743 |
| -2,36393 | 367 | 295,761 | 0,018729 | 1 | 0,217108 |
| 0,100611 | 280 | 248,7142 | 0,91994 | 1 | 0,977051 |
| -2,48624 | 421 | 317,1169 | 0,013424 | 1 | 0,138566 |
| -0,41857 | 421 | 179,7545 | 0,676028 | 1 | 0,854676 |
| -0,49984 | 420 | 348,8382 | 0,617504 | 1 | 0,964878 |
| -1,11062 | 367 | 299,6148 | 0,267621 | 1 | 0,609149 |
| 0,40663 | 280 | 252,0154 | 0,684625 | 1 | 0,913844 |
| -0,81548 | 421 | 361,3447 | 0,415336 | 1 | 0,695339 |
| -0,24181 | 421 | 194,4808 | 0,809186 | 1 | 0,926063 |
| -0,53232 | 420 | 393,4155 | 0,594803 | 1 | 0,964878 |
| 1,774089 | 367 | 332,8189 | 0,076963 | 1 | 0,379028 |
| 0,64402 | 280 | 261,3544 | 0,520127 | 1 | 0,833721 |
| -0,64886 | 421 | 402,7289 | 0,516799 | 1 | 0,767364 |
| -0,46216 | 421 | 218,4403 | 0,644428 | 1 | 0,838647 |
| 1,61821 | 420 | 415,8375 | 0,106375 | 1 | 0,848246 |
| 0,65624 | 367 | 362,9035 | 0,512086 | 1 | 0,797829 |
| -0,47944 | 280 | 275,8884 | 0,632009 | 1 | 0,903355 |
| -1,11059 | 421 | 359,8615 | 0,267488 | 1 | 0,563516 |
| 0,110726 | 421 | 194,7577 | 0,911948 | 1 | 0,96646 |
| -2,32362 | 420 | 386,3033 | 0,020664 | 1 | 0,75904 |
| 0,979306 | 367 | 346,6134 | 0,328112 | 1 | 0,655788 |
| 0,642914 | 280 | 262,1129 | 0,520842 | 1 | 0,833721 |
| 0,922934 | 421 | 342,7607 | 0,356691 | 1 | 0,646073 |
| 0,389368 | 421 | 189,2085 | 0,697442 | 1 | 0,865528 |
| 0,785752 | 420 | 375,9867 | 0,432508 | 1 | 0,960096 |
| 0,417705 | 367 | 333,9545 | 0,676431 | 1 | 0,898926 |
| -0,56678 | 280 | 261,4124 | 0,571352 | 1 | 0,869862 |
| -1,09184 | 421 | 325,9774 | 0,275709 | 1 | 0,57291 |
| -1,09862 | 421 | 184,5723 | 0,273366 | 1 | 0,574564 |
| -0,68831 | 420 | 361,2318 | 0,491698 | 1 | 0,960096 |
| 1,389816 | 367 | 324,5064 | 0,165537 | 1 | 0,512248 |
| -1,04233 | 280 | 258,8689 | 0,298232 | 1 | 0,723324 |
| -0,20428 | 421 | 381,2053 | 0,838241 | 1 | 0,93721 |
| 0,845743 | 421 | 202,4886 | 0,398694 | 1 | 0,670572 |
| -1,14639 | 420 | 407,333 | 0,252305 | 1 | 0,943897 |
| 0,452599 | 367 | 351,0887 | 0,651117 | 1 | 0,887994 |
| 0,783773 | 280 | 270,8801 | 0,433858 | 1 | 0,803161 |
| 1,544888 | 421 | 346,3149 | 0,123287 | 1 | 0,385082 |
| 0,589855 | 421 | 189,5034 | 0,555991 | 1 | 0,778696 |
| 0,204009 | 420 | 378,8723 | 0,838456 | 1 | 0,973894 |
| 0,629373 | 367 | 319,6714 | 0,529554 | 1 | 0,812666 |
| 1,950312 | 280 | 249,6034 | 0,052258 | 1 | 0,401537 |
| 0,878517 | 421 | 306,1091 | 0,380352 | 1 | 0,671614 |

| | | | | | |
|---|---|---|---|---|---|
| 1,807084 | 421 | 175,5292 | 0,072462 | 1 | 0,341588 |
| 0,065993 | 420 | 339,4882 | 0,947422 | 1 | 0,992811 |
| -0,77383 | 367 | 304,3371 | 0,439633 | 1 | 0,742744 |
| 1,304061 | 280 | 251,1761 | 0,193407 | 1 | 0,62543 |
| -0,92358 | 421 | 343,5314 | 0,356353 | 1 | 0,646073 |
| -0,90412 | 421 | 187,7874 | 0,367089 | 1 | 0,647779 |
| -1,52497 | 420 | 380,6371 | 0,128097 | 1 | 0,887798 |
| -0,82114 | 367 | 343,2455 | 0,412136 | 1 | 0,724248 |
| 0,308608 | 280 | 260,8469 | 0,757866 | 1 | 0,94273 |
| 3,075338 | 421 | 361,5109 | 0,002263 | 1 | 0,069718 |
| 1,45211 | 421 | 193,2008 | 0,148093 | 1 | 0,428784 |
| 1,585437 | 420 | 393,0718 | 0,113671 | 1 | 0,848246 |
| 1,674371 | 367 | 340,571 | 0,094976 | 1 | 0,412696 |
| 1,223912 | 280 | 262,4858 | 0,222083 | 1 | 0,654014 |
| 0,690828 | 421 | 334,752 | 0,490152 | 1 | 0,748388 |
| 0,565663 | 421 | 181,3768 | 0,572322 | 1 | 0,792729 |
| -0,46757 | 420 | 372,2681 | 0,640365 | 1 | 0,964878 |
| -0,39295 | 367 | 332,2464 | 0,694612 | 1 | 0,904597 |
| 0,240806 | 280 | 266,1087 | 0,809891 | 1 | 0,95747 |
| -1,31201 | 421 | 403,6511 | 0,190263 | 1 | 0,474713 |
| 1,073674 | 421 | 214,6501 | 0,284174 | 1 | 0,582654 |
| -0,0036 | 420 | 415,4142 | 0,99713 | 1 | 0,997643 |
| 0,316823 | 367 | 362,4135 | 0,751561 | 1 | 0,924056 |
| -0,06984 | 280 | 272,2956 | 0,944374 | 1 | 0,982175 |
| -0,30748 | 421 | 404,9282 | 0,758637 | 1 | 0,900919 |
| -0,9308 | 421 | 220,5825 | 0,352974 | 1 | 0,643758 |
| 0,074184 | 420 | 415,9231 | 0,9409 | 1 | 0,992811 |
| -1,7348 | 367 | 362,8302 | 0,083625 | 1 | 0,386574 |
| -1,28441 | 280 | 275,4935 | 0,200076 | 1 | 0,631529 |
| -0,6213 | 421 | 382,2091 | 0,534774 | 1 | 0,778195 |
| -0,98632 | 421 | 202,7481 | 0,325153 | 1 | 0,619085 |
| -0,24848 | 420 | 406,9909 | 0,803887 | 1 | 0,969427 |
| 0,072518 | 367 | 351,2125 | 0,942231 | 1 | 0,979539 |
| -0,73039 | 280 | 267,1675 | 0,465791 | 1 | 0,824256 |
| -0,3757 | 421 | 367,1192 | 0,707355 | 1 | 0,875021 |
| -1,17041 | 421 | 191,7944 | 0,243288 | 1 | 0,542422 |
| -0,84835 | 420 | 399,3326 | 0,396753 | 1 | 0,95743 |
| -1,33985 | 367 | 349,1757 | 0,181166 | 1 | 0,527769 |
| -2,8342 | 280 | 264,3323 | 0,004949 | 1 | 0,177558 |
| -1,53845 | 421 | 345,4213 | 0,124854 | 1 | 0,38857 |
| -1,63819 | 421 | 190,6362 | 0,103031 | 1 | 0,383322 |
| -0,29265 | 420 | 380,2743 | 0,769953 | 1 | 0,969427 |
| -1,33616 | 367 | 352,7469 | 0,182358 | 1 | 0,527769 |
| -2,95228 | 280 | 266,4006 | 0,003436 | 1 | 0,143818 |
| 0,893946 | 421 | 384,4086 | 0,37191 | 1 | 0,663276 |
| 1,017986 | 421 | 205,305 | 0,309883 | 1 | 0,607274 |
| 2,009395 | 420 | 409,0968 | 0,045152 | 1 | 0,846132 |
| -0,55479 | 367 | 354,3473 | 0,579386 | 1 | 0,844137 |

| | | | | | |
|---|---|---|---|---|---|
| -0,8053 | 280 | 266,746 | 0,421366 | 1 | 0,796701 |
| -2,82945 | 421 | 412,3571 | 0,004891 | 1 | 0,087964 |
| -3,10753 | 421 | 232,2592 | 0,002122 | 1 | 0,099921 |
| -0,97716 | 420 | 413,622 | 0,329061 | 1 | 0,947373 |
| -1,43961 | 367 | 358,6006 | 0,150851 | 1 | 0,499998 |
| -1,05782 | 280 | 275,8943 | 0,291062 | 1 | 0,720248 |
| -0,98261 | 421 | 333,9896 | 0,326512 | 1 | 0,61749 |
| -1,5248 | 421 | 183,6554 | 0,129029 | 1 | 0,410224 |
| -1,23799 | 420 | 363,532 | 0,216517 | 1 | 0,943055 |
| -0,64604 | 367 | 312,9026 | 0,518727 | 1 | 0,801766 |
| 0,267938 | 280 | 258,527 | 0,788961 | 1 | 0,953435 |
| -1,69464 | 421 | 366,7113 | 0,090993 | 1 | 0,32265 |
| -2,03988 | 421 | 197,6985 | 0,042692 | 1 | 0,273775 |
| -0,89086 | 420 | 401,3289 | 0,37354 | 1 | 0,95743 |
| -0,7215 | 367 | 345,0804 | 0,471091 | 1 | 0,77576 |
| -0,82375 | 280 | 268,3824 | 0,410811 | 1 | 0,788151 |
| 1,15703 | 421 | 335,4402 | 0,248083 | 1 | 0,544195 |
| 0,563527 | 421 | 185,0919 | 0,573758 | 1 | 0,79326 |
| -1,73879 | 420 | 370,9826 | 0,082902 | 1 | 0,848246 |
| 3,222047 | 367 | 314,211 | 0,001406 | 1 | 0,176492 |
| 0,864642 | 280 | 258,5248 | 0,388037 | 1 | 0,775559 |
| 0,053806 | 421 | 325,9849 | 0,957123 | 1 | 0,987258 |
| -0,62679 | 421 | 185,5134 | 0,531568 | 1 | 0,769894 |
| -0,95691 | 420 | 359,1352 | 0,339257 | 1 | 0,947373 |
| -0,80045 | 367 | 302,716 | 0,424081 | 1 | 0,733267 |
| -1,09155 | 280 | 276 | 0,275984 | 1 | 0,701156 |
| -2,4255 | 421 | 348,5788 | 0,015795 | 1 | 0,147897 |
| -1,95802 | 421 | 192,0726 | 0,051676 | 1 | 0,296106 |
| -2,36439 | 420 | 386,7677 | 0,018553 | 1 | 0,75904 |
| -1,44106 | 367 | 348,7639 | 0,150465 | 1 | 0,499998 |
| -0,7218 | 280 | 269,5498 | 0,471045 | 1 | 0,824256 |
| -0,74155 | 421 | 354,1529 | 0,458854 | 1 | 0,724077 |
| 0,393357 | 421 | 192,5742 | 0,694491 | 1 | 0,865526 |
| 1,055988 | 420 | 387,2666 | 0,291632 | 1 | 0,943897 |
| -0,5835 | 367 | 337,7339 | 0,559948 | 1 | 0,833012 |
| -1,72793 | 280 | 267,8287 | 0,085154 | 1 | 0,45831 |
| -0,51776 | 421 | 342,5019 | 0,604961 | 1 | 0,822071 |
| 0,343628 | 421 | 187,5824 | 0,731511 | 1 | 0,882616 |
| -0,4769 | 420 | 376,7183 | 0,63371 | 1 | 0,964878 |
| -1,0894 | 367 | 328,8757 | 0,276774 | 1 | 0,611884 |
| 0,445242 | 280 | 266,7619 | 0,656506 | 1 | 0,908644 |
| -2,00042 | 421 | 349,3967 | 0,046228 | 1 | 0,238583 |
| -1,38701 | 421 | 188,1661 | 0,167079 | 1 | 0,453987 |
| -1,0897 | 420 | 385,199 | 0,276528 | 1 | 0,943897 |
| 0,323572 | 367 | 330,8476 | 0,746466 | 1 | 0,923669 |
| -2,57661 | 280 | 257,3843 | 0,010535 | 1 | 0,230098 |
| -1,38434 | 421 | 320,6223 | 0,167217 | 1 | 0,441901 |
| -1,2499 | 421 | 181,3554 | 0,212946 | 1 | 0,512644 |

| | | | | | |
|---|---|---|---|---|---|
| -0,89811 | 420 | 352,1508 | 0,369742 | 1 | 0,95743 |
| -0,67051 | 367 | 316,0825 | 0,50302 | 1 | 0,793601 |
| -0,92158 | 280 | 276 | 0,357554 | 1 | 0,761846 |
| -1,49305 | 421 | 365,64 | 0,136288 | 1 | 0,407576 |
| -0,85318 | 421 | 198,6994 | 0,394588 | 1 | 0,665895 |
| -0,85642 | 420 | 400,4861 | 0,392279 | 1 | 0,95743 |
| -0,96219 | 367 | 340,7691 | 0,336637 | 1 | 0,662616 |
| 0,9171 | 280 | 259,0943 | 0,359943 | 1 | 0,761846 |
| -1,72241 | 421 | 348,5686 | 0,085882 | 1 | 0,31262 |
| -1,59259 | 421 | 190,796 | 0,112907 | 1 | 0,391524 |
| 1,205212 | 420 | 386,3655 | 0,228859 | 1 | 0,943055 |
| -2,70722 | 367 | 346,043 | 0,007122 | 1 | 0,180403 |
| -1,96978 | 280 | 263,1488 | 0,049912 | 1 | 0,39442 |
| -1,25519 | 421 | 336,3748 | 0,21028 | 1 | 0,499043 |
| -1,86708 | 421 | 187,4607 | 0,063452 | 1 | 0,323047 |
| 0,286922 | 420 | 371,9979 | 0,774332 | 1 | 0,969427 |
| 0,898182 | 367 | 300,5555 | 0,369808 | 1 | 0,679634 |
| -0,05934 | 280 | 254,4199 | 0,952726 | 1 | 0,983396 |
| -0,10769 | 421 | 322,1036 | 0,914312 | 1 | 0,966828 |
| 0,027662 | 421 | 183,5769 | 0,977962 | 1 | 0,991789 |
| 0,702118 | 420 | 354,3023 | 0,483066 | 1 | 0,960096 |
| -0,20666 | 367 | 329,7669 | 0,836405 | 1 | 0,956935 |
| -0,43899 | 280 | 260,495 | 0,661036 | 1 | 0,908644 |
| 1,440573 | 421 | 333,7691 | 0,150643 | 1 | 0,424072 |
| 1,510256 | 421 | 183,809 | 0,132695 | 1 | 0,414338 |
| 0,468002 | 420 | 370,5698 | 0,640058 | 1 | 0,964878 |
| 0,432093 | 367 | 317,3803 | 0,665968 | 1 | 0,894765 |
| -0,65576 | 280 | 253,8079 | 0,51257 | 1 | 0,831912 |
| -1,72936 | 421 | 335,1813 | 0,084666 | 1 | 0,309688 |
| -1,8849 | 421 | 187,5289 | 0,06099 | 1 | 0,31585 |
| -1,98164 | 420 | 376,3541 | 0,048247 | 1 | 0,846627 |
| -0,59126 | 367 | 315,9076 | 0,554769 | 1 | 0,830257 |
| 0,658733 | 280 | 253,816 | 0,510664 | 1 | 0,831912 |
| -1,65448 | 421 | 299,4747 | 0,099079 | 1 | 0,339344 |
| -0,85433 | 421 | 178,9454 | 0,394065 | 1 | 0,665792 |
| -0,36615 | 420 | 328,0285 | 0,714489 | 1 | 0,968866 |
| -1,19523 | 367 | 300,4799 | 0,23294 | 1 | 0,582158 |
| -1,34747 | 280 | 276 | 0,178935 | 1 | 0,605081 |
| -0,90297 | 421 | 331,3802 | 0,367198 | 1 | 0,658771 |
| -0,25064 | 421 | 186,6549 | 0,802366 | 1 | 0,922323 |
| -0,66106 | 420 | 366,4243 | 0,508989 | 1 | 0,960096 |
| 0,683311 | 367 | 318,9748 | 0,494906 | 1 | 0,793601 |
| -1,30029 | 280 | 262,5247 | 0,194643 | 1 | 0,62543 |
| -0,15526 | 421 | 373,9497 | 0,876698 | 1 | 0,95861 |
| -0,44367 | 421 | 199,7456 | 0,657763 | 1 | 0,841515 |
| 2,290948 | 420 | 403,7838 | 0,022481 | 1 | 0,75904 |
| -0,01363 | 367 | 351,2969 | 0,989135 | 1 | 0,996248 |
| 1,974315 | 280 | 271,7826 | 0,049358 | 1 | 0,39442 |

| | | | | | |
|---|---|---|---|---|---|
| 2,016618 | 421 | 384,1685 | 0,044431 | 1 | 0,236315 |
| 1,660994 | 421 | 204,0019 | 0,098251 | 1 | 0,379251 |
| 0,211717 | 420 | 409,5479 | 0,832433 | 1 | 0,973894 |
| 2,364092 | 367 | 359,104 | 0,018606 | 1 | 0,217108 |
| 0,990429 | 280 | 275,539 | 0,322833 | 1 | 0,745172 |
| 0,322431 | 421 | 311,3955 | 0,747343 | 1 | 0,898122 |
| 0,030555 | 421 | 179,8437 | 0,975658 | 1 | 0,991789 |
| 1,545063 | 420 | 344,0874 | 0,12325 | 1 | 0,87201 |
| 0,282296 | 367 | 306,2039 | 0,777907 | 1 | 0,942368 |
| -0,19788 | 280 | 276 | 0,843283 | 1 | 0,965181 |
| -1,74711 | 421 | 379,8534 | 0,081426 | 1 | 0,30376 |
| -2,13005 | 421 | 201,8678 | 0,034378 | 1 | 0,252759 |
| -0,07232 | 420 | 406,1289 | 0,942381 | 1 | 0,992811 |
| -1,18833 | 367 | 359,7339 | 0,235487 | 1 | 0,585627 |
| -2,8884 | 280 | 265,6284 | 0,004191 | 1 | 0,157901 |
| -0,64952 | 421 | 321,5999 | 0,516467 | 1 | 0,767364 |
| -2,82307 | 421 | 183,563 | 0,005282 | 1 | 0,139638 |
| -1,91871 | 420 | 353,2891 | 0,055827 | 1 | 0,848246 |
| -1,51964 | 367 | 308,4635 | 0,129627 | 1 | 0,473621 |
| -1,13917 | 280 | 249,864 | 0,255723 | 1 | 0,679673 |
| 2,356133 | 421 | 360,4913 | 0,019001 | 1 | 0,159771 |
| 2,076425 | 421 | 191,8405 | 0,039187 | 1 | 0,26998 |
| 1,330916 | 420 | 395,9954 | 0,183982 | 1 | 0,923408 |
| 1,726266 | 367 | 340,5851 | 0,085206 | 1 | 0,388246 |
| 3,044095 | 280 | 271,6828 | 0,002563 | 1 | 0,135787 |
| 3,412009 | 421 | 387,9929 | 7,13E-04 | 0,758315 | 0,051145 |
| 2,60506 | 421 | 211,185 | 0,009839 | 1 | 0,149771 |
| 1,182479 | 420 | 413,6583 | 0,237695 | 1 | 0,943055 |
| 2,786282 | 367 | 358,8268 | 0,005615 | 1 | 0,178183 |
| 2,015045 | 280 | 275,9795 | 0,044869 | 1 | 0,387749 |
| 0,3421 | 421 | 387,5638 | 0,732461 | 1 | 0,893058 |
| 1,400941 | 421 | 207,6276 | 0,162724 | 1 | 0,446677 |
| 1,396311 | 420 | 410,5616 | 0,163375 | 1 | 0,914005 |
| -0,60964 | 367 | 356,9467 | 0,542487 | 1 | 0,824121 |
| 0,578757 | 280 | 267,5444 | 0,56324 | 1 | 0,861728 |
| -2,46131 | 421 | 300,8974 | 0,014404 | 1 | 0,146671 |
| -1,09399 | 421 | 175,6288 | 0,275456 | 1 | 0,576759 |
| -0,59448 | 420 | 342,4395 | 0,552583 | 1 | 0,960096 |
| -3,18578 | 367 | 303,1749 | 0,001594 | 1 | 0,176492 |
| -2,14713 | 280 | 254,1944 | 0,032729 | 1 | 0,329568 |
| -0,36548 | 421 | 320,6501 | 0,714996 | 1 | 0,881149 |
| 0,294444 | 421 | 179,2458 | 0,768759 | 1 | 0,905801 |
| -0,07947 | 420 | 355,4728 | 0,936706 | 1 | 0,992059 |
| 0,574832 | 367 | 308,6863 | 0,565824 | 1 | 0,837838 |
| -0,0988 | 280 | 257,8714 | 0,921374 | 1 | 0,977136 |
| -3,0293 | 421 | 386,7898 | 0,002616 | 1 | 0,073944 |
| -1,82879 | 421 | 206,4779 | 0,068874 | 1 | 0,335504 |
| -0,78531 | 420 | 410,9803 | 0,432726 | 1 | 0,960096 |

| | | | | | |
|---|---|---|---|---|---|
| -1,76559 | 367 | 359,8192 | 0,078313 | 1 | 0,381638 |
| -1,87443 | 280 | 274,3596 | 0,061933 | 1 | 0,422605 |
| -0,57211 | 421 | 356,2084 | 0,56761 | 1 | 0,796637 |
| -0,4211 | 421 | 189,9094 | 0,674159 | 1 | 0,853746 |
| -0,21736 | 420 | 388,6587 | 0,828045 | 1 | 0,973894 |
| 0,376707 | 367 | 330,735 | 0,706633 | 1 | 0,91071 |
| -0,90131 | 280 | 261,2182 | 0,368255 | 1 | 0,763914 |
| -0,10753 | 421 | 349,9041 | 0,914431 | 1 | 0,966828 |
| 0,773194 | 421 | 187,9072 | 0,440379 | 1 | 0,705407 |
| -0,25458 | 420 | 382,9977 | 0,799186 | 1 | 0,969427 |
| 0,405144 | 367 | 337,0403 | 0,685629 | 1 | 0,902395 |
| 1,359468 | 280 | 254,4292 | 0,175202 | 1 | 0,598625 |
| -0,13847 | 421 | 351,9567 | 0,889952 | 1 | 0,962 |
| 2,637997 | 421 | 191,8419 | 0,009024 | 1 | 0,14815 |
| 1,781552 | 420 | 384,6462 | 0,075611 | 1 | 0,848246 |
| 3,033374 | 367 | 334,0667 | 0,002608 | 1 | 0,176492 |
| 1,472479 | 280 | 250,2707 | 0,142148 | 1 | 0,554427 |
| -0,1199 | 421 | 399,7748 | 0,904624 | 1 | 0,965673 |
| 3,26158 | 421 | 213,0474 | 0,00129 | 1 | 0,092971 |
| 0,804868 | 420 | 414,9107 | 0,421357 | 1 | 0,95743 |
| 1,259769 | 367 | 362,2098 | 0,208564 | 1 | 0,56126 |
| -0,31628 | 280 | 273,2312 | 0,752031 | 1 | 0,942071 |
| 0,705982 | 421 | 326,0142 | 0,480703 | 1 | 0,740716 |
| 0,399493 | 421 | 181,6587 | 0,69 | 1 | 0,864363 |
| -0,37164 | 420 | 359,9845 | 0,710377 | 1 | 0,968866 |
| 0,669377 | 367 | 320,5871 | 0,503737 | 1 | 0,793601 |
| 0,704706 | 280 | 254,9061 | 0,481637 | 1 | 0,824339 |
| -0,11508 | 421 | 323,8627 | 0,90845 | 1 | 0,966828 |
| 2,012363 | 421 | 182,9412 | 0,045648 | 1 | 0,281076 |
| 0,953172 | 420 | 358,2234 | 0,341146 | 1 | 0,947373 |
| 0,274221 | 367 | 315,0477 | 0,784094 | 1 | 0,94337 |
| 0,343537 | 280 | 252,2405 | 0,731481 | 1 | 0,936637 |
| 0,014042 | 421 | 358,7818 | 0,988804 | 1 | 0,998054 |
| 0,018613 | 421 | 192,9728 | 0,985169 | 1 | 0,995074 |
| -0,47339 | 420 | 392,5891 | 0,636198 | 1 | 0,964878 |
| -0,30297 | 367 | 339,4662 | 0,762096 | 1 | 0,929966 |
| 0,009766 | 280 | 261,7127 | 0,992215 | 1 | 0,996269 |
| -1,63598 | 421 | 334,1571 | 0,102785 | 1 | 0,344557 |
| -0,85747 | 421 | 187,1859 | 0,392284 | 1 | 0,665733 |
| -1,17277 | 420 | 369,3536 | 0,241643 | 1 | 0,943659 |
| -1,29743 | 367 | 324,748 | 0,195405 | 1 | 0,549567 |
| 0,367672 | 280 | 257,5062 | 0,71342 | 1 | 0,928432 |
| -0,13171 | 421 | 328,7364 | 0,895291 | 1 | 0,962 |
| -0,05187 | 421 | 182,4923 | 0,958691 | 1 | 0,98414 |
| -0,76787 | 420 | 362,9392 | 0,443064 | 1 | 0,960096 |
| 2,180701 | 367 | 306,4423 | 0,029966 | 1 | 0,261032 |
| 0,710641 | 280 | 252,2844 | 0,477963 | 1 | 0,824256 |
| 1,444376 | 421 | 340,4516 | 0,149553 | 1 | 0,422053 |

| | | | | | | |
|---|---|---|---|---|---|---|
| 0,264981 | 421 | 186,5692 | 0,791317 | | 1 | 0,918572 |
| 1,227244 | 420 | 369,9486 | 0,220511 | | 1 | 0,943055 |
| 0,228973 | 367 | 327,2746 | 0,819033 | | 1 | 0,951644 |
| -2,44601 | 280 | 255,211 | 0,015121 | | 1 | 0,263522 |

Model formula fit
y ~ current_feature + Sex + (1 | Subject) + storage_time
y ~ current_feature + Sex + (1 | Subject) + storage_time
y ~ current_feature + Sex + (1 | Subject) + storage_time
y ~ current_feature + Sex + (1 | Subject) + storage_time
y ~ current_feature + Sex + (1 | Subject) + storage_time
y ~ current_feature + Sex + (1 | Subject) + storage_time
y ~ current_feature + Sex + (1 | Subject) + storage_time
y ~ current_feature + Sex + (1 | Subject) + storage_time
y ~ current_feature + Sex + (1 | Subject) + storage_time
y ~ current_feature + Sex + (1 | Subject) + storage_time
y ~ current_feature + Sex + (1 | Subject) + storage_time
y ~ current_feature + Sex + (1 | Subject) + storage_time
y ~ current_feature + Sex + (1 | Subject) + storage_time
y ~ current_feature + Sex + (1 | Subject) + storage_time
y ~ current_feature + Sex + (1 | Subject) + storage_time
y ~ current_feature + Sex + (1 | Subject) + storage_time
y ~ current_feature + Sex + (1 | Subject) + storage_time
y ~ current_feature + Sex + (1 | Subject) + storage_time
y ~ current_feature + Sex + (1 | Subject) + storage_time
y ~ current_feature + Sex + (1 | Subject) + storage_time
y ~ current_feature + Sex + (1 | Subject) + storage_time
y ~ current_feature + Sex + (1 | Subject) + storage_time
y ~ current_feature + Sex + (1 | Subject) + storage_time
y ~ current_feature + Sex + (1 | Subject) + storage_time
y ~ current_feature + Sex + (1 | Subject) + storage_time
y ~ current_feature + Sex + (1 | Subject) + storage_time
y ~ current_feature + Sex + (1 | Subject) + storage_time
y ~ current_feature + Sex + (1 | Subject) + storage_time
y ~ current_feature + Sex + (1 | Subject) + storage_time
y ~ current_feature + Sex + (1 | Subject) + storage_time
y ~ current_feature + Sex + (1 | Subject) + storage_time
y ~ current_feature + Sex + (1 | Subject) + storage_time
y ~ current_feature + Sex + (1 | Subject) + storage_time
y ~ current_feature + Sex + (1 | Subject) + storage_time
y ~ current_feature + Sex + (1 | Subject) + storage_time
y ~ current_feature + Sex + (1 | Subject) + storage_time
y ~ current_feature + Sex + (1 | Subject) + storage_time
y ~ current_feature + Sex + (1 | Subject) + storage_time
y ~ current_feature + Sex + (1 | Subject) + storage_time
y ~ current_feature + Sex + (1 | Subject) + storage_time
y ~ current_feature + Sex + (1 | Subject) + storage_time
y ~ current_feature + Sex + (1 | Subject) + storage_time
y ~ current_feature + Sex + (1 | Subject) + storage_time
y ~ current_feature + Sex + (1 | Subject) + storage_time
y ~ current_feature + Sex + (1 | Subject) + storage_time
y ~ current_feature + Sex + (1 | Subject) + storage_time
y ~ current_feature + Sex + (1 | Subject) + storage_time

y ~ current_feature + Sex + (1 | Subject) + storage_time
y ~ current_feature + Sex + (1 | Subject) + storage_time
y ~ current_feature + Sex + (1 | Subject) + storage_time
y ~ current_feature + Sex + (1 | Subject) + storage_time
y ~ current_feature + Sex + (1 | Subject) + storage_time
y ~ current_feature + Sex + (1 | Subject) + storage_time
y ~ current_feature + Sex + (1 | Subject) + storage_time
y ~ current_feature + Sex + (1 | Subject) + storage_time
y ~ current_feature + Sex + (1 | Subject) + storage_time
y ~ current_feature + Sex + (1 | Subject) + storage_time
y ~ current_feature + Sex + (1 | Subject) + storage_time
y ~ current_feature + Sex + (1 | Subject) + storage_time
y ~ current_feature + Sex + (1 | Subject) + storage_time
y ~ current_feature + Sex + (1 | Subject) + storage_time
y ~ current_feature + Sex + (1 | Subject) + storage_time
y ~ current_feature + Sex + (1 | Subject) + storage_time
y ~ current_feature + Sex + (1 | Subject) + storage_time
y ~ current_feature + Sex + (1 | Subject) + storage_time
y ~ current_feature + Sex + (1 | Subject) + storage_time
y ~ current_feature + Sex + (1 | Subject) + storage_time
y ~ current_feature + Sex + (1 | Subject) + storage_time
y ~ current_feature + Sex + (1 | Subject) + storage_time
y ~ current_feature + Sex + (1 | Subject) + storage_time
y ~ current_feature + Sex + (1 | Subject) + storage_time
y ~ current_feature + Sex + (1 | Subject) + storage_time
y ~ current_feature + Sex + (1 | Subject) + storage_time
y ~ current_feature + Sex + (1 | Subject) + storage_time
y ~ current_feature + Sex + (1 | Subject) + storage_time
y ~ current_feature + Sex + (1 | Subject) + storage_time
y ~ current_feature + Sex + (1 | Subject) + storage_time
y ~ current_feature + Sex + (1 | Subject) + storage_time
y ~ current_feature + Sex + (1 | Subject) + storage_time
y ~ current_feature + Sex + (1 | Subject) + storage_time
y ~ current_feature + Sex + (1 | Subject) + storage_time
y ~ current_feature + Sex + (1 | Subject) + storage_time
y ~ current_feature + Sex + (1 | Subject) + storage_time
y ~ current_feature + Sex + (1 | Subject) + storage_time
y ~ current_feature + Sex + (1 | Subject) + storage_time
y ~ current_feature + Sex + (1 | Subject) + storage_time
y ~ current_feature + Sex + (1 | Subject) + storage_time
y ~ current_feature + Sex + (1 | Subject) + storage_time
y ~ current_feature + Sex + (1 | Subject) + storage_time
y ~ current_feature + Sex + (1 | Subject) + storage_time
y ~ current_feature + Sex + (1 | Subject) + storage_time
y ~ current_feature + Sex + (1 | Subject) + storage_time
y ~ current_feature + Sex + (1 | Subject) + storage_time
y ~ current_feature + Sex + (1 | Subject) + storage_time
y ~ current_feature + Sex + (1 | Subject) + storage_time

y ~ current_feature + Sex + (1 | Subject) + storage_time
y ~ current_feature + Sex + (1 | Subject) + storage_time
y ~ current_feature + Sex + (1 | Subject) + storage_time
y ~ current_feature + Sex + (1 | Subject) + storage_time
y ~ current_feature + Sex + (1 | Subject) + storage_time
y ~ current_feature + Sex + (1 | Subject) + storage_time
y ~ current_feature + Sex + (1 | Subject) + storage_time
y ~ current_feature + Sex + (1 | Subject) + storage_time
y ~ current_feature + Sex + (1 | Subject) + storage_time
y ~ current_feature + Sex + (1 | Subject) + storage_time
y ~ current_feature + Sex + (1 | Subject) + storage_time
y ~ current_feature + Sex + (1 | Subject) + storage_time
y ~ current_feature + Sex + (1 | Subject) + storage_time
y ~ current_feature + Sex + (1 | Subject) + storage_time
y ~ current_feature + Sex + (1 | Subject) + storage_time
y ~ current_feature + Sex + (1 | Subject) + storage_time
y ~ current_feature + Sex + (1 | Subject) + storage_time
y ~ current_feature + Sex + (1 | Subject) + storage_time
y ~ current_feature + Sex + (1 | Subject) + storage_time
y ~ current_feature + Sex + (1 | Subject) + storage_time
y ~ current_feature + Sex + (1 | Subject) + storage_time
y ~ current_feature + Sex + (1 | Subject) + storage_time
y ~ current_feature + Sex + (1 | Subject) + storage_time
y ~ current_feature + Sex + (1 | Subject) + storage_time
y ~ current_feature + Sex + (1 | Subject) + storage_time
y ~ current_feature + Sex + (1 | Subject) + storage_time
y ~ current_feature + Sex + (1 | Subject) + storage_time
y ~ current_feature + Sex + (1 | Subject) + storage_time
y ~ current_feature + Sex + (1 | Subject) + storage_time
y ~ current_feature + Sex + (1 | Subject) + storage_time
y ~ current_feature + Sex + (1 | Subject) + storage_time
y ~ current_feature + Sex + (1 | Subject) + storage_time
y ~ current_feature + Sex + (1 | Subject) + storage_time
y ~ current_feature + Sex + (1 | Subject) + storage_time
y ~ current_feature + Sex + (1 | Subject) + storage_time
y ~ current_feature + Sex + (1 | Subject) + storage_time
y ~ current_feature + Sex + (1 | Subject) + storage_time
y ~ current_feature + Sex + (1 | Subject) + storage_time
y ~ current_feature + Sex + (1 | Subject) + storage_time
y ~ current_feature + Sex + (1 | Subject) + storage_time
y ~ current_feature + Sex + (1 | Subject) + storage_time
y ~ current_feature + Sex + (1 | Subject) + storage_time
y ~ current_feature + Sex + (1 | Subject) + storage_time
y ~ current_feature + Sex + (1 | Subject) + storage_time
y ~ current_feature + Sex + (1 | Subject) + storage_time
y ~ current_feature + Sex + (1 | Subject) + storage_time
y ~ current_feature + Sex + (1 | Subject) + storage_time
y ~ current_feature + Sex + (1 | Subject) + storage_time

y ~ current_feature + Sex + (1 | Subject) + storage_time
y ~ current_feature + Sex + (1 | Subject) + storage_time
y ~ current_feature + Sex + (1 | Subject) + storage_time
y ~ current_feature + Sex + (1 | Subject) + storage_time
y ~ current_feature + Sex + (1 | Subject) + storage_time
y ~ current_feature + Sex + (1 | Subject) + storage_time
y ~ current_feature + Sex + (1 | Subject) + storage_time
y ~ current_feature + Sex + (1 | Subject) + storage_time
y ~ current_feature + Sex + (1 | Subject) + storage_time
y ~ current_feature + Sex + (1 | Subject) + storage_time
y ~ current_feature + Sex + (1 | Subject) + storage_time
y ~ current_feature + Sex + (1 | Subject) + storage_time
y ~ current_feature + Sex + (1 | Subject) + storage_time
y ~ current_feature + Sex + (1 | Subject) + storage_time
y ~ current_feature + Sex + (1 | Subject) + storage_time
y ~ current_feature + Sex + (1 | Subject) + storage_time
y ~ current_feature + Sex + (1 | Subject) + storage_time
y ~ current_feature + Sex + (1 | Subject) + storage_time
y ~ current_feature + Sex + (1 | Subject) + storage_time
y ~ current_feature + Sex + (1 | Subject) + storage_time
y ~ current_feature + Sex + (1 | Subject) + storage_time
y ~ current_feature + Sex + (1 | Subject) + storage_time
y ~ current_feature + Sex + (1 | Subject) + storage_time
y ~ current_feature + Sex + (1 | Subject) + storage_time
y ~ current_feature + Sex + (1 | Subject) + storage_time
y ~ current_feature + Sex + (1 | Subject) + storage_time
y ~ current_feature + Sex + (1 | Subject) + storage_time
y ~ current_feature + Sex + (1 | Subject) + storage_time
y ~ current_feature + Sex + (1 | Subject) + storage_time
y ~ current_feature + Sex + (1 | Subject) + storage_time
y ~ current_feature + Sex + (1 | Subject) + storage_time
y ~ current_feature + Sex + (1 | Subject) + storage_time
y ~ current_feature + Sex + (1 | Subject) + storage_time
y ~ current_feature + Sex + (1 | Subject) + storage_time
y ~ current_feature + Sex + (1 | Subject) + storage_time
y ~ current_feature + Sex + (1 | Subject) + storage_time
y ~ current_feature + Sex + (1 | Subject) + storage_time
y ~ current_feature + Sex + (1 | Subject) + storage_time
y ~ current_feature + Sex + (1 | Subject) + storage_time
y ~ current_feature + Sex + (1 | Subject) + storage_time
y ~ current_feature + Sex + (1 | Subject) + storage_time
y ~ current_feature + Sex + (1 | Subject) + storage_time
y ~ current_feature + Sex + (1 | Subject) + storage_time
y ~ current_feature + Sex + (1 | Subject) + storage_time
y ~ current_feature + Sex + (1 | Subject) + storage_time
y ~ current_feature + Sex + (1 | Subject) + storage_time
y ~ current_feature + Sex + (1 | Subject) + storage_time
y ~ current_feature + Sex + (1 | Subject) + storage_time

y ~ current_feature + Sex + (1 | Subject) + storage_time
y ~ current_feature + Sex + (1 | Subject) + storage_time
y ~ current_feature + Sex + (1 | Subject) + storage_time
y ~ current_feature + Sex + (1 | Subject) + storage_time
y ~ current_feature + Sex + (1 | Subject) + storage_time
y ~ current_feature + Sex + (1 | Subject) + storage_time
y ~ current_feature + Sex + (1 | Subject) + storage_time
y ~ current_feature + Sex + (1 | Subject) + storage_time
y ~ current_feature + Sex + (1 | Subject) + storage_time
y ~ current_feature + Sex + (1 | Subject) + storage_time
y ~ current_feature + Sex + (1 | Subject) + storage_time
y ~ current_feature + Sex + (1 | Subject) + storage_time
y ~ current_feature + Sex + (1 | Subject) + storage_time
y ~ current_feature + Sex + (1 | Subject) + storage_time
y ~ current_feature + Sex + (1 | Subject) + storage_time
y ~ current_feature + Sex + (1 | Subject) + storage_time
y ~ current_feature + Sex + (1 | Subject) + storage_time
y ~ current_feature + Sex + (1 | Subject) + storage_time
y ~ current_feature + Sex + (1 | Subject) + storage_time
y ~ current_feature + Sex + (1 | Subject) + storage_time
y ~ current_feature + Sex + (1 | Subject) + storage_time
y ~ current_feature + Sex + (1 | Subject) + storage_time
y ~ current_feature + Sex + (1 | Subject) + storage_time
y ~ current_feature + Sex + (1 | Subject) + storage_time
y ~ current_feature + Sex + (1 | Subject) + storage_time
y ~ current_feature + Sex + (1 | Subject) + storage_time
y ~ current_feature + Sex + (1 | Subject) + storage_time
y ~ current_feature + Sex + (1 | Subject) + storage_time
y ~ current_feature + Sex + (1 | Subject) + storage_time
y ~ current_feature + Sex + (1 | Subject) + storage_time
y ~ current_feature + Sex + (1 | Subject) + storage_time
y ~ current_feature + Sex + (1 | Subject) + storage_time
y ~ current_feature + Sex + (1 | Subject) + storage_time
y ~ current_feature + Sex + (1 | Subject) + storage_time
y ~ current_feature + Sex + (1 | Subject) + storage_time
y ~ current_feature + Sex + (1 | Subject) + storage_time
y ~ current_feature + Sex + (1 | Subject) + storage_time
y ~ current_feature + Sex + (1 | Subject) + storage_time
y ~ current_feature + Sex + (1 | Subject) + storage_time
y ~ current_feature + Sex + (1 | Subject) + storage_time
y ~ current_feature + Sex + (1 | Subject) + storage_time
y ~ current_feature + Sex + (1 | Subject) + storage_time
y ~ current_feature + Sex + (1 | Subject) + storage_time
y ~ current_feature + Sex + (1 | Subject) + storage_time
y ~ current_feature + Sex + (1 | Subject) + storage_time
y ~ current_feature + Sex + (1 | Subject) + storage_time
y ~ current_feature + Sex + (1 | Subject) + storage_time
y ~ current_feature + Sex + (1 | Subject) + storage_time

y ~ current_feature + Sex + (1 | Subject) + storage_time
y ~ current_feature + Sex + (1 | Subject) + storage_time
y ~ current_feature + Sex + (1 | Subject) + storage_time
y ~ current_feature + Sex + (1 | Subject) + storage_time
y ~ current_feature + Sex + (1 | Subject) + storage_time
y ~ current_feature + Sex + (1 | Subject) + storage_time
y ~ current_feature + Sex + (1 | Subject) + storage_time
y ~ current_feature + Sex + (1 | Subject) + storage_time
y ~ current_feature + Sex + (1 | Subject) + storage_time
y ~ current_feature + Sex + (1 | Subject) + storage_time
y ~ current_feature + Sex + (1 | Subject) + storage_time
y ~ current_feature + Sex + (1 | Subject) + storage_time
y ~ current_feature + Sex + (1 | Subject) + storage_time
y ~ current_feature + Sex + (1 | Subject) + storage_time
y ~ current_feature + Sex + (1 | Subject) + storage_time
y ~ current_feature + Sex + (1 | Subject) + storage_time
y ~ current_feature + Sex + (1 | Subject) + storage_time
y ~ current_feature + Sex + (1 | Subject) + storage_time
y ~ current_feature + Sex + (1 | Subject) + storage_time
y ~ current_feature + Sex + (1 | Subject) + storage_time
y ~ current_feature + Sex + (1 | Subject) + storage_time
y ~ current_feature + Sex + (1 | Subject) + storage_time
y ~ current_feature + Sex + (1 | Subject) + storage_time
y ~ current_feature + Sex + (1 | Subject) + storage_time
y ~ current_feature + Sex + (1 | Subject) + storage_time
y ~ current_feature + Sex + (1 | Subject) + storage_time
y ~ current_feature + Sex + (1 | Subject) + storage_time
y ~ current_feature + Sex + (1 | Subject) + storage_time
y ~ current_feature + Sex + (1 | Subject) + storage_time
y ~ current_feature + Sex + (1 | Subject) + storage_time
y ~ current_feature + Sex + (1 | Subject) + storage_time
y ~ current_feature + Sex + (1 | Subject) + storage_time
y ~ current_feature + Sex + (1 | Subject) + storage_time
y ~ current_feature + Sex + (1 | Subject) + storage_time
y ~ current_feature + Sex + (1 | Subject) + storage_time
y ~ current_feature + Sex + (1 | Subject) + storage_time
y ~ current_feature + Sex + (1 | Subject) + storage_time
y ~ current_feature + Sex + (1 | Subject) + storage_time
y ~ current_feature + Sex + (1 | Subject) + storage_time
y ~ current_feature + Sex + (1 | Subject) + storage_time
y ~ current_feature + Sex + (1 | Subject) + storage_time
y ~ current_feature + Sex + (1 | Subject) + storage_time
y ~ current_feature + Sex + (1 | Subject) + storage_time
y ~ current_feature + Sex + (1 | Subject) + storage_time
y ~ current_feature + Sex + (1 | Subject) + storage_time
y ~ current_feature + Sex + (1 | Subject) + storage_time
y ~ current_feature + Sex + (1 | Subject) + storage_time
y ~ current_feature + Sex + (1 | Subject) + storage_time

y ~ current_feature + Sex + (1 | Subject) + storage_time
y ~ current_feature + Sex + (1 | Subject) + storage_time
y ~ current_feature + Sex + (1 | Subject) + storage_time
y ~ current_feature + Sex + (1 | Subject) + storage_time
y ~ current_feature + Sex + (1 | Subject) + storage_time
y ~ current_feature + Sex + (1 | Subject) + storage_time
y ~ current_feature + Sex + (1 | Subject) + storage_time
y ~ current_feature + Sex + (1 | Subject) + storage_time
y ~ current_feature + Sex + (1 | Subject) + storage_time
y ~ current_feature + Sex + (1 | Subject) + storage_time
y ~ current_feature + Sex + (1 | Subject) + storage_time
y ~ current_feature + Sex + (1 | Subject) + storage_time
y ~ current_feature + Sex + (1 | Subject) + storage_time
y ~ current_feature + Sex + (1 | Subject) + storage_time
y ~ current_feature + Sex + (1 | Subject) + storage_time
y ~ current_feature + Sex + (1 | Subject) + storage_time
y ~ current_feature + Sex + (1 | Subject) + storage_time
y ~ current_feature + Sex + (1 | Subject) + storage_time
y ~ current_feature + Sex + (1 | Subject) + storage_time
y ~ current_feature + Sex + (1 | Subject) + storage_time
y ~ current_feature + Sex + (1 | Subject) + storage_time
y ~ current_feature + Sex + (1 | Subject) + storage_time
y ~ current_feature + Sex + (1 | Subject) + storage_time
y ~ current_feature + Sex + (1 | Subject) + storage_time
y ~ current_feature + Sex + (1 | Subject) + storage_time
y ~ current_feature + Sex + (1 | Subject) + storage_time
y ~ current_feature + Sex + (1 | Subject) + storage_time
y ~ current_feature + Sex + (1 | Subject) + storage_time
y ~ current_feature + Sex + (1 | Subject) + storage_time
y ~ current_feature + Sex + (1 | Subject) + storage_time
y ~ current_feature + Sex + (1 | Subject) + storage_time
y ~ current_feature + Sex + (1 | Subject) + storage_time
y ~ current_feature + Sex + (1 | Subject) + storage_time
y ~ current_feature + Sex + (1 | Subject) + storage_time
y ~ current_feature + Sex + (1 | Subject) + storage_time
y ~ current_feature + Sex + (1 | Subject) + storage_time
y ~ current_feature + Sex + (1 | Subject) + storage_time
y ~ current_feature + Sex + (1 | Subject) + storage_time
y ~ current_feature + Sex + (1 | Subject) + storage_time
y ~ current_feature + Sex + (1 | Subject) + storage_time
y ~ current_feature + Sex + (1 | Subject) + storage_time
y ~ current_feature + Sex + (1 | Subject) + storage_time
y ~ current_feature + Sex + (1 | Subject) + storage_time
y ~ current_feature + Sex + (1 | Subject) + storage_time
y ~ current_feature + Sex + (1 | Subject) + storage_time
y ~ current_feature + Sex + (1 | Subject) + storage_time
y ~ current_feature + Sex + (1 | Subject) + storage_time
y ~ current_feature + Sex + (1 | Subject) + storage_time

y ~ current_feature + Sex + (1 | Subject) + storage_time
y ~ current_feature + Sex + (1 | Subject) + storage_time
y ~ current_feature + Sex + (1 | Subject) + storage_time
y ~ current_feature + Sex + (1 | Subject) + storage_time
y ~ current_feature + Sex + (1 | Subject) + storage_time
y ~ current_feature + Sex + (1 | Subject) + storage_time
y ~ current_feature + Sex + (1 | Subject) + storage_time
y ~ current_feature + Sex + (1 | Subject) + storage_time
y ~ current_feature + Sex + (1 | Subject) + storage_time
y ~ current_feature + Sex + (1 | Subject) + storage_time
y ~ current_feature + Sex + (1 | Subject) + storage_time
y ~ current_feature + Sex + (1 | Subject) + storage_time
y ~ current_feature + Sex + (1 | Subject) + storage_time
y ~ current_feature + Sex + (1 | Subject) + storage_time
y ~ current_feature + Sex + (1 | Subject) + storage_time
y ~ current_feature + Sex + (1 | Subject) + storage_time
y ~ current_feature + Sex + (1 | Subject) + storage_time
y ~ current_feature + Sex + (1 | Subject) + storage_time
y ~ current_feature + Sex + (1 | Subject) + storage_time
y ~ current_feature + Sex + (1 | Subject) + storage_time
y ~ current_feature + Sex + (1 | Subject) + storage_time
y ~ current_feature + Sex + (1 | Subject) + storage_time
y ~ current_feature + Sex + (1 | Subject) + storage_time
y ~ current_feature + Sex + (1 | Subject) + storage_time
y ~ current_feature + Sex + (1 | Subject) + storage_time
y ~ current_feature + Sex + (1 | Subject) + storage_time
y ~ current_feature + Sex + (1 | Subject) + storage_time
y ~ current_feature + Sex + (1 | Subject) + storage_time
y ~ current_feature + Sex + (1 | Subject) + storage_time
y ~ current_feature + Sex + (1 | Subject) + storage_time
y ~ current_feature + Sex + (1 | Subject) + storage_time
y ~ current_feature + Sex + (1 | Subject) + storage_time
y ~ current_feature + Sex + (1 | Subject) + storage_time
y ~ current_feature + Sex + (1 | Subject) + storage_time
y ~ current_feature + Sex + (1 | Subject) + storage_time
y ~ current_feature + Sex + (1 | Subject) + storage_time
y ~ current_feature + Sex + (1 | Subject) + storage_time
y ~ current_feature + Sex + (1 | Subject) + storage_time
y ~ current_feature + Sex + (1 | Subject) + storage_time
y ~ current_feature + Sex + (1 | Subject) + storage_time
y ~ current_feature + Sex + (1 | Subject) + storage_time
y ~ current_feature + Sex + (1 | Subject) + storage_time
y ~ current_feature + Sex + (1 | Subject) + storage_time
y ~ current_feature + Sex + (1 | Subject) + storage_time
y ~ current_feature + Sex + (1 | Subject) + storage_time
y ~ current_feature + Sex + (1 | Subject) + storage_time
y ~ current_feature + Sex + (1 | Subject) + storage_time
y ~ current_feature + Sex + (1 | Subject) + storage_time

y ~ current_feature + Sex + (1 | Subject) + storage_time
y ~ current_feature + Sex + (1 | Subject) + storage_time
y ~ current_feature + Sex + (1 | Subject) + storage_time
y ~ current_feature + Sex + (1 | Subject) + storage_time
y ~ current_feature + Sex + (1 | Subject) + storage_time
y ~ current_feature + Sex + (1 | Subject) + storage_time
y ~ current_feature + Sex + (1 | Subject) + storage_time
y ~ current_feature + Sex + (1 | Subject) + storage_time
y ~ current_feature + Sex + (1 | Subject) + storage_time
y ~ current_feature + Sex + (1 | Subject) + storage_time
y ~ current_feature + Sex + (1 | Subject) + storage_time
y ~ current_feature + Sex + (1 | Subject) + storage_time
y ~ current_feature + Sex + (1 | Subject) + storage_time
y ~ current_feature + Sex + (1 | Subject) + storage_time
y ~ current_feature + Sex + (1 | Subject) + storage_time
y ~ current_feature + Sex + (1 | Subject) + storage_time
y ~ current_feature + Sex + (1 | Subject) + storage_time
y ~ current_feature + Sex + (1 | Subject) + storage_time
y ~ current_feature + Sex + (1 | Subject) + storage_time
y ~ current_feature + Sex + (1 | Subject) + storage_time
y ~ current_feature + Sex + (1 | Subject) + storage_time
y ~ current_feature + Sex + (1 | Subject) + storage_time
y ~ current_feature + Sex + (1 | Subject) + storage_time
y ~ current_feature + Sex + (1 | Subject) + storage_time
y ~ current_feature + Sex + (1 | Subject) + storage_time
y ~ current_feature + Sex + (1 | Subject) + storage_time
y ~ current_feature + Sex + (1 | Subject) + storage_time
y ~ current_feature + Sex + (1 | Subject) + storage_time
y ~ current_feature + Sex + (1 | Subject) + storage_time
y ~ current_feature + Sex + (1 | Subject) + storage_time
y ~ current_feature + Sex + (1 | Subject) + storage_time
y ~ current_feature + Sex + (1 | Subject) + storage_time
y ~ current_feature + Sex + (1 | Subject) + storage_time
y ~ current_feature + Sex + (1 | Subject) + storage_time
y ~ current_feature + Sex + (1 | Subject) + storage_time
y ~ current_feature + Sex + (1 | Subject) + storage_time
y ~ current_feature + Sex + (1 | Subject) + storage_time
y ~ current_feature + Sex + (1 | Subject) + storage_time
y ~ current_feature + Sex + (1 | Subject) + storage_time
y ~ current_feature + Sex + (1 | Subject) + storage_time
y ~ current_feature + Sex + (1 | Subject) + storage_time
y ~ current_feature + Sex + (1 | Subject) + storage_time
y ~ current_feature + Sex + (1 | Subject) + storage_time
y ~ current_feature + Sex + (1 | Subject) + storage_time
y ~ current_feature + Sex + (1 | Subject) + storage_time
y ~ current_feature + Sex + (1 | Subject) + storage_time
y ~ current_feature + Sex + (1 | Subject) + storage_time
y ~ current_feature + Sex + (1 | Subject) + storage_time

y ~ current_feature + Sex + (1 | Subject) + storage_time
y ~ current_feature + Sex + (1 | Subject) + storage_time
y ~ current_feature + Sex + (1 | Subject) + storage_time
y ~ current_feature + Sex + (1 | Subject) + storage_time
y ~ current_feature + Sex + (1 | Subject) + storage_time
y ~ current_feature + Sex + (1 | Subject) + storage_time
y ~ current_feature + Sex + (1 | Subject) + storage_time
y ~ current_feature + Sex + (1 | Subject) + storage_time
y ~ current_feature + Sex + (1 | Subject) + storage_time
y ~ current_feature + Sex + (1 | Subject) + storage_time
y ~ current_feature + Sex + (1 | Subject) + storage_time
y ~ current_feature + Sex + (1 | Subject) + storage_time
y ~ current_feature + Sex + (1 | Subject) + storage_time
y ~ current_feature + Sex + (1 | Subject) + storage_time
y ~ current_feature + Sex + (1 | Subject) + storage_time
y ~ current_feature + Sex + (1 | Subject) + storage_time
y ~ current_feature + Sex + (1 | Subject) + storage_time
y ~ current_feature + Sex + (1 | Subject) + storage_time
y ~ current_feature + Sex + (1 | Subject) + storage_time
y ~ current_feature + Sex + (1 | Subject) + storage_time
y ~ current_feature + Sex + (1 | Subject) + storage_time
y ~ current_feature + Sex + (1 | Subject) + storage_time
y ~ current_feature + Sex + (1 | Subject) + storage_time
y ~ current_feature + Sex + (1 | Subject) + storage_time
y ~ current_feature + Sex + (1 | Subject) + storage_time
y ~ current_feature + Sex + (1 | Subject) + storage_time
y ~ current_feature + Sex + (1 | Subject) + storage_time
y ~ current_feature + Sex + (1 | Subject) + storage_time
y ~ current_feature + Sex + (1 | Subject) + storage_time
y ~ current_feature + Sex + (1 | Subject) + storage_time
y ~ current_feature + Sex + (1 | Subject) + storage_time
y ~ current_feature + Sex + (1 | Subject) + storage_time
y ~ current_feature + Sex + (1 | Subject) + storage_time
y ~ current_feature + Sex + (1 | Subject) + storage_time
y ~ current_feature + Sex + (1 | Subject) + storage_time
y ~ current_feature + Sex + (1 | Subject) + storage_time
y ~ current_feature + Sex + (1 | Subject) + storage_time
y ~ current_feature + Sex + (1 | Subject) + storage_time
y ~ current_feature + Sex + (1 | Subject) + storage_time
y ~ current_feature + Sex + (1 | Subject) + storage_time
y ~ current_feature + Sex + (1 | Subject) + storage_time
y ~ current_feature + Sex + (1 | Subject) + storage_time
y ~ current_feature + Sex + (1 | Subject) + storage_time
y ~ current_feature + Sex + (1 | Subject) + storage_time
y ~ current_feature + Sex + (1 | Subject) + storage_time
y ~ current_feature + Sex + (1 | Subject) + storage_time
y ~ current_feature + Sex + (1 | Subject) + storage_time
y ~ current_feature + Sex + (1 | Subject) + storage_time

y ~ current_feature + Sex + (1 | Subject) + storage_time
y ~ current_feature + Sex + (1 | Subject) + storage_time
y ~ current_feature + Sex + (1 | Subject) + storage_time
y ~ current_feature + Sex + (1 | Subject) + storage_time
y ~ current_feature + Sex + (1 | Subject) + storage_time
y ~ current_feature + Sex + (1 | Subject) + storage_time
y ~ current_feature + Sex + (1 | Subject) + storage_time
y ~ current_feature + Sex + (1 | Subject) + storage_time
y ~ current_feature + Sex + (1 | Subject) + storage_time
y ~ current_feature + Sex + (1 | Subject) + storage_time
y ~ current_feature + Sex + (1 | Subject) + storage_time
y ~ current_feature + Sex + (1 | Subject) + storage_time
y ~ current_feature + Sex + (1 | Subject) + storage_time
y ~ current_feature + Sex + (1 | Subject) + storage_time
y ~ current_feature + Sex + (1 | Subject) + storage_time
y ~ current_feature + Sex + (1 | Subject) + storage_time
y ~ current_feature + Sex + (1 | Subject) + storage_time
y ~ current_feature + Sex + (1 | Subject) + storage_time
y ~ current_feature + Sex + (1 | Subject) + storage_time
y ~ current_feature + Sex + (1 | Subject) + storage_time
y ~ current_feature + Sex + (1 | Subject) + storage_time
y ~ current_feature + Sex + (1 | Subject) + storage_time
y ~ current_feature + Sex + (1 | Subject) + storage_time
y ~ current_feature + Sex + (1 | Subject) + storage_time
y ~ current_feature + Sex + (1 | Subject) + storage_time
y ~ current_feature + Sex + (1 | Subject) + storage_time
y ~ current_feature + Sex + (1 | Subject) + storage_time
y ~ current_feature + Sex + (1 | Subject) + storage_time
y ~ current_feature + Sex + (1 | Subject) + storage_time
y ~ current_feature + Sex + (1 | Subject) + storage_time
y ~ current_feature + Sex + (1 | Subject) + storage_time
y ~ current_feature + Sex + (1 | Subject) + storage_time
y ~ current_feature + Sex + (1 | Subject) + storage_time
y ~ current_feature + Sex + (1 | Subject) + storage_time
y ~ current_feature + Sex + (1 | Subject) + storage_time
y ~ current_feature + Sex + (1 | Subject) + storage_time
y ~ current_feature + Sex + (1 | Subject) + storage_time
y ~ current_feature + Sex + (1 | Subject) + storage_time
y ~ current_feature + Sex + (1 | Subject) + storage_time
y ~ current_feature + Sex + (1 | Subject) + storage_time
y ~ current_feature + Sex + (1 | Subject) + storage_time
y ~ current_feature + Sex + (1 | Subject) + storage_time
y ~ current_feature + Sex + (1 | Subject) + storage_time
y ~ current_feature + Sex + (1 | Subject) + storage_time
y ~ current_feature + Sex + (1 | Subject) + storage_time
y ~ current_feature + Sex + (1 | Subject) + storage_time
y ~ current_feature + Sex + (1 | Subject) + storage_time
y ~ current_feature + Sex + (1 | Subject) + storage_time

y ~ current_feature + Sex + (1 | Subject) + storage_time
y ~ current_feature + Sex + (1 | Subject) + storage_time
y ~ current_feature + Sex + (1 | Subject) + storage_time
y ~ current_feature + Sex + (1 | Subject) + storage_time
y ~ current_feature + Sex + (1 | Subject) + storage_time
y ~ current_feature + Sex + (1 | Subject) + storage_time
y ~ current_feature + Sex + (1 | Subject) + storage_time
y ~ current_feature + Sex + (1 | Subject) + storage_time
y ~ current_feature + Sex + (1 | Subject) + storage_time
y ~ current_feature + Sex + (1 | Subject) + storage_time
y ~ current_feature + Sex + (1 | Subject) + storage_time
y ~ current_feature + Sex + (1 | Subject) + storage_time
y ~ current_feature + Sex + (1 | Subject) + storage_time
y ~ current_feature + Sex + (1 | Subject) + storage_time
y ~ current_feature + Sex + (1 | Subject) + storage_time
y ~ current_feature + Sex + (1 | Subject) + storage_time
y ~ current_feature + Sex + (1 | Subject) + storage_time
y ~ current_feature + Sex + (1 | Subject) + storage_time
y ~ current_feature + Sex + (1 | Subject) + storage_time
y ~ current_feature + Sex + (1 | Subject) + storage_time
y ~ current_feature + Sex + (1 | Subject) + storage_time
y ~ current_feature + Sex + (1 | Subject) + storage_time
y ~ current_feature + Sex + (1 | Subject) + storage_time
y ~ current_feature + Sex + (1 | Subject) + storage_time
y ~ current_feature + Sex + (1 | Subject) + storage_time
y ~ current_feature + Sex + (1 | Subject) + storage_time
y ~ current_feature + Sex + (1 | Subject) + storage_time
y ~ current_feature + Sex + (1 | Subject) + storage_time
y ~ current_feature + Sex + (1 | Subject) + storage_time
y ~ current_feature + Sex + (1 | Subject) + storage_time
y ~ current_feature + Sex + (1 | Subject) + storage_time
y ~ current_feature + Sex + (1 | Subject) + storage_time
y ~ current_feature + Sex + (1 | Subject) + storage_time
y ~ current_feature + Sex + (1 | Subject) + storage_time
y ~ current_feature + Sex + (1 | Subject) + storage_time
y ~ current_feature + Sex + (1 | Subject) + storage_time
y ~ current_feature + Sex + (1 | Subject) + storage_time
y ~ current_feature + Sex + (1 | Subject) + storage_time
y ~ current_feature + Sex + (1 | Subject) + storage_time
y ~ current_feature + Sex + (1 | Subject) + storage_time
y ~ current_feature + Sex + (1 | Subject) + storage_time
y ~ current_feature + Sex + (1 | Subject) + storage_time
y ~ current_feature + Sex + (1 | Subject) + storage_time
y ~ current_feature + Sex + (1 | Subject) + storage_time
y ~ current_feature + Sex + (1 | Subject) + storage_time
y ~ current_feature + Sex + (1 | Subject) + storage_time
y ~ current_feature + Sex + (1 | Subject) + storage_time
y ~ current_feature + Sex + (1 | Subject) + storage_time

y ~ current_feature + Sex + (1 | Subject) + storage_time
y ~ current_feature + Sex + (1 | Subject) + storage_time
y ~ current_feature + Sex + (1 | Subject) + storage_time
y ~ current_feature + Sex + (1 | Subject) + storage_time
y ~ current_feature + Sex + (1 | Subject) + storage_time
y ~ current_feature + Sex + (1 | Subject) + storage_time
y ~ current_feature + Sex + (1 | Subject) + storage_time
y ~ current_feature + Sex + (1 | Subject) + storage_time
y ~ current_feature + Sex + (1 | Subject) + storage_time
y ~ current_feature + Sex + (1 | Subject) + storage_time
y ~ current_feature + Sex + (1 | Subject) + storage_time
y ~ current_feature + Sex + (1 | Subject) + storage_time
y ~ current_feature + Sex + (1 | Subject) + storage_time
y ~ current_feature + Sex + (1 | Subject) + storage_time
y ~ current_feature + Sex + (1 | Subject) + storage_time
y ~ current_feature + Sex + (1 | Subject) + storage_time
y ~ current_feature + Sex + (1 | Subject) + storage_time
y ~ current_feature + Sex + (1 | Subject) + storage_time
y ~ current_feature + Sex + (1 | Subject) + storage_time
y ~ current_feature + Sex + (1 | Subject) + storage_time
y ~ current_feature + Sex + (1 | Subject) + storage_time
y ~ current_feature + Sex + (1 | Subject) + storage_time
y ~ current_feature + Sex + (1 | Subject) + storage_time
y ~ current_feature + Sex + (1 | Subject) + storage_time
y ~ current_feature + Sex + (1 | Subject) + storage_time
y ~ current_feature + Sex + (1 | Subject) + storage_time
y ~ current_feature + Sex + (1 | Subject) + storage_time
y ~ current_feature + Sex + (1 | Subject) + storage_time
y ~ current_feature + Sex + (1 | Subject) + storage_time
y ~ current_feature + Sex + (1 | Subject) + storage_time
y ~ current_feature + Sex + (1 | Subject) + storage_time
y ~ current_feature + Sex + (1 | Subject) + storage_time
y ~ current_feature + Sex + (1 | Subject) + storage_time
y ~ current_feature + Sex + (1 | Subject) + storage_time
y ~ current_feature + Sex + (1 | Subject) + storage_time
y ~ current_feature + Sex + (1 | Subject) + storage_time
y ~ current_feature + Sex + (1 | Subject) + storage_time
y ~ current_feature + Sex + (1 | Subject) + storage_time
y ~ current_feature + Sex + (1 | Subject) + storage_time
y ~ current_feature + Sex + (1 | Subject) + storage_time
y ~ current_feature + Sex + (1 | Subject) + storage_time
y ~ current_feature + Sex + (1 | Subject) + storage_time
y ~ current_feature + Sex + (1 | Subject) + storage_time
y ~ current_feature + Sex + (1 | Subject) + storage_time
y ~ current_feature + Sex + (1 | Subject) + storage_time
y ~ current_feature + Sex + (1 | Subject) + storage_time
y ~ current_feature + Sex + (1 | Subject) + storage_time
y ~ current_feature + Sex + (1 | Subject) + storage_time

y ~ current_feature + Sex + (1 | Subject) + storage_time
y ~ current_feature + Sex + (1 | Subject) + storage_time
y ~ current_feature + Sex + (1 | Subject) + storage_time
y ~ current_feature + Sex + (1 | Subject) + storage_time
y ~ current_feature + Sex + (1 | Subject) + storage_time
y ~ current_feature + Sex + (1 | Subject) + storage_time
y ~ current_feature + Sex + (1 | Subject) + storage_time
y ~ current_feature + Sex + (1 | Subject) + storage_time
y ~ current_feature + Sex + (1 | Subject) + storage_time
y ~ current_feature + Sex + (1 | Subject) + storage_time
y ~ current_feature + Sex + (1 | Subject) + storage_time
y ~ current_feature + Sex + (1 | Subject) + storage_time
y ~ current_feature + Sex + (1 | Subject) + storage_time
y ~ current_feature + Sex + (1 | Subject) + storage_time
y ~ current_feature + Sex + (1 | Subject) + storage_time
y ~ current_feature + Sex + (1 | Subject) + storage_time
y ~ current_feature + Sex + (1 | Subject) + storage_time
y ~ current_feature + Sex + (1 | Subject) + storage_time
y ~ current_feature + Sex + (1 | Subject) + storage_time
y ~ current_feature + Sex + (1 | Subject) + storage_time
y ~ current_feature + Sex + (1 | Subject) + storage_time
y ~ current_feature + Sex + (1 | Subject) + storage_time
y ~ current_feature + Sex + (1 | Subject) + storage_time
y ~ current_feature + Sex + (1 | Subject) + storage_time
y ~ current_feature + Sex + (1 | Subject) + storage_time
y ~ current_feature + Sex + (1 | Subject) + storage_time
y ~ current_feature + Sex + (1 | Subject) + storage_time
y ~ current_feature + Sex + (1 | Subject) + storage_time
y ~ current_feature + Sex + (1 | Subject) + storage_time
y ~ current_feature + Sex + (1 | Subject) + storage_time
y ~ current_feature + Sex + (1 | Subject) + storage_time
y ~ current_feature + Sex + (1 | Subject) + storage_time
y ~ current_feature + Sex + (1 | Subject) + storage_time
y ~ current_feature + Sex + (1 | Subject) + storage_time
y ~ current_feature + Sex + (1 | Subject) + storage_time
y ~ current_feature + Sex + (1 | Subject) + storage_time
y ~ current_feature + Sex + (1 | Subject) + storage_time
y ~ current_feature + Sex + (1 | Subject) + storage_time
y ~ current_feature + Sex + (1 | Subject) + storage_time
y ~ current_feature + Sex + (1 | Subject) + storage_time
y ~ current_feature + Sex + (1 | Subject) + storage_time
y ~ current_feature + Sex + (1 | Subject) + storage_time
y ~ current_feature + Sex + (1 | Subject) + storage_time
y ~ current_feature + Sex + (1 | Subject) + storage_time
y ~ current_feature + Sex + (1 | Subject) + storage_time
y ~ current_feature + Sex + (1 | Subject) + storage_time
y ~ current_feature + Sex + (1 | Subject) + storage_time
y ~ current_feature + Sex + (1 | Subject) + storage_time

y ~ current_feature + Sex + (1 | Subject) + storage_time
y ~ current_feature + Sex + (1 | Subject) + storage_time
y ~ current_feature + Sex + (1 | Subject) + storage_time
y ~ current_feature + Sex + (1 | Subject) + storage_time
y ~ current_feature + Sex + (1 | Subject) + storage_time
y ~ current_feature + Sex + (1 | Subject) + storage_time
y ~ current_feature + Sex + (1 | Subject) + storage_time
y ~ current_feature + Sex + (1 | Subject) + storage_time
y ~ current_feature + Sex + (1 | Subject) + storage_time
y ~ current_feature + Sex + (1 | Subject) + storage_time
y ~ current_feature + Sex + (1 | Subject) + storage_time
y ~ current_feature + Sex + (1 | Subject) + storage_time
y ~ current_feature + Sex + (1 | Subject) + storage_time
y ~ current_feature + Sex + (1 | Subject) + storage_time
y ~ current_feature + Sex + (1 | Subject) + storage_time
y ~ current_feature + Sex + (1 | Subject) + storage_time
y ~ current_feature + Sex + (1 | Subject) + storage_time
y ~ current_feature + Sex + (1 | Subject) + storage_time
y ~ current_feature + Sex + (1 | Subject) + storage_time
y ~ current_feature + Sex + (1 | Subject) + storage_time
y ~ current_feature + Sex + (1 | Subject) + storage_time
y ~ current_feature + Sex + (1 | Subject) + storage_time
y ~ current_feature + Sex + (1 | Subject) + storage_time
y ~ current_feature + Sex + (1 | Subject) + storage_time
y ~ current_feature + Sex + (1 | Subject) + storage_time
y ~ current_feature + Sex + (1 | Subject) + storage_time
y ~ current_feature + Sex + (1 | Subject) + storage_time
y ~ current_feature + Sex + (1 | Subject) + storage_time
y ~ current_feature + Sex + (1 | Subject) + storage_time
y ~ current_feature + Sex + (1 | Subject) + storage_time
y ~ current_feature + Sex + (1 | Subject) + storage_time
y ~ current_feature + Sex + (1 | Subject) + storage_time
y ~ current_feature + Sex + (1 | Subject) + storage_time
y ~ current_feature + Sex + (1 | Subject) + storage_time
y ~ current_feature + Sex + (1 | Subject) + storage_time
y ~ current_feature + Sex + (1 | Subject) + storage_time
y ~ current_feature + Sex + (1 | Subject) + storage_time
y ~ current_feature + Sex + (1 | Subject) + storage_time
y ~ current_feature + Sex + (1 | Subject) + storage_time
y ~ current_feature + Sex + (1 | Subject) + storage_time
y ~ current_feature + Sex + (1 | Subject) + storage_time
y ~ current_feature + Sex + (1 | Subject) + storage_time
y ~ current_feature + Sex + (1 | Subject) + storage_time
y ~ current_feature + Sex + (1 | Subject) + storage_time
y ~ current_feature + Sex + (1 | Subject) + storage_time
y ~ current_feature + Sex + (1 | Subject) + storage_time
y ~ current_feature + Sex + (1 | Subject) + storage_time
y ~ current_feature + Sex + (1 | Subject) + storage_time

y ~ current_feature + Sex + (1 | Subject) + storage_time
y ~ current_feature + Sex + (1 | Subject) + storage_time
y ~ current_feature + Sex + (1 | Subject) + storage_time
y ~ current_feature + Sex + (1 | Subject) + storage_time
y ~ current_feature + Sex + (1 | Subject) + storage_time
y ~ current_feature + Sex + (1 | Subject) + storage_time
y ~ current_feature + Sex + (1 | Subject) + storage_time
y ~ current_feature + Sex + (1 | Subject) + storage_time
y ~ current_feature + Sex + (1 | Subject) + storage_time
y ~ current_feature + Sex + (1 | Subject) + storage_time
y ~ current_feature + Sex + (1 | Subject) + storage_time
y ~ current_feature + Sex + (1 | Subject) + storage_time
y ~ current_feature + Sex + (1 | Subject) + storage_time
y ~ current_feature + Sex + (1 | Subject) + storage_time
y ~ current_feature + Sex + (1 | Subject) + storage_time
y ~ current_feature + Sex + (1 | Subject) + storage_time
y ~ current_feature + Sex + (1 | Subject) + storage_time
y ~ current_feature + Sex + (1 | Subject) + storage_time
y ~ current_feature + Sex + (1 | Subject) + storage_time
y ~ current_feature + Sex + (1 | Subject) + storage_time
y ~ current_feature + Sex + (1 | Subject) + storage_time
y ~ current_feature + Sex + (1 | Subject) + storage_time
y ~ current_feature + Sex + (1 | Subject) + storage_time
y ~ current_feature + Sex + (1 | Subject) + storage_time
y ~ current_feature + Sex + (1 | Subject) + storage_time
y ~ current_feature + Sex + (1 | Subject) + storage_time
y ~ current_feature + Sex + (1 | Subject) + storage_time
y ~ current_feature + Sex + (1 | Subject) + storage_time
y ~ current_feature + Sex + (1 | Subject) + storage_time
y ~ current_feature + Sex + (1 | Subject) + storage_time
y ~ current_feature + Sex + (1 | Subject) + storage_time
y ~ current_feature + Sex + (1 | Subject) + storage_time
y ~ current_feature + Sex + (1 | Subject) + storage_time
y ~ current_feature + Sex + (1 | Subject) + storage_time
y ~ current_feature + Sex + (1 | Subject) + storage_time
y ~ current_feature + Sex + (1 | Subject) + storage_time
y ~ current_feature + Sex + (1 | Subject) + storage_time
y ~ current_feature + Sex + (1 | Subject) + storage_time
y ~ current_feature + Sex + (1 | Subject) + storage_time
y ~ current_feature + Sex + (1 | Subject) + storage_time
y ~ current_feature + Sex + (1 | Subject) + storage_time
y ~ current_feature + Sex + (1 | Subject) + storage_time
y ~ current_feature + Sex + (1 | Subject) + storage_time
y ~ current_feature + Sex + (1 | Subject) + storage_time
y ~ current_feature + Sex + (1 | Subject) + storage_time
y ~ current_feature + Sex + (1 | Subject) + storage_time
y ~ current_feature + Sex + (1 | Subject) + storage_time
y ~ current_feature + Sex + (1 | Subject) + storage_time

y ~ current_feature + Sex + (1 | Subject) + storage_time
y ~ current_feature + Sex + (1 | Subject) + storage_time
y ~ current_feature + Sex + (1 | Subject) + storage_time
y ~ current_feature + Sex + (1 | Subject) + storage_time
y ~ current_feature + Sex + (1 | Subject) + storage_time
y ~ current_feature + Sex + (1 | Subject) + storage_time
y ~ current_feature + Sex + (1 | Subject) + storage_time
y ~ current_feature + Sex + (1 | Subject) + storage_time
y ~ current_feature + Sex + (1 | Subject) + storage_time
y ~ current_feature + Sex + (1 | Subject) + storage_time
y ~ current_feature + Sex + (1 | Subject) + storage_time
y ~ current_feature + Sex + (1 | Subject) + storage_time
y ~ current_feature + Sex + (1 | Subject) + storage_time
y ~ current_feature + Sex + (1 | Subject) + storage_time
y ~ current_feature + Sex + (1 | Subject) + storage_time
y ~ current_feature + Sex + (1 | Subject) + storage_time
y ~ current_feature + Sex + (1 | Subject) + storage_time
y ~ current_feature + Sex + (1 | Subject) + storage_time
y ~ current_feature + Sex + (1 | Subject) + storage_time
y ~ current_feature + Sex + (1 | Subject) + storage_time
y ~ current_feature + Sex + (1 | Subject) + storage_time
y ~ current_feature + Sex + (1 | Subject) + storage_time
y ~ current_feature + Sex + (1 | Subject) + storage_time
y ~ current_feature + Sex + (1 | Subject) + storage_time
y ~ current_feature + Sex + (1 | Subject) + storage_time
y ~ current_feature + Sex + (1 | Subject) + storage_time
y ~ current_feature + Sex + (1 | Subject) + storage_time
y ~ current_feature + Sex + (1 | Subject) + storage_time
y ~ current_feature + Sex + (1 | Subject) + storage_time
y ~ current_feature + Sex + (1 | Subject) + storage_time
y ~ current_feature + Sex + (1 | Subject) + storage_time
y ~ current_feature + Sex + (1 | Subject) + storage_time
y ~ current_feature + Sex + (1 | Subject) + storage_time
y ~ current_feature + Sex + (1 | Subject) + storage_time
y ~ current_feature + Sex + (1 | Subject) + storage_time
y ~ current_feature + Sex + (1 | Subject) + storage_time
y ~ current_feature + Sex + (1 | Subject) + storage_time
y ~ current_feature + Sex + (1 | Subject) + storage_time
y ~ current_feature + Sex + (1 | Subject) + storage_time
y ~ current_feature + Sex + (1 | Subject) + storage_time
y ~ current_feature + Sex + (1 | Subject) + storage_time
y ~ current_feature + Sex + (1 | Subject) + storage_time
y ~ current_feature + Sex + (1 | Subject) + storage_time
y ~ current_feature + Sex + (1 | Subject) + storage_time
y ~ current_feature + Sex + (1 | Subject) + storage_time
y ~ current_feature + Sex + (1 | Subject) + storage_time
y ~ current_feature + Sex + (1 | Subject) + storage_time
y ~ current_feature + Sex + (1 | Subject) + storage_time

y ~ current_feature + Sex + (1 | Subject) + storage_time
y ~ current_feature + Sex + (1 | Subject) + storage_time
y ~ current_feature + Sex + (1 | Subject) + storage_time
y ~ current_feature + Sex + (1 | Subject) + storage_time
y ~ current_feature + Sex + (1 | Subject) + storage_time
y ~ current_feature + Sex + (1 | Subject) + storage_time
y ~ current_feature + Sex + (1 | Subject) + storage_time
y ~ current_feature + Sex + (1 | Subject) + storage_time
y ~ current_feature + Sex + (1 | Subject) + storage_time
y ~ current_feature + Sex + (1 | Subject) + storage_time
y ~ current_feature + Sex + (1 | Subject) + storage_time
y ~ current_feature + Sex + (1 | Subject) + storage_time
y ~ current_feature + Sex + (1 | Subject) + storage_time
y ~ current_feature + Sex + (1 | Subject) + storage_time
y ~ current_feature + Sex + (1 | Subject) + storage_time
y ~ current_feature + Sex + (1 | Subject) + storage_time
y ~ current_feature + Sex + (1 | Subject) + storage_time
y ~ current_feature + Sex + (1 | Subject) + storage_time
y ~ current_feature + Sex + (1 | Subject) + storage_time
y ~ current_feature + Sex + (1 | Subject) + storage_time
y ~ current_feature + Sex + (1 | Subject) + storage_time
y ~ current_feature + Sex + (1 | Subject) + storage_time
y ~ current_feature + Sex + (1 | Subject) + storage_time
y ~ current_feature + Sex + (1 | Subject) + storage_time
y ~ current_feature + Sex + (1 | Subject) + storage_time
y ~ current_feature + Sex + (1 | Subject) + storage_time
y ~ current_feature + Sex + (1 | Subject) + storage_time
y ~ current_feature + Sex + (1 | Subject) + storage_time
y ~ current_feature + Sex + (1 | Subject) + storage_time
y ~ current_feature + Sex + (1 | Subject) + storage_time
y ~ current_feature + Sex + (1 | Subject) + storage_time
y ~ current_feature + Sex + (1 | Subject) + storage_time
y ~ current_feature + Sex + (1 | Subject) + storage_time
y ~ current_feature + Sex + (1 | Subject) + storage_time
y ~ current_feature + Sex + (1 | Subject) + storage_time
y ~ current_feature + Sex + (1 | Subject) + storage_time
y ~ current_feature + Sex + (1 | Subject) + storage_time
y ~ current_feature + Sex + (1 | Subject) + storage_time
y ~ current_feature + Sex + (1 | Subject) + storage_time
y ~ current_feature + Sex + (1 | Subject) + storage_time
y ~ current_feature + Sex + (1 | Subject) + storage_time
y ~ current_feature + Sex + (1 | Subject) + storage_time
y ~ current_feature + Sex + (1 | Subject) + storage_time
y ~ current_feature + Sex + (1 | Subject) + storage_time
y ~ current_feature + Sex + (1 | Subject) + storage_time
y ~ current_feature + Sex + (1 | Subject) + storage_time
y ~ current_feature + Sex + (1 | Subject) + storage_time
y ~ current_feature + Sex + (1 | Subject) + storage_time

y ~ current_feature + Sex + (1 | Subject) + storage_time
y ~ current_feature + Sex + (1 | Subject) + storage_time
y ~ current_feature + Sex + (1 | Subject) + storage_time
y ~ current_feature + Sex + (1 | Subject) + storage_time
y ~ current_feature + Sex + (1 | Subject) + storage_time
y ~ current_feature + Sex + (1 | Subject) + storage_time
y ~ current_feature + Sex + (1 | Subject) + storage_time
y ~ current_feature + Sex + (1 | Subject) + storage_time
y ~ current_feature + Sex + (1 | Subject) + storage_time
y ~ current_feature + Sex + (1 | Subject) + storage_time
y ~ current_feature + Sex + (1 | Subject) + storage_time
y ~ current_feature + Sex + (1 | Subject) + storage_time
y ~ current_feature + Sex + (1 | Subject) + storage_time
y ~ current_feature + Sex + (1 | Subject) + storage_time
y ~ current_feature + Sex + (1 | Subject) + storage_time
y ~ current_feature + Sex + (1 | Subject) + storage_time
y ~ current_feature + Sex + (1 | Subject) + storage_time
y ~ current_feature + Sex + (1 | Subject) + storage_time
y ~ current_feature + Sex + (1 | Subject) + storage_time
y ~ current_feature + Sex + (1 | Subject) + storage_time
y ~ current_feature + Sex + (1 | Subject) + storage_time
y ~ current_feature + Sex + (1 | Subject) + storage_time
y ~ current_feature + Sex + (1 | Subject) + storage_time
y ~ current_feature + Sex + (1 | Subject) + storage_time
y ~ current_feature + Sex + (1 | Subject) + storage_time
y ~ current_feature + Sex + (1 | Subject) + storage_time
y ~ current_feature + Sex + (1 | Subject) + storage_time
y ~ current_feature + Sex + (1 | Subject) + storage_time
y ~ current_feature + Sex + (1 | Subject) + storage_time
y ~ current_feature + Sex + (1 | Subject) + storage_time
y ~ current_feature + Sex + (1 | Subject) + storage_time
y ~ current_feature + Sex + (1 | Subject) + storage_time
y ~ current_feature + Sex + (1 | Subject) + storage_time
y ~ current_feature + Sex + (1 | Subject) + storage_time
y ~ current_feature + Sex + (1 | Subject) + storage_time
y ~ current_feature + Sex + (1 | Subject) + storage_time
y ~ current_feature + Sex + (1 | Subject) + storage_time
y ~ current_feature + Sex + (1 | Subject) + storage_time
y ~ current_feature + Sex + (1 | Subject) + storage_time
y ~ current_feature + Sex + (1 | Subject) + storage_time
y ~ current_feature + Sex + (1 | Subject) + storage_time
y ~ current_feature + Sex + (1 | Subject) + storage_time
y ~ current_feature + Sex + (1 | Subject) + storage_time
y ~ current_feature + Sex + (1 | Subject) + storage_time
y ~ current_feature + Sex + (1 | Subject) + storage_time
y ~ current_feature + Sex + (1 | Subject) + storage_time
y ~ current_feature + Sex + (1 | Subject) + storage_time
y ~ current_feature + Sex + (1 | Subject) + storage_time

y ~ current_feature + Sex + (1 | Subject) + storage_time
y ~ current_feature + Sex + (1 | Subject) + storage_time
y ~ current_feature + Sex + (1 | Subject) + storage_time
y ~ current_feature + Sex + (1 | Subject) + storage_time
y ~ current_feature + Sex + (1 | Subject) + storage_time
y ~ current_feature + Sex + (1 | Subject) + storage_time
y ~ current_feature + Sex + (1 | Subject) + storage_time
y ~ current_feature + Sex + (1 | Subject) + storage_time
y ~ current_feature + Sex + (1 | Subject) + storage_time
y ~ current_feature + Sex + (1 | Subject) + storage_time
y ~ current_feature + Sex + (1 | Subject) + storage_time
y ~ current_feature + Sex + (1 | Subject) + storage_time
y ~ current_feature + Sex + (1 | Subject) + storage_time
y ~ current_feature + Sex + (1 | Subject) + storage_time
y ~ current_feature + Sex + (1 | Subject) + storage_time
y ~ current_feature + Sex + (1 | Subject) + storage_time
y ~ current_feature + Sex + (1 | Subject) + storage_time
y ~ current_feature + Sex + (1 | Subject) + storage_time
y ~ current_feature + Sex + (1 | Subject) + storage_time
y ~ current_feature + Sex + (1 | Subject) + storage_time
y ~ current_feature + Sex + (1 | Subject) + storage_time
y ~ current_feature + Sex + (1 | Subject) + storage_time
y ~ current_feature + Sex + (1 | Subject) + storage_time
y ~ current_feature + Sex + (1 | Subject) + storage_time
y ~ current_feature + Sex + (1 | Subject) + storage_time
y ~ current_feature + Sex + (1 | Subject) + storage_time
y ~ current_feature + Sex + (1 | Subject) + storage_time
y ~ current_feature + Sex + (1 | Subject) + storage_time
y ~ current_feature + Sex + (1 | Subject) + storage_time
y ~ current_feature + Sex + (1 | Subject) + storage_time
y ~ current_feature + Sex + (1 | Subject) + storage_time
y ~ current_feature + Sex + (1 | Subject) + storage_time
y ~ current_feature + Sex + (1 | Subject) + storage_time
y ~ current_feature + Sex + (1 | Subject) + storage_time
y ~ current_feature + Sex + (1 | Subject) + storage_time
y ~ current_feature + Sex + (1 | Subject) + storage_time
y ~ current_feature + Sex + (1 | Subject) + storage_time
y ~ current_feature + Sex + (1 | Subject) + storage_time
y ~ current_feature + Sex + (1 | Subject) + storage_time
y ~ current_feature + Sex + (1 | Subject) + storage_time
y ~ current_feature + Sex + (1 | Subject) + storage_time
y ~ current_feature + Sex + (1 | Subject) + storage_time
y ~ current_feature + Sex + (1 | Subject) + storage_time
y ~ current_feature + Sex + (1 | Subject) + storage_time
y ~ current_feature + Sex + (1 | Subject) + storage_time
y ~ current_feature + Sex + (1 | Subject) + storage_time
y ~ current_feature + Sex + (1 | Subject) + storage_time
y ~ current_feature + Sex + (1 | Subject) + storage_time

y ~ current_feature + Sex + (1 | Subject) + storage_time
y ~ current_feature + Sex + (1 | Subject) + storage_time
y ~ current_feature + Sex + (1 | Subject) + storage_time
y ~ current_feature + Sex + (1 | Subject) + storage_time
y ~ current_feature + Sex + (1 | Subject) + storage_time
y ~ current_feature + Sex + (1 | Subject) + storage_time
y ~ current_feature + Sex + (1 | Subject) + storage_time
y ~ current_feature + Sex + (1 | Subject) + storage_time
y ~ current_feature + Sex + (1 | Subject) + storage_time
y ~ current_feature + Sex + (1 | Subject) + storage_time
y ~ current_feature + Sex + (1 | Subject) + storage_time
y ~ current_feature + Sex + (1 | Subject) + storage_time
y ~ current_feature + Sex + (1 | Subject) + storage_time
y ~ current_feature + Sex + (1 | Subject) + storage_time
y ~ current_feature + Sex + (1 | Subject) + storage_time
y ~ current_feature + Sex + (1 | Subject) + storage_time
y ~ current_feature + Sex + (1 | Subject) + storage_time
y ~ current_feature + Sex + (1 | Subject) + storage_time
y ~ current_feature + Sex + (1 | Subject) + storage_time
y ~ current_feature + Sex + (1 | Subject) + storage_time
y ~ current_feature + Sex + (1 | Subject) + storage_time
y ~ current_feature + Sex + (1 | Subject) + storage_time
y ~ current_feature + Sex + (1 | Subject) + storage_time
y ~ current_feature + Sex + (1 | Subject) + storage_time
y ~ current_feature + Sex + (1 | Subject) + storage_time
y ~ current_feature + Sex + (1 | Subject) + storage_time
y ~ current_feature + Sex + (1 | Subject) + storage_time
y ~ current_feature + Sex + (1 | Subject) + storage_time
y ~ current_feature + Sex + (1 | Subject) + storage_time
y ~ current_feature + Sex + (1 | Subject) + storage_time
y ~ current_feature + Sex + (1 | Subject) + storage_time
y ~ current_feature + Sex + (1 | Subject) + storage_time
y ~ current_feature + Sex + (1 | Subject) + storage_time
y ~ current_feature + Sex + (1 | Subject) + storage_time
y ~ current_feature + Sex + (1 | Subject) + storage_time
y ~ current_feature + Sex + (1 | Subject) + storage_time
y ~ current_feature + Sex + (1 | Subject) + storage_time
y ~ current_feature + Sex + (1 | Subject) + storage_time
y ~ current_feature + Sex + (1 | Subject) + storage_time
y ~ current_feature + Sex + (1 | Subject) + storage_time
y ~ current_feature + Sex + (1 | Subject) + storage_time
y ~ current_feature + Sex + (1 | Subject) + storage_time
y ~ current_feature + Sex + (1 | Subject) + storage_time
y ~ current_feature + Sex + (1 | Subject) + storage_time
y ~ current_feature + Sex + (1 | Subject) + storage_time
y ~ current_feature + Sex + (1 | Subject) + storage_time
y ~ current_feature + Sex + (1 | Subject) + storage_time
y ~ current_feature + Sex + (1 | Subject) + storage_time

y ~ current_feature + Sex + (1 | Subject) + storage_time
y ~ current_feature + Sex + (1 | Subject) + storage_time
y ~ current_feature + Sex + (1 | Subject) + storage_time
y ~ current_feature + Sex + (1 | Subject) + storage_time
y ~ current_feature + Sex + (1 | Subject) + storage_time
y ~ current_feature + Sex + (1 | Subject) + storage_time
y ~ current_feature + Sex + (1 | Subject) + storage_time
y ~ current_feature + Sex + (1 | Subject) + storage_time
y ~ current_feature + Sex + (1 | Subject) + storage_time
y ~ current_feature + Sex + (1 | Subject) + storage_time
y ~ current_feature + Sex + (1 | Subject) + storage_time
y ~ current_feature + Sex + (1 | Subject) + storage_time
y ~ current_feature + Sex + (1 | Subject) + storage_time
y ~ current_feature + Sex + (1 | Subject) + storage_time
y ~ current_feature + Sex + (1 | Subject) + storage_time
y ~ current_feature + Sex + (1 | Subject) + storage_time
y ~ current_feature + Sex + (1 | Subject) + storage_time
y ~ current_feature + Sex + (1 | Subject) + storage_time
y ~ current_feature + Sex + (1 | Subject) + storage_time
y ~ current_feature + Sex + (1 | Subject) + storage_time
y ~ current_feature + Sex + (1 | Subject) + storage_time
y ~ current_feature + Sex + (1 | Subject) + storage_time
y ~ current_feature + Sex + (1 | Subject) + storage_time
y ~ current_feature + Sex + (1 | Subject) + storage_time
y ~ current_feature + Sex + (1 | Subject) + storage_time
y ~ current_feature + Sex + (1 | Subject) + storage_time
y ~ current_feature + Sex + (1 | Subject) + storage_time
y ~ current_feature + Sex + (1 | Subject) + storage_time
y ~ current_feature + Sex + (1 | Subject) + storage_time
y ~ current_feature + Sex + (1 | Subject) + storage_time
y ~ current_feature + Sex + (1 | Subject) + storage_time
y ~ current_feature + Sex + (1 | Subject) + storage_time
y ~ current_feature + Sex + (1 | Subject) + storage_time
y ~ current_feature + Sex + (1 | Subject) + storage_time
y ~ current_feature + Sex + (1 | Subject) + storage_time
y ~ current_feature + Sex + (1 | Subject) + storage_time
y ~ current_feature + Sex + (1 | Subject) + storage_time
y ~ current_feature + Sex + (1 | Subject) + storage_time
y ~ current_feature + Sex + (1 | Subject) + storage_time
y ~ current_feature + Sex + (1 | Subject) + storage_time
y ~ current_feature + Sex + (1 | Subject) + storage_time
y ~ current_feature + Sex + (1 | Subject) + storage_time
y ~ current_feature + Sex + (1 | Subject) + storage_time
y ~ current_feature + Sex + (1 | Subject) + storage_time
y ~ current_feature + Sex + (1 | Subject) + storage_time
y ~ current_feature + Sex + (1 | Subject) + storage_time
y ~ current_feature + Sex + (1 | Subject) + storage_time
y ~ current_feature + Sex + (1 | Subject) + storage_time

y ~ current_feature + Sex + (1 | Subject) + storage_time
y ~ current_feature + Sex + (1 | Subject) + storage_time
y ~ current_feature + Sex + (1 | Subject) + storage_time
y ~ current_feature + Sex + (1 | Subject) + storage_time
y ~ current_feature + Sex + (1 | Subject) + storage_time
y ~ current_feature + Sex + (1 | Subject) + storage_time
y ~ current_feature + Sex + (1 | Subject) + storage_time
y ~ current_feature + Sex + (1 | Subject) + storage_time
y ~ current_feature + Sex + (1 | Subject) + storage_time
y ~ current_feature + Sex + (1 | Subject) + storage_time
y ~ current_feature + Sex + (1 | Subject) + storage_time
y ~ current_feature + Sex + (1 | Subject) + storage_time
y ~ current_feature + Sex + (1 | Subject) + storage_time
y ~ current_feature + Sex + (1 | Subject) + storage_time
y ~ current_feature + Sex + (1 | Subject) + storage_time
y ~ current_feature + Sex + (1 | Subject) + storage_time
y ~ current_feature + Sex + (1 | Subject) + storage_time
y ~ current_feature + Sex + (1 | Subject) + storage_time
y ~ current_feature + Sex + (1 | Subject) + storage_time
y ~ current_feature + Sex + (1 | Subject) + storage_time
y ~ current_feature + Sex + (1 | Subject) + storage_time
y ~ current_feature + Sex + (1 | Subject) + storage_time
y ~ current_feature + Sex + (1 | Subject) + storage_time
y ~ current_feature + Sex + (1 | Subject) + storage_time
y ~ current_feature + Sex + (1 | Subject) + storage_time
y ~ current_feature + Sex + (1 | Subject) + storage_time
y ~ current_feature + Sex + (1 | Subject) + storage_time
y ~ current_feature + Sex + (1 | Subject) + storage_time
y ~ current_feature + Sex + (1 | Subject) + storage_time
y ~ current_feature + Sex + (1 | Subject) + storage_time
y ~ current_feature + Sex + (1 | Subject) + storage_time
y ~ current_feature + Sex + (1 | Subject) + storage_time
y ~ current_feature + Sex + (1 | Subject) + storage_time
y ~ current_feature + Sex + (1 | Subject) + storage_time
y ~ current_feature + Sex + (1 | Subject) + storage_time
y ~ current_feature + Sex + (1 | Subject) + storage_time
y ~ current_feature + Sex + (1 | Subject) + storage_time
y ~ current_feature + Sex + (1 | Subject) + storage_time
y ~ current_feature + Sex + (1 | Subject) + storage_time
y ~ current_feature + Sex + (1 | Subject) + storage_time
y ~ current_feature + Sex + (1 | Subject) + storage_time
y ~ current_feature + Sex + (1 | Subject) + storage_time
y ~ current_feature + Sex + (1 | Subject) + storage_time
y ~ current_feature + Sex + (1 | Subject) + storage_time
y ~ current_feature + Sex + (1 | Subject) + storage_time
y ~ current_feature + Sex + (1 | Subject) + storage_time
y ~ current_feature + Sex + (1 | Subject) + storage_time
y ~ current_feature + Sex + (1 | Subject) + storage_time

```
y ~ current_feature + Sex + (1 | Subject) + storage_time
y ~ current_feature + Sex + (1 | Subject) + storage_time
y ~ current_feature + Sex + (1 | Subject) + storage_time
y ~ current_feature + Sex + (1 | Subject) + storage_time
y ~ current_feature + Sex + (1 | Subject) + storage_time
y ~ current_feature + Sex + (1 | Subject) + storage_time
y ~ current_feature + Sex + (1 | Subject) + storage_time
y ~ current_feature + Sex + (1 | Subject) + storage_time
y ~ current_feature + Sex + (1 | Subject) + storage_time
y ~ current_feature + Sex + (1 | Subject) + storage_time
y ~ current_feature + Sex + (1 | Subject) + storage_time
y ~ current_feature + Sex + (1 | Subject) + storage_time
y ~ current_feature + Sex + (1 | Subject) + storage_time
y ~ current_feature + Sex + (1 | Subject) + storage_time
y ~ current_feature + Sex + (1 | Subject) + storage_time
y ~ current_feature + Sex + (1 | Subject) + storage_time
y ~ current_feature + Sex + (1 | Subject) + storage_time
y ~ current_feature + Sex + (1 | Subject) + storage_time
y ~ current_feature + Sex + (1 | Subject) + storage_time
y ~ current_feature + Sex + (1 | Subject) + storage_time
y ~ current_feature + Sex + (1 | Subject) + storage_time
y ~ current_feature + Sex + (1 | Subject) + storage_time
y ~ current_feature + Sex + (1 | Subject) + storage_time
y ~ current_feature + Sex + (1 | Subject) + storage_time
y ~ current_feature + Sex + (1 | Subject) + storage_time
y ~ current_feature + Sex + (1 | Subject) + storage_time
y ~ current_feature + Sex + (1 | Subject) + storage_time
y ~ current_feature + Sex + (1 | Subject) + storage_time
y ~ current_feature + Sex + (1 | Subject) + storage_time
y ~ current_feature + Sex + (1 | Subject) + storage_time
y ~ current_feature + Sex + (1 | Subject) + storage_time
y ~ current_feature + Sex + (1 | Subject) + storage_time
y ~ current_feature + Sex + (1 | Subject) + storage_time
y ~ current_feature + Sex + (1 | Subject) + storage_time
y ~ current_feature + Sex + (1 | Subject) + storage_time
y ~ current_feature + Sex + (1 | Subject) + storage_time
y ~ current_feature + Sex + (1 | Subject) + storage_time
y ~ current_feature + Sex + (1 | Subject) + storage_time
y ~ current_feature + Sex + (1 | Subject) + storage_time
y ~ current_feature + Sex + (1 | Subject) + storage_time
y ~ current_feature + Sex + (1 | Subject) + storage_time
y ~ current_feature + Sex + (1 | Subject) + storage_time
y ~ current_feature + Sex + (1 | Subject) + storage_time
y ~ current_feature + Sex + (1 | Subject) + storage_time
y ~ current_feature + Sex + (1 | Subject) + storage_time
y ~ current_feature + Sex + (1 | Subject) + storage_time
y ~ current_feature + Sex + (1 | Subject) + storage_time
y ~ current_feature + Sex + (1 | Subject) + storage_time
```

y ~ current_feature + Sex + (1 | Subject) + storage_time
y ~ current_feature + Sex + (1 | Subject) + storage_time
y ~ current_feature + Sex + (1 | Subject) + storage_time
y ~ current_feature + Sex + (1 | Subject) + storage_time
y ~ current_feature + Sex + (1 | Subject) + storage_time
y ~ current_feature + Sex + (1 | Subject) + storage_time
y ~ current_feature + Sex + (1 | Subject) + storage_time
y ~ current_feature + Sex + (1 | Subject) + storage_time
y ~ current_feature + Sex + (1 | Subject) + storage_time
y ~ current_feature + Sex + (1 | Subject) + storage_time
y ~ current_feature + Sex + (1 | Subject) + storage_time
y ~ current_feature + Sex + (1 | Subject) + storage_time
y ~ current_feature + Sex + (1 | Subject) + storage_time
y ~ current_feature + Sex + (1 | Subject) + storage_time
y ~ current_feature + Sex + (1 | Subject) + storage_time
y ~ current_feature + Sex + (1 | Subject) + storage_time
y ~ current_feature + Sex + (1 | Subject) + storage_time
y ~ current_feature + Sex + (1 | Subject) + storage_time
y ~ current_feature + Sex + (1 | Subject) + storage_time
y ~ current_feature + Sex + (1 | Subject) + storage_time
y ~ current_feature + Sex + (1 | Subject) + storage_time
y ~ current_feature + Sex + (1 | Subject) + storage_time
y ~ current_feature + Sex + (1 | Subject) + storage_time
y ~ current_feature + Sex + (1 | Subject) + storage_time
y ~ current_feature + Sex + (1 | Subject) + storage_time
y ~ current_feature + Sex + (1 | Subject) + storage_time
y ~ current_feature + Sex + (1 | Subject) + storage_time
y ~ current_feature + Sex + (1 | Subject) + storage_time
y ~ current_feature + Sex + (1 | Subject) + storage_time
y ~ current_feature + Sex + (1 | Subject) + storage_time
y ~ current_feature + Sex + (1 | Subject) + storage_time
y ~ current_feature + Sex + (1 | Subject) + storage_time
y ~ current_feature + Sex + (1 | Subject) + storage_time
y ~ current_feature + Sex + (1 | Subject) + storage_time
y ~ current_feature + Sex + (1 | Subject) + storage_time
y ~ current_feature + Sex + (1 | Subject) + storage_time
y ~ current_feature + Sex + (1 | Subject) + storage_time
y ~ current_feature + Sex + (1 | Subject) + storage_time
y ~ current_feature + Sex + (1 | Subject) + storage_time
y ~ current_feature + Sex + (1 | Subject) + storage_time
y ~ current_feature + Sex + (1 | Subject) + storage_time
y ~ current_feature + Sex + (1 | Subject) + storage_time
y ~ current_feature + Sex + (1 | Subject) + storage_time
y ~ current_feature + Sex + (1 | Subject) + storage_time
y ~ current_feature + Sex + (1 | Subject) + storage_time
y ~ current_feature + Sex + (1 | Subject) + storage_time
y ~ current_feature + Sex + (1 | Subject) + storage_time
y ~ current_feature + Sex + (1 | Subject) + storage_time

```
y ~ current_feature + Sex + (1 | Subject) + storage_time
y ~ current_feature + Sex + (1 | Subject) + storage_time
y ~ current_feature + Sex + (1 | Subject) + storage_time
y ~ current_feature + Sex + (1 | Subject) + storage_time
y ~ current_feature + Sex + (1 | Subject) + storage_time
y ~ current_feature + Sex + (1 | Subject) + storage_time
y ~ current_feature + Sex + (1 | Subject) + storage_time
y ~ current_feature + Sex + (1 | Subject) + storage_time
y ~ current_feature + Sex + (1 | Subject) + storage_time
y ~ current_feature + Sex + (1 | Subject) + storage_time
y ~ current_feature + Sex + (1 | Subject) + storage_time
y ~ current_feature + Sex + (1 | Subject) + storage_time
y ~ current_feature + Sex + (1 | Subject) + storage_time
y ~ current_feature + Sex + (1 | Subject) + storage_time
y ~ current_feature + Sex + (1 | Subject) + storage_time
y ~ current_feature + Sex + (1 | Subject) + storage_time
y ~ current_feature + Sex + (1 | Subject) + storage_time
y ~ current_feature + Sex + (1 | Subject) + storage_time
y ~ current_feature + Sex + (1 | Subject) + storage_time
y ~ current_feature + Sex + (1 | Subject) + storage_time
y ~ current_feature + Sex + (1 | Subject) + storage_time
y ~ current_feature + Sex + (1 | Subject) + storage_time
y ~ current_feature + Sex + (1 | Subject) + storage_time
y ~ current_feature + Sex + (1 | Subject) + storage_time
y ~ current_feature + Sex + (1 | Subject) + storage_time
y ~ current_feature + Sex + (1 | Subject) + storage_time
y ~ current_feature + Sex + (1 | Subject) + storage_time
y ~ current_feature + Sex + (1 | Subject) + storage_time
y ~ current_feature + Sex + (1 | Subject) + storage_time
y ~ current_feature + Sex + (1 | Subject) + storage_time
y ~ current_feature + Sex + (1 | Subject) + storage_time
y ~ current_feature + Sex + (1 | Subject) + storage_time
y ~ current_feature + Sex + (1 | Subject) + storage_time
y ~ current_feature + Sex + (1 | Subject) + storage_time
y ~ current_feature + Sex + (1 | Subject) + storage_time
y ~ current_feature + Sex + (1 | Subject) + storage_time
y ~ current_feature + Sex + (1 | Subject) + storage_time
y ~ current_feature + Sex + (1 | Subject) + storage_time
y ~ current_feature + Sex + (1 | Subject) + storage_time
y ~ current_feature + Sex + (1 | Subject) + storage_time
y ~ current_feature + Sex + (1 | Subject) + storage_time
y ~ current_feature + Sex + (1 | Subject) + storage_time
y ~ current_feature + Sex + (1 | Subject) + storage_time
y ~ current_feature + Sex + (1 | Subject) + storage_time
y ~ current_feature + Sex + (1 | Subject) + storage_time
y ~ current_feature + Sex + (1 | Subject) + storage_time
y ~ current_feature + Sex + (1 | Subject) + storage_time
y ~ current_feature + Sex + (1 | Subject) + storage_time
```

y ~ current_feature + Sex + (1 | Subject) + storage_time
y ~ current_feature + Sex + (1 | Subject) + storage_time
y ~ current_feature + Sex + (1 | Subject) + storage_time
y ~ current_feature + Sex + (1 | Subject) + storage_time
y ~ current_feature + Sex + (1 | Subject) + storage_time
y ~ current_feature + Sex + (1 | Subject) + storage_time
y ~ current_feature + Sex + (1 | Subject) + storage_time
y ~ current_feature + Sex + (1 | Subject) + storage_time
y ~ current_feature + Sex + (1 | Subject) + storage_time
y ~ current_feature + Sex + (1 | Subject) + storage_time
y ~ current_feature + Sex + (1 | Subject) + storage_time
y ~ current_feature + Sex + (1 | Subject) + storage_time
y ~ current_feature + Sex + (1 | Subject) + storage_time
y ~ current_feature + Sex + (1 | Subject) + storage_time
y ~ current_feature + Sex + (1 | Subject) + storage_time
y ~ current_feature + Sex + (1 | Subject) + storage_time
y ~ current_feature + Sex + (1 | Subject) + storage_time
y ~ current_feature + Sex + (1 | Subject) + storage_time
y ~ current_feature + Sex + (1 | Subject) + storage_time
y ~ current_feature + Sex + (1 | Subject) + storage_time
y ~ current_feature + Sex + (1 | Subject) + storage_time
y ~ current_feature + Sex + (1 | Subject) + storage_time
y ~ current_feature + Sex + (1 | Subject) + storage_time
y ~ current_feature + Sex + (1 | Subject) + storage_time
y ~ current_feature + Sex + (1 | Subject) + storage_time
y ~ current_feature + Sex + (1 | Subject) + storage_time
y ~ current_feature + Sex + (1 | Subject) + storage_time
y ~ current_feature + Sex + (1 | Subject) + storage_time
y ~ current_feature + Sex + (1 | Subject) + storage_time
y ~ current_feature + Sex + (1 | Subject) + storage_time
y ~ current_feature + Sex + (1 | Subject) + storage_time
y ~ current_feature + Sex + (1 | Subject) + storage_time
y ~ current_feature + Sex + (1 | Subject) + storage_time
y ~ current_feature + Sex + (1 | Subject) + storage_time
y ~ current_feature + Sex + (1 | Subject) + storage_time
y ~ current_feature + Sex + (1 | Subject) + storage_time
y ~ current_feature + Sex + (1 | Subject) + storage_time
y ~ current_feature + Sex + (1 | Subject) + storage_time
y ~ current_feature + Sex + (1 | Subject) + storage_time
y ~ current_feature + Sex + (1 | Subject) + storage_time
y ~ current_feature + Sex + (1 | Subject) + storage_time
y ~ current_feature + Sex + (1 | Subject) + storage_time
y ~ current_feature + Sex + (1 | Subject) + storage_time
y ~ current_feature + Sex + (1 | Subject) + storage_time
y ~ current_feature + Sex + (1 | Subject) + storage_time
y ~ current_feature + Sex + (1 | Subject) + storage_time
y ~ current_feature + Sex + (1 | Subject) + storage_time
y ~ current_feature + Sex + (1 | Subject) + storage_time

y ~ current_feature + Sex + (1 | Subject) + storage_time
y ~ current_feature + Sex + (1 | Subject) + storage_time
y ~ current_feature + Sex + (1 | Subject) + storage_time
y ~ current_feature + Sex + (1 | Subject) + storage_time
y ~ current_feature + Sex + (1 | Subject) + storage_time
y ~ current_feature + Sex + (1 | Subject) + storage_time
y ~ current_feature + Sex + (1 | Subject) + storage_time
y ~ current_feature + Sex + (1 | Subject) + storage_time
y ~ current_feature + Sex + (1 | Subject) + storage_time
y ~ current_feature + Sex + (1 | Subject) + storage_time
y ~ current_feature + Sex + (1 | Subject) + storage_time
y ~ current_feature + Sex + (1 | Subject) + storage_time
y ~ current_feature + Sex + (1 | Subject) + storage_time
y ~ current_feature + Sex + (1 | Subject) + storage_time
y ~ current_feature + Sex + (1 | Subject) + storage_time
y ~ current_feature + Sex + (1 | Subject) + storage_time
y ~ current_feature + Sex + (1 | Subject) + storage_time
y ~ current_feature + Sex + (1 | Subject) + storage_time
y ~ current_feature + Sex + (1 | Subject) + storage_time
y ~ current_feature + Sex + (1 | Subject) + storage_time
y ~ current_feature + Sex + (1 | Subject) + storage_time
y ~ current_feature + Sex + (1 | Subject) + storage_time
y ~ current_feature + Sex + (1 | Subject) + storage_time
y ~ current_feature + Sex + (1 | Subject) + storage_time
y ~ current_feature + Sex + (1 | Subject) + storage_time
y ~ current_feature + Sex + (1 | Subject) + storage_time
y ~ current_feature + Sex + (1 | Subject) + storage_time
y ~ current_feature + Sex + (1 | Subject) + storage_time
y ~ current_feature + Sex + (1 | Subject) + storage_time
y ~ current_feature + Sex + (1 | Subject) + storage_time
y ~ current_feature + Sex + (1 | Subject) + storage_time
y ~ current_feature + Sex + (1 | Subject) + storage_time
y ~ current_feature + Sex + (1 | Subject) + storage_time
y ~ current_feature + Sex + (1 | Subject) + storage_time
y ~ current_feature + Sex + (1 | Subject) + storage_time
y ~ current_feature + Sex + (1 | Subject) + storage_time
y ~ current_feature + Sex + (1 | Subject) + storage_time
y ~ current_feature + Sex + (1 | Subject) + storage_time
y ~ current_feature + Sex + (1 | Subject) + storage_time
y ~ current_feature + Sex + (1 | Subject) + storage_time
y ~ current_feature + Sex + (1 | Subject) + storage_time
y ~ current_feature + Sex + (1 | Subject) + storage_time
y ~ current_feature + Sex + (1 | Subject) + storage_time
y ~ current_feature + Sex + (1 | Subject) + storage_time
y ~ current_feature + Sex + (1 | Subject) + storage_time
y ~ current_feature + Sex + (1 | Subject) + storage_time
y ~ current_feature + Sex + (1 | Subject) + storage_time
y ~ current_feature + Sex + (1 | Subject) + storage_time

y ~ current_feature + Sex + (1 | Subject) + storage_time
y ~ current_feature + Sex + (1 | Subject) + storage_time
y ~ current_feature + Sex + (1 | Subject) + storage_time
y ~ current_feature + Sex + (1 | Subject) + storage_time
y ~ current_feature + Sex + (1 | Subject) + storage_time
y ~ current_feature + Sex + (1 | Subject) + storage_time
y ~ current_feature + Sex + (1 | Subject) + storage_time
y ~ current_feature + Sex + (1 | Subject) + storage_time
y ~ current_feature + Sex + (1 | Subject) + storage_time
y ~ current_feature + Sex + (1 | Subject) + storage_time
y ~ current_feature + Sex + (1 | Subject) + storage_time
y ~ current_feature + Sex + (1 | Subject) + storage_time
y ~ current_feature + Sex + (1 | Subject) + storage_time
y ~ current_feature + Sex + (1 | Subject) + storage_time
y ~ current_feature + Sex + (1 | Subject) + storage_time
y ~ current_feature + Sex + (1 | Subject) + storage_time
y ~ current_feature + Sex + (1 | Subject) + storage_time
y ~ current_feature + Sex + (1 | Subject) + storage_time
y ~ current_feature + Sex + (1 | Subject) + storage_time
y ~ current_feature + Sex + (1 | Subject) + storage_time
y ~ current_feature + Sex + (1 | Subject) + storage_time
y ~ current_feature + Sex + (1 | Subject) + storage_time
y ~ current_feature + Sex + (1 | Subject) + storage_time
y ~ current_feature + Sex + (1 | Subject) + storage_time
y ~ current_feature + Sex + (1 | Subject) + storage_time
y ~ current_feature + Sex + (1 | Subject) + storage_time
y ~ current_feature + Sex + (1 | Subject) + storage_time
y ~ current_feature + Sex + (1 | Subject) + storage_time
y ~ current_feature + Sex + (1 | Subject) + storage_time
y ~ current_feature + Sex + (1 | Subject) + storage_time
y ~ current_feature + Sex + (1 | Subject) + storage_time
y ~ current_feature + Sex + (1 | Subject) + storage_time
y ~ current_feature + Sex + (1 | Subject) + storage_time
y ~ current_feature + Sex + (1 | Subject) + storage_time
y ~ current_feature + Sex + (1 | Subject) + storage_time
y ~ current_feature + Sex + (1 | Subject) + storage_time
y ~ current_feature + Sex + (1 | Subject) + storage_time
y ~ current_feature + Sex + (1 | Subject) + storage_time
y ~ current_feature + Sex + (1 | Subject) + storage_time
y ~ current_feature + Sex + (1 | Subject) + storage_time
y ~ current_feature + Sex + (1 | Subject) + storage_time
y ~ current_feature + Sex + (1 | Subject) + storage_time
y ~ current_feature + Sex + (1 | Subject) + storage_time
y ~ current_feature + Sex + (1 | Subject) + storage_time
y ~ current_feature + Sex + (1 | Subject) + storage_time
y ~ current_feature + Sex + (1 | Subject) + storage_time
y ~ current_feature + Sex + (1 | Subject) + storage_time
y ~ current_feature + Sex + (1 | Subject) + storage_time

y ~ current_feature + Sex + (1 | Subject) + storage_time
y ~ current_feature + Sex + (1 | Subject) + storage_time
y ~ current_feature + Sex + (1 | Subject) + storage_time
y ~ current_feature + Sex + (1 | Subject) + storage_time
y ~ current_feature + Sex + (1 | Subject) + storage_time
y ~ current_feature + Sex + (1 | Subject) + storage_time
y ~ current_feature + Sex + (1 | Subject) + storage_time
y ~ current_feature + Sex + (1 | Subject) + storage_time
y ~ current_feature + Sex + (1 | Subject) + storage_time
y ~ current_feature + Sex + (1 | Subject) + storage_time
y ~ current_feature + Sex + (1 | Subject) + storage_time
y ~ current_feature + Sex + (1 | Subject) + storage_time
y ~ current_feature + Sex + (1 | Subject) + storage_time
y ~ current_feature + Sex + (1 | Subject) + storage_time
y ~ current_feature + Sex + (1 | Subject) + storage_time
y ~ current_feature + Sex + (1 | Subject) + storage_time
y ~ current_feature + Sex + (1 | Subject) + storage_time
y ~ current_feature + Sex + (1 | Subject) + storage_time
y ~ current_feature + Sex + (1 | Subject) + storage_time
y ~ current_feature + Sex + (1 | Subject) + storage_time
y ~ current_feature + Sex + (1 | Subject) + storage_time
y ~ current_feature + Sex + (1 | Subject) + storage_time
y ~ current_feature + Sex + (1 | Subject) + storage_time
y ~ current_feature + Sex + (1 | Subject) + storage_time
y ~ current_feature + Sex + (1 | Subject) + storage_time
y ~ current_feature + Sex + (1 | Subject) + storage_time
y ~ current_feature + Sex + (1 | Subject) + storage_time
y ~ current_feature + Sex + (1 | Subject) + storage_time
y ~ current_feature + Sex + (1 | Subject) + storage_time
y ~ current_feature + Sex + (1 | Subject) + storage_time
y ~ current_feature + Sex + (1 | Subject) + storage_time
y ~ current_feature + Sex + (1 | Subject) + storage_time
y ~ current_feature + Sex + (1 | Subject) + storage_time
y ~ current_feature + Sex + (1 | Subject) + storage_time
y ~ current_feature + Sex + (1 | Subject) + storage_time
y ~ current_feature + Sex + (1 | Subject) + storage_time
y ~ current_feature + Sex + (1 | Subject) + storage_time
y ~ current_feature + Sex + (1 | Subject) + storage_time
y ~ current_feature + Sex + (1 | Subject) + storage_time
y ~ current_feature + Sex + (1 | Subject) + storage_time
y ~ current_feature + Sex + (1 | Subject) + storage_time
y ~ current_feature + Sex + (1 | Subject) + storage_time
y ~ current_feature + Sex + (1 | Subject) + storage_time
y ~ current_feature + Sex + (1 | Subject) + storage_time
y ~ current_feature + Sex + (1 | Subject) + storage_time
y ~ current_feature + Sex + (1 | Subject) + storage_time
y ~ current_feature + Sex + (1 | Subject) + storage_time
y ~ current_feature + Sex + (1 | Subject) + storage_time

y ~ current_feature + Sex + (1 | Subject) + storage_time
y ~ current_feature + Sex + (1 | Subject) + storage_time
y ~ current_feature + Sex + (1 | Subject) + storage_time
y ~ current_feature + Sex + (1 | Subject) + storage_time
y ~ current_feature + Sex + (1 | Subject) + storage_time
y ~ current_feature + Sex + (1 | Subject) + storage_time
y ~ current_feature + Sex + (1 | Subject) + storage_time
y ~ current_feature + Sex + (1 | Subject) + storage_time
y ~ current_feature + Sex + (1 | Subject) + storage_time
y ~ current_feature + Sex + (1 | Subject) + storage_time
y ~ current_feature + Sex + (1 | Subject) + storage_time
y ~ current_feature + Sex + (1 | Subject) + storage_time
y ~ current_feature + Sex + (1 | Subject) + storage_time
y ~ current_feature + Sex + (1 | Subject) + storage_time
y ~ current_feature + Sex + (1 | Subject) + storage_time
y ~ current_feature + Sex + (1 | Subject) + storage_time
y ~ current_feature + Sex + (1 | Subject) + storage_time
y ~ current_feature + Sex + (1 | Subject) + storage_time
y ~ current_feature + Sex + (1 | Subject) + storage_time
y ~ current_feature + Sex + (1 | Subject) + storage_time
y ~ current_feature + Sex + (1 | Subject) + storage_time
y ~ current_feature + Sex + (1 | Subject) + storage_time
y ~ current_feature + Sex + (1 | Subject) + storage_time
y ~ current_feature + Sex + (1 | Subject) + storage_time
y ~ current_feature + Sex + (1 | Subject) + storage_time
y ~ current_feature + Sex + (1 | Subject) + storage_time
y ~ current_feature + Sex + (1 | Subject) + storage_time
y ~ current_feature + Sex + (1 | Subject) + storage_time
y ~ current_feature + Sex + (1 | Subject) + storage_time
y ~ current_feature + Sex + (1 | Subject) + storage_time
y ~ current_feature + Sex + (1 | Subject) + storage_time
y ~ current_feature + Sex + (1 | Subject) + storage_time
y ~ current_feature + Sex + (1 | Subject) + storage_time
y ~ current_feature + Sex + (1 | Subject) + storage_time
y ~ current_feature + Sex + (1 | Subject) + storage_time
y ~ current_feature + Sex + (1 | Subject) + storage_time
y ~ current_feature + Sex + (1 | Subject) + storage_time
y ~ current_feature + Sex + (1 | Subject) + storage_time
y ~ current_feature + Sex + (1 | Subject) + storage_time
y ~ current_feature + Sex + (1 | Subject) + storage_time
y ~ current_feature + Sex + (1 | Subject) + storage_time
y ~ current_feature + Sex + (1 | Subject) + storage_time
y ~ current_feature + Sex + (1 | Subject) + storage_time
y ~ current_feature + Sex + (1 | Subject) + storage_time
y ~ current_feature + Sex + (1 | Subject) + storage_time
y ~ current_feature + Sex + (1 | Subject) + storage_time
y ~ current_feature + Sex + (1 | Subject) + storage_time
y ~ current_feature + Sex + (1 | Subject) + storage_time

y ~ current_feature + Sex + (1 | Subject) + storage_time
y ~ current_feature + Sex + (1 | Subject) + storage_time
y ~ current_feature + Sex + (1 | Subject) + storage_time
y ~ current_feature + Sex + (1 | Subject) + storage_time
y ~ current_feature + Sex + (1 | Subject) + storage_time
y ~ current_feature + Sex + (1 | Subject) + storage_time
y ~ current_feature + Sex + (1 | Subject) + storage_time
y ~ current_feature + Sex + (1 | Subject) + storage_time
y ~ current_feature + Sex + (1 | Subject) + storage_time
y ~ current_feature + Sex + (1 | Subject) + storage_time
y ~ current_feature + Sex + (1 | Subject) + storage_time
y ~ current_feature + Sex + (1 | Subject) + storage_time
y ~ current_feature + Sex + (1 | Subject) + storage_time
y ~ current_feature + Sex + (1 | Subject) + storage_time
y ~ current_feature + Sex + (1 | Subject) + storage_time
y ~ current_feature + Sex + (1 | Subject) + storage_time
y ~ current_feature + Sex + (1 | Subject) + storage_time
y ~ current_feature + Sex + (1 | Subject) + storage_time
y ~ current_feature + Sex + (1 | Subject) + storage_time
y ~ current_feature + Sex + (1 | Subject) + storage_time
y ~ current_feature + Sex + (1 | Subject) + storage_time
y ~ current_feature + Sex + (1 | Subject) + storage_time
y ~ current_feature + Sex + (1 | Subject) + storage_time
y ~ current_feature + Sex + (1 | Subject) + storage_time
y ~ current_feature + Sex + (1 | Subject) + storage_time
y ~ current_feature + Sex + (1 | Subject) + storage_time
y ~ current_feature + Sex + (1 | Subject) + storage_time
y ~ current_feature + Sex + (1 | Subject) + storage_time
y ~ current_feature + Sex + (1 | Subject) + storage_time
y ~ current_feature + Sex + (1 | Subject) + storage_time
y ~ current_feature + Sex + (1 | Subject) + storage_time
y ~ current_feature + Sex + (1 | Subject) + storage_time
y ~ current_feature + Sex + (1 | Subject) + storage_time
y ~ current_feature + Sex + (1 | Subject) + storage_time
y ~ current_feature + Sex + (1 | Subject) + storage_time
y ~ current_feature + Sex + (1 | Subject) + storage_time
y ~ current_feature + Sex + (1 | Subject) + storage_time
y ~ current_feature + Sex + (1 | Subject) + storage_time
y ~ current_feature + Sex + (1 | Subject) + storage_time
y ~ current_feature + Sex + (1 | Subject) + storage_time
y ~ current_feature + Sex + (1 | Subject) + storage_time
y ~ current_feature + Sex + (1 | Subject) + storage_time
y ~ current_feature + Sex + (1 | Subject) + storage_time
y ~ current_feature + Sex + (1 | Subject) + storage_time
y ~ current_feature + Sex + (1 | Subject) + storage_time
y ~ current_feature + Sex + (1 | Subject) + storage_time
y ~ current_feature + Sex + (1 | Subject) + storage_time
y ~ current_feature + Sex + (1 | Subject) + storage_time

y ~ current_feature + Sex + (1 | Subject) + storage_time
y ~ current_feature + Sex + (1 | Subject) + storage_time
y ~ current_feature + Sex + (1 | Subject) + storage_time
y ~ current_feature + Sex + (1 | Subject) + storage_time
y ~ current_feature + Sex + (1 | Subject) + storage_time
y ~ current_feature + Sex + (1 | Subject) + storage_time
y ~ current_feature + Sex + (1 | Subject) + storage_time
y ~ current_feature + Sex + (1 | Subject) + storage_time
y ~ current_feature + Sex + (1 | Subject) + storage_time
y ~ current_feature + Sex + (1 | Subject) + storage_time
y ~ current_feature + Sex + (1 | Subject) + storage_time
y ~ current_feature + Sex + (1 | Subject) + storage_time
y ~ current_feature + Sex + (1 | Subject) + storage_time
y ~ current_feature + Sex + (1 | Subject) + storage_time
y ~ current_feature + Sex + (1 | Subject) + storage_time
y ~ current_feature + Sex + (1 | Subject) + storage_time
y ~ current_feature + Sex + (1 | Subject) + storage_time
y ~ current_feature + Sex + (1 | Subject) + storage_time
y ~ current_feature + Sex + (1 | Subject) + storage_time
y ~ current_feature + Sex + (1 | Subject) + storage_time
y ~ current_feature + Sex + (1 | Subject) + storage_time
y ~ current_feature + Sex + (1 | Subject) + storage_time
y ~ current_feature + Sex + (1 | Subject) + storage_time
y ~ current_feature + Sex + (1 | Subject) + storage_time
y ~ current_feature + Sex + (1 | Subject) + storage_time
y ~ current_feature + Sex + (1 | Subject) + storage_time
y ~ current_feature + Sex + (1 | Subject) + storage_time
y ~ current_feature + Sex + (1 | Subject) + storage_time
y ~ current_feature + Sex + (1 | Subject) + storage_time
y ~ current_feature + Sex + (1 | Subject) + storage_time
y ~ current_feature + Sex + (1 | Subject) + storage_time
y ~ current_feature + Sex + (1 | Subject) + storage_time
y ~ current_feature + Sex + (1 | Subject) + storage_time
y ~ current_feature + Sex + (1 | Subject) + storage_time
y ~ current_feature + Sex + (1 | Subject) + storage_time
y ~ current_feature + Sex + (1 | Subject) + storage_time
y ~ current_feature + Sex + (1 | Subject) + storage_time
y ~ current_feature + Sex + (1 | Subject) + storage_time
y ~ current_feature + Sex + (1 | Subject) + storage_time
y ~ current_feature + Sex + (1 | Subject) + storage_time
y ~ current_feature + Sex + (1 | Subject) + storage_time
y ~ current_feature + Sex + (1 | Subject) + storage_time
y ~ current_feature + Sex + (1 | Subject) + storage_time
y ~ current_feature + Sex + (1 | Subject) + storage_time
y ~ current_feature + Sex + (1 | Subject) + storage_time
y ~ current_feature + Sex + (1 | Subject) + storage_time
y ~ current_feature + Sex + (1 | Subject) + storage_time
y ~ current_feature + Sex + (1 | Subject) + storage_time

y ~ current_feature + Sex + (1 | Subject) + storage_time
y ~ current_feature + Sex + (1 | Subject) + storage_time
y ~ current_feature + Sex + (1 | Subject) + storage_time
y ~ current_feature + Sex + (1 | Subject) + storage_time
y ~ current_feature + Sex + (1 | Subject) + storage_time
y ~ current_feature + Sex + (1 | Subject) + storage_time
y ~ current_feature + Sex + (1 | Subject) + storage_time
y ~ current_feature + Sex + (1 | Subject) + storage_time
y ~ current_feature + Sex + (1 | Subject) + storage_time
y ~ current_feature + Sex + (1 | Subject) + storage_time
y ~ current_feature + Sex + (1 | Subject) + storage_time
y ~ current_feature + Sex + (1 | Subject) + storage_time
y ~ current_feature + Sex + (1 | Subject) + storage_time
y ~ current_feature + Sex + (1 | Subject) + storage_time
y ~ current_feature + Sex + (1 | Subject) + storage_time
y ~ current_feature + Sex + (1 | Subject) + storage_time
y ~ current_feature + Sex + (1 | Subject) + storage_time
y ~ current_feature + Sex + (1 | Subject) + storage_time
y ~ current_feature + Sex + (1 | Subject) + storage_time
y ~ current_feature + Sex + (1 | Subject) + storage_time
y ~ current_feature + Sex + (1 | Subject) + storage_time
y ~ current_feature + Sex + (1 | Subject) + storage_time
y ~ current_feature + Sex + (1 | Subject) + storage_time
y ~ current_feature + Sex + (1 | Subject) + storage_time
y ~ current_feature + Sex + (1 | Subject) + storage_time
y ~ current_feature + Sex + (1 | Subject) + storage_time
y ~ current_feature + Sex + (1 | Subject) + storage_time
y ~ current_feature + Sex + (1 | Subject) + storage_time
y ~ current_feature + Sex + (1 | Subject) + storage_time
y ~ current_feature + Sex + (1 | Subject) + storage_time
y ~ current_feature + Sex + (1 | Subject) + storage_time
y ~ current_feature + Sex + (1 | Subject) + storage_time
y ~ current_feature + Sex + (1 | Subject) + storage_time
y ~ current_feature + Sex + (1 | Subject) + storage_time
y ~ current_feature + Sex + (1 | Subject) + storage_time
y ~ current_feature + Sex + (1 | Subject) + storage_time
y ~ current_feature + Sex + (1 | Subject) + storage_time
y ~ current_feature + Sex + (1 | Subject) + storage_time
y ~ current_feature + Sex + (1 | Subject) + storage_time
y ~ current_feature + Sex + (1 | Subject) + storage_time
y ~ current_feature + Sex + (1 | Subject) + storage_time
y ~ current_feature + Sex + (1 | Subject) + storage_time
y ~ current_feature + Sex + (1 | Subject) + storage_time
y ~ current_feature + Sex + (1 | Subject) + storage_time
y ~ current_feature + Sex + (1 | Subject) + storage_time
y ~ current_feature + Sex + (1 | Subject) + storage_time
y ~ current_feature + Sex + (1 | Subject) + storage_time
y ~ current_feature + Sex + (1 | Subject) + storage_time

```
y ~ current_feature + Sex + (1 | Subject) + storage_time
y ~ current_feature + Sex + (1 | Subject) + storage_time
y ~ current_feature + Sex + (1 | Subject) + storage_time
y ~ current_feature + Sex + (1 | Subject) + storage_time
y ~ current_feature + Sex + (1 | Subject) + storage_time
y ~ current_feature + Sex + (1 | Subject) + storage_time
y ~ current_feature + Sex + (1 | Subject) + storage_time
y ~ current_feature + Sex + (1 | Subject) + storage_time
y ~ current_feature + Sex + (1 | Subject) + storage_time
y ~ current_feature + Sex + (1 | Subject) + storage_time
y ~ current_feature + Sex + (1 | Subject) + storage_time
y ~ current_feature + Sex + (1 | Subject) + storage_time
y ~ current_feature + Sex + (1 | Subject) + storage_time
y ~ current_feature + Sex + (1 | Subject) + storage_time
y ~ current_feature + Sex + (1 | Subject) + storage_time
y ~ current_feature + Sex + (1 | Subject) + storage_time
y ~ current_feature + Sex + (1 | Subject) + storage_time
y ~ current_feature + Sex + (1 | Subject) + storage_time
y ~ current_feature + Sex + (1 | Subject) + storage_time
y ~ current_feature + Sex + (1 | Subject) + storage_time
y ~ current_feature + Sex + (1 | Subject) + storage_time
y ~ current_feature + Sex + (1 | Subject) + storage_time
y ~ current_feature + Sex + (1 | Subject) + storage_time
y ~ current_feature + Sex + (1 | Subject) + storage_time
y ~ current_feature + Sex + (1 | Subject) + storage_time
y ~ current_feature + Sex + (1 | Subject) + storage_time
y ~ current_feature + Sex + (1 | Subject) + storage_time
y ~ current_feature + Sex + (1 | Subject) + storage_time
y ~ current_feature + Sex + (1 | Subject) + storage_time
y ~ current_feature + Sex + (1 | Subject) + storage_time
y ~ current_feature + Sex + (1 | Subject) + storage_time
y ~ current_feature + Sex + (1 | Subject) + storage_time
y ~ current_feature + Sex + (1 | Subject) + storage_time
y ~ current_feature + Sex + (1 | Subject) + storage_time
y ~ current_feature + Sex + (1 | Subject) + storage_time
y ~ current_feature + Sex + (1 | Subject) + storage_time
y ~ current_feature + Sex + (1 | Subject) + storage_time
y ~ current_feature + Sex + (1 | Subject) + storage_time
y ~ current_feature + Sex + (1 | Subject) + storage_time
y ~ current_feature + Sex + (1 | Subject) + storage_time
y ~ current_feature + Sex + (1 | Subject) + storage_time
y ~ current_feature + Sex + (1 | Subject) + storage_time
y ~ current_feature + Sex + (1 | Subject) + storage_time
y ~ current_feature + Sex + (1 | Subject) + storage_time
y ~ current_feature + Sex + (1 | Subject) + storage_time
y ~ current_feature + Sex + (1 | Subject) + storage_time
y ~ current_feature + Sex + (1 | Subject) + storage_time
y ~ current_feature + Sex + (1 | Subject) + storage_time
```

y ~ current_feature + Sex + (1 | Subject) + storage_time
y ~ current_feature + Sex + (1 | Subject) + storage_time
y ~ current_feature + Sex + (1 | Subject) + storage_time
y ~ current_feature + Sex + (1 | Subject) + storage_time
y ~ current_feature + Sex + (1 | Subject) + storage_time
y ~ current_feature + Sex + (1 | Subject) + storage_time
y ~ current_feature + Sex + (1 | Subject) + storage_time
y ~ current_feature + Sex + (1 | Subject) + storage_time
y ~ current_feature + Sex + (1 | Subject) + storage_time
y ~ current_feature + Sex + (1 | Subject) + storage_time
y ~ current_feature + Sex + (1 | Subject) + storage_time
y ~ current_feature + Sex + (1 | Subject) + storage_time
y ~ current_feature + Sex + (1 | Subject) + storage_time
y ~ current_feature + Sex + (1 | Subject) + storage_time
y ~ current_feature + Sex + (1 | Subject) + storage_time
y ~ current_feature + Sex + (1 | Subject) + storage_time
y ~ current_feature + Sex + (1 | Subject) + storage_time
y ~ current_feature + Sex + (1 | Subject) + storage_time
y ~ current_feature + Sex + (1 | Subject) + storage_time
y ~ current_feature + Sex + (1 | Subject) + storage_time
y ~ current_feature + Sex + (1 | Subject) + storage_time
y ~ current_feature + Sex + (1 | Subject) + storage_time
y ~ current_feature + Sex + (1 | Subject) + storage_time
y ~ current_feature + Sex + (1 | Subject) + storage_time
y ~ current_feature + Sex + (1 | Subject) + storage_time
y ~ current_feature + Sex + (1 | Subject) + storage_time
y ~ current_feature + Sex + (1 | Subject) + storage_time
y ~ current_feature + Sex + (1 | Subject) + storage_time
y ~ current_feature + Sex + (1 | Subject) + storage_time
y ~ current_feature + Sex + (1 | Subject) + storage_time
y ~ current_feature + Sex + (1 | Subject) + storage_time
y ~ current_feature + Sex + (1 | Subject) + storage_time
y ~ current_feature + Sex + (1 | Subject) + storage_time
y ~ current_feature + Sex + (1 | Subject) + storage_time
y ~ current_feature + Sex + (1 | Subject) + storage_time
y ~ current_feature + Sex + (1 | Subject) + storage_time
y ~ current_feature + Sex + (1 | Subject) + storage_time
y ~ current_feature + Sex + (1 | Subject) + storage_time
y ~ current_feature + Sex + (1 | Subject) + storage_time
y ~ current_feature + Sex + (1 | Subject) + storage_time
y ~ current_feature + Sex + (1 | Subject) + storage_time
y ~ current_feature + Sex + (1 | Subject) + storage_time
y ~ current_feature + Sex + (1 | Subject) + storage_time
y ~ current_feature + Sex + (1 | Subject) + storage_time
y ~ current_feature + Sex + (1 | Subject) + storage_time
y ~ current_feature + Sex + (1 | Subject) + storage_time
y ~ current_feature + Sex + (1 | Subject) + storage_time
y ~ current_feature + Sex + (1 | Subject) + storage_time

y ~ current_feature + Sex + (1 | Subject) + storage_time
y ~ current_feature + Sex + (1 | Subject) + storage_time
y ~ current_feature + Sex + (1 | Subject) + storage_time
y ~ current_feature + Sex + (1 | Subject) + storage_time
y ~ current_feature + Sex + (1 | Subject) + storage_time
y ~ current_feature + Sex + (1 | Subject) + storage_time
y ~ current_feature + Sex + (1 | Subject) + storage_time
y ~ current_feature + Sex + (1 | Subject) + storage_time
y ~ current_feature + Sex + (1 | Subject) + storage_time
y ~ current_feature + Sex + (1 | Subject) + storage_time
y ~ current_feature + Sex + (1 | Subject) + storage_time
y ~ current_feature + Sex + (1 | Subject) + storage_time
y ~ current_feature + Sex + (1 | Subject) + storage_time
y ~ current_feature + Sex + (1 | Subject) + storage_time
y ~ current_feature + Sex + (1 | Subject) + storage_time
y ~ current_feature + Sex + (1 | Subject) + storage_time
y ~ current_feature + Sex + (1 | Subject) + storage_time
y ~ current_feature + Sex + (1 | Subject) + storage_time
y ~ current_feature + Sex + (1 | Subject) + storage_time
y ~ current_feature + Sex + (1 | Subject) + storage_time
y ~ current_feature + Sex + (1 | Subject) + storage_time
y ~ current_feature + Sex + (1 | Subject) + storage_time
y ~ current_feature + Sex + (1 | Subject) + storage_time
y ~ current_feature + Sex + (1 | Subject) + storage_time
y ~ current_feature + Sex + (1 | Subject) + storage_time
y ~ current_feature + Sex + (1 | Subject) + storage_time
y ~ current_feature + Sex + (1 | Subject) + storage_time
y ~ current_feature + Sex + (1 | Subject) + storage_time
y ~ current_feature + Sex + (1 | Subject) + storage_time
y ~ current_feature + Sex + (1 | Subject) + storage_time
y ~ current_feature + Sex + (1 | Subject) + storage_time
y ~ current_feature + Sex + (1 | Subject) + storage_time
y ~ current_feature + Sex + (1 | Subject) + storage_time
y ~ current_feature + Sex + (1 | Subject) + storage_time
y ~ current_feature + Sex + (1 | Subject) + storage_time
y ~ current_feature + Sex + (1 | Subject) + storage_time
y ~ current_feature + Sex + (1 | Subject) + storage_time
y ~ current_feature + Sex + (1 | Subject) + storage_time
y ~ current_feature + Sex + (1 | Subject) + storage_time
y ~ current_feature + Sex + (1 | Subject) + storage_time
y ~ current_feature + Sex + (1 | Subject) + storage_time
y ~ current_feature + Sex + (1 | Subject) + storage_time
y ~ current_feature + Sex + (1 | Subject) + storage_time
y ~ current_feature + Sex + (1 | Subject) + storage_time
y ~ current_feature + Sex + (1 | Subject) + storage_time
y ~ current_feature + Sex + (1 | Subject) + storage_time
y ~ current_feature + Sex + (1 | Subject) + storage_time
y ~ current_feature + Sex + (1 | Subject) + storage_time

y ~ current_feature + Sex + (1 | Subject) + storage_time
y ~ current_feature + Sex + (1 | Subject) + storage_time
y ~ current_feature + Sex + (1 | Subject) + storage_time
y ~ current_feature + Sex + (1 | Subject) + storage_time
y ~ current_feature + Sex + (1 | Subject) + storage_time
y ~ current_feature + Sex + (1 | Subject) + storage_time
y ~ current_feature + Sex + (1 | Subject) + storage_time
y ~ current_feature + Sex + (1 | Subject) + storage_time
y ~ current_feature + Sex + (1 | Subject) + storage_time
y ~ current_feature + Sex + (1 | Subject) + storage_time
y ~ current_feature + Sex + (1 | Subject) + storage_time
y ~ current_feature + Sex + (1 | Subject) + storage_time
y ~ current_feature + Sex + (1 | Subject) + storage_time
y ~ current_feature + Sex + (1 | Subject) + storage_time
y ~ current_feature + Sex + (1 | Subject) + storage_time
y ~ current_feature + Sex + (1 | Subject) + storage_time
y ~ current_feature + Sex + (1 | Subject) + storage_time
y ~ current_feature + Sex + (1 | Subject) + storage_time
y ~ current_feature + Sex + (1 | Subject) + storage_time
y ~ current_feature + Sex + (1 | Subject) + storage_time
y ~ current_feature + Sex + (1 | Subject) + storage_time
y ~ current_feature + Sex + (1 | Subject) + storage_time
y ~ current_feature + Sex + (1 | Subject) + storage_time
y ~ current_feature + Sex + (1 | Subject) + storage_time
y ~ current_feature + Sex + (1 | Subject) + storage_time
y ~ current_feature + Sex + (1 | Subject) + storage_time
y ~ current_feature + Sex + (1 | Subject) + storage_time
y ~ current_feature + Sex + (1 | Subject) + storage_time
y ~ current_feature + Sex + (1 | Subject) + storage_time
y ~ current_feature + Sex + (1 | Subject) + storage_time
y ~ current_feature + Sex + (1 | Subject) + storage_time
y ~ current_feature + Sex + (1 | Subject) + storage_time
y ~ current_feature + Sex + (1 | Subject) + storage_time
y ~ current_feature + Sex + (1 | Subject) + storage_time
y ~ current_feature + Sex + (1 | Subject) + storage_time
y ~ current_feature + Sex + (1 | Subject) + storage_time
y ~ current_feature + Sex + (1 | Subject) + storage_time
y ~ current_feature + Sex + (1 | Subject) + storage_time
y ~ current_feature + Sex + (1 | Subject) + storage_time
y ~ current_feature + Sex + (1 | Subject) + storage_time
y ~ current_feature + Sex + (1 | Subject) + storage_time
y ~ current_feature + Sex + (1 | Subject) + storage_time
y ~ current_feature + Sex + (1 | Subject) + storage_time
y ~ current_feature + Sex + (1 | Subject) + storage_time
y ~ current_feature + Sex + (1 | Subject) + storage_time
y ~ current_feature + Sex + (1 | Subject) + storage_time
y ~ current_feature + Sex + (1 | Subject) + storage_time
y ~ current_feature + Sex + (1 | Subject) + storage_time

y ~ current_feature + Sex + (1 | Subject) + storage_time
y ~ current_feature + Sex + (1 | Subject) + storage_time
y ~ current_feature + Sex + (1 | Subject) + storage_time
y ~ current_feature + Sex + (1 | Subject) + storage_time
y ~ current_feature + Sex + (1 | Subject) + storage_time
y ~ current_feature + Sex + (1 | Subject) + storage_time
y ~ current_feature + Sex + (1 | Subject) + storage_time
y ~ current_feature + Sex + (1 | Subject) + storage_time
y ~ current_feature + Sex + (1 | Subject) + storage_time
y ~ current_feature + Sex + (1 | Subject) + storage_time
y ~ current_feature + Sex + (1 | Subject) + storage_time
y ~ current_feature + Sex + (1 | Subject) + storage_time
y ~ current_feature + Sex + (1 | Subject) + storage_time
y ~ current_feature + Sex + (1 | Subject) + storage_time
y ~ current_feature + Sex + (1 | Subject) + storage_time
y ~ current_feature + Sex + (1 | Subject) + storage_time
y ~ current_feature + Sex + (1 | Subject) + storage_time
y ~ current_feature + Sex + (1 | Subject) + storage_time
y ~ current_feature + Sex + (1 | Subject) + storage_time
y ~ current_feature + Sex + (1 | Subject) + storage_time
y ~ current_feature + Sex + (1 | Subject) + storage_time
y ~ current_feature + Sex + (1 | Subject) + storage_time
y ~ current_feature + Sex + (1 | Subject) + storage_time
y ~ current_feature + Sex + (1 | Subject) + storage_time
y ~ current_feature + Sex + (1 | Subject) + storage_time
y ~ current_feature + Sex + (1 | Subject) + storage_time
y ~ current_feature + Sex + (1 | Subject) + storage_time
y ~ current_feature + Sex + (1 | Subject) + storage_time
y ~ current_feature + Sex + (1 | Subject) + storage_time
y ~ current_feature + Sex + (1 | Subject) + storage_time
y ~ current_feature + Sex + (1 | Subject) + storage_time
y ~ current_feature + Sex + (1 | Subject) + storage_time
y ~ current_feature + Sex + (1 | Subject) + storage_time
y ~ current_feature + Sex + (1 | Subject) + storage_time
y ~ current_feature + Sex + (1 | Subject) + storage_time
y ~ current_feature + Sex + (1 | Subject) + storage_time
y ~ current_feature + Sex + (1 | Subject) + storage_time
y ~ current_feature + Sex + (1 | Subject) + storage_time
y ~ current_feature + Sex + (1 | Subject) + storage_time
y ~ current_feature + Sex + (1 | Subject) + storage_time
y ~ current_feature + Sex + (1 | Subject) + storage_time
y ~ current_feature + Sex + (1 | Subject) + storage_time
y ~ current_feature + Sex + (1 | Subject) + storage_time
y ~ current_feature + Sex + (1 | Subject) + storage_time
y ~ current_feature + Sex + (1 | Subject) + storage_time
y ~ current_feature + Sex + (1 | Subject) + storage_time
y ~ current_feature + Sex + (1 | Subject) + storage_time
y ~ current_feature + Sex + (1 | Subject) + storage_time

y ~ current_feature + Sex + (1 | Subject) + storage_time
y ~ current_feature + Sex + (1 | Subject) + storage_time
y ~ current_feature + Sex + (1 | Subject) + storage_time
y ~ current_feature + Sex + (1 | Subject) + storage_time
y ~ current_feature + Sex + (1 | Subject) + storage_time
y ~ current_feature + Sex + (1 | Subject) + storage_time
y ~ current_feature + Sex + (1 | Subject) + storage_time
y ~ current_feature + Sex + (1 | Subject) + storage_time
y ~ current_feature + Sex + (1 | Subject) + storage_time
y ~ current_feature + Sex + (1 | Subject) + storage_time
y ~ current_feature + Sex + (1 | Subject) + storage_time
y ~ current_feature + Sex + (1 | Subject) + storage_time
y ~ current_feature + Sex + (1 | Subject) + storage_time
y ~ current_feature + Sex + (1 | Subject) + storage_time
y ~ current_feature + Sex + (1 | Subject) + storage_time
y ~ current_feature + Sex + (1 | Subject) + storage_time
y ~ current_feature + Sex + (1 | Subject) + storage_time
y ~ current_feature + Sex + (1 | Subject) + storage_time
y ~ current_feature + Sex + (1 | Subject) + storage_time
y ~ current_feature + Sex + (1 | Subject) + storage_time
y ~ current_feature + Sex + (1 | Subject) + storage_time
y ~ current_feature + Sex + (1 | Subject) + storage_time
y ~ current_feature + Sex + (1 | Subject) + storage_time
y ~ current_feature + Sex + (1 | Subject) + storage_time
y ~ current_feature + Sex + (1 | Subject) + storage_time
y ~ current_feature + Sex + (1 | Subject) + storage_time
y ~ current_feature + Sex + (1 | Subject) + storage_time
y ~ current_feature + Sex + (1 | Subject) + storage_time
y ~ current_feature + Sex + (1 | Subject) + storage_time
y ~ current_feature + Sex + (1 | Subject) + storage_time
y ~ current_feature + Sex + (1 | Subject) + storage_time
y ~ current_feature + Sex + (1 | Subject) + storage_time
y ~ current_feature + Sex + (1 | Subject) + storage_time
y ~ current_feature + Sex + (1 | Subject) + storage_time
y ~ current_feature + Sex + (1 | Subject) + storage_time
y ~ current_feature + Sex + (1 | Subject) + storage_time
y ~ current_feature + Sex + (1 | Subject) + storage_time
y ~ current_feature + Sex + (1 | Subject) + storage_time
y ~ current_feature + Sex + (1 | Subject) + storage_time
y ~ current_feature + Sex + (1 | Subject) + storage_time
y ~ current_feature + Sex + (1 | Subject) + storage_time
y ~ current_feature + Sex + (1 | Subject) + storage_time
y ~ current_feature + Sex + (1 | Subject) + storage_time
y ~ current_feature + Sex + (1 | Subject) + storage_time
y ~ current_feature + Sex + (1 | Subject) + storage_time
y ~ current_feature + Sex + (1 | Subject) + storage_time
y ~ current_feature + Sex + (1 | Subject) + storage_time
y ~ current_feature + Sex + (1 | Subject) + storage_time

y ~ current_feature + Sex + (1 | Subject) + storage_time
y ~ current_feature + Sex + (1 | Subject) + storage_time
y ~ current_feature + Sex + (1 | Subject) + storage_time
y ~ current_feature + Sex + (1 | Subject) + storage_time
y ~ current_feature + Sex + (1 | Subject) + storage_time
y ~ current_feature + Sex + (1 | Subject) + storage_time
y ~ current_feature + Sex + (1 | Subject) + storage_time
y ~ current_feature + Sex + (1 | Subject) + storage_time
y ~ current_feature + Sex + (1 | Subject) + storage_time
y ~ current_feature + Sex + (1 | Subject) + storage_time
y ~ current_feature + Sex + (1 | Subject) + storage_time
y ~ current_feature + Sex + (1 | Subject) + storage_time
y ~ current_feature + Sex + (1 | Subject) + storage_time
y ~ current_feature + Sex + (1 | Subject) + storage_time
y ~ current_feature + Sex + (1 | Subject) + storage_time
y ~ current_feature + Sex + (1 | Subject) + storage_time
y ~ current_feature + Sex + (1 | Subject) + storage_time
y ~ current_feature + Sex + (1 | Subject) + storage_time
y ~ current_feature + Sex + (1 | Subject) + storage_time
y ~ current_feature + Sex + (1 | Subject) + storage_time
y ~ current_feature + Sex + (1 | Subject) + storage_time
y ~ current_feature + Sex + (1 | Subject) + storage_time
y ~ current_feature + Sex + (1 | Subject) + storage_time
y ~ current_feature + Sex + (1 | Subject) + storage_time
y ~ current_feature + Sex + (1 | Subject) + storage_time
y ~ current_feature + Sex + (1 | Subject) + storage_time
y ~ current_feature + Sex + (1 | Subject) + storage_time
y ~ current_feature + Sex + (1 | Subject) + storage_time
y ~ current_feature + Sex + (1 | Subject) + storage_time
y ~ current_feature + Sex + (1 | Subject) + storage_time
y ~ current_feature + Sex + (1 | Subject) + storage_time
y ~ current_feature + Sex + (1 | Subject) + storage_time
y ~ current_feature + Sex + (1 | Subject) + storage_time
y ~ current_feature + Sex + (1 | Subject) + storage_time
y ~ current_feature + Sex + (1 | Subject) + storage_time
y ~ current_feature + Sex + (1 | Subject) + storage_time
y ~ current_feature + Sex + (1 | Subject) + storage_time
y ~ current_feature + Sex + (1 | Subject) + storage_time
y ~ current_feature + Sex + (1 | Subject) + storage_time
y ~ current_feature + Sex + (1 | Subject) + storage_time
y ~ current_feature + Sex + (1 | Subject) + storage_time
y ~ current_feature + Sex + (1 | Subject) + storage_time
y ~ current_feature + Sex + (1 | Subject) + storage_time
y ~ current_feature + Sex + (1 | Subject) + storage_time
y ~ current_feature + Sex + (1 | Subject) + storage_time
y ~ current_feature + Sex + (1 | Subject) + storage_time
y ~ current_feature + Sex + (1 | Subject) + storage_time
y ~ current_feature + Sex + (1 | Subject) + storage_time

y ~ current_feature + Sex + (1 | Subject) + storage_time
y ~ current_feature + Sex + (1 | Subject) + storage_time
y ~ current_feature + Sex + (1 | Subject) + storage_time
y ~ current_feature + Sex + (1 | Subject) + storage_time
y ~ current_feature + Sex + (1 | Subject) + storage_time
y ~ current_feature + Sex + (1 | Subject) + storage_time
y ~ current_feature + Sex + (1 | Subject) + storage_time
y ~ current_feature + Sex + (1 | Subject) + storage_time
y ~ current_feature + Sex + (1 | Subject) + storage_time
y ~ current_feature + Sex + (1 | Subject) + storage_time
y ~ current_feature + Sex + (1 | Subject) + storage_time
y ~ current_feature + Sex + (1 | Subject) + storage_time
y ~ current_feature + Sex + (1 | Subject) + storage_time
y ~ current_feature + Sex + (1 | Subject) + storage_time
y ~ current_feature + Sex + (1 | Subject) + storage_time
y ~ current_feature + Sex + (1 | Subject) + storage_time
y ~ current_feature + Sex + (1 | Subject) + storage_time
y ~ current_feature + Sex + (1 | Subject) + storage_time
y ~ current_feature + Sex + (1 | Subject) + storage_time
y ~ current_feature + Sex + (1 | Subject) + storage_time
y ~ current_feature + Sex + (1 | Subject) + storage_time
y ~ current_feature + Sex + (1 | Subject) + storage_time
y ~ current_feature + Sex + (1 | Subject) + storage_time
y ~ current_feature + Sex + (1 | Subject) + storage_time
y ~ current_feature + Sex + (1 | Subject) + storage_time
y ~ current_feature + Sex + (1 | Subject) + storage_time
y ~ current_feature + Sex + (1 | Subject) + storage_time
y ~ current_feature + Sex + (1 | Subject) + storage_time
y ~ current_feature + Sex + (1 | Subject) + storage_time
y ~ current_feature + Sex + (1 | Subject) + storage_time
y ~ current_feature + Sex + (1 | Subject) + storage_time
y ~ current_feature + Sex + (1 | Subject) + storage_time
y ~ current_feature + Sex + (1 | Subject) + storage_time
y ~ current_feature + Sex + (1 | Subject) + storage_time
y ~ current_feature + Sex + (1 | Subject) + storage_time
y ~ current_feature + Sex + (1 | Subject) + storage_time
y ~ current_feature + Sex + (1 | Subject) + storage_time
y ~ current_feature + Sex + (1 | Subject) + storage_time
y ~ current_feature + Sex + (1 | Subject) + storage_time
y ~ current_feature + Sex + (1 | Subject) + storage_time
y ~ current_feature + Sex + (1 | Subject) + storage_time
y ~ current_feature + Sex + (1 | Subject) + storage_time
y ~ current_feature + Sex + (1 | Subject) + storage_time
y ~ current_feature + Sex + (1 | Subject) + storage_time
y ~ current_feature + Sex + (1 | Subject) + storage_time
y ~ current_feature + Sex + (1 | Subject) + storage_time
y ~ current_feature + Sex + (1 | Subject) + storage_time
y ~ current_feature + Sex + (1 | Subject) + storage_time

y ~ current_feature + Sex + (1 | Subject) + storage_time
y ~ current_feature + Sex + (1 | Subject) + storage_time
y ~ current_feature + Sex + (1 | Subject) + storage_time
y ~ current_feature + Sex + (1 | Subject) + storage_time
y ~ current_feature + Sex + (1 | Subject) + storage_time
y ~ current_feature + Sex + (1 | Subject) + storage_time
y ~ current_feature + Sex + (1 | Subject) + storage_time
y ~ current_feature + Sex + (1 | Subject) + storage_time
y ~ current_feature + Sex + (1 | Subject) + storage_time
y ~ current_feature + Sex + (1 | Subject) + storage_time
y ~ current_feature + Sex + (1 | Subject) + storage_time
y ~ current_feature + Sex + (1 | Subject) + storage_time
y ~ current_feature + Sex + (1 | Subject) + storage_time
y ~ current_feature + Sex + (1 | Subject) + storage_time
y ~ current_feature + Sex + (1 | Subject) + storage_time
y ~ current_feature + Sex + (1 | Subject) + storage_time
y ~ current_feature + Sex + (1 | Subject) + storage_time
y ~ current_feature + Sex + (1 | Subject) + storage_time
y ~ current_feature + Sex + (1 | Subject) + storage_time
y ~ current_feature + Sex + (1 | Subject) + storage_time
y ~ current_feature + Sex + (1 | Subject) + storage_time
y ~ current_feature + Sex + (1 | Subject) + storage_time
y ~ current_feature + Sex + (1 | Subject) + storage_time
y ~ current_feature + Sex + (1 | Subject) + storage_time
y ~ current_feature + Sex + (1 | Subject) + storage_time
y ~ current_feature + Sex + (1 | Subject) + storage_time
y ~ current_feature + Sex + (1 | Subject) + storage_time
y ~ current_feature + Sex + (1 | Subject) + storage_time
y ~ current_feature + Sex + (1 | Subject) + storage_time
y ~ current_feature + Sex + (1 | Subject) + storage_time
y ~ current_feature + Sex + (1 | Subject) + storage_time
y ~ current_feature + Sex + (1 | Subject) + storage_time
y ~ current_feature + Sex + (1 | Subject) + storage_time
y ~ current_feature + Sex + (1 | Subject) + storage_time
y ~ current_feature + Sex + (1 | Subject) + storage_time
y ~ current_feature + Sex + (1 | Subject) + storage_time
y ~ current_feature + Sex + (1 | Subject) + storage_time
y ~ current_feature + Sex + (1 | Subject) + storage_time
y ~ current_feature + Sex + (1 | Subject) + storage_time
y ~ current_feature + Sex + (1 | Subject) + storage_time
y ~ current_feature + Sex + (1 | Subject) + storage_time
y ~ current_feature + Sex + (1 | Subject) + storage_time
y ~ current_feature + Sex + (1 | Subject) + storage_time
y ~ current_feature + Sex + (1 | Subject) + storage_time
y ~ current_feature + Sex + (1 | Subject) + storage_time
y ~ current_feature + Sex + (1 | Subject) + storage_time
y ~ current_feature + Sex + (1 | Subject) + storage_time
y ~ current_feature + Sex + (1 | Subject) + storage_time

y ~ current_feature + Sex + (1 | Subject) + storage_time
y ~ current_feature + Sex + (1 | Subject) + storage_time
y ~ current_feature + Sex + (1 | Subject) + storage_time
y ~ current_feature + Sex + (1 | Subject) + storage_time
y ~ current_feature + Sex + (1 | Subject) + storage_time
y ~ current_feature + Sex + (1 | Subject) + storage_time
y ~ current_feature + Sex + (1 | Subject) + storage_time
y ~ current_feature + Sex + (1 | Subject) + storage_time
y ~ current_feature + Sex + (1 | Subject) + storage_time
y ~ current_feature + Sex + (1 | Subject) + storage_time
y ~ current_feature + Sex + (1 | Subject) + storage_time
y ~ current_feature + Sex + (1 | Subject) + storage_time
y ~ current_feature + Sex + (1 | Subject) + storage_time
y ~ current_feature + Sex + (1 | Subject) + storage_time
y ~ current_feature + Sex + (1 | Subject) + storage_time
y ~ current_feature + Sex + (1 | Subject) + storage_time
y ~ current_feature + Sex + (1 | Subject) + storage_time
y ~ current_feature + Sex + (1 | Subject) + storage_time
y ~ current_feature + Sex + (1 | Subject) + storage_time
y ~ current_feature + Sex + (1 | Subject) + storage_time
y ~ current_feature + Sex + (1 | Subject) + storage_time
y ~ current_feature + Sex + (1 | Subject) + storage_time
y ~ current_feature + Sex + (1 | Subject) + storage_time
y ~ current_feature + Sex + (1 | Subject) + storage_time
y ~ current_feature + Sex + (1 | Subject) + storage_time
y ~ current_feature + Sex + (1 | Subject) + storage_time
y ~ current_feature + Sex + (1 | Subject) + storage_time
y ~ current_feature + Sex + (1 | Subject) + storage_time
y ~ current_feature + Sex + (1 | Subject) + storage_time
y ~ current_feature + Sex + (1 | Subject) + storage_time
y ~ current_feature + Sex + (1 | Subject) + storage_time
y ~ current_feature + Sex + (1 | Subject) + storage_time
y ~ current_feature + Sex + (1 | Subject) + storage_time
y ~ current_feature + Sex + (1 | Subject) + storage_time
y ~ current_feature + Sex + (1 | Subject) + storage_time
y ~ current_feature + Sex + (1 | Subject) + storage_time
y ~ current_feature + Sex + (1 | Subject) + storage_time
y ~ current_feature + Sex + (1 | Subject) + storage_time
y ~ current_feature + Sex + (1 | Subject) + storage_time
y ~ current_feature + Sex + (1 | Subject) + storage_time
y ~ current_feature + Sex + (1 | Subject) + storage_time
y ~ current_feature + Sex + (1 | Subject) + storage_time
y ~ current_feature + Sex + (1 | Subject) + storage_time
y ~ current_feature + Sex + (1 | Subject) + storage_time
y ~ current_feature + Sex + (1 | Subject) + storage_time
y ~ current_feature + Sex + (1 | Subject) + storage_time
y ~ current_feature + Sex + (1 | Subject) + storage_time
y ~ current_feature + Sex + (1 | Subject) + storage_time

y ~ current_feature + Sex + (1 | Subject) + storage_time
y ~ current_feature + Sex + (1 | Subject) + storage_time
y ~ current_feature + Sex + (1 | Subject) + storage_time
y ~ current_feature + Sex + (1 | Subject) + storage_time
y ~ current_feature + Sex + (1 | Subject) + storage_time
y ~ current_feature + Sex + (1 | Subject) + storage_time
y ~ current_feature + Sex + (1 | Subject) + storage_time
y ~ current_feature + Sex + (1 | Subject) + storage_time
y ~ current_feature + Sex + (1 | Subject) + storage_time
y ~ current_feature + Sex + (1 | Subject) + storage_time
y ~ current_feature + Sex + (1 | Subject) + storage_time
y ~ current_feature + Sex + (1 | Subject) + storage_time
y ~ current_feature + Sex + (1 | Subject) + storage_time
y ~ current_feature + Sex + (1 | Subject) + storage_time
y ~ current_feature + Sex + (1 | Subject) + storage_time
y ~ current_feature + Sex + (1 | Subject) + storage_time
y ~ current_feature + Sex + (1 | Subject) + storage_time
y ~ current_feature + Sex + (1 | Subject) + storage_time
y ~ current_feature + Sex + (1 | Subject) + storage_time
y ~ current_feature + Sex + (1 | Subject) + storage_time
y ~ current_feature + Sex + (1 | Subject) + storage_time
y ~ current_feature + Sex + (1 | Subject) + storage_time
y ~ current_feature + Sex + (1 | Subject) + storage_time
y ~ current_feature + Sex + (1 | Subject) + storage_time
y ~ current_feature + Sex + (1 | Subject) + storage_time
y ~ current_feature + Sex + (1 | Subject) + storage_time
y ~ current_feature + Sex + (1 | Subject) + storage_time
y ~ current_feature + Sex + (1 | Subject) + storage_time
y ~ current_feature + Sex + (1 | Subject) + storage_time
y ~ current_feature + Sex + (1 | Subject) + storage_time
y ~ current_feature + Sex + (1 | Subject) + storage_time
y ~ current_feature + Sex + (1 | Subject) + storage_time
y ~ current_feature + Sex + (1 | Subject) + storage_time
y ~ current_feature + Sex + (1 | Subject) + storage_time
y ~ current_feature + Sex + (1 | Subject) + storage_time
y ~ current_feature + Sex + (1 | Subject) + storage_time
y ~ current_feature + Sex + (1 | Subject) + storage_time
y ~ current_feature + Sex + (1 | Subject) + storage_time
y ~ current_feature + Sex + (1 | Subject) + storage_time
y ~ current_feature + Sex + (1 | Subject) + storage_time
y ~ current_feature + Sex + (1 | Subject) + storage_time
y ~ current_feature + Sex + (1 | Subject) + storage_time
y ~ current_feature + Sex + (1 | Subject) + storage_time
y ~ current_feature + Sex + (1 | Subject) + storage_time
y ~ current_feature + Sex + (1 | Subject) + storage_time
y ~ current_feature + Sex + (1 | Subject) + storage_time
y ~ current_feature + Sex + (1 | Subject) + storage_time
y ~ current_feature + Sex + (1 | Subject) + storage_time

y ~ current_feature + Sex + (1 | Subject) + storage_time
y ~ current_feature + Sex + (1 | Subject) + storage_time
y ~ current_feature + Sex + (1 | Subject) + storage_time
y ~ current_feature + Sex + (1 | Subject) + storage_time
y ~ current_feature + Sex + (1 | Subject) + storage_time
y ~ current_feature + Sex + (1 | Subject) + storage_time
y ~ current_feature + Sex + (1 | Subject) + storage_time
y ~ current_feature + Sex + (1 | Subject) + storage_time
y ~ current_feature + Sex + (1 | Subject) + storage_time
y ~ current_feature + Sex + (1 | Subject) + storage_time
y ~ current_feature + Sex + (1 | Subject) + storage_time
y ~ current_feature + Sex + (1 | Subject) + storage_time
y ~ current_feature + Sex + (1 | Subject) + storage_time
y ~ current_feature + Sex + (1 | Subject) + storage_time
y ~ current_feature + Sex + (1 | Subject) + storage_time
y ~ current_feature + Sex + (1 | Subject) + storage_time
y ~ current_feature + Sex + (1 | Subject) + storage_time
y ~ current_feature + Sex + (1 | Subject) + storage_time
y ~ current_feature + Sex + (1 | Subject) + storage_time
y ~ current_feature + Sex + (1 | Subject) + storage_time
y ~ current_feature + Sex + (1 | Subject) + storage_time
y ~ current_feature + Sex + (1 | Subject) + storage_time
y ~ current_feature + Sex + (1 | Subject) + storage_time
y ~ current_feature + Sex + (1 | Subject) + storage_time
y ~ current_feature + Sex + (1 | Subject) + storage_time
y ~ current_feature + Sex + (1 | Subject) + storage_time
y ~ current_feature + Sex + (1 | Subject) + storage_time
y ~ current_feature + Sex + (1 | Subject) + storage_time
y ~ current_feature + Sex + (1 | Subject) + storage_time
y ~ current_feature + Sex + (1 | Subject) + storage_time
y ~ current_feature + Sex + (1 | Subject) + storage_time
y ~ current_feature + Sex + (1 | Subject) + storage_time
y ~ current_feature + Sex + (1 | Subject) + storage_time
y ~ current_feature + Sex + (1 | Subject) + storage_time
y ~ current_feature + Sex + (1 | Subject) + storage_time
y ~ current_feature + Sex + (1 | Subject) + storage_time
y ~ current_feature + Sex + (1 | Subject) + storage_time
y ~ current_feature + Sex + (1 | Subject) + storage_time
y ~ current_feature + Sex + (1 | Subject) + storage_time
y ~ current_feature + Sex + (1 | Subject) + storage_time
y ~ current_feature + Sex + (1 | Subject) + storage_time
y ~ current_feature + Sex + (1 | Subject) + storage_time
y ~ current_feature + Sex + (1 | Subject) + storage_time
y ~ current_feature + Sex + (1 | Subject) + storage_time
y ~ current_feature + Sex + (1 | Subject) + storage_time
y ~ current_feature + Sex + (1 | Subject) + storage_time
y ~ current_feature + Sex + (1 | Subject) + storage_time
y ~ current_feature + Sex + (1 | Subject) + storage_time

y ~ current_feature + Sex + (1 | Subject) + storage_time
y ~ current_feature + Sex + (1 | Subject) + storage_time
y ~ current_feature + Sex + (1 | Subject) + storage_time
y ~ current_feature + Sex + (1 | Subject) + storage_time
y ~ current_feature + Sex + (1 | Subject) + storage_time
y ~ current_feature + Sex + (1 | Subject) + storage_time
y ~ current_feature + Sex + (1 | Subject) + storage_time
y ~ current_feature + Sex + (1 | Subject) + storage_time
y ~ current_feature + Sex + (1 | Subject) + storage_time
y ~ current_feature + Sex + (1 | Subject) + storage_time
y ~ current_feature + Sex + (1 | Subject) + storage_time
y ~ current_feature + Sex + (1 | Subject) + storage_time
y ~ current_feature + Sex + (1 | Subject) + storage_time
y ~ current_feature + Sex + (1 | Subject) + storage_time
y ~ current_feature + Sex + (1 | Subject) + storage_time
y ~ current_feature + Sex + (1 | Subject) + storage_time
y ~ current_feature + Sex + (1 | Subject) + storage_time
y ~ current_feature + Sex + (1 | Subject) + storage_time
y ~ current_feature + Sex + (1 | Subject) + storage_time
y ~ current_feature + Sex + (1 | Subject) + storage_time
y ~ current_feature + Sex + (1 | Subject) + storage_time
y ~ current_feature + Sex + (1 | Subject) + storage_time
y ~ current_feature + Sex + (1 | Subject) + storage_time
y ~ current_feature + Sex + (1 | Subject) + storage_time
y ~ current_feature + Sex + (1 | Subject) + storage_time
y ~ current_feature + Sex + (1 | Subject) + storage_time
y ~ current_feature + Sex + (1 | Subject) + storage_time
y ~ current_feature + Sex + (1 | Subject) + storage_time
y ~ current_feature + Sex + (1 | Subject) + storage_time
y ~ current_feature + Sex + (1 | Subject) + storage_time
y ~ current_feature + Sex + (1 | Subject) + storage_time
y ~ current_feature + Sex + (1 | Subject) + storage_time
y ~ current_feature + Sex + (1 | Subject) + storage_time
y ~ current_feature + Sex + (1 | Subject) + storage_time
y ~ current_feature + Sex + (1 | Subject) + storage_time
y ~ current_feature + Sex + (1 | Subject) + storage_time
y ~ current_feature + Sex + (1 | Subject) + storage_time
y ~ current_feature + Sex + (1 | Subject) + storage_time
y ~ current_feature + Sex + (1 | Subject) + storage_time
y ~ current_feature + Sex + (1 | Subject) + storage_time
y ~ current_feature + Sex + (1 | Subject) + storage_time
y ~ current_feature + Sex + (1 | Subject) + storage_time
y ~ current_feature + Sex + (1 | Subject) + storage_time
y ~ current_feature + Sex + (1 | Subject) + storage_time
y ~ current_feature + Sex + (1 | Subject) + storage_time
y ~ current_feature + Sex + (1 | Subject) + storage_time
y ~ current_feature + Sex + (1 | Subject) + storage_time
y ~ current_feature + Sex + (1 | Subject) + storage_time

y ~ current_feature + Sex + (1 | Subject) + storage_time
y ~ current_feature + Sex + (1 | Subject) + storage_time
y ~ current_feature + Sex + (1 | Subject) + storage_time
y ~ current_feature + Sex + (1 | Subject) + storage_time
y ~ current_feature + Sex + (1 | Subject) + storage_time
y ~ current_feature + Sex + (1 | Subject) + storage_time
y ~ current_feature + Sex + (1 | Subject) + storage_time
y ~ current_feature + Sex + (1 | Subject) + storage_time
y ~ current_feature + Sex + (1 | Subject) + storage_time
y ~ current_feature + Sex + (1 | Subject) + storage_time
y ~ current_feature + Sex + (1 | Subject) + storage_time
y ~ current_feature + Sex + (1 | Subject) + storage_time
y ~ current_feature + Sex + (1 | Subject) + storage_time
y ~ current_feature + Sex + (1 | Subject) + storage_time
y ~ current_feature + Sex + (1 | Subject) + storage_time
y ~ current_feature + Sex + (1 | Subject) + storage_time
y ~ current_feature + Sex + (1 | Subject) + storage_time
y ~ current_feature + Sex + (1 | Subject) + storage_time
y ~ current_feature + Sex + (1 | Subject) + storage_time
y ~ current_feature + Sex + (1 | Subject) + storage_time
y ~ current_feature + Sex + (1 | Subject) + storage_time
y ~ current_feature + Sex + (1 | Subject) + storage_time
y ~ current_feature + Sex + (1 | Subject) + storage_time
y ~ current_feature + Sex + (1 | Subject) + storage_time
y ~ current_feature + Sex + (1 | Subject) + storage_time
y ~ current_feature + Sex + (1 | Subject) + storage_time
y ~ current_feature + Sex + (1 | Subject) + storage_time
y ~ current_feature + Sex + (1 | Subject) + storage_time
y ~ current_feature + Sex + (1 | Subject) + storage_time
y ~ current_feature + Sex + (1 | Subject) + storage_time
y ~ current_feature + Sex + (1 | Subject) + storage_time
y ~ current_feature + Sex + (1 | Subject) + storage_time
y ~ current_feature + Sex + (1 | Subject) + storage_time
y ~ current_feature + Sex + (1 | Subject) + storage_time
y ~ current_feature + Sex + (1 | Subject) + storage_time
y ~ current_feature + Sex + (1 | Subject) + storage_time
y ~ current_feature + Sex + (1 | Subject) + storage_time
y ~ current_feature + Sex + (1 | Subject) + storage_time
y ~ current_feature + Sex + (1 | Subject) + storage_time
y ~ current_feature + Sex + (1 | Subject) + storage_time
y ~ current_feature + Sex + (1 | Subject) + storage_time
y ~ current_feature + Sex + (1 | Subject) + storage_time
y ~ current_feature + Sex + (1 | Subject) + storage_time
y ~ current_feature + Sex + (1 | Subject) + storage_time
y ~ current_feature + Sex + (1 | Subject) + storage_time
y ~ current_feature + Sex + (1 | Subject) + storage_time
y ~ current_feature + Sex + (1 | Subject) + storage_time
y ~ current_feature + Sex + (1 | Subject) + storage_time

y ~ current_feature + Sex + (1 | Subject) + storage_time
y ~ current_feature + Sex + (1 | Subject) + storage_time
y ~ current_feature + Sex + (1 | Subject) + storage_time
y ~ current_feature + Sex + (1 | Subject) + storage_time
y ~ current_feature + Sex + (1 | Subject) + storage_time
y ~ current_feature + Sex + (1 | Subject) + storage_time
y ~ current_feature + Sex + (1 | Subject) + storage_time
y ~ current_feature + Sex + (1 | Subject) + storage_time
y ~ current_feature + Sex + (1 | Subject) + storage_time
y ~ current_feature + Sex + (1 | Subject) + storage_time
y ~ current_feature + Sex + (1 | Subject) + storage_time
y ~ current_feature + Sex + (1 | Subject) + storage_time
y ~ current_feature + Sex + (1 | Subject) + storage_time
y ~ current_feature + Sex + (1 | Subject) + storage_time
y ~ current_feature + Sex + (1 | Subject) + storage_time
y ~ current_feature + Sex + (1 | Subject) + storage_time
y ~ current_feature + Sex + (1 | Subject) + storage_time
y ~ current_feature + Sex + (1 | Subject) + storage_time
y ~ current_feature + Sex + (1 | Subject) + storage_time
y ~ current_feature + Sex + (1 | Subject) + storage_time
y ~ current_feature + Sex + (1 | Subject) + storage_time
y ~ current_feature + Sex + (1 | Subject) + storage_time
y ~ current_feature + Sex + (1 | Subject) + storage_time
y ~ current_feature + Sex + (1 | Subject) + storage_time
y ~ current_feature + Sex + (1 | Subject) + storage_time
y ~ current_feature + Sex + (1 | Subject) + storage_time
y ~ current_feature + Sex + (1 | Subject) + storage_time
y ~ current_feature + Sex + (1 | Subject) + storage_time
y ~ current_feature + Sex + (1 | Subject) + storage_time
y ~ current_feature + Sex + (1 | Subject) + storage_time
y ~ current_feature + Sex + (1 | Subject) + storage_time
y ~ current_feature + Sex + (1 | Subject) + storage_time
y ~ current_feature + Sex + (1 | Subject) + storage_time
y ~ current_feature + Sex + (1 | Subject) + storage_time
y ~ current_feature + Sex + (1 | Subject) + storage_time
y ~ current_feature + Sex + (1 | Subject) + storage_time
y ~ current_feature + Sex + (1 | Subject) + storage_time
y ~ current_feature + Sex + (1 | Subject) + storage_time
y ~ current_feature + Sex + (1 | Subject) + storage_time
y ~ current_feature + Sex + (1 | Subject) + storage_time
y ~ current_feature + Sex + (1 | Subject) + storage_time
y ~ current_feature + Sex + (1 | Subject) + storage_time
y ~ current_feature + Sex + (1 | Subject) + storage_time
y ~ current_feature + Sex + (1 | Subject) + storage_time
y ~ current_feature + Sex + (1 | Subject) + storage_time
y ~ current_feature + Sex + (1 | Subject) + storage_time
y ~ current_feature + Sex + (1 | Subject) + storage_time
y ~ current_feature + Sex + (1 | Subject) + storage_time

y ~ current_feature + Sex + (1 | Subject) + storage_time
y ~ current_feature + Sex + (1 | Subject) + storage_time
y ~ current_feature + Sex + (1 | Subject) + storage_time
y ~ current_feature + Sex + (1 | Subject) + storage_time
y ~ current_feature + Sex + (1 | Subject) + storage_time
y ~ current_feature + Sex + (1 | Subject) + storage_time
y ~ current_feature + Sex + (1 | Subject) + storage_time
y ~ current_feature + Sex + (1 | Subject) + storage_time
y ~ current_feature + Sex + (1 | Subject) + storage_time
y ~ current_feature + Sex + (1 | Subject) + storage_time
y ~ current_feature + Sex + (1 | Subject) + storage_time
y ~ current_feature + Sex + (1 | Subject) + storage_time
y ~ current_feature + Sex + (1 | Subject) + storage_time
y ~ current_feature + Sex + (1 | Subject) + storage_time
y ~ current_feature + Sex + (1 | Subject) + storage_time
y ~ current_feature + Sex + (1 | Subject) + storage_time
y ~ current_feature + Sex + (1 | Subject) + storage_time
y ~ current_feature + Sex + (1 | Subject) + storage_time
y ~ current_feature + Sex + (1 | Subject) + storage_time
y ~ current_feature + Sex + (1 | Subject) + storage_time
y ~ current_feature + Sex + (1 | Subject) + storage_time
y ~ current_feature + Sex + (1 | Subject) + storage_time
y ~ current_feature + Sex + (1 | Subject) + storage_time
y ~ current_feature + Sex + (1 | Subject) + storage_time
y ~ current_feature + Sex + (1 | Subject) + storage_time
y ~ current_feature + Sex + (1 | Subject) + storage_time
y ~ current_feature + Sex + (1 | Subject) + storage_time
y ~ current_feature + Sex + (1 | Subject) + storage_time
y ~ current_feature + Sex + (1 | Subject) + storage_time
y ~ current_feature + Sex + (1 | Subject) + storage_time
y ~ current_feature + Sex + (1 | Subject) + storage_time
y ~ current_feature + Sex + (1 | Subject) + storage_time
y ~ current_feature + Sex + (1 | Subject) + storage_time
y ~ current_feature + Sex + (1 | Subject) + storage_time
y ~ current_feature + Sex + (1 | Subject) + storage_time
y ~ current_feature + Sex + (1 | Subject) + storage_time
y ~ current_feature + Sex + (1 | Subject) + storage_time
y ~ current_feature + Sex + (1 | Subject) + storage_time
y ~ current_feature + Sex + (1 | Subject) + storage_time
y ~ current_feature + Sex + (1 | Subject) + storage_time
y ~ current_feature + Sex + (1 | Subject) + storage_time
y ~ current_feature + Sex + (1 | Subject) + storage_time
y ~ current_feature + Sex + (1 | Subject) + storage_time
y ~ current_feature + Sex + (1 | Subject) + storage_time
y ~ current_feature + Sex + (1 | Subject) + storage_time
y ~ current_feature + Sex + (1 | Subject) + storage_time
y ~ current_feature + Sex + (1 | Subject) + storage_time
y ~ current_feature + Sex + (1 | Subject) + storage_time

y ~ current_feature + Sex + (1 | Subject) + storage_time
y ~ current_feature + Sex + (1 | Subject) + storage_time
y ~ current_feature + Sex + (1 | Subject) + storage_time
y ~ current_feature + Sex + (1 | Subject) + storage_time
y ~ current_feature + Sex + (1 | Subject) + storage_time
y ~ current_feature + Sex + (1 | Subject) + storage_time
y ~ current_feature + Sex + (1 | Subject) + storage_time
y ~ current_feature + Sex + (1 | Subject) + storage_time
y ~ current_feature + Sex + (1 | Subject) + storage_time
y ~ current_feature + Sex + (1 | Subject) + storage_time
y ~ current_feature + Sex + (1 | Subject) + storage_time
y ~ current_feature + Sex + (1 | Subject) + storage_time
y ~ current_feature + Sex + (1 | Subject) + storage_time
y ~ current_feature + Sex + (1 | Subject) + storage_time
y ~ current_feature + Sex + (1 | Subject) + storage_time
y ~ current_feature + Sex + (1 | Subject) + storage_time
y ~ current_feature + Sex + (1 | Subject) + storage_time
y ~ current_feature + Sex + (1 | Subject) + storage_time
y ~ current_feature + Sex + (1 | Subject) + storage_time
y ~ current_feature + Sex + (1 | Subject) + storage_time
y ~ current_feature + Sex + (1 | Subject) + storage_time
y ~ current_feature + Sex + (1 | Subject) + storage_time
y ~ current_feature + Sex + (1 | Subject) + storage_time
y ~ current_feature + Sex + (1 | Subject) + storage_time
y ~ current_feature + Sex + (1 | Subject) + storage_time
y ~ current_feature + Sex + (1 | Subject) + storage_time
y ~ current_feature + Sex + (1 | Subject) + storage_time
y ~ current_feature + Sex + (1 | Subject) + storage_time
y ~ current_feature + Sex + (1 | Subject) + storage_time
y ~ current_feature + Sex + (1 | Subject) + storage_time
y ~ current_feature + Sex + (1 | Subject) + storage_time
y ~ current_feature + Sex + (1 | Subject) + storage_time
y ~ current_feature + Sex + (1 | Subject) + storage_time
y ~ current_feature + Sex + (1 | Subject) + storage_time
y ~ current_feature + Sex + (1 | Subject) + storage_time
y ~ current_feature + Sex + (1 | Subject) + storage_time
y ~ current_feature + Sex + (1 | Subject) + storage_time
y ~ current_feature + Sex + (1 | Subject) + storage_time
y ~ current_feature + Sex + (1 | Subject) + storage_time
y ~ current_feature + Sex + (1 | Subject) + storage_time
y ~ current_feature + Sex + (1 | Subject) + storage_time
y ~ current_feature + Sex + (1 | Subject) + storage_time
y ~ current_feature + Sex + (1 | Subject) + storage_time
y ~ current_feature + Sex + (1 | Subject) + storage_time
y ~ current_feature + Sex + (1 | Subject) + storage_time
y ~ current_feature + Sex + (1 | Subject) + storage_time
y ~ current_feature + Sex + (1 | Subject) + storage_time
y ~ current_feature + Sex + (1 | Subject) + storage_time

y ~ current_feature + Sex + (1 | Subject) + storage_time
y ~ current_feature + Sex + (1 | Subject) + storage_time
y ~ current_feature + Sex + (1 | Subject) + storage_time
y ~ current_feature + Sex + (1 | Subject) + storage_time
y ~ current_feature + Sex + (1 | Subject) + storage_time
y ~ current_feature + Sex + (1 | Subject) + storage_time
y ~ current_feature + Sex + (1 | Subject) + storage_time
y ~ current_feature + Sex + (1 | Subject) + storage_time
y ~ current_feature + Sex + (1 | Subject) + storage_time
y ~ current_feature + Sex + (1 | Subject) + storage_time
y ~ current_feature + Sex + (1 | Subject) + storage_time
y ~ current_feature + Sex + (1 | Subject) + storage_time
y ~ current_feature + Sex + (1 | Subject) + storage_time
y ~ current_feature + Sex + (1 | Subject) + storage_time
y ~ current_feature + Sex + (1 | Subject) + storage_time
y ~ current_feature + Sex + (1 | Subject) + storage_time
y ~ current_feature + Sex + (1 | Subject) + storage_time
y ~ current_feature + Sex + (1 | Subject) + storage_time
y ~ current_feature + Sex + (1 | Subject) + storage_time
y ~ current_feature + Sex + (1 | Subject) + storage_time
y ~ current_feature + Sex + (1 | Subject) + storage_time
y ~ current_feature + Sex + (1 | Subject) + storage_time
y ~ current_feature + Sex + (1 | Subject) + storage_time
y ~ current_feature + Sex + (1 | Subject) + storage_time
y ~ current_feature + Sex + (1 | Subject) + storage_time
y ~ current_feature + Sex + (1 | Subject) + storage_time
y ~ current_feature + Sex + (1 | Subject) + storage_time
y ~ current_feature + Sex + (1 | Subject) + storage_time
y ~ current_feature + Sex + (1 | Subject) + storage_time
y ~ current_feature + Sex + (1 | Subject) + storage_time
y ~ current_feature + Sex + (1 | Subject) + storage_time
y ~ current_feature + Sex + (1 | Subject) + storage_time
y ~ current_feature + Sex + (1 | Subject) + storage_time
y ~ current_feature + Sex + (1 | Subject) + storage_time
y ~ current_feature + Sex + (1 | Subject) + storage_time
y ~ current_feature + Sex + (1 | Subject) + storage_time
y ~ current_feature + Sex + (1 | Subject) + storage_time
y ~ current_feature + Sex + (1 | Subject) + storage_time
y ~ current_feature + Sex + (1 | Subject) + storage_time
y ~ current_feature + Sex + (1 | Subject) + storage_time
y ~ current_feature + Sex + (1 | Subject) + storage_time
y ~ current_feature + Sex + (1 | Subject) + storage_time
y ~ current_feature + Sex + (1 | Subject) + storage_time
y ~ current_feature + Sex + (1 | Subject) + storage_time
y ~ current_feature + Sex + (1 | Subject) + storage_time
y ~ current_feature + Sex + (1 | Subject) + storage_time
y ~ current_feature + Sex + (1 | Subject) + storage_time
y ~ current_feature + Sex + (1 | Subject) + storage_time

y ~ current_feature + Sex + (1 | Subject) + storage_time
y ~ current_feature + Sex + (1 | Subject) + storage_time
y ~ current_feature + Sex + (1 | Subject) + storage_time
y ~ current_feature + Sex + (1 | Subject) + storage_time
y ~ current_feature + Sex + (1 | Subject) + storage_time
y ~ current_feature + Sex + (1 | Subject) + storage_time
y ~ current_feature + Sex + (1 | Subject) + storage_time
y ~ current_feature + Sex + (1 | Subject) + storage_time
y ~ current_feature + Sex + (1 | Subject) + storage_time
y ~ current_feature + Sex + (1 | Subject) + storage_time
y ~ current_feature + Sex + (1 | Subject) + storage_time
y ~ current_feature + Sex + (1 | Subject) + storage_time
y ~ current_feature + Sex + (1 | Subject) + storage_time
y ~ current_feature + Sex + (1 | Subject) + storage_time
y ~ current_feature + Sex + (1 | Subject) + storage_time
y ~ current_feature + Sex + (1 | Subject) + storage_time
y ~ current_feature + Sex + (1 | Subject) + storage_time
y ~ current_feature + Sex + (1 | Subject) + storage_time
y ~ current_feature + Sex + (1 | Subject) + storage_time
y ~ current_feature + Sex + (1 | Subject) + storage_time
y ~ current_feature + Sex + (1 | Subject) + storage_time
y ~ current_feature + Sex + (1 | Subject) + storage_time
y ~ current_feature + Sex + (1 | Subject) + storage_time
y ~ current_feature + Sex + (1 | Subject) + storage_time
y ~ current_feature + Sex + (1 | Subject) + storage_time
y ~ current_feature + Sex + (1 | Subject) + storage_time
y ~ current_feature + Sex + (1 | Subject) + storage_time
y ~ current_feature + Sex + (1 | Subject) + storage_time
y ~ current_feature + Sex + (1 | Subject) + storage_time
y ~ current_feature + Sex + (1 | Subject) + storage_time
y ~ current_feature + Sex + (1 | Subject) + storage_time
y ~ current_feature + Sex + (1 | Subject) + storage_time
y ~ current_feature + Sex + (1 | Subject) + storage_time
y ~ current_feature + Sex + (1 | Subject) + storage_time
y ~ current_feature + Sex + (1 | Subject) + storage_time
y ~ current_feature + Sex + (1 | Subject) + storage_time
y ~ current_feature + Sex + (1 | Subject) + storage_time
y ~ current_feature + Sex + (1 | Subject) + storage_time
y ~ current_feature + Sex + (1 | Subject) + storage_time
y ~ current_feature + Sex + (1 | Subject) + storage_time
y ~ current_feature + Sex + (1 | Subject) + storage_time
y ~ current_feature + Sex + (1 | Subject) + storage_time
y ~ current_feature + Sex + (1 | Subject) + storage_time
y ~ current_feature + Sex + (1 | Subject) + storage_time
y ~ current_feature + Sex + (1 | Subject) + storage_time
y ~ current_feature + Sex + (1 | Subject) + storage_time
y ~ current_feature + Sex + (1 | Subject) + storage_time
y ~ current_feature + Sex + (1 | Subject) + storage_time

y ~ current_feature + Sex + (1 | Subject) + storage_time
y ~ current_feature + Sex + (1 | Subject) + storage_time
y ~ current_feature + Sex + (1 | Subject) + storage_time
y ~ current_feature + Sex + (1 | Subject) + storage_time
y ~ current_feature + Sex + (1 | Subject) + storage_time
y ~ current_feature + Sex + (1 | Subject) + storage_time
y ~ current_feature + Sex + (1 | Subject) + storage_time
y ~ current_feature + Sex + (1 | Subject) + storage_time
y ~ current_feature + Sex + (1 | Subject) + storage_time
y ~ current_feature + Sex + (1 | Subject) + storage_time
y ~ current_feature + Sex + (1 | Subject) + storage_time
y ~ current_feature + Sex + (1 | Subject) + storage_time
y ~ current_feature + Sex + (1 | Subject) + storage_time
y ~ current_feature + Sex + (1 | Subject) + storage_time
y ~ current_feature + Sex + (1 | Subject) + storage_time
y ~ current_feature + Sex + (1 | Subject) + storage_time
y ~ current_feature + Sex + (1 | Subject) + storage_time
y ~ current_feature + Sex + (1 | Subject) + storage_time
y ~ current_feature + Sex + (1 | Subject) + storage_time
y ~ current_feature + Sex + (1 | Subject) + storage_time
y ~ current_feature + Sex + (1 | Subject) + storage_time
y ~ current_feature + Sex + (1 | Subject) + storage_time
y ~ current_feature + Sex + (1 | Subject) + storage_time
y ~ current_feature + Sex + (1 | Subject) + storage_time
y ~ current_feature + Sex + (1 | Subject) + storage_time
y ~ current_feature + Sex + (1 | Subject) + storage_time
y ~ current_feature + Sex + (1 | Subject) + storage_time
y ~ current_feature + Sex + (1 | Subject) + storage_time
y ~ current_feature + Sex + (1 | Subject) + storage_time
y ~ current_feature + Sex + (1 | Subject) + storage_time
y ~ current_feature + Sex + (1 | Subject) + storage_time
y ~ current_feature + Sex + (1 | Subject) + storage_time
y ~ current_feature + Sex + (1 | Subject) + storage_time
y ~ current_feature + Sex + (1 | Subject) + storage_time
y ~ current_feature + Sex + (1 | Subject) + storage_time
y ~ current_feature + Sex + (1 | Subject) + storage_time
y ~ current_feature + Sex + (1 | Subject) + storage_time
y ~ current_feature + Sex + (1 | Subject) + storage_time
y ~ current_feature + Sex + (1 | Subject) + storage_time
y ~ current_feature + Sex + (1 | Subject) + storage_time
y ~ current_feature + Sex + (1 | Subject) + storage_time
y ~ current_feature + Sex + (1 | Subject) + storage_time
y ~ current_feature + Sex + (1 | Subject) + storage_time
y ~ current_feature + Sex + (1 | Subject) + storage_time
y ~ current_feature + Sex + (1 | Subject) + storage_time
y ~ current_feature + Sex + (1 | Subject) + storage_time
y ~ current_feature + Sex + (1 | Subject) + storage_time
y ~ current_feature + Sex + (1 | Subject) + storage_time

y ~ current_feature + Sex + (1 | Subject) + storage_time
y ~ current_feature + Sex + (1 | Subject) + storage_time
y ~ current_feature + Sex + (1 | Subject) + storage_time
y ~ current_feature + Sex + (1 | Subject) + storage_time
y ~ current_feature + Sex + (1 | Subject) + storage_time
y ~ current_feature + Sex + (1 | Subject) + storage_time
y ~ current_feature + Sex + (1 | Subject) + storage_time
y ~ current_feature + Sex + (1 | Subject) + storage_time
y ~ current_feature + Sex + (1 | Subject) + storage_time
y ~ current_feature + Sex + (1 | Subject) + storage_time
y ~ current_feature + Sex + (1 | Subject) + storage_time
y ~ current_feature + Sex + (1 | Subject) + storage_time
y ~ current_feature + Sex + (1 | Subject) + storage_time
y ~ current_feature + Sex + (1 | Subject) + storage_time
y ~ current_feature + Sex + (1 | Subject) + storage_time
y ~ current_feature + Sex + (1 | Subject) + storage_time
y ~ current_feature + Sex + (1 | Subject) + storage_time
y ~ current_feature + Sex + (1 | Subject) + storage_time
y ~ current_feature + Sex + (1 | Subject) + storage_time
y ~ current_feature + Sex + (1 | Subject) + storage_time
y ~ current_feature + Sex + (1 | Subject) + storage_time
y ~ current_feature + Sex + (1 | Subject) + storage_time
y ~ current_feature + Sex + (1 | Subject) + storage_time
y ~ current_feature + Sex + (1 | Subject) + storage_time
y ~ current_feature + Sex + (1 | Subject) + storage_time
y ~ current_feature + Sex + (1 | Subject) + storage_time
y ~ current_feature + Sex + (1 | Subject) + storage_time
y ~ current_feature + Sex + (1 | Subject) + storage_time
y ~ current_feature + Sex + (1 | Subject) + storage_time
y ~ current_feature + Sex + (1 | Subject) + storage_time
y ~ current_feature + Sex + (1 | Subject) + storage_time
y ~ current_feature + Sex + (1 | Subject) + storage_time
y ~ current_feature + Sex + (1 | Subject) + storage_time
y ~ current_feature + Sex + (1 | Subject) + storage_time
y ~ current_feature + Sex + (1 | Subject) + storage_time
y ~ current_feature + Sex + (1 | Subject) + storage_time
y ~ current_feature + Sex + (1 | Subject) + storage_time
y ~ current_feature + Sex + (1 | Subject) + storage_time
y ~ current_feature + Sex + (1 | Subject) + storage_time
y ~ current_feature + Sex + (1 | Subject) + storage_time
y ~ current_feature + Sex + (1 | Subject) + storage_time
y ~ current_feature + Sex + (1 | Subject) + storage_time
y ~ current_feature + Sex + (1 | Subject) + storage_time
y ~ current_feature + Sex + (1 | Subject) + storage_time
y ~ current_feature + Sex + (1 | Subject) + storage_time
y ~ current_feature + Sex + (1 | Subject) + storage_time
y ~ current_feature + Sex + (1 | Subject) + storage_time
y ~ current_feature + Sex + (1 | Subject) + storage_time

y ~ current_feature + Sex + (1 | Subject) + storage_time
y ~ current_feature + Sex + (1 | Subject) + storage_time
y ~ current_feature + Sex + (1 | Subject) + storage_time
y ~ current_feature + Sex + (1 | Subject) + storage_time
y ~ current_feature + Sex + (1 | Subject) + storage_time
y ~ current_feature + Sex + (1 | Subject) + storage_time
y ~ current_feature + Sex + (1 | Subject) + storage_time
y ~ current_feature + Sex + (1 | Subject) + storage_time
y ~ current_feature + Sex + (1 | Subject) + storage_time
y ~ current_feature + Sex + (1 | Subject) + storage_time
y ~ current_feature + Sex + (1 | Subject) + storage_time
y ~ current_feature + Sex + (1 | Subject) + storage_time
y ~ current_feature + Sex + (1 | Subject) + storage_time
y ~ current_feature + Sex + (1 | Subject) + storage_time
y ~ current_feature + Sex + (1 | Subject) + storage_time
y ~ current_feature + Sex + (1 | Subject) + storage_time
y ~ current_feature + Sex + (1 | Subject) + storage_time
y ~ current_feature + Sex + (1 | Subject) + storage_time
y ~ current_feature + Sex + (1 | Subject) + storage_time
y ~ current_feature + Sex + (1 | Subject) + storage_time
y ~ current_feature + Sex + (1 | Subject) + storage_time
y ~ current_feature + Sex + (1 | Subject) + storage_time
y ~ current_feature + Sex + (1 | Subject) + storage_time
y ~ current_feature + Sex + (1 | Subject) + storage_time
y ~ current_feature + Sex + (1 | Subject) + storage_time
y ~ current_feature + Sex + (1 | Subject) + storage_time
y ~ current_feature + Sex + (1 | Subject) + storage_time
y ~ current_feature + Sex + (1 | Subject) + storage_time
y ~ current_feature + Sex + (1 | Subject) + storage_time
y ~ current_feature + Sex + (1 | Subject) + storage_time
y ~ current_feature + Sex + (1 | Subject) + storage_time
y ~ current_feature + Sex + (1 | Subject) + storage_time
y ~ current_feature + Sex + (1 | Subject) + storage_time
y ~ current_feature + Sex + (1 | Subject) + storage_time
y ~ current_feature + Sex + (1 | Subject) + storage_time
y ~ current_feature + Sex + (1 | Subject) + storage_time
y ~ current_feature + Sex + (1 | Subject) + storage_time
y ~ current_feature + Sex + (1 | Subject) + storage_time
y ~ current_feature + Sex + (1 | Subject) + storage_time
y ~ current_feature + Sex + (1 | Subject) + storage_time
y ~ current_feature + Sex + (1 | Subject) + storage_time
y ~ current_feature + Sex + (1 | Subject) + storage_time
y ~ current_feature + Sex + (1 | Subject) + storage_time
y ~ current_feature + Sex + (1 | Subject) + storage_time
y ~ current_feature + Sex + (1 | Subject) + storage_time
y ~ current_feature + Sex + (1 | Subject) + storage_time
y ~ current_feature + Sex + (1 | Subject) + storage_time
y ~ current_feature + Sex + (1 | Subject) + storage_time

y ~ current_feature + Sex + (1 | Subject) + storage_time
y ~ current_feature + Sex + (1 | Subject) + storage_time
y ~ current_feature + Sex + (1 | Subject) + storage_time
y ~ current_feature + Sex + (1 | Subject) + storage_time
y ~ current_feature + Sex + (1 | Subject) + storage_time
y ~ current_feature + Sex + (1 | Subject) + storage_time
y ~ current_feature + Sex + (1 | Subject) + storage_time
y ~ current_feature + Sex + (1 | Subject) + storage_time
y ~ current_feature + Sex + (1 | Subject) + storage_time
y ~ current_feature + Sex + (1 | Subject) + storage_time
y ~ current_feature + Sex + (1 | Subject) + storage_time
y ~ current_feature + Sex + (1 | Subject) + storage_time
y ~ current_feature + Sex + (1 | Subject) + storage_time
y ~ current_feature + Sex + (1 | Subject) + storage_time
y ~ current_feature + Sex + (1 | Subject) + storage_time
y ~ current_feature + Sex + (1 | Subject) + storage_time
y ~ current_feature + Sex + (1 | Subject) + storage_time
y ~ current_feature + Sex + (1 | Subject) + storage_time
y ~ current_feature + Sex + (1 | Subject) + storage_time
y ~ current_feature + Sex + (1 | Subject) + storage_time
y ~ current_feature + Sex + (1 | Subject) + storage_time
y ~ current_feature + Sex + (1 | Subject) + storage_time
y ~ current_feature + Sex + (1 | Subject) + storage_time
y ~ current_feature + Sex + (1 | Subject) + storage_time
y ~ current_feature + Sex + (1 | Subject) + storage_time
y ~ current_feature + Sex + (1 | Subject) + storage_time
y ~ current_feature + Sex + (1 | Subject) + storage_time
y ~ current_feature + Sex + (1 | Subject) + storage_time
y ~ current_feature + Sex + (1 | Subject) + storage_time
y ~ current_feature + Sex + (1 | Subject) + storage_time
y ~ current_feature + Sex + (1 | Subject) + storage_time
y ~ current_feature + Sex + (1 | Subject) + storage_time
y ~ current_feature + Sex + (1 | Subject) + storage_time
y ~ current_feature + Sex + (1 | Subject) + storage_time
y ~ current_feature + Sex + (1 | Subject) + storage_time
y ~ current_feature + Sex + (1 | Subject) + storage_time
y ~ current_feature + Sex + (1 | Subject) + storage_time
y ~ current_feature + Sex + (1 | Subject) + storage_time
y ~ current_feature + Sex + (1 | Subject) + storage_time
y ~ current_feature + Sex + (1 | Subject) + storage_time
y ~ current_feature + Sex + (1 | Subject) + storage_time
y ~ current_feature + Sex + (1 | Subject) + storage_time
y ~ current_feature + Sex + (1 | Subject) + storage_time
y ~ current_feature + Sex + (1 | Subject) + storage_time
y ~ current_feature + Sex + (1 | Subject) + storage_time
y ~ current_feature + Sex + (1 | Subject) + storage_time
y ~ current_feature + Sex + (1 | Subject) + storage_time
y ~ current_feature + Sex + (1 | Subject) + storage_time

y ~ current_feature + Sex + (1 | Subject) + storage_time
y ~ current_feature + Sex + (1 | Subject) + storage_time
y ~ current_feature + Sex + (1 | Subject) + storage_time
y ~ current_feature + Sex + (1 | Subject) + storage_time
y ~ current_feature + Sex + (1 | Subject) + storage_time
y ~ current_feature + Sex + (1 | Subject) + storage_time
y ~ current_feature + Sex + (1 | Subject) + storage_time
y ~ current_feature + Sex + (1 | Subject) + storage_time
y ~ current_feature + Sex + (1 | Subject) + storage_time
y ~ current_feature + Sex + (1 | Subject) + storage_time
y ~ current_feature + Sex + (1 | Subject) + storage_time
y ~ current_feature + Sex + (1 | Subject) + storage_time
y ~ current_feature + Sex + (1 | Subject) + storage_time
y ~ current_feature + Sex + (1 | Subject) + storage_time
y ~ current_feature + Sex + (1 | Subject) + storage_time
y ~ current_feature + Sex + (1 | Subject) + storage_time
y ~ current_feature + Sex + (1 | Subject) + storage_time
y ~ current_feature + Sex + (1 | Subject) + storage_time
y ~ current_feature + Sex + (1 | Subject) + storage_time
y ~ current_feature + Sex + (1 | Subject) + storage_time
y ~ current_feature + Sex + (1 | Subject) + storage_time
y ~ current_feature + Sex + (1 | Subject) + storage_time
y ~ current_feature + Sex + (1 | Subject) + storage_time
y ~ current_feature + Sex + (1 | Subject) + storage_time
y ~ current_feature + Sex + (1 | Subject) + storage_time
y ~ current_feature + Sex + (1 | Subject) + storage_time
y ~ current_feature + Sex + (1 | Subject) + storage_time
y ~ current_feature + Sex + (1 | Subject) + storage_time
y ~ current_feature + Sex + (1 | Subject) + storage_time
y ~ current_feature + Sex + (1 | Subject) + storage_time
y ~ current_feature + Sex + (1 | Subject) + storage_time
y ~ current_feature + Sex + (1 | Subject) + storage_time
y ~ current_feature + Sex + (1 | Subject) + storage_time
y ~ current_feature + Sex + (1 | Subject) + storage_time
y ~ current_feature + Sex + (1 | Subject) + storage_time
y ~ current_feature + Sex + (1 | Subject) + storage_time
y ~ current_feature + Sex + (1 | Subject) + storage_time
y ~ current_feature + Sex + (1 | Subject) + storage_time
y ~ current_feature + Sex + (1 | Subject) + storage_time
y ~ current_feature + Sex + (1 | Subject) + storage_time
y ~ current_feature + Sex + (1 | Subject) + storage_time
y ~ current_feature + Sex + (1 | Subject) + storage_time
y ~ current_feature + Sex + (1 | Subject) + storage_time
y ~ current_feature + Sex + (1 | Subject) + storage_time
y ~ current_feature + Sex + (1 | Subject) + storage_time
y ~ current_feature + Sex + (1 | Subject) + storage_time
y ~ current_feature + Sex + (1 | Subject) + storage_time
y ~ current_feature + Sex + (1 | Subject) + storage_time

y ~ current_feature + Sex + (1 | Subject) + storage_time
y ~ current_feature + Sex + (1 | Subject) + storage_time
y ~ current_feature + Sex + (1 | Subject) + storage_time
y ~ current_feature + Sex + (1 | Subject) + storage_time
y ~ current_feature + Sex + (1 | Subject) + storage_time
y ~ current_feature + Sex + (1 | Subject) + storage_time
y ~ current_feature + Sex + (1 | Subject) + storage_time
y ~ current_feature + Sex + (1 | Subject) + storage_time
y ~ current_feature + Sex + (1 | Subject) + storage_time
y ~ current_feature + Sex + (1 | Subject) + storage_time
y ~ current_feature + Sex + (1 | Subject) + storage_time
y ~ current_feature + Sex + (1 | Subject) + storage_time
y ~ current_feature + Sex + (1 | Subject) + storage_time
y ~ current_feature + Sex + (1 | Subject) + storage_time
y ~ current_feature + Sex + (1 | Subject) + storage_time
y ~ current_feature + Sex + (1 | Subject) + storage_time
y ~ current_feature + Sex + (1 | Subject) + storage_time
y ~ current_feature + Sex + (1 | Subject) + storage_time
y ~ current_feature + Sex + (1 | Subject) + storage_time
y ~ current_feature + Sex + (1 | Subject) + storage_time
y ~ current_feature + Sex + (1 | Subject) + storage_time
y ~ current_feature + Sex + (1 | Subject) + storage_time
y ~ current_feature + Sex + (1 | Subject) + storage_time
y ~ current_feature + Sex + (1 | Subject) + storage_time
y ~ current_feature + Sex + (1 | Subject) + storage_time
y ~ current_feature + Sex + (1 | Subject) + storage_time
y ~ current_feature + Sex + (1 | Subject) + storage_time
y ~ current_feature + Sex + (1 | Subject) + storage_time
y ~ current_feature + Sex + (1 | Subject) + storage_time
y ~ current_feature + Sex + (1 | Subject) + storage_time
y ~ current_feature + Sex + (1 | Subject) + storage_time
y ~ current_feature + Sex + (1 | Subject) + storage_time
y ~ current_feature + Sex + (1 | Subject) + storage_time
y ~ current_feature + Sex + (1 | Subject) + storage_time
y ~ current_feature + Sex + (1 | Subject) + storage_time
y ~ current_feature + Sex + (1 | Subject) + storage_time
y ~ current_feature + Sex + (1 | Subject) + storage_time
y ~ current_feature + Sex + (1 | Subject) + storage_time
y ~ current_feature + Sex + (1 | Subject) + storage_time
y ~ current_feature + Sex + (1 | Subject) + storage_time
y ~ current_feature + Sex + (1 | Subject) + storage_time
y ~ current_feature + Sex + (1 | Subject) + storage_time
y ~ current_feature + Sex + (1 | Subject) + storage_time
y ~ current_feature + Sex + (1 | Subject) + storage_time
y ~ current_feature + Sex + (1 | Subject) + storage_time
y ~ current_feature + Sex + (1 | Subject) + storage_time
y ~ current_feature + Sex + (1 | Subject) + storage_time
y ~ current_feature + Sex + (1 | Subject) + storage_time

y ~ current_feature + Sex + (1 | Subject) + storage_time
y ~ current_feature + Sex + (1 | Subject) + storage_time
y ~ current_feature + Sex + (1 | Subject) + storage_time
y ~ current_feature + Sex + (1 | Subject) + storage_time
y ~ current_feature + Sex + (1 | Subject) + storage_time
y ~ current_feature + Sex + (1 | Subject) + storage_time
y ~ current_feature + Sex + (1 | Subject) + storage_time
y ~ current_feature + Sex + (1 | Subject) + storage_time
y ~ current_feature + Sex + (1 | Subject) + storage_time
y ~ current_feature + Sex + (1 | Subject) + storage_time
y ~ current_feature + Sex + (1 | Subject) + storage_time
y ~ current_feature + Sex + (1 | Subject) + storage_time
y ~ current_feature + Sex + (1 | Subject) + storage_time
y ~ current_feature + Sex + (1 | Subject) + storage_time
y ~ current_feature + Sex + (1 | Subject) + storage_time
y ~ current_feature + Sex + (1 | Subject) + storage_time
y ~ current_feature + Sex + (1 | Subject) + storage_time
y ~ current_feature + Sex + (1 | Subject) + storage_time
y ~ current_feature + Sex + (1 | Subject) + storage_time
y ~ current_feature + Sex + (1 | Subject) + storage_time
y ~ current_feature + Sex + (1 | Subject) + storage_time
y ~ current_feature + Sex + (1 | Subject) + storage_time
y ~ current_feature + Sex + (1 | Subject) + storage_time
y ~ current_feature + Sex + (1 | Subject) + storage_time
y ~ current_feature + Sex + (1 | Subject) + storage_time
y ~ current_feature + Sex + (1 | Subject) + storage_time
y ~ current_feature + Sex + (1 | Subject) + storage_time
y ~ current_feature + Sex + (1 | Subject) + storage_time
y ~ current_feature + Sex + (1 | Subject) + storage_time
y ~ current_feature + Sex + (1 | Subject) + storage_time
y ~ current_feature + Sex + (1 | Subject) + storage_time
y ~ current_feature + Sex + (1 | Subject) + storage_time
y ~ current_feature + Sex + (1 | Subject) + storage_time
y ~ current_feature + Sex + (1 | Subject) + storage_time
y ~ current_feature + Sex + (1 | Subject) + storage_time
y ~ current_feature + Sex + (1 | Subject) + storage_time
y ~ current_feature + Sex + (1 | Subject) + storage_time
y ~ current_feature + Sex + (1 | Subject) + storage_time
y ~ current_feature + Sex + (1 | Subject) + storage_time
y ~ current_feature + Sex + (1 | Subject) + storage_time
y ~ current_feature + Sex + (1 | Subject) + storage_time
y ~ current_feature + Sex + (1 | Subject) + storage_time
y ~ current_feature + Sex + (1 | Subject) + storage_time
y ~ current_feature + Sex + (1 | Subject) + storage_time
y ~ current_feature + Sex + (1 | Subject) + storage_time
y ~ current_feature + Sex + (1 | Subject) + storage_time
y ~ current_feature + Sex + (1 | Subject) + storage_time
y ~ current_feature + Sex + (1 | Subject) + storage_time

y ~ current_feature + Sex + (1 | Subject) + storage_time
y ~ current_feature + Sex + (1 | Subject) + storage_time
y ~ current_feature + Sex + (1 | Subject) + storage_time
y ~ current_feature + Sex + (1 | Subject) + storage_time
y ~ current_feature + Sex + (1 | Subject) + storage_time
y ~ current_feature + Sex + (1 | Subject) + storage_time
y ~ current_feature + Sex + (1 | Subject) + storage_time
y ~ current_feature + Sex + (1 | Subject) + storage_time
y ~ current_feature + Sex + (1 | Subject) + storage_time
y ~ current_feature + Sex + (1 | Subject) + storage_time
y ~ current_feature + Sex + (1 | Subject) + storage_time
y ~ current_feature + Sex + (1 | Subject) + storage_time
y ~ current_feature + Sex + (1 | Subject) + storage_time
y ~ current_feature + Sex + (1 | Subject) + storage_time
y ~ current_feature + Sex + (1 | Subject) + storage_time
y ~ current_feature + Sex + (1 | Subject) + storage_time
y ~ current_feature + Sex + (1 | Subject) + storage_time
y ~ current_feature + Sex + (1 | Subject) + storage_time
y ~ current_feature + Sex + (1 | Subject) + storage_time
y ~ current_feature + Sex + (1 | Subject) + storage_time
y ~ current_feature + Sex + (1 | Subject) + storage_time
y ~ current_feature + Sex + (1 | Subject) + storage_time
y ~ current_feature + Sex + (1 | Subject) + storage_time
y ~ current_feature + Sex + (1 | Subject) + storage_time
y ~ current_feature + Sex + (1 | Subject) + storage_time
y ~ current_feature + Sex + (1 | Subject) + storage_time
y ~ current_feature + Sex + (1 | Subject) + storage_time
y ~ current_feature + Sex + (1 | Subject) + storage_time
y ~ current_feature + Sex + (1 | Subject) + storage_time
y ~ current_feature + Sex + (1 | Subject) + storage_time
y ~ current_feature + Sex + (1 | Subject) + storage_time
y ~ current_feature + Sex + (1 | Subject) + storage_time
y ~ current_feature + Sex + (1 | Subject) + storage_time
y ~ current_feature + Sex + (1 | Subject) + storage_time
y ~ current_feature + Sex + (1 | Subject) + storage_time
y ~ current_feature + Sex + (1 | Subject) + storage_time
y ~ current_feature + Sex + (1 | Subject) + storage_time
y ~ current_feature + Sex + (1 | Subject) + storage_time
y ~ current_feature + Sex + (1 | Subject) + storage_time
y ~ current_feature + Sex + (1 | Subject) + storage_time
y ~ current_feature + Sex + (1 | Subject) + storage_time
y ~ current_feature + Sex + (1 | Subject) + storage_time
y ~ current_feature + Sex + (1 | Subject) + storage_time
y ~ current_feature + Sex + (1 | Subject) + storage_time
y ~ current_feature + Sex + (1 | Subject) + storage_time
y ~ current_feature + Sex + (1 | Subject) + storage_time
y ~ current_feature + Sex + (1 | Subject) + storage_time
y ~ current_feature + Sex + (1 | Subject) + storage_time

y ~ current_feature + Sex + (1 | Subject) + storage_time
y ~ current_feature + Sex + (1 | Subject) + storage_time
y ~ current_feature + Sex + (1 | Subject) + storage_time
y ~ current_feature + Sex + (1 | Subject) + storage_time
y ~ current_feature + Sex + (1 | Subject) + storage_time
y ~ current_feature + Sex + (1 | Subject) + storage_time
y ~ current_feature + Sex + (1 | Subject) + storage_time
y ~ current_feature + Sex + (1 | Subject) + storage_time
y ~ current_feature + Sex + (1 | Subject) + storage_time
y ~ current_feature + Sex + (1 | Subject) + storage_time
y ~ current_feature + Sex + (1 | Subject) + storage_time
y ~ current_feature + Sex + (1 | Subject) + storage_time
y ~ current_feature + Sex + (1 | Subject) + storage_time
y ~ current_feature + Sex + (1 | Subject) + storage_time
y ~ current_feature + Sex + (1 | Subject) + storage_time
y ~ current_feature + Sex + (1 | Subject) + storage_time
y ~ current_feature + Sex + (1 | Subject) + storage_time
y ~ current_feature + Sex + (1 | Subject) + storage_time
y ~ current_feature + Sex + (1 | Subject) + storage_time
y ~ current_feature + Sex + (1 | Subject) + storage_time
y ~ current_feature + Sex + (1 | Subject) + storage_time
y ~ current_feature + Sex + (1 | Subject) + storage_time
y ~ current_feature + Sex + (1 | Subject) + storage_time
y ~ current_feature + Sex + (1 | Subject) + storage_time
y ~ current_feature + Sex + (1 | Subject) + storage_time
y ~ current_feature + Sex + (1 | Subject) + storage_time
y ~ current_feature + Sex + (1 | Subject) + storage_time
y ~ current_feature + Sex + (1 | Subject) + storage_time
y ~ current_feature + Sex + (1 | Subject) + storage_time
y ~ current_feature + Sex + (1 | Subject) + storage_time
y ~ current_feature + Sex + (1 | Subject) + storage_time
y ~ current_feature + Sex + (1 | Subject) + storage_time
y ~ current_feature + Sex + (1 | Subject) + storage_time
y ~ current_feature + Sex + (1 | Subject) + storage_time
y ~ current_feature + Sex + (1 | Subject) + storage_time
y ~ current_feature + Sex + (1 | Subject) + storage_time
y ~ current_feature + Sex + (1 | Subject) + storage_time
y ~ current_feature + Sex + (1 | Subject) + storage_time
y ~ current_feature + Sex + (1 | Subject) + storage_time
y ~ current_feature + Sex + (1 | Subject) + storage_time
y ~ current_feature + Sex + (1 | Subject) + storage_time
y ~ current_feature + Sex + (1 | Subject) + storage_time
y ~ current_feature + Sex + (1 | Subject) + storage_time
y ~ current_feature + Sex + (1 | Subject) + storage_time
y ~ current_feature + Sex + (1 | Subject) + storage_time
y ~ current_feature + Sex + (1 | Subject) + storage_time
y ~ current_feature + Sex + (1 | Subject) + storage_time
y ~ current_feature + Sex + (1 | Subject) + storage_time

y ~ current_feature + Sex + (1 | Subject) + storage_time
y ~ current_feature + Sex + (1 | Subject) + storage_time
y ~ current_feature + Sex + (1 | Subject) + storage_time
y ~ current_feature + Sex + (1 | Subject) + storage_time
y ~ current_feature + Sex + (1 | Subject) + storage_time
y ~ current_feature + Sex + (1 | Subject) + storage_time
y ~ current_feature + Sex + (1 | Subject) + storage_time
y ~ current_feature + Sex + (1 | Subject) + storage_time
y ~ current_feature + Sex + (1 | Subject) + storage_time
y ~ current_feature + Sex + (1 | Subject) + storage_time
y ~ current_feature + Sex + (1 | Subject) + storage_time
y ~ current_feature + Sex + (1 | Subject) + storage_time
y ~ current_feature + Sex + (1 | Subject) + storage_time
y ~ current_feature + Sex + (1 | Subject) + storage_time
y ~ current_feature + Sex + (1 | Subject) + storage_time
y ~ current_feature + Sex + (1 | Subject) + storage_time
y ~ current_feature + Sex + (1 | Subject) + storage_time
y ~ current_feature + Sex + (1 | Subject) + storage_time
y ~ current_feature + Sex + (1 | Subject) + storage_time
y ~ current_feature + Sex + (1 | Subject) + storage_time
y ~ current_feature + Sex + (1 | Subject) + storage_time
y ~ current_feature + Sex + (1 | Subject) + storage_time
y ~ current_feature + Sex + (1 | Subject) + storage_time
y ~ current_feature + Sex + (1 | Subject) + storage_time
y ~ current_feature + Sex + (1 | Subject) + storage_time
y ~ current_feature + Sex + (1 | Subject) + storage_time
y ~ current_feature + Sex + (1 | Subject) + storage_time
y ~ current_feature + Sex + (1 | Subject) + storage_time
y ~ current_feature + Sex + (1 | Subject) + storage_time
y ~ current_feature + Sex + (1 | Subject) + storage_time
y ~ current_feature + Sex + (1 | Subject) + storage_time
y ~ current_feature + Sex + (1 | Subject) + storage_time
y ~ current_feature + Sex + (1 | Subject) + storage_time
y ~ current_feature + Sex + (1 | Subject) + storage_time
y ~ current_feature + Sex + (1 | Subject) + storage_time
y ~ current_feature + Sex + (1 | Subject) + storage_time
y ~ current_feature + Sex + (1 | Subject) + storage_time
y ~ current_feature + Sex + (1 | Subject) + storage_time
y ~ current_feature + Sex + (1 | Subject) + storage_time
y ~ current_feature + Sex + (1 | Subject) + storage_time
y ~ current_feature + Sex + (1 | Subject) + storage_time
y ~ current_feature + Sex + (1 | Subject) + storage_time
y ~ current_feature + Sex + (1 | Subject) + storage_time
y ~ current_feature + Sex + (1 | Subject) + storage_time
y ~ current_feature + Sex + (1 | Subject) + storage_time
y ~ current_feature + Sex + (1 | Subject) + storage_time
y ~ current_feature + Sex + (1 | Subject) + storage_time
y ~ current_feature + Sex + (1 | Subject) + storage_time

y ~ current_feature + Sex + (1 | Subject) + storage_time
y ~ current_feature + Sex + (1 | Subject) + storage_time
y ~ current_feature + Sex + (1 | Subject) + storage_time
y ~ current_feature + Sex + (1 | Subject) + storage_time
y ~ current_feature + Sex + (1 | Subject) + storage_time
y ~ current_feature + Sex + (1 | Subject) + storage_time
y ~ current_feature + Sex + (1 | Subject) + storage_time
y ~ current_feature + Sex + (1 | Subject) + storage_time
y ~ current_feature + Sex + (1 | Subject) + storage_time
y ~ current_feature + Sex + (1 | Subject) + storage_time
y ~ current_feature + Sex + (1 | Subject) + storage_time
y ~ current_feature + Sex + (1 | Subject) + storage_time
y ~ current_feature + Sex + (1 | Subject) + storage_time
y ~ current_feature + Sex + (1 | Subject) + storage_time
y ~ current_feature + Sex + (1 | Subject) + storage_time
y ~ current_feature + Sex + (1 | Subject) + storage_time
y ~ current_feature + Sex + (1 | Subject) + storage_time
y ~ current_feature + Sex + (1 | Subject) + storage_time
y ~ current_feature + Sex + (1 | Subject) + storage_time
y ~ current_feature + Sex + (1 | Subject) + storage_time
y ~ current_feature + Sex + (1 | Subject) + storage_time
y ~ current_feature + Sex + (1 | Subject) + storage_time
y ~ current_feature + Sex + (1 | Subject) + storage_time
y ~ current_feature + Sex + (1 | Subject) + storage_time
y ~ current_feature + Sex + (1 | Subject) + storage_time
y ~ current_feature + Sex + (1 | Subject) + storage_time
y ~ current_feature + Sex + (1 | Subject) + storage_time
y ~ current_feature + Sex + (1 | Subject) + storage_time
y ~ current_feature + Sex + (1 | Subject) + storage_time
y ~ current_feature + Sex + (1 | Subject) + storage_time
y ~ current_feature + Sex + (1 | Subject) + storage_time
y ~ current_feature + Sex + (1 | Subject) + storage_time
y ~ current_feature + Sex + (1 | Subject) + storage_time
y ~ current_feature + Sex + (1 | Subject) + storage_time
y ~ current_feature + Sex + (1 | Subject) + storage_time
y ~ current_feature + Sex + (1 | Subject) + storage_time
y ~ current_feature + Sex + (1 | Subject) + storage_time
y ~ current_feature + Sex + (1 | Subject) + storage_time
y ~ current_feature + Sex + (1 | Subject) + storage_time
y ~ current_feature + Sex + (1 | Subject) + storage_time
y ~ current_feature + Sex + (1 | Subject) + storage_time
y ~ current_feature + Sex + (1 | Subject) + storage_time
y ~ current_feature + Sex + (1 | Subject) + storage_time
y ~ current_feature + Sex + (1 | Subject) + storage_time
y ~ current_feature + Sex + (1 | Subject) + storage_time
y ~ current_feature + Sex + (1 | Subject) + storage_time
y ~ current_feature + Sex + (1 | Subject) + storage_time
y ~ current_feature + Sex + (1 | Subject) + storage_time

y ~ current_feature + Sex + (1 | Subject) + storage_time
y ~ current_feature + Sex + (1 | Subject) + storage_time
y ~ current_feature + Sex + (1 | Subject) + storage_time
y ~ current_feature + Sex + (1 | Subject) + storage_time
y ~ current_feature + Sex + (1 | Subject) + storage_time
y ~ current_feature + Sex + (1 | Subject) + storage_time
y ~ current_feature + Sex + (1 | Subject) + storage_time
y ~ current_feature + Sex + (1 | Subject) + storage_time
y ~ current_feature + Sex + (1 | Subject) + storage_time
y ~ current_feature + Sex + (1 | Subject) + storage_time
y ~ current_feature + Sex + (1 | Subject) + storage_time
y ~ current_feature + Sex + (1 | Subject) + storage_time
y ~ current_feature + Sex + (1 | Subject) + storage_time
y ~ current_feature + Sex + (1 | Subject) + storage_time
y ~ current_feature + Sex + (1 | Subject) + storage_time
y ~ current_feature + Sex + (1 | Subject) + storage_time
y ~ current_feature + Sex + (1 | Subject) + storage_time
y ~ current_feature + Sex + (1 | Subject) + storage_time
y ~ current_feature + Sex + (1 | Subject) + storage_time
y ~ current_feature + Sex + (1 | Subject) + storage_time
y ~ current_feature + Sex + (1 | Subject) + storage_time
y ~ current_feature + Sex + (1 | Subject) + storage_time
y ~ current_feature + Sex + (1 | Subject) + storage_time
y ~ current_feature + Sex + (1 | Subject) + storage_time
y ~ current_feature + Sex + (1 | Subject) + storage_time
y ~ current_feature + Sex + (1 | Subject) + storage_time
y ~ current_feature + Sex + (1 | Subject) + storage_time
y ~ current_feature + Sex + (1 | Subject) + storage_time
y ~ current_feature + Sex + (1 | Subject) + storage_time
y ~ current_feature + Sex + (1 | Subject) + storage_time
y ~ current_feature + Sex + (1 | Subject) + storage_time
y ~ current_feature + Sex + (1 | Subject) + storage_time
y ~ current_feature + Sex + (1 | Subject) + storage_time
y ~ current_feature + Sex + (1 | Subject) + storage_time
y ~ current_feature + Sex + (1 | Subject) + storage_time
y ~ current_feature + Sex + (1 | Subject) + storage_time
y ~ current_feature + Sex + (1 | Subject) + storage_time
y ~ current_feature + Sex + (1 | Subject) + storage_time
y ~ current_feature + Sex + (1 | Subject) + storage_time
y ~ current_feature + Sex + (1 | Subject) + storage_time
y ~ current_feature + Sex + (1 | Subject) + storage_time
y ~ current_feature + Sex + (1 | Subject) + storage_time
y ~ current_feature + Sex + (1 | Subject) + storage_time
y ~ current_feature + Sex + (1 | Subject) + storage_time
y ~ current_feature + Sex + (1 | Subject) + storage_time
y ~ current_feature + Sex + (1 | Subject) + storage_time
y ~ current_feature + Sex + (1 | Subject) + storage_time
y ~ current_feature + Sex + (1 | Subject) + storage_time

y ~ current_feature + Sex + (1 | Subject) + storage_time
y ~ current_feature + Sex + (1 | Subject) + storage_time
y ~ current_feature + Sex + (1 | Subject) + storage_time
y ~ current_feature + Sex + (1 | Subject) + storage_time
y ~ current_feature + Sex + (1 | Subject) + storage_time
y ~ current_feature + Sex + (1 | Subject) + storage_time
y ~ current_feature + Sex + (1 | Subject) + storage_time
y ~ current_feature + Sex + (1 | Subject) + storage_time
y ~ current_feature + Sex + (1 | Subject) + storage_time
y ~ current_feature + Sex + (1 | Subject) + storage_time
y ~ current_feature + Sex + (1 | Subject) + storage_time
y ~ current_feature + Sex + (1 | Subject) + storage_time
y ~ current_feature + Sex + (1 | Subject) + storage_time
y ~ current_feature + Sex + (1 | Subject) + storage_time
y ~ current_feature + Sex + (1 | Subject) + storage_time
y ~ current_feature + Sex + (1 | Subject) + storage_time
y ~ current_feature + Sex + (1 | Subject) + storage_time
y ~ current_feature + Sex + (1 | Subject) + storage_time
y ~ current_feature + Sex + (1 | Subject) + storage_time
y ~ current_feature + Sex + (1 | Subject) + storage_time
y ~ current_feature + Sex + (1 | Subject) + storage_time
y ~ current_feature + Sex + (1 | Subject) + storage_time
y ~ current_feature + Sex + (1 | Subject) + storage_time
y ~ current_feature + Sex + (1 | Subject) + storage_time
y ~ current_feature + Sex + (1 | Subject) + storage_time
y ~ current_feature + Sex + (1 | Subject) + storage_time
y ~ current_feature + Sex + (1 | Subject) + storage_time
y ~ current_feature + Sex + (1 | Subject) + storage_time
y ~ current_feature + Sex + (1 | Subject) + storage_time
y ~ current_feature + Sex + (1 | Subject) + storage_time
y ~ current_feature + Sex + (1 | Subject) + storage_time
y ~ current_feature + Sex + (1 | Subject) + storage_time
y ~ current_feature + Sex + (1 | Subject) + storage_time
y ~ current_feature + Sex + (1 | Subject) + storage_time
y ~ current_feature + Sex + (1 | Subject) + storage_time
y ~ current_feature + Sex + (1 | Subject) + storage_time
y ~ current_feature + Sex + (1 | Subject) + storage_time
y ~ current_feature + Sex + (1 | Subject) + storage_time
y ~ current_feature + Sex + (1 | Subject) + storage_time
y ~ current_feature + Sex + (1 | Subject) + storage_time
y ~ current_feature + Sex + (1 | Subject) + storage_time
y ~ current_feature + Sex + (1 | Subject) + storage_time
y ~ current_feature + Sex + (1 | Subject) + storage_time
y ~ current_feature + Sex + (1 | Subject) + storage_time
y ~ current_feature + Sex + (1 | Subject) + storage_time
y ~ current_feature + Sex + (1 | Subject) + storage_time
y ~ current_feature + Sex + (1 | Subject) + storage_time
y ~ current_feature + Sex + (1 | Subject) + storage_time

y ~ current_feature + Sex + (1 | Subject) + storage_time
y ~ current_feature + Sex + (1 | Subject) + storage_time
y ~ current_feature + Sex + (1 | Subject) + storage_time
y ~ current_feature + Sex + (1 | Subject) + storage_time
y ~ current_feature + Sex + (1 | Subject) + storage_time
y ~ current_feature + Sex + (1 | Subject) + storage_time
y ~ current_feature + Sex + (1 | Subject) + storage_time
y ~ current_feature + Sex + (1 | Subject) + storage_time
y ~ current_feature + Sex + (1 | Subject) + storage_time
y ~ current_feature + Sex + (1 | Subject) + storage_time
y ~ current_feature + Sex + (1 | Subject) + storage_time
y ~ current_feature + Sex + (1 | Subject) + storage_time
y ~ current_feature + Sex + (1 | Subject) + storage_time
y ~ current_feature + Sex + (1 | Subject) + storage_time
y ~ current_feature + Sex + (1 | Subject) + storage_time
y ~ current_feature + Sex + (1 | Subject) + storage_time
y ~ current_feature + Sex + (1 | Subject) + storage_time
y ~ current_feature + Sex + (1 | Subject) + storage_time
y ~ current_feature + Sex + (1 | Subject) + storage_time
y ~ current_feature + Sex + (1 | Subject) + storage_time
y ~ current_feature + Sex + (1 | Subject) + storage_time
y ~ current_feature + Sex + (1 | Subject) + storage_time
y ~ current_feature + Sex + (1 | Subject) + storage_time
y ~ current_feature + Sex + (1 | Subject) + storage_time
y ~ current_feature + Sex + (1 | Subject) + storage_time
y ~ current_feature + Sex + (1 | Subject) + storage_time
y ~ current_feature + Sex + (1 | Subject) + storage_time
y ~ current_feature + Sex + (1 | Subject) + storage_time
y ~ current_feature + Sex + (1 | Subject) + storage_time
y ~ current_feature + Sex + (1 | Subject) + storage_time
y ~ current_feature + Sex + (1 | Subject) + storage_time
y ~ current_feature + Sex + (1 | Subject) + storage_time
y ~ current_feature + Sex + (1 | Subject) + storage_time
y ~ current_feature + Sex + (1 | Subject) + storage_time
y ~ current_feature + Sex + (1 | Subject) + storage_time
y ~ current_feature + Sex + (1 | Subject) + storage_time
y ~ current_feature + Sex + (1 | Subject) + storage_time
y ~ current_feature + Sex + (1 | Subject) + storage_time
y ~ current_feature + Sex + (1 | Subject) + storage_time
y ~ current_feature + Sex + (1 | Subject) + storage_time
y ~ current_feature + Sex + (1 | Subject) + storage_time
y ~ current_feature + Sex + (1 | Subject) + storage_time
y ~ current_feature + Sex + (1 | Subject) + storage_time
y ~ current_feature + Sex + (1 | Subject) + storage_time
y ~ current_feature + Sex + (1 | Subject) + storage_time
y ~ current_feature + Sex + (1 | Subject) + storage_time
y ~ current_feature + Sex + (1 | Subject) + storage_time
y ~ current_feature + Sex + (1 | Subject) + storage_time

y ~ current_feature + Sex + (1 | Subject) + storage_time
y ~ current_feature + Sex + (1 | Subject) + storage_time
y ~ current_feature + Sex + (1 | Subject) + storage_time
y ~ current_feature + Sex + (1 | Subject) + storage_time
y ~ current_feature + Sex + (1 | Subject) + storage_time
y ~ current_feature + Sex + (1 | Subject) + storage_time
y ~ current_feature + Sex + (1 | Subject) + storage_time
y ~ current_feature + Sex + (1 | Subject) + storage_time
y ~ current_feature + Sex + (1 | Subject) + storage_time
y ~ current_feature + Sex + (1 | Subject) + storage_time
y ~ current_feature + Sex + (1 | Subject) + storage_time
y ~ current_feature + Sex + (1 | Subject) + storage_time
y ~ current_feature + Sex + (1 | Subject) + storage_time
y ~ current_feature + Sex + (1 | Subject) + storage_time
y ~ current_feature + Sex + (1 | Subject) + storage_time
y ~ current_feature + Sex + (1 | Subject) + storage_time
y ~ current_feature + Sex + (1 | Subject) + storage_time
y ~ current_feature + Sex + (1 | Subject) + storage_time
y ~ current_feature + Sex + (1 | Subject) + storage_time
y ~ current_feature + Sex + (1 | Subject) + storage_time
y ~ current_feature + Sex + (1 | Subject) + storage_time
y ~ current_feature + Sex + (1 | Subject) + storage_time
y ~ current_feature + Sex + (1 | Subject) + storage_time
y ~ current_feature + Sex + (1 | Subject) + storage_time
y ~ current_feature + Sex + (1 | Subject) + storage_time
y ~ current_feature + Sex + (1 | Subject) + storage_time
y ~ current_feature + Sex + (1 | Subject) + storage_time
y ~ current_feature + Sex + (1 | Subject) + storage_time
y ~ current_feature + Sex + (1 | Subject) + storage_time
y ~ current_feature + Sex + (1 | Subject) + storage_time
y ~ current_feature + Sex + (1 | Subject) + storage_time
y ~ current_feature + Sex + (1 | Subject) + storage_time
y ~ current_feature + Sex + (1 | Subject) + storage_time
y ~ current_feature + Sex + (1 | Subject) + storage_time
y ~ current_feature + Sex + (1 | Subject) + storage_time
y ~ current_feature + Sex + (1 | Subject) + storage_time
y ~ current_feature + Sex + (1 | Subject) + storage_time
y ~ current_feature + Sex + (1 | Subject) + storage_time
y ~ current_feature + Sex + (1 | Subject) + storage_time
y ~ current_feature + Sex + (1 | Subject) + storage_time
y ~ current_feature + Sex + (1 | Subject) + storage_time
y ~ current_feature + Sex + (1 | Subject) + storage_time
y ~ current_feature + Sex + (1 | Subject) + storage_time
y ~ current_feature + Sex + (1 | Subject) + storage_time
y ~ current_feature + Sex + (1 | Subject) + storage_time
y ~ current_feature + Sex + (1 | Subject) + storage_time
y ~ current_feature + Sex + (1 | Subject) + storage_time
y ~ current_feature + Sex + (1 | Subject) + storage_time

y ~ current_feature + Sex + (1 | Subject) + storage_time
y ~ current_feature + Sex + (1 | Subject) + storage_time
y ~ current_feature + Sex + (1 | Subject) + storage_time
y ~ current_feature + Sex + (1 | Subject) + storage_time
y ~ current_feature + Sex + (1 | Subject) + storage_time
y ~ current_feature + Sex + (1 | Subject) + storage_time
y ~ current_feature + Sex + (1 | Subject) + storage_time
y ~ current_feature + Sex + (1 | Subject) + storage_time
y ~ current_feature + Sex + (1 | Subject) + storage_time
y ~ current_feature + Sex + (1 | Subject) + storage_time
y ~ current_feature + Sex + (1 | Subject) + storage_time
y ~ current_feature + Sex + (1 | Subject) + storage_time
y ~ current_feature + Sex + (1 | Subject) + storage_time
y ~ current_feature + Sex + (1 | Subject) + storage_time
y ~ current_feature + Sex + (1 | Subject) + storage_time
y ~ current_feature + Sex + (1 | Subject) + storage_time
y ~ current_feature + Sex + (1 | Subject) + storage_time
y ~ current_feature + Sex + (1 | Subject) + storage_time
y ~ current_feature + Sex + (1 | Subject) + storage_time
y ~ current_feature + Sex + (1 | Subject) + storage_time
y ~ current_feature + Sex + (1 | Subject) + storage_time
y ~ current_feature + Sex + (1 | Subject) + storage_time
y ~ current_feature + Sex + (1 | Subject) + storage_time
y ~ current_feature + Sex + (1 | Subject) + storage_time
y ~ current_feature + Sex + (1 | Subject) + storage_time
y ~ current_feature + Sex + (1 | Subject) + storage_time
y ~ current_feature + Sex + (1 | Subject) + storage_time
y ~ current_feature + Sex + (1 | Subject) + storage_time
y ~ current_feature + Sex + (1 | Subject) + storage_time
y ~ current_feature + Sex + (1 | Subject) + storage_time
y ~ current_feature + Sex + (1 | Subject) + storage_time
y ~ current_feature + Sex + (1 | Subject) + storage_time
y ~ current_feature + Sex + (1 | Subject) + storage_time
y ~ current_feature + Sex + (1 | Subject) + storage_time
y ~ current_feature + Sex + (1 | Subject) + storage_time
y ~ current_feature + Sex + (1 | Subject) + storage_time
y ~ current_feature + Sex + (1 | Subject) + storage_time
y ~ current_feature + Sex + (1 | Subject) + storage_time
y ~ current_feature + Sex + (1 | Subject) + storage_time
y ~ current_feature + Sex + (1 | Subject) + storage_time
y ~ current_feature + Sex + (1 | Subject) + storage_time
y ~ current_feature + Sex + (1 | Subject) + storage_time
y ~ current_feature + Sex + (1 | Subject) + storage_time
y ~ current_feature + Sex + (1 | Subject) + storage_time
y ~ current_feature + Sex + (1 | Subject) + storage_time
y ~ current_feature + Sex + (1 | Subject) + storage_time
y ~ current_feature + Sex + (1 | Subject) + storage_time
y ~ current_feature + Sex + (1 | Subject) + storage_time

y ~ current_feature + Sex + (1 | Subject) + storage_time
y ~ current_feature + Sex + (1 | Subject) + storage_time
y ~ current_feature + Sex + (1 | Subject) + storage_time
y ~ current_feature + Sex + (1 | Subject) + storage_time
y ~ current_feature + Sex + (1 | Subject) + storage_time
y ~ current_feature + Sex + (1 | Subject) + storage_time
y ~ current_feature + Sex + (1 | Subject) + storage_time
y ~ current_feature + Sex + (1 | Subject) + storage_time
y ~ current_feature + Sex + (1 | Subject) + storage_time
y ~ current_feature + Sex + (1 | Subject) + storage_time
y ~ current_feature + Sex + (1 | Subject) + storage_time
y ~ current_feature + Sex + (1 | Subject) + storage_time
y ~ current_feature + Sex + (1 | Subject) + storage_time
y ~ current_feature + Sex + (1 | Subject) + storage_time
y ~ current_feature + Sex + (1 | Subject) + storage_time
y ~ current_feature + Sex + (1 | Subject) + storage_time
y ~ current_feature + Sex + (1 | Subject) + storage_time
y ~ current_feature + Sex + (1 | Subject) + storage_time
y ~ current_feature + Sex + (1 | Subject) + storage_time
y ~ current_feature + Sex + (1 | Subject) + storage_time
y ~ current_feature + Sex + (1 | Subject) + storage_time
y ~ current_feature + Sex + (1 | Subject) + storage_time
y ~ current_feature + Sex + (1 | Subject) + storage_time
y ~ current_feature + Sex + (1 | Subject) + storage_time
y ~ current_feature + Sex + (1 | Subject) + storage_time
y ~ current_feature + Sex + (1 | Subject) + storage_time
y ~ current_feature + Sex + (1 | Subject) + storage_time
y ~ current_feature + Sex + (1 | Subject) + storage_time
y ~ current_feature + Sex + (1 | Subject) + storage_time
y ~ current_feature + Sex + (1 | Subject) + storage_time
y ~ current_feature + Sex + (1 | Subject) + storage_time
y ~ current_feature + Sex + (1 | Subject) + storage_time
y ~ current_feature + Sex + (1 | Subject) + storage_time
y ~ current_feature + Sex + (1 | Subject) + storage_time
y ~ current_feature + Sex + (1 | Subject) + storage_time
y ~ current_feature + Sex + (1 | Subject) + storage_time
y ~ current_feature + Sex + (1 | Subject) + storage_time
y ~ current_feature + Sex + (1 | Subject) + storage_time
y ~ current_feature + Sex + (1 | Subject) + storage_time
y ~ current_feature + Sex + (1 | Subject) + storage_time
y ~ current_feature + Sex + (1 | Subject) + storage_time
y ~ current_feature + Sex + (1 | Subject) + storage_time
y ~ current_feature + Sex + (1 | Subject) + storage_time
y ~ current_feature + Sex + (1 | Subject) + storage_time
y ~ current_feature + Sex + (1 | Subject) + storage_time
y ~ current_feature + Sex + (1 | Subject) + storage_time
y ~ current_feature + Sex + (1 | Subject) + storage_time
y ~ current_feature + Sex + (1 | Subject) + storage_time

y ~ current_feature + Sex + (1 | Subject) + storage_time
y ~ current_feature + Sex + (1 | Subject) + storage_time
y ~ current_feature + Sex + (1 | Subject) + storage_time
y ~ current_feature + Sex + (1 | Subject) + storage_time
y ~ current_feature + Sex + (1 | Subject) + storage_time
y ~ current_feature + Sex + (1 | Subject) + storage_time
y ~ current_feature + Sex + (1 | Subject) + storage_time
y ~ current_feature + Sex + (1 | Subject) + storage_time
y ~ current_feature + Sex + (1 | Subject) + storage_time
y ~ current_feature + Sex + (1 | Subject) + storage_time
y ~ current_feature + Sex + (1 | Subject) + storage_time
y ~ current_feature + Sex + (1 | Subject) + storage_time
y ~ current_feature + Sex + (1 | Subject) + storage_time
y ~ current_feature + Sex + (1 | Subject) + storage_time
y ~ current_feature + Sex + (1 | Subject) + storage_time
y ~ current_feature + Sex + (1 | Subject) + storage_time
y ~ current_feature + Sex + (1 | Subject) + storage_time
y ~ current_feature + Sex + (1 | Subject) + storage_time
y ~ current_feature + Sex + (1 | Subject) + storage_time
y ~ current_feature + Sex + (1 | Subject) + storage_time
y ~ current_feature + Sex + (1 | Subject) + storage_time
y ~ current_feature + Sex + (1 | Subject) + storage_time
y ~ current_feature + Sex + (1 | Subject) + storage_time
y ~ current_feature + Sex + (1 | Subject) + storage_time
y ~ current_feature + Sex + (1 | Subject) + storage_time
y ~ current_feature + Sex + (1 | Subject) + storage_time
y ~ current_feature + Sex + (1 | Subject) + storage_time
y ~ current_feature + Sex + (1 | Subject) + storage_time
y ~ current_feature + Sex + (1 | Subject) + storage_time
y ~ current_feature + Sex + (1 | Subject) + storage_time
y ~ current_feature + Sex + (1 | Subject) + storage_time
y ~ current_feature + Sex + (1 | Subject) + storage_time
y ~ current_feature + Sex + (1 | Subject) + storage_time
y ~ current_feature + Sex + (1 | Subject) + storage_time
y ~ current_feature + Sex + (1 | Subject) + storage_time
y ~ current_feature + Sex + (1 | Subject) + storage_time
y ~ current_feature + Sex + (1 | Subject) + storage_time
y ~ current_feature + Sex + (1 | Subject) + storage_time
y ~ current_feature + Sex + (1 | Subject) + storage_time
y ~ current_feature + Sex + (1 | Subject) + storage_time
y ~ current_feature + Sex + (1 | Subject) + storage_time
y ~ current_feature + Sex + (1 | Subject) + storage_time
y ~ current_feature + Sex + (1 | Subject) + storage_time
y ~ current_feature + Sex + (1 | Subject) + storage_time
y ~ current_feature + Sex + (1 | Subject) + storage_time
y ~ current_feature + Sex + (1 | Subject) + storage_time
y ~ current_feature + Sex + (1 | Subject) + storage_time
y ~ current_feature + Sex + (1 | Subject) + storage_time

```
y ~ current_feature + Sex + (1 | Subject) + storage_time
y ~ current_feature + Sex + (1 | Subject) + storage_time
y ~ current_feature + Sex + (1 | Subject) + storage_time
y ~ current_feature + Sex + (1 | Subject) + storage_time
y ~ current_feature + Sex + (1 | Subject) + storage_time
y ~ current_feature + Sex + (1 | Subject) + storage_time
y ~ current_feature + Sex + (1 | Subject) + storage_time
y ~ current_feature + Sex + (1 | Subject) + storage_time
y ~ current_feature + Sex + (1 | Subject) + storage_time
y ~ current_feature + Sex + (1 | Subject) + storage_time
y ~ current_feature + Sex + (1 | Subject) + storage_time
y ~ current_feature + Sex + (1 | Subject) + storage_time
y ~ current_feature + Sex + (1 | Subject) + storage_time
y ~ current_feature + Sex + (1 | Subject) + storage_time
y ~ current_feature + Sex + (1 | Subject) + storage_time
y ~ current_feature + Sex + (1 | Subject) + storage_time
y ~ current_feature + Sex + (1 | Subject) + storage_time
y ~ current_feature + Sex + (1 | Subject) + storage_time
y ~ current_feature + Sex + (1 | Subject) + storage_time
y ~ current_feature + Sex + (1 | Subject) + storage_time
y ~ current_feature + Sex + (1 | Subject) + storage_time
y ~ current_feature + Sex + (1 | Subject) + storage_time
y ~ current_feature + Sex + (1 | Subject) + storage_time
y ~ current_feature + Sex + (1 | Subject) + storage_time
y ~ current_feature + Sex + (1 | Subject) + storage_time
y ~ current_feature + Sex + (1 | Subject) + storage_time
y ~ current_feature + Sex + (1 | Subject) + storage_time
y ~ current_feature + Sex + (1 | Subject) + storage_time
y ~ current_feature + Sex + (1 | Subject) + storage_time
y ~ current_feature + Sex + (1 | Subject) + storage_time
y ~ current_feature + Sex + (1 | Subject) + storage_time
y ~ current_feature + Sex + (1 | Subject) + storage_time
y ~ current_feature + Sex + (1 | Subject) + storage_time
y ~ current_feature + Sex + (1 | Subject) + storage_time
y ~ current_feature + Sex + (1 | Subject) + storage_time
y ~ current_feature + Sex + (1 | Subject) + storage_time
y ~ current_feature + Sex + (1 | Subject) + storage_time
y ~ current_feature + Sex + (1 | Subject) + storage_time
y ~ current_feature + Sex + (1 | Subject) + storage_time
y ~ current_feature + Sex + (1 | Subject) + storage_time
y ~ current_feature + Sex + (1 | Subject) + storage_time
y ~ current_feature + Sex + (1 | Subject) + storage_time
y ~ current_feature + Sex + (1 | Subject) + storage_time
y ~ current_feature + Sex + (1 | Subject) + storage_time
y ~ current_feature + Sex + (1 | Subject) + storage_time
y ~ current_feature + Sex + (1 | Subject) + storage_time
y ~ current_feature + Sex + (1 | Subject) + storage_time
y ~ current_feature + Sex + (1 | Subject) + storage_time
```

```
y ~ current_feature + Sex + (1 | Subject) + storage_time
y ~ current_feature + Sex + (1 | Subject) + storage_time
y ~ current_feature + Sex + (1 | Subject) + storage_time
y ~ current_feature + Sex + (1 | Subject) + storage_time
y ~ current_feature + Sex + (1 | Subject) + storage_time
y ~ current_feature + Sex + (1 | Subject) + storage_time
y ~ current_feature + Sex + (1 | Subject) + storage_time
y ~ current_feature + Sex + (1 | Subject) + storage_time
y ~ current_feature + Sex + (1 | Subject) + storage_time
y ~ current_feature + Sex + (1 | Subject) + storage_time
y ~ current_feature + Sex + (1 | Subject) + storage_time
y ~ current_feature + Sex + (1 | Subject) + storage_time
y ~ current_feature + Sex + (1 | Subject) + storage_time
y ~ current_feature + Sex + (1 | Subject) + storage_time
y ~ current_feature + Sex + (1 | Subject) + storage_time
y ~ current_feature + Sex + (1 | Subject) + storage_time
y ~ current_feature + Sex + (1 | Subject) + storage_time
y ~ current_feature + Sex + (1 | Subject) + storage_time
y ~ current_feature + Sex + (1 | Subject) + storage_time
y ~ current_feature + Sex + (1 | Subject) + storage_time
y ~ current_feature + Sex + (1 | Subject) + storage_time
y ~ current_feature + Sex + (1 | Subject) + storage_time
y ~ current_feature + Sex + (1 | Subject) + storage_time
y ~ current_feature + Sex + (1 | Subject) + storage_time
y ~ current_feature + Sex + (1 | Subject) + storage_time
y ~ current_feature + Sex + (1 | Subject) + storage_time
y ~ current_feature + Sex + (1 | Subject) + storage_time
y ~ current_feature + Sex + (1 | Subject) + storage_time
y ~ current_feature + Sex + (1 | Subject) + storage_time
y ~ current_feature + Sex + (1 | Subject) + storage_time
y ~ current_feature + Sex + (1 | Subject) + storage_time
y ~ current_feature + Sex + (1 | Subject) + storage_time
y ~ current_feature + Sex + (1 | Subject) + storage_time
y ~ current_feature + Sex + (1 | Subject) + storage_time
y ~ current_feature + Sex + (1 | Subject) + storage_time
y ~ current_feature + Sex + (1 | Subject) + storage_time
y ~ current_feature + Sex + (1 | Subject) + storage_time
y ~ current_feature + Sex + (1 | Subject) + storage_time
y ~ current_feature + Sex + (1 | Subject) + storage_time
y ~ current_feature + Sex + (1 | Subject) + storage_time
y ~ current_feature + Sex + (1 | Subject) + storage_time
y ~ current_feature + Sex + (1 | Subject) + storage_time
y ~ current_feature + Sex + (1 | Subject) + storage_time
y ~ current_feature + Sex + (1 | Subject) + storage_time
y ~ current_feature + Sex + (1 | Subject) + storage_time
y ~ current_feature + Sex + (1 | Subject) + storage_time
y ~ current_feature + Sex + (1 | Subject) + storage_time
y ~ current_feature + Sex + (1 | Subject) + storage_time
```

y ~ current_feature + Sex + (1 | Subject) + storage_time
y ~ current_feature + Sex + (1 | Subject) + storage_time
y ~ current_feature + Sex + (1 | Subject) + storage_time
y ~ current_feature + Sex + (1 | Subject) + storage_time
y ~ current_feature + Sex + (1 | Subject) + storage_time
y ~ current_feature + Sex + (1 | Subject) + storage_time
y ~ current_feature + Sex + (1 | Subject) + storage_time
y ~ current_feature + Sex + (1 | Subject) + storage_time
y ~ current_feature + Sex + (1 | Subject) + storage_time
y ~ current_feature + Sex + (1 | Subject) + storage_time
y ~ current_feature + Sex + (1 | Subject) + storage_time
y ~ current_feature + Sex + (1 | Subject) + storage_time
y ~ current_feature + Sex + (1 | Subject) + storage_time
y ~ current_feature + Sex + (1 | Subject) + storage_time
y ~ current_feature + Sex + (1 | Subject) + storage_time
y ~ current_feature + Sex + (1 | Subject) + storage_time
y ~ current_feature + Sex + (1 | Subject) + storage_time
y ~ current_feature + Sex + (1 | Subject) + storage_time
y ~ current_feature + Sex + (1 | Subject) + storage_time
y ~ current_feature + Sex + (1 | Subject) + storage_time
y ~ current_feature + Sex + (1 | Subject) + storage_time
y ~ current_feature + Sex + (1 | Subject) + storage_time
y ~ current_feature + Sex + (1 | Subject) + storage_time
y ~ current_feature + Sex + (1 | Subject) + storage_time
y ~ current_feature + Sex + (1 | Subject) + storage_time
y ~ current_feature + Sex + (1 | Subject) + storage_time
y ~ current_feature + Sex + (1 | Subject) + storage_time
y ~ current_feature + Sex + (1 | Subject) + storage_time
y ~ current_feature + Sex + (1 | Subject) + storage_time
y ~ current_feature + Sex + (1 | Subject) + storage_time
y ~ current_feature + Sex + (1 | Subject) + storage_time
y ~ current_feature + Sex + (1 | Subject) + storage_time
y ~ current_feature + Sex + (1 | Subject) + storage_time
y ~ current_feature + Sex + (1 | Subject) + storage_time
y ~ current_feature + Sex + (1 | Subject) + storage_time
y ~ current_feature + Sex + (1 | Subject) + storage_time
y ~ current_feature + Sex + (1 | Subject) + storage_time
y ~ current_feature + Sex + (1 | Subject) + storage_time
y ~ current_feature + Sex + (1 | Subject) + storage_time
y ~ current_feature + Sex + (1 | Subject) + storage_time
y ~ current_feature + Sex + (1 | Subject) + storage_time
y ~ current_feature + Sex + (1 | Subject) + storage_time
y ~ current_feature + Sex + (1 | Subject) + storage_time
y ~ current_feature + Sex + (1 | Subject) + storage_time
y ~ current_feature + Sex + (1 | Subject) + storage_time
y ~ current_feature + Sex + (1 | Subject) + storage_time
y ~ current_feature + Sex + (1 | Subject) + storage_time
y ~ current_feature + Sex + (1 | Subject) + storage_time

y ~ current_feature + Sex + (1 | Subject) + storage_time
y ~ current_feature + Sex + (1 | Subject) + storage_time
y ~ current_feature + Sex + (1 | Subject) + storage_time
y ~ current_feature + Sex + (1 | Subject) + storage_time
y ~ current_feature + Sex + (1 | Subject) + storage_time
y ~ current_feature + Sex + (1 | Subject) + storage_time
y ~ current_feature + Sex + (1 | Subject) + storage_time
y ~ current_feature + Sex + (1 | Subject) + storage_time
y ~ current_feature + Sex + (1 | Subject) + storage_time
y ~ current_feature + Sex + (1 | Subject) + storage_time
y ~ current_feature + Sex + (1 | Subject) + storage_time
y ~ current_feature + Sex + (1 | Subject) + storage_time
y ~ current_feature + Sex + (1 | Subject) + storage_time
y ~ current_feature + Sex + (1 | Subject) + storage_time
y ~ current_feature + Sex + (1 | Subject) + storage_time
y ~ current_feature + Sex + (1 | Subject) + storage_time
y ~ current_feature + Sex + (1 | Subject) + storage_time
y ~ current_feature + Sex + (1 | Subject) + storage_time
y ~ current_feature + Sex + (1 | Subject) + storage_time
y ~ current_feature + Sex + (1 | Subject) + storage_time
y ~ current_feature + Sex + (1 | Subject) + storage_time
y ~ current_feature + Sex + (1 | Subject) + storage_time
y ~ current_feature + Sex + (1 | Subject) + storage_time
y ~ current_feature + Sex + (1 | Subject) + storage_time
y ~ current_feature + Sex + (1 | Subject) + storage_time
y ~ current_feature + Sex + (1 | Subject) + storage_time
y ~ current_feature + Sex + (1 | Subject) + storage_time
y ~ current_feature + Sex + (1 | Subject) + storage_time
y ~ current_feature + Sex + (1 | Subject) + storage_time
y ~ current_feature + Sex + (1 | Subject) + storage_time
y ~ current_feature + Sex + (1 | Subject) + storage_time
y ~ current_feature + Sex + (1 | Subject) + storage_time
y ~ current_feature + Sex + (1 | Subject) + storage_time
y ~ current_feature + Sex + (1 | Subject) + storage_time
y ~ current_feature + Sex + (1 | Subject) + storage_time
y ~ current_feature + Sex + (1 | Subject) + storage_time
y ~ current_feature + Sex + (1 | Subject) + storage_time
y ~ current_feature + Sex + (1 | Subject) + storage_time
y ~ current_feature + Sex + (1 | Subject) + storage_time
y ~ current_feature + Sex + (1 | Subject) + storage_time
y ~ current_feature + Sex + (1 | Subject) + storage_time
y ~ current_feature + Sex + (1 | Subject) + storage_time
y ~ current_feature + Sex + (1 | Subject) + storage_time
y ~ current_feature + Sex + (1 | Subject) + storage_time
y ~ current_feature + Sex + (1 | Subject) + storage_time
y ~ current_feature + Sex + (1 | Subject) + storage_time
y ~ current_feature + Sex + (1 | Subject) + storage_time
y ~ current_feature + Sex + (1 | Subject) + storage_time

y ~ current_feature + Sex + (1 | Subject) + storage_time
y ~ current_feature + Sex + (1 | Subject) + storage_time
y ~ current_feature + Sex + (1 | Subject) + storage_time
y ~ current_feature + Sex + (1 | Subject) + storage_time
y ~ current_feature + Sex + (1 | Subject) + storage_time
y ~ current_feature + Sex + (1 | Subject) + storage_time
y ~ current_feature + Sex + (1 | Subject) + storage_time
y ~ current_feature + Sex + (1 | Subject) + storage_time
y ~ current_feature + Sex + (1 | Subject) + storage_time
y ~ current_feature + Sex + (1 | Subject) + storage_time
y ~ current_feature + Sex + (1 | Subject) + storage_time
y ~ current_feature + Sex + (1 | Subject) + storage_time
y ~ current_feature + Sex + (1 | Subject) + storage_time
y ~ current_feature + Sex + (1 | Subject) + storage_time
y ~ current_feature + Sex + (1 | Subject) + storage_time
y ~ current_feature + Sex + (1 | Subject) + storage_time
y ~ current_feature + Sex + (1 | Subject) + storage_time
y ~ current_feature + Sex + (1 | Subject) + storage_time
y ~ current_feature + Sex + (1 | Subject) + storage_time
y ~ current_feature + Sex + (1 | Subject) + storage_time
y ~ current_feature + Sex + (1 | Subject) + storage_time
y ~ current_feature + Sex + (1 | Subject) + storage_time
y ~ current_feature + Sex + (1 | Subject) + storage_time
y ~ current_feature + Sex + (1 | Subject) + storage_time
y ~ current_feature + Sex + (1 | Subject) + storage_time
y ~ current_feature + Sex + (1 | Subject) + storage_time
y ~ current_feature + Sex + (1 | Subject) + storage_time
y ~ current_feature + Sex + (1 | Subject) + storage_time
y ~ current_feature + Sex + (1 | Subject) + storage_time
y ~ current_feature + Sex + (1 | Subject) + storage_time
y ~ current_feature + Sex + (1 | Subject) + storage_time
y ~ current_feature + Sex + (1 | Subject) + storage_time
y ~ current_feature + Sex + (1 | Subject) + storage_time
y ~ current_feature + Sex + (1 | Subject) + storage_time
y ~ current_feature + Sex + (1 | Subject) + storage_time
y ~ current_feature + Sex + (1 | Subject) + storage_time
y ~ current_feature + Sex + (1 | Subject) + storage_time
y ~ current_feature + Sex + (1 | Subject) + storage_time
y ~ current_feature + Sex + (1 | Subject) + storage_time
y ~ current_feature + Sex + (1 | Subject) + storage_time
y ~ current_feature + Sex + (1 | Subject) + storage_time
y ~ current_feature + Sex + (1 | Subject) + storage_time
y ~ current_feature + Sex + (1 | Subject) + storage_time
y ~ current_feature + Sex + (1 | Subject) + storage_time
y ~ current_feature + Sex + (1 | Subject) + storage_time
y ~ current_feature + Sex + (1 | Subject) + storage_time
y ~ current_feature + Sex + (1 | Subject) + storage_time
y ~ current_feature + Sex + (1 | Subject) + storage_time

y ~ current_feature + Sex + (1 | Subject) + storage_time
y ~ current_feature + Sex + (1 | Subject) + storage_time
y ~ current_feature + Sex + (1 | Subject) + storage_time
y ~ current_feature + Sex + (1 | Subject) + storage_time
y ~ current_feature + Sex + (1 | Subject) + storage_time
y ~ current_feature + Sex + (1 | Subject) + storage_time
y ~ current_feature + Sex + (1 | Subject) + storage_time
y ~ current_feature + Sex + (1 | Subject) + storage_time
y ~ current_feature + Sex + (1 | Subject) + storage_time
y ~ current_feature + Sex + (1 | Subject) + storage_time
y ~ current_feature + Sex + (1 | Subject) + storage_time
y ~ current_feature + Sex + (1 | Subject) + storage_time
y ~ current_feature + Sex + (1 | Subject) + storage_time
y ~ current_feature + Sex + (1 | Subject) + storage_time
y ~ current_feature + Sex + (1 | Subject) + storage_time
y ~ current_feature + Sex + (1 | Subject) + storage_time
y ~ current_feature + Sex + (1 | Subject) + storage_time
y ~ current_feature + Sex + (1 | Subject) + storage_time
y ~ current_feature + Sex + (1 | Subject) + storage_time
y ~ current_feature + Sex + (1 | Subject) + storage_time
y ~ current_feature + Sex + (1 | Subject) + storage_time
y ~ current_feature + Sex + (1 | Subject) + storage_time
y ~ current_feature + Sex + (1 | Subject) + storage_time
y ~ current_feature + Sex + (1 | Subject) + storage_time
y ~ current_feature + Sex + (1 | Subject) + storage_time
y ~ current_feature + Sex + (1 | Subject) + storage_time
y ~ current_feature + Sex + (1 | Subject) + storage_time
y ~ current_feature + Sex + (1 | Subject) + storage_time
y ~ current_feature + Sex + (1 | Subject) + storage_time
y ~ current_feature + Sex + (1 | Subject) + storage_time
y ~ current_feature + Sex + (1 | Subject) + storage_time
y ~ current_feature + Sex + (1 | Subject) + storage_time
y ~ current_feature + Sex + (1 | Subject) + storage_time
y ~ current_feature + Sex + (1 | Subject) + storage_time
y ~ current_feature + Sex + (1 | Subject) + storage_time
y ~ current_feature + Sex + (1 | Subject) + storage_time
y ~ current_feature + Sex + (1 | Subject) + storage_time
y ~ current_feature + Sex + (1 | Subject) + storage_time
y ~ current_feature + Sex + (1 | Subject) + storage_time
y ~ current_feature + Sex + (1 | Subject) + storage_time
y ~ current_feature + Sex + (1 | Subject) + storage_time
y ~ current_feature + Sex + (1 | Subject) + storage_time
y ~ current_feature + Sex + (1 | Subject) + storage_time
y ~ current_feature + Sex + (1 | Subject) + storage_time
y ~ current_feature + Sex + (1 | Subject) + storage_time
y ~ current_feature + Sex + (1 | Subject) + storage_time
y ~ current_feature + Sex + (1 | Subject) + storage_time
y ~ current_feature + Sex + (1 | Subject) + storage_time

y ~ current_feature + Sex + (1 | Subject) + storage_time
y ~ current_feature + Sex + (1 | Subject) + storage_time
y ~ current_feature + Sex + (1 | Subject) + storage_time
y ~ current_feature + Sex + (1 | Subject) + storage_time
y ~ current_feature + Sex + (1 | Subject) + storage_time
y ~ current_feature + Sex + (1 | Subject) + storage_time
y ~ current_feature + Sex + (1 | Subject) + storage_time
y ~ current_feature + Sex + (1 | Subject) + storage_time
y ~ current_feature + Sex + (1 | Subject) + storage_time
y ~ current_feature + Sex + (1 | Subject) + storage_time
y ~ current_feature + Sex + (1 | Subject) + storage_time
y ~ current_feature + Sex + (1 | Subject) + storage_time
y ~ current_feature + Sex + (1 | Subject) + storage_time
y ~ current_feature + Sex + (1 | Subject) + storage_time
y ~ current_feature + Sex + (1 | Subject) + storage_time
y ~ current_feature + Sex + (1 | Subject) + storage_time
y ~ current_feature + Sex + (1 | Subject) + storage_time
y ~ current_feature + Sex + (1 | Subject) + storage_time
y ~ current_feature + Sex + (1 | Subject) + storage_time
y ~ current_feature + Sex + (1 | Subject) + storage_time
y ~ current_feature + Sex + (1 | Subject) + storage_time
y ~ current_feature + Sex + (1 | Subject) + storage_time
y ~ current_feature + Sex + (1 | Subject) + storage_time
y ~ current_feature + Sex + (1 | Subject) + storage_time
y ~ current_feature + Sex + (1 | Subject) + storage_time
y ~ current_feature + Sex + (1 | Subject) + storage_time
y ~ current_feature + Sex + (1 | Subject) + storage_time
y ~ current_feature + Sex + (1 | Subject) + storage_time
y ~ current_feature + Sex + (1 | Subject) + storage_time
y ~ current_feature + Sex + (1 | Subject) + storage_time
y ~ current_feature + Sex + (1 | Subject) + storage_time
y ~ current_feature + Sex + (1 | Subject) + storage_time
y ~ current_feature + Sex + (1 | Subject) + storage_time
y ~ current_feature + Sex + (1 | Subject) + storage_time
y ~ current_feature + Sex + (1 | Subject) + storage_time
y ~ current_feature + Sex + (1 | Subject) + storage_time
y ~ current_feature + Sex + (1 | Subject) + storage_time
y ~ current_feature + Sex + (1 | Subject) + storage_time
y ~ current_feature + Sex + (1 | Subject) + storage_time
y ~ current_feature + Sex + (1 | Subject) + storage_time
y ~ current_feature + Sex + (1 | Subject) + storage_time
y ~ current_feature + Sex + (1 | Subject) + storage_time
y ~ current_feature + Sex + (1 | Subject) + storage_time
y ~ current_feature + Sex + (1 | Subject) + storage_time
y ~ current_feature + Sex + (1 | Subject) + storage_time
y ~ current_feature + Sex + (1 | Subject) + storage_time
y ~ current_feature + Sex + (1 | Subject) + storage_time
y ~ current_feature + Sex + (1 | Subject) + storage_time

y ~ current_feature + Sex + (1 | Subject) + storage_time
y ~ current_feature + Sex + (1 | Subject) + storage_time
y ~ current_feature + Sex + (1 | Subject) + storage_time
y ~ current_feature + Sex + (1 | Subject) + storage_time
y ~ current_feature + Sex + (1 | Subject) + storage_time
y ~ current_feature + Sex + (1 | Subject) + storage_time
y ~ current_feature + Sex + (1 | Subject) + storage_time
y ~ current_feature + Sex + (1 | Subject) + storage_time
y ~ current_feature + Sex + (1 | Subject) + storage_time
y ~ current_feature + Sex + (1 | Subject) + storage_time
y ~ current_feature + Sex + (1 | Subject) + storage_time
y ~ current_feature + Sex + (1 | Subject) + storage_time
y ~ current_feature + Sex + (1 | Subject) + storage_time
y ~ current_feature + Sex + (1 | Subject) + storage_time
y ~ current_feature + Sex + (1 | Subject) + storage_time
y ~ current_feature + Sex + (1 | Subject) + storage_time
y ~ current_feature + Sex + (1 | Subject) + storage_time
y ~ current_feature + Sex + (1 | Subject) + storage_time
y ~ current_feature + Sex + (1 | Subject) + storage_time
y ~ current_feature + Sex + (1 | Subject) + storage_time
y ~ current_feature + Sex + (1 | Subject) + storage_time
y ~ current_feature + Sex + (1 | Subject) + storage_time
y ~ current_feature + Sex + (1 | Subject) + storage_time
y ~ current_feature + Sex + (1 | Subject) + storage_time
y ~ current_feature + Sex + (1 | Subject) + storage_time
y ~ current_feature + Sex + (1 | Subject) + storage_time
y ~ current_feature + Sex + (1 | Subject) + storage_time
y ~ current_feature + Sex + (1 | Subject) + storage_time
y ~ current_feature + Sex + (1 | Subject) + storage_time
y ~ current_feature + Sex + (1 | Subject) + storage_time
y ~ current_feature + Sex + (1 | Subject) + storage_time
y ~ current_feature + Sex + (1 | Subject) + storage_time
y ~ current_feature + Sex + (1 | Subject) + storage_time
y ~ current_feature + Sex + (1 | Subject) + storage_time
y ~ current_feature + Sex + (1 | Subject) + storage_time
y ~ current_feature + Sex + (1 | Subject) + storage_time
y ~ current_feature + Sex + (1 | Subject) + storage_time
y ~ current_feature + Sex + (1 | Subject) + storage_time
y ~ current_feature + Sex + (1 | Subject) + storage_time
y ~ current_feature + Sex + (1 | Subject) + storage_time
y ~ current_feature + Sex + (1 | Subject) + storage_time
y ~ current_feature + Sex + (1 | Subject) + storage_time
y ~ current_feature + Sex + (1 | Subject) + storage_time
y ~ current_feature + Sex + (1 | Subject) + storage_time
y ~ current_feature + Sex + (1 | Subject) + storage_time
y ~ current_feature + Sex + (1 | Subject) + storage_time
y ~ current_feature + Sex + (1 | Subject) + storage_time
y ~ current_feature + Sex + (1 | Subject) + storage_time

y ~ current_feature + Sex + (1 | Subject) + storage_time
y ~ current_feature + Sex + (1 | Subject) + storage_time
y ~ current_feature + Sex + (1 | Subject) + storage_time
y ~ current_feature + Sex + (1 | Subject) + storage_time
y ~ current_feature + Sex + (1 | Subject) + storage_time
y ~ current_feature + Sex + (1 | Subject) + storage_time
y ~ current_feature + Sex + (1 | Subject) + storage_time
y ~ current_feature + Sex + (1 | Subject) + storage_time
y ~ current_feature + Sex + (1 | Subject) + storage_time
y ~ current_feature + Sex + (1 | Subject) + storage_time
y ~ current_feature + Sex + (1 | Subject) + storage_time
y ~ current_feature + Sex + (1 | Subject) + storage_time
y ~ current_feature + Sex + (1 | Subject) + storage_time
y ~ current_feature + Sex + (1 | Subject) + storage_time
y ~ current_feature + Sex + (1 | Subject) + storage_time
y ~ current_feature + Sex + (1 | Subject) + storage_time
y ~ current_feature + Sex + (1 | Subject) + storage_time
y ~ current_feature + Sex + (1 | Subject) + storage_time
y ~ current_feature + Sex + (1 | Subject) + storage_time
y ~ current_feature + Sex + (1 | Subject) + storage_time
y ~ current_feature + Sex + (1 | Subject) + storage_time
y ~ current_feature + Sex + (1 | Subject) + storage_time
y ~ current_feature + Sex + (1 | Subject) + storage_time
y ~ current_feature + Sex + (1 | Subject) + storage_time
y ~ current_feature + Sex + (1 | Subject) + storage_time
y ~ current_feature + Sex + (1 | Subject) + storage_time
y ~ current_feature + Sex + (1 | Subject) + storage_time
y ~ current_feature + Sex + (1 | Subject) + storage_time
y ~ current_feature + Sex + (1 | Subject) + storage_time
y ~ current_feature + Sex + (1 | Subject) + storage_time
y ~ current_feature + Sex + (1 | Subject) + storage_time
y ~ current_feature + Sex + (1 | Subject) + storage_time
y ~ current_feature + Sex + (1 | Subject) + storage_time
y ~ current_feature + Sex + (1 | Subject) + storage_time
y ~ current_feature + Sex + (1 | Subject) + storage_time
y ~ current_feature + Sex + (1 | Subject) + storage_time
y ~ current_feature + Sex + (1 | Subject) + storage_time
y ~ current_feature + Sex + (1 | Subject) + storage_time
y ~ current_feature + Sex + (1 | Subject) + storage_time
y ~ current_feature + Sex + (1 | Subject) + storage_time
y ~ current_feature + Sex + (1 | Subject) + storage_time
y ~ current_feature + Sex + (1 | Subject) + storage_time
y ~ current_feature + Sex + (1 | Subject) + storage_time
y ~ current_feature + Sex + (1 | Subject) + storage_time
y ~ current_feature + Sex + (1 | Subject) + storage_time
y ~ current_feature + Sex + (1 | Subject) + storage_time
y ~ current_feature + Sex + (1 | Subject) + storage_time
y ~ current_feature + Sex + (1 | Subject) + storage_time

y ~ current_feature + Sex + (1 | Subject) + storage_time
y ~ current_feature + Sex + (1 | Subject) + storage_time
y ~ current_feature + Sex + (1 | Subject) + storage_time
y ~ current_feature + Sex + (1 | Subject) + storage_time
y ~ current_feature + Sex + (1 | Subject) + storage_time
y ~ current_feature + Sex + (1 | Subject) + storage_time
y ~ current_feature + Sex + (1 | Subject) + storage_time
y ~ current_feature + Sex + (1 | Subject) + storage_time
y ~ current_feature + Sex + (1 | Subject) + storage_time
y ~ current_feature + Sex + (1 | Subject) + storage_time
y ~ current_feature + Sex + (1 | Subject) + storage_time
y ~ current_feature + Sex + (1 | Subject) + storage_time
y ~ current_feature + Sex + (1 | Subject) + storage_time
y ~ current_feature + Sex + (1 | Subject) + storage_time
y ~ current_feature + Sex + (1 | Subject) + storage_time
y ~ current_feature + Sex + (1 | Subject) + storage_time
y ~ current_feature + Sex + (1 | Subject) + storage_time
y ~ current_feature + Sex + (1 | Subject) + storage_time
y ~ current_feature + Sex + (1 | Subject) + storage_time
y ~ current_feature + Sex + (1 | Subject) + storage_time
y ~ current_feature + Sex + (1 | Subject) + storage_time
y ~ current_feature + Sex + (1 | Subject) + storage_time
y ~ current_feature + Sex + (1 | Subject) + storage_time
y ~ current_feature + Sex + (1 | Subject) + storage_time
y ~ current_feature + Sex + (1 | Subject) + storage_time
y ~ current_feature + Sex + (1 | Subject) + storage_time
y ~ current_feature + Sex + (1 | Subject) + storage_time
y ~ current_feature + Sex + (1 | Subject) + storage_time
y ~ current_feature + Sex + (1 | Subject) + storage_time
y ~ current_feature + Sex + (1 | Subject) + storage_time
y ~ current_feature + Sex + (1 | Subject) + storage_time
y ~ current_feature + Sex + (1 | Subject) + storage_time
y ~ current_feature + Sex + (1 | Subject) + storage_time
y ~ current_feature + Sex + (1 | Subject) + storage_time
y ~ current_feature + Sex + (1 | Subject) + storage_time
y ~ current_feature + Sex + (1 | Subject) + storage_time
y ~ current_feature + Sex + (1 | Subject) + storage_time
y ~ current_feature + Sex + (1 | Subject) + storage_time
y ~ current_feature + Sex + (1 | Subject) + storage_time
y ~ current_feature + Sex + (1 | Subject) + storage_time
y ~ current_feature + Sex + (1 | Subject) + storage_time
y ~ current_feature + Sex + (1 | Subject) + storage_time
y ~ current_feature + Sex + (1 | Subject) + storage_time
y ~ current_feature + Sex + (1 | Subject) + storage_time
y ~ current_feature + Sex + (1 | Subject) + storage_time
y ~ current_feature + Sex + (1 | Subject) + storage_time
y ~ current_feature + Sex + (1 | Subject) + storage_time
y ~ current_feature + Sex + (1 | Subject) + storage_time

y ~ current_feature + Sex + (1 | Subject) + storage_time
y ~ current_feature + Sex + (1 | Subject) + storage_time
y ~ current_feature + Sex + (1 | Subject) + storage_time
y ~ current_feature + Sex + (1 | Subject) + storage_time
y ~ current_feature + Sex + (1 | Subject) + storage_time
y ~ current_feature + Sex + (1 | Subject) + storage_time
y ~ current_feature + Sex + (1 | Subject) + storage_time
y ~ current_feature + Sex + (1 | Subject) + storage_time
y ~ current_feature + Sex + (1 | Subject) + storage_time
y ~ current_feature + Sex + (1 | Subject) + storage_time
y ~ current_feature + Sex + (1 | Subject) + storage_time
y ~ current_feature + Sex + (1 | Subject) + storage_time
y ~ current_feature + Sex + (1 | Subject) + storage_time
y ~ current_feature + Sex + (1 | Subject) + storage_time
y ~ current_feature + Sex + (1 | Subject) + storage_time
y ~ current_feature + Sex + (1 | Subject) + storage_time
y ~ current_feature + Sex + (1 | Subject) + storage_time
y ~ current_feature + Sex + (1 | Subject) + storage_time
y ~ current_feature + Sex + (1 | Subject) + storage_time
y ~ current_feature + Sex + (1 | Subject) + storage_time
y ~ current_feature + Sex + (1 | Subject) + storage_time
y ~ current_feature + Sex + (1 | Subject) + storage_time
y ~ current_feature + Sex + (1 | Subject) + storage_time
y ~ current_feature + Sex + (1 | Subject) + storage_time
y ~ current_feature + Sex + (1 | Subject) + storage_time
y ~ current_feature + Sex + (1 | Subject) + storage_time
y ~ current_feature + Sex + (1 | Subject) + storage_time
y ~ current_feature + Sex + (1 | Subject) + storage_time
y ~ current_feature + Sex + (1 | Subject) + storage_time
y ~ current_feature + Sex + (1 | Subject) + storage_time
y ~ current_feature + Sex + (1 | Subject) + storage_time
y ~ current_feature + Sex + (1 | Subject) + storage_time
y ~ current_feature + Sex + (1 | Subject) + storage_time
y ~ current_feature + Sex + (1 | Subject) + storage_time
y ~ current_feature + Sex + (1 | Subject) + storage_time
y ~ current_feature + Sex + (1 | Subject) + storage_time
y ~ current_feature + Sex + (1 | Subject) + storage_time
y ~ current_feature + Sex + (1 | Subject) + storage_time
y ~ current_feature + Sex + (1 | Subject) + storage_time
y ~ current_feature + Sex + (1 | Subject) + storage_time
y ~ current_feature + Sex + (1 | Subject) + storage_time
y ~ current_feature + Sex + (1 | Subject) + storage_time
y ~ current_feature + Sex + (1 | Subject) + storage_time
y ~ current_feature + Sex + (1 | Subject) + storage_time
y ~ current_feature + Sex + (1 | Subject) + storage_time
y ~ current_feature + Sex + (1 | Subject) + storage_time
y ~ current_feature + Sex + (1 | Subject) + storage_time
y ~ current_feature + Sex + (1 | Subject) + storage_time

y ~ current_feature + Sex + (1 | Subject) + storage_time
y ~ current_feature + Sex + (1 | Subject) + storage_time
y ~ current_feature + Sex + (1 | Subject) + storage_time
y ~ current_feature + Sex + (1 | Subject) + storage_time
y ~ current_feature + Sex + (1 | Subject) + storage_time
y ~ current_feature + Sex + (1 | Subject) + storage_time
y ~ current_feature + Sex + (1 | Subject) + storage_time
y ~ current_feature + Sex + (1 | Subject) + storage_time
y ~ current_feature + Sex + (1 | Subject) + storage_time
y ~ current_feature + Sex + (1 | Subject) + storage_time
y ~ current_feature + Sex + (1 | Subject) + storage_time
y ~ current_feature + Sex + (1 | Subject) + storage_time
y ~ current_feature + Sex + (1 | Subject) + storage_time
y ~ current_feature + Sex + (1 | Subject) + storage_time
y ~ current_feature + Sex + (1 | Subject) + storage_time
y ~ current_feature + Sex + (1 | Subject) + storage_time
y ~ current_feature + Sex + (1 | Subject) + storage_time
y ~ current_feature + Sex + (1 | Subject) + storage_time
y ~ current_feature + Sex + (1 | Subject) + storage_time
y ~ current_feature + Sex + (1 | Subject) + storage_time
y ~ current_feature + Sex + (1 | Subject) + storage_time
y ~ current_feature + Sex + (1 | Subject) + storage_time
y ~ current_feature + Sex + (1 | Subject) + storage_time
y ~ current_feature + Sex + (1 | Subject) + storage_time
y ~ current_feature + Sex + (1 | Subject) + storage_time
y ~ current_feature + Sex + (1 | Subject) + storage_time
y ~ current_feature + Sex + (1 | Subject) + storage_time
y ~ current_feature + Sex + (1 | Subject) + storage_time
y ~ current_feature + Sex + (1 | Subject) + storage_time
y ~ current_feature + Sex + (1 | Subject) + storage_time
y ~ current_feature + Sex + (1 | Subject) + storage_time
y ~ current_feature + Sex + (1 | Subject) + storage_time
y ~ current_feature + Sex + (1 | Subject) + storage_time
y ~ current_feature + Sex + (1 | Subject) + storage_time
y ~ current_feature + Sex + (1 | Subject) + storage_time
y ~ current_feature + Sex + (1 | Subject) + storage_time
y ~ current_feature + Sex + (1 | Subject) + storage_time
y ~ current_feature + Sex + (1 | Subject) + storage_time
y ~ current_feature + Sex + (1 | Subject) + storage_time
y ~ current_feature + Sex + (1 | Subject) + storage_time
y ~ current_feature + Sex + (1 | Subject) + storage_time
y ~ current_feature + Sex + (1 | Subject) + storage_time
y ~ current_feature + Sex + (1 | Subject) + storage_time
y ~ current_feature + Sex + (1 | Subject) + storage_time
y ~ current_feature + Sex + (1 | Subject) + storage_time
y ~ current_feature + Sex + (1 | Subject) + storage_time
y ~ current_feature + Sex + (1 | Subject) + storage_time
y ~ current_feature + Sex + (1 | Subject) + storage_time

```
y ~ current_feature + Sex + (1 | Subject) + storage_time
y ~ current_feature + Sex + (1 | Subject) + storage_time
y ~ current_feature + Sex + (1 | Subject) + storage_time
y ~ current_feature + Sex + (1 | Subject) + storage_time
y ~ current_feature + Sex + (1 | Subject) + storage_time
y ~ current_feature + Sex + (1 | Subject) + storage_time
y ~ current_feature + Sex + (1 | Subject) + storage_time
y ~ current_feature + Sex + (1 | Subject) + storage_time
y ~ current_feature + Sex + (1 | Subject) + storage_time
y ~ current_feature + Sex + (1 | Subject) + storage_time
y ~ current_feature + Sex + (1 | Subject) + storage_time
y ~ current_feature + Sex + (1 | Subject) + storage_time
y ~ current_feature + Sex + (1 | Subject) + storage_time
y ~ current_feature + Sex + (1 | Subject) + storage_time
y ~ current_feature + Sex + (1 | Subject) + storage_time
y ~ current_feature + Sex + (1 | Subject) + storage_time
y ~ current_feature + Sex + (1 | Subject) + storage_time
y ~ current_feature + Sex + (1 | Subject) + storage_time
y ~ current_feature + Sex + (1 | Subject) + storage_time
y ~ current_feature + Sex + (1 | Subject) + storage_time
y ~ current_feature + Sex + (1 | Subject) + storage_time
y ~ current_feature + Sex + (1 | Subject) + storage_time
y ~ current_feature + Sex + (1 | Subject) + storage_time
y ~ current_feature + Sex + (1 | Subject) + storage_time
y ~ current_feature + Sex + (1 | Subject) + storage_time
y ~ current_feature + Sex + (1 | Subject) + storage_time
y ~ current_feature + Sex + (1 | Subject) + storage_time
y ~ current_feature + Sex + (1 | Subject) + storage_time
y ~ current_feature + Sex + (1 | Subject) + storage_time
y ~ current_feature + Sex + (1 | Subject) + storage_time
y ~ current_feature + Sex + (1 | Subject) + storage_time
y ~ current_feature + Sex + (1 | Subject) + storage_time
y ~ current_feature + Sex + (1 | Subject) + storage_time
y ~ current_feature + Sex + (1 | Subject) + storage_time
y ~ current_feature + Sex + (1 | Subject) + storage_time
y ~ current_feature + Sex + (1 | Subject) + storage_time
y ~ current_feature + Sex + (1 | Subject) + storage_time
y ~ current_feature + Sex + (1 | Subject) + storage_time
y ~ current_feature + Sex + (1 | Subject) + storage_time
y ~ current_feature + Sex + (1 | Subject) + storage_time
y ~ current_feature + Sex + (1 | Subject) + storage_time
y ~ current_feature + Sex + (1 | Subject) + storage_time
y ~ current_feature + Sex + (1 | Subject) + storage_time
y ~ current_feature + Sex + (1 | Subject) + storage_time
y ~ current_feature + Sex + (1 | Subject) + storage_time
y ~ current_feature + Sex + (1 | Subject) + storage_time
y ~ current_feature + Sex + (1 | Subject) + storage_time
y ~ current_feature + Sex + (1 | Subject) + storage_time
```

y ~ current_feature + Sex + (1 | Subject) + storage_time
y ~ current_feature + Sex + (1 | Subject) + storage_time
y ~ current_feature + Sex + (1 | Subject) + storage_time
y ~ current_feature + Sex + (1 | Subject) + storage_time
y ~ current_feature + Sex + (1 | Subject) + storage_time
y ~ current_feature + Sex + (1 | Subject) + storage_time
y ~ current_feature + Sex + (1 | Subject) + storage_time
y ~ current_feature + Sex + (1 | Subject) + storage_time
y ~ current_feature + Sex + (1 | Subject) + storage_time
y ~ current_feature + Sex + (1 | Subject) + storage_time
y ~ current_feature + Sex + (1 | Subject) + storage_time
y ~ current_feature + Sex + (1 | Subject) + storage_time
y ~ current_feature + Sex + (1 | Subject) + storage_time
y ~ current_feature + Sex + (1 | Subject) + storage_time
y ~ current_feature + Sex + (1 | Subject) + storage_time
y ~ current_feature + Sex + (1 | Subject) + storage_time
y ~ current_feature + Sex + (1 | Subject) + storage_time
y ~ current_feature + Sex + (1 | Subject) + storage_time
y ~ current_feature + Sex + (1 | Subject) + storage_time
y ~ current_feature + Sex + (1 | Subject) + storage_time
y ~ current_feature + Sex + (1 | Subject) + storage_time
y ~ current_feature + Sex + (1 | Subject) + storage_time
y ~ current_feature + Sex + (1 | Subject) + storage_time
y ~ current_feature + Sex + (1 | Subject) + storage_time
y ~ current_feature + Sex + (1 | Subject) + storage_time
y ~ current_feature + Sex + (1 | Subject) + storage_time
y ~ current_feature + Sex + (1 | Subject) + storage_time
y ~ current_feature + Sex + (1 | Subject) + storage_time
y ~ current_feature + Sex + (1 | Subject) + storage_time
y ~ current_feature + Sex + (1 | Subject) + storage_time
y ~ current_feature + Sex + (1 | Subject) + storage_time
y ~ current_feature + Sex + (1 | Subject) + storage_time
y ~ current_feature + Sex + (1 | Subject) + storage_time
y ~ current_feature + Sex + (1 | Subject) + storage_time
y ~ current_feature + Sex + (1 | Subject) + storage_time
y ~ current_feature + Sex + (1 | Subject) + storage_time
y ~ current_feature + Sex + (1 | Subject) + storage_time
y ~ current_feature + Sex + (1 | Subject) + storage_time
y ~ current_feature + Sex + (1 | Subject) + storage_time
y ~ current_feature + Sex + (1 | Subject) + storage_time
y ~ current_feature + Sex + (1 | Subject) + storage_time
y ~ current_feature + Sex + (1 | Subject) + storage_time
y ~ current_feature + Sex + (1 | Subject) + storage_time
y ~ current_feature + Sex + (1 | Subject) + storage_time
y ~ current_feature + Sex + (1 | Subject) + storage_time
y ~ current_feature + Sex + (1 | Subject) + storage_time
y ~ current_feature + Sex + (1 | Subject) + storage_time
y ~ current_feature + Sex + (1 | Subject) + storage_time

y ~ current_feature + Sex + (1 | Subject) + storage_time
y ~ current_feature + Sex + (1 | Subject) + storage_time
y ~ current_feature + Sex + (1 | Subject) + storage_time
y ~ current_feature + Sex + (1 | Subject) + storage_time
y ~ current_feature + Sex + (1 | Subject) + storage_time
y ~ current_feature + Sex + (1 | Subject) + storage_time
y ~ current_feature + Sex + (1 | Subject) + storage_time
y ~ current_feature + Sex + (1 | Subject) + storage_time
y ~ current_feature + Sex + (1 | Subject) + storage_time
y ~ current_feature + Sex + (1 | Subject) + storage_time
y ~ current_feature + Sex + (1 | Subject) + storage_time
y ~ current_feature + Sex + (1 | Subject) + storage_time
y ~ current_feature + Sex + (1 | Subject) + storage_time
y ~ current_feature + Sex + (1 | Subject) + storage_time
y ~ current_feature + Sex + (1 | Subject) + storage_time
y ~ current_feature + Sex + (1 | Subject) + storage_time
y ~ current_feature + Sex + (1 | Subject) + storage_time
y ~ current_feature + Sex + (1 | Subject) + storage_time
y ~ current_feature + Sex + (1 | Subject) + storage_time
y ~ current_feature + Sex + (1 | Subject) + storage_time
y ~ current_feature + Sex + (1 | Subject) + storage_time
y ~ current_feature + Sex + (1 | Subject) + storage_time
y ~ current_feature + Sex + (1 | Subject) + storage_time
y ~ current_feature + Sex + (1 | Subject) + storage_time
y ~ current_feature + Sex + (1 | Subject) + storage_time
y ~ current_feature + Sex + (1 | Subject) + storage_time
y ~ current_feature + Sex + (1 | Subject) + storage_time
y ~ current_feature + Sex + (1 | Subject) + storage_time
y ~ current_feature + Sex + (1 | Subject) + storage_time
y ~ current_feature + Sex + (1 | Subject) + storage_time
y ~ current_feature + Sex + (1 | Subject) + storage_time
y ~ current_feature + Sex + (1 | Subject) + storage_time
y ~ current_feature + Sex + (1 | Subject) + storage_time
y ~ current_feature + Sex + (1 | Subject) + storage_time
y ~ current_feature + Sex + (1 | Subject) + storage_time
y ~ current_feature + Sex + (1 | Subject) + storage_time
y ~ current_feature + Sex + (1 | Subject) + storage_time
y ~ current_feature + Sex + (1 | Subject) + storage_time
y ~ current_feature + Sex + (1 | Subject) + storage_time
y ~ current_feature + Sex + (1 | Subject) + storage_time
y ~ current_feature + Sex + (1 | Subject) + storage_time
y ~ current_feature + Sex + (1 | Subject) + storage_time
y ~ current_feature + Sex + (1 | Subject) + storage_time
y ~ current_feature + Sex + (1 | Subject) + storage_time
y ~ current_feature + Sex + (1 | Subject) + storage_time
y ~ current_feature + Sex + (1 | Subject) + storage_time
y ~ current_feature + Sex + (1 | Subject) + storage_time
y ~ current_feature + Sex + (1 | Subject) + storage_time

```
y ~ current_feature + Sex + (1 | Subject) + storage_time
y ~ current_feature + Sex + (1 | Subject) + storage_time
y ~ current_feature + Sex + (1 | Subject) + storage_time
y ~ current_feature + Sex + (1 | Subject) + storage_time
y ~ current_feature + Sex + (1 | Subject) + storage_time
y ~ current_feature + Sex + (1 | Subject) + storage_time
y ~ current_feature + Sex + (1 | Subject) + storage_time
y ~ current_feature + Sex + (1 | Subject) + storage_time
y ~ current_feature + Sex + (1 | Subject) + storage_time
y ~ current_feature + Sex + (1 | Subject) + storage_time
y ~ current_feature + Sex + (1 | Subject) + storage_time
y ~ current_feature + Sex + (1 | Subject) + storage_time
y ~ current_feature + Sex + (1 | Subject) + storage_time
y ~ current_feature + Sex + (1 | Subject) + storage_time
y ~ current_feature + Sex + (1 | Subject) + storage_time
y ~ current_feature + Sex + (1 | Subject) + storage_time
y ~ current_feature + Sex + (1 | Subject) + storage_time
y ~ current_feature + Sex + (1 | Subject) + storage_time
y ~ current_feature + Sex + (1 | Subject) + storage_time
y ~ current_feature + Sex + (1 | Subject) + storage_time
y ~ current_feature + Sex + (1 | Subject) + storage_time
y ~ current_feature + Sex + (1 | Subject) + storage_time
y ~ current_feature + Sex + (1 | Subject) + storage_time
y ~ current_feature + Sex + (1 | Subject) + storage_time
y ~ current_feature + Sex + (1 | Subject) + storage_time
y ~ current_feature + Sex + (1 | Subject) + storage_time
y ~ current_feature + Sex + (1 | Subject) + storage_time
y ~ current_feature + Sex + (1 | Subject) + storage_time
y ~ current_feature + Sex + (1 | Subject) + storage_time
y ~ current_feature + Sex + (1 | Subject) + storage_time
y ~ current_feature + Sex + (1 | Subject) + storage_time
y ~ current_feature + Sex + (1 | Subject) + storage_time
y ~ current_feature + Sex + (1 | Subject) + storage_time
y ~ current_feature + Sex + (1 | Subject) + storage_time
y ~ current_feature + Sex + (1 | Subject) + storage_time
y ~ current_feature + Sex + (1 | Subject) + storage_time
y ~ current_feature + Sex + (1 | Subject) + storage_time
y ~ current_feature + Sex + (1 | Subject) + storage_time
y ~ current_feature + Sex + (1 | Subject) + storage_time
y ~ current_feature + Sex + (1 | Subject) + storage_time
y ~ current_feature + Sex + (1 | Subject) + storage_time
y ~ current_feature + Sex + (1 | Subject) + storage_time
y ~ current_feature + Sex + (1 | Subject) + storage_time
y ~ current_feature + Sex + (1 | Subject) + storage_time
y ~ current_feature + Sex + (1 | Subject) + storage_time
y ~ current_feature + Sex + (1 | Subject) + storage_time
y ~ current_feature + Sex + (1 | Subject) + storage_time
y ~ current_feature + Sex + (1 | Subject) + storage_time
```

y ~ current_feature + Sex + (1 | Subject) + storage_time
y ~ current_feature + Sex + (1 | Subject) + storage_time
y ~ current_feature + Sex + (1 | Subject) + storage_time
y ~ current_feature + Sex + (1 | Subject) + storage_time
y ~ current_feature + Sex + (1 | Subject) + storage_time
y ~ current_feature + Sex + (1 | Subject) + storage_time
y ~ current_feature + Sex + (1 | Subject) + storage_time
y ~ current_feature + Sex + (1 | Subject) + storage_time
y ~ current_feature + Sex + (1 | Subject) + storage_time
y ~ current_feature + Sex + (1 | Subject) + storage_time
y ~ current_feature + Sex + (1 | Subject) + storage_time
y ~ current_feature + Sex + (1 | Subject) + storage_time
y ~ current_feature + Sex + (1 | Subject) + storage_time
y ~ current_feature + Sex + (1 | Subject) + storage_time
y ~ current_feature + Sex + (1 | Subject) + storage_time
y ~ current_feature + Sex + (1 | Subject) + storage_time
y ~ current_feature + Sex + (1 | Subject) + storage_time
y ~ current_feature + Sex + (1 | Subject) + storage_time
y ~ current_feature + Sex + (1 | Subject) + storage_time
y ~ current_feature + Sex + (1 | Subject) + storage_time
y ~ current_feature + Sex + (1 | Subject) + storage_time
y ~ current_feature + Sex + (1 | Subject) + storage_time
y ~ current_feature + Sex + (1 | Subject) + storage_time
y ~ current_feature + Sex + (1 | Subject) + storage_time
y ~ current_feature + Sex + (1 | Subject) + storage_time
y ~ current_feature + Sex + (1 | Subject) + storage_time
y ~ current_feature + Sex + (1 | Subject) + storage_time
y ~ current_feature + Sex + (1 | Subject) + storage_time
y ~ current_feature + Sex + (1 | Subject) + storage_time
y ~ current_feature + Sex + (1 | Subject) + storage_time
y ~ current_feature + Sex + (1 | Subject) + storage_time
y ~ current_feature + Sex + (1 | Subject) + storage_time
y ~ current_feature + Sex + (1 | Subject) + storage_time
y ~ current_feature + Sex + (1 | Subject) + storage_time
y ~ current_feature + Sex + (1 | Subject) + storage_time
y ~ current_feature + Sex + (1 | Subject) + storage_time
y ~ current_feature + Sex + (1 | Subject) + storage_time
y ~ current_feature + Sex + (1 | Subject) + storage_time
y ~ current_feature + Sex + (1 | Subject) + storage_time
y ~ current_feature + Sex + (1 | Subject) + storage_time
y ~ current_feature + Sex + (1 | Subject) + storage_time
y ~ current_feature + Sex + (1 | Subject) + storage_time
y ~ current_feature + Sex + (1 | Subject) + storage_time
y ~ current_feature + Sex + (1 | Subject) + storage_time
y ~ current_feature + Sex + (1 | Subject) + storage_time
y ~ current_feature + Sex + (1 | Subject) + storage_time
y ~ current_feature + Sex + (1 | Subject) + storage_time
y ~ current_feature + Sex + (1 | Subject) + storage_time

y ~ current_feature + Sex + (1 | Subject) + storage_time
y ~ current_feature + Sex + (1 | Subject) + storage_time
y ~ current_feature + Sex + (1 | Subject) + storage_time
y ~ current_feature + Sex + (1 | Subject) + storage_time
y ~ current_feature + Sex + (1 | Subject) + storage_time
y ~ current_feature + Sex + (1 | Subject) + storage_time
y ~ current_feature + Sex + (1 | Subject) + storage_time
y ~ current_feature + Sex + (1 | Subject) + storage_time
y ~ current_feature + Sex + (1 | Subject) + storage_time
y ~ current_feature + Sex + (1 | Subject) + storage_time
y ~ current_feature + Sex + (1 | Subject) + storage_time
y ~ current_feature + Sex + (1 | Subject) + storage_time
y ~ current_feature + Sex + (1 | Subject) + storage_time
y ~ current_feature + Sex + (1 | Subject) + storage_time
y ~ current_feature + Sex + (1 | Subject) + storage_time
y ~ current_feature + Sex + (1 | Subject) + storage_time
y ~ current_feature + Sex + (1 | Subject) + storage_time
y ~ current_feature + Sex + (1 | Subject) + storage_time
y ~ current_feature + Sex + (1 | Subject) + storage_time
y ~ current_feature + Sex + (1 | Subject) + storage_time
y ~ current_feature + Sex + (1 | Subject) + storage_time
y ~ current_feature + Sex + (1 | Subject) + storage_time
y ~ current_feature + Sex + (1 | Subject) + storage_time
y ~ current_feature + Sex + (1 | Subject) + storage_time
y ~ current_feature + Sex + (1 | Subject) + storage_time
y ~ current_feature + Sex + (1 | Subject) + storage_time
y ~ current_feature + Sex + (1 | Subject) + storage_time
y ~ current_feature + Sex + (1 | Subject) + storage_time
y ~ current_feature + Sex + (1 | Subject) + storage_time
y ~ current_feature + Sex + (1 | Subject) + storage_time
y ~ current_feature + Sex + (1 | Subject) + storage_time
y ~ current_feature + Sex + (1 | Subject) + storage_time
y ~ current_feature + Sex + (1 | Subject) + storage_time
y ~ current_feature + Sex + (1 | Subject) + storage_time
y ~ current_feature + Sex + (1 | Subject) + storage_time
y ~ current_feature + Sex + (1 | Subject) + storage_time
y ~ current_feature + Sex + (1 | Subject) + storage_time
y ~ current_feature + Sex + (1 | Subject) + storage_time
y ~ current_feature + Sex + (1 | Subject) + storage_time
y ~ current_feature + Sex + (1 | Subject) + storage_time
y ~ current_feature + Sex + (1 | Subject) + storage_time
y ~ current_feature + Sex + (1 | Subject) + storage_time
y ~ current_feature + Sex + (1 | Subject) + storage_time
y ~ current_feature + Sex + (1 | Subject) + storage_time
y ~ current_feature + Sex + (1 | Subject) + storage_time
y ~ current_feature + Sex + (1 | Subject) + storage_time
y ~ current_feature + Sex + (1 | Subject) + storage_time
y ~ current_feature + Sex + (1 | Subject) + storage_time

y ~ current_feature + Sex + (1 | Subject) + storage_time
y ~ current_feature + Sex + (1 | Subject) + storage_time
y ~ current_feature + Sex + (1 | Subject) + storage_time
y ~ current_feature + Sex + (1 | Subject) + storage_time
y ~ current_feature + Sex + (1 | Subject) + storage_time
y ~ current_feature + Sex + (1 | Subject) + storage_time
y ~ current_feature + Sex + (1 | Subject) + storage_time
y ~ current_feature + Sex + (1 | Subject) + storage_time
y ~ current_feature + Sex + (1 | Subject) + storage_time
y ~ current_feature + Sex + (1 | Subject) + storage_time
y ~ current_feature + Sex + (1 | Subject) + storage_time
y ~ current_feature + Sex + (1 | Subject) + storage_time
y ~ current_feature + Sex + (1 | Subject) + storage_time
y ~ current_feature + Sex + (1 | Subject) + storage_time
y ~ current_feature + Sex + (1 | Subject) + storage_time
y ~ current_feature + Sex + (1 | Subject) + storage_time
y ~ current_feature + Sex + (1 | Subject) + storage_time
y ~ current_feature + Sex + (1 | Subject) + storage_time
y ~ current_feature + Sex + (1 | Subject) + storage_time
y ~ current_feature + Sex + (1 | Subject) + storage_time
y ~ current_feature + Sex + (1 | Subject) + storage_time
y ~ current_feature + Sex + (1 | Subject) + storage_time
y ~ current_feature + Sex + (1 | Subject) + storage_time
y ~ current_feature + Sex + (1 | Subject) + storage_time
y ~ current_feature + Sex + (1 | Subject) + storage_time
y ~ current_feature + Sex + (1 | Subject) + storage_time
y ~ current_feature + Sex + (1 | Subject) + storage_time
y ~ current_feature + Sex + (1 | Subject) + storage_time
y ~ current_feature + Sex + (1 | Subject) + storage_time
y ~ current_feature + Sex + (1 | Subject) + storage_time
y ~ current_feature + Sex + (1 | Subject) + storage_time
y ~ current_feature + Sex + (1 | Subject) + storage_time
y ~ current_feature + Sex + (1 | Subject) + storage_time
y ~ current_feature + Sex + (1 | Subject) + storage_time
y ~ current_feature + Sex + (1 | Subject) + storage_time
y ~ current_feature + Sex + (1 | Subject) + storage_time
y ~ current_feature + Sex + (1 | Subject) + storage_time
y ~ current_feature + Sex + (1 | Subject) + storage_time
y ~ current_feature + Sex + (1 | Subject) + storage_time
y ~ current_feature + Sex + (1 | Subject) + storage_time
y ~ current_feature + Sex + (1 | Subject) + storage_time
y ~ current_feature + Sex + (1 | Subject) + storage_time
y ~ current_feature + Sex + (1 | Subject) + storage_time
y ~ current_feature + Sex + (1 | Subject) + storage_time
y ~ current_feature + Sex + (1 | Subject) + storage_time
y ~ current_feature + Sex + (1 | Subject) + storage_time
y ~ current_feature + Sex + (1 | Subject) + storage_time
y ~ current_feature + Sex + (1 | Subject) + storage_time

```
y ~ current_feature + Sex + (1 | Subject) + storage_time
y ~ current_feature + Sex + (1 | Subject) + storage_time
y ~ current_feature + Sex + (1 | Subject) + storage_time
y ~ current_feature + Sex + (1 | Subject) + storage_time
y ~ current_feature + Sex + (1 | Subject) + storage_time
y ~ current_feature + Sex + (1 | Subject) + storage_time
y ~ current_feature + Sex + (1 | Subject) + storage_time
y ~ current_feature + Sex + (1 | Subject) + storage_time
y ~ current_feature + Sex + (1 | Subject) + storage_time
y ~ current_feature + Sex + (1 | Subject) + storage_time
y ~ current_feature + Sex + (1 | Subject) + storage_time
y ~ current_feature + Sex + (1 | Subject) + storage_time
y ~ current_feature + Sex + (1 | Subject) + storage_time
y ~ current_feature + Sex + (1 | Subject) + storage_time
y ~ current_feature + Sex + (1 | Subject) + storage_time
y ~ current_feature + Sex + (1 | Subject) + storage_time
y ~ current_feature + Sex + (1 | Subject) + storage_time
y ~ current_feature + Sex + (1 | Subject) + storage_time
y ~ current_feature + Sex + (1 | Subject) + storage_time
y ~ current_feature + Sex + (1 | Subject) + storage_time
y ~ current_feature + Sex + (1 | Subject) + storage_time
y ~ current_feature + Sex + (1 | Subject) + storage_time
y ~ current_feature + Sex + (1 | Subject) + storage_time
y ~ current_feature + Sex + (1 | Subject) + storage_time
y ~ current_feature + Sex + (1 | Subject) + storage_time
y ~ current_feature + Sex + (1 | Subject) + storage_time
y ~ current_feature + Sex + (1 | Subject) + storage_time
y ~ current_feature + Sex + (1 | Subject) + storage_time
y ~ current_feature + Sex + (1 | Subject) + storage_time
y ~ current_feature + Sex + (1 | Subject) + storage_time
y ~ current_feature + Sex + (1 | Subject) + storage_time
y ~ current_feature + Sex + (1 | Subject) + storage_time
y ~ current_feature + Sex + (1 | Subject) + storage_time
y ~ current_feature + Sex + (1 | Subject) + storage_time
y ~ current_feature + Sex + (1 | Subject) + storage_time
y ~ current_feature + Sex + (1 | Subject) + storage_time
y ~ current_feature + Sex + (1 | Subject) + storage_time
y ~ current_feature + Sex + (1 | Subject) + storage_time
y ~ current_feature + Sex + (1 | Subject) + storage_time
y ~ current_feature + Sex + (1 | Subject) + storage_time
y ~ current_feature + Sex + (1 | Subject) + storage_time
y ~ current_feature + Sex + (1 | Subject) + storage_time
y ~ current_feature + Sex + (1 | Subject) + storage_time
y ~ current_feature + Sex + (1 | Subject) + storage_time
y ~ current_feature + Sex + (1 | Subject) + storage_time
y ~ current_feature + Sex + (1 | Subject) + storage_time
y ~ current_feature + Sex + (1 | Subject) + storage_time
y ~ current_feature + Sex + (1 | Subject) + storage_time
```

y ~ current_feature + Sex + (1 | Subject) + storage_time
y ~ current_feature + Sex + (1 | Subject) + storage_time
y ~ current_feature + Sex + (1 | Subject) + storage_time
y ~ current_feature + Sex + (1 | Subject) + storage_time
y ~ current_feature + Sex + (1 | Subject) + storage_time
y ~ current_feature + Sex + (1 | Subject) + storage_time
y ~ current_feature + Sex + (1 | Subject) + storage_time
y ~ current_feature + Sex + (1 | Subject) + storage_time
y ~ current_feature + Sex + (1 | Subject) + storage_time
y ~ current_feature + Sex + (1 | Subject) + storage_time
y ~ current_feature + Sex + (1 | Subject) + storage_time
y ~ current_feature + Sex + (1 | Subject) + storage_time
y ~ current_feature + Sex + (1 | Subject) + storage_time
y ~ current_feature + Sex + (1 | Subject) + storage_time
y ~ current_feature + Sex + (1 | Subject) + storage_time
y ~ current_feature + Sex + (1 | Subject) + storage_time
y ~ current_feature + Sex + (1 | Subject) + storage_time
y ~ current_feature + Sex + (1 | Subject) + storage_time
y ~ current_feature + Sex + (1 | Subject) + storage_time
y ~ current_feature + Sex + (1 | Subject) + storage_time
y ~ current_feature + Sex + (1 | Subject) + storage_time
y ~ current_feature + Sex + (1 | Subject) + storage_time
y ~ current_feature + Sex + (1 | Subject) + storage_time
y ~ current_feature + Sex + (1 | Subject) + storage_time
y ~ current_feature + Sex + (1 | Subject) + storage_time
y ~ current_feature + Sex + (1 | Subject) + storage_time
y ~ current_feature + Sex + (1 | Subject) + storage_time
y ~ current_feature + Sex + (1 | Subject) + storage_time
y ~ current_feature + Sex + (1 | Subject) + storage_time
y ~ current_feature + Sex + (1 | Subject) + storage_time
y ~ current_feature + Sex + (1 | Subject) + storage_time
y ~ current_feature + Sex + (1 | Subject) + storage_time
y ~ current_feature + Sex + (1 | Subject) + storage_time
y ~ current_feature + Sex + (1 | Subject) + storage_time
y ~ current_feature + Sex + (1 | Subject) + storage_time
y ~ current_feature + Sex + (1 | Subject) + storage_time
y ~ current_feature + Sex + (1 | Subject) + storage_time
y ~ current_feature + Sex + (1 | Subject) + storage_time
y ~ current_feature + Sex + (1 | Subject) + storage_time
y ~ current_feature + Sex + (1 | Subject) + storage_time
y ~ current_feature + Sex + (1 | Subject) + storage_time
y ~ current_feature + Sex + (1 | Subject) + storage_time
y ~ current_feature + Sex + (1 | Subject) + storage_time
y ~ current_feature + Sex + (1 | Subject) + storage_time
y ~ current_feature + Sex + (1 | Subject) + storage_time
y ~ current_feature + Sex + (1 | Subject) + storage_time
y ~ current_feature + Sex + (1 | Subject) + storage_time
y ~ current_feature + Sex + (1 | Subject) + storage_time

y ~ current_feature + Sex + (1 | Subject) + storage_time
y ~ current_feature + Sex + (1 | Subject) + storage_time
y ~ current_feature + Sex + (1 | Subject) + storage_time
y ~ current_feature + Sex + (1 | Subject) + storage_time
y ~ current_feature + Sex + (1 | Subject) + storage_time
y ~ current_feature + Sex + (1 | Subject) + storage_time
y ~ current_feature + Sex + (1 | Subject) + storage_time
y ~ current_feature + Sex + (1 | Subject) + storage_time
y ~ current_feature + Sex + (1 | Subject) + storage_time
y ~ current_feature + Sex + (1 | Subject) + storage_time
y ~ current_feature + Sex + (1 | Subject) + storage_time
y ~ current_feature + Sex + (1 | Subject) + storage_time
y ~ current_feature + Sex + (1 | Subject) + storage_time
y ~ current_feature + Sex + (1 | Subject) + storage_time
y ~ current_feature + Sex + (1 | Subject) + storage_time
y ~ current_feature + Sex + (1 | Subject) + storage_time
y ~ current_feature + Sex + (1 | Subject) + storage_time
y ~ current_feature + Sex + (1 | Subject) + storage_time
y ~ current_feature + Sex + (1 | Subject) + storage_time
y ~ current_feature + Sex + (1 | Subject) + storage_time
y ~ current_feature + Sex + (1 | Subject) + storage_time
y ~ current_feature + Sex + (1 | Subject) + storage_time
y ~ current_feature + Sex + (1 | Subject) + storage_time
y ~ current_feature + Sex + (1 | Subject) + storage_time
y ~ current_feature + Sex + (1 | Subject) + storage_time
y ~ current_feature + Sex + (1 | Subject) + storage_time
y ~ current_feature + Sex + (1 | Subject) + storage_time
y ~ current_feature + Sex + (1 | Subject) + storage_time
y ~ current_feature + Sex + (1 | Subject) + storage_time
y ~ current_feature + Sex + (1 | Subject) + storage_time
y ~ current_feature + Sex + (1 | Subject) + storage_time
y ~ current_feature + Sex + (1 | Subject) + storage_time
y ~ current_feature + Sex + (1 | Subject) + storage_time
y ~ current_feature + Sex + (1 | Subject) + storage_time
y ~ current_feature + Sex + (1 | Subject) + storage_time
y ~ current_feature + Sex + (1 | Subject) + storage_time
y ~ current_feature + Sex + (1 | Subject) + storage_time
y ~ current_feature + Sex + (1 | Subject) + storage_time
y ~ current_feature + Sex + (1 | Subject) + storage_time
y ~ current_feature + Sex + (1 | Subject) + storage_time
y ~ current_feature + Sex + (1 | Subject) + storage_time
y ~ current_feature + Sex + (1 | Subject) + storage_time
y ~ current_feature + Sex + (1 | Subject) + storage_time
y ~ current_feature + Sex + (1 | Subject) + storage_time
y ~ current_feature + Sex + (1 | Subject) + storage_time
y ~ current_feature + Sex + (1 | Subject) + storage_time
y ~ current_feature + Sex + (1 | Subject) + storage_time
y ~ current_feature + Sex + (1 | Subject) + storage_time

y ~ current_feature + Sex + (1 | Subject) + storage_time
y ~ current_feature + Sex + (1 | Subject) + storage_time
y ~ current_feature + Sex + (1 | Subject) + storage_time
y ~ current_feature + Sex + (1 | Subject) + storage_time
y ~ current_feature + Sex + (1 | Subject) + storage_time
y ~ current_feature + Sex + (1 | Subject) + storage_time
y ~ current_feature + Sex + (1 | Subject) + storage_time
y ~ current_feature + Sex + (1 | Subject) + storage_time
y ~ current_feature + Sex + (1 | Subject) + storage_time
y ~ current_feature + Sex + (1 | Subject) + storage_time
y ~ current_feature + Sex + (1 | Subject) + storage_time
y ~ current_feature + Sex + (1 | Subject) + storage_time
y ~ current_feature + Sex + (1 | Subject) + storage_time
y ~ current_feature + Sex + (1 | Subject) + storage_time
y ~ current_feature + Sex + (1 | Subject) + storage_time
y ~ current_feature + Sex + (1 | Subject) + storage_time
y ~ current_feature + Sex + (1 | Subject) + storage_time
y ~ current_feature + Sex + (1 | Subject) + storage_time
y ~ current_feature + Sex + (1 | Subject) + storage_time
y ~ current_feature + Sex + (1 | Subject) + storage_time
y ~ current_feature + Sex + (1 | Subject) + storage_time
y ~ current_feature + Sex + (1 | Subject) + storage_time
y ~ current_feature + Sex + (1 | Subject) + storage_time
y ~ current_feature + Sex + (1 | Subject) + storage_time
y ~ current_feature + Sex + (1 | Subject) + storage_time
y ~ current_feature + Sex + (1 | Subject) + storage_time
y ~ current_feature + Sex + (1 | Subject) + storage_time
y ~ current_feature + Sex + (1 | Subject) + storage_time
y ~ current_feature + Sex + (1 | Subject) + storage_time
y ~ current_feature + Sex + (1 | Subject) + storage_time
y ~ current_feature + Sex + (1 | Subject) + storage_time
y ~ current_feature + Sex + (1 | Subject) + storage_time
y ~ current_feature + Sex + (1 | Subject) + storage_time
y ~ current_feature + Sex + (1 | Subject) + storage_time
y ~ current_feature + Sex + (1 | Subject) + storage_time
y ~ current_feature + Sex + (1 | Subject) + storage_time
y ~ current_feature + Sex + (1 | Subject) + storage_time
y ~ current_feature + Sex + (1 | Subject) + storage_time
y ~ current_feature + Sex + (1 | Subject) + storage_time
y ~ current_feature + Sex + (1 | Subject) + storage_time
y ~ current_feature + Sex + (1 | Subject) + storage_time
y ~ current_feature + Sex + (1 | Subject) + storage_time
y ~ current_feature + Sex + (1 | Subject) + storage_time
y ~ current_feature + Sex + (1 | Subject) + storage_time
y ~ current_feature + Sex + (1 | Subject) + storage_time
y ~ current_feature + Sex + (1 | Subject) + storage_time
y ~ current_feature + Sex + (1 | Subject) + storage_time
y ~ current_feature + Sex + (1 | Subject) + storage_time

y ~ current_feature + Sex + (1 | Subject) + storage_time
y ~ current_feature + Sex + (1 | Subject) + storage_time
y ~ current_feature + Sex + (1 | Subject) + storage_time
y ~ current_feature + Sex + (1 | Subject) + storage_time
y ~ current_feature + Sex + (1 | Subject) + storage_time
y ~ current_feature + Sex + (1 | Subject) + storage_time
y ~ current_feature + Sex + (1 | Subject) + storage_time
y ~ current_feature + Sex + (1 | Subject) + storage_time
y ~ current_feature + Sex + (1 | Subject) + storage_time
y ~ current_feature + Sex + (1 | Subject) + storage_time
y ~ current_feature + Sex + (1 | Subject) + storage_time
y ~ current_feature + Sex + (1 | Subject) + storage_time
y ~ current_feature + Sex + (1 | Subject) + storage_time
y ~ current_feature + Sex + (1 | Subject) + storage_time
y ~ current_feature + Sex + (1 | Subject) + storage_time
y ~ current_feature + Sex + (1 | Subject) + storage_time
y ~ current_feature + Sex + (1 | Subject) + storage_time
y ~ current_feature + Sex + (1 | Subject) + storage_time
y ~ current_feature + Sex + (1 | Subject) + storage_time
y ~ current_feature + Sex + (1 | Subject) + storage_time
y ~ current_feature + Sex + (1 | Subject) + storage_time
y ~ current_feature + Sex + (1 | Subject) + storage_time
y ~ current_feature + Sex + (1 | Subject) + storage_time
y ~ current_feature + Sex + (1 | Subject) + storage_time
y ~ current_feature + Sex + (1 | Subject) + storage_time
y ~ current_feature + Sex + (1 | Subject) + storage_time
y ~ current_feature + Sex + (1 | Subject) + storage_time
y ~ current_feature + Sex + (1 | Subject) + storage_time
y ~ current_feature + Sex + (1 | Subject) + storage_time
y ~ current_feature + Sex + (1 | Subject) + storage_time
y ~ current_feature + Sex + (1 | Subject) + storage_time
y ~ current_feature + Sex + (1 | Subject) + storage_time
y ~ current_feature + Sex + (1 | Subject) + storage_time
y ~ current_feature + Sex + (1 | Subject) + storage_time
y ~ current_feature + Sex + (1 | Subject) + storage_time
y ~ current_feature + Sex + (1 | Subject) + storage_time
y ~ current_feature + Sex + (1 | Subject) + storage_time
y ~ current_feature + Sex + (1 | Subject) + storage_time
y ~ current_feature + Sex + (1 | Subject) + storage_time
y ~ current_feature + Sex + (1 | Subject) + storage_time
y ~ current_feature + Sex + (1 | Subject) + storage_time
y ~ current_feature + Sex + (1 | Subject) + storage_time
y ~ current_feature + Sex + (1 | Subject) + storage_time
y ~ current_feature + Sex + (1 | Subject) + storage_time
y ~ current_feature + Sex + (1 | Subject) + storage_time
y ~ current_feature + Sex + (1 | Subject) + storage_time
y ~ current_feature + Sex + (1 | Subject) + storage_time
y ~ current_feature + Sex + (1 | Subject) + storage_time

y ~ current_feature + Sex + (1 | Subject) + storage_time
y ~ current_feature + Sex + (1 | Subject) + storage_time
y ~ current_feature + Sex + (1 | Subject) + storage_time
y ~ current_feature + Sex + (1 | Subject) + storage_time
y ~ current_feature + Sex + (1 | Subject) + storage_time
y ~ current_feature + Sex + (1 | Subject) + storage_time
y ~ current_feature + Sex + (1 | Subject) + storage_time
y ~ current_feature + Sex + (1 | Subject) + storage_time
y ~ current_feature + Sex + (1 | Subject) + storage_time
y ~ current_feature + Sex + (1 | Subject) + storage_time
y ~ current_feature + Sex + (1 | Subject) + storage_time
y ~ current_feature + Sex + (1 | Subject) + storage_time
y ~ current_feature + Sex + (1 | Subject) + storage_time
y ~ current_feature + Sex + (1 | Subject) + storage_time
y ~ current_feature + Sex + (1 | Subject) + storage_time
y ~ current_feature + Sex + (1 | Subject) + storage_time
y ~ current_feature + Sex + (1 | Subject) + storage_time
y ~ current_feature + Sex + (1 | Subject) + storage_time
y ~ current_feature + Sex + (1 | Subject) + storage_time
y ~ current_feature + Sex + (1 | Subject) + storage_time
y ~ current_feature + Sex + (1 | Subject) + storage_time
y ~ current_feature + Sex + (1 | Subject) + storage_time
y ~ current_feature + Sex + (1 | Subject) + storage_time
y ~ current_feature + Sex + (1 | Subject) + storage_time
y ~ current_feature + Sex + (1 | Subject) + storage_time
y ~ current_feature + Sex + (1 | Subject) + storage_time
y ~ current_feature + Sex + (1 | Subject) + storage_time
y ~ current_feature + Sex + (1 | Subject) + storage_time
y ~ current_feature + Sex + (1 | Subject) + storage_time
y ~ current_feature + Sex + (1 | Subject) + storage_time
y ~ current_feature + Sex + (1 | Subject) + storage_time
y ~ current_feature + Sex + (1 | Subject) + storage_time
y ~ current_feature + Sex + (1 | Subject) + storage_time
y ~ current_feature + Sex + (1 | Subject) + storage_time
y ~ current_feature + Sex + (1 | Subject) + storage_time
y ~ current_feature + Sex + (1 | Subject) + storage_time
y ~ current_feature + Sex + (1 | Subject) + storage_time
y ~ current_feature + Sex + (1 | Subject) + storage_time
y ~ current_feature + Sex + (1 | Subject) + storage_time
y ~ current_feature + Sex + (1 | Subject) + storage_time
y ~ current_feature + Sex + (1 | Subject) + storage_time
y ~ current_feature + Sex + (1 | Subject) + storage_time
y ~ current_feature + Sex + (1 | Subject) + storage_time
y ~ current_feature + Sex + (1 | Subject) + storage_time
y ~ current_feature + Sex + (1 | Subject) + storage_time
y ~ current_feature + Sex + (1 | Subject) + storage_time
y ~ current_feature + Sex + (1 | Subject) + storage_time
y ~ current_feature + Sex + (1 | Subject) + storage_time

y ~ current_feature + Sex + (1 | Subject) + storage_time
y ~ current_feature + Sex + (1 | Subject) + storage_time
y ~ current_feature + Sex + (1 | Subject) + storage_time
y ~ current_feature + Sex + (1 | Subject) + storage_time
y ~ current_feature + Sex + (1 | Subject) + storage_time
y ~ current_feature + Sex + (1 | Subject) + storage_time
y ~ current_feature + Sex + (1 | Subject) + storage_time
y ~ current_feature + Sex + (1 | Subject) + storage_time
y ~ current_feature + Sex + (1 | Subject) + storage_time
y ~ current_feature + Sex + (1 | Subject) + storage_time
y ~ current_feature + Sex + (1 | Subject) + storage_time
y ~ current_feature + Sex + (1 | Subject) + storage_time
y ~ current_feature + Sex + (1 | Subject) + storage_time
y ~ current_feature + Sex + (1 | Subject) + storage_time
y ~ current_feature + Sex + (1 | Subject) + storage_time
y ~ current_feature + Sex + (1 | Subject) + storage_time
y ~ current_feature + Sex + (1 | Subject) + storage_time
y ~ current_feature + Sex + (1 | Subject) + storage_time
y ~ current_feature + Sex + (1 | Subject) + storage_time
y ~ current_feature + Sex + (1 | Subject) + storage_time
y ~ current_feature + Sex + (1 | Subject) + storage_time
y ~ current_feature + Sex + (1 | Subject) + storage_time
y ~ current_feature + Sex + (1 | Subject) + storage_time
y ~ current_feature + Sex + (1 | Subject) + storage_time
y ~ current_feature + Sex + (1 | Subject) + storage_time
y ~ current_feature + Sex + (1 | Subject) + storage_time
y ~ current_feature + Sex + (1 | Subject) + storage_time
y ~ current_feature + Sex + (1 | Subject) + storage_time
y ~ current_feature + Sex + (1 | Subject) + storage_time
y ~ current_feature + Sex + (1 | Subject) + storage_time
y ~ current_feature + Sex + (1 | Subject) + storage_time
y ~ current_feature + Sex + (1 | Subject) + storage_time
y ~ current_feature + Sex + (1 | Subject) + storage_time
y ~ current_feature + Sex + (1 | Subject) + storage_time
y ~ current_feature + Sex + (1 | Subject) + storage_time
y ~ current_feature + Sex + (1 | Subject) + storage_time
y ~ current_feature + Sex + (1 | Subject) + storage_time
y ~ current_feature + Sex + (1 | Subject) + storage_time
y ~ current_feature + Sex + (1 | Subject) + storage_time
y ~ current_feature + Sex + (1 | Subject) + storage_time
y ~ current_feature + Sex + (1 | Subject) + storage_time
y ~ current_feature + Sex + (1 | Subject) + storage_time
y ~ current_feature + Sex + (1 | Subject) + storage_time
y ~ current_feature + Sex + (1 | Subject) + storage_time
y ~ current_feature + Sex + (1 | Subject) + storage_time
y ~ current_feature + Sex + (1 | Subject) + storage_time
y ~ current_feature + Sex + (1 | Subject) + storage_time
y ~ current_feature + Sex + (1 | Subject) + storage_time

y ~ current_feature + Sex + (1 | Subject) + storage_time
y ~ current_feature + Sex + (1 | Subject) + storage_time
y ~ current_feature + Sex + (1 | Subject) + storage_time
y ~ current_feature + Sex + (1 | Subject) + storage_time
y ~ current_feature + Sex + (1 | Subject) + storage_time
y ~ current_feature + Sex + (1 | Subject) + storage_time
y ~ current_feature + Sex + (1 | Subject) + storage_time
y ~ current_feature + Sex + (1 | Subject) + storage_time
y ~ current_feature + Sex + (1 | Subject) + storage_time
y ~ current_feature + Sex + (1 | Subject) + storage_time
y ~ current_feature + Sex + (1 | Subject) + storage_time
y ~ current_feature + Sex + (1 | Subject) + storage_time
y ~ current_feature + Sex + (1 | Subject) + storage_time
y ~ current_feature + Sex + (1 | Subject) + storage_time
y ~ current_feature + Sex + (1 | Subject) + storage_time
y ~ current_feature + Sex + (1 | Subject) + storage_time
y ~ current_feature + Sex + (1 | Subject) + storage_time
y ~ current_feature + Sex + (1 | Subject) + storage_time
y ~ current_feature + Sex + (1 | Subject) + storage_time
y ~ current_feature + Sex + (1 | Subject) + storage_time
y ~ current_feature + Sex + (1 | Subject) + storage_time
y ~ current_feature + Sex + (1 | Subject) + storage_time
y ~ current_feature + Sex + (1 | Subject) + storage_time
y ~ current_feature + Sex + (1 | Subject) + storage_time
y ~ current_feature + Sex + (1 | Subject) + storage_time
y ~ current_feature + Sex + (1 | Subject) + storage_time
y ~ current_feature + Sex + (1 | Subject) + storage_time
y ~ current_feature + Sex + (1 | Subject) + storage_time
y ~ current_feature + Sex + (1 | Subject) + storage_time
y ~ current_feature + Sex + (1 | Subject) + storage_time
y ~ current_feature + Sex + (1 | Subject) + storage_time
y ~ current_feature + Sex + (1 | Subject) + storage_time
y ~ current_feature + Sex + (1 | Subject) + storage_time
y ~ current_feature + Sex + (1 | Subject) + storage_time
y ~ current_feature + Sex + (1 | Subject) + storage_time
y ~ current_feature + Sex + (1 | Subject) + storage_time
y ~ current_feature + Sex + (1 | Subject) + storage_time
y ~ current_feature + Sex + (1 | Subject) + storage_time
y ~ current_feature + Sex + (1 | Subject) + storage_time
y ~ current_feature + Sex + (1 | Subject) + storage_time
y ~ current_feature + Sex + (1 | Subject) + storage_time
y ~ current_feature + Sex + (1 | Subject) + storage_time
y ~ current_feature + Sex + (1 | Subject) + storage_time
y ~ current_feature + Sex + (1 | Subject) + storage_time
y ~ current_feature + Sex + (1 | Subject) + storage_time
y ~ current_feature + Sex + (1 | Subject) + storage_time
y ~ current_feature + Sex + (1 | Subject) + storage_time
y ~ current_feature + Sex + (1 | Subject) + storage_time

y ~ current_feature + Sex + (1 | Subject) + storage_time
y ~ current_feature + Sex + (1 | Subject) + storage_time
y ~ current_feature + Sex + (1 | Subject) + storage_time
y ~ current_feature + Sex + (1 | Subject) + storage_time
y ~ current_feature + Sex + (1 | Subject) + storage_time
y ~ current_feature + Sex + (1 | Subject) + storage_time
y ~ current_feature + Sex + (1 | Subject) + storage_time
y ~ current_feature + Sex + (1 | Subject) + storage_time
y ~ current_feature + Sex + (1 | Subject) + storage_time
y ~ current_feature + Sex + (1 | Subject) + storage_time
y ~ current_feature + Sex + (1 | Subject) + storage_time
y ~ current_feature + Sex + (1 | Subject) + storage_time
y ~ current_feature + Sex + (1 | Subject) + storage_time
y ~ current_feature + Sex + (1 | Subject) + storage_time
y ~ current_feature + Sex + (1 | Subject) + storage_time
y ~ current_feature + Sex + (1 | Subject) + storage_time
y ~ current_feature + Sex + (1 | Subject) + storage_time
y ~ current_feature + Sex + (1 | Subject) + storage_time
y ~ current_feature + Sex + (1 | Subject) + storage_time
y ~ current_feature + Sex + (1 | Subject) + storage_time
y ~ current_feature + Sex + (1 | Subject) + storage_time
y ~ current_feature + Sex + (1 | Subject) + storage_time
y ~ current_feature + Sex + (1 | Subject) + storage_time
y ~ current_feature + Sex + (1 | Subject) + storage_time
y ~ current_feature + Sex + (1 | Subject) + storage_time
y ~ current_feature + Sex + (1 | Subject) + storage_time
y ~ current_feature + Sex + (1 | Subject) + storage_time
y ~ current_feature + Sex + (1 | Subject) + storage_time
y ~ current_feature + Sex + (1 | Subject) + storage_time
y ~ current_feature + Sex + (1 | Subject) + storage_time
y ~ current_feature + Sex + (1 | Subject) + storage_time
y ~ current_feature + Sex + (1 | Subject) + storage_time
y ~ current_feature + Sex + (1 | Subject) + storage_time
y ~ current_feature + Sex + (1 | Subject) + storage_time
y ~ current_feature + Sex + (1 | Subject) + storage_time
y ~ current_feature + Sex + (1 | Subject) + storage_time
y ~ current_feature + Sex + (1 | Subject) + storage_time
y ~ current_feature + Sex + (1 | Subject) + storage_time
y ~ current_feature + Sex + (1 | Subject) + storage_time
y ~ current_feature + Sex + (1 | Subject) + storage_time
y ~ current_feature + Sex + (1 | Subject) + storage_time
y ~ current_feature + Sex + (1 | Subject) + storage_time
y ~ current_feature + Sex + (1 | Subject) + storage_time
y ~ current_feature + Sex + (1 | Subject) + storage_time
y ~ current_feature + Sex + (1 | Subject) + storage_time
y ~ current_feature + Sex + (1 | Subject) + storage_time
y ~ current_feature + Sex + (1 | Subject) + storage_time
y ~ current_feature + Sex + (1 | Subject) + storage_time

y ~ current_feature + Sex + (1 | Subject) + storage_time
y ~ current_feature + Sex + (1 | Subject) + storage_time
y ~ current_feature + Sex + (1 | Subject) + storage_time
y ~ current_feature + Sex + (1 | Subject) + storage_time
y ~ current_feature + Sex + (1 | Subject) + storage_time
y ~ current_feature + Sex + (1 | Subject) + storage_time
y ~ current_feature + Sex + (1 | Subject) + storage_time
y ~ current_feature + Sex + (1 | Subject) + storage_time
y ~ current_feature + Sex + (1 | Subject) + storage_time
y ~ current_feature + Sex + (1 | Subject) + storage_time
y ~ current_feature + Sex + (1 | Subject) + storage_time
y ~ current_feature + Sex + (1 | Subject) + storage_time
y ~ current_feature + Sex + (1 | Subject) + storage_time
y ~ current_feature + Sex + (1 | Subject) + storage_time
y ~ current_feature + Sex + (1 | Subject) + storage_time
y ~ current_feature + Sex + (1 | Subject) + storage_time
y ~ current_feature + Sex + (1 | Subject) + storage_time
y ~ current_feature + Sex + (1 | Subject) + storage_time
y ~ current_feature + Sex + (1 | Subject) + storage_time
y ~ current_feature + Sex + (1 | Subject) + storage_time
y ~ current_feature + Sex + (1 | Subject) + storage_time
y ~ current_feature + Sex + (1 | Subject) + storage_time
y ~ current_feature + Sex + (1 | Subject) + storage_time
y ~ current_feature + Sex + (1 | Subject) + storage_time
y ~ current_feature + Sex + (1 | Subject) + storage_time
y ~ current_feature + Sex + (1 | Subject) + storage_time
y ~ current_feature + Sex + (1 | Subject) + storage_time
y ~ current_feature + Sex + (1 | Subject) + storage_time
y ~ current_feature + Sex + (1 | Subject) + storage_time
y ~ current_feature + Sex + (1 | Subject) + storage_time
y ~ current_feature + Sex + (1 | Subject) + storage_time
y ~ current_feature + Sex + (1 | Subject) + storage_time
y ~ current_feature + Sex + (1 | Subject) + storage_time
y ~ current_feature + Sex + (1 | Subject) + storage_time
y ~ current_feature + Sex + (1 | Subject) + storage_time
y ~ current_feature + Sex + (1 | Subject) + storage_time
y ~ current_feature + Sex + (1 | Subject) + storage_time
y ~ current_feature + Sex + (1 | Subject) + storage_time
y ~ current_feature + Sex + (1 | Subject) + storage_time
y ~ current_feature + Sex + (1 | Subject) + storage_time
y ~ current_feature + Sex + (1 | Subject) + storage_time
y ~ current_feature + Sex + (1 | Subject) + storage_time
y ~ current_feature + Sex + (1 | Subject) + storage_time
y ~ current_feature + Sex + (1 | Subject) + storage_time
y ~ current_feature + Sex + (1 | Subject) + storage_time
y ~ current_feature + Sex + (1 | Subject) + storage_time
y ~ current_feature + Sex + (1 | Subject) + storage_time
y ~ current_feature + Sex + (1 | Subject) + storage_time

y ~ current_feature + Sex + (1 | Subject) + storage_time
y ~ current_feature + Sex + (1 | Subject) + storage_time
y ~ current_feature + Sex + (1 | Subject) + storage_time
y ~ current_feature + Sex + (1 | Subject) + storage_time
y ~ current_feature + Sex + (1 | Subject) + storage_time
y ~ current_feature + Sex + (1 | Subject) + storage_time
y ~ current_feature + Sex + (1 | Subject) + storage_time
y ~ current_feature + Sex + (1 | Subject) + storage_time
y ~ current_feature + Sex + (1 | Subject) + storage_time
y ~ current_feature + Sex + (1 | Subject) + storage_time
y ~ current_feature + Sex + (1 | Subject) + storage_time
y ~ current_feature + Sex + (1 | Subject) + storage_time
y ~ current_feature + Sex + (1 | Subject) + storage_time
y ~ current_feature + Sex + (1 | Subject) + storage_time
y ~ current_feature + Sex + (1 | Subject) + storage_time
y ~ current_feature + Sex + (1 | Subject) + storage_time
y ~ current_feature + Sex + (1 | Subject) + storage_time
y ~ current_feature + Sex + (1 | Subject) + storage_time
y ~ current_feature + Sex + (1 | Subject) + storage_time
y ~ current_feature + Sex + (1 | Subject) + storage_time
y ~ current_feature + Sex + (1 | Subject) + storage_time
y ~ current_feature + Sex + (1 | Subject) + storage_time
y ~ current_feature + Sex + (1 | Subject) + storage_time
y ~ current_feature + Sex + (1 | Subject) + storage_time
y ~ current_feature + Sex + (1 | Subject) + storage_time
y ~ current_feature + Sex + (1 | Subject) + storage_time
y ~ current_feature + Sex + (1 | Subject) + storage_time
y ~ current_feature + Sex + (1 | Subject) + storage_time
y ~ current_feature + Sex + (1 | Subject) + storage_time
y ~ current_feature + Sex + (1 | Subject) + storage_time
y ~ current_feature + Sex + (1 | Subject) + storage_time
y ~ current_feature + Sex + (1 | Subject) + storage_time
y ~ current_feature + Sex + (1 | Subject) + storage_time
y ~ current_feature + Sex + (1 | Subject) + storage_time
y ~ current_feature + Sex + (1 | Subject) + storage_time
y ~ current_feature + Sex + (1 | Subject) + storage_time
y ~ current_feature + Sex + (1 | Subject) + storage_time
y ~ current_feature + Sex + (1 | Subject) + storage_time
y ~ current_feature + Sex + (1 | Subject) + storage_time
y ~ current_feature + Sex + (1 | Subject) + storage_time
y ~ current_feature + Sex + (1 | Subject) + storage_time
y ~ current_feature + Sex + (1 | Subject) + storage_time
y ~ current_feature + Sex + (1 | Subject) + storage_time
y ~ current_feature + Sex + (1 | Subject) + storage_time
y ~ current_feature + Sex + (1 | Subject) + storage_time
y ~ current_feature + Sex + (1 | Subject) + storage_time
y ~ current_feature + Sex + (1 | Subject) + storage_time
y ~ current_feature + Sex + (1 | Subject) + storage_time

```
y ~ current_feature + Sex + (1 | Subject) + storage_time
y ~ current_feature + Sex + (1 | Subject) + storage_time
y ~ current_feature + Sex + (1 | Subject) + storage_time
y ~ current_feature + Sex + (1 | Subject) + storage_time
y ~ current_feature + Sex + (1 | Subject) + storage_time
y ~ current_feature + Sex + (1 | Subject) + storage_time
y ~ current_feature + Sex + (1 | Subject) + storage_time
y ~ current_feature + Sex + (1 | Subject) + storage_time
y ~ current_feature + Sex + (1 | Subject) + storage_time
y ~ current_feature + Sex + (1 | Subject) + storage_time
y ~ current_feature + Sex + (1 | Subject) + storage_time
y ~ current_feature + Sex + (1 | Subject) + storage_time
y ~ current_feature + Sex + (1 | Subject) + storage_time
y ~ current_feature + Sex + (1 | Subject) + storage_time
y ~ current_feature + Sex + (1 | Subject) + storage_time
y ~ current_feature + Sex + (1 | Subject) + storage_time
y ~ current_feature + Sex + (1 | Subject) + storage_time
y ~ current_feature + Sex + (1 | Subject) + storage_time
y ~ current_feature + Sex + (1 | Subject) + storage_time
y ~ current_feature + Sex + (1 | Subject) + storage_time
y ~ current_feature + Sex + (1 | Subject) + storage_time
y ~ current_feature + Sex + (1 | Subject) + storage_time
y ~ current_feature + Sex + (1 | Subject) + storage_time
y ~ current_feature + Sex + (1 | Subject) + storage_time
y ~ current_feature + Sex + (1 | Subject) + storage_time
y ~ current_feature + Sex + (1 | Subject) + storage_time
y ~ current_feature + Sex + (1 | Subject) + storage_time
y ~ current_feature + Sex + (1 | Subject) + storage_time
y ~ current_feature + Sex + (1 | Subject) + storage_time
y ~ current_feature + Sex + (1 | Subject) + storage_time
y ~ current_feature + Sex + (1 | Subject) + storage_time
y ~ current_feature + Sex + (1 | Subject) + storage_time
y ~ current_feature + Sex + (1 | Subject) + storage_time
y ~ current_feature + Sex + (1 | Subject) + storage_time
y ~ current_feature + Sex + (1 | Subject) + storage_time
y ~ current_feature + Sex + (1 | Subject) + storage_time
y ~ current_feature + Sex + (1 | Subject) + storage_time
y ~ current_feature + Sex + (1 | Subject) + storage_time
y ~ current_feature + Sex + (1 | Subject) + storage_time
y ~ current_feature + Sex + (1 | Subject) + storage_time
y ~ current_feature + Sex + (1 | Subject) + storage_time
y ~ current_feature + Sex + (1 | Subject) + storage_time
y ~ current_feature + Sex + (1 | Subject) + storage_time
y ~ current_feature + Sex + (1 | Subject) + storage_time
y ~ current_feature + Sex + (1 | Subject) + storage_time
y ~ current_feature + Sex + (1 | Subject) + storage_time
y ~ current_feature + Sex + (1 | Subject) + storage_time
y ~ current_feature + Sex + (1 | Subject) + storage_time
```

```
y ~ current_feature + Sex + (1 | Subject) + storage_time
y ~ current_feature + Sex + (1 | Subject) + storage_time
y ~ current_feature + Sex + (1 | Subject) + storage_time
y ~ current_feature + Sex + (1 | Subject) + storage_time
y ~ current_feature + Sex + (1 | Subject) + storage_time
y ~ current_feature + Sex + (1 | Subject) + storage_time
y ~ current_feature + Sex + (1 | Subject) + storage_time
y ~ current_feature + Sex + (1 | Subject) + storage_time
y ~ current_feature + Sex + (1 | Subject) + storage_time
y ~ current_feature + Sex + (1 | Subject) + storage_time
y ~ current_feature + Sex + (1 | Subject) + storage_time
y ~ current_feature + Sex + (1 | Subject) + storage_time
y ~ current_feature + Sex + (1 | Subject) + storage_time
y ~ current_feature + Sex + (1 | Subject) + storage_time
y ~ current_feature + Sex + (1 | Subject) + storage_time
y ~ current_feature + Sex + (1 | Subject) + storage_time
y ~ current_feature + Sex + (1 | Subject) + storage_time
y ~ current_feature + Sex + (1 | Subject) + storage_time
y ~ current_feature + Sex + (1 | Subject) + storage_time
y ~ current_feature + Sex + (1 | Subject) + storage_time
```

```
y ~ current_feature + Sex + (1 | Subject) + storage_time
```

y ~ current_feature + Sex + (1 | Subject) + storage_time

y ~ current_feature + Sex + (1 | Subject) + storage_time

y ~ current_feature + Sex + (1 | Subject) + storage_time

```
y ~ current_feature + Sex + (1 | Subject) + storage_time
y ~ current_feature + Sex + (1 | Subject) + storage_time
y ~ current_feature + Sex + (1 | Subject) + storage_time
y ~ current_feature + Sex + (1 | Subject) + storage_time
y ~ current_feature + Sex + (1 | Subject) + storage_time
y ~ current_feature + Sex + (1 | Subject) + storage_time
y ~ current_feature + Sex + (1 | Subject) + storage_time
y ~ current_feature + Sex + (1 | Subject) + storage_time
y ~ current_feature + Sex + (1 | Subject) + storage_time
y ~ current_feature + Sex + (1 | Subject) + storage_time
y ~ current_feature + Sex + (1 | Subject) + storage_time
y ~ current_feature + Sex + (1 | Subject) + storage_time
y ~ current_feature + Sex + (1 | Subject) + storage_time
y ~ current_feature + Sex + (1 | Subject) + storage_time
y ~ current_feature + Sex + (1 | Subject) + storage_time
y ~ current_feature + Sex + (1 | Subject) + storage_time
y ~ current_feature + Sex + (1 | Subject) + storage_time
y ~ current_feature + Sex + (1 | Subject) + storage_time
y ~ current_feature + Sex + (1 | Subject) + storage_time
y ~ current_feature + Sex + (1 | Subject) + storage_time
```

```
y ~ current_feature + Sex + (1 | Subject) + storage_time
y ~ current_feature + Sex + (1 | Subject) + storage_time
y ~ current_feature + Sex + (1 | Subject) + storage_time
y ~ current_feature + Sex + (1 | Subject) + storage_time
y ~ current_feature + Sex + (1 | Subject) + storage_time
y ~ current_feature + Sex + (1 | Subject) + storage_time
y ~ current_feature + Sex + (1 | Subject) + storage_time
y ~ current_feature + Sex + (1 | Subject) + storage_time
y ~ current_feature + Sex + (1 | Subject) + storage_time
y ~ current_feature + Sex + (1 | Subject) + storage_time
y ~ current_feature + Sex + (1 | Subject) + storage_time
y ~ current_feature + Sex + (1 | Subject) + storage_time
y ~ current_feature + Sex + (1 | Subject) + storage_time
y ~ current_feature + Sex + (1 | Subject) + storage_time
y ~ current_feature + Sex + (1 | Subject) + storage_time
y ~ current_feature + Sex + (1 | Subject) + storage_time
y ~ current_feature + Sex + (1 | Subject) + storage_time
y ~ current_feature + Sex + (1 | Subject) + storage_time
y ~ current_feature + Sex + (1 | Subject) + storage_time
y ~ current_feature + Sex + (1 | Subject) + storage_time
y ~ current_feature + Sex + (1 | Subject) + storage_time
y ~ current_feature + Sex + (1 | Subject) + storage_time
y ~ current_feature + Sex + (1 | Subject) + storage_time
y ~ current_feature + Sex + (1 | Subject) + storage_time
y ~ current_feature + Sex + (1 | Subject) + storage_time
y ~ current_feature + Sex + (1 | Subject) + storage_time
y ~ current_feature + Sex + (1 | Subject) + storage_time
y ~ current_feature + Sex + (1 | Subject) + storage_time
y ~ current_feature + Sex + (1 | Subject) + storage_time
y ~ current_feature + Sex + (1 | Subject) + storage_time
y ~ current_feature + Sex + (1 | Subject) + storage_time
y ~ current_feature + Sex + (1 | Subject) + storage_time
y ~ current_feature + Sex + (1 | Subject) + storage_time
y ~ current_feature + Sex + (1 | Subject) + storage_time
y ~ current_feature + Sex + (1 | Subject) + storage_time
y ~ current_feature + Sex + (1 | Subject) + storage_time
y ~ current_feature + Sex + (1 | Subject) + storage_time
y ~ current_feature + Sex + (1 | Subject) + storage_time
y ~ current_feature + Sex + (1 | Subject) + storage_time
y ~ current_feature + Sex + (1 | Subject) + storage_time
y ~ current_feature + Sex + (1 | Subject) + storage_time
y ~ current_feature + Sex + (1 | Subject) + storage_time
y ~ current_feature + Sex + (1 | Subject) + storage_time
y ~ current_feature + Sex + (1 | Subject) + storage_time
y ~ current_feature + Sex + (1 | Subject) + storage_time
y ~ current_feature + Sex + (1 | Subject) + storage_time
y ~ current_feature + Sex + (1 | Subject) + storage_time
y ~ current_feature + Sex + (1 | Subject) + storage_time
```

y ~ current_feature + Sex + (1 | Subject) + storage_time
y ~ current_feature + Sex + (1 | Subject) + storage_time
y ~ current_feature + Sex + (1 | Subject) + storage_time
y ~ current_feature + Sex + (1 | Subject) + storage_time
y ~ current_feature + Sex + (1 | Subject) + storage_time
y ~ current_feature + Sex + (1 | Subject) + storage_time
y ~ current_feature + Sex + (1 | Subject) + storage_time
y ~ current_feature + Sex + (1 | Subject) + storage_time
y ~ current_feature + Sex + (1 | Subject) + storage_time
y ~ current_feature + Sex + (1 | Subject) + storage_time
y ~ current_feature + Sex + (1 | Subject) + storage_time
y ~ current_feature + Sex + (1 | Subject) + storage_time
y ~ current_feature + Sex + (1 | Subject) + storage_time
y ~ current_feature + Sex + (1 | Subject) + storage_time
y ~ current_feature + Sex + (1 | Subject) + storage_time
y ~ current_feature + Sex + (1 | Subject) + storage_time
y ~ current_feature + Sex + (1 | Subject) + storage_time
y ~ current_feature + Sex + (1 | Subject) + storage_time
y ~ current_feature + Sex + (1 | Subject) + storage_time
y ~ current_feature + Sex + (1 | Subject) + storage_time
y ~ current_feature + Sex + (1 | Subject) + storage_time
y ~ current_feature + Sex + (1 | Subject) + storage_time
y ~ current_feature + Sex + (1 | Subject) + storage_time
y ~ current_feature + Sex + (1 | Subject) + storage_time
y ~ current_feature + Sex + (1 | Subject) + storage_time
y ~ current_feature + Sex + (1 | Subject) + storage_time
y ~ current_feature + Sex + (1 | Subject) + storage_time
y ~ current_feature + Sex + (1 | Subject) + storage_time
y ~ current_feature + Sex + (1 | Subject) + storage_time
y ~ current_feature + Sex + (1 | Subject) + storage_time
y ~ current_feature + Sex + (1 | Subject) + storage_time
y ~ current_feature + Sex + (1 | Subject) + storage_time
y ~ current_feature + Sex + (1 | Subject) + storage_time
y ~ current_feature + Sex + (1 | Subject) + storage_time
y ~ current_feature + Sex + (1 | Subject) + storage_time
y ~ current_feature + Sex + (1 | Subject) + storage_time
y ~ current_feature + Sex + (1 | Subject) + storage_time
y ~ current_feature + Sex + (1 | Subject) + storage_time
y ~ current_feature + Sex + (1 | Subject) + storage_time
y ~ current_feature + Sex + (1 | Subject) + storage_time
y ~ current_feature + Sex + (1 | Subject) + storage_time
y ~ current_feature + Sex + (1 | Subject) + storage_time
y ~ current_feature + Sex + (1 | Subject) + storage_time
y ~ current_feature + Sex + (1 | Subject) + storage_time
y ~ current_feature + Sex + (1 | Subject) + storage_time
y ~ current_feature + Sex + (1 | Subject) + storage_time
y ~ current_feature + Sex + (1 | Subject) + storage_time
y ~ current_feature + Sex + (1 | Subject) + storage_time

```
y ~ current_feature + Sex + (1 | Subject) + storage_time
y ~ current_feature + Sex + (1 | Subject) + storage_time
y ~ current_feature + Sex + (1 | Subject) + storage_time
y ~ current_feature + Sex + (1 | Subject) + storage_time
y ~ current_feature + Sex + (1 | Subject) + storage_time
y ~ current_feature + Sex + (1 | Subject) + storage_time
y ~ current_feature + Sex + (1 | Subject) + storage_time
y ~ current_feature + Sex + (1 | Subject) + storage_time
y ~ current_feature + Sex + (1 | Subject) + storage_time
y ~ current_feature + Sex + (1 | Subject) + storage_time
y ~ current_feature + Sex + (1 | Subject) + storage_time
y ~ current_feature + Sex + (1 | Subject) + storage_time
y ~ current_feature + Sex + (1 | Subject) + storage_time
y ~ current_feature + Sex + (1 | Subject) + storage_time
y ~ current_feature + Sex + (1 | Subject) + storage_time
y ~ current_feature + Sex + (1 | Subject) + storage_time
y ~ current_feature + Sex + (1 | Subject) + storage_time
y ~ current_feature + Sex + (1 | Subject) + storage_time
y ~ current_feature + Sex + (1 | Subject) + storage_time
y ~ current_feature + Sex + (1 | Subject) + storage_time
y ~ current_feature + Sex + (1 | Subject) + storage_time
y ~ current_feature + Sex + (1 | Subject) + storage_time
y ~ current_feature + Sex + (1 | Subject) + storage_time
y ~ current_feature + Sex + (1 | Subject) + storage_time
y ~ current_feature + Sex + (1 | Subject) + storage_time
y ~ current_feature + Sex + (1 | Subject) + storage_time
y ~ current_feature + Sex + (1 | Subject) + storage_time
y ~ current_feature + Sex + (1 | Subject) + storage_time
y ~ current_feature + Sex + (1 | Subject) + storage_time
y ~ current_feature + Sex + (1 | Subject) + storage_time
y ~ current_feature + Sex + (1 | Subject) + storage_time
y ~ current_feature + Sex + (1 | Subject) + storage_time
y ~ current_feature + Sex + (1 | Subject) + storage_time
y ~ current_feature + Sex + (1 | Subject) + storage_time
y ~ current_feature + Sex + (1 | Subject) + storage_time
y ~ current_feature + Sex + (1 | Subject) + storage_time
y ~ current_feature + Sex + (1 | Subject) + storage_time
y ~ current_feature + Sex + (1 | Subject) + storage_time
y ~ current_feature + Sex + (1 | Subject) + storage_time
y ~ current_feature + Sex + (1 | Subject) + storage_time
y ~ current_feature + Sex + (1 | Subject) + storage_time
y ~ current_feature + Sex + (1 | Subject) + storage_time
y ~ current_feature + Sex + (1 | Subject) + storage_time
y ~ current_feature + Sex + (1 | Subject) + storage_time
y ~ current_feature + Sex + (1 | Subject) + storage_time
y ~ current_feature + Sex + (1 | Subject) + storage_time
y ~ current_feature + Sex + (1 | Subject) + storage_time
y ~ current_feature + Sex + (1 | Subject) + storage_time
```

y ~ current_feature + Sex + (1 | Subject) + storage_time
y ~ current_feature + Sex + (1 | Subject) + storage_time
y ~ current_feature + Sex + (1 | Subject) + storage_time
y ~ current_feature + Sex + (1 | Subject) + storage_time
y ~ current_feature + Sex + (1 | Subject) + storage_time
y ~ current_feature + Sex + (1 | Subject) + storage_time
y ~ current_feature + Sex + (1 | Subject) + storage_time
y ~ current_feature + Sex + (1 | Subject) + storage_time
y ~ current_feature + Sex + (1 | Subject) + storage_time
y ~ current_feature + Sex + (1 | Subject) + storage_time
y ~ current_feature + Sex + (1 | Subject) + storage_time
y ~ current_feature + Sex + (1 | Subject) + storage_time
y ~ current_feature + Sex + (1 | Subject) + storage_time
y ~ current_feature + Sex + (1 | Subject) + storage_time
y ~ current_feature + Sex + (1 | Subject) + storage_time
y ~ current_feature + Sex + (1 | Subject) + storage_time
y ~ current_feature + Sex + (1 | Subject) + storage_time
y ~ current_feature + Sex + (1 | Subject) + storage_time
y ~ current_feature + Sex + (1 | Subject) + storage_time
y ~ current_feature + Sex + (1 | Subject) + storage_time
y ~ current_feature + Sex + (1 | Subject) + storage_time
y ~ current_feature + Sex + (1 | Subject) + storage_time
y ~ current_feature + Sex + (1 | Subject) + storage_time
y ~ current_feature + Sex + (1 | Subject) + storage_time
y ~ current_feature + Sex + (1 | Subject) + storage_time
y ~ current_feature + Sex + (1 | Subject) + storage_time
y ~ current_feature + Sex + (1 | Subject) + storage_time
y ~ current_feature + Sex + (1 | Subject) + storage_time
y ~ current_feature + Sex + (1 | Subject) + storage_time
y ~ current_feature + Sex + (1 | Subject) + storage_time
y ~ current_feature + Sex + (1 | Subject) + storage_time
y ~ current_feature + Sex + (1 | Subject) + storage_time
y ~ current_feature + Sex + (1 | Subject) + storage_time
y ~ current_feature + Sex + (1 | Subject) + storage_time
y ~ current_feature + Sex + (1 | Subject) + storage_time
y ~ current_feature + Sex + (1 | Subject) + storage_time
y ~ current_feature + Sex + (1 | Subject) + storage_time
y ~ current_feature + Sex + (1 | Subject) + storage_time
y ~ current_feature + Sex + (1 | Subject) + storage_time
y ~ current_feature + Sex + (1 | Subject) + storage_time
y ~ current_feature + Sex + (1 | Subject) + storage_time
y ~ current_feature + Sex + (1 | Subject) + storage_time
y ~ current_feature + Sex + (1 | Subject) + storage_time
y ~ current_feature + Sex + (1 | Subject) + storage_time
y ~ current_feature + Sex + (1 | Subject) + storage_time
y ~ current_feature + Sex + (1 | Subject) + storage_time
y ~ current_feature + Sex + (1 | Subject) + storage_time
y ~ current_feature + Sex + (1 | Subject) + storage_time

y ~ current_feature + Sex + (1 | Subject) + storage_time
y ~ current_feature + Sex + (1 | Subject) + storage_time
y ~ current_feature + Sex + (1 | Subject) + storage_time
y ~ current_feature + Sex + (1 | Subject) + storage_time
y ~ current_feature + Sex + (1 | Subject) + storage_time
y ~ current_feature + Sex + (1 | Subject) + storage_time
y ~ current_feature + Sex + (1 | Subject) + storage_time
y ~ current_feature + Sex + (1 | Subject) + storage_time
y ~ current_feature + Sex + (1 | Subject) + storage_time
y ~ current_feature + Sex + (1 | Subject) + storage_time
y ~ current_feature + Sex + (1 | Subject) + storage_time
y ~ current_feature + Sex + (1 | Subject) + storage_time
y ~ current_feature + Sex + (1 | Subject) + storage_time
y ~ current_feature + Sex + (1 | Subject) + storage_time
y ~ current_feature + Sex + (1 | Subject) + storage_time
y ~ current_feature + Sex + (1 | Subject) + storage_time
y ~ current_feature + Sex + (1 | Subject) + storage_time
y ~ current_feature + Sex + (1 | Subject) + storage_time
y ~ current_feature + Sex + (1 | Subject) + storage_time
y ~ current_feature + Sex + (1 | Subject) + storage_time
y ~ current_feature + Sex + (1 | Subject) + storage_time
y ~ current_feature + Sex + (1 | Subject) + storage_time
y ~ current_feature + Sex + (1 | Subject) + storage_time
y ~ current_feature + Sex + (1 | Subject) + storage_time
y ~ current_feature + Sex + (1 | Subject) + storage_time
y ~ current_feature + Sex + (1 | Subject) + storage_time
y ~ current_feature + Sex + (1 | Subject) + storage_time
y ~ current_feature + Sex + (1 | Subject) + storage_time
y ~ current_feature + Sex + (1 | Subject) + storage_time
y ~ current_feature + Sex + (1 | Subject) + storage_time
y ~ current_feature + Sex + (1 | Subject) + storage_time
y ~ current_feature + Sex + (1 | Subject) + storage_time
y ~ current_feature + Sex + (1 | Subject) + storage_time
y ~ current_feature + Sex + (1 | Subject) + storage_time
y ~ current_feature + Sex + (1 | Subject) + storage_time
y ~ current_feature + Sex + (1 | Subject) + storage_time
y ~ current_feature + Sex + (1 | Subject) + storage_time
y ~ current_feature + Sex + (1 | Subject) + storage_time
y ~ current_feature + Sex + (1 | Subject) + storage_time
y ~ current_feature + Sex + (1 | Subject) + storage_time
y ~ current_feature + Sex + (1 | Subject) + storage_time
y ~ current_feature + Sex + (1 | Subject) + storage_time
y ~ current_feature + Sex + (1 | Subject) + storage_time
y ~ current_feature + Sex + (1 | Subject) + storage_time
y ~ current_feature + Sex + (1 | Subject) + storage_time
y ~ current_feature + Sex + (1 | Subject) + storage_time
y ~ current_feature + Sex + (1 | Subject) + storage_time
y ~ current_feature + Sex + (1 | Subject) + storage_time

y ~ current_feature + Sex + (1 | Subject) + storage_time
y ~ current_feature + Sex + (1 | Subject) + storage_time
y ~ current_feature + Sex + (1 | Subject) + storage_time
y ~ current_feature + Sex + (1 | Subject) + storage_time
y ~ current_feature + Sex + (1 | Subject) + storage_time
y ~ current_feature + Sex + (1 | Subject) + storage_time
y ~ current_feature + Sex + (1 | Subject) + storage_time
y ~ current_feature + Sex + (1 | Subject) + storage_time
y ~ current_feature + Sex + (1 | Subject) + storage_time
y ~ current_feature + Sex + (1 | Subject) + storage_time
y ~ current_feature + Sex + (1 | Subject) + storage_time
y ~ current_feature + Sex + (1 | Subject) + storage_time
y ~ current_feature + Sex + (1 | Subject) + storage_time
y ~ current_feature + Sex + (1 | Subject) + storage_time
y ~ current_feature + Sex + (1 | Subject) + storage_time
y ~ current_feature + Sex + (1 | Subject) + storage_time
y ~ current_feature + Sex + (1 | Subject) + storage_time
y ~ current_feature + Sex + (1 | Subject) + storage_time
y ~ current_feature + Sex + (1 | Subject) + storage_time
y ~ current_feature + Sex + (1 | Subject) + storage_time
y ~ current_feature + Sex + (1 | Subject) + storage_time
y ~ current_feature + Sex + (1 | Subject) + storage_time
y ~ current_feature + Sex + (1 | Subject) + storage_time
y ~ current_feature + Sex + (1 | Subject) + storage_time
y ~ current_feature + Sex + (1 | Subject) + storage_time
y ~ current_feature + Sex + (1 | Subject) + storage_time
y ~ current_feature + Sex + (1 | Subject) + storage_time
y ~ current_feature + Sex + (1 | Subject) + storage_time
y ~ current_feature + Sex + (1 | Subject) + storage_time
y ~ current_feature + Sex + (1 | Subject) + storage_time
y ~ current_feature + Sex + (1 | Subject) + storage_time
y ~ current_feature + Sex + (1 | Subject) + storage_time
y ~ current_feature + Sex + (1 | Subject) + storage_time
y ~ current_feature + Sex + (1 | Subject) + storage_time
y ~ current_feature + Sex + (1 | Subject) + storage_time
y ~ current_feature + Sex + (1 | Subject) + storage_time
y ~ current_feature + Sex + (1 | Subject) + storage_time
y ~ current_feature + Sex + (1 | Subject) + storage_time
y ~ current_feature + Sex + (1 | Subject) + storage_time
y ~ current_feature + Sex + (1 | Subject) + storage_time
y ~ current_feature + Sex + (1 | Subject) + storage_time
y ~ current_feature + Sex + (1 | Subject) + storage_time
y ~ current_feature + Sex + (1 | Subject) + storage_time
y ~ current_feature + Sex + (1 | Subject) + storage_time
y ~ current_feature + Sex + (1 | Subject) + storage_time
y ~ current_feature + Sex + (1 | Subject) + storage_time
y ~ current_feature + Sex + (1 | Subject) + storage_time
y ~ current_feature + Sex + (1 | Subject) + storage_time

```
y ~ current_feature + Sex + (1 | Subject) + storage_time
y ~ current_feature + Sex + (1 | Subject) + storage_time
y ~ current_feature + Sex + (1 | Subject) + storage_time
y ~ current_feature + Sex + (1 | Subject) + storage_time
y ~ current_feature + Sex + (1 | Subject) + storage_time
y ~ current_feature + Sex + (1 | Subject) + storage_time
y ~ current_feature + Sex + (1 | Subject) + storage_time
y ~ current_feature + Sex + (1 | Subject) + storage_time
y ~ current_feature + Sex + (1 | Subject) + storage_time
y ~ current_feature + Sex + (1 | Subject) + storage_time
y ~ current_feature + Sex + (1 | Subject) + storage_time
y ~ current_feature + Sex + (1 | Subject) + storage_time
y ~ current_feature + Sex + (1 | Subject) + storage_time
y ~ current_feature + Sex + (1 | Subject) + storage_time
y ~ current_feature + Sex + (1 | Subject) + storage_time
y ~ current_feature + Sex + (1 | Subject) + storage_time
y ~ current_feature + Sex + (1 | Subject) + storage_time
y ~ current_feature + Sex + (1 | Subject) + storage_time
y ~ current_feature + Sex + (1 | Subject) + storage_time
y ~ current_feature + Sex + (1 | Subject) + storage_time
y ~ current_feature + Sex + (1 | Subject) + storage_time
y ~ current_feature + Sex + (1 | Subject) + storage_time
y ~ current_feature + Sex + (1 | Subject) + storage_time
y ~ current_feature + Sex + (1 | Subject) + storage_time
y ~ current_feature + Sex + (1 | Subject) + storage_time
y ~ current_feature + Sex + (1 | Subject) + storage_time
y ~ current_feature + Sex + (1 | Subject) + storage_time
y ~ current_feature + Sex + (1 | Subject) + storage_time
y ~ current_feature + Sex + (1 | Subject) + storage_time
y ~ current_feature + Sex + (1 | Subject) + storage_time
y ~ current_feature + Sex + (1 | Subject) + storage_time
y ~ current_feature + Sex + (1 | Subject) + storage_time
y ~ current_feature + Sex + (1 | Subject) + storage_time
y ~ current_feature + Sex + (1 | Subject) + storage_time
y ~ current_feature + Sex + (1 | Subject) + storage_time
y ~ current_feature + Sex + (1 | Subject) + storage_time
y ~ current_feature + Sex + (1 | Subject) + storage_time
y ~ current_feature + Sex + (1 | Subject) + storage_time
y ~ current_feature + Sex + (1 | Subject) + storage_time
y ~ current_feature + Sex + (1 | Subject) + storage_time
y ~ current_feature + Sex + (1 | Subject) + storage_time
y ~ current_feature + Sex + (1 | Subject) + storage_time
y ~ current_feature + Sex + (1 | Subject) + storage_time
y ~ current_feature + Sex + (1 | Subject) + storage_time
y ~ current_feature + Sex + (1 | Subject) + storage_time
y ~ current_feature + Sex + (1 | Subject) + storage_time
y ~ current_feature + Sex + (1 | Subject) + storage_time
y ~ current_feature + Sex + (1 | Subject) + storage_time
```

```
y ~ current_feature + Sex + (1 | Subject) + storage_time
y ~ current_feature + Sex + (1 | Subject) + storage_time
y ~ current_feature + Sex + (1 | Subject) + storage_time
y ~ current_feature + Sex + (1 | Subject) + storage_time
y ~ current_feature + Sex + (1 | Subject) + storage_time
y ~ current_feature + Sex + (1 | Subject) + storage_time
y ~ current_feature + Sex + (1 | Subject) + storage_time
y ~ current_feature + Sex + (1 | Subject) + storage_time
y ~ current_feature + Sex + (1 | Subject) + storage_time
y ~ current_feature + Sex + (1 | Subject) + storage_time
y ~ current_feature + Sex + (1 | Subject) + storage_time
y ~ current_feature + Sex + (1 | Subject) + storage_time
y ~ current_feature + Sex + (1 | Subject) + storage_time
y ~ current_feature + Sex + (1 | Subject) + storage_time
y ~ current_feature + Sex + (1 | Subject) + storage_time
y ~ current_feature + Sex + (1 | Subject) + storage_time
y ~ current_feature + Sex + (1 | Subject) + storage_time
y ~ current_feature + Sex + (1 | Subject) + storage_time
y ~ current_feature + Sex + (1 | Subject) + storage_time
y ~ current_feature + Sex + (1 | Subject) + storage_time
y ~ current_feature + Sex + (1 | Subject) + storage_time
y ~ current_feature + Sex + (1 | Subject) + storage_time
y ~ current_feature + Sex + (1 | Subject) + storage_time
y ~ current_feature + Sex + (1 | Subject) + storage_time
y ~ current_feature + Sex + (1 | Subject) + storage_time
y ~ current_feature + Sex + (1 | Subject) + storage_time
y ~ current_feature + Sex + (1 | Subject) + storage_time
y ~ current_feature + Sex + (1 | Subject) + storage_time
y ~ current_feature + Sex + (1 | Subject) + storage_time
y ~ current_feature + Sex + (1 | Subject) + storage_time
y ~ current_feature + Sex + (1 | Subject) + storage_time
y ~ current_feature + Sex + (1 | Subject) + storage_time
y ~ current_feature + Sex + (1 | Subject) + storage_time
y ~ current_feature + Sex + (1 | Subject) + storage_time
y ~ current_feature + Sex + (1 | Subject) + storage_time
y ~ current_feature + Sex + (1 | Subject) + storage_time
y ~ current_feature + Sex + (1 | Subject) + storage_time
y ~ current_feature + Sex + (1 | Subject) + storage_time
y ~ current_feature + Sex + (1 | Subject) + storage_time
y ~ current_feature + Sex + (1 | Subject) + storage_time
y ~ current_feature + Sex + (1 | Subject) + storage_time
y ~ current_feature + Sex + (1 | Subject) + storage_time
y ~ current_feature + Sex + (1 | Subject) + storage_time
y ~ current_feature + Sex + (1 | Subject) + storage_time
y ~ current_feature + Sex + (1 | Subject) + storage_time
y ~ current_feature + Sex + (1 | Subject) + storage_time
y ~ current_feature + Sex + (1 | Subject) + storage_time
y ~ current_feature + Sex + (1 | Subject) + storage_time
```

y ~ current_feature + Sex + (1 | Subject) + storage_time
y ~ current_feature + Sex + (1 | Subject) + storage_time
y ~ current_feature + Sex + (1 | Subject) + storage_time
y ~ current_feature + Sex + (1 | Subject) + storage_time
y ~ current_feature + Sex + (1 | Subject) + storage_time
y ~ current_feature + Sex + (1 | Subject) + storage_time
y ~ current_feature + Sex + (1 | Subject) + storage_time
y ~ current_feature + Sex + (1 | Subject) + storage_time
y ~ current_feature + Sex + (1 | Subject) + storage_time
y ~ current_feature + Sex + (1 | Subject) + storage_time
y ~ current_feature + Sex + (1 | Subject) + storage_time
y ~ current_feature + Sex + (1 | Subject) + storage_time
y ~ current_feature + Sex + (1 | Subject) + storage_time
y ~ current_feature + Sex + (1 | Subject) + storage_time
y ~ current_feature + Sex + (1 | Subject) + storage_time
y ~ current_feature + Sex + (1 | Subject) + storage_time
y ~ current_feature + Sex + (1 | Subject) + storage_time
y ~ current_feature + Sex + (1 | Subject) + storage_time
y ~ current_feature + Sex + (1 | Subject) + storage_time
y ~ current_feature + Sex + (1 | Subject) + storage_time
y ~ current_feature + Sex + (1 | Subject) + storage_time
y ~ current_feature + Sex + (1 | Subject) + storage_time
y ~ current_feature + Sex + (1 | Subject) + storage_time
y ~ current_feature + Sex + (1 | Subject) + storage_time
y ~ current_feature + Sex + (1 | Subject) + storage_time
y ~ current_feature + Sex + (1 | Subject) + storage_time
y ~ current_feature + Sex + (1 | Subject) + storage_time
y ~ current_feature + Sex + (1 | Subject) + storage_time
y ~ current_feature + Sex + (1 | Subject) + storage_time
y ~ current_feature + Sex + (1 | Subject) + storage_time
y ~ current_feature + Sex + (1 | Subject) + storage_time
y ~ current_feature + Sex + (1 | Subject) + storage_time
y ~ current_feature + Sex + (1 | Subject) + storage_time
y ~ current_feature + Sex + (1 | Subject) + storage_time
y ~ current_feature + Sex + (1 | Subject) + storage_time
y ~ current_feature + Sex + (1 | Subject) + storage_time
y ~ current_feature + Sex + (1 | Subject) + storage_time
y ~ current_feature + Sex + (1 | Subject) + storage_time
y ~ current_feature + Sex + (1 | Subject) + storage_time
y ~ current_feature + Sex + (1 | Subject) + storage_time
y ~ current_feature + Sex + (1 | Subject) + storage_time
y ~ current_feature + Sex + (1 | Subject) + storage_time
y ~ current_feature + Sex + (1 | Subject) + storage_time
y ~ current_feature + Sex + (1 | Subject) + storage_time
y ~ current_feature + Sex + (1 | Subject) + storage_time
y ~ current_feature + Sex + (1 | Subject) + storage_time
y ~ current_feature + Sex + (1 | Subject) + storage_time
y ~ current_feature + Sex + (1 | Subject) + storage_time

y ~ current_feature + Sex + (1 | Subject) + storage_time
y ~ current_feature + Sex + (1 | Subject) + storage_time
y ~ current_feature + Sex + (1 | Subject) + storage_time
y ~ current_feature + Sex + (1 | Subject) + storage_time
y ~ current_feature + Sex + (1 | Subject) + storage_time
y ~ current_feature + Sex + (1 | Subject) + storage_time
y ~ current_feature + Sex + (1 | Subject) + storage_time
y ~ current_feature + Sex + (1 | Subject) + storage_time
y ~ current_feature + Sex + (1 | Subject) + storage_time
y ~ current_feature + Sex + (1 | Subject) + storage_time
y ~ current_feature + Sex + (1 | Subject) + storage_time
y ~ current_feature + Sex + (1 | Subject) + storage_time
y ~ current_feature + Sex + (1 | Subject) + storage_time
y ~ current_feature + Sex + (1 | Subject) + storage_time
y ~ current_feature + Sex + (1 | Subject) + storage_time
y ~ current_feature + Sex + (1 | Subject) + storage_time
y ~ current_feature + Sex + (1 | Subject) + storage_time
y ~ current_feature + Sex + (1 | Subject) + storage_time
y ~ current_feature + Sex + (1 | Subject) + storage_time
y ~ current_feature + Sex + (1 | Subject) + storage_time
y ~ current_feature + Sex + (1 | Subject) + storage_time
y ~ current_feature + Sex + (1 | Subject) + storage_time
y ~ current_feature + Sex + (1 | Subject) + storage_time
y ~ current_feature + Sex + (1 | Subject) + storage_time
y ~ current_feature + Sex + (1 | Subject) + storage_time
y ~ current_feature + Sex + (1 | Subject) + storage_time
y ~ current_feature + Sex + (1 | Subject) + storage_time
y ~ current_feature + Sex + (1 | Subject) + storage_time
y ~ current_feature + Sex + (1 | Subject) + storage_time
y ~ current_feature + Sex + (1 | Subject) + storage_time
y ~ current_feature + Sex + (1 | Subject) + storage_time
y ~ current_feature + Sex + (1 | Subject) + storage_time
y ~ current_feature + Sex + (1 | Subject) + storage_time
y ~ current_feature + Sex + (1 | Subject) + storage_time
y ~ current_feature + Sex + (1 | Subject) + storage_time
y ~ current_feature + Sex + (1 | Subject) + storage_time
y ~ current_feature + Sex + (1 | Subject) + storage_time
y ~ current_feature + Sex + (1 | Subject) + storage_time
y ~ current_feature + Sex + (1 | Subject) + storage_time
y ~ current_feature + Sex + (1 | Subject) + storage_time
y ~ current_feature + Sex + (1 | Subject) + storage_time
y ~ current_feature + Sex + (1 | Subject) + storage_time
y ~ current_feature + Sex + (1 | Subject) + storage_time
y ~ current_feature + Sex + (1 | Subject) + storage_time
y ~ current_feature + Sex + (1 | Subject) + storage_time
y ~ current_feature + Sex + (1 | Subject) + storage_time
y ~ current_feature + Sex + (1 | Subject) + storage_time
y ~ current_feature + Sex + (1 | Subject) + storage_time

y ~ current_feature + Sex + (1 | Subject) + storage_time
y ~ current_feature + Sex + (1 | Subject) + storage_time
y ~ current_feature + Sex + (1 | Subject) + storage_time
y ~ current_feature + Sex + (1 | Subject) + storage_time
y ~ current_feature + Sex + (1 | Subject) + storage_time
y ~ current_feature + Sex + (1 | Subject) + storage_time
y ~ current_feature + Sex + (1 | Subject) + storage_time
y ~ current_feature + Sex + (1 | Subject) + storage_time
y ~ current_feature + Sex + (1 | Subject) + storage_time
y ~ current_feature + Sex + (1 | Subject) + storage_time
y ~ current_feature + Sex + (1 | Subject) + storage_time
y ~ current_feature + Sex + (1 | Subject) + storage_time
y ~ current_feature + Sex + (1 | Subject) + storage_time
y ~ current_feature + Sex + (1 | Subject) + storage_time
y ~ current_feature + Sex + (1 | Subject) + storage_time
y ~ current_feature + Sex + (1 | Subject) + storage_time
y ~ current_feature + Sex + (1 | Subject) + storage_time
y ~ current_feature + Sex + (1 | Subject) + storage_time
y ~ current_feature + Sex + (1 | Subject) + storage_time
y ~ current_feature + Sex + (1 | Subject) + storage_time
y ~ current_feature + Sex + (1 | Subject) + storage_time
y ~ current_feature + Sex + (1 | Subject) + storage_time
y ~ current_feature + Sex + (1 | Subject) + storage_time
y ~ current_feature + Sex + (1 | Subject) + storage_time
y ~ current_feature + Sex + (1 | Subject) + storage_time
y ~ current_feature + Sex + (1 | Subject) + storage_time
y ~ current_feature + Sex + (1 | Subject) + storage_time
y ~ current_feature + Sex + (1 | Subject) + storage_time
y ~ current_feature + Sex + (1 | Subject) + storage_time
y ~ current_feature + Sex + (1 | Subject) + storage_time
y ~ current_feature + Sex + (1 | Subject) + storage_time
y ~ current_feature + Sex + (1 | Subject) + storage_time
y ~ current_feature + Sex + (1 | Subject) + storage_time
y ~ current_feature + Sex + (1 | Subject) + storage_time
y ~ current_feature + Sex + (1 | Subject) + storage_time
y ~ current_feature + Sex + (1 | Subject) + storage_time
y ~ current_feature + Sex + (1 | Subject) + storage_time
y ~ current_feature + Sex + (1 | Subject) + storage_time
y ~ current_feature + Sex + (1 | Subject) + storage_time
y ~ current_feature + Sex + (1 | Subject) + storage_time
y ~ current_feature + Sex + (1 | Subject) + storage_time
y ~ current_feature + Sex + (1 | Subject) + storage_time
y ~ current_feature + Sex + (1 | Subject) + storage_time
y ~ current_feature + Sex + (1 | Subject) + storage_time
y ~ current_feature + Sex + (1 | Subject) + storage_time
y ~ current_feature + Sex + (1 | Subject) + storage_time
y ~ current_feature + Sex + (1 | Subject) + storage_time
y ~ current_feature + Sex + (1 | Subject) + storage_time

y ~ current_feature + Sex + (1 | Subject) + storage_time
y ~ current_feature + Sex + (1 | Subject) + storage_time
y ~ current_feature + Sex + (1 | Subject) + storage_time
y ~ current_feature + Sex + (1 | Subject) + storage_time
y ~ current_feature + Sex + (1 | Subject) + storage_time
y ~ current_feature + Sex + (1 | Subject) + storage_time
y ~ current_feature + Sex + (1 | Subject) + storage_time
y ~ current_feature + Sex + (1 | Subject) + storage_time
y ~ current_feature + Sex + (1 | Subject) + storage_time
y ~ current_feature + Sex + (1 | Subject) + storage_time
y ~ current_feature + Sex + (1 | Subject) + storage_time
y ~ current_feature + Sex + (1 | Subject) + storage_time
y ~ current_feature + Sex + (1 | Subject) + storage_time
y ~ current_feature + Sex + (1 | Subject) + storage_time
y ~ current_feature + Sex + (1 | Subject) + storage_time
y ~ current_feature + Sex + (1 | Subject) + storage_time
y ~ current_feature + Sex + (1 | Subject) + storage_time
y ~ current_feature + Sex + (1 | Subject) + storage_time
y ~ current_feature + Sex + (1 | Subject) + storage_time
y ~ current_feature + Sex + (1 | Subject) + storage_time
y ~ current_feature + Sex + (1 | Subject) + storage_time
y ~ current_feature + Sex + (1 | Subject) + storage_time
y ~ current_feature + Sex + (1 | Subject) + storage_time
y ~ current_feature + Sex + (1 | Subject) + storage_time
y ~ current_feature + Sex + (1 | Subject) + storage_time
y ~ current_feature + Sex + (1 | Subject) + storage_time
y ~ current_feature + Sex + (1 | Subject) + storage_time
y ~ current_feature + Sex + (1 | Subject) + storage_time
y ~ current_feature + Sex + (1 | Subject) + storage_time
y ~ current_feature + Sex + (1 | Subject) + storage_time
y ~ current_feature + Sex + (1 | Subject) + storage_time
y ~ current_feature + Sex + (1 | Subject) + storage_time
y ~ current_feature + Sex + (1 | Subject) + storage_time
y ~ current_feature + Sex + (1 | Subject) + storage_time
y ~ current_feature + Sex + (1 | Subject) + storage_time
y ~ current_feature + Sex + (1 | Subject) + storage_time
y ~ current_feature + Sex + (1 | Subject) + storage_time
y ~ current_feature + Sex + (1 | Subject) + storage_time
y ~ current_feature + Sex + (1 | Subject) + storage_time
y ~ current_feature + Sex + (1 | Subject) + storage_time
y ~ current_feature + Sex + (1 | Subject) + storage_time
y ~ current_feature + Sex + (1 | Subject) + storage_time
y ~ current_feature + Sex + (1 | Subject) + storage_time
y ~ current_feature + Sex + (1 | Subject) + storage_time
y ~ current_feature + Sex + (1 | Subject) + storage_time
y ~ current_feature + Sex + (1 | Subject) + storage_time
y ~ current_feature + Sex + (1 | Subject) + storage_time
y ~ current_feature + Sex + (1 | Subject) + storage_time

y ~ current_feature + Sex + (1 | Subject) + storage_time
y ~ current_feature + Sex + (1 | Subject) + storage_time
y ~ current_feature + Sex + (1 | Subject) + storage_time
y ~ current_feature + Sex + (1 | Subject) + storage_time
y ~ current_feature + Sex + (1 | Subject) + storage_time
y ~ current_feature + Sex + (1 | Subject) + storage_time
y ~ current_feature + Sex + (1 | Subject) + storage_time
y ~ current_feature + Sex + (1 | Subject) + storage_time
y ~ current_feature + Sex + (1 | Subject) + storage_time
y ~ current_feature + Sex + (1 | Subject) + storage_time
y ~ current_feature + Sex + (1 | Subject) + storage_time
y ~ current_feature + Sex + (1 | Subject) + storage_time
y ~ current_feature + Sex + (1 | Subject) + storage_time
y ~ current_feature + Sex + (1 | Subject) + storage_time
y ~ current_feature + Sex + (1 | Subject) + storage_time
y ~ current_feature + Sex + (1 | Subject) + storage_time
y ~ current_feature + Sex + (1 | Subject) + storage_time
y ~ current_feature + Sex + (1 | Subject) + storage_time
y ~ current_feature + Sex + (1 | Subject) + storage_time
y ~ current_feature + Sex + (1 | Subject) + storage_time
y ~ current_feature + Sex + (1 | Subject) + storage_time
y ~ current_feature + Sex + (1 | Subject) + storage_time
y ~ current_feature + Sex + (1 | Subject) + storage_time
y ~ current_feature + Sex + (1 | Subject) + storage_time
y ~ current_feature + Sex + (1 | Subject) + storage_time
y ~ current_feature + Sex + (1 | Subject) + storage_time
y ~ current_feature + Sex + (1 | Subject) + storage_time
y ~ current_feature + Sex + (1 | Subject) + storage_time
y ~ current_feature + Sex + (1 | Subject) + storage_time
y ~ current_feature + Sex + (1 | Subject) + storage_time
y ~ current_feature + Sex + (1 | Subject) + storage_time
y ~ current_feature + Sex + (1 | Subject) + storage_time
y ~ current_feature + Sex + (1 | Subject) + storage_time
y ~ current_feature + Sex + (1 | Subject) + storage_time
y ~ current_feature + Sex + (1 | Subject) + storage_time
y ~ current_feature + Sex + (1 | Subject) + storage_time
y ~ current_feature + Sex + (1 | Subject) + storage_time
y ~ current_feature + Sex + (1 | Subject) + storage_time
y ~ current_feature + Sex + (1 | Subject) + storage_time
y ~ current_feature + Sex + (1 | Subject) + storage_time
y ~ current_feature + Sex + (1 | Subject) + storage_time
y ~ current_feature + Sex + (1 | Subject) + storage_time
y ~ current_feature + Sex + (1 | Subject) + storage_time
y ~ current_feature + Sex + (1 | Subject) + storage_time
y ~ current_feature + Sex + (1 | Subject) + storage_time
y ~ current_feature + Sex + (1 | Subject) + storage_time
y ~ current_feature + Sex + (1 | Subject) + storage_time
y ~ current_feature + Sex + (1 | Subject) + storage_time

```
y ~ current_feature + Sex + (1 | Subject) + storage_time
y ~ current_feature + Sex + (1 | Subject) + storage_time
y ~ current_feature + Sex + (1 | Subject) + storage_time
y ~ current_feature + Sex + (1 | Subject) + storage_time
y ~ current_feature + Sex + (1 | Subject) + storage_time
y ~ current_feature + Sex + (1 | Subject) + storage_time
y ~ current_feature + Sex + (1 | Subject) + storage_time
y ~ current_feature + Sex + (1 | Subject) + storage_time
y ~ current_feature + Sex + (1 | Subject) + storage_time
y ~ current_feature + Sex + (1 | Subject) + storage_time
y ~ current_feature + Sex + (1 | Subject) + storage_time
y ~ current_feature + Sex + (1 | Subject) + storage_time
y ~ current_feature + Sex + (1 | Subject) + storage_time
y ~ current_feature + Sex + (1 | Subject) + storage_time
y ~ current_feature + Sex + (1 | Subject) + storage_time
y ~ current_feature + Sex + (1 | Subject) + storage_time
y ~ current_feature + Sex + (1 | Subject) + storage_time
y ~ current_feature + Sex + (1 | Subject) + storage_time
y ~ current_feature + Sex + (1 | Subject) + storage_time
y ~ current_feature + Sex + (1 | Subject) + storage_time
y ~ current_feature + Sex + (1 | Subject) + storage_time
y ~ current_feature + Sex + (1 | Subject) + storage_time
y ~ current_feature + Sex + (1 | Subject) + storage_time
y ~ current_feature + Sex + (1 | Subject) + storage_time
y ~ current_feature + Sex + (1 | Subject) + storage_time
y ~ current_feature + Sex + (1 | Subject) + storage_time
y ~ current_feature + Sex + (1 | Subject) + storage_time
y ~ current_feature + Sex + (1 | Subject) + storage_time
y ~ current_feature + Sex + (1 | Subject) + storage_time
y ~ current_feature + Sex + (1 | Subject) + storage_time
y ~ current_feature + Sex + (1 | Subject) + storage_time
y ~ current_feature + Sex + (1 | Subject) + storage_time
y ~ current_feature + Sex + (1 | Subject) + storage_time
y ~ current_feature + Sex + (1 | Subject) + storage_time
y ~ current_feature + Sex + (1 | Subject) + storage_time
y ~ current_feature + Sex + (1 | Subject) + storage_time
y ~ current_feature + Sex + (1 | Subject) + storage_time
y ~ current_feature + Sex + (1 | Subject) + storage_time
y ~ current_feature + Sex + (1 | Subject) + storage_time
y ~ current_feature + Sex + (1 | Subject) + storage_time
y ~ current_feature + Sex + (1 | Subject) + storage_time
y ~ current_feature + Sex + (1 | Subject) + storage_time
y ~ current_feature + Sex + (1 | Subject) + storage_time
y ~ current_feature + Sex + (1 | Subject) + storage_time
y ~ current_feature + Sex + (1 | Subject) + storage_time
y ~ current_feature + Sex + (1 | Subject) + storage_time
y ~ current_feature + Sex + (1 | Subject) + storage_time
y ~ current_feature + Sex + (1 | Subject) + storage_time
```

y ~ current_feature + Sex + (1 | Subject) + storage_time
y ~ current_feature + Sex + (1 | Subject) + storage_time
y ~ current_feature + Sex + (1 | Subject) + storage_time
y ~ current_feature + Sex + (1 | Subject) + storage_time
y ~ current_feature + Sex + (1 | Subject) + storage_time
y ~ current_feature + Sex + (1 | Subject) + storage_time
y ~ current_feature + Sex + (1 | Subject) + storage_time
y ~ current_feature + Sex + (1 | Subject) + storage_time
y ~ current_feature + Sex + (1 | Subject) + storage_time
y ~ current_feature + Sex + (1 | Subject) + storage_time
y ~ current_feature + Sex + (1 | Subject) + storage_time
y ~ current_feature + Sex + (1 | Subject) + storage_time
y ~ current_feature + Sex + (1 | Subject) + storage_time
y ~ current_feature + Sex + (1 | Subject) + storage_time
y ~ current_feature + Sex + (1 | Subject) + storage_time
y ~ current_feature + Sex + (1 | Subject) + storage_time
y ~ current_feature + Sex + (1 | Subject) + storage_time
y ~ current_feature + Sex + (1 | Subject) + storage_time
y ~ current_feature + Sex + (1 | Subject) + storage_time
y ~ current_feature + Sex + (1 | Subject) + storage_time
y ~ current_feature + Sex + (1 | Subject) + storage_time
y ~ current_feature + Sex + (1 | Subject) + storage_time
y ~ current_feature + Sex + (1 | Subject) + storage_time
y ~ current_feature + Sex + (1 | Subject) + storage_time
y ~ current_feature + Sex + (1 | Subject) + storage_time
y ~ current_feature + Sex + (1 | Subject) + storage_time
y ~ current_feature + Sex + (1 | Subject) + storage_time
y ~ current_feature + Sex + (1 | Subject) + storage_time
y ~ current_feature + Sex + (1 | Subject) + storage_time
y ~ current_feature + Sex + (1 | Subject) + storage_time
y ~ current_feature + Sex + (1 | Subject) + storage_time
y ~ current_feature + Sex + (1 | Subject) + storage_time
y ~ current_feature + Sex + (1 | Subject) + storage_time
y ~ current_feature + Sex + (1 | Subject) + storage_time
y ~ current_feature + Sex + (1 | Subject) + storage_time
y ~ current_feature + Sex + (1 | Subject) + storage_time
y ~ current_feature + Sex + (1 | Subject) + storage_time
y ~ current_feature + Sex + (1 | Subject) + storage_time
y ~ current_feature + Sex + (1 | Subject) + storage_time
y ~ current_feature + Sex + (1 | Subject) + storage_time
y ~ current_feature + Sex + (1 | Subject) + storage_time
y ~ current_feature + Sex + (1 | Subject) + storage_time
y ~ current_feature + Sex + (1 | Subject) + storage_time
y ~ current_feature + Sex + (1 | Subject) + storage_time
y ~ current_feature + Sex + (1 | Subject) + storage_time
y ~ current_feature + Sex + (1 | Subject) + storage_time
y ~ current_feature + Sex + (1 | Subject) + storage_time
y ~ current_feature + Sex + (1 | Subject) + storage_time

y ~ current_feature + Sex + (1 | Subject) + storage_time
y ~ current_feature + Sex + (1 | Subject) + storage_time
y ~ current_feature + Sex + (1 | Subject) + storage_time
y ~ current_feature + Sex + (1 | Subject) + storage_time
y ~ current_feature + Sex + (1 | Subject) + storage_time
y ~ current_feature + Sex + (1 | Subject) + storage_time
y ~ current_feature + Sex + (1 | Subject) + storage_time
y ~ current_feature + Sex + (1 | Subject) + storage_time
y ~ current_feature + Sex + (1 | Subject) + storage_time
y ~ current_feature + Sex + (1 | Subject) + storage_time
y ~ current_feature + Sex + (1 | Subject) + storage_time
y ~ current_feature + Sex + (1 | Subject) + storage_time
y ~ current_feature + Sex + (1 | Subject) + storage_time
y ~ current_feature + Sex + (1 | Subject) + storage_time
y ~ current_feature + Sex + (1 | Subject) + storage_time
y ~ current_feature + Sex + (1 | Subject) + storage_time
y ~ current_feature + Sex + (1 | Subject) + storage_time
y ~ current_feature + Sex + (1 | Subject) + storage_time
y ~ current_feature + Sex + (1 | Subject) + storage_time
y ~ current_feature + Sex + (1 | Subject) + storage_time
y ~ current_feature + Sex + (1 | Subject) + storage_time
y ~ current_feature + Sex + (1 | Subject) + storage_time
y ~ current_feature + Sex + (1 | Subject) + storage_time
y ~ current_feature + Sex + (1 | Subject) + storage_time
y ~ current_feature + Sex + (1 | Subject) + storage_time
y ~ current_feature + Sex + (1 | Subject) + storage_time
y ~ current_feature + Sex + (1 | Subject) + storage_time
y ~ current_feature + Sex + (1 | Subject) + storage_time
y ~ current_feature + Sex + (1 | Subject) + storage_time
y ~ current_feature + Sex + (1 | Subject) + storage_time
y ~ current_feature + Sex + (1 | Subject) + storage_time
y ~ current_feature + Sex + (1 | Subject) + storage_time
y ~ current_feature + Sex + (1 | Subject) + storage_time
y ~ current_feature + Sex + (1 | Subject) + storage_time
y ~ current_feature + Sex + (1 | Subject) + storage_time
y ~ current_feature + Sex + (1 | Subject) + storage_time
y ~ current_feature + Sex + (1 | Subject) + storage_time
y ~ current_feature + Sex + (1 | Subject) + storage_time
y ~ current_feature + Sex + (1 | Subject) + storage_time
y ~ current_feature + Sex + (1 | Subject) + storage_time
y ~ current_feature + Sex + (1 | Subject) + storage_time
y ~ current_feature + Sex + (1 | Subject) + storage_time
y ~ current_feature + Sex + (1 | Subject) + storage_time
y ~ current_feature + Sex + (1 | Subject) + storage_time
y ~ current_feature + Sex + (1 | Subject) + storage_time
y ~ current_feature + Sex + (1 | Subject) + storage_time
y ~ current_feature + Sex + (1 | Subject) + storage_time
y ~ current_feature + Sex + (1 | Subject) + storage_time

y ~ current_feature + Sex + (1 | Subject) + storage_time
y ~ current_feature + Sex + (1 | Subject) + storage_time
y ~ current_feature + Sex + (1 | Subject) + storage_time
y ~ current_feature + Sex + (1 | Subject) + storage_time
y ~ current_feature + Sex + (1 | Subject) + storage_time
y ~ current_feature + Sex + (1 | Subject) + storage_time
y ~ current_feature + Sex + (1 | Subject) + storage_time
y ~ current_feature + Sex + (1 | Subject) + storage_time
y ~ current_feature + Sex + (1 | Subject) + storage_time
y ~ current_feature + Sex + (1 | Subject) + storage_time
y ~ current_feature + Sex + (1 | Subject) + storage_time
y ~ current_feature + Sex + (1 | Subject) + storage_time
y ~ current_feature + Sex + (1 | Subject) + storage_time
y ~ current_feature + Sex + (1 | Subject) + storage_time
y ~ current_feature + Sex + (1 | Subject) + storage_time
y ~ current_feature + Sex + (1 | Subject) + storage_time
y ~ current_feature + Sex + (1 | Subject) + storage_time
y ~ current_feature + Sex + (1 | Subject) + storage_time
y ~ current_feature + Sex + (1 | Subject) + storage_time
y ~ current_feature + Sex + (1 | Subject) + storage_time
y ~ current_feature + Sex + (1 | Subject) + storage_time
y ~ current_feature + Sex + (1 | Subject) + storage_time
y ~ current_feature + Sex + (1 | Subject) + storage_time
y ~ current_feature + Sex + (1 | Subject) + storage_time
y ~ current_feature + Sex + (1 | Subject) + storage_time
y ~ current_feature + Sex + (1 | Subject) + storage_time
y ~ current_feature + Sex + (1 | Subject) + storage_time
y ~ current_feature + Sex + (1 | Subject) + storage_time
y ~ current_feature + Sex + (1 | Subject) + storage_time
y ~ current_feature + Sex + (1 | Subject) + storage_time
y ~ current_feature + Sex + (1 | Subject) + storage_time
y ~ current_feature + Sex + (1 | Subject) + storage_time
y ~ current_feature + Sex + (1 | Subject) + storage_time
y ~ current_feature + Sex + (1 | Subject) + storage_time
y ~ current_feature + Sex + (1 | Subject) + storage_time
y ~ current_feature + Sex + (1 | Subject) + storage_time
y ~ current_feature + Sex + (1 | Subject) + storage_time
y ~ current_feature + Sex + (1 | Subject) + storage_time
y ~ current_feature + Sex + (1 | Subject) + storage_time
y ~ current_feature + Sex + (1 | Subject) + storage_time
y ~ current_feature + Sex + (1 | Subject) + storage_time
y ~ current_feature + Sex + (1 | Subject) + storage_time
y ~ current_feature + Sex + (1 | Subject) + storage_time
y ~ current_feature + Sex + (1 | Subject) + storage_time
y ~ current_feature + Sex + (1 | Subject) + storage_time
y ~ current_feature + Sex + (1 | Subject) + storage_time
y ~ current_feature + Sex + (1 | Subject) + storage_time
y ~ current_feature + Sex + (1 | Subject) + storage_time

```
y ~ current_feature + Sex + (1 | Subject) + storage_time
y ~ current_feature + Sex + (1 | Subject) + storage_time
y ~ current_feature + Sex + (1 | Subject) + storage_time
y ~ current_feature + Sex + (1 | Subject) + storage_time
y ~ current_feature + Sex + (1 | Subject) + storage_time
y ~ current_feature + Sex + (1 | Subject) + storage_time
y ~ current_feature + Sex + (1 | Subject) + storage_time
y ~ current_feature + Sex + (1 | Subject) + storage_time
y ~ current_feature + Sex + (1 | Subject) + storage_time
y ~ current_feature + Sex + (1 | Subject) + storage_time
y ~ current_feature + Sex + (1 | Subject) + storage_time
y ~ current_feature + Sex + (1 | Subject) + storage_time
y ~ current_feature + Sex + (1 | Subject) + storage_time
y ~ current_feature + Sex + (1 | Subject) + storage_time
y ~ current_feature + Sex + (1 | Subject) + storage_time
y ~ current_feature + Sex + (1 | Subject) + storage_time
y ~ current_feature + Sex + (1 | Subject) + storage_time
y ~ current_feature + Sex + (1 | Subject) + storage_time
y ~ current_feature + Sex + (1 | Subject) + storage_time
y ~ current_feature + Sex + (1 | Subject) + storage_time
y ~ current_feature + Sex + (1 | Subject) + storage_time
y ~ current_feature + Sex + (1 | Subject) + storage_time
y ~ current_feature + Sex + (1 | Subject) + storage_time
y ~ current_feature + Sex + (1 | Subject) + storage_time
y ~ current_feature + Sex + (1 | Subject) + storage_time
y ~ current_feature + Sex + (1 | Subject) + storage_time
y ~ current_feature + Sex + (1 | Subject) + storage_time
y ~ current_feature + Sex + (1 | Subject) + storage_time
y ~ current_feature + Sex + (1 | Subject) + storage_time
y ~ current_feature + Sex + (1 | Subject) + storage_time
y ~ current_feature + Sex + (1 | Subject) + storage_time
y ~ current_feature + Sex + (1 | Subject) + storage_time
y ~ current_feature + Sex + (1 | Subject) + storage_time
y ~ current_feature + Sex + (1 | Subject) + storage_time
y ~ current_feature + Sex + (1 | Subject) + storage_time
y ~ current_feature + Sex + (1 | Subject) + storage_time
y ~ current_feature + Sex + (1 | Subject) + storage_time
y ~ current_feature + Sex + (1 | Subject) + storage_time
y ~ current_feature + Sex + (1 | Subject) + storage_time
y ~ current_feature + Sex + (1 | Subject) + storage_time
y ~ current_feature + Sex + (1 | Subject) + storage_time
y ~ current_feature + Sex + (1 | Subject) + storage_time
y ~ current_feature + Sex + (1 | Subject) + storage_time
y ~ current_feature + Sex + (1 | Subject) + storage_time
y ~ current_feature + Sex + (1 | Subject) + storage_time
y ~ current_feature + Sex + (1 | Subject) + storage_time
y ~ current_feature + Sex + (1 | Subject) + storage_time
y ~ current_feature + Sex + (1 | Subject) + storage_time
```

```
y ~ current_feature + Sex + (1 | Subject) + storage_time
y ~ current_feature + Sex + (1 | Subject) + storage_time
y ~ current_feature + Sex + (1 | Subject) + storage_time
y ~ current_feature + Sex + (1 | Subject) + storage_time
y ~ current_feature + Sex + (1 | Subject) + storage_time
y ~ current_feature + Sex + (1 | Subject) + storage_time
y ~ current_feature + Sex + (1 | Subject) + storage_time
y ~ current_feature + Sex + (1 | Subject) + storage_time
y ~ current_feature + Sex + (1 | Subject) + storage_time
y ~ current_feature + Sex + (1 | Subject) + storage_time
y ~ current_feature + Sex + (1 | Subject) + storage_time
y ~ current_feature + Sex + (1 | Subject) + storage_time
y ~ current_feature + Sex + (1 | Subject) + storage_time
y ~ current_feature + Sex + (1 | Subject) + storage_time
y ~ current_feature + Sex + (1 | Subject) + storage_time
y ~ current_feature + Sex + (1 | Subject) + storage_time
y ~ current_feature + Sex + (1 | Subject) + storage_time
y ~ current_feature + Sex + (1 | Subject) + storage_time
y ~ current_feature + Sex + (1 | Subject) + storage_time
y ~ current_feature + Sex + (1 | Subject) + storage_time
y ~ current_feature + Sex + (1 | Subject) + storage_time
y ~ current_feature + Sex + (1 | Subject) + storage_time
y ~ current_feature + Sex + (1 | Subject) + storage_time
y ~ current_feature + Sex + (1 | Subject) + storage_time
y ~ current_feature + Sex + (1 | Subject) + storage_time
y ~ current_feature + Sex + (1 | Subject) + storage_time
y ~ current_feature + Sex + (1 | Subject) + storage_time
y ~ current_feature + Sex + (1 | Subject) + storage_time
y ~ current_feature + Sex + (1 | Subject) + storage_time
y ~ current_feature + Sex + (1 | Subject) + storage_time
y ~ current_feature + Sex + (1 | Subject) + storage_time
y ~ current_feature + Sex + (1 | Subject) + storage_time
y ~ current_feature + Sex + (1 | Subject) + storage_time
y ~ current_feature + Sex + (1 | Subject) + storage_time
y ~ current_feature + Sex + (1 | Subject) + storage_time
y ~ current_feature + Sex + (1 | Subject) + storage_time
y ~ current_feature + Sex + (1 | Subject) + storage_time
y ~ current_feature + Sex + (1 | Subject) + storage_time
y ~ current_feature + Sex + (1 | Subject) + storage_time
y ~ current_feature + Sex + (1 | Subject) + storage_time
y ~ current_feature + Sex + (1 | Subject) + storage_time
y ~ current_feature + Sex + (1 | Subject) + storage_time
y ~ current_feature + Sex + (1 | Subject) + storage_time
y ~ current_feature + Sex + (1 | Subject) + storage_time
y ~ current_feature + Sex + (1 | Subject) + storage_time
y ~ current_feature + Sex + (1 | Subject) + storage_time
y ~ current_feature + Sex + (1 | Subject) + storage_time
y ~ current_feature + Sex + (1 | Subject) + storage_time
```

y ~ current_feature + Sex + (1 | Subject) + storage_time
y ~ current_feature + Sex + (1 | Subject) + storage_time
y ~ current_feature + Sex + (1 | Subject) + storage_time
y ~ current_feature + Sex + (1 | Subject) + storage_time
y ~ current_feature + Sex + (1 | Subject) + storage_time
y ~ current_feature + Sex + (1 | Subject) + storage_time
y ~ current_feature + Sex + (1 | Subject) + storage_time
y ~ current_feature + Sex + (1 | Subject) + storage_time
y ~ current_feature + Sex + (1 | Subject) + storage_time
y ~ current_feature + Sex + (1 | Subject) + storage_time
y ~ current_feature + Sex + (1 | Subject) + storage_time
y ~ current_feature + Sex + (1 | Subject) + storage_time
y ~ current_feature + Sex + (1 | Subject) + storage_time
y ~ current_feature + Sex + (1 | Subject) + storage_time
y ~ current_feature + Sex + (1 | Subject) + storage_time
y ~ current_feature + Sex + (1 | Subject) + storage_time
y ~ current_feature + Sex + (1 | Subject) + storage_time
y ~ current_feature + Sex + (1 | Subject) + storage_time
y ~ current_feature + Sex + (1 | Subject) + storage_time
y ~ current_feature + Sex + (1 | Subject) + storage_time
y ~ current_feature + Sex + (1 | Subject) + storage_time
y ~ current_feature + Sex + (1 | Subject) + storage_time
y ~ current_feature + Sex + (1 | Subject) + storage_time
y ~ current_feature + Sex + (1 | Subject) + storage_time
y ~ current_feature + Sex + (1 | Subject) + storage_time
y ~ current_feature + Sex + (1 | Subject) + storage_time
y ~ current_feature + Sex + (1 | Subject) + storage_time
y ~ current_feature + Sex + (1 | Subject) + storage_time
y ~ current_feature + Sex + (1 | Subject) + storage_time
y ~ current_feature + Sex + (1 | Subject) + storage_time
y ~ current_feature + Sex + (1 | Subject) + storage_time
y ~ current_feature + Sex + (1 | Subject) + storage_time
y ~ current_feature + Sex + (1 | Subject) + storage_time
y ~ current_feature + Sex + (1 | Subject) + storage_time
y ~ current_feature + Sex + (1 | Subject) + storage_time
y ~ current_feature + Sex + (1 | Subject) + storage_time
y ~ current_feature + Sex + (1 | Subject) + storage_time
y ~ current_feature + Sex + (1 | Subject) + storage_time
y ~ current_feature + Sex + (1 | Subject) + storage_time
y ~ current_feature + Sex + (1 | Subject) + storage_time
y ~ current_feature + Sex + (1 | Subject) + storage_time
y ~ current_feature + Sex + (1 | Subject) + storage_time
y ~ current_feature + Sex + (1 | Subject) + storage_time
y ~ current_feature + Sex + (1 | Subject) + storage_time
y ~ current_feature + Sex + (1 | Subject) + storage_time
y ~ current_feature + Sex + (1 | Subject) + storage_time
y ~ current_feature + Sex + (1 | Subject) + storage_time
y ~ current_feature + Sex + (1 | Subject) + storage_time

y ~ current_feature + Sex + (1 | Subject) + storage_time
y ~ current_feature + Sex + (1 | Subject) + storage_time
y ~ current_feature + Sex + (1 | Subject) + storage_time
y ~ current_feature + Sex + (1 | Subject) + storage_time
y ~ current_feature + Sex + (1 | Subject) + storage_time
y ~ current_feature + Sex + (1 | Subject) + storage_time
y ~ current_feature + Sex + (1 | Subject) + storage_time
y ~ current_feature + Sex + (1 | Subject) + storage_time
y ~ current_feature + Sex + (1 | Subject) + storage_time
y ~ current_feature + Sex + (1 | Subject) + storage_time
y ~ current_feature + Sex + (1 | Subject) + storage_time
y ~ current_feature + Sex + (1 | Subject) + storage_time
y ~ current_feature + Sex + (1 | Subject) + storage_time
y ~ current_feature + Sex + (1 | Subject) + storage_time
y ~ current_feature + Sex + (1 | Subject) + storage_time
y ~ current_feature + Sex + (1 | Subject) + storage_time
y ~ current_feature + Sex + (1 | Subject) + storage_time
y ~ current_feature + Sex + (1 | Subject) + storage_time
y ~ current_feature + Sex + (1 | Subject) + storage_time
y ~ current_feature + Sex + (1 | Subject) + storage_time
y ~ current_feature + Sex + (1 | Subject) + storage_time
y ~ current_feature + Sex + (1 | Subject) + storage_time
y ~ current_feature + Sex + (1 | Subject) + storage_time
y ~ current_feature + Sex + (1 | Subject) + storage_time
y ~ current_feature + Sex + (1 | Subject) + storage_time
y ~ current_feature + Sex + (1 | Subject) + storage_time
y ~ current_feature + Sex + (1 | Subject) + storage_time
y ~ current_feature + Sex + (1 | Subject) + storage_time
y ~ current_feature + Sex + (1 | Subject) + storage_time
y ~ current_feature + Sex + (1 | Subject) + storage_time
y ~ current_feature + Sex + (1 | Subject) + storage_time
y ~ current_feature + Sex + (1 | Subject) + storage_time
y ~ current_feature + Sex + (1 | Subject) + storage_time
y ~ current_feature + Sex + (1 | Subject) + storage_time
y ~ current_feature + Sex + (1 | Subject) + storage_time
y ~ current_feature + Sex + (1 | Subject) + storage_time
y ~ current_feature + Sex + (1 | Subject) + storage_time
y ~ current_feature + Sex + (1 | Subject) + storage_time
y ~ current_feature + Sex + (1 | Subject) + storage_time
y ~ current_feature + Sex + (1 | Subject) + storage_time
y ~ current_feature + Sex + (1 | Subject) + storage_time
y ~ current_feature + Sex + (1 | Subject) + storage_time
y ~ current_feature + Sex + (1 | Subject) + storage_time
y ~ current_feature + Sex + (1 | Subject) + storage_time
y ~ current_feature + Sex + (1 | Subject) + storage_time
y ~ current_feature + Sex + (1 | Subject) + storage_time
y ~ current_feature + Sex + (1 | Subject) + storage_time
y ~ current_feature + Sex + (1 | Subject) + storage_time

y ~ current_feature + Sex + (1 | Subject) + storage_time
y ~ current_feature + Sex + (1 | Subject) + storage_time
y ~ current_feature + Sex + (1 | Subject) + storage_time
y ~ current_feature + Sex + (1 | Subject) + storage_time
y ~ current_feature + Sex + (1 | Subject) + storage_time
y ~ current_feature + Sex + (1 | Subject) + storage_time
y ~ current_feature + Sex + (1 | Subject) + storage_time
y ~ current_feature + Sex + (1 | Subject) + storage_time
y ~ current_feature + Sex + (1 | Subject) + storage_time
y ~ current_feature + Sex + (1 | Subject) + storage_time
y ~ current_feature + Sex + (1 | Subject) + storage_time
y ~ current_feature + Sex + (1 | Subject) + storage_time
y ~ current_feature + Sex + (1 | Subject) + storage_time
y ~ current_feature + Sex + (1 | Subject) + storage_time
y ~ current_feature + Sex + (1 | Subject) + storage_time
y ~ current_feature + Sex + (1 | Subject) + storage_time
y ~ current_feature + Sex + (1 | Subject) + storage_time
y ~ current_feature + Sex + (1 | Subject) + storage_time
y ~ current_feature + Sex + (1 | Subject) + storage_time
y ~ current_feature + Sex + (1 | Subject) + storage_time
y ~ current_feature + Sex + (1 | Subject) + storage_time
y ~ current_feature + Sex + (1 | Subject) + storage_time
y ~ current_feature + Sex + (1 | Subject) + storage_time
y ~ current_feature + Sex + (1 | Subject) + storage_time
y ~ current_feature + Sex + (1 | Subject) + storage_time
y ~ current_feature + Sex + (1 | Subject) + storage_time
y ~ current_feature + Sex + (1 | Subject) + storage_time
y ~ current_feature + Sex + (1 | Subject) + storage_time
y ~ current_feature + Sex + (1 | Subject) + storage_time
y ~ current_feature + Sex + (1 | Subject) + storage_time
y ~ current_feature + Sex + (1 | Subject) + storage_time
y ~ current_feature + Sex + (1 | Subject) + storage_time
y ~ current_feature + Sex + (1 | Subject) + storage_time
y ~ current_feature + Sex + (1 | Subject) + storage_time
y ~ current_feature + Sex + (1 | Subject) + storage_time
y ~ current_feature + Sex + (1 | Subject) + storage_time
y ~ current_feature + Sex + (1 | Subject) + storage_time
y ~ current_feature + Sex + (1 | Subject) + storage_time
y ~ current_feature + Sex + (1 | Subject) + storage_time
y ~ current_feature + Sex + (1 | Subject) + storage_time
y ~ current_feature + Sex + (1 | Subject) + storage_time
y ~ current_feature + Sex + (1 | Subject) + storage_time
y ~ current_feature + Sex + (1 | Subject) + storage_time
y ~ current_feature + Sex + (1 | Subject) + storage_time
y ~ current_feature + Sex + (1 | Subject) + storage_time
y ~ current_feature + Sex + (1 | Subject) + storage_time
y ~ current_feature + Sex + (1 | Subject) + storage_time
y ~ current_feature + Sex + (1 | Subject) + storage_time

y ~ current_feature + Sex + (1 | Subject) + storage_time
y ~ current_feature + Sex + (1 | Subject) + storage_time
y ~ current_feature + Sex + (1 | Subject) + storage_time
y ~ current_feature + Sex + (1 | Subject) + storage_time
y ~ current_feature + Sex + (1 | Subject) + storage_time
y ~ current_feature + Sex + (1 | Subject) + storage_time
y ~ current_feature + Sex + (1 | Subject) + storage_time
y ~ current_feature + Sex + (1 | Subject) + storage_time
y ~ current_feature + Sex + (1 | Subject) + storage_time
y ~ current_feature + Sex + (1 | Subject) + storage_time
y ~ current_feature + Sex + (1 | Subject) + storage_time
y ~ current_feature + Sex + (1 | Subject) + storage_time
y ~ current_feature + Sex + (1 | Subject) + storage_time
y ~ current_feature + Sex + (1 | Subject) + storage_time
y ~ current_feature + Sex + (1 | Subject) + storage_time
y ~ current_feature + Sex + (1 | Subject) + storage_time
y ~ current_feature + Sex + (1 | Subject) + storage_time
y ~ current_feature + Sex + (1 | Subject) + storage_time
y ~ current_feature + Sex + (1 | Subject) + storage_time
y ~ current_feature + Sex + (1 | Subject) + storage_time
y ~ current_feature + Sex + (1 | Subject) + storage_time
y ~ current_feature + Sex + (1 | Subject) + storage_time
y ~ current_feature + Sex + (1 | Subject) + storage_time
y ~ current_feature + Sex + (1 | Subject) + storage_time
y ~ current_feature + Sex + (1 | Subject) + storage_time
y ~ current_feature + Sex + (1 | Subject) + storage_time
y ~ current_feature + Sex + (1 | Subject) + storage_time
y ~ current_feature + Sex + (1 | Subject) + storage_time
y ~ current_feature + Sex + (1 | Subject) + storage_time
y ~ current_feature + Sex + (1 | Subject) + storage_time
y ~ current_feature + Sex + (1 | Subject) + storage_time
y ~ current_feature + Sex + (1 | Subject) + storage_time
y ~ current_feature + Sex + (1 | Subject) + storage_time
y ~ current_feature + Sex + (1 | Subject) + storage_time
y ~ current_feature + Sex + (1 | Subject) + storage_time
y ~ current_feature + Sex + (1 | Subject) + storage_time
y ~ current_feature + Sex + (1 | Subject) + storage_time
y ~ current_feature + Sex + (1 | Subject) + storage_time
y ~ current_feature + Sex + (1 | Subject) + storage_time
y ~ current_feature + Sex + (1 | Subject) + storage_time
y ~ current_feature + Sex + (1 | Subject) + storage_time
y ~ current_feature + Sex + (1 | Subject) + storage_time
y ~ current_feature + Sex + (1 | Subject) + storage_time
y ~ current_feature + Sex + (1 | Subject) + storage_time
y ~ current_feature + Sex + (1 | Subject) + storage_time
y ~ current_feature + Sex + (1 | Subject) + storage_time
y ~ current_feature + Sex + (1 | Subject) + storage_time
y ~ current_feature + Sex + (1 | Subject) + storage_time

```
y ~ current_feature + Sex + (1 | Subject) + storage_time
y ~ current_feature + Sex + (1 | Subject) + storage_time
y ~ current_feature + Sex + (1 | Subject) + storage_time
y ~ current_feature + Sex + (1 | Subject) + storage_time
y ~ current_feature + Sex + (1 | Subject) + storage_time
y ~ current_feature + Sex + (1 | Subject) + storage_time
y ~ current_feature + Sex + (1 | Subject) + storage_time
y ~ current_feature + Sex + (1 | Subject) + storage_time
y ~ current_feature + Sex + (1 | Subject) + storage_time
y ~ current_feature + Sex + (1 | Subject) + storage_time
y ~ current_feature + Sex + (1 | Subject) + storage_time
y ~ current_feature + Sex + (1 | Subject) + storage_time
y ~ current_feature + Sex + (1 | Subject) + storage_time
y ~ current_feature + Sex + (1 | Subject) + storage_time
y ~ current_feature + Sex + (1 | Subject) + storage_time
y ~ current_feature + Sex + (1 | Subject) + storage_time
y ~ current_feature + Sex + (1 | Subject) + storage_time
y ~ current_feature + Sex + (1 | Subject) + storage_time
y ~ current_feature + Sex + (1 | Subject) + storage_time
y ~ current_feature + Sex + (1 | Subject) + storage_time
y ~ current_feature + Sex + (1 | Subject) + storage_time
y ~ current_feature + Sex + (1 | Subject) + storage_time
y ~ current_feature + Sex + (1 | Subject) + storage_time
y ~ current_feature + Sex + (1 | Subject) + storage_time
y ~ current_feature + Sex + (1 | Subject) + storage_time
y ~ current_feature + Sex + (1 | Subject) + storage_time
y ~ current_feature + Sex + (1 | Subject) + storage_time
y ~ current_feature + Sex + (1 | Subject) + storage_time
y ~ current_feature + Sex + (1 | Subject) + storage_time
y ~ current_feature + Sex + (1 | Subject) + storage_time
y ~ current_feature + Sex + (1 | Subject) + storage_time
y ~ current_feature + Sex + (1 | Subject) + storage_time
y ~ current_feature + Sex + (1 | Subject) + storage_time
y ~ current_feature + Sex + (1 | Subject) + storage_time
y ~ current_feature + Sex + (1 | Subject) + storage_time
y ~ current_feature + Sex + (1 | Subject) + storage_time
y ~ current_feature + Sex + (1 | Subject) + storage_time
y ~ current_feature + Sex + (1 | Subject) + storage_time
y ~ current_feature + Sex + (1 | Subject) + storage_time
y ~ current_feature + Sex + (1 | Subject) + storage_time
y ~ current_feature + Sex + (1 | Subject) + storage_time
y ~ current_feature + Sex + (1 | Subject) + storage_time
y ~ current_feature + Sex + (1 | Subject) + storage_time
y ~ current_feature + Sex + (1 | Subject) + storage_time
y ~ current_feature + Sex + (1 | Subject) + storage_time
y ~ current_feature + Sex + (1 | Subject) + storage_time
y ~ current_feature + Sex + (1 | Subject) + storage_time
y ~ current_feature + Sex + (1 | Subject) + storage_time
```

y ~ current_feature + Sex + (1 | Subject) + storage_time
y ~ current_feature + Sex + (1 | Subject) + storage_time
y ~ current_feature + Sex + (1 | Subject) + storage_time
y ~ current_feature + Sex + (1 | Subject) + storage_time
y ~ current_feature + Sex + (1 | Subject) + storage_time
y ~ current_feature + Sex + (1 | Subject) + storage_time
y ~ current_feature + Sex + (1 | Subject) + storage_time
y ~ current_feature + Sex + (1 | Subject) + storage_time
y ~ current_feature + Sex + (1 | Subject) + storage_time
y ~ current_feature + Sex + (1 | Subject) + storage_time
y ~ current_feature + Sex + (1 | Subject) + storage_time
y ~ current_feature + Sex + (1 | Subject) + storage_time
y ~ current_feature + Sex + (1 | Subject) + storage_time
y ~ current_feature + Sex + (1 | Subject) + storage_time
y ~ current_feature + Sex + (1 | Subject) + storage_time
y ~ current_feature + Sex + (1 | Subject) + storage_time
y ~ current_feature + Sex + (1 | Subject) + storage_time
y ~ current_feature + Sex + (1 | Subject) + storage_time
y ~ current_feature + Sex + (1 | Subject) + storage_time
y ~ current_feature + Sex + (1 | Subject) + storage_time
y ~ current_feature + Sex + (1 | Subject) + storage_time
y ~ current_feature + Sex + (1 | Subject) + storage_time
y ~ current_feature + Sex + (1 | Subject) + storage_time
y ~ current_feature + Sex + (1 | Subject) + storage_time
y ~ current_feature + Sex + (1 | Subject) + storage_time
y ~ current_feature + Sex + (1 | Subject) + storage_time
y ~ current_feature + Sex + (1 | Subject) + storage_time
y ~ current_feature + Sex + (1 | Subject) + storage_time
y ~ current_feature + Sex + (1 | Subject) + storage_time
y ~ current_feature + Sex + (1 | Subject) + storage_time
y ~ current_feature + Sex + (1 | Subject) + storage_time
y ~ current_feature + Sex + (1 | Subject) + storage_time
y ~ current_feature + Sex + (1 | Subject) + storage_time
y ~ current_feature + Sex + (1 | Subject) + storage_time
y ~ current_feature + Sex + (1 | Subject) + storage_time
y ~ current_feature + Sex + (1 | Subject) + storage_time
y ~ current_feature + Sex + (1 | Subject) + storage_time
y ~ current_feature + Sex + (1 | Subject) + storage_time
y ~ current_feature + Sex + (1 | Subject) + storage_time
y ~ current_feature + Sex + (1 | Subject) + storage_time
y ~ current_feature + Sex + (1 | Subject) + storage_time
y ~ current_feature + Sex + (1 | Subject) + storage_time
y ~ current_feature + Sex + (1 | Subject) + storage_time
y ~ current_feature + Sex + (1 | Subject) + storage_time
y ~ current_feature + Sex + (1 | Subject) + storage_time
y ~ current_feature + Sex + (1 | Subject) + storage_time
y ~ current_feature + Sex + (1 | Subject) + storage_time
y ~ current_feature + Sex + (1 | Subject) + storage_time

y ~ current_feature + Sex + (1 | Subject) + storage_time
y ~ current_feature + Sex + (1 | Subject) + storage_time
y ~ current_feature + Sex + (1 | Subject) + storage_time
y ~ current_feature + Sex + (1 | Subject) + storage_time
y ~ current_feature + Sex + (1 | Subject) + storage_time
y ~ current_feature + Sex + (1 | Subject) + storage_time
y ~ current_feature + Sex + (1 | Subject) + storage_time
y ~ current_feature + Sex + (1 | Subject) + storage_time
y ~ current_feature + Sex + (1 | Subject) + storage_time
y ~ current_feature + Sex + (1 | Subject) + storage_time
y ~ current_feature + Sex + (1 | Subject) + storage_time
y ~ current_feature + Sex + (1 | Subject) + storage_time
y ~ current_feature + Sex + (1 | Subject) + storage_time
y ~ current_feature + Sex + (1 | Subject) + storage_time
y ~ current_feature + Sex + (1 | Subject) + storage_time
y ~ current_feature + Sex + (1 | Subject) + storage_time
y ~ current_feature + Sex + (1 | Subject) + storage_time
y ~ current_feature + Sex + (1 | Subject) + storage_time
y ~ current_feature + Sex + (1 | Subject) + storage_time
y ~ current_feature + Sex + (1 | Subject) + storage_time
y ~ current_feature + Sex + (1 | Subject) + storage_time
y ~ current_feature + Sex + (1 | Subject) + storage_time
y ~ current_feature + Sex + (1 | Subject) + storage_time
y ~ current_feature + Sex + (1 | Subject) + storage_time
y ~ current_feature + Sex + (1 | Subject) + storage_time
y ~ current_feature + Sex + (1 | Subject) + storage_time
y ~ current_feature + Sex + (1 | Subject) + storage_time
y ~ current_feature + Sex + (1 | Subject) + storage_time
y ~ current_feature + Sex + (1 | Subject) + storage_time
y ~ current_feature + Sex + (1 | Subject) + storage_time
y ~ current_feature + Sex + (1 | Subject) + storage_time
y ~ current_feature + Sex + (1 | Subject) + storage_time
y ~ current_feature + Sex + (1 | Subject) + storage_time
y ~ current_feature + Sex + (1 | Subject) + storage_time
y ~ current_feature + Sex + (1 | Subject) + storage_time
y ~ current_feature + Sex + (1 | Subject) + storage_time
y ~ current_feature + Sex + (1 | Subject) + storage_time
y ~ current_feature + Sex + (1 | Subject) + storage_time
y ~ current_feature + Sex + (1 | Subject) + storage_time
y ~ current_feature + Sex + (1 | Subject) + storage_time
y ~ current_feature + Sex + (1 | Subject) + storage_time
y ~ current_feature + Sex + (1 | Subject) + storage_time
y ~ current_feature + Sex + (1 | Subject) + storage_time
y ~ current_feature + Sex + (1 | Subject) + storage_time
y ~ current_feature + Sex + (1 | Subject) + storage_time
y ~ current_feature + Sex + (1 | Subject) + storage_time
y ~ current_feature + Sex + (1 | Subject) + storage_time
y ~ current_feature + Sex + (1 | Subject) + storage_time

y ~ current_feature + Sex + (1 | Subject) + storage_time
y ~ current_feature + Sex + (1 | Subject) + storage_time
y ~ current_feature + Sex + (1 | Subject) + storage_time
y ~ current_feature + Sex + (1 | Subject) + storage_time
y ~ current_feature + Sex + (1 | Subject) + storage_time
y ~ current_feature + Sex + (1 | Subject) + storage_time
y ~ current_feature + Sex + (1 | Subject) + storage_time
y ~ current_feature + Sex + (1 | Subject) + storage_time
y ~ current_feature + Sex + (1 | Subject) + storage_time
y ~ current_feature + Sex + (1 | Subject) + storage_time
y ~ current_feature + Sex + (1 | Subject) + storage_time
y ~ current_feature + Sex + (1 | Subject) + storage_time
y ~ current_feature + Sex + (1 | Subject) + storage_time
y ~ current_feature + Sex + (1 | Subject) + storage_time
y ~ current_feature + Sex + (1 | Subject) + storage_time
y ~ current_feature + Sex + (1 | Subject) + storage_time
y ~ current_feature + Sex + (1 | Subject) + storage_time
y ~ current_feature + Sex + (1 | Subject) + storage_time
y ~ current_feature + Sex + (1 | Subject) + storage_time
y ~ current_feature + Sex + (1 | Subject) + storage_time
y ~ current_feature + Sex + (1 | Subject) + storage_time
y ~ current_feature + Sex + (1 | Subject) + storage_time
y ~ current_feature + Sex + (1 | Subject) + storage_time
y ~ current_feature + Sex + (1 | Subject) + storage_time
y ~ current_feature + Sex + (1 | Subject) + storage_time
y ~ current_feature + Sex + (1 | Subject) + storage_time
y ~ current_feature + Sex + (1 | Subject) + storage_time
y ~ current_feature + Sex + (1 | Subject) + storage_time
y ~ current_feature + Sex + (1 | Subject) + storage_time
y ~ current_feature + Sex + (1 | Subject) + storage_time
y ~ current_feature + Sex + (1 | Subject) + storage_time
y ~ current_feature + Sex + (1 | Subject) + storage_time
y ~ current_feature + Sex + (1 | Subject) + storage_time
y ~ current_feature + Sex + (1 | Subject) + storage_time
y ~ current_feature + Sex + (1 | Subject) + storage_time
y ~ current_feature + Sex + (1 | Subject) + storage_time
y ~ current_feature + Sex + (1 | Subject) + storage_time
y ~ current_feature + Sex + (1 | Subject) + storage_time
y ~ current_feature + Sex + (1 | Subject) + storage_time
y ~ current_feature + Sex + (1 | Subject) + storage_time
y ~ current_feature + Sex + (1 | Subject) + storage_time
y ~ current_feature + Sex + (1 | Subject) + storage_time
y ~ current_feature + Sex + (1 | Subject) + storage_time
y ~ current_feature + Sex + (1 | Subject) + storage_time
y ~ current_feature + Sex + (1 | Subject) + storage_time
y ~ current_feature + Sex + (1 | Subject) + storage_time
y ~ current_feature + Sex + (1 | Subject) + storage_time
y ~ current_feature + Sex + (1 | Subject) + storage_time

y ~ current_feature + Sex + (1 | Subject) + storage_time
y ~ current_feature + Sex + (1 | Subject) + storage_time
y ~ current_feature + Sex + (1 | Subject) + storage_time
y ~ current_feature + Sex + (1 | Subject) + storage_time
y ~ current_feature + Sex + (1 | Subject) + storage_time
y ~ current_feature + Sex + (1 | Subject) + storage_time
y ~ current_feature + Sex + (1 | Subject) + storage_time
y ~ current_feature + Sex + (1 | Subject) + storage_time
y ~ current_feature + Sex + (1 | Subject) + storage_time
y ~ current_feature + Sex + (1 | Subject) + storage_time
y ~ current_feature + Sex + (1 | Subject) + storage_time
y ~ current_feature + Sex + (1 | Subject) + storage_time
y ~ current_feature + Sex + (1 | Subject) + storage_time
y ~ current_feature + Sex + (1 | Subject) + storage_time
y ~ current_feature + Sex + (1 | Subject) + storage_time
y ~ current_feature + Sex + (1 | Subject) + storage_time
y ~ current_feature + Sex + (1 | Subject) + storage_time
y ~ current_feature + Sex + (1 | Subject) + storage_time
y ~ current_feature + Sex + (1 | Subject) + storage_time
y ~ current_feature + Sex + (1 | Subject) + storage_time
y ~ current_feature + Sex + (1 | Subject) + storage_time
y ~ current_feature + Sex + (1 | Subject) + storage_time
y ~ current_feature + Sex + (1 | Subject) + storage_time
y ~ current_feature + Sex + (1 | Subject) + storage_time
y ~ current_feature + Sex + (1 | Subject) + storage_time
y ~ current_feature + Sex + (1 | Subject) + storage_time
y ~ current_feature + Sex + (1 | Subject) + storage_time
y ~ current_feature + Sex + (1 | Subject) + storage_time
y ~ current_feature + Sex + (1 | Subject) + storage_time
y ~ current_feature + Sex + (1 | Subject) + storage_time
y ~ current_feature + Sex + (1 | Subject) + storage_time
y ~ current_feature + Sex + (1 | Subject) + storage_time
y ~ current_feature + Sex + (1 | Subject) + storage_time
y ~ current_feature + Sex + (1 | Subject) + storage_time
y ~ current_feature + Sex + (1 | Subject) + storage_time
y ~ current_feature + Sex + (1 | Subject) + storage_time
y ~ current_feature + Sex + (1 | Subject) + storage_time
y ~ current_feature + Sex + (1 | Subject) + storage_time
y ~ current_feature + Sex + (1 | Subject) + storage_time
y ~ current_feature + Sex + (1 | Subject) + storage_time
y ~ current_feature + Sex + (1 | Subject) + storage_time
y ~ current_feature + Sex + (1 | Subject) + storage_time
y ~ current_feature + Sex + (1 | Subject) + storage_time
y ~ current_feature + Sex + (1 | Subject) + storage_time
y ~ current_feature + Sex + (1 | Subject) + storage_time
y ~ current_feature + Sex + (1 | Subject) + storage_time
y ~ current_feature + Sex + (1 | Subject) + storage_time
y ~ current_feature + Sex + (1 | Subject) + storage_time

y ~ current_feature + Sex + (1 | Subject) + storage_time
y ~ current_feature + Sex + (1 | Subject) + storage_time
y ~ current_feature + Sex + (1 | Subject) + storage_time
y ~ current_feature + Sex + (1 | Subject) + storage_time
y ~ current_feature + Sex + (1 | Subject) + storage_time
y ~ current_feature + Sex + (1 | Subject) + storage_time
y ~ current_feature + Sex + (1 | Subject) + storage_time
y ~ current_feature + Sex + (1 | Subject) + storage_time
y ~ current_feature + Sex + (1 | Subject) + storage_time
y ~ current_feature + Sex + (1 | Subject) + storage_time
y ~ current_feature + Sex + (1 | Subject) + storage_time
y ~ current_feature + Sex + (1 | Subject) + storage_time
y ~ current_feature + Sex + (1 | Subject) + storage_time
y ~ current_feature + Sex + (1 | Subject) + storage_time
y ~ current_feature + Sex + (1 | Subject) + storage_time
y ~ current_feature + Sex + (1 | Subject) + storage_time
y ~ current_feature + Sex + (1 | Subject) + storage_time
y ~ current_feature + Sex + (1 | Subject) + storage_time
y ~ current_feature + Sex + (1 | Subject) + storage_time
y ~ current_feature + Sex + (1 | Subject) + storage_time
y ~ current_feature + Sex + (1 | Subject) + storage_time
y ~ current_feature + Sex + (1 | Subject) + storage_time
y ~ current_feature + Sex + (1 | Subject) + storage_time
y ~ current_feature + Sex + (1 | Subject) + storage_time
y ~ current_feature + Sex + (1 | Subject) + storage_time
y ~ current_feature + Sex + (1 | Subject) + storage_time
y ~ current_feature + Sex + (1 | Subject) + storage_time
y ~ current_feature + Sex + (1 | Subject) + storage_time
y ~ current_feature + Sex + (1 | Subject) + storage_time
y ~ current_feature + Sex + (1 | Subject) + storage_time
y ~ current_feature + Sex + (1 | Subject) + storage_time
y ~ current_feature + Sex + (1 | Subject) + storage_time
y ~ current_feature + Sex + (1 | Subject) + storage_time
y ~ current_feature + Sex + (1 | Subject) + storage_time
y ~ current_feature + Sex + (1 | Subject) + storage_time
y ~ current_feature + Sex + (1 | Subject) + storage_time
y ~ current_feature + Sex + (1 | Subject) + storage_time
y ~ current_feature + Sex + (1 | Subject) + storage_time
y ~ current_feature + Sex + (1 | Subject) + storage_time
y ~ current_feature + Sex + (1 | Subject) + storage_time
y ~ current_feature + Sex + (1 | Subject) + storage_time
y ~ current_feature + Sex + (1 | Subject) + storage_time
y ~ current_feature + Sex + (1 | Subject) + storage_time
y ~ current_feature + Sex + (1 | Subject) + storage_time
y ~ current_feature + Sex + (1 | Subject) + storage_time
y ~ current_feature + Sex + (1 | Subject) + storage_time
y ~ current_feature + Sex + (1 | Subject) + storage_time
y ~ current_feature + Sex + (1 | Subject) + storage_time

```
y ~ current_feature + Sex + (1 | Subject) + storage_time
y ~ current_feature + Sex + (1 | Subject) + storage_time
y ~ current_feature + Sex + (1 | Subject) + storage_time
y ~ current_feature + Sex + (1 | Subject) + storage_time
y ~ current_feature + Sex + (1 | Subject) + storage_time
y ~ current_feature + Sex + (1 | Subject) + storage_time
y ~ current_feature + Sex + (1 | Subject) + storage_time
y ~ current_feature + Sex + (1 | Subject) + storage_time
y ~ current_feature + Sex + (1 | Subject) + storage_time
y ~ current_feature + Sex + (1 | Subject) + storage_time
y ~ current_feature + Sex + (1 | Subject) + storage_time
y ~ current_feature + Sex + (1 | Subject) + storage_time
y ~ current_feature + Sex + (1 | Subject) + storage_time
y ~ current_feature + Sex + (1 | Subject) + storage_time
y ~ current_feature + Sex + (1 | Subject) + storage_time
y ~ current_feature + Sex + (1 | Subject) + storage_time
y ~ current_feature + Sex + (1 | Subject) + storage_time
y ~ current_feature + Sex + (1 | Subject) + storage_time
y ~ current_feature + Sex + (1 | Subject) + storage_time
y ~ current_feature + Sex + (1 | Subject) + storage_time
y ~ current_feature + Sex + (1 | Subject) + storage_time
y ~ current_feature + Sex + (1 | Subject) + storage_time
y ~ current_feature + Sex + (1 | Subject) + storage_time
y ~ current_feature + Sex + (1 | Subject) + storage_time
y ~ current_feature + Sex + (1 | Subject) + storage_time
y ~ current_feature + Sex + (1 | Subject) + storage_time
y ~ current_feature + Sex + (1 | Subject) + storage_time
y ~ current_feature + Sex + (1 | Subject) + storage_time
y ~ current_feature + Sex + (1 | Subject) + storage_time
y ~ current_feature + Sex + (1 | Subject) + storage_time
y ~ current_feature + Sex + (1 | Subject) + storage_time
y ~ current_feature + Sex + (1 | Subject) + storage_time
y ~ current_feature + Sex + (1 | Subject) + storage_time
y ~ current_feature + Sex + (1 | Subject) + storage_time
y ~ current_feature + Sex + (1 | Subject) + storage_time
y ~ current_feature + Sex + (1 | Subject) + storage_time
y ~ current_feature + Sex + (1 | Subject) + storage_time
y ~ current_feature + Sex + (1 | Subject) + storage_time
y ~ current_feature + Sex + (1 | Subject) + storage_time
y ~ current_feature + Sex + (1 | Subject) + storage_time
y ~ current_feature + Sex + (1 | Subject) + storage_time
y ~ current_feature + Sex + (1 | Subject) + storage_time
y ~ current_feature + Sex + (1 | Subject) + storage_time
y ~ current_feature + Sex + (1 | Subject) + storage_time
y ~ current_feature + Sex + (1 | Subject) + storage_time
y ~ current_feature + Sex + (1 | Subject) + storage_time
y ~ current_feature + Sex + (1 | Subject) + storage_time
y ~ current_feature + Sex + (1 | Subject) + storage_time
```

y ~ current_feature + Sex + (1 | Subject) + storage_time
y ~ current_feature + Sex + (1 | Subject) + storage_time
y ~ current_feature + Sex + (1 | Subject) + storage_time
y ~ current_feature + Sex + (1 | Subject) + storage_time
y ~ current_feature + Sex + (1 | Subject) + storage_time
y ~ current_feature + Sex + (1 | Subject) + storage_time
y ~ current_feature + Sex + (1 | Subject) + storage_time
y ~ current_feature + Sex + (1 | Subject) + storage_time
y ~ current_feature + Sex + (1 | Subject) + storage_time
y ~ current_feature + Sex + (1 | Subject) + storage_time
y ~ current_feature + Sex + (1 | Subject) + storage_time
y ~ current_feature + Sex + (1 | Subject) + storage_time
y ~ current_feature + Sex + (1 | Subject) + storage_time
y ~ current_feature + Sex + (1 | Subject) + storage_time
y ~ current_feature + Sex + (1 | Subject) + storage_time
y ~ current_feature + Sex + (1 | Subject) + storage_time
y ~ current_feature + Sex + (1 | Subject) + storage_time
y ~ current_feature + Sex + (1 | Subject) + storage_time
y ~ current_feature + Sex + (1 | Subject) + storage_time
y ~ current_feature + Sex + (1 | Subject) + storage_time
y ~ current_feature + Sex + (1 | Subject) + storage_time
y ~ current_feature + Sex + (1 | Subject) + storage_time
y ~ current_feature + Sex + (1 | Subject) + storage_time
y ~ current_feature + Sex + (1 | Subject) + storage_time
y ~ current_feature + Sex + (1 | Subject) + storage_time
y ~ current_feature + Sex + (1 | Subject) + storage_time
y ~ current_feature + Sex + (1 | Subject) + storage_time
y ~ current_feature + Sex + (1 | Subject) + storage_time
y ~ current_feature + Sex + (1 | Subject) + storage_time
y ~ current_feature + Sex + (1 | Subject) + storage_time
y ~ current_feature + Sex + (1 | Subject) + storage_time
y ~ current_feature + Sex + (1 | Subject) + storage_time
y ~ current_feature + Sex + (1 | Subject) + storage_time
y ~ current_feature + Sex + (1 | Subject) + storage_time
y ~ current_feature + Sex + (1 | Subject) + storage_time
y ~ current_feature + Sex + (1 | Subject) + storage_time
y ~ current_feature + Sex + (1 | Subject) + storage_time
y ~ current_feature + Sex + (1 | Subject) + storage_time
y ~ current_feature + Sex + (1 | Subject) + storage_time
y ~ current_feature + Sex + (1 | Subject) + storage_time
y ~ current_feature + Sex + (1 | Subject) + storage_time
y ~ current_feature + Sex + (1 | Subject) + storage_time
y ~ current_feature + Sex + (1 | Subject) + storage_time
y ~ current_feature + Sex + (1 | Subject) + storage_time
y ~ current_feature + Sex + (1 | Subject) + storage_time
y ~ current_feature + Sex + (1 | Subject) + storage_time
y ~ current_feature + Sex + (1 | Subject) + storage_time
y ~ current_feature + Sex + (1 | Subject) + storage_time

y ~ current_feature + Sex + (1 | Subject) + storage_time
y ~ current_feature + Sex + (1 | Subject) + storage_time
y ~ current_feature + Sex + (1 | Subject) + storage_time
y ~ current_feature + Sex + (1 | Subject) + storage_time
y ~ current_feature + Sex + (1 | Subject) + storage_time
y ~ current_feature + Sex + (1 | Subject) + storage_time
y ~ current_feature + Sex + (1 | Subject) + storage_time
y ~ current_feature + Sex + (1 | Subject) + storage_time
y ~ current_feature + Sex + (1 | Subject) + storage_time
y ~ current_feature + Sex + (1 | Subject) + storage_time
y ~ current_feature + Sex + (1 | Subject) + storage_time
y ~ current_feature + Sex + (1 | Subject) + storage_time
y ~ current_feature + Sex + (1 | Subject) + storage_time
y ~ current_feature + Sex + (1 | Subject) + storage_time
y ~ current_feature + Sex + (1 | Subject) + storage_time
y ~ current_feature + Sex + (1 | Subject) + storage_time
y ~ current_feature + Sex + (1 | Subject) + storage_time
y ~ current_feature + Sex + (1 | Subject) + storage_time
y ~ current_feature + Sex + (1 | Subject) + storage_time
y ~ current_feature + Sex + (1 | Subject) + storage_time
y ~ current_feature + Sex + (1 | Subject) + storage_time
y ~ current_feature + Sex + (1 | Subject) + storage_time
y ~ current_feature + Sex + (1 | Subject) + storage_time
y ~ current_feature + Sex + (1 | Subject) + storage_time
y ~ current_feature + Sex + (1 | Subject) + storage_time
y ~ current_feature + Sex + (1 | Subject) + storage_time
y ~ current_feature + Sex + (1 | Subject) + storage_time
y ~ current_feature + Sex + (1 | Subject) + storage_time
y ~ current_feature + Sex + (1 | Subject) + storage_time
y ~ current_feature + Sex + (1 | Subject) + storage_time
y ~ current_feature + Sex + (1 | Subject) + storage_time
y ~ current_feature + Sex + (1 | Subject) + storage_time
y ~ current_feature + Sex + (1 | Subject) + storage_time
y ~ current_feature + Sex + (1 | Subject) + storage_time
y ~ current_feature + Sex + (1 | Subject) + storage_time
y ~ current_feature + Sex + (1 | Subject) + storage_time
y ~ current_feature + Sex + (1 | Subject) + storage_time
y ~ current_feature + Sex + (1 | Subject) + storage_time
y ~ current_feature + Sex + (1 | Subject) + storage_time
y ~ current_feature + Sex + (1 | Subject) + storage_time
y ~ current_feature + Sex + (1 | Subject) + storage_time
y ~ current_feature + Sex + (1 | Subject) + storage_time
y ~ current_feature + Sex + (1 | Subject) + storage_time
y ~ current_feature + Sex + (1 | Subject) + storage_time
y ~ current_feature + Sex + (1 | Subject) + storage_time
y ~ current_feature + Sex + (1 | Subject) + storage_time
y ~ current_feature + Sex + (1 | Subject) + storage_time
y ~ current_feature + Sex + (1 | Subject) + storage_time

y ~ current_feature + Sex + (1 | Subject) + storage_time
y ~ current_feature + Sex + (1 | Subject) + storage_time
y ~ current_feature + Sex + (1 | Subject) + storage_time
y ~ current_feature + Sex + (1 | Subject) + storage_time
y ~ current_feature + Sex + (1 | Subject) + storage_time
y ~ current_feature + Sex + (1 | Subject) + storage_time
y ~ current_feature + Sex + (1 | Subject) + storage_time
y ~ current_feature + Sex + (1 | Subject) + storage_time
y ~ current_feature + Sex + (1 | Subject) + storage_time
y ~ current_feature + Sex + (1 | Subject) + storage_time
y ~ current_feature + Sex + (1 | Subject) + storage_time
y ~ current_feature + Sex + (1 | Subject) + storage_time
y ~ current_feature + Sex + (1 | Subject) + storage_time
y ~ current_feature + Sex + (1 | Subject) + storage_time
y ~ current_feature + Sex + (1 | Subject) + storage_time
y ~ current_feature + Sex + (1 | Subject) + storage_time
y ~ current_feature + Sex + (1 | Subject) + storage_time
y ~ current_feature + Sex + (1 | Subject) + storage_time
y ~ current_feature + Sex + (1 | Subject) + storage_time
y ~ current_feature + Sex + (1 | Subject) + storage_time
y ~ current_feature + Sex + (1 | Subject) + storage_time
y ~ current_feature + Sex + (1 | Subject) + storage_time
y ~ current_feature + Sex + (1 | Subject) + storage_time
y ~ current_feature + Sex + (1 | Subject) + storage_time
y ~ current_feature + Sex + (1 | Subject) + storage_time
y ~ current_feature + Sex + (1 | Subject) + storage_time
y ~ current_feature + Sex + (1 | Subject) + storage_time
y ~ current_feature + Sex + (1 | Subject) + storage_time
y ~ current_feature + Sex + (1 | Subject) + storage_time
y ~ current_feature + Sex + (1 | Subject) + storage_time
y ~ current_feature + Sex + (1 | Subject) + storage_time
y ~ current_feature + Sex + (1 | Subject) + storage_time
y ~ current_feature + Sex + (1 | Subject) + storage_time
y ~ current_feature + Sex + (1 | Subject) + storage_time
y ~ current_feature + Sex + (1 | Subject) + storage_time
y ~ current_feature + Sex + (1 | Subject) + storage_time
y ~ current_feature + Sex + (1 | Subject) + storage_time
y ~ current_feature + Sex + (1 | Subject) + storage_time
y ~ current_feature + Sex + (1 | Subject) + storage_time
y ~ current_feature + Sex + (1 | Subject) + storage_time
y ~ current_feature + Sex + (1 | Subject) + storage_time
y ~ current_feature + Sex + (1 | Subject) + storage_time
y ~ current_feature + Sex + (1 | Subject) + storage_time
y ~ current_feature + Sex + (1 | Subject) + storage_time
y ~ current_feature + Sex + (1 | Subject) + storage_time
y ~ current_feature + Sex + (1 | Subject) + storage_time
y ~ current_feature + Sex + (1 | Subject) + storage_time
y ~ current_feature + Sex + (1 | Subject) + storage_time

y ~ current_feature + Sex + (1 | Subject) + storage_time
y ~ current_feature + Sex + (1 | Subject) + storage_time
y ~ current_feature + Sex + (1 | Subject) + storage_time
y ~ current_feature + Sex + (1 | Subject) + storage_time
y ~ current_feature + Sex + (1 | Subject) + storage_time
y ~ current_feature + Sex + (1 | Subject) + storage_time
y ~ current_feature + Sex + (1 | Subject) + storage_time
y ~ current_feature + Sex + (1 | Subject) + storage_time
y ~ current_feature + Sex + (1 | Subject) + storage_time
y ~ current_feature + Sex + (1 | Subject) + storage_time
y ~ current_feature + Sex + (1 | Subject) + storage_time
y ~ current_feature + Sex + (1 | Subject) + storage_time
y ~ current_feature + Sex + (1 | Subject) + storage_time
y ~ current_feature + Sex + (1 | Subject) + storage_time
y ~ current_feature + Sex + (1 | Subject) + storage_time
y ~ current_feature + Sex + (1 | Subject) + storage_time
y ~ current_feature + Sex + (1 | Subject) + storage_time
y ~ current_feature + Sex + (1 | Subject) + storage_time
y ~ current_feature + Sex + (1 | Subject) + storage_time
y ~ current_feature + Sex + (1 | Subject) + storage_time
y ~ current_feature + Sex + (1 | Subject) + storage_time
y ~ current_feature + Sex + (1 | Subject) + storage_time
y ~ current_feature + Sex + (1 | Subject) + storage_time
y ~ current_feature + Sex + (1 | Subject) + storage_time
y ~ current_feature + Sex + (1 | Subject) + storage_time
y ~ current_feature + Sex + (1 | Subject) + storage_time
y ~ current_feature + Sex + (1 | Subject) + storage_time
y ~ current_feature + Sex + (1 | Subject) + storage_time
y ~ current_feature + Sex + (1 | Subject) + storage_time
y ~ current_feature + Sex + (1 | Subject) + storage_time
y ~ current_feature + Sex + (1 | Subject) + storage_time
y ~ current_feature + Sex + (1 | Subject) + storage_time
y ~ current_feature + Sex + (1 | Subject) + storage_time
y ~ current_feature + Sex + (1 | Subject) + storage_time
y ~ current_feature + Sex + (1 | Subject) + storage_time
y ~ current_feature + Sex + (1 | Subject) + storage_time
y ~ current_feature + Sex + (1 | Subject) + storage_time
y ~ current_feature + Sex + (1 | Subject) + storage_time
y ~ current_feature + Sex + (1 | Subject) + storage_time
y ~ current_feature + Sex + (1 | Subject) + storage_time
y ~ current_feature + Sex + (1 | Subject) + storage_time
y ~ current_feature + Sex + (1 | Subject) + storage_time
y ~ current_feature + Sex + (1 | Subject) + storage_time
y ~ current_feature + Sex + (1 | Subject) + storage_time
y ~ current_feature + Sex + (1 | Subject) + storage_time
y ~ current_feature + Sex + (1 | Subject) + storage_time
y ~ current_feature + Sex + (1 | Subject) + storage_time
y ~ current_feature + Sex + (1 | Subject) + storage_time

```
y ~ current_feature + Sex + (1 | Subject) + storage_time
y ~ current_feature + Sex + (1 | Subject) + storage_time
y ~ current_feature + Sex + (1 | Subject) + storage_time
y ~ current_feature + Sex + (1 | Subject) + storage_time
y ~ current_feature + Sex + (1 | Subject) + storage_time
y ~ current_feature + Sex + (1 | Subject) + storage_time
y ~ current_feature + Sex + (1 | Subject) + storage_time
y ~ current_feature + Sex + (1 | Subject) + storage_time
y ~ current_feature + Sex + (1 | Subject) + storage_time
y ~ current_feature + Sex + (1 | Subject) + storage_time
y ~ current_feature + Sex + (1 | Subject) + storage_time
y ~ current_feature + Sex + (1 | Subject) + storage_time
y ~ current_feature + Sex + (1 | Subject) + storage_time
y ~ current_feature + Sex + (1 | Subject) + storage_time
y ~ current_feature + Sex + (1 | Subject) + storage_time
y ~ current_feature + Sex + (1 | Subject) + storage_time
y ~ current_feature + Sex + (1 | Subject) + storage_time
y ~ current_feature + Sex + (1 | Subject) + storage_time
y ~ current_feature + Sex + (1 | Subject) + storage_time
y ~ current_feature + Sex + (1 | Subject) + storage_time
y ~ current_feature + Sex + (1 | Subject) + storage_time
y ~ current_feature + Sex + (1 | Subject) + storage_time
y ~ current_feature + Sex + (1 | Subject) + storage_time
y ~ current_feature + Sex + (1 | Subject) + storage_time
y ~ current_feature + Sex + (1 | Subject) + storage_time
y ~ current_feature + Sex + (1 | Subject) + storage_time
y ~ current_feature + Sex + (1 | Subject) + storage_time
y ~ current_feature + Sex + (1 | Subject) + storage_time
y ~ current_feature + Sex + (1 | Subject) + storage_time
y ~ current_feature + Sex + (1 | Subject) + storage_time
y ~ current_feature + Sex + (1 | Subject) + storage_time
y ~ current_feature + Sex + (1 | Subject) + storage_time
y ~ current_feature + Sex + (1 | Subject) + storage_time
y ~ current_feature + Sex + (1 | Subject) + storage_time
y ~ current_feature + Sex + (1 | Subject) + storage_time
y ~ current_feature + Sex + (1 | Subject) + storage_time
y ~ current_feature + Sex + (1 | Subject) + storage_time
y ~ current_feature + Sex + (1 | Subject) + storage_time
y ~ current_feature + Sex + (1 | Subject) + storage_time
y ~ current_feature + Sex + (1 | Subject) + storage_time
y ~ current_feature + Sex + (1 | Subject) + storage_time
y ~ current_feature + Sex + (1 | Subject) + storage_time
y ~ current_feature + Sex + (1 | Subject) + storage_time
y ~ current_feature + Sex + (1 | Subject) + storage_time
y ~ current_feature + Sex + (1 | Subject) + storage_time
y ~ current_feature + Sex + (1 | Subject) + storage_time
y ~ current_feature + Sex + (1 | Subject) + storage_time
y ~ current_feature + Sex + (1 | Subject) + storage_time
```

y ~ current_feature + Sex + (1 | Subject) + storage_time
y ~ current_feature + Sex + (1 | Subject) + storage_time
y ~ current_feature + Sex + (1 | Subject) + storage_time
y ~ current_feature + Sex + (1 | Subject) + storage_time
y ~ current_feature + Sex + (1 | Subject) + storage_time
y ~ current_feature + Sex + (1 | Subject) + storage_time
y ~ current_feature + Sex + (1 | Subject) + storage_time
y ~ current_feature + Sex + (1 | Subject) + storage_time
y ~ current_feature + Sex + (1 | Subject) + storage_time
y ~ current_feature + Sex + (1 | Subject) + storage_time
y ~ current_feature + Sex + (1 | Subject) + storage_time
y ~ current_feature + Sex + (1 | Subject) + storage_time
y ~ current_feature + Sex + (1 | Subject) + storage_time
y ~ current_feature + Sex + (1 | Subject) + storage_time
y ~ current_feature + Sex + (1 | Subject) + storage_time
y ~ current_feature + Sex + (1 | Subject) + storage_time
y ~ current_feature + Sex + (1 | Subject) + storage_time
y ~ current_feature + Sex + (1 | Subject) + storage_time
y ~ current_feature + Sex + (1 | Subject) + storage_time
y ~ current_feature + Sex + (1 | Subject) + storage_time
y ~ current_feature + Sex + (1 | Subject) + storage_time
y ~ current_feature + Sex + (1 | Subject) + storage_time
y ~ current_feature + Sex + (1 | Subject) + storage_time
y ~ current_feature + Sex + (1 | Subject) + storage_time
y ~ current_feature + Sex + (1 | Subject) + storage_time
y ~ current_feature + Sex + (1 | Subject) + storage_time
y ~ current_feature + Sex + (1 | Subject) + storage_time
y ~ current_feature + Sex + (1 | Subject) + storage_time
y ~ current_feature + Sex + (1 | Subject) + storage_time
y ~ current_feature + Sex + (1 | Subject) + storage_time
y ~ current_feature + Sex + (1 | Subject) + storage_time
y ~ current_feature + Sex + (1 | Subject) + storage_time
y ~ current_feature + Sex + (1 | Subject) + storage_time
y ~ current_feature + Sex + (1 | Subject) + storage_time
y ~ current_feature + Sex + (1 | Subject) + storage_time
y ~ current_feature + Sex + (1 | Subject) + storage_time
y ~ current_feature + Sex + (1 | Subject) + storage_time
y ~ current_feature + Sex + (1 | Subject) + storage_time
y ~ current_feature + Sex + (1 | Subject) + storage_time
y ~ current_feature + Sex + (1 | Subject) + storage_time
y ~ current_feature + Sex + (1 | Subject) + storage_time
y ~ current_feature + Sex + (1 | Subject) + storage_time
y ~ current_feature + Sex + (1 | Subject) + storage_time
y ~ current_feature + Sex + (1 | Subject) + storage_time
y ~ current_feature + Sex + (1 | Subject) + storage_time
y ~ current_feature + Sex + (1 | Subject) + storage_time
y ~ current_feature + Sex + (1 | Subject) + storage_time
y ~ current_feature + Sex + (1 | Subject) + storage_time

y ~ current_feature + Sex + (1 | Subject) + storage_time
y ~ current_feature + Sex + (1 | Subject) + storage_time
y ~ current_feature + Sex + (1 | Subject) + storage_time
y ~ current_feature + Sex + (1 | Subject) + storage_time
y ~ current_feature + Sex + (1 | Subject) + storage_time
y ~ current_feature + Sex + (1 | Subject) + storage_time
y ~ current_feature + Sex + (1 | Subject) + storage_time
y ~ current_feature + Sex + (1 | Subject) + storage_time
y ~ current_feature + Sex + (1 | Subject) + storage_time
y ~ current_feature + Sex + (1 | Subject) + storage_time
y ~ current_feature + Sex + (1 | Subject) + storage_time
y ~ current_feature + Sex + (1 | Subject) + storage_time
y ~ current_feature + Sex + (1 | Subject) + storage_time
y ~ current_feature + Sex + (1 | Subject) + storage_time
y ~ current_feature + Sex + (1 | Subject) + storage_time
y ~ current_feature + Sex + (1 | Subject) + storage_time
y ~ current_feature + Sex + (1 | Subject) + storage_time
y ~ current_feature + Sex + (1 | Subject) + storage_time
y ~ current_feature + Sex + (1 | Subject) + storage_time
y ~ current_feature + Sex + (1 | Subject) + storage_time
y ~ current_feature + Sex + (1 | Subject) + storage_time
y ~ current_feature + Sex + (1 | Subject) + storage_time
y ~ current_feature + Sex + (1 | Subject) + storage_time
y ~ current_feature + Sex + (1 | Subject) + storage_time
y ~ current_feature + Sex + (1 | Subject) + storage_time
y ~ current_feature + Sex + (1 | Subject) + storage_time
y ~ current_feature + Sex + (1 | Subject) + storage_time
y ~ current_feature + Sex + (1 | Subject) + storage_time
y ~ current_feature + Sex + (1 | Subject) + storage_time
y ~ current_feature + Sex + (1 | Subject) + storage_time
y ~ current_feature + Sex + (1 | Subject) + storage_time
y ~ current_feature + Sex + (1 | Subject) + storage_time
y ~ current_feature + Sex + (1 | Subject) + storage_time
y ~ current_feature + Sex + (1 | Subject) + storage_time
y ~ current_feature + Sex + (1 | Subject) + storage_time
y ~ current_feature + Sex + (1 | Subject) + storage_time
y ~ current_feature + Sex + (1 | Subject) + storage_time
y ~ current_feature + Sex + (1 | Subject) + storage_time
y ~ current_feature + Sex + (1 | Subject) + storage_time
y ~ current_feature + Sex + (1 | Subject) + storage_time
y ~ current_feature + Sex + (1 | Subject) + storage_time
y ~ current_feature + Sex + (1 | Subject) + storage_time
y ~ current_feature + Sex + (1 | Subject) + storage_time
y ~ current_feature + Sex + (1 | Subject) + storage_time
y ~ current_feature + Sex + (1 | Subject) + storage_time
y ~ current_feature + Sex + (1 | Subject) + storage_time
y ~ current_feature + Sex + (1 | Subject) + storage_time
y ~ current_feature + Sex + (1 | Subject) + storage_time

```
y ~ current_feature + Sex + (1 | Subject) + storage_time
y ~ current_feature + Sex + (1 | Subject) + storage_time
y ~ current_feature + Sex + (1 | Subject) + storage_time
y ~ current_feature + Sex + (1 | Subject) + storage_time
y ~ current_feature + Sex + (1 | Subject) + storage_time
y ~ current_feature + Sex + (1 | Subject) + storage_time
y ~ current_feature + Sex + (1 | Subject) + storage_time
y ~ current_feature + Sex + (1 | Subject) + storage_time
y ~ current_feature + Sex + (1 | Subject) + storage_time
y ~ current_feature + Sex + (1 | Subject) + storage_time
y ~ current_feature + Sex + (1 | Subject) + storage_time
y ~ current_feature + Sex + (1 | Subject) + storage_time
y ~ current_feature + Sex + (1 | Subject) + storage_time
y ~ current_feature + Sex + (1 | Subject) + storage_time
y ~ current_feature + Sex + (1 | Subject) + storage_time
y ~ current_feature + Sex + (1 | Subject) + storage_time
y ~ current_feature + Sex + (1 | Subject) + storage_time
y ~ current_feature + Sex + (1 | Subject) + storage_time
y ~ current_feature + Sex + (1 | Subject) + storage_time
y ~ current_feature + Sex + (1 | Subject) + storage_time
y ~ current_feature + Sex + (1 | Subject) + storage_time
y ~ current_feature + Sex + (1 | Subject) + storage_time
y ~ current_feature + Sex + (1 | Subject) + storage_time
y ~ current_feature + Sex + (1 | Subject) + storage_time
y ~ current_feature + Sex + (1 | Subject) + storage_time
y ~ current_feature + Sex + (1 | Subject) + storage_time
y ~ current_feature + Sex + (1 | Subject) + storage_time
y ~ current_feature + Sex + (1 | Subject) + storage_time
y ~ current_feature + Sex + (1 | Subject) + storage_time
y ~ current_feature + Sex + (1 | Subject) + storage_time
y ~ current_feature + Sex + (1 | Subject) + storage_time
y ~ current_feature + Sex + (1 | Subject) + storage_time
y ~ current_feature + Sex + (1 | Subject) + storage_time
y ~ current_feature + Sex + (1 | Subject) + storage_time
y ~ current_feature + Sex + (1 | Subject) + storage_time
y ~ current_feature + Sex + (1 | Subject) + storage_time
y ~ current_feature + Sex + (1 | Subject) + storage_time
y ~ current_feature + Sex + (1 | Subject) + storage_time
y ~ current_feature + Sex + (1 | Subject) + storage_time
y ~ current_feature + Sex + (1 | Subject) + storage_time
y ~ current_feature + Sex + (1 | Subject) + storage_time
y ~ current_feature + Sex + (1 | Subject) + storage_time
y ~ current_feature + Sex + (1 | Subject) + storage_time
y ~ current_feature + Sex + (1 | Subject) + storage_time
y ~ current_feature + Sex + (1 | Subject) + storage_time
y ~ current_feature + Sex + (1 | Subject) + storage_time
y ~ current_feature + Sex + (1 | Subject) + storage_time
y ~ current_feature + Sex + (1 | Subject) + storage_time
```

y ~ current_feature + Sex + (1 | Subject) + storage_time
y ~ current_feature + Sex + (1 | Subject) + storage_time
y ~ current_feature + Sex + (1 | Subject) + storage_time
y ~ current_feature + Sex + (1 | Subject) + storage_time
y ~ current_feature + Sex + (1 | Subject) + storage_time
y ~ current_feature + Sex + (1 | Subject) + storage_time
y ~ current_feature + Sex + (1 | Subject) + storage_time
y ~ current_feature + Sex + (1 | Subject) + storage_time
y ~ current_feature + Sex + (1 | Subject) + storage_time
y ~ current_feature + Sex + (1 | Subject) + storage_time
y ~ current_feature + Sex + (1 | Subject) + storage_time
y ~ current_feature + Sex + (1 | Subject) + storage_time
y ~ current_feature + Sex + (1 | Subject) + storage_time
y ~ current_feature + Sex + (1 | Subject) + storage_time
y ~ current_feature + Sex + (1 | Subject) + storage_time
y ~ current_feature + Sex + (1 | Subject) + storage_time
y ~ current_feature + Sex + (1 | Subject) + storage_time
y ~ current_feature + Sex + (1 | Subject) + storage_time
y ~ current_feature + Sex + (1 | Subject) + storage_time
y ~ current_feature + Sex + (1 | Subject) + storage_time
y ~ current_feature + Sex + (1 | Subject) + storage_time
y ~ current_feature + Sex + (1 | Subject) + storage_time
y ~ current_feature + Sex + (1 | Subject) + storage_time
y ~ current_feature + Sex + (1 | Subject) + storage_time
y ~ current_feature + Sex + (1 | Subject) + storage_time
y ~ current_feature + Sex + (1 | Subject) + storage_time
y ~ current_feature + Sex + (1 | Subject) + storage_time
y ~ current_feature + Sex + (1 | Subject) + storage_time
y ~ current_feature + Sex + (1 | Subject) + storage_time
y ~ current_feature + Sex + (1 | Subject) + storage_time
y ~ current_feature + Sex + (1 | Subject) + storage_time
y ~ current_feature + Sex + (1 | Subject) + storage_time
y ~ current_feature + Sex + (1 | Subject) + storage_time
y ~ current_feature + Sex + (1 | Subject) + storage_time
y ~ current_feature + Sex + (1 | Subject) + storage_time
y ~ current_feature + Sex + (1 | Subject) + storage_time
y ~ current_feature + Sex + (1 | Subject) + storage_time
y ~ current_feature + Sex + (1 | Subject) + storage_time
y ~ current_feature + Sex + (1 | Subject) + storage_time
y ~ current_feature + Sex + (1 | Subject) + storage_time
y ~ current_feature + Sex + (1 | Subject) + storage_time
y ~ current_feature + Sex + (1 | Subject) + storage_time
y ~ current_feature + Sex + (1 | Subject) + storage_time
y ~ current_feature + Sex + (1 | Subject) + storage_time
y ~ current_feature + Sex + (1 | Subject) + storage_time
y ~ current_feature + Sex + (1 | Subject) + storage_time
y ~ current_feature + Sex + (1 | Subject) + storage_time
y ~ current_feature + Sex + (1 | Subject) + storage_time

y ~ current_feature + Sex + (1 | Subject) + storage_time
y ~ current_feature + Sex + (1 | Subject) + storage_time
y ~ current_feature + Sex + (1 | Subject) + storage_time
y ~ current_feature + Sex + (1 | Subject) + storage_time
y ~ current_feature + Sex + (1 | Subject) + storage_time
y ~ current_feature + Sex + (1 | Subject) + storage_time
y ~ current_feature + Sex + (1 | Subject) + storage_time
y ~ current_feature + Sex + (1 | Subject) + storage_time
y ~ current_feature + Sex + (1 | Subject) + storage_time
y ~ current_feature + Sex + (1 | Subject) + storage_time
y ~ current_feature + Sex + (1 | Subject) + storage_time
y ~ current_feature + Sex + (1 | Subject) + storage_time
y ~ current_feature + Sex + (1 | Subject) + storage_time
y ~ current_feature + Sex + (1 | Subject) + storage_time
y ~ current_feature + Sex + (1 | Subject) + storage_time
y ~ current_feature + Sex + (1 | Subject) + storage_time
y ~ current_feature + Sex + (1 | Subject) + storage_time
y ~ current_feature + Sex + (1 | Subject) + storage_time
y ~ current_feature + Sex + (1 | Subject) + storage_time
y ~ current_feature + Sex + (1 | Subject) + storage_time
y ~ current_feature + Sex + (1 | Subject) + storage_time
y ~ current_feature + Sex + (1 | Subject) + storage_time
y ~ current_feature + Sex + (1 | Subject) + storage_time
y ~ current_feature + Sex + (1 | Subject) + storage_time
y ~ current_feature + Sex + (1 | Subject) + storage_time
y ~ current_feature + Sex + (1 | Subject) + storage_time
y ~ current_feature + Sex + (1 | Subject) + storage_time
y ~ current_feature + Sex + (1 | Subject) + storage_time
y ~ current_feature + Sex + (1 | Subject) + storage_time
y ~ current_feature + Sex + (1 | Subject) + storage_time
y ~ current_feature + Sex + (1 | Subject) + storage_time
y ~ current_feature + Sex + (1 | Subject) + storage_time
y ~ current_feature + Sex + (1 | Subject) + storage_time
y ~ current_feature + Sex + (1 | Subject) + storage_time
y ~ current_feature + Sex + (1 | Subject) + storage_time
y ~ current_feature + Sex + (1 | Subject) + storage_time
y ~ current_feature + Sex + (1 | Subject) + storage_time
y ~ current_feature + Sex + (1 | Subject) + storage_time
y ~ current_feature + Sex + (1 | Subject) + storage_time
y ~ current_feature + Sex + (1 | Subject) + storage_time
y ~ current_feature + Sex + (1 | Subject) + storage_time
y ~ current_feature + Sex + (1 | Subject) + storage_time
y ~ current_feature + Sex + (1 | Subject) + storage_time
y ~ current_feature + Sex + (1 | Subject) + storage_time
y ~ current_feature + Sex + (1 | Subject) + storage_time
y ~ current_feature + Sex + (1 | Subject) + storage_time
y ~ current_feature + Sex + (1 | Subject) + storage_time
y ~ current_feature + Sex + (1 | Subject) + storage_time

y ~ current_feature + Sex + (1 | Subject) + storage_time
y ~ current_feature + Sex + (1 | Subject) + storage_time
y ~ current_feature + Sex + (1 | Subject) + storage_time
y ~ current_feature + Sex + (1 | Subject) + storage_time
y ~ current_feature + Sex + (1 | Subject) + storage_time
y ~ current_feature + Sex + (1 | Subject) + storage_time
y ~ current_feature + Sex + (1 | Subject) + storage_time
y ~ current_feature + Sex + (1 | Subject) + storage_time
y ~ current_feature + Sex + (1 | Subject) + storage_time
y ~ current_feature + Sex + (1 | Subject) + storage_time
y ~ current_feature + Sex + (1 | Subject) + storage_time
y ~ current_feature + Sex + (1 | Subject) + storage_time
y ~ current_feature + Sex + (1 | Subject) + storage_time
y ~ current_feature + Sex + (1 | Subject) + storage_time
y ~ current_feature + Sex + (1 | Subject) + storage_time
y ~ current_feature + Sex + (1 | Subject) + storage_time
y ~ current_feature + Sex + (1 | Subject) + storage_time
y ~ current_feature + Sex + (1 | Subject) + storage_time
y ~ current_feature + Sex + (1 | Subject) + storage_time
y ~ current_feature + Sex + (1 | Subject) + storage_time
y ~ current_feature + Sex + (1 | Subject) + storage_time
y ~ current_feature + Sex + (1 | Subject) + storage_time
y ~ current_feature + Sex + (1 | Subject) + storage_time
y ~ current_feature + Sex + (1 | Subject) + storage_time
y ~ current_feature + Sex + (1 | Subject) + storage_time
y ~ current_feature + Sex + (1 | Subject) + storage_time
y ~ current_feature + Sex + (1 | Subject) + storage_time
y ~ current_feature + Sex + (1 | Subject) + storage_time
y ~ current_feature + Sex + (1 | Subject) + storage_time
y ~ current_feature + Sex + (1 | Subject) + storage_time
y ~ current_feature + Sex + (1 | Subject) + storage_time
y ~ current_feature + Sex + (1 | Subject) + storage_time
y ~ current_feature + Sex + (1 | Subject) + storage_time
y ~ current_feature + Sex + (1 | Subject) + storage_time
y ~ current_feature + Sex + (1 | Subject) + storage_time
y ~ current_feature + Sex + (1 | Subject) + storage_time
y ~ current_feature + Sex + (1 | Subject) + storage_time
y ~ current_feature + Sex + (1 | Subject) + storage_time
y ~ current_feature + Sex + (1 | Subject) + storage_time
y ~ current_feature + Sex + (1 | Subject) + storage_time
y ~ current_feature + Sex + (1 | Subject) + storage_time
y ~ current_feature + Sex + (1 | Subject) + storage_time
y ~ current_feature + Sex + (1 | Subject) + storage_time
y ~ current_feature + Sex + (1 | Subject) + storage_time
y ~ current_feature + Sex + (1 | Subject) + storage_time
y ~ current_feature + Sex + (1 | Subject) + storage_time
y ~ current_feature + Sex + (1 | Subject) + storage_time
y ~ current_feature + Sex + (1 | Subject) + storage_time

y ~ current_feature + Sex + (1 | Subject) + storage_time
y ~ current_feature + Sex + (1 | Subject) + storage_time
y ~ current_feature + Sex + (1 | Subject) + storage_time
y ~ current_feature + Sex + (1 | Subject) + storage_time
y ~ current_feature + Sex + (1 | Subject) + storage_time
y ~ current_feature + Sex + (1 | Subject) + storage_time
y ~ current_feature + Sex + (1 | Subject) + storage_time
y ~ current_feature + Sex + (1 | Subject) + storage_time
y ~ current_feature + Sex + (1 | Subject) + storage_time
y ~ current_feature + Sex + (1 | Subject) + storage_time
y ~ current_feature + Sex + (1 | Subject) + storage_time
y ~ current_feature + Sex + (1 | Subject) + storage_time
y ~ current_feature + Sex + (1 | Subject) + storage_time
y ~ current_feature + Sex + (1 | Subject) + storage_time
y ~ current_feature + Sex + (1 | Subject) + storage_time
y ~ current_feature + Sex + (1 | Subject) + storage_time
y ~ current_feature + Sex + (1 | Subject) + storage_time
y ~ current_feature + Sex + (1 | Subject) + storage_time
y ~ current_feature + Sex + (1 | Subject) + storage_time
y ~ current_feature + Sex + (1 | Subject) + storage_time
y ~ current_feature + Sex + (1 | Subject) + storage_time
y ~ current_feature + Sex + (1 | Subject) + storage_time
y ~ current_feature + Sex + (1 | Subject) + storage_time
y ~ current_feature + Sex + (1 | Subject) + storage_time
y ~ current_feature + Sex + (1 | Subject) + storage_time
y ~ current_feature + Sex + (1 | Subject) + storage_time
y ~ current_feature + Sex + (1 | Subject) + storage_time
y ~ current_feature + Sex + (1 | Subject) + storage_time
y ~ current_feature + Sex + (1 | Subject) + storage_time
y ~ current_feature + Sex + (1 | Subject) + storage_time
y ~ current_feature + Sex + (1 | Subject) + storage_time
y ~ current_feature + Sex + (1 | Subject) + storage_time
y ~ current_feature + Sex + (1 | Subject) + storage_time
y ~ current_feature + Sex + (1 | Subject) + storage_time
y ~ current_feature + Sex + (1 | Subject) + storage_time
y ~ current_feature + Sex + (1 | Subject) + storage_time
y ~ current_feature + Sex + (1 | Subject) + storage_time
y ~ current_feature + Sex + (1 | Subject) + storage_time
y ~ current_feature + Sex + (1 | Subject) + storage_time
y ~ current_feature + Sex + (1 | Subject) + storage_time
y ~ current_feature + Sex + (1 | Subject) + storage_time
y ~ current_feature + Sex + (1 | Subject) + storage_time
y ~ current_feature + Sex + (1 | Subject) + storage_time
y ~ current_feature + Sex + (1 | Subject) + storage_time
y ~ current_feature + Sex + (1 | Subject) + storage_time
y ~ current_feature + Sex + (1 | Subject) + storage_time
y ~ current_feature + Sex + (1 | Subject) + storage_time
y ~ current_feature + Sex + (1 | Subject) + storage_time

y ~ current_feature + Sex + (1 | Subject) + storage_time
y ~ current_feature + Sex + (1 | Subject) + storage_time
y ~ current_feature + Sex + (1 | Subject) + storage_time
y ~ current_feature + Sex + (1 | Subject) + storage_time
y ~ current_feature + Sex + (1 | Subject) + storage_time
y ~ current_feature + Sex + (1 | Subject) + storage_time
y ~ current_feature + Sex + (1 | Subject) + storage_time
y ~ current_feature + Sex + (1 | Subject) + storage_time
y ~ current_feature + Sex + (1 | Subject) + storage_time
y ~ current_feature + Sex + (1 | Subject) + storage_time
y ~ current_feature + Sex + (1 | Subject) + storage_time
y ~ current_feature + Sex + (1 | Subject) + storage_time
y ~ current_feature + Sex + (1 | Subject) + storage_time
y ~ current_feature + Sex + (1 | Subject) + storage_time
y ~ current_feature + Sex + (1 | Subject) + storage_time
y ~ current_feature + Sex + (1 | Subject) + storage_time
y ~ current_feature + Sex + (1 | Subject) + storage_time
y ~ current_feature + Sex + (1 | Subject) + storage_time
y ~ current_feature + Sex + (1 | Subject) + storage_time
y ~ current_feature + Sex + (1 | Subject) + storage_time
y ~ current_feature + Sex + (1 | Subject) + storage_time
y ~ current_feature + Sex + (1 | Subject) + storage_time
y ~ current_feature + Sex + (1 | Subject) + storage_time
y ~ current_feature + Sex + (1 | Subject) + storage_time
y ~ current_feature + Sex + (1 | Subject) + storage_time
y ~ current_feature + Sex + (1 | Subject) + storage_time
y ~ current_feature + Sex + (1 | Subject) + storage_time
y ~ current_feature + Sex + (1 | Subject) + storage_time
y ~ current_feature + Sex + (1 | Subject) + storage_time
y ~ current_feature + Sex + (1 | Subject) + storage_time
y ~ current_feature + Sex + (1 | Subject) + storage_time
y ~ current_feature + Sex + (1 | Subject) + storage_time
y ~ current_feature + Sex + (1 | Subject) + storage_time
y ~ current_feature + Sex + (1 | Subject) + storage_time
y ~ current_feature + Sex + (1 | Subject) + storage_time
y ~ current_feature + Sex + (1 | Subject) + storage_time
y ~ current_feature + Sex + (1 | Subject) + storage_time
y ~ current_feature + Sex + (1 | Subject) + storage_time
y ~ current_feature + Sex + (1 | Subject) + storage_time
y ~ current_feature + Sex + (1 | Subject) + storage_time
y ~ current_feature + Sex + (1 | Subject) + storage_time
y ~ current_feature + Sex + (1 | Subject) + storage_time
y ~ current_feature + Sex + (1 | Subject) + storage_time
y ~ current_feature + Sex + (1 | Subject) + storage_time
y ~ current_feature + Sex + (1 | Subject) + storage_time
y ~ current_feature + Sex + (1 | Subject) + storage_time
y ~ current_feature + Sex + (1 | Subject) + storage_time
y ~ current_feature + Sex + (1 | Subject) + storage_time

y ~ current_feature + Sex + (1 | Subject) + storage_time
y ~ current_feature + Sex + (1 | Subject) + storage_time
y ~ current_feature + Sex + (1 | Subject) + storage_time
y ~ current_feature + Sex + (1 | Subject) + storage_time
y ~ current_feature + Sex + (1 | Subject) + storage_time
y ~ current_feature + Sex + (1 | Subject) + storage_time
y ~ current_feature + Sex + (1 | Subject) + storage_time
y ~ current_feature + Sex + (1 | Subject) + storage_time
y ~ current_feature + Sex + (1 | Subject) + storage_time
y ~ current_feature + Sex + (1 | Subject) + storage_time
y ~ current_feature + Sex + (1 | Subject) + storage_time
y ~ current_feature + Sex + (1 | Subject) + storage_time
y ~ current_feature + Sex + (1 | Subject) + storage_time
y ~ current_feature + Sex + (1 | Subject) + storage_time
y ~ current_feature + Sex + (1 | Subject) + storage_time
y ~ current_feature + Sex + (1 | Subject) + storage_time
y ~ current_feature + Sex + (1 | Subject) + storage_time
y ~ current_feature + Sex + (1 | Subject) + storage_time
y ~ current_feature + Sex + (1 | Subject) + storage_time
y ~ current_feature + Sex + (1 | Subject) + storage_time
y ~ current_feature + Sex + (1 | Subject) + storage_time
y ~ current_feature + Sex + (1 | Subject) + storage_time
y ~ current_feature + Sex + (1 | Subject) + storage_time
y ~ current_feature + Sex + (1 | Subject) + storage_time
y ~ current_feature + Sex + (1 | Subject) + storage_time
y ~ current_feature + Sex + (1 | Subject) + storage_time
y ~ current_feature + Sex + (1 | Subject) + storage_time
y ~ current_feature + Sex + (1 | Subject) + storage_time
y ~ current_feature + Sex + (1 | Subject) + storage_time
y ~ current_feature + Sex + (1 | Subject) + storage_time
y ~ current_feature + Sex + (1 | Subject) + storage_time
y ~ current_feature + Sex + (1 | Subject) + storage_time
y ~ current_feature + Sex + (1 | Subject) + storage_time
y ~ current_feature + Sex + (1 | Subject) + storage_time
y ~ current_feature + Sex + (1 | Subject) + storage_time
y ~ current_feature + Sex + (1 | Subject) + storage_time
y ~ current_feature + Sex + (1 | Subject) + storage_time
y ~ current_feature + Sex + (1 | Subject) + storage_time
y ~ current_feature + Sex + (1 | Subject) + storage_time
y ~ current_feature + Sex + (1 | Subject) + storage_time
y ~ current_feature + Sex + (1 | Subject) + storage_time
y ~ current_feature + Sex + (1 | Subject) + storage_time
y ~ current_feature + Sex + (1 | Subject) + storage_time
y ~ current_feature + Sex + (1 | Subject) + storage_time
y ~ current_feature + Sex + (1 | Subject) + storage_time
y ~ current_feature + Sex + (1 | Subject) + storage_time
y ~ current_feature + Sex + (1 | Subject) + storage_time
y ~ current_feature + Sex + (1 | Subject) + storage_time

y ~ current_feature + Sex + (1 | Subject) + storage_time
y ~ current_feature + Sex + (1 | Subject) + storage_time
y ~ current_feature + Sex + (1 | Subject) + storage_time
y ~ current_feature + Sex + (1 | Subject) + storage_time
y ~ current_feature + Sex + (1 | Subject) + storage_time
y ~ current_feature + Sex + (1 | Subject) + storage_time
y ~ current_feature + Sex + (1 | Subject) + storage_time
y ~ current_feature + Sex + (1 | Subject) + storage_time
y ~ current_feature + Sex + (1 | Subject) + storage_time
y ~ current_feature + Sex + (1 | Subject) + storage_time
y ~ current_feature + Sex + (1 | Subject) + storage_time
y ~ current_feature + Sex + (1 | Subject) + storage_time
y ~ current_feature + Sex + (1 | Subject) + storage_time
y ~ current_feature + Sex + (1 | Subject) + storage_time
y ~ current_feature + Sex + (1 | Subject) + storage_time
y ~ current_feature + Sex + (1 | Subject) + storage_time
y ~ current_feature + Sex + (1 | Subject) + storage_time
y ~ current_feature + Sex + (1 | Subject) + storage_time
y ~ current_feature + Sex + (1 | Subject) + storage_time
y ~ current_feature + Sex + (1 | Subject) + storage_time
y ~ current_feature + Sex + (1 | Subject) + storage_time
y ~ current_feature + Sex + (1 | Subject) + storage_time
y ~ current_feature + Sex + (1 | Subject) + storage_time
y ~ current_feature + Sex + (1 | Subject) + storage_time
y ~ current_feature + Sex + (1 | Subject) + storage_time
y ~ current_feature + Sex + (1 | Subject) + storage_time
y ~ current_feature + Sex + (1 | Subject) + storage_time
y ~ current_feature + Sex + (1 | Subject) + storage_time
y ~ current_feature + Sex + (1 | Subject) + storage_time
y ~ current_feature + Sex + (1 | Subject) + storage_time
y ~ current_feature + Sex + (1 | Subject) + storage_time
y ~ current_feature + Sex + (1 | Subject) + storage_time
y ~ current_feature + Sex + (1 | Subject) + storage_time
y ~ current_feature + Sex + (1 | Subject) + storage_time
y ~ current_feature + Sex + (1 | Subject) + storage_time
y ~ current_feature + Sex + (1 | Subject) + storage_time
y ~ current_feature + Sex + (1 | Subject) + storage_time
y ~ current_feature + Sex + (1 | Subject) + storage_time
y ~ current_feature + Sex + (1 | Subject) + storage_time
y ~ current_feature + Sex + (1 | Subject) + storage_time
y ~ current_feature + Sex + (1 | Subject) + storage_time
y ~ current_feature + Sex + (1 | Subject) + storage_time
y ~ current_feature + Sex + (1 | Subject) + storage_time
y ~ current_feature + Sex + (1 | Subject) + storage_time
y ~ current_feature + Sex + (1 | Subject) + storage_time
y ~ current_feature + Sex + (1 | Subject) + storage_time
y ~ current_feature + Sex + (1 | Subject) + storage_time
y ~ current_feature + Sex + (1 | Subject) + storage_time

y ~ current_feature + Sex + (1 | Subject) + storage_time
y ~ current_feature + Sex + (1 | Subject) + storage_time
y ~ current_feature + Sex + (1 | Subject) + storage_time
y ~ current_feature + Sex + (1 | Subject) + storage_time
y ~ current_feature + Sex + (1 | Subject) + storage_time
y ~ current_feature + Sex + (1 | Subject) + storage_time
y ~ current_feature + Sex + (1 | Subject) + storage_time
y ~ current_feature + Sex + (1 | Subject) + storage_time
y ~ current_feature + Sex + (1 | Subject) + storage_time
y ~ current_feature + Sex + (1 | Subject) + storage_time
y ~ current_feature + Sex + (1 | Subject) + storage_time
y ~ current_feature + Sex + (1 | Subject) + storage_time
y ~ current_feature + Sex + (1 | Subject) + storage_time
y ~ current_feature + Sex + (1 | Subject) + storage_time
y ~ current_feature + Sex + (1 | Subject) + storage_time
y ~ current_feature + Sex + (1 | Subject) + storage_time
y ~ current_feature + Sex + (1 | Subject) + storage_time
y ~ current_feature + Sex + (1 | Subject) + storage_time
y ~ current_feature + Sex + (1 | Subject) + storage_time
y ~ current_feature + Sex + (1 | Subject) + storage_time
y ~ current_feature + Sex + (1 | Subject) + storage_time
y ~ current_feature + Sex + (1 | Subject) + storage_time
y ~ current_feature + Sex + (1 | Subject) + storage_time
y ~ current_feature + Sex + (1 | Subject) + storage_time
y ~ current_feature + Sex + (1 | Subject) + storage_time
y ~ current_feature + Sex + (1 | Subject) + storage_time
y ~ current_feature + Sex + (1 | Subject) + storage_time
y ~ current_feature + Sex + (1 | Subject) + storage_time
y ~ current_feature + Sex + (1 | Subject) + storage_time
y ~ current_feature + Sex + (1 | Subject) + storage_time
y ~ current_feature + Sex + (1 | Subject) + storage_time
y ~ current_feature + Sex + (1 | Subject) + storage_time
y ~ current_feature + Sex + (1 | Subject) + storage_time
y ~ current_feature + Sex + (1 | Subject) + storage_time
y ~ current_feature + Sex + (1 | Subject) + storage_time
y ~ current_feature + Sex + (1 | Subject) + storage_time
y ~ current_feature + Sex + (1 | Subject) + storage_time
y ~ current_feature + Sex + (1 | Subject) + storage_time
y ~ current_feature + Sex + (1 | Subject) + storage_time
y ~ current_feature + Sex + (1 | Subject) + storage_time
y ~ current_feature + Sex + (1 | Subject) + storage_time
y ~ current_feature + Sex + (1 | Subject) + storage_time
y ~ current_feature + Sex + (1 | Subject) + storage_time
y ~ current_feature + Sex + (1 | Subject) + storage_time
y ~ current_feature + Sex + (1 | Subject) + storage_time
y ~ current_feature + Sex + (1 | Subject) + storage_time
y ~ current_feature + Sex + (1 | Subject) + storage_time
y ~ current_feature + Sex + (1 | Subject) + storage_time

y ~ current_feature + Sex + (1 | Subject) + storage_time
y ~ current_feature + Sex + (1 | Subject) + storage_time
y ~ current_feature + Sex + (1 | Subject) + storage_time
y ~ current_feature + Sex + (1 | Subject) + storage_time
y ~ current_feature + Sex + (1 | Subject) + storage_time
y ~ current_feature + Sex + (1 | Subject) + storage_time
y ~ current_feature + Sex + (1 | Subject) + storage_time
y ~ current_feature + Sex + (1 | Subject) + storage_time
y ~ current_feature + Sex + (1 | Subject) + storage_time
y ~ current_feature + Sex + (1 | Subject) + storage_time
y ~ current_feature + Sex + (1 | Subject) + storage_time
y ~ current_feature + Sex + (1 | Subject) + storage_time
y ~ current_feature + Sex + (1 | Subject) + storage_time
y ~ current_feature + Sex + (1 | Subject) + storage_time
y ~ current_feature + Sex + (1 | Subject) + storage_time
y ~ current_feature + Sex + (1 | Subject) + storage_time
y ~ current_feature + Sex + (1 | Subject) + storage_time
y ~ current_feature + Sex + (1 | Subject) + storage_time
y ~ current_feature + Sex + (1 | Subject) + storage_time
y ~ current_feature + Sex + (1 | Subject) + storage_time
y ~ current_feature + Sex + (1 | Subject) + storage_time
y ~ current_feature + Sex + (1 | Subject) + storage_time
y ~ current_feature + Sex + (1 | Subject) + storage_time
y ~ current_feature + Sex + (1 | Subject) + storage_time
y ~ current_feature + Sex + (1 | Subject) + storage_time
y ~ current_feature + Sex + (1 | Subject) + storage_time
y ~ current_feature + Sex + (1 | Subject) + storage_time
y ~ current_feature + Sex + (1 | Subject) + storage_time
y ~ current_feature + Sex + (1 | Subject) + storage_time
y ~ current_feature + Sex + (1 | Subject) + storage_time
y ~ current_feature + Sex + (1 | Subject) + storage_time
y ~ current_feature + Sex + (1 | Subject) + storage_time
y ~ current_feature + Sex + (1 | Subject) + storage_time
y ~ current_feature + Sex + (1 | Subject) + storage_time
y ~ current_feature + Sex + (1 | Subject) + storage_time
y ~ current_feature + Sex + (1 | Subject) + storage_time
y ~ current_feature + Sex + (1 | Subject) + storage_time
y ~ current_feature + Sex + (1 | Subject) + storage_time
y ~ current_feature + Sex + (1 | Subject) + storage_time
y ~ current_feature + Sex + (1 | Subject) + storage_time
y ~ current_feature + Sex + (1 | Subject) + storage_time
y ~ current_feature + Sex + (1 | Subject) + storage_time
y ~ current_feature + Sex + (1 | Subject) + storage_time
y ~ current_feature + Sex + (1 | Subject) + storage_time
y ~ current_feature + Sex + (1 | Subject) + storage_time
y ~ current_feature + Sex + (1 | Subject) + storage_time
y ~ current_feature + Sex + (1 | Subject) + storage_time
y ~ current_feature + Sex + (1 | Subject) + storage_time

y ~ current_feature + Sex + (1 | Subject) + storage_time
y ~ current_feature + Sex + (1 | Subject) + storage_time
y ~ current_feature + Sex + (1 | Subject) + storage_time
y ~ current_feature + Sex + (1 | Subject) + storage_time
y ~ current_feature + Sex + (1 | Subject) + storage_time
y ~ current_feature + Sex + (1 | Subject) + storage_time
y ~ current_feature + Sex + (1 | Subject) + storage_time
y ~ current_feature + Sex + (1 | Subject) + storage_time
y ~ current_feature + Sex + (1 | Subject) + storage_time
y ~ current_feature + Sex + (1 | Subject) + storage_time
y ~ current_feature + Sex + (1 | Subject) + storage_time
y ~ current_feature + Sex + (1 | Subject) + storage_time
y ~ current_feature + Sex + (1 | Subject) + storage_time
y ~ current_feature + Sex + (1 | Subject) + storage_time
y ~ current_feature + Sex + (1 | Subject) + storage_time
y ~ current_feature + Sex + (1 | Subject) + storage_time
y ~ current_feature + Sex + (1 | Subject) + storage_time
y ~ current_feature + Sex + (1 | Subject) + storage_time
y ~ current_feature + Sex + (1 | Subject) + storage_time
y ~ current_feature + Sex + (1 | Subject) + storage_time
y ~ current_feature + Sex + (1 | Subject) + storage_time
y ~ current_feature + Sex + (1 | Subject) + storage_time
y ~ current_feature + Sex + (1 | Subject) + storage_time
y ~ current_feature + Sex + (1 | Subject) + storage_time
y ~ current_feature + Sex + (1 | Subject) + storage_time
y ~ current_feature + Sex + (1 | Subject) + storage_time
y ~ current_feature + Sex + (1 | Subject) + storage_time
y ~ current_feature + Sex + (1 | Subject) + storage_time
y ~ current_feature + Sex + (1 | Subject) + storage_time
y ~ current_feature + Sex + (1 | Subject) + storage_time
y ~ current_feature + Sex + (1 | Subject) + storage_time
y ~ current_feature + Sex + (1 | Subject) + storage_time
y ~ current_feature + Sex + (1 | Subject) + storage_time
y ~ current_feature + Sex + (1 | Subject) + storage_time
y ~ current_feature + Sex + (1 | Subject) + storage_time
y ~ current_feature + Sex + (1 | Subject) + storage_time
y ~ current_feature + Sex + (1 | Subject) + storage_time
y ~ current_feature + Sex + (1 | Subject) + storage_time
y ~ current_feature + Sex + (1 | Subject) + storage_time
y ~ current_feature + Sex + (1 | Subject) + storage_time
y ~ current_feature + Sex + (1 | Subject) + storage_time
y ~ current_feature + Sex + (1 | Subject) + storage_time
y ~ current_feature + Sex + (1 | Subject) + storage_time
y ~ current_feature + Sex + (1 | Subject) + storage_time
y ~ current_feature + Sex + (1 | Subject) + storage_time
y ~ current_feature + Sex + (1 | Subject) + storage_time
y ~ current_feature + Sex + (1 | Subject) + storage_time
y ~ current_feature + Sex + (1 | Subject) + storage_time

y ~ current_feature + Sex + (1 | Subject) + storage_time
y ~ current_feature + Sex + (1 | Subject) + storage_time
y ~ current_feature + Sex + (1 | Subject) + storage_time
y ~ current_feature + Sex + (1 | Subject) + storage_time
y ~ current_feature + Sex + (1 | Subject) + storage_time
y ~ current_feature + Sex + (1 | Subject) + storage_time
y ~ current_feature + Sex + (1 | Subject) + storage_time
y ~ current_feature + Sex + (1 | Subject) + storage_time
y ~ current_feature + Sex + (1 | Subject) + storage_time
y ~ current_feature + Sex + (1 | Subject) + storage_time
y ~ current_feature + Sex + (1 | Subject) + storage_time
y ~ current_feature + Sex + (1 | Subject) + storage_time
y ~ current_feature + Sex + (1 | Subject) + storage_time
y ~ current_feature + Sex + (1 | Subject) + storage_time
y ~ current_feature + Sex + (1 | Subject) + storage_time
y ~ current_feature + Sex + (1 | Subject) + storage_time
y ~ current_feature + Sex + (1 | Subject) + storage_time
y ~ current_feature + Sex + (1 | Subject) + storage_time
y ~ current_feature + Sex + (1 | Subject) + storage_time
y ~ current_feature + Sex + (1 | Subject) + storage_time
y ~ current_feature + Sex + (1 | Subject) + storage_time
y ~ current_feature + Sex + (1 | Subject) + storage_time
y ~ current_feature + Sex + (1 | Subject) + storage_time
y ~ current_feature + Sex + (1 | Subject) + storage_time
y ~ current_feature + Sex + (1 | Subject) + storage_time
y ~ current_feature + Sex + (1 | Subject) + storage_time
y ~ current_feature + Sex + (1 | Subject) + storage_time
y ~ current_feature + Sex + (1 | Subject) + storage_time
y ~ current_feature + Sex + (1 | Subject) + storage_time
y ~ current_feature + Sex + (1 | Subject) + storage_time
y ~ current_feature + Sex + (1 | Subject) + storage_time
y ~ current_feature + Sex + (1 | Subject) + storage_time
y ~ current_feature + Sex + (1 | Subject) + storage_time
y ~ current_feature + Sex + (1 | Subject) + storage_time
y ~ current_feature + Sex + (1 | Subject) + storage_time
y ~ current_feature + Sex + (1 | Subject) + storage_time
y ~ current_feature + Sex + (1 | Subject) + storage_time
y ~ current_feature + Sex + (1 | Subject) + storage_time
y ~ current_feature + Sex + (1 | Subject) + storage_time
y ~ current_feature + Sex + (1 | Subject) + storage_time
y ~ current_feature + Sex + (1 | Subject) + storage_time
y ~ current_feature + Sex + (1 | Subject) + storage_time
y ~ current_feature + Sex + (1 | Subject) + storage_time
y ~ current_feature + Sex + (1 | Subject) + storage_time
y ~ current_feature + Sex + (1 | Subject) + storage_time
y ~ current_feature + Sex + (1 | Subject) + storage_time
y ~ current_feature + Sex + (1 | Subject) + storage_time
y ~ current_feature + Sex + (1 | Subject) + storage_time

y ~ current_feature + Sex + (1 | Subject) + storage_time
y ~ current_feature + Sex + (1 | Subject) + storage_time
y ~ current_feature + Sex + (1 | Subject) + storage_time
y ~ current_feature + Sex + (1 | Subject) + storage_time
y ~ current_feature + Sex + (1 | Subject) + storage_time
y ~ current_feature + Sex + (1 | Subject) + storage_time
y ~ current_feature + Sex + (1 | Subject) + storage_time
y ~ current_feature + Sex + (1 | Subject) + storage_time
y ~ current_feature + Sex + (1 | Subject) + storage_time
y ~ current_feature + Sex + (1 | Subject) + storage_time
y ~ current_feature + Sex + (1 | Subject) + storage_time
y ~ current_feature + Sex + (1 | Subject) + storage_time
y ~ current_feature + Sex + (1 | Subject) + storage_time
y ~ current_feature + Sex + (1 | Subject) + storage_time
y ~ current_feature + Sex + (1 | Subject) + storage_time
y ~ current_feature + Sex + (1 | Subject) + storage_time
y ~ current_feature + Sex + (1 | Subject) + storage_time
y ~ current_feature + Sex + (1 | Subject) + storage_time
y ~ current_feature + Sex + (1 | Subject) + storage_time
y ~ current_feature + Sex + (1 | Subject) + storage_time
y ~ current_feature + Sex + (1 | Subject) + storage_time
y ~ current_feature + Sex + (1 | Subject) + storage_time
y ~ current_feature + Sex + (1 | Subject) + storage_time
y ~ current_feature + Sex + (1 | Subject) + storage_time
y ~ current_feature + Sex + (1 | Subject) + storage_time
y ~ current_feature + Sex + (1 | Subject) + storage_time
y ~ current_feature + Sex + (1 | Subject) + storage_time
y ~ current_feature + Sex + (1 | Subject) + storage_time
y ~ current_feature + Sex + (1 | Subject) + storage_time
y ~ current_feature + Sex + (1 | Subject) + storage_time
y ~ current_feature + Sex + (1 | Subject) + storage_time
y ~ current_feature + Sex + (1 | Subject) + storage_time
y ~ current_feature + Sex + (1 | Subject) + storage_time
y ~ current_feature + Sex + (1 | Subject) + storage_time
y ~ current_feature + Sex + (1 | Subject) + storage_time
y ~ current_feature + Sex + (1 | Subject) + storage_time
y ~ current_feature + Sex + (1 | Subject) + storage_time
y ~ current_feature + Sex + (1 | Subject) + storage_time
y ~ current_feature + Sex + (1 | Subject) + storage_time
y ~ current_feature + Sex + (1 | Subject) + storage_time
y ~ current_feature + Sex + (1 | Subject) + storage_time
y ~ current_feature + Sex + (1 | Subject) + storage_time
y ~ current_feature + Sex + (1 | Subject) + storage_time
y ~ current_feature + Sex + (1 | Subject) + storage_time
y ~ current_feature + Sex + (1 | Subject) + storage_time
y ~ current_feature + Sex + (1 | Subject) + storage_time
y ~ current_feature + Sex + (1 | Subject) + storage_time
y ~ current_feature + Sex + (1 | Subject) + storage_time

y ~ current_feature + Sex + (1 | Subject) + storage_time
y ~ current_feature + Sex + (1 | Subject) + storage_time
y ~ current_feature + Sex + (1 | Subject) + storage_time
y ~ current_feature + Sex + (1 | Subject) + storage_time
y ~ current_feature + Sex + (1 | Subject) + storage_time
y ~ current_feature + Sex + (1 | Subject) + storage_time
y ~ current_feature + Sex + (1 | Subject) + storage_time
y ~ current_feature + Sex + (1 | Subject) + storage_time
y ~ current_feature + Sex + (1 | Subject) + storage_time
y ~ current_feature + Sex + (1 | Subject) + storage_time
y ~ current_feature + Sex + (1 | Subject) + storage_time
y ~ current_feature + Sex + (1 | Subject) + storage_time
y ~ current_feature + Sex + (1 | Subject) + storage_time
y ~ current_feature + Sex + (1 | Subject) + storage_time
y ~ current_feature + Sex + (1 | Subject) + storage_time
y ~ current_feature + Sex + (1 | Subject) + storage_time
y ~ current_feature + Sex + (1 | Subject) + storage_time
y ~ current_feature + Sex + (1 | Subject) + storage_time
y ~ current_feature + Sex + (1 | Subject) + storage_time
y ~ current_feature + Sex + (1 | Subject) + storage_time
y ~ current_feature + Sex + (1 | Subject) + storage_time
y ~ current_feature + Sex + (1 | Subject) + storage_time
y ~ current_feature + Sex + (1 | Subject) + storage_time
y ~ current_feature + Sex + (1 | Subject) + storage_time
y ~ current_feature + Sex + (1 | Subject) + storage_time
y ~ current_feature + Sex + (1 | Subject) + storage_time
y ~ current_feature + Sex + (1 | Subject) + storage_time
y ~ current_feature + Sex + (1 | Subject) + storage_time
y ~ current_feature + Sex + (1 | Subject) + storage_time
y ~ current_feature + Sex + (1 | Subject) + storage_time
y ~ current_feature + Sex + (1 | Subject) + storage_time
y ~ current_feature + Sex + (1 | Subject) + storage_time
y ~ current_feature + Sex + (1 | Subject) + storage_time
y ~ current_feature + Sex + (1 | Subject) + storage_time
y ~ current_feature + Sex + (1 | Subject) + storage_time
y ~ current_feature + Sex + (1 | Subject) + storage_time
y ~ current_feature + Sex + (1 | Subject) + storage_time
y ~ current_feature + Sex + (1 | Subject) + storage_time
y ~ current_feature + Sex + (1 | Subject) + storage_time
y ~ current_feature + Sex + (1 | Subject) + storage_time
y ~ current_feature + Sex + (1 | Subject) + storage_time
y ~ current_feature + Sex + (1 | Subject) + storage_time
y ~ current_feature + Sex + (1 | Subject) + storage_time
y ~ current_feature + Sex + (1 | Subject) + storage_time
y ~ current_feature + Sex + (1 | Subject) + storage_time
y ~ current_feature + Sex + (1 | Subject) + storage_time
y ~ current_feature + Sex + (1 | Subject) + storage_time
y ~ current_feature + Sex + (1 | Subject) + storage_time

y ~ current_feature + Sex + (1 | Subject) + storage_time
y ~ current_feature + Sex + (1 | Subject) + storage_time
y ~ current_feature + Sex + (1 | Subject) + storage_time
y ~ current_feature + Sex + (1 | Subject) + storage_time
y ~ current_feature + Sex + (1 | Subject) + storage_time
y ~ current_feature + Sex + (1 | Subject) + storage_time
y ~ current_feature + Sex + (1 | Subject) + storage_time
y ~ current_feature + Sex + (1 | Subject) + storage_time
y ~ current_feature + Sex + (1 | Subject) + storage_time
y ~ current_feature + Sex + (1 | Subject) + storage_time
y ~ current_feature + Sex + (1 | Subject) + storage_time
y ~ current_feature + Sex + (1 | Subject) + storage_time
y ~ current_feature + Sex + (1 | Subject) + storage_time
y ~ current_feature + Sex + (1 | Subject) + storage_time
y ~ current_feature + Sex + (1 | Subject) + storage_time
y ~ current_feature + Sex + (1 | Subject) + storage_time
y ~ current_feature + Sex + (1 | Subject) + storage_time
y ~ current_feature + Sex + (1 | Subject) + storage_time
y ~ current_feature + Sex + (1 | Subject) + storage_time
y ~ current_feature + Sex + (1 | Subject) + storage_time
y ~ current_feature + Sex + (1 | Subject) + storage_time
y ~ current_feature + Sex + (1 | Subject) + storage_time
y ~ current_feature + Sex + (1 | Subject) + storage_time
y ~ current_feature + Sex + (1 | Subject) + storage_time
y ~ current_feature + Sex + (1 | Subject) + storage_time
y ~ current_feature + Sex + (1 | Subject) + storage_time
y ~ current_feature + Sex + (1 | Subject) + storage_time
y ~ current_feature + Sex + (1 | Subject) + storage_time
y ~ current_feature + Sex + (1 | Subject) + storage_time
y ~ current_feature + Sex + (1 | Subject) + storage_time
y ~ current_feature + Sex + (1 | Subject) + storage_time
y ~ current_feature + Sex + (1 | Subject) + storage_time
y ~ current_feature + Sex + (1 | Subject) + storage_time
y ~ current_feature + Sex + (1 | Subject) + storage_time
y ~ current_feature + Sex + (1 | Subject) + storage_time
y ~ current_feature + Sex + (1 | Subject) + storage_time
y ~ current_feature + Sex + (1 | Subject) + storage_time
y ~ current_feature + Sex + (1 | Subject) + storage_time
y ~ current_feature + Sex + (1 | Subject) + storage_time
y ~ current_feature + Sex + (1 | Subject) + storage_time
y ~ current_feature + Sex + (1 | Subject) + storage_time
y ~ current_feature + Sex + (1 | Subject) + storage_time
y ~ current_feature + Sex + (1 | Subject) + storage_time
y ~ current_feature + Sex + (1 | Subject) + storage_time
y ~ current_feature + Sex + (1 | Subject) + storage_time
y ~ current_feature + Sex + (1 | Subject) + storage_time
y ~ current_feature + Sex + (1 | Subject) + storage_time
y ~ current_feature + Sex + (1 | Subject) + storage_time

y ~ current_feature + Sex + (1 | Subject) + storage_time
y ~ current_feature + Sex + (1 | Subject) + storage_time
y ~ current_feature + Sex + (1 | Subject) + storage_time
y ~ current_feature + Sex + (1 | Subject) + storage_time
y ~ current_feature + Sex + (1 | Subject) + storage_time
y ~ current_feature + Sex + (1 | Subject) + storage_time
y ~ current_feature + Sex + (1 | Subject) + storage_time
y ~ current_feature + Sex + (1 | Subject) + storage_time
y ~ current_feature + Sex + (1 | Subject) + storage_time
y ~ current_feature + Sex + (1 | Subject) + storage_time
y ~ current_feature + Sex + (1 | Subject) + storage_time
y ~ current_feature + Sex + (1 | Subject) + storage_time
y ~ current_feature + Sex + (1 | Subject) + storage_time
y ~ current_feature + Sex + (1 | Subject) + storage_time
y ~ current_feature + Sex + (1 | Subject) + storage_time
y ~ current_feature + Sex + (1 | Subject) + storage_time
y ~ current_feature + Sex + (1 | Subject) + storage_time
y ~ current_feature + Sex + (1 | Subject) + storage_time
y ~ current_feature + Sex + (1 | Subject) + storage_time
y ~ current_feature + Sex + (1 | Subject) + storage_time
y ~ current_feature + Sex + (1 | Subject) + storage_time
y ~ current_feature + Sex + (1 | Subject) + storage_time
y ~ current_feature + Sex + (1 | Subject) + storage_time
y ~ current_feature + Sex + (1 | Subject) + storage_time
y ~ current_feature + Sex + (1 | Subject) + storage_time
y ~ current_feature + Sex + (1 | Subject) + storage_time
y ~ current_feature + Sex + (1 | Subject) + storage_time
y ~ current_feature + Sex + (1 | Subject) + storage_time
y ~ current_feature + Sex + (1 | Subject) + storage_time
y ~ current_feature + Sex + (1 | Subject) + storage_time
y ~ current_feature + Sex + (1 | Subject) + storage_time
y ~ current_feature + Sex + (1 | Subject) + storage_time
y ~ current_feature + Sex + (1 | Subject) + storage_time
y ~ current_feature + Sex + (1 | Subject) + storage_time
y ~ current_feature + Sex + (1 | Subject) + storage_time
y ~ current_feature + Sex + (1 | Subject) + storage_time
y ~ current_feature + Sex + (1 | Subject) + storage_time
y ~ current_feature + Sex + (1 | Subject) + storage_time
y ~ current_feature + Sex + (1 | Subject) + storage_time
y ~ current_feature + Sex + (1 | Subject) + storage_time
y ~ current_feature + Sex + (1 | Subject) + storage_time
y ~ current_feature + Sex + (1 | Subject) + storage_time
y ~ current_feature + Sex + (1 | Subject) + storage_time
y ~ current_feature + Sex + (1 | Subject) + storage_time
y ~ current_feature + Sex + (1 | Subject) + storage_time
y ~ current_feature + Sex + (1 | Subject) + storage_time
y ~ current_feature + Sex + (1 | Subject) + storage_time
y ~ current_feature + Sex + (1 | Subject) + storage_time

y ~ current_feature + Sex + (1 | Subject) + storage_time
y ~ current_feature + Sex + (1 | Subject) + storage_time
y ~ current_feature + Sex + (1 | Subject) + storage_time
y ~ current_feature + Sex + (1 | Subject) + storage_time
y ~ current_feature + Sex + (1 | Subject) + storage_time
y ~ current_feature + Sex + (1 | Subject) + storage_time
y ~ current_feature + Sex + (1 | Subject) + storage_time
y ~ current_feature + Sex + (1 | Subject) + storage_time
y ~ current_feature + Sex + (1 | Subject) + storage_time
y ~ current_feature + Sex + (1 | Subject) + storage_time
y ~ current_feature + Sex + (1 | Subject) + storage_time
y ~ current_feature + Sex + (1 | Subject) + storage_time
y ~ current_feature + Sex + (1 | Subject) + storage_time
y ~ current_feature + Sex + (1 | Subject) + storage_time
y ~ current_feature + Sex + (1 | Subject) + storage_time
y ~ current_feature + Sex + (1 | Subject) + storage_time
y ~ current_feature + Sex + (1 | Subject) + storage_time
y ~ current_feature + Sex + (1 | Subject) + storage_time
y ~ current_feature + Sex + (1 | Subject) + storage_time
y ~ current_feature + Sex + (1 | Subject) + storage_time
y ~ current_feature + Sex + (1 | Subject) + storage_time
y ~ current_feature + Sex + (1 | Subject) + storage_time
y ~ current_feature + Sex + (1 | Subject) + storage_time
y ~ current_feature + Sex + (1 | Subject) + storage_time
y ~ current_feature + Sex + (1 | Subject) + storage_time
y ~ current_feature + Sex + (1 | Subject) + storage_time
y ~ current_feature + Sex + (1 | Subject) + storage_time
y ~ current_feature + Sex + (1 | Subject) + storage_time
y ~ current_feature + Sex + (1 | Subject) + storage_time
y ~ current_feature + Sex + (1 | Subject) + storage_time
y ~ current_feature + Sex + (1 | Subject) + storage_time
y ~ current_feature + Sex + (1 | Subject) + storage_time
y ~ current_feature + Sex + (1 | Subject) + storage_time
y ~ current_feature + Sex + (1 | Subject) + storage_time
y ~ current_feature + Sex + (1 | Subject) + storage_time
y ~ current_feature + Sex + (1 | Subject) + storage_time
y ~ current_feature + Sex + (1 | Subject) + storage_time
y ~ current_feature + Sex + (1 | Subject) + storage_time
y ~ current_feature + Sex + (1 | Subject) + storage_time
y ~ current_feature + Sex + (1 | Subject) + storage_time
y ~ current_feature + Sex + (1 | Subject) + storage_time
y ~ current_feature + Sex + (1 | Subject) + storage_time
y ~ current_feature + Sex + (1 | Subject) + storage_time
y ~ current_feature + Sex + (1 | Subject) + storage_time
y ~ current_feature + Sex + (1 | Subject) + storage_time
y ~ current_feature + Sex + (1 | Subject) + storage_time
y ~ current_feature + Sex + (1 | Subject) + storage_time
y ~ current_feature + Sex + (1 | Subject) + storage_time

```
y ~ current_feature + Sex + (1 | Subject) + storage_time
y ~ current_feature + Sex + (1 | Subject) + storage_time
y ~ current_feature + Sex + (1 | Subject) + storage_time
y ~ current_feature + Sex + (1 | Subject) + storage_time
```

| Name unique |
| --- |
| 2-Methylpropanoic acid |
| 2-Methylpropanoic acid |
| 2-Methylpropanoic acid |
| 2-Methylpropanoic acid |
| 2-Methylpropanoic acid |
| 3-Methylbutanoic acid |
| 3-Methylbutanoic acid |
| 3-Methylbutanoic acid |
| 3-Methylbutanoic acid |
| 3-Methylbutanoic acid |
| Acetic acid |
| Acetic acid |
| Acetic acid |
| Acetic acid |
| Acetic acid |
| Butanoic acid |
| Butanoic acid |
| Butanoic acid |
| Butanoic acid |
| Butanoic acid |
| Hexanoic acid |
| Hexanoic acid |
| Hexanoic acid |
| Hexanoic acid |
| Hexanoic acid |
| M1 |
| M1 |
| M1 |
| M1 |
| M1 |
| M10 |
| M10 |
| M10 |
| M10 |
| M10 |
| M11 |
| M11 |
| M11 |
| M11 |
| M11 |
| M12 |
| M12 |
| M12 |
| M12 |
| M12 |
| M13 |
| M13 |

M13
M13
M13
M14
M14
M14
M14
M14
M15
M15
M15
M15
M15
M16
M16
M16
M16
M16
M17
M17
M17
M17
M17
M18
M18
M18
M18
M18
M19
M19
M19
M19
M19
M2
M2
M2
M2
M2
M20
M20
M20
M20
M20
M21
M21
M21
M21
M21

M22
M22
M22
M22
M22
M23
M23
M23
M23
M23
M24
M24
M24
M24
M24
M25
M25
M25
M25
M25
M26
M26
M26
M26
M26
M27
M27
M27
M27
M27
M28
M28
M28
M28
M28
M29
M29
M29
M29
M29
M3
M3
M3
M3
M3
M30
M30
M30

M30
M30
M31
M31
M31
M31
M31
M32
M32
M32
M32
M32
M33
M33
M33
M33
M33
M34
M34
M34
M34
M34
M4
M4
M4
M4
M4
M5
M5
M5
M5
M5
M6
M6
M6
M6
M6
M7
M7
M7
M7
M7
M8
M8
M8
M8
M8
M9

M9
M9
M9
M9
Pentanoic acid
Pentanoic acid
Pentanoic acid
Pentanoic acid
Pentanoic acid
Propanoic acid
Propanoic acid
Propanoic acid
Propanoic acid
Propanoic acid
Deoxycytidine
Deoxycytidine
Deoxycytidine
Deoxycytidine
Deoxycytidine
N-Acetylhistidine
N-Acetylhistidine
N-Acetylhistidine
N-Acetylhistidine
N-Acetylhistidine
Aspartic acid
Aspartic acid
Aspartic acid
Aspartic acid
Aspartic acid
Carnitine
Carnitine
Carnitine
Carnitine
Carnitine
Creatine
Creatine
Creatine
Creatine
Creatine
Glycylglycine
Glycylglycine
Glycylglycine
Glycylglycine
Glycylglycine
Kynurenine
Kynurenine
Kynurenine
Kynurenine

Kynurenine
N-Acetylarginine
N-Acetylarginine
N-Acetylarginine
N-Acetylarginine
N-Acetylarginine
N-Isovalerylglycine
N-Isovalerylglycine
N-Isovalerylglycine
N-Isovalerylglycine
N-Isovalerylglycine
Ornithine
Ornithine
Ornithine
Ornithine
Ornithine
Proline
Proline
Proline
Proline
Proline
Pyridoxal
Pyridoxal
Pyridoxal
Pyridoxal
Pyridoxal
Valine/ 5-Aminovaleric acid
Valine/ 5-Aminovaleric acid
Valine/ 5-Aminovaleric acid
Valine/ 5-Aminovaleric acid
Valine/ 5-Aminovaleric acid
5-Methylcytosine
5-Methylcytosine
5-Methylcytosine
5-Methylcytosine
5-Methylcytosine
Allopurinol
Allopurinol
Allopurinol
Allopurinol
Allopurinol
Dissaccarides I
Dissaccarides I
Dissaccarides I
Dissaccarides I
Dissaccarides I
Hexoses III
Hexoses III

Hexoses III
Hexoses III
Hexoses III
Hexoses II
Hexoses II
Hexoses II
Hexoses II
Hexoses II
Gulonic acid Î³-lactone
Gulonic acid Î³-lactone
Gulonic acid Î³-lactone
Gulonic acid Î³-lactone
Gulonic acid Î³-lactone
Hexoses I
Hexoses I
Hexoses I
Hexoses I
Hexoses I
Histidinol
Histidinol
Histidinol
Histidinol
Histidinol
Dissaccarides III
Dissaccarides III
Dissaccarides III
Dissaccarides III
Dissaccarides III
N-Acetylalanine
N-Acetylalanine
N-Acetylalanine
N-Acetylalanine
N-Acetylalanine
N-Acetylglutamine
N-Acetylglutamine
N-Acetylglutamine
N-Acetylglutamine
N-Acetylglutamine
Phenylacetylglutamine
Phenylacetylglutamine
Phenylacetylglutamine
Phenylacetylglutamine
Phenylacetylglutamine
Pyridoxamine
Pyridoxamine
Pyridoxamine
Pyridoxamine
Pyridoxamine

Dissaccarides II
Dissaccarides II
Dissaccarides II
Dissaccarides II
Dissaccarides II
Trigonelline
Trigonelline
Trigonelline
Trigonelline
Trigonelline
Uracil
Uracil
Uracil
Uracil
Uracil
Uridine
Uridine
Uridine
Uridine
Uridine
2-Oxo-3-phenylpropanoic acid
2-Oxo-3-phenylpropanoic acid
2-Oxo-3-phenylpropanoic acid
2-Oxo-3-phenylpropanoic acid
2-Oxo-3-phenylpropanoic acid
3-Hydroxybutyric acid
3-Hydroxybutyric acid
3-Hydroxybutyric acid
3-Hydroxybutyric acid
3-Hydroxybutyric acid
4-Methyl-2-oxovaleric acid
4-Methyl-2-oxovaleric acid
4-Methyl-2-oxovaleric acid
4-Methyl-2-oxovaleric acid
4-Methyl-2-oxovaleric acid
5-Hydroxyindole-3-acetic acid
5-Hydroxyindole-3-acetic acid
5-Hydroxyindole-3-acetic acid
5-Hydroxyindole-3-acetic acid
5-Hydroxyindole-3-acetic acid
8-Aminooctanoic acid
8-Aminooctanoic acid
8-Aminooctanoic acid
8-Aminooctanoic acid
8-Aminooctanoic acid
Pentose II
Pentose II
Pentose II

Pentose II
Pentose II
Citraconic acid
Citraconic acid
Citraconic acid
Citraconic acid
Citraconic acid
Galactonic acid
Galactonic acid
Galactonic acid
Galactonic acid
Galactonic acid
Galactosamine
Galactosamine
Galactosamine
Galactosamine
Galactosamine
Glycolic acid
Glycolic acid
Glycolic acid
Glycolic acid
Glycolic acid
Glyoxylic acid
Glyoxylic acid
Glyoxylic acid
Glyoxylic acid
Glyoxylic acid
Lactic acid
Lactic acid
Lactic acid
Lactic acid
Lactic acid
N-Acetylglycine
N-Acetylglycine
N-Acetylglycine
N-Acetylglycine
N-Acetylglycine
Pinitol
Pinitol
Pinitol
Pinitol
Pinitol
Pyruvic acid
Pyruvic acid
Pyruvic acid
Pyruvic acid
Pyruvic acid
Deoxysugar I (possibly Rhamnose/2-Deoxyglucose)

Deoxysugar I (possibly Rhamnose/2-Deoxyglucose)
Deoxysugar I (possibly Rhamnose/2-Deoxyglucose)
Deoxysugar I (possibly Rhamnose/2-Deoxyglucose)
Deoxysugar I (possibly Rhamnose/2-Deoxyglucose)
sugar alcohol (C5)
sugar alcohol (C5)
sugar alcohol (C5)
sugar alcohol (C5)
sugar alcohol (C5)
Pentose III
Pentose III
Pentose III
Pentose III
Pentose III
Shikimic acid
Shikimic acid
Shikimic acid
Shikimic acid
Shikimic acid
Gluconic acid
Gluconic acid
Gluconic acid
Gluconic acid
Gluconic acid
Pentose I
Pentose I
Pentose I
Pentose I
Pentose I
cis-Aconitic acid
cis-Aconitic acid
cis-Aconitic acid
cis-Aconitic acid
cis-Aconitic acid
Methyl acetoacetate
Methyl acetoacetate
Methyl acetoacetate
Methyl acetoacetate
Methyl acetoacetate
Purine
Purine
Purine
Purine
Purine
Thymine
Thymine
Thymine
Thymine

Thymine
3-hydroxy-4,5-dimethyl-3(5H)-furanone
3-hydroxy-4,5-dimethyl-3(5H)-furanone
3-hydroxy-4,5-dimethyl-3(5H)-furanone
3-hydroxy-4,5-dimethyl-3(5H)-furanone
3-hydroxy-4,5-dimethyl-3(5H)-furanone
Dopamine
Dopamine
Dopamine
Dopamine
Dopamine
3-Hydroxyhippuric acid
3-Hydroxyhippuric acid
3-Hydroxyhippuric acid
3-Hydroxyhippuric acid
3-Hydroxyhippuric acid
Dihydroferulic acid
Dihydroferulic acid
Dihydroferulic acid
Dihydroferulic acid
Dihydroferulic acid
DOPA
DOPA
DOPA
DOPA
DOPA
Propionylcarnitine
Propionylcarnitine
Propionylcarnitine
Propionylcarnitine
Propionylcarnitine
N-Acetylmannosamine
N-Acetylmannosamine
N-Acetylmannosamine
N-Acetylmannosamine
N-Acetylmannosamine
Butyrylcarnitine
Butyrylcarnitine
Butyrylcarnitine
Butyrylcarnitine
Butyrylcarnitine
5-Methyluridine
5-Methyluridine
5-Methyluridine
5-Methyluridine
5-Methyluridine
Acetylmuramic acid
Acetylmuramic acid

Acetylmuramic acid
Acetylmuramic acid
Acetylmuramic acid
Î²-Muricholic acid/ Ï‰-Muricholic acid
Î²-Muricholic acid/ Ï‰-Muricholic acid
Î²-Muricholic acid/ Ï‰-Muricholic acid
Î²-Muricholic acid/ Ï‰-Muricholic acid
Î²-Muricholic acid/ Ï‰-Muricholic acid
1-Aminocyclopropanecarboxylic acid
1-Aminocyclopropanecarboxylic acid
1-Aminocyclopropanecarboxylic acid
1-Aminocyclopropanecarboxylic acid
1-Aminocyclopropanecarboxylic acid
N,N-Dimethylglycine
N,N-Dimethylglycine
N,N-Dimethylglycine
N,N-Dimethylglycine
N,N-Dimethylglycine
2-Aminoisobutyric acid
2-Aminoisobutyric acid
2-Aminoisobutyric acid
2-Aminoisobutyric acid
2-Aminoisobutyric acid
Malonic acid
Malonic acid
Malonic acid
Malonic acid
Malonic acid
4-Aminophenol
4-Aminophenol
4-Aminophenol
4-Aminophenol
4-Aminophenol
Î³-Caprolactone
Î³-Caprolactone
Î³-Caprolactone
Î³-Caprolactone
Î³-Caprolactone
2-Methylmalonic acid
2-Methylmalonic acid
2-Methylmalonic acid
2-Methylmalonic acid
2-Methylmalonic acid
2-(hydroxymethyl)butanoic acid
2-(hydroxymethyl)butanoic acid
2-(hydroxymethyl)butanoic acid
2-(hydroxymethyl)butanoic acid
2-(hydroxymethyl)butanoic acid

2-Hydroxy-3-methylbutyric acid
2-Hydroxy-3-methylbutyric acid
2-Hydroxy-3-methylbutyric acid
2-Hydroxy-3-methylbutyric acid
2-Hydroxy-3-methylbutyric acid
Imidazoleacetic acid
Imidazoleacetic acid
Imidazoleacetic acid
Imidazoleacetic acid
Imidazoleacetic acid
N-Methylnicotinamide
N-Methylnicotinamide
N-Methylnicotinamide
N-Methylnicotinamide
N-Methylnicotinamide
1-Aminocyclohexanecarboxylic acid
1-Aminocyclohexanecarboxylic acid
1-Aminocyclohexanecarboxylic acid
1-Aminocyclohexanecarboxylic acid
1-Aminocyclohexanecarboxylic acid
Stachydrine
Stachydrine
Stachydrine
Stachydrine
Stachydrine
Adipic acid
Adipic acid
Adipic acid
Adipic acid
Adipic acid
N-(5-Aminopentyl)acetamide
N-(5-Aminopentyl)acetamide
N-(5-Aminopentyl)acetamide
N-(5-Aminopentyl)acetamide
N-(5-Aminopentyl)acetamide
N-Methyltyramine
N-Methyltyramine
N-Methyltyramine
N-Methyltyramine
N-Methyltyramine
N1-Methyl-2-pyridone-5-carboxamide
N1-Methyl-2-pyridone-5-carboxamide
N1-Methyl-2-pyridone-5-carboxamide
N1-Methyl-2-pyridone-5-carboxamide
N1-Methyl-2-pyridone-5-carboxamide
N-AcetylProline
N-AcetylProline
N-AcetylProline

N-AcetylProline
N-AcetylProline
2-Aminoadipic acid
2-Aminoadipic acid
2-Aminoadipic acid
2-Aminoadipic acid
2-Aminoadipic acid
3-(2-Hydroxyethyl)indole
3-(2-Hydroxyethyl)indole
3-(2-Hydroxyethyl)indole
3-(2-Hydroxyethyl)indole
3-(2-Hydroxyethyl)indole
p-Coumaric acid
p-Coumaric acid
p-Coumaric acid
p-Coumaric acid
p-Coumaric acid
Gallic acid
Gallic acid
Gallic acid
Gallic acid
Gallic acid
N-Acetylleucine/ Hexanoylglycine
N-Acetylleucine/ Hexanoylglycine
N-Acetylleucine/ Hexanoylglycine
N-Acetylleucine/ Hexanoylglycine
N-Acetylleucine/ Hexanoylglycine
Theophylline
Theophylline
Theophylline
Theophylline
Theophylline
Homovanilic acid
Homovanilic acid
Homovanilic acid
Homovanilic acid
Homovanilic acid
4-Hydroxy-3-methoxymandelic acid
4-Hydroxy-3-methoxymandelic acid
4-Hydroxy-3-methoxymandelic acid
4-Hydroxy-3-methoxymandelic acid
4-Hydroxy-3-methoxymandelic acid
Asymmetric dimethylarginine
Asymmetric dimethylarginine
Asymmetric dimethylarginine
Asymmetric dimethylarginine
Asymmetric dimethylarginine
Homocarnosine/ Anserine

Homocarnosine/ Anserine
Homocarnosine/ Anserine
Homocarnosine/ Anserine
Homocarnosine/ Anserine
Tiglylcarnitine
Tiglylcarnitine
Tiglylcarnitine
Tiglylcarnitine
Tiglylcarnitine
Isovalerylcarnitine
Isovalerylcarnitine
Isovalerylcarnitine
Isovalerylcarnitine
Isovalerylcarnitine
1-Carboxyethyltyrosine
1-Carboxyethyltyrosine
1-Carboxyethyltyrosine
1-Carboxyethyltyrosine
1-Carboxyethyltyrosine
Daidzein
Daidzein
Daidzein
Daidzein
Daidzein
5-Methylcytidine
5-Methylcytidine
5-Methylcytidine
5-Methylcytidine
5-Methylcytidine
Glucosamine-6-phosphate
Glucosamine-6-phosphate
Glucosamine-6-phosphate
Glucosamine-6-phosphate
Glucosamine-6-phosphate
Apigenin
Apigenin
Apigenin
Apigenin
Apigenin
12,13-DHOME
12,13-DHOME
12,13-DHOME
12,13-DHOME
12,13-DHOME
Sucralose
Sucralose
Sucralose
Sucralose

Sucralose
2-Ketobutyric acid
2-Ketobutyric acid
2-Ketobutyric acid
2-Ketobutyric acid
2-Ketobutyric acid
Glutamine
Glutamine
Glutamine
Glutamine
Glutamine
Succinic acid
Succinic acid
Succinic acid
Succinic acid
Succinic acid
sugar alcohol (C6)
sugar alcohol (C6)
sugar alcohol (C6)
sugar alcohol (C6)
sugar alcohol (C6)
N-Methylaspartic acid
N-Methylaspartic acid
N-Methylaspartic acid
N-Methylaspartic acid
N-Methylaspartic acid
N-Formylmethionine
N-Formylmethionine
N-Formylmethionine
N-Formylmethionine
N-Formylmethionine
Acesulfame
Acesulfame
Acesulfame
Acesulfame
Acesulfame
Inosine
Inosine
Inosine
Inosine
Inosine
N-Acetylglutamic acid
N-Acetylglutamic acid
N-Acetylglutamic acid
N-Acetylglutamic acid
N-Acetylglutamic acid
Urocanic acid
Urocanic acid

Urocanic acid
Urocanic acid
Urocanic acid
3,4-Dihydroxybenzoic acid
3,4-Dihydroxybenzoic acid
3,4-Dihydroxybenzoic acid
3,4-Dihydroxybenzoic acid
3,4-Dihydroxybenzoic acid
Indole-3-propionic acid
Indole-3-propionic acid
Indole-3-propionic acid
Indole-3-propionic acid
Indole-3-propionic acid
Deoxycholic acid
Deoxycholic acid
Deoxycholic acid
Deoxycholic acid
Deoxycholic acid
Pyroglutamic acid
Pyroglutamic acid
Pyroglutamic acid
Pyroglutamic acid
Pyroglutamic acid
Choline
Choline
Choline
Choline
Choline
Serine
Serine
Serine
Serine
Serine
Histamine
Histamine
Histamine
Histamine
Histamine
N-Acetyltyrosine
N-Acetyltyrosine
N-Acetyltyrosine
N-Acetyltyrosine
N-Acetyltyrosine
Glucuronic acid
Glucuronic acid
Glucuronic acid
Glucuronic acid
Glucuronic acid

Threonine
Threonine
Threonine
Threonine
Threonine
Î²-Hydroxyisovaleric acid
Î²-Hydroxyisovaleric acid
Î²-Hydroxyisovaleric acid
Î²-Hydroxyisovaleric acid
Î²-Hydroxyisovaleric acid
Î²-D-Glucopyranuronic acid
Î²-D-Glucopyranuronic acid
Î²-D-Glucopyranuronic acid
Î²-D-Glucopyranuronic acid
Î²-D-Glucopyranuronic acid
3-(4-hydroxyphenyl)propionic acid
3-(4-hydroxyphenyl)propionic acid
3-(4-hydroxyphenyl)propionic acid
3-(4-hydroxyphenyl)propionic acid
3-(4-hydroxyphenyl)propionic acid
N-Acetylornithine
N-Acetylornithine
N-Acetylornithine
N-Acetylornithine
N-Acetylornithine
3,4-Dihydroxyhydrocinnamic acid
3,4-Dihydroxyhydrocinnamic acid
3,4-Dihydroxyhydrocinnamic acid
3,4-Dihydroxyhydrocinnamic acid
3,4-Dihydroxyhydrocinnamic acid
Ethylmalonic acid
Ethylmalonic acid
Ethylmalonic acid
Ethylmalonic acid
Ethylmalonic acid
Thiamine
Thiamine
Thiamine
Thiamine
Thiamine
2-Hydroxycaproic acid
2-Hydroxycaproic acid
2-Hydroxycaproic acid
2-Hydroxycaproic acid
2-Hydroxycaproic acid
Traumatic acid
Traumatic acid
Traumatic acid

Traumatic acid
Traumatic acid
Tryptamine
Tryptamine
Tryptamine
Tryptamine
Tryptamine
Hypoxanthine
Hypoxanthine
Hypoxanthine
Hypoxanthine
Hypoxanthine
N-Acetylmethionine
N-Acetylmethionine
N-Acetylmethionine
N-Acetylmethionine
N-Acetylmethionine
Î±-aminobutyric acid
Î±-aminobutyric acid
Î±-aminobutyric acid
Î±-aminobutyric acid
Î±-aminobutyric acid
Tyrosine
Tyrosine
Tyrosine
Tyrosine
Tyrosine
3,5-Dihydroxybenzoic acid
3,5-Dihydroxybenzoic acid
3,5-Dihydroxybenzoic acid
3,5-Dihydroxybenzoic acid
3,5-Dihydroxybenzoic acid
Indole-3-lactic acid
Indole-3-lactic acid
Indole-3-lactic acid
Indole-3-lactic acid
Indole-3-lactic acid
Methylsuccinic acid
Methylsuccinic acid
Methylsuccinic acid
Methylsuccinic acid
Methylsuccinic acid
Isoleucine
Isoleucine
Isoleucine
Isoleucine
Isoleucine
Acetylagmatine

Acetylagmatine
Acetylagmatine
Acetylagmatine
Acetylagmatine
Cholic acid
Cholic acid
Cholic acid
Cholic acid
Cholic acid
1,7-Dimethylxanthine
1,7-Dimethylxanthine
1,7-Dimethylxanthine
1,7-Dimethylxanthine
1,7-Dimethylxanthine
Tricarballylic acid
Tricarballylic acid
Tricarballylic acid
Tricarballylic acid
Tricarballylic acid
Pantothenic acid
Pantothenic acid
Pantothenic acid
Pantothenic acid
Pantothenic acid
4-Hydroxybenzoic acid
4-Hydroxybenzoic acid
4-Hydroxybenzoic acid
4-Hydroxybenzoic acid
4-Hydroxybenzoic acid
Malic acid
Malic acid
Malic acid
Malic acid
Malic acid
3-Methylhistidine
3-Methylhistidine
3-Methylhistidine
3-Methylhistidine
3-Methylhistidine
Deoxyinosine
Deoxyinosine
Deoxyinosine
Deoxyinosine
Deoxyinosine
Methionine
Methionine
Methionine
Methionine

Methionine
2,6-Dihydroxybenzoic acid
2,6-Dihydroxybenzoic acid
2,6-Dihydroxybenzoic acid
2,6-Dihydroxybenzoic acid
2,6-Dihydroxybenzoic acid
7-Methylguanine
7-Methylguanine
7-Methylguanine
7-Methylguanine
7-Methylguanine
Threonic acid
Threonic acid
Threonic acid
Threonic acid
Threonic acid
Tryptophan
Tryptophan
Tryptophan
Tryptophan
Tryptophan
N-Acetyltryptophan
N-Acetyltryptophan
N-Acetyltryptophan
N-Acetyltryptophan
N-Acetyltryptophan
N6-Acetyllysine
N6-Acetyllysine
N6-Acetyllysine
N6-Acetyllysine
N6-Acetyllysine
Xanthosine
Xanthosine
Xanthosine
Xanthosine
Xanthosine
N-Acetylphenylalanine
N-Acetylphenylalanine
N-Acetylphenylalanine
N-Acetylphenylalanine
N-Acetylphenylalanine
Indole-3-methyl acetate
Indole-3-methyl acetate
Indole-3-methyl acetate
Indole-3-methyl acetate
Indole-3-methyl acetate
Histidine
Histidine

Histidine
Histidine
Histidine
Cytidine
Cytidine
Cytidine
Cytidine
Cytidine
Leucylalanine
Leucylalanine
Leucylalanine
Leucylalanine
Leucylalanine
Phenylalanine
Phenylalanine
Phenylalanine
Phenylalanine
Phenylalanine
Leucine
Leucine
Leucine
Leucine
Leucine
Taurine
Taurine
Taurine
Taurine
Taurine
Nicotinic acid
Nicotinic acid
Nicotinic acid
Nicotinic acid
Nicotinic acid
Quinic acid
Quinic acid
Quinic acid
Quinic acid
Quinic acid
Deoxyuridine
Deoxyuridine
Deoxyuridine
Deoxyuridine
Deoxyuridine
Glyceric acid
Glyceric acid
Glyceric acid
Glyceric acid
Glyceric acid

N-Acetylglucosamine
N-Acetylglucosamine
N-Acetylglucosamine
N-Acetylglucosamine
N-Acetylglucosamine
leu-gln_a
leu-gln_a
leu-gln_a
leu-gln_a
leu-gln_a
primidone_a
primidone_a
primidone_a
primidone_a
primidone_a
1-(2-Carboxyethyl)-2,3,4,9-tetrahydro-1H-beta-carboline-3-carboxylic acid_a
1-(2-Carboxyethyl)-2,3,4,9-tetrahydro-1H-beta-carboline-3-carboxylic acid_a
1-(2-Carboxyethyl)-2,3,4,9-tetrahydro-1H-beta-carboline-3-carboxylic acid_a
1-(2-Carboxyethyl)-2,3,4,9-tetrahydro-1H-beta-carboline-3-carboxylic acid_a
1-(2-Carboxyethyl)-2,3,4,9-tetrahydro-1H-beta-carboline-3-carboxylic acid_a
3-(Butylsulfinyl)-L-alanin
3-(Butylsulfinyl)-L-alanin
3-(Butylsulfinyl)-L-alanin
3-(Butylsulfinyl)-L-alanin
3-(Butylsulfinyl)-L-alanin
tert-Butyl 3-amino-1-methyl-2,3-dioxopropylcarbamate_b
tert-Butyl 3-amino-1-methyl-2,3-dioxopropylcarbamate_b
tert-Butyl 3-amino-1-methyl-2,3-dioxopropylcarbamate_b
tert-Butyl 3-amino-1-methyl-2,3-dioxopropylcarbamate_b
tert-Butyl 3-amino-1-methyl-2,3-dioxopropylcarbamate_b
4-O-{3-O-[(1S,2R,3S,4S,5R)-2,3,4-Trihydroxy-5-(hydroxymethyl)cyclohexyl]-beta-D-galactopyranosyl}-D-glucose
4-O-{3-O-[(1S,2R,3S,4S,5R)-2,3,4-Trihydroxy-5-(hydroxymethyl)cyclohexyl]-beta-D-galactopyranosyl}-D-glucose
4-O-{3-O-[(1S,2R,3S,4S,5R)-2,3,4-Trihydroxy-5-(hydroxymethyl)cyclohexyl]-beta-D-galactopyranosyl}-D-glucose
4-O-{3-O-[(1S,2R,3S,4S,5R)-2,3,4-Trihydroxy-5-(hydroxymethyl)cyclohexyl]-beta-D-galactopyranosyl}-D-glucose
4-O-{3-O-[(1S,2R,3S,4S,5R)-2,3,4-Trihydroxy-5-(hydroxymethyl)cyclohexyl]-beta-D-galactopyranosyl}-D-glucose
Astemizole_b
Astemizole_b
Astemizole_b
Astemizole_b
Astemizole_b
Nifedipine
Nifedipine
Nifedipine
Nifedipine
Nifedipine
g-Aminobutyryl-lysine_d
g-Aminobutyryl-lysine_d
g-Aminobutyryl-lysine_d

g-Aminobutyryl-lysine_d
g-Aminobutyryl-lysine_d
OI1700000
OI1700000
OI1700000
OI1700000
OI1700000
IN00258
IN00258
IN00258
IN00258
IN00258
TDP-2_a
TDP-2_a
TDP-2_a
TDP-2_a
TDP-2_a
Mexiletine
Mexiletine
Mexiletine
Mexiletine
Mexiletine
PEG n12
PEG n12
PEG n12
PEG n12
PEG n12
2-Oxo-3-(phosphonooxy)propyl decanoate
2-Oxo-3-(phosphonooxy)propyl decanoate
2-Oxo-3-(phosphonooxy)propyl decanoate
2-Oxo-3-(phosphonooxy)propyl decanoate
2-Oxo-3-(phosphonooxy)propyl decanoate
Formylkynurenine_b
Formylkynurenine_b
Formylkynurenine_b
Formylkynurenine_b
Formylkynurenine_b
Panthenol_b
Panthenol_b
Panthenol_b
Panthenol_b
Panthenol_b
3-Hydroxy-2-(hydroxymethyl)-2-[(sulfooxy)methyl]propanoic acid
3-Hydroxy-2-(hydroxymethyl)-2-[(sulfooxy)methyl]propanoic acid
3-Hydroxy-2-(hydroxymethyl)-2-[(sulfooxy)methyl]propanoic acid
3-Hydroxy-2-(hydroxymethyl)-2-[(sulfooxy)methyl]propanoic acid
3-Hydroxy-2-(hydroxymethyl)-2-[(sulfooxy)methyl]propanoic acid
4-(9H-beta-Carbolin-1-yl)-1,2,4-butanetriol_b

4-(9H-beta-Carbolin-1-yl)-1,2,4-butanetriol_b
4-(9H-beta-Carbolin-1-yl)-1,2,4-butanetriol_b
4-(9H-beta-Carbolin-1-yl)-1,2,4-butanetriol_b
4-(9H-beta-Carbolin-1-yl)-1,2,4-butanetriol_b
SECONAL_f
SECONAL_f
SECONAL_f
SECONAL_f
SECONAL_f
nitecapone
nitecapone
nitecapone
nitecapone
nitecapone
hexobarbital_a
hexobarbital_a
hexobarbital_a
hexobarbital_a
hexobarbital_a
N-[(2S)-2-Hydroxypropanoyl]methionine_c
N-[(2S)-2-Hydroxypropanoyl]methionine_c
N-[(2S)-2-Hydroxypropanoyl]methionine_c
N-[(2S)-2-Hydroxypropanoyl]methionine_c
N-[(2S)-2-Hydroxypropanoyl]methionine_c
pretazettine
pretazettine
pretazettine
pretazettine
pretazettine
3-Formyl-2a-methoxy-6,6,7b-trimethyl-2,2a,4a,5,6,7,7a,7b-octahydro-1H-cyclobuta[e]inden-2-yl 3-chloro-6-hyd[illegible]
3-Formyl-2a-methoxy-6,6,7b-trimethyl-2,2a,4a,5,6,7,7a,7b-octahydro-1H-cyclobuta[e]inden-2-yl 3-chloro-6-hyd[illegible]
3-Formyl-2a-methoxy-6,6,7b-trimethyl-2,2a,4a,5,6,7,7a,7b-octahydro-1H-cyclobuta[e]inden-2-yl 3-chloro-6-hyd[illegible]
3-Formyl-2a-methoxy-6,6,7b-trimethyl-2,2a,4a,5,6,7,7a,7b-octahydro-1H-cyclobuta[e]inden-2-yl 3-chloro-6-hyd[illegible]
3-Formyl-2a-methoxy-6,6,7b-trimethyl-2,2a,4a,5,6,7,7a,7b-octahydro-1H-cyclobuta[e]inden-2-yl 3-chloro-6-hyd[illegible]
7alpha-Hydroxy-3-oxochol-4-en-24-oic acid_a
7alpha-Hydroxy-3-oxochol-4-en-24-oic acid_a
7alpha-Hydroxy-3-oxochol-4-en-24-oic acid_a
7alpha-Hydroxy-3-oxochol-4-en-24-oic acid_a
7alpha-Hydroxy-3-oxochol-4-en-24-oic acid_a
piscidic acid_a
piscidic acid_a
piscidic acid_a
piscidic acid_a
piscidic acid_a
8-(Methylsulfinyl)octyl isothiocyanate
8-(Methylsulfinyl)octyl isothiocyanate
8-(Methylsulfinyl)octyl isothiocyanate
8-(Methylsulfinyl)octyl isothiocyanate

8-(Methylsulfinyl)octyl isothiocyanate
Gly-Ser
Gly-Ser
Gly-Ser
Gly-Ser
Gly-Ser
Sparfloxacin
Sparfloxacin
Sparfloxacin
Sparfloxacin
Sparfloxacin
Nisinic acid_a
Nisinic acid_a
Nisinic acid_a
Nisinic acid_a
Nisinic acid_a
R-(+)-Etiracetam_b
R-(+)-Etiracetam_b
R-(+)-Etiracetam_b
R-(+)-Etiracetam_b
R-(+)-Etiracetam_b
3-hydroxyoctanoylcarnitine
3-hydroxyoctanoylcarnitine
3-hydroxyoctanoylcarnitine
3-hydroxyoctanoylcarnitine
3-hydroxyoctanoylcarnitine
3-[(2Z)-1-Oxo-2-buten-2-yl]pentanedioic acid_a
3-[(2Z)-1-Oxo-2-buten-2-yl]pentanedioic acid_a
3-[(2Z)-1-Oxo-2-buten-2-yl]pentanedioic acid_a
3-[(2Z)-1-Oxo-2-buten-2-yl]pentanedioic acid_a
3-[(2Z)-1-Oxo-2-buten-2-yl]pentanedioic acid_a
miglustat
miglustat
miglustat
miglustat
miglustat
Nicotinamide
Nicotinamide
Nicotinamide
Nicotinamide
Nicotinamide
4-(5,6-Dihydroxy-6-methyl-1-hepten-2-yl)-1-methyl-1,2-cyclohexanediol
4-(5,6-Dihydroxy-6-methyl-1-hepten-2-yl)-1-methyl-1,2-cyclohexanediol
4-(5,6-Dihydroxy-6-methyl-1-hepten-2-yl)-1-methyl-1,2-cyclohexanediol
4-(5,6-Dihydroxy-6-methyl-1-hepten-2-yl)-1-methyl-1,2-cyclohexanediol
4-(5,6-Dihydroxy-6-methyl-1-hepten-2-yl)-1-methyl-1,2-cyclohexanediol
N~6~,N~6~-Dimethyllysine_a
N~6~,N~6~-Dimethyllysine_a

N~6~,N~6~-Dimethyllysine_a
N~6~,N~6~-Dimethyllysine_a
N~6~,N~6~-Dimethyllysine_a
Deacetyldiltiazem
Deacetyldiltiazem
Deacetyldiltiazem
Deacetyldiltiazem
Deacetyldiltiazem
7-Chloro-5-(2-fluorophenyl)-1-(2-methoxyethyl)-1,3-dihydro-2H-1,4-benzodiazepin-2-one_b
7-Chloro-5-(2-fluorophenyl)-1-(2-methoxyethyl)-1,3-dihydro-2H-1,4-benzodiazepin-2-one_b
7-Chloro-5-(2-fluorophenyl)-1-(2-methoxyethyl)-1,3-dihydro-2H-1,4-benzodiazepin-2-one_b
7-Chloro-5-(2-fluorophenyl)-1-(2-methoxyethyl)-1,3-dihydro-2H-1,4-benzodiazepin-2-one_b
7-Chloro-5-(2-fluorophenyl)-1-(2-methoxyethyl)-1,3-dihydro-2H-1,4-benzodiazepin-2-one_b
Zinecard_a
Zinecard_a
Zinecard_a
Zinecard_a
Zinecard_a
n-Ribosylhistidine
n-Ribosylhistidine
n-Ribosylhistidine
n-Ribosylhistidine
n-Ribosylhistidine
valganciclovir
valganciclovir
valganciclovir
valganciclovir
valganciclovir
N-[(2S)-2-Hydroxypropanoyl]methionine_b
N-[(2S)-2-Hydroxypropanoyl]methionine_b
N-[(2S)-2-Hydroxypropanoyl]methionine_b
N-[(2S)-2-Hydroxypropanoyl]methionine_b
N-[(2S)-2-Hydroxypropanoyl]methionine_b
(3S,5R,6E)-7-[3-(4-Fluorophenyl)-5-hydroxy-1-isopropyl-1H-indol-2-yl]-3,5-dihydroxy-6-heptenoic acid
(3S,5R,6E)-7-[3-(4-Fluorophenyl)-5-hydroxy-1-isopropyl-1H-indol-2-yl]-3,5-dihydroxy-6-heptenoic acid
(3S,5R,6E)-7-[3-(4-Fluorophenyl)-5-hydroxy-1-isopropyl-1H-indol-2-yl]-3,5-dihydroxy-6-heptenoic acid
(3S,5R,6E)-7-[3-(4-Fluorophenyl)-5-hydroxy-1-isopropyl-1H-indol-2-yl]-3,5-dihydroxy-6-heptenoic acid
(3S,5R,6E)-7-[3-(4-Fluorophenyl)-5-hydroxy-1-isopropyl-1H-indol-2-yl]-3,5-dihydroxy-6-heptenoic acid
Aspartyl-L-proline_c
Aspartyl-L-proline_c
Aspartyl-L-proline_c
Aspartyl-L-proline_c
Aspartyl-L-proline_c
MFCD18695608_a
MFCD18695608_a
MFCD18695608_a
MFCD18695608_a
MFCD18695608_a

11beta,13-dihydro-8-deoxylactucin
11beta,13-dihydro-8-deoxylactucin
11beta,13-dihydro-8-deoxylactucin
11beta,13-dihydro-8-deoxylactucin
11beta,13-dihydro-8-deoxylactucin
(1R,2S)-1-(7,8-Dihydro-6-pteridinyl)-1,2-propanediol
(1R,2S)-1-(7,8-Dihydro-6-pteridinyl)-1,2-propanediol
(1R,2S)-1-(7,8-Dihydro-6-pteridinyl)-1,2-propanediol
(1R,2S)-1-(7,8-Dihydro-6-pteridinyl)-1,2-propanediol
(1R,2S)-1-(7,8-Dihydro-6-pteridinyl)-1,2-propanediol
3-[3-Methoxy-4-(sulfooxy)phenyl]propanoic acid
3-[3-Methoxy-4-(sulfooxy)phenyl]propanoic acid
3-[3-Methoxy-4-(sulfooxy)phenyl]propanoic acid
3-[3-Methoxy-4-(sulfooxy)phenyl]propanoic acid
3-[3-Methoxy-4-(sulfooxy)phenyl]propanoic acid
5-Allyl-5-sec-butyl-1,3-dimethyl-2,4,6(1H,3H,5H)-pyrimidinetrione_c
5-Allyl-5-sec-butyl-1,3-dimethyl-2,4,6(1H,3H,5H)-pyrimidinetrione_c
5-Allyl-5-sec-butyl-1,3-dimethyl-2,4,6(1H,3H,5H)-pyrimidinetrione_c
5-Allyl-5-sec-butyl-1,3-dimethyl-2,4,6(1H,3H,5H)-pyrimidinetrione_c
5-Allyl-5-sec-butyl-1,3-dimethyl-2,4,6(1H,3H,5H)-pyrimidinetrione_c
Midodrine_a
Midodrine_a
Midodrine_a
Midodrine_a
Midodrine_a
Fenoterol
Fenoterol
Fenoterol
Fenoterol
Fenoterol
3-Mercaptopropionic acid
3-Mercaptopropionic acid
3-Mercaptopropionic acid
3-Mercaptopropionic acid
3-Mercaptopropionic acid
4-O-beta-D-xylo-Hexopyranosyl-3-ulose-D-glucopyranose
4-O-beta-D-xylo-Hexopyranosyl-3-ulose-D-glucopyranose
4-O-beta-D-xylo-Hexopyranosyl-3-ulose-D-glucopyranose
4-O-beta-D-xylo-Hexopyranosyl-3-ulose-D-glucopyranose
4-O-beta-D-xylo-Hexopyranosyl-3-ulose-D-glucopyranose
LW8000000_a
LW8000000_a
LW8000000_a
LW8000000_a
LW8000000_a
N-(2,3,4-Trimethoxybenzoyl)glycine_a
N-(2,3,4-Trimethoxybenzoyl)glycine_a
N-(2,3,4-Trimethoxybenzoyl)glycine_a

N-(2,3,4-Trimethoxybenzoyl)glycine_a
N-(2,3,4-Trimethoxybenzoyl)glycine_a
1,2,3,4-Tetrahydro-ÃŽÂ²-carboline-3-carboxylic acid
1,2,3,4-Tetrahydro-ÃŽÂ²-carboline-3-carboxylic acid
1,2,3,4-Tetrahydro-ÃŽÂ²-carboline-3-carboxylic acid
1,2,3,4-Tetrahydro-ÃŽÂ²-carboline-3-carboxylic acid
1,2,3,4-Tetrahydro-ÃŽÂ²-carboline-3-carboxylic acid
(7R)-7-(5-carboxy-5-oxopentanamido)cephalosporanic acid
(7R)-7-(5-carboxy-5-oxopentanamido)cephalosporanic acid
(7R)-7-(5-carboxy-5-oxopentanamido)cephalosporanic acid
(7R)-7-(5-carboxy-5-oxopentanamido)cephalosporanic acid
(7R)-7-(5-carboxy-5-oxopentanamido)cephalosporanic acid
Linamarin
Linamarin
Linamarin
Linamarin
Linamarin
primidone_b
primidone_b
primidone_b
primidone_b
primidone_b
2-Phenylethyl ÃŽÂ²-D-glucopyranosiduronic acid
2-Phenylethyl ÃŽÂ²-D-glucopyranosiduronic acid
2-Phenylethyl ÃŽÂ²-D-glucopyranosiduronic acid
2-Phenylethyl ÃŽÂ²-D-glucopyranosiduronic acid
2-Phenylethyl ÃŽÂ²-D-glucopyranosiduronic acid
(4S)-4-[(2E)-2-Octenoyloxy]-4-(trimethylammonio)butanoate_b
(4S)-4-[(2E)-2-Octenoyloxy]-4-(trimethylammonio)butanoate_b
(4S)-4-[(2E)-2-Octenoyloxy]-4-(trimethylammonio)butanoate_b
(4S)-4-[(2E)-2-Octenoyloxy]-4-(trimethylammonio)butanoate_b
(4S)-4-[(2E)-2-Octenoyloxy]-4-(trimethylammonio)butanoate_b
1-(4-Amino-4-carboxybutanoyl)-2-piperidinecarboxylic acid_b
1-(4-Amino-4-carboxybutanoyl)-2-piperidinecarboxylic acid_b
1-(4-Amino-4-carboxybutanoyl)-2-piperidinecarboxylic acid_b
1-(4-Amino-4-carboxybutanoyl)-2-piperidinecarboxylic acid_b
1-(4-Amino-4-carboxybutanoyl)-2-piperidinecarboxylic acid_b
epsilon-(gamma-Glutamyl)-lysine_a
epsilon-(gamma-Glutamyl)-lysine_a
epsilon-(gamma-Glutamyl)-lysine_a
epsilon-(gamma-Glutamyl)-lysine_a
epsilon-(gamma-Glutamyl)-lysine_a
pentobarbital_c
pentobarbital_c
pentobarbital_c
pentobarbital_c
pentobarbital_c
DIBEHENIN

DIBEHENIN
DIBEHENIN
DIBEHENIN
DIBEHENIN
hexobarbital_b
hexobarbital_b
hexobarbital_b
hexobarbital_b
hexobarbital_b
3-(14-Ethyl-4,8,13,18-tetramethyl-20-oxo-9-vinyl-3-phorbinyl)propanoic acid
3-(14-Ethyl-4,8,13,18-tetramethyl-20-oxo-9-vinyl-3-phorbinyl)propanoic acid
3-(14-Ethyl-4,8,13,18-tetramethyl-20-oxo-9-vinyl-3-phorbinyl)propanoic acid
3-(14-Ethyl-4,8,13,18-tetramethyl-20-oxo-9-vinyl-3-phorbinyl)propanoic acid
3-(14-Ethyl-4,8,13,18-tetramethyl-20-oxo-9-vinyl-3-phorbinyl)propanoic acid
Ethyl maltol
Ethyl maltol
Ethyl maltol
Ethyl maltol
Ethyl maltol
coronatine
coronatine
coronatine
coronatine
coronatine
g-Aminobutyryl-lysine_a
g-Aminobutyryl-lysine_a
g-Aminobutyryl-lysine_a
g-Aminobutyryl-lysine_a
g-Aminobutyryl-lysine_a
Dihydrouracil_b
Dihydrouracil_b
Dihydrouracil_b
Dihydrouracil_b
Dihydrouracil_b
Methyl alpha-aspartylphenylalaninate_a
Methyl alpha-aspartylphenylalaninate_a
Methyl alpha-aspartylphenylalaninate_a
Methyl alpha-aspartylphenylalaninate_a
Methyl alpha-aspartylphenylalaninate_a
17,21-Dihydroxypregnenolone
17,21-Dihydroxypregnenolone
17,21-Dihydroxypregnenolone
17,21-Dihydroxypregnenolone
17,21-Dihydroxypregnenolone
(-)-Physostigmine_b
(-)-Physostigmine_b
(-)-Physostigmine_b
(-)-Physostigmine_b

(-)-Physostigmine_b
(betaS)-beta,3-Dihydroxy-D-tyrosine
(betaS)-beta,3-Dihydroxy-D-tyrosine
(betaS)-beta,3-Dihydroxy-D-tyrosine
(betaS)-beta,3-Dihydroxy-D-tyrosine
(betaS)-beta,3-Dihydroxy-D-tyrosine
N-(2,3,4-Trimethoxybenzoyl)glycine_c
N-(2,3,4-Trimethoxybenzoyl)glycine_c
N-(2,3,4-Trimethoxybenzoyl)glycine_c
N-(2,3,4-Trimethoxybenzoyl)glycine_c
N-(2,3,4-Trimethoxybenzoyl)glycine_c
Prephenic acid
Prephenic acid
Prephenic acid
Prephenic acid
Prephenic acid
DL-Carbocysteine
DL-Carbocysteine
DL-Carbocysteine
DL-Carbocysteine
DL-Carbocysteine
2-(3,4-Dimethoxyphenyl)-5,7-dihydroxy-6,8-dimethoxy-2,3-dihydro-4H-chromen-4-one
2-(3,4-Dimethoxyphenyl)-5,7-dihydroxy-6,8-dimethoxy-2,3-dihydro-4H-chromen-4-one
2-(3,4-Dimethoxyphenyl)-5,7-dihydroxy-6,8-dimethoxy-2,3-dihydro-4H-chromen-4-one
2-(3,4-Dimethoxyphenyl)-5,7-dihydroxy-6,8-dimethoxy-2,3-dihydro-4H-chromen-4-one
2-(3,4-Dimethoxyphenyl)-5,7-dihydroxy-6,8-dimethoxy-2,3-dihydro-4H-chromen-4-one
asn-val_b
asn-val_b
asn-val_b
asn-val_b
asn-val_b
Lys-Pro_c
Lys-Pro_c
Lys-Pro_c
Lys-Pro_c
Lys-Pro_c
DL-Mevalonic acid
DL-Mevalonic acid
DL-Mevalonic acid
DL-Mevalonic acid
DL-Mevalonic acid
3-[(3-Hydroxytridecanoyl)oxy]-4-(trimethylammonio)butanoate
3-[(3-Hydroxytridecanoyl)oxy]-4-(trimethylammonio)butanoate
3-[(3-Hydroxytridecanoyl)oxy]-4-(trimethylammonio)butanoate
3-[(3-Hydroxytridecanoyl)oxy]-4-(trimethylammonio)butanoate
3-[(3-Hydroxytridecanoyl)oxy]-4-(trimethylammonio)butanoate
bis(4-isothiocyanatobutyl) disulfide
bis(4-isothiocyanatobutyl) disulfide

bis(4-isothiocyanatobutyl) disulfide
bis(4-isothiocyanatobutyl) disulfide
bis(4-isothiocyanatobutyl) disulfide
3-Morpholino-4-tetrahydro-1H-pyrrol-1-ylcyclobut-3-ene-1,2-dione
3-Morpholino-4-tetrahydro-1H-pyrrol-1-ylcyclobut-3-ene-1,2-dione
3-Morpholino-4-tetrahydro-1H-pyrrol-1-ylcyclobut-3-ene-1,2-dione
3-Morpholino-4-tetrahydro-1H-pyrrol-1-ylcyclobut-3-ene-1,2-dione
3-Morpholino-4-tetrahydro-1H-pyrrol-1-ylcyclobut-3-ene-1,2-dione
(2R)-1-{[(2-Aminoethoxy)(hydroxy)phosphoryl]oxy}-3-hydroxy-2-propanyl formate
(2R)-1-{[(2-Aminoethoxy)(hydroxy)phosphoryl]oxy}-3-hydroxy-2-propanyl formate
(2R)-1-{[(2-Aminoethoxy)(hydroxy)phosphoryl]oxy}-3-hydroxy-2-propanyl formate
(2R)-1-{[(2-Aminoethoxy)(hydroxy)phosphoryl]oxy}-3-hydroxy-2-propanyl formate
(2R)-1-{[(2-Aminoethoxy)(hydroxy)phosphoryl]oxy}-3-hydroxy-2-propanyl formate
(âˆ’)-nabilone_a
(âˆ’)-nabilone_a
(âˆ’)-nabilone_a
(âˆ’)-nabilone_a
(âˆ’)-nabilone_a
FB9500000_a
FB9500000_a
FB9500000_a
FB9500000_a
FB9500000_a
(2,4-Dihydroxyphenyl)(oxo)acetic acid
(2,4-Dihydroxyphenyl)(oxo)acetic acid
(2,4-Dihydroxyphenyl)(oxo)acetic acid
(2,4-Dihydroxyphenyl)(oxo)acetic acid
(2,4-Dihydroxyphenyl)(oxo)acetic acid
L-gamma-Glutamyl-L-valine_i
L-gamma-Glutamyl-L-valine_i
L-gamma-Glutamyl-L-valine_i
L-gamma-Glutamyl-L-valine_i
L-gamma-Glutamyl-L-valine_i
5-Hydantoinpropionic acid
5-Hydantoinpropionic acid
5-Hydantoinpropionic acid
5-Hydantoinpropionic acid
5-Hydantoinpropionic acid
threonylphenylalanine_c
threonylphenylalanine_c
threonylphenylalanine_c
threonylphenylalanine_c
threonylphenylalanine_c
Cadralazine
Cadralazine
Cadralazine
Cadralazine
Cadralazine

metixene
metixene
metixene
metixene
metixene
IN00260_a
IN00260_a
IN00260_a
IN00260_a
IN00260_a
Seryltyrosine_a
Seryltyrosine_a
Seryltyrosine_a
Seryltyrosine_a
Seryltyrosine_a
Oleuropein aglycon
Oleuropein aglycon
Oleuropein aglycon
Oleuropein aglycon
Oleuropein aglycon
MFCD18695608_d
MFCD18695608_d
MFCD18695608_d
MFCD18695608_d
MFCD18695608_d
Spermic acid_a
Spermic acid_a
Spermic acid_a
Spermic acid_a
Spermic acid_a
Methyl 4-(4,6-dimethyl-9-oxo-4,9-dihydro-1H-imidazo[1,2-a]purin-7-yl)-3-hydroperoxy-2-[(methoxycarbonyl)amir
Methyl 4-(4,6-dimethyl-9-oxo-4,9-dihydro-1H-imidazo[1,2-a]purin-7-yl)-3-hydroperoxy-2-[(methoxycarbonyl)amir
Methyl 4-(4,6-dimethyl-9-oxo-4,9-dihydro-1H-imidazo[1,2-a]purin-7-yl)-3-hydroperoxy-2-[(methoxycarbonyl)amir
Methyl 4-(4,6-dimethyl-9-oxo-4,9-dihydro-1H-imidazo[1,2-a]purin-7-yl)-3-hydroperoxy-2-[(methoxycarbonyl)amir
Methyl 4-(4,6-dimethyl-9-oxo-4,9-dihydro-1H-imidazo[1,2-a]purin-7-yl)-3-hydroperoxy-2-[(methoxycarbonyl)amir
1-{3-Carboxy-3-[(3-carboxy-3-hydroxypropyl)amino]propyl}-2-azetidinecarboxylic acid
1-{3-Carboxy-3-[(3-carboxy-3-hydroxypropyl)amino]propyl}-2-azetidinecarboxylic acid
1-{3-Carboxy-3-[(3-carboxy-3-hydroxypropyl)amino]propyl}-2-azetidinecarboxylic acid
1-{3-Carboxy-3-[(3-carboxy-3-hydroxypropyl)amino]propyl}-2-azetidinecarboxylic acid
1-{3-Carboxy-3-[(3-carboxy-3-hydroxypropyl)amino]propyl}-2-azetidinecarboxylic acid
4-Thiapentanoic acid
4-Thiapentanoic acid
4-Thiapentanoic acid
4-Thiapentanoic acid
4-Thiapentanoic acid
Glycerophosphoglycerol
Glycerophosphoglycerol
Glycerophosphoglycerol

Glycerophosphoglycerol
Glycerophosphoglycerol
L-gamma-Glutamyl-L-leucine_g
L-gamma-Glutamyl-L-leucine_g
L-gamma-Glutamyl-L-leucine_g
L-gamma-Glutamyl-L-leucine_g
L-gamma-Glutamyl-L-leucine_g
Leucyltryptophan
Leucyltryptophan
Leucyltryptophan
Leucyltryptophan
Leucyltryptophan
butalbital_a
butalbital_a
butalbital_a
butalbital_a
butalbital_a
INK (Peptide Ile-Asn-Lys)
INK (Peptide Ile-Asn-Lys)
INK (Peptide Ile-Asn-Lys)
INK (Peptide Ile-Asn-Lys)
INK (Peptide Ile-Asn-Lys)
3-(Sulfooxy)-L-tyrosine_a
3-(Sulfooxy)-L-tyrosine_a
3-(Sulfooxy)-L-tyrosine_a
3-(Sulfooxy)-L-tyrosine_a
3-(Sulfooxy)-L-tyrosine_a
uridine 5â€²-diphosphate
uridine 5â€²-diphosphate
uridine 5â€²-diphosphate
uridine 5â€²-diphosphate
uridine 5â€²-diphosphate
2-Furoylglycine
2-Furoylglycine
2-Furoylglycine
2-Furoylglycine
2-Furoylglycine
2-Ammonio-4-{4-[(2S)-2-ammonio-2-carboxylatoethyl]-1H-imidazol-2-yl}butanoate
2-Ammonio-4-{4-[(2S)-2-ammonio-2-carboxylatoethyl]-1H-imidazol-2-yl}butanoate
2-Ammonio-4-{4-[(2S)-2-ammonio-2-carboxylatoethyl]-1H-imidazol-2-yl}butanoate
2-Ammonio-4-{4-[(2S)-2-ammonio-2-carboxylatoethyl]-1H-imidazol-2-yl}butanoate
2-Ammonio-4-{4-[(2S)-2-ammonio-2-carboxylatoethyl]-1H-imidazol-2-yl}butanoate
carglumic acid
carglumic acid
carglumic acid
carglumic acid
carglumic acid
N-Acetylprocainamide_b

N-Acetylprocainamide_b
N-Acetylprocainamide_b
N-Acetylprocainamide_b
N-Acetylprocainamide_b
Prunasin
Prunasin
Prunasin
Prunasin
Prunasin
2-(2,4-Dihydroxyphenyl)-3-[(2Z)-3,7-dimethyl-2,6-octadien-1-yl]-5,7-dihydroxy-6-[(2E)-4-hydroxy-3-methyl-2-but
2-(2,4-Dihydroxyphenyl)-3-[(2Z)-3,7-dimethyl-2,6-octadien-1-yl]-5,7-dihydroxy-6-[(2E)-4-hydroxy-3-methyl-2-but
2-(2,4-Dihydroxyphenyl)-3-[(2Z)-3,7-dimethyl-2,6-octadien-1-yl]-5,7-dihydroxy-6-[(2E)-4-hydroxy-3-methyl-2-but
2-(2,4-Dihydroxyphenyl)-3-[(2Z)-3,7-dimethyl-2,6-octadien-1-yl]-5,7-dihydroxy-6-[(2E)-4-hydroxy-3-methyl-2-but
2-(2,4-Dihydroxyphenyl)-3-[(2Z)-3,7-dimethyl-2,6-octadien-1-yl]-5,7-dihydroxy-6-[(2E)-4-hydroxy-3-methyl-2-but
Ro 20-1724_b
Ro 20-1724_b
Ro 20-1724_b
Ro 20-1724_b
Ro 20-1724_b
Guanadrel
Guanadrel
Guanadrel
Guanadrel
Guanadrel
5-methylthioribose
5-methylthioribose
5-methylthioribose
5-methylthioribose
5-methylthioribose
N-(3,5-Dimethoxybenzoyl)glycine_d
N-(3,5-Dimethoxybenzoyl)glycine_d
N-(3,5-Dimethoxybenzoyl)glycine_d
N-(3,5-Dimethoxybenzoyl)glycine_d
N-(3,5-Dimethoxybenzoyl)glycine_d
Triethyl citrate_a
Triethyl citrate_a
Triethyl citrate_a
Triethyl citrate_a
Triethyl citrate_a
meprobamate_b
meprobamate_b
meprobamate_b
meprobamate_b
meprobamate_b
Ethyl malate_d
Ethyl malate_d
Ethyl malate_d
Ethyl malate_d

Ethyl malate_d
2-Hydroxy-3,4,5-trimethoxybenzoic acid
2-Hydroxy-3,4,5-trimethoxybenzoic acid
2-Hydroxy-3,4,5-trimethoxybenzoic acid
2-Hydroxy-3,4,5-trimethoxybenzoic acid
2-Hydroxy-3,4,5-trimethoxybenzoic acid
2-Acetamido-2-deoxy-3-O-(6-deoxy-alpha-L-galactopyranosyl)-D-glucose
2-Acetamido-2-deoxy-3-O-(6-deoxy-alpha-L-galactopyranosyl)-D-glucose
2-Acetamido-2-deoxy-3-O-(6-deoxy-alpha-L-galactopyranosyl)-D-glucose
2-Acetamido-2-deoxy-3-O-(6-deoxy-alpha-L-galactopyranosyl)-D-glucose
2-Acetamido-2-deoxy-3-O-(6-deoxy-alpha-L-galactopyranosyl)-D-glucose
Astemizole_a
Astemizole_a
Astemizole_a
Astemizole_a
Astemizole_a
Butabarbital_b
Butabarbital_b
Butabarbital_b
Butabarbital_b
Butabarbital_b
N-(Carboxymethyl)norleucine_c
N-(Carboxymethyl)norleucine_c
N-(Carboxymethyl)norleucine_c
N-(Carboxymethyl)norleucine_c
N-(Carboxymethyl)norleucine_c
Lys-phe_b
Lys-phe_b
Lys-phe_b
Lys-phe_b
Lys-phe_b
Erythorbic acid
Erythorbic acid
Erythorbic acid
Erythorbic acid
Erythorbic acid
2,3,4,9-Tetrahydro-1H-Î²-carboline-3-carboxylic acid
2,3,4,9-Tetrahydro-1H-Î²-carboline-3-carboxylic acid
2,3,4,9-Tetrahydro-1H-Î²-carboline-3-carboxylic acid
2,3,4,9-Tetrahydro-1H-Î²-carboline-3-carboxylic acid
2,3,4,9-Tetrahydro-1H-Î²-carboline-3-carboxylic acid
Methyl 2,3-dihydro-3-hydroxy-2-oxo-1H-indole-3-acetate_a
Methyl 2,3-dihydro-3-hydroxy-2-oxo-1H-indole-3-acetate_a
Methyl 2,3-dihydro-3-hydroxy-2-oxo-1H-indole-3-acetate_a
Methyl 2,3-dihydro-3-hydroxy-2-oxo-1H-indole-3-acetate_a
Methyl 2,3-dihydro-3-hydroxy-2-oxo-1H-indole-3-acetate_a
Tetramethylpyrazine
Tetramethylpyrazine

Tetramethylpyrazine
Tetramethylpyrazine
Tetramethylpyrazine
(2S)-3-Methyl-2-({[(3S,4S,5R)-2,3,4-trihydroxy-5-(hydroxymethyl)tetrahydro-2-furanyl]methyl}amino)butanoic aci
(2S)-3-Methyl-2-({[(3S,4S,5R)-2,3,4-trihydroxy-5-(hydroxymethyl)tetrahydro-2-furanyl]methyl}amino)butanoic aci
(2S)-3-Methyl-2-({[(3S,4S,5R)-2,3,4-trihydroxy-5-(hydroxymethyl)tetrahydro-2-furanyl]methyl}amino)butanoic aci
(2S)-3-Methyl-2-({[(3S,4S,5R)-2,3,4-trihydroxy-5-(hydroxymethyl)tetrahydro-2-furanyl]methyl}amino)butanoic aci
(2S)-3-Methyl-2-({[(3S,4S,5R)-2,3,4-trihydroxy-5-(hydroxymethyl)tetrahydro-2-furanyl]methyl}amino)butanoic aci
Roxane
Roxane
Roxane
Roxane
Roxane
lys-tyr_b
lys-tyr_b
lys-tyr_b
lys-tyr_b
lys-tyr_b
4-(METHYLNITROSAMINO)-1-(3-PYRIDYL-N-OXIDE)-1-BUTANOL_d
4-(METHYLNITROSAMINO)-1-(3-PYRIDYL-N-OXIDE)-1-BUTANOL_d
4-(METHYLNITROSAMINO)-1-(3-PYRIDYL-N-OXIDE)-1-BUTANOL_d
4-(METHYLNITROSAMINO)-1-(3-PYRIDYL-N-OXIDE)-1-BUTANOL_d
4-(METHYLNITROSAMINO)-1-(3-PYRIDYL-N-OXIDE)-1-BUTANOL_d
3-(2,3-Dihydroxy-3-methylbutyl)-4-methoxy-1-methyl-2(1H)-quinolinone_b
3-(2,3-Dihydroxy-3-methylbutyl)-4-methoxy-1-methyl-2(1H)-quinolinone_b
3-(2,3-Dihydroxy-3-methylbutyl)-4-methoxy-1-methyl-2(1H)-quinolinone_b
3-(2,3-Dihydroxy-3-methylbutyl)-4-methoxy-1-methyl-2(1H)-quinolinone_b
3-(2,3-Dihydroxy-3-methylbutyl)-4-methoxy-1-methyl-2(1H)-quinolinone_b
N-(Carboxymethyl)norleucine_b
N-(Carboxymethyl)norleucine_b
N-(Carboxymethyl)norleucine_b
N-(Carboxymethyl)norleucine_b
N-(Carboxymethyl)norleucine_b
asn-val_c
asn-val_c
asn-val_c
asn-val_c
asn-val_c
Tocainide
Tocainide
Tocainide
Tocainide
Tocainide
Histidylglycine_b
Histidylglycine_b
Histidylglycine_b
Histidylglycine_b
Histidylglycine_b

7-Chloro-5-(2-fluorophenyl)-1-(2-methoxyethyl)-1,3-dihydro-2H-1,4-benzodiazepin-2-one_a
7-Chloro-5-(2-fluorophenyl)-1-(2-methoxyethyl)-1,3-dihydro-2H-1,4-benzodiazepin-2-one_a
7-Chloro-5-(2-fluorophenyl)-1-(2-methoxyethyl)-1,3-dihydro-2H-1,4-benzodiazepin-2-one_a
7-Chloro-5-(2-fluorophenyl)-1-(2-methoxyethyl)-1,3-dihydro-2H-1,4-benzodiazepin-2-one_a
7-Chloro-5-(2-fluorophenyl)-1-(2-methoxyethyl)-1,3-dihydro-2H-1,4-benzodiazepin-2-one_a
3-Hydroxy-3-methyl-1-[(7-oxo-7H-furo[3,2-g]chromen-9-yl)oxy]-2-butanyl 3-methyl-2-butenoate
3-Hydroxy-3-methyl-1-[(7-oxo-7H-furo[3,2-g]chromen-9-yl)oxy]-2-butanyl 3-methyl-2-butenoate
3-Hydroxy-3-methyl-1-[(7-oxo-7H-furo[3,2-g]chromen-9-yl)oxy]-2-butanyl 3-methyl-2-butenoate
3-Hydroxy-3-methyl-1-[(7-oxo-7H-furo[3,2-g]chromen-9-yl)oxy]-2-butanyl 3-methyl-2-butenoate
3-Hydroxy-3-methyl-1-[(7-oxo-7H-furo[3,2-g]chromen-9-yl)oxy]-2-butanyl 3-methyl-2-butenoate
11-(4-Hydroxyphenyl)-3-methyl-6,7,8,9-tetrahydropyridazino[1,2-a]indazol-10-ium-1-olate
11-(4-Hydroxyphenyl)-3-methyl-6,7,8,9-tetrahydropyridazino[1,2-a]indazol-10-ium-1-olate
11-(4-Hydroxyphenyl)-3-methyl-6,7,8,9-tetrahydropyridazino[1,2-a]indazol-10-ium-1-olate
11-(4-Hydroxyphenyl)-3-methyl-6,7,8,9-tetrahydropyridazino[1,2-a]indazol-10-ium-1-olate
11-(4-Hydroxyphenyl)-3-methyl-6,7,8,9-tetrahydropyridazino[1,2-a]indazol-10-ium-1-olate
Arctiopicrin
Arctiopicrin
Arctiopicrin
Arctiopicrin
Arctiopicrin
Ile-cys
Ile-cys
Ile-cys
Ile-cys
Ile-cys
Val-Ser_b
Val-Ser_b
Val-Ser_b
Val-Ser_b
Val-Ser_b
beta-D-Ethyl glucuronide_a
beta-D-Ethyl glucuronide_a
beta-D-Ethyl glucuronide_a
beta-D-Ethyl glucuronide_a
beta-D-Ethyl glucuronide_a
N-acetyl-9-O-acetylneuraminic acid
N-acetyl-9-O-acetylneuraminic acid
N-acetyl-9-O-acetylneuraminic acid
N-acetyl-9-O-acetylneuraminic acid
N-acetyl-9-O-acetylneuraminic acid
(-)-Aspidospermine
(-)-Aspidospermine
(-)-Aspidospermine
(-)-Aspidospermine
(-)-Aspidospermine
6-Hydroxymelatonin
6-Hydroxymelatonin
6-Hydroxymelatonin

6-Hydroxymelatonin
6-Hydroxymelatonin
Succinic anhydride
Succinic anhydride
Succinic anhydride
Succinic anhydride
Succinic anhydride
Homovanillic acid sulfate
Homovanillic acid sulfate
Homovanillic acid sulfate
Homovanillic acid sulfate
Homovanillic acid sulfate
2-Acetamidohexanedioic acid_e
2-Acetamidohexanedioic acid_e
2-Acetamidohexanedioic acid_e
2-Acetamidohexanedioic acid_e
2-Acetamidohexanedioic acid_e
SECONAL_b
SECONAL_b
SECONAL_b
SECONAL_b
SECONAL_b
3'-Hydroxycotinine, Cis-
3'-Hydroxycotinine, Cis-
3'-Hydroxycotinine, Cis-
3'-Hydroxycotinine, Cis-
3'-Hydroxycotinine, Cis-
Asarone
Asarone
Asarone
Asarone
Asarone
YWA1
YWA1
YWA1
YWA1
YWA1
Piperonylonitrile
Piperonylonitrile
Piperonylonitrile
Piperonylonitrile
Piperonylonitrile
alliin
alliin
alliin
alliin
alliin
D-2-Amino-hexano-6-lactam

D-2-Amino-hexano-6-lactam
D-2-Amino-hexano-6-lactam
D-2-Amino-hexano-6-lactam
D-2-Amino-hexano-6-lactam
SECONAL_c
SECONAL_c
SECONAL_c
SECONAL_c
SECONAL_c
(E)-4-Methoxycinnamic acid
(E)-4-Methoxycinnamic acid
(E)-4-Methoxycinnamic acid
(E)-4-Methoxycinnamic acid
(E)-4-Methoxycinnamic acid
Homoanserine_c
Homoanserine_c
Homoanserine_c
Homoanserine_c
Homoanserine_c
Ala-Tyr
Ala-Tyr
Ala-Tyr
Ala-Tyr
Ala-Tyr
6-Myoporol
6-Myoporol
6-Myoporol
6-Myoporol
6-Myoporol
Hostmaniane
Hostmaniane
Hostmaniane
Hostmaniane
Hostmaniane
hexobarbital_c
hexobarbital_c
hexobarbital_c
hexobarbital_c
hexobarbital_c
ophthalmic acid_a
ophthalmic acid_a
ophthalmic acid_a
ophthalmic acid_a
ophthalmic acid_a
5-Hydroxy-6-[(2E)-4-hydroxy-3-methyl-2-buten-1-yl]-2-methyl-4-oxo-4H-chromen-7-yl hexopyranoside
5-Hydroxy-6-[(2E)-4-hydroxy-3-methyl-2-buten-1-yl]-2-methyl-4-oxo-4H-chromen-7-yl hexopyranoside
5-Hydroxy-6-[(2E)-4-hydroxy-3-methyl-2-buten-1-yl]-2-methyl-4-oxo-4H-chromen-7-yl hexopyranoside
5-Hydroxy-6-[(2E)-4-hydroxy-3-methyl-2-buten-1-yl]-2-methyl-4-oxo-4H-chromen-7-yl hexopyranoside

5-Hydroxy-6-[(2E)-4-hydroxy-3-methyl-2-buten-1-yl]-2-methyl-4-oxo-4H-chromen-7-yl hexopyranoside
tert-Butyl 3-amino-1-methyl-2,3-dioxopropylcarbamate_c
tert-Butyl 3-amino-1-methyl-2,3-dioxopropylcarbamate_c
tert-Butyl 3-amino-1-methyl-2,3-dioxopropylcarbamate_c
tert-Butyl 3-amino-1-methyl-2,3-dioxopropylcarbamate_c
tert-Butyl 3-amino-1-methyl-2,3-dioxopropylcarbamate_c
tert-Butyl 3-amino-1-methyl-2,3-dioxopropylcarbamate_f
tert-Butyl 3-amino-1-methyl-2,3-dioxopropylcarbamate_f
tert-Butyl 3-amino-1-methyl-2,3-dioxopropylcarbamate_f
tert-Butyl 3-amino-1-methyl-2,3-dioxopropylcarbamate_f
tert-Butyl 3-amino-1-methyl-2,3-dioxopropylcarbamate_f
9-Methyluric acid_a
9-Methyluric acid_a
9-Methyluric acid_a
9-Methyluric acid_a
9-Methyluric acid_a
Kynurenic acid
Kynurenic acid
Kynurenic acid
Kynurenic acid
Kynurenic acid
2-Aminooctanedioic acid_a
2-Aminooctanedioic acid_a
2-Aminooctanedioic acid_a
2-Aminooctanedioic acid_a
2-Aminooctanedioic acid_a
1H-Pyrazol-4-ylmethanol
1H-Pyrazol-4-ylmethanol
1H-Pyrazol-4-ylmethanol
1H-Pyrazol-4-ylmethanol
1H-Pyrazol-4-ylmethanol
N-Propionylmethionine_b
N-Propionylmethionine_b
N-Propionylmethionine_b
N-Propionylmethionine_b
N-Propionylmethionine_b
Methyl 1-hydroxy-7-(hydroxymethyl)-1,4a,5,7a-tetrahydrocyclopenta[c]pyran-4-carboxylate_a
Methyl 1-hydroxy-7-(hydroxymethyl)-1,4a,5,7a-tetrahydrocyclopenta[c]pyran-4-carboxylate_a
Methyl 1-hydroxy-7-(hydroxymethyl)-1,4a,5,7a-tetrahydrocyclopenta[c]pyran-4-carboxylate_a
Methyl 1-hydroxy-7-(hydroxymethyl)-1,4a,5,7a-tetrahydrocyclopenta[c]pyran-4-carboxylate_a
Methyl 1-hydroxy-7-(hydroxymethyl)-1,4a,5,7a-tetrahydrocyclopenta[c]pyran-4-carboxylate_a
N-Benzoylaspartic acid_a
N-Benzoylaspartic acid_a
N-Benzoylaspartic acid_a
N-Benzoylaspartic acid_a
N-Benzoylaspartic acid_a
Indole-3-carbidol
Indole-3-carbidol

Indole-3-carbidol
Indole-3-carbidol
Indole-3-carbidol
Propafenone
Propafenone
Propafenone
Propafenone
Propafenone
Glu-Glu
Glu-Glu
Glu-Glu
Glu-Glu
Glu-Glu
Raltitrexed
Raltitrexed
Raltitrexed
Raltitrexed
Raltitrexed
2-(3,5-dimethyl-1H-pyrazol-4-yl)-5-methoxybenzoic acid
2-(3,5-dimethyl-1H-pyrazol-4-yl)-5-methoxybenzoic acid
2-(3,5-dimethyl-1H-pyrazol-4-yl)-5-methoxybenzoic acid
2-(3,5-dimethyl-1H-pyrazol-4-yl)-5-methoxybenzoic acid
2-(3,5-dimethyl-1H-pyrazol-4-yl)-5-methoxybenzoic acid
3-(Sulfooxy)-L-tyrosine_b
3-(Sulfooxy)-L-tyrosine_b
3-(Sulfooxy)-L-tyrosine_b
3-(Sulfooxy)-L-tyrosine_b
3-(Sulfooxy)-L-tyrosine_b
Validamycin A
Validamycin A
Validamycin A
Validamycin A
Validamycin A
N,N-Diethyl-4-methyl-1-piperazinecarboxamide 4-oxide_a
N,N-Diethyl-4-methyl-1-piperazinecarboxamide 4-oxide_a
N,N-Diethyl-4-methyl-1-piperazinecarboxamide 4-oxide_a
N,N-Diethyl-4-methyl-1-piperazinecarboxamide 4-oxide_a
N,N-Diethyl-4-methyl-1-piperazinecarboxamide 4-oxide_a
n-Propyl Gallate
n-Propyl Gallate
n-Propyl Gallate
n-Propyl Gallate
n-Propyl Gallate
N-(3,5-Dimethoxybenzoyl)glycine_c
N-(3,5-Dimethoxybenzoyl)glycine_c
N-(3,5-Dimethoxybenzoyl)glycine_c
N-(3,5-Dimethoxybenzoyl)glycine_c
N-(3,5-Dimethoxybenzoyl)glycine_c

Butylphthalide
Butylphthalide
Butylphthalide
Butylphthalide
Butylphthalide
DLK (Peptide Asp-Leu-Lys)
DLK (Peptide Asp-Leu-Lys)
DLK (Peptide Asp-Leu-Lys)
DLK (Peptide Asp-Leu-Lys)
DLK (Peptide Asp-Leu-Lys)
7-Methyladenine
7-Methyladenine
7-Methyladenine
7-Methyladenine
7-Methyladenine
3-Ureidopropionic acid
3-Ureidopropionic acid
3-Ureidopropionic acid
3-Ureidopropionic acid
3-Ureidopropionic acid
6-(1-Hydroxyethyl)-3-(hydroxymethyl)-2,7-dioxabicyclo[4.1.0]hept-3-en-5-one_b
6-(1-Hydroxyethyl)-3-(hydroxymethyl)-2,7-dioxabicyclo[4.1.0]hept-3-en-5-one_b
6-(1-Hydroxyethyl)-3-(hydroxymethyl)-2,7-dioxabicyclo[4.1.0]hept-3-en-5-one_b
6-(1-Hydroxyethyl)-3-(hydroxymethyl)-2,7-dioxabicyclo[4.1.0]hept-3-en-5-one_b
6-(1-Hydroxyethyl)-3-(hydroxymethyl)-2,7-dioxabicyclo[4.1.0]hept-3-en-5-one_b
tenivastatin
tenivastatin
tenivastatin
tenivastatin
tenivastatin
Leu-pro_a
Leu-pro_a
Leu-pro_a
Leu-pro_a
Leu-pro_a
Dopamine 3-O-sulfate
Dopamine 3-O-sulfate
Dopamine 3-O-sulfate
Dopamine 3-O-sulfate
Dopamine 3-O-sulfate
Ethylvanillin propylene glycol acetal
Ethylvanillin propylene glycol acetal
Ethylvanillin propylene glycol acetal
Ethylvanillin propylene glycol acetal
Ethylvanillin propylene glycol acetal
Dihydrothymine
Dihydrothymine
Dihydrothymine

Dihydrothymine
Dihydrothymine
(7E,7'E)-5,5'-diferulic acid_c
(7E,7'E)-5,5'-diferulic acid_c
(7E,7'E)-5,5'-diferulic acid_c
(7E,7'E)-5,5'-diferulic acid_c
(7E,7'E)-5,5'-diferulic acid_c
4-Amino-1-(4-{[(3R)-3-amino-5-(N-methylcarbamimidamido)pentanoyl]amino}-2,3,4-trideoxy-beta-D-erythro-he›
4-Amino-1-(4-{[(3R)-3-amino-5-(N-methylcarbamimidamido)pentanoyl]amino}-2,3,4-trideoxy-beta-D-erythro-he›
4-Amino-1-(4-{[(3R)-3-amino-5-(N-methylcarbamimidamido)pentanoyl]amino}-2,3,4-trideoxy-beta-D-erythro-he›
4-Amino-1-(4-{[(3R)-3-amino-5-(N-methylcarbamimidamido)pentanoyl]amino}-2,3,4-trideoxy-beta-D-erythro-he›
4-Amino-1-(4-{[(3R)-3-amino-5-(N-methylcarbamimidamido)pentanoyl]amino}-2,3,4-trideoxy-beta-D-erythro-he›
6-APA_a
6-APA_a
6-APA_a
6-APA_a
6-APA_a
thyronine
thyronine
thyronine
thyronine
thyronine
L-gamma-Glutamyl-L-leucine_h
L-gamma-Glutamyl-L-leucine_h
L-gamma-Glutamyl-L-leucine_h
L-gamma-Glutamyl-L-leucine_h
L-gamma-Glutamyl-L-leucine_h
Leucyltyrosine_d
Leucyltyrosine_d
Leucyltyrosine_d
Leucyltyrosine_d
Leucyltyrosine_d
4-(3-Oxopentyl)phenyl hydrogen sulfate
4-(3-Oxopentyl)phenyl hydrogen sulfate
4-(3-Oxopentyl)phenyl hydrogen sulfate
4-(3-Oxopentyl)phenyl hydrogen sulfate
4-(3-Oxopentyl)phenyl hydrogen sulfate
Val-Trp_a
Val-Trp_a
Val-Trp_a
Val-Trp_a
Val-Trp_a
S-Propylcysteine
S-Propylcysteine
S-Propylcysteine
S-Propylcysteine
S-Propylcysteine
piscidic acid_b

piscidic acid_b
piscidic acid_b
piscidic acid_b
piscidic acid_b
R-(+)-Etiracetam_a
R-(+)-Etiracetam_a
R-(+)-Etiracetam_a
R-(+)-Etiracetam_a
R-(+)-Etiracetam_a
ala-ser_b
ala-ser_b
ala-ser_b
ala-ser_b
ala-ser_b
N-Phenylacetylglutamic acid_a
N-Phenylacetylglutamic acid_a
N-Phenylacetylglutamic acid_a
N-Phenylacetylglutamic acid_a
N-Phenylacetylglutamic acid_a
Calcitriol
Calcitriol
Calcitriol
Calcitriol
Calcitriol
N,N-Diethyl-4-methyl-1-piperazinecarboxamide 4-oxide_b
N,N-Diethyl-4-methyl-1-piperazinecarboxamide 4-oxide_b
N,N-Diethyl-4-methyl-1-piperazinecarboxamide 4-oxide_b
N,N-Diethyl-4-methyl-1-piperazinecarboxamide 4-oxide_b
N,N-Diethyl-4-methyl-1-piperazinecarboxamide 4-oxide_b
6-hydroxypseudooxynicotine_a
6-hydroxypseudooxynicotine_a
6-hydroxypseudooxynicotine_a
6-hydroxypseudooxynicotine_a
6-hydroxypseudooxynicotine_a
L-Pyrrolysine
L-Pyrrolysine
L-Pyrrolysine
L-Pyrrolysine
L-Pyrrolysine
6-APA_b
6-APA_b
6-APA_b
6-APA_b
6-APA_b
Tetraacetylethylenediamine_g
Tetraacetylethylenediamine_g
Tetraacetylethylenediamine_g
Tetraacetylethylenediamine_g

Tetraacetylethylenediamine_g
9-ribosylzeatin
9-ribosylzeatin
9-ribosylzeatin
9-ribosylzeatin
9-ribosylzeatin
3-Methoxy-4-hydroxyhippuric acid_e
3-Methoxy-4-hydroxyhippuric acid_e
3-Methoxy-4-hydroxyhippuric acid_e
3-Methoxy-4-hydroxyhippuric acid_e
3-Methoxy-4-hydroxyhippuric acid_e
FC2505000
FC2505000
FC2505000
FC2505000
FC2505000
6,8-Dimethoxy-1,2,3a,12c-tetrahydro-7H-furo[3',2':4,5]furo[2,3-c]xanthen-7-one
6,8-Dimethoxy-1,2,3a,12c-tetrahydro-7H-furo[3',2':4,5]furo[2,3-c]xanthen-7-one
6,8-Dimethoxy-1,2,3a,12c-tetrahydro-7H-furo[3',2':4,5]furo[2,3-c]xanthen-7-one
6,8-Dimethoxy-1,2,3a,12c-tetrahydro-7H-furo[3',2':4,5]furo[2,3-c]xanthen-7-one
6,8-Dimethoxy-1,2,3a,12c-tetrahydro-7H-furo[3',2':4,5]furo[2,3-c]xanthen-7-one
Eslicarbazepine
Eslicarbazepine
Eslicarbazepine
Eslicarbazepine
Eslicarbazepine
(+/-)-2-Hydroxyglutaric acid_b
(+/-)-2-Hydroxyglutaric acid_b
(+/-)-2-Hydroxyglutaric acid_b
(+/-)-2-Hydroxyglutaric acid_b
(+/-)-2-Hydroxyglutaric acid_b
Kyotorphin
Kyotorphin
Kyotorphin
Kyotorphin
Kyotorphin
4-(1-Hydroxy-2-{[6-(4-hydroxy-4-phenylbutoxy)hexyl]amino}ethyl)-2-(hydroxymethyl)phenol
4-(1-Hydroxy-2-{[6-(4-hydroxy-4-phenylbutoxy)hexyl]amino}ethyl)-2-(hydroxymethyl)phenol
4-(1-Hydroxy-2-{[6-(4-hydroxy-4-phenylbutoxy)hexyl]amino}ethyl)-2-(hydroxymethyl)phenol
4-(1-Hydroxy-2-{[6-(4-hydroxy-4-phenylbutoxy)hexyl]amino}ethyl)-2-(hydroxymethyl)phenol
4-(1-Hydroxy-2-{[6-(4-hydroxy-4-phenylbutoxy)hexyl]amino}ethyl)-2-(hydroxymethyl)phenol
7alpha-Hydroxy-3-oxochol-4-en-24-oic acid_d
7alpha-Hydroxy-3-oxochol-4-en-24-oic acid_d
7alpha-Hydroxy-3-oxochol-4-en-24-oic acid_d
7alpha-Hydroxy-3-oxochol-4-en-24-oic acid_d
7alpha-Hydroxy-3-oxochol-4-en-24-oic acid_d
Marimastat
Marimastat

Marimastat
Marimastat
Marimastat
2-(3-Hydroxy-3,4,5,6-tetrahydro-1H-cyclopenta[c]furan-4-yl)-3-methoxy-3-oxopropanoic acid
2-(3-Hydroxy-3,4,5,6-tetrahydro-1H-cyclopenta[c]furan-4-yl)-3-methoxy-3-oxopropanoic acid
2-(3-Hydroxy-3,4,5,6-tetrahydro-1H-cyclopenta[c]furan-4-yl)-3-methoxy-3-oxopropanoic acid
2-(3-Hydroxy-3,4,5,6-tetrahydro-1H-cyclopenta[c]furan-4-yl)-3-methoxy-3-oxopropanoic acid
2-(3-Hydroxy-3,4,5,6-tetrahydro-1H-cyclopenta[c]furan-4-yl)-3-methoxy-3-oxopropanoic acid
folinic acid
folinic acid
folinic acid
folinic acid
folinic acid
N~6~-[5-(1,2-Dithiolan-3-yl)pentanoyl]-D-lysine
N~6~-[5-(1,2-Dithiolan-3-yl)pentanoyl]-D-lysine
N~6~-[5-(1,2-Dithiolan-3-yl)pentanoyl]-D-lysine
N~6~-[5-(1,2-Dithiolan-3-yl)pentanoyl]-D-lysine
N~6~-[5-(1,2-Dithiolan-3-yl)pentanoyl]-D-lysine
N-(2,3,4-Trimethoxybenzoyl)glycine_b
N-(2,3,4-Trimethoxybenzoyl)glycine_b
N-(2,3,4-Trimethoxybenzoyl)glycine_b
N-(2,3,4-Trimethoxybenzoyl)glycine_b
N-(2,3,4-Trimethoxybenzoyl)glycine_b
Toluene_a
Toluene_a
Toluene_a
Toluene_a
Toluene_a
MFCD00055782
MFCD00055782
MFCD00055782
MFCD00055782
MFCD00055782
L-gamma-Glutamyl-L-leucine_a
L-gamma-Glutamyl-L-leucine_a
L-gamma-Glutamyl-L-leucine_a
L-gamma-Glutamyl-L-leucine_a
L-gamma-Glutamyl-L-leucine_a
(2E)-3-Methyl-4-(sulfooxy)-2-butenoic acid_b
(2E)-3-Methyl-4-(sulfooxy)-2-butenoic acid_b
(2E)-3-Methyl-4-(sulfooxy)-2-butenoic acid_b
(2E)-3-Methyl-4-(sulfooxy)-2-butenoic acid_b
(2E)-3-Methyl-4-(sulfooxy)-2-butenoic acid_b
N-Acetylprocainamide_a
N-Acetylprocainamide_a
N-Acetylprocainamide_a
N-Acetylprocainamide_a
N-Acetylprocainamide_a

3-Methoxy-4-hydroxyhippuric acid_a
3-Methoxy-4-hydroxyhippuric acid_a
3-Methoxy-4-hydroxyhippuric acid_a
3-Methoxy-4-hydroxyhippuric acid_a
3-Methoxy-4-hydroxyhippuric acid_a
Valylvaline_f
Valylvaline_f
Valylvaline_f
Valylvaline_f
Valylvaline_f
N-(2-Cyanoethyl)-L-glutamine
N-(2-Cyanoethyl)-L-glutamine
N-(2-Cyanoethyl)-L-glutamine
N-(2-Cyanoethyl)-L-glutamine
N-(2-Cyanoethyl)-L-glutamine
Vorinostat_b
Vorinostat_b
Vorinostat_b
Vorinostat_b
Vorinostat_b
Ro 20-1724_a
Ro 20-1724_a
Ro 20-1724_a
Ro 20-1724_a
Ro 20-1724_a
3-(Sulfooxy)benzenepropanoic acid_b
3-(Sulfooxy)benzenepropanoic acid_b
3-(Sulfooxy)benzenepropanoic acid_b
3-(Sulfooxy)benzenepropanoic acid_b
3-(Sulfooxy)benzenepropanoic acid_b
Diacetin_b
Diacetin_b
Diacetin_b
Diacetin_b
Diacetin_b
Pentoxifylline
Pentoxifylline
Pentoxifylline
Pentoxifylline
Pentoxifylline
Pseudouridine
Pseudouridine
Pseudouridine
Pseudouridine
Pseudouridine
Bicine_b
Bicine_b
Bicine_b

Bicine_b
Bicine_b
Pro-tyr
Pro-tyr
Pro-tyr
Pro-tyr
Pro-tyr
NPK (Peptide Asn-Pro-Lys)
NPK (Peptide Asn-Pro-Lys)
NPK (Peptide Asn-Pro-Lys)
NPK (Peptide Asn-Pro-Lys)
NPK (Peptide Asn-Pro-Lys)
Formiminoglutamic Acid
Formiminoglutamic Acid
Formiminoglutamic Acid
Formiminoglutamic Acid
Formiminoglutamic Acid
N-COUMAROYL-L-ASPARTIC ACID
N-COUMAROYL-L-ASPARTIC ACID
N-COUMAROYL-L-ASPARTIC ACID
N-COUMAROYL-L-ASPARTIC ACID
N-COUMAROYL-L-ASPARTIC ACID
2-Methoxy-2-methyldihydro-3(2H)-thiophenone_a
2-Methoxy-2-methyldihydro-3(2H)-thiophenone_a
2-Methoxy-2-methyldihydro-3(2H)-thiophenone_a
2-Methoxy-2-methyldihydro-3(2H)-thiophenone_a
2-Methoxy-2-methyldihydro-3(2H)-thiophenone_a
nicotianamine_b
nicotianamine_b
nicotianamine_b
nicotianamine_b
nicotianamine_b
6-hydroxymethylpterin
6-hydroxymethylpterin
6-hydroxymethylpterin
6-hydroxymethylpterin
6-hydroxymethylpterin
Guanidinoethyl methyl phosphate
Guanidinoethyl methyl phosphate
Guanidinoethyl methyl phosphate
Guanidinoethyl methyl phosphate
Guanidinoethyl methyl phosphate
(3S,4S)-7,12-Dimethyl-3,4-dihydro-3,4-tetraphenediol
(3S,4S)-7,12-Dimethyl-3,4-dihydro-3,4-tetraphenediol
(3S,4S)-7,12-Dimethyl-3,4-dihydro-3,4-tetraphenediol
(3S,4S)-7,12-Dimethyl-3,4-dihydro-3,4-tetraphenediol
(3S,4S)-7,12-Dimethyl-3,4-dihydro-3,4-tetraphenediol
Tetrahydro-2,5-furan-diacetic acid

Tetrahydro-2,5-furan-diacetic acid
Tetrahydro-2,5-furan-diacetic acid
Tetrahydro-2,5-furan-diacetic acid
Tetrahydro-2,5-furan-diacetic acid
5-Hydroxy-7-methoxy-2,2-dimethyl-6-(3-phenylpropanoyl)-3,4-dihydro-2H-chromen-4-yl hydrogen sulfate
5-Hydroxy-7-methoxy-2,2-dimethyl-6-(3-phenylpropanoyl)-3,4-dihydro-2H-chromen-4-yl hydrogen sulfate
5-Hydroxy-7-methoxy-2,2-dimethyl-6-(3-phenylpropanoyl)-3,4-dihydro-2H-chromen-4-yl hydrogen sulfate
5-Hydroxy-7-methoxy-2,2-dimethyl-6-(3-phenylpropanoyl)-3,4-dihydro-2H-chromen-4-yl hydrogen sulfate
5-Hydroxy-7-methoxy-2,2-dimethyl-6-(3-phenylpropanoyl)-3,4-dihydro-2H-chromen-4-yl hydrogen sulfate
1-(2-Carboxyethyl)-2,3,4,9-tetrahydro-1H-beta-carboline-3-carboxylic acid_b
1-(2-Carboxyethyl)-2,3,4,9-tetrahydro-1H-beta-carboline-3-carboxylic acid_b
1-(2-Carboxyethyl)-2,3,4,9-tetrahydro-1H-beta-carboline-3-carboxylic acid_b
1-(2-Carboxyethyl)-2,3,4,9-tetrahydro-1H-beta-carboline-3-carboxylic acid_b
1-(2-Carboxyethyl)-2,3,4,9-tetrahydro-1H-beta-carboline-3-carboxylic acid_b
Gly-Trp_b
Gly-Trp_b
Gly-Trp_b
Gly-Trp_b
Gly-Trp_b
Dihydrouridine_b
Dihydrouridine_b
Dihydrouridine_b
Dihydrouridine_b
Dihydrouridine_b
1ÃŽÂ²-Hydroxycholic acid_b
1ÃŽÂ²-Hydroxycholic acid_b
1ÃŽÂ²-Hydroxycholic acid_b
1ÃŽÂ²-Hydroxycholic acid_b
1ÃŽÂ²-Hydroxycholic acid_b
Methdilazine
Methdilazine
Methdilazine
Methdilazine
Methdilazine
2'-Deoxyadenosine
2'-Deoxyadenosine
2'-Deoxyadenosine
2'-Deoxyadenosine
2'-Deoxyadenosine
Hydroxyphenobarbital
Hydroxyphenobarbital
Hydroxyphenobarbital
Hydroxyphenobarbital
Hydroxyphenobarbital
4,4'-Thiobis(butan-2-one)
4,4'-Thiobis(butan-2-one)
4,4'-Thiobis(butan-2-one)
4,4'-Thiobis(butan-2-one)

4,4'-Thiobis(butan-2-one)
His-pro_b
His-pro_b
His-pro_b
His-pro_b
His-pro_b
L-alpha-Aspartyl-L-phenylalanine
L-alpha-Aspartyl-L-phenylalanine
L-alpha-Aspartyl-L-phenylalanine
L-alpha-Aspartyl-L-phenylalanine
L-alpha-Aspartyl-L-phenylalanine
2-Hydroxyhepta-2_4-dienedioate
2-Hydroxyhepta-2_4-dienedioate
2-Hydroxyhepta-2_4-dienedioate
2-Hydroxyhepta-2_4-dienedioate
2-Hydroxyhepta-2_4-dienedioate
Choline Alfoscerate
Choline Alfoscerate
Choline Alfoscerate
Choline Alfoscerate
Choline Alfoscerate
4-(9H-beta-Carbolin-1-yl)-1,2,4-butanetriol_a
4-(9H-beta-Carbolin-1-yl)-1,2,4-butanetriol_a
4-(9H-beta-Carbolin-1-yl)-1,2,4-butanetriol_a
4-(9H-beta-Carbolin-1-yl)-1,2,4-butanetriol_a
4-(9H-beta-Carbolin-1-yl)-1,2,4-butanetriol_a
Val-Trp_b
Val-Trp_b
Val-Trp_b
Val-Trp_b
Val-Trp_b
9-(alpha-D-glucosyl)dihydrozeatin
9-(alpha-D-glucosyl)dihydrozeatin
9-(alpha-D-glucosyl)dihydrozeatin
9-(alpha-D-glucosyl)dihydrozeatin
9-(alpha-D-glucosyl)dihydrozeatin
SECONAL_d
SECONAL_d
SECONAL_d
SECONAL_d
SECONAL_d
epsilon-(gamma-Glutamyl)-lysine_d
epsilon-(gamma-Glutamyl)-lysine_d
epsilon-(gamma-Glutamyl)-lysine_d
epsilon-(gamma-Glutamyl)-lysine_d
epsilon-(gamma-Glutamyl)-lysine_d
3,4-Dihydroxyphenylglycol
3,4-Dihydroxyphenylglycol

3,4-Dihydroxyphenylglycol
3,4-Dihydroxyphenylglycol
3,4-Dihydroxyphenylglycol
(1S,3R,4s)-1,3,4,5-Tetrahydroxycyclohexanecarboxylic acid
(1S,3R,4s)-1,3,4,5-Tetrahydroxycyclohexanecarboxylic acid
(1S,3R,4s)-1,3,4,5-Tetrahydroxycyclohexanecarboxylic acid
(1S,3R,4s)-1,3,4,5-Tetrahydroxycyclohexanecarboxylic acid
(1S,3R,4s)-1,3,4,5-Tetrahydroxycyclohexanecarboxylic acid
4-Hydroxyaminoquinoline N-oxide
4-Hydroxyaminoquinoline N-oxide
4-Hydroxyaminoquinoline N-oxide
4-Hydroxyaminoquinoline N-oxide
4-Hydroxyaminoquinoline N-oxide
6-imino-5-oxocyclohexa-1,3-dienecarboxylic acid
6-imino-5-oxocyclohexa-1,3-dienecarboxylic acid
6-imino-5-oxocyclohexa-1,3-dienecarboxylic acid
6-imino-5-oxocyclohexa-1,3-dienecarboxylic acid
6-imino-5-oxocyclohexa-1,3-dienecarboxylic acid
3-[(2Z)-1-Oxo-2-buten-2-yl]pentanedioic acid_b
3-[(2Z)-1-Oxo-2-buten-2-yl]pentanedioic acid_b
3-[(2Z)-1-Oxo-2-buten-2-yl]pentanedioic acid_b
3-[(2Z)-1-Oxo-2-buten-2-yl]pentanedioic acid_b
3-[(2Z)-1-Oxo-2-buten-2-yl]pentanedioic acid_b
2-glyceryl 14,15-epoxy-(5Z,8Z,11Z)-icosatrienoate
2-glyceryl 14,15-epoxy-(5Z,8Z,11Z)-icosatrienoate
2-glyceryl 14,15-epoxy-(5Z,8Z,11Z)-icosatrienoate
2-glyceryl 14,15-epoxy-(5Z,8Z,11Z)-icosatrienoate
2-glyceryl 14,15-epoxy-(5Z,8Z,11Z)-icosatrienoate
Methylol Dimethylhydantoin_b
Methylol Dimethylhydantoin_b
Methylol Dimethylhydantoin_b
Methylol Dimethylhydantoin_b
Methylol Dimethylhydantoin_b
o-Succinylbenzoate
o-Succinylbenzoate
o-Succinylbenzoate
o-Succinylbenzoate
o-Succinylbenzoate
Gly-Trp_a
Gly-Trp_a
Gly-Trp_a
Gly-Trp_a
Gly-Trp_a
Glycylglycylglycine
Glycylglycylglycine
Glycylglycylglycine
Glycylglycylglycine
Glycylglycylglycine

trp-ser
trp-ser
trp-ser
trp-ser
trp-ser
asp-gln_a
asp-gln_a
asp-gln_a
asp-gln_a
asp-gln_a
N,N-Dimethyl-L-histidine_b
N,N-Dimethyl-L-histidine_b
N,N-Dimethyl-L-histidine_b
N,N-Dimethyl-L-histidine_b
N,N-Dimethyl-L-histidine_b
9-[(5R)-5-Ethyl-alpha-D-xylopyranosyl]-N-(3-methyl-2-buten-1-yl)-9H-purin-6-amine
9-[(5R)-5-Ethyl-alpha-D-xylopyranosyl]-N-(3-methyl-2-buten-1-yl)-9H-purin-6-amine
9-[(5R)-5-Ethyl-alpha-D-xylopyranosyl]-N-(3-methyl-2-buten-1-yl)-9H-purin-6-amine
9-[(5R)-5-Ethyl-alpha-D-xylopyranosyl]-N-(3-methyl-2-buten-1-yl)-9H-purin-6-amine
9-[(5R)-5-Ethyl-alpha-D-xylopyranosyl]-N-(3-methyl-2-buten-1-yl)-9H-purin-6-amine
Guanfacine
Guanfacine
Guanfacine
Guanfacine
Guanfacine
Sinapinic acid_a
Sinapinic acid_a
Sinapinic acid_a
Sinapinic acid_a
Sinapinic acid_a
Propantheline
Propantheline
Propantheline
Propantheline
Propantheline
Losalen
Losalen
Losalen
Losalen
Losalen
2-Methoxy-2-methyldihydro-3(2H)-thiophenone_b
2-Methoxy-2-methyldihydro-3(2H)-thiophenone_b
2-Methoxy-2-methyldihydro-3(2H)-thiophenone_b
2-Methoxy-2-methyldihydro-3(2H)-thiophenone_b
2-Methoxy-2-methyldihydro-3(2H)-thiophenone_b
Desonide
Desonide
Desonide

Desonide
Desonide
2-Methoxy-8-(7-methoxy-2-oxo-2H-chromen-6-yl)-6-methylnaphthoquinone
2-Methoxy-8-(7-methoxy-2-oxo-2H-chromen-6-yl)-6-methylnaphthoquinone
2-Methoxy-8-(7-methoxy-2-oxo-2H-chromen-6-yl)-6-methylnaphthoquinone
2-Methoxy-8-(7-methoxy-2-oxo-2H-chromen-6-yl)-6-methylnaphthoquinone
2-Methoxy-8-(7-methoxy-2-oxo-2H-chromen-6-yl)-6-methylnaphthoquinone
2-{(2S,4S)-4-[(3-Amino-2,3,6-trideoxyhexopyranosyl)oxy]-2,5,12-trihydroxy-7-methoxy-6,11-dioxo-1,2,3,4,6,11-h
2-{(2S,4S)-4-[(3-Amino-2,3,6-trideoxyhexopyranosyl)oxy]-2,5,12-trihydroxy-7-methoxy-6,11-dioxo-1,2,3,4,6,11-h
2-{(2S,4S)-4-[(3-Amino-2,3,6-trideoxyhexopyranosyl)oxy]-2,5,12-trihydroxy-7-methoxy-6,11-dioxo-1,2,3,4,6,11-h
2-{(2S,4S)-4-[(3-Amino-2,3,6-trideoxyhexopyranosyl)oxy]-2,5,12-trihydroxy-7-methoxy-6,11-dioxo-1,2,3,4,6,11-h
2-{(2S,4S)-4-[(3-Amino-2,3,6-trideoxyhexopyranosyl)oxy]-2,5,12-trihydroxy-7-methoxy-6,11-dioxo-1,2,3,4,6,11-h
2-BUTYL PROPENYL DISULFIDE, E(R)-_b
2-BUTYL PROPENYL DISULFIDE, E(R)-_b
2-BUTYL PROPENYL DISULFIDE, E(R)-_b
2-BUTYL PROPENYL DISULFIDE, E(R)-_b
2-BUTYL PROPENYL DISULFIDE, E(R)-_b
shinorine
shinorine
shinorine
shinorine
shinorine
ferrileghemoglobin
ferrileghemoglobin
ferrileghemoglobin
ferrileghemoglobin
ferrileghemoglobin
2-(1-Ethoxyethoxy)propanoic acid_c
2-(1-Ethoxyethoxy)propanoic acid_c
2-(1-Ethoxyethoxy)propanoic acid_c
2-(1-Ethoxyethoxy)propanoic acid_c
2-(1-Ethoxyethoxy)propanoic acid_c
Uramustine
Uramustine
Uramustine
Uramustine
Uramustine
2-Aminomuconic acid
2-Aminomuconic acid
2-Aminomuconic acid
2-Aminomuconic acid
2-Aminomuconic acid
(3R)-2-(3,4-Dihydroxyphenyl)-6,8-dihydroxy-3,4-dihydro-2H-chromen-3-yl sulfate
(3R)-2-(3,4-Dihydroxyphenyl)-6,8-dihydroxy-3,4-dihydro-2H-chromen-3-yl sulfate
(3R)-2-(3,4-Dihydroxyphenyl)-6,8-dihydroxy-3,4-dihydro-2H-chromen-3-yl sulfate
(3R)-2-(3,4-Dihydroxyphenyl)-6,8-dihydroxy-3,4-dihydro-2H-chromen-3-yl sulfate
(3R)-2-(3,4-Dihydroxyphenyl)-6,8-dihydroxy-3,4-dihydro-2H-chromen-3-yl sulfate
Artesunate

Artesunate
Artesunate
Artesunate
Artesunate
Bentazone
Bentazone
Bentazone
Bentazone
Bentazone
3-(3,4-dihydroxypyridinium-1-yl)-L-alanine
3-(3,4-dihydroxypyridinium-1-yl)-L-alanine
3-(3,4-dihydroxypyridinium-1-yl)-L-alanine
3-(3,4-dihydroxypyridinium-1-yl)-L-alanine
3-(3,4-dihydroxypyridinium-1-yl)-L-alanine
2,3,4,5-tetrahydrodipicolinic acid
2,3,4,5-tetrahydrodipicolinic acid
2,3,4,5-tetrahydrodipicolinic acid
2,3,4,5-tetrahydrodipicolinic acid
2,3,4,5-tetrahydrodipicolinic acid
Tetraacetylethylenediamine_e
Tetraacetylethylenediamine_e
Tetraacetylethylenediamine_e
Tetraacetylethylenediamine_e
Tetraacetylethylenediamine_e
L-fucopyranose 1-phosphate
L-fucopyranose 1-phosphate
L-fucopyranose 1-phosphate
L-fucopyranose 1-phosphate
L-fucopyranose 1-phosphate
4-(METHYLNITROSAMINO)-1-(3-PYRIDYL-N-OXIDE)-1-BUTANOL_e
4-(METHYLNITROSAMINO)-1-(3-PYRIDYL-N-OXIDE)-1-BUTANOL_e
4-(METHYLNITROSAMINO)-1-(3-PYRIDYL-N-OXIDE)-1-BUTANOL_e
4-(METHYLNITROSAMINO)-1-(3-PYRIDYL-N-OXIDE)-1-BUTANOL_e
4-(METHYLNITROSAMINO)-1-(3-PYRIDYL-N-OXIDE)-1-BUTANOL_e
3-Methoxy-4-hydroxyhippuric acid_c
3-Methoxy-4-hydroxyhippuric acid_c
3-Methoxy-4-hydroxyhippuric acid_c
3-Methoxy-4-hydroxyhippuric acid_c
3-Methoxy-4-hydroxyhippuric acid_c
3-Methoxy-4-hydroxyhippuric acid_f
3-Methoxy-4-hydroxyhippuric acid_f
3-Methoxy-4-hydroxyhippuric acid_f
3-Methoxy-4-hydroxyhippuric acid_f
3-Methoxy-4-hydroxyhippuric acid_f
Spaglumic acid
Spaglumic acid
Spaglumic acid
Spaglumic acid

Spaglumic acid
gamma-L-glutamyl-L-tyrosine
gamma-L-glutamyl-L-tyrosine
gamma-L-glutamyl-L-tyrosine
gamma-L-glutamyl-L-tyrosine
gamma-L-glutamyl-L-tyrosine
3-Hydroxytetradecanedioic acid
3-Hydroxytetradecanedioic acid
3-Hydroxytetradecanedioic acid
3-Hydroxytetradecanedioic acid
3-Hydroxytetradecanedioic acid
6-[(Z)-2-(3,4-Dimethoxyphenyl)vinyl]-4-methoxy-5,6-dihydro-2H-pyran-2-one
6-[(Z)-2-(3,4-Dimethoxyphenyl)vinyl]-4-methoxy-5,6-dihydro-2H-pyran-2-one
6-[(Z)-2-(3,4-Dimethoxyphenyl)vinyl]-4-methoxy-5,6-dihydro-2H-pyran-2-one
6-[(Z)-2-(3,4-Dimethoxyphenyl)vinyl]-4-methoxy-5,6-dihydro-2H-pyran-2-one
6-[(Z)-2-(3,4-Dimethoxyphenyl)vinyl]-4-methoxy-5,6-dihydro-2H-pyran-2-one
Zinecard_b
Zinecard_b
Zinecard_b
Zinecard_b
Zinecard_b
Aminohippuric acid_a
Aminohippuric acid_a
Aminohippuric acid_a
Aminohippuric acid_a
Aminohippuric acid_a
Benzamide
Benzamide
Benzamide
Benzamide
Benzamide
3-Benzyl-6-isobutyl-2,5-piperazinedione_c
3-Benzyl-6-isobutyl-2,5-piperazinedione_c
3-Benzyl-6-isobutyl-2,5-piperazinedione_c
3-Benzyl-6-isobutyl-2,5-piperazinedione_c
3-Benzyl-6-isobutyl-2,5-piperazinedione_c
L-Glutamic acid, 5-[2-(4-carboxyphenyl)hydrazide]
L-Glutamic acid, 5-[2-(4-carboxyphenyl)hydrazide]
L-Glutamic acid, 5-[2-(4-carboxyphenyl)hydrazide]
L-Glutamic acid, 5-[2-(4-carboxyphenyl)hydrazide]
L-Glutamic acid, 5-[2-(4-carboxyphenyl)hydrazide]
Pyrimidine 5'-nucleotide
Pyrimidine 5'-nucleotide
Pyrimidine 5'-nucleotide
Pyrimidine 5'-nucleotide
Pyrimidine 5'-nucleotide
Dihydrouracil_a
Dihydrouracil_a

Dihydrouracil_a
Dihydrouracil_a
Dihydrouracil_a
9,11-Dihydroxy-2-methoxy-12H-benzo[a]xanthen-12-one
9,11-Dihydroxy-2-methoxy-12H-benzo[a]xanthen-12-one
9,11-Dihydroxy-2-methoxy-12H-benzo[a]xanthen-12-one
9,11-Dihydroxy-2-methoxy-12H-benzo[a]xanthen-12-one
9,11-Dihydroxy-2-methoxy-12H-benzo[a]xanthen-12-one
2-Amino-6-[(E)-(5-amino-5-carboxy-2-hydroxypentylidene)amino]-5-hydroxyhexanoic acid (non-preferred name)
2-Amino-6-[(E)-(5-amino-5-carboxy-2-hydroxypentylidene)amino]-5-hydroxyhexanoic acid (non-preferred name)
2-Amino-6-[(E)-(5-amino-5-carboxy-2-hydroxypentylidene)amino]-5-hydroxyhexanoic acid (non-preferred name)
2-Amino-6-[(E)-(5-amino-5-carboxy-2-hydroxypentylidene)amino]-5-hydroxyhexanoic acid (non-preferred name)
2-Amino-6-[(E)-(5-amino-5-carboxy-2-hydroxypentylidene)amino]-5-hydroxyhexanoic acid (non-preferred name)
Mono(3-carboxypropyl) phthalate
Mono(3-carboxypropyl) phthalate
Mono(3-carboxypropyl) phthalate
Mono(3-carboxypropyl) phthalate
Mono(3-carboxypropyl) phthalate
3-Methyladenine_b
3-Methyladenine_b
3-Methyladenine_b
3-Methyladenine_b
3-Methyladenine_b
(2E)-5-Hydroxyferulic acid
(2E)-5-Hydroxyferulic acid
(2E)-5-Hydroxyferulic acid
(2E)-5-Hydroxyferulic acid
(2E)-5-Hydroxyferulic acid
Tyrosol
Tyrosol
Tyrosol
Tyrosol
Tyrosol
N-Nonanoylglycine_b
N-Nonanoylglycine_b
N-Nonanoylglycine_b
N-Nonanoylglycine_b
N-Nonanoylglycine_b
Valylvaline_b
Valylvaline_b
Valylvaline_b
Valylvaline_b
Valylvaline_b
N-Acetyl-S-(allylcarbamothioyl)cysteine
N-Acetyl-S-(allylcarbamothioyl)cysteine
N-Acetyl-S-(allylcarbamothioyl)cysteine
N-Acetyl-S-(allylcarbamothioyl)cysteine
N-Acetyl-S-(allylcarbamothioyl)cysteine

L-gamma-Glutamyl-L-valine_d
L-gamma-Glutamyl-L-valine_d
L-gamma-Glutamyl-L-valine_d
L-gamma-Glutamyl-L-valine_d
L-gamma-Glutamyl-L-valine_d
N-(4-Amino-1-carboxybutyl)glutamic acid_b
N-(4-Amino-1-carboxybutyl)glutamic acid_b
N-(4-Amino-1-carboxybutyl)glutamic acid_b
N-(4-Amino-1-carboxybutyl)glutamic acid_b
N-(4-Amino-1-carboxybutyl)glutamic acid_b
1,4-Naphthoquinone
1,4-Naphthoquinone
1,4-Naphthoquinone
1,4-Naphthoquinone
1,4-Naphthoquinone
Lisdexamfetamine
Lisdexamfetamine
Lisdexamfetamine
Lisdexamfetamine
Lisdexamfetamine
asp-gln_b
asp-gln_b
asp-gln_b
asp-gln_b
asp-gln_b
pterin
pterin
pterin
pterin
pterin
13a-Hydroxy-2,3-dimethoxy-6a,13a-dihydrochromeno[3,4-b]furo[3,2-g]chromen-13(6H)-one
13a-Hydroxy-2,3-dimethoxy-6a,13a-dihydrochromeno[3,4-b]furo[3,2-g]chromen-13(6H)-one
13a-Hydroxy-2,3-dimethoxy-6a,13a-dihydrochromeno[3,4-b]furo[3,2-g]chromen-13(6H)-one
13a-Hydroxy-2,3-dimethoxy-6a,13a-dihydrochromeno[3,4-b]furo[3,2-g]chromen-13(6H)-one
13a-Hydroxy-2,3-dimethoxy-6a,13a-dihydrochromeno[3,4-b]furo[3,2-g]chromen-13(6H)-one
6-(alpha-D-glucosaminyl)-1D-myo-inositol_b
6-(alpha-D-glucosaminyl)-1D-myo-inositol_b
6-(alpha-D-glucosaminyl)-1D-myo-inositol_b
6-(alpha-D-glucosaminyl)-1D-myo-inositol_b
6-(alpha-D-glucosaminyl)-1D-myo-inositol_b
Dinoseb
Dinoseb
Dinoseb
Dinoseb
Dinoseb
Methional
Methional
Methional

Methional
Methional
epsilon-(gamma-Glutamyl)-lysine_c
epsilon-(gamma-Glutamyl)-lysine_c
epsilon-(gamma-Glutamyl)-lysine_c
epsilon-(gamma-Glutamyl)-lysine_c
epsilon-(gamma-Glutamyl)-lysine_c
hydroxyhexamide
hydroxyhexamide
hydroxyhexamide
hydroxyhexamide
hydroxyhexamide
1-Methylinosine
1-Methylinosine
1-Methylinosine
1-Methylinosine
1-Methylinosine
AAMU_b
AAMU_b
AAMU_b
AAMU_b
AAMU_b
Minoxidil
Minoxidil
Minoxidil
Minoxidil
Minoxidil
N-[(4-Methoxy-1-benzofuran-5-yl)carbonyl]glycine
N-[(4-Methoxy-1-benzofuran-5-yl)carbonyl]glycine
N-[(4-Methoxy-1-benzofuran-5-yl)carbonyl]glycine
N-[(4-Methoxy-1-benzofuran-5-yl)carbonyl]glycine
N-[(4-Methoxy-1-benzofuran-5-yl)carbonyl]glycine
Ethosuximide
Ethosuximide
Ethosuximide
Ethosuximide
Ethosuximide
5-Methoxy-3-indoleaceate
5-Methoxy-3-indoleaceate
5-Methoxy-3-indoleaceate
5-Methoxy-3-indoleaceate
5-Methoxy-3-indoleaceate
16alpha-hydroxydehydroepiandrosterone 3-sulfate
16alpha-hydroxydehydroepiandrosterone 3-sulfate
16alpha-hydroxydehydroepiandrosterone 3-sulfate
16alpha-hydroxydehydroepiandrosterone 3-sulfate
16alpha-hydroxydehydroepiandrosterone 3-sulfate
7-Aminomethyl-7-deazaguanine

7-Aminomethyl-7-deazaguanine
7-Aminomethyl-7-deazaguanine
7-Aminomethyl-7-deazaguanine
7-Aminomethyl-7-deazaguanine
2-(4-Isopropyl-4-methyl-5-oxo-4,5-dihydro-1H-imidazol-2-yl)-4-methylbenzoic acid_b
2-(4-Isopropyl-4-methyl-5-oxo-4,5-dihydro-1H-imidazol-2-yl)-4-methylbenzoic acid_b
2-(4-Isopropyl-4-methyl-5-oxo-4,5-dihydro-1H-imidazol-2-yl)-4-methylbenzoic acid_b
2-(4-Isopropyl-4-methyl-5-oxo-4,5-dihydro-1H-imidazol-2-yl)-4-methylbenzoic acid_b
2-(4-Isopropyl-4-methyl-5-oxo-4,5-dihydro-1H-imidazol-2-yl)-4-methylbenzoic acid_b
his-asn
his-asn
his-asn
his-asn
his-asn
meprobamate_d
meprobamate_d
meprobamate_d
meprobamate_d
meprobamate_d
6-Sulfatoxymelatonin
6-Sulfatoxymelatonin
6-Sulfatoxymelatonin
6-Sulfatoxymelatonin
6-Sulfatoxymelatonin
Toluene_b
Toluene_b
Toluene_b
Toluene_b
Toluene_b
Trifluoromethyl-bismethyl ketone
Trifluoromethyl-bismethyl ketone
Trifluoromethyl-bismethyl ketone
Trifluoromethyl-bismethyl ketone
Trifluoromethyl-bismethyl ketone
Taxifolin
Taxifolin
Taxifolin
Taxifolin
Taxifolin
17-Hydroxypregnenolone sulfate
17-Hydroxypregnenolone sulfate
17-Hydroxypregnenolone sulfate
17-Hydroxypregnenolone sulfate
17-Hydroxypregnenolone sulfate
5-O-alpha-L-Arabinofuranosyl-alpha-L-arabinofuranose
5-O-alpha-L-Arabinofuranosyl-alpha-L-arabinofuranose
5-O-alpha-L-Arabinofuranosyl-alpha-L-arabinofuranose
5-O-alpha-L-Arabinofuranosyl-alpha-L-arabinofuranose

5-O-alpha-L-Arabinofuranosyl-alpha-L-arabinofuranose
Sinapinic acid_b
Sinapinic acid_b
Sinapinic acid_b
Sinapinic acid_b
Sinapinic acid_b
N-Desalkylflurazepam
N-Desalkylflurazepam
N-Desalkylflurazepam
N-Desalkylflurazepam
N-Desalkylflurazepam
MFCD09953737_a
MFCD09953737_a
MFCD09953737_a
MFCD09953737_a
MFCD09953737_a
L-Proline, 4-hydroxy-5-oxo-4-(tetrahydro-2,3,4-trihydroxy-2-furanyl)-
L-Proline, 4-hydroxy-5-oxo-4-(tetrahydro-2,3,4-trihydroxy-2-furanyl)-
L-Proline, 4-hydroxy-5-oxo-4-(tetrahydro-2,3,4-trihydroxy-2-furanyl)-
L-Proline, 4-hydroxy-5-oxo-4-(tetrahydro-2,3,4-trihydroxy-2-furanyl)-
L-Proline, 4-hydroxy-5-oxo-4-(tetrahydro-2,3,4-trihydroxy-2-furanyl)-
Cilazapril
Cilazapril
Cilazapril
Cilazapril
Cilazapril
MFCD02728197_c
MFCD02728197_c
MFCD02728197_c
MFCD02728197_c
MFCD02728197_c
2-Hydroxyphenylacetic acid
2-Hydroxyphenylacetic acid
2-Hydroxyphenylacetic acid
2-Hydroxyphenylacetic acid
2-Hydroxyphenylacetic acid
N-(1-{[Methyl(2-methyl-2-propanyl)carbamoyl]amino}ethyl)-alpha-asparagine
N-(1-{[Methyl(2-methyl-2-propanyl)carbamoyl]amino}ethyl)-alpha-asparagine
N-(1-{[Methyl(2-methyl-2-propanyl)carbamoyl]amino}ethyl)-alpha-asparagine
N-(1-{[Methyl(2-methyl-2-propanyl)carbamoyl]amino}ethyl)-alpha-asparagine
N-(1-{[Methyl(2-methyl-2-propanyl)carbamoyl]amino}ethyl)-alpha-asparagine
Hypericin
Hypericin
Hypericin
Hypericin
Hypericin
(2E)-N-3,7-dimethyl-2,6-octadienyl cyclopropylcarboxamide
(2E)-N-3,7-dimethyl-2,6-octadienyl cyclopropylcarboxamide

(2E)-N-3,7-dimethyl-2,6-octadienyl cyclopropylcarboxamide
(2E)-N-3,7-dimethyl-2,6-octadienyl cyclopropylcarboxamide
(2E)-N-3,7-dimethyl-2,6-octadienyl cyclopropylcarboxamide
Casimiroin
Casimiroin
Casimiroin
Casimiroin
Casimiroin
pro-gln_c
pro-gln_c
pro-gln_c
pro-gln_c
pro-gln_c
Histidylphenylalanine
Histidylphenylalanine
Histidylphenylalanine
Histidylphenylalanine
Histidylphenylalanine
L-gamma-Glutamyl-L-leucine_d
L-gamma-Glutamyl-L-leucine_d
L-gamma-Glutamyl-L-leucine_d
L-gamma-Glutamyl-L-leucine_d
L-gamma-Glutamyl-L-leucine_d
ELK (Peptide Glu-Leu-Lys)_b
ELK (Peptide Glu-Leu-Lys)_b
ELK (Peptide Glu-Leu-Lys)_b
ELK (Peptide Glu-Leu-Lys)_b
ELK (Peptide Glu-Leu-Lys)_b
(3aS,5S,6R,7aR,7bS,9aS,10R,12aS,12bS)-10-[(2S,3R,4R)-3,4-Dihydroxy-6-methyl-2-heptanyl]-5,6-dihydroxy-7a,[illegible]
(3aS,5S,6R,7aR,7bS,9aS,10R,12aS,12bS)-10-[(2S,3R,4R)-3,4-Dihydroxy-6-methyl-2-heptanyl]-5,6-dihydroxy-7a,[illegible]
(3aS,5S,6R,7aR,7bS,9aS,10R,12aS,12bS)-10-[(2S,3R,4R)-3,4-Dihydroxy-6-methyl-2-heptanyl]-5,6-dihydroxy-7a,[illegible]
(3aS,5S,6R,7aR,7bS,9aS,10R,12aS,12bS)-10-[(2S,3R,4R)-3,4-Dihydroxy-6-methyl-2-heptanyl]-5,6-dihydroxy-7a,[illegible]
(3aS,5S,6R,7aR,7bS,9aS,10R,12aS,12bS)-10-[(2S,3R,4R)-3,4-Dihydroxy-6-methyl-2-heptanyl]-5,6-dihydroxy-7a,[illegible]
UQ3675000
UQ3675000
UQ3675000
UQ3675000
UQ3675000
S-Sulfocysteine
S-Sulfocysteine
S-Sulfocysteine
S-Sulfocysteine
S-Sulfocysteine
cis-3-Hexenyl Pyruvate
cis-3-Hexenyl Pyruvate
cis-3-Hexenyl Pyruvate
cis-3-Hexenyl Pyruvate
cis-3-Hexenyl Pyruvate

3,6-Dichloro-3,5-cyclohexadiene-1,2-diol
3,6-Dichloro-3,5-cyclohexadiene-1,2-diol
3,6-Dichloro-3,5-cyclohexadiene-1,2-diol
3,6-Dichloro-3,5-cyclohexadiene-1,2-diol
3,6-Dichloro-3,5-cyclohexadiene-1,2-diol
Lys-phe_a
Lys-phe_a
Lys-phe_a
Lys-phe_a
Lys-phe_a
felbamate
felbamate
felbamate
felbamate
felbamate
Urothion
Urothion
Urothion
Urothion
Urothion
[7-Hydroxy-1-(4-hydroxy-3-methoxyphenyl)-3-(hydroxymethyl)-6-methoxy-1,2,3,4-tetrahydro-2-naphthalenyl]met
[7-Hydroxy-1-(4-hydroxy-3-methoxyphenyl)-3-(hydroxymethyl)-6-methoxy-1,2,3,4-tetrahydro-2-naphthalenyl]met
[7-Hydroxy-1-(4-hydroxy-3-methoxyphenyl)-3-(hydroxymethyl)-6-methoxy-1,2,3,4-tetrahydro-2-naphthalenyl]met
[7-Hydroxy-1-(4-hydroxy-3-methoxyphenyl)-3-(hydroxymethyl)-6-methoxy-1,2,3,4-tetrahydro-2-naphthalenyl]met
[7-Hydroxy-1-(4-hydroxy-3-methoxyphenyl)-3-(hydroxymethyl)-6-methoxy-1,2,3,4-tetrahydro-2-naphthalenyl]met
4-(METHYLNITROSAMINO)-1-(3-PYRIDYL-N-OXIDE)-1-BUTANOL_b
4-(METHYLNITROSAMINO)-1-(3-PYRIDYL-N-OXIDE)-1-BUTANOL_b
4-(METHYLNITROSAMINO)-1-(3-PYRIDYL-N-OXIDE)-1-BUTANOL_b
4-(METHYLNITROSAMINO)-1-(3-PYRIDYL-N-OXIDE)-1-BUTANOL_b
4-(METHYLNITROSAMINO)-1-(3-PYRIDYL-N-OXIDE)-1-BUTANOL_b
3,4-dihydroxyphenylpyruvic acid
3,4-dihydroxyphenylpyruvic acid
3,4-dihydroxyphenylpyruvic acid
3,4-dihydroxyphenylpyruvic acid
3,4-dihydroxyphenylpyruvic acid
imazamethabenz-methyl_a
imazamethabenz-methyl_a
imazamethabenz-methyl_a
imazamethabenz-methyl_a
imazamethabenz-methyl_a
Dimeric mercapto propanone
Dimeric mercapto propanone
Dimeric mercapto propanone
Dimeric mercapto propanone
Dimeric mercapto propanone
N-(4-Hydroxy-3,5-dimethoxybenzoyl)glycine_b
N-(4-Hydroxy-3,5-dimethoxybenzoyl)glycine_b
N-(4-Hydroxy-3,5-dimethoxybenzoyl)glycine_b

N-(4-Hydroxy-3,5-dimethoxybenzoyl)glycine_b
N-(4-Hydroxy-3,5-dimethoxybenzoyl)glycine_b
H-DL-MET-DL-MET-OH
H-DL-MET-DL-MET-OH
H-DL-MET-DL-MET-OH
H-DL-MET-DL-MET-OH
H-DL-MET-DL-MET-OH
3-(4,7-Dimethoxy-1,3-benzodioxol-5-yl)propanoic acid
3-(4,7-Dimethoxy-1,3-benzodioxol-5-yl)propanoic acid
3-(4,7-Dimethoxy-1,3-benzodioxol-5-yl)propanoic acid
3-(4,7-Dimethoxy-1,3-benzodioxol-5-yl)propanoic acid
3-(4,7-Dimethoxy-1,3-benzodioxol-5-yl)propanoic acid
2-methoxyacetaminophen sulfate
2-methoxyacetaminophen sulfate
2-methoxyacetaminophen sulfate
2-methoxyacetaminophen sulfate
2-methoxyacetaminophen sulfate
NPYR
NPYR
NPYR
NPYR
NPYR
2-Acetamidohexanedioic acid_c
2-Acetamidohexanedioic acid_c
2-Acetamidohexanedioic acid_c
2-Acetamidohexanedioic acid_c
2-Acetamidohexanedioic acid_c
DIBOA
DIBOA
DIBOA
DIBOA
DIBOA
3-(Sulfooxy)benzenepropanoic acid_a
3-(Sulfooxy)benzenepropanoic acid_a
3-(Sulfooxy)benzenepropanoic acid_a
3-(Sulfooxy)benzenepropanoic acid_a
3-(Sulfooxy)benzenepropanoic acid_a
Redul
Redul
Redul
Redul
Redul
(S)-2-hydrazino-3-(4-hydroxy-3-methoxyphenyl)-2-methylpropionic acid
(S)-2-hydrazino-3-(4-hydroxy-3-methoxyphenyl)-2-methylpropionic acid
(S)-2-hydrazino-3-(4-hydroxy-3-methoxyphenyl)-2-methylpropionic acid
(S)-2-hydrazino-3-(4-hydroxy-3-methoxyphenyl)-2-methylpropionic acid
(S)-2-hydrazino-3-(4-hydroxy-3-methoxyphenyl)-2-methylpropionic acid
S-Allylcysteine_b

S-Allylcysteine_b
S-Allylcysteine_b
S-Allylcysteine_b
S-Allylcysteine_b
Scopoletin acetate
Scopoletin acetate
Scopoletin acetate
Scopoletin acetate
Scopoletin acetate
Azulfidine
Azulfidine
Azulfidine
Azulfidine
Azulfidine
S(8)-aminomethyldihydrolipoamide
S(8)-aminomethyldihydrolipoamide
S(8)-aminomethyldihydrolipoamide
S(8)-aminomethyldihydrolipoamide
S(8)-aminomethyldihydrolipoamide
GLY-MET
GLY-MET
GLY-MET
GLY-MET
GLY-MET
3-Benzyl-6-isobutyl-2,5-piperazinedione_b
3-Benzyl-6-isobutyl-2,5-piperazinedione_b
3-Benzyl-6-isobutyl-2,5-piperazinedione_b
3-Benzyl-6-isobutyl-2,5-piperazinedione_b
3-Benzyl-6-isobutyl-2,5-piperazinedione_b
(3aS,5S,6R,7aR,7bS,9aS,10R,12aS,12bS)-10-[(2S,3R,4R)-3,4-Dihydroxy-6-methyl-2-heptanyl]-5,6-dihydroxy-7a,[illegible]
(3aS,5S,6R,7aR,7bS,9aS,10R,12aS,12bS)-10-[(2S,3R,4R)-3,4-Dihydroxy-6-methyl-2-heptanyl]-5,6-dihydroxy-7a,[illegible]
(3aS,5S,6R,7aR,7bS,9aS,10R,12aS,12bS)-10-[(2S,3R,4R)-3,4-Dihydroxy-6-methyl-2-heptanyl]-5,6-dihydroxy-7a,[illegible]
(3aS,5S,6R,7aR,7bS,9aS,10R,12aS,12bS)-10-[(2S,3R,4R)-3,4-Dihydroxy-6-methyl-2-heptanyl]-5,6-dihydroxy-7a,[illegible]
(3aS,5S,6R,7aR,7bS,9aS,10R,12aS,12bS)-10-[(2S,3R,4R)-3,4-Dihydroxy-6-methyl-2-heptanyl]-5,6-dihydroxy-7a,[illegible]
Trolox
Trolox
Trolox
Trolox
Trolox
1ÃŽÂ²-Hydroxycholic acid_a
1ÃŽÂ²-Hydroxycholic acid_a
1ÃŽÂ²-Hydroxycholic acid_a
1ÃŽÂ²-Hydroxycholic acid_a
1ÃŽÂ²-Hydroxycholic acid_a
Xanthurenic acid
Xanthurenic acid
Xanthurenic acid
Xanthurenic acid

Xanthurenic acid
N-[(10Z)-7-Isobutyl-3-isopropyl-5,8-dioxo-2-oxa-6,9-diazabicyclo[10.2.2]hexadeca-1(14),10,12,15-tetraen-4-yl]-
N-[(10Z)-7-Isobutyl-3-isopropyl-5,8-dioxo-2-oxa-6,9-diazabicyclo[10.2.2]hexadeca-1(14),10,12,15-tetraen-4-yl]-
N-[(10Z)-7-Isobutyl-3-isopropyl-5,8-dioxo-2-oxa-6,9-diazabicyclo[10.2.2]hexadeca-1(14),10,12,15-tetraen-4-yl]-
N-[(10Z)-7-Isobutyl-3-isopropyl-5,8-dioxo-2-oxa-6,9-diazabicyclo[10.2.2]hexadeca-1(14),10,12,15-tetraen-4-yl]-
N-[(10Z)-7-Isobutyl-3-isopropyl-5,8-dioxo-2-oxa-6,9-diazabicyclo[10.2.2]hexadeca-1(14),10,12,15-tetraen-4-yl]-
Serotonin
Serotonin
Serotonin
Serotonin
Serotonin
mesifurane_c
mesifurane_c
mesifurane_c
mesifurane_c
mesifurane_c
2,3-Dihydroxypropyl 3,4,5-trihydroxybenzoate
2,3-Dihydroxypropyl 3,4,5-trihydroxybenzoate
2,3-Dihydroxypropyl 3,4,5-trihydroxybenzoate
2,3-Dihydroxypropyl 3,4,5-trihydroxybenzoate
2,3-Dihydroxypropyl 3,4,5-trihydroxybenzoate
methocarbamol
methocarbamol
methocarbamol
methocarbamol
methocarbamol
N-[(2S)-2-Hydroxypropanoyl]-L-tryptophan
N-[(2S)-2-Hydroxypropanoyl]-L-tryptophan
N-[(2S)-2-Hydroxypropanoyl]-L-tryptophan
N-[(2S)-2-Hydroxypropanoyl]-L-tryptophan
N-[(2S)-2-Hydroxypropanoyl]-L-tryptophan
g-Aminobutyryl-lysine_c
g-Aminobutyryl-lysine_c
g-Aminobutyryl-lysine_c
g-Aminobutyryl-lysine_c
g-Aminobutyryl-lysine_c
quinol sulfate_a
quinol sulfate_a
quinol sulfate_a
quinol sulfate_a
quinol sulfate_a
Nitrendipine
Nitrendipine
Nitrendipine
Nitrendipine
Nitrendipine
(2S)-3-(1H-Indol-3-yl)-2-({[(3S,4S,5R)-2,3,4-trihydroxy-5-(hydroxymethyl)tetrahydro-2-furanyl]methyl}amino)prop
(2S)-3-(1H-Indol-3-yl)-2-({[(3S,4S,5R)-2,3,4-trihydroxy-5-(hydroxymethyl)tetrahydro-2-furanyl]methyl}amino)prop

(2S)-3-(1H-Indol-3-yl)-2-({[(3S,4S,5R)-2,3,4-trihydroxy-5-(hydroxymethyl)tetrahydro-2-furanyl]methyl}amino)prop
(2S)-3-(1H-Indol-3-yl)-2-({[(3S,4S,5R)-2,3,4-trihydroxy-5-(hydroxymethyl)tetrahydro-2-furanyl]methyl}amino)prop
(2S)-3-(1H-Indol-3-yl)-2-({[(3S,4S,5R)-2,3,4-trihydroxy-5-(hydroxymethyl)tetrahydro-2-furanyl]methyl}amino)prop
N-D-Glucosylarylamine
N-D-Glucosylarylamine
N-D-Glucosylarylamine
N-D-Glucosylarylamine
N-D-Glucosylarylamine
3,7,12,17-Tetramethyl-21H,23H-porphine-2,18-dipropanoic Acid
3,7,12,17-Tetramethyl-21H,23H-porphine-2,18-dipropanoic Acid
3,7,12,17-Tetramethyl-21H,23H-porphine-2,18-dipropanoic Acid
3,7,12,17-Tetramethyl-21H,23H-porphine-2,18-dipropanoic Acid
3,7,12,17-Tetramethyl-21H,23H-porphine-2,18-dipropanoic Acid
1,2-dihydroxy-3-keto-5-methylthiopentene
1,2-dihydroxy-3-keto-5-methylthiopentene
1,2-dihydroxy-3-keto-5-methylthiopentene
1,2-dihydroxy-3-keto-5-methylthiopentene
1,2-dihydroxy-3-keto-5-methylthiopentene
(5Z)-2-Amino-4,5-dihydroxy-5-{[3-(hydroxymethyl)-2,5-dihydro-2-furanyl]imino}pentanoic acid (non-preferred nar
(5Z)-2-Amino-4,5-dihydroxy-5-{[3-(hydroxymethyl)-2,5-dihydro-2-furanyl]imino}pentanoic acid (non-preferred nar
(5Z)-2-Amino-4,5-dihydroxy-5-{[3-(hydroxymethyl)-2,5-dihydro-2-furanyl]imino}pentanoic acid (non-preferred nar
(5Z)-2-Amino-4,5-dihydroxy-5-{[3-(hydroxymethyl)-2,5-dihydro-2-furanyl]imino}pentanoic acid (non-preferred nar
(5Z)-2-Amino-4,5-dihydroxy-5-{[3-(hydroxymethyl)-2,5-dihydro-2-furanyl]imino}pentanoic acid (non-preferred nar
2-(2-Amino-2-carboxyethyl)-5-oxotetrahydro-2-furancarboxylic acid
2-(2-Amino-2-carboxyethyl)-5-oxotetrahydro-2-furancarboxylic acid
2-(2-Amino-2-carboxyethyl)-5-oxotetrahydro-2-furancarboxylic acid
2-(2-Amino-2-carboxyethyl)-5-oxotetrahydro-2-furancarboxylic acid
2-(2-Amino-2-carboxyethyl)-5-oxotetrahydro-2-furancarboxylic acid
Tetraacetylethylenediamine_a
Tetraacetylethylenediamine_a
Tetraacetylethylenediamine_a
Tetraacetylethylenediamine_a
Tetraacetylethylenediamine_a
4-(METHYLNITROSAMINO)-1-(3-PYRIDYL-N-OXIDE)-1-BUTANONE_a
4-(METHYLNITROSAMINO)-1-(3-PYRIDYL-N-OXIDE)-1-BUTANONE_a
4-(METHYLNITROSAMINO)-1-(3-PYRIDYL-N-OXIDE)-1-BUTANONE_a
4-(METHYLNITROSAMINO)-1-(3-PYRIDYL-N-OXIDE)-1-BUTANONE_a
4-(METHYLNITROSAMINO)-1-(3-PYRIDYL-N-OXIDE)-1-BUTANONE_a
L-gamma-Glutamyl-L-valine_a
L-gamma-Glutamyl-L-valine_a
L-gamma-Glutamyl-L-valine_a
L-gamma-Glutamyl-L-valine_a
L-gamma-Glutamyl-L-valine_a
(19R,25S)-22,25,28,28-Tetrahydroxy-22,28-dioxido-16-oxo-17,21,23,27-tetraoxa-22lambda~5~,28lambda~5~-d
(19R,25S)-22,25,28,28-Tetrahydroxy-22,28-dioxido-16-oxo-17,21,23,27-tetraoxa-22lambda~5~,28lambda~5~-d
(19R,25S)-22,25,28,28-Tetrahydroxy-22,28-dioxido-16-oxo-17,21,23,27-tetraoxa-22lambda~5~,28lambda~5~-d
(19R,25S)-22,25,28,28-Tetrahydroxy-22,28-dioxido-16-oxo-17,21,23,27-tetraoxa-22lambda~5~,28lambda~5~-d
(19R,25S)-22,25,28,28-Tetrahydroxy-22,28-dioxido-16-oxo-17,21,23,27-tetraoxa-22lambda~5~,28lambda~5~-d

1-(4-Methylphenyl)pyrrolidine-2,5-dione
1-(4-Methylphenyl)pyrrolidine-2,5-dione
1-(4-Methylphenyl)pyrrolidine-2,5-dione
1-(4-Methylphenyl)pyrrolidine-2,5-dione
1-(4-Methylphenyl)pyrrolidine-2,5-dione
Selsun
Selsun
Selsun
Selsun
Selsun
Diacetin_a
Diacetin_a
Diacetin_a
Diacetin_a
Diacetin_a
FB9500000_c
FB9500000_c
FB9500000_c
FB9500000_c
FB9500000_c
Coprine_a
Coprine_a
Coprine_a
Coprine_a
Coprine_a
N,N-Dimethyl-L-histidine_c
N,N-Dimethyl-L-histidine_c
N,N-Dimethyl-L-histidine_c
N,N-Dimethyl-L-histidine_c
N,N-Dimethyl-L-histidine_c
N-[(2S)-2-Hydroxypropanoyl]methionine_a
N-[(2S)-2-Hydroxypropanoyl]methionine_a
N-[(2S)-2-Hydroxypropanoyl]methionine_a
N-[(2S)-2-Hydroxypropanoyl]methionine_a
N-[(2S)-2-Hydroxypropanoyl]methionine_a
gamma-Glu-gln
gamma-Glu-gln
gamma-Glu-gln
gamma-Glu-gln
gamma-Glu-gln
(DL)-3-O-Methyldopa_b
(DL)-3-O-Methyldopa_b
(DL)-3-O-Methyldopa_b
(DL)-3-O-Methyldopa_b
(DL)-3-O-Methyldopa_b
vinyl sulfide
vinyl sulfide
vinyl sulfide

vinyl sulfide
vinyl sulfide
Valylvaline_a
Valylvaline_a
Valylvaline_a
Valylvaline_a
Valylvaline_a
1-(2,3-Dihydro-1H-pyrrolizin-5-yl)-1,4-pentanedione
1-(2,3-Dihydro-1H-pyrrolizin-5-yl)-1,4-pentanedione
1-(2,3-Dihydro-1H-pyrrolizin-5-yl)-1,4-pentanedione
1-(2,3-Dihydro-1H-pyrrolizin-5-yl)-1,4-pentanedione
1-(2,3-Dihydro-1H-pyrrolizin-5-yl)-1,4-pentanedione
Glycylleucine
Glycylleucine
Glycylleucine
Glycylleucine
Glycylleucine
O-heptanoylcarnitine
O-heptanoylcarnitine
O-heptanoylcarnitine
O-heptanoylcarnitine
O-heptanoylcarnitine
Tetraacetylethylenediamine_d
Tetraacetylethylenediamine_d
Tetraacetylethylenediamine_d
Tetraacetylethylenediamine_d
Tetraacetylethylenediamine_d
riboprine
riboprine
riboprine
riboprine
riboprine
Midodrine_c
Midodrine_c
Midodrine_c
Midodrine_c
Midodrine_c
Hydroxycarteolol_c
Hydroxycarteolol_c
Hydroxycarteolol_c
Hydroxycarteolol_c
Hydroxycarteolol_c
Val-Ser_a
Val-Ser_a
Val-Ser_a
Val-Ser_a
Val-Ser_a
MFCD09953737_b

MFCD09953737_b
MFCD09953737_b
MFCD09953737_b
MFCD09953737_b
2-Methoxy-3,4,13-trimethyl-12-oxo-11,14-dioxatetracyclo[8.3.1.0~1,10~.0~3,8~]tetradec-5-yl (2E)-3-(methylsul[illegible]
2-Methoxy-3,4,13-trimethyl-12-oxo-11,14-dioxatetracyclo[8.3.1.0~1,10~.0~3,8~]tetradec-5-yl (2E)-3-(methylsul[illegible]
2-Methoxy-3,4,13-trimethyl-12-oxo-11,14-dioxatetracyclo[8.3.1.0~1,10~.0~3,8~]tetradec-5-yl (2E)-3-(methylsul[illegible]
2-Methoxy-3,4,13-trimethyl-12-oxo-11,14-dioxatetracyclo[8.3.1.0~1,10~.0~3,8~]tetradec-5-yl (2E)-3-(methylsul[illegible]
2-Methoxy-3,4,13-trimethyl-12-oxo-11,14-dioxatetracyclo[8.3.1.0~1,10~.0~3,8~]tetradec-5-yl (2E)-3-(methylsul[illegible]
2-Methylbutyrylglycine
2-Methylbutyrylglycine
2-Methylbutyrylglycine
2-Methylbutyrylglycine
2-Methylbutyrylglycine
S-Methyl-1-thio-D-glycerate
S-Methyl-1-thio-D-glycerate
S-Methyl-1-thio-D-glycerate
S-Methyl-1-thio-D-glycerate
S-Methyl-1-thio-D-glycerate
Yangonin
Yangonin
Yangonin
Yangonin
Yangonin
Zalcitabine_c
Zalcitabine_c
Zalcitabine_c
Zalcitabine_c
Zalcitabine_c
alpha-ketoadipic acid
alpha-ketoadipic acid
alpha-ketoadipic acid
alpha-ketoadipic acid
alpha-ketoadipic acid
N-Acetylaspartic acid
N-Acetylaspartic acid
N-Acetylaspartic acid
N-Acetylaspartic acid
N-Acetylaspartic acid
leu-gln_b
leu-gln_b
leu-gln_b
leu-gln_b
leu-gln_b
Nisinic acid_b
Nisinic acid_b
Nisinic acid_b
Nisinic acid_b

Nisinic acid_b
delta-Guanidinovalericacid
delta-Guanidinovalericacid
delta-Guanidinovalericacid
delta-Guanidinovalericacid
delta-Guanidinovalericacid
4-(Nitrosoamino)-1-(3-pyridinyl)-1-butanol_a
4-(Nitrosoamino)-1-(3-pyridinyl)-1-butanol_a
4-(Nitrosoamino)-1-(3-pyridinyl)-1-butanol_a
4-(Nitrosoamino)-1-(3-pyridinyl)-1-butanol_a
4-(Nitrosoamino)-1-(3-pyridinyl)-1-butanol_a
Tetrahydropapaveroline_a
Tetrahydropapaveroline_a
Tetrahydropapaveroline_a
Tetrahydropapaveroline_a
Tetrahydropapaveroline_a
Leucylasparagine_b
Leucylasparagine_b
Leucylasparagine_b
Leucylasparagine_b
Leucylasparagine_b
lys-tyr_a
lys-tyr_a
lys-tyr_a
lys-tyr_a
lys-tyr_a
Glu-Gly
Glu-Gly
Glu-Gly
Glu-Gly
Glu-Gly
His-pro_a
His-pro_a
His-pro_a
His-pro_a
His-pro_a
[3-({3-[(Cyclopropylmethyl)amino]-3-oxetanyl}methyl)-1,2-oxazol-5-yl]methanol
[3-({3-[(Cyclopropylmethyl)amino]-3-oxetanyl}methyl)-1,2-oxazol-5-yl]methanol
[3-({3-[(Cyclopropylmethyl)amino]-3-oxetanyl}methyl)-1,2-oxazol-5-yl]methanol
[3-({3-[(Cyclopropylmethyl)amino]-3-oxetanyl}methyl)-1,2-oxazol-5-yl]methanol
[3-({3-[(Cyclopropylmethyl)amino]-3-oxetanyl}methyl)-1,2-oxazol-5-yl]methanol
N-(4-Amino-1-carboxybutyl)glutamic acid_a
N-(4-Amino-1-carboxybutyl)glutamic acid_a
N-(4-Amino-1-carboxybutyl)glutamic acid_a
N-(4-Amino-1-carboxybutyl)glutamic acid_a
N-(4-Amino-1-carboxybutyl)glutamic acid_a
D-Alanyl-D-alanine
D-Alanyl-D-alanine

D-Alanyl-D-alanine
D-Alanyl-D-alanine
D-Alanyl-D-alanine
(S)-?-glycerophosphorylethanolamine
(S)-?-glycerophosphorylethanolamine
(S)-?-glycerophosphorylethanolamine
(S)-?-glycerophosphorylethanolamine
(S)-?-glycerophosphorylethanolamine
mesifurane_b
mesifurane_b
mesifurane_b
mesifurane_b
mesifurane_b
9-Methyluric acid_c
9-Methyluric acid_c
9-Methyluric acid_c
9-Methyluric acid_c
9-Methyluric acid_c
asn-pro_b
asn-pro_b
asn-pro_b
asn-pro_b
asn-pro_b
3-Hydroxy-3-[(3-methylbutanoyl)oxy]-4-(trimethylammonio)butanoate
3-Hydroxy-3-[(3-methylbutanoyl)oxy]-4-(trimethylammonio)butanoate
3-Hydroxy-3-[(3-methylbutanoyl)oxy]-4-(trimethylammonio)butanoate
3-Hydroxy-3-[(3-methylbutanoyl)oxy]-4-(trimethylammonio)butanoate
3-Hydroxy-3-[(3-methylbutanoyl)oxy]-4-(trimethylammonio)butanoate
NicotinateD-ribonucleoside
NicotinateD-ribonucleoside
NicotinateD-ribonucleoside
NicotinateD-ribonucleoside
NicotinateD-ribonucleoside
Corticosterone
Corticosterone
Corticosterone
Corticosterone
Corticosterone
mesifurane_a
mesifurane_a
mesifurane_a
mesifurane_a
mesifurane_a
(4R)-4-{[3-(2-Aminoethyl)-1H-indol-5-yl]methyl}-1,3-oxazolidin-2-one
(4R)-4-{[3-(2-Aminoethyl)-1H-indol-5-yl]methyl}-1,3-oxazolidin-2-one
(4R)-4-{[3-(2-Aminoethyl)-1H-indol-5-yl]methyl}-1,3-oxazolidin-2-one
(4R)-4-{[3-(2-Aminoethyl)-1H-indol-5-yl]methyl}-1,3-oxazolidin-2-one
(4R)-4-{[3-(2-Aminoethyl)-1H-indol-5-yl]methyl}-1,3-oxazolidin-2-one

Isoquinoline_a
Isoquinoline_a
Isoquinoline_a
Isoquinoline_a
Isoquinoline_a
Propamocarb
Propamocarb
Propamocarb
Propamocarb
Propamocarb
7alpha-Hydroxy-3-oxochol-4-en-24-oic acid_b
7alpha-Hydroxy-3-oxochol-4-en-24-oic acid_b
7alpha-Hydroxy-3-oxochol-4-en-24-oic acid_b
7alpha-Hydroxy-3-oxochol-4-en-24-oic acid_b
7alpha-Hydroxy-3-oxochol-4-en-24-oic acid_b
Leu-Leu_d
Leu-Leu_d
Leu-Leu_d
Leu-Leu_d
Leu-Leu_d
Bis-D-fructose 2',1:2,1'-dianhydride_b
Bis-D-fructose 2',1:2,1'-dianhydride_b
Bis-D-fructose 2',1:2,1'-dianhydride_b
Bis-D-fructose 2',1:2,1'-dianhydride_b
Bis-D-fructose 2',1:2,1'-dianhydride_b
Zalcitabine_e
Zalcitabine_e
Zalcitabine_e
Zalcitabine_e
Zalcitabine_e
trans-Zeatin
trans-Zeatin
trans-Zeatin
trans-Zeatin
trans-Zeatin
Oxprenolol
Oxprenolol
Oxprenolol
Oxprenolol
Oxprenolol
TDP-2_b
TDP-2_b
TDP-2_b
TDP-2_b
TDP-2_b
L-gamma-Glutamyl-L-valine_e
L-gamma-Glutamyl-L-valine_e
L-gamma-Glutamyl-L-valine_e

L-gamma-Glutamyl-L-valine_e
L-gamma-Glutamyl-L-valine_e
N,N-dimethyl-9H-purin-6-amine
N,N-dimethyl-9H-purin-6-amine
N,N-dimethyl-9H-purin-6-amine
N,N-dimethyl-9H-purin-6-amine
N,N-dimethyl-9H-purin-6-amine
MFCD18695608_c
MFCD18695608_c
MFCD18695608_c
MFCD18695608_c
MFCD18695608_c
Caffeic acid
Caffeic acid
Caffeic acid
Caffeic acid
Caffeic acid
Phloionolic acid
Phloionolic acid
Phloionolic acid
Phloionolic acid
Phloionolic acid
Isophthalic acid
Isophthalic acid
Isophthalic acid
Isophthalic acid
Isophthalic acid
Flemichapparin B
Flemichapparin B
Flemichapparin B
Flemichapparin B
Flemichapparin B
Asparaginyl-4-hydroxyproline_b
Asparaginyl-4-hydroxyproline_b
Asparaginyl-4-hydroxyproline_b
Asparaginyl-4-hydroxyproline_b
Asparaginyl-4-hydroxyproline_b
Glycylproline
Glycylproline
Glycylproline
Glycylproline
Glycylproline
Butenylcarnitine
Butenylcarnitine
Butenylcarnitine
Butenylcarnitine
Butenylcarnitine
KYNURAMINE

KYNURAMINE
KYNURAMINE
KYNURAMINE
KYNURAMINE
Spermic acid_c
Spermic acid_c
Spermic acid_c
Spermic acid_c
Spermic acid_c
4-(2,5-Difluorophenyl)-5-(3-isopropyl-[1,2,4]triazolo[4,3-a]pyridin-6-yl)oxazole
4-(2,5-Difluorophenyl)-5-(3-isopropyl-[1,2,4]triazolo[4,3-a]pyridin-6-yl)oxazole
4-(2,5-Difluorophenyl)-5-(3-isopropyl-[1,2,4]triazolo[4,3-a]pyridin-6-yl)oxazole
4-(2,5-Difluorophenyl)-5-(3-isopropyl-[1,2,4]triazolo[4,3-a]pyridin-6-yl)oxazole
4-(2,5-Difluorophenyl)-5-(3-isopropyl-[1,2,4]triazolo[4,3-a]pyridin-6-yl)oxazole
butyrin
butyrin
butyrin
butyrin
butyrin
2-Acetamidohexanedioic acid_b
2-Acetamidohexanedioic acid_b
2-Acetamidohexanedioic acid_b
2-Acetamidohexanedioic acid_b
2-Acetamidohexanedioic acid_b
3,8,9-trihydroxy-10-propyl-3,4,5,8,9,10-hexahydro-2H-oxecin-2-one_b
3,8,9-trihydroxy-10-propyl-3,4,5,8,9,10-hexahydro-2H-oxecin-2-one_b
3,8,9-trihydroxy-10-propyl-3,4,5,8,9,10-hexahydro-2H-oxecin-2-one_b
3,8,9-trihydroxy-10-propyl-3,4,5,8,9,10-hexahydro-2H-oxecin-2-one_b
3,8,9-trihydroxy-10-propyl-3,4,5,8,9,10-hexahydro-2H-oxecin-2-one_b
8-Amino-7-oxononanoic acid_a
8-Amino-7-oxononanoic acid_a
8-Amino-7-oxononanoic acid_a
8-Amino-7-oxononanoic acid_a
8-Amino-7-oxononanoic acid_a
4-Hydroxyprolylleucine_c
4-Hydroxyprolylleucine_c
4-Hydroxyprolylleucine_c
4-Hydroxyprolylleucine_c
4-Hydroxyprolylleucine_c
DNOP_d
DNOP_d
DNOP_d
DNOP_d
DNOP_d
8-Amino-7-oxononanoic acid_b
8-Amino-7-oxononanoic acid_b
8-Amino-7-oxononanoic acid_b
8-Amino-7-oxononanoic acid_b

8-Amino-7-oxononanoic acid_b
N(alpha)-Benzyloxycarbonyl-L-leucine
N(alpha)-Benzyloxycarbonyl-L-leucine
N(alpha)-Benzyloxycarbonyl-L-leucine
N(alpha)-Benzyloxycarbonyl-L-leucine
N(alpha)-Benzyloxycarbonyl-L-leucine
MFCD00025555_c
MFCD00025555_c
MFCD00025555_c
MFCD00025555_c
MFCD00025555_c
APM_b
APM_b
APM_b
APM_b
APM_b
Caffeic acid 3-glucoside
Caffeic acid 3-glucoside
Caffeic acid 3-glucoside
Caffeic acid 3-glucoside
Caffeic acid 3-glucoside
glu-pro
glu-pro
glu-pro
glu-pro
glu-pro
Leu-Val_b
Leu-Val_b
Leu-Val_b
Leu-Val_b
Leu-Val_b
2-Acetamidohexanedioic acid_f
2-Acetamidohexanedioic acid_f
2-Acetamidohexanedioic acid_f
2-Acetamidohexanedioic acid_f
2-Acetamidohexanedioic acid_f
MFCD28369208
MFCD28369208
MFCD28369208
MFCD28369208
MFCD28369208
L-Homocysteic acid
L-Homocysteic acid
L-Homocysteic acid
L-Homocysteic acid
L-Homocysteic acid
Gly-Lys
Gly-Lys

Gly-Lys
Gly-Lys
Gly-Lys
Asparaginyl-4-hydroxyproline_a
Asparaginyl-4-hydroxyproline_a
Asparaginyl-4-hydroxyproline_a
Asparaginyl-4-hydroxyproline_a
Asparaginyl-4-hydroxyproline_a
N-{4-[(2R,3R)-3-(Hydroxymethyl)-4-methyl-5-oxo-2-morpholinyl]phenyl}acetamide
N-{4-[(2R,3R)-3-(Hydroxymethyl)-4-methyl-5-oxo-2-morpholinyl]phenyl}acetamide
N-{4-[(2R,3R)-3-(Hydroxymethyl)-4-methyl-5-oxo-2-morpholinyl]phenyl}acetamide
N-{4-[(2R,3R)-3-(Hydroxymethyl)-4-methyl-5-oxo-2-morpholinyl]phenyl}acetamide
N-{4-[(2R,3R)-3-(Hydroxymethyl)-4-methyl-5-oxo-2-morpholinyl]phenyl}acetamide
Methanesulfonic acid
Methanesulfonic acid
Methanesulfonic acid
Methanesulfonic acid
Methanesulfonic acid
3-(1-hydroxyethyl)-2,3,6,7,8,8a-hexahydropyrrolo[1,2-a]pyrazine-1,4-dione_b
3-(1-hydroxyethyl)-2,3,6,7,8,8a-hexahydropyrrolo[1,2-a]pyrazine-1,4-dione_b
3-(1-hydroxyethyl)-2,3,6,7,8,8a-hexahydropyrrolo[1,2-a]pyrazine-1,4-dione_b
3-(1-hydroxyethyl)-2,3,6,7,8,8a-hexahydropyrrolo[1,2-a]pyrazine-1,4-dione_b
3-(1-hydroxyethyl)-2,3,6,7,8,8a-hexahydropyrrolo[1,2-a]pyrazine-1,4-dione_b
Leupeptin
Leupeptin
Leupeptin
Leupeptin
Leupeptin
4-Acetamidobutanoic acid
4-Acetamidobutanoic acid
4-Acetamidobutanoic acid
4-Acetamidobutanoic acid
4-Acetamidobutanoic acid
Panthenol_a
Panthenol_a
Panthenol_a
Panthenol_a
Panthenol_a
Methyl 1-hydroxy-7-(hydroxymethyl)-1,4a,5,7a-tetrahydrocyclopenta[c]pyran-4-carboxylate_b
Methyl 1-hydroxy-7-(hydroxymethyl)-1,4a,5,7a-tetrahydrocyclopenta[c]pyran-4-carboxylate_b
Methyl 1-hydroxy-7-(hydroxymethyl)-1,4a,5,7a-tetrahydrocyclopenta[c]pyran-4-carboxylate_b
Methyl 1-hydroxy-7-(hydroxymethyl)-1,4a,5,7a-tetrahydrocyclopenta[c]pyran-4-carboxylate_b
Methyl 1-hydroxy-7-(hydroxymethyl)-1,4a,5,7a-tetrahydrocyclopenta[c]pyran-4-carboxylate_b
2,6-Dimethylpyrazine
2,6-Dimethylpyrazine
2,6-Dimethylpyrazine
2,6-Dimethylpyrazine
2,6-Dimethylpyrazine

3-hydroxy-3-methyloxindole_b
3-hydroxy-3-methyloxindole_b
3-hydroxy-3-methyloxindole_b
3-hydroxy-3-methyloxindole_b
3-hydroxy-3-methyloxindole_b
MFCD00142316
MFCD00142316
MFCD00142316
MFCD00142316
MFCD00142316
Leu-Val_d
Leu-Val_d
Leu-Val_d
Leu-Val_d
Leu-Val_d
Spermic acid_d
Spermic acid_d
Spermic acid_d
Spermic acid_d
Spermic acid_d
Lys-Pro_b
Lys-Pro_b
Lys-Pro_b
Lys-Pro_b
Lys-Pro_b
quinol sulfate_b
quinol sulfate_b
quinol sulfate_b
quinol sulfate_b
quinol sulfate_b
N-Butyryl-L-homoserinelactone
N-Butyryl-L-homoserinelactone
N-Butyryl-L-homoserinelactone
N-Butyryl-L-homoserinelactone
N-Butyryl-L-homoserinelactone
4-(METHYLNITROSAMINO)-1-(3-PYRIDYL-N-OXIDE)-1-BUTANOL_c
4-(METHYLNITROSAMINO)-1-(3-PYRIDYL-N-OXIDE)-1-BUTANOL_c
4-(METHYLNITROSAMINO)-1-(3-PYRIDYL-N-OXIDE)-1-BUTANOL_c
4-(METHYLNITROSAMINO)-1-(3-PYRIDYL-N-OXIDE)-1-BUTANOL_c
4-(METHYLNITROSAMINO)-1-(3-PYRIDYL-N-OXIDE)-1-BUTANOL_c
2-Isopropylmalic acid
2-Isopropylmalic acid
2-Isopropylmalic acid
2-Isopropylmalic acid
2-Isopropylmalic acid
Glycitein
Glycitein
Glycitein

Glycitein
Glycitein
Homocitrulline
Homocitrulline
Homocitrulline
Homocitrulline
Homocitrulline
(S)-3-sulfolactic acid
(S)-3-sulfolactic acid
(S)-3-sulfolactic acid
(S)-3-sulfolactic acid
(S)-3-sulfolactic acid
Hydroxycarteolol_a
Hydroxycarteolol_a
Hydroxycarteolol_a
Hydroxycarteolol_a
Hydroxycarteolol_a
Valylvaline_h
Valylvaline_h
Valylvaline_h
Valylvaline_h
Valylvaline_h
Leucyltyrosine_b
Leucyltyrosine_b
Leucyltyrosine_b
Leucyltyrosine_b
Leucyltyrosine_b
L-gamma-Glutamyl-L-valine_b
L-gamma-Glutamyl-L-valine_b
L-gamma-Glutamyl-L-valine_b
L-gamma-Glutamyl-L-valine_b
L-gamma-Glutamyl-L-valine_b
7-Hydroxy-2,11,11-trimethyl-3-oxotricyclo[4.3.2.01,5]undecane-9-carboxylic acid
7-Hydroxy-2,11,11-trimethyl-3-oxotricyclo[4.3.2.01,5]undecane-9-carboxylic acid
7-Hydroxy-2,11,11-trimethyl-3-oxotricyclo[4.3.2.01,5]undecane-9-carboxylic acid
7-Hydroxy-2,11,11-trimethyl-3-oxotricyclo[4.3.2.01,5]undecane-9-carboxylic acid
7-Hydroxy-2,11,11-trimethyl-3-oxotricyclo[4.3.2.01,5]undecane-9-carboxylic acid
Scymnol
Scymnol
Scymnol
Scymnol
Scymnol
Bromazine
Bromazine
Bromazine
Bromazine
Bromazine
4-Methylcatechol

4-Methylcatechol
4-Methylcatechol
4-Methylcatechol
4-Methylcatechol
entecavir
entecavir
entecavir
entecavir
entecavir
N6-METHYLLYSINE_a
N6-METHYLLYSINE_a
N6-METHYLLYSINE_a
N6-METHYLLYSINE_a
N6-METHYLLYSINE_a
feruloylagmatine
feruloylagmatine
feruloylagmatine
feruloylagmatine
feruloylagmatine
Tetraacetylethylenediamine_b
Tetraacetylethylenediamine_b
Tetraacetylethylenediamine_b
Tetraacetylethylenediamine_b
Tetraacetylethylenediamine_b
LW8000000_c
LW8000000_c
LW8000000_c
LW8000000_c
LW8000000_c
3-Hydroxy-3-(2,3,4-trimethoxyphenyl)propanoic acid
3-Hydroxy-3-(2,3,4-trimethoxyphenyl)propanoic acid
3-Hydroxy-3-(2,3,4-trimethoxyphenyl)propanoic acid
3-Hydroxy-3-(2,3,4-trimethoxyphenyl)propanoic acid
3-Hydroxy-3-(2,3,4-trimethoxyphenyl)propanoic acid
MFCD00870462
MFCD00870462
MFCD00870462
MFCD00870462
MFCD00870462
7-Methylxanthine
7-Methylxanthine
7-Methylxanthine
7-Methylxanthine
7-Methylxanthine
Diethylpyrocarbonate_b
Diethylpyrocarbonate_b
Diethylpyrocarbonate_b
Diethylpyrocarbonate_b

Diethylpyrocarbonate_b
1-PYRENYL B-D-GLUCURONIDE
1-PYRENYL B-D-GLUCURONIDE
1-PYRENYL B-D-GLUCURONIDE
1-PYRENYL B-D-GLUCURONIDE
1-PYRENYL B-D-GLUCURONIDE
N-(4-Hydroxy-3,5-dimethoxybenzoyl)glycine_d
N-(4-Hydroxy-3,5-dimethoxybenzoyl)glycine_d
N-(4-Hydroxy-3,5-dimethoxybenzoyl)glycine_d
N-(4-Hydroxy-3,5-dimethoxybenzoyl)glycine_d
N-(4-Hydroxy-3,5-dimethoxybenzoyl)glycine_d
2-Hydroxybutyric acid
2-Hydroxybutyric acid
2-Hydroxybutyric acid
2-Hydroxybutyric acid
2-Hydroxybutyric acid
Sinapyl alcohol
Sinapyl alcohol
Sinapyl alcohol
Sinapyl alcohol
Sinapyl alcohol
MFCD00153469
MFCD00153469
MFCD00153469
MFCD00153469
MFCD00153469
N~6~-Octanoyllysine_a
N~6~-Octanoyllysine_a
N~6~-Octanoyllysine_a
N~6~-Octanoyllysine_a
N~6~-Octanoyllysine_a
pro-gln_a
pro-gln_a
pro-gln_a
pro-gln_a
pro-gln_a
(9cis)-O~15~-[(2S,3R,4R,5S,6R)-6-Carboxy-3,4,5-trihydroxytetrahydro-2H-pyran-2-yl]-2-oxoretinoic acid
(9cis)-O~15~-[(2S,3R,4R,5S,6R)-6-Carboxy-3,4,5-trihydroxytetrahydro-2H-pyran-2-yl]-2-oxoretinoic acid
(9cis)-O~15~-[(2S,3R,4R,5S,6R)-6-Carboxy-3,4,5-trihydroxytetrahydro-2H-pyran-2-yl]-2-oxoretinoic acid
(9cis)-O~15~-[(2S,3R,4R,5S,6R)-6-Carboxy-3,4,5-trihydroxytetrahydro-2H-pyran-2-yl]-2-oxoretinoic acid
(9cis)-O~15~-[(2S,3R,4R,5S,6R)-6-Carboxy-3,4,5-trihydroxytetrahydro-2H-pyran-2-yl]-2-oxoretinoic acid
1,1'-[1,12-Dodecanediylbis(oxy)]dibenzene_d
1,1'-[1,12-Dodecanediylbis(oxy)]dibenzene_d
1,1'-[1,12-Dodecanediylbis(oxy)]dibenzene_d
1,1'-[1,12-Dodecanediylbis(oxy)]dibenzene_d
1,1'-[1,12-Dodecanediylbis(oxy)]dibenzene_d
2-Hydroxy-4-(methylthio)butanoic acid
2-Hydroxy-4-(methylthio)butanoic acid

2-Hydroxy-4-(methylthio)butanoic acid
2-Hydroxy-4-(methylthio)butanoic acid
2-Hydroxy-4-(methylthio)butanoic acid
N-Benzoylaspartic acid_b
N-Benzoylaspartic acid_b
N-Benzoylaspartic acid_b
N-Benzoylaspartic acid_b
N-Benzoylaspartic acid_b
Homoanserine_d
Homoanserine_d
Homoanserine_d
Homoanserine_d
Homoanserine_d
Primaquine
Primaquine
Primaquine
Primaquine
Primaquine
1-(3,4-dimethoxyphenyl)ethan-1-one oxime
1-(3,4-dimethoxyphenyl)ethan-1-one oxime
1-(3,4-dimethoxyphenyl)ethan-1-one oxime
1-(3,4-dimethoxyphenyl)ethan-1-one oxime
1-(3,4-dimethoxyphenyl)ethan-1-one oxime
asn-lys
asn-lys
asn-lys
asn-lys
asn-lys
Leu-Val_c
Leu-Val_c
Leu-Val_c
Leu-Val_c
Leu-Val_c
Tiglic acid_b
Tiglic acid_b
Tiglic acid_b
Tiglic acid_b
Tiglic acid_b
tert-Butyl 3-amino-1-methyl-2,3-dioxopropylcarbamate_d
tert-Butyl 3-amino-1-methyl-2,3-dioxopropylcarbamate_d
tert-Butyl 3-amino-1-methyl-2,3-dioxopropylcarbamate_d
tert-Butyl 3-amino-1-methyl-2,3-dioxopropylcarbamate_d
tert-Butyl 3-amino-1-methyl-2,3-dioxopropylcarbamate_d
Methylol Dimethylhydantoin_c
Methylol Dimethylhydantoin_c
Methylol Dimethylhydantoin_c
Methylol Dimethylhydantoin_c
Methylol Dimethylhydantoin_c

4-(4-Deoxy-alpha-D-gluc-4-enuronosyl)-D-galacturonate
4-(4-Deoxy-alpha-D-gluc-4-enuronosyl)-D-galacturonate
4-(4-Deoxy-alpha-D-gluc-4-enuronosyl)-D-galacturonate
4-(4-Deoxy-alpha-D-gluc-4-enuronosyl)-D-galacturonate
4-(4-Deoxy-alpha-D-gluc-4-enuronosyl)-D-galacturonate
tert-Butyl 3-amino-1-methyl-2,3-dioxopropylcarbamate_e
tert-Butyl 3-amino-1-methyl-2,3-dioxopropylcarbamate_e
tert-Butyl 3-amino-1-methyl-2,3-dioxopropylcarbamate_e
tert-Butyl 3-amino-1-methyl-2,3-dioxopropylcarbamate_e
tert-Butyl 3-amino-1-methyl-2,3-dioxopropylcarbamate_e
Menadiol
Menadiol
Menadiol
Menadiol
Menadiol
N-Acetyl-5-oxo-L-norvaline_b
N-Acetyl-5-oxo-L-norvaline_b
N-Acetyl-5-oxo-L-norvaline_b
N-Acetyl-5-oxo-L-norvaline_b
N-Acetyl-5-oxo-L-norvaline_b
Nicotine glucuronide
Nicotine glucuronide
Nicotine glucuronide
Nicotine glucuronide
Nicotine glucuronide
pentobarbital_f
pentobarbital_f
pentobarbital_f
pentobarbital_f
pentobarbital_f
Agomelatine
Agomelatine
Agomelatine
Agomelatine
Agomelatine
3,4-Dimethoxyphenylacetic acid
3,4-Dimethoxyphenylacetic acid
3,4-Dimethoxyphenylacetic acid
3,4-Dimethoxyphenylacetic acid
3,4-Dimethoxyphenylacetic acid
Pirbuterol
Pirbuterol
Pirbuterol
Pirbuterol
Pirbuterol
Homocysteine thiolactone
Homocysteine thiolactone
Homocysteine thiolactone

Homocysteine thiolactone
Homocysteine thiolactone
Esculin
Esculin
Esculin
Esculin
Esculin
Ectoine
Ectoine
Ectoine
Ectoine
Ectoine
Arg-pro
Arg-pro
Arg-pro
Arg-pro
Arg-pro
Alanyltryptophan
Alanyltryptophan
Alanyltryptophan
Alanyltryptophan
Alanyltryptophan
3-Methoxy-4-hydroxyhippuric acid_b
3-Methoxy-4-hydroxyhippuric acid_b
3-Methoxy-4-hydroxyhippuric acid_b
3-Methoxy-4-hydroxyhippuric acid_b
3-Methoxy-4-hydroxyhippuric acid_b
NSC 92778
NSC 92778
NSC 92778
NSC 92778
NSC 92778
SECONAL_a
SECONAL_a
SECONAL_a
SECONAL_a
SECONAL_a
(1R,3R,5R)-2-[(2S)-2-Amino-2-(3,5-dihydroxyadamantan-1-yl)acetyl]-2-azabicyclo[3.1.0]hexane-3-carbonitrile
(1R,3R,5R)-2-[(2S)-2-Amino-2-(3,5-dihydroxyadamantan-1-yl)acetyl]-2-azabicyclo[3.1.0]hexane-3-carbonitrile
(1R,3R,5R)-2-[(2S)-2-Amino-2-(3,5-dihydroxyadamantan-1-yl)acetyl]-2-azabicyclo[3.1.0]hexane-3-carbonitrile
(1R,3R,5R)-2-[(2S)-2-Amino-2-(3,5-dihydroxyadamantan-1-yl)acetyl]-2-azabicyclo[3.1.0]hexane-3-carbonitrile
(1R,3R,5R)-2-[(2S)-2-Amino-2-(3,5-dihydroxyadamantan-1-yl)acetyl]-2-azabicyclo[3.1.0]hexane-3-carbonitrile
2-[(carboxymethyl)methylamino]-5-methoxy-Benzoic acid_a
2-[(carboxymethyl)methylamino]-5-methoxy-Benzoic acid_a
2-[(carboxymethyl)methylamino]-5-methoxy-Benzoic acid_a
2-[(carboxymethyl)methylamino]-5-methoxy-Benzoic acid_a
2-[(carboxymethyl)methylamino]-5-methoxy-Benzoic acid_a
Zalcitabine_f

Zalcitabine_f
Zalcitabine_f
Zalcitabine_f
Zalcitabine_f
4-Hydroxyprolylleucine_a
4-Hydroxyprolylleucine_a
4-Hydroxyprolylleucine_a
4-Hydroxyprolylleucine_a
4-Hydroxyprolylleucine_a
N-Ethylpropanamide_b
N-Ethylpropanamide_b
N-Ethylpropanamide_b
N-Ethylpropanamide_b
N-Ethylpropanamide_b
Paraldehyde
Paraldehyde
Paraldehyde
Paraldehyde
Paraldehyde
pimethixene
pimethixene
pimethixene
pimethixene
pimethixene
Octyl benzoate
Octyl benzoate
Octyl benzoate
Octyl benzoate
Octyl benzoate
7Î±-Hydroxytestosterone
7Î±-Hydroxytestosterone
7Î±-Hydroxytestosterone
7Î±-Hydroxytestosterone
7Î±-Hydroxytestosterone
Allyl mercaptan
Allyl mercaptan
Allyl mercaptan
Allyl mercaptan
Allyl mercaptan
N-Pentanoylphenylalanine_a
N-Pentanoylphenylalanine_a
N-Pentanoylphenylalanine_a
N-Pentanoylphenylalanine_a
N-Pentanoylphenylalanine_a
Piperidine_c
Piperidine_c
Piperidine_c
Piperidine_c

Piperidine_c
6-Hydroxypentadecanedioic acid
6-Hydroxypentadecanedioic acid
6-Hydroxypentadecanedioic acid
6-Hydroxypentadecanedioic acid
6-Hydroxypentadecanedioic acid
2-Methylthiazolidine_a
2-Methylthiazolidine_a
2-Methylthiazolidine_a
2-Methylthiazolidine_a
2-Methylthiazolidine_a
O-succinylcarnitine
O-succinylcarnitine
O-succinylcarnitine
O-succinylcarnitine
O-succinylcarnitine
Valylproline
Valylproline
Valylproline
Valylproline
Valylproline
Leu-Leu_c
Leu-Leu_c
Leu-Leu_c
Leu-Leu_c
Leu-Leu_c
asn-val_a
asn-val_a
asn-val_a
asn-val_a
asn-val_a
Hyodeoxycholic acid
Hyodeoxycholic acid
Hyodeoxycholic acid
Hyodeoxycholic acid
Hyodeoxycholic acid
N-Pentanoylphenylalanine_b
N-Pentanoylphenylalanine_b
N-Pentanoylphenylalanine_b
N-Pentanoylphenylalanine_b
N-Pentanoylphenylalanine_b
N-LACTOYL ETHANOLAMINE PHOSPHATE
N-LACTOYL ETHANOLAMINE PHOSPHATE
N-LACTOYL ETHANOLAMINE PHOSPHATE
N-LACTOYL ETHANOLAMINE PHOSPHATE
N-LACTOYL ETHANOLAMINE PHOSPHATE
heptabarbital
heptabarbital

heptabarbital
heptabarbital
heptabarbital
Pregabalin
Pregabalin
Pregabalin
Pregabalin
Pregabalin
Dibutyl malate
Dibutyl malate
Dibutyl malate
Dibutyl malate
Dibutyl malate
Lysylvaline_b
Lysylvaline_b
Lysylvaline_b
Lysylvaline_b
Lysylvaline_b
MFCD02728197_b
MFCD02728197_b
MFCD02728197_b
MFCD02728197_b
MFCD02728197_b
Sulfoacetate
Sulfoacetate
Sulfoacetate
Sulfoacetate
Sulfoacetate
Zalcitabine_a
Zalcitabine_a
Zalcitabine_a
Zalcitabine_a
Zalcitabine_a
Leu-Val_f
Leu-Val_f
Leu-Val_f
Leu-Val_f
Leu-Val_f
N~5~-[P-Amino-P-(sulfoamino)phosphorimidoyl]-L-ornithyl-L-alanyl-L-arginine
N~5~-[P-Amino-P-(sulfoamino)phosphorimidoyl]-L-ornithyl-L-alanyl-L-arginine
N~5~-[P-Amino-P-(sulfoamino)phosphorimidoyl]-L-ornithyl-L-alanyl-L-arginine
N~5~-[P-Amino-P-(sulfoamino)phosphorimidoyl]-L-ornithyl-L-alanyl-L-arginine
N~5~-[P-Amino-P-(sulfoamino)phosphorimidoyl]-L-ornithyl-L-alanyl-L-arginine
4-Phenolsulfonic acid
4-Phenolsulfonic acid
4-Phenolsulfonic acid
4-Phenolsulfonic acid
4-Phenolsulfonic acid

hexobarbital_d
hexobarbital_d
hexobarbital_d
hexobarbital_d
hexobarbital_d
N,N-Diethyl-4-methyl-1-piperazinecarboxamide 4-oxide_c
N,N-Diethyl-4-methyl-1-piperazinecarboxamide 4-oxide_c
N,N-Diethyl-4-methyl-1-piperazinecarboxamide 4-oxide_c
N,N-Diethyl-4-methyl-1-piperazinecarboxamide 4-oxide_c
N,N-Diethyl-4-methyl-1-piperazinecarboxamide 4-oxide_c
meprobamate_e
meprobamate_e
meprobamate_e
meprobamate_e
meprobamate_e
Naphthalen-2-amine
Naphthalen-2-amine
Naphthalen-2-amine
Naphthalen-2-amine
Naphthalen-2-amine
N-Acetyl-5-oxo-L-norvaline_a
N-Acetyl-5-oxo-L-norvaline_a
N-Acetyl-5-oxo-L-norvaline_a
N-Acetyl-5-oxo-L-norvaline_a
N-Acetyl-5-oxo-L-norvaline_a
pro-gln_b
pro-gln_b
pro-gln_b
pro-gln_b
pro-gln_b
L-(+)-Erythrulose_b
L-(+)-Erythrulose_b
L-(+)-Erythrulose_b
L-(+)-Erythrulose_b
L-(+)-Erythrulose_b
N-(1-Methyl-4-oxo-4,5-dihydro-1H-imidazol-2-yl)alanine_a
N-(1-Methyl-4-oxo-4,5-dihydro-1H-imidazol-2-yl)alanine_a
N-(1-Methyl-4-oxo-4,5-dihydro-1H-imidazol-2-yl)alanine_a
N-(1-Methyl-4-oxo-4,5-dihydro-1H-imidazol-2-yl)alanine_a
N-(1-Methyl-4-oxo-4,5-dihydro-1H-imidazol-2-yl)alanine_a
2,4-Quinolinediol_a
2,4-Quinolinediol_a
2,4-Quinolinediol_a
2,4-Quinolinediol_a
2,4-Quinolinediol_a
pentobarbital_a
pentobarbital_a
pentobarbital_a

pentobarbital_a
pentobarbital_a
3,3-Dimethylglutaric acid_a
3,3-Dimethylglutaric acid_a
3,3-Dimethylglutaric acid_a
3,3-Dimethylglutaric acid_a
3,3-Dimethylglutaric acid_a
Glycylprolylhydroxyproline_b
Glycylprolylhydroxyproline_b
Glycylprolylhydroxyproline_b
Glycylprolylhydroxyproline_b
Glycylprolylhydroxyproline_b
threonylphenylalanine_a
threonylphenylalanine_a
threonylphenylalanine_a
threonylphenylalanine_a
threonylphenylalanine_a
2-methylcitric acid
2-methylcitric acid
2-methylcitric acid
2-methylcitric acid
2-methylcitric acid
Leucylasparagine_c
Leucylasparagine_c
Leucylasparagine_c
Leucylasparagine_c
Leucylasparagine_c
(7E,7'E)-5,5'-diferulic acid_a
(7E,7'E)-5,5'-diferulic acid_a
(7E,7'E)-5,5'-diferulic acid_a
(7E,7'E)-5,5'-diferulic acid_a
(7E,7'E)-5,5'-diferulic acid_a
3,3-Dimethylglutaric acid_b
3,3-Dimethylglutaric acid_b
3,3-Dimethylglutaric acid_b
3,3-Dimethylglutaric acid_b
3,3-Dimethylglutaric acid_b
(3aR,4R,5R,6aS)-5-Hydroxy-4-[(1E,3S)-3-hydroxy-1-octen-1-yl]hexahydro-2H-cyclopenta[b]furan-2-one
(3aR,4R,5R,6aS)-5-Hydroxy-4-[(1E,3S)-3-hydroxy-1-octen-1-yl]hexahydro-2H-cyclopenta[b]furan-2-one
(3aR,4R,5R,6aS)-5-Hydroxy-4-[(1E,3S)-3-hydroxy-1-octen-1-yl]hexahydro-2H-cyclopenta[b]furan-2-one
(3aR,4R,5R,6aS)-5-Hydroxy-4-[(1E,3S)-3-hydroxy-1-octen-1-yl]hexahydro-2H-cyclopenta[b]furan-2-one
(3aR,4R,5R,6aS)-5-Hydroxy-4-[(1E,3S)-3-hydroxy-1-octen-1-yl]hexahydro-2H-cyclopenta[b]furan-2-one
5-(2-Carboxyethyl)-4-hydroxy-6-oxo-5,6-dihydro-2-pyridinecarboxylic acid
5-(2-Carboxyethyl)-4-hydroxy-6-oxo-5,6-dihydro-2-pyridinecarboxylic acid
5-(2-Carboxyethyl)-4-hydroxy-6-oxo-5,6-dihydro-2-pyridinecarboxylic acid
5-(2-Carboxyethyl)-4-hydroxy-6-oxo-5,6-dihydro-2-pyridinecarboxylic acid
5-(2-Carboxyethyl)-4-hydroxy-6-oxo-5,6-dihydro-2-pyridinecarboxylic acid
5-Phospho-beta-D-ribosylamine

5-Phospho-beta-D-ribosylamine
5-Phospho-beta-D-ribosylamine
5-Phospho-beta-D-ribosylamine
5-Phospho-beta-D-ribosylamine
8-Hydroxyhexadecanedioic acid
8-Hydroxyhexadecanedioic acid
8-Hydroxyhexadecanedioic acid
8-Hydroxyhexadecanedioic acid
8-Hydroxyhexadecanedioic acid
2-(Carboxyacetamido)benzoic acid_b
2-(Carboxyacetamido)benzoic acid_b
2-(Carboxyacetamido)benzoic acid_b
2-(Carboxyacetamido)benzoic acid_b
2-(Carboxyacetamido)benzoic acid_b
6-Acetamido-2-oxohexanoic acid
6-Acetamido-2-oxohexanoic acid
6-Acetamido-2-oxohexanoic acid
6-Acetamido-2-oxohexanoic acid
6-Acetamido-2-oxohexanoic acid
Triacetin
Triacetin
Triacetin
Triacetin
Triacetin
Nicotinic acid ribonucleoside
Nicotinic acid ribonucleoside
Nicotinic acid ribonucleoside
Nicotinic acid ribonucleoside
Nicotinic acid ribonucleoside
3,4,15-Trihydroxy-12,13-epoxytrichothec-9-en-8-yl 3-methylbutanoate
3,4,15-Trihydroxy-12,13-epoxytrichothec-9-en-8-yl 3-methylbutanoate
3,4,15-Trihydroxy-12,13-epoxytrichothec-9-en-8-yl 3-methylbutanoate
3,4,15-Trihydroxy-12,13-epoxytrichothec-9-en-8-yl 3-methylbutanoate
3,4,15-Trihydroxy-12,13-epoxytrichothec-9-en-8-yl 3-methylbutanoate
Rivastigmine
Rivastigmine
Rivastigmine
Rivastigmine
Rivastigmine
asn-pro_d
asn-pro_d
asn-pro_d
asn-pro_d
asn-pro_d
Aspartyl-L-proline_a
Aspartyl-L-proline_a
Aspartyl-L-proline_a
Aspartyl-L-proline_a

Aspartyl-L-proline_a
2,2-Bis(hydroxymethyl)propionic acid_b
2,2-Bis(hydroxymethyl)propionic acid_b
2,2-Bis(hydroxymethyl)propionic acid_b
2,2-Bis(hydroxymethyl)propionic acid_b
2,2-Bis(hydroxymethyl)propionic acid_b
Threonylserine
Threonylserine
Threonylserine
Threonylserine
Threonylserine
(âˆ’)-nabilone_b
(âˆ’)-nabilone_b
(âˆ’)-nabilone_b
(âˆ’)-nabilone_b
(âˆ’)-nabilone_b
Phenyl D-glucopyranosiduronic acid_a
Phenyl D-glucopyranosiduronic acid_a
Phenyl D-glucopyranosiduronic acid_a
Phenyl D-glucopyranosiduronic acid_a
Phenyl D-glucopyranosiduronic acid_a
porphobilinogen_a
porphobilinogen_a
porphobilinogen_a
porphobilinogen_a
porphobilinogen_a
Bis-D-fructose 2',1:2,1'-dianhydride_a
Bis-D-fructose 2',1:2,1'-dianhydride_a
Bis-D-fructose 2',1:2,1'-dianhydride_a
Bis-D-fructose 2',1:2,1'-dianhydride_a
Bis-D-fructose 2',1:2,1'-dianhydride_a
Hydroxypropionylcarnitine_a
Hydroxypropionylcarnitine_a
Hydroxypropionylcarnitine_a
Hydroxypropionylcarnitine_a
Hydroxypropionylcarnitine_a
Homoanserine_a
Homoanserine_a
Homoanserine_a
Homoanserine_a
Homoanserine_a
Tranexamic acid
Tranexamic acid
Tranexamic acid
Tranexamic acid
Tranexamic acid
Chenodeoxycholic acid
Chenodeoxycholic acid

Chenodeoxycholic acid
Chenodeoxycholic acid
Chenodeoxycholic acid
2,2-Bis(hydroxymethyl)propionic acid_a
2,2-Bis(hydroxymethyl)propionic acid_a
2,2-Bis(hydroxymethyl)propionic acid_a
2,2-Bis(hydroxymethyl)propionic acid_a
2,2-Bis(hydroxymethyl)propionic acid_a
Î´-Valerolactam_c
Î´-Valerolactam_c
Î´-Valerolactam_c
Î´-Valerolactam_c
Î´-Valerolactam_c
DNOP_h
DNOP_h
DNOP_h
DNOP_h
DNOP_h
Rutinose (beta-anomer)
Rutinose (beta-anomer)
Rutinose (beta-anomer)
Rutinose (beta-anomer)
Rutinose (beta-anomer)
N-Acetylasparagine
N-Acetylasparagine
N-Acetylasparagine
N-Acetylasparagine
N-Acetylasparagine
N6-METHYLLYSINE_b
N6-METHYLLYSINE_b
N6-METHYLLYSINE_b
N6-METHYLLYSINE_b
N6-METHYLLYSINE_b
DNOP_c
DNOP_c
DNOP_c
DNOP_c
DNOP_c
N'-Hydroxy-4-pentylbenzenecarboximidamide
N'-Hydroxy-4-pentylbenzenecarboximidamide
N'-Hydroxy-4-pentylbenzenecarboximidamide
N'-Hydroxy-4-pentylbenzenecarboximidamide
N'-Hydroxy-4-pentylbenzenecarboximidamide
4-pyridoxic acid
4-pyridoxic acid
4-pyridoxic acid
4-pyridoxic acid
4-pyridoxic acid

coenzyme Q2
coenzyme Q2
coenzyme Q2
coenzyme Q2
coenzyme Q2
Butylparaben
Butylparaben
Butylparaben
Butylparaben
Butylparaben
3-(Sulfooxy)butanoic acid
3-(Sulfooxy)butanoic acid
3-(Sulfooxy)butanoic acid
3-(Sulfooxy)butanoic acid
3-(Sulfooxy)butanoic acid
Valylvaline_e
Valylvaline_e
Valylvaline_e
Valylvaline_e
Valylvaline_e
4-Amino-1-piperidinecarboxylic acid
4-Amino-1-piperidinecarboxylic acid
4-Amino-1-piperidinecarboxylic acid
4-Amino-1-piperidinecarboxylic acid
4-Amino-1-piperidinecarboxylic acid
2,4-Quinolinediol_b
2,4-Quinolinediol_b
2,4-Quinolinediol_b
2,4-Quinolinediol_b
2,4-Quinolinediol_b
Glucosamine
Glucosamine
Glucosamine
Glucosamine
Glucosamine
1-pyrroline
1-pyrroline
1-pyrroline
1-pyrroline
1-pyrroline
TO0127900_b
TO0127900_b
TO0127900_b
TO0127900_b
TO0127900_b
1,1'-[1,12-Dodecanediylbis(oxy)]dibenzene_g
1,1'-[1,12-Dodecanediylbis(oxy)]dibenzene_g
1,1'-[1,12-Dodecanediylbis(oxy)]dibenzene_g

1,1'-[1,12-Dodecanediylbis(oxy)]dibenzene_g
1,1'-[1,12-Dodecanediylbis(oxy)]dibenzene_g
2'-Deoxy-5-hydroxymethylcytidine-5'-diphosphate
2'-Deoxy-5-hydroxymethylcytidine-5'-diphosphate
2'-Deoxy-5-hydroxymethylcytidine-5'-diphosphate
2'-Deoxy-5-hydroxymethylcytidine-5'-diphosphate
2'-Deoxy-5-hydroxymethylcytidine-5'-diphosphate
4-Hydroxybenzaldehyde
4-Hydroxybenzaldehyde
4-Hydroxybenzaldehyde
4-Hydroxybenzaldehyde
4-Hydroxybenzaldehyde
Naringenin
Naringenin
Naringenin
Naringenin
Naringenin
MFCD00025555_b
MFCD00025555_b
MFCD00025555_b
MFCD00025555_b
MFCD00025555_b
Oxypeucedanin
Oxypeucedanin
Oxypeucedanin
Oxypeucedanin
Oxypeucedanin
1-Vinylimidazole_a
1-Vinylimidazole_a
1-Vinylimidazole_a
1-Vinylimidazole_a
1-Vinylimidazole_a
4-Acetamidobenzaldehyde
4-Acetamidobenzaldehyde
4-Acetamidobenzaldehyde
4-Acetamidobenzaldehyde
4-Acetamidobenzaldehyde
Procaine_a
Procaine_a
Procaine_a
Procaine_a
Procaine_a
Piperidine_b
Piperidine_b
Piperidine_b
Piperidine_b
Piperidine_b
4-{[(3-Hydroxy-3-methylbutanoyl)oxy]methyl}-1-[(3-methylbutanoyl)oxy]-6,7a-dihydro-1H-spiro[cyclopenta[c]pyr[illegible]

4-{[(3-Hydroxy-3-methylbutanoyl)oxy]methyl}-1-[(3-methylbutanoyl)oxy]-6,7a-dihydro-1H-spiro[cyclopenta[c]pyra
4-{[(3-Hydroxy-3-methylbutanoyl)oxy]methyl}-1-[(3-methylbutanoyl)oxy]-6,7a-dihydro-1H-spiro[cyclopenta[c]pyra
4-{[(3-Hydroxy-3-methylbutanoyl)oxy]methyl}-1-[(3-methylbutanoyl)oxy]-6,7a-dihydro-1H-spiro[cyclopenta[c]pyra
4-{[(3-Hydroxy-3-methylbutanoyl)oxy]methyl}-1-[(3-methylbutanoyl)oxy]-6,7a-dihydro-1H-spiro[cyclopenta[c]pyra
(âˆ’)-nabilone_e
(âˆ’)-nabilone_e
(âˆ’)-nabilone_e
(âˆ’)-nabilone_e
(âˆ’)-nabilone_e
L-(+)-Erythrulose_a
L-(+)-Erythrulose_a
L-(+)-Erythrulose_a
L-(+)-Erythrulose_a
L-(+)-Erythrulose_a
(+/-)-2-Hydroxyglutaric acid_a
(+/-)-2-Hydroxyglutaric acid_a
(+/-)-2-Hydroxyglutaric acid_a
(+/-)-2-Hydroxyglutaric acid_a
(+/-)-2-Hydroxyglutaric acid_a
Pyrrolidine
Pyrrolidine
Pyrrolidine
Pyrrolidine
Pyrrolidine
Ursodeoxycholic acid
Ursodeoxycholic acid
Ursodeoxycholic acid
Ursodeoxycholic acid
Ursodeoxycholic acid
Prolinamide
Prolinamide
Prolinamide
Prolinamide
Prolinamide
1,7-Dimethyluric acid
1,7-Dimethyluric acid
1,7-Dimethyluric acid
1,7-Dimethyluric acid
1,7-Dimethyluric acid
Alanine
Alanine
Alanine
Alanine
Alanine
alpha-Chaconine
alpha-Chaconine
alpha-Chaconine
alpha-Chaconine

alpha-Chaconine
Homoanserine_e
Homoanserine_e
Homoanserine_e
Homoanserine_e
Homoanserine_e
Saccharin
Saccharin
Saccharin
Saccharin
Saccharin
Ethyl sulfate
Ethyl sulfate
Ethyl sulfate
Ethyl sulfate
Ethyl sulfate
Leucylproline
Leucylproline
Leucylproline
Leucylproline
Leucylproline
Xanthine
Xanthine
Xanthine
Xanthine
Xanthine
5-Hydroxyindole
5-Hydroxyindole
5-Hydroxyindole
5-Hydroxyindole
5-Hydroxyindole
5-Allyl-5-sec-butyl-1,3-dimethyl-2,4,6(1H,3H,5H)-pyrimidinetrione_a
5-Allyl-5-sec-butyl-1,3-dimethyl-2,4,6(1H,3H,5H)-pyrimidinetrione_a
5-Allyl-5-sec-butyl-1,3-dimethyl-2,4,6(1H,3H,5H)-pyrimidinetrione_a
5-Allyl-5-sec-butyl-1,3-dimethyl-2,4,6(1H,3H,5H)-pyrimidinetrione_a
5-Allyl-5-sec-butyl-1,3-dimethyl-2,4,6(1H,3H,5H)-pyrimidinetrione_a
8-(3-Furyl)-5-hydroxy-1,1,5a,7a,11b-pentamethyldecahydrooxireno[4,4a]isochromeno[6,5-g][2]benzoxepine-3,1[illegible]
8-(3-Furyl)-5-hydroxy-1,1,5a,7a,11b-pentamethyldecahydrooxireno[4,4a]isochromeno[6,5-g][2]benzoxepine-3,1[illegible]
8-(3-Furyl)-5-hydroxy-1,1,5a,7a,11b-pentamethyldecahydrooxireno[4,4a]isochromeno[6,5-g][2]benzoxepine-3,1[illegible]
8-(3-Furyl)-5-hydroxy-1,1,5a,7a,11b-pentamethyldecahydrooxireno[4,4a]isochromeno[6,5-g][2]benzoxepine-3,1[illegible]
8-(3-Furyl)-5-hydroxy-1,1,5a,7a,11b-pentamethyldecahydrooxireno[4,4a]isochromeno[6,5-g][2]benzoxepine-3,1[illegible]
7-ketodeoxycholic acid_b
7-ketodeoxycholic acid_b
7-ketodeoxycholic acid_b
7-ketodeoxycholic acid_b
7-ketodeoxycholic acid_b
2-[(Sulfooxy)methyl]butanoic acid
2-[(Sulfooxy)methyl]butanoic acid

2-[(Sulfooxy)methyl]butanoic acid
2-[(Sulfooxy)methyl]butanoic acid
2-[(Sulfooxy)methyl]butanoic acid
5-Allyl-5-sec-butyl-1,3-dimethyl-2,4,6(1H,3H,5H)-pyrimidinetrione_b
5-Allyl-5-sec-butyl-1,3-dimethyl-2,4,6(1H,3H,5H)-pyrimidinetrione_b
5-Allyl-5-sec-butyl-1,3-dimethyl-2,4,6(1H,3H,5H)-pyrimidinetrione_b
5-Allyl-5-sec-butyl-1,3-dimethyl-2,4,6(1H,3H,5H)-pyrimidinetrione_b
5-Allyl-5-sec-butyl-1,3-dimethyl-2,4,6(1H,3H,5H)-pyrimidinetrione_b
N-Pentanoylphenylalanine_c
N-Pentanoylphenylalanine_c
N-Pentanoylphenylalanine_c
N-Pentanoylphenylalanine_c
N-Pentanoylphenylalanine_c
N-Acetylvaline_d
N-Acetylvaline_d
N-Acetylvaline_d
N-Acetylvaline_d
N-Acetylvaline_d
4-methylpyridine-3-sulfonic acid
4-methylpyridine-3-sulfonic acid
4-methylpyridine-3-sulfonic acid
4-methylpyridine-3-sulfonic acid
4-methylpyridine-3-sulfonic acid
5-amino-2-(dimethylamino)benzoic acid_a
5-amino-2-(dimethylamino)benzoic acid_a
5-amino-2-(dimethylamino)benzoic acid_a
5-amino-2-(dimethylamino)benzoic acid_a
5-amino-2-(dimethylamino)benzoic acid_a
p-Cresylsulfate
p-Cresylsulfate
p-Cresylsulfate
p-Cresylsulfate
p-Cresylsulfate
Asp-lys
Asp-lys
Asp-lys
Asp-lys
Asp-lys
1-(4-Aminobutyl)urea_a
1-(4-Aminobutyl)urea_a
1-(4-Aminobutyl)urea_a
1-(4-Aminobutyl)urea_a
1-(4-Aminobutyl)urea_a
N-(3-acetamidopropyl)pyrrolidin-2-one_d
N-(3-acetamidopropyl)pyrrolidin-2-one_d
N-(3-acetamidopropyl)pyrrolidin-2-one_d
N-(3-acetamidopropyl)pyrrolidin-2-one_d
N-(3-acetamidopropyl)pyrrolidin-2-one_d

Methionine sulfoxide
Methionine sulfoxide
Methionine sulfoxide
Methionine sulfoxide
Methionine sulfoxide
3,8,9-trihydroxy-10-propyl-3,4,5,8,9,10-hexahydro-2H-oxecin-2-one_a
3,8,9-trihydroxy-10-propyl-3,4,5,8,9,10-hexahydro-2H-oxecin-2-one_a
3,8,9-trihydroxy-10-propyl-3,4,5,8,9,10-hexahydro-2H-oxecin-2-one_a
3,8,9-trihydroxy-10-propyl-3,4,5,8,9,10-hexahydro-2H-oxecin-2-one_a
3,8,9-trihydroxy-10-propyl-3,4,5,8,9,10-hexahydro-2H-oxecin-2-one_a
7-ketodeoxycholic acid_d
7-ketodeoxycholic acid_d
7-ketodeoxycholic acid_d
7-ketodeoxycholic acid_d
7-ketodeoxycholic acid_d
O-propenoyl-D-carnitine
O-propenoyl-D-carnitine
O-propenoyl-D-carnitine
O-propenoyl-D-carnitine
O-propenoyl-D-carnitine
(2Z)-2-({[(6S,7S)-7-Hydroxy-2,3,10,11,12-pentamethoxy-6,7-dimethyl-5,6,7,8-tetrahydrodibenzo[a,c][8]annulen-
(2Z)-2-({[(6S,7S)-7-Hydroxy-2,3,10,11,12-pentamethoxy-6,7-dimethyl-5,6,7,8-tetrahydrodibenzo[a,c][8]annulen-
(2Z)-2-({[(6S,7S)-7-Hydroxy-2,3,10,11,12-pentamethoxy-6,7-dimethyl-5,6,7,8-tetrahydrodibenzo[a,c][8]annulen-
(2Z)-2-({[(6S,7S)-7-Hydroxy-2,3,10,11,12-pentamethoxy-6,7-dimethyl-5,6,7,8-tetrahydrodibenzo[a,c][8]annulen-
(2Z)-2-({[(6S,7S)-7-Hydroxy-2,3,10,11,12-pentamethoxy-6,7-dimethyl-5,6,7,8-tetrahydrodibenzo[a,c][8]annulen-
Genistein
Genistein
Genistein
Genistein
Genistein
NL8513000
NL8513000
NL8513000
NL8513000
NL8513000
Docosahexaenoic acid ethyl ester_a
Docosahexaenoic acid ethyl ester_a
Docosahexaenoic acid ethyl ester_a
Docosahexaenoic acid ethyl ester_a
Docosahexaenoic acid ethyl ester_a
Î´-Gluconolactone
Î´-Gluconolactone
Î´-Gluconolactone
Î´-Gluconolactone
Î´-Gluconolactone
Lysine
Lysine
Lysine

Lysine
Lysine
(2E)-3-(3,4-dimethoxyphenyl)prop-2-enoic acid
(2E)-3-(3,4-dimethoxyphenyl)prop-2-enoic acid
(2E)-3-(3,4-dimethoxyphenyl)prop-2-enoic acid
(2E)-3-(3,4-dimethoxyphenyl)prop-2-enoic acid
(2E)-3-(3,4-dimethoxyphenyl)prop-2-enoic acid
Indole-3-acetic acid
Indole-3-acetic acid
Indole-3-acetic acid
Indole-3-acetic acid
Indole-3-acetic acid
DNOP_a
DNOP_a
DNOP_a
DNOP_a
DNOP_a
N-(1-Methyl-4-oxo-4,5-dihydro-1H-imidazol-2-yl)alanine_b
N-(1-Methyl-4-oxo-4,5-dihydro-1H-imidazol-2-yl)alanine_b
N-(1-Methyl-4-oxo-4,5-dihydro-1H-imidazol-2-yl)alanine_b
N-(1-Methyl-4-oxo-4,5-dihydro-1H-imidazol-2-yl)alanine_b
N-(1-Methyl-4-oxo-4,5-dihydro-1H-imidazol-2-yl)alanine_b
3-Benzyl-6-isobutyl-2,5-piperazinedione_a
3-Benzyl-6-isobutyl-2,5-piperazinedione_a
3-Benzyl-6-isobutyl-2,5-piperazinedione_a
3-Benzyl-6-isobutyl-2,5-piperazinedione_a
3-Benzyl-6-isobutyl-2,5-piperazinedione_a
3-O-beta-D-galactosyl-sn-glycerol
3-O-beta-D-galactosyl-sn-glycerol
3-O-beta-D-galactosyl-sn-glycerol
3-O-beta-D-galactosyl-sn-glycerol
3-O-beta-D-galactosyl-sn-glycerol
paracetamol sulfate
paracetamol sulfate
paracetamol sulfate
paracetamol sulfate
paracetamol sulfate
3-[4-methyl-1-(2-methylpropanoyl)-3-oxocyclohexyl]butanoic acid
3-[4-methyl-1-(2-methylpropanoyl)-3-oxocyclohexyl]butanoic acid
3-[4-methyl-1-(2-methylpropanoyl)-3-oxocyclohexyl]butanoic acid
3-[4-methyl-1-(2-methylpropanoyl)-3-oxocyclohexyl]butanoic acid
3-[4-methyl-1-(2-methylpropanoyl)-3-oxocyclohexyl]butanoic acid
N-Acetylvaline_a
N-Acetylvaline_a
N-Acetylvaline_a
N-Acetylvaline_a
N-Acetylvaline_a
D-Alanine methyl ester

D-Alanine methyl ester
D-Alanine methyl ester
D-Alanine methyl ester
D-Alanine methyl ester
APM_c
APM_c
APM_c
APM_c
APM_c
3-(2-Oxo-2,3-dihydro-1,3-benzoxazol-3-yl)propanoic acid_a
3-(2-Oxo-2,3-dihydro-1,3-benzoxazol-3-yl)propanoic acid_a
3-(2-Oxo-2,3-dihydro-1,3-benzoxazol-3-yl)propanoic acid_a
3-(2-Oxo-2,3-dihydro-1,3-benzoxazol-3-yl)propanoic acid_a
3-(2-Oxo-2,3-dihydro-1,3-benzoxazol-3-yl)propanoic acid_a
Diethylpyrocarbonate_a
Diethylpyrocarbonate_a
Diethylpyrocarbonate_a
Diethylpyrocarbonate_a
Diethylpyrocarbonate_a
(DL)-3-O-Methyldopa_a
(DL)-3-O-Methyldopa_a
(DL)-3-O-Methyldopa_a
(DL)-3-O-Methyldopa_a
(DL)-3-O-Methyldopa_a
Lovastatin_a
Lovastatin_a
Lovastatin_a
Lovastatin_a
Lovastatin_a
hexobarbital_e
hexobarbital_e
hexobarbital_e
hexobarbital_e
hexobarbital_e
(S)-2-methylbutanal
(S)-2-methylbutanal
(S)-2-methylbutanal
(S)-2-methylbutanal
(S)-2-methylbutanal
Piperine
Piperine
Piperine
Piperine
Piperine
Caprolactam
Caprolactam
Caprolactam
Caprolactam

Caprolactam
7alpha-Hydroxy-3-oxochol-4-en-24-oic acid_c
7alpha-Hydroxy-3-oxochol-4-en-24-oic acid_c
7alpha-Hydroxy-3-oxochol-4-en-24-oic acid_c
7alpha-Hydroxy-3-oxochol-4-en-24-oic acid_c
7alpha-Hydroxy-3-oxochol-4-en-24-oic acid_c
Arginine
Arginine
Arginine
Arginine
Arginine
Cytosine
Cytosine
Cytosine
Cytosine
Cytosine
1,3-dimethyluracil
1,3-dimethyluracil
1,3-dimethyluracil
1,3-dimethyluracil
1,3-dimethyluracil
N-Acetylhistamine
N-Acetylhistamine
N-Acetylhistamine
N-Acetylhistamine
N-Acetylhistamine
Prolylleucine_a
Prolylleucine_a
Prolylleucine_a
Prolylleucine_a
Prolylleucine_a
N-lauroylglycine
N-lauroylglycine
N-lauroylglycine
N-lauroylglycine
N-lauroylglycine
Capryloylglycine_a
Capryloylglycine_a
Capryloylglycine_a
Capryloylglycine_a
Capryloylglycine_a
Metirosine
Metirosine
Metirosine
Metirosine
Metirosine
Triethyl citrate_b
Triethyl citrate_b

Triethyl citrate_b
Triethyl citrate_b
Triethyl citrate_b
3-[2-[(Z)-[3-(2-carboxyethyl)-4-methyl-5-[(3-methyl-5-oxo-4-vinyl-1,2-dihydropyrrol-2-yl)methyl]pyrrol-2-ylidene]r
3-[2-[(Z)-[3-(2-carboxyethyl)-4-methyl-5-[(3-methyl-5-oxo-4-vinyl-1,2-dihydropyrrol-2-yl)methyl]pyrrol-2-ylidene]r
3-[2-[(Z)-[3-(2-carboxyethyl)-4-methyl-5-[(3-methyl-5-oxo-4-vinyl-1,2-dihydropyrrol-2-yl)methyl]pyrrol-2-ylidene]r
3-[2-[(Z)-[3-(2-carboxyethyl)-4-methyl-5-[(3-methyl-5-oxo-4-vinyl-1,2-dihydropyrrol-2-yl)methyl]pyrrol-2-ylidene]r
3-[2-[(Z)-[3-(2-carboxyethyl)-4-methyl-5-[(3-methyl-5-oxo-4-vinyl-1,2-dihydropyrrol-2-yl)methyl]pyrrol-2-ylidene]r
Creatinine
Creatinine
Creatinine
Creatinine
Creatinine
Maltotriose
Maltotriose
Maltotriose
Maltotriose
Maltotriose
1,5-Isoquinolinediol_c
1,5-Isoquinolinediol_c
1,5-Isoquinolinediol_c
1,5-Isoquinolinediol_c
1,5-Isoquinolinediol_c
Choline sulfate
Choline sulfate
Choline sulfate
Choline sulfate
Choline sulfate
7-ketodeoxycholic acid_c
7-ketodeoxycholic acid_c
7-ketodeoxycholic acid_c
7-ketodeoxycholic acid_c
7-ketodeoxycholic acid_c
2-(5-Benzyl-3,6-dioxopiperazin-2-yl)acetic acid
2-(5-Benzyl-3,6-dioxopiperazin-2-yl)acetic acid
2-(5-Benzyl-3,6-dioxopiperazin-2-yl)acetic acid
2-(5-Benzyl-3,6-dioxopiperazin-2-yl)acetic acid
2-(5-Benzyl-3,6-dioxopiperazin-2-yl)acetic acid
Piperidine_a
Piperidine_a
Piperidine_a
Piperidine_a
Piperidine_a
Carbofuran
Carbofuran
Carbofuran
Carbofuran
Carbofuran

(âˆ’)-nabilone_c
(âˆ’)-nabilone_c
(âˆ’)-nabilone_c
(âˆ’)-nabilone_c
(âˆ’)-nabilone_c
Crotamiton
Crotamiton
Crotamiton
Crotamiton
Crotamiton
N6,N6,N6-Trimethyl-L-lysine_c
N6,N6,N6-Trimethyl-L-lysine_c
N6,N6,N6-Trimethyl-L-lysine_c
N6,N6,N6-Trimethyl-L-lysine_c
N6,N6,N6-Trimethyl-L-lysine_c
TO0127900_a
TO0127900_a
TO0127900_a
TO0127900_a
TO0127900_a

(3R,4S,5S,6R,7R,9R,11R,12R,13S,14R)-6-{[(2S,3R,4S,6R)-4-(Dimethylamino)-3-hydroxy-6-methyltetrahydro-2H-pyran-2-yl]oxy}-12,13-dihydroxy-14-[(1S)-1-hydroxyethyl]-4-{[(2R,4R,5S,6S)-5-hydroxy-4-methoxy-4,6-dimethyltetrahydro-2H-pyran-2-yl]oxy}-7-methoxy-3,5,7,9,11,13-hexamethyloxacyclotetradecane-2,10-dione

(3R,4S,5S,6R,7R,9R,11R,12R,13S,14R)-6-{[(2S,3R,4S,6R)-4-(Dimethylamino)-3-hydroxy-6-methyltetrahydro-2H-pyran-2-yl]oxy}-12,13-dihydroxy-14-[(1S)-1-hydroxyethyl]-4-{[(2R,4R,5S,6S)-5-hydroxy-4-methoxy-4,6-dimethyltetrahydro-2H-pyran-2-yl]oxy}-7-methoxy-3,5,7,9,11,13-hexamethyloxacyclotetradecane-2,10-dione

(3R,4S,5S,6R,7R,9R,11R,12R,13S,14R)-6-{[(2S,3R,4S,6R)-4-(Dimethylamino)-3-hydroxy-6-methyltetrahydro-2H-pyran-2-yl]oxy}-12,13-dihydroxy-14-[(1S)-1-hydroxyethyl]-4-{[(2R,4R,5S,6S)-5-hydroxy-4-methoxy-4,6-dimethyltetrahydro-2H-pyran-2-yl]oxy}-7-methoxy-3,5,7,9,11,13-hexamethyloxacyclotetradecane-2,10-dione

(3R,4S,5S,6R,7R,9R,11R,12R,13S,14R)-6-{[(2S,3R,4S,6R)-4-(Dimethylamino)-3-hydroxy-6-methyltetrahydro-2H-pyran-2-yl]oxy}-12,13-dihydroxy-14-[(1S)-1-hydroxyethyl]-4-{[(2R,4R,5S,6S)-5-hydroxy-4-methoxy-4,6-dimethyltetrahydro-2H-pyran-2-yl]oxy}-7-methoxy-3,5,7,9,11,13-hexamethyloxacyclotetradecane-2,10-dione

(3R,4S,5S,6R,7R,9R,11R,12R,13S,14R)-6-{[(2S,3R,4S,6R)-4-(Dimethylamino)-3-hydroxy-6-methyltetrahydro-2H-pyran-2-yl]oxy}-12,13-dihydroxy-14-[(1S)-1-hydroxyethyl]-4-{[(2R,4R,5S,6S)-5-hydroxy-4-methoxy-4,6-dimethyltetrahydro-2H-pyran-2-yl]oxy}-7-methoxy-3,5,7,9,11,13-hexamethyloxacyclotetradecane-2,10-dione
3,3,5,5-Tetramethylpyrroline-N-oxide
3,3,5,5-Tetramethylpyrroline-N-oxide
3,3,5,5-Tetramethylpyrroline-N-oxide
3,3,5,5-Tetramethylpyrroline-N-oxide
3,3,5,5-Tetramethylpyrroline-N-oxide
N-Acetylvaline_b
N-Acetylvaline_b
N-Acetylvaline_b
N-Acetylvaline_b
N-Acetylvaline_b
Biotin
Biotin
Biotin
Biotin
Biotin
Hexamethylene bisacetamide
Hexamethylene bisacetamide
Hexamethylene bisacetamide
Hexamethylene bisacetamide

Hexamethylene bisacetamide
(3Z,6Z,9Z,12Z,21R)-27-Amino-24-hydroxy-24-oxido-18-oxo-19,23,25-trioxa-24lambda~5~-phosphaheptacosa-3,
(3Z,6Z,9Z,12Z,21R)-27-Amino-24-hydroxy-24-oxido-18-oxo-19,23,25-trioxa-24lambda~5~-phosphaheptacosa-3,
(3Z,6Z,9Z,12Z,21R)-27-Amino-24-hydroxy-24-oxido-18-oxo-19,23,25-trioxa-24lambda~5~-phosphaheptacosa-3,
(3Z,6Z,9Z,12Z,21R)-27-Amino-24-hydroxy-24-oxido-18-oxo-19,23,25-trioxa-24lambda~5~-phosphaheptacosa-3,
(3Z,6Z,9Z,12Z,21R)-27-Amino-24-hydroxy-24-oxido-18-oxo-19,23,25-trioxa-24lambda~5~-phosphaheptacosa-3,
Î´-Valerolactam_a
Î´-Valerolactam_a
Î´-Valerolactam_a
Î´-Valerolactam_a
Î´-Valerolactam_a
L-Urobilin
L-Urobilin
L-Urobilin
L-Urobilin
L-Urobilin
UROBILIN, (-)-_a
UROBILIN, (-)-_a
UROBILIN, (-)-_a
UROBILIN, (-)-_a
UROBILIN, (-)-_a
N-(5-acetamidopentyl)acetamide
N-(5-acetamidopentyl)acetamide
N-(5-acetamidopentyl)acetamide
N-(5-acetamidopentyl)acetamide
N-(5-acetamidopentyl)acetamide
Cyclamic acid
Cyclamic acid
Cyclamic acid
Cyclamic acid
Cyclamic acid
urobilinogen
urobilinogen
urobilinogen
urobilinogen
urobilinogen
N-Acetylputrescine
N-Acetylputrescine
N-Acetylputrescine
N-Acetylputrescine
N-Acetylputrescine
presqualene diphosphate
presqualene diphosphate
presqualene diphosphate
presqualene diphosphate
presqualene diphosphate
Acetophenone
Acetophenone

Acetophenone
Acetophenone
Acetophenone
Prilocaine
Prilocaine
Prilocaine
Prilocaine
Prilocaine
4-Acetamidophenol
4-Acetamidophenol
4-Acetamidophenol
4-Acetamidophenol
4-Acetamidophenol
(Â±)-Albuterol
(Â±)-Albuterol
(Â±)-Albuterol
(Â±)-Albuterol
(Â±)-Albuterol
(-)-Erythromycin
(-)-Erythromycin
(-)-Erythromycin
(-)-Erythromycin
(-)-Erythromycin
N6,N6,N6-Trimethyl-L-lysine_b
N6,N6,N6-Trimethyl-L-lysine_b
N6,N6,N6-Trimethyl-L-lysine_b
N6,N6,N6-Trimethyl-L-lysine_b
N6,N6,N6-Trimethyl-L-lysine_b
Pipecolinic acid
Pipecolinic acid
Pipecolinic acid
Pipecolinic acid
Pipecolinic acid
DNOP_f
DNOP_f
DNOP_f
DNOP_f
DNOP_f
N-Methylcaprolactam
N-Methylcaprolactam
N-Methylcaprolactam
N-Methylcaprolactam
N-Methylcaprolactam
Limonin
Limonin
Limonin
Limonin
Limonin

Atenolol
Atenolol
Atenolol
Atenolol
Atenolol
Styrene
Styrene
Styrene
Styrene
Styrene
Valylvaline_g
Valylvaline_g
Valylvaline_g
Valylvaline_g
Valylvaline_g
2-(Hydroxymethyl)-1-methyl-3,4,5-piperidinetriol
2-(Hydroxymethyl)-1-methyl-3,4,5-piperidinetriol
2-(Hydroxymethyl)-1-methyl-3,4,5-piperidinetriol
2-(Hydroxymethyl)-1-methyl-3,4,5-piperidinetriol
2-(Hydroxymethyl)-1-methyl-3,4,5-piperidinetriol
Theobromine
Theobromine
Theobromine
Theobromine
Theobromine
6-hydroxypseudooxynicotine_b
6-hydroxypseudooxynicotine_b
6-hydroxypseudooxynicotine_b
6-hydroxypseudooxynicotine_b
6-hydroxypseudooxynicotine_b
Isoprene
Isoprene
Isoprene
Isoprene
Isoprene
Tyramine
Tyramine
Tyramine
Tyramine
Tyramine
1-[(4E)-4-(4-Methyl-5-oxo-2(5H)-furanylidene)butyl]-2-pyrrolidinone
1-[(4E)-4-(4-Methyl-5-oxo-2(5H)-furanylidene)butyl]-2-pyrrolidinone
1-[(4E)-4-(4-Methyl-5-oxo-2(5H)-furanylidene)butyl]-2-pyrrolidinone
1-[(4E)-4-(4-Methyl-5-oxo-2(5H)-furanylidene)butyl]-2-pyrrolidinone
1-[(4E)-4-(4-Methyl-5-oxo-2(5H)-furanylidene)butyl]-2-pyrrolidinone
Methylimidazoleacetic acid
Methylimidazoleacetic acid
Methylimidazoleacetic acid

Methylimidazoleacetic acid
Methylimidazoleacetic acid
butalbital_d
butalbital_d
butalbital_d
butalbital_d
butalbital_d
Piperidine_d
Piperidine_d
Piperidine_d
Piperidine_d
Piperidine_d
UROBILIN, (-)-_b
UROBILIN, (-)-_b
UROBILIN, (-)-_b
UROBILIN, (-)-_b
UROBILIN, (-)-_b
(2S)-6-Amino-2-[(E)-(hydroxymethylene)amino]hexanimidic acid_b
(2S)-6-Amino-2-[(E)-(hydroxymethylene)amino]hexanimidic acid_b
(2S)-6-Amino-2-[(E)-(hydroxymethylene)amino]hexanimidic acid_b
(2S)-6-Amino-2-[(E)-(hydroxymethylene)amino]hexanimidic acid_b
(2S)-6-Amino-2-[(E)-(hydroxymethylene)amino]hexanimidic acid_b
Î´-Valerolactam_b
Î´-Valerolactam_b
Î´-Valerolactam_b
Î´-Valerolactam_b
Î´-Valerolactam_b
MFCD00025555_d
MFCD00025555_d
MFCD00025555_d
MFCD00025555_d
MFCD00025555_d
N-Acetylneuraminic acid
N-Acetylneuraminic acid
N-Acetylneuraminic acid
N-Acetylneuraminic acid
N-Acetylneuraminic acid
Methylol Dimethylhydantoin_a
Methylol Dimethylhydantoin_a
Methylol Dimethylhydantoin_a
Methylol Dimethylhydantoin_a
Methylol Dimethylhydantoin_a
Deoxysugar II (possibly Fucose)
Deoxysugar II (possibly Fucose)
Deoxysugar II (possibly Fucose)
Deoxysugar II (possibly Fucose)
Deoxysugar II (possibly Fucose)
DNOP_e

DNOP_e
DNOP_e
DNOP_e
DNOP_e
Tropinone
Tropinone
Tropinone
Tropinone
Tropinone
Thymidine
Thymidine
Thymidine
Thymidine
Thymidine
8-Methyl-8-azabicyclo[3.2.1]octane-1,2,3,4,6-pentol_a
8-Methyl-8-azabicyclo[3.2.1]octane-1,2,3,4,6-pentol_a
8-Methyl-8-azabicyclo[3.2.1]octane-1,2,3,4,6-pentol_a
8-Methyl-8-azabicyclo[3.2.1]octane-1,2,3,4,6-pentol_a
8-Methyl-8-azabicyclo[3.2.1]octane-1,2,3,4,6-pentol_a
Solanidine
Solanidine
Solanidine
Solanidine
Solanidine
Tetrahydrofuran
Tetrahydrofuran
Tetrahydrofuran
Tetrahydrofuran
Tetrahydrofuran
Crotonic acid
Crotonic acid
Crotonic acid
Crotonic acid
Crotonic acid
N-{6-[(7-Chloro-4-quinazolinyl)oxy]-3-pyridinyl}-2-thiophenesulfonamide
N-{6-[(7-Chloro-4-quinazolinyl)oxy]-3-pyridinyl}-2-thiophenesulfonamide
N-{6-[(7-Chloro-4-quinazolinyl)oxy]-3-pyridinyl}-2-thiophenesulfonamide
N-{6-[(7-Chloro-4-quinazolinyl)oxy]-3-pyridinyl}-2-thiophenesulfonamide
N-{6-[(7-Chloro-4-quinazolinyl)oxy]-3-pyridinyl}-2-thiophenesulfonamide
Prolylleucine_b
Prolylleucine_b
Prolylleucine_b
Prolylleucine_b
Prolylleucine_b
3-[2-[(Z)-[3-(2-carboxyethyl)-4-methyl-5-[(3-methyl-5-oxo-4-vinyl-1,2-dihydropyrrol-2-yl)methyl]pyrrol-2-ylidene]m
3-[2-[(Z)-[3-(2-carboxyethyl)-4-methyl-5-[(3-methyl-5-oxo-4-vinyl-1,2-dihydropyrrol-2-yl)methyl]pyrrol-2-ylidene]m
3-[2-[(Z)-[3-(2-carboxyethyl)-4-methyl-5-[(3-methyl-5-oxo-4-vinyl-1,2-dihydropyrrol-2-yl)methyl]pyrrol-2-ylidene]m
3-[2-[(Z)-[3-(2-carboxyethyl)-4-methyl-5-[(3-methyl-5-oxo-4-vinyl-1,2-dihydropyrrol-2-yl)methyl]pyrrol-2-ylidene]m

3-[2-[(Z)-[3-(2-carboxyethyl)-4-methyl-5-[(3-methyl-5-oxo-4-vinyl-1,2-dihydropyrrol-2-yl)methyl]pyrrol-2-ylidene]r
3-Hydroxy-2-methyl-2-[(sulfooxy)methyl]propanoic acid
3-Hydroxy-2-methyl-2-[(sulfooxy)methyl]propanoic acid
3-Hydroxy-2-methyl-2-[(sulfooxy)methyl]propanoic acid
3-Hydroxy-2-methyl-2-[(sulfooxy)methyl]propanoic acid
3-Hydroxy-2-methyl-2-[(sulfooxy)methyl]propanoic acid
Docosahexaenoic acid ethyl ester_b
Docosahexaenoic acid ethyl ester_b
Docosahexaenoic acid ethyl ester_b
Docosahexaenoic acid ethyl ester_b
Docosahexaenoic acid ethyl ester_b
3-(2-Oxo-2,3-dihydro-1,3-benzoxazol-3-yl)propanoic acid_b
3-(2-Oxo-2,3-dihydro-1,3-benzoxazol-3-yl)propanoic acid_b
3-(2-Oxo-2,3-dihydro-1,3-benzoxazol-3-yl)propanoic acid_b
3-(2-Oxo-2,3-dihydro-1,3-benzoxazol-3-yl)propanoic acid_b
3-(2-Oxo-2,3-dihydro-1,3-benzoxazol-3-yl)propanoic acid_b
N-{3-Carboxy-3-[(3-carboxy-3-hydroxypropyl)amino]propyl}homoserine
N-{3-Carboxy-3-[(3-carboxy-3-hydroxypropyl)amino]propyl}homoserine
N-{3-Carboxy-3-[(3-carboxy-3-hydroxypropyl)amino]propyl}homoserine
N-{3-Carboxy-3-[(3-carboxy-3-hydroxypropyl)amino]propyl}homoserine
N-{3-Carboxy-3-[(3-carboxy-3-hydroxypropyl)amino]propyl}homoserine
2-Hydroxy-13,20-dimethoxy-4,7,17,22,22-pentamethyl-11-oxo-5,10,21,23-tetraoxahexacyclo[18.2.1.0~1,17~.0~
2-Hydroxy-13,20-dimethoxy-4,7,17,22,22-pentamethyl-11-oxo-5,10,21,23-tetraoxahexacyclo[18.2.1.0~1,17~.0~
2-Hydroxy-13,20-dimethoxy-4,7,17,22,22-pentamethyl-11-oxo-5,10,21,23-tetraoxahexacyclo[18.2.1.0~1,17~.0~
2-Hydroxy-13,20-dimethoxy-4,7,17,22,22-pentamethyl-11-oxo-5,10,21,23-tetraoxahexacyclo[18.2.1.0~1,17~.0~
2-Hydroxy-13,20-dimethoxy-4,7,17,22,22-pentamethyl-11-oxo-5,10,21,23-tetraoxahexacyclo[18.2.1.0~1,17~.0~
Cadaverine
Cadaverine
Cadaverine
Cadaverine
Cadaverine
Indole-3-carboxyaldehyde
Indole-3-carboxyaldehyde
Indole-3-carboxyaldehyde
Indole-3-carboxyaldehyde
Indole-3-carboxyaldehyde
4-Hydroxyprolylleucine_d
4-Hydroxyprolylleucine_d
4-Hydroxyprolylleucine_d
4-Hydroxyprolylleucine_d
4-Hydroxyprolylleucine_d
3',5,7-Trihydroxy-4'-methoxyflavanone
3',5,7-Trihydroxy-4'-methoxyflavanone
3',5,7-Trihydroxy-4'-methoxyflavanone
3',5,7-Trihydroxy-4'-methoxyflavanone
3',5,7-Trihydroxy-4'-methoxyflavanone
6-(alpha-D-glucosaminyl)-1D-myo-inositol_a
6-(alpha-D-glucosaminyl)-1D-myo-inositol_a

6-(alpha-D-glucosaminyl)-1D-myo-inositol_a
6-(alpha-D-glucosaminyl)-1D-myo-inositol_a
6-(alpha-D-glucosaminyl)-1D-myo-inositol_a
Ethyl malate_c
Ethyl malate_c
Ethyl malate_c
Ethyl malate_c
Ethyl malate_c
Safrole
Safrole
Safrole
Safrole
Safrole
1,1'-[1,12-Dodecanediylbis(oxy)]dibenzene_b
1,1'-[1,12-Dodecanediylbis(oxy)]dibenzene_b
1,1'-[1,12-Dodecanediylbis(oxy)]dibenzene_b
1,1'-[1,12-Dodecanediylbis(oxy)]dibenzene_b
1,1'-[1,12-Dodecanediylbis(oxy)]dibenzene_b
DNOP_g
DNOP_g
DNOP_g
DNOP_g
DNOP_g
(2E)-3-Methyl-4-(sulfooxy)-2-butenoic acid_a
(2E)-3-Methyl-4-(sulfooxy)-2-butenoic acid_a
(2E)-3-Methyl-4-(sulfooxy)-2-butenoic acid_a
(2E)-3-Methyl-4-(sulfooxy)-2-butenoic acid_a
(2E)-3-Methyl-4-(sulfooxy)-2-butenoic acid_a
Citrulline
Citrulline
Citrulline
Citrulline
Citrulline
g-Butyrobetaine
g-Butyrobetaine
g-Butyrobetaine
g-Butyrobetaine
g-Butyrobetaine
Piperidine_e
Piperidine_e
Piperidine_e
Piperidine_e
Piperidine_e
Isopelletierine
Isopelletierine
Isopelletierine
Isopelletierine
Isopelletierine

2-Acetamidohexanedioic acid_d
2-Acetamidohexanedioic acid_d
2-Acetamidohexanedioic acid_d
2-Acetamidohexanedioic acid_d
2-Acetamidohexanedioic acid_d
4-Vinylphenol
4-Vinylphenol
4-Vinylphenol
4-Vinylphenol
4-Vinylphenol
2-Hydroxycinnamic acid
2-Hydroxycinnamic acid
2-Hydroxycinnamic acid
2-Hydroxycinnamic acid
2-Hydroxycinnamic acid
Methyl (2Z,3E,5Z)-6-{4,4-dimethyl-3-[(3-methyl-2-buten-1-yl)oxy]-3,4-dihydro-2H-1,5-benzodioxepin-7-yl}-3-(hyd
Methyl (2Z,3E,5Z)-6-{4,4-dimethyl-3-[(3-methyl-2-buten-1-yl)oxy]-3,4-dihydro-2H-1,5-benzodioxepin-7-yl}-3-(hyd
Methyl (2Z,3E,5Z)-6-{4,4-dimethyl-3-[(3-methyl-2-buten-1-yl)oxy]-3,4-dihydro-2H-1,5-benzodioxepin-7-yl}-3-(hyd
Methyl (2Z,3E,5Z)-6-{4,4-dimethyl-3-[(3-methyl-2-buten-1-yl)oxy]-3,4-dihydro-2H-1,5-benzodioxepin-7-yl}-3-(hyd
Methyl (2Z,3E,5Z)-6-{4,4-dimethyl-3-[(3-methyl-2-buten-1-yl)oxy]-3,4-dihydro-2H-1,5-benzodioxepin-7-yl}-3-(hyd
Tiglic acid_a
Tiglic acid_a
Tiglic acid_a
Tiglic acid_a
Tiglic acid_a
O-Ethyl (4-hydroxyphenyl)carbamothioate
O-Ethyl (4-hydroxyphenyl)carbamothioate
O-Ethyl (4-hydroxyphenyl)carbamothioate
O-Ethyl (4-hydroxyphenyl)carbamothioate
O-Ethyl (4-hydroxyphenyl)carbamothioate
porphobilinogen_b
porphobilinogen_b
porphobilinogen_b
porphobilinogen_b
porphobilinogen_b
DL-4-Hydroxyphenyllactic acid
DL-4-Hydroxyphenyllactic acid
DL-4-Hydroxyphenyllactic acid
DL-4-Hydroxyphenyllactic acid
DL-4-Hydroxyphenyllactic acid
4-(METHYLNITROSAMINO)-1-(3-PYRIDYL-N-OXIDE)-1-BUTANONE_b
4-(METHYLNITROSAMINO)-1-(3-PYRIDYL-N-OXIDE)-1-BUTANONE_b
4-(METHYLNITROSAMINO)-1-(3-PYRIDYL-N-OXIDE)-1-BUTANONE_b
4-(METHYLNITROSAMINO)-1-(3-PYRIDYL-N-OXIDE)-1-BUTANONE_b
4-(METHYLNITROSAMINO)-1-(3-PYRIDYL-N-OXIDE)-1-BUTANONE_b
Leu-Val_e
Leu-Val_e
Leu-Val_e

Leu-Val_e
Leu-Val_e
Valylvaline_d
Valylvaline_d
Valylvaline_d
Valylvaline_d
Valylvaline_d
Phenyl D-glucopyranosiduronic acid_d
Phenyl D-glucopyranosiduronic acid_d
Phenyl D-glucopyranosiduronic acid_d
Phenyl D-glucopyranosiduronic acid_d
Phenyl D-glucopyranosiduronic acid_d
Lanthionine ketimine_a
Lanthionine ketimine_a
Lanthionine ketimine_a
Lanthionine ketimine_a
Lanthionine ketimine_a
9-Methyluric acid_b
9-Methyluric acid_b
9-Methyluric acid_b
9-Methyluric acid_b
9-Methyluric acid_b
2-Hydroxy-4-methylthiobutanoic acid
2-Hydroxy-4-methylthiobutanoic acid
2-Hydroxy-4-methylthiobutanoic acid
2-Hydroxy-4-methylthiobutanoic acid
2-Hydroxy-4-methylthiobutanoic acid
Seryltyrosine_b
Seryltyrosine_b
Seryltyrosine_b
Seryltyrosine_b
Seryltyrosine_b
2-Amino-4-methylpyrimidine
2-Amino-4-methylpyrimidine
2-Amino-4-methylpyrimidine
2-Amino-4-methylpyrimidine
2-Amino-4-methylpyrimidine
LU3453000
LU3453000
LU3453000
LU3453000
LU3453000
Brilliant blue FCF
Brilliant blue FCF
Brilliant blue FCF
Brilliant blue FCF
Brilliant blue FCF
Coumarone

Coumarone
Coumarone
Coumarone
Coumarone
1-Methylhistidine
1-Methylhistidine
1-Methylhistidine
1-Methylhistidine
1-Methylhistidine
Setoclavine
Setoclavine
Setoclavine
Setoclavine
Setoclavine
Butabarbital_a
Butabarbital_a
Butabarbital_a
Butabarbital_a
Butabarbital_a
Mevalonic acid
Mevalonic acid
Mevalonic acid
Mevalonic acid
Mevalonic acid
adrenaline
adrenaline
adrenaline
adrenaline
adrenaline
DNOP_b
DNOP_b
DNOP_b
DNOP_b
DNOP_b
Capryloylglycine_b
Capryloylglycine_b
Capryloylglycine_b
Capryloylglycine_b
Capryloylglycine_b
Lidocaine
Lidocaine
Lidocaine
Lidocaine
Lidocaine
N6,N6,N6-Trimethyl-L-lysine_a
N6,N6,N6-Trimethyl-L-lysine_a
N6,N6,N6-Trimethyl-L-lysine_a
N6,N6,N6-Trimethyl-L-lysine_a

N6,N6,N6-Trimethyl-L-lysine_a
Indole-3-acetaldehyde
Indole-3-acetaldehyde
Indole-3-acetaldehyde
Indole-3-acetaldehyde
Indole-3-acetaldehyde
Prolylhydroxyproline
Prolylhydroxyproline
Prolylhydroxyproline
Prolylhydroxyproline
Prolylhydroxyproline
1-Methylxanthine
1-Methylxanthine
1-Methylxanthine
1-Methylxanthine
1-Methylxanthine
Methyl 3-formyl-1H-indole-2-sulfinate
Methyl 3-formyl-1H-indole-2-sulfinate
Methyl 3-formyl-1H-indole-2-sulfinate
Methyl 3-formyl-1H-indole-2-sulfinate
Methyl 3-formyl-1H-indole-2-sulfinate
8-Hydroxyquinoline
8-Hydroxyquinoline
8-Hydroxyquinoline
8-Hydroxyquinoline
8-Hydroxyquinoline
Glycylprolylhydroxyproline_a
Glycylprolylhydroxyproline_a
Glycylprolylhydroxyproline_a
Glycylprolylhydroxyproline_a
Glycylprolylhydroxyproline_a
(15Z)-9,12,13-Trihydroxy-15-octadecenoic acid
(15Z)-9,12,13-Trihydroxy-15-octadecenoic acid
(15Z)-9,12,13-Trihydroxy-15-octadecenoic acid
(15Z)-9,12,13-Trihydroxy-15-octadecenoic acid
(15Z)-9,12,13-Trihydroxy-15-octadecenoic acid
2-(1,3-Benzodioxol-5-yl)-5,7-dimethoxy-3-chromanol
2-(1,3-Benzodioxol-5-yl)-5,7-dimethoxy-3-chromanol
2-(1,3-Benzodioxol-5-yl)-5,7-dimethoxy-3-chromanol
2-(1,3-Benzodioxol-5-yl)-5,7-dimethoxy-3-chromanol
2-(1,3-Benzodioxol-5-yl)-5,7-dimethoxy-3-chromanol
Methyl [9-(3-furyl)-4-hydroxy-3,3,5a,9a,11b-pentamethyl-5,7-dioxotetradecahydro[2]benzofuro[5,4-f]oxireno[d]is
Methyl [9-(3-furyl)-4-hydroxy-3,3,5a,9a,11b-pentamethyl-5,7-dioxotetradecahydro[2]benzofuro[5,4-f]oxireno[d]is
Methyl [9-(3-furyl)-4-hydroxy-3,3,5a,9a,11b-pentamethyl-5,7-dioxotetradecahydro[2]benzofuro[5,4-f]oxireno[d]is
Methyl [9-(3-furyl)-4-hydroxy-3,3,5a,9a,11b-pentamethyl-5,7-dioxotetradecahydro[2]benzofuro[5,4-f]oxireno[d]is
Methyl [9-(3-furyl)-4-hydroxy-3,3,5a,9a,11b-pentamethyl-5,7-dioxotetradecahydro[2]benzofuro[5,4-f]oxireno[d]is
Midodrine_b
Midodrine_b

Midodrine_b
Midodrine_b
Midodrine_b
7-[4-(tert-butyl)phenyl]-2-methyl-5-(methylthio)pyrazolo[1,5-a]pyrimidine-6-carbonitrile
7-[4-(tert-butyl)phenyl]-2-methyl-5-(methylthio)pyrazolo[1,5-a]pyrimidine-6-carbonitrile
7-[4-(tert-butyl)phenyl]-2-methyl-5-(methylthio)pyrazolo[1,5-a]pyrimidine-6-carbonitrile
7-[4-(tert-butyl)phenyl]-2-methyl-5-(methylthio)pyrazolo[1,5-a]pyrimidine-6-carbonitrile
7-[4-(tert-butyl)phenyl]-2-methyl-5-(methylthio)pyrazolo[1,5-a]pyrimidine-6-carbonitrile
Lysylvaline_a
Lysylvaline_a
Lysylvaline_a
Lysylvaline_a
Lysylvaline_a
11-dehydro Thromboxane B2
11-dehydro Thromboxane B2
11-dehydro Thromboxane B2
11-dehydro Thromboxane B2
11-dehydro Thromboxane B2
2-Hydroxy-13,20-dimethoxy-4,7,17,22,22-pentamethyl-5,10,21,23-tetraoxahexacyclo[18.2.1.0~1,17~.0~4,16~.[illegible]
2-Hydroxy-13,20-dimethoxy-4,7,17,22,22-pentamethyl-5,10,21,23-tetraoxahexacyclo[18.2.1.0~1,17~.0~4,16~.[illegible]
2-Hydroxy-13,20-dimethoxy-4,7,17,22,22-pentamethyl-5,10,21,23-tetraoxahexacyclo[18.2.1.0~1,17~.0~4,16~.[illegible]
2-Hydroxy-13,20-dimethoxy-4,7,17,22,22-pentamethyl-5,10,21,23-tetraoxahexacyclo[18.2.1.0~1,17~.0~4,16~.[illegible]
2-Hydroxy-13,20-dimethoxy-4,7,17,22,22-pentamethyl-5,10,21,23-tetraoxahexacyclo[18.2.1.0~1,17~.0~4,16~.[illegible]
3b-Hydroxy-5-cholenoic acid_a
3b-Hydroxy-5-cholenoic acid_a
3b-Hydroxy-5-cholenoic acid_a
3b-Hydroxy-5-cholenoic acid_a
3b-Hydroxy-5-cholenoic acid_a
4-[(2E,4Z)-6-Carboxy-2,4-heptadien-2-yl]-3-(carboxymethyl)proline
4-[(2E,4Z)-6-Carboxy-2,4-heptadien-2-yl]-3-(carboxymethyl)proline
4-[(2E,4Z)-6-Carboxy-2,4-heptadien-2-yl]-3-(carboxymethyl)proline
4-[(2E,4Z)-6-Carboxy-2,4-heptadien-2-yl]-3-(carboxymethyl)proline
4-[(2E,4Z)-6-Carboxy-2,4-heptadien-2-yl]-3-(carboxymethyl)proline
2-Hydroxyphenylalanine
2-Hydroxyphenylalanine
2-Hydroxyphenylalanine
2-Hydroxyphenylalanine
2-Hydroxyphenylalanine
asn-pro_c
asn-pro_c
asn-pro_c
asn-pro_c
asn-pro_c
Dodecanedioic acid
Dodecanedioic acid
Dodecanedioic acid
Dodecanedioic acid
Dodecanedioic acid

Cys-tyr
Cys-tyr
Cys-tyr
Cys-tyr
Cys-tyr
N-(3-acetamidopropyl)pyrrolidin-2-one_c
N-(3-acetamidopropyl)pyrrolidin-2-one_c
N-(3-acetamidopropyl)pyrrolidin-2-one_c
N-(3-acetamidopropyl)pyrrolidin-2-one_c
N-(3-acetamidopropyl)pyrrolidin-2-one_c
MFCD12974029
MFCD12974029
MFCD12974029
MFCD12974029
MFCD12974029
N,N-Dimethyl-L-histidine_a
N,N-Dimethyl-L-histidine_a
N,N-Dimethyl-L-histidine_a
N,N-Dimethyl-L-histidine_a
N,N-Dimethyl-L-histidine_a
Leucyltyrosine_a
Leucyltyrosine_a
Leucyltyrosine_a
Leucyltyrosine_a
Leucyltyrosine_a
L-gamma-Glutamyl-L-leucine_b
L-gamma-Glutamyl-L-leucine_b
L-gamma-Glutamyl-L-leucine_b
L-gamma-Glutamyl-L-leucine_b
L-gamma-Glutamyl-L-leucine_b
Î±-Muricholic acid
Î±-Muricholic acid
Î±-Muricholic acid
Î±-Muricholic acid
Î±-Muricholic acid
Dihydrouridine_a
Dihydrouridine_a
Dihydrouridine_a
Dihydrouridine_a
Dihydrouridine_a
1-(4-Amino-4-carboxybutanoyl)-2-piperidinecarboxylic acid_a
1-(4-Amino-4-carboxybutanoyl)-2-piperidinecarboxylic acid_a
1-(4-Amino-4-carboxybutanoyl)-2-piperidinecarboxylic acid_a
1-(4-Amino-4-carboxybutanoyl)-2-piperidinecarboxylic acid_a
1-(4-Amino-4-carboxybutanoyl)-2-piperidinecarboxylic acid_a
Coprine_c
Coprine_c
Coprine_c

Coprine_c
Coprine_c
1-(beta-D-Ribofuranosyl)-1,4-dihydronicotinamide
1-(beta-D-Ribofuranosyl)-1,4-dihydronicotinamide
1-(beta-D-Ribofuranosyl)-1,4-dihydronicotinamide
1-(beta-D-Ribofuranosyl)-1,4-dihydronicotinamide
1-(beta-D-Ribofuranosyl)-1,4-dihydronicotinamide
Isoprenaline
Isoprenaline
Isoprenaline
Isoprenaline
Isoprenaline
(2E,6E)-9-[(2R)-6-Hydroxy-2,5,7,8-tetramethyl-3,4-dihydro-2H-chromen-2-yl]-2,6-dimethyl-2,6-nonadienoic acid
(2E,6E)-9-[(2R)-6-Hydroxy-2,5,7,8-tetramethyl-3,4-dihydro-2H-chromen-2-yl]-2,6-dimethyl-2,6-nonadienoic acid
(2E,6E)-9-[(2R)-6-Hydroxy-2,5,7,8-tetramethyl-3,4-dihydro-2H-chromen-2-yl]-2,6-dimethyl-2,6-nonadienoic acid
(2E,6E)-9-[(2R)-6-Hydroxy-2,5,7,8-tetramethyl-3,4-dihydro-2H-chromen-2-yl]-2,6-dimethyl-2,6-nonadienoic acid
(2E,6E)-9-[(2R)-6-Hydroxy-2,5,7,8-tetramethyl-3,4-dihydro-2H-chromen-2-yl]-2,6-dimethyl-2,6-nonadienoic acid
Indole-3-carboxylate
Indole-3-carboxylate
Indole-3-carboxylate
Indole-3-carboxylate
Indole-3-carboxylate
Piceid
Piceid
Piceid
Piceid
Piceid
Bile acid I (similar to Deoxycholic acid)
Bile acid I (similar to Deoxycholic acid)
Bile acid I (similar to Deoxycholic acid)
Bile acid I (similar to Deoxycholic acid)
Bile acid I (similar to Deoxycholic acid)
N-Ethylpropanamide_a
N-Ethylpropanamide_a
N-Ethylpropanamide_a
N-Ethylpropanamide_a
N-Ethylpropanamide_a
MFCD18695608_b
MFCD18695608_b
MFCD18695608_b
MFCD18695608_b
MFCD18695608_b
7,8-Diaminononanoic acid
7,8-Diaminononanoic acid
7,8-Diaminononanoic acid
7,8-Diaminononanoic acid
7,8-Diaminononanoic acid
tyramine sulfate_a

tyramine sulfate_a
tyramine sulfate_a
tyramine sulfate_a
tyramine sulfate_a
Nitrosoheptamethyleneimine
Nitrosoheptamethyleneimine
Nitrosoheptamethyleneimine
Nitrosoheptamethyleneimine
Nitrosoheptamethyleneimine
1,3,7-Trimethyluric acid
1,3,7-Trimethyluric acid
1,3,7-Trimethyluric acid
1,3,7-Trimethyluric acid
1,3,7-Trimethyluric acid
N2-Acetyllysine
N2-Acetyllysine
N2-Acetyllysine
N2-Acetyllysine
N2-Acetyllysine
3-Methylsulfolene
3-Methylsulfolene
3-Methylsulfolene
3-Methylsulfolene
3-Methylsulfolene
N-(Carboxymethyl)norleucine_a
N-(Carboxymethyl)norleucine_a
N-(Carboxymethyl)norleucine_a
N-(Carboxymethyl)norleucine_a
N-(Carboxymethyl)norleucine_a
tert-Butyl 3-amino-1-methyl-2,3-dioxopropylcarbamate_a
tert-Butyl 3-amino-1-methyl-2,3-dioxopropylcarbamate_a
tert-Butyl 3-amino-1-methyl-2,3-dioxopropylcarbamate_a
tert-Butyl 3-amino-1-methyl-2,3-dioxopropylcarbamate_a
tert-Butyl 3-amino-1-methyl-2,3-dioxopropylcarbamate_a
meticillin
meticillin
meticillin
meticillin
meticillin
2-Acetamidohexanedioic acid_a
2-Acetamidohexanedioic acid_a
2-Acetamidohexanedioic acid_a
2-Acetamidohexanedioic acid_a
2-Acetamidohexanedioic acid_a
Uric acid
Uric acid
Uric acid
Uric acid

Uric acid
7-ketodeoxycholic acid_a
7-ketodeoxycholic acid_a
7-ketodeoxycholic acid_a
7-ketodeoxycholic acid_a
7-ketodeoxycholic acid_a
N-Propionylmethionine_c
N-Propionylmethionine_c
N-Propionylmethionine_c
N-Propionylmethionine_c
N-Propionylmethionine_c
Butabarbital_c
Butabarbital_c
Butabarbital_c
Butabarbital_c
Butabarbital_c
Leucyltyrosine_c
Leucyltyrosine_c
Leucyltyrosine_c
Leucyltyrosine_c
Leucyltyrosine_c
MFCD00025555_a
MFCD00025555_a
MFCD00025555_a
MFCD00025555_a
MFCD00025555_a
Hept-2-ulose
Hept-2-ulose
Hept-2-ulose
Hept-2-ulose
Hept-2-ulose
1,5-Isoquinolinediol_b
1,5-Isoquinolinediol_b
1,5-Isoquinolinediol_b
1,5-Isoquinolinediol_b
1,5-Isoquinolinediol_b
hypaphorine
hypaphorine
hypaphorine
hypaphorine
hypaphorine
meprobamate_c
meprobamate_c
meprobamate_c
meprobamate_c
meprobamate_c
2,5-Dimethylphenol
2,5-Dimethylphenol

2,5-Dimethylphenol
2,5-Dimethylphenol
2,5-Dimethylphenol
N-Stearoyltyrosine
N-Stearoyltyrosine
N-Stearoyltyrosine
N-Stearoyltyrosine
N-Stearoyltyrosine
1-(4-Aminobutyl)urea_b
1-(4-Aminobutyl)urea_b
1-(4-Aminobutyl)urea_b
1-(4-Aminobutyl)urea_b
1-(4-Aminobutyl)urea_b
3,3-Dimethylglutaric acid_c
3,3-Dimethylglutaric acid_c
3,3-Dimethylglutaric acid_c
3,3-Dimethylglutaric acid_c
3,3-Dimethylglutaric acid_c
Pilocarpine
Pilocarpine
Pilocarpine
Pilocarpine
Pilocarpine
Hydroxypropionylcarnitine_b
Hydroxypropionylcarnitine_b
Hydroxypropionylcarnitine_b
Hydroxypropionylcarnitine_b
Hydroxypropionylcarnitine_b
NPC
NPC
NPC
NPC
NPC
1-methylhypoxanthine
1-methylhypoxanthine
1-methylhypoxanthine
1-methylhypoxanthine
1-methylhypoxanthine
Ethanoic anhydride
Ethanoic anhydride
Ethanoic anhydride
Ethanoic anhydride
Ethanoic anhydride
7-Sulfocholic acid
7-Sulfocholic acid
7-Sulfocholic acid
7-Sulfocholic acid
7-Sulfocholic acid

3b-Hydroxy-5-cholenoic acid_b
3b-Hydroxy-5-cholenoic acid_b
3b-Hydroxy-5-cholenoic acid_b
3b-Hydroxy-5-cholenoic acid_b
3b-Hydroxy-5-cholenoic acid_b
Aurorix
Aurorix
Aurorix
Aurorix
Aurorix
2-(1-Ethoxyethoxy)propanoic acid_b
2-(1-Ethoxyethoxy)propanoic acid_b
2-(1-Ethoxyethoxy)propanoic acid_b
2-(1-Ethoxyethoxy)propanoic acid_b
2-(1-Ethoxyethoxy)propanoic acid_b
2-Acetamidohexanedioic acid_g
2-Acetamidohexanedioic acid_g
2-Acetamidohexanedioic acid_g
2-Acetamidohexanedioic acid_g
2-Acetamidohexanedioic acid_g
4-Iodoanisole
4-Iodoanisole
4-Iodoanisole
4-Iodoanisole
4-Iodoanisole
Phenethylamine
Phenethylamine
Phenethylamine
Phenethylamine
Phenethylamine
Arabic acid
Arabic acid
Arabic acid
Arabic acid
Arabic acid
6-Methylquinoline
6-Methylquinoline
6-Methylquinoline
6-Methylquinoline
6-Methylquinoline
S-Allylcysteine_c
S-Allylcysteine_c
S-Allylcysteine_c
S-Allylcysteine_c
S-Allylcysteine_c
YV8195000
YV8195000
YV8195000

YV8195000
YV8195000
N-Methyl-1H-indole-3-propanamide
N-Methyl-1H-indole-3-propanamide
N-Methyl-1H-indole-3-propanamide
N-Methyl-1H-indole-3-propanamide
N-Methyl-1H-indole-3-propanamide
1,1'-[1,12-Dodecanediylbis(oxy)]dibenzene_a
1,1'-[1,12-Dodecanediylbis(oxy)]dibenzene_a
1,1'-[1,12-Dodecanediylbis(oxy)]dibenzene_a
1,1'-[1,12-Dodecanediylbis(oxy)]dibenzene_a
1,1'-[1,12-Dodecanediylbis(oxy)]dibenzene_a
Methohexital_b
Methohexital_b
Methohexital_b
Methohexital_b
Methohexital_b
3-Aminosalicylic acid
3-Aminosalicylic acid
3-Aminosalicylic acid
3-Aminosalicylic acid
3-Aminosalicylic acid
3-(1-hydroxyethyl)-2,3,6,7,8,8a-hexahydropyrrolo[1,2-a]pyrazine-1,4-dione_a
3-(1-hydroxyethyl)-2,3,6,7,8,8a-hexahydropyrrolo[1,2-a]pyrazine-1,4-dione_a
3-(1-hydroxyethyl)-2,3,6,7,8,8a-hexahydropyrrolo[1,2-a]pyrazine-1,4-dione_a
3-(1-hydroxyethyl)-2,3,6,7,8,8a-hexahydropyrrolo[1,2-a]pyrazine-1,4-dione_a
3-(1-hydroxyethyl)-2,3,6,7,8,8a-hexahydropyrrolo[1,2-a]pyrazine-1,4-dione_a
Procaine_b
Procaine_b
Procaine_b
Procaine_b
Procaine_b
Acrylic acid
Acrylic acid
Acrylic acid
Acrylic acid
Acrylic acid
3,4-Methylenedioxyamphetamine
3,4-Methylenedioxyamphetamine
3,4-Methylenedioxyamphetamine
3,4-Methylenedioxyamphetamine
3,4-Methylenedioxyamphetamine
N-Acetylvaline_e
N-Acetylvaline_e
N-Acetylvaline_e
N-Acetylvaline_e
N-Acetylvaline_e
3,7-Dimethyluric acid

3,7-Dimethyluric acid
3,7-Dimethyluric acid
3,7-Dimethyluric acid
3,7-Dimethyluric acid
N~6~,N~6~-Dimethyllysine_b
N~6~,N~6~-Dimethyllysine_b
N~6~,N~6~-Dimethyllysine_b
N~6~,N~6~-Dimethyllysine_b
N~6~,N~6~-Dimethyllysine_b
CYS-ASP
CYS-ASP
CYS-ASP
CYS-ASP
CYS-ASP
Chenodeoxycholic acid 3-sulfate
Chenodeoxycholic acid 3-sulfate
Chenodeoxycholic acid 3-sulfate
Chenodeoxycholic acid 3-sulfate
Chenodeoxycholic acid 3-sulfate
Spermic acid_f
Spermic acid_f
Spermic acid_f
Spermic acid_f
Spermic acid_f
(2R,3S)-3-Hydroxy-8-methyl-8-azabicyclo[3.2.1]octane-2-carboxylic acid
(2R,3S)-3-Hydroxy-8-methyl-8-azabicyclo[3.2.1]octane-2-carboxylic acid
(2R,3S)-3-Hydroxy-8-methyl-8-azabicyclo[3.2.1]octane-2-carboxylic acid
(2R,3S)-3-Hydroxy-8-methyl-8-azabicyclo[3.2.1]octane-2-carboxylic acid
(2R,3S)-3-Hydroxy-8-methyl-8-azabicyclo[3.2.1]octane-2-carboxylic acid
4-Hydroxy-5-methylfuran-3(2H)-one
4-Hydroxy-5-methylfuran-3(2H)-one
4-Hydroxy-5-methylfuran-3(2H)-one
4-Hydroxy-5-methylfuran-3(2H)-one
4-Hydroxy-5-methylfuran-3(2H)-one
14-Hydroxy-3,9-dimethyl-12-methylene-5,8,10-trioxatetracyclo[9.3.1.0~1,9~.0~2,7~]pentadecane-6,13-dione
14-Hydroxy-3,9-dimethyl-12-methylene-5,8,10-trioxatetracyclo[9.3.1.0~1,9~.0~2,7~]pentadecane-6,13-dione
14-Hydroxy-3,9-dimethyl-12-methylene-5,8,10-trioxatetracyclo[9.3.1.0~1,9~.0~2,7~]pentadecane-6,13-dione
14-Hydroxy-3,9-dimethyl-12-methylene-5,8,10-trioxatetracyclo[9.3.1.0~1,9~.0~2,7~]pentadecane-6,13-dione
14-Hydroxy-3,9-dimethyl-12-methylene-5,8,10-trioxatetracyclo[9.3.1.0~1,9~.0~2,7~]pentadecane-6,13-dione
1-Vinylimidazole_b
1-Vinylimidazole_b
1-Vinylimidazole_b
1-Vinylimidazole_b
1-Vinylimidazole_b
Lanthionine ketimine_b
Lanthionine ketimine_b
Lanthionine ketimine_b
Lanthionine ketimine_b

Lanthionine ketimine_b
Tetraacetylethylenediamine_c
Tetraacetylethylenediamine_c
Tetraacetylethylenediamine_c
Tetraacetylethylenediamine_c
Tetraacetylethylenediamine_c
3-Phenylpropyl hydrogen sulfate
3-Phenylpropyl hydrogen sulfate
3-Phenylpropyl hydrogen sulfate
3-Phenylpropyl hydrogen sulfate
3-Phenylpropyl hydrogen sulfate
5beta-Cholestane-3alpha,7alpha,12alpha,25,26-pentol
5beta-Cholestane-3alpha,7alpha,12alpha,25,26-pentol
5beta-Cholestane-3alpha,7alpha,12alpha,25,26-pentol
5beta-Cholestane-3alpha,7alpha,12alpha,25,26-pentol
5beta-Cholestane-3alpha,7alpha,12alpha,25,26-pentol
pentoxyl
pentoxyl
pentoxyl
pentoxyl
pentoxyl
Vorinostat_a
Vorinostat_a
Vorinostat_a
Vorinostat_a
Vorinostat_a
GLK (Peptide  Gly-Leu-Lys))
GLK (Peptide  Gly-Leu-Lys))
GLK (Peptide  Gly-Leu-Lys))
GLK (Peptide  Gly-Leu-Lys))
GLK (Peptide  Gly-Leu-Lys))
Semilicoisoflavone B
Semilicoisoflavone B
Semilicoisoflavone B
Semilicoisoflavone B
Semilicoisoflavone B
Nonivamide
Nonivamide
Nonivamide
Nonivamide
Nonivamide
Dipivefrin
Dipivefrin
Dipivefrin
Dipivefrin
Dipivefrin
1,9-Nonanedithiol
1,9-Nonanedithiol

1,9-Nonanedithiol
1,9-Nonanedithiol
1,9-Nonanedithiol
Aminohippuric acid_b
Aminohippuric acid_b
Aminohippuric acid_b
Aminohippuric acid_b
Aminohippuric acid_b
IN00150
IN00150
IN00150
IN00150
IN00150
feruloylserotonin
feruloylserotonin
feruloylserotonin
feruloylserotonin
feruloylserotonin
Procaine_c
Procaine_c
Procaine_c
Procaine_c
Procaine_c
N(2)-succinyl-L-ornithine_a
N(2)-succinyl-L-ornithine_a
N(2)-succinyl-L-ornithine_a
N(2)-succinyl-L-ornithine_a
N(2)-succinyl-L-ornithine_a
Varanic acid
Varanic acid
Varanic acid
Varanic acid
Varanic acid
Arenaine
Arenaine
Arenaine
Arenaine
Arenaine
5-guanidino-2-oxopentanoic acid
5-guanidino-2-oxopentanoic acid
5-guanidino-2-oxopentanoic acid
5-guanidino-2-oxopentanoic acid
5-guanidino-2-oxopentanoic acid
4-Hydroxyproline
4-Hydroxyproline
4-Hydroxyproline
4-Hydroxyproline
4-Hydroxyproline

N-[(1R,2S,6R)-4-Azatricyclo[5.2.1.0~2,6~]dec-4-yl]-4-chloro-3-sulfamoylbenzamide
N-[(1R,2S,6R)-4-Azatricyclo[5.2.1.0~2,6~]dec-4-yl]-4-chloro-3-sulfamoylbenzamide
N-[(1R,2S,6R)-4-Azatricyclo[5.2.1.0~2,6~]dec-4-yl]-4-chloro-3-sulfamoylbenzamide
N-[(1R,2S,6R)-4-Azatricyclo[5.2.1.0~2,6~]dec-4-yl]-4-chloro-3-sulfamoylbenzamide
N-[(1R,2S,6R)-4-Azatricyclo[5.2.1.0~2,6~]dec-4-yl]-4-chloro-3-sulfamoylbenzamide
Tetrahydropapaveroline_b
Tetrahydropapaveroline_b
Tetrahydropapaveroline_b
Tetrahydropapaveroline_b
Tetrahydropapaveroline_b
N,N-Diethyl-4-methyl-1-piperazinecarboxamide 4-oxide_d
N,N-Diethyl-4-methyl-1-piperazinecarboxamide 4-oxide_d
N,N-Diethyl-4-methyl-1-piperazinecarboxamide 4-oxide_d
N,N-Diethyl-4-methyl-1-piperazinecarboxamide 4-oxide_d
N,N-Diethyl-4-methyl-1-piperazinecarboxamide 4-oxide_d
1_2-Dihydroxydibenzothiophene
1_2-Dihydroxydibenzothiophene
1_2-Dihydroxydibenzothiophene
1_2-Dihydroxydibenzothiophene
1_2-Dihydroxydibenzothiophene
N-(3-acetamidopropyl)pyrrolidin-2-one_a
N-(3-acetamidopropyl)pyrrolidin-2-one_a
N-(3-acetamidopropyl)pyrrolidin-2-one_a
N-(3-acetamidopropyl)pyrrolidin-2-one_a
N-(3-acetamidopropyl)pyrrolidin-2-one_a
LysoSM(d18:1)
LysoSM(d18:1)
LysoSM(d18:1)
LysoSM(d18:1)
LysoSM(d18:1)
1-(2-Hydroxy-4,6-dimethoxyphenyl)-3-(4-methoxyphenyl)-1-propanone
1-(2-Hydroxy-4,6-dimethoxyphenyl)-3-(4-methoxyphenyl)-1-propanone
1-(2-Hydroxy-4,6-dimethoxyphenyl)-3-(4-methoxyphenyl)-1-propanone
1-(2-Hydroxy-4,6-dimethoxyphenyl)-3-(4-methoxyphenyl)-1-propanone
1-(2-Hydroxy-4,6-dimethoxyphenyl)-3-(4-methoxyphenyl)-1-propanone
Ethyl acetate
Ethyl acetate
Ethyl acetate
Ethyl acetate
Ethyl acetate
Valylvaline_i
Valylvaline_i
Valylvaline_i
Valylvaline_i
Valylvaline_i
Valyl-4-hydroxyproline_b
Valyl-4-hydroxyproline_b
Valyl-4-hydroxyproline_b

Valyl-4-hydroxyproline_b
Valyl-4-hydroxyproline_b
Coprine_d
Coprine_d
Coprine_d
Coprine_d
Coprine_d
Ferulic acid
Ferulic acid
Ferulic acid
Ferulic acid
Ferulic acid
Prenisteine
Prenisteine
Prenisteine
Prenisteine
Prenisteine
N-[1-Carboxy-3-(methylsulfinyl)propyl]glutamine
N-[1-Carboxy-3-(methylsulfinyl)propyl]glutamine
N-[1-Carboxy-3-(methylsulfinyl)propyl]glutamine
N-[1-Carboxy-3-(methylsulfinyl)propyl]glutamine
N-[1-Carboxy-3-(methylsulfinyl)propyl]glutamine
Salicylic acid
Salicylic acid
Salicylic acid
Salicylic acid
Salicylic acid
4-[(E)-2-(3,4,5-Trimethoxyphenyl)vinyl]phenol_b
4-[(E)-2-(3,4,5-Trimethoxyphenyl)vinyl]phenol_b
4-[(E)-2-(3,4,5-Trimethoxyphenyl)vinyl]phenol_b
4-[(E)-2-(3,4,5-Trimethoxyphenyl)vinyl]phenol_b
4-[(E)-2-(3,4,5-Trimethoxyphenyl)vinyl]phenol_b
AAMU_a
AAMU_a
AAMU_a
AAMU_a
AAMU_a
IN00260_b
IN00260_b
IN00260_b
IN00260_b
IN00260_b
L-gamma-Glutamyl-L-valine_k
L-gamma-Glutamyl-L-valine_k
L-gamma-Glutamyl-L-valine_k
L-gamma-Glutamyl-L-valine_k
L-gamma-Glutamyl-L-valine_k
2-Keto-glutaramic acid

2-Keto-glutaramic acid
2-Keto-glutaramic acid
2-Keto-glutaramic acid
2-Keto-glutaramic acid
gamma-Glu-His
gamma-Glu-His
gamma-Glu-His
gamma-Glu-His
gamma-Glu-His
L-gamma-Glutamyl-L-valine_f
L-gamma-Glutamyl-L-valine_f
L-gamma-Glutamyl-L-valine_f
L-gamma-Glutamyl-L-valine_f
L-gamma-Glutamyl-L-valine_f
Aspartyl-L-proline_b
Aspartyl-L-proline_b
Aspartyl-L-proline_b
Aspartyl-L-proline_b
Aspartyl-L-proline_b
(4S)-4-[(2E)-2-Octenoyloxy]-4-(trimethylammonio)butanoate_a
(4S)-4-[(2E)-2-Octenoyloxy]-4-(trimethylammonio)butanoate_a
(4S)-4-[(2E)-2-Octenoyloxy]-4-(trimethylammonio)butanoate_a
(4S)-4-[(2E)-2-Octenoyloxy]-4-(trimethylammonio)butanoate_a
(4S)-4-[(2E)-2-Octenoyloxy]-4-(trimethylammonio)butanoate_a
4-(Nitrosoamino)-1-(3-pyridinyl)-1-butanol_b
4-(Nitrosoamino)-1-(3-pyridinyl)-1-butanol_b
4-(Nitrosoamino)-1-(3-pyridinyl)-1-butanol_b
4-(Nitrosoamino)-1-(3-pyridinyl)-1-butanol_b
4-(Nitrosoamino)-1-(3-pyridinyl)-1-butanol_b
MFCD02728197_a
MFCD02728197_a
MFCD02728197_a
MFCD02728197_a
MFCD02728197_a
Sular
Sular
Sular
Sular
Sular
butalbital_c
butalbital_c
butalbital_c
butalbital_c
butalbital_c
1,1'-[1,12-Dodecanediylbis(oxy)]dibenzene_e
1,1'-[1,12-Dodecanediylbis(oxy)]dibenzene_e
1,1'-[1,12-Dodecanediylbis(oxy)]dibenzene_e
1,1'-[1,12-Dodecanediylbis(oxy)]dibenzene_e

1,1'-[1,12-Dodecanediylbis(oxy)]dibenzene_e
5-(5-Methyl-2-furyl)-2,3-dihydro-1H-pyrrolizine
5-(5-Methyl-2-furyl)-2,3-dihydro-1H-pyrrolizine
5-(5-Methyl-2-furyl)-2,3-dihydro-1H-pyrrolizine
5-(5-Methyl-2-furyl)-2,3-dihydro-1H-pyrrolizine
5-(5-Methyl-2-furyl)-2,3-dihydro-1H-pyrrolizine
3-Deoxy-D-manno-octulosonic acid
3-Deoxy-D-manno-octulosonic acid
3-Deoxy-D-manno-octulosonic acid
3-Deoxy-D-manno-octulosonic acid
3-Deoxy-D-manno-octulosonic acid
2-(1-Naphthyl)acetamide_b
2-(1-Naphthyl)acetamide_b
2-(1-Naphthyl)acetamide_b
2-(1-Naphthyl)acetamide_b
2-(1-Naphthyl)acetamide_b
asn-pro_a
asn-pro_a
asn-pro_a
asn-pro_a
asn-pro_a
ophthalmic acid_b
ophthalmic acid_b
ophthalmic acid_b
ophthalmic acid_b
ophthalmic acid_b
2-Isopropyl-4,6-dimethyl-1,3,5-dithiazinane
2-Isopropyl-4,6-dimethyl-1,3,5-dithiazinane
2-Isopropyl-4,6-dimethyl-1,3,5-dithiazinane
2-Isopropyl-4,6-dimethyl-1,3,5-dithiazinane
2-Isopropyl-4,6-dimethyl-1,3,5-dithiazinane
L-gamma-Glutamyl-L-valine_c
L-gamma-Glutamyl-L-valine_c
L-gamma-Glutamyl-L-valine_c
L-gamma-Glutamyl-L-valine_c
L-gamma-Glutamyl-L-valine_c
2-(2-Hydroxy-3,4,5-trimethoxyphenyl)-7-methoxy-8-chromanol
2-(2-Hydroxy-3,4,5-trimethoxyphenyl)-7-methoxy-8-chromanol
2-(2-Hydroxy-3,4,5-trimethoxyphenyl)-7-methoxy-8-chromanol
2-(2-Hydroxy-3,4,5-trimethoxyphenyl)-7-methoxy-8-chromanol
2-(2-Hydroxy-3,4,5-trimethoxyphenyl)-7-methoxy-8-chromanol
N(1),N(8)-bis(coumaroyl)spermidine
N(1),N(8)-bis(coumaroyl)spermidine
N(1),N(8)-bis(coumaroyl)spermidine
N(1),N(8)-bis(coumaroyl)spermidine
N(1),N(8)-bis(coumaroyl)spermidine
Spermic acid_g
Spermic acid_g

Spermic acid_g
Spermic acid_g
Spermic acid_g
2-Hydroxy-5-[(5-oxotetrahydro-2-furanyl)methyl]phenyl hydrogen sulfate
2-Hydroxy-5-[(5-oxotetrahydro-2-furanyl)methyl]phenyl hydrogen sulfate
2-Hydroxy-5-[(5-oxotetrahydro-2-furanyl)methyl]phenyl hydrogen sulfate
2-Hydroxy-5-[(5-oxotetrahydro-2-furanyl)methyl]phenyl hydrogen sulfate
2-Hydroxy-5-[(5-oxotetrahydro-2-furanyl)methyl]phenyl hydrogen sulfate
4-(METHYLNITROSAMINO)-1-(3-PYRIDYL-N-OXIDE)-1-BUTANOL_a
4-(METHYLNITROSAMINO)-1-(3-PYRIDYL-N-OXIDE)-1-BUTANOL_a
4-(METHYLNITROSAMINO)-1-(3-PYRIDYL-N-OXIDE)-1-BUTANOL_a
4-(METHYLNITROSAMINO)-1-(3-PYRIDYL-N-OXIDE)-1-BUTANOL_a
4-(METHYLNITROSAMINO)-1-(3-PYRIDYL-N-OXIDE)-1-BUTANOL_a
(-)-Physostigmine_a
(-)-Physostigmine_a
(-)-Physostigmine_a
(-)-Physostigmine_a
(-)-Physostigmine_a
N-(3,5-Dimethoxybenzoyl)glycine_a
N-(3,5-Dimethoxybenzoyl)glycine_a
N-(3,5-Dimethoxybenzoyl)glycine_a
N-(3,5-Dimethoxybenzoyl)glycine_a
N-(3,5-Dimethoxybenzoyl)glycine_a
Homoanserine_b
Homoanserine_b
Homoanserine_b
Homoanserine_b
Homoanserine_b
S(6)-acetyldihydrolipoamide
S(6)-acetyldihydrolipoamide
S(6)-acetyldihydrolipoamide
S(6)-acetyldihydrolipoamide
S(6)-acetyldihydrolipoamide
4-Methyleneglutamic acid
4-Methyleneglutamic acid
4-Methyleneglutamic acid
4-Methyleneglutamic acid
4-Methyleneglutamic acid
asn-phe
asn-phe
asn-phe
asn-phe
asn-phe
2-O-beta-D-Galactopyranosyl-D-xylopyranose
2-O-beta-D-Galactopyranosyl-D-xylopyranose
2-O-beta-D-Galactopyranosyl-D-xylopyranose
2-O-beta-D-Galactopyranosyl-D-xylopyranose
2-O-beta-D-Galactopyranosyl-D-xylopyranose

N-Propionylmethionine_a
N-Propionylmethionine_a
N-Propionylmethionine_a
N-Propionylmethionine_a
N-Propionylmethionine_a
N4-(beta-N-Acetyl-D-glucosaminyl)-L-asparagine
N4-(beta-N-Acetyl-D-glucosaminyl)-L-asparagine
N4-(beta-N-Acetyl-D-glucosaminyl)-L-asparagine
N4-(beta-N-Acetyl-D-glucosaminyl)-L-asparagine
N4-(beta-N-Acetyl-D-glucosaminyl)-L-asparagine
3-Methoxy-4-hydroxyhippuric acid_d
3-Methoxy-4-hydroxyhippuric acid_d
3-Methoxy-4-hydroxyhippuric acid_d
3-Methoxy-4-hydroxyhippuric acid_d
3-Methoxy-4-hydroxyhippuric acid_d
Spermic acid_e
Spermic acid_e
Spermic acid_e
Spermic acid_e
Spermic acid_e
L-gamma-Glutamyl-L-valine_l
L-gamma-Glutamyl-L-valine_l
L-gamma-Glutamyl-L-valine_l
L-gamma-Glutamyl-L-valine_l
L-gamma-Glutamyl-L-valine_l
N-(1H-Pyrrol-2-ylcarbonyl)glycine
N-(1H-Pyrrol-2-ylcarbonyl)glycine
N-(1H-Pyrrol-2-ylcarbonyl)glycine
N-(1H-Pyrrol-2-ylcarbonyl)glycine
N-(1H-Pyrrol-2-ylcarbonyl)glycine
N-(3,5-Dimethoxybenzoyl)glycine_e
N-(3,5-Dimethoxybenzoyl)glycine_e
N-(3,5-Dimethoxybenzoyl)glycine_e
N-(3,5-Dimethoxybenzoyl)glycine_e
N-(3,5-Dimethoxybenzoyl)glycine_e
N-{3-[(4-Acetamidobutyl)amino]propyl}acetamide_a
N-{3-[(4-Acetamidobutyl)amino]propyl}acetamide_a
N-{3-[(4-Acetamidobutyl)amino]propyl}acetamide_a
N-{3-[(4-Acetamidobutyl)amino]propyl}acetamide_a
N-{3-[(4-Acetamidobutyl)amino]propyl}acetamide_a
3-hydroxy-3-methyloxindole_a
3-hydroxy-3-methyloxindole_a
3-hydroxy-3-methyloxindole_a
3-hydroxy-3-methyloxindole_a
3-hydroxy-3-methyloxindole_a
tyramine sulfate_b
tyramine sulfate_b
tyramine sulfate_b

tyramine sulfate_b
tyramine sulfate_b
N-Hydroxy-2-acetamidofluorene
N-Hydroxy-2-acetamidofluorene
N-Hydroxy-2-acetamidofluorene
N-Hydroxy-2-acetamidofluorene
N-Hydroxy-2-acetamidofluorene
Endothal
Endothal
Endothal
Endothal
Endothal
2,4,6-Triisobutyl-1,3,5-dithiazinane
2,4,6-Triisobutyl-1,3,5-dithiazinane
2,4,6-Triisobutyl-1,3,5-dithiazinane
2,4,6-Triisobutyl-1,3,5-dithiazinane
2,4,6-Triisobutyl-1,3,5-dithiazinane
N-(3,5-Dimethoxybenzoyl)glycine_b
N-(3,5-Dimethoxybenzoyl)glycine_b
N-(3,5-Dimethoxybenzoyl)glycine_b
N-(3,5-Dimethoxybenzoyl)glycine_b
N-(3,5-Dimethoxybenzoyl)glycine_b
TOLMETIN
TOLMETIN
TOLMETIN
TOLMETIN
TOLMETIN
linatine
linatine
linatine
linatine
linatine
2,3,4,5,6-Pentahydroxy-N-(2-hydroxyethyl)hexanamide
2,3,4,5,6-Pentahydroxy-N-(2-hydroxyethyl)hexanamide
2,3,4,5,6-Pentahydroxy-N-(2-hydroxyethyl)hexanamide
2,3,4,5,6-Pentahydroxy-N-(2-hydroxyethyl)hexanamide
2,3,4,5,6-Pentahydroxy-N-(2-hydroxyethyl)hexanamide
UK3870000
UK3870000
UK3870000
UK3870000
UK3870000
(+)-Etomidate_b
(+)-Etomidate_b
(+)-Etomidate_b
(+)-Etomidate_b
(+)-Etomidate_b
7,8-Didehydro-4,5-epoxymorphinan-3,6-diol

7,8-Didehydro-4,5-epoxymorphinan-3,6-diol
7,8-Didehydro-4,5-epoxymorphinan-3,6-diol
7,8-Didehydro-4,5-epoxymorphinan-3,6-diol
7,8-Didehydro-4,5-epoxymorphinan-3,6-diol
LW8000000_b
LW8000000_b
LW8000000_b
LW8000000_b
LW8000000_b
N(2)-succinyl-L-ornithine_b
N(2)-succinyl-L-ornithine_b
N(2)-succinyl-L-ornithine_b
N(2)-succinyl-L-ornithine_b
N(2)-succinyl-L-ornithine_b
3-(2,3-Dihydroxy-3-methylbutyl)-4-methoxy-1-methyl-2(1H)-quinolinone_a
3-(2,3-Dihydroxy-3-methylbutyl)-4-methoxy-1-methyl-2(1H)-quinolinone_a
3-(2,3-Dihydroxy-3-methylbutyl)-4-methoxy-1-methyl-2(1H)-quinolinone_a
3-(2,3-Dihydroxy-3-methylbutyl)-4-methoxy-1-methyl-2(1H)-quinolinone_a
3-(2,3-Dihydroxy-3-methylbutyl)-4-methoxy-1-methyl-2(1H)-quinolinone_a
Formylkynurenine_a
Formylkynurenine_a
Formylkynurenine_a
Formylkynurenine_a
Formylkynurenine_a
indoline-2-carboxylic acid
indoline-2-carboxylic acid
indoline-2-carboxylic acid
indoline-2-carboxylic acid
indoline-2-carboxylic acid
{[(15-Hydroxy-3-methoxy-1,2,6,7-tetradehydroerythrinan-16-yl)oxy]sulfonyl}acetic acid
{[(15-Hydroxy-3-methoxy-1,2,6,7-tetradehydroerythrinan-16-yl)oxy]sulfonyl}acetic acid
{[(15-Hydroxy-3-methoxy-1,2,6,7-tetradehydroerythrinan-16-yl)oxy]sulfonyl}acetic acid
{[(15-Hydroxy-3-methoxy-1,2,6,7-tetradehydroerythrinan-16-yl)oxy]sulfonyl}acetic acid
{[(15-Hydroxy-3-methoxy-1,2,6,7-tetradehydroerythrinan-16-yl)oxy]sulfonyl}acetic acid
(7E,7'E)-5,5'-diferulic acid_b
(7E,7'E)-5,5'-diferulic acid_b
(7E,7'E)-5,5'-diferulic acid_b
(7E,7'E)-5,5'-diferulic acid_b
(7E,7'E)-5,5'-diferulic acid_b
MFCD01664827
MFCD01664827
MFCD01664827
MFCD01664827
MFCD01664827
Estrone glucuronide
Estrone glucuronide
Estrone glucuronide
Estrone glucuronide

Estrone glucuronide
ELK (Peptide Glu-Leu-Lys)_a
ELK (Peptide Glu-Leu-Lys)_a
ELK (Peptide Glu-Leu-Lys)_a
ELK (Peptide Glu-Leu-Lys)_a
ELK (Peptide Glu-Leu-Lys)_a
Tetrahydro-2-furanylmethyl hydrogen sulfate
Tetrahydro-2-furanylmethyl hydrogen sulfate
Tetrahydro-2-furanylmethyl hydrogen sulfate
Tetrahydro-2-furanylmethyl hydrogen sulfate
Tetrahydro-2-furanylmethyl hydrogen sulfate
2-(3-CARBOXYPROPIONYL)-6-HYDROXY-CYCLOHEXA-2,4-DIENE CARBOXYLIC ACID_a
2-(3-CARBOXYPROPIONYL)-6-HYDROXY-CYCLOHEXA-2,4-DIENE CARBOXYLIC ACID_a
2-(3-CARBOXYPROPIONYL)-6-HYDROXY-CYCLOHEXA-2,4-DIENE CARBOXYLIC ACID_a
2-(3-CARBOXYPROPIONYL)-6-HYDROXY-CYCLOHEXA-2,4-DIENE CARBOXYLIC ACID_a
2-(3-CARBOXYPROPIONYL)-6-HYDROXY-CYCLOHEXA-2,4-DIENE CARBOXYLIC ACID_a
Queuosine
Queuosine
Queuosine
Queuosine
Queuosine
Methyl alpha-aspartylphenylalaninate_b
Methyl alpha-aspartylphenylalaninate_b
Methyl alpha-aspartylphenylalaninate_b
Methyl alpha-aspartylphenylalaninate_b
Methyl alpha-aspartylphenylalaninate_b
Zalcitabine_g
Zalcitabine_g
Zalcitabine_g
Zalcitabine_g
Zalcitabine_g
(2S)-6-Amino-2-[(E)-(hydroxymethylene)amino]hexanimidic acid_a
(2S)-6-Amino-2-[(E)-(hydroxymethylene)amino]hexanimidic acid_a
(2S)-6-Amino-2-[(E)-(hydroxymethylene)amino]hexanimidic acid_a
(2S)-6-Amino-2-[(E)-(hydroxymethylene)amino]hexanimidic acid_a
(2S)-6-Amino-2-[(E)-(hydroxymethylene)amino]hexanimidic acid_a
3-Methyladenine_c
3-Methyladenine_c
3-Methyladenine_c
3-Methyladenine_c
3-Methyladenine_c
N-Benzyl-3-[(2S,5aS,8aR)-6-(1H-imidazol-2-ylmethyl)-1-methyl-5-oxodecahydropyrrolo[3,2-E][1,4]diazepin-2-yl]
N-Benzyl-3-[(2S,5aS,8aR)-6-(1H-imidazol-2-ylmethyl)-1-methyl-5-oxodecahydropyrrolo[3,2-E][1,4]diazepin-2-yl]
N-Benzyl-3-[(2S,5aS,8aR)-6-(1H-imidazol-2-ylmethyl)-1-methyl-5-oxodecahydropyrrolo[3,2-E][1,4]diazepin-2-yl]
N-Benzyl-3-[(2S,5aS,8aR)-6-(1H-imidazol-2-ylmethyl)-1-methyl-5-oxodecahydropyrrolo[3,2-E][1,4]diazepin-2-yl]
N-Benzyl-3-[(2S,5aS,8aR)-6-(1H-imidazol-2-ylmethyl)-1-methyl-5-oxodecahydropyrrolo[3,2-E][1,4]diazepin-2-yl]
4-[(E)-2-(3,4,5-Trimethoxyphenyl)vinyl]phenol_a
4-[(E)-2-(3,4,5-Trimethoxyphenyl)vinyl]phenol_a

4-[(E)-2-(3,4,5-Trimethoxyphenyl)vinyl]phenol_a
4-[(E)-2-(3,4,5-Trimethoxyphenyl)vinyl]phenol_a
4-[(E)-2-(3,4,5-Trimethoxyphenyl)vinyl]phenol_a
Harmane
Harmane
Harmane
Harmane
Harmane
1-(beta-D-ribofuranosyl)thymine_b
1-(beta-D-ribofuranosyl)thymine_b
1-(beta-D-ribofuranosyl)thymine_b
1-(beta-D-ribofuranosyl)thymine_b
1-(beta-D-ribofuranosyl)thymine_b
N(6),N(6)-Dimethyladenine
N(6),N(6)-Dimethyladenine
N(6),N(6)-Dimethyladenine
N(6),N(6)-Dimethyladenine
N(6),N(6)-Dimethyladenine
MC0555300
MC0555300
MC0555300
MC0555300
MC0555300
4-ethylphenylsulfonic acid
4-ethylphenylsulfonic acid
4-ethylphenylsulfonic acid
4-ethylphenylsulfonic acid
4-ethylphenylsulfonic acid
5-Hydroxy-N-[(6-oxo-2-piperidinyl)methyl]-2-(2,2,2-trifluoroethoxy)benzamide
5-Hydroxy-N-[(6-oxo-2-piperidinyl)methyl]-2-(2,2,2-trifluoroethoxy)benzamide
5-Hydroxy-N-[(6-oxo-2-piperidinyl)methyl]-2-(2,2,2-trifluoroethoxy)benzamide
5-Hydroxy-N-[(6-oxo-2-piperidinyl)methyl]-2-(2,2,2-trifluoroethoxy)benzamide
5-Hydroxy-N-[(6-oxo-2-piperidinyl)methyl]-2-(2,2,2-trifluoroethoxy)benzamide
g-Aminobutyryl-lysine_b
g-Aminobutyryl-lysine_b
g-Aminobutyryl-lysine_b
g-Aminobutyryl-lysine_b
g-Aminobutyryl-lysine_b
pentobarbital_e
pentobarbital_e
pentobarbital_e
pentobarbital_e
pentobarbital_e
Spermic acid_b
Spermic acid_b
Spermic acid_b
Spermic acid_b
Spermic acid_b

3-Hydroxyhexadecadienoylcarnitine
3-Hydroxyhexadecadienoylcarnitine
3-Hydroxyhexadecadienoylcarnitine
3-Hydroxyhexadecadienoylcarnitine
3-Hydroxyhexadecadienoylcarnitine
5beta-Cyprinolsulfate
5beta-Cyprinolsulfate
5beta-Cyprinolsulfate
5beta-Cyprinolsulfate
5beta-Cyprinolsulfate
Iminoglycine
Iminoglycine
Iminoglycine
Iminoglycine
Iminoglycine
Biocytin
Biocytin
Biocytin
Biocytin
Biocytin
3-Methyladenine_a
3-Methyladenine_a
3-Methyladenine_a
3-Methyladenine_a
3-Methyladenine_a
TLK (Peptide Thr-Leu-Lys)
TLK (Peptide Thr-Leu-Lys)
TLK (Peptide Thr-Leu-Lys)
TLK (Peptide Thr-Leu-Lys)
TLK (Peptide Thr-Leu-Lys)
N-(4-Heptanyl)-1,3-benzodioxole-5-carboxamide
N-(4-Heptanyl)-1,3-benzodioxole-5-carboxamide
N-(4-Heptanyl)-1,3-benzodioxole-5-carboxamide
N-(4-Heptanyl)-1,3-benzodioxole-5-carboxamide
N-(4-Heptanyl)-1,3-benzodioxole-5-carboxamide
N-Acetylvaline_c
N-Acetylvaline_c
N-Acetylvaline_c
N-Acetylvaline_c
N-Acetylvaline_c
N-[(10Z)-7-Isobutyl-3-isopropyl-5,8-dioxo-2-oxa-6,9-diazabicyclo[10.2.2]hexadeca-1(14),10,12,15-tetraen-4-yl]-[illegible]
N-[(10Z)-7-Isobutyl-3-isopropyl-5,8-dioxo-2-oxa-6,9-diazabicyclo[10.2.2]hexadeca-1(14),10,12,15-tetraen-4-yl]-[illegible]
N-[(10Z)-7-Isobutyl-3-isopropyl-5,8-dioxo-2-oxa-6,9-diazabicyclo[10.2.2]hexadeca-1(14),10,12,15-tetraen-4-yl]-[illegible]
N-[(10Z)-7-Isobutyl-3-isopropyl-5,8-dioxo-2-oxa-6,9-diazabicyclo[10.2.2]hexadeca-1(14),10,12,15-tetraen-4-yl]-[illegible]
N-[(10Z)-7-Isobutyl-3-isopropyl-5,8-dioxo-2-oxa-6,9-diazabicyclo[10.2.2]hexadeca-1(14),10,12,15-tetraen-4-yl]-[illegible]
Midodrine_e
Midodrine_e
Midodrine_e

Midodrine_e
Midodrine_e
2-[4-(3-Hydroxypropyl)-2-methoxyphenoxy]-1,3-propanediol
2-[4-(3-Hydroxypropyl)-2-methoxyphenoxy]-1,3-propanediol
2-[4-(3-Hydroxypropyl)-2-methoxyphenoxy]-1,3-propanediol
2-[4-(3-Hydroxypropyl)-2-methoxyphenoxy]-1,3-propanediol
2-[4-(3-Hydroxypropyl)-2-methoxyphenoxy]-1,3-propanediol
4,9a-Dimethoxy-3,4a,5-trimethyl-2-oxo-2,4,4a,5,6,7,8,8a,9,9a-decahydronaphtho[2,3-b]furan-6-yl (2E)-3-(methy
4,9a-Dimethoxy-3,4a,5-trimethyl-2-oxo-2,4,4a,5,6,7,8,8a,9,9a-decahydronaphtho[2,3-b]furan-6-yl (2E)-3-(methy
4,9a-Dimethoxy-3,4a,5-trimethyl-2-oxo-2,4,4a,5,6,7,8,8a,9,9a-decahydronaphtho[2,3-b]furan-6-yl (2E)-3-(methy
4,9a-Dimethoxy-3,4a,5-trimethyl-2-oxo-2,4,4a,5,6,7,8,8a,9,9a-decahydronaphtho[2,3-b]furan-6-yl (2E)-3-(methy
4,9a-Dimethoxy-3,4a,5-trimethyl-2-oxo-2,4,4a,5,6,7,8,8a,9,9a-decahydronaphtho[2,3-b]furan-6-yl (2E)-3-(methy
epsilon-(gamma-Glutamyl)-lysine_b
epsilon-(gamma-Glutamyl)-lysine_b
epsilon-(gamma-Glutamyl)-lysine_b
epsilon-(gamma-Glutamyl)-lysine_b
epsilon-(gamma-Glutamyl)-lysine_b
Nimodipine
Nimodipine
Nimodipine
Nimodipine
Nimodipine
his-gln_a
his-gln_a
his-gln_a
his-gln_a
his-gln_a
Dehydroacetic acid
Dehydroacetic acid
Dehydroacetic acid
Dehydroacetic acid
Dehydroacetic acid
trimethadione
trimethadione
trimethadione
trimethadione
trimethadione
(2E,6E)-9-[(2R)-6-Hydroxy-2,5,7,8-tetramethyl-3,4-dihydro-2H-chromen-2-yl]-2,6-dimethyl-2,6-nonadienoic acid
(2E,6E)-9-[(2R)-6-Hydroxy-2,5,7,8-tetramethyl-3,4-dihydro-2H-chromen-2-yl]-2,6-dimethyl-2,6-nonadienoic acid
(2E,6E)-9-[(2R)-6-Hydroxy-2,5,7,8-tetramethyl-3,4-dihydro-2H-chromen-2-yl]-2,6-dimethyl-2,6-nonadienoic acid
(2E,6E)-9-[(2R)-6-Hydroxy-2,5,7,8-tetramethyl-3,4-dihydro-2H-chromen-2-yl]-2,6-dimethyl-2,6-nonadienoic acid
(2E,6E)-9-[(2R)-6-Hydroxy-2,5,7,8-tetramethyl-3,4-dihydro-2H-chromen-2-yl]-2,6-dimethyl-2,6-nonadienoic acid
Coixol
Coixol
Coixol
Coixol
Coixol
Methyl 2,3-dihydro-3-hydroxy-2-oxo-1H-indole-3-acetate_b

Methyl 2,3-dihydro-3-hydroxy-2-oxo-1H-indole-3-acetate_b
Methyl 2,3-dihydro-3-hydroxy-2-oxo-1H-indole-3-acetate_b
Methyl 2,3-dihydro-3-hydroxy-2-oxo-1H-indole-3-acetate_b
Methyl 2,3-dihydro-3-hydroxy-2-oxo-1H-indole-3-acetate_b
ala-met
ala-met
ala-met
ala-met
ala-met
Leu-pro_b
Leu-pro_b
Leu-pro_b
Leu-pro_b
Leu-pro_b
SECONAL_e
SECONAL_e
SECONAL_e
SECONAL_e
SECONAL_e
N-(4-Hydroxy-3,5-dimethoxybenzoyl)glycine_a
N-(4-Hydroxy-3,5-dimethoxybenzoyl)glycine_a
N-(4-Hydroxy-3,5-dimethoxybenzoyl)glycine_a
N-(4-Hydroxy-3,5-dimethoxybenzoyl)glycine_a
N-(4-Hydroxy-3,5-dimethoxybenzoyl)glycine_a
1-Methyl-1,2,3,4-tetrahydro-ÃŽÂ$^{2}$-carboline-3-carboxylic acid_a
1-Methyl-1,2,3,4-tetrahydro-ÃŽÂ$^{2}$-carboline-3-carboxylic acid_a
1-Methyl-1,2,3,4-tetrahydro-ÃŽÂ$^{2}$-carboline-3-carboxylic acid_a
1-Methyl-1,2,3,4-tetrahydro-ÃŽÂ$^{2}$-carboline-3-carboxylic acid_a
1-Methyl-1,2,3,4-tetrahydro-ÃŽÂ$^{2}$-carboline-3-carboxylic acid_a
imazamethabenz-methyl_b
imazamethabenz-methyl_b
imazamethabenz-methyl_b
imazamethabenz-methyl_b
imazamethabenz-methyl_b
cys-met
cys-met
cys-met
cys-met
cys-met
2-(3-CARBOXYPROPIONYL)-6-HYDROXY-CYCLOHEXA-2,4-DIENE CARBOXYLIC ACID_b
2-(3-CARBOXYPROPIONYL)-6-HYDROXY-CYCLOHEXA-2,4-DIENE CARBOXYLIC ACID_b
2-(3-CARBOXYPROPIONYL)-6-HYDROXY-CYCLOHEXA-2,4-DIENE CARBOXYLIC ACID_b
2-(3-CARBOXYPROPIONYL)-6-HYDROXY-CYCLOHEXA-2,4-DIENE CARBOXYLIC ACID_b
2-(3-CARBOXYPROPIONYL)-6-HYDROXY-CYCLOHEXA-2,4-DIENE CARBOXYLIC ACID_b
Quinaldic acid
Quinaldic acid
Quinaldic acid
Quinaldic acid

Quinaldic acid
Gly-DL-Phe
Gly-DL-Phe
Gly-DL-Phe
Gly-DL-Phe
Gly-DL-Phe
4-Formyl-2-methoxyphenyl hydrogen sulfate
4-Formyl-2-methoxyphenyl hydrogen sulfate
4-Formyl-2-methoxyphenyl hydrogen sulfate
4-Formyl-2-methoxyphenyl hydrogen sulfate
4-Formyl-2-methoxyphenyl hydrogen sulfate
L-gamma-Glutamyl-L-valine_h
L-gamma-Glutamyl-L-valine_h
L-gamma-Glutamyl-L-valine_h
L-gamma-Glutamyl-L-valine_h
L-gamma-Glutamyl-L-valine_h
Imidazolelactic acid
Imidazolelactic acid
Imidazolelactic acid
Imidazolelactic acid
Imidazolelactic acid
Ethynodiol diacetate
Ethynodiol diacetate
Ethynodiol diacetate
Ethynodiol diacetate
Ethynodiol diacetate
(+)-Etomidate_a
(+)-Etomidate_a
(+)-Etomidate_a
(+)-Etomidate_a
(+)-Etomidate_a
3-methyl-4-(methylamino)-1,2-diphenylbutan-2-yl propanoate
3-methyl-4-(methylamino)-1,2-diphenylbutan-2-yl propanoate
3-methyl-4-(methylamino)-1,2-diphenylbutan-2-yl propanoate
3-methyl-4-(methylamino)-1,2-diphenylbutan-2-yl propanoate
3-methyl-4-(methylamino)-1,2-diphenylbutan-2-yl propanoate
Maleamate
Maleamate
Maleamate
Maleamate
Maleamate
Lys-Pro_d
Lys-Pro_d
Lys-Pro_d
Lys-Pro_d
Lys-Pro_d
2,3,8,9-Tetrahydroxybenzo[b][1]benzofuro[2,3-f][1]benzofuran-6,12-dione
2,3,8,9-Tetrahydroxybenzo[b][1]benzofuro[2,3-f][1]benzofuran-6,12-dione

2,3,8,9-Tetrahydroxybenzo[b][1]benzofuro[2,3-f][1]benzofuran-6,12-dione
2,3,8,9-Tetrahydroxybenzo[b][1]benzofuro[2,3-f][1]benzofuran-6,12-dione
2,3,8,9-Tetrahydroxybenzo[b][1]benzofuro[2,3-f][1]benzofuran-6,12-dione
4-morpholinobenzoic acid
4-morpholinobenzoic acid
4-morpholinobenzoic acid
4-morpholinobenzoic acid
4-morpholinobenzoic acid
1-Methyl-1,2,3,4-tetrahydro-ÃŽÂ²-carboline-3-carboxylic acid_b
1-Methyl-1,2,3,4-tetrahydro-ÃŽÂ²-carboline-3-carboxylic acid_b
1-Methyl-1,2,3,4-tetrahydro-ÃŽÂ²-carboline-3-carboxylic acid_b
1-Methyl-1,2,3,4-tetrahydro-ÃŽÂ²-carboline-3-carboxylic acid_b
1-Methyl-1,2,3,4-tetrahydro-ÃŽÂ²-carboline-3-carboxylic acid_b
Lovastatin_b
Lovastatin_b
Lovastatin_b
Lovastatin_b
Lovastatin_b
Sulfurous acid
Sulfurous acid
Sulfurous acid
Sulfurous acid
Sulfurous acid
4H-1-Benzopyran-4-one, 6-Î²-D-glucopyranosyl-2,3-dihydro-5,7-dihydroxy-2-(4-hydroxyphenyl)-, (S)-
4H-1-Benzopyran-4-one, 6-Î²-D-glucopyranosyl-2,3-dihydro-5,7-dihydroxy-2-(4-hydroxyphenyl)-, (S)-
4H-1-Benzopyran-4-one, 6-Î²-D-glucopyranosyl-2,3-dihydro-5,7-dihydroxy-2-(4-hydroxyphenyl)-, (S)-
4H-1-Benzopyran-4-one, 6-Î²-D-glucopyranosyl-2,3-dihydro-5,7-dihydroxy-2-(4-hydroxyphenyl)-, (S)-
4H-1-Benzopyran-4-one, 6-Î²-D-glucopyranosyl-2,3-dihydro-5,7-dihydroxy-2-(4-hydroxyphenyl)-, (S)-
S-Allylcysteine_a
S-Allylcysteine_a
S-Allylcysteine_a
S-Allylcysteine_a
S-Allylcysteine_a
L-gamma-Glutamyl-L-leucine_c
L-gamma-Glutamyl-L-leucine_c
L-gamma-Glutamyl-L-leucine_c
L-gamma-Glutamyl-L-leucine_c
L-gamma-Glutamyl-L-leucine_c
2-(1-Naphthyl)acetamide_a
2-(1-Naphthyl)acetamide_a
2-(1-Naphthyl)acetamide_a
2-(1-Naphthyl)acetamide_a
2-(1-Naphthyl)acetamide_a
L-Arogenate
L-Arogenate
L-Arogenate
L-Arogenate
L-Arogenate

4-Indolecarbaldehyde
4-Indolecarbaldehyde
4-Indolecarbaldehyde
4-Indolecarbaldehyde
4-Indolecarbaldehyde
Phenyl D-glucopyranosiduronic acid_c
Phenyl D-glucopyranosiduronic acid_c
Phenyl D-glucopyranosiduronic acid_c
Phenyl D-glucopyranosiduronic acid_c
Phenyl D-glucopyranosiduronic acid_c
Zalcitabine_d
Zalcitabine_d
Zalcitabine_d
Zalcitabine_d
Zalcitabine_d
L-N2-(2-Carboxyethyl)arginine_b
L-N2-(2-Carboxyethyl)arginine_b
L-N2-(2-Carboxyethyl)arginine_b
L-N2-(2-Carboxyethyl)arginine_b
L-N2-(2-Carboxyethyl)arginine_b
Coenzyme M
Coenzyme M
Coenzyme M
Coenzyme M
Coenzyme M
Leucylasparagine_a
Leucylasparagine_a
Leucylasparagine_a
Leucylasparagine_a
Leucylasparagine_a
Ro 20-1724_d
Ro 20-1724_d
Ro 20-1724_d
Ro 20-1724_d
Ro 20-1724_d
3-Methylxanthine
3-Methylxanthine
3-Methylxanthine
3-Methylxanthine
3-Methylxanthine
beta-D-GlcpA-(1->6)-beta-D-Galp
beta-D-GlcpA-(1->6)-beta-D-Galp
beta-D-GlcpA-(1->6)-beta-D-Galp
beta-D-GlcpA-(1->6)-beta-D-Galp
beta-D-GlcpA-(1->6)-beta-D-Galp
L-Saccharopine
L-Saccharopine
L-Saccharopine

L-Saccharopine
L-Saccharopine
1,5-Isoquinolinediol_a
1,5-Isoquinolinediol_a
1,5-Isoquinolinediol_a
1,5-Isoquinolinediol_a
1,5-Isoquinolinediol_a
(6alpha,11beta)-6,9-Difluoro-11,21-dihydroxy-3,20-dioxopregna-1,4-dien-17-yl butyrate
(6alpha,11beta)-6,9-Difluoro-11,21-dihydroxy-3,20-dioxopregna-1,4-dien-17-yl butyrate
(6alpha,11beta)-6,9-Difluoro-11,21-dihydroxy-3,20-dioxopregna-1,4-dien-17-yl butyrate
(6alpha,11beta)-6,9-Difluoro-11,21-dihydroxy-3,20-dioxopregna-1,4-dien-17-yl butyrate
(6alpha,11beta)-6,9-Difluoro-11,21-dihydroxy-3,20-dioxopregna-1,4-dien-17-yl butyrate
2-(Carboxyacetamido)benzoic acid_a
2-(Carboxyacetamido)benzoic acid_a
2-(Carboxyacetamido)benzoic acid_a
2-(Carboxyacetamido)benzoic acid_a
2-(Carboxyacetamido)benzoic acid_a
Ro 20-1724_c
Ro 20-1724_c
Ro 20-1724_c
Ro 20-1724_c
Ro 20-1724_c
8-Amino-7-oxononanoic acid_d
8-Amino-7-oxononanoic acid_d
8-Amino-7-oxononanoic acid_d
8-Amino-7-oxononanoic acid_d
8-Amino-7-oxononanoic acid_d
glu-ser
glu-ser
glu-ser
glu-ser
glu-ser
GAMMA-HYDROXYHOMOARGININE
GAMMA-HYDROXYHOMOARGININE
GAMMA-HYDROXYHOMOARGININE
GAMMA-HYDROXYHOMOARGININE
GAMMA-HYDROXYHOMOARGININE
Coumarin
Coumarin
Coumarin
Coumarin
Coumarin
4-Guanidinobutyric acid
4-Guanidinobutyric acid
4-Guanidinobutyric acid
4-Guanidinobutyric acid
4-Guanidinobutyric acid
26Q0EO75R3

26Q0EO75R3
26Q0EO75R3
26Q0EO75R3
26Q0EO75R3
4-Guanidinobutanal
4-Guanidinobutanal
4-Guanidinobutanal
4-Guanidinobutanal
4-Guanidinobutanal
APM_a
APM_a
APM_a
APM_a
APM_a
dopaquinone
dopaquinone
dopaquinone
dopaquinone
dopaquinone
ophthalmic acid_c
ophthalmic acid_c
ophthalmic acid_c
ophthalmic acid_c
ophthalmic acid_c
MFCD00025555_e
MFCD00025555_e
MFCD00025555_e
MFCD00025555_e
MFCD00025555_e
L-N2-(2-Carboxyethyl)arginine_a
L-N2-(2-Carboxyethyl)arginine_a
L-N2-(2-Carboxyethyl)arginine_a
L-N2-(2-Carboxyethyl)arginine_a
L-N2-(2-Carboxyethyl)arginine_a
CMPF
CMPF
CMPF
CMPF
CMPF
Glaucine
Glaucine
Glaucine
Glaucine
Glaucine
pentobarbital_b
pentobarbital_b
pentobarbital_b
pentobarbital_b

pentobarbital_b
Glycine
Glycine
Glycine
Glycine
Glycine
2-(1-Ethoxyethoxy)propanoic acid_a
2-(1-Ethoxyethoxy)propanoic acid_a
2-(1-Ethoxyethoxy)propanoic acid_a
2-(1-Ethoxyethoxy)propanoic acid_a
2-(1-Ethoxyethoxy)propanoic acid_a
Leu-Leu_a
Leu-Leu_a
Leu-Leu_a
Leu-Leu_a
Leu-Leu_a
(8)-Gingerol
(8)-Gingerol
(8)-Gingerol
(8)-Gingerol
(8)-Gingerol
Vorinostat_c
Vorinostat_c
Vorinostat_c
Vorinostat_c
Vorinostat_c
Glycocyamine
Glycocyamine
Glycocyamine
Glycocyamine
Glycocyamine
2-Acrylamido-2-methyl-1-propane sulfonic acid
2-Acrylamido-2-methyl-1-propane sulfonic acid
2-Acrylamido-2-methyl-1-propane sulfonic acid
2-Acrylamido-2-methyl-1-propane sulfonic acid
2-Acrylamido-2-methyl-1-propane sulfonic acid
Monomethyl glutarate
Monomethyl glutarate
Monomethyl glutarate
Monomethyl glutarate
Monomethyl glutarate
2-methyl-1H-benzimidazole-5-carboxylic acid
2-methyl-1H-benzimidazole-5-carboxylic acid
2-methyl-1H-benzimidazole-5-carboxylic acid
2-methyl-1H-benzimidazole-5-carboxylic acid
2-methyl-1H-benzimidazole-5-carboxylic acid
N-Nonanoylglycine_c
N-Nonanoylglycine_c

N-Nonanoylglycine_c
N-Nonanoylglycine_c
N-Nonanoylglycine_c
8-hydroxy-11-(hydroxymethyl)-1,5,11-trimethyltricyclo[6.2.1.02,6]undec-2-en-9-one
8-hydroxy-11-(hydroxymethyl)-1,5,11-trimethyltricyclo[6.2.1.02,6]undec-2-en-9-one
8-hydroxy-11-(hydroxymethyl)-1,5,11-trimethyltricyclo[6.2.1.02,6]undec-2-en-9-one
8-hydroxy-11-(hydroxymethyl)-1,5,11-trimethyltricyclo[6.2.1.02,6]undec-2-en-9-one
8-hydroxy-11-(hydroxymethyl)-1,5,11-trimethyltricyclo[6.2.1.02,6]undec-2-en-9-one
4-Hydroxyprolylleucine_b
4-Hydroxyprolylleucine_b
4-Hydroxyprolylleucine_b
4-Hydroxyprolylleucine_b
4-Hydroxyprolylleucine_b
Methyl 1-hydroxy-7-(hydroxymethyl)-1,4a,5,7a-tetrahydrocyclopenta[c]pyran-4-carboxylate_c
Methyl 1-hydroxy-7-(hydroxymethyl)-1,4a,5,7a-tetrahydrocyclopenta[c]pyran-4-carboxylate_c
Methyl 1-hydroxy-7-(hydroxymethyl)-1,4a,5,7a-tetrahydrocyclopenta[c]pyran-4-carboxylate_c
Methyl 1-hydroxy-7-(hydroxymethyl)-1,4a,5,7a-tetrahydrocyclopenta[c]pyran-4-carboxylate_c
Methyl 1-hydroxy-7-(hydroxymethyl)-1,4a,5,7a-tetrahydrocyclopenta[c]pyran-4-carboxylate_c
2-Methylthiazolidine_c
2-Methylthiazolidine_c
2-Methylthiazolidine_c
2-Methylthiazolidine_c
2-Methylthiazolidine_c
N1-(5-methylisoxazol-3-yl)-2-morpholinoacetamide
N1-(5-methylisoxazol-3-yl)-2-morpholinoacetamide
N1-(5-methylisoxazol-3-yl)-2-morpholinoacetamide
N1-(5-methylisoxazol-3-yl)-2-morpholinoacetamide
N1-(5-methylisoxazol-3-yl)-2-morpholinoacetamide
Aprobarbital
Aprobarbital
Aprobarbital
Aprobarbital
Aprobarbital
N-Phenylacetylglutamic acid_b
N-Phenylacetylglutamic acid_b
N-Phenylacetylglutamic acid_b
N-Phenylacetylglutamic acid_b
N-Phenylacetylglutamic acid_b
6-(1-Hydroxyethyl)-3-(hydroxymethyl)-2,7-dioxabicyclo[4.1.0]hept-3-en-5-one_a
6-(1-Hydroxyethyl)-3-(hydroxymethyl)-2,7-dioxabicyclo[4.1.0]hept-3-en-5-one_a
6-(1-Hydroxyethyl)-3-(hydroxymethyl)-2,7-dioxabicyclo[4.1.0]hept-3-en-5-one_a
6-(1-Hydroxyethyl)-3-(hydroxymethyl)-2,7-dioxabicyclo[4.1.0]hept-3-en-5-one_a
6-(1-Hydroxyethyl)-3-(hydroxymethyl)-2,7-dioxabicyclo[4.1.0]hept-3-en-5-one_a
8-hydroxy-7-methylguanine
8-hydroxy-7-methylguanine
8-hydroxy-7-methylguanine
8-hydroxy-7-methylguanine
8-hydroxy-7-methylguanine

Glycylvaline
Glycylvaline
Glycylvaline
Glycylvaline
Glycylvaline
Gln-Gln
Gln-Gln
Gln-Gln
Gln-Gln
Gln-Gln
(âˆ’)-nabilone_d
(âˆ’)-nabilone_d
(âˆ’)-nabilone_d
(âˆ’)-nabilone_d
(âˆ’)-nabilone_d
Furaneol
Furaneol
Furaneol
Furaneol
Furaneol
norhaman
norhaman
norhaman
norhaman
norhaman
Acetanilide
Acetanilide
Acetanilide
Acetanilide
Acetanilide
Ethyl malate_b
Ethyl malate_b
Ethyl malate_b
Ethyl malate_b
Ethyl malate_b
Toxopyrimidine
Toxopyrimidine
Toxopyrimidine
Toxopyrimidine
Toxopyrimidine
5-Nitro-2-propoxyaniline
5-Nitro-2-propoxyaniline
5-Nitro-2-propoxyaniline
5-Nitro-2-propoxyaniline
5-Nitro-2-propoxyaniline
Glycyrin
Glycyrin
Glycyrin

Glycyrin
Glycyrin
Cystine
Cystine
Cystine
Cystine
Cystine
2-(4-Isopropyl-4-methyl-5-oxo-4,5-dihydro-1H-imidazol-2-yl)-4-methylbenzoic acid_a
2-(4-Isopropyl-4-methyl-5-oxo-4,5-dihydro-1H-imidazol-2-yl)-4-methylbenzoic acid_a
2-(4-Isopropyl-4-methyl-5-oxo-4,5-dihydro-1H-imidazol-2-yl)-4-methylbenzoic acid_a
2-(4-Isopropyl-4-methyl-5-oxo-4,5-dihydro-1H-imidazol-2-yl)-4-methylbenzoic acid_a
2-(4-Isopropyl-4-methyl-5-oxo-4,5-dihydro-1H-imidazol-2-yl)-4-methylbenzoic acid_a
Bicine_a
Bicine_a
Bicine_a
Bicine_a
Bicine_a
Histidylglycine_a
Histidylglycine_a
Histidylglycine_a
Histidylglycine_a
Histidylglycine_a
Zalcitabine_b
Zalcitabine_b
Zalcitabine_b
Zalcitabine_b
Zalcitabine_b
butalbital_b
butalbital_b
butalbital_b
butalbital_b
butalbital_b
Hyocholic acid
Hyocholic acid
Hyocholic acid
Hyocholic acid
Hyocholic acid
5-amino-2-(dimethylamino)benzoic acid_b
5-amino-2-(dimethylamino)benzoic acid_b
5-amino-2-(dimethylamino)benzoic acid_b
5-amino-2-(dimethylamino)benzoic acid_b
5-amino-2-(dimethylamino)benzoic acid_b
Melatonin
Melatonin
Melatonin
Melatonin
Melatonin
L-gamma-Glutamyl-L-leucine_e

L-gamma-Glutamyl-L-leucine_e
L-gamma-Glutamyl-L-leucine_e
L-gamma-Glutamyl-L-leucine_e
L-gamma-Glutamyl-L-leucine_e
L-gamma-Glutamyl-L-valine_g
L-gamma-Glutamyl-L-valine_g
L-gamma-Glutamyl-L-valine_g
L-gamma-Glutamyl-L-valine_g
L-gamma-Glutamyl-L-valine_g
N-acetyl-beta-D-glucosaminylamine
N-acetyl-beta-D-glucosaminylamine
N-acetyl-beta-D-glucosaminylamine
N-acetyl-beta-D-glucosaminylamine
N-acetyl-beta-D-glucosaminylamine
6-Hydroxynicotinic acid
6-Hydroxynicotinic acid
6-Hydroxynicotinic acid
6-Hydroxynicotinic acid
6-Hydroxynicotinic acid
bis-noryangonin
bis-noryangonin
bis-noryangonin
bis-noryangonin
bis-noryangonin
Isoquinoline_b
Isoquinoline_b
Isoquinoline_b
Isoquinoline_b
Isoquinoline_b
meprobamate_a
meprobamate_a
meprobamate_a
meprobamate_a
meprobamate_a
Temozolomide
Temozolomide
Temozolomide
Temozolomide
Temozolomide
MFCD09953737_c
MFCD09953737_c
MFCD09953737_c
MFCD09953737_c
MFCD09953737_c
clavulanic acid
clavulanic acid
clavulanic acid
clavulanic acid

clavulanic acid
1,1'-[1,12-Dodecanediylbis(oxy)]dibenzene_f
1,1'-[1,12-Dodecanediylbis(oxy)]dibenzene_f
1,1'-[1,12-Dodecanediylbis(oxy)]dibenzene_f
1,1'-[1,12-Dodecanediylbis(oxy)]dibenzene_f
1,1'-[1,12-Dodecanediylbis(oxy)]dibenzene_f
8-Methyl-8-azabicyclo[3.2.1]octane-1,2,3,4,6-pentol_b
8-Methyl-8-azabicyclo[3.2.1]octane-1,2,3,4,6-pentol_b
8-Methyl-8-azabicyclo[3.2.1]octane-1,2,3,4,6-pentol_b
8-Methyl-8-azabicyclo[3.2.1]octane-1,2,3,4,6-pentol_b
8-Methyl-8-azabicyclo[3.2.1]octane-1,2,3,4,6-pentol_b
Vorinostat_d
Vorinostat_d
Vorinostat_d
Vorinostat_d
Vorinostat_d
Threonylglutamine
Threonylglutamine
Threonylglutamine
Threonylglutamine
Threonylglutamine
pentobarbital_d
pentobarbital_d
pentobarbital_d
pentobarbital_d
pentobarbital_d
acetyltaurine
acetyltaurine
acetyltaurine
acetyltaurine
acetyltaurine
8-Methyl-8-azabicyclo[3.2.1]octane-1,2,3,4,6-pentol_c
8-Methyl-8-azabicyclo[3.2.1]octane-1,2,3,4,6-pentol_c
8-Methyl-8-azabicyclo[3.2.1]octane-1,2,3,4,6-pentol_c
8-Methyl-8-azabicyclo[3.2.1]octane-1,2,3,4,6-pentol_c
8-Methyl-8-azabicyclo[3.2.1]octane-1,2,3,4,6-pentol_c
2-BUTYL PROPENYL DISULFIDE, E(R)-_a
2-BUTYL PROPENYL DISULFIDE, E(R)-_a
2-BUTYL PROPENYL DISULFIDE, E(R)-_a
2-BUTYL PROPENYL DISULFIDE, E(R)-_a
2-BUTYL PROPENYL DISULFIDE, E(R)-_a
2-[(carboxymethyl)methylamino]-5-methoxy-Benzoic acid_b
2-[(carboxymethyl)methylamino]-5-methoxy-Benzoic acid_b
2-[(carboxymethyl)methylamino]-5-methoxy-Benzoic acid_b
2-[(carboxymethyl)methylamino]-5-methoxy-Benzoic acid_b
2-[(carboxymethyl)methylamino]-5-methoxy-Benzoic acid_b
Furan
Furan

Furan
Furan
Furan
butalbital_e
butalbital_e
butalbital_e
butalbital_e
butalbital_e
2,4-Bis(3-methyl-2-buten-1-yl)-1,3,5-benzenetriol
2,4-Bis(3-methyl-2-buten-1-yl)-1,3,5-benzenetriol
2,4-Bis(3-methyl-2-buten-1-yl)-1,3,5-benzenetriol
2,4-Bis(3-methyl-2-buten-1-yl)-1,3,5-benzenetriol
2,4-Bis(3-methyl-2-buten-1-yl)-1,3,5-benzenetriol
N-Acetyl-5-oxo-L-norvaline_c
N-Acetyl-5-oxo-L-norvaline_c
N-Acetyl-5-oxo-L-norvaline_c
N-Acetyl-5-oxo-L-norvaline_c
N-Acetyl-5-oxo-L-norvaline_c
1-(3-Amino-2,3-dideoxypentofuranosyl)-5-methyl-2,4(1H,3H)-pyrimidinedione
1-(3-Amino-2,3-dideoxypentofuranosyl)-5-methyl-2,4(1H,3H)-pyrimidinedione
1-(3-Amino-2,3-dideoxypentofuranosyl)-5-methyl-2,4(1H,3H)-pyrimidinedione
1-(3-Amino-2,3-dideoxypentofuranosyl)-5-methyl-2,4(1H,3H)-pyrimidinedione
1-(3-Amino-2,3-dideoxypentofuranosyl)-5-methyl-2,4(1H,3H)-pyrimidinedione
3-Hydroxypyridine
3-Hydroxypyridine
3-Hydroxypyridine
3-Hydroxypyridine
3-Hydroxypyridine
(2E)-3-(3,4-Dimethoxyphenyl)acrylic acid
(2E)-3-(3,4-Dimethoxyphenyl)acrylic acid
(2E)-3-(3,4-Dimethoxyphenyl)acrylic acid
(2E)-3-(3,4-Dimethoxyphenyl)acrylic acid
(2E)-3-(3,4-Dimethoxyphenyl)acrylic acid
Leu-arg
Leu-arg
Leu-arg
Leu-arg
Leu-arg
Dihydrocortisol
Dihydrocortisol
Dihydrocortisol
Dihydrocortisol
Dihydrocortisol
Resveratrol-4'-O-Glucuronide
Resveratrol-4'-O-Glucuronide
Resveratrol-4'-O-Glucuronide
Resveratrol-4'-O-Glucuronide
Resveratrol-4'-O-Glucuronide

N,N'-Bis[4-(4,5-dihydro-1H-imidazol-2-yl)phenyl]-2-methoxyterephthalamide
N,N'-Bis[4-(4,5-dihydro-1H-imidazol-2-yl)phenyl]-2-methoxyterephthalamide
N,N'-Bis[4-(4,5-dihydro-1H-imidazol-2-yl)phenyl]-2-methoxyterephthalamide
N,N'-Bis[4-(4,5-dihydro-1H-imidazol-2-yl)phenyl]-2-methoxyterephthalamide
N,N'-Bis[4-(4,5-dihydro-1H-imidazol-2-yl)phenyl]-2-methoxyterephthalamide
Tauropine
Tauropine
Tauropine
Tauropine
Tauropine
Hydroxycarteolol_b
Hydroxycarteolol_b
Hydroxycarteolol_b
Hydroxycarteolol_b
Hydroxycarteolol_b
Coprine_b
Coprine_b
Coprine_b
Coprine_b
Coprine_b
5,8,12-Trihydroxy-2-oxododecanoic acid
5,8,12-Trihydroxy-2-oxododecanoic acid
5,8,12-Trihydroxy-2-oxododecanoic acid
5,8,12-Trihydroxy-2-oxododecanoic acid
5,8,12-Trihydroxy-2-oxododecanoic acid
Leu-Leu_e
Leu-Leu_e
Leu-Leu_e
Leu-Leu_e
Leu-Leu_e
Lys-Pro_a
Lys-Pro_a
Lys-Pro_a
Lys-Pro_a
Lys-Pro_a
QJ9720000
QJ9720000
QJ9720000
QJ9720000
QJ9720000
Fluocinolone Acetonide
Fluocinolone Acetonide
Fluocinolone Acetonide
Fluocinolone Acetonide
Fluocinolone Acetonide
Leu-Leu_b
Leu-Leu_b
Leu-Leu_b

Leu-Leu_b
Leu-Leu_b
Ethyl malate_a
Ethyl malate_a
Ethyl malate_a
Ethyl malate_a
Ethyl malate_a
MFCD00037215
MFCD00037215
MFCD00037215
MFCD00037215
MFCD00037215
N-Nonanoylglycine_a
N-Nonanoylglycine_a
N-Nonanoylglycine_a
N-Nonanoylglycine_a
N-Nonanoylglycine_a
(4R,5S,9S,10R,12S,13S)-1,5,9-Trimethyl-11,14,15,16-tetraoxatetracyclo[10.3.1.0~4,13~.0~8,13~]hexadecan-10
(4R,5S,9S,10R,12S,13S)-1,5,9-Trimethyl-11,14,15,16-tetraoxatetracyclo[10.3.1.0~4,13~.0~8,13~]hexadecan-10
(4R,5S,9S,10R,12S,13S)-1,5,9-Trimethyl-11,14,15,16-tetraoxatetracyclo[10.3.1.0~4,13~.0~8,13~]hexadecan-10
(4R,5S,9S,10R,12S,13S)-1,5,9-Trimethyl-11,14,15,16-tetraoxatetracyclo[10.3.1.0~4,13~.0~8,13~]hexadecan-10
(4R,5S,9S,10R,12S,13S)-1,5,9-Trimethyl-11,14,15,16-tetraoxatetracyclo[10.3.1.0~4,13~.0~8,13~]hexadecan-10
1-Methylguanine
1-Methylguanine
1-Methylguanine
1-Methylguanine
1-Methylguanine
Valyl-4-hydroxyproline_a
Valyl-4-hydroxyproline_a
Valyl-4-hydroxyproline_a
Valyl-4-hydroxyproline_a
Valyl-4-hydroxyproline_a
Sulfurol
Sulfurol
Sulfurol
Sulfurol
Sulfurol
trp-pro
trp-pro
trp-pro
trp-pro
trp-pro
lys-leu
lys-leu
lys-leu
lys-leu
lys-leu
Penbutolol

Penbutolol
Penbutolol
Penbutolol
Penbutolol
(6S)-2-Amino-6-[(3-carboxypropanoyl)amino]heptanedioic acid
(6S)-2-Amino-6-[(3-carboxypropanoyl)amino]heptanedioic acid
(6S)-2-Amino-6-[(3-carboxypropanoyl)amino]heptanedioic acid
(6S)-2-Amino-6-[(3-carboxypropanoyl)amino]heptanedioic acid
(6S)-2-Amino-6-[(3-carboxypropanoyl)amino]heptanedioic acid
Leu-Val_a
Leu-Val_a
Leu-Val_a
Leu-Val_a
Leu-Val_a
Methohexital_a
Methohexital_a
Methohexital_a
Methohexital_a
Methohexital_a
dihydroxybenzylamine
dihydroxybenzylamine
dihydroxybenzylamine
dihydroxybenzylamine
dihydroxybenzylamine
N~6~-Octanoyllysine_b
N~6~-Octanoyllysine_b
N~6~-Octanoyllysine_b
N~6~-Octanoyllysine_b
N~6~-Octanoyllysine_b
L-gamma-Glutamyl-L-leucine_f
L-gamma-Glutamyl-L-leucine_f
L-gamma-Glutamyl-L-leucine_f
L-gamma-Glutamyl-L-leucine_f
L-gamma-Glutamyl-L-leucine_f
Voglibose
Voglibose
Voglibose
Voglibose
Voglibose
Butabarbital_d
Butabarbital_d
Butabarbital_d
Butabarbital_d
Butabarbital_d
3-Oxo-4,6-choladienoic acid
3-Oxo-4,6-choladienoic acid
3-Oxo-4,6-choladienoic acid
3-Oxo-4,6-choladienoic acid

3-Oxo-4,6-choladienoic acid
1-Methylhistamine
1-Methylhistamine
1-Methylhistamine
1-Methylhistamine
1-Methylhistamine
3-Sulfino-L-alanine
3-Sulfino-L-alanine
3-Sulfino-L-alanine
3-Sulfino-L-alanine
3-Sulfino-L-alanine
3-Succinoylpyridine
3-Succinoylpyridine
3-Succinoylpyridine
3-Succinoylpyridine
3-Succinoylpyridine
L-gamma-Glutamyl-L-valine_j
L-gamma-Glutamyl-L-valine_j
L-gamma-Glutamyl-L-valine_j
L-gamma-Glutamyl-L-valine_j
L-gamma-Glutamyl-L-valine_j
N-{3-[(4-Acetamidobutyl)amino]propyl}acetamide_b
N-{3-[(4-Acetamidobutyl)amino]propyl}acetamide_b
N-{3-[(4-Acetamidobutyl)amino]propyl}acetamide_b
N-{3-[(4-Acetamidobutyl)amino]propyl}acetamide_b
N-{3-[(4-Acetamidobutyl)amino]propyl}acetamide_b
5-Amino-6-(4-hydroxybutanoyl)-2,2-dimethyl-2,3-dihydro-4H-chromen-4-one
5-Amino-6-(4-hydroxybutanoyl)-2,2-dimethyl-2,3-dihydro-4H-chromen-4-one
5-Amino-6-(4-hydroxybutanoyl)-2,2-dimethyl-2,3-dihydro-4H-chromen-4-one
5-Amino-6-(4-hydroxybutanoyl)-2,2-dimethyl-2,3-dihydro-4H-chromen-4-one
5-Amino-6-(4-hydroxybutanoyl)-2,2-dimethyl-2,3-dihydro-4H-chromen-4-one
8-Amino-7-oxononanoic acid_c
8-Amino-7-oxononanoic acid_c
8-Amino-7-oxononanoic acid_c
8-Amino-7-oxononanoic acid_c
8-Amino-7-oxononanoic acid_c
pro-met
pro-met
pro-met
pro-met
pro-met
beta-D-Ethyl glucuronide_b
beta-D-Ethyl glucuronide_b
beta-D-Ethyl glucuronide_b
beta-D-Ethyl glucuronide_b
beta-D-Ethyl glucuronide_b
gamma-Glutamyl-S-[1-(6,7-dimethoxy-1,3-benzodioxol-5-yl)-1,3-dihydroxy-2-propanyl]cysteinylglycine
gamma-Glutamyl-S-[1-(6,7-dimethoxy-1,3-benzodioxol-5-yl)-1,3-dihydroxy-2-propanyl]cysteinylglycine

gamma-Glutamyl-S-[1-(6,7-dimethoxy-1,3-benzodioxol-5-yl)-1,3-dihydroxy-2-propanyl]cysteinylglycine
gamma-Glutamyl-S-[1-(6,7-dimethoxy-1,3-benzodioxol-5-yl)-1,3-dihydroxy-2-propanyl]cysteinylglycine
gamma-Glutamyl-S-[1-(6,7-dimethoxy-1,3-benzodioxol-5-yl)-1,3-dihydroxy-2-propanyl]cysteinylglycine
his-gln_b
his-gln_b
his-gln_b
his-gln_b
his-gln_b
3-Hydroxysebacic acid
3-Hydroxysebacic acid
3-Hydroxysebacic acid
3-Hydroxysebacic acid
3-Hydroxysebacic acid
Valylvaline_c
Valylvaline_c
Valylvaline_c
Valylvaline_c
Valylvaline_c
FB9500000_b
FB9500000_b
FB9500000_b
FB9500000_b
FB9500000_b
13(S)-HOTrE
13(S)-HOTrE
13(S)-HOTrE
13(S)-HOTrE
13(S)-HOTrE
thr-trp
thr-trp
thr-trp
thr-trp
thr-trp
6-hydroxypseudooxynicotine_c
6-hydroxypseudooxynicotine_c
6-hydroxypseudooxynicotine_c
6-hydroxypseudooxynicotine_c
6-hydroxypseudooxynicotine_c
2-Aminooctanedioic acid_b
2-Aminooctanedioic acid_b
2-Aminooctanedioic acid_b
2-Aminooctanedioic acid_b
2-Aminooctanedioic acid_b
1,1'-[1,12-Dodecanediylbis(oxy)]dibenzene_c
1,1'-[1,12-Dodecanediylbis(oxy)]dibenzene_c
1,1'-[1,12-Dodecanediylbis(oxy)]dibenzene_c
1,1'-[1,12-Dodecanediylbis(oxy)]dibenzene_c
1,1'-[1,12-Dodecanediylbis(oxy)]dibenzene_c

N-[(2E)-3-(4-Hydroxyphenyl)-2-propenoyl]tryptophan
N-[(2E)-3-(4-Hydroxyphenyl)-2-propenoyl]tryptophan
N-[(2E)-3-(4-Hydroxyphenyl)-2-propenoyl]tryptophan
N-[(2E)-3-(4-Hydroxyphenyl)-2-propenoyl]tryptophan
N-[(2E)-3-(4-Hydroxyphenyl)-2-propenoyl]tryptophan
ALA-PRO
ALA-PRO
ALA-PRO
ALA-PRO
ALA-PRO
threonylphenylalanine_b
threonylphenylalanine_b
threonylphenylalanine_b
threonylphenylalanine_b
threonylphenylalanine_b
Phenyl D-glucopyranosiduronic acid_b
Phenyl D-glucopyranosiduronic acid_b
Phenyl D-glucopyranosiduronic acid_b
Phenyl D-glucopyranosiduronic acid_b
Phenyl D-glucopyranosiduronic acid_b
3-Methyl-2-buten-1-yl hydrogen sulfate
3-Methyl-2-buten-1-yl hydrogen sulfate
3-Methyl-2-buten-1-yl hydrogen sulfate
3-Methyl-2-buten-1-yl hydrogen sulfate
3-Methyl-2-buten-1-yl hydrogen sulfate
2-Methylthiazolidine_b
2-Methylthiazolidine_b
2-Methylthiazolidine_b
2-Methylthiazolidine_b
2-Methylthiazolidine_b
N-(3-acetamidopropyl)pyrrolidin-2-one_b
N-(3-acetamidopropyl)pyrrolidin-2-one_b
N-(3-acetamidopropyl)pyrrolidin-2-one_b
N-(3-acetamidopropyl)pyrrolidin-2-one_b
N-(3-acetamidopropyl)pyrrolidin-2-one_b
Midodrine_d
Midodrine_d
Midodrine_d
Midodrine_d
Midodrine_d
glu-thr
glu-thr
glu-thr
glu-thr
glu-thr
N-(4-Hydroxy-3,5-dimethoxybenzoyl)glycine_c
N-(4-Hydroxy-3,5-dimethoxybenzoyl)glycine_c
N-(4-Hydroxy-3,5-dimethoxybenzoyl)glycine_c

N-(4-Hydroxy-3,5-dimethoxybenzoyl)glycine_c
N-(4-Hydroxy-3,5-dimethoxybenzoyl)glycine_c
Tetraacetylethylenediamine_f
Tetraacetylethylenediamine_f
Tetraacetylethylenediamine_f
Tetraacetylethylenediamine_f
Tetraacetylethylenediamine_f
2_7-Anhydro-alpha-N-acetylneuraminicacid
2_7-Anhydro-alpha-N-acetylneuraminicacid
2_7-Anhydro-alpha-N-acetylneuraminicacid
2_7-Anhydro-alpha-N-acetylneuraminicacid
2_7-Anhydro-alpha-N-acetylneuraminicacid
1-(beta-D-ribofuranosyl)thymine_a
1-(beta-D-ribofuranosyl)thymine_a
1-(beta-D-ribofuranosyl)thymine_a
1-(beta-D-ribofuranosyl)thymine_a
1-(beta-D-ribofuranosyl)thymine_a
{2-[2-(Isobutyryloxy)-4-methylphenyl]-2-oxiranyl}methyl 2-methylbutanoate
{2-[2-(Isobutyryloxy)-4-methylphenyl]-2-oxiranyl}methyl 2-methylbutanoate
{2-[2-(Isobutyryloxy)-4-methylphenyl]-2-oxiranyl}methyl 2-methylbutanoate
{2-[2-(Isobutyryloxy)-4-methylphenyl]-2-oxiranyl}methyl 2-methylbutanoate
{2-[2-(Isobutyryloxy)-4-methylphenyl]-2-oxiranyl}methyl 2-methylbutanoate
ala-ser_a
ala-ser_a
ala-ser_a
ala-ser_a
ala-ser_a
nicotianamine_a
nicotianamine_a
nicotianamine_a
nicotianamine_a
nicotianamine_a
1,3-Dihydroxy-2-propanyl (7Z,10Z,13Z,16Z,19Z)-7,10,13,16,19-docosapentaenoate
1,3-Dihydroxy-2-propanyl (7Z,10Z,13Z,16Z,19Z)-7,10,13,16,19-docosapentaenoate
1,3-Dihydroxy-2-propanyl (7Z,10Z,13Z,16Z,19Z)-7,10,13,16,19-docosapentaenoate
1,3-Dihydroxy-2-propanyl (7Z,10Z,13Z,16Z,19Z)-7,10,13,16,19-docosapentaenoate
1,3-Dihydroxy-2-propanyl (7Z,10Z,13Z,16Z,19Z)-7,10,13,16,19-docosapentaenoate
2-Despiperidyl-2-amino Repaglinide
2-Despiperidyl-2-amino Repaglinide
2-Despiperidyl-2-amino Repaglinide
2-Despiperidyl-2-amino Repaglinide
2-Despiperidyl-2-amino Repaglinide
Furfuranol
Furfuranol
Furfuranol
Furfuranol
Furfuranol
p-Cresol

p-Cresol
p-Cresol
p-Cresol
p-Cresol

nethyl]-5-[(3-ethyl-4-methyl-5-oxo-1,2-dihydropyrrol-2-yl)methyl]-4-methyl-1H-pyrrol-3-yl]propanoic acid_a
nethyl]-5-[(3-ethyl-4-methyl-5-oxo-1,2-dihydropyrrol-2-yl)methyl]-4-methyl-1H-pyrrol-3-yl]propanoic acid_a
nethyl]-5-[(3-ethyl-4-methyl-5-oxo-1,2-dihydropyrrol-2-yl)methyl]-4-methyl-1H-pyrrol-3-yl]propanoic acid_a
nethyl]-5-[(3-ethyl-4-methyl-5-oxo-1,2-dihydropyrrol-2-yl)methyl]-4-methyl-1H-pyrrol-3-yl]propanoic acid_a
nethyl]-5-[(3-ethyl-4-methyl-5-oxo-1,2-dihydropyrrol-2-yl)methyl]-4-methyl-1H-pyrrol-3-yl]propanoic acid_a

nethyl]-5-[(3-ethyl-4-methyl-5-oxo-1,2-dihydropyrrol-2-yl)methyl]-4-methyl-1H-pyrrol-3-yl]propanoic acid_b
nethyl]-5-[(3-ethyl-4-methyl-5-oxo-1,2-dihydropyrrol-2-yl)methyl]-4-methyl-1H-pyrrol-3-yl]propanoic acid_b
nethyl]-5-[(3-ethyl-4-methyl-5-oxo-1,2-dihydropyrrol-2-yl)methyl]-4-methyl-1H-pyrrol-3-yl]propanoic acid_b
nethyl]-5-[(3-ethyl-4-methyl-5-oxo-1,2-dihydropyrrol-2-yl)methyl]-4-methyl-1H-pyrrol-3-yl]propanoic acid_b

nethyl]-5-[(3-ethyl-4-methyl-5-oxo-1,2-dihydropyrrol-2-yl)methyl]-4-methyl-1H-pyrrol-3-yl]propanoic acid_b
