## Supplemental Data 1 for "A YOUNG CHILD FORMULA SUPPLEMENTED WITH L. REUTERI AND GALACTO-OLIGOSACCHARIDES MODULATES THE COMPOSITION AND FUNCTION OF THE GUT MICROBIOME SUPPORTING BONE AND MUSCLE DEVELOPMENT IN TODDLERS"

| Clinical variable | Grouping | Characteristic | Beta | 95% CI low |
|---|---|---|---|---|
| Tibia SOS | Experimental + Control | Increase / Non-increase | 55 | 9,6 |
| Tibia SOS | Experimental + Control | Tibia SOS (V1) | 0.34 | 0,2 |
| Tibia SOS | Experimental + Control | Sex | 62 | 21 |
| Tibia SOS | Experimental + Control | Computed BMI | -16 | -30 |
| Tibia SOS | Experimental + Control | Vitamin D (V1) | -0.49 | -3 |
| Tibia SOS | Experimental + Control + Reference | Increase / Non-increase | 46 | 4,7 |
| Tibia SOS | Experimental + Control + Reference | Tibia SOS (V1) | 0.33 | 0,21 |
| Tibia SOS | Experimental + Control + Reference | Sex | 32 | -0,65 |
| Tibia SOS | Experimental + Control + Reference | Computed BMI | -12 | -24 |
| Tibia SOS | Experimental + Control + Reference | Vitamin D (V1) | -0.49 | -2,5 |
| Vitamin D | Experimental + Control | Vitamin D (V1) | 0.02 | 0,02 |
| Vitamin D | Experimental + Control | Increase / Non-increase | 0.10 | 0,03 |
| Vitamin D | Experimental + Control | Sex | -0.03 | -0,1 |
| Vitamin D | Experimental + Control + Reference | Vitamin D (V1) | 0.02 | 0,02 |
| Vitamin D | Experimental + Control + Reference | Increase / Non-increase | 0.07 | 0,01 |
| Vitamin D | Experimental + Control + Reference | Sex | -0.04 | -0,09 |
| Tibia length | Experimental + Control | Increase / Non-increase | -0.29 | -0,81 |
| Tibia length | Experimental + Control | Tibia length (V1) | 0.16 | 0,09 |
| Tibia length | Experimental + Control | Sex | 0.17 | -0,3 |
| Tibia length | Experimental + Control | Computed BMI | 0.17 | 0,01 |
| Tibia length | Experimental + Control | Vitamin D (V1) | -0.01 | -0,04 |
| Tibia length | Experimental + Control + Reference | Increase / Non-increase | 0.02 | -0,41 |
| Tibia length | Experimental + Control + Reference | Tibia length (V1) | 0.18 | 0,13 |
| Tibia length | Experimental + Control + Reference | Sex | 0.20 | -0,14 |
| Tibia length | Experimental + Control + Reference | Computed BMI | 0.20 | 0,07 |
| Tibia length | Experimental + Control + Reference | Vitamin D (V1) | -0.01 | -0,03 |
| Radius SOS | Experimental + Control | Increase / Non-increase | 19 | -26 |
| Radius SOS | Experimental + Control | Radius SOS (V1) | 0.34 | 0,21 |
| Radius SOS | Experimental + Control | Sex | 51 | 11 |
| Radius SOS | Experimental + Control | Computed BMI | -3.2 | -17 |
| Radius SOS | Experimental + Control | Vitamin D (V1) | -0.99 | -3,4 |
| Radius SOS | Experimental + Control + Reference | Increase / Non-increase | 15 | -27 |
| Radius SOS | Experimental + Control + Reference | Radius SOS (V1) | 0.46 | 0,35 |
| Radius SOS | Experimental + Control + Reference | Sex | 32 | -1,3 |
| Radius SOS | Experimental + Control + Reference | Computed BMI | -0.84 | -13 |
| Radius SOS | Experimental + Control + Reference | Vitamin D (V1) | 0.33 | -1,7 |
| Radius length | Experimental + Control | Increase / Non-increase | -0.02 | -0,24 |
| Radius length | Experimental + Control | Radius length (V1) | 0.87 | 0,78 |
| Radius length | Experimental + Control | Sex | 0.27 | 0,07 |
| Radius length | Experimental + Control | Computed BMI | -0.01 | -0,08 |
| Radius length | Experimental + Control | Vitamin D (V1) | -0.01 | -0,02 |
| Radius length | Experimental + Control + Reference | Increase / Non-increase | 0.07 | -0,12 |
| Radius length | Experimental + Control + Reference | Radius length (V1) | 0.83 | 0,76 |
| Radius length | Experimental + Control + Reference | Sex | 0.20 | 0,05 |
| Radius length | Experimental + Control + Reference | Computed BMI | 0.00 | -0,05 |
| Radius length | Experimental + Control + Reference | Vitamin D (V1) | -0.01 | -0,02 |

| 95% CI high | P-value |
|---|---|
| 101 | 0.018 |
| 0,49 | <0.001 |
| 103 | 0.004 |
| -1,1 | 0.035 |
| 2 | 0.7 |
| 87 | 0.029 |
| 0,46 | <0.001 |
| 65 | 0.055 |
| -0,59 | 0.040 |
| 1,5 | 0.6 |
| 0,03 | <0.001 |
| 0,18 | 0.005 |
| 0,03 | 0.3 |
| 0,02 | <0.001 |
| 0,13 | 0.023 |
| 0,01 | 0.086 |
| 0,22 | 0.3 |
| 0,24 | <0.001 |
| 0,63 | 0.5 |
| 0,33 | 0.042 |
| 0,01 | 0.3 |
| 0,45 | >0.9 |
| 0,24 | <0.001 |
| 0,55 | 0.3 |
| 0,32 | 0.002 |
| 0,01 | 0.4 |
| 63 | 0.4 |
| 0,48 | <0.001 |
| 91 | 0.014 |
| 11 | 0.7 |
| 1,4 | 0.4 |
| 58 | 0.5 |
| 0,57 | <0.001 |
| 66 | 0.059 |
| 11 | 0.9 |
| 2,4 | 0.8 |
| 0,2 | 0.9 |
| 0,96 | <0.001 |
| 0,47 | 0.009 |
| 0,06 | 0.8 |
| 0 | 0.046 |
| 0,25 | 0.5 |
| 0,91 | <0.001 |
| 0,35 | 0.010 |
| 0,06 | >0.9 |
| 0 | 0.13 |
