## Supplemental table S15. Stool consistency and Descriptive statistics for the TCGQ questionnaire. for "A YOUNG CHILD FORMULA SUPPLEMENTED WITH L. REUTERI AND GALACTO-OLIGOSACCHARIDES MODULATES THE COMPOSITION AND FUNCTION OF THE GUT MICROBIOME SUPPORTING BONE AND MUSCLE DEVELOPMENT IN TODDLERS"

| Category | Item | Visit | Arm | n | min | Q1 | Median |
|---|---|---|---|---|---|---|---|
| Average consistency of stools averaged over 3 days | | V2 | CM | 72 | 3 | 3,65 | 4 |
| | | V2 | EYCF | 70 | 1,29 | 3 | 3,75 |
| | | V3 | CM | 69 | 2,5 | 4 | 4 |
| | | V3 | EYCF | 69 | 2,67 | 3,5 | 4 |
| Gastro intestinal TOTAL SCORE | | V1 | REF | 91 | 9 | 10 | 10 |
| | | V1 | CM | 91 | 9 | 10 | 11 |
| | | V1 | EYCF | 91 | 9 | 10 | 11 |
| | | V2 | REF | 87 | 9 | 9 | 10 |
| | | V2 | CM | 72 | 9 | 10 | 10 |
| | | V2 | EYCF | 70 | 9 | 9,25 | 10 |
| | | V3 | REF | 87 | 9 | 9 | 10 |
| | | V3 | CM | 69 | 9 | 10 | 10 |
| | | V3 | EYCF | 69 | 9 | 9 | 10 |
| testinal related Q | Gastro intestinal symptoms | V1 | REF | 91 | 6 | 6 | 6 |
| | | V1 | CM | 91 | 6 | 6 | 6 |
| | | V1 | EYCF | 91 | 6 | 6 | 6 |
| | | V2 | REF | 87 | 6 | 6 | 6 |
| | | V2 | CM | 72 | 6 | 6 | 6 |
| | | V2 | EYCF | 70 | 6 | 6 | 6 |
| | | V3 | REF | 87 | 6 | 6 | 6 |
| | | V3 | CM | 69 | 6 | 6 | 6 |
| | | V3 | EYCF | 69 | 6 | 6 | 6 |
| | Q1. Did your child have stooling issues | V1 | REF | 91 | 1 | 1 | 1 |
| | | V1 | CM | 91 | 1 | 1 | 1 |
| | | V1 | EYCF | 91 | 1 | 1 | 1 |
| | | V2 | REF | 87 | 1 | 1 | 1 |
| | | V2 | CM | 72 | 1 | 1 | 1 |
| | | V2 | EYCF | 70 | 1 | 1 | 1 |
| | | V3 | REF | 87 | 1 | 1 | 1 |
| | | V3 | CM | 69 | 1 | 1 | 1 |
| | | V3 | EYCF | 69 | 1 | 1 | 1 |
| | Q1.a Did your child have constipation? | V1 | REF | 91 | 1 | 1 | 1 |
| | | V1 | CM | 91 | 1 | 1 | 1 |
| | | V1 | EYCF | 91 | 1 | 1 | 1 |
| | | V2 | REF | 87 | 1 | 1 | 1 |
| | | V2 | CM | 72 | 1 | 1 | 1 |
| | | V2 | EYCF | 70 | 1 | 1 | 1 |
| | | V3 | REF | 87 | 1 | 1 | 1 |
| | | V3 | CM | 69 | 1 | 1 | 1 |
| | | V3 | EYCF | 69 | 1 | 1 | 1 |
| | Q1.b Did your child have diarrhea? | V1 | REF | 91 | 1 | 1 | 1 |
| | | V1 | CM | 91 | 1 | 1 | 1 |
| | | V1 | EYCF | 91 | 1 | 1 | 1 |
| | | V2 | REF | 87 | 1 | 1 | 1 |
| | | V2 | CM | 72 | 1 | 1 | 1 |
| | | V2 | EYCF | 70 | 1 | 1 | 1 |

| | | | | | | | |
|---|---|---|---|---|---|---|---|
| | | V3 | REF | 87 | 1 | 1 | 1 |
| | | V3 | CM | 69 | 1 | 1 | 1 |
| | | V3 | EYCF | 69 | 1 | 1 | 1 |
| | Q2. Did your child experience gassiness? | V1 | REF | 91 | 1 | 1 | 1 |
| | | V1 | CM | 91 | 1 | 1 | 1 |
| | | V1 | EYCF | 91 | 1 | 1 | 1 |
| | | V2 | REF | 87 | 1 | 1 | 1 |
| | | V2 | CM | 72 | 1 | 1 | 1 |
| | | V2 | EYCF | 70 | 1 | 1 | 1 |
| | | V3 | REF | 87 | 1 | 1 | 1 |
| | | V3 | CM | 69 | 1 | 1 | 1 |
| | | V3 | EYCF | 69 | 1 | 1 | 1 |
| | Q3. Did your child have abdominal pain? | V1 | REF | 91 | 1 | 1 | 1 |
| | | V1 | CM | 91 | 1 | 1 | 1 |
| | | V1 | EYCF | 91 | 1 | 1 | 1 |
| | | V2 | REF | 87 | 1 | 1 | 1 |
| | | V2 | CM | 72 | 1 | 1 | 1 |
| | | V2 | EYCF | 70 | 1 | 1 | 1 |
| | | V3 | REF | 87 | 1 | 1 | 1 |
| | | V3 | CM | 69 | 1 | 1 | 1 |
| | | V3 | EYCF | 69 | 1 | 1 | 1 |
| | Q4. Did your child feel bloated? | V1 | REF | 91 | 1 | 1 | 1 |
| | | V1 | CM | 91 | 1 | 1 | 1 |
| | | V1 | EYCF | 91 | 1 | 1 | 1 |
| | | V2 | REF | 87 | 1 | 1 | 1 |
| | | V2 | CM | 72 | 1 | 1 | 1 |
| | | V2 | EYCF | 70 | 1 | 1 | 1 |
| | | V3 | REF | 87 | 1 | 1 | 1 |
| | | V3 | CM | 69 | 1 | 1 | 1 |
| | | V3 | EYCF | 69 | 1 | 1 | 1 |
| | Gastro intestinal-RELATED BEHAVIORS domain | V1 | REF | 91 | 3 | 4 | 4 |
| | | V1 | CM | 91 | 3 | 4 | 5 |
| | | V1 | EYCF | 91 | 3 | 4 | 5 |
| | | V2 | REF | 87 | 3 | 3 | 4 |
| | | V2 | CM | 72 | 3 | 3 | 4 |
| | | V2 | EYCF | 70 | 3 | 3 | 4 |
| | | V3 | REF | 87 | 3 | 3 | 4 |
| | | V3 | CM | 69 | 3 | 4 | 4 |
| | | V3 | EYCF | 69 | 3 | 3 | 4 |
| | Q1. Did your child seem fussy and irritable? | V1 | REF | 91 | 1 | 1 | 1 |
| | | V1 | CM | 91 | 1 | 1 | 1 |
| | | V1 | EYCF | 91 | 1 | 1 | 1 |
| | | V2 | REF | 87 | 1 | 1 | 1 |
| | | V2 | CM | 72 | 1 | 1 | 1 |
| | | V2 | EYCF | 70 | 1 | 1 | 1 |
| | | V3 | REF | 87 | 1 | 1 | 1 |
| | | V3 | CM | 69 | 1 | 1 | 1 |

| | | | | | | | |
|---|---|---|---|---|---|---|---|
| Gastro intestinal related behavior questions | | V3 | EYCF | 69 | 1 | 1 | 1 |
| | Q2. Do you consider the sleep of your toddler a problem? | V1 | REF | 91 | 1 | 1 | 1 |
| | | V1 | CM | 91 | 1 | 1 | 1 |
| | | V1 | EYCF | 91 | 1 | 1 | 1 |
| | | V2 | REF | 87 | 1 | 1 | 1 |
| | | V2 | CM | 72 | 1 | 1 | 1 |
| | | V2 | EYCF | 70 | 1 | 1 | 1 |
| | | V3 | REF | 87 | 1 | 1 | 1 |
| | | V3 | CM | 69 | 1 | 1 | 1 |
| | | V3 | EYCF | 69 | 1 | 1 | 1 |
| | Q3. Generally, how sleepy did your toddler get during the day? | V1 | REF | 91 | 1 | 1 | 1 |
| | | V1 | CM | 91 | 1 | 1 | 1 |
| | | V1 | EYCF | 91 | 1 | 1 | 1 |
| | | V2 | REF | 87 | 1 | 1 | 1 |
| | | V2 | CM | 72 | 1 | 1 | 1 |
| | | V2 | EYCF | 70 | 1 | 1 | 1 |
| | | V3 | REF | 87 | 1 | 1 | 1 |
| | | V3 | CM | 69 | 1 | 1 | 1 |
| | | V3 | EYCF | 69 | 1 | 1 | 1 |
| | Q4. How many times did your toddler wake up during the night (between 7 in the evening and 7 in the morning)? | V1 | REF | 91 | 0 | 0 | 1 |
| | | V1 | CM | 91 | 0 | 1 | 1 |
| | | V1 | EYCF | 91 | 0 | 1 | 1 |
| | | V2 | REF | 87 | 0 | 0 | 1 |
| | | V2 | CM | 72 | 0 | 0 | 1 |
| | | V2 | EYCF | 70 | 0 | 0 | 1 |
| | | V3 | REF | 87 | 0 | 0 | 1 |
| | | V3 | CM | 69 | 0 | 0 | 1 |
| | | V3 | EYCF | 69 | 0 | 0 | 1 |

**questionnaire**

| Q3 | max | mean | sd |
|---|---|---|---|
| 4 | 5 | 3,78 | 0,39 |
| 4 | 4 | 3,5 | 0,62 |
| 4 | 5 | 3,87 | 0,37 |
| 4 | 4 | 3,72 | 0,4 |
| 11,5 | 23 | 11,21 | 2,75 |
| 12 | 26 | 11,37 | 2,62 |
| 12 | 29 | 12,05 | 3,43 |
| 11 | 15 | 10,15 | 1,25 |
| 11 | 18 | 10,85 | 1,96 |
| 11 | 24 | 10,64 | 2,28 |
| 11 | 13 | 10,17 | 0,97 |
| 11 | 18 | 10,57 | 1,44 |
| 11 | 17 | 10,62 | 1,63 |
| 6 | 16 | 6,55 | 1,72 |
| 6 | 18 | 6,45 | 1,56 |
| 6 | 17 | 6,79 | 2,04 |
| 6 | 7 | 6,05 | 0,21 |
| 6 | 12 | 6,49 | 1,19 |
| 6 | 13 | 6,5 | 1,25 |
| 6 | 7 | 6,09 | 0,29 |
| 6 | 11 | 6,22 | 0,68 |
| 6 | 9 | 6,14 | 0,49 |
| 1 | 3 | 1,12 | 0,44 |
| 1 | 4 | 1,05 | 0,35 |
| 1 | 4 | 1,13 | 0,52 |
| 1 | 1 | 1 | 0 |
| 1 | 4 | 1,14 | 0,48 |
| 1 | 3 | 1,1 | 0,35 |
| 1 | 2 | 1,01 | 0,11 |
| 1 | 2 | 1,01 | 0,12 |
| 1 | 2 | 1,01 | 0,12 |
| 1 | 2 | 1,04 | 0,21 |
| 1 | 3 | 1,05 | 0,27 |
| 1 | 5 | 1,15 | 0,59 |
| 1 | 1 | 1 | 0 |
| 1 | 3 | 1,12 | 0,37 |
| 1 | 3 | 1,1 | 0,35 |
| 1 | 2 | 1,05 | 0,21 |
| 1 | 2 | 1,09 | 0,28 |
| 1 | 2 | 1,06 | 0,24 |
| 1 | 3 | 1,09 | 0,32 |
| 1 | 3 | 1,1 | 0,37 |
| 1 | 3 | 1,14 | 0,44 |
| 1 | 1 | 1 | 0 |
| 1 | 2 | 1,03 | 0,17 |
| 1 | 4 | 1,13 | 0,51 |

| | | | |
|---|---|---|---|
| 1 | 1 | 1 | 0 |
| 1 | 4 | 1,04 | 0,36 |
| 1 | 1 | 1 | 0 |
| 1 | 3 | 1,07 | 0,29 |
| 1 | 4 | 1,07 | 0,36 |
| 1 | 3 | 1,12 | 0,39 |
| 1 | 1 | 1 | 0 |
| 1 | 3 | 1,03 | 0,24 |
| 1 | 3 | 1,04 | 0,27 |
| 1 | 1 | 1 | 0 |
| 1 | 1 | 1 | 0 |
| 1 | 2 | 1,01 | 0,12 |
| 1 | 3 | 1,08 | 0,31 |
| 1 | 2 | 1,05 | 0,23 |
| 1 | 3 | 1,09 | 0,32 |
| 1 | 2 | 1,01 | 0,11 |
| 1 | 3 | 1,04 | 0,26 |
| 1 | 2 | 1,03 | 0,17 |
| 1 | 2 | 1,01 | 0,11 |
| 1 | 3 | 1,03 | 0,24 |
| 1 | 1 | 1 | 0 |
| 1 | 3 | 1,15 | 0,45 |
| 1 | 5 | 1,12 | 0,55 |
| 1 | 3 | 1,15 | 0,45 |
| 1 | 2 | 1,03 | 0,18 |
| 1 | 3 | 1,12 | 0,47 |
| 1 | 4 | 1,1 | 0,46 |
| 1 | 2 | 1,02 | 0,15 |
| 1 | 2 | 1,04 | 0,21 |
| 1 | 2 | 1,06 | 0,24 |
| 5 | 11 | 4,66 | 1,57 |
| 6 | 10 | 4,92 | 1,53 |
| 6 | 12 | 5,26 | 1,88 |
| 5 | 9 | 4,1 | 1,21 |
| 5 | 9 | 4,36 | 1,26 |
| 4 | 11 | 4,14 | 1,37 |
| 5 | 7 | 4,08 | 0,92 |
| 5 | 7 | 4,35 | 1,07 |
| 5 | 11 | 4,48 | 1,48 |
| 1 | 3 | 1,14 | 0,46 |
| 1 | 4 | 1,13 | 0,48 |
| 1 | 5 | 1,22 | 0,66 |
| 1 | 2 | 1,01 | 0,11 |
| 1 | 2 | 1,01 | 0,12 |
| 1 | 2 | 1,01 | 0,12 |
| 1 | 1 | 1 | 0 |
| 1 | 2 | 1,01 | 0,12 |

| | | | |
|---|---|---|---|
| 1 | 1 | 1 | 0 |
| 1 | 5 | 1,14 | 0,61 |
| 1 | 2 | 1,02 | 0,15 |
| 1 | 5 | 1,05 | 0,43 |
| 1 | 2 | 1,01 | 0,11 |
| 1 | 4 | 1,11 | 0,46 |
| 1 | 2 | 1,01 | 0,12 |
| 1 | 2 | 1,01 | 0,11 |
| 1 | 2 | 1,03 | 0,17 |
| 1 | 6 | 1,1 | 0,62 |
| 2 | 4 | 1,54 | 0,72 |
| 2 | 6 | 1,68 | 0,89 |
| 2 | 6 | 1,77 | 1,11 |
| 1 | 6 | 1,32 | 0,74 |
| 2 | 3 | 1,33 | 0,53 |
| 1 | 6 | 1,37 | 0,89 |
| 2 | 3 | 1,4 | 0,56 |
| 2 | 3 | 1,45 | 0,58 |
| 2 | 5 | 1,65 | 0,84 |
| 1 | 3 | 0,84 | 0,76 |
| 2 | 3 | 1,09 | 0,75 |
| 2 | 4 | 1,22 | 0,95 |
| 1 | 5 | 0,76 | 0,86 |
| 1 | 3 | 0,9 | 0,89 |
| 1 | 3 | 0,74 | 0,77 |
| 1 | 2 | 0,67 | 0,6 |
| 1 | 3 | 0,86 | 0,81 |
| 1 | 2 | 0,72 | 0,7 |
