## Supplemental table S16. Comparison of stool consistency and Gastrointestinal total score. for "A YOUNG CHILD FORMULA SUPPLEMENTED WITH L. REUTERI AND GALACTO-OLIGOSACCHARIDES MODULATES THE COMPOSITION AND FUNCTION OF THE GUT MICROBIOME SUPPORTING BONE AND MUSCLE DEVELOPMENT IN TODDLERS"

| | Treatment | Visit | Estimate | CI95% | p-value |
|---|---|---|---|---|---|
| Stool consistency average | (EYCF/CM-1)% | V2 | -8% | [-12%; -4%] | < 0.001 |
| | (EYCF/CM-1)% | V3 | -4% | [-8%; 0%] | 0,079 |
| Gastrointestinal total score | (EYCF/CM-1)% | V2 | -3% | [-8%; 2%] | 0,236 |
| | (EYCF/CM-1)% | V3 | 0% | [-5%; 5%] | 1 |
